# Effect of vaccination on time till Long COVID, a comparison of two ways to model effect of vaccination and two outcome definitions

**DOI:** 10.1101/2023.04.28.23289271

**Authors:** Peter D Smits, Patricia J Rodriguez, Samuel Gratzl, Brianna M Goodwin Cartwright, Sarah Gilson, Ryan Lee, Charlotte Baker, Nicholas L Stucky

## Abstract

Long COVID, or post-COVID syndrome, is a constellation of symptoms observed in patients at least four weeks after COVID-19 infection. We analyzed the effect of COVID-19 vaccination status on risk of either developing Long COVID symptoms or being diagnosed with Long COVID. In separate analyses we compared the effect of vaccination status at time of COVID-19 infection and the effect of vaccination status as a time-dependent covariate where vaccination could occur at any point with respect to COVID-19 infection.

To address this question, we identified a subset of adult patients from Truveta Data who experienced a COVID-19 infection as indicated by a positive laboratory test between 2021-10-01 and 2022-11-31. We considered two distinct ways of modeling the effect of vaccination status (time-independent and time-dependent) and two distinct outcomes of interest (Long COVID symptoms or diagnosis with Long COVID), representing four distinct analyses. The presence of Long COVID symptoms was defined as the presence of one or more new symptoms consistent with COVID-19/Long COVID at least four weeks post COVID-19 infection. Diagnosis of Long COVID was determined by the presence of one or more ICD-10-CM or SNOMED-CT codes explicitly identifying a patient as having been diagnosed with Long COVID.

Our analysis focusing on the effect of COVID-19 vaccination status at time of COVID-19 infection found that patients who had completed a primary COVID-19 vaccination sequence or had completed a primary vaccination sequence and received a booster dose at time of COVID-19 infection were on average at lower risk of either developing Long COVID symptoms or being diagnosed with Long COVID than unvaccinated patients (vaccinated versus unvaccinated HR of symptoms 0.9 [0.87-0.94], HR of diagnosis 0.86 [0.74-0.99]; vaccinated and boosted versus unvaccinated HR of symptoms 0.87 [0.83-0.91], HR of diagnosis 0.81 [0.69-0.95]). We do not find evidence that having received a booster dose in addition to having completed a primary vaccination sequence offers additional protection over having completed the primary sequence alone (vaccinated and boosted versus vaccinated HR of symptoms 0.96 [0.91-1.01], HR of diagnosis 0.94-1.13).

Our analysis of COVID-19 vaccination status modeled as a time-dependent covariate yielded similar results for patients who had completed a primary COVID-19 vaccination sequence or had completed a primary vaccination sequence and a booster dose. Both groups were on average at lower risk of developing Long COVID symptoms or being diagnosed with Long COVID than patients who where never vaccinated (vaccinated versus unvaccinated HR of symptoms 0.91 [0.88-0.95], HR of diagnosis 0.86 [0.75-0.99]; vaccinated and boosted versus unvaccinated HR of symptoms 0.88 [0.85-0.91], HR of diagnosis 0.77 [0.67-0.9]). As with the time-independent analysis, we also find that having completed a booster dose in addition to a primary COVID-19 vaccination sequence does not provide additional protection from developing Long COVID symptoms or being diagnosed with Long COVID over having completed the primary sequence alone (vaccinated and boosted versus vaccinated HR of symptoms 0.96 [0.92-1.01], HR of diagnosis 0.89 [0.76-1.06]).

We find that completing a primary vaccination sequence is associated with a decreased risk of developing Long COVID symptoms or being diagnosed with Long COVID compared with no vaccination regardless of whether vaccination status is modeled as a time-independent or time-dependent covariate. We find a similar protective effect in patients who have completed a primary vaccination sequence and a booster dose when compared to the those who are unvaccinated. However, we do not find evidence for a difference in protective effect between patients who have completed a primary vaccination sequence and a booster dose and those patients who have only completed a primary vaccination sequence.

Our results support the growing evidence that having complete a primary vaccination sequence is protective against the development of Long COVID symptoms or the diagnosis of Long COVID.

## 1 Introduction

Long COVID is a constellation of symptoms that otherwise resist a clear understanding or diagnosis [1], and is a condition which is primarily a diagnosis of exclusion [2]. As of 2021-10-01, the World Health Organization (WHO) provided a definition of Long COVID and the associated constellation of symptoms [2]. At the same time, ICD-10-CM codes (U09.9) and SNOMED-CT codes (1119303003, 1119304009) were introduced in order to better classify which patients have been diagnosed with Long COVID versus having symptoms consistent with Long COVID [3–5].

In practice, however, Long COVID is defined in many ways and not necessarily in a consistent manner[6]. One common difference in studies of Long COVID is that some studies analyze time from infection until the abatement of COVID-19 symptoms while others focus on the time to development of new Long COVID symptoms following a COVID-19 infection [7–9]. Another common variation among studies of Long COVID is a difference in observation window, which varies between 4 weeks to 1 year following COVID-19 infection, with further variation on whether the study period includes or starts after the recommended 28 days post-COVID-19 infection wait period before identifying a patient as having Long COVID [7–10].

Studies of the effects of COVID-19 vaccination status on development of Long COVID or persistence of COVID-19 symptoms have primarily focused on two comparisons: 1) difference in outcome rate or probability between people who were unvaccinated at time of COVID-19 infection versus those people who were vaccinated (in some form) prior to having a COVID-19 infection [6, 10–27] and 2) difference in outcome rate or probability between people who were unvaccinated at time of COVID-19 infection versus those people who were vaccinated after having a COVID-19 infection [28–38]. Though see Simon et al. [39] for an example of an analysis of the effect of vaccination on the development of Long COVID which considers vaccination before and after COVID-19 infection. The majority of studies and meta-analyses of the effects of vaccination prior to COVID-19 infection on risk of Long COVID have found that vaccination is protective against development of Long COVID symptoms and the persistence of COVID-19 symptoms [1, 6–9, 11, 27, 30]. In contrast, studies and subsequent meta-analyses of the effect of vaccination after COVID-19 on risk of Long COVID are fewer in number. Additionally, there is great heterogeneity in results, with the effect of vaccination after COVID-19 on risk of Long COVID having been show to either decrease, increase, or have no effect on the incidence or likelihood of Long COVID [7–9].

Here we estimate the effect of completing a primary COVID-19 vaccination sequence and the effect of a COVID-19 booster dose on either the risk of developing Long COVID symptoms or being diagnosed with Long COVID as separate analyses. We consider two ways of analyzing vaccination state: time-independent where we only considered completion of primary vaccination sequence and booster doses received prior to COVID-19 infection, and completion of primary vaccination sequence and booster doses as time-dependent covariates where these events can have occurred at any point relative to when a patient experienced a COVID-19 infection.

## 2 Methods

### Study Setting

The study population included a subset of Truveta Data focusing on patients who had a positive COVID-19 PCR test between 2021-10-01 and 2022-11-31 [40]. We used Truveta Studio to access the de-identified medical records used in this study on 2023-04-05. Truveta is a consortium of healthcare systems which have combined their electronic health record (EHR) data to enable medical research. Currently this consortium includes 28 members who provide patient care in over 20,000 clinics and 700 hospitals across 43 states. Updated data is provided daily to Truveta. Similar data fields across systems are mapped though syntactic normalization to a common schema referred to as the Truveta Data Model (TDM). Once organized into common fields, values are then semantically normalized to common ontologies such as ICD-10-CM, SNOMED-CT, LOINC, RxNorm, CVX, etc. These normalization procedures employ an expert-led, artificial intelligence driven process to accomplish high-confidence modeling at scale. The data are then de-identified by expert determination under the HIPAA Privacy Rule. Once de-identified, the data are then made available for analysis using Truveta Studio.

### 2.1 Population

Patients were included if they had their first positive COVID-19 laboratory test indicating infection between 2021-10-01 through 2022-11-30. The start of this study period was chosen because it represents when the ICD-10-CM code for a Long COVID diagnosis were created and put into effect [41].

Patients were excluded from analysis if they have had evidence of more than one COVID-19 infection in their medical history. We also excluded any patients who were younger than 18 years old at time of COVID-19 infection. Additionally, we also excluded patients who were only partially vaccinated with an mRNA vaccine sequence and never completed their primary sequence (see 2.1.1). Finally, patients were excluded for various data hygiene reasons (diagnosed with Long COVID before testing positive for COVID-19, having a booster dose before completing a primary vaccination sequence, or an impossibly long time between vaccination and COVID-19 infection (e.g., being vaccinated more than 2.5 years prior to COVID-19 infection, or being diagnosed with COVID-19 more than 3 years before getting vaccinated).

Time zero (T0) for this study was the time of positive COVID-19 laboratory test plus 28 days. This 28 day period was implemented as Long COVID is only diagnosable 28 days after COVID-19 infection [7–10]. If we did not implement this 28 day wait, then we would be artificially and uniformly increasing the time to event.

We considered two treatments, or exposures, in our study: completion of primary COVID-19 vaccination sequence, and a booster COVID-19 vaccine dose (see 2.1.1). Below we present two ways of analyze the effect of either of these exposures: a time-independent approach where only vaccination events prior to COVID-19 infection are considered, and a time-dependent approach where vaccination events were allowed to have occurred at any point relative to that patient’s COVID-19 infection.

#### 2.1.1 Vaccination sequence logic

Vaccination sequence algorithm is based on the COVID-19 vaccination sequence recommendations provided by the CDC [42] with the modification that we allowed for discordance in the primary sequence (mixture of either a Moderna, Pfizer, or Novavax vaccine doses).

If patients’ first dose was an mRNA vaccine, a check is performed for a second dose of mRNA vaccine within 3 to 8 weeks of the first dose. If a person has a second mRNA dose, but it does not fall within this 3 to 8 week window, this person is excluded from our analysis. While Pfizer and Moderna have different wait periods between doses, this window encompasses both definitions and allows for discordance between dose manufacturers. If the patients first dose was a traditional vaccine (i.e. Janssen), then only that single dose is required to have completed the doses for the primary sequence.

A patient’s time of having completed their primary vaccination sequence is two weeks after either their second mRNA dose or two weeks after their first traditional vaccine dose.

For a list of all CVX codes used to identify a vaccination, please see the Supplemental Material.

### 2.2 Outcomes

We considered two different definitions of Long COVID as our event of interest. These responses were analyzed independently with their own models as they describe very different aspects of our patient population.

Our first definition of Long COVID is based on the definition provided by the CDC where a constellation of symptoms are used as diagnostic criteria. We modified this definition slightly to only consider patients who tested positive for COVID-19 based on PCR test results.

Patients with this phenotype have or had long COVID based on the CDC’s published guidelines. A patient who had a positive COVID-19 PCR test and a specific symptom four or more weeks after the COVID-19 infection with no diagnosis code for the specific sign or symptom in the year prior to the COVID-19 infection, excluding the week prior. Symptoms of Long COVID include abdominal pain, anosmia, anxiety and/or depression, arthralgia, dyspnea, chest pain, cognitive impairment, cough, diarrhea, fatigue, fever, headache, impaired daily function, insomnia, lightheadedness, menstrual cycle irregularities. mood changes, myalgia, pain, palpitations and tachycardia, paresthesia, post-exertional malaise, and rash. For a complete set of codes associated with Long COVID and its symptoms, please see the Supplemental Material.

For analyses of the presence of Long COVID symptoms as the outcome of interest, a patient’s outcome time was defined as the minimum of the following: time of first development of new Long COVID symptoms, last recorded encounter in the EHR after their positive COVID-19 test, or 365 days (if their last encounter in the EHR was greater than 365 days after T0). Time till event was expressed in weeks (continuous). If a person did not experience the development of Long COVID symptoms, then they were right-censored at their last recorded time as described above. We considered all censoring to be uninformative. In the event a patient has no EHR events after their positive test result, that patient’s outcome time was defined as the time of positive COVID-19 test plus 28 days (e.g., time zero) plus 5 *×* 10*^−^*^7^ weeks. The fractional amount of time is added to the outcome time of patients who’s last encounter in the EHR was their positive COVID-19 test because, by definition, patient’s cannot have an outcome time of 0 as *S*(*t* = 0) = 1 [43]. This fractional amount of time is the minimum amount of time which does not cause the R package survival to error when attempting to fit a Cox regression model, as values smaller than 5 *×* 10*^−^*^7^ are below the tolerance for detecting if outcome time is 0.

Our second definition of Long COVID only considered patients who were diagnosed with Long COVID as indicated by relevant ICD-10-CM or SNOMED-CT diagnostic codes being present as a diagnosis. See the Supplemental Material for a full list of the ICD-10-CM and SNOMED-CT diagnostic codes used to identify if a patient was diagnosed with Long COVID in this study.

For the analysis of time till diagnosis with Long COVID, a patient’s outcome time was defined as the minimum of the following: time of Long COVID diagnosis, last recorded encounter in the EHR after their positive COVID-19 test, or 365 days (if their last encounter in the EHR was greater than 365 days after T0). Time till event was expressed in weeks. In the event a patient has no EHR events after their positive test result, that patient’s outcome time was defined as the time of positive COVID-19 test plus 28 days (e.g., time zero) plus 5 *×* 10*^−^*^7^ weeks. The logic for these decisions was identical as our choices described above for our analysis of time till development of Long COVID symptoms.

We performed sensitivity analyses to determine if excluding those patients with effectively zero follow-up time had a meaningful impact on our results. Please see the Supplemental Material for those results.

### 2.3 Comorbidities, demographics, and descriptive covariates

In addition to vaccination and booster status, we considered the following conditions as potential confounding features: anxiety, cardiovascular disease, cancer, cerebrovascular disease or stroke or transient ischemic attack (as one condition), chronic kidney disease, chronic obstructive pulmonary disease, dementia, depression, diabetes, immunocompromised, and peripheral artery disease. A patient was considered to have a comorbidity if an associated diagnostic code was present in their record within the two years prior to T0. The definitions of anxiety and depression used for our comorbidities are more expansive than our definition of anxiety or depression used as symptoms of Long COVID. Please see the Supplementary Material for complete list of all diagnostic codes used to identify these comorbidities based on patient information.

We also considered the following demographic and descriptive features as potential confounding features: sex (Female, Male, Unknown), race (American Indian or Alaska Native, Asian, Black or African American, Native Hawaiian or Other Pacific Islander, White, and Other), ethnicity (Not Hispanic or Latino, Hispanic or Latino, and Unknown), age in years at time of COVID-19 infection, year-month of COVID-19 infection, smoking status, one or more influenza vaccines within the two years prior to T0, number of inpatient encounters within the two years prior to T0, and number of outpatient encounters within the two years prior to T0.

Year-month was included as a covariate to reflect the COVID-19 “environment” (e.g., variant, infected population size, etc.) experienced by the patient at the time of their infection. The presence of a previous influenza vaccine, along with number of inpatient and outpatient encounters, were considered proxies for a patient’s likelihood to request or receive care as well as their overall health.

For a full list of diagnostic concept codes used to define the comorbidities and smoking status of our patients see the Supplemental Material.

### 2.4 Model of time from 28 days after COVID-19 infection till Long COVID

We transformed some of the covariates prior to fitting our Cox regression models. The count of inpatient encounters within the last two years, and the count of outpatient encounters within the last two years were both square-root transformed prior to being included in the model. This transform stabilizes the variance of the covariate and attenuates the effect of large observations. Age in years at time of COVID-19 infection was modeled using a natural cubic spline with 5 degrees of freedom. All categorical covariates with more than 2 levels were transformed into multiple indicator variables, or one-hot encoded, with the most frequently occurring state being the “reference” category. Hazard ratios comparing different combinations of COVID-19 vaccinated or boosted states to unvaccinated or vaccinated states were calculated using the `emmeans` R package [44].

#### 2.4.1 Time-independent treatment

Our first set of analyses of the effect of completing a primary COVID-19 vaccination sequence and receiving a booster dose versus being unvaccinated on time till Long COVID symptom development or diagnosis considered only patients who are either unvaccinated, have completed a primary vaccination sequence, or have completed a primary vaccination sequence and a booster dose prior to COVID-19 infection. Patients were excluded if they completed a primary vaccination sequence or received a booster dose after their COVID-19 infection (Section 2.2). This requirement is an additional exclusion criteria unique to the time-independent treatment analysis. This model was fit using the `survival` R package [45].

#### 2.4.2 Time-dependent treatment

In addition to the time-independent approach considered above, we also performed an analysis where we allowed patients to complete their primary vaccination sequence or receive a booster dose at any point relative to their COVID-19 infection. To allow for time dependent covariates, we modeled time from 28 days after COVID-19 infection till Long COVID using an extended Cox regression model in order to account for the time-dependence of the vaccination events, meaning a person could have completed their primary vaccination sequence or booster dose before or after T0. Unlike the time-independent analysis described above, there are no additional exclusion criteria as all patients can be included regardless of when they completed their primary COVID-19 vaccination sequence or received a booster dose. Note, as before we’re still excluding patients who were vaccinated during the 28 day waiting period between COVID-19 infection and this studies T0. These patients are excluded because they are not biologically directly comparable to patients who were vaccinated prior to a COVID-19 infection and the 28 day waiting period means we cannot distinguish, statistically, between vaccinated before or vaccinated within 28 days.

This model was fit using the `survival` R package [45]. See Terry M. Therneau and Patricia M. Grambsch [43] for further explanation of how time-dependent covariates are modeled in an extended Cox model.

All analyses were done using the R programming language [46] with a particular emphasis on the following packages: `survival` [45], `emmeans` [44], `broom` [47], `survminer` [48], `splines` [46], `dplyr` [49], `lubridate` [50], `rlang` [51], tidyr [52], arrow [53], table1 [54], and `xtable` [55].

## 3 Results

### 3.1 Population

438,431 patients tested positive for COVID-19 between 2021-10-01 and 2022-11-30 and met our inclusion and exclusion criteria (Table 1). Of the patients in our study, 95,228 completed a COVID-19 primary vaccination sequence at any point, 49,204 received a booster dose at any point, and 343,203 were never vaccinated. 93,505 were vaccinated prior to testing positive for COVID-19, and 1,723 were later vaccinated after testing positive for COVID-19. Similarly, of those patients who received a booster vaccine dose, 39,911 received that booster prior to testing positive for COVID-19, and 9,293 later received that booster after testing positive for COVID-19.

**Table 1:** Overall summary statistics of our analyzed population of patients who experienced a COVID-19 infection, stratified by vaccination status at time of COVID-19 infection.

| Feature | Unvaccinated<br>(N=344,926) | Vaccinated<br>(N=53,594) | Vaccinated and boosted<br>(N=39,911) | Overall<br>(N=438,431) |
| --- | --- | --- | --- | --- |
| Sex |  |  |  |  |
| Female | 196,659 (57.0%) | 32,618 (60.9%) | 23,121 (57.9%) | 252,398 (57.6%) |
| Male | 147,442 (42.7%) | 20,894 (39.0%) | 16,713 (41.9%) | 185,049 (42.2%) |
| Unknown | 825 (0.2%) | 82 (0.2%) | 77 (0.2%) | 984 (0.2%) |
| Age (y) |  |  |  |  |
| Mean (SD) | 49.4 (19.9) | 50.8 (19.3) | 61.1 (18.5) | 50.6 (20.0) |
| Median [Min, Max] | 47.5 [17.9, 98.9] | 49.7 [17.9, 98.8] | 63.5 [17.9, 98.9] | 49.4 [17.9, 98.9] |
| Year-month of COVID infection |  |  |  |  |
| 2021_10 | 20,051 (5.8%) | 3,364 (6.3%) | 138 (0.3%) | 23,553 (5.4%) |
| 2021_11 | 15,967 (4.6%) | 2,655 (5.0%) | 272 (0.7%) | 18,894 (4.3%) |
| 2021_12 | 51,835 (15.0%) | 6,503 (12.1%) | 1,562 (3.9%) | 59,900 (13.7%) |
| 2022_01 | 110,238 (32.0%) | 20,514 (38.3%) | 8,453 (21.2%) | 139,205 (31.8%) |
| 2022_02 | 19,200 (5.6%) | 3,092 (5.8%) | 2,138 (5.4%) | 24,430 (5.6%) |
| 2022_03 | 5,436 (1.6%) | 787 (1.5%) | 811 (2.0%) | 7,034 (1.6%) |
| 2022_04 | 7,520 (2.2%) | 916 (1.7%) | 1,307 (3.3%) | 9,743 (2.2%) |
| 2022_05 | 16,879 (4.9%) | 2,507 (4.7%) | 3,568 (8.9%) | 22,954 (5.2%) |
| 2022_06 | 21,268 (6.2%) | 2,985 (5.6%) | 4,723 (11.8%) | 28,976 (6.6%) |
| 2022_07 | 28,102 (8.1%) | 3,631 (6.8%) | 5,466 (13.7%) | 37,199 (8.5%) |
| 2022_08 | 20,525 (6.0%) | 2,702 (5.0%) | 4,264 (10.7%) | 27,491 (6.3%) |
| 2022_09 | 11,920 (3.5%) | 1,419 (2.6%) | 2,670 (6.7%) | 16,009 (3.7%) |
| 2022_10 | 7,620 (2.2%) | 1,075 (2.0%) | 2,053 (5.1%) | 10,748 (2.5%) |
| 2022_11 | 8,365 (2.4%) | 1,444 (2.7%) | 2,486 (6.2%) | 12,295 (2.8%) |
| Race |  |  |  |  |
| American Indian or Alaska Native | 2,050 (0.6%) | 633 (1.2%) | 413 (1.0%) | 3,096 (0.7%) |
| Asian | 7,552 (2.2%) | 2,111 (3.9%) | 2,389 (6.0%) | 12,052 (2.7%) |
| Black or African American | 60,578 (17.6%) | 3,030 (5.7%) | 1,443 (3.6%) | 65,051 (14.8%) |
| Native Hawaiian or Other Pacific Islander | 924 (0.3%) | 319 (0.6%) | 141 (0.4%) | 1,384 (0.3%) |
| Other | 58,676 (17.0%) | 9,373 (17.5%) | 5,758 (14.4%) | 73,807 (16.8%) |
| White | 215,146 (62.4%) | 38,128 (71.1%) | 29,767 (74.6%) | 283,041 (64.6%) |
| Ethnicity |  |  |  |  |
| Not Hispanic or Latino | 266,171 (77.2%) | 42,166 (78.7%) | 33,400 (83.7%) | 341,737 (77.9%) |
| Hispanic or Latino | 49,535 (14.4%) | 8,134 (15.2%) | 4,323 (10.8%) | 61,992 (14.1%) |
| Unknown | 29,220 (8.5%) | 3,294 (6.1%) | 2,188 (5.5%) | 34,702 (7.9%) |
| Anxiety |  |  |  |  |
| Yes | 52,122 (15.1%) | 11,563 (21.6%) | 8,769 (22.0%) | 72,454 (16.5%) |
| Continued on next page |  |  |  |  |

**Table1 – continued from previous page**
| Feature | Unvaccinated | Vaccinated | Vaccinated and boosted | Overall |
| --- | --- | --- | --- | --- |
| No | 292,804 (84.9%) | 42,031 (78.4%) | 31,142 (78.0%) | 365,977 (83.5%) |
| Cardiovascular Disease |  |  |  |  |
| Yes | 122,775 (35.6%) | 21,548 (40.2%) | 20,631 (51.7%) | 164,954 (37.6%) |
| No | 222,151 (64.4%) | 32,046 (59.8%) | 19,280 (48.3%) | 273,477 (62.4%) |
| Cancer |  |  |  |  |
| Yes | 16,673 (4.8%) | 3,776 (7.0%) | 5,299 (13.3%) | 25,748 (5.9%) |
| No | 328,253 (95.2%) | 49,818 (93.0%) | 34,612 (86.7%) | 412,683 (94.1%) |
| Cerebrovascular Disease/Stroke/TIA |  |  |  |  |
| Yes | 20,137 (5.8%) | 3,816 (7.1%) | 4,686 (11.7%) | 28,639 (6.5%) |
| No | 324,789 (94.2%) | 49,778 (92.9%) | 35,225 (88.3%) | 409,792 (93.5%) |
| CKD |  |  |  |  |
| Yes | 28,319 (8.2%) | 5,248 (9.8%) | 6,306 (15.8%) | 39,873 (9.1%) |
| No | 316,607 (91.8%) | 48,346 (90.2%) | 33,605 (84.2%) | 398,558 (90.9%) |
| COPD |  |  |  |  |
| Yes | 20,660 (6.0%) | 3,458 (6.5%) | 3,455 (8.7%) | 27,573 (6.3%) |
| No | 324,266 (94.0%) | 50,136 (93.5%) | 36,456 (91.3%) | 410,858 (93.7%) |
| Dementia |  |  |  |  |
| Yes | 13,828 (4.0%) | 2,266 (4.2%) | 2,641 (6.6%) | 18,735 (4.3%) |
| No | 331,098 (96.0%) | 51,328 (95.8%) | 37,270 (93.4%) | 419,696 (95.7%) |
| Depression |  |  |  |  |
| Yes | 76,451 (22.2%) | 18,873 (35.2%) | 15,396 (38.6%) | 110,720 (25.3%) |
| No | 268,475 (77.8%) | 34,721 (64.8%) | 24,515 (61.4%) | 327,711 (74.7%) |
| Diabetes |  |  |  |  |
| Yes | 48,932 (14.2%) | 8,918 (16.6%) | 8,855 (22.2%) | 66,705 (15.2%) |
| No | 295,994 (85.8%) | 44,676 (83.4%) | 31,056 (77.8%) | 371,726 (84.8%) |
| Immunocompromised |  |  |  |  |
| Yes | 74,243 (21.5%) | 13,919 (26.0%) | 14,514 (36.4%) | 102,676 (23.4%) |
| No | 270,683 (78.5%) | 39,675 (74.0%) | 25,397 (63.6%) | 335,755 (76.6%) |
| PAD |  |  |  |  |
| Yes | 11,133 (3.2%) | 2,691 (5.0%) | 3,379 (8.5%) | 17,203 (3.9%) |
| No | 333,793 (96.8%) | 50,903 (95.0%) | 36,532 (91.5%) | 421,228 (96.1%) |
| Smoking Status |  |  |  |  |
| Yes | 66,103 (19.2%) | 11,320 (21.1%) | 8,793 (22.0%) | 86,216 (19.7%) |
| No | 278,823 (80.8%) | 42,274 (78.9%) | 31,118 (78.0%) | 352,215 (80.3%) |
| 1+ influenza vaccines within 2 years prior |  |  |  |  |
| Yes | 24,399 (7.1%) | 20,358 (38.0%) | 22,465 (56.3%) | 67,222 (15.3%) |
| Continued on next page |  |  |  |  |

| Feature | Unvaccinated | Vaccinated | Vaccinated and boosted | Overall |
| --- | --- | --- | --- | --- |
| No | 320,527 (92.9%) | 33,236 (62.0%) | 17,446 (43.7%) | 371,209 (84.7%) |
| Number of inpatient encounters within last 2 years |  |  |  |  |
| Mean (SD) | 0.295 (2.43) | 0.568 (3.93) | 0.649 (3.80) | 0.360 (2.81) |
| Median [Min, Max] | 0 [0, 382] | 0 [0, 278] | 0 [0, 246] | 0 [0, 382] |
| Number of outpatient encounters within last 2 years |  |  |  |  |
| Mean (SD) | 15.5 (35.5) | 39.6 (67.4) | 48.8 (75.4) | 21.5 (46.9) |
| Median [Min, Max] | 2.00 [0, 1,200] | 16.0 [0, 1,400] | 23.0 [0, 1,730] | 4.00 [0, 1,730] |
| Number of unique blood panel labs within 2 years prior |  |  |  |  |
| Mean (SD) | 29.0 (110) | 43.0 (129) | 50.0 (136) | 32.7 (115) |
| Median [Min, Max] | 0 [0, 10,800] | 0 [0, 4,880] | 11.0 [0, 6,410] | 0 [0, 10,800] |
| Developed Long COVID symptoms |  |  |  |  |
| Yes | 46,108 (13.4%) | 8,334 (15.6%) | 5,197 (13.0%) | 59,639 (13.6%) |
| No | 298,818 (86.6%) | 45,260 (84.4%) | 34,714 (87.0%) | 378,792 (86.4%) |
| Diagnosed with Long COVID |  |  |  |  |
| Yes | 2,170 (0.6%) | 603 (1.1%) | 437 (1.1%) | 3,210 (0.7%) |
| No | 342,756 (99.4%) | 52,991 (98.9%) | 39,474 (98.9%) | 435,221 (99.3%) |

When we compare which patients had either of our outcomes of interest we observe that there is surprisingly little overlap between the population for which we observe either the presence of Long COVID symptoms or a diagnosis with Long COVID (Table 2).

**Table 2:**
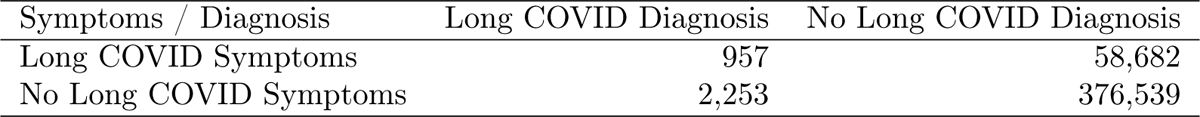
Contingecy table comparing overlap in differeing Long COVID outcomes among patients.

Of our 438,431 patients, 59,639 experienced Long COVID symptoms at least 28 days after their COVID-19 infection while 3,210 were given a diagnosis on Long COVID at least 28 days after their COVID-19 infection.

### 3.2 Risk of Long COVID associated with vaccination status

#### 3.2.1 Time-independent

We present here the results from our analysis of patients who were vaccinated, vaccinated and boosted, and unvaccinated prior to experiencing a COVID-19 infection, excluding any patients who either completed their primary COVID-19 vaccination sequence or received a booster dose after experiencing a COVID-19 infection. There were a total of 427,703 patients in this analysis.

First, we present our results for the analysis of time till development of Long COVID symptoms. Kaplan-Meier estimated survival curves demonstrate obvious differences in time till outcome between persons with the unvaccinated state compared those with the vaccinated or vaccinated and boosted states (Fig. 1). In contrast, there is little perceptible difference between the estimated survival curves for those patients who were vaccinated versus those patients who were vaccinated and boosted.

**Figure 1:**
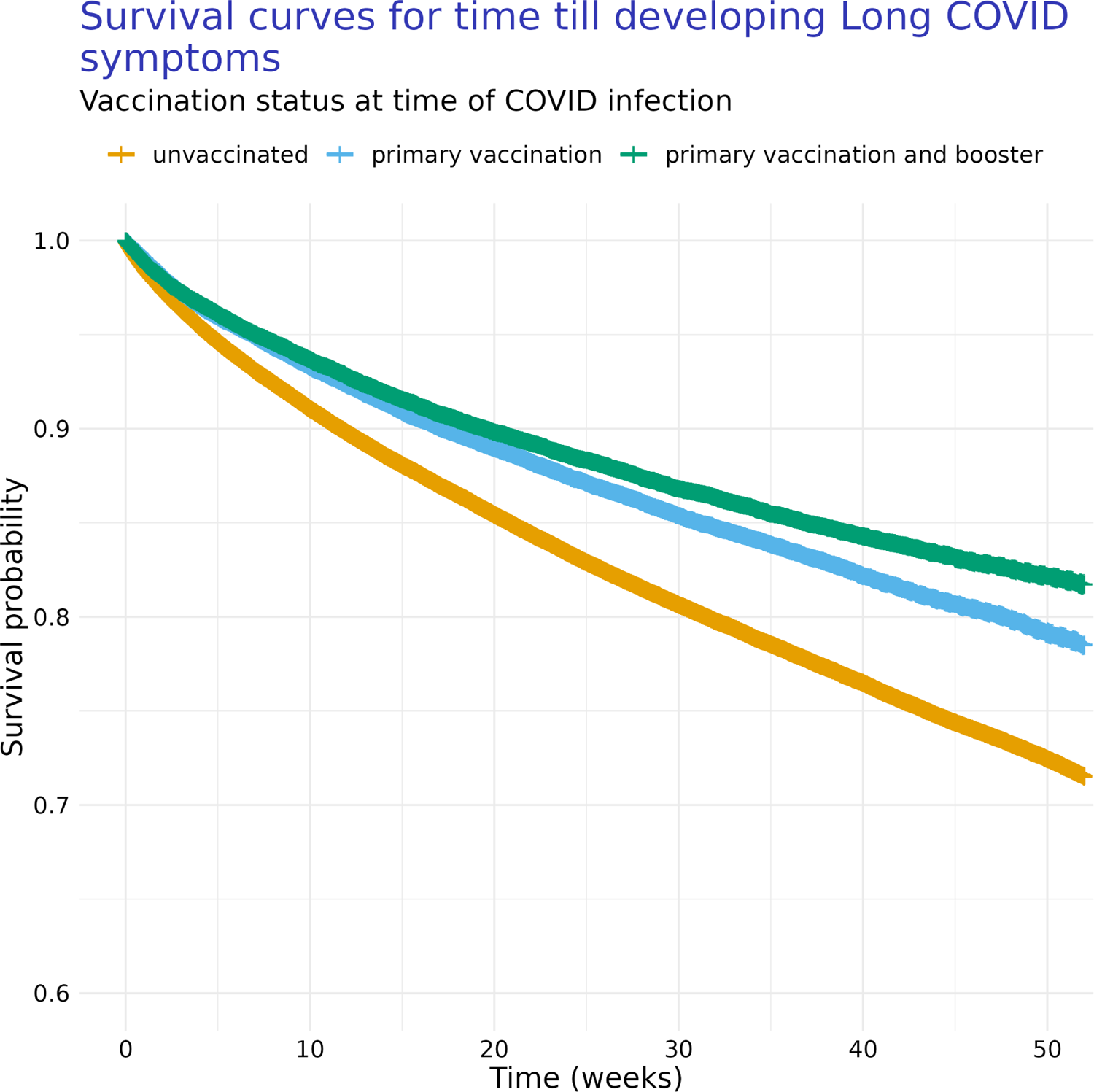
Kaplan-Meier curves of time till development of Long COVID symptoms for patients with different COVID-19 vaccination states (i.e., unvaccinated, completed primary vaccination sequence, and vaccinated plus having received a booster doses). For this analysis, we considered only vaccination state at time of COVID-19 infection.

The patterns from the Kaplan-Meier curves are consistent with hazard ratios estimated as part of our Cox regression model where time till event is further conditioned on multiple comorbidities and demographic features (Fig. 2, Table 3). We find that, when considering only those vaccination events prior to COVID-19 infection, patients who are vaccinated are at a lower risk of developing Long COVID symptoms than those who are unvaccinated (Fig.2, Table 3). Similarly, those who are vaccinated and boosted are at a lower risk of developing Long COVID symptoms than those who are unvaccinated. Finally, we find no evidence of a difference in risk of developing symptoms of Long COVID between patients who are vaccinated and boosted versus those patients who are vaccinated (Fig.2, Table 3).

**Figure 2:**
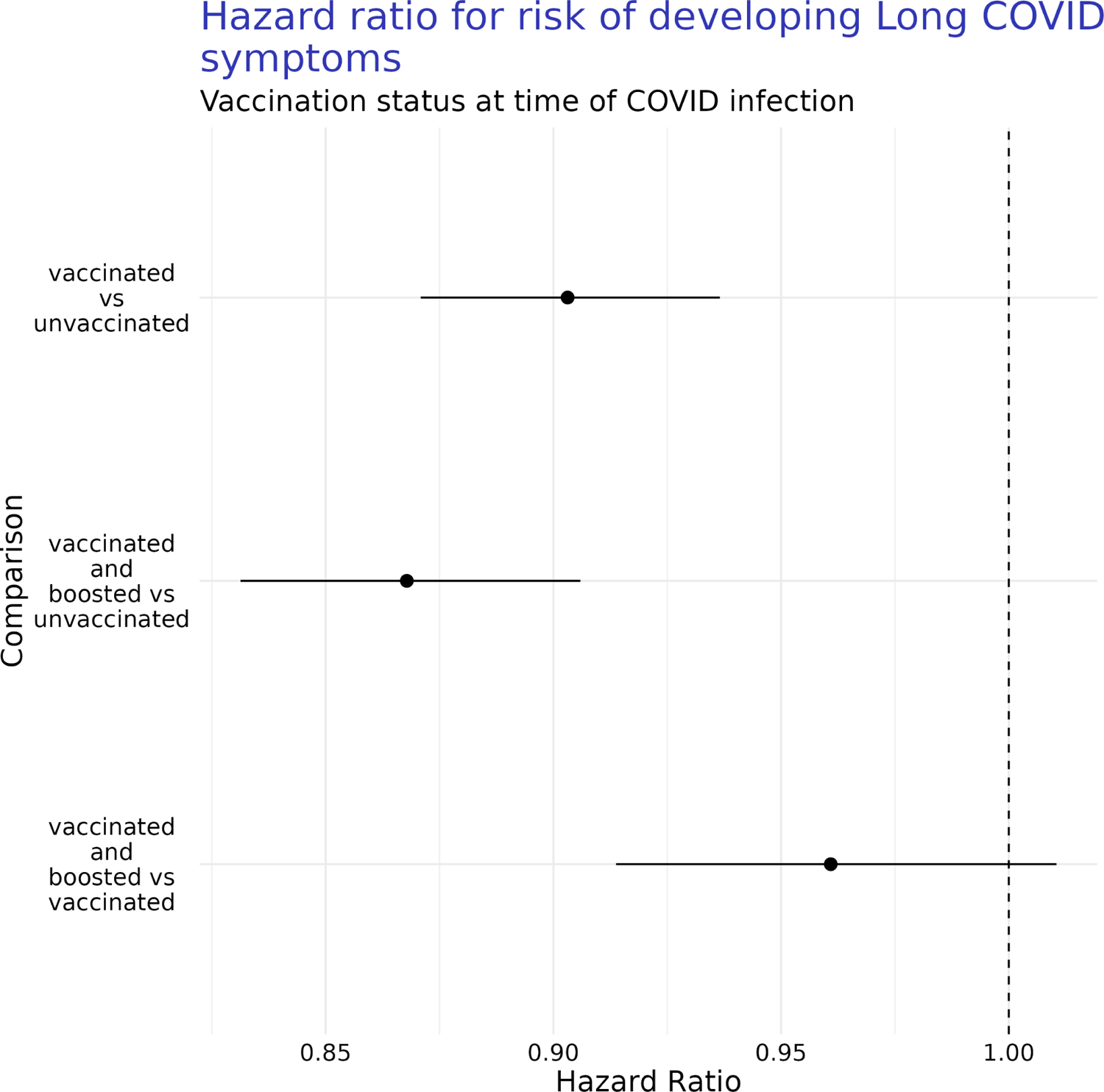
Graphical comparison of estimated hazard ratios for risk of developing Long COVID symptoms 28 days after COVID-19 infection depending on a person’s vaccination state at time of COVID-19 infection (i.e., unvaccinated, completed primary vaccination sequence, and vaccinated plus having received a booster doses). Estimates are presented with 95% confidence intervals.

**Table 3:** Estimated hazard ratios for risk of developing Long COVID symptoms based on vaccination status at time of COVID infection. Hazard ratios are presented with 95% confidence intervals.

| Comparison | Hazard Ratio [95% CI] |
| --- | --- |
| vaccinated vs unvaccinated | 0.9 [0.87, 0.94] |
| vaccinated and boosted vs unvaccinated | 0.87 [0.83, 0.91] |
| vaccinated and boosted vs vaccinated | 0.96 [0.91, 1.01] |

In contrast to the results with presence of Long COVID symptoms as our outcome of interest, when we consider a diagnosis of Long COVID as our response of interest, the estimated survival are nearly flat over time (Fig. 3), which is consistent with how rare this diagnosis is in our population (Tables 1, 2). Whatever difference in time till diagnosis that exists between the unvaccinated, vaccinated, and vaccinated and boosted populations are extremely small in terms of absolute effect.

**Figure 3:**
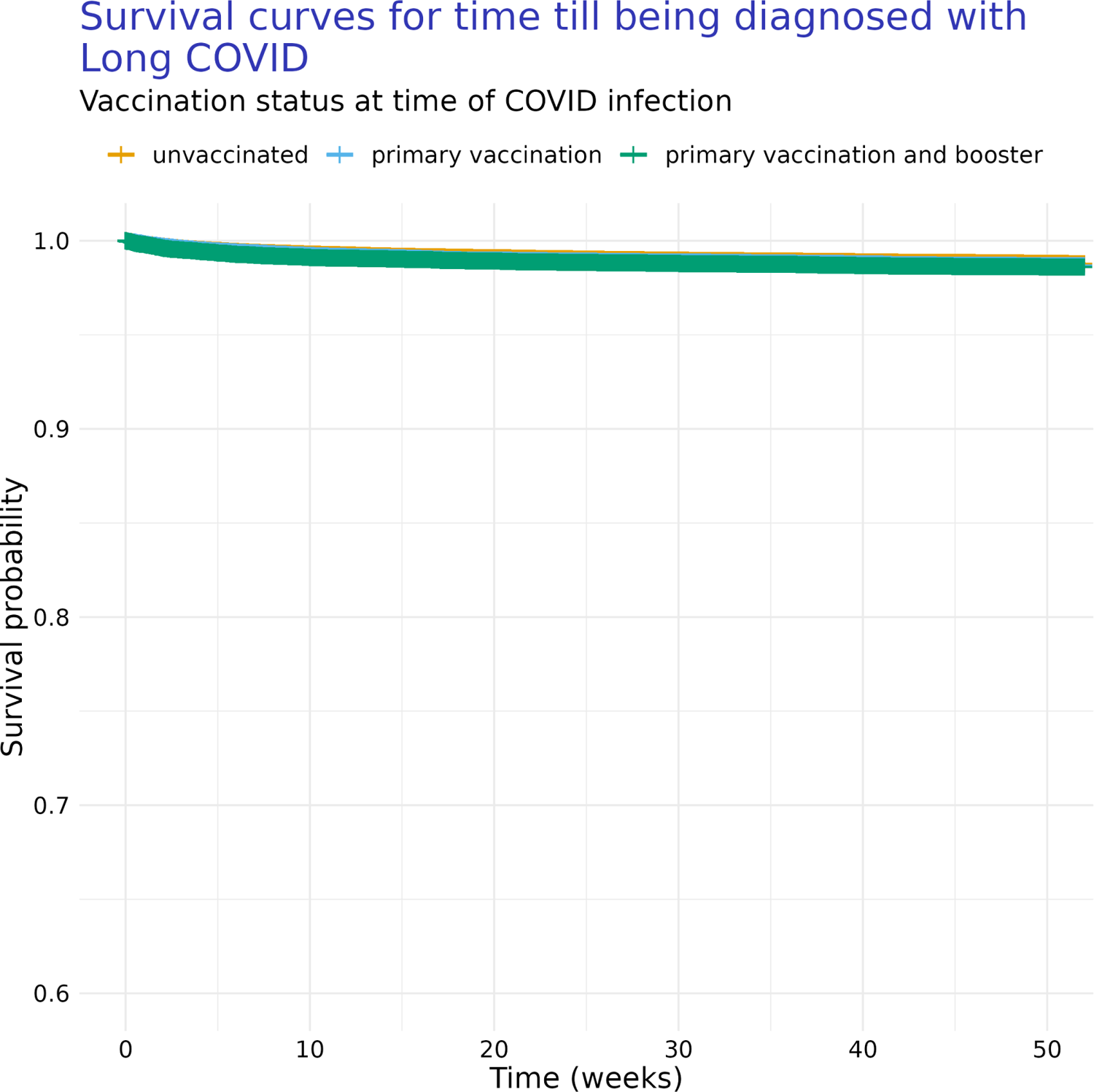
Kaplan-Meier curves of time till diagnosis with Long COVID for patients with different COVID-19 vaccination states (i.e., unvaccinated, completed primary vaccination sequence, and vaccinated plus having received a booster doses). For this analysis, we considered only vaccination state at time of COVID-19 infection, and all subsequent vaccination events are ignored.

The pattern from the Kaplan-Meier curves is effectively retained when time till development of Long COVID symptoms is further conditioned on the comorbidities and patient descriptive features described above, as evidenced by the hazard ratios estimated as part of our Cox regression model (Fig.4, Table 4).

**Figure 4:**
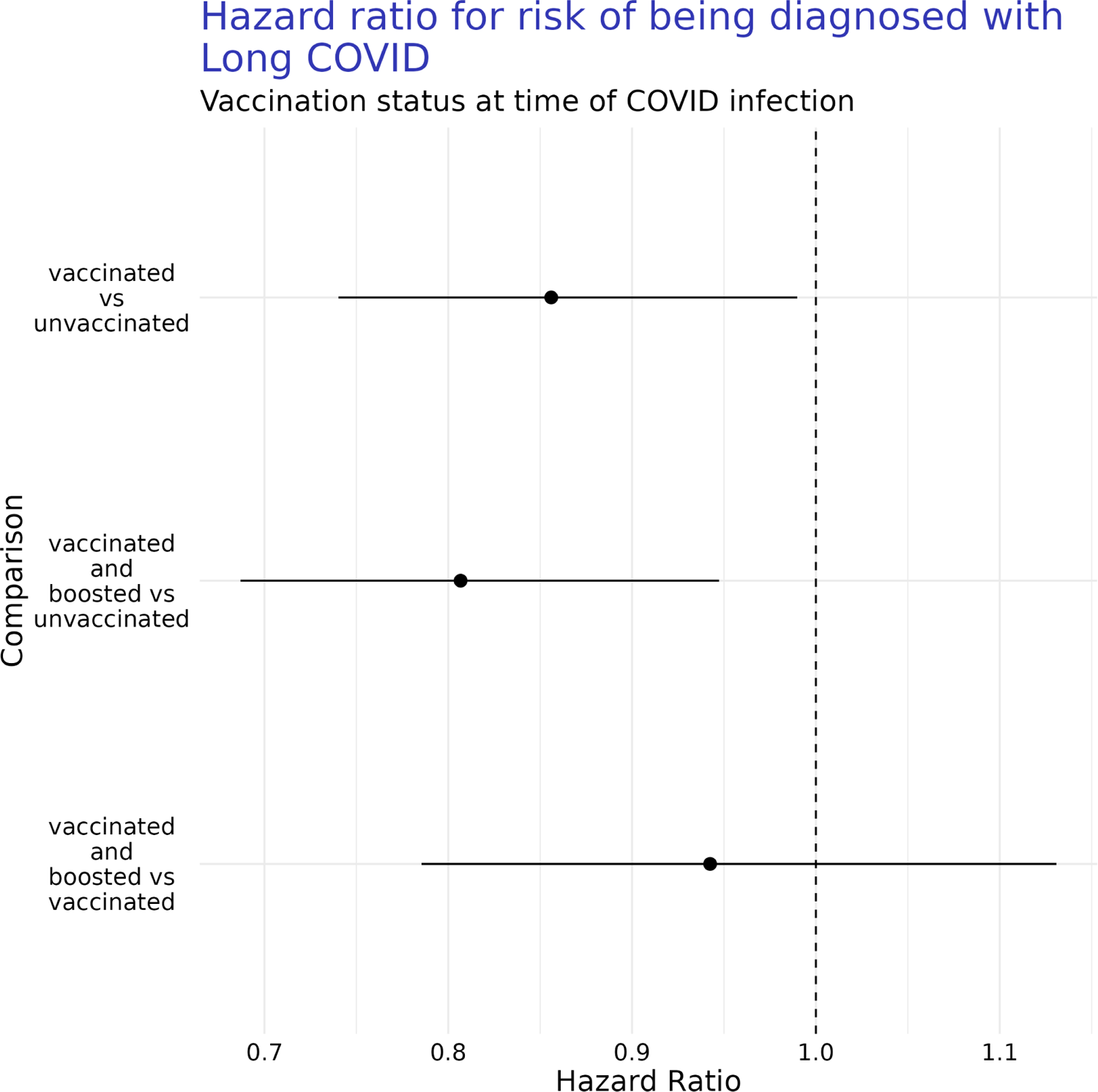
Comparison of estimated hazard ratios for risk of being diagnosed with Long COVID after COVID-19 infection depending on a person’s vaccination state at time of COVID-19 infection (i.e., unvaccinated, completed primary vaccination sequence, and vaccinated plus having received a booster doses). Estimates are presented with 95% confidence intervals. For this analysis, we considered only vaccination state at time of COVID-19 infection, and all subsequent vaccination events are ignored.

**Table 4:** Estimated hazard ratios for risk of being diagnosed with Long COVID based on vaccination status at time of COVID infection. Hazard ratios are presented with 95% confidence intervals.

| Comparison | Hazard Ratio [95% CI] |
| --- | --- |
| vaccinated vs unvaccinated | 0.86 [0.74, 0.99] |
| vaccinated and boosted vs unvaccinated | 0.81 [0.69, 0.95] |
| vaccinated and boosted vs vaccinated | 0.94 [0.79, 1.13] |

We find that, when considering only those vaccination events prior to COVID-19 infection, patients who are vaccinated are at a lower risk of being diagnosed with Long COVID than those who are unvaccinated (Fig. 4, Table 4). Similarly, we find that patients who are vaccinated and boosted have a lower risk of being diagnosed with Long COVID over time than the unvaccinated. Additionally, we do not find evidence of a difference in risk in receiving a Long COVID diagnosis between patients who are vaccinated and boosted versus those patients who were only vaccinated (Fig. 4, Table 4).

#### 3.2.2 Time-dependent

We present here the results from our analysis of patients who were vaccinated, vaccinated and boosted, and unvaccinated at any time relative to their COVID-19 infection, with vaccination events considered as time-dependent covariates.

We had a total of 438,431 people when we allowed vaccination and booster timing to vary with respect to COVID-19 infection, accounting for individuals who were completed a primary vaccination sequence or received a booster dose after their COVID-19 infection.

Estimated survival functions for the time from T0 till development of Long COVID symptoms with vaccination state treated as a time-dependent covariate (Fig. 5) have a similar pattern to the survival curves estimated from the time-independent analysis (Fig. 1). We see obvious differences in the time till outcome between persons with the unvaccinated state compared those with the vaccinated or vaccinated and boosted states (Fig. 5).

**Figure 5:**
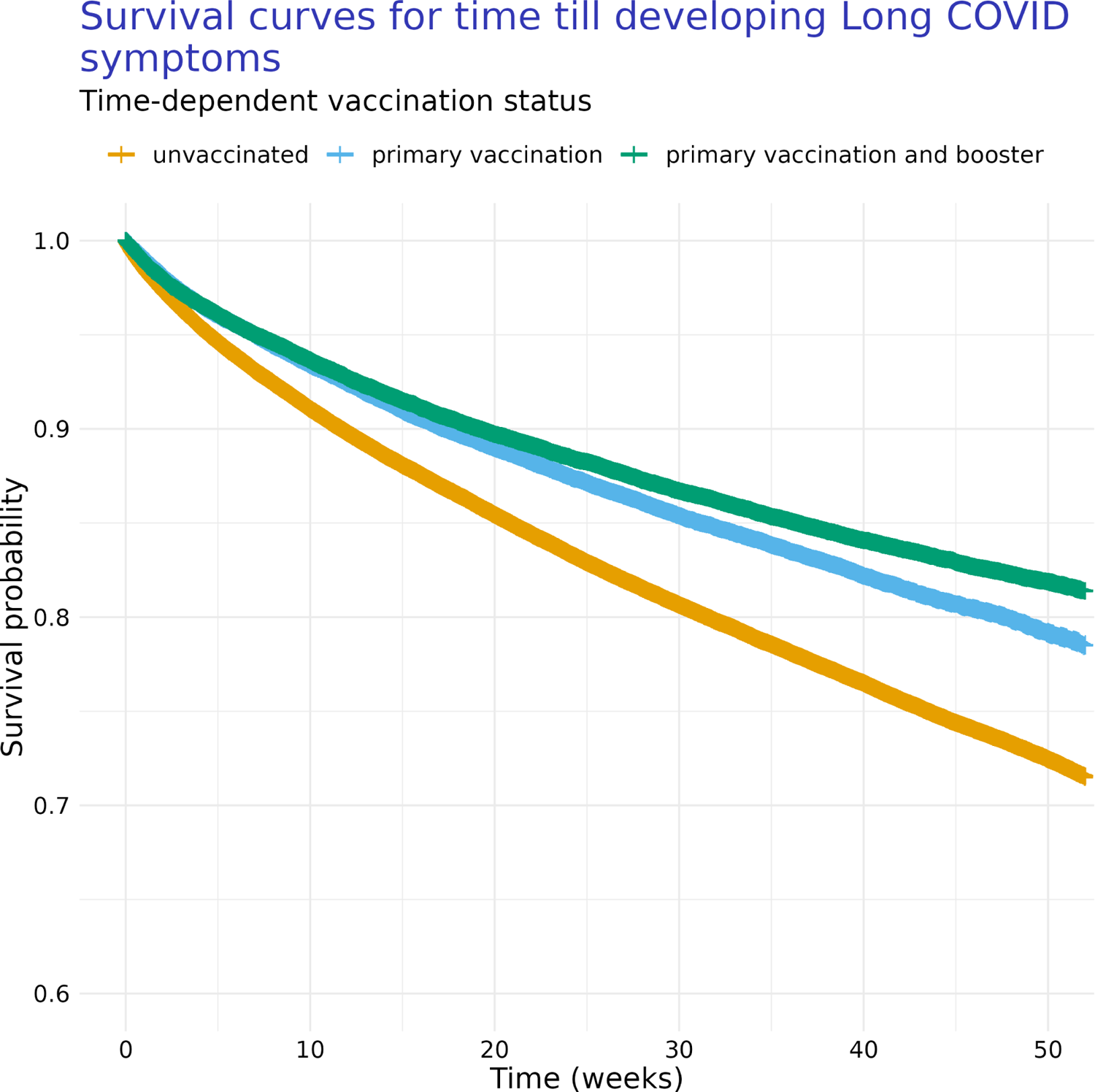
Kaplan-Meier curves of time till development of Long COVID symptoms depending on patient’s COVID-19 vaccination status (i.e., unvaccinated, completed primary vaccination sequence, and vaccinated plus having received a booster doses). For this analysis, we considered vaccination and booster states as time-dependent covariates, meaning they can happen at any point relative to T0.

The pattern from the Kaplan-Meier curves is effectively retained when time till development of Long COVID symptoms is further conditioned on the comorbidities and patient descriptive features described above, as evidenced by the hazard ratios estimated as part of our Cox regression model (Fig.6, Table 5). We find that patients who are vaccinated are at a lower risk of developing Long COVID symptoms than those who are unvaccinated (Fig. 6, Table 5). Similarly, those who are vaccinated and boosted are at a lower risk of developing Long COVID symptoms than those who are unvaccinated. Finally, we also find no evidence of a difference in risk of developing Long COVID symptoms between patients vaccinated and boosted versus those patients who are vaccinated (Fig. 6, Table 5).

**Figure 6:**
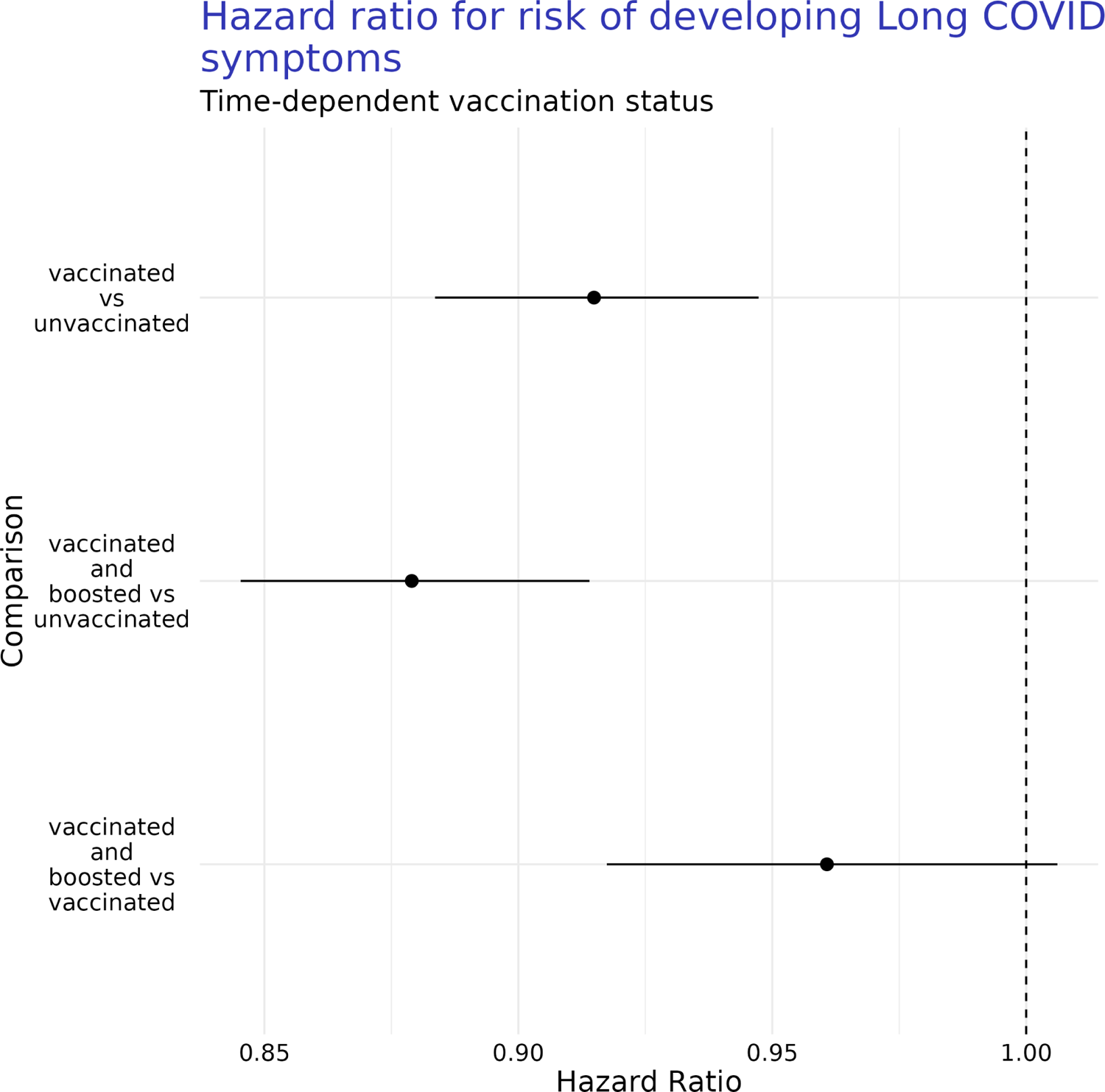
Comparison of estimated hazard ratios for risk of developing Long COVID symptoms after COVID-19 infection based on a person’s vaccination state at time of COVID-19 infection (i.e., unvaccinated, completed primary vaccination sequence, and vaccinated plus having received a booster doses). Estimates are presented with 95% confidence intervals. For this analysis, we considered vaccination and booster states as time-dependent covariates, meaning they can happen at any point relative to T0.

**Table 5:** Estimated hazard ratios for risk of developing Long COVID symptoms associated with vaccination status where vaccination status is modeled as time-dependent covariates. Hazard ratios are presented with 95% confidence intervals.

| Comparison | Hazard Ratio [95% CI] |
| --- | --- |
| vaccinated vs unvaccinated | 0.91 [0.88, 0.95] |
| vaccinated and boosted vs unvaccinated | 0.88 [0.85, 0.91] |
| vaccinated and boosted vs vaccinated | 0.96 [0.92, 1.01] |

In contrast, when we consider a diagnosis of Long COVID as our response of interest along with treating vaccination status as a time-dependent covariate, the estimated survival are nearly flat over time (Fig. 7), which is consistent with how rare this diagnosis is in our population (Table 1). Whatever difference exists between the unvaccinated, vaccinated, and vaccinated and boosted populations they are extremely small in absolute effect on time till diagnosis. These results are consistent with the previous analysis where only vaccination events prior to T0 were considered.

**Figure 7:**
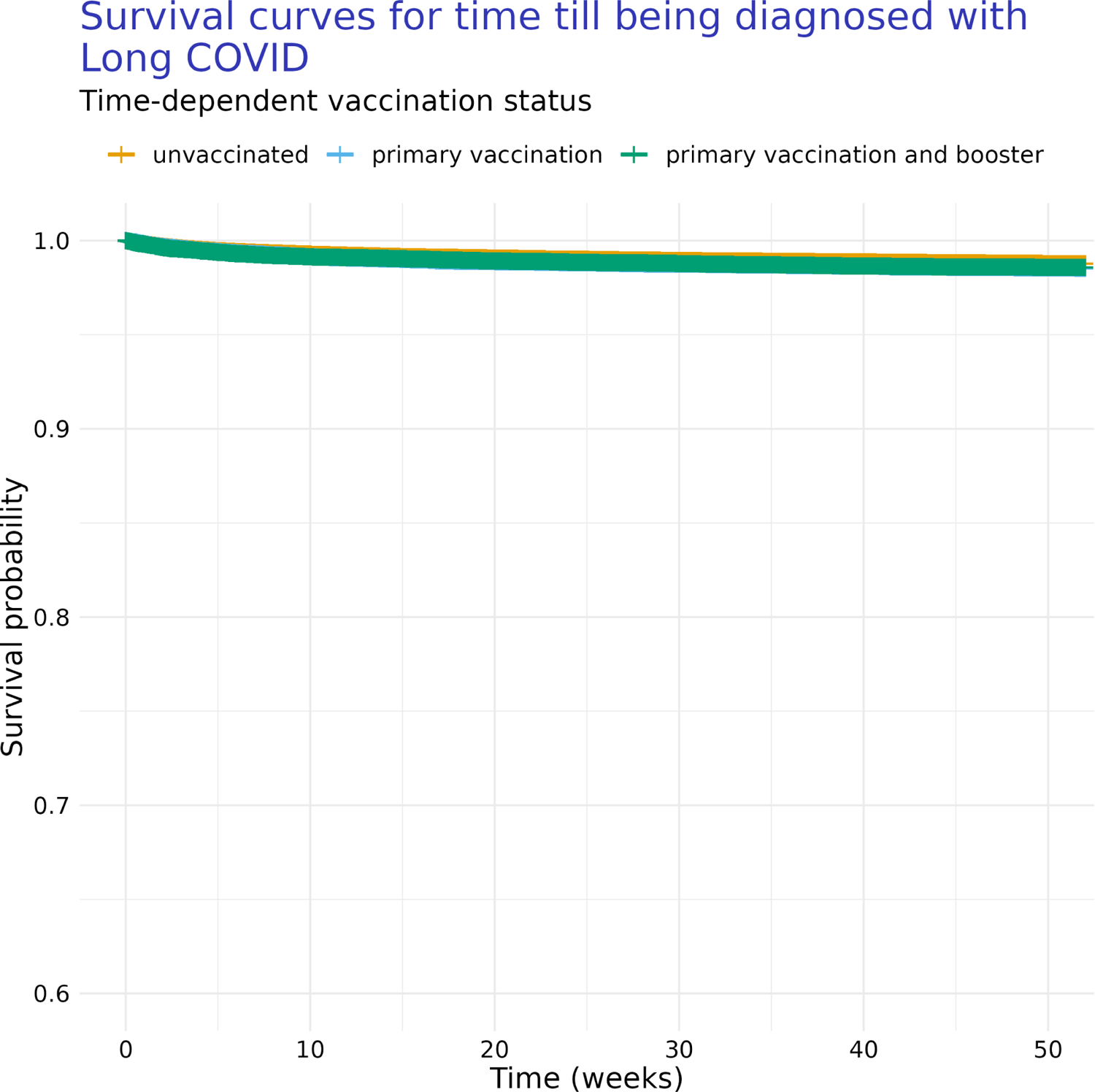
Kaplan-Meier curves for time till diagnosis with Long COVID depending on a patient’s COVID-19 vaccination states (i.e., unvaccinated, completed primary vaccination sequence, and vaccinated plus having received a booster doses). For this analysis, we considered vaccination and booster states as time-dependent covariates, meaning they can happen at any point relative to T0.

The pattern from the Kaplan-Meier curves is effectively retained when time till development of Long COVID symptoms is further conditioned on the comorbidities and patient descriptive features described above, as evidenced by the hazard ratios estimated as part of our Cox regression model (Fig.8, Table 6). These results are consistent with the previous analysis where only vaccination events prior to T0 were considered.

**Figure 8:**
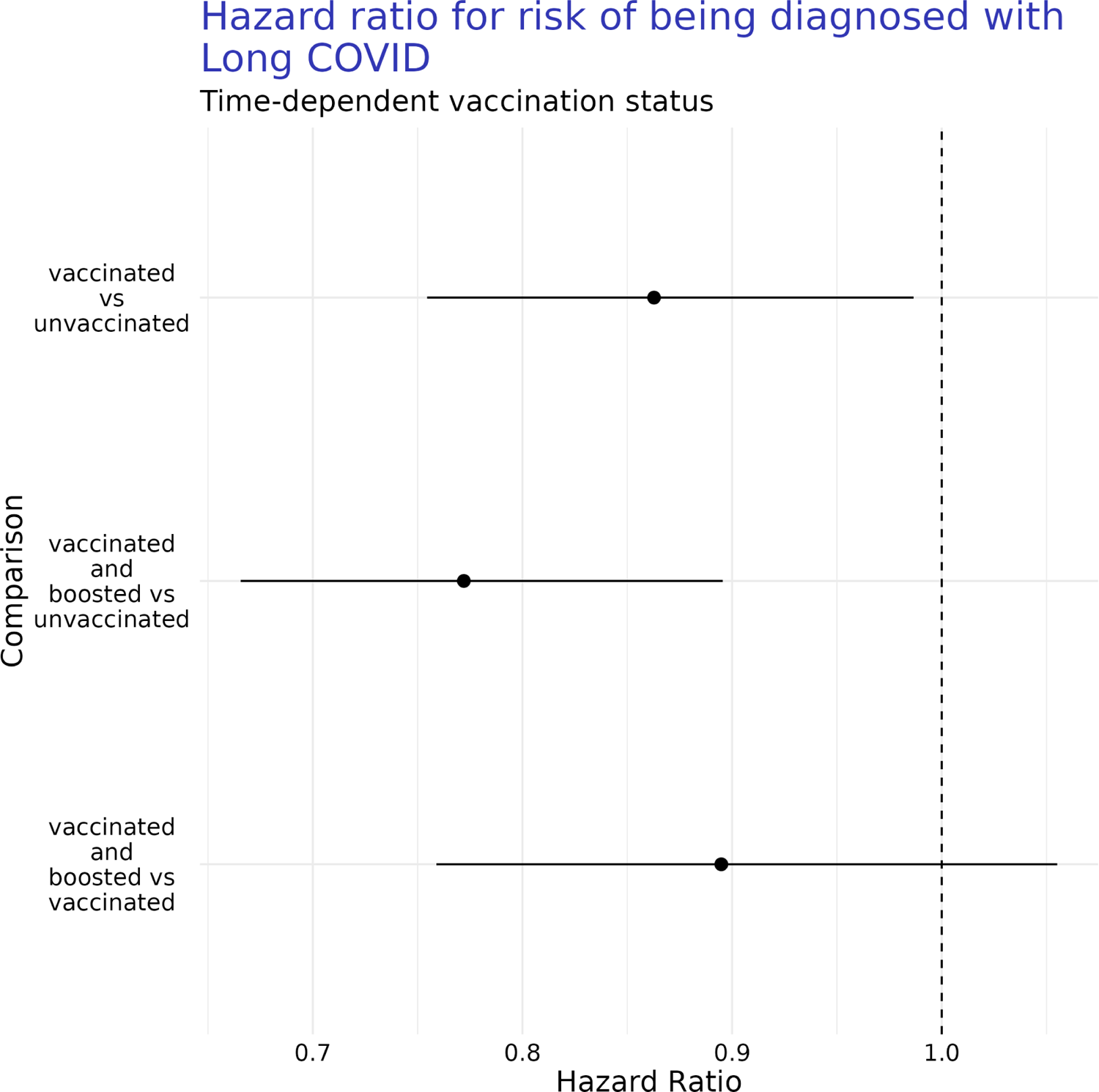
Comparison of estimated hazard ratios for risk of diagnosis with Long COVID symptoms after COVID-19 infection based on a person’s vaccination state at time of COVID-19 infection (i.e., unvaccinated, completed primary vaccination sequence, and vaccinated plus having received a booster doses). Estimates are presented with 95% confidence intervals. For this analysis, we considered vaccination and booster states as time-dependent covariates, meaning they can happen at any point relative to T0.

**Table 6:** Estimated hazard ratios for risk of being diagnosed with Long COVID associated with vaccination status where vaccination status is modeled as time-dependent covariates. Hazard ratios are presented with 95% confidence intervals.

| Comparison | Hazard Ratio [95% CI] |
| --- | --- |
| vaccinated vs unvaccinated | 0.86 [0.75, 0.99] |
| vaccinated and boosted vs unvaccinated | 0.77 [0.67, 0.9] |
| vaccinated and boosted vs vaccinated | 0.89 [0.76, 1.06] |

We find that, when vaccination and booster status are modeled as time-dependent covariates, patients who are vaccinated are at a lower risk of being diagnosed with Long COVID than those who are unvaccinated (Fig. 8). Similarly, we find that patients who are vaccinated and boosted have a lower risk of being diagnosed with Long COVID over time than the unvaccinated. Finally, we find no evidence of a difference in risk of developing Long COVID symptoms between patients vaccinated and boosted versus those patients who are vaccinated (Fig. 8, Table 6).

## 4 Discussion

Our analyses support the overall conclusion that completing a primary vaccination sequence is protective against the development or persistence of Long COVID defined either as the presence of Long COVID symptoms four weeks after COVID-19 infection or diagnosis with Long COVID in adults. These results are consistent with much of the established literature [1, 6–9, 11, 27, 30].

We also find that the effect of a booster dose of COVID-19 vaccination does not necessarily provide additional protective effect against the persistence or development of Long COVID symptoms four weeks after COVID-19 infection or diagnosis with Long COVID (Fig. 2, Fig. 4, Fig. 6, Fig. 8). We find that the magnitude of any protective effects are dependent on how the vaccination is modelled (time-independent versus time-dependent) as well as the definition of Long COVID being used (development of Long COVID symptoms versus diagnosis with Long COVID). This result is consistent with existing literature indicating an inconsistent and unclear effect, if any, of booster doses of COVID-19 vaccine against Long COVID [7–9]. Future work can assess if this result still holds as the population of people who have received a COVID-19 booster vaccine dose increases.

One of our most striking results is the lack of overlap in patients experiencing persistent or who developed Long COVID symptoms four weeks after COVID-19 infection versus those patients who were diagnosed with Long COVID (Table 2). Some degree of bias has previously been found in the assignment of ICD-10-CM codes for Long COVID [5], which may indicate that adoption of these codes is heterogeneous across providers and within health care systems. Additionally, Long COVID is a diagnosis of exclusion [2], which may lead to a delay in the addition of a Long COVID diagnostic code to a patient’s chart when compared to the addition of a Long COVID related symptom. Similarly, the symptoms based definition may be capturing patients who may not eventually be diagnosed with Long COVID. Symptom based definitions of Long COVID may have high sensitivity but low specificity for the condition while diagnosis based definitions may have high specificity but low sensitivity. More work should be done to define a gold standard for defining this condition. Finally, because there has been limited adoption of a standard definition of Long COVID the individual provider application of these codes and changing practices over time might account for substantial variation between those with Long COVID-like symptoms and those diagnosed with Long COVID [3].

Like all studies of EHR data, ours is subject to a variety of known limitations [56–61]. We are only able to identify events that are captured by the constituent health care systems that are a part of the Truveta member system. This means we will not capture COVID-19 infections or vaccinations which were recorded in a health care system that is not a part of Truveta. Similarly, we will not capture COVID-19 infections or vaccinations which were never reported to a health care system. This limitation means patients with a precedent COVID-19 infection may be missed as part of our inclusion and exclusion criteria. Another example limitation is that a patient’s COVID-19 vaccination status may not captured in our data because some member HCS may not reconcile their records with state health registries and other locations where patients receive care. Finally, comorbidity status may be misclassified in our data set because it is captured in a different, non-member HCS or are classified in the EHR using codes that were not present in our codesets. These are common and well understood limitations associated with using EHR data.

An additional limitation of our study was that we did not account for any potential differences in effect of vaccinations having to do with manufacturer or the type of booster dose. For example, we did not distinguish if a booster dose was from a bivalent formulation or not. This limitation means that we cannot distinguish if there is differences in protective effect against Long COVID associated with a particular make of vaccine. Future work can assess if there differences associated with different makes of COVID-19 vaccine that able to be estimated and if there are further meaningful differences associated with the type of booster dose a patient received. Additionally, our study focused on an adult population, so our results may not generatlize to pediatric populations, so a potential future avenue of study is a focused analysis of pediatric patients.

In the context of this study, these inherent limitations will most likely lead to an underestimation of the number of patients who experienced a COVID-19 infection and an underestimation of how many of those patients completed a primary COVID-19 vaccination sequence or received a booster dose and when. Under counting can result in underestimation of the effect of vaccination on persistence or development of Long COVID symptoms or diagnosis with Long COVID because individuals with unknown vaccination status will be incorrectly treated as unvaccinated which reduces the observed difference between patients who are unvaccinated versus patients who are vaccinated, and cause us to underestimate the protective effect of vaccination on risk of developing Long COVID symptoms or being diagnosed with Long COVID. By focusing our study on patients who experienced a positive COVID-19 test as recorded by the hospital systems, we hoped to limit the possibility of underestimating the vaccinated population as it represents individuals who are potentially more likely to interact with their hospital system than not.

This study adds to the growing literature demonstrating the protective effects of vaccination against Long COVID. Additionally, this study highlights how the definition of Long COVID impacts the estimated protective effect, if any, of completing a primary COVID-19 vaccination sequence or receiving a booster dose on the risk of persistent Long COVID symptoms or diagnosis with Long COVID. Inconsistency in defining this condition hinders study of this important topic.

## Acknowledgements

The authors thank Mackenzie Bogiages, Grace Turner, Jesse Weiss, Mohan Dharmarajan, and the clinical informaticists at Truveta for their assistance and input during the design and development of this study.

## Institutional Review Board Approval

This study performs analysis of de-identified electronic health records (EHR) data accessed via Truveta Studio. Truveta Studio only contains data that has been de-identified by expert determination in accordance with HIPAA Privacy Rule, and therefore this study was exempt from Institutional Review Board approval.

## Data Availability Statement

The data used in this study is available to all Truveta subscribers and may be accessed at studio.truveta.com. The R code used to perform all analyses and generate all tables and figures is available on GitHub at https://github.com/Truveta/smits_et_al_vaccines_long_covid.

