## Supplemental Material for "Effect of vaccination on time till Long COVID, a comparison of two ways to model effect of vaccination and two outcome definitions"

#### Contents

**1 COVID-19 laboratory tests** **2**

**2 COVID-19 vaccinations** **4**

**3 Long COVID diagnosis** **8**

**4 Long COVID symptoms** **9**

### 1 COVID-19 laboratory tests

Table S1: Concept codes used to identify if a patient had tested positive for COVID-19.

| CodeSystem | ConceptCode | ConceptName |
|---|---|---|
| LOINC | 94307-6 | SARS-CoV-2 (COVID-19) N gene [Presence] in Specimen by Nucleic acid amplification using CDC primer-probe set N1 |
| LOINC | 94308-4 | SARS-CoV-2 (COVID-19) N gene [Presence] in Specimen by Nucleic acid amplification using CDC primer-probe set N2 |
| LOINC | 94309-2 | SARS-CoV-2 (COVID-19) RNA [Presence] in Specimen by NAA with probe detection |
| LOINC | 94314-2 | SARS-CoV-2 (COVID-19) RdRp gene [Presence] in Specimen by NAA with probe detection |
| LOINC | 94316-7 | SARS-CoV-2 (COVID-19) N gene [Presence] in Specimen by NAA with probe detection |

Continued on next page

**TableS1 – continued from previous page**

| CodeSystem | ConceptCode | ConceptName |
|---|---|---|
| LOINC | 94500-6 | SARS-CoV-2 (COVID-19) RNA [Presence] in Respiratory specimen by NAA with probe detection |
| LOINC | 94533-7 | SARS-CoV-2 (COVID-19) N gene [Presence] in Respiratory specimen by NAA with probe detection |
| LOINC | 94534-5 | SARS-CoV-2 (COVID-19) RdRp gene [Presence] in Respiratory specimen by NAA with probe detection |
| LOINC | 94559-2 | SARS-CoV-2 (COVID-19) ORF1ab region [Presence] in Respiratory specimen by NAA with probe detection |
| LOINC | 94565-9 | SARS-CoV-2 (COVID-19) RNA [Presence] in Nasopharynx by NAA with non-probe detection |
| LOINC | 94639-2 | SARS-CoV-2 (COVID-19) ORF1ab region [Presence] in Specimen by NAA with probe detection |
| LOINC | 94640-0 | SARS-CoV-2 (COVID-19) S gene [Presence] in Respiratory specimen by NAA with probe detection |
| LOINC | 94641-8 | SARS-CoV-2 (COVID-19) S gene [Presence] in Specimen by NAA with probe detection |
| LOINC | 94660-8 | SARS-CoV-2 (COVID-19) RNA [Presence] in Serum or Plasma by NAA with probe detection |
| LOINC | 94756-4 | SARS-CoV-2 (COVID-19) N gene [Presence] in Respiratory specimen by Nucleic acid amplification using CDC primer-probe set N1 |
| LOINC | 94757-2 | SARS-CoV-2 (COVID-19) N gene [Presence] in Respiratory specimen by Nucleic acid amplification using CDC primer-probe set N2 |
| LOINC | 94759-8 | SARS-CoV-2 (COVID-19) RNA [Presence] in Nasopharynx by NAA with probe detection |
| LOINC | 94766-3 | SARS-CoV-2 (COVID-19) N gene [Presence] in Serum or Plasma by NAA with probe detection |
| LOINC | 94767-1 | SARS-CoV-2 (COVID-19) S gene [Presence] in Serum or Plasma by NAA with probe detection |
| LOINC | 94845-5 | SARS-CoV-2 (COVID-19) RNA [Presence] in Saliva (oral fluid) by NAA with probe detection |
| LOINC | 95406-5 | SARS-CoV-2 (COVID-19) RNA [Presence] in Nose by NAA with probe detection |
| LOINC | 95409-9 | SARS-CoV-2 (COVID-19) N gene [Presence] in Nose by NAA with probe detection |
| LOINC | 95425-5 | SARS-CoV-2 (COVID-19) N gene [Presence] in Saliva (oral fluid) by NAA with probe detection |
| LOINC | 95608-6 | SARS-CoV-2 (COVID-19) RNA [Presence] in Respiratory specimen by NAA with non-probe detection |
| LOINC | 95824-9 | SARS-CoV-2 (COVID-19) ORF1ab region [Presence] in Saliva (oral fluid) by NAA with probe detection |
| LOINC | 96091-4 | SARS-CoV-2 (COVID-19) RdRp gene [Presence] in Saliva (oral fluid) by NAA with probe detection |
| LOINC | 96120-1 | SARS-CoV-2 (COVID-19) RdRp gene [Presence] in Lower respiratory specimen by NAA with probe detection |

Continued on next page

**TableS1 – continued from previous page**

| CodeSystem | ConceptCode | ConceptName |
|---|---|---|
| LOINC | 96123-5 | SARS-CoV-2 (COVID-19) RdRp gene [Presence] in Upper respiratory specimen by NAA with probe detection |
| LOINC | 96448-6 | SARS-CoV-2 (COVID-19) N gene [Presence] in Saliva (oral fluid) by Nucleic acid amplification using CDC primer-probe set N1 |
| LOINC | 98131-6 | SARS-CoV-2 (COVID-19) ORF1b region [Presence] in Respiratory specimen by NAA with probe detection |
| LOINC | 98132-4 | SARS-CoV-2 (COVID-19) ORF1a region [Presence] in Respiratory specimen by NAA with probe detection |
| LOINC | 98493-0 | SARS-CoV-2 (COVID-19) ORF1b region [Presence] in Saliva (oral fluid) by NAA with probe detection |
| LOINC | 98494-8 | SARS-CoV-2 (COVID-19) ORF1a region [Presence] in Saliva (oral fluid) by NAA with probe detection |
| LOINC | 96763-8 | SARS-CoV-2 (COVID-19) E gene [Presence] in Respiratory specimen by NAA with probe detection |
| LOINC | 96765-3 | SARS-CoV-2 (COVID-19) S gene [Presence] in Saliva (oral fluid) by NAA with probe detection |
| LOINC | 96797-6 | SARS-CoV-2 (COVID-19) RNA [Presence] in Oropharyngeal wash by NAA with probe detection |
| LOINC | 96829-7 | SARS-CoV-2 (COVID-19) RNA [Presence] in Specimen from Donor by NAA with probe detection |
| LOINC | 96957-6 | SARS-CoV-2 (COVID-19) M gene [Presence] in Upper respiratory specimen by NAA with probe detection |
| LOINC | 96958-4 | SARS-CoV-2 (COVID-19) N gene [Presence] in Saliva (oral fluid) by Nucleic acid amplification using CDC primer-probe set N2 |
| LOINC | 96986-5 | SARS-CoV-2 (COVID-19) N gene [Presence] in Nose by NAA with non-probe detection |
| LOINC | 97098-8 | SARS-CoV-2 (COVID-19) Nsp2 gene [Presence] in Upper respiratory specimen by NAA with probe detection |

### 2 COVID-19 vaccinations

Table S2: Concept codes used to identify if a patient had a COVID-19 vaccination event.

| CodeSystem | ConceptCode | ConceptName |
|---|---|---|
| CVX | 207 | SARS-COV-2 (COVID-19) vaccine, mRNA, spike protein, LNP, preservative free, 100 mcg/0.5mL dose |
| CVX | 208 | SARS-COV-2 (COVID-19) vaccine, mRNA, spike protein, LNP, preservative free, 30 mcg/0.3mL dose |
| CVX | 212 | SARS-COV-2 (COVID-19) vaccine, vector non-replicating, recombinant spike protein-Ad26, preservative free, 0.5 mL |
| CVX | 217 | SARS-COV-2 (COVID-19) vaccine, mRNA, spike protein, LNP, preservative free, 30 mcg/0.3 mL dose, tris-sucrose formulation |
| CVX | 218 | SARS-COV-2 (COVID-19) vaccine, mRNA, spike protein, LNP, preservative free, 10 mcg/0.2 mL dose, tris-sucrose formulation |

Continued on next page

**TableS2 – continued from previous page**

| CodeSystem | ConceptCode | ConceptName |
|---|---|---|
| CVX | 219 | SARS-COV-2 (COVID-19) vaccine, mRNA, spike protein, LNP, preservative free, 3 mcg/0.2 mL dose, tris-sucrose formulation |
| CVX | 300 | SARS-COV-2 (COVID-19) vaccine, mRNA, spike protein, LNP, bivalent booster, preservative free, 30 mcg/0.3mL dose, tris-sucrose formulation |
| CVX | 301 | SARS-COV-2 (COVID-19) vaccine, mRNA, spike protein, LNP, bivalent booster, preservative free, 10 mcg/0.2 mL dose, tris-sucrose formulation |
| CVX | 221 | COVID-19, mRNA, LNP-S, PF, 50 mcg/0.5 mL dose |
| CVX | 228 | COVID-19, mRNA, LNP-S, PF, pediatric 25 mcg/0.25 mL dose |
| RxNorm | 2470234 | SARS-CoV-2 (COVID-19) vaccine, mRNA-1273 0.2 MG/ML Injectable Suspension |
| RxNorm | 2468235 | SARS-CoV-2 (COVID-19) vaccine, mRNA-BNT162b2 0.1 MG/ML Injectable Suspension |
| RxNorm | 2479835 | SARS-COV-2 (COVID-19) vaccine, vector - Ad26 100000000000 UNT/ML Injectable Suspension |
| RxNorm | 2583743 | SARS-CoV-2 (COVID-19) vaccine, mRNA-BNT162b2 0.05 MG/ML Injectable Suspension |
| RxNorm | 2606078 | SARS-CoV-2 (COVID-19) vaccine, protein NVX-CoV2373 0.01 MG/ML Injectable Suspension |
| RxNorm | 2598700 | SARS-CoV-2 (COVID-19) vaccine, mRNA-1273 0.1 MG/ML Injectable Suspension |
| RxNorm | 2593847 | SARS-CoV-2 (COVID-19) vaccine, mRNA-BNT162b2 0.1 MG/ML Injectable Suspension [Comirnaty] |
| RxNorm | 2601552 | SARS-CoV-2 (COVID-19) vaccine, mRNA-1273 0.2 MG/ML Injectable Suspension [Spikevax] |
| NDC | 59267007801 | PFIZER-BIONTECH COVID-19 VACCINE - bnt162b2 injection, suspension |
| NDC | 59267007804 | bnt162b2 3ug/.2mL INTRAMUSCULAR INJECTION, SUSPENSION |
| NDC | 59267100001 | SARS-CoV-2 (COVID-19) vaccine, mRNA-BNT162b2 0.1 MG/ML Injectable Suspension |
| NDC | 59267100002 | SARS-CoV-2 (COVID-19) vaccine, mRNA-BNT162b2 0.1 MG/ML Injectable Suspension |
| NDC | 59267100003 | SARS-CoV-2 (COVID-19) vaccine, mRNA-BNT162b2 0.1 MG/ML Injectable Suspension |
| NDC | 59267102501 | SARS-CoV-2 (COVID-19) vaccine, mRNA-BNT162b2 0.1 MG/ML Injectable Suspension |
| NDC | 59267102502 | SARS-CoV-2 (COVID-19) vaccine, mRNA-BNT162b2 0.1 MG/ML Injectable Suspension |
| NDC | 59267102503 | SARS-CoV-2 (COVID-19) vaccine, mRNA-BNT162b2 0.1 MG/ML Injectable Suspension |
| NDC | 59267102504 | SARS-CoV-2 (COVID-19) vaccine, mRNA-BNT162b2 0.1 MG/ML Injectable Suspension |
| NDC | 59267105501 | SARS-CoV-2 (COVID-19) vaccine, mRNA-BNT162b2 0.05 MG/ML Injectable Suspension |

Continued on next page

**TableS2 – continued from previous page**

| CodeSystem | ConceptCode | ConceptName |
|---|---|---|
| NDC | 59267105502 | SARS-CoV-2 (COVID-19) vaccine, mRNA-BNT162b2 0.05 MG/ML Injectable Suspension |
| NDC | 59267105504 | SARS-CoV-2 (COVID-19) vaccine, mRNA-BNT162b2 0.05 MG/ML Injectable Suspension |
| NDC | 59676058005 | Janssen COVID-19 vaccine, DNA, spike protein, adenovirus type 26 (Ad26) vector, preservative free, 5x1010 viral particles/0.5mL dosage, for intramuscular use |
| NDC | 59676058015 | SARS-COV-2 (COVID-19) vaccine, vector - Ad26 100000000000 UNT/ML Injectable Suspension |
| NDC | 80777027310 | SARS-CoV-2 (COVID-19) vaccine, mRNA-1273 0.2 MG/ML Injectable Suspension |
| NDC | 80777027315 | SARS-CoV-2 (COVID-19) vaccine, mRNA-1273 0.2 MG/ML Injectable Suspension |
| NDC | 80777027398 | SARS-CoV-2 (COVID-19) vaccine, mRNA-1273 0.2 MG/ML Injectable Suspension |
| NDC | 80777027399 | SARS-CoV-2 (COVID-19) vaccine, mRNA-1273 0.2 MG/ML Injectable Suspension |
| CPT | 0094A | Administration of coronavirus vaccine 10, reserved |
| CPT | 0093A | Administration of coronavirus vaccine 10, booster |
| CPT | 0091A | Administration of coronavirus vaccine 10, dose 1 |
| CPT | 0002A | Immunization administration by intramuscular injection of severe acute respiratory syndrome coronavirus 2 (SARS-CoV-2) (coronavirus disease [COVID-19]) vaccine, mRNA-LNP, spike protein, preservative free, 30 mcg/0.3 mL dosage, diluent reconstituted; second dose |
| CPT | 0011A | Immunization administration by intramuscular injection of severe acute respiratory syndrome coronavirus 2 (SARS-CoV-2) (coronavirus disease [COVID-19]) vaccine, mRNA-LNP, spike protein, preservative free, 100 mcg/0.5 mL dosage; first dose |
| CPT | 0022A | Immunization administration by intramuscular injection of severe acute respiratory syndrome coronavirus 2 (SARS-CoV-2) (coronavirus disease [COVID-19]) vaccine, DNA, spike protein, chimpanzee adenovirus Oxford 1 (ChAdOx1) vector, preservative free, 5x1010 viral particles/0.5 mL dosage; second dose |
| CPT | 0021A | Immunization administration by intramuscular injection of severe acute respiratory syndrome coronavirus 2 (SARS-CoV-2) (coronavirus disease [COVID-19]) vaccine, DNA, spike protein, chimpanzee adenovirus Oxford 1 (ChAdOx1) vector, preservative free, 5x1010 viral particles/0.5 mL dosage; first dose |
| CPT | 0034A | Administration of coronavirus vaccine 4, reserved |
| CPT | 0042A | Immunization administration by intramuscular injection of severe acute respiratory syndrome coronavirus 2 (SARS-CoV-2) (coronavirus disease [COVID-19]) vaccine, recombinant spike protein nanoparticle, saponin-based adjuvant, preservative free, 5 mcg/0.5 mL dosage; second dose |

Continued on next page

**TableS2 – continued from previous page**

| CodeSystem | ConceptCode | ConceptName |
|---|---|---|
| CPT | 0041A | Immunization administration by intramuscular injection of severe acute respiratory syndrome coronavirus 2 (SARS-CoV-2) (coronavirus disease [COVID-19]) vaccine, recombinant spike protein nanoparticle, saponin-based adjuvant, preservative free, 5 mcg/0.5 mL dosage; first dose |
| CPT | 0072A | Pfizer-BioNTech Covid-19 Pediatric Vaccine - Administration - Second dose |
| CPT | 0064A | Moderna Covid-19 Vaccine (Low Dose) Administration - Booster |
| CPT | 0054A | Pfizer-BioNTech Covid-19 Vaccine (Ready to Use) Administration - Booster |
| CPT | 0071A | Pfizer-BioNTech Covid-19 Pediatric Vaccine - Administration - First dose |
| CPT | 0012A | Immunization administration by intramuscular injection of severe acute respiratory syndrome coronavirus 2 (SARS-CoV-2) (coronavirus disease [COVID-19]) vaccine, mRNA-LNP, spike protein, preservative free, 100 mcg/0.5 mL dosage; second dose |
| CPT | 0031A | Immunization administration by intramuscular injection of severe acute respiratory syndrome coronavirus 2 (SARS-CoV-2) (coronavirus disease [COVID-19]) vaccine, DNA, spike protein, adenovirus type 26 (Ad26) vector, preservative free, 5x1010 viral particles/0.5 mL dosage, single dose |
| CPT | 0001A | Immunization administration by intramuscular injection of severe acute respiratory syndrome coronavirus 2 (SARS-CoV-2) (coronavirus disease [COVID-19]) vaccine, mRNA-LNP, spike protein, preservative free, 30 mcg/0.3 mL dosage, diluent reconstituted; first dose |
| CPT | 91302 | Severe acute respiratory syndrome coronavirus 2 (SARS-CoV-2) (coronavirus disease [COVID-19]) vaccine, DNA, spike protein, chimpanzee adenovirus Oxford 1 (ChAdOx1) vector, preservative free, 5x1010 viral particles/0.5 mL dosage, for intramuscular use |
| CPT | 91303 | Severe acute respiratory syndrome coronavirus 2 (SARS-CoV-2) (coronavirus disease [COVID-19]) vaccine, DNA, spike protein, adenovirus type 26 (Ad26) vector, preservative free, 5x1010 viral particles/0.5 mL dosage, for intramuscular use |
| CPT | 91300 | Severe acute respiratory syndrome coronavirus 2 (SARS-CoV-2) (coronavirus disease [COVID-19]) vaccine, mRNA-LNP, spike protein, preservative free, 30 mcg/0.3 mL dosage, diluent reconstituted, for intramuscular use |
| CPT | 91301 | Severe acute respiratory syndrome coronavirus 2 (SARS-CoV-2) (coronavirus disease [COVID-19]) vaccine, mRNA-LNP, spike protein, preservative free, 100 mcg/0.5 mL dosage, for intramuscular use |
| CPT | 0081A | Administration of coronavirus vaccine 9, dose 1 |

Continued on next page

**TableS2 – continued from previous page**

| CodeSystem | ConceptCode | ConceptName |
|---|---|---|
| CPT | 0003A | Immunization administration by intramuscular injection of severe acute respiratory syndrome coronavirus 2 (SARS-CoV2) (coronavirus disease [COVID-19]) vaccine, mRNA-LNP, spike protein, preservative free, 30 mcg/0.3 mL dosage, diluent reconstituted; third dose |
| CPT | 0051A | Pfizer-BioNTech Covid-19 Vaccine (Ready to Use) Administration - First dose |
| CPT | 0053A | Pfizer-BioNTech Covid-19 Vaccine (Ready to Use) Administration - Third dose |
| CPT | 0083A | Administration of coronavirus vaccine 9, booster |
| CPT | 0052A | Pfizer-BioNTech Covid-19 Vaccine (Ready to Use) Administration - Second dose |
| CPT | 0073A | Administration of coronavirus vaccine 8, booster |
| CPT | 0104A | Administration of coronavirus vaccine 11, reserved |
| CPT | 0004A | Pfizer-BioNTech Covid-19 Vaccine Administration - Booster |
| CPT | 0074A | Administration of coronavirus vaccine 8, reserved |
| CPT | 0013A | Immunization administration by intramuscular injection of severe acute respiratory syndrome coronavirus 2 (SARS-CoV-2) (coronavirus disease [COVID-19]) vaccine, mRNA-LNP, spike protein, preservative free, 100 mcg/0.5 mL dosage; third dose |
| CPT | 0092A | Administration of coronavirus vaccine 10, dose 2 |
| CPT | 0082A | Administration of coronavirus vaccine 9, dose 2 |
| CPT | 91305 | Pfizer-BioNTech Covid-19 Vaccine (Ready to Use) |
| CPT | M0201 | Covid-19 vaccine administration inside a patient's home; reported only once per individual home per date of service when only covid-19 vaccine administration is performed at the patient's home |
| CPT | 91306 | Moderna Covid-19 Vaccine (Low Dose) |
| CPT | 91307 | Pfizer-BioNTech Covid-19 Pediatric Vaccine |
| CPT | 91308 | Coronavirus vaccine 9 |
| CPT | 91309 | Coronavirus vaccine 10 |
| CPT | 91310 | Coronavirus vaccine 11 |
| CPT | 91304 | Severe acute respiratory syndrome coronavirus 2 (SARS-CoV-2) (coronavirus disease [COVID-19]) vaccine, recombinant spike protein nanoparticle, saponin-based adjuvant, preservative free, 5 mcg/0.5 mL dosage, for intramuscular use |

### 3 Long COVID diagnosis

Table S3: Concept codes used to identify if a patient was diagnosed with Long COVID via an ICD10-CM and SNOMED CT code.

| CodeSystem | ConceptCode | ConceptName |
|---|---|---|
| ICD10CM | U09.9 | Post COVID-19 condition, unspecified |

Continued on next page

**TableS3 – continued from previous page**

| CodeSystem | ConceptCode | ConceptName |
|---|---|---|
| SNOMED CT | 1119303003 | Post-acute COVID-19 |
| SNOMED CT | 1119304009 | Chronic post-COVID-19 syndrome |
| Truveta | 1215955 | Post-COVID-19 chronic anxiety |
| Truveta | 1215956 | Post-COVID-19 chronic concentration deficit |
| Truveta | 1215957 | Post-COVID-19 chronic cough |
| Truveta | 1215958 | Post-COVID-19 chronic decreased mobility and endurance |
| Truveta | 1215959 | Post-COVID-19 chronic dyspnea |
| Truveta | 1215960 | Post-COVID-19 chronic fatigue |
| Truveta | 1215961 | Post-COVID-19 chronic headache |
| Truveta | 1215962 | Post-COVID-19 chronic joint pain |
| Truveta | 1215963 | Post-COVID-19 chronic loss of smell |
| Truveta | 1215964 | Post-COVID-19 chronic loss of smell and taste |
| Truveta | 1215965 | Post-COVID-19 chronic loss of taste |
| Truveta | 1215966 | Post-COVID-19 chronic muscle pain |
| Truveta | 1215967 | Post-COVID-19 chronic neurologic symptoms |
| Truveta | 1215968 | Post-COVID-19 chronic palpitations |

### 4 Long COVID symptoms

#### 4.1 Abdominal pain

Table S4: Concept codes used to identify if a patient was diagnosed with abdominal pain.

| CodeSystem | ConceptCode | ConceptName |
|---|---|---|
| ICD10CM | R10.1 | Pain localized to upper abdomen |
| ICD10CM | R10.3 | Pain localized to other parts of lower abdomen |
| ICD10CM | R10.8 | Other abdominal pain |
| ICD10CM | R10.9 | Unspecified abdominal pain |
| ICD10CM | R10.10 | Upper abdominal pain, unspecified |
| ICD10CM | R10.11 | Right upper quadrant pain |
| ICD10CM | R10.12 | Left upper quadrant pain |
| ICD10CM | R10.13 | Epigastric pain |
| ICD10CM | R10.30 | Lower abdominal pain, unspecified |
| ICD10CM | R10.31 | Right lower quadrant pain |
| ICD10CM | R10.32 | Left lower quadrant pain |
| ICD10CM | R10.33 | Periumbilical pain |
| ICD10CM | R10.81 | Abdominal tenderness |
| ICD10CM | R10.811 | Right upper quadrant abdominal tenderness |
| ICD10CM | R10.812 | Left upper quadrant abdominal tenderness |
| ICD10CM | R10.813 | Right lower quadrant abdominal tenderness |
| ICD10CM | R10.814 | Left lower quadrant abdominal tenderness |
| ICD10CM | R10.815 | Periumbilic abdominal tenderness |
| ICD10CM | R10.816 | Epigastric abdominal tenderness |

Continued on next page

**TableS4 – continued from previous page**

| CodeSystem | ConceptCode | ConceptName |
|---|---|---|
| ICD10CM | R10.817 | Generalized abdominal tenderness |
| ICD10CM | R10.819 | Abdominal tenderness, unspecified site |
| ICD10CM | R10.82 | Rebound abdominal tenderness |
| ICD10CM | R10.821 | Right upper quadrant rebound abdominal tenderness |
| ICD10CM | R10.822 | Left upper quadrant rebound abdominal tenderness |
| ICD10CM | R10.823 | Right lower quadrant rebound abdominal tenderness |
| ICD10CM | R10.824 | Left lower quadrant rebound abdominal tenderness |
| ICD10CM | R10.825 | Periumbilic rebound abdominal tenderness |
| ICD10CM | R10.826 | Epigastric rebound abdominal tenderness |
| ICD10CM | R10.827 | Generalized rebound abdominal tenderness |
| ICD10CM | R10.829 | Rebound abdominal tenderness, unspecified site |
| ICD10CM | R10.83 | Colic |
| ICD10CM | R10.84 | Generalized abdominal pain |
| SNOMED CT | 21522001 | Abdominal pain |
| SNOMED CT | 266599000 | Dysmenorrhea |
| SNOMED CT | 271681002 | Stomach ache |
| SNOMED CT | 271832001 | Flatulence, eructation and gas pain |
| SNOMED CT | 271853005 | Hypochondrial pain |
| SNOMED CT | 271857006 | Loin pain |
| SNOMED CT | 271858001 | Recurrent acute abdominal pain |
| SNOMED CT | 272021002 | C/O - ureteric colic |
| SNOMED CT | 272047006 | C/O - loin pain |
| SNOMED CT | 272049009 | C/O - renal pain |
| SNOMED CT | 274277005 | C/O right iliac fossa pain |
| SNOMED CT | 274278000 | C/O left iliac fossa pain |
| SNOMED CT | 274279008 | Renal pain |
| SNOMED CT | 274280006 | Ureteric pain |
| SNOMED CT | 274287009 | O/E - abdominal pain |
| SNOMED CT | 274288004 | O/E - epigastric pain |
| SNOMED CT | 274289007 | On examination - iliac pain - abdominal |
| SNOMED CT | 274290003 | On examination - lumbar pain abdominal |
| SNOMED CT | 275314000 | Anal stump pain |
| SNOMED CT | 275406005 | Appendicular pain |
| SNOMED CT | 276498006 | C/O - ureteric pain |
| SNOMED CT | 279028009 | Ovarian pain |
| SNOMED CT | 285387005 | Left sided abdominal pain |
| SNOMED CT | 285388000 | Right sided abdominal pain |
| SNOMED CT | 286993001 | Dysmenorrhea - non-psychogenic |
| SNOMED CT | 289669005 | Pain of uterus |
| SNOMED CT | 289670006 | Pain on movement of uterus |
| SNOMED CT | 289732000 | Painful uterine contractions |
| SNOMED CT | 289817003 | Pain of cervix |
| SNOMED CT | 289818008 | Pain on movement of cervix |
| SNOMED CT | 289845009 | Pain on movement of ovary |
| SNOMED CT | 301367001 | Right hypochondrial pain |
| SNOMED CT | 301368006 | Left hypochondrial pain |
| SNOMED CT | 301715003 | Left upper quadrant pain |

Continued on next page

**TableS4 – continued from previous page**

| CodeSystem | ConceptCode | ConceptName |
|---|---|---|
| SNOMED CT | 301716002 | Left lower quadrant pain |
| SNOMED CT | 301717006 | Right upper quadrant pain |
| SNOMED CT | 301754002 | Right lower quadrant pain |
| SNOMED CT | 302770003 | Tenesmus - anal |
| SNOMED CT | 304542004 | Nonspecific abdominal pain |
| SNOMED CT | 308903002 | On examination - epigastric pain on palpation |
| SNOMED CT | 309737007 | Abdominal pain in pregnancy |
| SNOMED CT | 314041007 | Abdominal pain in early pregnancy |
| SNOMED CT | 314212008 | Abdominal pain - cause unknown |
| SNOMED CT | 371094000 | Colicky hypogastric pain |
| SNOMED CT | 371102005 | Generalized colicky abdominal pain |
| SNOMED CT | 414991007 | Painful haemorrhage of rectum |
| SNOMED CT | 426135001 | Chronic prostatitis - chronic pelvic pain syndrome |
| SNOMED CT | 426628005 | Chronic vaginal pain |
| SNOMED CT | 438506002 | Visceral abdominal pain |
| SNOMED CT | 439469002 | Recurrent abdominal pain |
| SNOMED CT | 443503005 | Periumbilical pain |
| SNOMED CT | 444746004 | Acute exacerbation of chronic abdominal pain |
| SNOMED CT | 449890002 | Functional abdominal pain syndrome |
| SNOMED CT | 707597009 | Continuous abdominal pain of left lower quadrant |
| SNOMED CT | 69186005 | Broad ligament laceration syndrome |
| SNOMED CT | 71850005 | Abdominal pain worse on motion |
| SNOMED CT | 74704000 | Abdominal pain through to back |
| SNOMED CT | 75879005 | Abdominal migraine |
| SNOMED CT | 77880009 | Rectal pain |
| SNOMED CT | 79922009 | Epigastric pain |
| SNOMED CT | 83132003 | Upper abdominal pain |
| SNOMED CT | 86208007 | Loin pain-hematuria syndrome |
| SNOMED CT | 88522004 | Umbilical pain |
| SNOMED CT | 102613000 | Localized abdominal pain |
| SNOMED CT | 102614006 | Generalized abdominal pain |
| SNOMED CT | 102615007 | Ulcer-type pain |
| SNOMED CT | 102626001 | Liver pain |
| SNOMED CT | 102628000 | Gallbladder pain |
| SNOMED CT | 102631004 | Pancreatic pain |
| SNOMED CT | 102831002 | Renal angle pain |
| SNOMED CT | 111985007 | Chronic abdominal pain |
| SNOMED CT | 116290004 | Acute abdominal pain |
| SNOMED CT | 162038003 | Non-colicky abdominal pain |
| SNOMED CT | 162040008 | Abdominal migraine - symptom |
| SNOMED CT | 162042000 | Abdominal wall pain |
| SNOMED CT | 162046002 | Central abdominal pain |
| SNOMED CT | 162047006 | Left subcostal pain |
| SNOMED CT | 162048001 | Right subcostal pain |
| SNOMED CT | 162049009 | Left flank pain |
| SNOMED CT | 162050009 | Right flank pain |
| SNOMED CT | 162051008 | Right iliac fossa pain |

Continued on next page

**TableS4 – continued from previous page**

| CodeSystem | ConceptCode | ConceptName |
|---|---|---|
| SNOMED CT | 162052001 | Left iliac fossa pain |
| SNOMED CT | 162053006 | Suprapubic pain |
| SNOMED CT | 163214004 | On examination - abdominal pain - right hypochondrium |
| SNOMED CT | 163215003 | On examination - abdominal pain - epigastrium |
| SNOMED CT | 163216002 | On examination - abdominal pain - left hypochondrium |
| SNOMED CT | 163217006 | On examination - abdominal pain - right lumbar |
| SNOMED CT | 163218001 | On examination - abdominal pain - umbilical |
| SNOMED CT | 163219009 | On examination - abdominal pain - left lumbar |
| SNOMED CT | 163220003 | On examination - abdominal pain - right iliac |
| SNOMED CT | 163221004 | On examination - abdominal pain - hypogastrium |
| SNOMED CT | 163222006 | On examination - abdominal pain - left iliac |
| SNOMED CT | 164167004 | O/E - painful splenomegaly |
| SNOMED CT | 196791007 | Appendicular colic |
| SNOMED CT | 197232005 | Anorectal pain |
| SNOMED CT | 230648001 | Abdominal cutaneous nerve entrapment syndrome |
| SNOMED CT | 235841007 | Chronic nonspecific abdominal pain |
| SNOMED CT | 236554001 | Pain during inflow of dialysate |
| SNOMED CT | 236555000 | Pain during outflow of dialysate |
| SNOMED CT | 238970000 | Proctodynia |
| SNOMED CT | 247352008 | Subcostal pain |
| SNOMED CT | 247353003 | Site of abdominal pain |
| SNOMED CT | 247354009 | Iliac fossa pain |
| SNOMED CT | 247355005 | Flank pain |
| SNOMED CT | 247361008 | Evening colic |
| SNOMED CT | 247362001 | Pain on abdominal wall movement |
| SNOMED CT | 5268006 | Mechanical dysmenorrhea |
| SNOMED CT | 6548007 | Rectal tenesmus |
| SNOMED CT | 6744007 | Painful bladder spasm |
| SNOMED CT | 7093002 | Renal colic |
| SNOMED CT | 9991008 | Abdominal colic |
| SNOMED CT | 1197719006 | Functional abdominal pain of childhood |
| SNOMED CT | 15803009 | Bladder pain |
| SNOMED CT | 16844001 | Painful urging to urinate |
| SNOMED CT | 17329003 | Ureteric colic |
| SNOMED CT | 21005005 | Burning epigastric pain |
| SNOMED CT | 32096006 | Secondary dysmenorrhea |
| SNOMED CT | 35074008 | Chronic idiopathic anal pain |
| SNOMED CT | 35363006 | Infantile colic |
| SNOMED CT | 36729000 | Pain of prostate |
| SNOMED CT | 37389005 | Biliary colic |
| SNOMED CT | 37488003 | Vulvovaginal pain |
| SNOMED CT | 38343000 | Vaginal pain |
| SNOMED CT | 38654001 | Recurrent biliary colic |
| SNOMED CT | 39402007 | Pelvic congestion syndrome |
| SNOMED CT | 43548008 | Mittelschmerz |
| SNOMED CT | 45979003 | Abdominal wind pain |
| SNOMED CT | 54586004 | Lower abdominal pain |

Continued on next page

**TableS4 – continued from previous page**

| CodeSystem | ConceptCode | ConceptName |
|---|---|---|
| SNOMED CT | 60043000 | Midabdominal crampy pain |
| SNOMED CT | 62647006 | Painful spasm of anus |
| SNOMED CT | 68653001 | Anal pain |
| SNOMED CT | 722876002 | Functional heartburn |
| SNOMED CT | 784285002 | Vesical tenesmus |
| SNOMED CT | 838410008 | Pain in epigastric region on palpation |
| SNOMED CT | 838411007 | Pain in abdominal region on palpation |
| SNOMED CT | 122141000119109 | Refractory abdominal migraine |
| SNOMED CT | 125661000119107 | Biliary colic with obstruction |
| SNOMED CT | 136051000119105 | Abdominal colic in child less than or equal to 12 months |
| SNOMED CT | 136571000119109 | Adult colic |
| SNOMED CT | 137891000119105 | Abdominal colic in adult or child greater than 12 months |
| SNOMED CT | 10760581000119104 | Pain in round ligament in pregnancy |
| SNOMED CT | 14440001000004101 | Postoperative abdominal pain |
| SNOMED CT | 14700001000004102 | Intractable abdominal pain |
| SNOMED CT | 16580691000119107 | Chronic urinary bladder pain |
| SNOMED CT | 65754002 | Spasmodic dysmenorrhea |
| SNOMED CT | 1119217009 | Pain in right abdominal lumbar region |
| SNOMED CT | 1119218004 | Pain in left abdominal lumbar region |
| SNOMED CT | 1197734000 | Chronic proctalgia |

#### 4.2 Anosmia

Table S5: Concept codes used to identify if a patient was diagnosed with anosmia.

| CodeSystem | ConceptCode | ConceptName |
|---|---|---|
| ICD10CM | R43 | Disturbances of smell and taste |
| ICD10CM | R43.0 | Anosmia |
| ICD10CM | R43.1 | Parosmia |
| ICD10CM | R43.2 | Parageusia |
| ICD10CM | R43.8 | Other disturbances of smell and taste |
| ICD10CM | R43.9 | Unspecified disturbances of smell and taste |
| SNOMED CT | 76489005 | Finding of sense of taste |
| SNOMED CT | 106151004 | Finding of sense of smell |
| SNOMED CT | 271801002 | Taste sense altered |
| SNOMED CT | 272041007 | C/O - loss of taste sense |
| SNOMED CT | 275462005 | Disorder of smell |
| SNOMED CT | 299899002 | Problem of sense of smell |
| SNOMED CT | 299900007 | Sense of smell normal |
| SNOMED CT | 299901006 | Sense of taste normal |
| SNOMED CT | 302188001 | Bad taste in mouth |
| SNOMED CT | 346138009 | Neurologic unpleasant taste |
| SNOMED CT | 348083004 | Primary taste disorder |
| SNOMED CT | 349084003 | Unpleasant taste in mouth |

Continued on next page

**TableS5 – continued from previous page**

| CodeSystem | ConceptCode | ConceptName |
|---|---|---|
| SNOMED CT | 399993004 | Disorder of taste |
| SNOMED CT | 697990000 | Hypogeusia |
| SNOMED CT | 697992008 | Hemiageusia |
| SNOMED CT | 698002002 | Loss of taste anterior two thirds of tongue |
| SNOMED CT | 699220008 | Phantom taste |
| SNOMED CT | 699656000 | Radiation induced taste impairment |
| SNOMED CT | 702531002 | Loss of taste posterior one third of tongue |
| SNOMED CT | 708673009 | Parosmia |
| SNOMED CT | 722027009 | Kallman syndrome with heart disease |
| SNOMED CT | 83156004 | Sense of smell impaired |
| SNOMED CT | 93559003 | Hypogonadism with anosmia |
| SNOMED CT | 110365009 | Aftertaste |
| SNOMED CT | 112105008 | Sense of smell altered |
| SNOMED CT | 112107000 | Primary bitter taste disorder |
| SNOMED CT | 164055003 | On examination - taste loss anterior 2/3 tongue |
| SNOMED CT | 164056002 | O/E-taste loss post 1/3 tongue |
| SNOMED CT | 164083001 | O/E - smell tested |
| SNOMED CT | 164084007 | O/E - smell normal |
| SNOMED CT | 164085008 | O/E - smell abnormal |
| SNOMED CT | 164086009 | O/E - anosmia |
| SNOMED CT | 230502003 | Congenital anosmia |
| SNOMED CT | 247318005 | Abnormal taste in mouth |
| SNOMED CT | 247319002 | Taste of food lingers |
| SNOMED CT | 247320008 | Things smell different |
| SNOMED CT | 247321007 | Unusual smell in nose |
| SNOMED CT | 11193009 | Metallic taste |
| SNOMED CT | 12869004 | Primary salt taste disorder |
| SNOMED CT | 15732002 | Secondary taste disorder |
| SNOMED CT | 20801005 | Secondary acid taste disorder |
| SNOMED CT | 22281006 | Pseudogeusesthesia |
| SNOMED CT | 26440003 | Franklinic taste |
| SNOMED CT | 34680005 | Primary sweet taste disorder |
| SNOMED CT | 36905001 | Secondary bitter taste disorder |
| SNOMED CT | 36955009 | Loss of taste |
| SNOMED CT | 37303008 | Sensitivity to individual odor |
| SNOMED CT | 37481009 | Anterior tongue taste disorder |
| SNOMED CT | 38175008 | Garlic taste |
| SNOMED CT | 40307005 | Primary acid taste disorder |
| SNOMED CT | 41783004 | Traumatic anosmia |
| SNOMED CT | 44169009 | Loss of sense of smell |
| SNOMED CT | 44525005 | Abnormal unpleasant perception of strong scent |
| SNOMED CT | 45846002 | Sensitive to smells |
| SNOMED CT | 47339000 | Secondary sweet taste disorder |
| SNOMED CT | 57791006 | Taste-blindness |
| SNOMED CT | 66073004 | Glossopharyngeal taste disorder |
| SNOMED CT | 67724008 | Secondary salt taste disorder |
| SNOMED CT | 771147003 | Isolated arhinencephaly |

Continued on next page

**TableS5 – continued from previous page**

| CodeSystem | ConceptCode | ConceptName |
|---|---|---|
| SNOMED CT | 272028008 | Complaining of anosmia |
| Truveta | 1215963 | Post-COVID-19 chronic loss of smell |
| Truveta | 1215964 | Post-COVID-19 chronic loss of smell and taste |
| Truveta | 1215965 | Post-COVID-19 chronic loss of taste |

#### 4.3 Anxiety and depression relevant to Long COVID

Table S6: Concept codes used to identify if a patient was diagnosed with general anxiety or depression relevant to Long COVID.

| CodeSystem | ConceptCode | ConceptName |
|---|---|---|
| ICD10CM | N91 | Absent, scanty and rare menstruation |
| ICD10CM | N92 | Excessive, frequent and irregular menstruation |
| ICD10CM | N94.5 | Secondary dysmenorrhea |
| ICD10CM | N94.6 | Dysmenorrhea, unspecified |
| ICD10CM | N91.0 | Primary amenorrhea |
| ICD10CM | N91.1 | Secondary amenorrhea |
| ICD10CM | N91.2 | Amenorrhea, unspecified |
| ICD10CM | N91.3 | Primary oligomenorrhea |
| ICD10CM | N91.4 | Secondary oligomenorrhea |
| ICD10CM | N91.5 | Oligomenorrhea, unspecified |
| ICD10CM | N92.0 | Excessive and frequent menstruation with regular cycle |
| ICD10CM | N92.1 | Excessive and frequent menstruation with irregular cycle |
| ICD10CM | N92.2 | Excessive menstruation at puberty |
| ICD10CM | N92.3 | Ovulation bleeding |
| ICD10CM | N92.4 | Excessive bleeding in the premenopausal period |
| ICD10CM | N92.5 | Other specified irregular menstruation |
| ICD10CM | N92.6 | Irregular menstruation, unspecified |
| SNOMED CT | 248968007 | Finding of pattern of menstrual cycle |
| SNOMED CT | 266601003 | Excessive and frequent menstruation |
| SNOMED CT | 266603000 | Ovulation bleeding |
| SNOMED CT | 289887006 | Finding of frequency of menstruation |
| SNOMED CT | 289890000 | Finding of duration of menstruation |
| SNOMED CT | 289891001 | Finding of duration of menstrual cycle |
| SNOMED CT | 289892008 | Short menstrual cycle |
| SNOMED CT | 289893003 | Finding of duration of menstrual period |
| SNOMED CT | 302757007 | Regular periods |
| SNOMED CT | 314631008 | Menometrorrhagia |
| SNOMED CT | 351814001 | Intermenstrual heavy bleeding |
| SNOMED CT | 386704007 | Light and infrequent menstruation |
| SNOMED CT | 413487000 | Amenorrhea associated with obesity |
| SNOMED CT | 444560000 | Oligoovulatory dysfunctional uterine bleeding |
| SNOMED CT | 444769001 | Anovulatory amenorrhea |
| SNOMED CT | 702466002 | Normogonadotropic amenorrhea |
| SNOMED CT | 78456001 | Normal menstrual cycle |

Continued on next page

**TableS6 – continued from previous page**

| CodeSystem | ConceptCode | ConceptName |
|---|---|---|
| SNOMED CT | 80182007 | Irregular periods |
| SNOMED CT | 84292000 | Menstrual period late |
| SNOMED CT | 102868004 | Intermenstrual spotting due to intrauterine device |
| SNOMED CT | 156035004 | Primary amenorrhea |
| SNOMED CT | 156036003 | Secondary amenorrhea |
| SNOMED CT | 161718009 | Menstruation postponed |
| SNOMED CT | 161724003 | Length of cycle stable |
| SNOMED CT | 161725002 | Length of cycle increasing |
| SNOMED CT | 161726001 | Length of cycle decreasing |
| SNOMED CT | 237119008 | Exercise-related amenorrhea |
| SNOMED CT | 237122005 | Lactational amenorrhea |
| SNOMED CT | 237128009 | Metropathia hemorrhagica |
| SNOMED CT | 237130006 | Mid-cycle bleeding |
| SNOMED CT | 237132003 | Intermenstrual bleeding - regular |
| SNOMED CT | 248962008 | Period has come early |
| SNOMED CT | 248963003 | Long menstrual cycle |
| SNOMED CT | 248965005 | Finding of regularity of menstrual cycle |
| SNOMED CT | 248966006 | Prolonged periods |
| SNOMED CT | 248967002 | Short menstrual periods |
| SNOMED CT | 248969004 | Spotting precedes menstrual flow |
| SNOMED CT | 8913004 | Primary physiologic amenorrhea |
| SNOMED CT | 14302001 | Amenorrhea |
| SNOMED CT | 14884007 | Primary oligomenorrhea |
| SNOMED CT | 21669001 | Dysponderal amenorrhea |
| SNOMED CT | 27585009 | Anovular menstruation |
| SNOMED CT | 32590007 | Premenopausal amenorrhea |
| SNOMED CT | 34272008 | Secondary oligomenorrhea |
| SNOMED CT | 36521000 | Missed period |
| SNOMED CT | 39931000 | Postpill amenorrhea |
| SNOMED CT | 42131003 | Abnormal menstrual cycle |
| SNOMED CT | 44811000 | Physiologic amenorrhea |
| SNOMED CT | 52073004 | Oligomenorrhea |
| SNOMED CT | 52754008 | Polymenorrhea |
| SNOMED CT | 56169002 | Break-through bleeding |
| SNOMED CT | 56561006 | Pathologic amenorrhea |
| SNOMED CT | 57260004 | Dietary amenorrhea |
| SNOMED CT | 58851004 | Cryptomenorrhea |
| SNOMED CT | 64996003 | Intermenstrual bleeding - irregular |
| SNOMED CT | 788919002 | Excessive menstruation with irregular cycle |
| SNOMED CT | 455771000124105 | Normal menarche |
| SNOMED CT | 86030004 | Loss of menstrual period |
| SNOMED CT | 1142058003 | Amenorrhea due to obstetric fistula |
| SNOMED CT | 1144947008 | Amenorrhea due to congenital gonadotrophin releasing hormone deficiency |
| SNOMED CT | 1144987003 | Amenorrhea due to pituitary dysfunction |
| SNOMED CT | 1144991008 | Amenorrhea due to disorder of ovary |
| SNOMED CT | 1144993006 | Amenorrhea due to hypothalamic disorder |

Continued on next page

**TableS6 – continued from previous page**

| CodeSystem | ConceptCode | ConceptName |
|---|---|---|
| SNOMED CT | 1217062009 | Hyperhormonal amenorrhea |

#### 4.4 Arthralgia

Table S7: Concept codes used to identify if a patient was diagnosed with arthralgia.

| CodeSystem | ConceptCode | ConceptName |
|---|---|---|
| ICD10CM | M25.5 | Pain in joint |
| ICD10CM | M25.9 | Joint disorder, unspecified |
| ICD10CM | M25.50 | Pain in unspecified joint |
| ICD10CM | M25.51 | Pain in shoulder |
| ICD10CM | M25.511 | Pain in right shoulder |
| ICD10CM | M25.512 | Pain in left shoulder |
| ICD10CM | M25.519 | Pain in unspecified shoulder |
| ICD10CM | M25.52 | Pain in elbow |
| ICD10CM | M25.521 | Pain in right elbow |
| ICD10CM | M25.522 | Pain in left elbow |
| ICD10CM | M25.529 | Pain in unspecified elbow |
| ICD10CM | M25.53 | Pain in wrist |
| ICD10CM | M25.531 | Pain in right wrist |
| ICD10CM | M25.532 | Pain in left wrist |
| ICD10CM | M25.539 | Pain in unspecified wrist |
| ICD10CM | M25.54 | Pain in joints of hand |
| ICD10CM | M25.541 | Pain in joints of right hand |
| ICD10CM | M25.542 | Pain in joints of left hand |
| ICD10CM | M25.549 | Pain in joints of unspecified hand |
| ICD10CM | M25.55 | Pain in hip |
| ICD10CM | M25.551 | Pain in right hip |
| ICD10CM | M25.552 | Pain in left hip |
| ICD10CM | M25.559 | Pain in unspecified hip |
| ICD10CM | M25.56 | Pain in knee |
| ICD10CM | M25.561 | Pain in right knee |
| ICD10CM | M25.562 | Pain in left knee |
| ICD10CM | M25.569 | Pain in unspecified knee |
| ICD10CM | M25.57 | Pain in ankle and joints of foot |
| ICD10CM | M25.571 | Pain in right ankle and joints of right foot |
| ICD10CM | M25.572 | Pain in left ankle and joints of left foot |
| ICD10CM | M25.579 | Pain in unspecified ankle and joints of unspecified foot |
| ICD10CM | M25.59 | Pain in other specified joint |
| SNOMED CT | 57676002 | Joint pain |
| SNOMED CT | 267949000 | Shoulder joint pain |
| SNOMED CT | 267950000 | Arthralgia of the upper arm |
| SNOMED CT | 267952008 | Arthralgia of the pelvic region and thigh |
| SNOMED CT | 267954009 | Arthralgia of the ankle and/or foot |

Continued on next page

**TableS7 – continued from previous page**

| CodeSystem | ConceptCode | ConceptName |
|---|---|---|
| SNOMED CT | 277138006 | Rubella arthralgia |
| SNOMED CT | 279063004 | Lumbar facet joint pain |
| SNOMED CT | 279066007 | Foot joint pain |
| SNOMED CT | 279067003 | Metatarsophalangeal joint pain |
| SNOMED CT | 298253002 | Cervical facet joint pain |
| SNOMED CT | 298254008 | Thoracic facet joint pain |
| SNOMED CT | 298255009 | Pain on joint movement |
| SNOMED CT | 298376001 | Temporomandibular joint painful on movement |
| SNOMED CT | 298478001 | Pain on movement of cervical spine |
| SNOMED CT | 298579007 | Pain on movement of thoracic spine |
| SNOMED CT | 298674008 | Pain on movement of lumbar spine |
| SNOMED CT | 298857005 | Shoulder joint painful on movement |
| SNOMED CT | 298858000 | Shoulder joint - painful arc |
| SNOMED CT | 298929004 | Elbow joint - painful on movement |
| SNOMED CT | 299018007 | Wrist joint painful on movement |
| SNOMED CT | 299112005 | Finger joint painful on movement |
| SNOMED CT | 299199000 | Thumb joint painful on movement |
| SNOMED CT | 299308007 | Hip joint painful on movement |
| SNOMED CT | 299377003 | Knee joint painful on movement |
| SNOMED CT | 299447008 | Ankle joint - painful on movement |
| SNOMED CT | 299513007 | Foot joint - painful on movement |
| SNOMED CT | 299554004 | Subtalar joint painful on movement |
| SNOMED CT | 299633000 | Toe joint painful on movement |
| SNOMED CT | 309765009 | Pain due to hip joint prosthesis |
| SNOMED CT | 309767001 | Pain due to knee joint prosthesis |
| SNOMED CT | 309768006 | Pain due to shoulder joint prosthesis |
| SNOMED CT | 387638003 | Painful swelling of joint |
| SNOMED CT | 429531000 | Pain on passive stretch of joint |
| SNOMED CT | 433017009 | Pain in symphysis pubis in pregnancy |
| SNOMED CT | 703619001 | Pelvic girdle pain |
| SNOMED CT | 713413001 | Joint pain of pelvic region |
| SNOMED CT | 91943004 | Arthralgia of temporomandibular joint |
| SNOMED CT | 164539000 | O/E - joint movement painful |
| SNOMED CT | 202472008 | Hand joint pain |
| SNOMED CT | 202478007 | Sternoclavicular joint pain |
| SNOMED CT | 202479004 | Acromioclavicular joint pain |
| SNOMED CT | 202480001 | Elbow joint pain |
| SNOMED CT | 202481002 | Distal radioulnar joint pain |
| SNOMED CT | 202482009 | Wrist joint pain |
| SNOMED CT | 202483004 | Metacarpophalangeal joint pain |
| SNOMED CT | 202484005 | Proximal interphalangeal joint of finger pain |
| SNOMED CT | 202485006 | Distal interphalangeal joint of finger pain |
| SNOMED CT | 202487003 | Sacroiliac joint pain |
| SNOMED CT | 202489000 | Tibiofibular joint pain |
| SNOMED CT | 202490009 | Ankle joint pain |
| SNOMED CT | 202491008 | Subtalar joint pain |
| SNOMED CT | 202493006 | Talonavicular joint pain |

Continued on next page

**TableS7 – continued from previous page**

| CodeSystem | ConceptCode | ConceptName |
|---|---|---|
| SNOMED CT | 202495004 | First metatarsophalangeal joint pain |
| SNOMED CT | 202496003 | Lesser metatarsophalangeal joint pain |
| SNOMED CT | 202497007 | Interphalangeal joint of toe pain |
| SNOMED CT | 239733006 | Anterior knee pain |
| SNOMED CT | 240271006 | Persistent prosthetic joint pain |
| SNOMED CT | 247369005 | Facet joint pain |
| SNOMED CT | 30989003 | Knee pain |
| SNOMED CT | 35678005 | Multiple joint pain |
| SNOMED CT | 46960006 | Lumbago-sciatica due to displacement of lumbar intervertebral disc |
| SNOMED CT | 49218002 | Hip pain |
| SNOMED CT | 736428003 | Chronic arthralgia of temporomandibular joint |
| SNOMED CT | 736447000 | Infrequent episodic arthralgia of temporomandibular joint |
| SNOMED CT | 772826002 | Anti-cyclic citrullinated peptide antibody positive arthralgia |
| SNOMED CT | 774136009 | Pain of right acromioclavicular joint |
| SNOMED CT | 774137000 | Pain of left acromioclavicular joint |
| SNOMED CT | 782661001 | Chronic sacroiliac joint pain |
| SNOMED CT | 87381000119101 | Vertebral joint pain |
| SNOMED CT | 112581000119104 | Acute pain of joint of knee |
| SNOMED CT | 316781000119100 | Pain of left hip joint |
| SNOMED CT | 316791000119102 | Pain in left knee |
| SNOMED CT | 316921000119102 | Pain in right hip joint |
| SNOMED CT | 316931000119104 | Pain in right knee |
| SNOMED CT | 448701000124105 | Herniation of lumbar intervertebral disc with sciatica |
| SNOMED CT | 1075331000119103 | Patellofemoral syndrome of right knee |
| SNOMED CT | 1075341000119107 | Patellofemoral syndrome of left knee |
| SNOMED CT | 1075631000119107 | Pain of right sternoclavicular joint |
| SNOMED CT | 1075651000119101 | Pain of left sternoclavicular joint |
| SNOMED CT | 1076781000119107 | Pain of joint of right foot |
| SNOMED CT | 1076801000119106 | Pain of joint of left foot |
| SNOMED CT | 11874791000119104 | Bilateral knee pain |
| SNOMED CT | 12240271000119107 | Pain of left temporomandibular joint |
| SNOMED CT | 12240311000119107 | Pain of right temporomandibular joint |
| SNOMED CT | 12242551000119102 | Pain in left sacroiliac joint |
| SNOMED CT | 12242591000119107 | Pain in right sacroiliac joint |
| SNOMED CT | 12247221000119105 | Patellofemoral syndrome of bilateral knees |
| SNOMED CT | 12247571000119109 | Bilateral foot joint pain |
| SNOMED CT | 12247611000119100 | Bilateral hip joint pain |
| SNOMED CT | 12247731000119102 | Bilateral ankle joint pain |
| SNOMED CT | 12247771000119104 | Bilateral shoulder joint pain |
| SNOMED CT | 12247851000119103 | Bilateral elbow joint pain |
| SNOMED CT | 12400141000119103 | Bilateral chronic pain following total hip arthroplasty |
| SNOMED CT | 12400221000119100 | Chronic pain following left total hip arthroplasty |
| SNOMED CT | 12400261000119105 | Chronic pain following left total knee arthroplasty |
| SNOMED CT | 12400301000119102 | Chronic pain following right total hip arthroplasty |
| SNOMED CT | 12400341000119100 | Chronic pain following right total knee arthroplasty |
| SNOMED CT | 15634631000119104 | Bilateral sacroiliac joint pain |

Continued on next page

**TableS7 – continued from previous page**

| CodeSystem | ConceptCode | ConceptName |
|---|---|---|
| SNOMED CT | 15639561000119107 | Bilateral acromioclavicular joint pain |
| SNOMED CT | 15639721000119105 | Bilateral temporomandibular joint pain |
| SNOMED CT | 15743161000119106 | Pain of joint of bilateral lower legs |
| SNOMED CT | 15743201000119101 | Pain of joint of right lower leg |
| SNOMED CT | 15743241000119104 | Pain of joint of left lower leg |
| SNOMED CT | 15743641000119101 | Pain of bilateral sternoclavicular joints |
| SNOMED CT | 15916891000119103 | Pain of left ankle joint |
| SNOMED CT | 15916931000119106 | Pain of right ankle joint |
| SNOMED CT | 15916971000119109 | Pain of left shoulder joint |
| SNOMED CT | 15917011000119100 | Pain of right shoulder joint |
| SNOMED CT | 15917051000119104 | Pain of left elbow joint |
| SNOMED CT | 15917091000119109 | Pain of right elbow joint |
| SNOMED CT | 16899701000119102 | Bilateral pain of joint of hands |
| SNOMED CT | 1076721000119108 | Joint pain in right hand |
| SNOMED CT | 1076731000119106 | Joint pain in left hand |
| SNOMED CT | 12400181000119108 | Chronic pain following bilateral total knee arthroplasty |
| SNOMED CT | 1003721002 | Pain of joint of knee |
| SNOMED CT | 468231000124100 | Pain of right knee joint |
| SNOMED CT | 468251000124107 | Pain of left knee joint |
| SNOMED CT | 468281000124104 | Pain of bilateral knee joints |
| Truveta | 1215962 | Post-COVID-19 chronic joint pain |

#### 4.5 Chest pain

Table S8: Concept codes used to identify if a patient was diagnosed with chest pain.

| CodeSystem | ConceptCode | ConceptName |
|---|---|---|
| ICD10CM | R07 | Pain in throat and chest |
| ICD10CM | R07.0 | Pain in throat |
| ICD10CM | R07.1 | Chest pain on breathing |
| ICD10CM | R07.2 | Precordial pain |
| ICD10CM | R07.8 | Other chest pain |
| ICD10CM | R07.81 | Pleurodynia |
| ICD10CM | R07.82 | Intercostal pain |
| ICD10CM | R07.89 | Other chest pain |
| ICD10CM | R07.9 | Chest pain, unspecified |
| SNOMED CT | 29857009 | Chest pain |
| SNOMED CT | 267981009 | Pain in thoracic spine |
| SNOMED CT | 274664007 | Chest pain on breathing |
| SNOMED CT | 274668005 | Non-cardiac chest pain |
| SNOMED CT | 279019008 | Central crushing chest pain |
| SNOMED CT | 279035001 | Acute thoracic back pain |
| SNOMED CT | 279036000 | Myofascial pain syndrome of thorax |
| SNOMED CT | 279037009 | Thoracic segmental dysfunction |

Continued on next page

**TableS8 – continued from previous page**

| CodeSystem | ConceptCode | ConceptName |
|---|---|---|
| SNOMED CT | 279038004 | Thoracic back pain |
| SNOMED CT | 279048002 | Internal mammary artery syndrome |
| SNOMED CT | 279054001 | Pain when milk coming in |
| SNOMED CT | 281245003 | Musculoskeletal chest pain |
| SNOMED CT | 285385002 | Left sided chest pain |
| SNOMED CT | 285386001 | Right sided chest pain |
| SNOMED CT | 285389008 | Upper chest pain |
| SNOMED CT | 297217002 | Rib pain |
| SNOMED CT | 298254008 | Thoracic facet joint pain |
| SNOMED CT | 298579007 | Pain on movement of thoracic spine |
| SNOMED CT | 298731003 | Pain of sternum |
| SNOMED CT | 300995000 | Exercise-induced angina |
| SNOMED CT | 314116003 | Post infarct angina |
| SNOMED CT | 315025001 | Refractory angina |
| SNOMED CT | 315248002 | Intractable breast pain |
| SNOMED CT | 315250005 | Persistent mastalgia |
| SNOMED CT | 371030007 | Squeezing chest pain |
| SNOMED CT | 371806006 | Progressive angina |
| SNOMED CT | 371807002 | Atypical angina |
| SNOMED CT | 371808007 | Recurrent angina after percutaneous transluminal coronary angioplasty |
| SNOMED CT | 371809004 | Recurrent angina following placement of coronary artery stent |
| SNOMED CT | 371810009 | Recurrent angina after coronary artery bypass graft |
| SNOMED CT | 371811008 | Recurrent angina post rotational atherectomy |
| SNOMED CT | 371812001 | Recurrent angina after directional coronary atherectomy |
| SNOMED CT | 425991004 | Cervicothoracic segmental dysfunction |
| SNOMED CT | 426396005 | Cardiac chest pain |
| SNOMED CT | 429559004 | Typical angina |
| SNOMED CT | 444227004 | Acute postthoracotomy pain syndrome |
| SNOMED CT | 713829001 | Postinfective intercostal neuralgia |
| SNOMED CT | 713839007 | Intercostal neuralgia as late effect of trauma |
| SNOMED CT | 71884009 | Precordial pain |
| SNOMED CT | 81953000 | Chest pain on exertion |
| SNOMED CT | 83264000 | Epidemic pleurodynia |
| SNOMED CT | 85284003 | Angina, class III |
| SNOMED CT | 87343002 | Prinzmetal angina |
| SNOMED CT | 89323001 | Angina, class IV |
| SNOMED CT | 89638008 | Xiphodynia |
| SNOMED CT | 89874002 | Xiphoidalgia syndrome |
| SNOMED CT | 95421005 | Intercostal myalgia |
| SNOMED CT | 102587001 | Acute chest pain |
| SNOMED CT | 102588006 | Chest wall pain |
| SNOMED CT | 102589003 | Atypical chest pain |
| SNOMED CT | 103015000 | Thoracic nerve root pain |
| SNOMED CT | 135876005 | Mastalgia of puberty |
| SNOMED CT | 161972006 | Central chest pain |
| SNOMED CT | 161973001 | Anterior chest wall pain |

Continued on next page

**TableS8 – continued from previous page**

| CodeSystem | ConceptCode | ConceptName |
|---|---|---|
| SNOMED CT | 161974007 | Parasternal pain |
| SNOMED CT | 161977000 | Costal margin chest pain |
| SNOMED CT | 194823009 | Acute coronary insufficiency |
| SNOMED CT | 194828000 | Angina pectoris |
| SNOMED CT | 200430001 | Breastfeeding painful |
| SNOMED CT | 202478007 | Sternoclavicular joint pain |
| SNOMED CT | 225566008 | Ischemic chest pain |
| SNOMED CT | 230597003 | Intercostal post-herpetic neuralgia |
| SNOMED CT | 230648001 | Abdominal cutaneous nerve entrapment syndrome |
| SNOMED CT | 233819005 | Stable angina |
| SNOMED CT | 233821000 | New onset angina |
| SNOMED CT | 233845001 | Cardiac syndrome X |
| SNOMED CT | 237453001 | Cyclical mastalgia |
| SNOMED CT | 237454007 | Non-cyclical mastalgia |
| SNOMED CT | 237455008 | Trigger point mastalgia |
| SNOMED CT | 237477007 | Pain caused by breast implant |
| SNOMED CT | 239175003 | Post-thoracotomy pain syndrome |
| SNOMED CT | 247350000 | Sore nipple |
| SNOMED CT | 247389006 | Intercostal neuralgia |
| SNOMED CT | 247415009 | Painful lactation |
| SNOMED CT | 2237002 | Pleuritic pain |
| SNOMED CT | 3368006 | Dull chest pain |
| SNOMED CT | 4557003 | Preinfarction syndrome |
| SNOMED CT | 4568003 | Retrosternal pain |
| SNOMED CT | 9267009 | Chest pain at rest |
| SNOMED CT | 10000006 | Radiating chest pain |
| SNOMED CT | 13057000 | Pleuropericardial chest pain |
| SNOMED CT | 19057007 | Status anginosus |
| SNOMED CT | 20793008 | Scapulalgia |
| SNOMED CT | 21470009 | Syncope anginosa |
| SNOMED CT | 35928006 | Nocturnal angina |
| SNOMED CT | 36859004 | Esophageal chest pain |
| SNOMED CT | 41334000 | Angina, class II |
| SNOMED CT | 53430007 | Pain of breast |
| SNOMED CT | 59021001 | Angina decubitus |
| SNOMED CT | 59139008 | Crushing chest pain |
| SNOMED CT | 61490001 | Angina, class I |
| SNOMED CT | 735940001 | Pain of intercostal space |
| SNOMED CT | 866043000 | Neuralgia of twelfth thoracic nerve |
| SNOMED CT | 7921000119104 | Anterior pleuritic pain |
| SNOMED CT | 34791000119103 | Chest pain due to pericarditis |
| SNOMED CT | 41321000119101 | Myofascial pain syndrome of thoracic spine |
| SNOMED CT | 97001000119106 | Localized chest pain |
| SNOMED CT | 98611000119104 | Chronic post-thoracotomy pain syndrome |
| SNOMED CT | 136791000119103 | Chronic thoracic back pain |
| SNOMED CT | 434011000124101 | Chronic chest pain |
| SNOMED CT | 1075631000119107 | Pain of right sternoclavicular joint |

Continued on next page

**TableS8 – continued from previous page**

| CodeSystem | ConceptCode | ConceptName |
|---|---|---|
| SNOMED CT | 1075651000119101 | Pain of left sternoclavicular joint |
| SNOMED CT | 12242431000119109 | Mastodynia of bilateral breasts |
| SNOMED CT | 12242471000119107 | Mastodynia of left breast |
| SNOMED CT | 12242511000119103 | Mastodynia of right breast |
| SNOMED CT | 15743641000119101 | Pain of bilateral sternoclavicular joints |
| SNOMED CT | 15743681000119106 | Bilateral pain of shoulder blades |
| SNOMED CT | 15960061000119102 | Unstable angina co-occurrent and due to coronary arteriosclerosis |
| SNOMED CT | 15960141000119102 | Angina co-occurrent and due to coronary arteriosclerosis |
| SNOMED CT | 15960381000119109 | Angina co-occurrent and due to arteriosclerosis of coronary artery bypass graft |
| SNOMED CT | 15960541000119107 | Unstable angina due to arteriosclerosis of autologous vein coronary artery bypass graft |
| SNOMED CT | 15960581000119102 | Arteriosclerosis of autologous vein coronary artery bypass graft with angina |
| SNOMED CT | 15960661000119107 | Unstable angina co-occurrent and due to arteriosclerosis of coronary artery bypass graft |
| SNOMED CT | 15960781000119108 | Arteriosclerosis of autologous arterial coronary artery bypass graft with angina |
| SNOMED CT | 16754391000119100 | Stable angina due to coronary arteriosclerosis |
| SNOMED CT | 774134007 | Right scapulalgia |
| SNOMED CT | 774135008 | Left scapulalgia |
| SNOMED CT | 791000119109 | Angina due to type 2 diabetes mellitus |
| SNOMED CT | 1010235008 | Mastodynia of left breast |
| SNOMED CT | 1010237000 | Mastodynia of right breast |
| SNOMED CT | 15960341000119104 | Unstable angina due to arteriosclerosis of coronary artery bypass graft of transplanted heart |
| SNOMED CT | 15960461000119105 | Unstable angina due to arteriosclerosis of autologous arterial coronary artery bypass graft |

#### 4.6 Cognitive impairment

Table S9: Concept codes used to identify if a patient was diagnosed with cognitive impairement.

| CodeSystem | ConceptCode | ConceptName |
|---|---|---|
| ICD10CM | F09 | Unspecified mental disorder due to known physiological condition |
| ICD10CM | G31.84 | Mild cognitive impairment, so stated |
| ICD10CM | R41.89 | Other symptoms and signs involving cognitive functions and awareness |
| ICD10CM | R41.9 | Unspecified symptoms and signs involving cognitive functions and awareness |
| SNOMED CT | 386806002 | Impaired cognition |
| SNOMED CT | 443265004 | Cognitive disorder |

Continued on next page

**TableS9 – continued from previous page**

| CodeSystem | ConceptCode | ConceptName |
|---|---|---|
| SNOMED CT | 268612007 | Senile and presenile organic psychotic conditions |
| SNOMED CT | 275277000 | Post-traumatic amnesia |
| SNOMED CT | 278857002 | Dementia of frontal lobe type |
| SNOMED CT | 283878007 | Impairment of registration |
| SNOMED CT | 307413004 | Transient memory loss |
| SNOMED CT | 371024007 | Senile dementia with delusion |
| SNOMED CT | 371026009 | Senile dementia with psychosis |
| SNOMED CT | 386805003 | Mild cognitive disorder |
| SNOMED CT | 386807006 | Memory impairment |
| SNOMED CT | 395689002 | Transient epileptic amnesia |
| SNOMED CT | 416106008 | Disturbance of memory for order of events |
| SNOMED CT | 425805004 | Cognitive developmental delay |
| SNOMED CT | 429458009 | Dementia due to Creutzfeldt Jakob disease |
| SNOMED CT | 429998004 | Vascular dementia |
| SNOMED CT | 433081000 | Organic amnesia of language |
| SNOMED CT | 442212003 | Residual cognitive deficit as late effect of cerebrovascular accident |
| SNOMED CT | 698624003 | Dementia associated with cerebral lipidosis |
| SNOMED CT | 698625002 | Dementia associated with normal pressure hydrocephalus |
| SNOMED CT | 698626001 | Dementia associated with multiple sclerosis |
| SNOMED CT | 698687007 | Post-traumatic dementia with behavioral change |
| SNOMED CT | 698691002 | Cognitive disorder in remission |
| SNOMED CT | 698725008 | Dementia associated with neurosyphilis |
| SNOMED CT | 698726009 | Dementia associated with viral encephalitis |
| SNOMED CT | 698781002 | Dementia associated with cerebral anoxia |
| SNOMED CT | 698948009 | Vascular dementia in remission |
| SNOMED CT | 698949001 | Dementia in remission |
| SNOMED CT | 698954005 | Primary degenerative dementia of the Alzheimer type, senile onset in remission |
| SNOMED CT | 698955006 | Primary degenerative dementia of the Alzheimer type, presenile onset in remission |
| SNOMED CT | 702955000 | Moderate cognitive impairment |
| SNOMED CT | 702956004 | Severe cognitive impairment |
| SNOMED CT | 709073001 | Neurocognitive disorder |
| SNOMED CT | 713488003 | Presenile dementia co-occurrent with human immunodeficiency virus infection |
| SNOMED CT | 713844000 | Dementia co-occurrent with human immunodeficiency virus infection |
| SNOMED CT | 715737004 | Parkinsonism with dementia of Guadeloupe |
| SNOMED CT | 716306006 | Acquired language comprehension impairment |
| SNOMED CT | 70936005 | Multi-infarct dementia, uncomplicated |
| SNOMED CT | 76789006 | Information conversion problem |
| SNOMED CT | 82959004 | Dementia paralytica juvenilis |
| SNOMED CT | 88822006 | Anterograde amnesia |
| SNOMED CT | 90099008 | Subcortical leukoencephalopathy |
| SNOMED CT | 102891000 | Age-related cognitive decline |
| SNOMED CT | 110352000 | Minimal cognitive impairment |

Continued on next page

**TableS9 – continued from previous page**

| CodeSystem | ConceptCode | ConceptName |
|---|---|---|
| SNOMED CT | 110355003 | Lack of thinking ability |
| SNOMED CT | 111480006 | Psychoactive substance-induced organic dementia |
| SNOMED CT | 116340000 | Disturbance of cognitive learning |
| SNOMED CT | 130964008 | Impaired environmental interpretation syndrome |
| SNOMED CT | 162200009 | Temporary loss of memory |
| SNOMED CT | 191449005 | Uncomplicated senile dementia |
| SNOMED CT | 191451009 | Uncomplicated presenile dementia |
| SNOMED CT | 191452002 | Presenile dementia with delirium |
| SNOMED CT | 191454001 | Presenile dementia with paranoia |
| SNOMED CT | 191455000 | Presenile dementia with depression |
| SNOMED CT | 191457008 | Senile dementia with depressive or paranoid features |
| SNOMED CT | 191458003 | Senile dementia with paranoia |
| SNOMED CT | 191459006 | Senile dementia with depression |
| SNOMED CT | 191461002 | Senile dementia with delirium |
| SNOMED CT | 191463004 | Uncomplicated arteriosclerotic dementia |
| SNOMED CT | 191464005 | Arteriosclerotic dementia with delirium |
| SNOMED CT | 191465006 | Arteriosclerotic dementia with paranoia |
| SNOMED CT | 191466007 | Arteriosclerotic dementia with depression |
| SNOMED CT | 191493005 | Drug-induced dementia |
| SNOMED CT | 191519005 | Dementia associated with another disease |
| SNOMED CT | 192071009 | Mild memory disturbance |
| SNOMED CT | 225037001 | Minor memory lapses |
| SNOMED CT | 225038006 | Memory lapses |
| SNOMED CT | 225039003 | Mixes past with present |
| SNOMED CT | 229676007 | Language-related cognitive disorder |
| SNOMED CT | 230282000 | Traumatic encephalopathy |
| SNOMED CT | 230285003 | Vascular dementia of acute onset |
| SNOMED CT | 230286002 | Subcortical vascular dementia |
| SNOMED CT | 230287006 | Mixed cortical and subcortical vascular dementia |
| SNOMED CT | 230289009 | Patchy dementia |
| SNOMED CT | 230736007 | Transient global amnesia |
| SNOMED CT | 247607004 | Amnesia for day to day facts |
| SNOMED CT | 247611005 | Amnesia for important personal information |
| SNOMED CT | 281004 | Dementia associated with alcoholism |
| SNOMED CT | 6149008 | Amnesia for remote events |
| SNOMED CT | 9345005 | Dialysis dementia |
| SNOMED CT | 10349009 | Multi-infarct dementia with delirium |
| SNOMED CT | 12348006 | Presenile dementia |
| SNOMED CT | 14070001 | Multi-infarct dementia with depression |
| SNOMED CT | 15662003 | Senile dementia |
| SNOMED CT | 25772007 | Multi-infarct dementia with delusions |
| SNOMED CT | 26581009 | Retrospective falsification |
| SNOMED CT | 32541007 | Paramnesia |
| SNOMED CT | 32875003 | Inhalant-induced persisting dementia |
| SNOMED CT | 40425004 | Postconcussion syndrome |
| SNOMED CT | 42176003 | Amnesia for recent events |
| SNOMED CT | 48167000 | LOM - loss of memory |

Continued on next page

**TableS9 – continued from previous page**

| CodeSystem | ConceptCode | ConceptName |
|---|---|---|
| SNOMED CT | 51921000 | Retrograde amnesia |
| SNOMED CT | 51928006 | General paresis - neurosyphilis |
| SNOMED CT | 52448006 | Dementia |
| SNOMED CT | 54502004 | Primary degenerative dementia of the Alzheimer type, presenile onset, with delusions |
| SNOMED CT | 55533009 | Forgetful |
| SNOMED CT | 56267009 | Multi-infarct dementia |
| SNOMED CT | 59651006 | Sedative, hypnotic AND/OR anxiolytic-induced persisting dementia |
| SNOMED CT | 65096006 | Primary degenerative dementia of the Alzheimer type, presenile onset, with delirium |
| SNOMED CT | 722600006 | Non-amnestic Alzheimer disease |
| SNOMED CT | 722977005 | Dementia co-occurrent and due to neurocysticercosis |
| SNOMED CT | 722978000 | Dementia caused by toxin |
| SNOMED CT | 722979008 | Dementia due to metabolic abnormality |
| SNOMED CT | 722980006 | Dementia due to chromosomal anomaly |
| SNOMED CT | 723123001 | Ischemic vascular dementia |
| SNOMED CT | 723390000 | Rapidly progressive dementia |
| SNOMED CT | 724760003 | Cognitive impairment co-occurrent and due to primary psychotic disorder |
| SNOMED CT | 724776007 | Dementia due to disorder of central nervous system |
| SNOMED CT | 724777003 | Dementia due to infectious disease |
| SNOMED CT | 724992007 | Epilepsy co-occurrent and due to dementia |
| SNOMED CT | 725898002 | Delirium co-occurrent with dementia |
| SNOMED CT | 733184002 | Dementia caused by heavy metal exposure |
| SNOMED CT | 733185001 | Dementia following injury caused by exposure to ionizing radiation |
| SNOMED CT | 733190003 | Dementia due to primary malignant neoplasm of brain |
| SNOMED CT | 733191004 | Dementia due to chronic subdural hematoma |
| SNOMED CT | 733192006 | Dementia due to herpes encephalitis |
| SNOMED CT | 733194007 | Dementia with Down syndrome |
| SNOMED CT | 736314008 | Dissociative neurological symptom disorder co-occurrent with cognitive symptoms |
| SNOMED CT | 736317001 | Impaired executive functioning |
| SNOMED CT | 762298000 | Cognitive impairment due to toxicity of substance |
| SNOMED CT | 762350007 | Dementia due to prion disease |
| SNOMED CT | 762351006 | Dementia due to and following injury of head |
| SNOMED CT | 762707000 | Subcortical dementia |
| SNOMED CT | 771417007 | Postoperative cognitive dysfunction |
| SNOMED CT | 774069007 | PRKAR1B-related neurodegenerative dementia with intermediate filaments |
| SNOMED CT | 783161005 | ABri amyloidosis |
| SNOMED CT | 783258000 | ADan amyloidosis |
| SNOMED CT | 788898005 | Dementia caused by volatile inhalant |
| SNOMED CT | 788899002 | Dementia due to pellagra |
| SNOMED CT | 789170003 | Disinhibited behavior due to dementia |
| SNOMED CT | 833326008 | Cortical vascular dementia |

Continued on next page

**TableS9 – continued from previous page**

| CodeSystem | ConceptCode | ConceptName |
|---|---|---|
| SNOMED CT | 836301008 | Amnestic mild cognitive disorder |
| SNOMED CT | 838276009 | Amyotrophic lateral sclerosis, parkinsonism, dementia complex |
| SNOMED CT | 840449007 | Cognitive impairment caused by ingestible alcohol |
| SNOMED CT | 840464007 | Dementia due to carbon monoxide poisoning |
| SNOMED CT | 840465008 | Dementia due to iron deficiency |
| SNOMED CT | 870262000 | Pervasive developmental disorder with disorder of intellectual development without loss of previously acquired skills |
| SNOMED CT | 870264004 | Pervasive developmental disorder with disorder of intellectual development and pervasive impairment of functional language without loss of previously acquired skills |
| SNOMED CT | 870265003 | Pervasive developmental disorder with disorder of intellectual development with loss of previously acquired skills |
| SNOMED CT | 870266002 | Pervasive developmental disorder with disorder of intellectual development and marked impairment of functional language with loss of previously acquired skills |
| SNOMED CT | 870267006 | Pervasive developmental disorder with disorder of intellectual development and marked impairment of functional language without loss of previously acquired skills |
| SNOMED CT | 870268001 | Pervasive developmental disorder with disorder of intellectual development and complete impairment of functional language without loss of previously acquired skills |
| SNOMED CT | 870269009 | Pervasive developmental disorder with disorder of intellectual development and absence of functional language with loss of previously acquired skills |
| SNOMED CT | 870270005 | Pervasive developmental disorder with disorder of intellectual development and complete impairment of functional language with loss of previously acquired skills |
| SNOMED CT | 870305003 | Pervasive developmental disorder with cognitive developmental delay and marked impairment of functional language |
| SNOMED CT | 870308001 | Pervasive developmental disorder with cognitive developmental delay and complete impairment of functional language |
| SNOMED CT | 1581000119101 | Dementia of the Alzheimer type with behavioral disturbance |
| SNOMED CT | 2421000119107 | Hallucinations co-occurrent and due to late onset dementia |
| SNOMED CT | 18541000119100 | Borderline cognitive developmental delay |
| SNOMED CT | 22381000119105 | Primary degenerative dementia |
| SNOMED CT | 31081000119101 | Presenile dementia with delusions |
| SNOMED CT | 79341000119107 | Mixed dementia |
| SNOMED CT | 82371000119101 | Dementia due to multiple sclerosis with altered behavior |
| SNOMED CT | 97751000119108 | Altered behavior in Alzheimer's disease |
| SNOMED CT | 101421000119107 | Dementia due to Parkinson's disease |
| SNOMED CT | 105421000119105 | Early onset Alzheimer's disease with behavioral disturbance |
| SNOMED CT | 106021000119105 | Multi-infarct dementia due to atherosclerosis |
| SNOMED CT | 130121000119104 | Dementia due to Rett's syndrome |
| SNOMED CT | 141601000119107 | Cognitive changes due to organic disorder |
| SNOMED CT | 141991000119109 | Delusions in Alzheimer's disease |
| SNOMED CT | 142001000119106 | Depressed mood in Alzheimer's disease |
| SNOMED CT | 142011000119109 | Alzheimer's disease co-occurrent with delirium |

Continued on next page

**TableS9 – continued from previous page**

| CodeSystem | ConceptCode | ConceptName |
|---|---|---|
| SNOMED CT | 142261000119100 | Cognitive deficit in attention |
| SNOMED CT | 288631000119104 | Vascular dementia with behavioral disturbance |
| SNOMED CT | 290621000119101 | Cognitive deficit due to and following cerebrovascular disease |
| SNOMED CT | 291621000119109 | Cognitive deficit due to and following nontraumatic subarachnoid hemorrhage |
| SNOMED CT | 291711000119109 | Cognitive deficit due to and following nontraumatic intracerebral hemorrhage |
| SNOMED CT | 428051000124108 | Mild dementia |
| SNOMED CT | 428351000124105 | Severe dementia |
| SNOMED CT | 430771000124100 | Moderate dementia |
| SNOMED CT | 672561000119103 | Cognitive deficit due to and following ischemic cerebrovascular accident |
| SNOMED CT | 672571000119109 | Cognitive deficit due to and following hemorrhagic cerebrovascular accident |
| SNOMED CT | 690341000119100 | Cognitive deficit due to and following embolic cerebrovascular accident |
| SNOMED CT | 15928141000119107 | Human immunodeficiency virus infection with cognitive impairment |
| SNOMED CT | 16219201000119101 | Behavioural disturbance co-occurrent and due to late onset Alzheimer dementia |
| SNOMED CT | 16276361000119109 | Vascular dementia without behavioral disturbance |
| SNOMED CT | 16703551000119107 | Memory deficit due to and following cerebrovascular disease |
| SNOMED CT | 16703661000119105 | Memory deficit due to and following cerebrovascular accident |
| SNOMED CT | 16703761000119102 | Memory deficit due to and following ischemic cerebrovascular accident |
| SNOMED CT | 16703821000119101 | Memory deficit due to and following hemorrhagic cerebrovascular accident |
| SNOMED CT | 25470001000004105 | Cognitive impairment due to multiple sclerosis |
| SNOMED CT | 4817008 | Dementia Alzheimers type, late onset with delirium |
| SNOMED CT | 6475002 | Dementia in Alzheimer's disease - type 2 |
| SNOMED CT | 10532003 | Alzheimers dementia, early onset, with depressed mood |
| SNOMED CT | 26852004 | Alzheimers dementia, late onset, with depressive mood |
| SNOMED CT | 26929004 | Alzheimer dementia |
| SNOMED CT | 55009008 | Alzheimers dementia, late onset, with delusions |
| SNOMED CT | 58756001 | Huntington disease |
| SNOMED CT | 66108005 | Dementia in Alzheimer's disease - type 1 |
| SNOMED CT | 230265002 | Familial Alzheimer disease of early onset |
| SNOMED CT | 230266001 | Non-familial Alzheimer disease of early onset |
| SNOMED CT | 230267005 | Familial Alzheimer disease of late onset |
| SNOMED CT | 230268000 | Non-familial Alzheimer disease of late onset |
| SNOMED CT | 230269008 | Focal Alzheimer disease |
| SNOMED CT | 230280008 | Progressive aphasia in Alzheimer disease |
| SNOMED CT | 230299004 | Juvenile onset Huntington disease |
| SNOMED CT | 230300007 | Late onset Huntington disease |
| SNOMED CT | 230301006 | Akinetic-rigid form of Huntington disease |
| SNOMED CT | 312991009 | Senile Lewy body dementia |
| SNOMED CT | 416780008 | Dementia in Alzheimer's disease with early onset |

Continued on next page

**TableS9 – continued from previous page**

| CodeSystem | ConceptCode | ConceptName |
|---|---|---|
| SNOMED CT | 416975007 | Dementia of the Alzheimers type, late onset |
| SNOMED CT | 420614009 | Organic dementia with AIDS (acquired immunodeficiency syndrome) |
| SNOMED CT | 421023003 | Presenile dementia with AIDS (acquired immunodeficiency syndrome) |
| SNOMED CT | 421529006 | Dementia with AIDS (acquired immunodeficiency syndrome) |
| SNOMED CT | 425390006 | Dementia in Parkinsons disease |
| SNOMED CT | 442344002 | Dementia due to Huntingtons disease |
| SNOMED CT | 733193001 | Dementia with progressive multifocal leucoencephalopathy |
| SNOMED CT | 21921000119103 | Dementia co-occurrent and due to Pick's disease |
| SNOMED CT | 82361000119107 | Altered behavior in dementia due to Huntington chorea |
| SNOMED CT | 429161000124103 | Primary degenerative dementia of the Alzheimer type, senile onset, with behavioral disturbance |
| SNOMED CT | 16703491000119101 | Memory deficit due to and following spontaneous intracerebral hemorrhage |
| SNOMED CT | 16703601000119109 | Memory deficit due to and following spontaneous subarachnoid hemorrhage |
| SNOMED CT | 16703711000119100 | Memory deficit due to and following embolic cerebrovascular accident |
| SNOMED CT | 1144748009 | Impaired concentration |
| SNOMED CT | 1148924004 | Dementia due to deficiency of folic acid |
| SNOMED CT | 1156789004 | Autosomal dominant Alzheimer disease due to mutation of amyloid precursor protein |
| SNOMED CT | 1156798001 | Autosomal dominant Alzheimer disease due to mutation of presenilin 2 |
| SNOMED CT | 1156800008 | Autosomal dominant Alzheimer disease due to mutation of presenilin 1 |
| SNOMED CT | 1186724002 | HTRA1-related autosomal dominant cerebral small vessel disease |
| SNOMED CT | 1187004001 | Chronic traumatic encephalopathy |
| SNOMED CT | 1186839009 | Cognitive impairment due to lead toxicity |
| SNOMED CT | 293671000119109 | Behavioral disturbance due to multi-infarct dementia |
| SNOMED CT | 1187126002 | ITM2B-related amyloidosis |
| SNOMED CT | 1186887004 | Dementia caused by manganese |
| SNOMED CT | 1186880002 | Dementia due to cobalamin deficiency |
| SNOMED CT | 1186879000 | Dementia due to thiamine deficiency |
| SNOMED CT | 1186877003 | Dementia due to vitamin E deficiency |
| SNOMED CT | 1186881003 | Dementia due to niacin deficiency |
| SNOMED CT | 1186883000 | Dementia due to nutritional deficiency |

#### 4.7 Cough

Table S10: Concept codes used to identify if a patient was diagnosed with a cough.

| CodeSystem | ConceptCode | ConceptName |
|---|---|---|
| ICD10CM | R05 | Cough |
| ICD10CM | R05.2 | Subacute cough |
| ICD10CM | R05.3 | Chronic cough |
| ICD10CM | R05.9 | Cough, unspecified |
| ICD10CM | R05.4 | Cough syncope |
| ICD10CM | R05.1 | Acute cough |
| ICD10CM | R05.8 | Other specified cough |
| SNOMED CT | 263731006 | Coughing |
| SNOMED CT | 49727002 | Cough |
| SNOMED CT | 272039006 | C/O - cough |
| SNOMED CT | 276314008 | Coughing ineffective |
| SNOMED CT | 277357006 | Sputum retention |
| SNOMED CT | 284523002 | Persistent cough |
| SNOMED CT | 289113002 | Able to cough |
| SNOMED CT | 289114008 | Does cough |
| SNOMED CT | 289115009 | Does not cough |
| SNOMED CT | 289116005 | Difficulty coughing |
| SNOMED CT | 289965001 | Croupy cough |
| SNOMED CT | 300959008 | Allergic cough |
| SNOMED CT | 301236000 | Effective cough |
| SNOMED CT | 301245004 | Difficulty in coughing up sputum |
| SNOMED CT | 301246003 | Does cough up sputum |
| SNOMED CT | 301247007 | Does not cough up sputum |
| SNOMED CT | 306780002 | Able to cough voluntarily |
| SNOMED CT | 306781003 | Unable to cough voluntarily |
| SNOMED CT | 306782005 | Difficulty coughing voluntarily |
| SNOMED CT | 315246003 | Unexplained cough |
| SNOMED CT | 366124008 | Finding related to ability to cough |
| SNOMED CT | 366125009 | Finding related to ability to cough voluntarily |
| SNOMED CT | 366126005 | Finding related to ability to cough up sputum |
| SNOMED CT | 417850002 | Respiratory tract congestion and cough |
| SNOMED CT | 445241004 | Postviral cough |
| SNOMED CT | 102580004 | Cough suppression |
| SNOMED CT | 111962006 | Postural cough |
| SNOMED CT | 123819004 | Cough reflex absent |
| SNOMED CT | 135883003 | Cough with fever |
| SNOMED CT | 161923004 | Productive cough -clear sputum |
| SNOMED CT | 161924005 | Productive cough -green sputum |
| SNOMED CT | 161925006 | Productive cough-yellow sputum |
| SNOMED CT | 161932002 | Morning cough |
| SNOMED CT | 161933007 | Evening cough |
| SNOMED CT | 161947006 | Nocturnal cough / wheeze |
| SNOMED CT | 225575005 | Unable to cough up sputum |
| SNOMED CT | 225576006 | Able to cough up sputum |
| SNOMED CT | 247410004 | Painful cough |

Continued on next page

**TableS10 – continued from previous page**

| CodeSystem | ConceptCode | ConceptName |
|---|---|---|
| SNOMED CT | 248589007 | Clearing throat - hawking |
| SNOMED CT | 248590003 | Unable to cough |
| SNOMED CT | 248593001 | Cough when swallowing |
| SNOMED CT | 248594007 | Tracheal esophageal fistula cough |
| SNOMED CT | 7142008 | Cough at rest |
| SNOMED CT | 11833005 | Dry cough |
| SNOMED CT | 17986004 | Barking cough |
| SNOMED CT | 19282004 | Cough on exercise |
| SNOMED CT | 20670007 | Brassy cough |
| SNOMED CT | 43025008 | Paroxysmal cough |
| SNOMED CT | 46789001 | Cough after eating |
| SNOMED CT | 46802002 | Smokers' cough |
| SNOMED CT | 52673003 | Decreased coughing |
| SNOMED CT | 59994004 | Hacking cough |
| SNOMED CT | 62427007 | Bovine cough |
| SNOMED CT | 62548007 | Nocturnal cough |
| SNOMED CT | 62618004 | Early morning cough |
| SNOMED CT | 62731002 | Increasing frequency of cough |
| SNOMED CT | 63000007 | Spasmodic cough |
| SNOMED CT | 68154008 | Chronic cough |
| SNOMED CT | 769211009 | No cough strength |
| SNOMED CT | 28743005 | Bronchial cough |
| Truveta | 1215927 | Cough with exposure to severe acute respiratory syndrome coronavirus 2 (SARS-CoV-2) |
| Truveta | 1215957 | Post-COVID-19 chronic cough |
| SNOMED CT | 2561000112106 | Chronic refractory cough |
| SNOMED CT | 1204369007 | Unexplained chronic cough |

#### 4.8 Diarrhea

Table S11: Concept codes used to identify if a patient was diagnosed with diarrhea.

| CodeSystem | ConceptCode | ConceptName |
|---|---|---|
| ICD10CM | R19.7 | Diarrhea, unspecified |
| SNOMED CT | 62315008 | Diarrhea |
| SNOMED CT | 267060006 | Diarrhea symptom |
| SNOMED CT | 268651002 | Psychogenic diarrhea |
| SNOMED CT | 268850004 | Neonatal diarrhea |
| SNOMED CT | 274081004 | Bacillary dysentery |
| SNOMED CT | 275297005 | Diarrhea and vomiting, symptom |
| SNOMED CT | 276524004 | Neonatal chloridorrhea |
| SNOMED CT | 288199005 | Diarrhea and vomiting after gastrointestinal tract surgery |
| SNOMED CT | 301469002 | Antibiotic-associated diarrhea |
| SNOMED CT | 312109000 | Bacterial dysentery |

Continued on next page

**TableS11 – continued from previous page**

| CodeSystem | ConceptCode | ConceptName |
|---|---|---|
| SNOMED CT | 387754006 | Amebic dysentery |
| SNOMED CT | 398570005 | Diarrhea due to staphylococcus |
| SNOMED CT | 409587002 | Severe diarrhea |
| SNOMED CT | 409966000 | Acute diarrhea |
| SNOMED CT | 425739008 | Brainerd diarrhea |
| SNOMED CT | 428867008 | Diarrhea due to drug |
| SNOMED CT | 703406006 | Trichohepatoenteric syndrome |
| SNOMED CT | 716277000 | Chronic diarrhea due to glucoamylase deficiency |
| SNOMED CT | 720009004 | Intractable diarrhea with choanal atresia and eye anomaly syndrome |
| SNOMED CT | 721615009 | Non-infective neonatal diarrhea |
| SNOMED CT | 69980003 | Non-infective diarrhea |
| SNOMED CT | 82047000 | Diarrhea due to alcohol intake |
| SNOMED CT | 83134002 | Dietetic diarrhea |
| SNOMED CT | 86615009 | Epidemic diarrhea |
| SNOMED CT | 95544006 | Inflammatory diarrhea |
| SNOMED CT | 95545007 | Hemorrhagic diarrhea |
| SNOMED CT | 95879001 | Diarrheic shellfish poisoning |
| SNOMED CT | 111911008 | Chronic intestinal amebiasis without abscess |
| SNOMED CT | 111912001 | Acute amebic dysentery without abscess |
| SNOMED CT | 186116005 | Acute amebic dysentery |
| SNOMED CT | 197125005 | Irritable bowel syndrome with diarrhea |
| SNOMED CT | 197132001 | Diarrhea after gastrointestinal tract surgery |
| SNOMED CT | 235839006 | Irritable bowel syndrome variant of childhood with diarrhea |
| SNOMED CT | 236066002 | Prototheca diarrhea |
| SNOMED CT | 236071009 | Chronic diarrhea |
| SNOMED CT | 236074001 | Diarrhea due to laxative abuse |
| SNOMED CT | 236075000 | Diarrhea due to ingestion of unabsorbable substances |
| SNOMED CT | 236077008 | Protracted diarrhea |
| SNOMED CT | 236078003 | Post-vagotomy diarrhea |
| SNOMED CT | 249517009 | Constipation alternates with diarrhea |
| SNOMED CT | 249519007 | Diarrhea and vomiting |
| SNOMED CT | 2919008 | Nausea, vomiting and diarrhea |
| SNOMED CT | 2946003 | Osmotic diarrhea |
| SNOMED CT | 11003002 | Nervous diarrhea |
| SNOMED CT | 14384003 | Postprandial diarrhea |
| SNOMED CT | 15699003 | Secretory diarrhea |
| SNOMED CT | 17551007 | Chronic diarrhea of unknown origin |
| SNOMED CT | 18168004 | Raw-milk associated diarrhea |
| SNOMED CT | 18425006 | Passage of rice water stools |
| SNOMED CT | 18805001 | Congenital secretory diarrhea, sodium type |
| SNOMED CT | 19213003 | Infectious diarrheal disease |
| SNOMED CT | 23874000 | Chronic amebiasis |
| SNOMED CT | 24412005 | Congenital secretory diarrhea, chloride type |
| SNOMED CT | 25319005 | Chronic diarrhea of infants AND/OR young children |
| SNOMED CT | 25898005 | Congenital secretory diarrhea |
| SNOMED CT | 31499008 | Chronic constipation with overflow |

Continued on next page

**TableS11 – continued from previous page**

| CodeSystem | ConceptCode | ConceptName |
|---|---|---|
| SNOMED CT | 36188001 | Shigellosis |
| SNOMED CT | 38205001 | Diarrhea due to diabetes mellitus |
| SNOMED CT | 39963006 | Toddler diarrhea |
| SNOMED CT | 43240000 | Diarrhea of presumed infectious origin |
| SNOMED CT | 46799006 | Dysenteric diarrhea |
| SNOMED CT | 49237006 | Allergic diarrhea |
| SNOMED CT | 53156005 | Postcholecystectomy diarrhea |
| SNOMED CT | 57725006 | Balantidiasis |
| SNOMED CT | 722392003 | Congenital malabsorptive diarrhea due to paucity of enteroendocrine cells |
| SNOMED CT | 734019006 | Chronic diarrhea with villous atrophy syndrome |
| SNOMED CT | 734884008 | Carcinoid syndrome diarrhea |
| SNOMED CT | 773579007 | Congenital chronic diarrhea with protein-losing enteropathy |
| SNOMED CT | 785727000 | Chronic infantile diarrhea due to guanylate cyclase 2C overactivity |
| SNOMED CT | 5891000119102 | Clostridium difficile diarrhea |
| SNOMED CT | 11718971000119100 | Diarrhea in pregnancy |
| SNOMED CT | 11840006 | Traveler's diarrhoea |
| SNOMED CT | 128333008 | Diarrhoeal disorder |
| Truveta | 1215942 | Diarrhea due to COVID-19 |
| SNOMED CT | 1222709003 | Syndromic congenital sodium diarrhea |

#### 4.9 Dyspnea

Table S12: Concept codes used to identify if a patient was diagnosed with dyspnea.

| CodeSystem | ConceptCode | ConceptName |
|---|---|---|
| ICD10CM | R06.0 | Dyspnea |
| ICD10CM | R06.00 | Dyspnea, unspecified |
| ICD10CM | R06.01 | Orthopnea |
| ICD10CM | R06.02 | Shortness of breath |
| ICD10CM | R06.03 | Acute respiratory distress |
| ICD10CM | R06.09 | Other forms of dyspnea |
| SNOMED CT | 267036007 | Dyspnea |
| SNOMED CT | 297216006 | Increasing breathlessness |
| SNOMED CT | 390870001 | Short of breath dressing/undressing |
| SNOMED CT | 390871002 | Breathless - strenuous exertion |
| SNOMED CT | 391120009 | Medical Research Council Dyspnoea scale grade 1 |
| SNOMED CT | 391123006 | Medical Research Council Dyspnoea scale grade 2 |
| SNOMED CT | 391124000 | Medical Research Council Dyspnoea scale grade 3 |
| SNOMED CT | 391125004 | Medical Research Council Dyspnoea scale grade 4 |
| SNOMED CT | 391126003 | Medical Research Council Dyspnoea scale grade 5 |
| SNOMED CT | 397702004 | Borg Breathlessness Score finding |
| SNOMED CT | 401275008 | Borg Breathlessness Score: 0 none at all |

Continued on next page

**TableS12 – continued from previous page**

| CodeSystem | ConceptCode | ConceptName |
|---|---|---|
| SNOMED CT | 401279002 | Borg Breathlessness Score: 1 very slight |
| SNOMED CT | 401280004 | Borg Breathlessness Score: 2 slight |
| SNOMED CT | 401281000 | Borg Breathlessness Score: 3 moderate |
| SNOMED CT | 401282007 | Borg Breathlessness Score: 4 somewhat severe |
| SNOMED CT | 401284008 | Borg Breathlessness Score: 5 severe |
| SNOMED CT | 401286005 | Borg Breathlessness Score: 6 severe (+) |
| SNOMED CT | 401290007 | Borg Breathlessness Score: 7 very severe |
| SNOMED CT | 401291006 | Borg Breathlessness Score: 8 very severe (+) |
| SNOMED CT | 401292004 | Borg Breathlessness Score: 9 very, very severe (almost maximal) |
| SNOMED CT | 401293009 | Borg Breathlessness Score: 10 maximal |
| SNOMED CT | 401323002 | Borg Breathlessness Score: 0.5 very, very slight (just noticeable) |
| SNOMED CT | 407588003 | Unable to complete a sentence in one breath |
| SNOMED CT | 102577000 | Trepopnea |
| SNOMED CT | 161939006 | Breathless - moderate exertion |
| SNOMED CT | 161940008 | Breathless - mild exertion |
| SNOMED CT | 161941007 | Dyspnea at rest |
| SNOMED CT | 162890008 | O/E - dyspnea |
| SNOMED CT | 162891007 | O/E - orthopnea |
| SNOMED CT | 248548009 | Nocturnal dyspnea |
| SNOMED CT | 17216000 | Dyspnea, class I |
| SNOMED CT | 20112008 | Dyspnea leaning over |
| SNOMED CT | 23141003 | Gasping for breath |
| SNOMED CT | 24921003 | Dyspnea raising arms |
| SNOMED CT | 25209001 | Inspiratory dyspnea |
| SNOMED CT | 30744009 | Platypnea |
| SNOMED CT | 34560001 | Expiratory dyspnea |
| SNOMED CT | 55442000 | Paroxysmal nocturnal dyspnea |
| SNOMED CT | 57769004 | Dyspnea after eating |
| SNOMED CT | 59265000 | Paroxysmal dyspnea |
| SNOMED CT | 60845006 | Dyspnea on exertion |
| SNOMED CT | 62744007 | Orthopnea |
| SNOMED CT | 870535009 | Chronic dyspnea |
| SNOMED CT | 119981000146107 | Dyspnea caused by severe acute respiratory syndrome coronavirus 2 |
| SNOMED CT | 39950000 | Dyspnea, class 3 |
| SNOMED CT | 72365000 | Dyspnea, class 2 |
| SNOMED CT | 73322006 | Dyspnea, class 4 |
| SNOMED CT | 422177004 | Dyspnea with AIDS (acquired immunodeficiency syndrome) |
| Truveta | 1215952 | Persistent dyspnea after COVID-19 |
| Truveta | 1215959 | Post-COVID-19 chronic dyspnea |
| Truveta | 1215975 | Shortness of breath with exposure to severe acute respiratory syndrome coronavirus 2 (SARS-CoV-2) |
| SNOMED CT | 1217110005 | Bendopnea |

##### 4.10 Fatigue

Table S13: Concept codes used to identify if a patient was diagnosed with fatigue.

| CodeSystem | ConceptCode | ConceptName |
|---|---|---|
| ICD10CM | G93.3 | Postviral fatigue syndrome |
| ICD10CM | R53.8 | Other malaise and fatigue |
| ICD10CM | R53.81 | Other malaise |
| ICD10CM | R53.82 | Chronic fatigue, unspecified |
| ICD10CM | R53.83 | Other fatigue |
| SNOMED CT | 271795006 | Malaise and fatigue |
| SNOMED CT | 224960004 | Tired |
| SNOMED CT | 52702003 | Chronic fatigue syndrome |
| SNOMED CT | 267032009 | Tired all the time |
| SNOMED CT | 444042007 | Postexertional fatigue |
| SNOMED CT | 713568000 | Occasionally tired |
| SNOMED CT | 248269005 | Tired on least exertion |

#### 4.11 Fever

Table S14: Concept codes used to identify if a patient was diagnosed with a fever

| CodeSystem | ConceptCode | ConceptName |
|---|---|---|
| ICD10CM | R50.81 | Fever presenting with conditions classified elsewhere |
| ICD10CM | R50.9 | Fever, unspecified |
| SNOMED CT | 386661006 | Fever |
| SNOMED CT | 267340006 | Maternal pyrexia in labor |
| SNOMED CT | 271749004 | Acute rise of fever |
| SNOMED CT | 271750004 | Gradual rise of fever |
| SNOMED CT | 271751000 | Continuous fever |
| SNOMED CT | 271752007 | Staircase fever |
| SNOMED CT | 271753002 | Irregular fever |
| SNOMED CT | 271754008 | Rapid fall of fever |
| SNOMED CT | 271755009 | Gradual fall of fever |
| SNOMED CT | 271897009 | O/E - fever |
| SNOMED CT | 274308003 | O/E - hyperpyrexia |
| SNOMED CT | 274640006 | Fever with chills |
| SNOMED CT | 304213008 | Low grade pyrexia |
| SNOMED CT | 365977007 | Pattern of fever - finding |
| SNOMED CT | 365989009 | Phase of fever - finding |
| SNOMED CT | 405501007 | Malignant hyperthermia |
| SNOMED CT | 405543000 | Drug-induced hyperpyrexia |
| SNOMED CT | 409702008 | Hyperpyrexia |
| SNOMED CT | 420079008 | Relapsing fever |
| SNOMED CT | 449129007 | Paraneoplastic fever |
| SNOMED CT | 698578005 | Post diphtheria, tetanus and pertussis vaccination fever |
| SNOMED CT | 698579002 | Post diphtheria vaccination fever |
| SNOMED CT | 698580004 | Post tetanus vaccination fever |

Continued on next page

**TableS14 – continued from previous page**

| CodeSystem | ConceptCode | ConceptName |
|---|---|---|
| SNOMED CT | 698581000 | Post pertussis vaccination fever |
| SNOMED CT | 704425001 | Chronic fever |
| SNOMED CT | 713014002 | Infection caused by Borrelia miyamotoi |
| SNOMED CT | 713731001 | Pyrexia of unknown origin co-occurrent with human immunodeficiency virus infection |
| SNOMED CT | 719398004 | Malignant hyperthermia with arthrogryposis and torticollis syndrome |
| SNOMED CT | 73301000 | Febrile transfusion reaction |
| SNOMED CT | 74873003 | Relapsing fever of Central AND/OR South America |
| SNOMED CT | 77957000 | Intermittent fever |
| SNOMED CT | 78168002 | Relapsing fever of Western North America |
| SNOMED CT | 88808006 | Relapsing fever of Iran AND/OR Central Asia |
| SNOMED CT | 95627000 | Fever of newborn |
| SNOMED CT | 102496004 | Spiking fever |
| SNOMED CT | 111950007 | Factitious fever |
| SNOMED CT | 135883003 | Cough with fever |
| SNOMED CT | 164288004 | O/E - pyrexia of unknown origin |
| SNOMED CT | 164299002 | O/E - level of fever |
| SNOMED CT | 164301009 | O/E - temperature low |
| SNOMED CT | 164304001 | O/E - hyperpyrexia - greater than 40.5 degrees Celsius |
| SNOMED CT | 164307008 | O/E - fever - acute rise |
| SNOMED CT | 164308003 | O/E - fever - gradual rise |
| SNOMED CT | 164309006 | O/E - fever - continuous |
| SNOMED CT | 164311002 | O/E - fever - remittent |
| SNOMED CT | 164312009 | O/E - fever - intermittent |
| SNOMED CT | 164314005 | O/E - fever - irregular |
| SNOMED CT | 164315006 | O/E - fever - fast fall-crisis |
| SNOMED CT | 164316007 | O/E - fever-gradual fall-lysis |
| SNOMED CT | 186694006 | Sweating fever |
| SNOMED CT | 200277008 | Puerperal pyrexia of unknown origin |
| SNOMED CT | 200280009 | Puerperal pyrexia of unknown origin - delivered with postnatal complication |
| SNOMED CT | 200281008 | Puerperal pyrexia of unknown origin with postnatal complication |
| SNOMED CT | 201796004 | Systemic onset juvenile chronic arthritis |
| SNOMED CT | 213026003 | Malignant hyperpyrexia due to anesthetic |
| SNOMED CT | 237348005 | Puerperal pyrexia |
| SNOMED CT | 240822009 | Bancroftian filarial fever |
| SNOMED CT | 240827003 | Malayan filarial fever |
| SNOMED CT | 248432005 | Rising phase of fever |
| SNOMED CT | 248433000 | Plateau phase of fever |
| SNOMED CT | 248434006 | Falling phase of fever |
| SNOMED CT | 248435007 | Prolonged fever |
| SNOMED CT | 248436008 | Slightly remittent fever |
| SNOMED CT | 248437004 | Malarial fever |
| SNOMED CT | 248438009 | Quotidien fever |
| SNOMED CT | 248439001 | Double quotidien fever |
| SNOMED CT | 248440004 | Tertian fever |

Continued on next page

**TableS14 – continued from previous page**

| CodeSystem | ConceptCode | ConceptName |
| --- | --- | --- |
| SNOMED CT | 248441000 | Malignant tertian fever |
| SNOMED CT | 248442007 | Quartan fever |
| SNOMED CT | 248443002 | Intermittent hepatic fever |
| SNOMED CT | 248444008 | Biphasic fever |
| SNOMED CT | 248445009 | Fever, diurnal variation |
| SNOMED CT | 248446005 | Reversed diurnal fever |
| SNOMED CT | 248447001 | Fever defervescence |
| SNOMED CT | 248449003 | Central fever |
| SNOMED CT | 248450003 | Postoperative fever |
| SNOMED CT | 248451004 | Postpartum fever |
| SNOMED CT | 7520000 | Pyrexia of unknown origin |
| SNOMED CT | 8554002 | Brazilian purpuric fever |
| SNOMED CT | 9619006 | Aseptic fever |
| SNOMED CT | 10301003 | Tick-borne relapsing fever |
| SNOMED CT | 14683004 | Louse-borne relapsing fever |
| SNOMED CT | 27578005 | Relapsing fever of the Caucasus |
| SNOMED CT | 31120005 | Relapsing fever of Asia AND/OR Africa |
| SNOMED CT | 39002001 | Relapsing fever of Western United States |
| SNOMED CT | 42136008 | Swinging fever |
| SNOMED CT | 43626008 | Relapsing fever of Central AND/OR South Africa |
| SNOMED CT | 49513005 | Dehydration fever in newborn |
| SNOMED CT | 52635002 | Relapsing fever of Iberian Peninsula AND/OR Northwest Africa |
| SNOMED CT | 52715007 | Pel Ebstein fever |
| SNOMED CT | 60025004 | Transitory fever of newborn |
| SNOMED CT | 63993003 | Remittent fever |
| SNOMED CT | 722892007 | Fever due to infection |
| SNOMED CT | 10151000132103 | Viral fever |
| SNOMED CT | 119751000146104 | Fever caused by Severe acute respiratory syndrome coronavirus 2 |
| SNOMED CT | 123471000119103 | Post vaccination fever |
| SNOMED CT | 130091000119103 | Postprocedural fever |
| SNOMED CT | 435001000124103 | Febrile transfusion reaction without hemolysis |
| SNOMED CT | 58827009 | Relapsing fever of Southern United States, Mexico, Central AND/OR South America |
| SNOMED CT | 239920006 | Wissler-Fanconi syndrome |
| SNOMED CT | 426000000 | Fever greater than 38 Celsius |
| Truveta | 1215943 | Fever with exposure to severe acute respiratory syndrome coronavirus 2 (SARS-CoV-2) |
| SNOMED CT | 1187615007 | TAFRO syndrome |

#### 4.12 Headache

Table S15: Concept codes used to identify if a patient was diagnosed with a headache

| CodeSystem | ConceptCode | ConceptName |
|---|---|---|
| ICD10CM | G44.83 | Primary cough headache |
| ICD10CM | G44.89 | Other headache syndrome |
| ICD10CM | R51 | Headache |
| ICD10CM | R51.0 | Headache with orthostatic component, not elsewhere classified |
| ICD10CM | R51.9 | Headache, unspecified |
| SNOMED CT | 25064002 | Headache |
| SNOMED CT | 267096005 | Frontal headache |
| SNOMED CT | 272027003 | C/O - a headache |
| SNOMED CT | 274305000 | On examination - maxillary sinus pain |
| SNOMED CT | 274306004 | O/E -frontal sinus pain |
| SNOMED CT | 279016001 | Cervicogenic headache |
| SNOMED CT | 301356002 | Maxillary sinus pain |
| SNOMED CT | 301357006 | Frontal sinus pain |
| SNOMED CT | 364760007 | Headache character - finding |
| SNOMED CT | 395688005 | Benign coital headache |
| SNOMED CT | 398057008 | Tension-type headache |
| SNOMED CT | 398126006 | Muscular headache |
| SNOMED CT | 398987004 | Headache following lumbar puncture |
| SNOMED CT | 423279000 | Refractory migraine without aura |
| SNOMED CT | 423683008 | Refractory migraine with aura |
| SNOMED CT | 423894005 | Refractory migraine |
| SNOMED CT | 424699007 | Migraine variants, not intractable |
| SNOMED CT | 425007008 | Migraine without aura, not refractory |
| SNOMED CT | 425365009 | Refractory migraine variants |
| SNOMED CT | 425422007 | Headache due to high altitude |
| SNOMED CT | 425936006 | Intractable ophthalmic migraine |
| SNOMED CT | 427419006 | Transformed migraine |
| SNOMED CT | 431237007 | Chronic headache disorder |
| SNOMED CT | 443094001 | Paroxysmal hemicrania |
| SNOMED CT | 443095000 | Hemicrania continua |
| SNOMED CT | 445322004 | Migraine variant with headache |
| SNOMED CT | 445511002 | Orthostatic headache |
| SNOMED CT | 449814007 | Trigeminal autonomic cephalalgia |
| SNOMED CT | 698803006 | Medication overuse headache |
| SNOMED CT | 699314009 | Migraine with persistent visual aura |
| SNOMED CT | 699694000 | Chronic post-concussion headache |
| SNOMED CT | 703182002 | Headache due to reversible cerebral vasoconstriction syndrome |
| SNOMED CT | 711545001 | Migraine due to estrogen contraceptive |
| SNOMED CT | 712826000 | Intermittent headache |
| SNOMED CT | 712831003 | Frequent headache |
| SNOMED CT | 79267007 | Retinal migraine |
| SNOMED CT | 83351003 | Basilar migraine |
| SNOMED CT | 86925001 | Ocular headache |
| SNOMED CT | 95653008 | Acute confusional migraine |
| SNOMED CT | 95654002 | Chronic paroxysmal hemicrania |

Continued on next page

**TableS15 – continued from previous page**

| CodeSystem | ConceptCode | ConceptName |
|---|---|---|
| SNOMED CT | 95655001 | Ophthalmic migraine |
| SNOMED CT | 95656000 | Familial hemiplegic migraine |
| SNOMED CT | 95657009 | Chronic mixed headache syndrome |
| SNOMED CT | 95658004 | Cough headache syndrome |
| SNOMED CT | 95659007 | Hindbrain hernia headache |
| SNOMED CT | 95660002 | Thunderclap headache |
| SNOMED CT | 95670000 | Nervus intermedius neuralgia |
| SNOMED CT | 103007003 | Headache due to external compression of head |
| SNOMED CT | 103008008 | Postpartum headache |
| SNOMED CT | 103009000 | Headache due to cold exposure |
| SNOMED CT | 103010005 | Headache associated with sexual activity |
| SNOMED CT | 103011009 | Benign exertional headache |
| SNOMED CT | 103012002 | Analgesic overuse headache |
| SNOMED CT | 112101004 | Dental headache |
| SNOMED CT | 128187005 | Vascular headache |
| SNOMED CT | 129610005 | Headache due to intracranial disease |
| SNOMED CT | 129612002 | Anterior ethmoidal nerve syndrome |
| SNOMED CT | 162211001 | Viral headache |
| SNOMED CT | 162299003 | Generalized headache |
| SNOMED CT | 162300006 | Unilateral headache |
| SNOMED CT | 162301005 | Bilateral headache |
| SNOMED CT | 162304002 | Parietal headache |
| SNOMED CT | 162307009 | Aching headache |
| SNOMED CT | 162308004 | Throbbing headache |
| SNOMED CT | 162309007 | Shooting headache |
| SNOMED CT | 162310002 | Morning headache |
| SNOMED CT | 162311003 | Heavy head |
| SNOMED CT | 169464001 | Headache caused by oral contraceptive pill |
| SNOMED CT | 193028008 | Sick headache |
| SNOMED CT | 193030005 | Migraine variants |
| SNOMED CT | 193031009 | Cluster headache |
| SNOMED CT | 193039006 | Complicated migraine |
| SNOMED CT | 198438009 | Menopausal headache |
| SNOMED CT | 200071001 | Obstetric spinal and epidural anesthesia-induced headache |
| SNOMED CT | 200072008 | Spinal and epidural anesthesia-induced headache during pregnancy |
| SNOMED CT | 200073003 | Spinal and epidural anesthesia-induced headache during the puerperium |
| SNOMED CT | 200076006 | Spinal and epidural anesthesia-induced headache during labor and delivery |
| SNOMED CT | 230461009 | Headache disorder |
| SNOMED CT | 230462002 | Migraine with typical aura |
| SNOMED CT | 230464001 | Non-familial hemiplegic migraine |
| SNOMED CT | 230465000 | Migraine aura without headache |
| SNOMED CT | 230467008 | Status migrainosus |
| SNOMED CT | 230468003 | Migraine with ischemic complication |
| SNOMED CT | 230470007 | Episodic tension-type headache |

Continued on next page

**TableS15 – continued from previous page**

| CodeSystem | ConceptCode | ConceptName |
|---|---|---|
| SNOMED CT | 230471006 | Chronic tension-type headache |
| SNOMED CT | 230472004 | Episodic cluster headache |
| SNOMED CT | 230473009 | Chronic cluster headache |
| SNOMED CT | 230474003 | Chronic cluster headache unremitting from onset |
| SNOMED CT | 230475002 | Chronic cluster headache evolved from episodic cluster headache |
| SNOMED CT | 230476001 | Atypical cluster headache |
| SNOMED CT | 230477005 | Chronic post-traumatic headache |
| SNOMED CT | 230478000 | Low pressure headache |
| SNOMED CT | 230479008 | Post dural puncture headache |
| SNOMED CT | 230480006 | Headache following myelography |
| SNOMED CT | 230481005 | Idiopathic stabbing headache |
| SNOMED CT | 230483008 | Headache associated with substance abuse or withdrawal |
| SNOMED CT | 330007 | Occipital headache |
| SNOMED CT | 4448006 | Allergic headache |
| SNOMED CT | 4473006 | Migraine with aura |
| SNOMED CT | 4969004 | Sinus headache |
| SNOMED CT | 13322008 | Postseizure headache |
| SNOMED CT | 23186000 | Menstrual migraine |
| SNOMED CT | 26150009 | Lower half migraine |
| SNOMED CT | 37796009 | Migraine |
| SNOMED CT | 38823002 | Aural headache |
| SNOMED CT | 41413006 | Temporal headache |
| SNOMED CT | 43242008 | Drug withdrawal headache |
| SNOMED CT | 44538002 | Nasal headache |
| SNOMED CT | 49605003 | Ophthalmoplegic migraine |
| SNOMED CT | 54012000 | Posttraumatic headache |
| SNOMED CT | 56097005 | Migraine without aura |
| SNOMED CT | 59292006 | Hemiplegic migraine |
| SNOMED CT | 722981005 | Infrequent episodic tension-type headache |
| SNOMED CT | 722982003 | Frequent episodic tension-type headache |
| SNOMED CT | 722983008 | Headache due to injury of head and neck |
| SNOMED CT | 724429004 | Stroke co-occurrent with migraine |
| SNOMED CT | 725058003 | Short-lasting unilateral neuralgiform headache attacks with conjunctival injection and tearing syndrome |
| SNOMED CT | 735938006 | Acute headache |
| SNOMED CT | 762353009 | Acute headache due to traumatic injury of head |
| SNOMED CT | 762354003 | Persistent headache due to and following injury of head |
| SNOMED CT | 762592003 | Chronic primary headache |
| SNOMED CT | 571000119103 | Daily headache |
| SNOMED CT | 121021000119105 | New daily persistent headache |
| SNOMED CT | 121881000119102 | Intractable allergic migraine |
| SNOMED CT | 122021000119104 | Ophthalmic status migrainosus |
| SNOMED CT | 122051000119107 | Retinal status migrainosus |
| SNOMED CT | 122061000119109 | Intractable retinal migraine |
| SNOMED CT | 122141000119109 | Refractory abdominal migraine |
| SNOMED CT | 122711000119109 | Hypnic headache |
| SNOMED CT | 122731000119104 | Primary thunderclap headache |

Continued on next page

**TableS15 – continued from previous page**

| CodeSystem | ConceptCode | ConceptName |
|---|---|---|
| SNOMED CT | 122751000119105 | Acute posttraumatic headache |
| SNOMED CT | 124001000119104 | Common migraine with status migrainosus |
| SNOMED CT | 124041000119102 | Intractable hemiplegic migraine |
| SNOMED CT | 124051000119100 | Hemiplegic status migrainosus |
| SNOMED CT | 124061000119103 | Intractable hemiplegic status migrainosus |
| SNOMED CT | 124071000119109 | Intractable menstrual migraine |
| SNOMED CT | 124081000119107 | Menstrual status migrainosus |
| SNOMED CT | 124091000119105 | Intractable menstrual status migrainosus |
| SNOMED CT | 124161000119104 | Chronic migraine without aura with status migrainosus |
| SNOMED CT | 124171000119105 | Chronic intractable migraine without aura |
| SNOMED CT | 124391000119108 | Status migrainosus with aura |
| SNOMED CT | 145611000119107 | Allergic migraine |
| SNOMED CT | 290431000119101 | Intractable headache following trauma |
| SNOMED CT | 290441000119105 | Intractable chronic headache following trauma |
| SNOMED CT | 290531000119102 | Intractable chronic tension headache |
| SNOMED CT | 290541000119106 | Intractable episodic tension-type headache |
| SNOMED CT | 290731000119106 | Idiopathic partial status epilepticus |
| SNOMED CT | 293901000119108 | Intractable episodic cluster headache |
| SNOMED CT | 294031000119103 | Episodic paroxysmal hemicrania |
| SNOMED CT | 294081000119102 | Intractable headache caused by drug |
| SNOMED CT | 294091000119104 | Headache caused by drug |
| SNOMED CT | 429181000124108 | Refractory acute confusional migraine |
| SNOMED CT | 431061000124100 | Ophthalmoplegic migraine, refractory |
| SNOMED CT | 431601000124105 | Chronic migraine without aura |
| SNOMED CT | 436771000124102 | Basilar artery migraine, refractory |
| SNOMED CT | 437931000124100 | Chronic migraine without aura, non-refractory |
| SNOMED CT | 445491000124104 | Postural headache |
| SNOMED CT | 458241000124102 | Chronic daily headache |
| SNOMED CT | 1119294003 | Unilateral left sided headache |
| SNOMED CT | 1119295002 | Unilateral right sided headache |
| SNOMED CT | 466841000124105 | Headache after cough |
| SNOMED CT | 145621000146106 | Headache due to exertion |
| SNOMED CT | 471651000124106 | Episodic migraine |
| SNOMED CT | 471661000124108 | Acute migraine |
| Truveta | 1215961 | Post-COVID-19 chronic headache |

#### 4.13 Insomnia

Table S16: Concept codes used to identify if a patient was diagnosed with insomnia.

| CodeSystem | ConceptCode | ConceptName |
|---|---|---|
| ICD10CM | F51 | Sleep disorders not due to a substance or known physiological condition |
| ICD10CM | G47 | Sleep disorders |

Continued on next page

**TableS16 – continued from previous page**

| CodeSystem | ConceptCode | ConceptName |
|---|---|---|
| ICD10CM | F51.0 | Insomnia not due to a substance or known physiological condition |
| ICD10CM | F51.01 | Primary insomnia |
| ICD10CM | F51.02 | Adjustment insomnia |
| ICD10CM | F51.03 | Paradoxical insomnia |
| ICD10CM | F51.04 | Psychophysiologic insomnia |
| ICD10CM | F51.05 | Insomnia due to other mental disorder |
| ICD10CM | F51.09 | Other insomnia not due to a substance or known physiological condition |
| ICD10CM | F51.1 | Hypersomnia not due to a substance or known physiological condition |
| ICD10CM | F51.11 | Primary hypersomnia |
| ICD10CM | F51.12 | Insufficient sleep syndrome |
| ICD10CM | F51.13 | Hypersomnia due to other mental disorder |
| ICD10CM | F51.19 | Other hypersomnia not due to a substance or known physiological condition |
| ICD10CM | F51.3 | Sleepwalking [somnambulism] |
| ICD10CM | F51.4 | Sleep terrors [night terrors] |
| ICD10CM | F51.5 | Nightmare disorder |
| ICD10CM | F51.8 | Other sleep disorders not due to a substance or known physiological condition |
| ICD10CM | F51.9 | Sleep disorder not due to a substance or known physiological condition, unspecified |
| ICD10CM | G47.0 | Insomnia |
| ICD10CM | G47.00 | Insomnia, unspecified |
| ICD10CM | G47.01 | Insomnia due to medical condition |
| ICD10CM | G47.09 | Other insomnia |
| ICD10CM | G47.1 | Hypersomnia |
| ICD10CM | G47.10 | Hypersomnia, unspecified |
| ICD10CM | G47.11 | Idiopathic hypersomnia with long sleep time |
| ICD10CM | G47.12 | Idiopathic hypersomnia without long sleep time |
| ICD10CM | G47.13 | Recurrent hypersomnia |
| ICD10CM | G47.14 | Hypersomnia due to medical condition |
| ICD10CM | G47.19 | Other hypersomnia |
| ICD10CM | G47.2 | Circadian rhythm sleep disorders |
| ICD10CM | G47.20 | Circadian rhythm sleep disorder, unspecified type |
| ICD10CM | G47.21 | Circadian rhythm sleep disorder, delayed sleep phase type |
| ICD10CM | G47.22 | Circadian rhythm sleep disorder, advanced sleep phase type |
| ICD10CM | G47.23 | Circadian rhythm sleep disorder, irregular sleep wake type |
| ICD10CM | G47.24 | Circadian rhythm sleep disorder, free running type |
| ICD10CM | G47.25 | Circadian rhythm sleep disorder, jet lag type |
| ICD10CM | G47.26 | Circadian rhythm sleep disorder, shift work type |
| ICD10CM | G47.27 | Circadian rhythm sleep disorder in conditions classified elsewhere |
| ICD10CM | G47.29 | Other circadian rhythm sleep disorder |
| ICD10CM | G47.3 | Sleep apnea |
| ICD10CM | G47.30 | Sleep apnea, unspecified |
| ICD10CM | G47.31 | Primary central sleep apnea |

Continued on next page

**TableS16 – continued from previous page**

| CodeSystem | ConceptCode | ConceptName |
|---|---|---|
| ICD10CM | G47.32 | High altitude periodic breathing |
| ICD10CM | G47.33 | Obstructive sleep apnea (adult) (pediatric) |
| ICD10CM | G47.34 | Idiopathic sleep related nonobstructive alveolar hypoventilation |
| ICD10CM | G47.35 | Congenital central alveolar hypoventilation syndrome |
| ICD10CM | G47.36 | Sleep related hypoventilation in conditions classified elsewhere |
| ICD10CM | G47.37 | Central sleep apnea in conditions classified elsewhere |
| ICD10CM | G47.39 | Other sleep apnea |
| ICD10CM | G47.4 | Narcolepsy and cataplexy |
| ICD10CM | G47.41 | Narcolepsy |
| ICD10CM | G47.411 | Narcolepsy with cataplexy |
| ICD10CM | G47.419 | Narcolepsy without cataplexy |
| ICD10CM | G47.42 | Narcolepsy in conditions classified elsewhere |
| ICD10CM | G47.421 | Narcolepsy in conditions classified elsewhere with cataplexy |
| ICD10CM | G47.429 | Narcolepsy in conditions classified elsewhere without cataplexy |
| ICD10CM | G47.5 | Parasomnia |
| ICD10CM | G47.50 | Parasomnia, unspecified |
| ICD10CM | G47.51 | Confusional arousals |
| ICD10CM | G47.52 | REM sleep behavior disorder |
| ICD10CM | G47.53 | Recurrent isolated sleep paralysis |
| ICD10CM | G47.54 | Parasomnia in conditions classified elsewhere |
| ICD10CM | G47.59 | Other parasomnia |
| ICD10CM | G47.6 | Sleep related movement disorders |
| ICD10CM | G47.61 | Periodic limb movement disorder |
| ICD10CM | G47.62 | Sleep related leg cramps |
| ICD10CM | G47.63 | Sleep related bruxism |
| ICD10CM | G47.69 | Other sleep related movement disorders |
| ICD10CM | G47.8 | Other sleep disorders |
| ICD10CM | G47.9 | Sleep disorder, unspecified |
| SNOMED CT | 39898005 | Sleep disorder |
| SNOMED CT | 268652009 | Transient insomnia |
| SNOMED CT | 268653004 | Transient hypersomnia |
| SNOMED CT | 268654005 | Repetitive intrusions of sleep |
| SNOMED CT | 268722008 | Non-organic disorder of the sleep-wake schedule |
| SNOMED CT | 270487001 | Non-organic sleep disorder |
| SNOMED CT | 271793004 | Irregular sleep-wake pattern |
| SNOMED CT | 271794005 | Disorder of sleep-wake cycle |
| SNOMED CT | 274950005 | Sleep related bruxism |
| SNOMED CT | 277180005 | Sleep paralysis |
| SNOMED CT | 277181009 | Sleep starts |
| SNOMED CT | 361208003 | Primary sleep apnea of newborn |
| SNOMED CT | 363314000 | Substance-induced sleep disorder |
| SNOMED CT | 370971007 | Somnolence syndrome |
| SNOMED CT | 401236004 | Early morning waking |
| SNOMED CT | 407666007 | Nocturnal sleep-related eating disorder |
| SNOMED CT | 413638006 | Benign neonatal sleep myoclonus |
| SNOMED CT | 415238003 | REM sleep behavior disorder |
| SNOMED CT | 418424006 | Post-radiotherapy somnolence syndrome |

Continued on next page

**TableS16 – continued from previous page**

| CodeSystem | ConceptCode | ConceptName |
|---|---|---|
| SNOMED CT | 418475009 | Drug-induced sleep disorder |
| SNOMED CT | 418763003 | Periodic limb movement disorder |
| SNOMED CT | 418955009 | Organic sleep disorder |
| SNOMED CT | 425476007 | Non-organic parasomnia |
| SNOMED CT | 425832009 | Psychophysiologic insomnia |
| SNOMED CT | 425881006 | Limit-setting sleep disorder |
| SNOMED CT | 426451004 | Recurrent hypersomnia |
| SNOMED CT | 426542005 | Sleep hypoventilation due to lower airway obstruction |
| SNOMED CT | 426824009 | Sleep-onset association disorder |
| SNOMED CT | 426943005 | Hypersomnia disorder related to menstruation |
| SNOMED CT | 427426006 | Sleep attack |
| SNOMED CT | 428708005 | Secondary parasomnia |
| SNOMED CT | 429266002 | Sleep-related groaning |
| SNOMED CT | 429276004 | Environmental sleep disorder |
| SNOMED CT | 429456008 | Hypoventilation during sleep due to neuromuscular disorder |
| SNOMED CT | 429571005 | Sleep-related dissociative disorder |
| SNOMED CT | 430390000 | Sleep-related neurogenic tachypnea |
| SNOMED CT | 430893009 | Sleep related rhythmic movement disorder |
| SNOMED CT | 441830002 | Confusional arousal disorder |
| SNOMED CT | 441877007 | Sleep dysfunction with sleep stage disturbance |
| SNOMED CT | 441910000 | Idiopathic sleep related non-obstructive alveolar hypoventilation |
| SNOMED CT | 441976002 | Organic parasomnia |
| SNOMED CT | 442120007 | Recurrent isolated sleep paralysis |
| SNOMED CT | 442164004 | Organic sleep apnea |
| SNOMED CT | 442176004 | Sleep dysfunction with arousal disturbance |
| SNOMED CT | 442292004 | Idiopathic hypersomnia without long sleep time |
| SNOMED CT | 442416002 | Idiopathic hypersomnia associated with long sleep time |
| SNOMED CT | 443664006 | Long sleeper syndrome |
| SNOMED CT | 443760008 | Sleep hypoventilation |
| SNOMED CT | 449160007 | Exploding head syndrome |
| SNOMED CT | 472819006 | Adjustment insomnia |
| SNOMED CT | 711565008 | Dream enactment behavior |
| SNOMED CT | 713498009 | Circadian rhythm sleep disorder of shift work type |
| SNOMED CT | 713524002 | Sleep disorder caused by reverse transcriptase inhibitor |
| SNOMED CT | 715829003 | Familial advanced sleep phase syndrome |
| SNOMED CT | 719972004 | Haddad syndrome |
| SNOMED CT | 719976001 | Glaucoma and sleep apnea syndrome |
| SNOMED CT | 73430006 | Sleep apnea |
| SNOMED CT | 77692006 | Hypersomnia |
| SNOMED CT | 78275009 | Obstructive sleep apnea syndrome |
| SNOMED CT | 79280005 | Hypersomnia with sleep apnea |
| SNOMED CT | 80495009 | Sleep walking disorder |
| SNOMED CT | 81608000 | Insomnia disorder related to known organic factor |
| SNOMED CT | 83157008 | Fatal familial insomnia |
| SNOMED CT | 88926005 | Opioid-induced sleep disorder |
| SNOMED CT | 88982005 | Rebound insomnia |
| SNOMED CT | 89415002 | Hypersomnia disorder related to another mental disorder |

Continued on next page

**TableS16 – continued from previous page**

| CodeSystem | ConceptCode | ConceptName |
|---|---|---|
| SNOMED CT | 89675003 | Sleep terror disorder |
| SNOMED CT | 111488004 | Kleine-Levin syndrome |
| SNOMED CT | 111489007 | Breathing-related sleep disorder |
| SNOMED CT | 162204000 | Late insomnia |
| SNOMED CT | 191997003 | Persistent insomnia |
| SNOMED CT | 191999000 | Persistent hypersomnia |
| SNOMED CT | 192003008 | Sleep drunkenness |
| SNOMED CT | 192004002 | Repeated rapid eye movement sleep interruptions |
| SNOMED CT | 192008004 | Reversed sleep-wake cycle |
| SNOMED CT | 192454004 | Nonorganic insomnia |
| SNOMED CT | 193042000 | Cataplexy and narcolepsy |
| SNOMED CT | 193462001 | Insomnia |
| SNOMED CT | 194437008 | Disorders of initiating and maintaining sleep |
| SNOMED CT | 194439006 | Disorder of excessive somnolence |
| SNOMED CT | 198437004 | Menopausal sleeplessness |
| SNOMED CT | 230488004 | Hypersomnia of non-organic origin |
| SNOMED CT | 230489007 | Excessive daytime sleepiness - normal night sleep |
| SNOMED CT | 230490003 | Excessive day and night-time sleepiness |
| SNOMED CT | 230491004 | Postviral excessive daytime sleepiness |
| SNOMED CT | 230492006 | Excessive daytime sleepiness with sleep paralysis |
| SNOMED CT | 230494007 | Alveolar sleep apnea |
| SNOMED CT | 230495008 | Transient sleep-wake rhythm disorder |
| SNOMED CT | 230496009 | Non-24 hour sleep-wake cycle |
| SNOMED CT | 230497000 | Sleep-related head banging |
| SNOMED CT | 230498005 | Sleep-related painful erections |
| SNOMED CT | 230499002 | Sleep-related respiratory failure |
| SNOMED CT | 230500006 | Sleep-related dystonia |
| SNOMED CT | 247951006 | Reaction to sudden wakening |
| SNOMED CT | 247962006 | Sleep automatism |
| SNOMED CT | 248256006 | Not getting enough sleep |
| SNOMED CT | 248258007 | Circumstances interfere with sleep |
| SNOMED CT | 248259004 | Symptoms interfere with sleep |
| SNOMED CT | 248260009 | Unrefreshed by sleep |
| SNOMED CT | 248261008 | Oversleeps |
| SNOMED CT | 248262001 | Always sleepy |
| SNOMED CT | 3972004 | Primary insomnia |
| SNOMED CT | 17402007 | Pavor diurnus |
| SNOMED CT | 22574000 | Cocaine-induced sleep disorder |
| SNOMED CT | 24121004 | Insomnia disorder related to another mental disorder |
| SNOMED CT | 24825006 | Central alveolar hypoventilation syndrome |
| SNOMED CT | 25753007 | Amphetamine-induced sleep disorder |
| SNOMED CT | 27405005 | Central sleep apnea syndrome |
| SNOMED CT | 31537005 | Sleep-wake schedule disorder, advanced phase type |
| SNOMED CT | 32914008 | Restless legs |
| SNOMED CT | 36124002 | Primary hypersomnia |
| SNOMED CT | 41083005 | Sleep disorder caused by ingestible alcohol |
| SNOMED CT | 41975002 | Insomnia with sleep apnea |

Continued on next page

**TableS16 – continued from previous page**

| CodeSystem | ConceptCode | ConceptName |
|---|---|---|
| SNOMED CT | 44186003 | Dyssomnia |
| SNOMED CT | 44455001 | Hypersomnia disorder related to a known organic factor |
| SNOMED CT | 54230003 | Mixed insomnia |
| SNOMED CT | 54532007 | Sleep-wake schedule disorder, frequently changing type |
| SNOMED CT | 56194001 | Sleep disorder caused by caffeine |
| SNOMED CT | 57588009 | Sedative, hypnotic AND/OR anxiolytic-induced sleep disorder |
| SNOMED CT | 58690002 | Parasomnia |
| SNOMED CT | 59050008 | Initial insomnia |
| SNOMED CT | 60380001 | Narcolepsy |
| SNOMED CT | 67062000 | Terminal insomnia |
| SNOMED CT | 67233009 | Nocturnal awakening |
| SNOMED CT | 722293005 | Autosomal dominant cerebellar ataxia, deafness and narcolepsy syndrome |
| SNOMED CT | 722963007 | Secondary restless legs syndrome |
| SNOMED CT | 723937009 | Sleep disorder caused by cannabis |
| SNOMED CT | 724504007 | Central sleep apnea caused by substance |
| SNOMED CT | 724506009 | Central sleep apnea co-occurrent with Cheyne Stokes respiration |
| SNOMED CT | 724507000 | Central sleep apnea without Cheyne-Stokes respiration |
| SNOMED CT | 724508005 | Late-onset central hypoventilation co-occurrent and due to hypothalamic dysfunction |
| SNOMED CT | 724509002 | Sleep-related hypoventilation caused by substance |
| SNOMED CT | 724699001 | Sleep disorder caused by nicotine |
| SNOMED CT | 724732000 | Sleep disorder caused by psychoactive substance |
| SNOMED CT | 724748004 | Chronic insomnia |
| SNOMED CT | 724749007 | Behaviorally induced hypersomnia |
| SNOMED CT | 724750007 | Hypersomnolence caused by substance |
| SNOMED CT | 724751006 | Sleep-wake schedule disorder due to jet lag |
| SNOMED CT | 724752004 | Hypnogogic exploding head syndrome |
| SNOMED CT | 724753009 | Recurrent isolated sleep-related hallucinations |
| SNOMED CT | 724754003 | Parasomnia disorder caused by substance |
| SNOMED CT | 735676003 | Narcolepsy type 1 |
| SNOMED CT | 737342004 | Sleep disorder caused by synthetic cannabinoid |
| SNOMED CT | 762334004 | Sleep disorder caused by stimulant |
| SNOMED CT | 762348004 | Adjustment insomnia |
| SNOMED CT | 762518003 | Sleep disorder caused by synthetic cathinone |
| SNOMED CT | 765751002 | Autoimmune encephalopathy with parasomnia and obstructive sleep apnea |
| SNOMED CT | 774068004 | AHDC1-related intellectual disability, obstructive sleep apnea, mild dysmorphism syndrome |
| SNOMED CT | 787175002 | ANK3-related intellectual disability, sleep disturbance syndrome |
| SNOMED CT | 789009003 | Primary central sleep apnea of prematurity |
| SNOMED CT | 789046004 | Sleep-related movement disorder caused by drug |
| SNOMED CT | 789047008 | Sleep-related movement disorder caused by substance |
| SNOMED CT | 789055001 | Treatment-emergent central sleep apnea |
| SNOMED CT | 1091000119108 | Obstructive sleep apnea of child |
| SNOMED CT | 1101000119103 | Obstructive sleep apnea of adult |
| SNOMED CT | 3731000119107 | Idiopathic hypersomnia |

Continued on next page

**TableS16 – continued from previous page**

| CodeSystem | ConceptCode | ConceptName |
|---|---|---|
| SNOMED CT | 4681000119100 | Sleep disturbance in infancy |
| SNOMED CT | 9741000119101 | Primary central sleep apnea |
| SNOMED CT | 23871000119106 | Somnambulism co-occurrent with sleep terror disorder |
| SNOMED CT | 27971000119105 | Insomnia co-occurrent and due to medical condition |
| SNOMED CT | 31491000119109 | Disruptions of 24 hour sleep-wake cycle |
| SNOMED CT | 31771000119102 | Daytime hypersomnia |
| SNOMED CT | 34101000119105 | Nocturnal myoclonus |
| SNOMED CT | 84991000119105 | Sleep related movement disorder |
| SNOMED CT | 85721000119105 | Central sleep apnea due to periodic breathing |
| SNOMED CT | 89911000119102 | Sleep related hypoventilation or hypoxemia |
| SNOMED CT | 90011000119105 | Organic sleep related movement disorder |
| SNOMED CT | 90361000119105 | Behavioral insomnia of childhood |
| SNOMED CT | 91411000119105 | Drug-induced insomnia |
| SNOMED CT | 91441000119109 | Central sleep apnea caused by high altitude |
| SNOMED CT | 91461000119108 | Circadian rhythm disorder caused by drug |
| SNOMED CT | 91471000119102 | Drug-induced hypersomnia |
| SNOMED CT | 91521000119104 | Narcolepsy without cataplexy |
| SNOMED CT | 91561000119109 | Parasomnia caused by drug |
| SNOMED CT | 93681000119102 | Insomnia co-occurrent and due to nocturnal myoclonus |
| SNOMED CT | 99711000119104 | Organic sleep-wake cycle disorder |
| SNOMED CT | 104831000119109 | Drug induced central sleep apnea |
| SNOMED CT | 105191000119100 | Acquired central alveolar hypoventilation |
| SNOMED CT | 105491000119107 | Primary hyposomnia |
| SNOMED CT | 105521000119109 | Organic mixed sleep disorder |
| SNOMED CT | 135301000119103 | Parasomnia caused by alcohol |
| SNOMED CT | 135311000119100 | Insomnia caused by alcohol |
| SNOMED CT | 135321000119107 | Hypersomnia caused by alcohol |
| SNOMED CT | 135331000119105 | Circadian rhythm sleep disorder caused by alcohol |
| SNOMED CT | 288081000119106 | Behavioral insomnia of childhood, combined type |
| SNOMED CT | 288091000119109 | Behavioral insomnia of childhood, limit setting type |
| SNOMED CT | 288101000119104 | Behavioral insomnia of childhood, sleep onset association type |
| SNOMED CT | 288421000119100 | Hyposomnia co-occurrent and due to psychological disorder |
| SNOMED CT | 288581000119102 | Sleep related hypoxemia |
| SNOMED CT | 434241000124107 | Secondary narcolepsy |
| SNOMED CT | 434251000124109 | Secondary narcolepsy without cataplexy |
| SNOMED CT | 434261000124106 | Secondary narcolepsy with cataplexy |
| SNOMED CT | 436001000124105 | Insomnia due to anxiety and fear |
| SNOMED CT | 12398651000119100 | Sleep disorder caused by methamphetamine |
| SNOMED CT | 16275741000119100 | Severe pediatric obstructive sleep apnea |
| SNOMED CT | 80623000 | Delayed sleep-wake phase disorder |
| SNOMED CT | 230493001 | Complex sleep apnoea |
| SNOMED CT | 399040002 | Ondine curse |
| SNOMED CT | 1052282008 | Restless leg syndrome due to uremia |
| SNOMED CT | 1052283003 | Restless leg syndrome due to iron deficiency anemia |
| SNOMED CT | 1052284009 | Restless leg syndrome in pregnancy |
| SNOMED CT | 1142084009 | Hypernychthemeral syndrome |

#### 4.14 Lightheadedness

Table S17: Concept codes used to identify if a patient was diagnosed with lightheadedness.

| CodeSystem | ConceptCode | ConceptName |
|---|---|---|
| ICD10CM | R42 | Dizziness and giddiness |
| SNOMED CT | 386705008 | Lightheadedness |

#### 4.15 Myalgia

Table S18: Concept codes used to identify if a patient was diagnosed with myalgia.

| CodeSystem | ConceptCode | ConceptName |
|---|---|---|
| ICD10CM | M79.1 | Myalgia |
| ICD10CM | M79.10 | Myalgia, unspecified site |
| ICD10CM | M79.11 | Myalgia of mastication muscle |
| ICD10CM | M79.12 | Myalgia of auxiliary muscles, head and neck |
| ICD10CM | M79.18 | Myalgia, other site |
| SNOMED CT | 68962001 | Muscle pain |
| SNOMED CT | 279030006 | Myofascial pain syndrome of neck |
| SNOMED CT | 279036000 | Myofascial pain syndrome of thorax |
| SNOMED CT | 279041008 | Myofascial pain syndrome of lower back |
| SNOMED CT | 279070004 | Muscle tension pain |
| SNOMED CT | 288225004 | Myalgia/myositis - multiple |
| SNOMED CT | 288226003 | Myalgia/myositis - shoulder |
| SNOMED CT | 288227007 | Myalgia/myositis - upper arm |
| SNOMED CT | 288228002 | Myalgia/myositis - forearm |
| SNOMED CT | 288229005 | Myalgia/myositis - hand |
| SNOMED CT | 288230000 | Myalgia/myositis -pelvis/thigh |
| SNOMED CT | 288231001 | Myalgia/myositis - lower leg |
| SNOMED CT | 288232008 | Myalgia/myositis - ankle/foot |
| SNOMED CT | 298292009 | Pain on movement of skeletal muscle |
| SNOMED CT | 403735006 | Eosinophilia-myalgia syndrome from tryptophan |
| SNOMED CT | 699682009 | Localized masticatory muscle soreness |
| SNOMED CT | 702549002 | Centrally mediated myalgia |
| SNOMED CT | 712752004 | Myalgia of pelvic floor |
| SNOMED CT | 76821002 | Epidemic cervical myalgia |
| SNOMED CT | 83264000 | Epidemic pleurodynia |
| SNOMED CT | 85879003 | Secondary fibrositis |
| SNOMED CT | 95415006 | Polymyalgia |
| SNOMED CT | 95416007 | Eosinophilia myalgia syndrome |
| SNOMED CT | 95417003 | Primary fibromyalgia syndrome |
| SNOMED CT | 95421005 | Intercostal myalgia |
| SNOMED CT | 95443002 | Venous intermittent claudication |

Continued on next page

**TableS18 – continued from previous page**

| CodeSystem | ConceptCode | ConceptName |
|---|---|---|
| SNOMED CT | 102557007 | Arm claudication |
| SNOMED CT | 240044003 | Fibrositis and nodular fasciitis |
| SNOMED CT | 240107001 | Viral myalgia |
| SNOMED CT | 30173002 | Primary fibrositis |
| SNOMED CT | 43922008 | Jaw claudication |
| SNOMED CT | 49605003 | Ophthalmoplegic migraine |
| SNOMED CT | 54981004 | Charleyhorse |
| SNOMED CT | 56557000 | Fibrositis |
| SNOMED CT | 792845002 | Intermittent claudication due to atherosclerosis of artery of limb |
| SNOMED CT | 28221000119103 | Abdominal muscle pain |
| SNOMED CT | 41321000119101 | Myofascial pain syndrome of thoracic spine |
| SNOMED CT | 113611000119100 | Myofascial pain syndrome of lumbar spine |
| SNOMED CT | 301161000119100 | Repetitive strain injury of left ankle |
| SNOMED CT | 301171000119106 | Repetitive strain injury of left foot |
| SNOMED CT | 301221000119101 | Repetitive strain injury of left thigh |
| SNOMED CT | 301241000119107 | Repetitive strain injury of right ankle |
| SNOMED CT | 301251000119109 | Repetitive strain injury of right foot |
| SNOMED CT | 301301000119103 | Repetitive strain injury of right thigh |
| SNOMED CT | 431061000124100 | Ophthalmoplegic migraine, refractory |
| SNOMED CT | 12236951000119108 | Intermittent claudication of bilateral lower limbs co-occurrent and due to atherosclerosis |
| SNOMED CT | 12237071000119100 | Intermittent claudication of left lower limb co-occurrent and due to atherosclerosis |
| SNOMED CT | 12237191000119103 | Intermittent claudication of right lower limb co-occurrent and due to atherosclerosis |
| SNOMED CT | 15704721000119106 | Repetitive strain injury of bilateral hands |
| SNOMED CT | 15704761000119101 | Repetitive strain injury of bilateral forearms |
| SNOMED CT | 16013711000119104 | Intermittent claudication of right lower limb co-occurrent and due to atherosclerosis of bypass graft |
| SNOMED CT | 16014031000119109 | Atherosclerosis of bypass graft of left lower limb with intermittent claudication |
| SNOMED CT | 16014071000119107 | Atherosclerosis of bypass graft of bilateral lower limbs with intermittent claudication |
| SNOMED CT | 16462851000119106 | Myalgia caused by statin |
| SNOMED CT | 63491006 | Claudication |
| SNOMED CT | 726531007 | MPS - myofascial pain syndrome |
| SNOMED CT | 16013431000119104 | Bilateral intermittent claudication of lower limbs due to atherosclerosis of nonbiological bypass graft |
| Truveta | 1215966 | Post-COVID-19 chronic muscle pain |
| SNOMED CT | 610921000124106 | Pain of erector spinae muscle |

#### 4.16 Palpitations/tachycardia

Table S19: Concept codes used to identify if a patient was diagnosed with heart palpitations and/or tachycardia.

| CodeSystem | ConceptCode | ConceptName |
|---|---|---|
| ICD10CM | I47 | Paroxysmal tachycardia |
| ICD10CM | R00.0 | Tachycardia, unspecified |
| ICD10CM | R00.2 | Palpitations |
| ICD10CM | I47.0 | Re-entry ventricular arrhythmia |
| ICD10CM | I47.1 | Supraventricular tachycardia |
| ICD10CM | I47.2 | Ventricular tachycardia |
| ICD10CM | I47.9 | Paroxysmal tachycardia, unspecified |
| SNOMED CT | 80313002 | Palpitations |
| SNOMED CT | 3424008 | Tachycardia |
| SNOMED CT | 278086000 | Baseline tachycardia |
| SNOMED CT | 278482008 | Atrioventricular tachycardia |
| SNOMED CT | 282825002 | Paroxysmal atrial fibrillation |
| SNOMED CT | 300996004 | Controlled atrial fibrillation |
| SNOMED CT | 314208002 | Rapid atrial fibrillation |
| SNOMED CT | 405806007 | Pulseless ventricular tachycardia |
| SNOMED CT | 413342000 | Neonatal tachycardia |
| SNOMED CT | 418493005 | Right atrial incisional tachycardia |
| SNOMED CT | 418872003 | Fascicular ventricular tachycardia |
| SNOMED CT | 419400008 | Incisional tachycardia |
| SNOMED CT | 419671004 | Catecholaminergic polymorphic ventricular tachycardia |
| SNOMED CT | 419752005 | Sinoatrial nodal reentrant tachycardia |
| SNOMED CT | 420002000 | Left atrial incisional tachycardia |
| SNOMED CT | 421348006 | Tachycardia - baseline fetal heart rate |
| SNOMED CT | 425582007 | Inappropriate sinus tachycardia |
| SNOMED CT | 426525004 | Sustained ventricular tachycardia |
| SNOMED CT | 426749004 | Chronic atrial fibrillation |
| SNOMED CT | 428919002 | Palpitations with regular rhythm |
| SNOMED CT | 440028005 | Permanent atrial fibrillation |
| SNOMED CT | 440059007 | Persistent atrial fibrillation |
| SNOMED CT | 442515000 | Reflex tachycardia |
| SNOMED CT | 444658006 | Nonsustained ventricular tachycardia |
| SNOMED CT | 462169004 | Fetal supraventricular tachycardia with short ventriculoatrial interval |
| SNOMED CT | 462170003 | Fetal supraventricular tachycardia with long ventriculoatrial interval |
| SNOMED CT | 706923002 | Longstanding persistent atrial fibrillation |
| SNOMED CT | 708124001 | Recurrent ventricular tachycardia |
| SNOMED CT | 709065005 | Postoperative fluttering heart |
| SNOMED CT | 710878005 | Mahaim fiber tachycardia |
| SNOMED CT | 715395008 | Familial atrial fibrillation |
| SNOMED CT | 718104007 | Torsade de pointes with short coupling interval syndrome |
| SNOMED CT | 74615001 | Tachycardia-bradycardia |
| SNOMED CT | 82838007 | Irregular tachycardia |
| SNOMED CT | 102590007 | Intermittent palpitations |
| SNOMED CT | 111288001 | Ventricular flutter |

Continued on next page

**TableS19 – continued from previous page**

| CodeSystem | ConceptCode | ConceptName |
|---|---|---|
| SNOMED CT | 161968007 | Bumping heart |
| SNOMED CT | 161969004 | Fluttering heart |
| SNOMED CT | 162992001 | O/E - pulse rate tachycardia |
| SNOMED CT | 195069001 | Atrial paroxysmal tachycardia |
| SNOMED CT | 195070000 | Paroxysmal atrioventricular tachycardia |
| SNOMED CT | 195071001 | Paroxysmal junctional tachycardia |
| SNOMED CT | 195072008 | Paroxysmal nodal tachycardia |
| SNOMED CT | 195080001 | Atrial fibrillation and flutter |
| SNOMED CT | 195083004 | Ventricular fibrillation and flutter |
| SNOMED CT | 195105007 | Re-entry ventricular arrhythmia |
| SNOMED CT | 233893007 | Re-entrant atrial tachycardia |
| SNOMED CT | 233895000 | Ectopic atrioventricular node tachycardia |
| SNOMED CT | 233896004 | Re-entrant atrioventricular node tachycardia |
| SNOMED CT | 233897008 | Re-entrant atrioventricular tachycardia |
| SNOMED CT | 233898003 | Orthodromic atrioventricular re-entrant tachycardia |
| SNOMED CT | 233899006 | Antidromic atrioventricular re-entrant tachycardia |
| SNOMED CT | 233900001 | Supraventricular tachycardia with functional bundle branch block |
| SNOMED CT | 233901002 | His bundle tachycardia |
| SNOMED CT | 233902009 | Congenital His bundle tachycardia |
| SNOMED CT | 233903004 | Postoperative His bundle tachycardia |
| SNOMED CT | 233905006 | Ventricular tachycardia with normal heart |
| SNOMED CT | 233906007 | Familial ventricular tachycardia |
| SNOMED CT | 233907003 | Induced ventricular tachycardia |
| SNOMED CT | 233908008 | Incessant infant ventricular tachycardia |
| SNOMED CT | 233909000 | Right ventricular outflow tract ventricular tachycardia |
| SNOMED CT | 233910005 | Lone atrial fibrillation |
| SNOMED CT | 233911009 | Non-rheumatic atrial fibrillation |
| SNOMED CT | 234225006 | Cardiac pacemaker re-entrant tachycardia |
| SNOMED CT | 240298005 | Fetal tachycardia |
| SNOMED CT | 248648003 | Palpitations - rapid |
| SNOMED CT | 248657009 | Pounding heart |
| SNOMED CT | 251155001 | Junctional ectopic tachycardia |
| SNOMED CT | 251156000 | Wide QRS ventricular tachycardia |
| SNOMED CT | 251157009 | Narrow QRS ventricular tachycardia |
| SNOMED CT | 251158004 | Ventricular tachycardia, monomorphic |
| SNOMED CT | 251159007 | Ventricular tachycardia, polymorphic |
| SNOMED CT | 251160002 | Ventricular tachycardia, polymorphic without Q-T prolongation |
| SNOMED CT | 251165007 | AV junctional (nodal) tachycardia |
| SNOMED CT | 251166008 | AV nodal re-entry tachycardia |
| SNOMED CT | 4006006 | Fetal tachycardia affecting management of mother |
| SNOMED CT | 6285003 | Tachyarrhythmia |
| SNOMED CT | 6456007 | Supraventricular tachycardia |
| SNOMED CT | 6624005 | Ventricular tachyarrhythmia |
| SNOMED CT | 11092001 | Sinus tachycardia |
| SNOMED CT | 12026006 | Paroxysmal tachycardia |
| SNOMED CT | 25569003 | Ventricular tachycardia |

Continued on next page

**TableS19 – continued from previous page**

| CodeSystem | ConceptCode | ConceptName |
|---|---|---|
| SNOMED CT | 31722008 | Torsades de pointes |
| SNOMED CT | 39357005 | Paroxysmal atrial tachycardia with block |
| SNOMED CT | 49436004 | Atrial fibrillation |
| SNOMED CT | 66657009 | Paroxysmal ventricular tachycardia |
| SNOMED CT | 67198005 | Paroxysmal supraventricular tachycardia |
| SNOMED CT | 725060001 | Torsades de pointe caused by drug |
| SNOMED CT | 735682000 | Macro re-entrant atrial tachycardia |
| SNOMED CT | 735683005 | Cavotricuspid isthmus dependent macroreentry tachycardia |
| SNOMED CT | 735685003 | Atrioventricular reciprocating tachycardia |
| SNOMED CT | 762247006 | Preexcited atrial fibrillation |
| SNOMED CT | 789039008 | Scar mediated macro re-entrant atrial tachycardia |
| SNOMED CT | 870695003 | Fetal heart baseline rate range 180 to 200 |
| SNOMED CT | 870696002 | Fetal heart baseline rate range above 200 |
| SNOMED CT | 870700005 | Fetal heart baseline rate range 160 to 180 |
| SNOMED CT | 5761000119100 | Nonsustained paroxysmal supraventricular tachycardia |
| SNOMED CT | 21421000119109 | Nonsustained paroxysmal ventricular tachycardia |
| SNOMED CT | 120041000119109 | Atrial fibrillation with rapid ventricular response |
| SNOMED CT | 39260000 | Nonparoxysmal atrioventricular nodal tachycardia |
| SNOMED CT | 1010405004 | Paroxysmal atrial fibrillation with rapid ventricular response |
| SNOMED CT | 1142040009 | Right atrial non-cavotricuspid isthmus dependent macro re-entrant atrial tachycardia |
| SNOMED CT | 1142041008 | Non-scar mediated right atrial non-cavotricuspid isthmus dependent macro re-entrant atrial tachycardia |
| SNOMED CT | 1142057008 | Anticlockwise cavotricuspid isthmus dependent macroreentry tachycardia |
| SNOMED CT | 1142064005 | Antidromic atrioventricular reciprocating tachycardia utilizing atrio-ventricular accessory pathway with antegrade unidirectional conduction |
| SNOMED CT | 1142118003 | Atypical atrioventricular nodal re-entry tachycardia |
| SNOMED CT | 1142066007 | Antidromic atrioventricular reciprocating tachycardia utilizing atrio-fascicular accessory pathway with antegrade unidirectional conduction |
| SNOMED CT | 1142067003 | Non scar mediated left atrial non-cavotricuspid isthmus dependent macro re-entrant atrial tachycardia |
| SNOMED CT | 1142068008 | Antidromic atrioventricular reciprocating tachycardia utilizing accessory pathway with antegrade unidirectional conduction |
| SNOMED CT | 1142069000 | Scar mediated left atrial non-cavotricuspid isthmus dependent macro re-entrant atrial tachycardia |
| SNOMED CT | 1142082008 | Idiopathic junctional ectopic tachycardia |
| SNOMED CT | 1142086006 | Congenital junctional ectopic tachycardia |
| SNOMED CT | 1142090008 | Clockwise cavotricuspid isthmus dependent macroreentry tachycardia |
| SNOMED CT | 1142093005 | Left atrial non-cavotricuspid isthmus dependent macro re-entrant atrial tachycardia |
| SNOMED CT | 1142098001 | Postoperative junctional ectopic tachycardia |

Continued on next page

**TableS19 – continued from previous page**

| CodeSystem | ConceptCode | ConceptName |
|---|---|---|
| SNOMED CT | 1142103004 | Antidromic atrioventricular reciprocating tachycardia utilizing nodo-fascicular accessory pathway with antegrade unidirectional conduction |
| SNOMED CT | 1142105006 | Typical atrioventricular nodal re-entry tachycardia |
| SNOMED CT | 1142114001 | Scar mediated right atrial non-cavotricuspid isthmus dependent macro re-entrant atrial tachycardia |
| SNOMED CT | 1142117008 | Atypical slow slow atrioventricular nodal re-entry tachycardia |
| SNOMED CT | 1142119006 | Atypical fast slow atrioventricular nodal re-entry tachycardia |
| SNOMED CT | 1142120000 | Orthodromic atrioventricular reciprocating tachycardia utilizing manifest accessory pathway |
| SNOMED CT | 1142121001 | Atrioventricular nodal reentry tachycardia with twin atrioventricular nodes |
| SNOMED CT | 1142122008 | Antidromic atrioventricular reciprocating tachycardia utilizing accessory pathway with bidirectional conduction |
| SNOMED CT | 1142204001 | Orthodromic atrioventricular reciprocating tachycardia utilizing concealed accessory pathway |
| SNOMED CT | 16415081000119104 | Supraventricular tachycardia following acute myocardial infarction |
| Truveta | 1215968 | Post-COVID-19 chronic palpitations |
| SNOMED CT | 1220643007 | Regular wide QRS complex tachycardia |
| SNOMED CT | 1204167003 | Catecholaminergic polymorphic ventricular tachycardia due to calsequestrin mutation |
| SNOMED CT | 1204129001 | Catecholaminergic polymorphic ventricular tachycardia due to RYR2 mutation |
| SNOMED CT | 1197418004 | Familial atrial tachyarrhythmia, infra-Hisian cardiac conduction disease |
| SNOMED CT | 1204205002 | Left outflow tract ventricular tachycardia |
| SNOMED CT | 1208899000 | Mitral annular ventricular tachycardia |
| SNOMED CT | 1208832006 | Pacemaker mediated tachycardia |
| SNOMED CT | 1204171000 | Bidirectional ventricular tachycardia |
| SNOMED CT | 1163438008 | Idiopathic fascicular ventricular tachycardia |
| SNOMED CT | 1163437003 | Idiopathic ventricular tachycardia |

#### 4.17 Paresthesia

Table S20: Concept codes used to identify if a patient was diagnosed with paresthesia.

| CodeSystem | ConceptCode | ConceptName |
|---|---|---|
| ICD10CM | R20.2 | Paresthesia of skin |
| SNOMED CT | 91019004 | Paresthesia |
| SNOMED CT | 309086004 | Paresthesia of hand |
| SNOMED CT | 309087008 | Paresthesia of foot |
| SNOMED CT | 310481001 | C/O paresthesia |
| SNOMED CT | 403600002 | Formication of skin |

Continued on next page

**TableS20 – continued from previous page**

| CodeSystem | ConceptCode | ConceptName |
|---|---|---|
| SNOMED CT | 429783005 | Paresthesia of lower extremity |
| SNOMED CT | 79256006 | Acroparesthesia |
| SNOMED CT | 95358009 | Paresthesia of mucous membrane |
| SNOMED CT | 95665007 | Facial paresthesia |
| SNOMED CT | 95673003 | Paresthesia of upper limb |
| SNOMED CT | 135873002 | Transient paresthesia |
| SNOMED CT | 163707002 | O/E - paresthesia present |
| SNOMED CT | 163708007 | O/E - paresthesia in hands |
| SNOMED CT | 57105000 | Simple acroparesthesia |
| SNOMED CT | 62507009 | Pins and needles |
| SNOMED CT | 762374005 | Paresthesia of finger |
| SNOMED CT | 840319006 | Paresthesia of bilateral hands |
| SNOMED CT | 101000119102 | Numbness and tingling sensation of skin |
| SNOMED CT | 15634791000119106 | Paresthesia of right lower limb |
| SNOMED CT | 15634841000119109 | Paresthesia of left lower limb |
| SNOMED CT | 15973661000119106 | Paresthesia of left upper limb |
| SNOMED CT | 15973701000119104 | Paresthesia of right upper limb |
| SNOMED CT | 1187255000 | Paresthesia of saddle area |
| SNOMED CT | 1172701002 | Facial diplegia with paresthesia |

##### 4.18 Post-exertional malaise

Table S21: Concept codes used to identify if a patient was diagnosed with post-exertional malaise.

| CodeSystem | ConceptCode | ConceptName |
|---|---|---|
| ICD10CM | R53.8 | Other malaise and fatigue |
| ICD10CM | R53.81 | Other malaise |
| ICD10CM | R53.82 | Chronic fatigue, unspecified |
| ICD10CM | R53.83 | Other fatigue |
| SNOMED CT | 248271005 | Exhausted on least exertion |

#### 5 Comorbid conditions

##### 5.1 Anxiety

Table S22: Concept codes used to identify if a patient was diagnosed with anxiety.

| CodeSystem | ConceptCode | ConceptName |
|---|---|---|
| ICD10CM | F41.0 | Panic disorder [episodic paroxysmal anxiety] |
| ICD10CM | F41.1 | Generalized anxiety disorder |

Continued on next page

**TableS22 – continued from previous page**

| CodeSystem | ConceptCode | ConceptName |
|---|---|---|
| ICD10CM | F41.3 | Other mixed anxiety disorders |
| ICD10CM | F41.8 | Other specified anxiety disorders |
| ICD10CM | F41.9 | Anxiety disorder, unspecified |
| SNOMED CT | 279611005 | Shell shock |
| SNOMED CT | 313182004 | Chronic post-traumatic stress disorder |
| SNOMED CT | 317816007 | Stockholm syndrome |
| SNOMED CT | 318784009 | Posttraumatic stress disorder, delayed onset |
| SNOMED CT | 371631005 | Panic disorder |
| SNOMED CT | 386810004 | Phobic disorder |
| SNOMED CT | 403593004 | Phobic fear of skin cancer |
| SNOMED CT | 426174008 | Chronic stress disorder |
| SNOMED CT | 428687006 | Nightmares associated with chronic post-traumatic stress disorder |
| SNOMED CT | 443919007 | Complex posttraumatic stress disorder |
| SNOMED CT | 446175003 | Acute posttraumatic stress disorder following military combat |
| SNOMED CT | 446180007 | Delayed posttraumatic stress disorder following military combat |
| SNOMED CT | 699241002 | Chronic post-traumatic stress disorder following military combat |
| SNOMED CT | 69479009 | Anxiety hyperventilation |
| SNOMED CT | 70691001 | Agoraphobia |
| SNOMED CT | 72861004 | Panic disorder without agoraphobia with mild panic attacks |
| SNOMED CT | 74010007 | Panic disorder with agoraphobia, severe agoraphobic avoidance AND panic attacks in partial remission |
| SNOMED CT | 76812003 | Panic disorder with agoraphobia, moderate agoraphobic avoidance AND panic attacks in partial remission |
| SNOMED CT | 76868007 | Panic disorder with agoraphobia, agoraphobic avoidance in full remission AND mild panic attacks |
| SNOMED CT | 82339009 | Anxiety disorder caused by amfetamine |
| SNOMED CT | 82415003 | Agoraphobia without history of panic disorder without limited symptom attacks |
| SNOMED CT | 82494000 | Panic disorder without agoraphobia with panic attacks in full remission |
| SNOMED CT | 82738004 | Panic disorder with agoraphobia, moderate agoraphobic avoidance AND moderate panic attacks |
| SNOMED CT | 83253003 | Shyness disorder of childhood |
| SNOMED CT | 83631006 | Panic disorder with agoraphobia, moderate agoraphobic avoidance AND severe panic attacks |
| SNOMED CT | 85061001 | Separation anxiety disorder of childhood, early onset |
| SNOMED CT | 87798009 | Panic disorder with agoraphobia, agoraphobic avoidance in full remission AND moderate panic attacks |
| SNOMED CT | 89948007 | Panic disorder with agoraphobia AND mild panic attacks |
| SNOMED CT | 90790003 | Avoidant disorder of adolescence |
| SNOMED CT | 111487009 | Dream anxiety disorder |
| SNOMED CT | 111490003 | Panic disorder with agoraphobia, agoraphobic avoidance in partial remission AND severe panic attacks |
| SNOMED CT | 111491004 | Panic disorder with agoraphobia, agoraphobic avoidance in full remission AND panic attacks in partial remission |
| SNOMED CT | 126943008 | Separation anxiety |

Continued on next page

**TableS22 – continued from previous page**

| CodeSystem | ConceptCode | ConceptName |
|---|---|---|
| SNOMED CT | 191722009 | Agoraphobia with panic attacks |
| SNOMED CT | 191724005 | Social phobia, fear of eating in public |
| SNOMED CT | 191725006 | Social phobia, fear of public speaking |
| SNOMED CT | 191726007 | Social phobia, fear of public washing |
| SNOMED CT | 191736004 | Obsessive-compulsive disorder |
| SNOMED CT | 191737008 | Compulsive neurosis |
| SNOMED CT | 191738003 | Obsessional neurosis |
| SNOMED CT | 192014006 | Psychogenic rumination |
| SNOMED CT | 192037000 | Acute panic state due to acute stress reaction |
| SNOMED CT | 192038005 | Acute fugue state due to acute stress reaction |
| SNOMED CT | 192039002 | Acute stupor state due to acute stress reaction |
| SNOMED CT | 192041001 | Acute situational disturbance |
| SNOMED CT | 192042008 | Acute post-trauma stress state |
| SNOMED CT | 192044009 | Stress reaction causing mixed disturbance of emotion and conduct |
| SNOMED CT | 192611004 | Childhood phobic anxiety disorder |
| SNOMED CT | 197480006 | Anxiety disorder |
| SNOMED CT | 231504006 | Mixed anxiety and depressive disorder |
| SNOMED CT | 238965007 | Venereophobia |
| SNOMED CT | 238966008 | Syphilophobia |
| SNOMED CT | 238976006 | Bromisodrophobia |
| SNOMED CT | 1380006 | Agoraphobia without history of panic disorder with limited symptom attacks |
| SNOMED CT | 1686006 | Sedative, hypnotic AND/OR anxiolytic-induced anxiety disorder |
| SNOMED CT | 1816003 | Panic disorder with agoraphobia, severe agoraphobic avoidance AND mild panic attacks |
| SNOMED CT | 3158007 | Panic disorder with agoraphobia, agoraphobic avoidance in partial remission AND panic attacks in partial remission |
| SNOMED CT | 109006 | Anxiety disorder of childhood OR adolescence |
| SNOMED CT | 4932002 | Panic disorder with agoraphobia, moderate agoraphobic avoidance AND mild panic attacks |
| SNOMED CT | 5509004 | Panic disorder with agoraphobia AND severe panic attacks |
| SNOMED CT | 8185002 | Panic disorder with agoraphobia AND moderate panic attacks |
| SNOMED CT | 10586006 | Occupation-related stress disorder |
| SNOMED CT | 11806006 | Separation anxiety disorder of childhood |
| SNOMED CT | 11941006 | Panic disorder with agoraphobia, agoraphobic avoidance in full remission AND panic attacks in full remission |
| SNOMED CT | 13438001 | Overanxious disorder of childhood |
| SNOMED CT | 17496003 | Organic anxiety disorder |
| SNOMED CT | 19766004 | Panic disorder with agoraphobia, mild agoraphobic avoidance AND severe panic attacks |
| SNOMED CT | 21897009 | Generalized anxiety disorder |
| SNOMED CT | 22230001 | Panic disorder with agoraphobia, agoraphobic avoidance in partial remission AND panic attacks in full remission |
| SNOMED CT | 24781009 | Panic disorder with agoraphobia, mild agoraphobic avoidance AND panic attacks in full remission |
| SNOMED CT | 25501002 | Social phobia |

Continued on next page

**TableS22 – continued from previous page**

| CodeSystem | ConceptCode | ConceptName |
|---|---|---|
| SNOMED CT | 30059008 | Panic disorder with agoraphobia, severe agoraphobic avoidance AND moderate panic attacks |
| SNOMED CT | 31781004 | Panic disorder with agoraphobia, agoraphobic avoidance in partial remission AND mild panic attacks |
| SNOMED CT | 32388005 | Panic disorder with agoraphobia, agoraphobic avoidance in partial remission AND moderate panic attacks |
| SNOMED CT | 34116005 | Panic disorder with agoraphobia, agoraphobic avoidance in full remission AND severe panic attacks |
| SNOMED CT | 34938008 | Anxiety disorder caused by alcohol |
| SNOMED CT | 35607004 | Panic disorder with agoraphobia |
| SNOMED CT | 37868008 | Anxiety disorder of adolescence |
| SNOMED CT | 37872007 | Avoidant disorder of childhood OR adolescence |
| SNOMED CT | 38328002 | Panic disorder with agoraphobia, severe agoraphobic avoidance AND panic attacks in full remission |
| SNOMED CT | 43150009 | Panic disorder without agoraphobia with severe panic attacks |
| SNOMED CT | 47505003 | Posttraumatic stress disorder |
| SNOMED CT | 49564006 | Panic disorder with agoraphobia, mild agoraphobic avoidance AND moderate panic attacks |
| SNOMED CT | 50026000 | Organic anxiety disorder caused by psychoactive substance |
| SNOMED CT | 50983008 | Panic disorder with agoraphobia, mild agoraphobic avoidance AND panic attacks in partial remission |
| SNOMED CT | 51493001 | Anxiety disorder caused by cocaine |
| SNOMED CT | 52910006 | Anxiety disorder due to a general medical condition |
| SNOMED CT | 53467004 | Anxiety disorder of childhood |
| SNOMED CT | 53956006 | Panic disorder without agoraphobia with panic attacks in partial remission |
| SNOMED CT | 54587008 | Simple phobia |
| SNOMED CT | 55967005 | Anxiety disorder caused by phencyclidine |
| SNOMED CT | 56576003 | Panic disorder without agoraphobia |
| SNOMED CT | 58535001 | Physical AND emotional exhaustion state |
| SNOMED CT | 59923000 | Panic disorder with agoraphobia AND panic attacks in full remission |
| SNOMED CT | 61157009 | Combat fatigue |
| SNOMED CT | 61212007 | Panic disorder with agoraphobia, severe agoraphobic avoidance AND severe panic attacks |
| SNOMED CT | 61569007 | Agoraphobia without history of panic disorder |
| SNOMED CT | 62351001 | Generalized social phobia |
| SNOMED CT | 63701002 | Panic disorder with agoraphobia, mild agoraphobic avoidance AND mild panic attacks |
| SNOMED CT | 63909006 | Panic disorder with agoraphobia AND panic attacks in partial remission |
| SNOMED CT | 64060000 | Panic disorder with agoraphobia, moderate agoraphobic avoidance AND panic attacks in full remission |
| SNOMED CT | 64165008 | Avoidant disorder of childhood |
| SNOMED CT | 65064003 | Panic disorder without agoraphobia with moderate panic attacks |
| SNOMED CT | 67195008 | Acute stress disorder |
| SNOMED CT | 723913009 | Olfactory reference disorder |

Continued on next page

**TableS22 – continued from previous page**

| CodeSystem | ConceptCode | ConceptName |
|---|---|---|
| SNOMED CT | 724654009 | Anxiety disorder caused by opioid |
| SNOMED CT | 724693000 | Obsessive compulsive disorder caused by cocaine |
| SNOMED CT | 724708007 | Anxiety disorder caused by methylenedioxymethamphetamine |
| SNOMED CT | 724722007 | Anxiety disorder caused by dissociative drug |
| SNOMED CT | 724723002 | Anxiety disorder caused by ketamine |
| SNOMED CT | 724730008 | Obsessive compulsive disorder caused by psychoactive substance |
| SNOMED CT | 737341006 | Anxiety disorder caused by synthetic cannabinoid |
| SNOMED CT | 762331007 | Anxiety disorder caused by stimulant |
| SNOMED CT | 762332000 | Obsessive compulsive disorder caused by stimulant |
| SNOMED CT | 762515000 | Anxiety disorder caused by synthetic cathinone |
| SNOMED CT | 762516004 | Obsessive compulsive disorder caused by synthetic cathinone |
| SNOMED CT | 22621000119103 | Anxiety disorder caused by drug |
| SNOMED CT | 10743001000119103 | Anxiety disorder in mother complicating childbirth |
| SNOMED CT | 12398201000119102 | Anxiety disorder caused by methamphetamine |
| SNOMED CT | 16264621000119109 | Recurrent mild major depressive disorder co-occurrent with anxiety |
| SNOMED CT | 16264821000119108 | Recurrent severe major depressive disorder co-occurrent with anxiety |
| SNOMED CT | 16264901000119109 | Recurrent moderate major depressive disorder co-occurrent with anxiety |
| SNOMED CT | 16265061000119105 | Recurrent major depressive disorder co-occurrent with anxiety in full remission |
| SNOMED CT | 16265301000119106 | Recurrent major depressive disorder in partial remission co-occurrent with anxiety |
| SNOMED CT | 16265951000119109 | Mild major depressive disorder co-occurrent with anxiety single episode |
| SNOMED CT | 16266831000119100 | Moderate major depressive disorder co-occurrent with anxiety single episode |
| SNOMED CT | 16266991000119108 | Severe major depressive disorder co-occurrent with anxiety single episode |

#### 5.2 Cardiovascular disease

Table S23: Concept codes used to identify if a patient was diagnosed with cardiovascular disease.

| CodeSystem | ConceptCode | ConceptName |
|---|---|---|
| ICD10CM | A36.81 | Diphtheritic cardiomyopathy |
| ICD10CM | A38.1 | Scarlet fever with myocarditis |
| ICD10CM | A39.50 | Meningococcal carditis, unspecified |
| ICD10CM | A39.51 | Meningococcal endocarditis |
| ICD10CM | A39.52 | Meningococcal myocarditis |
| ICD10CM | A39.53 | Meningococcal pericarditis |
| ICD10CM | B26.82 | Mumps myocarditis |
| ICD10CM | B33.20 | Viral carditis, unspecified |

Continued on next page

**TableS23 – continued from previous page**

| CodeSystem | ConceptCode | ConceptName |
|---|---|---|
| ICD10CM | B33.21 | Viral endocarditis |
| ICD10CM | B33.22 | Viral myocarditis |
| ICD10CM | B33.23 | Viral pericarditis |
| ICD10CM | B33.24 | Viral cardiomyopathy |
| ICD10CM | B37.6 | Candidal endocarditis |
| ICD10CM | B58.81 | Toxoplasma myocarditis |
| ICD10CM | G45 | Transient cerebral ischemic attacks and related syndromes |
| ICD10CM | G45.0 | Vertebro-basilar artery syndrome |
| ICD10CM | G45.1 | Carotid artery syndrome (hemispheric) |
| ICD10CM | G45.2 | Multiple and bilateral precerebral artery syndromes |
| ICD10CM | G45.3 | Amaurosis fugax |
| ICD10CM | G45.4 | Transient global amnesia |
| ICD10CM | G45.8 | Other transient cerebral ischemic attacks and related syndromes |
| ICD10CM | G45.9 | Transient cerebral ischemic attack, unspecified |
| ICD10CM | I01.0 | Acute rheumatic pericarditis |
| ICD10CM | I01.1 | Acute rheumatic endocarditis |
| ICD10CM | I01.2 | Acute rheumatic myocarditis |
| ICD10CM | I01.8 | Other acute rheumatic heart disease |
| ICD10CM | I01.9 | Acute rheumatic heart disease, unspecified |
| ICD10CM | I02.0 | Rheumatic chorea with heart involvement |
| ICD10CM | I05.0 | Rheumatic mitral stenosis |
| ICD10CM | I05.1 | Rheumatic mitral insufficiency |
| ICD10CM | I05.2 | Rheumatic mitral stenosis with insufficiency |
| ICD10CM | I05.8 | Other rheumatic mitral valve diseases |
| ICD10CM | I05.9 | Rheumatic mitral valve disease, unspecified |
| ICD10CM | I06.0 | Rheumatic aortic stenosis |
| ICD10CM | I06.1 | Rheumatic aortic insufficiency |
| ICD10CM | I06.2 | Rheumatic aortic stenosis with insufficiency |
| ICD10CM | I06.8 | Other rheumatic aortic valve diseases |
| ICD10CM | I06.9 | Rheumatic aortic valve disease, unspecified |
| ICD10CM | I07.0 | Rheumatic tricuspid stenosis |
| ICD10CM | I07.1 | Rheumatic tricuspid insufficiency |
| ICD10CM | I07.2 | Rheumatic tricuspid stenosis and insufficiency |
| ICD10CM | I07.8 | Other rheumatic tricuspid valve diseases |
| ICD10CM | I07.9 | Rheumatic tricuspid valve disease, unspecified |
| ICD10CM | I08.0 | Rheumatic disorders of both mitral and aortic valves |
| ICD10CM | I08.1 | Rheumatic disorders of both mitral and tricuspid valves |
| ICD10CM | I08.2 | Rheumatic disorders of both aortic and tricuspid valves |
| ICD10CM | I08.3 | Combined rheumatic disorders of mitral, aortic and tricuspid valves |
| ICD10CM | I08.8 | Other rheumatic multiple valve diseases |
| ICD10CM | I08.9 | Rheumatic multiple valve disease, unspecified |
| ICD10CM | I09.0 | Rheumatic myocarditis |
| ICD10CM | I09.1 | Rheumatic diseases of endocardium, valve unspecified |
| ICD10CM | I09.2 | Chronic rheumatic pericarditis |
| ICD10CM | I09.89 | Other specified rheumatic heart diseases |
| ICD10CM | I09.9 | Rheumatic heart disease, unspecified |

Continued on next page

**TableS23 – continued from previous page**

| CodeSystem | ConceptCode | ConceptName |
|---|---|---|
| ICD10CM | I20.0 | Unstable angina |
| ICD10CM | I20.1 | Angina pectoris with documented spasm |
| ICD10CM | I20.8 | Other forms of angina pectoris |
| ICD10CM | I20.9 | Angina pectoris, unspecified |
| ICD10CM | I21.01 | ST elevation (STEMI) myocardial infarction involving left main coronary artery |
| ICD10CM | I21.02 | ST elevation (STEMI) myocardial infarction involving left anterior descending coronary artery |
| ICD10CM | I21.09 | ST elevation (STEMI) myocardial infarction involving other coronary artery of anterior wall |
| ICD10CM | I21.11 | ST elevation (STEMI) myocardial infarction involving right coronary artery |
| ICD10CM | I21.19 | ST elevation (STEMI) myocardial infarction involving other coronary artery of inferior wall |
| ICD10CM | I21.21 | ST elevation (STEMI) myocardial infarction involving left circumflex coronary artery |
| ICD10CM | I21.29 | ST elevation (STEMI) myocardial infarction involving other sites |
| ICD10CM | I21.3 | ST elevation (STEMI) myocardial infarction of unspecified site |
| ICD10CM | I21.4 | Non-ST elevation (NSTEMI) myocardial infarction |
| ICD10CM | I21.9 | Acute myocardial infarction, unspecified |
| ICD10CM | I21.A1 | Myocardial infarction type 2 |
| ICD10CM | I21.A9 | Other myocardial infarction type |
| ICD10CM | I22.0 | Subsequent ST elevation (STEMI) myocardial infarction of anterior wall |
| ICD10CM | I22.1 | Subsequent ST elevation (STEMI) myocardial infarction of inferior wall |
| ICD10CM | I22.2 | Subsequent non-ST elevation (NSTEMI) myocardial infarction |
| ICD10CM | I22.8 | Subsequent ST elevation (STEMI) myocardial infarction of other sites |
| ICD10CM | I22.9 | Subsequent ST elevation (STEMI) myocardial infarction of unspecified site |
| ICD10CM | I23.0 | Hemopericardium as current complication following acute myocardial infarction |
| ICD10CM | I23.1 | Atrial septal defect as current complication following acute myocardial infarction |
| ICD10CM | I23.2 | Ventricular septal defect as current complication following acute myocardial infarction |
| ICD10CM | I23.3 | Rupture of cardiac wall without hemopericardium as current complication following acute myocardial infarction |
| ICD10CM | I23.4 | Rupture of chordae tendineae as current complication following acute myocardial infarction |
| ICD10CM | I23.5 | Rupture of papillary muscle as current complication following acute myocardial infarction |
| ICD10CM | I23.6 | Thrombosis of atrium, auricular appendage, and ventricle as current complications following acute myocardial infarction |
| ICD10CM | I23.7 | Postinfarction angina |

Continued on next page

**TableS23 – continued from previous page**

| CodeSystem | ConceptCode | ConceptName |
|---|---|---|
| ICD10CM | I23.8 | Other current complications following acute myocardial infarction |
| ICD10CM | I24 | Other acute ischemic heart diseases |
| ICD10CM | I24.0 | Acute coronary thrombosis not resulting in myocardial infarction |
| ICD10CM | I24.1 | Dressler's syndrome |
| ICD10CM | I24.8 | Other forms of acute ischemic heart disease |
| ICD10CM | I24.9 | Acute ischemic heart disease, unspecified |
| ICD10CM | I25.1 | Atherosclerotic heart disease of native coronary artery |
| ICD10CM | I25.10 | Atherosclerotic heart disease of native coronary artery without angina pectoris |
| ICD10CM | I25.11 | Atherosclerotic heart disease of native coronary artery with angina pectoris |
| ICD10CM | I25.110 | Atherosclerotic heart disease of native coronary artery with unstable angina pectoris |
| ICD10CM | I25.111 | Atherosclerotic heart disease of native coronary artery with angina pectoris with documented spasm |
| ICD10CM | I25.118 | Atherosclerotic heart disease of native coronary artery with other forms of angina pectoris |
| ICD10CM | I25.119 | Atherosclerotic heart disease of native coronary artery with unspecified angina pectoris |
| ICD10CM | I25.2 | Old myocardial infarction |
| ICD10CM | I25.3 | Aneurysm of heart |
| ICD10CM | I25.4 | Coronary artery aneurysm and dissection |
| ICD10CM | I25.41 | Coronary artery aneurysm |
| ICD10CM | I25.42 | Coronary artery dissection |
| ICD10CM | I25.5 | Ischemic cardiomyopathy |
| ICD10CM | I25.6 | Silent myocardial ischemia |
| ICD10CM | I25.7 | Atherosclerosis of coronary artery bypass graft(s) and coronary artery of transplanted heart with angina pectoris |
| ICD10CM | I25.70 | Atherosclerosis of coronary artery bypass graft(s), unspecified, with angina pectoris |
| ICD10CM | I25.700 | Atherosclerosis of coronary artery bypass graft(s), unspecified, with unstable angina pectoris |
| ICD10CM | I25.701 | Atherosclerosis of coronary artery bypass graft(s), unspecified, with angina pectoris with documented spasm |
| ICD10CM | I25.708 | Atherosclerosis of coronary artery bypass graft(s), unspecified, with other forms of angina pectoris |
| ICD10CM | I25.709 | Atherosclerosis of coronary artery bypass graft(s), unspecified, with unspecified angina pectoris |
| ICD10CM | I25.71 | Atherosclerosis of autologous vein coronary artery bypass graft(s) with angina pectoris |
| ICD10CM | I25.710 | Atherosclerosis of autologous vein coronary artery bypass graft(s) with unstable angina pectoris |
| ICD10CM | I25.711 | Atherosclerosis of autologous vein coronary artery bypass graft(s) with angina pectoris with documented spasm |
| ICD10CM | I25.718 | Atherosclerosis of autologous vein coronary artery bypass graft(s) with other forms of angina pectoris |

Continued on next page

**TableS23 – continued from previous page**

| CodeSystem | ConceptCode | ConceptName |
|---|---|---|
| ICD10CM | I25.719 | Atherosclerosis of autologous vein coronary artery bypass graft(s) with unspecified angina pectoris |
| ICD10CM | I25.72 | Atherosclerosis of autologous artery coronary artery bypass graft(s) with angina pectoris |
| ICD10CM | I25.720 | Atherosclerosis of autologous artery coronary artery bypass graft(s) with unstable angina pectoris |
| ICD10CM | I25.721 | Atherosclerosis of autologous artery coronary artery bypass graft(s) with angina pectoris with documented spasm |
| ICD10CM | I25.728 | Atherosclerosis of autologous artery coronary artery bypass graft(s) with other forms of angina pectoris |
| ICD10CM | I25.729 | Atherosclerosis of autologous artery coronary artery bypass graft(s) with unspecified angina pectoris |
| ICD10CM | I25.73 | Atherosclerosis of nonautologous biological coronary artery bypass graft(s) with angina pectoris |
| ICD10CM | I25.730 | Atherosclerosis of nonautologous biological coronary artery bypass graft(s) with unstable angina pectoris |
| ICD10CM | I25.731 | Atherosclerosis of nonautologous biological coronary artery bypass graft(s) with angina pectoris with documented spasm |
| ICD10CM | I25.738 | Atherosclerosis of nonautologous biological coronary artery bypass graft(s) with other forms of angina pectoris |
| ICD10CM | I25.739 | Atherosclerosis of nonautologous biological coronary artery bypass graft(s) with unspecified angina pectoris |
| ICD10CM | I25.75 | Atherosclerosis of native coronary artery of transplanted heart with angina pectoris |
| ICD10CM | I25.750 | Atherosclerosis of native coronary artery of transplanted heart with unstable angina |
| ICD10CM | I25.751 | Atherosclerosis of native coronary artery of transplanted heart with angina pectoris with documented spasm |
| ICD10CM | I25.758 | Atherosclerosis of native coronary artery of transplanted heart with other forms of angina pectoris |
| ICD10CM | I25.759 | Atherosclerosis of native coronary artery of transplanted heart with unspecified angina pectoris |
| ICD10CM | I25.76 | Atherosclerosis of bypass graft of coronary artery of transplanted heart with angina pectoris |
| ICD10CM | I25.760 | Atherosclerosis of bypass graft of coronary artery of transplanted heart with unstable angina |
| ICD10CM | I25.761 | Atherosclerosis of bypass graft of coronary artery of transplanted heart with angina pectoris with documented spasm |
| ICD10CM | I25.768 | Atherosclerosis of bypass graft of coronary artery of transplanted heart with other forms of angina pectoris |
| ICD10CM | I25.769 | Atherosclerosis of bypass graft of coronary artery of transplanted heart with unspecified angina pectoris |
| ICD10CM | I25.79 | Atherosclerosis of other coronary artery bypass graft(s) with angina pectoris |
| ICD10CM | I25.790 | Atherosclerosis of other coronary artery bypass graft(s) with unstable angina pectoris |

Continued on next page

**TableS23 – continued from previous page**

| CodeSystem | ConceptCode | ConceptName |
|---|---|---|
| ICD10CM | I25.791 | Atherosclerosis of other coronary artery bypass graft(s) with angina pectoris with documented spasm |
| ICD10CM | I25.798 | Atherosclerosis of other coronary artery bypass graft(s) with other forms of angina pectoris |
| ICD10CM | I25.799 | Atherosclerosis of other coronary artery bypass graft(s) with unspecified angina pectoris |
| ICD10CM | I25.8 | Other forms of chronic ischemic heart disease |
| ICD10CM | I25.81 | Atherosclerosis of other coronary vessels without angina pectoris |
| ICD10CM | I25.810 | Atherosclerosis of coronary artery bypass graft(s) without angina pectoris |
| ICD10CM | I25.811 | Atherosclerosis of native coronary artery of transplanted heart without angina pectoris |
| ICD10CM | I25.812 | Atherosclerosis of bypass graft of coronary artery of transplanted heart without angina pectoris |
| ICD10CM | I25.82 | Chronic total occlusion of coronary artery |
| ICD10CM | I25.83 | Coronary atherosclerosis due to lipid rich plaque |
| ICD10CM | I25.84 | Coronary atherosclerosis due to calcified coronary lesion |
| ICD10CM | I25.89 | Other forms of chronic ischemic heart disease |
| ICD10CM | I25.9 | Chronic ischemic heart disease, unspecified |
| ICD10CM | I30.0 | Acute nonspecific idiopathic pericarditis |
| ICD10CM | I30.1 | Infective pericarditis |
| ICD10CM | I30.8 | Other forms of acute pericarditis |
| ICD10CM | I30.9 | Acute pericarditis, unspecified |
| ICD10CM | I31.0 | Chronic adhesive pericarditis |
| ICD10CM | I31.1 | Chronic constrictive pericarditis |
| ICD10CM | I31.2 | Hemopericardium, not elsewhere classified |
| ICD10CM | I31.3 | Pericardial effusion (noninflammatory) |
| ICD10CM | I31.4 | Cardiac tamponade |
| ICD10CM | I31.8 | Other specified diseases of pericardium |
| ICD10CM | I31.9 | Disease of pericardium, unspecified |
| ICD10CM | I32 | Pericarditis in diseases classified elsewhere |
| ICD10CM | I33.0 | Acute and subacute infective endocarditis |
| ICD10CM | I33.9 | Acute and subacute endocarditis, unspecified |
| ICD10CM | I34.0 | Nonrheumatic mitral (valve) insufficiency |
| ICD10CM | I34.1 | Nonrheumatic mitral (valve) prolapse |
| ICD10CM | I34.2 | Nonrheumatic mitral (valve) stenosis |
| ICD10CM | I34.8 | Other nonrheumatic mitral valve disorders |
| ICD10CM | I34.9 | Nonrheumatic mitral valve disorder, unspecified |
| ICD10CM | I35.0 | Nonrheumatic aortic (valve) stenosis |
| ICD10CM | I35.1 | Nonrheumatic aortic (valve) insufficiency |
| ICD10CM | I35.2 | Nonrheumatic aortic (valve) stenosis with insufficiency |
| ICD10CM | I35.8 | Other nonrheumatic aortic valve disorders |
| ICD10CM | I35.9 | Nonrheumatic aortic valve disorder, unspecified |
| ICD10CM | I36.0 | Nonrheumatic tricuspid (valve) stenosis |
| ICD10CM | I36.1 | Nonrheumatic tricuspid (valve) insufficiency |
| ICD10CM | I36.2 | Nonrheumatic tricuspid (valve) stenosis with insufficiency |
| ICD10CM | I36.8 | Other nonrheumatic tricuspid valve disorders |

Continued on next page

**TableS23 – continued from previous page**

| CodeSystem | ConceptCode | ConceptName |
|---|---|---|
| ICD10CM | I36.9 | Nonrheumatic tricuspid valve disorder, unspecified |
| ICD10CM | I37.0 | Nonrheumatic pulmonary valve stenosis |
| ICD10CM | I37.1 | Nonrheumatic pulmonary valve insufficiency |
| ICD10CM | I37.2 | Nonrheumatic pulmonary valve stenosis with insufficiency |
| ICD10CM | I37.8 | Other nonrheumatic pulmonary valve disorders |
| ICD10CM | I37.9 | Nonrheumatic pulmonary valve disorder, unspecified |
| ICD10CM | I38 | Endocarditis, valve unspecified |
| ICD10CM | I39 | Endocarditis and heart valve disorders in diseases classified elsewhere |
| ICD10CM | I40.0 | Infective myocarditis |
| ICD10CM | I40.1 | Isolated myocarditis |
| ICD10CM | I40.8 | Other acute myocarditis |
| ICD10CM | I40.9 | Acute myocarditis, unspecified |
| ICD10CM | I41 | Myocarditis in diseases classified elsewhere |
| ICD10CM | I42.0 | Dilated cardiomyopathy |
| ICD10CM | I42.1 | Obstructive hypertrophic cardiomyopathy |
| ICD10CM | I42.2 | Other hypertrophic cardiomyopathy |
| ICD10CM | I42.3 | Endomyocardial (eosinophilic) disease |
| ICD10CM | I42.4 | Endocardial fibroelastosis |
| ICD10CM | I42.5 | Other restrictive cardiomyopathy |
| ICD10CM | I42.7 | Cardiomyopathy due to drug and external agent |
| ICD10CM | I42.8 | Other cardiomyopathies |
| ICD10CM | I42.9 | Cardiomyopathy, unspecified |
| ICD10CM | I43 | Cardiomyopathy in diseases classified elsewhere |
| ICD10CM | I44.0 | Atrioventricular block, first degree |
| ICD10CM | I44.1 | Atrioventricular block, second degree |
| ICD10CM | I44.2 | Atrioventricular block, complete |
| ICD10CM | I44.30 | Unspecified atrioventricular block |
| ICD10CM | I44.39 | Other atrioventricular block |
| ICD10CM | I44.4 | Left anterior fascicular block |
| ICD10CM | I44.5 | Left posterior fascicular block |
| ICD10CM | I44.60 | Unspecified fascicular block |
| ICD10CM | I44.69 | Other fascicular block |
| ICD10CM | I44.7 | Left bundle-branch block, unspecified |
| ICD10CM | I45.0 | Right fascicular block |
| ICD10CM | I45.10 | Unspecified right bundle-branch block |
| ICD10CM | I45.19 | Other right bundle-branch block |
| ICD10CM | I45.2 | Bifascicular block |
| ICD10CM | I45.3 | Trifascicular block |
| ICD10CM | I45.4 | Nonspecific intraventricular block |
| ICD10CM | I45.5 | Other specified heart block |
| ICD10CM | I45.6 | Pre-excitation syndrome |
| ICD10CM | I45.81 | Long QT syndrome |
| ICD10CM | I45.89 | Other specified conduction disorders |
| ICD10CM | I45.9 | Conduction disorder, unspecified |
| ICD10CM | I46.2 | Cardiac arrest due to underlying cardiac condition |
| ICD10CM | I46.8 | Cardiac arrest due to other underlying condition |

Continued on next page

**TableS23 – continued from previous page**

| CodeSystem | ConceptCode | ConceptName |
|---|---|---|
| ICD10CM | I46.9 | Cardiac arrest, cause unspecified |
| ICD10CM | I47.0 | Re-entry ventricular arrhythmia |
| ICD10CM | I47.1 | Supraventricular tachycardia |
| ICD10CM | I47.2 | Ventricular tachycardia |
| ICD10CM | I47.9 | Paroxysmal tachycardia, unspecified |
| ICD10CM | I48.0 | Paroxysmal atrial fibrillation |
| ICD10CM | I48.1 | Persistent atrial fibrillation |
| ICD10CM | I48.2 | Chronic atrial fibrillation |
| ICD10CM | I48.3 | Typical atrial flutter |
| ICD10CM | I48.4 | Atypical atrial flutter |
| ICD10CM | I48.91 | Unspecified atrial fibrillation |
| ICD10CM | I48.92 | Unspecified atrial flutter |
| ICD10CM | I49.01 | Ventricular fibrillation |
| ICD10CM | I49.02 | Ventricular flutter |
| ICD10CM | I49.1 | Atrial premature depolarization |
| ICD10CM | I49.2 | Junctional premature depolarization |
| ICD10CM | I49.3 | Ventricular premature depolarization |
| ICD10CM | I49.40 | Unspecified premature depolarization |
| ICD10CM | I49.49 | Other premature depolarization |
| ICD10CM | I49.5 | Sick sinus syndrome |
| ICD10CM | I49.8 | Other specified cardiac arrhythmias |
| ICD10CM | I49.9 | Cardiac arrhythmia, unspecified |
| ICD10CM | I51.0 | Cardiac septal defect, acquired |
| ICD10CM | I51.1 | Rupture of chordae tendineae, not elsewhere classified |
| ICD10CM | I51.2 | Rupture of papillary muscle, not elsewhere classified |
| ICD10CM | I51.3 | Intracardiac thrombosis, not elsewhere classified |
| ICD10CM | I51.4 | Myocarditis, unspecified |
| ICD10CM | I51.5 | Myocardial degeneration |
| ICD10CM | I51.7 | Cardiomegaly |
| ICD10CM | I51.81 | Takotsubo syndrome |
| ICD10CM | I51.89 | Other ill-defined heart diseases |
| ICD10CM | I51.9 | Heart disease, unspecified |
| ICD10CM | I52 | Other heart disorders in diseases classified elsewhere |
| ICD10CM | I63 | Cerebral infarction |
| ICD10CM | I63.0 | Cerebral infarction due to thrombosis of precerebral arteries |
| ICD10CM | I63.00 | Cerebral infarction due to thrombosis of unspecified precerebral artery |
| ICD10CM | I63.01 | Cerebral infarction due to thrombosis of vertebral artery |
| ICD10CM | I63.011 | Cerebral infarction due to thrombosis of right vertebral artery |
| ICD10CM | I63.012 | Cerebral infarction due to thrombosis of left vertebral artery |
| ICD10CM | I63.013 | Cerebral infarction due to thrombosis of bilateral vertebral arteries |
| ICD10CM | I63.019 | Cerebral infarction due to thrombosis of unspecified vertebral artery |
| ICD10CM | I63.02 | Cerebral infarction due to thrombosis of basilar artery |
| ICD10CM | I63.03 | Cerebral infarction due to thrombosis of carotid artery |
| ICD10CM | I63.031 | Cerebral infarction due to thrombosis of right carotid artery |

Continued on next page

**TableS23 – continued from previous page**

| CodeSystem | ConceptCode | ConceptName |
|---|---|---|
| ICD10CM | I63.032 | Cerebral infarction due to thrombosis of left carotid artery |
| ICD10CM | I63.033 | Cerebral infarction due to thrombosis of bilateral carotid arteries |
| ICD10CM | I63.039 | Cerebral infarction due to thrombosis of unspecified carotid artery |
| ICD10CM | I63.09 | Cerebral infarction due to thrombosis of other precerebral artery |
| ICD10CM | I63.1 | Cerebral infarction due to embolism of precerebral arteries |
| ICD10CM | I63.10 | Cerebral infarction due to embolism of unspecified precerebral artery |
| ICD10CM | I63.11 | Cerebral infarction due to embolism of vertebral artery |
| ICD10CM | I63.111 | Cerebral infarction due to embolism of right vertebral artery |
| ICD10CM | I63.112 | Cerebral infarction due to embolism of left vertebral artery |
| ICD10CM | I63.113 | Cerebral infarction due to embolism of bilateral vertebral arteries |
| ICD10CM | I63.119 | Cerebral infarction due to embolism of unspecified vertebral artery |
| ICD10CM | I63.12 | Cerebral infarction due to embolism of basilar artery |
| ICD10CM | I63.13 | Cerebral infarction due to embolism of carotid artery |
| ICD10CM | I63.131 | Cerebral infarction due to embolism of right carotid artery |
| ICD10CM | I63.132 | Cerebral infarction due to embolism of left carotid artery |
| ICD10CM | I63.133 | Cerebral infarction due to embolism of bilateral carotid arteries |
| ICD10CM | I63.139 | Cerebral infarction due to embolism of unspecified carotid artery |
| ICD10CM | I63.19 | Cerebral infarction due to embolism of other precerebral artery |
| ICD10CM | I63.2 | Cerebral infarction due to unspecified occlusion or stenosis of precerebral arteries |
| ICD10CM | I63.20 | Cerebral infarction due to unspecified occlusion or stenosis of unspecified precerebral arteries |
| ICD10CM | I63.21 | Cerebral infarction due to unspecified occlusion or stenosis of vertebral arteries |
| ICD10CM | I63.211 | Cerebral infarction due to unspecified occlusion or stenosis of right vertebral artery |
| ICD10CM | I63.212 | Cerebral infarction due to unspecified occlusion or stenosis of left vertebral artery |
| ICD10CM | I63.213 | Cerebral infarction due to unspecified occlusion or stenosis of bilateral vertebral arteries |
| ICD10CM | I63.219 | Cerebral infarction due to unspecified occlusion or stenosis of unspecified vertebral artery |
| ICD10CM | I63.22 | Cerebral infarction due to unspecified occlusion or stenosis of basilar artery |
| ICD10CM | I63.23 | Cerebral infarction due to unspecified occlusion or stenosis of carotid arteries |
| ICD10CM | I63.231 | Cerebral infarction due to unspecified occlusion or stenosis of right carotid arteries |
| ICD10CM | I63.232 | Cerebral infarction due to unspecified occlusion or stenosis of left carotid arteries |
| ICD10CM | I63.233 | Cerebral infarction due to unspecified occlusion or stenosis of bilateral carotid arteries |
| ICD10CM | I63.239 | Cerebral infarction due to unspecified occlusion or stenosis of unspecified carotid artery |

Continued on next page

**TableS23 – continued from previous page**

| CodeSystem | ConceptCode | ConceptName |
|---|---|---|
| ICD10CM | I63.29 | Cerebral infarction due to unspecified occlusion or stenosis of other precerebral arteries |
| ICD10CM | I63.3 | Cerebral infarction due to thrombosis of cerebral arteries |
| ICD10CM | I63.30 | Cerebral infarction due to thrombosis of unspecified cerebral artery |
| ICD10CM | I63.31 | Cerebral infarction due to thrombosis of middle cerebral artery |
| ICD10CM | I63.311 | Cerebral infarction due to thrombosis of right middle cerebral artery |
| ICD10CM | I63.312 | Cerebral infarction due to thrombosis of left middle cerebral artery |
| ICD10CM | I63.313 | Cerebral infarction due to thrombosis of bilateral middle cerebral arteries |
| ICD10CM | I63.319 | Cerebral infarction due to thrombosis of unspecified middle cerebral artery |
| ICD10CM | I63.32 | Cerebral infarction due to thrombosis of anterior cerebral artery |
| ICD10CM | I63.321 | Cerebral infarction due to thrombosis of right anterior cerebral artery |
| ICD10CM | I63.322 | Cerebral infarction due to thrombosis of left anterior cerebral artery |
| ICD10CM | I63.323 | Cerebral infarction due to thrombosis of bilateral anterior cerebral arteries |
| ICD10CM | I63.329 | Cerebral infarction due to thrombosis of unspecified anterior cerebral artery |
| ICD10CM | I63.33 | Cerebral infarction due to thrombosis of posterior cerebral artery |
| ICD10CM | I63.331 | Cerebral infarction due to thrombosis of right posterior cerebral artery |
| ICD10CM | I63.332 | Cerebral infarction due to thrombosis of left posterior cerebral artery |
| ICD10CM | I63.333 | Cerebral infarction due to thrombosis of bilateral posterior cerebral arteries |
| ICD10CM | I63.339 | Cerebral infarction due to thrombosis of unspecified posterior cerebral artery |
| ICD10CM | I63.34 | Cerebral infarction due to thrombosis of cerebellar artery |
| ICD10CM | I63.341 | Cerebral infarction due to thrombosis of right cerebellar artery |
| ICD10CM | I63.342 | Cerebral infarction due to thrombosis of left cerebellar artery |
| ICD10CM | I63.343 | Cerebral infarction due to thrombosis of bilateral cerebellar arteries |
| ICD10CM | I63.349 | Cerebral infarction due to thrombosis of unspecified cerebellar artery |
| ICD10CM | I63.39 | Cerebral infarction due to thrombosis of other cerebral artery |
| ICD10CM | I63.4 | Cerebral infarction due to embolism of cerebral arteries |
| ICD10CM | I63.40 | Cerebral infarction due to embolism of unspecified cerebral artery |
| ICD10CM | I63.41 | Cerebral infarction due to embolism of middle cerebral artery |
| ICD10CM | I63.411 | Cerebral infarction due to embolism of right middle cerebral artery |
| ICD10CM | I63.412 | Cerebral infarction due to embolism of left middle cerebral artery |

Continued on next page

**TableS23 – continued from previous page**

| CodeSystem | ConceptCode | ConceptName |
|---|---|---|
| ICD10CM | I63.413 | Cerebral infarction due to embolism of bilateral middle cerebral arteries |
| ICD10CM | I63.419 | Cerebral infarction due to embolism of unspecified middle cerebral artery |
| ICD10CM | I63.42 | Cerebral infarction due to embolism of anterior cerebral artery |
| ICD10CM | I63.421 | Cerebral infarction due to embolism of right anterior cerebral artery |
| ICD10CM | I63.422 | Cerebral infarction due to embolism of left anterior cerebral artery |
| ICD10CM | I63.423 | Cerebral infarction due to embolism of bilateral anterior cerebral arteries |
| ICD10CM | I63.429 | Cerebral infarction due to embolism of unspecified anterior cerebral artery |
| ICD10CM | I63.43 | Cerebral infarction due to embolism of posterior cerebral artery |
| ICD10CM | I63.431 | Cerebral infarction due to embolism of right posterior cerebral artery |
| ICD10CM | I63.432 | Cerebral infarction due to embolism of left posterior cerebral artery |
| ICD10CM | I63.433 | Cerebral infarction due to embolism of bilateral posterior cerebral arteries |
| ICD10CM | I63.439 | Cerebral infarction due to embolism of unspecified posterior cerebral artery |
| ICD10CM | I63.44 | Cerebral infarction due to embolism of cerebellar artery |
| ICD10CM | I63.441 | Cerebral infarction due to embolism of right cerebellar artery |
| ICD10CM | I63.442 | Cerebral infarction due to embolism of left cerebellar artery |
| ICD10CM | I63.443 | Cerebral infarction due to embolism of bilateral cerebellar arteries |
| ICD10CM | I63.449 | Cerebral infarction due to embolism of unspecified cerebellar artery |
| ICD10CM | I63.49 | Cerebral infarction due to embolism of other cerebral artery |
| ICD10CM | I63.5 | Cerebral infarction due to unspecified occlusion or stenosis of cerebral arteries |
| ICD10CM | I63.50 | Cerebral infarction due to unspecified occlusion or stenosis of unspecified cerebral artery |
| ICD10CM | I63.51 | Cerebral infarction due to unspecified occlusion or stenosis of middle cerebral artery |
| ICD10CM | I63.511 | Cerebral infarction due to unspecified occlusion or stenosis of right middle cerebral artery |
| ICD10CM | I63.512 | Cerebral infarction due to unspecified occlusion or stenosis of left middle cerebral artery |
| ICD10CM | I63.513 | Cerebral infarction due to unspecified occlusion or stenosis of bilateral middle cerebral arteries |
| ICD10CM | I63.519 | Cerebral infarction due to unspecified occlusion or stenosis of unspecified middle cerebral artery |
| ICD10CM | I63.52 | Cerebral infarction due to unspecified occlusion or stenosis of anterior cerebral artery |

Continued on next page

**TableS23 – continued from previous page**

| CodeSystem | ConceptCode | ConceptName |
|---|---|---|
| ICD10CM | I63.521 | Cerebral infarction due to unspecified occlusion or stenosis of right anterior cerebral artery |
| ICD10CM | I63.522 | Cerebral infarction due to unspecified occlusion or stenosis of left anterior cerebral artery |
| ICD10CM | I63.523 | Cerebral infarction due to unspecified occlusion or stenosis of bilateral anterior cerebral arteries |
| ICD10CM | I63.529 | Cerebral infarction due to unspecified occlusion or stenosis of unspecified anterior cerebral artery |
| ICD10CM | I63.53 | Cerebral infarction due to unspecified occlusion or stenosis of posterior cerebral artery |
| ICD10CM | I63.531 | Cerebral infarction due to unspecified occlusion or stenosis of right posterior cerebral artery |
| ICD10CM | I63.532 | Cerebral infarction due to unspecified occlusion or stenosis of left posterior cerebral artery |
| ICD10CM | I63.533 | Cerebral infarction due to unspecified occlusion or stenosis of bilateral posterior cerebral arteries |
| ICD10CM | I63.539 | Cerebral infarction due to unspecified occlusion or stenosis of unspecified posterior cerebral artery |
| ICD10CM | I63.54 | Cerebral infarction due to unspecified occlusion or stenosis of cerebellar artery |
| ICD10CM | I63.541 | Cerebral infarction due to unspecified occlusion or stenosis of right cerebellar artery |
| ICD10CM | I63.542 | Cerebral infarction due to unspecified occlusion or stenosis of left cerebellar artery |
| ICD10CM | I63.543 | Cerebral infarction due to unspecified occlusion or stenosis of bilateral cerebellar arteries |
| ICD10CM | I63.549 | Cerebral infarction due to unspecified occlusion or stenosis of unspecified cerebellar artery |
| ICD10CM | I63.59 | Cerebral infarction due to unspecified occlusion or stenosis of other cerebral artery |
| ICD10CM | I63.6 | Cerebral infarction due to cerebral venous thrombosis, nonpyogenic |
| ICD10CM | I63.8 | Other cerebral infarction |
| ICD10CM | I63.81 | Other cerebral infarction due to occlusion or stenosis of small artery |
| ICD10CM | I63.89 | Other cerebral infarction |
| ICD10CM | I63.9 | Cerebral infarction, unspecified |
| ICD10CM | I67.0 | Dissection of cerebral arteries, nonruptured |
| ICD10CM | I70.0 | Atherosclerosis of aorta |
| ICD10CM | I70.1 | Atherosclerosis of renal artery |
| ICD10CM | I70.2 | Atherosclerosis of native arteries of the extremities |
| ICD10CM | I70.20 | Unspecified atherosclerosis of native arteries of extremities |
| ICD10CM | I70.201 | Unspecified atherosclerosis of native arteries of extremities, right leg |
| ICD10CM | I70.202 | Unspecified atherosclerosis of native arteries of extremities, left leg |

Continued on next page

**TableS23 – continued from previous page**

| CodeSystem | ConceptCode | ConceptName |
|---|---|---|
| ICD10CM | I70.203 | Unspecified atherosclerosis of native arteries of extremities, bilateral legs |
| ICD10CM | I70.208 | Unspecified atherosclerosis of native arteries of extremities, other extremity |
| ICD10CM | I70.209 | Unspecified atherosclerosis of native arteries of extremities, unspecified extremity |
| ICD10CM | I70.21 | Atherosclerosis of native arteries of extremities with intermittent claudication |
| ICD10CM | I70.211 | Atherosclerosis of native arteries of extremities with intermittent claudication, right leg |
| ICD10CM | I70.212 | Atherosclerosis of native arteries of extremities with intermittent claudication, left leg |
| ICD10CM | I70.213 | Atherosclerosis of native arteries of extremities with intermittent claudication, bilateral legs |
| ICD10CM | I70.218 | Atherosclerosis of native arteries of extremities with intermittent claudication, other extremity |
| ICD10CM | I70.219 | Atherosclerosis of native arteries of extremities with intermittent claudication, unspecified extremity |
| ICD10CM | I70.22 | Atherosclerosis of native arteries of extremities with rest pain |
| ICD10CM | I70.221 | Atherosclerosis of native arteries of extremities with rest pain, right leg |
| ICD10CM | I70.222 | Atherosclerosis of native arteries of extremities with rest pain, left leg |
| ICD10CM | I70.223 | Atherosclerosis of native arteries of extremities with rest pain, bilateral legs |
| ICD10CM | I70.228 | Atherosclerosis of native arteries of extremities with rest pain, other extremity |
| ICD10CM | I70.229 | Atherosclerosis of native arteries of extremities with rest pain, unspecified extremity |
| ICD10CM | I70.23 | Atherosclerosis of native arteries of right leg with ulceration |
| ICD10CM | I70.231 | Atherosclerosis of native arteries of right leg with ulceration of thigh |
| ICD10CM | I70.232 | Atherosclerosis of native arteries of right leg with ulceration of calf |
| ICD10CM | I70.233 | Atherosclerosis of native arteries of right leg with ulceration of ankle |
| ICD10CM | I70.234 | Atherosclerosis of native arteries of right leg with ulceration of heel and midfoot |
| ICD10CM | I70.235 | Atherosclerosis of native arteries of right leg with ulceration of other part of foot |
| ICD10CM | I70.238 | Atherosclerosis of native arteries of right leg with ulceration of other part of lower leg |
| ICD10CM | I70.239 | Atherosclerosis of native arteries of right leg with ulceration of unspecified site |
| ICD10CM | I70.24 | Atherosclerosis of native arteries of left leg with ulceration |
| ICD10CM | I70.241 | Atherosclerosis of native arteries of left leg with ulceration of thigh |

Continued on next page

**TableS23 – continued from previous page**

| CodeSystem | ConceptCode | ConceptName |
|---|---|---|
| ICD10CM | I70.242 | Atherosclerosis of native arteries of left leg with ulceration of calf |
| ICD10CM | I70.243 | Atherosclerosis of native arteries of left leg with ulceration of ankle |
| ICD10CM | I70.244 | Atherosclerosis of native arteries of left leg with ulceration of heel and midfoot |
| ICD10CM | I70.245 | Atherosclerosis of native arteries of left leg with ulceration of other part of foot |
| ICD10CM | I70.248 | Atherosclerosis of native arteries of left leg with ulceration of other part of lower leg |
| ICD10CM | I70.249 | Atherosclerosis of native arteries of left leg with ulceration of unspecified site |
| ICD10CM | I70.25 | Atherosclerosis of native arteries of other extremities with ulceration |
| ICD10CM | I70.26 | Atherosclerosis of native arteries of extremities with gangrene |
| ICD10CM | I70.261 | Atherosclerosis of native arteries of extremities with gangrene, right leg |
| ICD10CM | I70.262 | Atherosclerosis of native arteries of extremities with gangrene, left leg |
| ICD10CM | I70.263 | Atherosclerosis of native arteries of extremities with gangrene, bilateral legs |
| ICD10CM | I70.268 | Atherosclerosis of native arteries of extremities with gangrene, other extremity |
| ICD10CM | I70.269 | Atherosclerosis of native arteries of extremities with gangrene, unspecified extremity |
| ICD10CM | I70.29 | Other atherosclerosis of native arteries of extremities |
| ICD10CM | I70.291 | Other atherosclerosis of native arteries of extremities, right leg |
| ICD10CM | I70.292 | Other atherosclerosis of native arteries of extremities, left leg |
| ICD10CM | I70.293 | Other atherosclerosis of native arteries of extremities, bilateral legs |
| ICD10CM | I70.298 | Other atherosclerosis of native arteries of extremities, other extremity |
| ICD10CM | I70.299 | Other atherosclerosis of native arteries of extremities, unspecified extremity |
| ICD10CM | I71.6 | Thoracoabdominal aortic aneurysm, without rupture |
| ICD10CM | I71.8 | Aortic aneurysm of unspecified site, ruptured |
| ICD10CM | I71.9 | Aortic aneurysm of unspecified site, without rupture |
| ICD10CM | I70.8 | Atherosclerosis of other arteries |
| ICD10CM | I70.90 | Unspecified atherosclerosis |
| ICD10CM | I70.91 | Generalized atherosclerosis |
| ICD10CM | I70.92 | Chronic total occlusion of artery of the extremities |
| ICD10CM | I71.00 | Dissection of unspecified site of aorta |
| ICD10CM | I71.01 | Dissection of thoracic aorta |
| ICD10CM | I71.02 | Dissection of abdominal aorta |
| ICD10CM | I71.03 | Dissection of thoracoabdominal aorta |
| ICD10CM | I71.1 | Thoracic aortic aneurysm, ruptured |
| ICD10CM | I71.2 | Thoracic aortic aneurysm, without rupture |
| ICD10CM | I71.3 | Abdominal aortic aneurysm, ruptured |

Continued on next page

**TableS23 – continued from previous page**

| CodeSystem | ConceptCode | ConceptName |
|---|---|---|
| ICD10CM | I71.4 | Abdominal aortic aneurysm, without rupture |
| ICD10CM | I71.5 | Thoracoabdominal aortic aneurysm, ruptured |
| ICD10CM | I72.0 | Aneurysm of carotid artery |
| ICD10CM | I72.1 | Aneurysm of artery of upper extremity |
| ICD10CM | I72.2 | Aneurysm of renal artery |
| ICD10CM | I72.3 | Aneurysm of iliac artery |
| ICD10CM | I72.4 | Aneurysm of artery of lower extremity |
| ICD10CM | I72.5 | Aneurysm of other precerebral arteries |
| ICD10CM | I72.6 | Aneurysm of vertebral artery |
| ICD10CM | I72.8 | Aneurysm of other specified arteries |
| ICD10CM | I72.9 | Aneurysm of unspecified site |
| ICD10CM | I73.00 | Raynaud's syndrome without gangrene |
| ICD10CM | I73.1 | Thromboangiitis obliterans [Buerger's disease] |
| ICD10CM | I73.81 | Erythromelalgia |
| ICD10CM | I73.89 | Other specified peripheral vascular diseases |
| ICD10CM | I73.9 | Peripheral vascular disease, unspecified |
| ICD10CM | I74.0 | Embolism and thrombosis of abdominal aorta |
| ICD10CM | I74.01 | Saddle embolus of abdominal aorta |
| ICD10CM | I74.09 | Other arterial embolism and thrombosis of abdominal aorta |
| ICD10CM | I74.10 | Embolism and thrombosis of unspecified parts of aorta |
| ICD10CM | I74.11 | Embolism and thrombosis of thoracic aorta |
| ICD10CM | I74.19 | Embolism and thrombosis of other parts of aorta |
| ICD10CM | I74.2 | Embolism and thrombosis of arteries of the upper extremities |
| ICD10CM | I74.3 | Embolism and thrombosis of arteries of the lower extremities |
| ICD10CM | I74.4 | Embolism and thrombosis of arteries of extremities, unspecified |
| ICD10CM | I74.5 | Embolism and thrombosis of iliac artery |
| ICD10CM | I74.8 | Embolism and thrombosis of other arteries |
| ICD10CM | I74.9 | Embolism and thrombosis of unspecified artery |
| ICD10CM | I75.0 | Atheroembolism of extremities |
| ICD10CM | I75.01 | Atheroembolism of upper extremity |
| ICD10CM | I75.011 | Atheroembolism of right upper extremity |
| ICD10CM | I75.012 | Atheroembolism of left upper extremity |
| ICD10CM | I75.013 | Atheroembolism of bilateral upper extremities |
| ICD10CM | I75.019 | Atheroembolism of unspecified upper extremity |
| ICD10CM | I75.02 | Atheroembolism of lower extremity |
| ICD10CM | I75.021 | Atheroembolism of right lower extremity |
| ICD10CM | I75.022 | Atheroembolism of left lower extremity |
| ICD10CM | I75.023 | Atheroembolism of bilateral lower extremities |
| ICD10CM | I75.029 | Atheroembolism of unspecified lower extremity |
| ICD10CM | I75.81 | Atheroembolism of kidney |
| ICD10CM | I75.89 | Atheroembolism of other site |
| ICD10CM | I77.0 | Arteriovenous fistula, acquired |
| ICD10CM | I77.1 | Stricture of artery |
| ICD10CM | I77.2 | Rupture of artery |
| ICD10CM | I77.3 | Arterial fibromuscular dysplasia |
| ICD10CM | I77.4 | Celiac artery compression syndrome |
| ICD10CM | I77.5 | Necrosis of artery |

Continued on next page

**TableS23 – continued from previous page**

| CodeSystem | ConceptCode | ConceptName |
|---|---|---|
| ICD10CM | I77.6 | Arteritis, unspecified |
| ICD10CM | I77.70 | Dissection of unspecified artery |
| ICD10CM | I77.71 | Dissection of carotid artery |
| ICD10CM | I77.72 | Dissection of iliac artery |
| ICD10CM | I77.73 | Dissection of renal artery |
| ICD10CM | I77.74 | Dissection of vertebral artery |
| ICD10CM | I77.75 | Dissection of other precerebral arteries |
| ICD10CM | I77.76 | Dissection of artery of upper extremity |
| ICD10CM | I77.77 | Dissection of artery of lower extremity |
| ICD10CM | I77.79 | Dissection of other specified artery |
| ICD10CM | I77.810 | Thoracic aortic ectasia |
| ICD10CM | I77.811 | Abdominal aortic ectasia |
| ICD10CM | I77.812 | Thoracoabdominal aortic ectasia |
| ICD10CM | I77.819 | Aortic ectasia, unspecified site |
| ICD10CM | I77.89 | Other specified disorders of arteries and arterioles |
| ICD10CM | I77.9 | Disorder of arteries and arterioles, unspecified |
| ICD10CM | I78.0 | Hereditary hemorrhagic telangiectasia |
| ICD10CM | I78.1 | Nevus, non-neoplastic |
| ICD10CM | I78.8 | Other diseases of capillaries |
| ICD10CM | I78.9 | Disease of capillaries, unspecified |
| ICD10CM | I79.0 | Aneurysm of aorta in diseases classified elsewhere |
| ICD10CM | I79.1 | Aortitis in diseases classified elsewhere |
| ICD10CM | I79.8 | Other disorders of arteries, arterioles and capillaries in diseases classified elsewhere |
| ICD10CM | I95.0 | Idiopathic hypotension |
| ICD10CM | I95.1 | Orthostatic hypotension |
| ICD10CM | I95.89 | Other hypotension |
| ICD10CM | I95.9 | Hypotension, unspecified |
| ICD10CM | I99.8 | Other disorder of circulatory system |
| ICD10CM | I99.9 | Unspecified disorder of circulatory system |
| ICD10CM | J10.82 | Influenza due to other identified influenza virus with myocarditis |
| ICD10CM | K55.0 | Acute vascular disorders of intestine |
| ICD10CM | K55.011 | Focal (segmental) acute (reversible) ischemia of small intestine |
| ICD10CM | K55.012 | Diffuse acute (reversible) ischemia of small intestine |
| ICD10CM | K55.019 | Acute (reversible) ischemia of small intestine, extent unspecified |
| ICD10CM | K55.021 | Focal (segmental) acute infarction of small intestine |
| ICD10CM | K55.022 | Diffuse acute infarction of small intestine |
| ICD10CM | K55.029 | Acute infarction of small intestine, extent unspecified |
| ICD10CM | K55.031 | Focal (segmental) acute (reversible) ischemia of large intestine |
| ICD10CM | K55.032 | Diffuse acute (reversible) ischemia of large intestine |
| ICD10CM | K55.039 | Acute (reversible) ischemia of large intestine, extent unspecified |
| ICD10CM | K55.041 | Focal (segmental) acute infarction of large intestine |
| ICD10CM | K55.042 | Diffuse acute infarction of large intestine |
| ICD10CM | K55.049 | Acute infarction of large intestine, extent unspecified |
| ICD10CM | K55.051 | Focal (segmental) acute (reversible) ischemia of intestine, part unspecified |
| ICD10CM | K55.052 | Diffuse acute (reversible) ischemia of intestine, part unspecified |

Continued on next page

**TableS23 – continued from previous page**

| CodeSystem | ConceptCode | ConceptName |
|---|---|---|
| ICD10CM | K55.059 | Acute (reversible) ischemia of intestine, part and extent unspecified |
| ICD10CM | K55.061 | Focal (segmental) acute infarction of intestine, part unspecified |
| ICD10CM | K55.062 | Diffuse acute infarction of intestine, part unspecified |
| ICD10CM | K55.069 | Acute infarction of intestine, part and extent unspecified |
| ICD10CM | K55.1 | Chronic vascular disorders of intestine |
| ICD10CM | K55.8 | Other vascular disorders of intestine |
| ICD10CM | K55.9 | Vascular disorder of intestine, unspecified |
| ICD10CM | M31.0 | Hypersensitivity angiitis |
| ICD10CM | P29.3 | Persistent fetal circulation |
| ICD10CM | P29.30 | Pulmonary hypertension of newborn |
| ICD10CM | P29.38 | Other persistent fetal circulation |
| ICD10CM | Q20.0 | Common arterial trunk |
| ICD10CM | Q20.1 | Double outlet right ventricle |
| ICD10CM | Q20.2 | Double outlet left ventricle |
| ICD10CM | Q20.3 | Discordant ventriculoarterial connection |
| ICD10CM | Q20.4 | Double inlet ventricle |
| ICD10CM | Q20.5 | Discordant atrioventricular connection |
| ICD10CM | Q20.6 | Isomerism of atrial appendages |
| ICD10CM | Q20.8 | Other congenital malformations of cardiac chambers and connections |
| ICD10CM | Q20.9 | Congenital malformation of cardiac chambers and connections, unspecified |
| ICD10CM | Q21.0 | Ventricular septal defect |
| ICD10CM | Q21.1 | Atrial septal defect |
| ICD10CM | Q21.2 | Atrioventricular septal defect |
| ICD10CM | Q21.3 | Tetralogy of Fallot |
| ICD10CM | Q21.4 | Aortopulmonary septal defect |
| ICD10CM | Q21.8 | Other congenital malformations of cardiac septa |
| ICD10CM | Q21.9 | Congenital malformation of cardiac septum, unspecified |
| ICD10CM | Q22.0 | Pulmonary valve atresia |
| ICD10CM | Q22.1 | Congenital pulmonary valve stenosis |
| ICD10CM | Q22.2 | Congenital pulmonary valve insufficiency |
| ICD10CM | Q22.3 | Other congenital malformations of pulmonary valve |
| ICD10CM | Q22.4 | Congenital tricuspid stenosis |
| ICD10CM | Q22.5 | Ebstein's anomaly |
| ICD10CM | Q22.6 | Hypoplastic right heart syndrome |
| ICD10CM | Q22.8 | Other congenital malformations of tricuspid valve |
| ICD10CM | Q22.9 | Congenital malformation of tricuspid valve, unspecified |
| ICD10CM | Q23.0 | Congenital stenosis of aortic valve |
| ICD10CM | Q23.1 | Congenital insufficiency of aortic valve |
| ICD10CM | Q23.2 | Congenital mitral stenosis |
| ICD10CM | Q23.3 | Congenital mitral insufficiency |
| ICD10CM | Q23.4 | Hypoplastic left heart syndrome |
| ICD10CM | Q23.8 | Other congenital malformations of aortic and mitral valves |
| ICD10CM | Q23.9 | Congenital malformation of aortic and mitral valves, unspecified |
| ICD10CM | Q24.0 | Dextrocardia |

Continued on next page

**TableS23 – continued from previous page**

| CodeSystem | ConceptCode | ConceptName |
|---|---|---|
| ICD10CM | Q24.1 | Levocardia |
| ICD10CM | Q24.2 | Cor triatriatum |
| ICD10CM | Q24.3 | Pulmonary infundibular stenosis |
| ICD10CM | Q24.4 | Congenital subaortic stenosis |
| ICD10CM | Q24.5 | Malformation of coronary vessels |
| ICD10CM | Q24.6 | Congenital heart block |
| ICD10CM | Q24.8 | Other specified congenital malformations of heart |
| ICD10CM | Q24.9 | Congenital malformation of heart, unspecified |
| ICD10CM | Q25.0 | Patent ductus arteriosus |
| ICD10CM | Q25.1 | Coarctation of aorta |
| ICD10CM | Q25.2 | Atresia of aorta |
| ICD10CM | Q25.21 | Interruption of aortic arch |
| ICD10CM | Q25.29 | Other atresia of aorta |
| ICD10CM | Q25.3 | Supravalvular aortic stenosis |
| ICD10CM | Q25.4 | Other congenital malformations of aorta |
| ICD10CM | Q25.40 | Congenital malformation of aorta unspecified |
| ICD10CM | Q25.41 | Absence and aplasia of aorta |
| ICD10CM | Q25.42 | Hypoplasia of aorta |
| ICD10CM | Q25.43 | Congenital aneurysm of aorta |
| ICD10CM | Q25.44 | Congenital dilation of aorta |
| ICD10CM | Q25.45 | Double aortic arch |
| ICD10CM | Q25.46 | Tortuous aortic arch |
| ICD10CM | Q25.47 | Right aortic arch |
| ICD10CM | Q25.48 | Anomalous origin of subclavian artery |
| ICD10CM | Q25.49 | Other congenital malformations of aorta |
| ICD10CM | Q25.5 | Atresia of pulmonary artery |
| ICD10CM | Q25.6 | Stenosis of pulmonary artery |
| ICD10CM | Q25.71 | Coarctation of pulmonary artery |
| ICD10CM | Q25.72 | Congenital pulmonary arteriovenous malformation |
| ICD10CM | Q25.79 | Other congenital malformations of pulmonary artery |
| ICD10CM | Q25.8 | Other congenital malformations of other great arteries |
| ICD10CM | Q25.9 | Congenital malformation of great arteries, unspecified |
| ICD10CM | Q26.0 | Congenital stenosis of vena cava |
| ICD10CM | Q26.1 | Persistent left superior vena cava |
| ICD10CM | Q26.2 | Total anomalous pulmonary venous connection |
| ICD10CM | Q26.3 | Partial anomalous pulmonary venous connection |
| ICD10CM | Q26.4 | Anomalous pulmonary venous connection, unspecified |
| ICD10CM | Q26.5 | Anomalous portal venous connection |
| ICD10CM | Q26.6 | Portal vein-hepatic artery fistula |
| ICD10CM | Q26.8 | Other congenital malformations of great veins |
| ICD10CM | Q26.9 | Congenital malformation of great vein, unspecified |
| ICD10CM | Q27.0 | Congenital absence and hypoplasia of umbilical artery |
| ICD10CM | Q27.1 | Congenital renal artery stenosis |
| ICD10CM | Q27.2 | Other congenital malformations of renal artery |
| ICD10CM | Q27.30 | Arteriovenous malformation, site unspecified |
| ICD10CM | Q27.31 | Arteriovenous malformation of vessel of upper limb |
| ICD10CM | Q27.32 | Arteriovenous malformation of vessel of lower limb |

Continued on next page

**TableS23 – continued from previous page**

| CodeSystem | ConceptCode | ConceptName |
|---|---|---|
| ICD10CM | Q27.33 | Arteriovenous malformation of digestive system vessel |
| ICD10CM | Q27.34 | Arteriovenous malformation of renal vessel |
| ICD10CM | Q27.39 | Arteriovenous malformation, other site |
| ICD10CM | Q27.4 | Congenital phlebectasia |
| ICD10CM | Q27.8 | Other specified congenital malformations of peripheral vascular system |
| ICD10CM | Q27.9 | Congenital malformation of peripheral vascular system, unspecified |
| ICD10CM | Q28.0 | Arteriovenous malformation of precerebral vessels |
| ICD10CM | Q28.1 | Other malformations of precerebral vessels |
| ICD10CM | Q28.2 | Arteriovenous malformation of cerebral vessels |
| ICD10CM | Q28.3 | Other malformations of cerebral vessels |
| ICD10CM | Q28.8 | Other specified congenital malformations of circulatory system |
| ICD10CM | Q28.9 | Congenital malformation of circulatory system, unspecified |
| ICD10CM | R00.0 | Tachycardia, unspecified |
| ICD10CM | R00.1 | Bradycardia, unspecified |
| ICD10CM | R00.2 | Palpitations |
| ICD10CM | R00.8 | Other abnormalities of heart beat |
| ICD10CM | R00.9 | Unspecified abnormalities of heart beat |
| ICD10CM | R01.0 | Benign and innocent cardiac murmurs |
| ICD10CM | R01.1 | Cardiac murmur, unspecified |
| ICD10CM | R01.2 | Other cardiac sounds |
| ICD10CM | R03.0 | Elevated blood-pressure reading, without diagnosis of hypertension |
| ICD10CM | R03.1 | Nonspecific low blood-pressure reading |
| ICD10CM | R07.2 | Precordial pain |
| ICD10CM | R07.82 | Intercostal pain |
| ICD10CM | R07.89 | Other chest pain |
| ICD10CM | R07.9 | Chest pain, unspecified |
| ICD10CM | R09.89 | Other specified symptoms and signs involving the circulatory and respiratory systems |
| ICD10CM | R58 | Hemorrhage, not elsewhere classified |
| ICD10CM | Z45.010 | Encounter for checking and testing of cardiac pacemaker pulse generator [battery] |
| ICD10CM | Z45.018 | Encounter for adjustment and management of other part of cardiac pacemaker |
| ICD10CM | Z45.02 | Encounter for adjustment and management of automatic implantable cardiac defibrillator |
| ICD10CM | Z45.09 | Encounter for adjustment and management of other cardiac device |
| ICD10CM | Z86.73 | Personal history of transient ischemic attack (TIA), and cerebral infarction without residual deficits |
| ICD10CM | Z86.74 | Personal history of sudden cardiac arrest |
| ICD10CM | Z86.79 | Personal history of other diseases of the circulatory system |
| ICD10CM | Z87.74 | Personal history of (corrected) congenital malformations of heart and circulatory system |
| ICD10CM | Z94.1 | Heart transplant status |

Continued on next page

**TableS23 – continued from previous page**

| CodeSystem | ConceptCode | ConceptName |
|---|---|---|
| ICD10CM | Z94.3 | Heart and lungs transplant status |
| ICD10CM | Z95.0 | Presence of cardiac pacemaker |
| ICD10CM | Z95.1 | Presence of aortocoronary bypass graft |
| ICD10CM | Z95.2 | Presence of prosthetic heart valve |
| ICD10CM | Z95.3 | Presence of xenogenic heart valve |
| ICD10CM | Z95.4 | Presence of other heart-valve replacement |
| ICD10CM | Z95.5 | Presence of coronary angioplasty implant and graft |
| ICD10CM | Z95.810 | Presence of automatic (implantable) cardiac defibrillator |
| ICD10CM | Z95.811 | Presence of heart assist device |
| ICD10CM | Z95.812 | Presence of fully implantable artificial heart |
| ICD10CM | Z95.818 | Presence of other cardiac implants and grafts |
| ICD10CM | Z95.820 | Peripheral vascular angioplasty status with implants and grafts |
| ICD10CM | Z95.828 | Presence of other vascular implants and grafts |
| ICD10CM | Z95.9 | Presence of cardiac and vascular implant and graft, unspecified |
| ICD10CM | Z98.61 | Coronary angioplasty status |
| ICD10CM | Z98.62 | Peripheral vascular angioplasty status |
| ICD10CM | I48.11 | Longstanding persistent atrial fibrillation |
| ICD10CM | I48.19 | Other persistent atrial fibrillation |
| ICD10CM | I48.20 | Chronic atrial fibrillation, unspecified |
| ICD10CM | I48.21 | Permanent atrial fibrillation |
| ICD10CM | K55.01 | Acute (reversible) ischemia of small intestine |
| ICD10CM | K55.02 | Acute infarction of small intestine |
| ICD10CM | K55.03 | Acute (reversible) ischemia of large intestine |
| ICD10CM | K55.04 | Acute infarction of large intestine |
| ICD10CM | K55.05 | Acute (reversible) ischemia of intestine, part unspecified |
| ICD10CM | K55.06 | Acute infarction of intestine, part unspecified |
| SNOMED CT | 266257000 | Transient cerebral ischemia |
| SNOMED CT | 275434003 | Stroke in the puerperium |
| SNOMED CT | 276220007 | Foville syndrome |
| SNOMED CT | 276222004 | Top of basilar syndrome |
| SNOMED CT | 276517000 | Transient myocardial ischemia of newborn |
| SNOMED CT | 281240008 | Extension of cerebrovascular accident |
| SNOMED CT | 300920004 | Carotid atherosclerosis |
| SNOMED CT | 300995000 | Exercise-induced angina |
| SNOMED CT | 302910002 | Atherosclerotic renal artery stenosis |
| SNOMED CT | 304914007 | Acute Q wave myocardial infarction |
| SNOMED CT | 307140009 | Acute non-Q wave infarction |
| SNOMED CT | 311792005 | Postoperative transmural myocardial infarction of anterior wall |
| SNOMED CT | 311793000 | Postoperative transmural myocardial infarction of inferior wall |
| SNOMED CT | 311796008 | Postoperative subendocardial myocardial infarction |
| SNOMED CT | 314116003 | Post infarct angina |
| SNOMED CT | 314207007 | Non-Q wave myocardial infarction |
| SNOMED CT | 315025001 | Refractory angina |
| SNOMED CT | 315348000 | Asymptomatic coronary heart disease |
| SNOMED CT | 361132001 | Senile arteriosclerosis |
| SNOMED CT | 361133006 | Arteriosclerosis obliterans |
| SNOMED CT | 371040005 | Thrombotic stroke |

Continued on next page

**TableS23 – continued from previous page**

| CodeSystem | ConceptCode | ConceptName |
|---|---|---|
| SNOMED CT | 371041009 | Embolic stroke |
| SNOMED CT | 371121002 | Neonatal stroke |
| SNOMED CT | 371803003 | Multi vessel coronary artery disease |
| SNOMED CT | 371804009 | Left main coronary artery disease |
| SNOMED CT | 371805005 | Significant coronary bypass graft disease |
| SNOMED CT | 371806006 | Progressive angina |
| SNOMED CT | 371807002 | Atypical angina |
| SNOMED CT | 371808007 | Recurrent angina after percutaneous transluminal coronary angioplasty |
| SNOMED CT | 371809004 | Recurrent angina following placement of coronary artery stent |
| SNOMED CT | 371810009 | Recurrent angina after coronary artery bypass graft |
| SNOMED CT | 371811008 | Recurrent angina post rotational atherectomy |
| SNOMED CT | 371812001 | Recurrent angina after directional coronary atherectomy |
| SNOMED CT | 373606000 | Occlusive stroke |
| SNOMED CT | 384993003 | Periventricular hemorrhagic venous infarct |
| SNOMED CT | 394659003 | Acute coronary syndrome |
| SNOMED CT | 394710008 | First myocardial infarction |
| SNOMED CT | 398274000 | Coronary artery thrombosis |
| SNOMED CT | 401303003 | Acute ST segment elevation myocardial infarction |
| SNOMED CT | 401314000 | Acute non-ST segment elevation myocardial infarction |
| SNOMED CT | 408546009 | Coronary artery bypass graft occlusion |
| SNOMED CT | 413102000 | Infarction of basal ganglia |
| SNOMED CT | 413439005 | Acute ischemic heart disease |
| SNOMED CT | 413444003 | Acute myocardial ischemia |
| SNOMED CT | 413758000 | Cardioembolic stroke |
| SNOMED CT | 418044006 | Myocardial infarction in recovery phase |
| SNOMED CT | 420006002 | Obliterative coronary artery disease |
| SNOMED CT | 421327009 | Coronary artery stent thrombosis |
| SNOMED CT | 422504002 | Ischemic stroke |
| SNOMED CT | 426814001 | Transient cerebral ischemia due to atrial fibrillation |
| SNOMED CT | 427919004 | Coronary arteriosclerosis due to radiation |
| SNOMED CT | 428196007 | Mixed myocardial ischemia and infarction |
| SNOMED CT | 428507003 | Atherosclerotic stenosis of brachiocephalic artery |
| SNOMED CT | 429245005 | Recurrent coronary arteriosclerosis after percutaneous transluminal coronary angioplasty |
| SNOMED CT | 429559004 | Typical angina |
| SNOMED CT | 441574008 | Atherosclerosis of artery |
| SNOMED CT | 442439008 | Atherosclerosis of bypass graft of limb |
| SNOMED CT | 442693003 | Atherosclerosis of autologous vein bypass graft of limb |
| SNOMED CT | 442701004 | Atherosclerosis of nonautologous biological bypass graft of limb |
| SNOMED CT | 442735001 | Atherosclerosis of nonautologous bypass graft of limb |
| SNOMED CT | 443502000 | Coronary atherosclerosis |
| SNOMED CT | 443971004 | Arteriosclerosis of artery of extremity |
| SNOMED CT | 444172003 | Recurrent transient cerebral ischemic attack |
| SNOMED CT | 444657001 | Superior cerebellar artery syndrome |
| SNOMED CT | 703164000 | Acute anterior ST segment elevation myocardial infarction |

Continued on next page

**TableS23 – continued from previous page**

| CodeSystem | ConceptCode | ConceptName |
|---|---|---|
| SNOMED CT | 703165004 | Acute ST segment elevation myocardial infarction of anterior wall involving right ventricle |
| SNOMED CT | 703209002 | Subsequent ST segment elevation myocardial infarction of inferior wall |
| SNOMED CT | 703210007 | Subsequent ST segment elevation myocardial infarction of anterior wall |
| SNOMED CT | 703211006 | Subsequent ST segment elevation myocardial infarction |
| SNOMED CT | 703212004 | Acute myocardial infarction during procedure |
| SNOMED CT | 703213009 | Acute ST segment elevation myocardial infarction of inferior wall |
| SNOMED CT | 703251009 | Acute myocardial infarction of inferior wall involving right ventricle |
| SNOMED CT | 703252002 | Acute myocardial infarction of anterior wall involving right ventricle |
| SNOMED CT | 703253007 | Acute ST segment elevation myocardial infarction of inferior wall involving right ventricle |
| SNOMED CT | 703356002 | Coronary artery occlusion due to neoplastic disease |
| SNOMED CT | 703360004 | Subsequent non-ST segment elevation myocardial infarction |
| SNOMED CT | 709584004 | Atherosclerosis of bypass graft of lower limb |
| SNOMED CT | 709585003 | Atherosclerosis of nonautologous biological bypass graft of lower limb |
| SNOMED CT | 709587006 | Atherosclerosis of autologous bypass graft of lower limb |
| SNOMED CT | 710575003 | Transient ischemic attack due to embolism |
| SNOMED CT | 713405002 | Subacute ischemic heart disease |
| SNOMED CT | 716051003 | CVA (cerebrovascular accident) during surgery |
| SNOMED CT | 719678003 | Non-obstructive atherosclerosis of coronary artery |
| SNOMED CT | 70211005 | Acute myocardial infarction of anterolateral wall |
| SNOMED CT | 70422006 | Acute subendocardial infarction |
| SNOMED CT | 70936005 | Multi-infarct dementia, uncomplicated |
| SNOMED CT | 70998009 | Acute myocardial infarction of posterobasal wall |
| SNOMED CT | 72092001 | Arteriosclerotic vascular disease |
| SNOMED CT | 73795002 | Acute myocardial infarction of inferior wall |
| SNOMED CT | 76593002 | Acute myocardial infarction of inferoposterior wall |
| SNOMED CT | 78569004 | Posterior inferior cerebellar artery syndrome |
| SNOMED CT | 79009004 | Acute myocardial infarction of septum |
| SNOMED CT | 81363003 | Malignant arteriolar nephrosclerosis |
| SNOMED CT | 81817003 | Atherosclerosis of aorta |
| SNOMED CT | 85284003 | Angina, class III |
| SNOMED CT | 87343002 | Prinzmetal angina |
| SNOMED CT | 88032003 | Amaurosis fugax |
| SNOMED CT | 89323001 | Angina, class IV |
| SNOMED CT | 90493000 | Arteriolar nephrosclerosis |
| SNOMED CT | 92517006 | Calcific coronary arteriosclerosis |
| SNOMED CT | 95691008 | Arteriosclerotic retinopathy |
| SNOMED CT | 111297002 | Nonparalytic stroke |
| SNOMED CT | 116288000 | Paralytic stroke |
| SNOMED CT | 123641001 | Left coronary artery occlusion |
| SNOMED CT | 123642008 | Right coronary artery occlusion |

Continued on next page

**TableS23 – continued from previous page**

| CodeSystem | ConceptCode | ConceptName |
|---|---|---|
| SNOMED CT | 129573006 | Atherosclerotic occlusive disease |
| SNOMED CT | 129574000 | Postoperative myocardial infarction |
| SNOMED CT | 194802003 | True posterior myocardial infarction |
| SNOMED CT | 194809007 | Acute myocardial infarction of atrium |
| SNOMED CT | 194821006 | Coronary thrombosis not resulting in myocardial infarction |
| SNOMED CT | 194823009 | Acute coronary insufficiency |
| SNOMED CT | 194828000 | Angina pectoris |
| SNOMED CT | 194842008 | Single coronary vessel disease |
| SNOMED CT | 194843003 | Double coronary vessel disease |
| SNOMED CT | 194856005 | Subsequent myocardial infarction |
| SNOMED CT | 194857001 | Subsequent myocardial infarction of anterior wall |
| SNOMED CT | 194858006 | Subsequent myocardial infarction of inferior wall |
| SNOMED CT | 195200006 | Carotid artery syndrome hemispheric |
| SNOMED CT | 195209007 | Middle cerebral artery syndrome |
| SNOMED CT | 195210002 | Anterior cerebral artery syndrome |
| SNOMED CT | 195211003 | Posterior cerebral artery syndrome |
| SNOMED CT | 195212005 | Brainstem stroke syndrome |
| SNOMED CT | 195213000 | Cerebellar stroke syndrome |
| SNOMED CT | 195216008 | Left sided cerebral hemisphere cerebrovascular accident |
| SNOMED CT | 195217004 | Right sided cerebral hemisphere cerebrovascular accident |
| SNOMED CT | 230690007 | Cerebrovascular accident |
| SNOMED CT | 230691006 | CVA - cerebrovascular accident due to cerebral artery occlusion |
| SNOMED CT | 230713003 | Stroke of uncertain pathology |
| SNOMED CT | 230714009 | Anterior circulation stroke of uncertain pathology |
| SNOMED CT | 230715005 | Posterior circulation stroke of uncertain pathology |
| SNOMED CT | 230716006 | Carotid territory transient ischemic attack |
| SNOMED CT | 230717002 | Vertebrobasilar territory transient ischemic attack |
| SNOMED CT | 233817007 | Triple vessel disease of the heart |
| SNOMED CT | 233819005 | Stable angina |
| SNOMED CT | 233821000 | New onset angina |
| SNOMED CT | 233825009 | Acute Q wave infarction - anteroseptal |
| SNOMED CT | 233826005 | Acute non-Q wave infarction - anteroseptal |
| SNOMED CT | 233827001 | Acute Q wave infarction - anterolateral |
| SNOMED CT | 233828006 | Acute non-Q wave infarction - anterolateral |
| SNOMED CT | 233829003 | Acute Q wave infarction - inferior |
| SNOMED CT | 233830008 | Acute non-Q wave infarction - inferior |
| SNOMED CT | 233831007 | Acute Q wave infarction - inferolateral |
| SNOMED CT | 233832000 | Acute non-Q wave infarction - inferolateral |
| SNOMED CT | 233833005 | Acute Q wave infarction - lateral |
| SNOMED CT | 233834004 | Acute non-Q wave infarction - lateral |
| SNOMED CT | 233835003 | Acute widespread myocardial infarction |
| SNOMED CT | 233836002 | Acute Q wave infarction - widespread |
| SNOMED CT | 233837006 | Acute non-Q wave infarction - widespread |
| SNOMED CT | 233838001 | Acute posterior myocardial infarction |
| SNOMED CT | 233839009 | Old anterior myocardial infarction |
| SNOMED CT | 233840006 | Old inferior myocardial infarction |
| SNOMED CT | 233841005 | Old lateral myocardial infarction |

Continued on next page

**TableS23 – continued from previous page**

| CodeSystem | ConceptCode | ConceptName |
|---|---|---|
| SNOMED CT | 233842003 | Old posterior myocardial infarction |
| SNOMED CT | 233843008 | Silent myocardial infarction |
| SNOMED CT | 233844002 | Accelerated coronary artery disease in transplanted heart |
| SNOMED CT | 233955003 | Abdominal aortic atherosclerosis |
| SNOMED CT | 233956002 | Aortoiliac atherosclerosis |
| SNOMED CT | 1755008 | Old myocardial infarction |
| SNOMED CT | 282006 | Acute myocardial infarction of basal-lateral wall |
| SNOMED CT | 4557003 | Preinfarction syndrome |
| SNOMED CT | 10273003 | Acute infarction of papillary muscle |
| SNOMED CT | 10349009 | Multi-infarct dementia with delirium |
| SNOMED CT | 10365005 | Right main coronary artery thrombosis |
| SNOMED CT | 14070001 | Multi-infarct dementia with depression |
| SNOMED CT | 14309005 | Anterior choroidal artery syndrome |
| SNOMED CT | 15990001 | Acute myocardial infarction of posterolateral wall |
| SNOMED CT | 19057007 | Status anginosus |
| SNOMED CT | 21470009 | Syncope anginosa |
| SNOMED CT | 22298006 | Myocardial infarction |
| SNOMED CT | 25106000 | Impending infarction |
| SNOMED CT | 25133001 | Completed stroke |
| SNOMED CT | 25772007 | Multi-infarct dementia with delusions |
| SNOMED CT | 28248000 | Left anterior descending coronary artery thrombosis |
| SNOMED CT | 29899005 | Coronary artery embolism |
| SNOMED CT | 30277009 | Rupture of ventricle due to acute myocardial infarction |
| SNOMED CT | 35928006 | Nocturnal angina |
| SNOMED CT | 39823006 | Generalized atherosclerosis |
| SNOMED CT | 41334000 | Angina, class II |
| SNOMED CT | 42531007 | Microinfarct of heart |
| SNOMED CT | 42866003 | Congenital coronary artery sclerosis |
| SNOMED CT | 45281005 | Atherosclerosis of renal artery |
| SNOMED CT | 46109009 | Subendocardial ischemia |
| SNOMED CT | 49176002 | Arteriosclerotic gangrene |
| SNOMED CT | 49778009 | Idiopathic pulmonary arteriosclerosis |
| SNOMED CT | 50570003 | Aneurysm of coronary vessels |
| SNOMED CT | 51274000 | Atherosclerosis of arteries of the extremities |
| SNOMED CT | 52035003 | Acute anteroapical myocardial infarction |
| SNOMED CT | 53741008 | Coronary arteriosclerosis |
| SNOMED CT | 54329005 | Acute myocardial infarction of anterior wall |
| SNOMED CT | 55382008 | Cerebral atherosclerosis |
| SNOMED CT | 56267009 | Multi-infarct dementia |
| SNOMED CT | 56276002 | Left main coronary artery thrombosis |
| SNOMED CT | 57054005 | Acute myocardial infarction |
| SNOMED CT | 57981008 | Progressing stroke |
| SNOMED CT | 58612006 | Acute myocardial infarction of lateral wall |
| SNOMED CT | 59021001 | Angina decubitus |
| SNOMED CT | 59063002 | Acute myocardial infarction of apical-lateral wall |
| SNOMED CT | 61490001 | Angina, class I |
| SNOMED CT | 62240004 | Benign arteriolar nephrosclerosis |

Continued on next page

**TableS23 – continued from previous page**

| CodeSystem | ConceptCode | ConceptName |
|---|---|---|
| SNOMED CT | 62695002 | Acute anteroseptal myocardial infarction |
| SNOMED CT | 63739005 | Coronary occlusion |
| SNOMED CT | 64009001 | Basilar artery syndrome |
| SNOMED CT | 64627002 | Acute myocardial infarction of high lateral wall |
| SNOMED CT | 65312002 | Cerebral arteriosclerosis |
| SNOMED CT | 65547006 | Acute myocardial infarction of inferolateral wall |
| SNOMED CT | 67682002 | Coronary artery atheroma |
| SNOMED CT | 722929005 | Perinatal arterial ischemic stroke of fetus and/or neonate |
| SNOMED CT | 723082006 | Silent cerebral infarct |
| SNOMED CT | 724424009 | Cerebral ischemic stroke due to small artery occlusion |
| SNOMED CT | 724425005 | Cerebral ischemic stroke due to intracranial large artery atherosclerosis |
| SNOMED CT | 724426006 | Cerebral ischemic stroke due to extracranial large artery atherosclerosis |
| SNOMED CT | 724429004 | Stroke co-occurrent with migraine |
| SNOMED CT | 724440007 | Atherosclerosis of artery of lower limb |
| SNOMED CT | 724993002 | Cerebral ischemic stroke due to occlusion of extracranial large artery |
| SNOMED CT | 724994008 | Cerebral ischemic stroke due to stenosis of extracranial large artery |
| SNOMED CT | 725132001 | Ischemic stroke without residual deficits |
| SNOMED CT | 788880006 | Cerebral ischemic stroke due to dissection of artery |
| SNOMED CT | 788881005 | Cerebral ischemic stroke due to aortic arch embolism |
| SNOMED CT | 788882003 | Cerebral ischemic stroke due to global hypoperfusion with watershed infarct |
| SNOMED CT | 788883008 | Cerebral ischemic stroke due to hypercoagulable state |
| SNOMED CT | 788884002 | Cerebral ischemic stroke due to subarachnoid hemorrhage |
| SNOMED CT | 789703007 | Arteriosclerosis of artery of lower extremity |
| SNOMED CT | 789704001 | Arteriosclerosis of artery of upper extremity |
| SNOMED CT | 836293000 | Acute myocardial infarction of right ventricle |
| SNOMED CT | 836294006 | Acute myocardial infarction of apex of heart |
| SNOMED CT | 836295007 | Acute myocardial infarction of inferolateral wall with posterior extension |
| SNOMED CT | 840309000 | Acute ST segment elevation myocardial infarction due to proximal left anterior descending coronary artery occlusion |
| SNOMED CT | 840310005 | Occlusion of proximal left anterior descending coronary artery |
| SNOMED CT | 840312002 | Acute ST segment elevation myocardial infarction due to mid left anterior descending coronary artery occlusion |
| SNOMED CT | 840313007 | Occlusion of mid left anterior descending coronary artery |
| SNOMED CT | 840315000 | Occlusion of distal left anterior descending coronary artery |
| SNOMED CT | 840316004 | Acute ST segment elevation myocardial infarction due to distal left anterior descending coronary artery occlusion |
| SNOMED CT | 840608004 | Occlusion of anterior descending branch of left coronary artery |
| SNOMED CT | 840609007 | Acute ST segment elevation myocardial infarction due to occlusion of anterior descending branch of left coronary artery |
| SNOMED CT | 840679006 | Occlusion of septal branch of anterior descending branch of left coronary artery |

Continued on next page

**TableS23 – continued from previous page**

| CodeSystem | ConceptCode | ConceptName |
|---|---|---|
| SNOMED CT | 840680009 | Acute ST segment elevation myocardial infarction due to occlusion of septal branch of anterior descending branch of left coronary artery |
| SNOMED CT | 846667001 | Occlusion of diagonal branch of anterior descending branch of left coronary artery |
| SNOMED CT | 846668006 | Acute ST segment elevation myocardial infarction due to occlusion of diagonal branch of anterior descending branch of left coronary artery |
| SNOMED CT | 846683001 | Acute ST segment elevation myocardial infarction due to occlusion of intermediate artery |
| SNOMED CT | 846684007 | Occlusion of intermediate artery |
| SNOMED CT | 868214006 | Acute ST segment elevation myocardial infarction due to occlusion of proximal portion of right coronary artery |
| SNOMED CT | 868215007 | Occlusion of proximal portion of right coronary artery |
| SNOMED CT | 868216008 | Occlusion of distal portion of right coronary artery |
| SNOMED CT | 868217004 | Acute ST segment elevation myocardial infarction due to occlusion of distal portion of right coronary artery |
| SNOMED CT | 868219001 | Occlusion of mid portion of right coronary artery |
| SNOMED CT | 868220007 | Acute ST segment elevation myocardial infarction due to occlusion of mid portion of right coronary artery |
| SNOMED CT | 868221006 | Occlusion of marginal branch of right coronary artery |
| SNOMED CT | 868222004 | Occlusion of posterior descending branch of right coronary artery |
| SNOMED CT | 868223009 | Occlusion of posterior lateral branch of right coronary artery |
| SNOMED CT | 868224003 | Acute ST segment elevation myocardial infarction due to occlusion of marginal branch of right coronary artery |
| SNOMED CT | 868225002 | Acute ST segment elevation myocardial infarction due to occlusion of posterior descending branch of right coronary artery |
| SNOMED CT | 868226001 | Acute ST segment elevation myocardial infarction due to occlusion of posterior lateral branch of right coronary artery |
| SNOMED CT | 5471000124102 | Ruptured aneurysm of intracranial artery |
| SNOMED CT | 5571000124103 | Cerebrovascular accident with intracranial hemorrhage |
| SNOMED CT | 8001000119106 | Atherosclerosis of aortoiliac bypass graft |
| SNOMED CT | 9901000119100 | Occlusion of cerebral artery with stroke |
| SNOMED CT | 17531000119105 | Acute myocardial infarction due to left coronary artery occlusion |
| SNOMED CT | 23311000119105 | Acute myocardial infarction due to right coronary artery occlusion |
| SNOMED CT | 78741000119103 | Acute coronary artery occlusion not resulting in myocardial infarction |
| SNOMED CT | 99451000119105 | Cerebral infarction due to stenosis of carotid artery |
| SNOMED CT | 106021000119105 | Multi-infarct dementia due to atherosclerosis |
| SNOMED CT | 117051000119103 | Chronic total occlusion of coronary artery |
| SNOMED CT | 139011000119104 | Coronary arteriosclerosis following coronary artery bypass graft |
| SNOMED CT | 140911000119109 | Ischemic stroke with coma |
| SNOMED CT | 140921000119102 | Ischemic stroke without coma |
| SNOMED CT | 284871000119105 | Atherosclerosis of left carotid artery |
| SNOMED CT | 284881000119108 | Atherosclerosis of right carotid artery |

Continued on next page

**TableS23 – continued from previous page**

| CodeSystem | ConceptCode | ConceptName |
|---|---|---|
| SNOMED CT | 285981000119103 | Acute ST segment elevation myocardial infarction involving left anterior descending coronary artery |
| SNOMED CT | 292661000119105 | Cerebrovascular accident due to stenosis of right vertebral artery |
| SNOMED CT | 292671000119104 | Cerebrovascular accident due to stenosis of left vertebral artery |
| SNOMED CT | 292681000119101 | Cerebrovascular accident due to right vertebral artery occlusion |
| SNOMED CT | 292691000119103 | Cerebrovascular accident due to occlusion of left vertebral artery |
| SNOMED CT | 329361000119107 | Cerebrovascular accident due to occlusion of right middle cerebral artery by embolus |
| SNOMED CT | 329371000119101 | Cerebrovascular accident due to occlusion of left middle cerebral artery by embolus |
| SNOMED CT | 329391000119100 | Right anterior cerebral artery embolism with stroke |
| SNOMED CT | 329401000119103 | Left anterior cerebral artery embolism with stroke |
| SNOMED CT | 329421000119107 | Cerebrovascular accident due to occlusion of right posterior cerebral artery by embolus |
| SNOMED CT | 329431000119105 | Cerebrovascular accident due to occlusion of left posterior cerebral artery by embolus |
| SNOMED CT | 329451000119104 | Cerebrovascular accident due to occlusion of right cerebellar artery by embolus |
| SNOMED CT | 329461000119102 | Cerebrovascular accident due to occlusion of left cerebellar artery by embolus |
| SNOMED CT | 329621000119105 | Cerebrovascular accident due to thrombus of right vertebral artery |
| SNOMED CT | 329631000119108 | Left vertebral artery thrombosis with stroke |
| SNOMED CT | 329641000119104 | Cerebrovascular accident due to thrombus of basilar artery |
| SNOMED CT | 329651000119102 | Cerebrovascular accident due to thrombus of right carotid artery |
| SNOMED CT | 330111000119103 | Ruptured acquired aneurysm of cerebral artery |
| SNOMED CT | 330791000119108 | Cerebrovascular accident due to thrombus of left carotid artery |
| SNOMED CT | 380001000004106 | Subendocardial myocardial infarction |
| SNOMED CT | 431391000124106 | Arteriosclerosis of aorta |
| SNOMED CT | 431401000124108 | Arteriosclerosis of abdominal aorta |
| SNOMED CT | 431411000124106 | Arteriosclerosis of thoracic aorta |
| SNOMED CT | 431421000124103 | Arteriosclerosis of carotid artery |
| SNOMED CT | 431431000124100 | Arteriosclerosis of renal artery |
| SNOMED CT | 434141000124103 | Chronic cerebrovascular accident |
| SNOMED CT | 436031000124102 | Anterior inferior cerebellar artery occlusion with infarction |
| SNOMED CT | 436041000124107 | Posterior inferior cerebellar artery occlusion with infarction |
| SNOMED CT | 451041000124103 | Atherosclerosis of coronary artery without angina pectoris |
| SNOMED CT | 451361000124102 | Lipid-rich atherosclerosis of coronary artery |
| SNOMED CT | 457551000124104 | Acute stroke |
| SNOMED CT | 461431000124108 | Vertebral artery atherosclerosis |
| SNOMED CT | 461441000124103 | Aortic arch atherosclerosis |
| SNOMED CT | 11018701000119109 | Coronary arteriosclerosis after percutaneous coronary angioplasty |
| SNOMED CT | 12237911000119109 | Amaurosis fugax of left eye |
| SNOMED CT | 12237951000119105 | Amaurosis fugax of right eye |
| SNOMED CT | 12238111000119106 | Acute ST segment elevation myocardial infarction of inferolateral wall |

Continued on next page

**TableS23 – continued from previous page**

| CodeSystem | ConceptCode | ConceptName |
|---|---|---|
| SNOMED CT | 12238151000119107 | Acute ST segment elevation myocardial infarction of inferoposterior wall |
| SNOMED CT | 15648641000119100 | Atherosclerosis of artery of bilateral upper limbs |
| SNOMED CT | 15712841000119100 | Acute ST segment elevation myocardial infarction of posterolateral wall |
| SNOMED CT | 15712881000119105 | Acute ST segment elevation myocardial infarction of anterolateral wall |
| SNOMED CT | 15712921000119103 | Acute ST segment elevation myocardial infarction of lateral wall |
| SNOMED CT | 15712961000119108 | Acute ST segment elevation myocardial infarction of anteroseptal wall |
| SNOMED CT | 15713041000119103 | Acute ST segment elevation myocardial infarction of posterior wall |
| SNOMED CT | 15713161000119100 | Acute ST segment elevation myocardial infarction of septum |
| SNOMED CT | 15713201000119105 | Acute ST segment elevation myocardial infarction of posterobasal wall |
| SNOMED CT | 15960061000119102 | Unstable angina co-occurrent and due to coronary arteriosclerosis |
| SNOMED CT | 15960141000119102 | Angina co-occurrent and due to coronary arteriosclerosis |
| SNOMED CT | 15960381000119109 | Angina co-occurrent and due to arteriosclerosis of coronary artery bypass graft |
| SNOMED CT | 15960541000119107 | Unstable angina due to arteriosclerosis of autologous vein coronary artery bypass graft |
| SNOMED CT | 15960581000119102 | Arteriosclerosis of autologous vein coronary artery bypass graft with angina |
| SNOMED CT | 15960661000119107 | Unstable angina co-occurrent and due to arteriosclerosis of coronary artery bypass graft |
| SNOMED CT | 15960781000119108 | Arteriosclerosis of autologous arterial coronary artery bypass graft with angina |
| SNOMED CT | 15962541000119106 | Acute ST segment elevation myocardial infarction of anteroapical wall |
| SNOMED CT | 16000351000119109 | Cerebrovascular accident due to occlusion of left posterior cerebral artery |
| SNOMED CT | 16000391000119104 | Cerebrovascular accident due to occlusion of right posterior cerebral artery |
| SNOMED CT | 16000431000119109 | Cerebrovascular accident due to occlusion of right middle cerebral artery |
| SNOMED CT | 16000511000119103 | Cerebrovascular accident due to occlusion of left middle cerebral artery |
| SNOMED CT | 16002031000119102 | Cerebrovascular accident due to thrombus of right middle cerebral artery |
| SNOMED CT | 16002071000119104 | Cerebrovascular accident due to thrombus of right cerebellar artery |
| SNOMED CT | 16002111000119106 | Cerebrovascular accident due to thrombus of left middle cerebral artery |
| SNOMED CT | 16002151000119107 | Nonpyogenic cerebral venous thrombosis with stroke |
| SNOMED CT | 16002191000119102 | Left cerebellar artery thrombosis with stroke |

Continued on next page

**TableS23 – continued from previous page**

| CodeSystem | ConceptCode | ConceptName |
|---|---|---|
| SNOMED CT | 16002231000119106 | Cerebrovascular accident due to thrombus of right posterior cerebral artery |
| SNOMED CT | 16002271000119109 | Left posterior cerebral artery thrombosis with stroke |
| SNOMED CT | 16002351000119105 | Right vertebral artery embolism with stroke |
| SNOMED CT | 16002391000119100 | Left vertebral artery embolism with stroke |
| SNOMED CT | 16002431000119105 | Right carotid artery embolism with stroke |
| SNOMED CT | 16002471000119108 | Left carotid artery embolism with stroke |
| SNOMED CT | 16002511000119104 | Basilar artery embolism with stroke |
| SNOMED CT | 16023911000119108 | Cerebrovascular accident due to right carotid artery occlusion |
| SNOMED CT | 16023991000119104 | Cerebrovascular accident due to occlusion of left pontine artery |
| SNOMED CT | 16024031000119100 | Cerebrovascular accident due to occlusion of right pontine artery |
| SNOMED CT | 16024111000119109 | Cerebrovascular accident due to occlusion of left carotid artery |
| SNOMED CT | 16024151000119105 | Cerebrovascular accident due to occlusion of left cerebellar artery |
| SNOMED CT | 16024271000119107 | Cerebrovascular accident due to occlusion of right cerebellar artery |
| SNOMED CT | 16026951000119102 | Cerebrovascular accident due to right carotid artery stenosis |
| SNOMED CT | 16371781000119100 | Cerebellar stroke |
| SNOMED CT | 16644681000119102 | Cerebrovascular accident due to occlusion of bilateral pontine arteries |
| SNOMED CT | 16661931000119102 | Cerebrovascular accident due to stenosis of bilateral vertebral arteries |
| SNOMED CT | 16661971000119104 | Cerebrovascular accident due to stenosis of bilateral carotid arteries |
| SNOMED CT | 16754391000119100 | Stable angina due to coronary arteriosclerosis |
| SNOMED CT | 16837681000119104 | Myocardial infarction due to demand ischemia |
| SNOMED CT | 16891151000119103 | Coronary artery disease due to type 2 diabetes mellitus |
| SNOMED CT | 19830001000004106 | Thrombosis of left circumflex artery |
| SNOMED CT | 24654003 | Weber syndrome |
| SNOMED CT | 32574007 | Past myocardial infarction diagnosed on electrocardiogram AND/OR other special investigation, but currently presenting no symptoms |
| SNOMED CT | 39925003 | Mitochondrial myopathy, encephalopathy, lactic acidosis and stroke-like episodes |
| SNOMED CT | 69742007 | Monckeberg medial sclerosis |
| SNOMED CT | 87555007 | Rubrospinal cerebellar peduncle syndrome |
| SNOMED CT | 276221006 | Gubler syndrome |
| SNOMED CT | 791000119109 | Angina due to type 2 diabetes mellitus |
| SNOMED CT | 284861000119104 | Atherosclerosis of bilateral carotid arteries |
| SNOMED CT | 15713081000119108 | Acute ST segment elevation myocardial infarction involving left main coronary artery |
| SNOMED CT | 15713121000119105 | Acute STEMI (ST elevation myocardial infarction) due to RCA (right coronary artery) occlusion |
| SNOMED CT | 16026991000119107 | Cerebrovascular accident due to stenosis of left carotid artery |
| SNOMED CT | 429673002 | Arteriosclerosis of coronary artery bypass graft |
| SNOMED CT | 442224005 | Arteriosclerosis of autologous vein coronary artery bypass graft |
| SNOMED CT | 442240008 | Arteriosclerosis of nonautologous coronary artery bypass graft |
| SNOMED CT | 442421004 | Arteriosclerosis of arterial coronary artery bypass graft |

Continued on next page

**TableS23 – continued from previous page**

| CodeSystem | ConceptCode | ConceptName |
|---|---|---|
| SNOMED CT | 444855007 | Arteriosclerosis of coronary artery bypass graft of transplanted heart |
| SNOMED CT | 444856008 | Arteriosclerosis of internal mammary artery coronary artery bypass graft |
| SNOMED CT | 720519003 | Atherosclerosis, deafness, diabetes, epilepsy, nephropathy syndrome |
| SNOMED CT | 204378009 | Congenital coronary aneurysm |
| SNOMED CT | 723862008 | Atherosclerosis of non-autologous coronary artery bypass graft |
| SNOMED CT | 724431008 | Atherosclerosis of autologous coronary artery bypass graft |
| SNOMED CT | 735565009 | Perforation of coronary artery co-occurrent and due to aneurysm of coronary artery |
| SNOMED CT | 735566005 | Rupture of coronary artery co-occurrent and due to aneurysm of coronary artery |
| SNOMED CT | 792844003 | Limb pain at rest due to atherosclerosis of artery of lower limb |
| SNOMED CT | 285141000119106 | Arteriosclerosis of autologous arterial coronary artery bypass graft |
| SNOMED CT | 285151000119108 | Arteriosclerosis of autologous coronary artery bypass graft |
| SNOMED CT | 329671000119106 | Traumatic subarachnoid hemorrhage with loss of consciousness |
| SNOMED CT | 430851000124104 | Multiple and bilateral precerebral artery stenosis with infarction |
| SNOMED CT | 434881000124100 | Multiple and bilateral precerebral arterial occlusion with infarction |
| SNOMED CT | 455941000124104 | Occlusion of circumflex coronary artery |
| SNOMED CT | 12236911000119107 | Bilateral atherosclerosis of arteries of lower limbs |
| SNOMED CT | 12236991000119103 | Bilateral lower limb atherosclerosis pain at rest co-occurrent and due to atherosclerosis |
| SNOMED CT | 12237031000119103 | Atherosclerosis of arteries of left lower limb |
| SNOMED CT | 12237111000119107 | Pain at rest of left lower limb co-occurrent and due to atherosclerosis |
| SNOMED CT | 12237151000119108 | Atherosclerosis of arteries of right lower limb |
| SNOMED CT | 12237231000119107 | Pain at rest of right lower limb co-occurrent and due to atherosclerosis |
| SNOMED CT | 15648601000119102 | Atherosclerosis of artery of right upper limb |
| SNOMED CT | 15648691000119108 | Atherosclerosis of artery of left upper limb |
| SNOMED CT | 15648731000119101 | Atherosclerosis of right renal artery |
| SNOMED CT | 15648771000119103 | Atherosclerosis of bilateral renal arteries |
| SNOMED CT | 15648821000119105 | Atherosclerosis of left renal artery |
| SNOMED CT | 15649941000119102 | Bilateral atherosclerosis of lower limbs with gangrene |
| SNOMED CT | 15710281000119108 | Atherosclerosis of autologous bypass graft of right lower extremity |
| SNOMED CT | 15710361000119108 | Atherosclerosis of nonautologous biological bypass graft of left lower limb |
| SNOMED CT | 15710401000119104 | Atherosclerosis of bypass graft of right lower limb |
| SNOMED CT | 15710441000119102 | Atherosclerosis of bypass graft of left lower limb |
| SNOMED CT | 15710481000119107 | Atherosclerosis of nonautologous biological bypass graft of right lower limb |
| SNOMED CT | 15710521000119107 | Atherosclerosis of autologous bypass graft of left lower extremity |
| SNOMED CT | 15710561000119102 | Atherosclerosis of autologous bypass graft of bilateral lower limbs |

Continued on next page

**TableS23 – continued from previous page**

| CodeSystem | ConceptCode | ConceptName |
|---|---|---|
| SNOMED CT | 15710601000119102 | Atherosclerosis of bypass graft of bilateral lower limbs |
| SNOMED CT | 15963181000119104 | Acute ST segment elevation myocardial infarction due to occlusion of circumflex coronary artery |
| SNOMED CT | 16000471000119107 | Cerebrovascular accident due to occlusion of left anterior cerebral artery |
| SNOMED CT | 16000551000119102 | Cerebrovascular accident due to occlusion of right anterior cerebral artery |
| SNOMED CT | 16013351000119104 | Claudication of bilateral upper limbs co-occurrent and due to atherosclerosis |
| SNOMED CT | 16013711000119104 | Intermittent claudication of right lower limb co-occurrent and due to atherosclerosis of bypass graft |
| SNOMED CT | 16013871000119104 | Claudication of right upper limb co-occurrent and due to atherosclerosis |
| SNOMED CT | 16014031000119109 | Atherosclerosis of bypass graft of left lower limb with intermittent claudication |
| SNOMED CT | 16014071000119107 | Atherosclerosis of bypass graft of bilateral lower limbs with intermittent claudication |
| SNOMED CT | 16014231000119100 | Claudication of left upper limb co-occurrent and due to atherosclerosis |
| SNOMED CT | 16018831000119106 | Atherosclerosis of bilateral iliac arteries |
| SNOMED CT | 16018911000119107 | Bilateral lower limb atherosclerosis of non-biological bypass graft |
| SNOMED CT | 16019471000119104 | Atherosclerosis of left iliac artery |
| SNOMED CT | 16019511000119108 | Atherosclerosis of right iliac artery |
| SNOMED CT | 16024191000119100 | Cerebrovascular accident due to occlusion of left posterior communicating artery |
| SNOMED CT | 16024231000119109 | Cerebrovascular accident due to occlusion of right posterior communicating artery |
| SNOMED CT | 16025471000119107 | Aneurysm of coronary artery due to and following acute febrile mucocutaneous lymph node syndrome |
| SNOMED CT | 16064811000119108 | Atherosclerosis of bypass graft of left lower limb with gangrene |
| SNOMED CT | 16066731000119102 | Pain at rest of right lower limb co-occurrent and due to atherosclerosis of bypass graft |
| SNOMED CT | 16899261000119106 | Atherosclerosis of superior mesenteric artery |
| SNOMED CT | 16065251000119109 | Atherosclerosis of bypass graft of right lower limb with gangrene |
| SNOMED CT | 15960341000119104 | Unstable angina due to arteriosclerosis of coronary artery bypass graft of transplanted heart |
| SNOMED CT | 15960461000119105 | Unstable angina due to arteriosclerosis of autologous arterial coronary artery bypass graft |
| SNOMED CT | 16018991000119103 | Atherosclerosis of autologous vein bypass graft of left upper limb |
| SNOMED CT | 16019031000119105 | Atherosclerosis of autologous vein bypass graft of right upper limb |
| SNOMED CT | 16019351000119102 | Atherosclerosis of bypass graft of left upper limb |
| SNOMED CT | 16019431000119102 | Atherosclerosis of bypass graft of right upper limb |
| SNOMED CT | 16891111000119104 | Idiopathic ischemic stroke |
| SNOMED CT | 16891191000119108 | Coronary artery disease due to type 1 diabetes mellitus |
| SNOMED CT | 595899961000119100 | Cerebrovascular accident of basal ganglia |
| SNOMED CT | 652287331000119104 | Cerebrovascular accident of brainstem |

Continued on next page

**TableS23 – continued from previous page**

| CodeSystem | ConceptCode | ConceptName |
|---|---|---|
| SNOMED CT | 726499301000119105 | Myocardial infarction due to atherothrombotic coronary artery disease |
| SNOMED CT | 879955009 | Myocardial infarction with non-obstructive coronary artery |
| SNOMED CT | 881694631000119107 | Cerebrovascular accident of medulla oblongata |
| SNOMED CT | 896689003 | Acute myocardial infarction due to occlusion of circumflex branch of left coronary artery |
| SNOMED CT | 896690007 | Occlusion of circumflex branch of left coronary artery |
| SNOMED CT | 896691006 | Acute ST segment elevation myocardial infarction due to occlusion of circumflex branch of left coronary artery |
| SNOMED CT | 896696001 | Acute ST segment elevation myocardial infarction of apex of heart |
| SNOMED CT | 896697005 | Acute ST segment elevation myocardial infarction of right ventricle |
| SNOMED CT | 915141931000119109 | Cerebrovascular accident due to embolism of basilar artery |
| SNOMED CT | 107557061000119108 | Cerebrovascular accident due to embolism of bilateral anterior cerebral arteries |
| SNOMED CT | 1142056004 | Neonatal ischemic stroke |
| SNOMED CT | 117776611000119102 | Cerebrovascular accident due to occlusion of left posterior communicating artery |
| SNOMED CT | 152148641000119104 | Cerebrovascular accident due to embolism of bilateral carotid arteries |
| SNOMED CT | 15710321000119103 | Bilateral atherosclerosis of nonautologous biological bypass graft of lower limbs |
| SNOMED CT | 16019311000119103 | Left lower limb atherosclerosis of nonbiological bypass graft |
| SNOMED CT | 16019391000119107 | Right lower limb atherosclerosis of nonbiological bypass graft |
| SNOMED CT | 168747591000119109 | Cerebrovascular accident due to embolism of bilateral posterior cerebral arteries |
| SNOMED CT | 182960891000119101 | Cerebrovascular accident due to occlusion of left anterior cerebral artery |
| SNOMED CT | 188174841000119103 | Cerebrovascular accident due to occlusion of bilateral middle cerebral arteries |
| SNOMED CT | 239965291000119107 | Cerebrovascular accident due to occlusion of basilar artery |
| SNOMED CT | 251770561000119107 | Cerebrovascular accident due to embolism of left anterior cerebral artery |
| SNOMED CT | 346674811000119104 | Cerebrovascular accident due to occlusion of bilateral cerebellar arteries |
| SNOMED CT | 384430101000119103 | Cerebrovascular accident due to embolism of right carotid artery |
| SNOMED CT | 38595071000119104 | Cerebrovascular accident due to thrombosis of right posterior cerebral artery |
| SNOMED CT | 496369931000119104 | Cerebrovascular accident due to thrombosis of right cerebellar artery |
| SNOMED CT | 511452481000119102 | Cerebrovascular accident due to thrombosis of right vertebral artery |
| SNOMED CT | 517253051000119105 | Cerebrovascular accident due to embolism of left vertebral artery |
| SNOMED CT | 58173271000119101 | Cerebrovascular accident due to embolism of bilateral cerebellar arteries |

Continued on next page

**TableS23 – continued from previous page**

| CodeSystem | ConceptCode | ConceptName |
|---|---|---|
| SNOMED CT | 655081461000119101 | Cerebrovascular accident due to occlusion of right anterior cerebral artery |
| SNOMED CT | 720261501000119107 | Cerebrovascular accident due to occlusion of bilateral anterior cerebral arteries |
| SNOMED CT | 732330391000119107 | Cerebrovascular accident due to embolism of bilateral vertebral arteries |
| SNOMED CT | 759950981000119101 | Cerebrovascular accident due to thrombosis of left vertebral artery |
| SNOMED CT | 769023031000119104 | Cerebrovascular accident of thalamus |
| SNOMED CT | 806161651000119106 | Cerebrovascular accident due to embolism of right vertebral artery |
| SNOMED CT | 849488701000119104 | Cerebrovascular accident due to embolism of right anterior cerebral artery |
| SNOMED CT | 849579281000119106 | Cerebrovascular accident due to occlusion of right posterior communicating artery |
| SNOMED CT | 851365731000119106 | Cerebrovascular accident due to thrombosis of left posterior cerebral artery |
| SNOMED CT | 859422751000119101 | Cerebrovascular accident due to embolism of left carotid artery |
| SNOMED CT | 86553761000119103 | Cerebrovascular accident due to occlusion of bilateral posterior cerebral arteries |
| SNOMED CT | 898941951000119108 | Cerebrovascular accident due to occlusion of bilateral vertebral arteries |
| SNOMED CT | 939885431000119109 | Cerebrovascular accident due to embolism of bilateral middle cerebral arteries |
| SNOMED CT | 957319791000119104 | Cerebrovascular accident due to thrombosis of left cerebellar artery |
| Truveta | 1215926 | Cerebrovascular accident (CVA) associated with severe acute respiratory syndrome coronavirus 2 (SARS-CoV-2) infection |
| SNOMED CT | 1169367003 | Multifocal atherosclerosis |
| SNOMED CT | 1204170004 | Atherosclerosis of abdominal visceral artery |
| SNOMED CT | 1204144000 | Atherosclerosis of axillary artery |
| SNOMED CT | 1204145004 | Atherosclerosis of brachial artery |
| SNOMED CT | 1204175009 | Atherosclerosis of crural artery |
| SNOMED CT | 1204176005 | Atherosclerosis of iliac artery |
| SNOMED CT | 1204173002 | Atherosclerosis of popliteal artery |
| SNOMED CT | 1204174008 | Atherosclerosis of subclavian artery |
| SNOMED CT | 1204172007 | Atherosclerosis of superficial femoral artery |
| SNOMED CT | 1204151009 | Acute inferior non-ST segment elevation myocardial infarction of right ventricle |
| SNOMED CT | 1204154001 | Acute anterior non-ST segment elevation myocardial infarction with right ventricular involvement |
| SNOMED CT | 1204155000 | Acute anterior non-ST segment elevation myocardial infarction |
| SNOMED CT | 1204152002 | Acute inferior non-ST segment elevation myocardial infarction |
| SNOMED CT | 1204222000 | Acute non-ST segment elevation myocardial infarction of right ventricle |
| SNOMED CT | 1197429000 | Cathepsin A-related arteriopathy, strokes, leukoencephalopathy |
| SNOMED CT | 1231168008 | Malignant middle cerebral artery syndrome |

Continued on next page

**TableS23 – continued from previous page**

| CodeSystem | ConceptCode | ConceptName |
|---|---|---|
| SNOMED CT | 1197363004 | Pediatric arterial ischemic stroke |
| SNOMED CT | 1208872002 | Subsequent anterior non-ST segment elevation myocardial infarction |
| SNOMED CT | 1208873007 | Subsequent inferior non-ST segment elevation myocardial infarction |
| SNOMED CT | 1208871009 | Transient ischemic attack co-occurrent with subarachnoid hemorrhage |
| SNOMED CT | 293671000119109 | Behavioral disturbance due to multi-infarct dementia |
| SNOMED CT | 1163440003 | Postoperative acute myocardial infarction |

#### 5.3 Cancer

Table S24: Concept codes used to identify if a patient was diagnosed with cancer.

| CodeSystem | ConceptCode | ConceptName |
|---|---|---|
| ICD10CM | C00.0 | Malignant neoplasm of external upper lip |
| ICD10CM | C00.1 | Malignant neoplasm of external lower lip |
| ICD10CM | C00.2 | Malignant neoplasm of external lip, unspecified |
| ICD10CM | C00.3 | Malignant neoplasm of upper lip, inner aspect |
| ICD10CM | C00.4 | Malignant neoplasm of lower lip, inner aspect |
| ICD10CM | C00.5 | Malignant neoplasm of lip, unspecified, inner aspect |
| ICD10CM | C00.6 | Malignant neoplasm of commissure of lip, unspecified |
| ICD10CM | C00.8 | Malignant neoplasm of overlapping sites of lip |
| ICD10CM | C00.9 | Malignant neoplasm of lip, unspecified |
| ICD10CM | C01 | Malignant neoplasm of base of tongue |
| ICD10CM | C02.0 | Malignant neoplasm of dorsal surface of tongue |
| ICD10CM | C02.1 | Malignant neoplasm of border of tongue |
| ICD10CM | C02.2 | Malignant neoplasm of ventral surface of tongue |
| ICD10CM | C02.3 | Malignant neoplasm of anterior two-thirds of tongue, part unspecified |
| ICD10CM | C02.4 | Malignant neoplasm of lingual tonsil |
| ICD10CM | C02.8 | Malignant neoplasm of overlapping sites of tongue |
| ICD10CM | C02.9 | Malignant neoplasm of tongue, unspecified |
| ICD10CM | C03.0 | Malignant neoplasm of upper gum |
| ICD10CM | C03.1 | Malignant neoplasm of lower gum |
| ICD10CM | C03.9 | Malignant neoplasm of gum, unspecified |
| ICD10CM | C04.0 | Malignant neoplasm of anterior floor of mouth |
| ICD10CM | C04.1 | Malignant neoplasm of lateral floor of mouth |
| ICD10CM | C04.8 | Malignant neoplasm of overlapping sites of floor of mouth |
| ICD10CM | C04.9 | Malignant neoplasm of floor of mouth, unspecified |
| ICD10CM | C05.0 | Malignant neoplasm of hard palate |
| ICD10CM | C05.1 | Malignant neoplasm of soft palate |
| ICD10CM | C05.2 | Malignant neoplasm of uvula |
| ICD10CM | C05.8 | Malignant neoplasm of overlapping sites of palate |

Continued on next page

**TableS24 – continued from previous page**

| CodeSystem | ConceptCode | ConceptName |
|---|---|---|
| ICD10CM | C05.9 | Malignant neoplasm of palate, unspecified |
| ICD10CM | C06.0 | Malignant neoplasm of cheek mucosa |
| ICD10CM | C06.1 | Malignant neoplasm of vestibule of mouth |
| ICD10CM | C06.2 | Malignant neoplasm of retromolar area |
| ICD10CM | C06.80 | Malignant neoplasm of overlapping sites of unspecified parts of mouth |
| ICD10CM | C06.89 | Malignant neoplasm of overlapping sites of other parts of mouth |
| ICD10CM | C06.9 | Malignant neoplasm of mouth, unspecified |
| ICD10CM | C07 | Malignant neoplasm of parotid gland |
| ICD10CM | C08.0 | Malignant neoplasm of submandibular gland |
| ICD10CM | C08.1 | Malignant neoplasm of sublingual gland |
| ICD10CM | C08.9 | Malignant neoplasm of major salivary gland, unspecified |
| ICD10CM | C09.0 | Malignant neoplasm of tonsillar fossa |
| ICD10CM | C09.1 | Malignant neoplasm of tonsillar pillar (anterior) (posterior) |
| ICD10CM | C09.8 | Malignant neoplasm of overlapping sites of tonsil |
| ICD10CM | C09.9 | Malignant neoplasm of tonsil, unspecified |
| ICD10CM | C10.0 | Malignant neoplasm of vallecula |
| ICD10CM | C10.1 | Malignant neoplasm of anterior surface of epiglottis |
| ICD10CM | C10.2 | Malignant neoplasm of lateral wall of oropharynx |
| ICD10CM | C10.3 | Malignant neoplasm of posterior wall of oropharynx |
| ICD10CM | C10.4 | Malignant neoplasm of branchial cleft |
| ICD10CM | C10.8 | Malignant neoplasm of overlapping sites of oropharynx |
| ICD10CM | C10.9 | Malignant neoplasm of oropharynx, unspecified |
| ICD10CM | C11.0 | Malignant neoplasm of superior wall of nasopharynx |
| ICD10CM | C11.1 | Malignant neoplasm of posterior wall of nasopharynx |
| ICD10CM | C11.2 | Malignant neoplasm of lateral wall of nasopharynx |
| ICD10CM | C11.3 | Malignant neoplasm of anterior wall of nasopharynx |
| ICD10CM | C11.8 | Malignant neoplasm of overlapping sites of nasopharynx |
| ICD10CM | C11.9 | Malignant neoplasm of nasopharynx, unspecified |
| ICD10CM | C12 | Malignant neoplasm of pyriform sinus |
| ICD10CM | C13.0 | Malignant neoplasm of postcricoid region |
| ICD10CM | C13.1 | Malignant neoplasm of aryepiglottic fold, hypopharyngeal aspect |
| ICD10CM | C13.2 | Malignant neoplasm of posterior wall of hypopharynx |
| ICD10CM | C13.8 | Malignant neoplasm of overlapping sites of hypopharynx |
| ICD10CM | C13.9 | Malignant neoplasm of hypopharynx, unspecified |
| ICD10CM | C14.0 | Malignant neoplasm of pharynx, unspecified |
| ICD10CM | C14.2 | Malignant neoplasm of Waldeyer's ring |
| ICD10CM | C14.8 | Malignant neoplasm of overlapping sites of lip, oral cavity and pharynx |
| ICD10CM | C15.3 | Malignant neoplasm of upper third of esophagus |
| ICD10CM | C15.4 | Malignant neoplasm of middle third of esophagus |
| ICD10CM | C15.5 | Malignant neoplasm of lower third of esophagus |
| ICD10CM | C15.8 | Malignant neoplasm of overlapping sites of esophagus |
| ICD10CM | C15.9 | Malignant neoplasm of esophagus, unspecified |
| ICD10CM | C16.0 | Malignant neoplasm of cardia |
| ICD10CM | C16.1 | Malignant neoplasm of fundus of stomach |
| ICD10CM | C16.2 | Malignant neoplasm of body of stomach |

Continued on next page

**TableS24 – continued from previous page**

| CodeSystem | ConceptCode | ConceptName |
|---|---|---|
| ICD10CM | C16.3 | Malignant neoplasm of pyloric antrum |
| ICD10CM | C16.4 | Malignant neoplasm of pylorus |
| ICD10CM | C16.5 | Malignant neoplasm of lesser curvature of stomach, unspecified |
| ICD10CM | C16.6 | Malignant neoplasm of greater curvature of stomach, unspecified |
| ICD10CM | C16.8 | Malignant neoplasm of overlapping sites of stomach |
| ICD10CM | C16.9 | Malignant neoplasm of stomach, unspecified |
| ICD10CM | C17.0 | Malignant neoplasm of duodenum |
| ICD10CM | C17.1 | Malignant neoplasm of jejunum |
| ICD10CM | C17.2 | Malignant neoplasm of ileum |
| ICD10CM | C17.3 | Meckel's diverticulum, malignant |
| ICD10CM | C17.8 | Malignant neoplasm of overlapping sites of small intestine |
| ICD10CM | C17.9 | Malignant neoplasm of small intestine, unspecified |
| ICD10CM | C18.0 | Malignant neoplasm of cecum |
| ICD10CM | C18.1 | Malignant neoplasm of appendix |
| ICD10CM | C18.2 | Malignant neoplasm of ascending colon |
| ICD10CM | C18.3 | Malignant neoplasm of hepatic flexure |
| ICD10CM | C18.4 | Malignant neoplasm of transverse colon |
| ICD10CM | C18.5 | Malignant neoplasm of splenic flexure |
| ICD10CM | C18.6 | Malignant neoplasm of descending colon |
| ICD10CM | C18.7 | Malignant neoplasm of sigmoid colon |
| ICD10CM | C18.8 | Malignant neoplasm of overlapping sites of colon |
| ICD10CM | C18.9 | Malignant neoplasm of colon, unspecified |
| ICD10CM | C19 | Malignant neoplasm of rectosigmoid junction |
| ICD10CM | C20 | Malignant neoplasm of rectum |
| ICD10CM | C21.2 | Malignant neoplasm of cloacogenic zone |
| ICD10CM | C21.8 | Malignant neoplasm of overlapping sites of rectum, anus and anal canal |
| ICD10CM | C26.0 | Malignant neoplasm of intestinal tract, part unspecified |
| ICD10CM | C26.1 | Malignant neoplasm of spleen |
| ICD10CM | C26.9 | Malignant neoplasm of ill-defined sites within the digestive system |
| ICD10CM | C30.0 | Malignant neoplasm of nasal cavity |
| ICD10CM | C30.1 | Malignant neoplasm of middle ear |
| ICD10CM | C31.0 | Malignant neoplasm of maxillary sinus |
| ICD10CM | C31.1 | Malignant neoplasm of ethmoidal sinus |
| ICD10CM | C31.2 | Malignant neoplasm of frontal sinus |
| ICD10CM | C31.3 | Malignant neoplasm of sphenoid sinus |
| ICD10CM | C31.8 | Malignant neoplasm of overlapping sites of accessory sinuses |
| ICD10CM | C31.9 | Malignant neoplasm of accessory sinus, unspecified |
| ICD10CM | C32.0 | Malignant neoplasm of glottis |
| ICD10CM | C32.1 | Malignant neoplasm of supraglottis |
| ICD10CM | C32.2 | Malignant neoplasm of subglottis |
| ICD10CM | C32.3 | Malignant neoplasm of laryngeal cartilage |
| ICD10CM | C32.8 | Malignant neoplasm of overlapping sites of larynx |
| ICD10CM | C32.9 | Malignant neoplasm of larynx, unspecified |
| ICD10CM | C33 | Malignant neoplasm of trachea |
| ICD10CM | C34.00 | Malignant neoplasm of unspecified main bronchus |

Continued on next page

**TableS24 – continued from previous page**

| CodeSystem | ConceptCode | ConceptName |
|---|---|---|
| ICD10CM | C34.01 | Malignant neoplasm of right main bronchus |
| ICD10CM | C34.02 | Malignant neoplasm of left main bronchus |
| ICD10CM | C34.10 | Malignant neoplasm of upper lobe, unspecified bronchus or lung |
| ICD10CM | C34.11 | Malignant neoplasm of upper lobe, right bronchus or lung |
| ICD10CM | C34.12 | Malignant neoplasm of upper lobe, left bronchus or lung |
| ICD10CM | C34.2 | Malignant neoplasm of middle lobe, bronchus or lung |
| ICD10CM | C34.30 | Malignant neoplasm of lower lobe, unspecified bronchus or lung |
| ICD10CM | C34.31 | Malignant neoplasm of lower lobe, right bronchus or lung |
| ICD10CM | C34.32 | Malignant neoplasm of lower lobe, left bronchus or lung |
| ICD10CM | C34.80 | Malignant neoplasm of overlapping sites of unspecified bronchus and lung |
| ICD10CM | C34.81 | Malignant neoplasm of overlapping sites of right bronchus and lung |
| ICD10CM | C34.82 | Malignant neoplasm of overlapping sites of left bronchus and lung |
| ICD10CM | C34.90 | Malignant neoplasm of unspecified part of unspecified bronchus or lung |
| ICD10CM | C34.91 | Malignant neoplasm of unspecified part of right bronchus or lung |
| ICD10CM | C34.92 | Malignant neoplasm of unspecified part of left bronchus or lung |
| ICD10CM | C37 | Malignant neoplasm of thymus |
| ICD10CM | C38.0 | Malignant neoplasm of heart |
| ICD10CM | C38.1 | Malignant neoplasm of anterior mediastinum |
| ICD10CM | C38.2 | Malignant neoplasm of posterior mediastinum |
| ICD10CM | C38.3 | Malignant neoplasm of mediastinum, part unspecified |
| ICD10CM | C38.4 | Malignant neoplasm of pleura |
| ICD10CM | C38.8 | Malignant neoplasm of overlapping sites of heart, mediastinum and pleura |
| ICD10CM | C39.0 | Malignant neoplasm of upper respiratory tract, part unspecified |
| ICD10CM | C39.9 | Malignant neoplasm of lower respiratory tract, part unspecified |
| ICD10CM | C40.00 | Malignant neoplasm of scapula and long bones of unspecified upper limb |
| ICD10CM | C40.01 | Malignant neoplasm of scapula and long bones of right upper limb |
| ICD10CM | C40.02 | Malignant neoplasm of scapula and long bones of left upper limb |
| ICD10CM | C40.10 | Malignant neoplasm of short bones of unspecified upper limb |
| ICD10CM | C40.11 | Malignant neoplasm of short bones of right upper limb |
| ICD10CM | C40.12 | Malignant neoplasm of short bones of left upper limb |
| ICD10CM | C40.20 | Malignant neoplasm of long bones of unspecified lower limb |
| ICD10CM | C40.21 | Malignant neoplasm of long bones of right lower limb |
| ICD10CM | C40.22 | Malignant neoplasm of long bones of left lower limb |
| ICD10CM | C40.30 | Malignant neoplasm of short bones of unspecified lower limb |
| ICD10CM | C40.31 | Malignant neoplasm of short bones of right lower limb |
| ICD10CM | C40.32 | Malignant neoplasm of short bones of left lower limb |
| ICD10CM | C40.80 | Malignant neoplasm of overlapping sites of bone and articular cartilage of unspecified limb |
| ICD10CM | C40.81 | Malignant neoplasm of overlapping sites of bone and articular cartilage of right limb |

Continued on next page

**TableS24 – continued from previous page**

| CodeSystem | ConceptCode | ConceptName |
|---|---|---|
| ICD10CM | C40.82 | Malignant neoplasm of overlapping sites of bone and articular cartilage of left limb |
| ICD10CM | C40.90 | Malignant neoplasm of unspecified bones and articular cartilage of unspecified limb |
| ICD10CM | C40.91 | Malignant neoplasm of unspecified bones and articular cartilage of right limb |
| ICD10CM | C40.92 | Malignant neoplasm of unspecified bones and articular cartilage of left limb |
| ICD10CM | C41.0 | Malignant neoplasm of bones of skull and face |
| ICD10CM | C41.1 | Malignant neoplasm of mandible |
| ICD10CM | C41.2 | Malignant neoplasm of vertebral column |
| ICD10CM | C41.3 | Malignant neoplasm of ribs, sternum and clavicle |
| ICD10CM | C41.4 | Malignant neoplasm of pelvic bones, sacrum and coccyx |
| ICD10CM | C41.9 | Malignant neoplasm of bone and articular cartilage, unspecified |
| ICD10CM | C43.0 | Malignant melanoma of lip |
| ICD10CM | C43.10 | Malignant melanoma of unspecified eyelid, including canthus |
| ICD10CM | C43.11 | Malignant melanoma of right eyelid, including canthus |
| ICD10CM | C43.12 | Malignant melanoma of left eyelid, including canthus |
| ICD10CM | C43.20 | Malignant melanoma of unspecified ear and external auricular canal |
| ICD10CM | C43.21 | Malignant melanoma of right ear and external auricular canal |
| ICD10CM | C43.22 | Malignant melanoma of left ear and external auricular canal |
| ICD10CM | C43.30 | Malignant melanoma of unspecified part of face |
| ICD10CM | C43.31 | Malignant melanoma of nose |
| ICD10CM | C43.39 | Malignant melanoma of other parts of face |
| ICD10CM | C43.4 | Malignant melanoma of scalp and neck |
| ICD10CM | C43.51 | Malignant melanoma of anal skin |
| ICD10CM | C43.52 | Malignant melanoma of skin of breast |
| ICD10CM | C43.59 | Malignant melanoma of other part of trunk |
| ICD10CM | C43.60 | Malignant melanoma of unspecified upper limb, including shoulder |
| ICD10CM | C43.61 | Malignant melanoma of right upper limb, including shoulder |
| ICD10CM | C43.62 | Malignant melanoma of left upper limb, including shoulder |
| ICD10CM | C43.70 | Malignant melanoma of unspecified lower limb, including hip |
| ICD10CM | C43.71 | Malignant melanoma of right lower limb, including hip |
| ICD10CM | C43.72 | Malignant melanoma of left lower limb, including hip |
| ICD10CM | C43.8 | Malignant melanoma of overlapping sites of skin |
| ICD10CM | C43.9 | Malignant melanoma of skin, unspecified |
| ICD10CM | C44.00 | Unspecified malignant neoplasm of skin of lip |
| ICD10CM | C44.01 | Basal cell carcinoma of skin of lip |
| ICD10CM | C44.02 | Squamous cell carcinoma of skin of lip |
| ICD10CM | C44.09 | Other specified malignant neoplasm of skin of lip |
| ICD10CM | C44.101 | Unspecified malignant neoplasm of skin of unspecified eyelid, including canthus |
| ICD10CM | C44.102 | Unspecified malignant neoplasm of skin of right eyelid, including canthus |

Continued on next page

**TableS24 – continued from previous page**

| CodeSystem | ConceptCode | ConceptName |
|---|---|---|
| ICD10CM | C44.109 | Unspecified malignant neoplasm of skin of left eyelid, including canthus |
| ICD10CM | C44.111 | Basal cell carcinoma of skin of unspecified eyelid, including canthus |
| ICD10CM | C44.112 | Basal cell carcinoma of skin of right eyelid, including canthus |
| ICD10CM | C44.119 | Basal cell carcinoma of skin of left eyelid, including canthus |
| ICD10CM | C44.121 | Squamous cell carcinoma of skin of unspecified eyelid, including canthus |
| ICD10CM | C44.122 | Squamous cell carcinoma of skin of right eyelid, including canthus |
| ICD10CM | C44.129 | Squamous cell carcinoma of skin of left eyelid, including canthus |
| ICD10CM | C44.191 | Other specified malignant neoplasm of skin of unspecified eyelid, including canthus |
| ICD10CM | C44.192 | Other specified malignant neoplasm of skin of right eyelid, including canthus |
| ICD10CM | C44.199 | Other specified malignant neoplasm of skin of left eyelid, including canthus |
| ICD10CM | C44.201 | Unspecified malignant neoplasm of skin of unspecified ear and external auricular canal |
| ICD10CM | C44.202 | Unspecified malignant neoplasm of skin of right ear and external auricular canal |
| ICD10CM | C44.209 | Unspecified malignant neoplasm of skin of left ear and external auricular canal |
| ICD10CM | C44.211 | Basal cell carcinoma of skin of unspecified ear and external auricular canal |
| ICD10CM | C44.212 | Basal cell carcinoma of skin of right ear and external auricular canal |
| ICD10CM | C44.219 | Basal cell carcinoma of skin of left ear and external auricular canal |
| ICD10CM | C44.221 | Squamous cell carcinoma of skin of unspecified ear and external auricular canal |
| ICD10CM | C44.222 | Squamous cell carcinoma of skin of right ear and external auricular canal |
| ICD10CM | C44.229 | Squamous cell carcinoma of skin of left ear and external auricular canal |
| ICD10CM | C44.291 | Other specified malignant neoplasm of skin of unspecified ear and external auricular canal |
| ICD10CM | C44.292 | Other specified malignant neoplasm of skin of right ear and external auricular canal |
| ICD10CM | C44.299 | Other specified malignant neoplasm of skin of left ear and external auricular canal |
| ICD10CM | C44.300 | Unspecified malignant neoplasm of skin of unspecified part of face |
| ICD10CM | C44.301 | Unspecified malignant neoplasm of skin of nose |
| ICD10CM | C44.309 | Unspecified malignant neoplasm of skin of other parts of face |
| ICD10CM | C44.310 | Basal cell carcinoma of skin of unspecified parts of face |
| ICD10CM | C44.311 | Basal cell carcinoma of skin of nose |

Continued on next page

**TableS24 – continued from previous page**

| CodeSystem | ConceptCode | ConceptName |
|---|---|---|
| ICD10CM | C44.319 | Basal cell carcinoma of skin of other parts of face |
| ICD10CM | C44.320 | Squamous cell carcinoma of skin of unspecified parts of face |
| ICD10CM | C44.321 | Squamous cell carcinoma of skin of nose |
| ICD10CM | C44.329 | Squamous cell carcinoma of skin of other parts of face |
| ICD10CM | C44.390 | Other specified malignant neoplasm of skin of unspecified parts of face |
| ICD10CM | C44.391 | Other specified malignant neoplasm of skin of nose |
| ICD10CM | C44.399 | Other specified malignant neoplasm of skin of other parts of face |
| ICD10CM | C44.40 | Unspecified malignant neoplasm of skin of scalp and neck |
| ICD10CM | C44.41 | Basal cell carcinoma of skin of scalp and neck |
| ICD10CM | C44.42 | Squamous cell carcinoma of skin of scalp and neck |
| ICD10CM | C44.49 | Other specified malignant neoplasm of skin of scalp and neck |
| ICD10CM | C44.500 | Unspecified malignant neoplasm of anal skin |
| ICD10CM | C44.501 | Unspecified malignant neoplasm of skin of breast |
| ICD10CM | C44.509 | Unspecified malignant neoplasm of skin of other part of trunk |
| ICD10CM | C44.510 | Basal cell carcinoma of anal skin |
| ICD10CM | C44.511 | Basal cell carcinoma of skin of breast |
| ICD10CM | C44.519 | Basal cell carcinoma of skin of other part of trunk |
| ICD10CM | C44.520 | Squamous cell carcinoma of anal skin |
| ICD10CM | C44.521 | Squamous cell carcinoma of skin of breast |
| ICD10CM | C44.529 | Squamous cell carcinoma of skin of other part of trunk |
| ICD10CM | C44.590 | Other specified malignant neoplasm of anal skin |
| ICD10CM | C44.591 | Other specified malignant neoplasm of skin of breast |
| ICD10CM | C44.599 | Other specified malignant neoplasm of skin of other part of trunk |
| ICD10CM | C44.601 | Unspecified malignant neoplasm of skin of unspecified upper limb, including shoulder |
| ICD10CM | C44.602 | Unspecified malignant neoplasm of skin of right upper limb, including shoulder |
| ICD10CM | C44.609 | Unspecified malignant neoplasm of skin of left upper limb, including shoulder |
| ICD10CM | C44.611 | Basal cell carcinoma of skin of unspecified upper limb, including shoulder |
| ICD10CM | C44.612 | Basal cell carcinoma of skin of right upper limb, including shoulder |
| ICD10CM | C44.619 | Basal cell carcinoma of skin of left upper limb, including shoulder |
| ICD10CM | C44.621 | Squamous cell carcinoma of skin of unspecified upper limb, including shoulder |
| ICD10CM | C44.622 | Squamous cell carcinoma of skin of right upper limb, including shoulder |
| ICD10CM | C44.629 | Squamous cell carcinoma of skin of left upper limb, including shoulder |
| ICD10CM | C44.691 | Other specified malignant neoplasm of skin of unspecified upper limb, including shoulder |
| ICD10CM | C44.692 | Other specified malignant neoplasm of skin of right upper limb, including shoulder |
| ICD10CM | C44.699 | Other specified malignant neoplasm of skin of left upper limb, including shoulder |

Continued on next page

**TableS24 – continued from previous page**

| CodeSystem | ConceptCode | ConceptName |
|---|---|---|
| ICD10CM | C44.701 | Unspecified malignant neoplasm of skin of unspecified lower limb, including hip |
| ICD10CM | C44.702 | Unspecified malignant neoplasm of skin of right lower limb, including hip |
| ICD10CM | C44.709 | Unspecified malignant neoplasm of skin of left lower limb, including hip |
| ICD10CM | C44.711 | Basal cell carcinoma of skin of unspecified lower limb, including hip |
| ICD10CM | C44.712 | Basal cell carcinoma of skin of right lower limb, including hip |
| ICD10CM | C44.719 | Basal cell carcinoma of skin of left lower limb, including hip |
| ICD10CM | C44.721 | Squamous cell carcinoma of skin of unspecified lower limb, including hip |
| ICD10CM | C44.722 | Squamous cell carcinoma of skin of right lower limb, including hip |
| ICD10CM | C44.729 | Squamous cell carcinoma of skin of left lower limb, including hip |
| ICD10CM | C44.791 | Other specified malignant neoplasm of skin of unspecified lower limb, including hip |
| ICD10CM | C44.792 | Other specified malignant neoplasm of skin of right lower limb, including hip |
| ICD10CM | C44.799 | Other specified malignant neoplasm of skin of left lower limb, including hip |
| ICD10CM | C44.80 | Unspecified malignant neoplasm of overlapping sites of skin |
| ICD10CM | C44.81 | Basal cell carcinoma of overlapping sites of skin |
| ICD10CM | C44.82 | Squamous cell carcinoma of overlapping sites of skin |
| ICD10CM | C44.89 | Other specified malignant neoplasm of overlapping sites of skin |
| ICD10CM | C44.90 | Unspecified malignant neoplasm of skin, unspecified |
| ICD10CM | C44.91 | Basal cell carcinoma of skin, unspecified |
| ICD10CM | C44.92 | Squamous cell carcinoma of skin, unspecified |
| ICD10CM | C44.99 | Other specified malignant neoplasm of skin, unspecified |
| ICD10CM | C45.0 | Mesothelioma of pleura |
| ICD10CM | C45.1 | Mesothelioma of peritoneum |
| ICD10CM | C45.2 | Mesothelioma of pericardium |
| ICD10CM | C45.7 | Mesothelioma of other sites |
| ICD10CM | C45.9 | Mesothelioma, unspecified |
| ICD10CM | C47.0 | Malignant neoplasm of peripheral nerves of head, face and neck |
| ICD10CM | C47.10 | Malignant neoplasm of peripheral nerves of unspecified upper limb, including shoulder |
| ICD10CM | C47.11 | Malignant neoplasm of peripheral nerves of right upper limb, including shoulder |
| ICD10CM | C47.12 | Malignant neoplasm of peripheral nerves of left upper limb, including shoulder |
| ICD10CM | C47.20 | Malignant neoplasm of peripheral nerves of unspecified lower limb, including hip |
| ICD10CM | C47.21 | Malignant neoplasm of peripheral nerves of right lower limb, including hip |
| ICD10CM | C47.22 | Malignant neoplasm of peripheral nerves of left lower limb, including hip |

Continued on next page

**TableS24 – continued from previous page**

| CodeSystem | ConceptCode | ConceptName |
|---|---|---|
| ICD10CM | C47.3 | Malignant neoplasm of peripheral nerves of thorax |
| ICD10CM | C47.4 | Malignant neoplasm of peripheral nerves of abdomen |
| ICD10CM | C47.5 | Malignant neoplasm of peripheral nerves of pelvis |
| ICD10CM | C47.6 | Malignant neoplasm of peripheral nerves of trunk, unspecified |
| ICD10CM | C47.8 | Malignant neoplasm of overlapping sites of peripheral nerves and autonomic nervous system |
| ICD10CM | C47.9 | Malignant neoplasm of peripheral nerves and autonomic nervous system, unspecified |
| ICD10CM | C48.0 | Malignant neoplasm of retroperitoneum |
| ICD10CM | C48.1 | Malignant neoplasm of specified parts of peritoneum |
| ICD10CM | C48.2 | Malignant neoplasm of peritoneum, unspecified |
| ICD10CM | C48.8 | Malignant neoplasm of overlapping sites of retroperitoneum and peritoneum |
| ICD10CM | C49.0 | Malignant neoplasm of connective and soft tissue of head, face and neck |
| ICD10CM | C49.10 | Malignant neoplasm of connective and soft tissue of unspecified upper limb, including shoulder |
| ICD10CM | C49.11 | Malignant neoplasm of connective and soft tissue of right upper limb, including shoulder |
| ICD10CM | C49.12 | Malignant neoplasm of connective and soft tissue of left upper limb, including shoulder |
| ICD10CM | C49.20 | Malignant neoplasm of connective and soft tissue of unspecified lower limb, including hip |
| ICD10CM | C49.21 | Malignant neoplasm of connective and soft tissue of right lower limb, including hip |
| ICD10CM | C49.22 | Malignant neoplasm of connective and soft tissue of left lower limb, including hip |
| ICD10CM | C49.3 | Malignant neoplasm of connective and soft tissue of thorax |
| ICD10CM | C49.4 | Malignant neoplasm of connective and soft tissue of abdomen |
| ICD10CM | C49.5 | Malignant neoplasm of connective and soft tissue of pelvis |
| ICD10CM | C49.6 | Malignant neoplasm of connective and soft tissue of trunk, unspecified |
| ICD10CM | C49.8 | Malignant neoplasm of overlapping sites of connective and soft tissue |
| ICD10CM | C49.9 | Malignant neoplasm of connective and soft tissue, unspecified |
| ICD10CM | C49.A0 | Gastrointestinal stromal tumor, unspecified site |
| ICD10CM | C49.A1 | Gastrointestinal stromal tumor of esophagus |
| ICD10CM | C49.A2 | Gastrointestinal stromal tumor of stomach |
| ICD10CM | C49.A3 | Gastrointestinal stromal tumor of small intestine |
| ICD10CM | C49.A4 | Gastrointestinal stromal tumor of large intestine |
| ICD10CM | C49.A5 | Gastrointestinal stromal tumor of rectum |
| ICD10CM | C49.A9 | Gastrointestinal stromal tumor of other sites |
| ICD10CM | C4A.0 | Merkel cell carcinoma of lip |
| ICD10CM | C4A.10 | Merkel cell carcinoma of unspecified eyelid, including canthus |
| ICD10CM | C4A.11 | Merkel cell carcinoma of right eyelid, including canthus |
| ICD10CM | C4A.12 | Merkel cell carcinoma of left eyelid, including canthus |

Continued on next page

**TableS24 – continued from previous page**

| CodeSystem | ConceptCode | ConceptName |
|---|---|---|
| ICD10CM | C4A.20 | Merkel cell carcinoma of unspecified ear and external auricular canal |
| ICD10CM | C4A.21 | Merkel cell carcinoma of right ear and external auricular canal |
| ICD10CM | C4A.22 | Merkel cell carcinoma of left ear and external auricular canal |
| ICD10CM | C4A.30 | Merkel cell carcinoma of unspecified part of face |
| ICD10CM | C4A.31 | Merkel cell carcinoma of nose |
| ICD10CM | C4A.39 | Merkel cell carcinoma of other parts of face |
| ICD10CM | C4A.4 | Merkel cell carcinoma of scalp and neck |
| ICD10CM | C4A.51 | Merkel cell carcinoma of anal skin |
| ICD10CM | C4A.52 | Merkel cell carcinoma of skin of breast |
| ICD10CM | C4A.59 | Merkel cell carcinoma of other part of trunk |
| ICD10CM | C4A.60 | Merkel cell carcinoma of unspecified upper limb, including shoulder |
| ICD10CM | C4A.61 | Merkel cell carcinoma of right upper limb, including shoulder |
| ICD10CM | C4A.62 | Merkel cell carcinoma of left upper limb, including shoulder |
| ICD10CM | C4A.70 | Merkel cell carcinoma of unspecified lower limb, including hip |
| ICD10CM | C4A.71 | Merkel cell carcinoma of right lower limb, including hip |
| ICD10CM | C4A.72 | Merkel cell carcinoma of left lower limb, including hip |
| ICD10CM | C4A.8 | Merkel cell carcinoma of overlapping sites |
| ICD10CM | C4A.9 | Merkel cell carcinoma, unspecified |
| ICD10CM | C50.011 | Malignant neoplasm of nipple and areola, right female breast |
| ICD10CM | C50.012 | Malignant neoplasm of nipple and areola, left female breast |
| ICD10CM | C50.019 | Malignant neoplasm of nipple and areola, unspecified female breast |
| ICD10CM | C50.021 | Malignant neoplasm of nipple and areola, right male breast |
| ICD10CM | C50.022 | Malignant neoplasm of nipple and areola, left male breast |
| ICD10CM | C50.029 | Malignant neoplasm of nipple and areola, unspecified male breast |
| ICD10CM | C50.111 | Malignant neoplasm of central portion of right female breast |
| ICD10CM | C50.112 | Malignant neoplasm of central portion of left female breast |
| ICD10CM | C50.119 | Malignant neoplasm of central portion of unspecified female breast |
| ICD10CM | C50.121 | Malignant neoplasm of central portion of right male breast |
| ICD10CM | C50.122 | Malignant neoplasm of central portion of left male breast |
| ICD10CM | C50.129 | Malignant neoplasm of central portion of unspecified male breast |
| ICD10CM | C50.211 | Malignant neoplasm of upper-inner quadrant of right female breast |
| ICD10CM | C50.212 | Malignant neoplasm of upper-inner quadrant of left female breast |
| ICD10CM | C50.219 | Malignant neoplasm of upper-inner quadrant of unspecified female breast |
| ICD10CM | C50.221 | Malignant neoplasm of upper-inner quadrant of right male breast |
| ICD10CM | C50.222 | Malignant neoplasm of upper-inner quadrant of left male breast |
| ICD10CM | C50.229 | Malignant neoplasm of upper-inner quadrant of unspecified male breast |
| ICD10CM | C50.311 | Malignant neoplasm of lower-inner quadrant of right female breast |
| ICD10CM | C50.312 | Malignant neoplasm of lower-inner quadrant of left female breast |

Continued on next page

**TableS24 – continued from previous page**

| CodeSystem | ConceptCode | ConceptName |
|---|---|---|
| ICD10CM | C50.319 | Malignant neoplasm of lower-inner quadrant of unspecified female breast |
| ICD10CM | C50.321 | Malignant neoplasm of lower-inner quadrant of right male breast |
| ICD10CM | C50.322 | Malignant neoplasm of lower-inner quadrant of left male breast |
| ICD10CM | C50.329 | Malignant neoplasm of lower-inner quadrant of unspecified male breast |
| ICD10CM | C50.411 | Malignant neoplasm of upper-outer quadrant of right female breast |
| ICD10CM | C50.412 | Malignant neoplasm of upper-outer quadrant of left female breast |
| ICD10CM | C50.419 | Malignant neoplasm of upper-outer quadrant of unspecified female breast |
| ICD10CM | C50.421 | Malignant neoplasm of upper-outer quadrant of right male breast |
| ICD10CM | C50.422 | Malignant neoplasm of upper-outer quadrant of left male breast |
| ICD10CM | C50.429 | Malignant neoplasm of upper-outer quadrant of unspecified male breast |
| ICD10CM | C50.511 | Malignant neoplasm of lower-outer quadrant of right female breast |
| ICD10CM | C50.512 | Malignant neoplasm of lower-outer quadrant of left female breast |
| ICD10CM | C50.519 | Malignant neoplasm of lower-outer quadrant of unspecified female breast |
| ICD10CM | C50.521 | Malignant neoplasm of lower-outer quadrant of right male breast |
| ICD10CM | C50.522 | Malignant neoplasm of lower-outer quadrant of left male breast |
| ICD10CM | C50.529 | Malignant neoplasm of lower-outer quadrant of unspecified male breast |
| ICD10CM | C50.611 | Malignant neoplasm of axillary tail of right female breast |
| ICD10CM | C50.612 | Malignant neoplasm of axillary tail of left female breast |
| ICD10CM | C50.619 | Malignant neoplasm of axillary tail of unspecified female breast |
| ICD10CM | C50.621 | Malignant neoplasm of axillary tail of right male breast |
| ICD10CM | C50.622 | Malignant neoplasm of axillary tail of left male breast |
| ICD10CM | C50.629 | Malignant neoplasm of axillary tail of unspecified male breast |
| ICD10CM | C50.811 | Malignant neoplasm of overlapping sites of right female breast |
| ICD10CM | C50.812 | Malignant neoplasm of overlapping sites of left female breast |
| ICD10CM | C50.819 | Malignant neoplasm of overlapping sites of unspecified female breast |
| ICD10CM | C50.821 | Malignant neoplasm of overlapping sites of right male breast |
| ICD10CM | C50.822 | Malignant neoplasm of overlapping sites of left male breast |
| ICD10CM | C50.829 | Malignant neoplasm of overlapping sites of unspecified male breast |
| ICD10CM | C50.911 | Malignant neoplasm of unspecified site of right female breast |
| ICD10CM | C50.912 | Malignant neoplasm of unspecified site of left female breast |
| ICD10CM | C50.919 | Malignant neoplasm of unspecified site of unspecified female breast |
| ICD10CM | C50.921 | Malignant neoplasm of unspecified site of right male breast |
| ICD10CM | C50.922 | Malignant neoplasm of unspecified site of left male breast |
| ICD10CM | C50.929 | Malignant neoplasm of unspecified site of unspecified male breast |
| ICD10CM | C51.0 | Malignant neoplasm of labium majus |
| ICD10CM | C51.1 | Malignant neoplasm of labium minus |

Continued on next page

**TableS24 – continued from previous page**

| CodeSystem | ConceptCode | ConceptName |
|---|---|---|
| ICD10CM | C51.2 | Malignant neoplasm of clitoris |
| ICD10CM | C51.8 | Malignant neoplasm of overlapping sites of vulva |
| ICD10CM | C51.9 | Malignant neoplasm of vulva, unspecified |
| ICD10CM | C52 | Malignant neoplasm of vagina |
| ICD10CM | C53.0 | Malignant neoplasm of endocervix |
| ICD10CM | C53.1 | Malignant neoplasm of exocervix |
| ICD10CM | C53.8 | Malignant neoplasm of overlapping sites of cervix uteri |
| ICD10CM | C53.9 | Malignant neoplasm of cervix uteri, unspecified |
| ICD10CM | C54.0 | Malignant neoplasm of isthmus uteri |
| ICD10CM | C54.1 | Malignant neoplasm of endometrium |
| ICD10CM | C54.2 | Malignant neoplasm of myometrium |
| ICD10CM | C54.3 | Malignant neoplasm of fundus uteri |
| ICD10CM | C54.8 | Malignant neoplasm of overlapping sites of corpus uteri |
| ICD10CM | C54.9 | Malignant neoplasm of corpus uteri, unspecified |
| ICD10CM | C55 | Malignant neoplasm of uterus, part unspecified |
| ICD10CM | C56.1 | Malignant neoplasm of right ovary |
| ICD10CM | C56.2 | Malignant neoplasm of left ovary |
| ICD10CM | C56.9 | Malignant neoplasm of unspecified ovary |
| ICD10CM | C57.00 | Malignant neoplasm of unspecified fallopian tube |
| ICD10CM | C57.01 | Malignant neoplasm of right fallopian tube |
| ICD10CM | C57.02 | Malignant neoplasm of left fallopian tube |
| ICD10CM | C57.10 | Malignant neoplasm of unspecified broad ligament |
| ICD10CM | C57.11 | Malignant neoplasm of right broad ligament |
| ICD10CM | C57.12 | Malignant neoplasm of left broad ligament |
| ICD10CM | C57.20 | Malignant neoplasm of unspecified round ligament |
| ICD10CM | C57.21 | Malignant neoplasm of right round ligament |
| ICD10CM | C57.22 | Malignant neoplasm of left round ligament |
| ICD10CM | C57.3 | Malignant neoplasm of parametrium |
| ICD10CM | C57.4 | Malignant neoplasm of uterine adnexa, unspecified |
| ICD10CM | C57.7 | Malignant neoplasm of other specified female genital organs |
| ICD10CM | C57.8 | Malignant neoplasm of overlapping sites of female genital organs |
| ICD10CM | C57.9 | Malignant neoplasm of female genital organ, unspecified |
| ICD10CM | C58 | Malignant neoplasm of placenta |
| ICD10CM | C60.0 | Malignant neoplasm of prepuce |
| ICD10CM | C60.1 | Malignant neoplasm of glans penis |
| ICD10CM | C60.2 | Malignant neoplasm of body of penis |
| ICD10CM | C60.8 | Malignant neoplasm of overlapping sites of penis |
| ICD10CM | C60.9 | Malignant neoplasm of penis, unspecified |
| ICD10CM | C61 | Malignant neoplasm of prostate |
| ICD10CM | C62.00 | Malignant neoplasm of unspecified undescended testis |
| ICD10CM | C62.01 | Malignant neoplasm of undescended right testis |
| ICD10CM | C62.02 | Malignant neoplasm of undescended left testis |
| ICD10CM | C62.10 | Malignant neoplasm of unspecified descended testis |
| ICD10CM | C62.11 | Malignant neoplasm of descended right testis |
| ICD10CM | C62.12 | Malignant neoplasm of descended left testis |
| ICD10CM | C62.90 | Malignant neoplasm of unspecified testis, unspecified whether descended or undescended |

Continued on next page

**TableS24 – continued from previous page**

| CodeSystem | ConceptCode | ConceptName |
|---|---|---|
| ICD10CM | C62.91 | Malignant neoplasm of right testis, unspecified whether descended or undescended |
| ICD10CM | C62.92 | Malignant neoplasm of left testis, unspecified whether descended or undescended |
| ICD10CM | C63.00 | Malignant neoplasm of unspecified epididymis |
| ICD10CM | C63.01 | Malignant neoplasm of right epididymis |
| ICD10CM | C63.02 | Malignant neoplasm of left epididymis |
| ICD10CM | C63.10 | Malignant neoplasm of unspecified spermatic cord |
| ICD10CM | C63.11 | Malignant neoplasm of right spermatic cord |
| ICD10CM | C63.12 | Malignant neoplasm of left spermatic cord |
| ICD10CM | C63.2 | Malignant neoplasm of scrotum |
| ICD10CM | C63.7 | Malignant neoplasm of other specified male genital organs |
| ICD10CM | C63.8 | Malignant neoplasm of overlapping sites of male genital organs |
| ICD10CM | C63.9 | Malignant neoplasm of male genital organ, unspecified |
| ICD10CM | C64.1 | Malignant neoplasm of right kidney, except renal pelvis |
| ICD10CM | C64.2 | Malignant neoplasm of left kidney, except renal pelvis |
| ICD10CM | C64.9 | Malignant neoplasm of unspecified kidney, except renal pelvis |
| ICD10CM | C65.1 | Malignant neoplasm of right renal pelvis |
| ICD10CM | C65.2 | Malignant neoplasm of left renal pelvis |
| ICD10CM | C65.9 | Malignant neoplasm of unspecified renal pelvis |
| ICD10CM | C66.1 | Malignant neoplasm of right ureter |
| ICD10CM | C66.2 | Malignant neoplasm of left ureter |
| ICD10CM | C66.9 | Malignant neoplasm of unspecified ureter |
| ICD10CM | C67.0 | Malignant neoplasm of trigone of bladder |
| ICD10CM | C67.1 | Malignant neoplasm of dome of bladder |
| ICD10CM | C67.2 | Malignant neoplasm of lateral wall of bladder |
| ICD10CM | C67.3 | Malignant neoplasm of anterior wall of bladder |
| ICD10CM | C67.4 | Malignant neoplasm of posterior wall of bladder |
| ICD10CM | C67.5 | Malignant neoplasm of bladder neck |
| ICD10CM | C67.6 | Malignant neoplasm of ureteric orifice |
| ICD10CM | C67.7 | Malignant neoplasm of urachus |
| ICD10CM | C67.8 | Malignant neoplasm of overlapping sites of bladder |
| ICD10CM | C67.9 | Malignant neoplasm of bladder, unspecified |
| ICD10CM | C68.0 | Malignant neoplasm of urethra |
| ICD10CM | C68.1 | Malignant neoplasm of paraurethral glands |
| ICD10CM | C68.8 | Malignant neoplasm of overlapping sites of urinary organs |
| ICD10CM | C68.9 | Malignant neoplasm of urinary organ, unspecified |
| ICD10CM | C69.00 | Malignant neoplasm of unspecified conjunctiva |
| ICD10CM | C69.01 | Malignant neoplasm of right conjunctiva |
| ICD10CM | C69.02 | Malignant neoplasm of left conjunctiva |
| ICD10CM | C69.10 | Malignant neoplasm of unspecified cornea |
| ICD10CM | C69.11 | Malignant neoplasm of right cornea |
| ICD10CM | C69.12 | Malignant neoplasm of left cornea |
| ICD10CM | C69.20 | Malignant neoplasm of unspecified retina |
| ICD10CM | C69.21 | Malignant neoplasm of right retina |
| ICD10CM | C69.22 | Malignant neoplasm of left retina |
| ICD10CM | C69.30 | Malignant neoplasm of unspecified choroid |

Continued on next page

**TableS24 – continued from previous page**

| CodeSystem | ConceptCode | ConceptName |
|---|---|---|
| ICD10CM | C69.31 | Malignant neoplasm of right choroid |
| ICD10CM | C69.32 | Malignant neoplasm of left choroid |
| ICD10CM | C69.40 | Malignant neoplasm of unspecified ciliary body |
| ICD10CM | C69.41 | Malignant neoplasm of right ciliary body |
| ICD10CM | C69.42 | Malignant neoplasm of left ciliary body |
| ICD10CM | C69.50 | Malignant neoplasm of unspecified lacrimal gland and duct |
| ICD10CM | C69.51 | Malignant neoplasm of right lacrimal gland and duct |
| ICD10CM | C69.52 | Malignant neoplasm of left lacrimal gland and duct |
| ICD10CM | C69.60 | Malignant neoplasm of unspecified orbit |
| ICD10CM | C69.61 | Malignant neoplasm of right orbit |
| ICD10CM | C69.62 | Malignant neoplasm of left orbit |
| ICD10CM | C69.80 | Malignant neoplasm of overlapping sites of unspecified eye and adnexa |
| ICD10CM | C69.81 | Malignant neoplasm of overlapping sites of right eye and adnexa |
| ICD10CM | C69.82 | Malignant neoplasm of overlapping sites of left eye and adnexa |
| ICD10CM | C69.90 | Malignant neoplasm of unspecified site of unspecified eye |
| ICD10CM | C69.91 | Malignant neoplasm of unspecified site of right eye |
| ICD10CM | C69.92 | Malignant neoplasm of unspecified site of left eye |
| ICD10CM | C70.0 | Malignant neoplasm of cerebral meninges |
| ICD10CM | C70.1 | Malignant neoplasm of spinal meninges |
| ICD10CM | C70.9 | Malignant neoplasm of meninges, unspecified |
| ICD10CM | C71.0 | Malignant neoplasm of cerebrum, except lobes and ventricles |
| ICD10CM | C71.1 | Malignant neoplasm of frontal lobe |
| ICD10CM | C71.2 | Malignant neoplasm of temporal lobe |
| ICD10CM | C71.3 | Malignant neoplasm of parietal lobe |
| ICD10CM | C71.4 | Malignant neoplasm of occipital lobe |
| ICD10CM | C71.5 | Malignant neoplasm of cerebral ventricle |
| ICD10CM | C71.6 | Malignant neoplasm of cerebellum |
| ICD10CM | C71.7 | Malignant neoplasm of brain stem |
| ICD10CM | C71.8 | Malignant neoplasm of overlapping sites of brain |
| ICD10CM | C71.9 | Malignant neoplasm of brain, unspecified |
| ICD10CM | C72.0 | Malignant neoplasm of spinal cord |
| ICD10CM | C72.1 | Malignant neoplasm of cauda equina |
| ICD10CM | C72.20 | Malignant neoplasm of unspecified olfactory nerve |
| ICD10CM | C72.21 | Malignant neoplasm of right olfactory nerve |
| ICD10CM | C72.22 | Malignant neoplasm of left olfactory nerve |
| ICD10CM | C72.30 | Malignant neoplasm of unspecified optic nerve |
| ICD10CM | C72.31 | Malignant neoplasm of right optic nerve |
| ICD10CM | C72.32 | Malignant neoplasm of left optic nerve |
| ICD10CM | C72.40 | Malignant neoplasm of unspecified acoustic nerve |
| ICD10CM | C72.41 | Malignant neoplasm of right acoustic nerve |
| ICD10CM | C72.42 | Malignant neoplasm of left acoustic nerve |
| ICD10CM | C72.50 | Malignant neoplasm of unspecified cranial nerve |
| ICD10CM | C72.59 | Malignant neoplasm of other cranial nerves |
| ICD10CM | C72.9 | Malignant neoplasm of central nervous system, unspecified |
| ICD10CM | C73 | Malignant neoplasm of thyroid gland |
| ICD10CM | C74.00 | Malignant neoplasm of cortex of unspecified adrenal gland |

Continued on next page

**TableS24 – continued from previous page**

| CodeSystem | ConceptCode | ConceptName |
|---|---|---|
| ICD10CM | C74.01 | Malignant neoplasm of cortex of right adrenal gland |
| ICD10CM | C74.02 | Malignant neoplasm of cortex of left adrenal gland |
| ICD10CM | C74.10 | Malignant neoplasm of medulla of unspecified adrenal gland |
| ICD10CM | C74.11 | Malignant neoplasm of medulla of right adrenal gland |
| ICD10CM | C74.12 | Malignant neoplasm of medulla of left adrenal gland |
| ICD10CM | C74.90 | Malignant neoplasm of unspecified part of unspecified adrenal gland |
| ICD10CM | C74.91 | Malignant neoplasm of unspecified part of right adrenal gland |
| ICD10CM | C74.92 | Malignant neoplasm of unspecified part of left adrenal gland |
| ICD10CM | C75.0 | Malignant neoplasm of parathyroid gland |
| ICD10CM | C75.1 | Malignant neoplasm of pituitary gland |
| ICD10CM | C75.2 | Malignant neoplasm of craniopharyngeal duct |
| ICD10CM | C75.3 | Malignant neoplasm of pineal gland |
| ICD10CM | C75.4 | Malignant neoplasm of carotid body |
| ICD10CM | C75.5 | Malignant neoplasm of aortic body and other paraganglia |
| ICD10CM | C75.8 | Malignant neoplasm with pluriglandular involvement, unspecified |
| ICD10CM | C75.9 | Malignant neoplasm of endocrine gland, unspecified |
| ICD10CM | C76.0 | Malignant neoplasm of head, face and neck |
| ICD10CM | C76.1 | Malignant neoplasm of thorax |
| ICD10CM | C76.2 | Malignant neoplasm of abdomen |
| ICD10CM | C76.3 | Malignant neoplasm of pelvis |
| ICD10CM | C76.40 | Malignant neoplasm of unspecified upper limb |
| ICD10CM | C76.41 | Malignant neoplasm of right upper limb |
| ICD10CM | C76.42 | Malignant neoplasm of left upper limb |
| ICD10CM | C76.50 | Malignant neoplasm of unspecified lower limb |
| ICD10CM | C76.51 | Malignant neoplasm of right lower limb |
| ICD10CM | C76.52 | Malignant neoplasm of left lower limb |
| ICD10CM | C76.8 | Malignant neoplasm of other specified ill-defined sites |
| ICD10CM | C77.0 | Secondary and unspecified malignant neoplasm of lymph nodes of head, face and neck |
| ICD10CM | C77.1 | Secondary and unspecified malignant neoplasm of intrathoracic lymph nodes |
| ICD10CM | C77.2 | Secondary and unspecified malignant neoplasm of intra-abdominal lymph nodes |
| ICD10CM | C77.3 | Secondary and unspecified malignant neoplasm of axilla and upper limb lymph nodes |
| ICD10CM | C77.4 | Secondary and unspecified malignant neoplasm of inguinal and lower limb lymph nodes |
| ICD10CM | C77.5 | Secondary and unspecified malignant neoplasm of intrapelvic lymph nodes |
| ICD10CM | C77.8 | Secondary and unspecified malignant neoplasm of lymph nodes of multiple regions |
| ICD10CM | C77.9 | Secondary and unspecified malignant neoplasm of lymph node, unspecified |
| ICD10CM | C78.00 | Secondary malignant neoplasm of unspecified lung |
| ICD10CM | C78.01 | Secondary malignant neoplasm of right lung |

Continued on next page

**TableS24 – continued from previous page**

| CodeSystem | ConceptCode | ConceptName |
|---|---|---|
| ICD10CM | C78.02 | Secondary malignant neoplasm of left lung |
| ICD10CM | C78.1 | Secondary malignant neoplasm of mediastinum |
| ICD10CM | C78.2 | Secondary malignant neoplasm of pleura |
| ICD10CM | C78.30 | Secondary malignant neoplasm of unspecified respiratory organ |
| ICD10CM | C78.39 | Secondary malignant neoplasm of other respiratory organs |
| ICD10CM | C78.4 | Secondary malignant neoplasm of small intestine |
| ICD10CM | C78.5 | Secondary malignant neoplasm of large intestine and rectum |
| ICD10CM | C78.6 | Secondary malignant neoplasm of retroperitoneum and peritoneum |
| ICD10CM | C78.7 | Secondary malignant neoplasm of liver and intrahepatic bile duct |
| ICD10CM | C78.80 | Secondary malignant neoplasm of unspecified digestive organ |
| ICD10CM | C78.89 | Secondary malignant neoplasm of other digestive organs |
| ICD10CM | C79.00 | Secondary malignant neoplasm of unspecified kidney and renal pelvis |
| ICD10CM | C79.01 | Secondary malignant neoplasm of right kidney and renal pelvis |
| ICD10CM | C79.02 | Secondary malignant neoplasm of left kidney and renal pelvis |
| ICD10CM | C79.10 | Secondary malignant neoplasm of unspecified urinary organs |
| ICD10CM | C79.11 | Secondary malignant neoplasm of bladder |
| ICD10CM | C79.19 | Secondary malignant neoplasm of other urinary organs |
| ICD10CM | C79.2 | Secondary malignant neoplasm of skin |
| ICD10CM | C79.31 | Secondary malignant neoplasm of brain |
| ICD10CM | C79.32 | Secondary malignant neoplasm of cerebral meninges |
| ICD10CM | C79.40 | Secondary malignant neoplasm of unspecified part of nervous system |
| ICD10CM | C79.49 | Secondary malignant neoplasm of other parts of nervous system |
| ICD10CM | C79.51 | Secondary malignant neoplasm of bone |
| ICD10CM | C79.52 | Secondary malignant neoplasm of bone marrow |
| ICD10CM | C79.60 | Secondary malignant neoplasm of unspecified ovary |
| ICD10CM | C79.61 | Secondary malignant neoplasm of right ovary |
| ICD10CM | C79.62 | Secondary malignant neoplasm of left ovary |
| ICD10CM | C79.70 | Secondary malignant neoplasm of unspecified adrenal gland |
| ICD10CM | C79.71 | Secondary malignant neoplasm of right adrenal gland |
| ICD10CM | C79.72 | Secondary malignant neoplasm of left adrenal gland |
| ICD10CM | C79.81 | Secondary malignant neoplasm of breast |
| ICD10CM | C79.82 | Secondary malignant neoplasm of genital organs |
| ICD10CM | C79.89 | Secondary malignant neoplasm of other specified sites |
| ICD10CM | C79.9 | Secondary malignant neoplasm of unspecified site |
| ICD10CM | C7A.00 | Malignant carcinoid tumor of unspecified site |
| ICD10CM | C7A.010 | Malignant carcinoid tumor of the duodenum |
| ICD10CM | C7A.011 | Malignant carcinoid tumor of the jejunum |
| ICD10CM | C7A.012 | Malignant carcinoid tumor of the ileum |
| ICD10CM | C7A.019 | Malignant carcinoid tumor of the small intestine, unspecified portion |
| ICD10CM | C7A.020 | Malignant carcinoid tumor of the appendix |
| ICD10CM | C7A.029 | Malignant carcinoid tumor of the large intestine, unspecified portion |
| ICD10CM | C7A.090 | Malignant carcinoid tumor of the bronchus and lung |

Continued on next page

**TableS24 – continued from previous page**

| CodeSystem | ConceptCode | ConceptName |
|---|---|---|
| ICD10CM | C7A.091 | Malignant carcinoid tumor of the thymus |
| ICD10CM | C7A.092 | Malignant carcinoid tumor of the stomach |
| ICD10CM | C7A.093 | Malignant carcinoid tumor of the kidney |
| ICD10CM | C7A.094 | Malignant carcinoid tumor of the foregut, unspecified |
| ICD10CM | C7A.095 | Malignant carcinoid tumor of the midgut, unspecified |
| ICD10CM | C7A.096 | Malignant carcinoid tumor of the hindgut, unspecified |
| ICD10CM | C7A.098 | Malignant carcinoid tumors of other sites |
| ICD10CM | C7A.1 | Malignant poorly differentiated neuroendocrine tumors |
| ICD10CM | C7A.8 | Other malignant neuroendocrine tumors |
| ICD10CM | C7B.00 | Secondary carcinoid tumors, unspecified site |
| ICD10CM | C7B.03 | Secondary carcinoid tumors of bone |
| ICD10CM | C7B.04 | Secondary carcinoid tumors of peritoneum |
| ICD10CM | C7B.09 | Secondary carcinoid tumors of other sites |
| ICD10CM | C7B.1 | Secondary Merkel cell carcinoma |
| ICD10CM | C7B.8 | Other secondary neuroendocrine tumors |
| ICD10CM | C80.0 | Disseminated malignant neoplasm, unspecified |
| ICD10CM | C80.1 | Malignant (primary) neoplasm, unspecified |
| ICD10CM | C80.2 | Malignant neoplasm associated with transplanted organ |
| ICD10CM | C81.00 | Nodular lymphocyte predominant Hodgkin lymphoma, unspecified site |
| ICD10CM | C81.01 | Nodular lymphocyte predominant Hodgkin lymphoma, lymph nodes of head, face, and neck |
| ICD10CM | C81.02 | Nodular lymphocyte predominant Hodgkin lymphoma, intrathoracic lymph nodes |
| ICD10CM | C81.03 | Nodular lymphocyte predominant Hodgkin lymphoma, intra-abdominal lymph nodes |
| ICD10CM | C81.04 | Nodular lymphocyte predominant Hodgkin lymphoma, lymph nodes of axilla and upper limb |
| ICD10CM | C81.05 | Nodular lymphocyte predominant Hodgkin lymphoma, lymph nodes of inguinal region and lower limb |
| ICD10CM | C81.06 | Nodular lymphocyte predominant Hodgkin lymphoma, intrapelvic lymph nodes |
| ICD10CM | C81.07 | Nodular lymphocyte predominant Hodgkin lymphoma, spleen |
| ICD10CM | C81.08 | Nodular lymphocyte predominant Hodgkin lymphoma, lymph nodes of multiple sites |
| ICD10CM | C81.09 | Nodular lymphocyte predominant Hodgkin lymphoma, extranodal and solid organ sites |
| ICD10CM | C81.10 | Nodular sclerosis Hodgkin lymphoma, unspecified site |
| ICD10CM | C81.11 | Nodular sclerosis Hodgkin lymphoma, lymph nodes of head, face, and neck |
| ICD10CM | C81.12 | Nodular sclerosis Hodgkin lymphoma, intrathoracic lymph nodes |
| ICD10CM | C81.13 | Nodular sclerosis Hodgkin lymphoma, intra-abdominal lymph nodes |
| ICD10CM | C81.14 | Nodular sclerosis Hodgkin lymphoma, lymph nodes of axilla and upper limb |
| ICD10CM | C81.15 | Nodular sclerosis Hodgkin lymphoma, lymph nodes of inguinal region and lower limb |

Continued on next page

**TableS24 – continued from previous page**

| CodeSystem | ConceptCode | ConceptName |
|---|---|---|
| ICD10CM | C81.16 | Nodular sclerosis Hodgkin lymphoma, intrapelvic lymph nodes |
| ICD10CM | C81.17 | Nodular sclerosis Hodgkin lymphoma, spleen |
| ICD10CM | C81.18 | Nodular sclerosis Hodgkin lymphoma, lymph nodes of multiple sites |
| ICD10CM | C81.19 | Nodular sclerosis Hodgkin lymphoma, extranodal and solid organ sites |
| ICD10CM | C81.20 | Mixed cellularity Hodgkin lymphoma, unspecified site |
| ICD10CM | C81.21 | Mixed cellularity Hodgkin lymphoma, lymph nodes of head, face, and neck |
| ICD10CM | C81.22 | Mixed cellularity Hodgkin lymphoma, intrathoracic lymph nodes |
| ICD10CM | C81.23 | Mixed cellularity Hodgkin lymphoma, intra-abdominal lymph nodes |
| ICD10CM | C81.24 | Mixed cellularity Hodgkin lymphoma, lymph nodes of axilla and upper limb |
| ICD10CM | C81.25 | Mixed cellularity Hodgkin lymphoma, lymph nodes of inguinal region and lower limb |
| ICD10CM | C81.26 | Mixed cellularity Hodgkin lymphoma, intrapelvic lymph nodes |
| ICD10CM | C81.27 | Mixed cellularity Hodgkin lymphoma, spleen |
| ICD10CM | C81.28 | Mixed cellularity Hodgkin lymphoma, lymph nodes of multiple sites |
| ICD10CM | C81.29 | Mixed cellularity Hodgkin lymphoma, extranodal and solid organ sites |
| ICD10CM | C81.30 | Lymphocyte depleted Hodgkin lymphoma, unspecified site |
| ICD10CM | C81.31 | Lymphocyte depleted Hodgkin lymphoma, lymph nodes of head, face, and neck |
| ICD10CM | C81.32 | Lymphocyte depleted Hodgkin lymphoma, intrathoracic lymph nodes |
| ICD10CM | C81.33 | Lymphocyte depleted Hodgkin lymphoma, intra-abdominal lymph nodes |
| ICD10CM | C81.34 | Lymphocyte depleted Hodgkin lymphoma, lymph nodes of axilla and upper limb |
| ICD10CM | C81.35 | Lymphocyte depleted Hodgkin lymphoma, lymph nodes of inguinal region and lower limb |
| ICD10CM | C81.36 | Lymphocyte depleted Hodgkin lymphoma, intrapelvic lymph nodes |
| ICD10CM | C81.37 | Lymphocyte depleted Hodgkin lymphoma, spleen |
| ICD10CM | C81.38 | Lymphocyte depleted Hodgkin lymphoma, lymph nodes of multiple sites |
| ICD10CM | C81.39 | Lymphocyte depleted Hodgkin lymphoma, extranodal and solid organ sites |
| ICD10CM | C81.40 | Lymphocyte-rich Hodgkin lymphoma, unspecified site |
| ICD10CM | C81.41 | Lymphocyte-rich Hodgkin lymphoma, lymph nodes of head, face, and neck |
| ICD10CM | C81.42 | Lymphocyte-rich Hodgkin lymphoma, intrathoracic lymph nodes |
| ICD10CM | C81.43 | Lymphocyte-rich Hodgkin lymphoma, intra-abdominal lymph nodes |

Continued on next page

**TableS24 – continued from previous page**

| CodeSystem | ConceptCode | ConceptName |
|---|---|---|
| ICD10CM | C81.44 | Lymphocyte-rich Hodgkin lymphoma, lymph nodes of axilla and upper limb |
| ICD10CM | C81.45 | Lymphocyte-rich Hodgkin lymphoma, lymph nodes of inguinal region and lower limb |
| ICD10CM | C81.46 | Lymphocyte-rich Hodgkin lymphoma, intrapelvic lymph nodes |
| ICD10CM | C81.47 | Lymphocyte-rich Hodgkin lymphoma, spleen |
| ICD10CM | C81.48 | Lymphocyte-rich Hodgkin lymphoma, lymph nodes of multiple sites |
| ICD10CM | C81.49 | Lymphocyte-rich Hodgkin lymphoma, extranodal and solid organ sites |
| ICD10CM | C81.70 | Other Hodgkin lymphoma, unspecified site |
| ICD10CM | C81.71 | Other Hodgkin lymphoma, lymph nodes of head, face, and neck |
| ICD10CM | C81.72 | Other Hodgkin lymphoma, intrathoracic lymph nodes |
| ICD10CM | C81.73 | Other Hodgkin lymphoma, intra-abdominal lymph nodes |
| ICD10CM | C81.74 | Other Hodgkin lymphoma, lymph nodes of axilla and upper limb |
| ICD10CM | C81.75 | Other Hodgkin lymphoma, lymph nodes of inguinal region and lower limb |
| ICD10CM | C81.76 | Other Hodgkin lymphoma, intrapelvic lymph nodes |
| ICD10CM | C81.77 | Other Hodgkin lymphoma, spleen |
| ICD10CM | C81.78 | Other Hodgkin lymphoma, lymph nodes of multiple sites |
| ICD10CM | C81.79 | Other Hodgkin lymphoma, extranodal and solid organ sites |
| ICD10CM | C81.90 | Hodgkin lymphoma, unspecified, unspecified site |
| ICD10CM | C81.91 | Hodgkin lymphoma, unspecified, lymph nodes of head, face, and neck |
| ICD10CM | C81.92 | Hodgkin lymphoma, unspecified, intrathoracic lymph nodes |
| ICD10CM | C81.93 | Hodgkin lymphoma, unspecified, intra-abdominal lymph nodes |
| ICD10CM | C81.94 | Hodgkin lymphoma, unspecified, lymph nodes of axilla and upper limb |
| ICD10CM | C81.95 | Hodgkin lymphoma, unspecified, lymph nodes of inguinal region and lower limb |
| ICD10CM | C81.96 | Hodgkin lymphoma, unspecified, intrapelvic lymph nodes |
| ICD10CM | C81.97 | Hodgkin lymphoma, unspecified, spleen |
| ICD10CM | C81.98 | Hodgkin lymphoma, unspecified, lymph nodes of multiple sites |
| ICD10CM | C81.99 | Hodgkin lymphoma, unspecified, extranodal and solid organ sites |
| ICD10CM | C82.00 | Follicular lymphoma grade I, unspecified site |
| ICD10CM | C82.01 | Follicular lymphoma grade I, lymph nodes of head, face, and neck |
| ICD10CM | C82.02 | Follicular lymphoma grade I, intrathoracic lymph nodes |
| ICD10CM | C82.03 | Follicular lymphoma grade I, intra-abdominal lymph nodes |
| ICD10CM | C82.04 | Follicular lymphoma grade I, lymph nodes of axilla and upper limb |
| ICD10CM | C82.05 | Follicular lymphoma grade I, lymph nodes of inguinal region and lower limb |
| ICD10CM | C82.06 | Follicular lymphoma grade I, intrapelvic lymph nodes |
| ICD10CM | C82.07 | Follicular lymphoma grade I, spleen |
| ICD10CM | C82.08 | Follicular lymphoma grade I, lymph nodes of multiple sites |
| ICD10CM | C82.09 | Follicular lymphoma grade I, extranodal and solid organ sites |

Continued on next page

**TableS24 – continued from previous page**

| CodeSystem | ConceptCode | ConceptName |
|---|---|---|
| ICD10CM | C82.10 | Follicular lymphoma grade II, unspecified site |
| ICD10CM | C82.11 | Follicular lymphoma grade II, lymph nodes of head, face, and neck |
| ICD10CM | C82.12 | Follicular lymphoma grade II, intrathoracic lymph nodes |
| ICD10CM | C82.13 | Follicular lymphoma grade II, intra-abdominal lymph nodes |
| ICD10CM | C82.14 | Follicular lymphoma grade II, lymph nodes of axilla and upper limb |
| ICD10CM | C82.15 | Follicular lymphoma grade II, lymph nodes of inguinal region and lower limb |
| ICD10CM | C82.16 | Follicular lymphoma grade II, intrapelvic lymph nodes |
| ICD10CM | C82.17 | Follicular lymphoma grade II, spleen |
| ICD10CM | C82.18 | Follicular lymphoma grade II, lymph nodes of multiple sites |
| ICD10CM | C82.19 | Follicular lymphoma grade II, extranodal and solid organ sites |
| ICD10CM | C82.20 | Follicular lymphoma grade III, unspecified, unspecified site |
| ICD10CM | C82.21 | Follicular lymphoma grade III, unspecified, lymph nodes of head, face, and neck |
| ICD10CM | C82.22 | Follicular lymphoma grade III, unspecified, intrathoracic lymph nodes |
| ICD10CM | C82.23 | Follicular lymphoma grade III, unspecified, intra-abdominal lymph nodes |
| ICD10CM | C82.24 | Follicular lymphoma grade III, unspecified, lymph nodes of axilla and upper limb |
| ICD10CM | C82.25 | Follicular lymphoma grade III, unspecified, lymph nodes of inguinal region and lower limb |
| ICD10CM | C82.26 | Follicular lymphoma grade III, unspecified, intrapelvic lymph nodes |
| ICD10CM | C82.27 | Follicular lymphoma grade III, unspecified, spleen |
| ICD10CM | C82.28 | Follicular lymphoma grade III, unspecified, lymph nodes of multiple sites |
| ICD10CM | C82.29 | Follicular lymphoma grade III, unspecified, extranodal and solid organ sites |
| ICD10CM | C82.30 | Follicular lymphoma grade IIIa, unspecified site |
| ICD10CM | C82.31 | Follicular lymphoma grade IIIa, lymph nodes of head, face, and neck |
| ICD10CM | C82.32 | Follicular lymphoma grade IIIa, intrathoracic lymph nodes |
| ICD10CM | C82.33 | Follicular lymphoma grade IIIa, intra-abdominal lymph nodes |
| ICD10CM | C82.34 | Follicular lymphoma grade IIIa, lymph nodes of axilla and upper limb |
| ICD10CM | C82.35 | Follicular lymphoma grade IIIa, lymph nodes of inguinal region and lower limb |
| ICD10CM | C82.36 | Follicular lymphoma grade IIIa, intrapelvic lymph nodes |
| ICD10CM | C82.37 | Follicular lymphoma grade IIIa, spleen |
| ICD10CM | C82.38 | Follicular lymphoma grade IIIa, lymph nodes of multiple sites |
| ICD10CM | C82.39 | Follicular lymphoma grade IIIa, extranodal and solid organ sites |
| ICD10CM | C82.40 | Follicular lymphoma grade IIIb, unspecified site |
| ICD10CM | C82.41 | Follicular lymphoma grade IIIb, lymph nodes of head, face, and neck |

Continued on next page

**TableS24 – continued from previous page**

| CodeSystem | ConceptCode | ConceptName |
|---|---|---|
| ICD10CM | C82.42 | Follicular lymphoma grade IIIb, intrathoracic lymph nodes |
| ICD10CM | C82.43 | Follicular lymphoma grade IIIb, intra-abdominal lymph nodes |
| ICD10CM | C82.44 | Follicular lymphoma grade IIIb, lymph nodes of axilla and upper limb |
| ICD10CM | C82.45 | Follicular lymphoma grade IIIb, lymph nodes of inguinal region and lower limb |
| ICD10CM | C82.46 | Follicular lymphoma grade IIIb, intrapelvic lymph nodes |
| ICD10CM | C82.47 | Follicular lymphoma grade IIIb, spleen |
| ICD10CM | C82.48 | Follicular lymphoma grade IIIb, lymph nodes of multiple sites |
| ICD10CM | C82.49 | Follicular lymphoma grade IIIb, extranodal and solid organ sites |
| ICD10CM | C82.50 | Diffuse follicle center lymphoma, unspecified site |
| ICD10CM | C82.51 | Diffuse follicle center lymphoma, lymph nodes of head, face, and neck |
| ICD10CM | C82.52 | Diffuse follicle center lymphoma, intrathoracic lymph nodes |
| ICD10CM | C82.53 | Diffuse follicle center lymphoma, intra-abdominal lymph nodes |
| ICD10CM | C82.54 | Diffuse follicle center lymphoma, lymph nodes of axilla and upper limb |
| ICD10CM | C82.55 | Diffuse follicle center lymphoma, lymph nodes of inguinal region and lower limb |
| ICD10CM | C82.56 | Diffuse follicle center lymphoma, intrapelvic lymph nodes |
| ICD10CM | C82.57 | Diffuse follicle center lymphoma, spleen |
| ICD10CM | C82.58 | Diffuse follicle center lymphoma, lymph nodes of multiple sites |
| ICD10CM | C82.59 | Diffuse follicle center lymphoma, extranodal and solid organ sites |
| ICD10CM | C82.60 | Cutaneous follicle center lymphoma, unspecified site |
| ICD10CM | C82.61 | Cutaneous follicle center lymphoma, lymph nodes of head, face, and neck |
| ICD10CM | C82.62 | Cutaneous follicle center lymphoma, intrathoracic lymph nodes |
| ICD10CM | C82.63 | Cutaneous follicle center lymphoma, intra-abdominal lymph nodes |
| ICD10CM | C82.64 | Cutaneous follicle center lymphoma, lymph nodes of axilla and upper limb |
| ICD10CM | C82.65 | Cutaneous follicle center lymphoma, lymph nodes of inguinal region and lower limb |
| ICD10CM | C82.66 | Cutaneous follicle center lymphoma, intrapelvic lymph nodes |
| ICD10CM | C82.67 | Cutaneous follicle center lymphoma, spleen |
| ICD10CM | C82.68 | Cutaneous follicle center lymphoma, lymph nodes of multiple sites |
| ICD10CM | C82.69 | Cutaneous follicle center lymphoma, extranodal and solid organ sites |
| ICD10CM | C82.80 | Other types of follicular lymphoma, unspecified site |
| ICD10CM | C82.81 | Other types of follicular lymphoma, lymph nodes of head, face, and neck |
| ICD10CM | C82.82 | Other types of follicular lymphoma, intrathoracic lymph nodes |
| ICD10CM | C82.83 | Other types of follicular lymphoma, intra-abdominal lymph nodes |
| ICD10CM | C82.84 | Other types of follicular lymphoma, lymph nodes of axilla and upper limb |

Continued on next page

**TableS24 – continued from previous page**

| CodeSystem | ConceptCode | ConceptName |
|---|---|---|
| ICD10CM | C82.85 | Other types of follicular lymphoma, lymph nodes of inguinal region and lower limb |
| ICD10CM | C82.86 | Other types of follicular lymphoma, intrapelvic lymph nodes |
| ICD10CM | C82.87 | Other types of follicular lymphoma, spleen |
| ICD10CM | C82.88 | Other types of follicular lymphoma, lymph nodes of multiple sites |
| ICD10CM | C82.89 | Other types of follicular lymphoma, extranodal and solid organ sites |
| ICD10CM | C82.90 | Follicular lymphoma, unspecified, unspecified site |
| ICD10CM | C82.91 | Follicular lymphoma, unspecified, lymph nodes of head, face, and neck |
| ICD10CM | C82.92 | Follicular lymphoma, unspecified, intrathoracic lymph nodes |
| ICD10CM | C82.93 | Follicular lymphoma, unspecified, intra-abdominal lymph nodes |
| ICD10CM | C82.94 | Follicular lymphoma, unspecified, lymph nodes of axilla and upper limb |
| ICD10CM | C82.95 | Follicular lymphoma, unspecified, lymph nodes of inguinal region and lower limb |
| ICD10CM | C82.96 | Follicular lymphoma, unspecified, intrapelvic lymph nodes |
| ICD10CM | C82.97 | Follicular lymphoma, unspecified, spleen |
| ICD10CM | C82.98 | Follicular lymphoma, unspecified, lymph nodes of multiple sites |
| ICD10CM | C82.99 | Follicular lymphoma, unspecified, extranodal and solid organ sites |
| ICD10CM | C83.00 | Small cell B-cell lymphoma, unspecified site |
| ICD10CM | C83.01 | Small cell B-cell lymphoma, lymph nodes of head, face, and neck |
| ICD10CM | C83.02 | Small cell B-cell lymphoma, intrathoracic lymph nodes |
| ICD10CM | C83.03 | Small cell B-cell lymphoma, intra-abdominal lymph nodes |
| ICD10CM | C83.04 | Small cell B-cell lymphoma, lymph nodes of axilla and upper limb |
| ICD10CM | C83.05 | Small cell B-cell lymphoma, lymph nodes of inguinal region and lower limb |
| ICD10CM | C83.06 | Small cell B-cell lymphoma, intrapelvic lymph nodes |
| ICD10CM | C83.07 | Small cell B-cell lymphoma, spleen |
| ICD10CM | C83.08 | Small cell B-cell lymphoma, lymph nodes of multiple sites |
| ICD10CM | C83.09 | Small cell B-cell lymphoma, extranodal and solid organ sites |
| ICD10CM | C83.10 | Mantle cell lymphoma, unspecified site |
| ICD10CM | C83.11 | Mantle cell lymphoma, lymph nodes of head, face, and neck |
| ICD10CM | C83.12 | Mantle cell lymphoma, intrathoracic lymph nodes |
| ICD10CM | C83.13 | Mantle cell lymphoma, intra-abdominal lymph nodes |
| ICD10CM | C83.14 | Mantle cell lymphoma, lymph nodes of axilla and upper limb |
| ICD10CM | C83.15 | Mantle cell lymphoma, lymph nodes of inguinal region and lower limb |
| ICD10CM | C83.16 | Mantle cell lymphoma, intrapelvic lymph nodes |
| ICD10CM | C83.17 | Mantle cell lymphoma, spleen |
| ICD10CM | C83.18 | Mantle cell lymphoma, lymph nodes of multiple sites |
| ICD10CM | C83.19 | Mantle cell lymphoma, extranodal and solid organ sites |
| ICD10CM | C83.30 | Diffuse large B-cell lymphoma, unspecified site |

Continued on next page

**TableS24 – continued from previous page**

| CodeSystem | ConceptCode | ConceptName |
|---|---|---|
| ICD10CM | C83.31 | Diffuse large B-cell lymphoma, lymph nodes of head, face, and neck |
| ICD10CM | C83.32 | Diffuse large B-cell lymphoma, intrathoracic lymph nodes |
| ICD10CM | C83.33 | Diffuse large B-cell lymphoma, intra-abdominal lymph nodes |
| ICD10CM | C83.34 | Diffuse large B-cell lymphoma, lymph nodes of axilla and upper limb |
| ICD10CM | C83.35 | Diffuse large B-cell lymphoma, lymph nodes of inguinal region and lower limb |
| ICD10CM | C83.36 | Diffuse large B-cell lymphoma, intrapelvic lymph nodes |
| ICD10CM | C83.37 | Diffuse large B-cell lymphoma, spleen |
| ICD10CM | C83.38 | Diffuse large B-cell lymphoma, lymph nodes of multiple sites |
| ICD10CM | C83.39 | Diffuse large B-cell lymphoma, extranodal and solid organ sites |
| ICD10CM | C83.50 | Lymphoblastic (diffuse) lymphoma, unspecified site |
| ICD10CM | C83.51 | Lymphoblastic (diffuse) lymphoma, lymph nodes of head, face, and neck |
| ICD10CM | C83.52 | Lymphoblastic (diffuse) lymphoma, intrathoracic lymph nodes |
| ICD10CM | C83.53 | Lymphoblastic (diffuse) lymphoma, intra-abdominal lymph nodes |
| ICD10CM | C83.54 | Lymphoblastic (diffuse) lymphoma, lymph nodes of axilla and upper limb |
| ICD10CM | C83.55 | Lymphoblastic (diffuse) lymphoma, lymph nodes of inguinal region and lower limb |
| ICD10CM | C83.56 | Lymphoblastic (diffuse) lymphoma, intrapelvic lymph nodes |
| ICD10CM | C83.57 | Lymphoblastic (diffuse) lymphoma, spleen |
| ICD10CM | C83.58 | Lymphoblastic (diffuse) lymphoma, lymph nodes of multiple sites |
| ICD10CM | C83.59 | Lymphoblastic (diffuse) lymphoma, extranodal and solid organ sites |
| ICD10CM | C83.70 | Burkitt lymphoma, unspecified site |
| ICD10CM | C83.71 | Burkitt lymphoma, lymph nodes of head, face, and neck |
| ICD10CM | C83.72 | Burkitt lymphoma, intrathoracic lymph nodes |
| ICD10CM | C83.73 | Burkitt lymphoma, intra-abdominal lymph nodes |
| ICD10CM | C83.74 | Burkitt lymphoma, lymph nodes of axilla and upper limb |
| ICD10CM | C83.75 | Burkitt lymphoma, lymph nodes of inguinal region and lower limb |
| ICD10CM | C83.76 | Burkitt lymphoma, intrapelvic lymph nodes |
| ICD10CM | C83.77 | Burkitt lymphoma, spleen |
| ICD10CM | C83.78 | Burkitt lymphoma, lymph nodes of multiple sites |
| ICD10CM | C83.79 | Burkitt lymphoma, extranodal and solid organ sites |
| ICD10CM | C83.80 | Other non-follicular lymphoma, unspecified site |
| ICD10CM | C83.81 | Other non-follicular lymphoma, lymph nodes of head, face, and neck |
| ICD10CM | C83.82 | Other non-follicular lymphoma, intrathoracic lymph nodes |
| ICD10CM | C83.83 | Other non-follicular lymphoma, intra-abdominal lymph nodes |
| ICD10CM | C83.84 | Other non-follicular lymphoma, lymph nodes of axilla and upper limb |

Continued on next page

**TableS24 – continued from previous page**

| CodeSystem | ConceptCode | ConceptName |
|---|---|---|
| ICD10CM | C83.85 | Other non-follicular lymphoma, lymph nodes of inguinal region and lower limb |
| ICD10CM | C83.86 | Other non-follicular lymphoma, intrapelvic lymph nodes |
| ICD10CM | C83.87 | Other non-follicular lymphoma, spleen |
| ICD10CM | C83.88 | Other non-follicular lymphoma, lymph nodes of multiple sites |
| ICD10CM | C83.89 | Other non-follicular lymphoma, extranodal and solid organ sites |
| ICD10CM | C83.90 | Non-follicular (diffuse) lymphoma, unspecified, unspecified site |
| ICD10CM | C83.91 | Non-follicular (diffuse) lymphoma, unspecified, lymph nodes of head, face, and neck |
| ICD10CM | C83.92 | Non-follicular (diffuse) lymphoma, unspecified, intrathoracic lymph nodes |
| ICD10CM | C83.93 | Non-follicular (diffuse) lymphoma, unspecified, intra-abdominal lymph nodes |
| ICD10CM | C83.94 | Non-follicular (diffuse) lymphoma, unspecified, lymph nodes of axilla and upper limb |
| ICD10CM | C83.95 | Non-follicular (diffuse) lymphoma, unspecified, lymph nodes of inguinal region and lower limb |
| ICD10CM | C83.96 | Non-follicular (diffuse) lymphoma, unspecified, intrapelvic lymph nodes |
| ICD10CM | C83.97 | Non-follicular (diffuse) lymphoma, unspecified, spleen |
| ICD10CM | C83.98 | Non-follicular (diffuse) lymphoma, unspecified, lymph nodes of multiple sites |
| ICD10CM | C83.99 | Non-follicular (diffuse) lymphoma, unspecified, extranodal and solid organ sites |
| ICD10CM | C84.00 | Mycosis fungoides, unspecified site |
| ICD10CM | C84.01 | Mycosis fungoides, lymph nodes of head, face, and neck |
| ICD10CM | C84.02 | Mycosis fungoides, intrathoracic lymph nodes |
| ICD10CM | C84.03 | Mycosis fungoides, intra-abdominal lymph nodes |
| ICD10CM | C84.04 | Mycosis fungoides, lymph nodes of axilla and upper limb |
| ICD10CM | C84.05 | Mycosis fungoides, lymph nodes of inguinal region and lower limb |
| ICD10CM | C84.06 | Mycosis fungoides, intrapelvic lymph nodes |
| ICD10CM | C84.07 | Mycosis fungoides, spleen |
| ICD10CM | C84.08 | Mycosis fungoides, lymph nodes of multiple sites |
| ICD10CM | C84.09 | Mycosis fungoides, extranodal and solid organ sites |
| ICD10CM | C84.10 | Sezary disease, unspecified site |
| ICD10CM | C84.11 | Sezary disease, lymph nodes of head, face, and neck |
| ICD10CM | C84.12 | Sezary disease, intrathoracic lymph nodes |
| ICD10CM | C84.13 | Sezary disease, intra-abdominal lymph nodes |
| ICD10CM | C84.14 | Sezary disease, lymph nodes of axilla and upper limb |
| ICD10CM | C84.15 | Sezary disease, lymph nodes of inguinal region and lower limb |
| ICD10CM | C84.16 | Sezary disease, intrapelvic lymph nodes |
| ICD10CM | C84.17 | Sezary disease, spleen |
| ICD10CM | C84.18 | Sezary disease, lymph nodes of multiple sites |
| ICD10CM | C84.19 | Sezary disease, extranodal and solid organ sites |
| ICD10CM | C84.40 | Peripheral T-cell lymphoma, not classified, unspecified site |
| ICD10CM | C84.41 | Peripheral T-cell lymphoma, not classified, lymph nodes of head, face, and neck |

Continued on next page

**TableS24 – continued from previous page**

| CodeSystem | ConceptCode | ConceptName |
|---|---|---|
| ICD10CM | C84.42 | Peripheral T-cell lymphoma, not classified, intrathoracic lymph nodes |
| ICD10CM | C84.43 | Peripheral T-cell lymphoma, not classified, intra-abdominal lymph nodes |
| ICD10CM | C84.44 | Peripheral T-cell lymphoma, not classified, lymph nodes of axilla and upper limb |
| ICD10CM | C84.45 | Peripheral T-cell lymphoma, not classified, lymph nodes of inguinal region and lower limb |
| ICD10CM | C84.46 | Peripheral T-cell lymphoma, not classified, intrapelvic lymph nodes |
| ICD10CM | C84.47 | Peripheral T-cell lymphoma, not classified, spleen |
| ICD10CM | C84.48 | Peripheral T-cell lymphoma, not classified, lymph nodes of multiple sites |
| ICD10CM | C84.49 | Peripheral T-cell lymphoma, not classified, extranodal and solid organ sites |
| ICD10CM | C84.60 | Anaplastic large cell lymphoma, ALK-positive, unspecified site |
| ICD10CM | C84.61 | Anaplastic large cell lymphoma, ALK-positive, lymph nodes of head, face, and neck |
| ICD10CM | C84.62 | Anaplastic large cell lymphoma, ALK-positive, intrathoracic lymph nodes |
| ICD10CM | C84.63 | Anaplastic large cell lymphoma, ALK-positive, intra-abdominal lymph nodes |
| ICD10CM | C84.64 | Anaplastic large cell lymphoma, ALK-positive, lymph nodes of axilla and upper limb |
| ICD10CM | C84.65 | Anaplastic large cell lymphoma, ALK-positive, lymph nodes of inguinal region and lower limb |
| ICD10CM | C84.66 | Anaplastic large cell lymphoma, ALK-positive, intrapelvic lymph nodes |
| ICD10CM | C84.67 | Anaplastic large cell lymphoma, ALK-positive, spleen |
| ICD10CM | C84.68 | Anaplastic large cell lymphoma, ALK-positive, lymph nodes of multiple sites |
| ICD10CM | C84.69 | Anaplastic large cell lymphoma, ALK-positive, extranodal and solid organ sites |
| ICD10CM | C84.70 | Anaplastic large cell lymphoma, ALK-negative, unspecified site |
| ICD10CM | C84.71 | Anaplastic large cell lymphoma, ALK-negative, lymph nodes of head, face, and neck |
| ICD10CM | C84.72 | Anaplastic large cell lymphoma, ALK-negative, intrathoracic lymph nodes |
| ICD10CM | C84.73 | Anaplastic large cell lymphoma, ALK-negative, intra-abdominal lymph nodes |
| ICD10CM | C84.74 | Anaplastic large cell lymphoma, ALK-negative, lymph nodes of axilla and upper limb |
| ICD10CM | C84.75 | Anaplastic large cell lymphoma, ALK-negative, lymph nodes of inguinal region and lower limb |
| ICD10CM | C84.76 | Anaplastic large cell lymphoma, ALK-negative, intrapelvic lymph nodes |
| ICD10CM | C84.77 | Anaplastic large cell lymphoma, ALK-negative, spleen |

Continued on next page

**TableS24 – continued from previous page**

| CodeSystem | ConceptCode | ConceptName |
|---|---|---|
| ICD10CM | C84.78 | Anaplastic large cell lymphoma, ALK-negative, lymph nodes of multiple sites |
| ICD10CM | C84.79 | Anaplastic large cell lymphoma, ALK-negative, extranodal and solid organ sites |
| ICD10CM | C84.90 | Mature T/NK-cell lymphomas, unspecified, unspecified site |
| ICD10CM | C84.91 | Mature T/NK-cell lymphomas, unspecified, lymph nodes of head, face, and neck |
| ICD10CM | C84.92 | Mature T/NK-cell lymphomas, unspecified, intrathoracic lymph nodes |
| ICD10CM | C84.93 | Mature T/NK-cell lymphomas, unspecified, intra-abdominal lymph nodes |
| ICD10CM | C84.94 | Mature T/NK-cell lymphomas, unspecified, lymph nodes of axilla and upper limb |
| ICD10CM | C84.95 | Mature T/NK-cell lymphomas, unspecified, lymph nodes of inguinal region and lower limb |
| ICD10CM | C84.96 | Mature T/NK-cell lymphomas, unspecified, intrapelvic lymph nodes |
| ICD10CM | C84.97 | Mature T/NK-cell lymphomas, unspecified, spleen |
| ICD10CM | C84.98 | Mature T/NK-cell lymphomas, unspecified, lymph nodes of multiple sites |
| ICD10CM | C84.99 | Mature T/NK-cell lymphomas, unspecified, extranodal and solid organ sites |
| ICD10CM | C84.A0 | Cutaneous T-cell lymphoma, unspecified, unspecified site |
| ICD10CM | C84.A1 | Cutaneous T-cell lymphoma, unspecified lymph nodes of head, face, and neck |
| ICD10CM | C84.A2 | Cutaneous T-cell lymphoma, unspecified, intrathoracic lymph nodes |
| ICD10CM | C84.A3 | Cutaneous T-cell lymphoma, unspecified, intra-abdominal lymph nodes |
| ICD10CM | C84.A4 | Cutaneous T-cell lymphoma, unspecified, lymph nodes of axilla and upper limb |
| ICD10CM | C84.A5 | Cutaneous T-cell lymphoma, unspecified, lymph nodes of inguinal region and lower limb |
| ICD10CM | C84.A6 | Cutaneous T-cell lymphoma, unspecified, intrapelvic lymph nodes |
| ICD10CM | C84.A7 | Cutaneous T-cell lymphoma, unspecified, spleen |
| ICD10CM | C84.A8 | Cutaneous T-cell lymphoma, unspecified, lymph nodes of multiple sites |
| ICD10CM | C84.A9 | Cutaneous T-cell lymphoma, unspecified, extranodal and solid organ sites |
| ICD10CM | C84.Z0 | Other mature T/NK-cell lymphomas, unspecified site |
| ICD10CM | C84.Z1 | Other mature T/NK-cell lymphomas, lymph nodes of head, face, and neck |
| ICD10CM | C84.Z2 | Other mature T/NK-cell lymphomas, intrathoracic lymph nodes |
| ICD10CM | C84.Z3 | Other mature T/NK-cell lymphomas, intra-abdominal lymph nodes |

Continued on next page

**TableS24 – continued from previous page**

| CodeSystem | ConceptCode | ConceptName |
|---|---|---|
| ICD10CM | C84.Z4 | Other mature T/NK-cell lymphomas, lymph nodes of axilla and upper limb |
| ICD10CM | C84.Z5 | Other mature T/NK-cell lymphomas, lymph nodes of inguinal region and lower limb |
| ICD10CM | C84.Z6 | Other mature T/NK-cell lymphomas, intrapelvic lymph nodes |
| ICD10CM | C84.Z7 | Other mature T/NK-cell lymphomas, spleen |
| ICD10CM | C84.Z8 | Other mature T/NK-cell lymphomas, lymph nodes of multiple sites |
| ICD10CM | C84.Z9 | Other mature T/NK-cell lymphomas, extranodal and solid organ sites |
| ICD10CM | C85.10 | Unspecified B-cell lymphoma, unspecified site |
| ICD10CM | C85.11 | Unspecified B-cell lymphoma, lymph nodes of head, face, and neck |
| ICD10CM | C85.12 | Unspecified B-cell lymphoma, intrathoracic lymph nodes |
| ICD10CM | C85.13 | Unspecified B-cell lymphoma, intra-abdominal lymph nodes |
| ICD10CM | C85.14 | Unspecified B-cell lymphoma, lymph nodes of axilla and upper limb |
| ICD10CM | C85.15 | Unspecified B-cell lymphoma, lymph nodes of inguinal region and lower limb |
| ICD10CM | C85.16 | Unspecified B-cell lymphoma, intrapelvic lymph nodes |
| ICD10CM | C85.17 | Unspecified B-cell lymphoma, spleen |
| ICD10CM | C85.18 | Unspecified B-cell lymphoma, lymph nodes of multiple sites |
| ICD10CM | C85.19 | Unspecified B-cell lymphoma, extranodal and solid organ sites |
| ICD10CM | C85.20 | Mediastinal (thymic) large B-cell lymphoma, unspecified site |
| ICD10CM | C85.21 | Mediastinal (thymic) large B-cell lymphoma, lymph nodes of head, face, and neck |
| ICD10CM | C85.22 | Mediastinal (thymic) large B-cell lymphoma, intrathoracic lymph nodes |
| ICD10CM | C85.23 | Mediastinal (thymic) large B-cell lymphoma, intra-abdominal lymph nodes |
| ICD10CM | C85.24 | Mediastinal (thymic) large B-cell lymphoma, lymph nodes of axilla and upper limb |
| ICD10CM | C85.25 | Mediastinal (thymic) large B-cell lymphoma, lymph nodes of inguinal region and lower limb |
| ICD10CM | C85.26 | Mediastinal (thymic) large B-cell lymphoma, intrapelvic lymph nodes |
| ICD10CM | C85.27 | Mediastinal (thymic) large B-cell lymphoma, spleen |
| ICD10CM | C85.28 | Mediastinal (thymic) large B-cell lymphoma, lymph nodes of multiple sites |
| ICD10CM | C85.29 | Mediastinal (thymic) large B-cell lymphoma, extranodal and solid organ sites |
| ICD10CM | C85.80 | Other specified types of non-Hodgkin lymphoma, unspecified site |
| ICD10CM | C85.81 | Other specified types of non-Hodgkin lymphoma, lymph nodes of head, face, and neck |
| ICD10CM | C85.82 | Other specified types of non-Hodgkin lymphoma, intrathoracic lymph nodes |

Continued on next page

**TableS24 – continued from previous page**

| CodeSystem | ConceptCode | ConceptName |
|---|---|---|
| ICD10CM | C85.83 | Other specified types of non-Hodgkin lymphoma, intra-abdominal lymph nodes |
| ICD10CM | C85.84 | Other specified types of non-Hodgkin lymphoma, lymph nodes of axilla and upper limb |
| ICD10CM | C85.85 | Other specified types of non-Hodgkin lymphoma, lymph nodes of inguinal region and lower limb |
| ICD10CM | C85.86 | Other specified types of non-Hodgkin lymphoma, intrapelvic lymph nodes |
| ICD10CM | C85.87 | Other specified types of non-Hodgkin lymphoma, spleen |
| ICD10CM | C85.88 | Other specified types of non-Hodgkin lymphoma, lymph nodes of multiple sites |
| ICD10CM | C85.89 | Other specified types of non-Hodgkin lymphoma, extranodal and solid organ sites |
| ICD10CM | C85.90 | Non-Hodgkin lymphoma, unspecified, unspecified site |
| ICD10CM | C85.91 | Non-Hodgkin lymphoma, unspecified, lymph nodes of head, face, and neck |
| ICD10CM | C85.92 | Non-Hodgkin lymphoma, unspecified, intrathoracic lymph nodes |
| ICD10CM | C85.93 | Non-Hodgkin lymphoma, unspecified, intra-abdominal lymph nodes |
| ICD10CM | C85.94 | Non-Hodgkin lymphoma, unspecified, lymph nodes of axilla and upper limb |
| ICD10CM | C85.95 | Non-Hodgkin lymphoma, unspecified, lymph nodes of inguinal region and lower limb |
| ICD10CM | C85.96 | Non-Hodgkin lymphoma, unspecified, intrapelvic lymph nodes |
| ICD10CM | C85.97 | Non-Hodgkin lymphoma, unspecified, spleen |
| ICD10CM | C85.98 | Non-Hodgkin lymphoma, unspecified, lymph nodes of multiple sites |
| ICD10CM | C85.99 | Non-Hodgkin lymphoma, unspecified, extranodal and solid organ sites |
| ICD10CM | C86.0 | Extranodal NK/T-cell lymphoma, nasal type |
| ICD10CM | C86.1 | Hepatosplenic T-cell lymphoma |
| ICD10CM | C86.2 | Enteropathy-type (intestinal) T-cell lymphoma |
| ICD10CM | C86.3 | Subcutaneous panniculitis-like T-cell lymphoma |
| ICD10CM | C86.4 | Blastic NK-cell lymphoma |
| ICD10CM | C86.5 | Angioimmunoblastic T-cell lymphoma |
| ICD10CM | C86.6 | Primary cutaneous CD30-positive T-cell proliferations |
| ICD10CM | C88.2 | Heavy chain disease |
| ICD10CM | C88.3 | Immunoproliferative small intestinal disease |
| ICD10CM | C88.4 | Extranodal marginal zone B-cell lymphoma of mucosa-associated lymphoid tissue [MALT-lymphoma] |
| ICD10CM | C88.8 | Other malignant immunoproliferative diseases |
| ICD10CM | C88.9 | Malignant immunoproliferative disease, unspecified |
| ICD10CM | C90.00 | Multiple myeloma not having achieved remission |
| ICD10CM | C90.01 | Multiple myeloma in remission |
| ICD10CM | C90.02 | Multiple myeloma in relapse |
| ICD10CM | C90.10 | Plasma cell leukemia not having achieved remission |
| ICD10CM | C90.11 | Plasma cell leukemia in remission |

Continued on next page

**TableS24 – continued from previous page**

| CodeSystem | ConceptCode | ConceptName |
|---|---|---|
| ICD10CM | C90.12 | Plasma cell leukemia in relapse |
| ICD10CM | C90.20 | Extramedullary plasmacytoma not having achieved remission |
| ICD10CM | C90.21 | Extramedullary plasmacytoma in remission |
| ICD10CM | C90.22 | Extramedullary plasmacytoma in relapse |
| ICD10CM | C90.30 | Solitary plasmacytoma not having achieved remission |
| ICD10CM | C90.31 | Solitary plasmacytoma in remission |
| ICD10CM | C90.32 | Solitary plasmacytoma in relapse |
| ICD10CM | C91.00 | Acute lymphoblastic leukemia not having achieved remission |
| ICD10CM | C91.01 | Acute lymphoblastic leukemia, in remission |
| ICD10CM | C91.02 | Acute lymphoblastic leukemia, in relapse |
| ICD10CM | C91.10 | Chronic lymphocytic leukemia of B-cell type not having achieved remission |
| ICD10CM | C91.11 | Chronic lymphocytic leukemia of B-cell type in remission |
| ICD10CM | C91.12 | Chronic lymphocytic leukemia of B-cell type in relapse |
| ICD10CM | C91.30 | Prolymphocytic leukemia of B-cell type not having achieved remission |
| ICD10CM | C91.31 | Prolymphocytic leukemia of B-cell type, in remission |
| ICD10CM | C91.32 | Prolymphocytic leukemia of B-cell type, in relapse |
| ICD10CM | C91.40 | Hairy cell leukemia not having achieved remission |
| ICD10CM | C91.41 | Hairy cell leukemia, in remission |
| ICD10CM | C91.42 | Hairy cell leukemia, in relapse |
| ICD10CM | C91.50 | Adult T-cell lymphoma/leukemia (HTLV-1-associated) not having achieved remission |
| ICD10CM | C91.51 | Adult T-cell lymphoma/leukemia (HTLV-1-associated), in remission |
| ICD10CM | C91.52 | Adult T-cell lymphoma/leukemia (HTLV-1-associated), in relapse |
| ICD10CM | C91.60 | Prolymphocytic leukemia of T-cell type not having achieved remission |
| ICD10CM | C91.61 | Prolymphocytic leukemia of T-cell type, in remission |
| ICD10CM | C91.62 | Prolymphocytic leukemia of T-cell type, in relapse |
| ICD10CM | C91.90 | Lymphoid leukemia, unspecified not having achieved remission |
| ICD10CM | C91.91 | Lymphoid leukemia, unspecified, in remission |
| ICD10CM | C91.92 | Lymphoid leukemia, unspecified, in relapse |
| ICD10CM | C91.A0 | Mature B-cell leukemia Burkitt-type not having achieved remission |
| ICD10CM | C91.A1 | Mature B-cell leukemia Burkitt-type, in remission |
| ICD10CM | C91.A2 | Mature B-cell leukemia Burkitt-type, in relapse |
| ICD10CM | C91.Z0 | Other lymphoid leukemia not having achieved remission |
| ICD10CM | C91.Z1 | Other lymphoid leukemia, in remission |
| ICD10CM | C91.Z2 | Other lymphoid leukemia, in relapse |
| ICD10CM | C92.00 | Acute myeloblastic leukemia, not having achieved remission |
| ICD10CM | C92.01 | Acute myeloblastic leukemia, in remission |
| ICD10CM | C92.02 | Acute myeloblastic leukemia, in relapse |
| ICD10CM | C92.10 | Chronic myeloid leukemia, BCR/ABL-positive, not having achieved remission |
| ICD10CM | C92.11 | Chronic myeloid leukemia, BCR/ABL-positive, in remission |

Continued on next page

**TableS24 – continued from previous page**

| CodeSystem | ConceptCode | ConceptName |
|---|---|---|
| ICD10CM | C92.12 | Chronic myeloid leukemia, BCR/ABL-positive, in relapse |
| ICD10CM | C92.20 | Atypical chronic myeloid leukemia, BCR/ABL-negative, not having achieved remission |
| ICD10CM | C92.21 | Atypical chronic myeloid leukemia, BCR/ABL-negative, in remission |
| ICD10CM | C92.22 | Atypical chronic myeloid leukemia, BCR/ABL-negative, in relapse |
| ICD10CM | C92.30 | Myeloid sarcoma, not having achieved remission |
| ICD10CM | C92.31 | Myeloid sarcoma, in remission |
| ICD10CM | C92.32 | Myeloid sarcoma, in relapse |
| ICD10CM | C92.40 | Acute promyelocytic leukemia, not having achieved remission |
| ICD10CM | C92.41 | Acute promyelocytic leukemia, in remission |
| ICD10CM | C92.42 | Acute promyelocytic leukemia, in relapse |
| ICD10CM | C92.50 | Acute myelomonocytic leukemia, not having achieved remission |
| ICD10CM | C92.51 | Acute myelomonocytic leukemia, in remission |
| ICD10CM | C92.52 | Acute myelomonocytic leukemia, in relapse |
| ICD10CM | C92.60 | Acute myeloid leukemia with 11q23-abnormality not having achieved remission |
| ICD10CM | C92.61 | Acute myeloid leukemia with 11q23-abnormality in remission |
| ICD10CM | C92.62 | Acute myeloid leukemia with 11q23-abnormality in relapse |
| ICD10CM | C92.90 | Myeloid leukemia, unspecified, not having achieved remission |
| ICD10CM | C92.91 | Myeloid leukemia, unspecified in remission |
| ICD10CM | C92.92 | Myeloid leukemia, unspecified in relapse |
| ICD10CM | C92.A0 | Acute myeloid leukemia with multilineage dysplasia, not having achieved remission |
| ICD10CM | C92.A1 | Acute myeloid leukemia with multilineage dysplasia, in remission |
| ICD10CM | C92.A2 | Acute myeloid leukemia with multilineage dysplasia, in relapse |
| ICD10CM | C92.Z0 | Other myeloid leukemia not having achieved remission |
| ICD10CM | C92.Z1 | Other myeloid leukemia, in remission |
| ICD10CM | C92.Z2 | Other myeloid leukemia, in relapse |
| ICD10CM | C93.00 | Acute monoblastic/monocytic leukemia, not having achieved remission |
| ICD10CM | C93.01 | Acute monoblastic/monocytic leukemia, in remission |
| ICD10CM | C93.02 | Acute monoblastic/monocytic leukemia, in relapse |
| ICD10CM | C93.10 | Chronic myelomonocytic leukemia not having achieved remission |
| ICD10CM | C93.11 | Chronic myelomonocytic leukemia, in remission |
| ICD10CM | C93.12 | Chronic myelomonocytic leukemia, in relapse |
| ICD10CM | C93.30 | Juvenile myelomonocytic leukemia, not having achieved remission |
| ICD10CM | C93.31 | Juvenile myelomonocytic leukemia, in remission |
| ICD10CM | C93.32 | Juvenile myelomonocytic leukemia, in relapse |
| ICD10CM | C93.90 | Monocytic leukemia, unspecified, not having achieved remission |
| ICD10CM | C93.91 | Monocytic leukemia, unspecified in remission |
| ICD10CM | C93.92 | Monocytic leukemia, unspecified in relapse |
| ICD10CM | C93.Z0 | Other monocytic leukemia, not having achieved remission |
| ICD10CM | C93.Z1 | Other monocytic leukemia, in remission |
| ICD10CM | C93.Z2 | Other monocytic leukemia, in relapse |

Continued on next page

**TableS24 – continued from previous page**

| CodeSystem | ConceptCode | ConceptName |
|---|---|---|
| ICD10CM | C94.00 | Acute erythroid leukemia, not having achieved remission |
| ICD10CM | C94.01 | Acute erythroid leukemia, in remission |
| ICD10CM | C94.02 | Acute erythroid leukemia, in relapse |
| ICD10CM | C94.20 | Acute megakaryoblastic leukemia not having achieved remission |
| ICD10CM | C94.21 | Acute megakaryoblastic leukemia, in remission |
| ICD10CM | C94.22 | Acute megakaryoblastic leukemia, in relapse |
| ICD10CM | C94.30 | Mast cell leukemia not having achieved remission |
| ICD10CM | C94.31 | Mast cell leukemia, in remission |
| ICD10CM | C94.32 | Mast cell leukemia, in relapse |
| ICD10CM | C94.40 | Acute panmyelosis with myelofibrosis not having achieved remission |
| ICD10CM | C94.41 | Acute panmyelosis with myelofibrosis, in remission |
| ICD10CM | C94.42 | Acute panmyelosis with myelofibrosis, in relapse |
| ICD10CM | C94.6 | Myelodysplastic disease, not classified |
| ICD10CM | C94.80 | Other specified leukemias not having achieved remission |
| ICD10CM | C94.81 | Other specified leukemias, in remission |
| ICD10CM | C94.82 | Other specified leukemias, in relapse |
| ICD10CM | C95.00 | Acute leukemia of unspecified cell type not having achieved remission |
| ICD10CM | C95.01 | Acute leukemia of unspecified cell type, in remission |
| ICD10CM | C95.02 | Acute leukemia of unspecified cell type, in relapse |
| ICD10CM | C95.10 | Chronic leukemia of unspecified cell type not having achieved remission |
| ICD10CM | C95.11 | Chronic leukemia of unspecified cell type, in remission |
| ICD10CM | C95.12 | Chronic leukemia of unspecified cell type, in relapse |
| ICD10CM | C95.90 | Leukemia, unspecified not having achieved remission |
| ICD10CM | C95.91 | Leukemia, unspecified, in remission |
| ICD10CM | C95.92 | Leukemia, unspecified, in relapse |
| ICD10CM | C96.0 | Multifocal and multisystemic (disseminated) Langerhans-cell histiocytosis |
| ICD10CM | C96.2 | Malignant mast cell neoplasm |
| ICD10CM | C96.20 | Malignant mast cell neoplasm, unspecified |
| ICD10CM | C96.21 | Aggressive systemic mastocytosis |
| ICD10CM | C96.22 | Mast cell sarcoma |
| ICD10CM | C96.29 | Other malignant mast cell neoplasm |
| ICD10CM | C96.4 | Sarcoma of dendritic cells (accessory cells) |
| ICD10CM | C96.9 | Malignant neoplasm of lymphoid, hematopoietic and related tissue, unspecified |
| ICD10CM | C96.A | Histiocytic sarcoma |
| ICD10CM | C96.Z | Other specified malignant neoplasms of lymphoid, hematopoietic and related tissue |
| ICD10CM | D03.0 | Melanoma in situ of lip |
| ICD10CM | D03.10 | Melanoma in situ of unspecified eyelid, including canthus |
| ICD10CM | D03.11 | Melanoma in situ of right eyelid, including canthus |
| ICD10CM | D03.12 | Melanoma in situ of left eyelid, including canthus |
| ICD10CM | D03.20 | Melanoma in situ of unspecified ear and external auricular canal |
| ICD10CM | D03.21 | Melanoma in situ of right ear and external auricular canal |

Continued on next page

**TableS24 – continued from previous page**

| CodeSystem | ConceptCode | ConceptName |
|---|---|---|
| ICD10CM | D03.22 | Melanoma in situ of left ear and external auricular canal |
| ICD10CM | D03.30 | Melanoma in situ of unspecified part of face |
| ICD10CM | D03.39 | Melanoma in situ of other parts of face |
| ICD10CM | D03.4 | Melanoma in situ of scalp and neck |
| ICD10CM | D03.51 | Melanoma in situ of anal skin |
| ICD10CM | D03.52 | Melanoma in situ of breast (skin) (soft tissue) |
| ICD10CM | D03.59 | Melanoma in situ of other part of trunk |
| ICD10CM | D03.60 | Melanoma in situ of unspecified upper limb, including shoulder |
| ICD10CM | D03.61 | Melanoma in situ of right upper limb, including shoulder |
| ICD10CM | D03.62 | Melanoma in situ of left upper limb, including shoulder |
| ICD10CM | D03.70 | Melanoma in situ of unspecified lower limb, including hip |
| ICD10CM | D03.71 | Melanoma in situ of right lower limb, including hip |
| ICD10CM | D03.72 | Melanoma in situ of left lower limb, including hip |
| ICD10CM | D03.8 | Melanoma in situ of other sites |
| ICD10CM | D03.9 | Melanoma in situ, unspecified |
| ICD10CM | D47.Z2 | Castleman disease |
| ICD10CM | D49.511 | Neoplasm of unspecified behavior of right kidney |
| ICD10CM | D49.512 | Neoplasm of unspecified behavior of left kidney |
| ICD10CM | D49.519 | Neoplasm of unspecified behavior of unspecified kidney |
| ICD10CM | D49.59 | Neoplasm of unspecified behavior of other genitourinary organ |
| ICD10CM | C43.111 | Malignant melanoma of right upper eyelid, including canthus |
| ICD10CM | C43.112 | Malignant melanoma of right lower eyelid, including canthus |
| ICD10CM | C43.121 | Malignant melanoma of left upper eyelid, including canthus |
| ICD10CM | C43.122 | Malignant melanoma of left lower eyelid, including canthus |
| ICD10CM | C44.1021 | Unspecified malignant neoplasm of skin of right upper eyelid, including canthus |
| ICD10CM | C44.1022 | Unspecified malignant neoplasm of skin of right lower eyelid, including canthus |
| ICD10CM | C44.1091 | Unspecified malignant neoplasm of skin of left upper eyelid, including canthus |
| ICD10CM | C44.1092 | Unspecified malignant neoplasm of skin of left lower eyelid, including canthus |
| ICD10CM | C44.1121 | Basal cell carcinoma of skin of right upper eyelid, including canthus |
| ICD10CM | C44.1122 | Basal cell carcinoma of skin of right lower eyelid, including canthus |
| ICD10CM | C44.1191 | Basal cell carcinoma of skin of left upper eyelid, including canthus |
| ICD10CM | C44.1192 | Basal cell carcinoma of skin of left lower eyelid, including canthus |
| ICD10CM | C44.1221 | Squamous cell carcinoma of skin of right upper eyelid, including canthus |
| ICD10CM | C44.1222 | Squamous cell carcinoma of skin of right lower eyelid, including canthus |
| ICD10CM | C44.1291 | Squamous cell carcinoma of skin of left upper eyelid, including canthus |
| ICD10CM | C44.1292 | Squamous cell carcinoma of skin of left lower eyelid, including canthus |

Continued on next page

**TableS24 – continued from previous page**

| CodeSystem | ConceptCode | ConceptName |
|---|---|---|
| ICD10CM | C44.1921 | Other specified malignant neoplasm of skin of right upper eyelid, including canthus |
| ICD10CM | C44.1922 | Other specified malignant neoplasm of skin of right lower eyelid, including canthus |
| ICD10CM | C44.1991 | Other specified malignant neoplasm of skin of left upper eyelid, including canthus |
| ICD10CM | C44.1992 | Other specified malignant neoplasm of skin of left lower eyelid, including canthus |
| ICD10CM | C4A.111 | Merkel cell carcinoma of right upper eyelid, including canthus |
| ICD10CM | C4A.112 | Merkel cell carcinoma of right lower eyelid, including canthus |
| ICD10CM | C4A.121 | Merkel cell carcinoma of left upper eyelid, including canthus |
| ICD10CM | C4A.122 | Merkel cell carcinoma of left lower eyelid, including canthus |
| ICD10CM | D03.111 | Melanoma in situ of right upper eyelid, including canthus |
| ICD10CM | D03.112 | Melanoma in situ of right lower eyelid, including canthus |
| ICD10CM | D03.121 | Melanoma in situ of left upper eyelid, including canthus |
| ICD10CM | D03.122 | Melanoma in situ of left lower eyelid, including canthus |
| ICD9CM | 140.0 | Malignant neoplasm of upper lip, vermilion border |
| ICD9CM | 140.1 | Malignant neoplasm of lower lip, vermilion border |
| ICD9CM | 140.3 | Malignant neoplasm of upper lip, inner aspect |
| ICD9CM | 140.4 | Malignant neoplasm of lower lip, inner aspect |
| ICD9CM | 140.5 | Malignant neoplasm of lip, unspecified, inner aspect |
| ICD9CM | 140.6 | Malignant neoplasm of commissure of lip |
| ICD9CM | 140.8 | Malignant neoplasm of other sites of lip |
| ICD9CM | 140.9 | Malignant neoplasm of lip, unspecified, vermilion border |
| ICD9CM | 141.0 | Malignant neoplasm of base of tongue |
| ICD9CM | 141.1 | Malignant neoplasm of dorsal surface of tongue |
| ICD9CM | 141.2 | Malignant neoplasm of tip and lateral border of tongue |
| ICD9CM | 141.3 | Malignant neoplasm of ventral surface of tongue |
| ICD9CM | 141.4 | Malignant neoplasm of anterior two-thirds of tongue, part unspecified |
| ICD9CM | 141.5 | Malignant neoplasm of junctional zone of tongue |
| ICD9CM | 141.6 | Malignant neoplasm of lingual tonsil |
| ICD9CM | 141.8 | Malignant neoplasm of other sites of tongue |
| ICD9CM | 141.9 | Malignant neoplasm of tongue, unspecified |
| ICD9CM | 142.0 | Malignant neoplasm of parotid gland |
| ICD9CM | 142.1 | Malignant neoplasm of submandibular gland |
| ICD9CM | 142.2 | Malignant neoplasm of sublingual gland |
| ICD9CM | 142.8 | Malignant neoplasm of other major salivary glands |
| ICD9CM | 142.9 | Malignant neoplasm of salivary gland, unspecified |
| ICD9CM | 143.0 | Malignant neoplasm of upper gum |
| ICD9CM | 143.1 | Malignant neoplasm of lower gum |
| ICD9CM | 143.8 | Malignant neoplasm of other sites of gum |
| ICD9CM | 143.9 | Malignant neoplasm of gum, unspecified |
| ICD9CM | 144.0 | Malignant neoplasm of anterior portion of floor of mouth |
| ICD9CM | 144.1 | Malignant neoplasm of lateral portion of floor of mouth |
| ICD9CM | 144.8 | Malignant neoplasm of other sites of floor of mouth |
| ICD9CM | 144.9 | Malignant neoplasm of floor of mouth, part unspecified |

Continued on next page

**TableS24 – continued from previous page**

| CodeSystem | ConceptCode | ConceptName |
|---|---|---|
| ICD9CM | 145.0 | Malignant neoplasm of cheek mucosa |
| ICD9CM | 145.1 | Malignant neoplasm of vestibule of mouth |
| ICD9CM | 145.2 | Malignant neoplasm of hard palate |
| ICD9CM | 145.3 | Malignant neoplasm of soft palate |
| ICD9CM | 145.4 | Malignant neoplasm of uvula |
| ICD9CM | 145.5 | Malignant neoplasm of palate, unspecified |
| ICD9CM | 145.6 | Malignant neoplasm of retromolar area |
| ICD9CM | 145.8 | Malignant neoplasm of other specified parts of mouth |
| ICD9CM | 145.9 | Malignant neoplasm of mouth, unspecified |
| ICD9CM | 146.0 | Malignant neoplasm of tonsil |
| ICD9CM | 146.1 | Malignant neoplasm of tonsillar fossa |
| ICD9CM | 146.2 | Malignant neoplasm of tonsillar pillars (anterior) (posterior) |
| ICD9CM | 146.3 | Malignant neoplasm of vallecula epiglottica |
| ICD9CM | 146.4 | Malignant neoplasm of anterior aspect of epiglottis |
| ICD9CM | 146.5 | Malignant neoplasm of junctional region of oropharynx |
| ICD9CM | 146.6 | Malignant neoplasm of lateral wall of oropharynx |
| ICD9CM | 146.7 | Malignant neoplasm of posterior wall of oropharynx |
| ICD9CM | 146.8 | Malignant neoplasm of other specified sites of oropharynx |
| ICD9CM | 146.9 | Malignant neoplasm of oropharynx, unspecified site |
| ICD9CM | 147.0 | Malignant neoplasm of superior wall of nasopharynx |
| ICD9CM | 147.1 | Malignant neoplasm of posterior wall of nasopharynx |
| ICD9CM | 147.2 | Malignant neoplasm of lateral wall of nasopharynx |
| ICD9CM | 147.3 | Malignant neoplasm of anterior wall of nasopharynx |
| ICD9CM | 147.8 | Malignant neoplasm of other specified sites of nasopharynx |
| ICD9CM | 147.9 | Malignant neoplasm of nasopharynx, unspecified site |
| ICD9CM | 148.0 | Malignant neoplasm of postcricoid region of hypopharynx |
| ICD9CM | 148.1 | Malignant neoplasm of pyriform sinus |
| ICD9CM | 148.2 | Malignant neoplasm of aryepiglottic fold, hypopharyngeal aspect |
| ICD9CM | 148.3 | Malignant neoplasm of posterior hypopharyngeal wall |
| ICD9CM | 148.8 | Malignant neoplasm of other specified sites of hypopharynx |
| ICD9CM | 148.9 | Malignant neoplasm of hypopharynx, unspecified site |
| ICD9CM | 149.0 | Malignant neoplasm of pharynx, unspecified |
| ICD9CM | 149.1 | Malignant neoplasm of waldeyer's ring |
| ICD9CM | 149.8 | Malignant neoplasm of other sites within the lip and oral cavity |
| ICD9CM | 149.9 | Malignant neoplasm of ill-defined sites within the lip and oral cavity |
| ICD9CM | 150.0 | Malignant neoplasm of cervical esophagus |
| ICD9CM | 150.1 | Malignant neoplasm of thoracic esophagus |
| ICD9CM | 150.2 | Malignant neoplasm of abdominal esophagus |
| ICD9CM | 150.3 | Malignant neoplasm of upper third of esophagus |
| ICD9CM | 150.4 | Malignant neoplasm of middle third of esophagus |
| ICD9CM | 150.5 | Malignant neoplasm of lower third of esophagus |
| ICD9CM | 150.8 | Malignant neoplasm of other specified part of esophagus |
| ICD9CM | 150.9 | Malignant neoplasm of esophagus, unspecified site |
| ICD9CM | 151.0 | Malignant neoplasm of cardia |
| ICD9CM | 151.1 | Malignant neoplasm of pylorus |
| ICD9CM | 151.2 | Malignant neoplasm of pyloric antrum |

Continued on next page

**TableS24 – continued from previous page**

| CodeSystem | ConceptCode | ConceptName |
|---|---|---|
| ICD9CM | 151.3 | Malignant neoplasm of fundus of stomach |
| ICD9CM | 151.4 | Malignant neoplasm of body of stomach |
| ICD9CM | 151.5 | Malignant neoplasm of lesser curvature of stomach, unspecified |
| ICD9CM | 151.6 | Malignant neoplasm of greater curvature of stomach, unspecified |
| ICD9CM | 151.8 | Malignant neoplasm of other specified sites of stomach |
| ICD9CM | 151.9 | Malignant neoplasm of stomach, unspecified site |
| ICD9CM | 152.0 | Malignant neoplasm of duodenum |
| ICD9CM | 152.1 | Malignant neoplasm of jejunum |
| ICD9CM | 152.2 | Malignant neoplasm of ileum |
| ICD9CM | 152.3 | Malignant neoplasm of Meckel's diverticulum |
| ICD9CM | 152.8 | Malignant neoplasm of other specified sites of small intestine |
| ICD9CM | 152.9 | Malignant neoplasm of small intestine, unspecified site |
| ICD9CM | 153.0 | Malignant neoplasm of hepatic flexure |
| ICD9CM | 153.1 | Malignant neoplasm of transverse colon |
| ICD9CM | 153.2 | Malignant neoplasm of descending colon |
| ICD9CM | 153.3 | Malignant neoplasm of sigmoid colon |
| ICD9CM | 153.4 | Malignant neoplasm of cecum |
| ICD9CM | 153.5 | Malignant neoplasm of appendix vermiformis |
| ICD9CM | 153.6 | Malignant neoplasm of ascending colon |
| ICD9CM | 153.7 | Malignant neoplasm of splenic flexure |
| ICD9CM | 153.8 | Malignant neoplasm of other specified sites of large intestine |
| ICD9CM | 153.9 | Malignant neoplasm of colon, unspecified site |
| ICD9CM | 154.0 | Malignant neoplasm of rectosigmoid junction |
| ICD9CM | 154.1 | Malignant neoplasm of rectum |
| ICD9CM | 154.8 | Malignant neoplasm of other sites of rectum, rectosigmoid junction, and anus |
| ICD9CM | 155.0 | Malignant neoplasm of liver, primary |
| ICD9CM | 155.1 | Malignant neoplasm of intrahepatic bile ducts |
| ICD9CM | 155.2 | Malignant neoplasm of liver, not specified as primary or secondary |
| ICD9CM | 156.0 | Malignant neoplasm of gallbladder |
| ICD9CM | 156.1 | Malignant neoplasm of extrahepatic bile ducts |
| ICD9CM | 156.2 | Malignant neoplasm of ampulla of vater |
| ICD9CM | 156.8 | Malignant neoplasm of other specified sites of gallbladder and extrahepatic bile ducts |
| ICD9CM | 156.9 | Malignant neoplasm of biliary tract, part unspecified site |
| ICD9CM | 157.0 | Malignant neoplasm of head of pancreas |
| ICD9CM | 157.1 | Malignant neoplasm of body of pancreas |
| ICD9CM | 157.2 | Malignant neoplasm of tail of pancreas |
| ICD9CM | 157.3 | Malignant neoplasm of pancreatic duct |
| ICD9CM | 157.4 | Malignant neoplasm of islets of langerhans |
| ICD9CM | 157.8 | Malignant neoplasm of other specified sites of pancreas |
| ICD9CM | 157.9 | Malignant neoplasm of pancreas, part unspecified |
| ICD9CM | 158.0 | Malignant neoplasm of retroperitoneum |
| ICD9CM | 158.8 | Malignant neoplasm of specified parts of peritoneum |
| ICD9CM | 158.9 | Malignant neoplasm of peritoneum, unspecified |
| ICD9CM | 159.0 | Malignant neoplasm of intestinal tract, part unspecified |

Continued on next page

**TableS24 – continued from previous page**

| CodeSystem | ConceptCode | ConceptName |
|---|---|---|
| ICD9CM | 159.1 | Malignant neoplasm of spleen, not elsewhere classified |
| ICD9CM | 159.8 | Malignant neoplasm of other sites of digestive system and intra-abdominal organs |
| ICD9CM | 159.9 | Malignant neoplasm of ill-defined sites within the digestive organs and peritoneum |
| ICD9CM | 160.0 | Malignant neoplasm of nasal cavities |
| ICD9CM | 160.1 | Malignant neoplasm of auditory tube, middle ear, and mastoid air cells |
| ICD9CM | 160.2 | Malignant neoplasm of maxillary sinus |
| ICD9CM | 160.3 | Malignant neoplasm of ethmoidal sinus |
| ICD9CM | 160.4 | Malignant neoplasm of frontal sinus |
| ICD9CM | 160.5 | Malignant neoplasm of sphenoidal sinus |
| ICD9CM | 160.8 | Malignant neoplasm of other accessory sinuses |
| ICD9CM | 160.9 | Malignant neoplasm of accessory sinus, unspecified |
| ICD9CM | 161.0 | Malignant neoplasm of glottis |
| ICD9CM | 161.1 | Malignant neoplasm of supraglottis |
| ICD9CM | 161.2 | Malignant neoplasm of subglottis |
| ICD9CM | 161.3 | Malignant neoplasm of laryngeal cartilages |
| ICD9CM | 161.8 | Malignant neoplasm of other specified sites of larynx |
| ICD9CM | 161.9 | Malignant neoplasm of larynx, unspecified |
| ICD9CM | 162.0 | Malignant neoplasm of trachea |
| ICD9CM | 162.2 | Malignant neoplasm of main bronchus |
| ICD9CM | 162.3 | Malignant neoplasm of upper lobe, bronchus or lung |
| ICD9CM | 162.4 | Malignant neoplasm of middle lobe, bronchus or lung |
| ICD9CM | 162.5 | Malignant neoplasm of lower lobe, bronchus or lung |
| ICD9CM | 162.8 | Malignant neoplasm of other parts of bronchus or lung |
| ICD9CM | 162.9 | Malignant neoplasm of bronchus and lung, unspecified |
| ICD9CM | 163.0 | Malignant neoplasm of parietal pleura |
| ICD9CM | 163.1 | Malignant neoplasm of visceral pleura |
| ICD9CM | 163.8 | Malignant neoplasm of other specified sites of pleura |
| ICD9CM | 163.9 | Malignant neoplasm of pleura, unspecified |
| ICD9CM | 164.0 | Malignant neoplasm of thymus |
| ICD9CM | 164.1 | Malignant neoplasm of heart |
| ICD9CM | 164.2 | Malignant neoplasm of anterior mediastinum |
| ICD9CM | 164.3 | Malignant neoplasm of posterior mediastinum |
| ICD9CM | 164.8 | Malignant neoplasm of other parts of mediastinum |
| ICD9CM | 164.9 | Malignant neoplasm of mediastinum, part unspecified |
| ICD9CM | 165.0 | Malignant neoplasm of upper respiratory tract, part unspecified |
| ICD9CM | 165.8 | Malignant neoplasm of other sites within the respiratory system and intrathoracic organs |
| ICD9CM | 165.9 | Malignant neoplasm of ill-defined sites within the respiratory system |
| ICD9CM | 170.0 | Malignant neoplasm of bones of skull and face, except mandible |
| ICD9CM | 170.1 | Malignant neoplasm of mandible |
| ICD9CM | 170.2 | Malignant neoplasm of vertebral column, excluding sacrum and coccyx |
| ICD9CM | 170.3 | Malignant neoplasm of ribs, sternum, and clavicle |

Continued on next page

**TableS24 – continued from previous page**

| CodeSystem | ConceptCode | ConceptName |
|---|---|---|
| ICD9CM | 170.4 | Malignant neoplasm of scapula and long bones of upper limb |
| ICD9CM | 170.5 | Malignant neoplasm of short bones of upper limb |
| ICD9CM | 170.6 | Malignant neoplasm of pelvic bones, sacrum, and coccyx |
| ICD9CM | 170.7 | Malignant neoplasm of long bones of lower limb |
| ICD9CM | 170.8 | Malignant neoplasm of short bones of lower limb |
| ICD9CM | 170.9 | Malignant neoplasm of bone and articular cartilage, site unspecified |
| ICD9CM | 171.0 | Malignant neoplasm of connective and other soft tissue of head, face, and neck |
| ICD9CM | 171.2 | Malignant neoplasm of connective and other soft tissue of upper limb, including shoulder |
| ICD9CM | 171.3 | Malignant neoplasm of connective and other soft tissue of lower limb, including hip |
| ICD9CM | 171.4 | Malignant neoplasm of connective and other soft tissue of thorax |
| ICD9CM | 171.5 | Malignant neoplasm of connective and other soft tissue of abdomen |
| ICD9CM | 171.6 | Malignant neoplasm of connective and other soft tissue of pelvis |
| ICD9CM | 171.7 | Malignant neoplasm of connective and other soft tissue of trunk, unspecified |
| ICD9CM | 171.8 | Malignant neoplasm of other specified sites of connective and other soft tissue |
| ICD9CM | 171.9 | Malignant neoplasm of connective and other soft tissue, site unspecified |
| ICD9CM | 172.0 | Malignant melanoma of skin of lip |
| ICD9CM | 172.1 | Malignant melanoma of skin of eyelid, including canthus |
| ICD9CM | 172.2 | Malignant melanoma of skin of ear and external auditory canal |
| ICD9CM | 172.3 | Malignant melanoma of skin of other and unspecified parts of face |
| ICD9CM | 172.4 | Malignant melanoma of skin of scalp and neck |
| ICD9CM | 172.5 | Malignant melanoma of skin of trunk, except scrotum |
| ICD9CM | 172.6 | Malignant melanoma of skin of upper limb, including shoulder |
| ICD9CM | 172.7 | Malignant melanoma of skin of lower limb, including hip |
| ICD9CM | 172.8 | Malignant melanoma of other specified sites of skin |
| ICD9CM | 172.9 | Melanoma of skin, site unspecified |
| ICD9CM | 173.00 | Unspecified malignant neoplasm of skin of lip |
| ICD9CM | 173.01 | Basal cell carcinoma of skin of lip |
| ICD9CM | 173.02 | Squamous cell carcinoma of skin of lip |
| ICD9CM | 173.09 | Other specified malignant neoplasm of skin of lip |
| ICD9CM | 173.10 | Unspecified malignant neoplasm of eyelid, including canthus |
| ICD9CM | 173.11 | Basal cell carcinoma of eyelid, including canthus |
| ICD9CM | 173.12 | Squamous cell carcinoma of eyelid, including canthus |
| ICD9CM | 173.19 | Other specified malignant neoplasm of eyelid, including canthus |
| ICD9CM | 173.20 | Unspecified malignant neoplasm of skin of ear and external auditory canal |
| ICD9CM | 173.21 | Basal cell carcinoma of skin of ear and external auditory canal |
| ICD9CM | 173.22 | Squamous cell carcinoma of skin of ear and external auditory canal |

Continued on next page

**TableS24 – continued from previous page**

| CodeSystem | ConceptCode | ConceptName |
|---|---|---|
| ICD9CM | 173.29 | Other specified malignant neoplasm of skin of ear and external auditory canal |
| ICD9CM | 173.30 | Unspecified malignant neoplasm of skin of other and unspecified parts of face |
| ICD9CM | 173.31 | Basal cell carcinoma of skin of other and unspecified parts of face |
| ICD9CM | 173.32 | Squamous cell carcinoma of skin of other and unspecified parts of face |
| ICD9CM | 173.39 | Other specified malignant neoplasm of skin of other and unspecified parts of face |
| ICD9CM | 173.40 | Unspecified malignant neoplasm of scalp and skin of neck |
| ICD9CM | 173.41 | Basal cell carcinoma of scalp and skin of neck |
| ICD9CM | 173.42 | Squamous cell carcinoma of scalp and skin of neck |
| ICD9CM | 173.49 | Other specified malignant neoplasm of scalp and skin of neck |
| ICD9CM | 173.50 | Unspecified malignant neoplasm of skin of trunk, except scrotum |
| ICD9CM | 173.51 | Basal cell carcinoma of skin of trunk, except scrotum |
| ICD9CM | 173.52 | Squamous cell carcinoma of skin of trunk, except scrotum |
| ICD9CM | 173.59 | Other specified malignant neoplasm of skin of trunk, except scrotum |
| ICD9CM | 173.60 | Unspecified malignant neoplasm of skin of upper limb, including shoulder |
| ICD9CM | 173.61 | Basal cell carcinoma of skin of upper limb, including shoulder |
| ICD9CM | 173.62 | Squamous cell carcinoma of skin of upper limb, including shoulder |
| ICD9CM | 173.69 | Other specified malignant neoplasm of skin of upper limb, including shoulder |
| ICD9CM | 173.70 | Unspecified malignant neoplasm of skin of lower limb, including hip |
| ICD9CM | 173.71 | Basal cell carcinoma of skin of lower limb, including hip |
| ICD9CM | 173.72 | Squamous cell carcinoma of skin of lower limb, including hip |
| ICD9CM | 173.79 | Other specified malignant neoplasm of skin of lower limb, including hip |
| ICD9CM | 173.80 | Unspecified malignant neoplasm of other specified sites of skin |
| ICD9CM | 173.81 | Basal cell carcinoma of other specified sites of skin |
| ICD9CM | 173.82 | Squamous cell carcinoma of other specified sites of skin |
| ICD9CM | 173.89 | Other specified malignant neoplasm of other specified sites of skin |
| ICD9CM | 173.90 | Unspecified malignant neoplasm of skin, site unspecified |
| ICD9CM | 173.91 | Basal cell carcinoma of skin, site unspecified |
| ICD9CM | 173.92 | Squamous cell carcinoma of skin, site unspecified |
| ICD9CM | 173.99 | Other specified malignant neoplasm of skin, site unspecified |
| ICD9CM | 174.0 | Malignant neoplasm of nipple and areola of female breast |
| ICD9CM | 174.1 | Malignant neoplasm of central portion of female breast |
| ICD9CM | 174.2 | Malignant neoplasm of upper-inner quadrant of female breast |
| ICD9CM | 174.3 | Malignant neoplasm of lower-inner quadrant of female breast |
| ICD9CM | 174.4 | Malignant neoplasm of upper-outer quadrant of female breast |
| ICD9CM | 174.5 | Malignant neoplasm of lower-outer quadrant of female breast |
| ICD9CM | 174.6 | Malignant neoplasm of axillary tail of female breast |

Continued on next page

**TableS24 – continued from previous page**

| CodeSystem | ConceptCode | ConceptName |
|---|---|---|
| ICD9CM | 174.8 | Malignant neoplasm of other specified sites of female breast |
| ICD9CM | 174.9 | Malignant neoplasm of breast (female), unspecified |
| ICD9CM | 175.0 | Malignant neoplasm of nipple and areola of male breast |
| ICD9CM | 175.9 | Malignant neoplasm of other and unspecified sites of male breast |
| ICD9CM | 176.0 | Kaposi's sarcoma, skin |
| ICD9CM | 176.1 | Kaposi's sarcoma, soft tissue |
| ICD9CM | 176.2 | Kaposi's sarcoma, palate |
| ICD9CM | 176.3 | Kaposi's sarcoma, gastrointestinal sites |
| ICD9CM | 176.4 | Kaposi's sarcoma, lung |
| ICD9CM | 176.5 | Kaposi's sarcoma, lymph nodes |
| ICD9CM | 176.8 | Kaposi's sarcoma, other specified sites |
| ICD9CM | 176.9 | Kaposi's sarcoma, unspecified site |
| ICD9CM | 179 | Malignant neoplasm of uterus, part unspecified |
| ICD9CM | 180.0 | Malignant neoplasm of endocervix |
| ICD9CM | 180.1 | Malignant neoplasm of exocervix |
| ICD9CM | 180.8 | Malignant neoplasm of other specified sites of cervix |
| ICD9CM | 180.9 | Malignant neoplasm of cervix uteri, unspecified site |
| ICD9CM | 181 | Malignant neoplasm of placenta |
| ICD9CM | 182.0 | Malignant neoplasm of corpus uteri, except isthmus |
| ICD9CM | 182.1 | Malignant neoplasm of isthmus |
| ICD9CM | 182.8 | Malignant neoplasm of other specified sites of body of uterus |
| ICD9CM | 183.0 | Malignant neoplasm of ovary |
| ICD9CM | 183.2 | Malignant neoplasm of fallopian tube |
| ICD9CM | 183.3 | Malignant neoplasm of broad ligament of uterus |
| ICD9CM | 183.4 | Malignant neoplasm of parametrium |
| ICD9CM | 183.5 | Malignant neoplasm of round ligament of uterus |
| ICD9CM | 183.8 | Malignant neoplasm of other specified sites of uterine adnexa |
| ICD9CM | 183.9 | Malignant neoplasm of uterine adnexa, unspecified site |
| ICD9CM | 184.0 | Malignant neoplasm of vagina |
| ICD9CM | 184.1 | Malignant neoplasm of labia majora |
| ICD9CM | 184.2 | Malignant neoplasm of labia minora |
| ICD9CM | 184.3 | Malignant neoplasm of clitoris |
| ICD9CM | 184.4 | Malignant neoplasm of vulva, unspecified site |
| ICD9CM | 184.8 | Malignant neoplasm of other specified sites of female genital organs |
| ICD9CM | 184.9 | Malignant neoplasm of female genital organ, site unspecified |
| ICD9CM | 185 | Malignant neoplasm of prostate |
| ICD9CM | 186.0 | Malignant neoplasm of undescended testis |
| ICD9CM | 186.9 | Malignant neoplasm of other and unspecified testis |
| ICD9CM | 187.1 | Malignant neoplasm of prepuce |
| ICD9CM | 187.2 | Malignant neoplasm of glans penis |
| ICD9CM | 187.3 | Malignant neoplasm of body of penis |
| ICD9CM | 187.4 | Malignant neoplasm of penis, part unspecified |
| ICD9CM | 187.5 | Malignant neoplasm of epididymis |
| ICD9CM | 187.6 | Malignant neoplasm of spermatic cord |
| ICD9CM | 187.7 | Malignant neoplasm of scrotum |
| ICD9CM | 187.8 | Malignant neoplasm of other specified sites of male genital organs |

Continued on next page

**TableS24 – continued from previous page**

| CodeSystem | ConceptCode | ConceptName |
|---|---|---|
| ICD9CM | 187.9 | Malignant neoplasm of male genital organ, site unspecified |
| ICD9CM | 188.0 | Malignant neoplasm of trigone of urinary bladder |
| ICD9CM | 188.1 | Malignant neoplasm of dome of urinary bladder |
| ICD9CM | 188.2 | Malignant neoplasm of lateral wall of urinary bladder |
| ICD9CM | 188.3 | Malignant neoplasm of anterior wall of urinary bladder |
| ICD9CM | 188.4 | Malignant neoplasm of posterior wall of urinary bladder |
| ICD9CM | 188.5 | Malignant neoplasm of bladder neck |
| ICD9CM | 188.6 | Malignant neoplasm of ureteric orifice |
| ICD9CM | 188.7 | Malignant neoplasm of urachus |
| ICD9CM | 188.8 | Malignant neoplasm of other specified sites of bladder |
| ICD9CM | 188.9 | Malignant neoplasm of bladder, part unspecified |
| ICD9CM | 189.0 | Malignant neoplasm of kidney, except pelvis |
| ICD9CM | 189.1 | Malignant neoplasm of renal pelvis |
| ICD9CM | 189.2 | Malignant neoplasm of ureter |
| ICD9CM | 189.3 | Malignant neoplasm of urethra |
| ICD9CM | 189.4 | Malignant neoplasm of paraurethral glands |
| ICD9CM | 189.8 | Malignant neoplasm of other specified sites of urinary organs |
| ICD9CM | 189.9 | Malignant neoplasm of urinary organ, site unspecified |
| ICD9CM | 190.0 | Malignant neoplasm of eyeball, except conjunctiva, cornea, retina, and choroid |
| ICD9CM | 190.1 | Malignant neoplasm of orbit |
| ICD9CM | 190.2 | Malignant neoplasm of lacrimal gland |
| ICD9CM | 190.3 | Malignant neoplasm of conjunctiva |
| ICD9CM | 190.4 | Malignant neoplasm of cornea |
| ICD9CM | 190.5 | Malignant neoplasm of retina |
| ICD9CM | 190.6 | Malignant neoplasm of choroid |
| ICD9CM | 190.7 | Malignant neoplasm of lacrimal duct |
| ICD9CM | 190.8 | Malignant neoplasm of other specified sites of eye |
| ICD9CM | 190.9 | Malignant neoplasm of eye, part unspecified |
| ICD9CM | 191.0 | Malignant neoplasm of cerebrum, except lobes and ventricles |
| ICD9CM | 191.1 | Malignant neoplasm of frontal lobe |
| ICD9CM | 191.2 | Malignant neoplasm of temporal lobe |
| ICD9CM | 191.3 | Malignant neoplasm of parietal lobe |
| ICD9CM | 191.4 | Malignant neoplasm of occipital lobe |
| ICD9CM | 191.5 | Malignant neoplasm of ventricles |
| ICD9CM | 191.6 | Malignant neoplasm of cerebellum nos |
| ICD9CM | 191.7 | Malignant neoplasm of brain stem |
| ICD9CM | 191.8 | Malignant neoplasm of other parts of brain |
| ICD9CM | 191.9 | Malignant neoplasm of brain, unspecified |
| ICD9CM | 192.0 | Malignant neoplasm of cranial nerves |
| ICD9CM | 192.1 | Malignant neoplasm of cerebral meninges |
| ICD9CM | 192.2 | Malignant neoplasm of spinal cord |
| ICD9CM | 192.3 | Malignant neoplasm of spinal meninges |
| ICD9CM | 192.8 | Malignant neoplasm of other specified sites of nervous system |
| ICD9CM | 192.9 | Malignant neoplasm of nervous system, part unspecified |
| ICD9CM | 194.0 | Malignant neoplasm of adrenal gland |
| ICD9CM | 194.1 | Malignant neoplasm of parathyroid gland |

Continued on next page

**TableS24 – continued from previous page**

| CodeSystem | ConceptCode | ConceptName |
|---|---|---|
| ICD9CM | 194.3 | Malignant neoplasm of pituitary gland and craniopharyngeal duct |
| ICD9CM | 194.4 | Malignant neoplasm of pineal gland |
| ICD9CM | 194.5 | Malignant neoplasm of carotid body |
| ICD9CM | 194.6 | Malignant neoplasm of aortic body and other paraganglia |
| ICD9CM | 194.8 | Malignant neoplasm of other endocrine glands and related structures |
| ICD9CM | 194.9 | Malignant neoplasm of endocrine gland, site unspecified |
| ICD9CM | 195.0 | Malignant neoplasm of head, face, and neck |
| ICD9CM | 195.1 | Malignant neoplasm of thorax |
| ICD9CM | 195.2 | Malignant neoplasm of abdomen |
| ICD9CM | 195.3 | Malignant neoplasm of pelvis |
| ICD9CM | 195.4 | Malignant neoplasm of upper limb |
| ICD9CM | 195.5 | Malignant neoplasm of lower limb |
| ICD9CM | 195.8 | Malignant neoplasm of other specified sites |
| ICD9CM | 196.0 | Secondary and unspecified malignant neoplasm of lymph nodes of head, face, and neck |
| ICD9CM | 196.1 | Secondary and unspecified malignant neoplasm of intrathoracic lymph nodes |
| ICD9CM | 196.2 | Secondary and unspecified malignant neoplasm of intra-abdominal lymph nodes |
| ICD9CM | 196.3 | Secondary and unspecified malignant neoplasm of lymph nodes of axilla and upper limb |
| ICD9CM | 196.5 | Secondary and unspecified malignant neoplasm of lymph nodes of inguinal region and lower limb |
| ICD9CM | 196.6 | Secondary and unspecified malignant neoplasm of intrapelvic lymph nodes |
| ICD9CM | 196.8 | Secondary and unspecified malignant neoplasm of lymph nodes of multiple sites |
| ICD9CM | 196.9 | Secondary and unspecified malignant neoplasm of lymph nodes, site unspecified |
| ICD9CM | 197.5 | Secondary malignant neoplasm of large intestine and rectum |
| ICD9CM | 200.00 | Reticulosarcoma, unspecified site, extranodal and solid organ sites |
| ICD9CM | 200.01 | Reticulosarcoma, lymph nodes of head, face, and neck |
| ICD9CM | 200.02 | Reticulosarcoma, intrathoracic lymph nodes |
| ICD9CM | 200.03 | Reticulosarcoma, intra-abdominal lymph nodes |
| ICD9CM | 200.04 | Reticulosarcoma, lymph nodes of axilla and upper limb |
| ICD9CM | 200.05 | Reticulosarcoma, lymph nodes of inguinal region and lower limb |
| ICD9CM | 200.06 | Reticulosarcoma, intrapelvic lymph nodes |
| ICD9CM | 200.07 | Reticulosarcoma, spleen |
| ICD9CM | 200.08 | Reticulosarcoma, lymph nodes of multiple sites |
| ICD9CM | 200.10 | Lymphosarcoma, unspecified site, extranodal and solid organ sites |
| ICD9CM | 200.11 | Lymphosarcoma, lymph nodes of head, face, and neck |
| ICD9CM | 200.12 | Lymphosarcoma, intrathoracic lymph nodes |
| ICD9CM | 200.13 | Lymphosarcoma, intra-abdominal lymph nodes |

Continued on next page

**TableS24 – continued from previous page**

| CodeSystem | ConceptCode | ConceptName |
|---|---|---|
| ICD9CM | 200.14 | Lymphosarcoma, lymph nodes of axilla and upper limb |
| ICD9CM | 200.15 | Lymphosarcoma, lymph nodes of inguinal region and lower limb |
| ICD9CM | 200.16 | Lymphosarcoma, intrapelvic lymph nodes |
| ICD9CM | 200.17 | Lymphosarcoma, spleen |
| ICD9CM | 200.18 | Lymphosarcoma, lymph nodes of multiple sites |
| ICD9CM | 200.20 | Burkitt's tumor or lymphoma, unspecified site, extranodal and solid organ sites |
| ICD9CM | 200.21 | Burkitt's tumor or lymphoma, lymph nodes of head, face, and neck |
| ICD9CM | 200.22 | Burkitt's tumor or lymphoma, intrathoracic lymph nodes |
| ICD9CM | 200.23 | Burkitt's tumor or lymphoma, intra-abdominal lymph nodes |
| ICD9CM | 200.24 | Burkitt's tumor or lymphoma, lymph nodes of axilla and upper limb |
| ICD9CM | 200.25 | Burkitt's tumor or lymphoma, lymph nodes of inguinal region and lower limb |
| ICD9CM | 200.26 | Burkitt's tumor or lymphoma, intrapelvic lymph nodes |
| ICD9CM | 200.27 | Burkitt's tumor or lymphoma, spleen |
| ICD9CM | 200.28 | Burkitt's tumor or lymphoma, lymph nodes of multiple sites |
| ICD9CM | 200.30 | Marginal zone lymphoma, unspecified site, extranodal and solid organ sites |
| ICD9CM | 200.31 | Marginal zone lymphoma, lymph nodes of head, face, and neck |
| ICD9CM | 200.32 | Marginal zone lymphoma, intrathoracic lymph nodes |
| ICD9CM | 200.33 | Marginal zone lymphoma, intraabdominal lymph nodes |
| ICD9CM | 200.34 | Marginal zone lymphoma, lymph nodes of axilla and upper limb |
| ICD9CM | 200.35 | Marginal zone lymphoma, lymph nodes of inguinal region and lower limb |
| ICD9CM | 200.36 | Marginal zone lymphoma, intrapelvic lymph nodes |
| ICD9CM | 200.37 | Marginal zone lymphoma, spleen |
| ICD9CM | 200.38 | Marginal zone lymphoma, lymph nodes of multiple sites |
| ICD9CM | 200.40 | Mantle cell lymphoma, unspecified site, extranodal and solid organ sites |
| ICD9CM | 200.41 | Mantle cell lymphoma, lymph nodes of head, face, and neck |
| ICD9CM | 200.42 | Mantle cell lymphoma, intrathoracic lymph nodes |
| ICD9CM | 200.43 | Mantle cell lymphoma, intra-abdominal lymph nodes |
| ICD9CM | 200.44 | Mantle cell lymphoma, lymph nodes of axilla and upper limb |
| ICD9CM | 200.45 | Mantle cell lymphoma, lymph nodes of inguinal region and lower limb |
| ICD9CM | 200.46 | Mantle cell lymphoma, intrapelvic lymph nodes |
| ICD9CM | 200.47 | Mantle cell lymphoma, spleen |
| ICD9CM | 200.48 | Mantle cell lymphoma, lymph nodes of multiple sites |
| ICD9CM | 200.50 | Primary central nervous system lymphoma, unspecified site, extranodal and solid organ sites |
| ICD9CM | 200.51 | Primary central nervous system lymphoma, lymph nodes of head, face, and neck |
| ICD9CM | 200.52 | Primary central nervous system lymphoma, intrathoracic lymph nodes |

Continued on next page

**TableS24 – continued from previous page**

| CodeSystem | ConceptCode | ConceptName |
|---|---|---|
| ICD9CM | 200.53 | Primary central nervous system lymphoma, intra-abdominal lymph nodes |
| ICD9CM | 200.54 | Primary central nervous system lymphoma, lymph nodes of axilla and upper limb |
| ICD9CM | 200.55 | Primary central nervous system lymphoma, lymph nodes of inguinal region and lower limb |
| ICD9CM | 200.56 | Primary central nervous system lymphoma, intrapelvic lymph nodes |
| ICD9CM | 200.57 | Primary central nervous system lymphoma, spleen |
| ICD9CM | 200.58 | Primary central nervous system lymphoma, lymph nodes of multiple sites |
| ICD9CM | 200.60 | Anaplastic large cell lymphoma, unspecified site, extranodal and solid organ sites |
| ICD9CM | 200.61 | Anaplastic large cell lymphoma, lymph nodes of head, face, and neck |
| ICD9CM | 200.62 | Anaplastic large cell lymphoma, intrathoracic lymph nodes |
| ICD9CM | 200.63 | Anaplastic large cell lymphoma, intra-abdominal lymph nodes |
| ICD9CM | 200.64 | Anaplastic large cell lymphoma, lymph nodes of axilla and upper limb |
| ICD9CM | 200.65 | Anaplastic large cell lymphoma, lymph nodes of inguinal region and lower limb |
| ICD9CM | 200.66 | Anaplastic large cell lymphoma, intrapelvic lymph nodes |
| ICD9CM | 200.67 | Anaplastic large cell lymphoma, spleen |
| ICD9CM | 200.68 | Anaplastic large cell lymphoma, lymph nodes of multiple sites |
| ICD9CM | 200.70 | Large cell lymphoma, unspecified site, extranodal and solid organ sites |
| ICD9CM | 200.71 | Large cell lymphoma, lymph nodes of head, face, and neck |
| ICD9CM | 200.72 | Large cell lymphoma, intrathoracic lymph nodes |
| ICD9CM | 200.73 | Large cell lymphoma, intra-abdominal lymph nodes |
| ICD9CM | 200.74 | Large cell lymphoma, lymph nodes of axilla and upper limb |
| ICD9CM | 200.75 | Large cell lymphoma, lymph nodes of inguinal region and lower limb |
| ICD9CM | 200.76 | Large cell lymphoma, intrapelvic lymph nodes |
| ICD9CM | 200.77 | Large cell lymphoma, spleen |
| ICD9CM | 200.78 | Large cell lymphoma, lymph nodes of multiple sites |
| ICD9CM | 200.80 | Other named variants of lymphosarcoma and reticulosarcoma, unspecified site, extranodal and solid organ sites |
| ICD9CM | 200.81 | Other named variants of lymphosarcoma and reticulosarcoma, lymph nodes of head, face, and neck |
| ICD9CM | 200.82 | Other named variants of lymphosarcoma and reticulosarcoma,intrathoracic lymph nodes |
| ICD9CM | 200.83 | Other named variants of lymphosarcoma and reticulosarcoma, intra-abdominal lymph nodes |
| ICD9CM | 200.84 | Other named variants of lymphosarcoma and reticulosarcoma, lymph nodes of axilla and upper limb |
| ICD9CM | 200.85 | Other named variants of lymphosarcoma and reticulosarcoma, lymph nodes of inguinal region and lower limb |

Continued on next page

**TableS24 – continued from previous page**

| CodeSystem | ConceptCode | ConceptName |
|---|---|---|
| ICD9CM | 200.86 | Other named variants of lymphosarcoma and reticulosarcoma, intrapelvic lymph nodes |
| ICD9CM | 200.87 | Other named variants of lymphosarcoma and reticulosarcoma, spleen |
| ICD9CM | 200.88 | Other named variants of lymphosarcoma and reticulosarcoma, lymph nodes of multiple sites |
| ICD9CM | 201.00 | Hodgkin's paragranuloma, unspecified site, extranodal and solid organ sites |
| ICD9CM | 201.01 | Hodgkin's paragranuloma, lymph nodes of head, face, and neck |
| ICD9CM | 201.02 | Hodgkin's paragranuloma, intrathoracic lymph nodes |
| ICD9CM | 201.03 | Hodgkin's paragranuloma, intra-abdominal lymph nodes |
| ICD9CM | 201.04 | Hodgkin's paragranuloma, lymph nodes of axilla and upper limb |
| ICD9CM | 201.05 | Hodgkin's paragranuloma, lymph nodes of inguinal region and lower limb |
| ICD9CM | 201.06 | Hodgkin's paragranuloma, intrapelvic lymph nodes |
| ICD9CM | 201.07 | Hodgkin's paragranuloma, spleen |
| ICD9CM | 201.08 | Hodgkin's paragranuloma, lymph nodes of multiple sites |
| ICD9CM | 201.10 | Hodgkin's granuloma, unspecified site, extranodal and solid organ sites |
| ICD9CM | 201.11 | Hodgkin's granuloma, lymph nodes of head, face, and neck |
| ICD9CM | 201.12 | Hodgkin's granuloma, intrathoracic lymph nodes |
| ICD9CM | 201.13 | Hodgkin's granuloma, intra-abdominal lymph nodes |
| ICD9CM | 201.14 | Hodgkin's granuloma, lymph nodes of axilla and upper limb |
| ICD9CM | 201.15 | Hodgkin's granuloma, lymph nodes of inguinal region and lower limb |
| ICD9CM | 201.16 | Hodgkin's granuloma, intrapelvic lymph nodes |
| ICD9CM | 201.17 | Hodgkin's granuloma, spleen |
| ICD9CM | 201.18 | Hodgkin's granuloma, lymph nodes of multiple sites |
| ICD9CM | 201.20 | Hodgkin's sarcoma, unspecified site, extranodal and solid organ sites |
| ICD9CM | 201.21 | Hodgkin's sarcoma, lymph nodes of head, face, and neck |
| ICD9CM | 201.22 | Hodgkin's sarcoma, intrathoracic lymph nodes |
| ICD9CM | 201.23 | Hodgkin's sarcoma, intra-abdominal lymph nodes |
| ICD9CM | 201.24 | Hodgkin's sarcoma, lymph nodes of axilla and upper limb |
| ICD9CM | 201.25 | Hodgkin's sarcoma, lymph nodes of inguinal region and lower limb |
| ICD9CM | 201.26 | Hodgkin's sarcoma, intrapelvic lymph nodes |
| ICD9CM | 201.27 | Hodgkin's sarcoma, spleen |
| ICD9CM | 201.28 | Hodgkin's sarcoma, lymph nodes of multiple sites |
| ICD9CM | 201.40 | Hodgkin's disease, lymphocytic-histiocytic predominance, unspecified site, extranodal and solid organ sites |
| ICD9CM | 201.41 | Hodgkin's disease, lymphocytic-histiocytic predominance, lymph nodes of head, face, and neck |
| ICD9CM | 201.42 | Hodgkin's disease, lymphocytic-histiocytic predominance, intrathoracic lymph nodes |
| ICD9CM | 201.43 | Hodgkin's disease, lymphocytic-histiocytic predominance, intra-abdominal lymph nodes |

Continued on next page

**TableS24 – continued from previous page**

| CodeSystem | ConceptCode | ConceptName |
|---|---|---|
| ICD9CM | 201.44 | Hodgkin's disease, lymphocytic-histiocytic predominance, lymph nodes of axilla and upper limb |
| ICD9CM | 201.45 | Hodgkin's disease, lymphocytic-histiocytic predominance, lymph nodes of inguinal region and lower limb |
| ICD9CM | 201.46 | Hodgkin's disease, lymphocytic-histiocytic predominance, intrapelvic lymph nodes |
| ICD9CM | 201.47 | Hodgkin's disease, lymphocytic-histiocytic predominance, spleen |
| ICD9CM | 201.48 | Hodgkin's disease, lymphocytic-histiocytic predominance, lymph nodes of multiple sites |
| ICD9CM | 201.50 | Hodgkin's disease, nodular sclerosis, unspecified site, extranodal and solid organ sites |
| ICD9CM | 201.51 | Hodgkin's disease, nodular sclerosis, lymph nodes of head, face, and neck |
| ICD9CM | 201.52 | Hodgkin's disease, nodular sclerosis, intrathoracic lymph nodes |
| ICD9CM | 201.53 | Hodgkin's disease, nodular sclerosis, intra-abdominal lymph nodes |
| ICD9CM | 201.54 | Hodgkin's disease, nodular sclerosis, lymph nodes of axilla and upper limb |
| ICD9CM | 201.55 | Hodgkin's disease, nodular sclerosis, lymph nodes of inguinal region and lower limb |
| ICD9CM | 201.56 | Hodgkin's disease, nodular sclerosis, intrapelvic lymph nodes |
| ICD9CM | 201.57 | Hodgkin's disease, nodular sclerosis, spleen |
| ICD9CM | 201.58 | Hodgkin's disease, nodular sclerosis, lymph nodes of multiple sites |
| ICD9CM | 201.60 | Hodgkin's disease, mixed cellularity, unspecified site, extranodal and solid organ sites |
| ICD9CM | 201.61 | Hodgkin's disease, mixed cellularity, lymph nodes of head, face, and neck |
| ICD9CM | 201.62 | Hodgkin's disease, mixed cellularity, intrathoracic lymph nodes |
| ICD9CM | 201.63 | Hodgkin's disease, mixed cellularity, intra-abdominal lymph nodes |
| ICD9CM | 201.64 | Hodgkin's disease, mixed cellularity, lymph nodes of axilla and upper limb |
| ICD9CM | 201.65 | Hodgkin's disease, mixed cellularity, lymph nodes of inguinal region and lower limb |
| ICD9CM | 201.66 | Hodgkin's disease, mixed cellularity, intrapelvic lymph nodes |
| ICD9CM | 201.67 | Hodgkin's disease, mixed cellularity, spleen |
| ICD9CM | 201.68 | Hodgkin's disease, mixed cellularity, lymph nodes of multiple sites |
| ICD9CM | 201.70 | Hodgkin's disease, lymphocytic depletion, unspecified site, extranodal and solid organ sites |
| ICD9CM | 201.71 | Hodgkin's disease, lymphocytic depletion, lymph nodes of head, face, and neck |
| ICD9CM | 201.72 | Hodgkin's disease, lymphocytic depletion, intrathoracic lymph nodes |
| ICD9CM | 201.73 | Hodgkin's disease, lymphocytic depletion, intra-abdominal lymph nodes |

Continued on next page

**TableS24 – continued from previous page**

| CodeSystem | ConceptCode | ConceptName |
|---|---|---|
| ICD9CM | 201.74 | Hodgkin's disease, lymphocytic depletion, lymph nodes of axilla and upper limb |
| ICD9CM | 201.75 | Hodgkin's disease, lymphocytic depletion, lymph nodes of inguinal region and lower limb |
| ICD9CM | 201.76 | Hodgkin's disease, lymphocytic depletion, intrapelvic lymph nodes |
| ICD9CM | 201.77 | Hodgkin's disease, lymphocytic depletion, spleen |
| ICD9CM | 201.78 | Hodgkin's disease, lymphocytic depletion, lymph nodes of multiple sites |
| ICD9CM | 201.90 | Hodgkin's disease, unspecified type, unspecified site, extranodal and solid organ sites |
| ICD9CM | 201.91 | Hodgkin's disease, unspecified type, lymph nodes of head, face, and neck |
| ICD9CM | 201.92 | Hodgkin's disease, unspecified type, intrathoracic lymph nodes |
| ICD9CM | 201.93 | Hodgkin's disease, unspecified type, intra-abdominal lymph nodes |
| ICD9CM | 201.94 | Hodgkin's disease, unspecified type, lymph nodes of axilla and upper limb |
| ICD9CM | 201.95 | Hodgkin's disease, unspecified type, lymph nodes of inguinal region and lower limb |
| ICD9CM | 201.96 | Hodgkin's disease, unspecified type, intrapelvic lymph nodes |
| ICD9CM | 201.97 | Hodgkin's disease, unspecified type, spleen |
| ICD9CM | 201.98 | Hodgkin's disease, unspecified type, lymph nodes of multiple sites |
| ICD9CM | 202.00 | Nodular lymphoma, unspecified site, extranodal and solid organ sites |
| ICD9CM | 202.01 | Nodular lymphoma, lymph nodes of head, face, and neck |
| ICD9CM | 202.02 | Nodular lymphoma, intrathoracic lymph nodes |
| ICD9CM | 202.03 | Nodular lymphoma, intra-abdominal lymph nodes |
| ICD9CM | 202.04 | Nodular lymphoma, lymph nodes of axilla and upper limb |
| ICD9CM | 202.05 | Nodular lymphoma, lymph nodes of inguinal region and lower limb |
| ICD9CM | 202.06 | Nodular lymphoma, intrapelvic lymph nodes |
| ICD9CM | 202.07 | Nodular lymphoma, spleen |
| ICD9CM | 202.08 | Nodular lymphoma, lymph nodes of multiple sites |
| ICD9CM | 202.10 | Mycosis fungoides, unspecified site, extranodal and solid organ sites |
| ICD9CM | 202.11 | Mycosis fungoides, lymph nodes of head, face, and neck |
| ICD9CM | 202.12 | Mycosis fungoides, intrathoracic lymph nodes |
| ICD9CM | 202.13 | Mycosis fungoides, intra-abdominal lymph nodes |
| ICD9CM | 202.14 | Mycosis fungoides, lymph nodes of axilla and upper limb |
| ICD9CM | 202.15 | Mycosis fungoides, lymph nodes of inguinal region and lower limb |
| ICD9CM | 202.16 | Mycosis fungoides, intrapelvic lymph nodes |
| ICD9CM | 202.17 | Mycosis fungoides, spleen |
| ICD9CM | 202.18 | Mycosis fungoides, lymph nodes of multiple sites |
| ICD9CM | 202.20 | Sezary's disease, unspecified site, extranodal and solid organ sites |
| ICD9CM | 202.21 | Sezary's disease, lymph nodes of head, face, and neck |

Continued on next page

**TableS24 – continued from previous page**

| CodeSystem | ConceptCode | ConceptName |
|---|---|---|
| ICD9CM | 202.22 | Sezary's disease, intrathoracic lymph nodes |
| ICD9CM | 202.23 | Sezary's disease, intra-abdominal lymph nodes |
| ICD9CM | 202.24 | Sezary's disease, lymph nodes of axilla and upper limb |
| ICD9CM | 202.25 | Sezary's disease, lymph nodes of inguinal region and lower limb |
| ICD9CM | 202.26 | Sezary's disease, intrapelvic lymph nodes |
| ICD9CM | 202.27 | Sezary's disease, spleen |
| ICD9CM | 202.28 | Sezary's disease, lymph nodes of multiple sites |
| ICD9CM | 202.30 | Malignant histiocytosis, unspecified site, extranodal and solid organ sites |
| ICD9CM | 202.31 | Malignant histiocytosis, lymph nodes of head, face, and neck |
| ICD9CM | 202.32 | Malignant histiocytosis, intrathoracic lymph nodes |
| ICD9CM | 202.33 | Malignant histiocytosis, intra-abdominal lymph nodes |
| ICD9CM | 202.34 | Malignant histiocytosis, lymph nodes of axilla and upper limb |
| ICD9CM | 202.35 | Malignant histiocytosis, lymph nodes of inguinal region and lower limb |
| ICD9CM | 202.36 | Malignant histiocytosis, intrapelvic lymph nodes |
| ICD9CM | 202.37 | Malignant histiocytosis, spleen |
| ICD9CM | 202.38 | Malignant histiocytosis, lymph nodes of multiple sites |
| ICD9CM | 202.40 | Leukemic reticuloendotheliosis, unspecified site, extranodal and solid organ sites |
| ICD9CM | 202.41 | Leukemic reticuloendotheliosis, lymph nodes of head, face, and neck |
| ICD9CM | 202.42 | Leukemic reticuloendotheliosis, intrathoracic lymph nodes |
| ICD9CM | 202.43 | Leukemic reticuloendotheliosis, intra-abdominal lymph nodes |
| ICD9CM | 202.44 | Leukemic reticuloendotheliosis, lymph nodes of axilla and upper arm |
| ICD9CM | 202.45 | Leukemic reticuloendotheliosis, lymph nodes of inguinal region and lower limb |
| ICD9CM | 202.46 | Leukemic reticuloendotheliosis, intrapelvic lymph nodes |
| ICD9CM | 202.47 | Leukemic reticuloendotheliosis, spleen |
| ICD9CM | 202.48 | Leukemic reticuloendotheliosis, lymph nodes of multiple sites |
| ICD9CM | 202.50 | Letterer-siwe disease, unspecified site, extranodal and solid organ sites |
| ICD9CM | 202.51 | Letterer-siwe disease, lymph nodes of head, face, and neck |
| ICD9CM | 202.52 | Letterer-siwe disease, intrathoracic lymph nodes |
| ICD9CM | 202.53 | Letterer-siwe disease, intra-abdominal lymph nodes |
| ICD9CM | 202.54 | Letterer-siwe disease, lymph nodes of axilla and upper limb |
| ICD9CM | 202.55 | Letterer-siwe disease, lymph nodes of inguinal region and lower limb |
| ICD9CM | 202.56 | Letterer-siwe disease, intrapelvic lymph nodes |
| ICD9CM | 202.57 | Letterer-siwe disease, spleen |
| ICD9CM | 202.58 | Letterer-siwe disease, lymph nodes of multiple sites |
| ICD9CM | 202.60 | Malignant mast cell tumors, unspecified site, extranodal and solid organ sites |
| ICD9CM | 202.61 | Malignant mast cell tumors, lymph nodes of head, face, and neck |
| ICD9CM | 202.62 | Malignant mast cell tumors, intrathoracic lymph nodes |
| ICD9CM | 202.63 | Malignant mast cell tumors, intra-abdominal lymph nodes |

Continued on next page

**TableS24 – continued from previous page**

| CodeSystem | ConceptCode | ConceptName |
|---|---|---|
| ICD9CM | 202.64 | Malignant mast cell tumors, lymph nodes of axilla and upper limb |
| ICD9CM | 202.65 | Malignant mast cell tumors, lymph nodes of inguinal region and lower limb |
| ICD9CM | 202.66 | Malignant mast cell tumors, intrapelvic lymph nodes |
| ICD9CM | 202.67 | Malignant mast cell tumors, spleen |
| ICD9CM | 202.68 | Malignant mast cell tumors, lymph nodes of multiple sites |
| ICD9CM | 202.70 | Peripheral T cell lymphoma, unspecified site, extranodal and solid organ sites |
| ICD9CM | 202.71 | Peripheral T cell lymphoma, lymph nodes of head, face, and neck |
| ICD9CM | 202.72 | Peripheral T cell lymphoma, intrathoracic lymph nodes |
| ICD9CM | 202.73 | Peripheral T cell lymphoma, intra-abdominal lymph nodes |
| ICD9CM | 202.74 | Peripheral T cell lymphoma, lymph nodes of axilla and upper limb |
| ICD9CM | 202.75 | Peripheral T cell lymphoma, lymph nodes of inguinal region and lower limb |
| ICD9CM | 202.76 | Peripheral T cell lymphoma, intrapelvic lymph nodes |
| ICD9CM | 202.77 | Peripheral T cell lymphoma, spleen |
| ICD9CM | 202.78 | Peripheral T cell lymphoma, lymph nodes of multiple sites |
| ICD9CM | 202.80 | Other malignant lymphomas, unspecified site, extranodal and solid organ sites |
| ICD9CM | 202.81 | Other malignant lymphomas, lymph nodes of head, face, and neck |
| ICD9CM | 202.82 | Other malignant lymphomas, intrathoracic lymph nodes |
| ICD9CM | 202.83 | Other malignant lymphomas, intra-abdominal lymph nodes |
| ICD9CM | 202.84 | Other malignant lymphomas, lymph nodes of axilla and upper limb |
| ICD9CM | 202.85 | Other malignant lymphomas, lymph nodes of inguinal region and lower limb |
| ICD9CM | 202.86 | Other malignant lymphomas, intrapelvic lymph nodes |
| ICD9CM | 202.87 | Other malignant lymphomas, spleen |
| ICD9CM | 202.88 | Other malignant lymphomas, lymph nodes of multiple sites |
| ICD9CM | 202.90 | Other and unspecified malignant neoplasms of lymphoid and histiocytic tissue, unspecified site, extranodal and solid organ sites |
| ICD9CM | 202.91 | Other and unspecified malignant neoplasms of lymphoid and histiocytic tissue, lymph nodes of head, face, and neck |
| ICD9CM | 202.92 | Other and unspecified malignant neoplasms of lymphoid and histiocytic tissue, intrathoracic lymph nodes |
| ICD9CM | 202.93 | Other and unspecified malignant neoplasms of lymphoid and histiocytic tissue, intra-abdominal lymph nodes |
| ICD9CM | 202.94 | Other and unspecified malignant neoplasms of lymphoid and histiocytic tissue, lymph nodes of axilla and upper limb |
| ICD9CM | 202.95 | Other and unspecified malignant neoplasms of lymphoid and histiocytic tissue, lymph nodes of inguinal region and lower limb |
| ICD9CM | 202.96 | Other and unspecified malignant neoplasms of lymphoid and histiocytic tissue, intrapelvic lymph nodes |

Continued on next page

**TableS24 – continued from previous page**

| CodeSystem | ConceptCode | ConceptName |
|---|---|---|
| ICD9CM | 202.97 | Other and unspecified malignant neoplasms of lymphoid and histiocytic tissue, spleen |
| ICD9CM | 202.98 | Other and unspecified malignant neoplasms of lymphoid and histiocytic tissue, lymph nodes of multiple sites |
| ICD9CM | 203.00 | Multiple myeloma, without mention of having achieved remission |
| ICD9CM | 203.01 | Multiple myeloma, in remission |
| ICD9CM | 203.02 | Multiple myeloma, in relapse |
| ICD9CM | 203.10 | Plasma cell leukemia, without mention of having achieved remission |
| ICD9CM | 203.11 | Plasma cell leukemia, in remission |
| ICD9CM | 203.12 | Plasma cell leukemia, in relapse |
| ICD9CM | 203.80 | Other immunoproliferative neoplasms, without mention of having achieved remission |
| ICD9CM | 203.81 | Other immunoproliferative neoplasms, in remission |
| ICD9CM | 203.82 | Other immunoproliferative neoplasms, in relapse |
| ICD9CM | 204.00 | Acute lymphoid leukemia, without mention of having achieved remission |
| ICD9CM | 204.01 | Acute lymphoid leukemia, in remission |
| ICD9CM | 204.10 | Chronic lymphoid leukemia, without mention of having achieved remission |
| ICD9CM | 204.11 | Chronic lymphoid leukemia, in remission |
| ICD9CM | 204.20 | Subacute lymphoid leukemia, without mention of having achieved remission |
| ICD9CM | 204.21 | Subacute lymphoid leukemia, in remission |
| ICD9CM | 204.80 | Other lymphoid leukemia, without mention of having achieved remission |
| ICD9CM | 204.81 | Other lymphoid leukemia, in remission |
| ICD9CM | 204.90 | Unspecified lymphoid leukemia, without mention of having achieved remission |
| ICD9CM | 204.91 | Unspecified lymphoid leukemia, in remission |
| ICD9CM | 205.00 | Acute myeloid leukemia, without mention of having achieved remission |
| ICD9CM | 205.01 | Acute myeloid leukemia, in remission |
| ICD9CM | 205.10 | Chronic myeloid leukemia, without mention of having achieved remission |
| ICD9CM | 205.11 | Chronic myeloid leukemia, in remission |
| ICD9CM | 205.20 | Subacute myeloid leukemia, without mention of having achieved remission |
| ICD9CM | 205.21 | Subacute myeloid leukemia,in remission |
| ICD9CM | 205.30 | Myeloid sarcoma, without mention of having achieved remission |
| ICD9CM | 205.31 | Myeloid sarcoma, in remission |
| ICD9CM | 205.80 | Other myeloid leukemia, without mention of having achieved remission |
| ICD9CM | 205.81 | Other myeloid leukemia, in remission |
| ICD9CM | 205.90 | Unspecified myeloid leukemia, without mention of having achieved remission |
| ICD9CM | 205.91 | Unspecified myeloid leukemia, in remission |

Continued on next page

**TableS24 – continued from previous page**

| CodeSystem | ConceptCode | ConceptName |
|---|---|---|
| ICD9CM | 206.00 | Acute monocytic leukemia, without mention of having achieved remission |
| ICD9CM | 206.01 | Acute monocytic leukemia,in remission |
| ICD9CM | 206.10 | Chronic monocytic leukemia, without mention of having achieved remission |
| ICD9CM | 206.11 | Chronic monocytic leukemia, in remission |
| ICD9CM | 206.20 | Subacute monocytic leukemia, without mention of having achieved remission |
| ICD9CM | 206.21 | Subacute monocytic leukemia, in remission |
| ICD9CM | 206.80 | Other monocytic leukemia, without mention of having achieved remission |
| ICD9CM | 206.81 | Other monocytic leukemia, in remission |
| ICD9CM | 206.90 | Unspecified monocytic leukemia, without mention of having achieved remission |
| ICD9CM | 206.91 | Unspecified monocytic leukemia, in remission |
| ICD9CM | 207.00 | Acute erythremia and erythroleukemia, without mention of having achieved remission |
| ICD9CM | 207.01 | Acute erythremia and erythroleukemia, in remission |
| ICD9CM | 207.10 | Chronic erythremia, without mention of having achieved remission |
| ICD9CM | 207.11 | Chronic erythremia, in remission |
| ICD9CM | 207.20 | Megakaryocytic leukemia, without mention of having achieved remission |
| ICD9CM | 207.21 | Megakaryocytic leukemia, in remission |
| ICD9CM | 207.80 | Other specified leukemia, without mention of having achieved remission |
| ICD9CM | 207.81 | Other specified leukemia, in remission |
| ICD9CM | 208.00 | Acute leukemia of unspecified cell type, without mention of having achieved remission |
| ICD9CM | 208.01 | Acute leukemia of unspecified cell type, in remission |
| ICD9CM | 208.10 | Chronic leukemia of unspecified cell type, without mention of having achieved remission |
| ICD9CM | 208.11 | Chronic leukemia of unspecified cell type, in remission |
| ICD9CM | 208.20 | Subacute leukemia of unspecified cell type, without mention of having achieved remission |
| ICD9CM | 208.21 | Subacute leukemia of unspecified cell type, in remission |
| ICD9CM | 208.80 | Other leukemia of unspecified cell type, without mention of having achieved remission |
| ICD9CM | 208.81 | Other leukemia of unspecified cell type, in remission |
| ICD9CM | 208.90 | Unspecified leukemia, without mention of having achieved remission |
| SNOMED CT | 264906008 | Pituitary macroadenoma with extrasellar extension |
| SNOMED CT | 269459004 | Malignant tumor of lesser curve of stomach |
| SNOMED CT | 269460009 | Malignant tumor of greater curve of stomach |
| SNOMED CT | 269463006 | Malignant tumor of middle ear and mastoid |
| SNOMED CT | 269464000 | Malignant neoplasm of upper lobe, bronchus or lung |
| SNOMED CT | 269467007 | Malignant neoplasm of hand bones |

Continued on next page

**TableS24 – continued from previous page**

| CodeSystem | ConceptCode | ConceptName |
|---|---|---|
| SNOMED CT | 269475001 | Malignant tumor of lymphoid hemopoietic and related tissue |
| SNOMED CT | 269476000 | Nodular lymphoma |
| SNOMED CT | 269515006 | Carcinoma of lip |
| SNOMED CT | 269516007 | Tongue carcinoma |
| SNOMED CT | 269533000 | Carcinoma of colon |
| SNOMED CT | 269544008 | Carcinoma of the rectosigmoid junction |
| SNOMED CT | 269581007 | Malignant melanoma of lower limb |
| SNOMED CT | 271568003 | Malignant tumor of lower labial mucosa |
| SNOMED CT | 271943005 | Carcinoma of base of tongue |
| SNOMED CT | 274084007 | Palate carcinoma |
| SNOMED CT | 274085008 | Tonsil carcinoma |
| SNOMED CT | 274087000 | Malignant melanoma of eye |
| SNOMED CT | 274905008 | Malignant lymphoma - lymphocytic, intermediate differentiation |
| SNOMED CT | 275394001 | Carcinoma ventral surface of tongue |
| SNOMED CT | 275395000 | Carcinoma anterior 2/3 tongue ventrum |
| SNOMED CT | 275396004 | Carcinoma of anterior two-thirds of tongue - dorsal surface |
| SNOMED CT | 275397008 | Carcinoma of midline of tongue |
| SNOMED CT | 275399006 | Malignant tumor of lipstick area of lip |
| SNOMED CT | 275490009 | Carcinoma of tongue base - dorsal surface |
| SNOMED CT | 276419004 | Malignant tumor of corpus spongiosum |
| SNOMED CT | 276420005 | Malignant tumor of corpus cavernosum |
| SNOMED CT | 276751004 | Amelanotic malignant melanoma of skin |
| SNOMED CT | 276803003 | Adenocarcinoma of esophagus |
| SNOMED CT | 276804009 | Squamous cell carcinoma of esophagus |
| SNOMED CT | 276808007 | Carcinoid tumor of stomach |
| SNOMED CT | 276809004 | Early gastric cancer |
| SNOMED CT | 276810009 | Late gastric cancer |
| SNOMED CT | 276811008 | Gastric lymphoma |
| SNOMED CT | 276815004 | Lymphoma of intestine |
| SNOMED CT | 276821000 | Malignant melanoma of anus |
| SNOMED CT | 276826005 | Malignant glioma of brain |
| SNOMED CT | 276827001 | Malignant glioma of spinal cord |
| SNOMED CT | 276828006 | Glioblastoma multiforme of brain |
| SNOMED CT | 276829003 | Glioblastoma multiforme of spinal cord |
| SNOMED CT | 276836002 | Primary cerebral lymphoma |
| SNOMED CT | 276852004 | Papillary tumor of ampulla of Vater |
| SNOMED CT | 276860003 | Squamous cell carcinoma of scrotum |
| SNOMED CT | 276870001 | Carcinoma of fallopian tube |
| SNOMED CT | 276871002 | Vaginal intraepithelial neoplasia grade 1 |
| SNOMED CT | 276872009 | Vaginal intraepithelial neoplasia grade 2 |
| SNOMED CT | 276952000 | Squamous cell carcinoma of tongue |
| SNOMED CT | 276953005 | Squamous cell carcinoma of gum |
| SNOMED CT | 276954004 | Squamous cell carcinoma of floor of mouth |
| SNOMED CT | 276962007 | Squamous cell carcinoma of palate |
| SNOMED CT | 276968006 | Odontogenic tumor of jaw |
| SNOMED CT | 276975007 | Carcinoma of larynx |
| SNOMED CT | 277456001 | Suprasellar germ cell tumor |

Continued on next page

**TableS24 – continued from previous page**

| CodeSystem | ConceptCode | ConceptName |
|---|---|---|
| SNOMED CT | 277461004 | Anaplastic astrocytoma of brain |
| SNOMED CT | 277473004 | B-cell chronic lymphocytic leukemia |
| SNOMED CT | 277474005 | B-cell chronic lymphocytic leukemia variant |
| SNOMED CT | 277505007 | Medulloblastoma of cerebellum |
| SNOMED CT | 277507004 | Pilocytic astrocytoma of cerebellum |
| SNOMED CT | 277508009 | Pineal germ cell tumor |
| SNOMED CT | 277523004 | Primary melanocytic lesion of meninges |
| SNOMED CT | 277526007 | Diffuse melanosis of meninges |
| SNOMED CT | 277527003 | Melanocytoma of meninges |
| SNOMED CT | 277530005 | Malignant melanoma of meninges |
| SNOMED CT | 277545003 | T-cell chronic lymphocytic leukemia |
| SNOMED CT | 277549009 | Chronic lymphocytic prolymphocytic leukemia syndrome |
| SNOMED CT | 277551008 | Splenic lymphoma with villous lymphocytes |
| SNOMED CT | 277567002 | T-cell prolymphocytic leukemia |
| SNOMED CT | 277568007 | Hairy cell leukemia variant |
| SNOMED CT | 277569004 | Large granular lymphocytic leukemia |
| SNOMED CT | 277570003 | Lymphoma with spill |
| SNOMED CT | 277571004 | B-cell acute lymphoblastic leukemia |
| SNOMED CT | 277572006 | Precursor B-cell acute lymphoblastic leukemia |
| SNOMED CT | 277573001 | Common acute lymphoblastic leukemia |
| SNOMED CT | 277574007 | Null cell acute lymphoblastic leukemia |
| SNOMED CT | 277575008 | T-cell acute lymphoblastic leukemia |
| SNOMED CT | 277587001 | Juvenile chronic myeloid leukemia |
| SNOMED CT | 277589003 | Atypical chronic myeloid leukemia |
| SNOMED CT | 277597005 | Myelodysplastic syndrome with isolated del(5q) |
| SNOMED CT | 277601005 | Acute monoblastic leukemia |
| SNOMED CT | 277602003 | Acute megakaryoblastic leukemia |
| SNOMED CT | 277604002 | Acute eosinophilic leukemia |
| SNOMED CT | 277613000 | Cutaneous/peripheral T-cell lymphoma |
| SNOMED CT | 277614006 | Prethymic and thymic T-cell lymphoma/leukemia |
| SNOMED CT | 277615007 | Low grade B-cell lymphoma |
| SNOMED CT | 277616008 | Diffuse low grade B-cell lymphoma |
| SNOMED CT | 277617004 | High grade B-cell lymphoma |
| SNOMED CT | 277618009 | Follicular low grade B-cell lymphoma |
| SNOMED CT | 277619001 | B-cell prolymphocytic leukemia |
| SNOMED CT | 277622004 | Mucosa-associated lymphoma |
| SNOMED CT | 277623009 | Monocytoid B-cell lymphoma |
| SNOMED CT | 277624003 | Follicular malignant lymphoma - mixed cell type |
| SNOMED CT | 277625002 | Follicular malignant lymphoma - small cleaved cell |
| SNOMED CT | 277626001 | Diffuse high grade B-cell lymphoma |
| SNOMED CT | 277627005 | Nodular high grade B-cell lymphoma |
| SNOMED CT | 277628000 | Diffuse malignant lymphoma - large cleaved cell |
| SNOMED CT | 277629008 | Diffuse malignant lymphoma - large non-cleaved cell |
| SNOMED CT | 277632006 | Diffuse malignant lymphoma - centroblastic polymorphic |
| SNOMED CT | 277637000 | Large cell anaplastic lymphoma |
| SNOMED CT | 277642008 | Low grade T-cell lymphoma |
| SNOMED CT | 277643003 | High grade T-cell lymphoma |

Continued on next page

**TableS24 – continued from previous page**

| CodeSystem | ConceptCode | ConceptName |
|---|---|---|
| SNOMED CT | 277651000 | Peripheral T-cell lymphoma - pleomorphic small cell |
| SNOMED CT | 277653002 | Peripheral T-cell lymphoma - pleomorphic medium and large cell |
| SNOMED CT | 277664004 | Malignant lymphoma of testis |
| SNOMED CT | 277782009 | Malignant peritoneal local recurrence |
| SNOMED CT | 278024000 | Rhabdomyosarcoma of bladder |
| SNOMED CT | 278042005 | Malignant teratoma of mediastinum |
| SNOMED CT | 278043000 | Malignant seminoma of mediastinum |
| SNOMED CT | 278044006 | Malignant neuroma of mediastinum |
| SNOMED CT | 278046008 | Sarcoma of bladder |
| SNOMED CT | 278051002 | Malignant lymphoma of thyroid gland |
| SNOMED CT | 278052009 | Malignant lymphoma of breast |
| SNOMED CT | 278055006 | Malignant Leydig cell tumor of testis |
| SNOMED CT | 278057003 | Sertoli cell tumor of testis |
| SNOMED CT | 278060005 | Endometrioid carcinoma of prostate |
| SNOMED CT | 278189009 | Hypergranular promyelocytic leukemia |
| SNOMED CT | 278453007 | Acute biphenotypic leukemia |
| SNOMED CT | 278491007 | Mixed seminoma teratoma of testis |
| SNOMED CT | 280116003 | Tumor of tunica vaginalis |
| SNOMED CT | 280959007 | Malignant tumor of lacrimal drainage structure |
| SNOMED CT | 281560004 | Neuroblastoma of brain |
| SNOMED CT | 281561000 | Sacrococcygeal teratoma |
| SNOMED CT | 281562007 | Adrenal neuroblastoma |
| SNOMED CT | 281563002 | Thoracic neuroblastoma |
| SNOMED CT | 281566005 | Abdominothoracic neuroblastoma |
| SNOMED CT | 281702006 | Tibial adamantinoma |
| SNOMED CT | 285307007 | Squamous cell carcinoma of skin of upper lip |
| SNOMED CT | 285308002 | Squamous cell carcinoma of skin of lower lip |
| SNOMED CT | 285309005 | Squamous cell carcinoma of skin of cheek |
| SNOMED CT | 285310000 | Carcinoma of anal canal |
| SNOMED CT | 285311001 | Ameloblastoma of jaw |
| SNOMED CT | 285312008 | Carcinoma of sigmoid colon |
| SNOMED CT | 285432005 | Carcinoma of cervix |
| SNOMED CT | 285598005 | Metastasis to trachea of unknown primary |
| SNOMED CT | 285603002 | Metastasis to bronchus of unknown primary |
| SNOMED CT | 285604008 | Metastasis to lung of unknown primary |
| SNOMED CT | 285605009 | Metastasis to pleura of unknown primary |
| SNOMED CT | 285606005 | Metastasis to heart of unknown primary |
| SNOMED CT | 285607001 | Metastasis to mediastinum of unknown primary |
| SNOMED CT | 285608006 | Metastasis to thymus of unknown primary |
| SNOMED CT | 285611007 | Metastasis to colon of unknown primary |
| SNOMED CT | 285616002 | Metastasis to peritoneum of unknown primary |
| SNOMED CT | 285618001 | Metastasis to bone of unknown primary |
| SNOMED CT | 285619009 | Metastasis to vertebral column of unknown primary |
| SNOMED CT | 285634003 | Metastasis to breast of unknown primary |
| SNOMED CT | 285635002 | Metastasis to uterus of unknown primary |
| SNOMED CT | 285636001 | Cervical intraepithelial neoplasia |
| SNOMED CT | 285637005 | Metastasis to ovary of unknown primary |

Continued on next page

**TableS24 – continued from previous page**

| CodeSystem | ConceptCode | ConceptName |
|---|---|---|
| SNOMED CT | 285639008 | Metastasis to kidney of unknown primary |
| SNOMED CT | 285640005 | Metastasis to bladder of unknown primary |
| SNOMED CT | 285641009 | Metastasis to brain of unknown primary |
| SNOMED CT | 285642002 | Metastasis to eye of unknown primary |
| SNOMED CT | 285643007 | Metastasis to adrenal gland of unknown primary |
| SNOMED CT | 285644001 | Metastasis to lymph node of unknown primary |
| SNOMED CT | 285769009 | Acute promyelocytic leukemia - hypogranular variant |
| SNOMED CT | 285776004 | Intermediate grade B-cell lymphoma |
| SNOMED CT | 285836003 | Cervical intraepithelial neoplasia grade 1 |
| SNOMED CT | 285838002 | Cervical intraepithelial neoplasia grade 2 |
| SNOMED CT | 285839005 | Acute myelomonocytic leukemia - eosinophilic variant |
| SNOMED CT | 286889008 | Carcinoma of upper limb bones/scapula |
| SNOMED CT | 286890004 | Carcinoma of lower limb bones |
| SNOMED CT | 300988009 | Transitional cell carcinoma of ureter |
| SNOMED CT | 301756000 | Adenocarcinoma of sigmoid colon |
| SNOMED CT | 302815008 | Malignant tumor of frenum of lip |
| SNOMED CT | 302820008 | Intracranial meningioma |
| SNOMED CT | 302823005 | Glucagonoma |
| SNOMED CT | 302824004 | Gastrinoma |
| SNOMED CT | 302826002 | Adrenal cortical adenoma |
| SNOMED CT | 302837001 | Lentigo maligna melanoma |
| SNOMED CT | 302841002 | Malignant lymphoma - small lymphocytic |
| SNOMED CT | 302842009 | Diffuse malignant lymphoma - centroblastic |
| SNOMED CT | 302845006 | Nodular malignant lymphoma, lymphocytic - well differentiated |
| SNOMED CT | 302848008 | Nodular malignant lymphoma, lymphocytic - intermediate differentiation |
| SNOMED CT | 302849000 | Nephroblastoma |
| SNOMED CT | 302855005 | Subacute leukemia |
| SNOMED CT | 302856006 | Aleukemic leukemia |
| SNOMED CT | 303012000 | Malignant tumor of posterior wall of hypopharynx |
| SNOMED CT | 303017006 | Malignant lymphoma, convoluted cell type |
| SNOMED CT | 303055001 | Malignant lymphoma, follicular center cell |
| SNOMED CT | 303056000 | Malignant lymphoma, follicular center cell, cleaved |
| SNOMED CT | 303057009 | Malignant lymphoma, follicular center cell, non-cleaved |
| SNOMED CT | 303194003 | Metastasis to head and neck lymph node |
| SNOMED CT | 303201005 | Metastasis to multiple lymph nodes |
| SNOMED CT | 307216009 | Perforated carcinoma of esophagus |
| SNOMED CT | 307340003 | Monosomy 7 syndrome |
| SNOMED CT | 307341004 | Atypical hairy cell leukemia |
| SNOMED CT | 307502000 | Squamous cell carcinoma of mouth |
| SNOMED CT | 307576001 | Osteosarcoma |
| SNOMED CT | 307592006 | Basophilic leukemia |
| SNOMED CT | 307601000 | Pseudomyxoma peritonei |
| SNOMED CT | 307603002 | Malignant blue nevus of skin |
| SNOMED CT | 307604008 | Mesoblastic nephroma |
| SNOMED CT | 307605009 | Osteoblastoma of bone |
| SNOMED CT | 307606005 | Osteochondroma of bone |

Continued on next page

**TableS24 – continued from previous page**

| CodeSystem | ConceptCode | ConceptName |
|---|---|---|
| SNOMED CT | 307607001 | Chondromyxoid fibroma of bone |
| SNOMED CT | 307609003 | Adamantinoma of long bone |
| SNOMED CT | 307617006 | Neutrophilic leukemia |
| SNOMED CT | 307618001 | Juxtaglomerular tumor |
| SNOMED CT | 307622006 | Prolymphocytic lymphosarcoma |
| SNOMED CT | 307623001 | Malignant lymphoma - lymphoplasmacytic |
| SNOMED CT | 307624007 | Diffuse malignant lymphoma - centroblastic-centrocytic |
| SNOMED CT | 307625008 | Malignant lymphoma - centrocytic |
| SNOMED CT | 307636001 | Malignant lymphoma, mixed lymphocytic-histiocytic, nodular |
| SNOMED CT | 307637005 | Malignant lymphoma, centroblastic-centrocytic, follicular |
| SNOMED CT | 307646004 | Malignant lymphoma, lymphocytic, poorly differentiated, nodular |
| SNOMED CT | 307647008 | Malignant lymphoma, centroblastic type, follicular |
| SNOMED CT | 307649006 | Primary central nervous system lymphoma |
| SNOMED CT | 307650006 | Histiocytic medullary reticulosis |
| SNOMED CT | 309245001 | Adenocarcinoma of uterus |
| SNOMED CT | 310599006 | Malignant neoplasm of canthus |
| SNOMED CT | 312111009 | Carcinoma of ascending colon |
| SNOMED CT | 312112002 | Carcinoma of transverse colon |
| SNOMED CT | 312113007 | Carcinoma of descending colon |
| SNOMED CT | 312114001 | Carcinoma of hepatic flexure |
| SNOMED CT | 312115000 | Carcinoma of splenic flexure |
| SNOMED CT | 312949007 | Retinal pigment epithelial adenocarcinoma |
| SNOMED CT | 313248004 | Malignant melanoma of chest wall |
| SNOMED CT | 313249007 | Malignant neoplasm of upper eyelid |
| SNOMED CT | 313353007 | Squamous cell carcinoma of bronchus in left lower lobe |
| SNOMED CT | 313354001 | Squamous cell carcinoma of bronchus in left upper lobe |
| SNOMED CT | 313355000 | Squamous cell carcinoma of bronchus in right lower lobe |
| SNOMED CT | 313356004 | Squamous cell carcinoma of bronchus in right middle lobe |
| SNOMED CT | 313357008 | Squamous cell carcinoma of bronchus in right upper lobe |
| SNOMED CT | 313428008 | Seminoma of undescended testis |
| SNOMED CT | 313429000 | Seminoma of descended testis |
| SNOMED CT | 314191009 | Cystadenocarcinoma of ovary |
| SNOMED CT | 314408000 | Leukemic infiltrate of choroid |
| SNOMED CT | 314418005 | Leukemic infiltrate of retina |
| SNOMED CT | 314419002 | Bilateral diffuse uveal melanocytic proliferation |
| SNOMED CT | 314947008 | Carcinoid tumor of ampulla of Vater |
| SNOMED CT | 314951005 | Local recurrence of malignant tumor of tongue |
| SNOMED CT | 314952003 | Local recurrence of malignant tumor of buccal cavity |
| SNOMED CT | 314953008 | Local recurrence of malignant tumor of thyroid gland |
| SNOMED CT | 314954002 | Local recurrence of malignant tumor of lung |
| SNOMED CT | 314960002 | Local recurrence of malignant tumor of esophagus |
| SNOMED CT | 314961003 | Local recurrence of malignant tumor of stomach |
| SNOMED CT | 314965007 | Local recurrence of malignant tumor of colon |
| SNOMED CT | 314967004 | Local recurrence of malignant tumor of kidney |
| SNOMED CT | 314968009 | Local recurrence of malignant tumor of urinary bladder |
| SNOMED CT | 314969001 | Local recurrence of malignant tumor of prostate |

Continued on next page

**TableS24 – continued from previous page**

| CodeSystem | ConceptCode | ConceptName |
|---|---|---|
| SNOMED CT | 314970000 | Local recurrence of malignant tumor of cervix |
| SNOMED CT | 314973003 | Local recurrence of malignant tumor of bone |
| SNOMED CT | 314976006 | Local recurrence of malignant melanoma of skin |
| SNOMED CT | 359782004 | Metastatic malignant neoplasm to apex of urinary bladder |
| SNOMED CT | 359785002 | Metastatic malignant neoplasm to dome of urinary bladder |
| SNOMED CT | 359987004 | Krukenberg tumor |
| SNOMED CT | 363348004 | Malignant tumor of lip |
| SNOMED CT | 363349007 | Malignant tumor of stomach |
| SNOMED CT | 363352004 | Malignant tumor of anal canal |
| SNOMED CT | 363354003 | Malignant tumor of cervix |
| SNOMED CT | 363355002 | Malignant tumor of adrenal gland |
| SNOMED CT | 363358000 | Malignant tumor of lung |
| SNOMED CT | 363359008 | Malignant tumor of middle ear |
| SNOMED CT | 363360003 | Malignant tumor of anterior two-thirds of tongue |
| SNOMED CT | 363365008 | Malignant tumor of soft tissue of thorax |
| SNOMED CT | 363370001 | Malignant neoplasm of mesentery |
| SNOMED CT | 363372009 | Malignant tumor of vermilion border of upper lip |
| SNOMED CT | 363373004 | Malignant tumor of vermilion border of lower lip |
| SNOMED CT | 363374005 | Malignant tumor of commissure of lip |
| SNOMED CT | 363375006 | Malignant tumor of tongue |
| SNOMED CT | 363376007 | Malignant tumor of base of tongue |
| SNOMED CT | 363377003 | Malignant tumor of lingual tonsil |
| SNOMED CT | 363378008 | Malignant tumor of major salivary gland |
| SNOMED CT | 363379000 | Malignant tumor of parotid gland |
| SNOMED CT | 363380002 | Malignant tumor of submandibular gland |
| SNOMED CT | 363381003 | Malignant tumor of sublingual gland |
| SNOMED CT | 363382005 | Malignant tumor of gum |
| SNOMED CT | 363383000 | Malignant tumor of upper gingiva |
| SNOMED CT | 363384006 | Malignant tumor of lower gingiva |
| SNOMED CT | 363385007 | Malignant tumor of floor of mouth |
| SNOMED CT | 363386008 | Malignant tumor of buccal mucosa |
| SNOMED CT | 363387004 | Malignant tumor of hard palate |
| SNOMED CT | 363388009 | Malignant tumor of soft palate |
| SNOMED CT | 363389001 | Malignant tumor of uvula |
| SNOMED CT | 363390005 | Malignant tumor of palate |
| SNOMED CT | 363391009 | Malignant tumor of retromolar area |
| SNOMED CT | 363392002 | Malignant tumor of oropharynx |
| SNOMED CT | 363393007 | Malignant tumor of tonsil |
| SNOMED CT | 363394001 | Malignant tumor of tonsillar fossa |
| SNOMED CT | 363395000 | Malignant tumor of vallecula |
| SNOMED CT | 363398003 | Malignant tumor of lateral wall of nasopharynx |
| SNOMED CT | 363399006 | Malignant tumor of hypopharynx |
| SNOMED CT | 363400004 | Malignant tumor of postcricoid region |
| SNOMED CT | 363401000 | Malignant tumor of pyriform fossa |
| SNOMED CT | 363402007 | Malignant tumor of esophagus |
| SNOMED CT | 363403002 | Malignant tumor of duodenum |
| SNOMED CT | 363406005 | Malignant tumor of colon |

Continued on next page

**TableS24 – continued from previous page**

| CodeSystem | ConceptCode | ConceptName |
|---|---|---|
| SNOMED CT | 363407001 | Malignant tumor of hepatic flexure |
| SNOMED CT | 363408006 | Malignant tumor of transverse colon |
| SNOMED CT | 363409003 | Malignant tumor of descending colon |
| SNOMED CT | 363410008 | Malignant tumor of sigmoid colon |
| SNOMED CT | 363412000 | Malignant tumor of ascending colon |
| SNOMED CT | 363413005 | Malignant tumor of splenic flexure |
| SNOMED CT | 363414004 | Malignant tumor of rectosigmoid junction |
| SNOMED CT | 363417006 | Malignant tumor of ampulla of Vater |
| SNOMED CT | 363421004 | Malignant neoplasm of omentum |
| SNOMED CT | 363422006 | Malignant tumor of nasal cavity |
| SNOMED CT | 363423001 | Malignant tumor of nasal septum |
| SNOMED CT | 363424007 | Malignant tumor of mastoid air cells |
| SNOMED CT | 363425008 | Malignant tumor of maxillary sinus |
| SNOMED CT | 363426009 | Malignant tumor of ethmoid sinus |
| SNOMED CT | 363427000 | Malignant tumor of frontal sinus |
| SNOMED CT | 363428005 | Malignant tumor of sphenoid sinus |
| SNOMED CT | 363429002 | Malignant tumor of larynx |
| SNOMED CT | 363430007 | Malignant tumor of subglottis |
| SNOMED CT | 363431006 | Malignant tumor of laryngeal cartilage |
| SNOMED CT | 363432004 | Malignant tumor of trachea |
| SNOMED CT | 363433009 | Malignant tumor of pleura |
| SNOMED CT | 363434003 | Malignant tumor of thymus |
| SNOMED CT | 363435002 | Malignant tumor of heart |
| SNOMED CT | 363436001 | Malignant tumor of endocardium |
| SNOMED CT | 363437005 | Malignant tumor of myocardium |
| SNOMED CT | 363438000 | Malignant neoplasm of vertebral column |
| SNOMED CT | 363443007 | Malignant tumor of ovary |
| SNOMED CT | 363444001 | Malignant tumor of fallopian tube |
| SNOMED CT | 363445000 | Malignant tumor of vagina |
| SNOMED CT | 363446004 | Malignant neoplasm of labia majora |
| SNOMED CT | 363447008 | Malignant neoplasm of labia minora |
| SNOMED CT | 363449006 | Malignant tumor of testis |
| SNOMED CT | 363451005 | Malignant tumor of glans penis |
| SNOMED CT | 363452003 | Malignant tumor of epididymis |
| SNOMED CT | 363453008 | Malignant tumor of spermatic cord |
| SNOMED CT | 363454002 | Malignant tumor of scrotum |
| SNOMED CT | 363456000 | Malignant tumor of urachus |
| SNOMED CT | 363457009 | Malignant tumor of renal pelvis |
| SNOMED CT | 363458004 | Malignant tumor of ureter |
| SNOMED CT | 363459007 | Malignant tumor of urethra |
| SNOMED CT | 363460002 | Malignant tumor of paraurethral gland |
| SNOMED CT | 363461003 | Malignant neoplasm of eye |
| SNOMED CT | 363462005 | Malignant tumor of orbit |
| SNOMED CT | 363463000 | Malignant tumor of conjunctiva |
| SNOMED CT | 363464006 | Malignant tumor of cornea |
| SNOMED CT | 363465007 | Malignant tumor of retina |
| SNOMED CT | 363466008 | Malignant tumor of choroid |

Continued on next page

**TableS24 – continued from previous page**

| CodeSystem | ConceptCode | ConceptName |
|---|---|---|
| SNOMED CT | 363467004 | Malignant neoplasm of frontal lobe |
| SNOMED CT | 363468009 | Malignant neoplasm of temporal lobe |
| SNOMED CT | 363469001 | Malignant neoplasm of parietal lobe |
| SNOMED CT | 363470000 | Malignant neoplasm of occipital lobe |
| SNOMED CT | 363471001 | Malignant neoplasm of cerebral ventricles |
| SNOMED CT | 363473003 | Malignant neoplasm of brainstem |
| SNOMED CT | 363474009 | Malignant neoplasm of cerebral meninges |
| SNOMED CT | 363475005 | Malignant tumor of spinal cord |
| SNOMED CT | 363476006 | Malignant neoplasm of spinal meninges |
| SNOMED CT | 363478007 | Malignant tumor of thyroid gland |
| SNOMED CT | 363481002 | Malignant tumor of parathyroid gland |
| SNOMED CT | 363482009 | Malignant tumor of pituitary gland |
| SNOMED CT | 363483004 | Malignant tumor of pineal gland |
| SNOMED CT | 363485006 | Malignant tumor of minor salivary gland |
| SNOMED CT | 363486007 | Malignant tumor of vocal cord |
| SNOMED CT | 363487003 | Malignant tumor of aryepiglottic fold - laryngeal aspect |
| SNOMED CT | 363488008 | Malignant tumor of false cord |
| SNOMED CT | 363490009 | Malignant tumor of anus |
| SNOMED CT | 363491008 | Malignant tumor of cloacogenic zone |
| SNOMED CT | 363492001 | Malignant tumor of peritoneum |
| SNOMED CT | 363493006 | Malignant tumor of bronchus |
| SNOMED CT | 363494000 | Malignant tumor of mediastinum |
| SNOMED CT | 363497007 | Malignant tumor of meninges |
| SNOMED CT | 363498002 | Malignant tumor of optic nerve |
| SNOMED CT | 363502009 | Malignant tumor of axilla |
| SNOMED CT | 363506007 | Malignant tumor of nasal sinuses |
| SNOMED CT | 363507003 | Malignant tumor of pharynx |
| SNOMED CT | 363514001 | Malignant tumor of female genital organ |
| SNOMED CT | 363515000 | Malignant tumor of male genital organ |
| SNOMED CT | 363516004 | Malignant tumor of penis |
| SNOMED CT | 363517008 | Malignant tumor of urinary tract proper |
| SNOMED CT | 363518003 | Malignant tumor of kidney |
| SNOMED CT | 369463005 | Malignant tumor involving ureter by direct extension from bladder |
| SNOMED CT | 369464004 | Malignant tumor involving ureter by separate metastasis from bladder |
| SNOMED CT | 369465003 | Malignant tumor involving urethra by direct extension from bladder |
| SNOMED CT | 369466002 | Malignant tumor involving urethra by direct extension from prostate |
| SNOMED CT | 369467006 | Malignant tumor involving urethra by separate metastasis from bladder |
| SNOMED CT | 369468001 | Malignant tumor involving urethra by separate metastasis from prostate |
| SNOMED CT | 369469009 | Malignant tumor involving bladder by direct extension from endometrium |

Continued on next page

**TableS24 – continued from previous page**

| CodeSystem | ConceptCode | ConceptName |
|---|---|---|
| SNOMED CT | 369470005 | Malignant tumor involving bladder by direct extension from fallopian tube |
| SNOMED CT | 369471009 | Malignant tumor involving bladder by direct extension from ovary |
| SNOMED CT | 369472002 | Malignant tumor involving bladder by direct extension from prostate |
| SNOMED CT | 369475000 | Malignant tumor involving bladder by direct extension from vagina |
| SNOMED CT | 369476004 | Malignant tumor involving bladder by separate metastasis from endometrium |
| SNOMED CT | 369477008 | Malignant tumor involving bladder by separate metastasis from fallopian tube |
| SNOMED CT | 369478003 | Malignant tumor involving bladder by separate metastasis from ovary |
| SNOMED CT | 369479006 | Malignant tumor involving bladder by separate metastasis from prostate |
| SNOMED CT | 369480009 | Malignant tumor involving bladder by separate metastasis from uterine cervix |
| SNOMED CT | 369481008 | Malignant tumor involving bladder by separate metastasis from uterus |
| SNOMED CT | 369482001 | Malignant tumor involving bladder by separate metastasis from vagina |
| SNOMED CT | 369484000 | Malignant tumor involving vasa deferentia by separate metastasis from prostate |
| SNOMED CT | 369485004 | Malignant tumor involving prostate by direct extension from bladder |
| SNOMED CT | 369486003 | Malignant tumor involving prostate by separate metastasis from bladder |
| SNOMED CT | 369487007 | Primary malignant neoplasm of seminal vesicle |
| SNOMED CT | 369489005 | Malignant tumor involving seminal vesicle by direct extension from bladder |
| SNOMED CT | 369490001 | Malignant tumor involving seminal vesicle by direct extension from prostate |
| SNOMED CT | 369491002 | Malignant tumor involving seminal vesicle by separate metastasis from bladder |
| SNOMED CT | 369492009 | Malignant tumor involving seminal vesicle by separate metastasis from prostate |
| SNOMED CT | 369493004 | Malignant tumor involving uterine corpus by direct extension from bladder |
| SNOMED CT | 369494005 | Malignant tumor involving uterine corpus by direct extension from ovary |
| SNOMED CT | 369495006 | Malignant tumor involving uterine corpus by direct extension from uterine cervix |
| SNOMED CT | 369496007 | Malignant tumor involving uterine corpus by direct extension from vagina |
| SNOMED CT | 369497003 | Malignant tumor involving uterine cervix by direct extension from fallopian tube |

Continued on next page

**TableS24 – continued from previous page**

| CodeSystem | ConceptCode | ConceptName |
|---|---|---|
| SNOMED CT | 369498008 | Malignant tumor involving uterine cervix by direct extension from ovary |
| SNOMED CT | 369499000 | Malignant tumor involving uterine cervix by direct extension from vagina |
| SNOMED CT | 369500009 | Malignant tumor involving uterine cervix by separate metastasis from fallopian tube |
| SNOMED CT | 369501008 | Malignant tumor involving uterine cervix by separate metastasis from ovary |
| SNOMED CT | 369502001 | Malignant tumor involving uterine corpus by separate metastasis from bladder |
| SNOMED CT | 369503006 | Malignant tumor involving vagina by direct extension from uterus |
| SNOMED CT | 369504000 | Malignant tumor involving vagina by direct extension from bladder |
| SNOMED CT | 369505004 | Malignant tumor involving vagina by direct extension from endometrium |
| SNOMED CT | 369506003 | Malignant tumor involving vagina by direct extension from fallopian tube |
| SNOMED CT | 369512008 | Malignant tumor involving vagina by direct extension from ovary |
| SNOMED CT | 369513003 | Primary malignant neoplasm of left fallopian tube |
| SNOMED CT | 369515005 | Malignant tumor involving left fallopian tube by direct extension from endometrium |
| SNOMED CT | 369516006 | Malignant tumor involving left fallopian tube by direct extension from ovary |
| SNOMED CT | 369517002 | Malignant tumor involving left fallopian tube by direct extension from right fallopian tube |
| SNOMED CT | 369518007 | Malignant tumor involving left fallopian tube by direct extension from uterine cervix |
| SNOMED CT | 369519004 | Malignant tumor involving left fallopian tube by direct extension from uterus |
| SNOMED CT | 369520005 | Primary malignant neoplasm of right fallopian tube |
| SNOMED CT | 369522002 | Primary malignant neoplasm of left ovary |
| SNOMED CT | 369524001 | Malignant tumor involving left ovary by direct extension from endometrium |
| SNOMED CT | 369525000 | Malignant tumor involving left ovary by direct extension from fallopian tube |
| SNOMED CT | 369526004 | Malignant tumor involving left ovary by direct extension from right ovary |
| SNOMED CT | 369527008 | Malignant tumor involving left ovary by direct extension from uterine cervix |
| SNOMED CT | 369528003 | Malignant tumor involving left ovary by direct extension from uterus |
| SNOMED CT | 369529006 | Primary malignant neoplasm of right ovary |
| SNOMED CT | 369531002 | Malignant tumor involving right ovary by direct extension from endometrium |
| SNOMED CT | 369532009 | Malignant tumor involving right ovary by direct extension from fallopian tube |

Continued on next page

**TableS24 – continued from previous page**

| CodeSystem | ConceptCode | ConceptName |
|---|---|---|
| SNOMED CT | 369533004 | Malignant tumor involving right ovary by direct extension from left ovary |
| SNOMED CT | 369534005 | Malignant tumor involving right ovary by direct extension from uterine cervix |
| SNOMED CT | 369537003 | Malignant tumor involving left broad ligament by direct extension from ovary |
| SNOMED CT | 369538008 | Malignant tumor involving left broad ligament by metastasis from ovary |
| SNOMED CT | 369539000 | Malignant tumor involving right broad ligament by direct extension from ovary |
| SNOMED CT | 369540003 | Malignant tumor involving right broad ligament by metastasis from ovary |
| SNOMED CT | 369541004 | Malignant tumor involving left fallopian tube by direct extension from vagina |
| SNOMED CT | 369547000 | Malignant tumor involving right fallopian tube by direct extension from endometrium |
| SNOMED CT | 369548005 | Malignant tumor involving right fallopian tube by direct extension from left fallopian tube |
| SNOMED CT | 369549002 | Malignant tumor involving right fallopian tube by direct extension from ovary |
| SNOMED CT | 369550002 | Malignant tumor involving right fallopian tube by direct extension from uterine cervix |
| SNOMED CT | 369551003 | Malignant tumor involving right fallopian tube by direct extension from uterus |
| SNOMED CT | 369552005 | Malignant tumor involving right fallopian tube by direct extension from vagina |
| SNOMED CT | 369559001 | Malignant tumor involving left ovary by direct extension from vagina |
| SNOMED CT | 369566000 | Malignant tumor involving right ovary by direct extension from uterus |
| SNOMED CT | 369567009 | Malignant tumor involving right ovary by direct extension from vagina |
| SNOMED CT | 369574004 | Malignant tumor involving uterine cervix by separate metastasis from vagina |
| SNOMED CT | 369575003 | Malignant tumor involving uterine corpus by separate metastasis from fallopian tube |
| SNOMED CT | 369576002 | Malignant tumor involving uterine corpus by separate metastasis from ovary |
| SNOMED CT | 369577006 | Malignant tumor involving uterine corpus by separate metastasis from uterine cervix |
| SNOMED CT | 369578001 | Malignant tumor involving uterine corpus by separate metastasis from vagina |
| SNOMED CT | 369579009 | Malignant tumor involving uterine corpus by direct extension from fallopian tube |
| SNOMED CT | 369580007 | Malignant tumor involving vagina by direct extension from uterine cervix |
| SNOMED CT | 370967009 | Retinoblastoma |

Continued on next page

**TableS24 – continued from previous page**

| CodeSystem | ConceptCode | ConceptName |
|---|---|---|
| SNOMED CT | 370987005 | Anaplastic astrocytoma of spinal cord |
| SNOMED CT | 371012000 | Acute lymphoblastic leukemia, transitional pre-B-cell |
| SNOMED CT | 371134001 | Malignant lymphoma, large cell, polymorphous, immunoblastic |
| SNOMED CT | 371962007 | Primary malignant neoplasm of abdominal esophagus |
| SNOMED CT | 371963002 | Primary malignant neoplasm of adrenal cortex |
| SNOMED CT | 371964008 | Malignant neoplasm of adrenal cortex |
| SNOMED CT | 371965009 | Malignant neoplasm of adrenal medulla |
| SNOMED CT | 371966005 | Primary malignant neoplasm of adrenal medulla |
| SNOMED CT | 371967001 | Primary malignant neoplasm of ampulla of Vater |
| SNOMED CT | 371968006 | Primary malignant neoplasm of anterior two-thirds of tongue |
| SNOMED CT | 371969003 | Primary malignant neoplasm of apex of urinary bladder |
| SNOMED CT | 371971003 | Primary malignant neoplasm of body of uterus |
| SNOMED CT | 371972005 | Malignant neoplasm of body of uterus |
| SNOMED CT | 371973000 | Malignant neoplasm of uterus |
| SNOMED CT | 371974006 | Malignant neoplasm of border of tongue |
| SNOMED CT | 371975007 | Primary malignant neoplasm of border of tongue |
| SNOMED CT | 371976008 | Primary malignant neoplasm of buccal mucosa |
| SNOMED CT | 371978009 | Primary malignant neoplasm of cervical esophagus |
| SNOMED CT | 371979001 | Malignant neoplasm of clitoris |
| SNOMED CT | 371980003 | Primary malignant neoplasm of clitoris |
| SNOMED CT | 371981004 | Primary malignant neoplasm of commissure of lip |
| SNOMED CT | 371982006 | Malignant neoplasm of endocrine gland |
| SNOMED CT | 371983001 | Primary malignant neoplasm of endocrine gland |
| SNOMED CT | 371984007 | Primary malignant neoplasm of esophagus |
| SNOMED CT | 371986009 | Primary malignant neoplasm of eye |
| SNOMED CT | 371987000 | Primary malignant neoplasm of fallopian tube |
| SNOMED CT | 371988005 | Primary malignant neoplasm of false vocal cord |
| SNOMED CT | 371989002 | Primary malignant neoplasm of glans penis |
| SNOMED CT | 371990006 | Primary malignant neoplasm of gum |
| SNOMED CT | 371991005 | Primary malignant neoplasm of hard palate |
| SNOMED CT | 371993008 | Primary malignant neoplasm of lacrimal gland |
| SNOMED CT | 371994002 | Primary malignant neoplasm of laryngeal aspect of aryepiglottic fold |
| SNOMED CT | 371995001 | Primary malignant neoplasm of larynx |
| SNOMED CT | 371996000 | Primary malignant neoplasm of lip |
| SNOMED CT | 371997009 | Primary malignant neoplasm of lower gum |
| SNOMED CT | 371998004 | Primary malignant neoplasm of lower third of esophagus |
| SNOMED CT | 371999007 | Primary malignant neoplasm of middle third of esophagus |
| SNOMED CT | 372000001 | Primary malignant neoplasm of minor salivary gland |
| SNOMED CT | 372001002 | Primary malignant neoplasm of oral cavity |
| SNOMED CT | 372002009 | Primary malignant neoplasm of palate |
| SNOMED CT | 372004005 | Primary malignant neoplasm of parotid gland |
| SNOMED CT | 372005006 | Primary malignant neoplasm of penis |
| SNOMED CT | 372006007 | Primary malignant neoplasm of prepuce |
| SNOMED CT | 372008008 | Primary malignant neoplasm of scaphoid bone |
| SNOMED CT | 372009000 | Primary malignant neoplasm of scrotum |
| SNOMED CT | 372013007 | Primary malignant neoplasm of spermatic cord |

Continued on next page

**TableS24 – continued from previous page**

| CodeSystem | ConceptCode | ConceptName |
|---|---|---|
| SNOMED CT | 372014001 | Primary malignant neoplasm of stomach |
| SNOMED CT | 372015000 | Primary malignant neoplasm of the mesentery |
| SNOMED CT | 372016004 | Primary malignant neoplasm of the peritoneum |
| SNOMED CT | 372017008 | Primary malignant neoplasm of thoracic esophagus |
| SNOMED CT | 372018003 | Malignant neoplasm of thoracic vertebral column |
| SNOMED CT | 372019006 | Primary malignant neoplasm of thoracic vertebral column |
| SNOMED CT | 372020000 | Primary malignant neoplasm of tonsil |
| SNOMED CT | 372021001 | Primary malignant neoplasm of trapezium |
| SNOMED CT | 372022008 | Primary malignant neoplasm of upper gum |
| SNOMED CT | 372024009 | Primary malignant neoplasm of uterine cervix |
| SNOMED CT | 372025005 | Primary malignant neoplasm of vagina |
| SNOMED CT | 372026006 | Primary malignant neoplasm of vermilion border of lower lip |
| SNOMED CT | 372027002 | Primary malignant neoplasm of vermilion border of upper lip |
| SNOMED CT | 372028007 | Primary malignant neoplasm of vertebral column |
| SNOMED CT | 372030009 | Primary malignant neoplasm of vocal cord |
| SNOMED CT | 372062007 | Malignant neoplasm of central nervous system |
| SNOMED CT | 372064008 | Malignant neoplasm of female breast |
| SNOMED CT | 372065009 | Malignant neoplasm of main bronchus |
| SNOMED CT | 372097009 | Malignant neoplasm of endocervix |
| SNOMED CT | 372098004 | Carcinoma of endocervix |
| SNOMED CT | 372099007 | Malignant neoplasm of exocervix |
| SNOMED CT | 372100004 | Carcinoma of exocervix |
| SNOMED CT | 372103002 | Carcinoma of glottis |
| SNOMED CT | 372104008 | Carcinoma of subglottis |
| SNOMED CT | 372105009 | Carcinoma of supraglottis |
| SNOMED CT | 372106005 | Carcinoma of penis |
| SNOMED CT | 372107001 | Primary malignant neoplasm of ribs and/or sternum and/or clavicle |
| SNOMED CT | 372108006 | Malignant neoplasm of bone of lower limb |
| SNOMED CT | 372110008 | Primary malignant neoplasm of lower lobe, bronchus or lung |
| SNOMED CT | 372111007 | Carcinoma of lower lobe, bronchus or lung |
| SNOMED CT | 372115003 | Primary malignant neoplasm of pelvic bones, sacrum and coccyx |
| SNOMED CT | 372116002 | Carcinoma of pelvic bones, sacrum and coccyx |
| SNOMED CT | 372120003 | Carcinoma of main bronchus |
| SNOMED CT | 372121004 | Carcinoma of ribs and/or sternum and/or clavicle |
| SNOMED CT | 372133009 | Primary malignant neoplasm of upper limb bones and scapula |
| SNOMED CT | 372138000 | Carcinoma of esophagus |
| SNOMED CT | 372141009 | Carcinoma of vocal cord |
| SNOMED CT | 372143007 | Carcinoma of stomach |
| SNOMED CT | 373168002 | Reticulosarcoma |
| SNOMED CT | 385478001 | Adenoma malignum |
| SNOMED CT | 388871003 | Primary optic nerve sheath meningioma |
| SNOMED CT | 396198006 | Small cell carcinoma of prostate |
| SNOMED CT | 397008008 | Aggressive lymphadenopathic mastocytosis with eosinophilia |
| SNOMED CT | 397009000 | Mast cell malignancy |
| SNOMED CT | 397011009 | Mast cell malignancy of lymph nodes |

Continued on next page

**TableS24 – continued from previous page**

| CodeSystem | ConceptCode | ConceptName |
|---|---|---|
| SNOMED CT | 397015000 | Systemic mastocytosis with associated clonal hematological non-mast cell lineage disease |
| SNOMED CT | 398623004 | Myelodysplastic syndrome with excess blasts |
| SNOMED CT | 398679002 | Bowenoid papulosis of penis |
| SNOMED CT | 398768004 | Queyrat's erythroplasia |
| SNOMED CT | 398831006 | Bowens disease of penis |
| SNOMED CT | 399068003 | Malignant tumor of prostate |
| SNOMED CT | 399326009 | Malignant tumor of urinary bladder |
| SNOMED CT | 399490008 | Adenocarcinoma of prostate |
| SNOMED CT | 399590005 | Squamous cell carcinoma of prostate |
| SNOMED CT | 399660006 | Ring melanoma of ciliary body |
| SNOMED CT | 399967006 | Neoplasm of uncertain behavior of stomach |
| SNOMED CT | 400001003 | Primary cutaneous lymphoma |
| SNOMED CT | 400092004 | Penile intraepithelial neoplasia grade III |
| SNOMED CT | 400118002 | Bowenoid papulosis of penis (PIN III) |
| SNOMED CT | 400122007 | Primary cutaneous T-cell lymphoma |
| SNOMED CT | 401312001 | Anal intraepithelial neoplasia grade II |
| SNOMED CT | 401313006 | Anal intraepithelial neoplasia grade I |
| SNOMED CT | 402494003 | Basal cell carcinoma of lower eyelid |
| SNOMED CT | 402495002 | Basal cell carcinoma of medial canthus |
| SNOMED CT | 402496001 | Basal cell carcinoma of lateral canthus |
| SNOMED CT | 402503002 | Basal cell carcinoma of nasal columella |
| SNOMED CT | 402504008 | Basal cell carcinoma of nasolabial groove |
| SNOMED CT | 402505009 | Basal cell carcinoma of upper lip |
| SNOMED CT | 402506005 | Basal cell carcinoma of lower lip |
| SNOMED CT | 402507001 | Basal cell carcinoma of cheek |
| SNOMED CT | 402515003 | Basal cell carcinoma of anterior chest |
| SNOMED CT | 402819001 | Basal cell carcinoma of skin of lip |
| SNOMED CT | 402880009 | Primary cutaneous large T-cell lymphoma |
| SNOMED CT | 402882001 | Hodgkin's disease affecting skin |
| SNOMED CT | 403467008 | Lichen sclerosus-associated penile intraepithelial neoplasia |
| SNOMED CT | 403468003 | Squamous cell carcinoma of penis |
| SNOMED CT | 403714009 | PUVA therapy-associated malignant melanoma |
| SNOMED CT | 403889000 | Verrucous carcinoma of oral cavity |
| SNOMED CT | 403919005 | Basal cell carcinoma of upper eyelid |
| SNOMED CT | 403926005 | Malignant melanoma of oral cavity |
| SNOMED CT | 403950007 | Moll's gland adenocarcinoma |
| SNOMED CT | 404090003 | Malignant infiltration of oral cavity by underlying tumor |
| SNOMED CT | 404094007 | Metastasis involving oral cavity |
| SNOMED CT | 404103007 | Lymphomatoid papulosis type A (CD-30 positive type) |
| SNOMED CT | 404104001 | Lymphomatoid papulosis type B - mycosis fungoides-like |
| SNOMED CT | 404105000 | Lymphomatoid papulosis type C (anaplastic large-cell lymphoma-like) |
| SNOMED CT | 404107008 | Patch/plaque stage mycosis fungoides |
| SNOMED CT | 404108003 | Poikilodermatous mycosis fungoides |
| SNOMED CT | 404109006 | Follicular mucinosis type mycosis fungoides |
| SNOMED CT | 404110001 | Hypomelanotic mycosis fungoides |

Continued on next page

**TableS24 – continued from previous page**

| CodeSystem | ConceptCode | ConceptName |
|---|---|---|
| SNOMED CT | 404111002 | Lymphomatoid papulosis-associated mycosis fungoides |
| SNOMED CT | 404112009 | Granulomatous mycosis fungoides |
| SNOMED CT | 404113004 | Tumor stage mycosis fungoides |
| SNOMED CT | 404114005 | Erythrodermic mycosis fungoides |
| SNOMED CT | 404115006 | Bullous mycosis fungoides |
| SNOMED CT | 404116007 | Mycosis fungoides with systemic infiltration |
| SNOMED CT | 404117003 | Spongiotic mycosis fungoides |
| SNOMED CT | 404118008 | Syringotropic mycosis fungoides |
| SNOMED CT | 404119000 | Pagetoid reticulosis |
| SNOMED CT | 404120006 | Localized pagetoid reticulosis |
| SNOMED CT | 404121005 | Generalized pagetoid reticulosis |
| SNOMED CT | 404126000 | CD-30 positive pleomorphic large T-cell cutaneous lymphoma |
| SNOMED CT | 404127009 | CD-30 positive T-immunoblastic cutaneous lymphoma |
| SNOMED CT | 404128004 | CD-30 negative cutaneous T-cell lymphoma |
| SNOMED CT | 404129007 | CD-30 negative anaplastic large T-cell cutaneous lymphoma |
| SNOMED CT | 404130002 | CD-30 negative pleomorphic large T-cell cutaneous lymphoma |
| SNOMED CT | 404131003 | CD-30 negative T-immunoblastic cutaneous lymphoma |
| SNOMED CT | 404133000 | Subcutaneous panniculitis-like T-cell lymphoma |
| SNOMED CT | 404134006 | Anaplastic large T-cell systemic malignant lymphoma |
| SNOMED CT | 404137004 | Precursor B-cell lymphoblastic lymphoma involving skin |
| SNOMED CT | 404138009 | Small lymphocytic B-cell lymphoma involving skin |
| SNOMED CT | 404139001 | Leukemic infiltration of skin in hairy-cell leukemia |
| SNOMED CT | 404140004 | Primary cutaneous marginal zone B-cell lymphoma |
| SNOMED CT | 404141000 | Primary cutaneous immunocytoma |
| SNOMED CT | 404142007 | Primary cutaneous plasmacytoma |
| SNOMED CT | 404144008 | Primary cutaneous diffuse large cell B-cell lymphoma |
| SNOMED CT | 404147001 | Follicular center B-cell lymphoma (nodal/systemic with skin involvement) |
| SNOMED CT | 404148006 | Diffuse large B-cell lymphoma (nodal/systemic with skin involvement) |
| SNOMED CT | 404149003 | Lymphoplasmacytic B-cell lymphoma, nodal/systemic with skin involvement |
| SNOMED CT | 404150003 | Mantle cell B-cell lymphoma (nodal/systemic with skin involvement) |
| SNOMED CT | 404151004 | Leukemic infiltration of skin in myeloid leukemia |
| SNOMED CT | 404152006 | Leukemic infiltration of skin in acute myeloid leukemia |
| SNOMED CT | 404153001 | Leukemic infiltration of skin in chronic myeloid leukemia |
| SNOMED CT | 404154007 | Leukemic infiltration of skin in monocytic leukemia |
| SNOMED CT | 404155008 | Granulocytic sarcoma affecting skin |
| SNOMED CT | 404169008 | Malignant histiocytosis involving skin |
| SNOMED CT | 404172001 | Mast cell leukemia affecting skin |
| SNOMED CT | 404653000 | Chiasmal glioma |
| SNOMED CT | 404664002 | Malignant optic glioma |
| SNOMED CT | 405546008 | Malignant pericardial effusion |
| SNOMED CT | 405822008 | Squamous cell carcinoma of larynx |
| SNOMED CT | 405945003 | Malignant neoplasm of metatarsal bone of foot |
| SNOMED CT | 408642003 | Transitional cell carcinoma of kidney |

Continued on next page

**TableS24 – continued from previous page**

| CodeSystem | ConceptCode | ConceptName |
|---|---|---|
| SNOMED CT | 408644002 | Adenocarcinoma of duodenum |
| SNOMED CT | 408647009 | Adenocarcinoma of stomach |
| SNOMED CT | 408648004 | Squamous cell carcinoma of epiglottis |
| SNOMED CT | 408649007 | Squamous cell carcinoma of pharynx |
| SNOMED CT | 413389003 | Accelerated phase chronic myeloid leukemia |
| SNOMED CT | 413441006 | Acute monocytic leukemia |
| SNOMED CT | 413442004 | Acute monocytic/monoblastic leukemia |
| SNOMED CT | 413537009 | Angioimmunoblastic T-cell lymphoma |
| SNOMED CT | 413656006 | Blastic phase chronic myeloid leukemia |
| SNOMED CT | 413842007 | Chronic myeloid leukemia in lymphoid blast crisis |
| SNOMED CT | 413843002 | Chronic myeloid leukemia in myeloid blast crisis |
| SNOMED CT | 413847001 | Chronic phase chronic myeloid leukemia |
| SNOMED CT | 414250001 | Fibrous histiocytoma of orbit |
| SNOMED CT | 414626005 | Lymphangioma of orbit |
| SNOMED CT | 414666009 | Meningioma of orbit |
| SNOMED CT | 414676007 | Metastatic neuroblastoma of orbit proper |
| SNOMED CT | 414785000 | Multiple solitary plasmacytomas |
| SNOMED CT | 414950005 | Optic nerve glioma of orbit |
| SNOMED CT | 415112005 | Plasmacytoma |
| SNOMED CT | 415283002 | Myelodysplastic syndrome with excess blasts-1 |
| SNOMED CT | 415284008 | Myelodysplastic syndrome with excess blasts-2 |
| SNOMED CT | 415285009 | Myelodysplastic syndrome with multilineage dysplasia |
| SNOMED CT | 415286005 | Myelodysplastic syndrome with ring sideroblasts and multilineage dysplasia |
| SNOMED CT | 415287001 | Relapsing chronic myeloid leukemia |
| SNOMED CT | 415653001 | Submucosal tumor of esophagus |
| SNOMED CT | 416351002 | Intraocular optic nerve glioma |
| SNOMED CT | 416510003 | Neoplasm of lacrimal system |
| SNOMED CT | 416712009 | Clear cell (mesonephric) neoplasm of ovary |
| SNOMED CT | 416753001 | Neoplasm of nasolacrimal duct |
| SNOMED CT | 416842003 | Malignant sacral teratoma |
| SNOMED CT | 416901002 | Malignant medulloepithelioma of ciliary body |
| SNOMED CT | 417417007 | Malignant teratoma of undescended testis |
| SNOMED CT | 417554000 | Malignant teratoma of descended testis |
| SNOMED CT | 417619001 | Intracranial optic nerve glioma |
| SNOMED CT | 418134006 | Conjunctival intraepithelial neoplasia |
| SNOMED CT | 418372008 | Squamous cell carcinoma of mucous membrane of lower lip |
| SNOMED CT | 419052002 | Malignant tumor of urinary system |
| SNOMED CT | 419240004 | Squamous cell carcinoma of mucous membrane of upper lip |
| SNOMED CT | 419317000 | Neoplasm of uncertain behavior of lacrimal drainage structure |
| SNOMED CT | 419327006 | Endometrial intraepithelial neoplasia |
| SNOMED CT | 419842002 | Squamous cell carcinoma of oral mucous membrane |
| SNOMED CT | 419889000 | Neoplasm of lacrimal drainage system |
| SNOMED CT | 420519005 | Malignant lymphoma of the eye region |
| SNOMED CT | 420788006 | Primary intraocular non-Hodgkin malignant lymphoma |
| SNOMED CT | 420835009 | Corneal intraepithelial neoplasia |
| SNOMED CT | 421246008 | Precursor T-cell lymphoblastic lymphoma |

Continued on next page

**TableS24 – continued from previous page**

| CodeSystem | ConceptCode | ConceptName |
|---|---|---|
| SNOMED CT | 421249001 | Malignant tumor of vermilion border of lip |
| SNOMED CT | 422541001 | Undifferentiated carcinoma of nasopharynx |
| SNOMED CT | 422691006 | Squamous cell carcinoma of nasopharynx |
| SNOMED CT | 422758009 | Carcinoma of nasal meatus |
| SNOMED CT | 422782004 | Ovarian cancer, disseminated |
| SNOMED CT | 422833009 | Adenoid cystic carcinoma of salivary gland |
| SNOMED CT | 422853008 | Lymphoma of retroperitoneal space |
| SNOMED CT | 422886007 | Olfactory neuroblastoma |
| SNOMED CT | 422968005 | Non-small cell carcinoma of lung, TNM stage 3 |
| SNOMED CT | 423005002 | Primary malignant neoplasm of lacrimal gland duct |
| SNOMED CT | 423032007 | Leukemic infiltration of orbit |
| SNOMED CT | 423038006 | Polymorphous low grade adenocarcinoma of salivary gland |
| SNOMED CT | 423050000 | Large cell carcinoma of lung, TNM stage 2 |
| SNOMED CT | 423106003 | Adenocarcinoma of nasopharynx |
| SNOMED CT | 423121009 | Non-small cell carcinoma of lung, TNM stage 4 |
| SNOMED CT | 423158009 | Hurthle cell carcinoma of thyroid |
| SNOMED CT | 423189008 | Adenoid cystic carcinoma of submandibular gland |
| SNOMED CT | 423195009 | Primary malignant neoplasm of lacrimal drainage system |
| SNOMED CT | 423278008 | Lentigo maligna of skin of eyelid |
| SNOMED CT | 423280002 | Malignant melanoma of skin of canthus of eye |
| SNOMED CT | 423295000 | Squamous cell carcinoma of lung, TNM stage 1 |
| SNOMED CT | 423424005 | Mucoepidermoid carcinoma of submandibular gland |
| SNOMED CT | 423425006 | Malignant neoplasm of skin of eyelid |
| SNOMED CT | 423447006 | Malignant melanoma of skin of lower eyelid |
| SNOMED CT | 423464009 | Squamous cell carcinoma of oropharynx |
| SNOMED CT | 423468007 | Squamous cell carcinoma of lung, TNM stage 2 |
| SNOMED CT | 423494003 | Malignant melanoma of skin of upper eyelid |
| SNOMED CT | 423600008 | Large cell carcinoma of lung, TNM stage 4 |
| SNOMED CT | 423607006 | Adenocarcinoma of anus |
| SNOMED CT | 423615009 | Adenoid cystic carcinoma of parotid gland |
| SNOMED CT | 423619003 | Teratoma of orbit |
| SNOMED CT | 423627007 | Sarcoma of ovary |
| SNOMED CT | 423673009 | Malignant melanoma of retina |
| SNOMED CT | 423691004 | Malignant neoplasm of gum and contiguous sites |
| SNOMED CT | 423708008 | Mucoepidermoid carcinoma of salivary gland |
| SNOMED CT | 423973006 | Carcinoma of uterine cervix, invasive |
| SNOMED CT | 423987006 | Cancer of vulva, disseminated |
| SNOMED CT | 424132000 | Non-small cell carcinoma of lung, TNM stage 1 |
| SNOMED CT | 424151006 | Anaplastic glioma of brain |
| SNOMED CT | 424260006 | Squamous cell carcinoma of nasolabial fold |
| SNOMED CT | 424276002 | Malignant glioma of brainstem |
| SNOMED CT | 424302003 | Malignant melanoma of skin of lower lip |
| SNOMED CT | 424334007 | Malignant tumor of spinal cord, intramedullary |
| SNOMED CT | 424408000 | Keratoacanthoma of eyelid |
| SNOMED CT | 424422000 | Osteoma of orbit |
| SNOMED CT | 424487008 | Malignant melanoma of skin of upper lip |
| SNOMED CT | 424549003 | Malignant tumor of spinal cord, extramedullary |

Continued on next page

**TableS24 – continued from previous page**

| CodeSystem | ConceptCode | ConceptName |
|---|---|---|
| SNOMED CT | 424849005 | Primary sarcoma of tongue |
| SNOMED CT | 424887002 | Thyroid cancer metastatic to bone |
| SNOMED CT | 424938000 | Large cell carcinoma of lung, TNM stage 1 |
| SNOMED CT | 424970000 | Large cell carcinoma of lung, TNM stage 3 |
| SNOMED CT | 425048006 | Non-small cell carcinoma of lung, TNM stage 2 |
| SNOMED CT | 425066001 | Carcinoma of urinary bladder, invasive |
| SNOMED CT | 425127006 | Carcinoma ex pleomorphic adenoma of parotid gland |
| SNOMED CT | 425178004 | Adenocarcinoma of rectosigmoid junction |
| SNOMED CT | 425225007 | Malignant mixed tumor of salivary gland |
| SNOMED CT | 425230006 | Squamous cell carcinoma of lung, TNM stage 3 |
| SNOMED CT | 425231005 | Carcinoma of urinary bladder, superficial |
| SNOMED CT | 425376008 | Squamous cell carcinoma of lung, TNM stage 4 |
| SNOMED CT | 425688002 | Philadelphia chromosome-positive acute lymphoblastic leukemia |
| SNOMED CT | 425749006 | Subacute myeloid leukemia in remission |
| SNOMED CT | 425869007 | Acute promyelocytic leukemia, FAB M3, in remission |
| SNOMED CT | 425941003 | Precursor B-cell acute lymphoblastic leukemia in remission |
| SNOMED CT | 426071002 | Hodgkin's disease in remission |
| SNOMED CT | 426124006 | Acute myeloid leukemia with maturation, FAB M2, in remission |
| SNOMED CT | 426191007 | Myxoma of heart |
| SNOMED CT | 426217000 | Aleukaemic leukaemia in remission |
| SNOMED CT | 426248008 | Aleukemic lymphoid leukemia in remission |
| SNOMED CT | 426336007 | Solitary osseous myeloma |
| SNOMED CT | 426370008 | Subacute lymphoid leukemia in remission |
| SNOMED CT | 426642002 | Erythroleukemia, FAB M6 in remission |
| SNOMED CT | 426885008 | Hodgkin's disease, lymphocytic depletion of lymph nodes of head |
| SNOMED CT | 426964009 | Epidermal growth factor receptor positive non-small cell lung cancer |
| SNOMED CT | 427038005 | Epidermal growth factor receptor negative non-small cell lung cancer |
| SNOMED CT | 427056005 | Subacute leukemia in remission |
| SNOMED CT | 427141003 | Malignant lymphoma in remission |
| SNOMED CT | 427492003 | Hormone refractory prostate cancer |
| SNOMED CT | 427642009 | T-cell acute lymphoblastic leukemia in remission |
| SNOMED CT | 427658007 | Acute myelomonocytic leukemia, FAB M4, in remission |
| SNOMED CT | 428061005 | Malignant neoplasm of brain |
| SNOMED CT | 428100006 | Malignant neoplasm of thoracic cavity structure |
| SNOMED CT | 428281000 | Malignant neoplasm of bone |
| SNOMED CT | 428753007 | Low grade glioma of cerebellum |
| SNOMED CT | 428960009 | Low grade glioma of brainstem |
| SNOMED CT | 428964000 | Low grade glioma of cerebrum |
| SNOMED CT | 428965004 | Low grade glioma of thalamus |
| SNOMED CT | 429033009 | Malignant neoplasm of cerebrum |
| SNOMED CT | 429408002 | Low grade glioma of brain |
| SNOMED CT | 429565004 | Germ cell tumor of the brain |
| SNOMED CT | 430338009 | Smoldering chronic lymphocytic leukemia |
| SNOMED CT | 430621000 | Malignant neoplasm of lower respiratory tract |
| SNOMED CT | 433067002 | Adamantinoma of femur |

Continued on next page

**TableS24 – continued from previous page**

| CodeSystem | ConceptCode | ConceptName |
|---|---|---|
| SNOMED CT | 438946002 | Siewert type I adenocarcinoma of esophagogastric junction |
| SNOMED CT | 439478008 | Siewert type III adenocarcinoma of esophagogastric junction |
| SNOMED CT | 440173001 | Nonsquamous nonsmall cell neoplasm of lung |
| SNOMED CT | 440397000 | Primary osteosarcoma of pelvis |
| SNOMED CT | 440501006 | Siewert type II adenocarcinoma of esophagogastric junction |
| SNOMED CT | 440525003 | Primary small cell neoplasm of thymus |
| SNOMED CT | 440527006 | Primary squamous cell carcinoma of naris |
| SNOMED CT | 441559006 | Mantle cell lymphoma of spleen |
| SNOMED CT | 441962003 | Large cell lymphoma of intrapelvic lymph nodes |
| SNOMED CT | 442537007 | Non-Hodgkin lymphoma associated with Human immunodeficiency virus infection |
| SNOMED CT | 443487006 | Mantle cell lymphoma |
| SNOMED CT | 444545003 | Glioma of brainstem |
| SNOMED CT | 444597005 | Mucosa-associated lymphoid tissue (MALT) lymphoma of stomach |
| SNOMED CT | 444869007 | Cavernous hemangioma of brain |
| SNOMED CT | 444910004 | Primary mediastinal (thymic) large B-cell lymphoma |
| SNOMED CT | 444911000 | Acute myeloid leukemia with t(9:11)(p22;q23); MLLT3-MLL |
| SNOMED CT | 445227008 | Juvenile myelomonocytic leukemia |
| SNOMED CT | 445406001 | Hepatosplenic T-cell lymphoma |
| SNOMED CT | 445448008 | Acute myeloid leukemia with myelodysplasia-related changes |
| SNOMED CT | 445513004 | Intracranial cavernous hemangioma |
| SNOMED CT | 445736006 | Gastrointestinal stromal tumor of stomach |
| SNOMED CT | 446022000 | Carcinoma of uterus |
| SNOMED CT | 446076000 | Extragastrointestinal stromal tumor of peritoneum |
| SNOMED CT | 446124001 | Gastrointestinal stromal tumor of esophagus |
| SNOMED CT | 446593006 | Bizarre parosteal osteochondromatous proliferation |
| SNOMED CT | 446710005 | Low grade prostatic intraepithelial neoplasia |
| SNOMED CT | 446711009 | High grade prostatic intraepithelial neoplasia |
| SNOMED CT | 446887007 | Endometrial stromal tumor |
| SNOMED CT | 447100004 | Marginal zone lymphoma |
| SNOMED CT | 447266004 | Sarcoma of endometrium |
| SNOMED CT | 447389009 | Leiomyosarcoma of uterus |
| SNOMED CT | 447390000 | Adenosarcoma of uterus |
| SNOMED CT | 447656001 | Lymphoma of pylorus of stomach |
| SNOMED CT | 447658000 | Lymphoma of fundus of stomach |
| SNOMED CT | 447705002 | Malignant germ cell neoplasm of posterior mediastinum |
| SNOMED CT | 447707005 | Leiomyosarcoma of cardia of stomach |
| SNOMED CT | 447708000 | Malignant germ cell neoplasm of anterior mediastinum |
| SNOMED CT | 447711004 | Follicular neoplasm of thyroid |
| SNOMED CT | 447712006 | Malignant melanoma of skin of anus |
| SNOMED CT | 447738006 | Paget's disease of skin of scrotum |
| SNOMED CT | 447757002 | Angiosarcoma of cheek |
| SNOMED CT | 447766003 | Lymphoma of pyloric antrum of stomach |
| SNOMED CT | 447768002 | Follicular adenoma of ectopic thyroid tissue |
| SNOMED CT | 447781009 | Carcinoma of peritoneum |
| SNOMED CT | 447782002 | Carcinoma of female breast |

Continued on next page

**TableS24 – continued from previous page**

| CodeSystem | ConceptCode | ConceptName |
|---|---|---|
| SNOMED CT | 447783007 | Sarcoma of peritoneum |
| SNOMED CT | 447784001 | Sarcoma of axillary tail of female breast |
| SNOMED CT | 447785000 | Leiomyosarcoma of stomach |
| SNOMED CT | 447800002 | Adenocarcinoma of scrotum |
| SNOMED CT | 447805007 | Lymphoma of greater curvature of stomach |
| SNOMED CT | 447806008 | Lymphoma of cardia of stomach |
| SNOMED CT | 447949005 | Carcinoma of cheek |
| SNOMED CT | 447989004 | Non-Hodgkin's lymphoma of extranodal site |
| SNOMED CT | 448148000 | Functioning pituitary neoplasm |
| SNOMED CT | 448212009 | Anaplastic large cell lymphoma, ALK negative |
| SNOMED CT | 448213004 | Diffuse non-Hodgkin's lymphoma of prostate |
| SNOMED CT | 448214005 | Malignant epithelial neoplasm of oropharynx |
| SNOMED CT | 448215006 | Carcinoma of renal pelvis |
| SNOMED CT | 448216007 | Carcinoma of thyroid |
| SNOMED CT | 448217003 | Follicular non-Hodgkin's lymphoma of prostate |
| SNOMED CT | 448218008 | Malignant neoplasm of cerebellopontine angle |
| SNOMED CT | 448220006 | Non-Hodgkin's lymphoma of bone |
| SNOMED CT | 448221005 | Sarcoma of fibula |
| SNOMED CT | 448229007 | Leiomyosarcoma of lower esophagus |
| SNOMED CT | 448231003 | Follicular non-Hodgkin's lymphoma of nose |
| SNOMED CT | 448233000 | Malignant neoplasm of urinary organ |
| SNOMED CT | 448248006 | Malignant neoplasm of axial suprasellar region of brain |
| SNOMED CT | 448250003 | Malignant teratoma of pineal region |
| SNOMED CT | 448254007 | Non-Hodgkin's lymphoma of central nervous system |
| SNOMED CT | 448258005 | Sarcoma of mesentery |
| SNOMED CT | 448259002 | Sarcoma of posterior mediastinum |
| SNOMED CT | 448269008 | Lymphoma of lesser curvature of stomach |
| SNOMED CT | 448273006 | Malignant melanoma of skin of scrotum |
| SNOMED CT | 448298007 | Malignant melanoma of skin of penis |
| SNOMED CT | 448299004 | Malignant neoplasm of mastoid |
| SNOMED CT | 448300007 | Malignant melanoma of skin of vulva |
| SNOMED CT | 448315008 | Carcinoma of anus |
| SNOMED CT | 448317000 | Follicular non-Hodgkin's lymphoma of soft tissue |
| SNOMED CT | 448319002 | Diffuse non-Hodgkin's lymphoma of nasopharynx |
| SNOMED CT | 448354009 | Non-Hodgkin's lymphoma of intestine |
| SNOMED CT | 448371005 | Non-Hodgkin's lymphoma of nasopharynx |
| SNOMED CT | 448372003 | Non-Hodgkin's lymphoma of lung |
| SNOMED CT | 448376000 | Non-Hodgkin's lymphoma of ovary |
| SNOMED CT | 448377009 | Sarcoma of sacrum |
| SNOMED CT | 448378004 | Sarcoma of bone of foot |
| SNOMED CT | 448384001 | Non-Hodgkin's lymphoma of nose |
| SNOMED CT | 448386004 | Non-Hodgkin's lymphoma of oral cavity |
| SNOMED CT | 448387008 | Non-Hodgkin's lymphoma of testis |
| SNOMED CT | 448388003 | Sarcoma of lower outer quadrant of female breast |
| SNOMED CT | 448401007 | Choriocarcinoma of placenta |
| SNOMED CT | 448408001 | Sarcoma of upper inner quadrant of female breast |
| SNOMED CT | 448435005 | Sarcoma lower inner quadrant of female breast |

Continued on next page

**TableS24 – continued from previous page**

| CodeSystem | ConceptCode | ConceptName |
|---|---|---|
| SNOMED CT | 448436006 | Sarcoma of central portion of female breast |
| SNOMED CT | 448447004 | Non-Hodgkin's lymphoma of skin |
| SNOMED CT | 448449001 | Sarcoma of female breast |
| SNOMED CT | 448450001 | Sarcoma of omentum |
| SNOMED CT | 448451002 | Sarcoma upper outer quadrant of female breast |
| SNOMED CT | 448465000 | Diffuse non-Hodgkin's lymphoma of testis |
| SNOMED CT | 448468003 | Diffuse non-Hodgkin's lymphoma of oral cavity |
| SNOMED CT | 448509007 | Transglottic malignant neoplasm of larynx |
| SNOMED CT | 448553002 | Lymphoma of pelvis |
| SNOMED CT | 448555009 | Lymphoma of body of stomach |
| SNOMED CT | 448558006 | Malignant neoplasm of maxillofacial bone |
| SNOMED CT | 448560008 | Diffuse non-Hodgkin's lymphoma of extranodal site |
| SNOMED CT | 448561007 | Follicular non-Hodgkin's lymphoma of extranodal site |
| SNOMED CT | 448607004 | Diffuse non-Hodgkin's lymphoma of uterine cervix |
| SNOMED CT | 448609001 | Diffuse non-Hodgkin's lymphoma of ovary |
| SNOMED CT | 448663003 | Diffuse non-Hodgkin's lymphoma of stomach |
| SNOMED CT | 448665005 | Malignant epithelial neoplasm of hypopharynx |
| SNOMED CT | 448666006 | Follicular non-Hodgkin's lymphoma of bone |
| SNOMED CT | 448668007 | Malignant neoplasm of mandible |
| SNOMED CT | 448669004 | Malignant neoplasm of soft tissue of orbit |
| SNOMED CT | 448670003 | Malignant neoplasm of posterior mediastinum |
| SNOMED CT | 448672006 | Follicular non-Hodgkin's lymphoma of lung |
| SNOMED CT | 448674007 | Malignant neoplasm of parametrium |
| SNOMED CT | 448709005 | Non-Hodgkin's lymphoma of stomach |
| SNOMED CT | 448710000 | Sarcoma of bone |
| SNOMED CT | 448712008 | Sarcoma of femur |
| SNOMED CT | 448738008 | Non-Hodgkin's lymphoma of soft tissue |
| SNOMED CT | 448774004 | Non-Hodgkin's lymphoma of uterine cervix |
| SNOMED CT | 448775003 | Sarcoma of radius |
| SNOMED CT | 448776002 | Sarcoma of vertebra |
| SNOMED CT | 448863000 | Carcinoma of pineal gland |
| SNOMED CT | 448864006 | Carcinoma of ureter |
| SNOMED CT | 448865007 | Follicular non-Hodgkin's lymphoma of skin |
| SNOMED CT | 448867004 | Diffuse non-Hodgkin's lymphoma of lung |
| SNOMED CT | 448868009 | Malignant neoplasm of lateral wall of oropharynx |
| SNOMED CT | 448911008 | Torsion of uterine fibroid |
| SNOMED CT | 448952004 | Infiltrating duct carcinoma of female breast |
| SNOMED CT | 448954003 | Carcinoma of urethra |
| SNOMED CT | 448989001 | Carcinoma of brain |
| SNOMED CT | 448990005 | Carcinoma of nasal cavity |
| SNOMED CT | 448993007 | Carcinoma of lung |
| SNOMED CT | 448995000 | Follicular non-Hodgkin's lymphoma of central nervous system |
| SNOMED CT | 449034009 | Malignant neoplasm of anterior and lateral floor of mouth |
| SNOMED CT | 449053004 | Lymphoma of lower esophagus |
| SNOMED CT | 449054005 | Carcinoma of fundus of uterus |
| SNOMED CT | 449055006 | Leiomyosarcoma of cardioesophageal junction |
| SNOMED CT | 449058008 | Follicular non-Hodgkin's lymphoma of tonsil |

Continued on next page

**TableS24 – continued from previous page**

| CodeSystem | ConceptCode | ConceptName |
|---|---|---|
| SNOMED CT | 449059000 | Follicular non-Hodgkin's lymphoma of uterine cervix |
| SNOMED CT | 449063007 | Follicular non-Hodgkin's lymphoma of oral cavity |
| SNOMED CT | 449065000 | Diffuse non-Hodgkin's lymphoma of nose |
| SNOMED CT | 449066004 | Malignant neoplasm of upper respiratory tract |
| SNOMED CT | 449067008 | Malignant neoplasm of parietal pleura |
| SNOMED CT | 449072004 | Gastrointestinal lymphoma |
| SNOMED CT | 449073009 | Carcinoma of corpus uteri |
| SNOMED CT | 449074003 | Lymphoma of small bowel |
| SNOMED CT | 449075002 | Lymphoma of cardioesophageal junction |
| SNOMED CT | 449077005 | Carcinoma of maxilla |
| SNOMED CT | 449097000 | Toxic thyroid adenoma |
| SNOMED CT | 449100005 | Sarcoma of bone of pelvis |
| SNOMED CT | 449101009 | Sarcoma of sternum |
| SNOMED CT | 449108003 | Philadelphia chromosome positive chronic myelogenous leukemia |
| SNOMED CT | 449153001 | Adenocarcinoma of lower esophagus |
| SNOMED CT | 449156009 | Carcinoma of floor of mouth |
| SNOMED CT | 449173006 | Diffuse non-Hodgkin's lymphoma of tonsil |
| SNOMED CT | 449176003 | Diffuse non-Hodgkin's lymphoma of intestine |
| SNOMED CT | 449177007 | Diffuse non-Hodgkin's lymphoma of bone |
| SNOMED CT | 449206007 | Sarcoma of tibia |
| SNOMED CT | 449207003 | Sarcoma of humerus |
| SNOMED CT | 449208008 | Sarcoma of coccyx |
| SNOMED CT | 449216004 | Diffuse non-Hodgkin's lymphoma of soft tissue |
| SNOMED CT | 449217008 | Diffuse non-Hodgkin's lymphoma of skin |
| SNOMED CT | 449218003 | Lymphoma of sigmoid colon |
| SNOMED CT | 449219006 | Follicular non-Hodgkin's lymphoma of nasopharynx |
| SNOMED CT | 449220000 | Diffuse follicle center lymphoma |
| SNOMED CT | 449221001 | Diffuse non-Hodgkin's lymphoma of central nervous system |
| SNOMED CT | 449222008 | Follicular non-Hodgkin's lymphoma of stomach |
| SNOMED CT | 449223003 | Malignant neoplasm of alveolus of maxilla |
| SNOMED CT | 449224009 | Malignant neoplasm of anterior mediastinum |
| SNOMED CT | 449248000 | Nasopharyngeal carcinoma |
| SNOMED CT | 449253005 | Carcinoma of hypothalamus |
| SNOMED CT | 449254004 | Carcinoma of pharynx |
| SNOMED CT | 449259009 | Malignant neoplasm of broad ligament of uterus |
| SNOMED CT | 449260004 | Malignant neoplasm of alveolus dentalis |
| SNOMED CT | 449267001 | Sarcoma of mandible |
| SNOMED CT | 449268006 | Sarcoma of clavicle |
| SNOMED CT | 449269003 | Sarcoma of rib |
| SNOMED CT | 449292003 | Non-Hodgkin's lymphoma of tonsil |
| SNOMED CT | 449294002 | Sarcoma of scapula |
| SNOMED CT | 449295001 | Sarcoma of skull |
| SNOMED CT | 449307001 | Follicular non-Hodgkin's lymphoma of ovary |
| SNOMED CT | 449308006 | Malignant neoplasm of visceral pleura |
| SNOMED CT | 449309003 | Malignant neoplasm of skin of scrotum |
| SNOMED CT | 449318001 | Non-Hodgkin's lymphoma of prostate |
| SNOMED CT | 449377002 | Malignant neoplasm of pelvic peritoneum |

Continued on next page

**TableS24 – continued from previous page**

| CodeSystem | ConceptCode | ConceptName |
|---|---|---|
| SNOMED CT | 449386007 | Philadelphia chromosome negative chronic myelogenous leukemia |
| SNOMED CT | 449416001 | Carcinoma of skin of anus |
| SNOMED CT | 449417005 | Carcinoma of nasal septum |
| SNOMED CT | 449418000 | Follicular non-Hodgkin's lymphoma of testis |
| SNOMED CT | 449419008 | Follicular non-Hodgkin's lymphoma of intestine |
| SNOMED CT | 449420002 | Malignant neoplasm of cerebellum |
| SNOMED CT | 449472007 | Malignant epithelial neoplasm of alveolus dentalis |
| SNOMED CT | 449487002 | Carcinoma of mandible |
| SNOMED CT | 449497006 | Sarcoma of pelvic peritoneum |
| SNOMED CT | 449578008 | Malignant neoplasm of alveolus of mandible |
| SNOMED CT | 449628003 | Malignant neoplasm of long bone of lower leg |
| SNOMED CT | 449636007 | Malignant melanoma of skin of lower leg |
| SNOMED CT | 449637003 | Malignant melanoma of skin of upper arm |
| SNOMED CT | 698646006 | Acute monoblastic leukemia in remission |
| SNOMED CT | 699657009 | Hepatosplenic gamma-delta cell lymphoma |
| SNOMED CT | 699818003 | T-cell large granular lymphocytosis |
| SNOMED CT | 702446006 | Core binding factor acute myeloid leukemia |
| SNOMED CT | 702476004 | Therapy-related myelodysplastic syndrome |
| SNOMED CT | 702785000 | Large cell (Ki-1 positive) lymphoma |
| SNOMED CT | 702786004 | Follicular non-Hodgkin's lymphoma diffuse follicle center sub-type grade 1 |
| SNOMED CT | 702977001 | Follicular non-Hodgkin's lymphoma diffuse follicle center cell sub-type grade 2 |
| SNOMED CT | 703387000 | Cytogenetically normal acute myeloid leukemia |
| SNOMED CT | 703626001 | Anaplastic large cell lymphoma, T/Null cell, primary systemic type |
| SNOMED CT | 705061009 | Childhood myelodysplastic syndrome |
| SNOMED CT | 709471005 | Periodontitis co-occurrent with leukemia |
| SNOMED CT | 713325002 | Primary cerebral lymphoma co-occurrent with human immunodeficiency virus infection |
| SNOMED CT | 713483007 | Reticulosarcoma co-occurrent with human immunodeficiency virus infection |
| SNOMED CT | 713516007 | Primary effusion lymphoma |
| SNOMED CT | 713718006 | Diffuse non-Hodgkin immunoblastic lymphoma co-occurrent with human immunodeficiency virus infection |
| SNOMED CT | 713897006 | Burkitt lymphoma co-occurrent with human immunodeficiency virus infection |
| SNOMED CT | 714251006 | Philadelphia chromosome-negative precursor B-cell acute lymphoblastic leukemia |
| SNOMED CT | 714463003 | Primary effusion lymphoma co-occurrent with infection caused by Human herpesvirus 8 |
| SNOMED CT | 715664005 | Interdigitating dendritic cell sarcoma |
| SNOMED CT | 715950008 | ALK-positive large B-cell lymphoma |
| SNOMED CT | 716788007 | Epstein-Barr virus positive diffuse large B-cell lymphoma of elderly |
| SNOMED CT | 718200007 | Primary pulmonary lymphoma |

Continued on next page

**TableS24 – continued from previous page**

| CodeSystem | ConceptCode | ConceptName |
|---|---|---|
| SNOMED CT | 721302006 | Myelodysplastic/myeloproliferative neoplasm with ring sideroblasts and thrombocytosis |
| SNOMED CT | 721303001 | Refractory neutropenia |
| SNOMED CT | 721304007 | Refractory thrombocytopenia |
| SNOMED CT | 721305008 | Acute myeloid leukemia due to recurrent genetic abnormality |
| SNOMED CT | 721306009 | Therapy related acute myeloid leukemia and myelodysplastic syndrome |
| SNOMED CT | 721308005 | Acute leukemia of ambiguous lineage |
| SNOMED CT | 721310007 | Aggressive natural killer-cell leukemia |
| SNOMED CT | 721313009 | Indeterminate dendritic cell neoplasm |
| SNOMED CT | 721314003 | Fibroblastic reticular cell neoplasm |
| SNOMED CT | 721555001 | Follicular lymphoma of small intestine |
| SNOMED CT | 721695008 | Primary adenocarcinoma of ascending colon and right flexure |
| SNOMED CT | 721696009 | Primary adenocarcinoma of transverse colon |
| SNOMED CT | 721699002 | Primary adenocarcinoma of descending colon and splenic flexure |
| SNOMED CT | 721762007 | Adult T-cell leukemia/lymphoma of skin |
| SNOMED CT | 76564002 | Catecholamine secretion by pheochromocytoma |
| SNOMED CT | 80914001 | Anaplasia of cervix |
| SNOMED CT | 81021006 | Myoma of prostate |
| SNOMED CT | 91854005 | Acute leukemia in remission |
| SNOMED CT | 91855006 | Acute leukemia |
| SNOMED CT | 91856007 | Acute lymphoid leukemia in remission |
| SNOMED CT | 91857003 | Acute lymphoid leukemia |
| SNOMED CT | 91858008 | Acute monocytic leukemia in remission |
| SNOMED CT | 91860005 | Acute myeloid leukemia in remission |
| SNOMED CT | 91861009 | Acute myeloid leukemia, disease |
| SNOMED CT | 92087008 | Benign neoplasm of epicardium |
| SNOMED CT | 92752000 | Carcinoma in situ of spermatic cord |
| SNOMED CT | 92793008 | Carcinoma in situ of vas deferens |
| SNOMED CT | 92811003 | Chronic leukemia in remission |
| SNOMED CT | 92812005 | Chronic leukemia |
| SNOMED CT | 92813000 | Chronic lymphoid leukemia in remission |
| SNOMED CT | 92814006 | Chronic lymphoid leukemia, disease |
| SNOMED CT | 92817004 | Chronic myeloid leukemia in remission |
| SNOMED CT | 92818009 | Chronic myeloid leukemia |
| SNOMED CT | 93133006 | Letterer-Siwe disease of intra-abdominal lymph nodes |
| SNOMED CT | 93134000 | Letterer-Siwe disease of intrapelvic lymph nodes |
| SNOMED CT | 93135004 | Letterer-Siwe disease of intrathoracic lymph nodes |
| SNOMED CT | 93136003 | Letterer-Siwe disease of lymph nodes of axilla AND/OR upper limb |
| SNOMED CT | 93137007 | Letterer-Siwe disease of lymph nodes of head, face AND/OR neck |
| SNOMED CT | 93138002 | Letterer-Siwe disease of lymph nodes of inguinal region AND/OR lower limb |
| SNOMED CT | 93139005 | Letterer-Siwe disease of lymph nodes of multiple sites |
| SNOMED CT | 93140007 | Letterer-Siwe disease of spleen |
| SNOMED CT | 93141006 | Letterer-Siwe disease of extranodal AND/OR solid organ site |

Continued on next page

**TableS24 – continued from previous page**

| CodeSystem | ConceptCode | ConceptName |
|---|---|---|
| SNOMED CT | 93142004 | Leukemia in remission |
| SNOMED CT | 93143009 | Leukemia |
| SNOMED CT | 93144003 | Leukemic reticuloendotheliosis of intra-abdominal lymph nodes |
| SNOMED CT | 93145002 | Leukemic reticuloendotheliosis of intrapelvic lymph nodes |
| SNOMED CT | 93146001 | Leukemic reticuloendotheliosis of intrathoracic lymph nodes |
| SNOMED CT | 93150008 | Leukemic reticuloendotheliosis of lymph nodes of multiple sites |
| SNOMED CT | 93151007 | Hairy cell leukemia of spleen |
| SNOMED CT | 93152000 | Leukemic reticuloendotheliosis of extranodal AND/OR solid organ site |
| SNOMED CT | 93169003 | Lymphoid leukemia in remission |
| SNOMED CT | 93182006 | Malignant histiocytosis of intra-abdominal lymph nodes |
| SNOMED CT | 93183001 | Malignant histiocytosis of intrapelvic lymph nodes |
| SNOMED CT | 93184007 | Malignant histiocytosis of intrathoracic lymph nodes |
| SNOMED CT | 93185008 | Malignant histiocytosis of lymph nodes of axilla AND/OR upper limb |
| SNOMED CT | 93186009 | Malignant histiocytosis of lymph nodes of head, face AND/OR neck |
| SNOMED CT | 93187000 | Malignant histiocytosis of lymph nodes of inguinal region AND/OR lower limb |
| SNOMED CT | 93188005 | Malignant histiocytosis of lymph nodes of multiple sites |
| SNOMED CT | 93189002 | Malignant histiocytosis of spleen |
| SNOMED CT | 93190006 | Malignant histiocytosis of extranodal AND/OR solid organ site |
| SNOMED CT | 93191005 | Malignant lymphoma of intra-abdominal lymph nodes |
| SNOMED CT | 93192003 | Malignant lymphoma of intrapelvic lymph nodes |
| SNOMED CT | 93193008 | Malignant lymphoma of intrathoracic lymph nodes |
| SNOMED CT | 93194002 | Malignant lymphoma of lymph nodes of axilla AND/OR upper limb |
| SNOMED CT | 93195001 | Malignant lymphoma of lymph nodes of head, face AND/OR neck |
| SNOMED CT | 93196000 | Malignant lymphoma of lymph nodes of inguinal region AND/OR lower limb |
| SNOMED CT | 93197009 | Malignant lymphoma of lymph nodes of multiple sites |
| SNOMED CT | 93198004 | Malignant lymphoma of spleen |
| SNOMED CT | 93199007 | Malignant lymphoma of extranodal AND/OR solid organ site |
| SNOMED CT | 93200005 | Malignant mast cell tumor of intra-abdominal lymph nodes |
| SNOMED CT | 93201009 | Malignant mast cell tumor of intrapelvic lymph nodes |
| SNOMED CT | 93202002 | Malignant mast cell tumor of intrathoracic lymph nodes |
| SNOMED CT | 93203007 | Malignant mast cell tumor of lymph nodes of axilla AND/OR upper limb |
| SNOMED CT | 93204001 | Malignant mast cell tumor of lymph nodes of head, face AND/OR neck |
| SNOMED CT | 93205000 | Malignant mast cell tumor of lymph nodes of inguinal region AND/OR lower limb |
| SNOMED CT | 93209006 | Malignant melanoma of perianal skin |
| SNOMED CT | 93210001 | Malignant melanoma of skin of abdomen |
| SNOMED CT | 93211002 | Malignant melanoma of skin of ankle |
| SNOMED CT | 93213004 | Malignant melanoma of skin of axilla |

Continued on next page

**TableS24 – continued from previous page**

| CodeSystem | ConceptCode | ConceptName |
|---|---|---|
| SNOMED CT | 93214005 | Malignant melanoma of skin of back |
| SNOMED CT | 93215006 | Malignant melanoma of skin of breast |
| SNOMED CT | 93216007 | Malignant melanoma of skin of buttock |
| SNOMED CT | 93217003 | Malignant melanoma of skin of cheek |
| SNOMED CT | 93218008 | Malignant melanoma of skin of chest |
| SNOMED CT | 93219000 | Malignant melanoma of skin of chin |
| SNOMED CT | 93220006 | Malignant melanoma of skin of ear |
| SNOMED CT | 93221005 | Malignant melanoma of skin of elbow |
| SNOMED CT | 93222003 | Malignant melanoma of skin of external auditory canal |
| SNOMED CT | 93223008 | Malignant melanoma of skin of eyebrow |
| SNOMED CT | 93224002 | Malignant melanoma of skin of eyelid |
| SNOMED CT | 93225001 | Malignant melanoma of skin of face |
| SNOMED CT | 93226000 | Malignant melanoma of skin of finger |
| SNOMED CT | 93227009 | Malignant melanoma of skin of foot |
| SNOMED CT | 93228004 | Malignant melanoma of skin of forearm |
| SNOMED CT | 93229007 | Malignant melanoma of skin of forehead |
| SNOMED CT | 93230002 | Malignant melanoma of skin of groin |
| SNOMED CT | 93636004 | Malignant melanoma of skin of hand |
| SNOMED CT | 93637008 | Malignant melanoma of skin of hip |
| SNOMED CT | 93638003 | Malignant melanoma of skin of knee |
| SNOMED CT | 93640008 | Malignant melanoma of skin of lip |
| SNOMED CT | 93641007 | Malignant melanoma of skin of lower limb |
| SNOMED CT | 93642000 | Malignant melanoma of skin of neck |
| SNOMED CT | 93643005 | Malignant melanoma of skin of nose |
| SNOMED CT | 93644004 | Malignant melanoma of skin of perineum |
| SNOMED CT | 93645003 | Malignant melanoma of skin of popliteal area |
| SNOMED CT | 93646002 | Malignant melanoma of skin of scalp |
| SNOMED CT | 93647006 | Malignant melanoma of skin of shoulder |
| SNOMED CT | 93648001 | Malignant melanoma of skin of temporal region |
| SNOMED CT | 93649009 | Malignant melanoma of skin of thigh |
| SNOMED CT | 93650009 | Malignant melanoma of skin of toe |
| SNOMED CT | 93651008 | Malignant melanoma of skin of trunk |
| SNOMED CT | 93652001 | Malignant melanoma of skin of umbilicus |
| SNOMED CT | 93653006 | Malignant melanoma of skin of upper limb |
| SNOMED CT | 93654000 | Malignant melanoma of skin of wrist |
| SNOMED CT | 93655004 | Malignant melanoma of skin |
| SNOMED CT | 93659005 | Primary malignant neoplasm of accessory sinus |
| SNOMED CT | 93661001 | Primary malignant neoplasm of acromion |
| SNOMED CT | 93662008 | Primary malignant neoplasm of adenoid |
| SNOMED CT | 93665005 | Primary malignant neoplasm of adrenal gland |
| SNOMED CT | 93667002 | Primary malignant neoplasm of alveolar ridge mucosa |
| SNOMED CT | 93669004 | Primary malignant neoplasm of anal canal |
| SNOMED CT | 93670003 | Primary malignant neoplasm of anterior aspect of epiglottis |
| SNOMED CT | 93671004 | Primary malignant neoplasm of anterior mediastinum |
| SNOMED CT | 93672006 | Primary malignant neoplasm of anterior portion of floor of mouth |
| SNOMED CT | 93674007 | Primary malignant neoplasm of anterior wall of nasopharynx |
| SNOMED CT | 93676009 | Primary malignant neoplasm of anus |

Continued on next page

**TableS24 – continued from previous page**

| CodeSystem | ConceptCode | ConceptName |
|---|---|---|
| SNOMED CT | 93680004 | Primary malignant neoplasm of areola of female breast |
| SNOMED CT | 93684008 | Primary malignant neoplasm of axilla |
| SNOMED CT | 93687001 | Primary malignant neoplasm of base of tongue |
| SNOMED CT | 93689003 | Primary malignant neoplasm of bladder |
| SNOMED CT | 93692004 | Primary malignant neoplasm of blood vessel of axilla |
| SNOMED CT | 93710000 | Primary malignant neoplasm of blood vessel of thorax |
| SNOMED CT | 93716006 | Primary malignant neoplasm of body of penis |
| SNOMED CT | 93717002 | Primary malignant neoplasm of body of stomach |
| SNOMED CT | 93721009 | Primary malignant neoplasm of bone of face |
| SNOMED CT | 93722002 | Primary malignant neoplasm of bone of lower limb |
| SNOMED CT | 93723007 | Primary malignant neoplasm of bone of skull |
| SNOMED CT | 93724001 | Primary malignant neoplasm of bone of upper limb |
| SNOMED CT | 93725000 | Primary malignant neoplasm of bone |
| SNOMED CT | 93726004 | Primary malignant neoplasm of brain stem |
| SNOMED CT | 93727008 | Primary malignant neoplasm of brain |
| SNOMED CT | 93728003 | Primary malignant neoplasm of broad ligament |
| SNOMED CT | 93729006 | Primary malignant neoplasm of bronchus of left lower lobe |
| SNOMED CT | 93730001 | Primary malignant neoplasm of bronchus of left upper lobe |
| SNOMED CT | 93731002 | Primary malignant neoplasm of bronchus of right lower lobe |
| SNOMED CT | 93732009 | Primary malignant neoplasm of bronchus of right middle lobe |
| SNOMED CT | 93733004 | Primary malignant neoplasm of bronchus of right upper lobe |
| SNOMED CT | 93734005 | Primary malignant neoplasm of bronchus |
| SNOMED CT | 93737003 | Primary malignant neoplasm of calcaneus |
| SNOMED CT | 93738008 | Primary malignant neoplasm of cardia of stomach |
| SNOMED CT | 93739000 | Primary malignant neoplasm of carina |
| SNOMED CT | 93741004 | Primary malignant neoplasm of carpal bone |
| SNOMED CT | 93742006 | Primary malignant neoplasm of cartilage of nose |
| SNOMED CT | 93744007 | Primary malignant neoplasm of central nervous system |
| SNOMED CT | 93745008 | Primary malignant neoplasm of central portion of female breast |
| SNOMED CT | 93746009 | Primary malignant neoplasm of cerebellum |
| SNOMED CT | 93747000 | Primary malignant neoplasm of cerebral meninges |
| SNOMED CT | 93748005 | Primary malignant neoplasm of cerebral ventricle |
| SNOMED CT | 93749002 | Primary malignant neoplasm of cerebrum |
| SNOMED CT | 93753000 | Primary malignant neoplasm of cheek |
| SNOMED CT | 93754006 | Primary malignant neoplasm of chest wall |
| SNOMED CT | 93755007 | Primary malignant neoplasm of choroid |
| SNOMED CT | 93756008 | Primary malignant neoplasm of ciliary body (primary) |
| SNOMED CT | 93757004 | Primary malignant neoplasm of clavicle |
| SNOMED CT | 93760006 | Primary malignant neoplasm of coccyx |
| SNOMED CT | 93761005 | Primary malignant neoplasm of colon |
| SNOMED CT | 93764002 | Primary malignant neoplasm of conjunctiva of eye |
| SNOMED CT | 93766000 | Primary malignant neoplasm of cornea of eye |
| SNOMED CT | 93768004 | Primary malignant neoplasm of craniopharyngeal duct |
| SNOMED CT | 93769007 | Primary malignant neoplasm of cuboid |
| SNOMED CT | 93771007 | Primary malignant neoplasm of descending colon |
| SNOMED CT | 93772000 | Primary malignant neoplasm of diaphragm |
| SNOMED CT | 93773005 | Primary malignant neoplasm of dorsal surface of tongue |

Continued on next page

**TableS24 – continued from previous page**

| CodeSystem | ConceptCode | ConceptName |
|---|---|---|
| SNOMED CT | 93775003 | Primary malignant neoplasm of duodenum |
| SNOMED CT | 93776002 | Primary malignant neoplasm of ectopic female breast tissue |
| SNOMED CT | 93778001 | Primary malignant neoplasm of endocardium |
| SNOMED CT | 93779009 | Primary malignant neoplasm of endocervix |
| SNOMED CT | 93781006 | Primary malignant neoplasm of endometrium |
| SNOMED CT | 93782004 | Primary malignant neoplasm of epicardium |
| SNOMED CT | 93783009 | Primary malignant neoplasm of epididymis |
| SNOMED CT | 93784003 | Primary malignant neoplasm of epiglottis |
| SNOMED CT | 93786001 | Primary malignant neoplasm of ethmoid bone |
| SNOMED CT | 93787005 | Primary malignant neoplasm of ethmoidal sinus |
| SNOMED CT | 93788000 | Primary malignant neoplasm of eustachian tube |
| SNOMED CT | 93789008 | Primary malignant neoplasm of exocervix |
| SNOMED CT | 93796005 | Primary malignant neoplasm of female breast |
| SNOMED CT | 93797001 | Primary malignant neoplasm of female genital organ |
| SNOMED CT | 93798006 | Primary malignant neoplasm of femur |
| SNOMED CT | 93799003 | Primary malignant neoplasm of fibula |
| SNOMED CT | 93800004 | Primary malignant neoplasm of first cuneiform bone of foot |
| SNOMED CT | 93802007 | Primary malignant neoplasm of floor of mouth |
| SNOMED CT | 93806005 | Primary malignant neoplasm of frontal bone |
| SNOMED CT | 93807001 | Primary malignant neoplasm of frontal lobe |
| SNOMED CT | 93808006 | Primary malignant neoplasm of frontal sinus |
| SNOMED CT | 93809003 | Primary malignant neoplasm of fundus of stomach |
| SNOMED CT | 93812000 | Primary malignant neoplasm of gingival mucosa |
| SNOMED CT | 93816002 | Primary malignant neoplasm of glottis |
| SNOMED CT | 93818001 | Primary malignant neoplasm of greater curvature of stomach |
| SNOMED CT | 93820003 | Primary malignant neoplasm of hamate bone |
| SNOMED CT | 93825008 | Primary malignant neoplasm of heart |
| SNOMED CT | 93826009 | Primary malignant neoplasm of hepatic flexure of colon |
| SNOMED CT | 93827000 | Primary malignant neoplasm of hilus of lung |
| SNOMED CT | 93829002 | Primary malignant neoplasm of hypopharyngeal aspect of aryepiglottic fold |
| SNOMED CT | 93830007 | Primary malignant neoplasm of hypopharyngeal aspect of interarytenoid fold |
| SNOMED CT | 93831006 | Primary malignant neoplasm of hypopharynx |
| SNOMED CT | 93833009 | Primary malignant neoplasm of ilium |
| SNOMED CT | 93835002 | Primary malignant neoplasm of inner aspect of lip |
| SNOMED CT | 93836001 | Primary malignant neoplasm of inner aspect of lower lip |
| SNOMED CT | 93837005 | Primary malignant neoplasm of inner aspect of upper lip |
| SNOMED CT | 93841009 | Primary malignant neoplasm of intrathoracic organs |
| SNOMED CT | 93842002 | Primary malignant neoplasm of ischium |
| SNOMED CT | 93843007 | Primary malignant neoplasm of islets of Langerhans |
| SNOMED CT | 93844001 | Primary malignant neoplasm of isthmus of uterus |
| SNOMED CT | 93845000 | Primary malignant neoplasm of jaw |
| SNOMED CT | 93848003 | Primary malignant neoplasm of junctional zone of tongue |
| SNOMED CT | 93849006 | Primary malignant neoplasm of kidney |
| SNOMED CT | 93850006 | Primary malignant neoplasm of labia majora |
| SNOMED CT | 93851005 | Primary malignant neoplasm of labia minora |

Continued on next page

**TableS24 – continued from previous page**

| CodeSystem | ConceptCode | ConceptName |
|---|---|---|
| SNOMED CT | 93857009 | Primary malignant neoplasm of laryngeal commissure |
| SNOMED CT | 93858004 | Primary malignant neoplasm of laryngeal surface of epiglottis |
| SNOMED CT | 93860002 | Primary malignant neoplasm of lateral portion of floor of mouth |
| SNOMED CT | 93861003 | Primary malignant neoplasm of lateral wall of nasopharynx |
| SNOMED CT | 93862005 | Primary malignant neoplasm of lateral wall of oropharynx |
| SNOMED CT | 93864006 | Primary malignant neoplasm of lower lobe of left lung |
| SNOMED CT | 93865007 | Primary malignant neoplasm of left upper lobe of lung |
| SNOMED CT | 93867004 | Primary malignant neoplasm of lesser curvature of stomach |
| SNOMED CT | 93868009 | Primary malignant neoplasm of lingual tonsil |
| SNOMED CT | 93871001 | Primary malignant neoplasm of long bone of lower limb |
| SNOMED CT | 93872008 | Primary malignant neoplasm of long bone of upper limb |
| SNOMED CT | 93874009 | Primary malignant neoplasm of lower inner quadrant of female breast |
| SNOMED CT | 93876006 | Primary malignant neoplasm of lower outer quadrant of female breast |
| SNOMED CT | 93879004 | Primary malignant neoplasm of lunate bone |
| SNOMED CT | 93880001 | Primary malignant neoplasm of lung |
| SNOMED CT | 93882009 | Primary malignant neoplasm of main bronchus |
| SNOMED CT | 93883004 | Primary malignant neoplasm of major salivary gland |
| SNOMED CT | 93885006 | Primary malignant neoplasm of male genital organ |
| SNOMED CT | 93886007 | Primary malignant neoplasm of mandible |
| SNOMED CT | 93887003 | Primary malignant neoplasm of mastoid air cells |
| SNOMED CT | 93888008 | Primary malignant neoplasm of maxilla |
| SNOMED CT | 93889000 | Primary malignant neoplasm of maxillary sinus |
| SNOMED CT | 93891008 | Primary malignant neoplasm of mediastinum |
| SNOMED CT | 93892001 | Primary malignant neoplasm of metacarpal bone |
| SNOMED CT | 93893006 | Primary malignant neoplasm of metatarsal bone |
| SNOMED CT | 93894000 | Primary malignant neoplasm of middle ear |
| SNOMED CT | 93898002 | Primary malignant neoplasm of multiple endocrine glands |
| SNOMED CT | 93910009 | Primary malignant neoplasm of muscle of thorax |
| SNOMED CT | 93915004 | Primary malignant neoplasm of myometrium |
| SNOMED CT | 93916003 | Primary malignant neoplasm of nasal bone |
| SNOMED CT | 93917007 | Primary malignant neoplasm of nasal cavity |
| SNOMED CT | 93918002 | Primary malignant neoplasm of nasal concha |
| SNOMED CT | 93920004 | Primary malignant neoplasm of navicular bone of foot |
| SNOMED CT | 93924008 | Primary malignant neoplasm of nipple of female breast |
| SNOMED CT | 93927001 | Primary malignant neoplasm of occipital bone |
| SNOMED CT | 93928006 | Primary malignant neoplasm of occipital lobe |
| SNOMED CT | 93930008 | Primary malignant neoplasm of olfactory nerve |
| SNOMED CT | 93931007 | Primary malignant neoplasm of optic nerve |
| SNOMED CT | 93932000 | Primary malignant neoplasm of orbit |
| SNOMED CT | 93933005 | Primary malignant neoplasm of oropharynx |
| SNOMED CT | 93934004 | Primary malignant neoplasm of ovary |
| SNOMED CT | 93936002 | Primary malignant neoplasm of palatine bone |
| SNOMED CT | 93942003 | Primary malignant neoplasm of parametrium |
| SNOMED CT | 93943008 | Primary malignant neoplasm of parathyroid gland |
| SNOMED CT | 93944002 | Primary malignant neoplasm of paraurethral glands |

Continued on next page

**TableS24 – continued from previous page**

| CodeSystem | ConceptCode | ConceptName |
|---|---|---|
| SNOMED CT | 93945001 | Primary malignant neoplasm of parietal bone |
| SNOMED CT | 93946000 | Primary malignant neoplasm of parietal lobe |
| SNOMED CT | 93947009 | Primary malignant neoplasm of parietal peritoneum |
| SNOMED CT | 93948004 | Primary malignant neoplasm of parietal pleura |
| SNOMED CT | 93950007 | Primary malignant neoplasm of patella |
| SNOMED CT | 93951006 | Primary malignant neoplasm of pelvic bone |
| SNOMED CT | 93952004 | Primary malignant neoplasm of pelvic peritoneum |
| SNOMED CT | 93955002 | Primary malignant neoplasm of periadrenal tissue |
| SNOMED CT | 93957005 | Primary malignant neoplasm of pericardium |
| SNOMED CT | 93958000 | Primary malignant neoplasm of perirenal tissue |
| SNOMED CT | 93959008 | Primary malignant neoplasm of phalanx of foot |
| SNOMED CT | 93960003 | Primary malignant neoplasm of phalanx of hand |
| SNOMED CT | 93961004 | Primary malignant neoplasm of pharynx |
| SNOMED CT | 93962006 | Primary malignant neoplasm of pineal gland |
| SNOMED CT | 93963001 | Primary malignant neoplasm of pisiform bone of hand |
| SNOMED CT | 93964007 | Primary malignant neoplasm of pituitary gland |
| SNOMED CT | 93966009 | Primary malignant neoplasm of pleura |
| SNOMED CT | 93967000 | Primary malignant neoplasm of postcricoid region |
| SNOMED CT | 93968005 | Primary malignant neoplasm of posterior hypopharyngeal wall |
| SNOMED CT | 93969002 | Primary malignant neoplasm of posterior mediastinum |
| SNOMED CT | 93970001 | Primary malignant neoplasm of posterior wall of nasopharynx |
| SNOMED CT | 93971002 | Primary malignant neoplasm of posterior wall of oropharynx |
| SNOMED CT | 93974005 | Primary malignant neoplasm of prostate |
| SNOMED CT | 93975006 | Primary malignant neoplasm of pubis |
| SNOMED CT | 93976007 | Primary malignant neoplasm of pyloric antrum |
| SNOMED CT | 93977003 | Primary malignant neoplasm of pylorus |
| SNOMED CT | 93978008 | Primary malignant neoplasm of pyriform sinus |
| SNOMED CT | 93979000 | Primary malignant neoplasm of radius |
| SNOMED CT | 93980002 | Primary malignant neoplasm of rectosigmoid junction |
| SNOMED CT | 93981003 | Primary malignant neoplasm of rectouterine pouch |
| SNOMED CT | 93982005 | Primary malignant neoplasm of rectovaginal septum |
| SNOMED CT | 93983000 | Primary malignant neoplasm of rectovesical septum |
| SNOMED CT | 93985007 | Primary malignant neoplasm of renal pelvis |
| SNOMED CT | 93986008 | Primary malignant neoplasm of respiratory tract |
| SNOMED CT | 93987004 | Primary malignant neoplasm of retina |
| SNOMED CT | 93989001 | Primary malignant neoplasm of retromolar area |
| SNOMED CT | 93990005 | Primary malignant neoplasm of rib |
| SNOMED CT | 93991009 | Primary malignant neoplasm of right lower lobe of lung |
| SNOMED CT | 93992002 | Primary malignant neoplasm of right middle lobe of lung |
| SNOMED CT | 93993007 | Primary malignant neoplasm of upper lobe of right lung |
| SNOMED CT | 93994001 | Primary malignant neoplasm of round ligament of uterus |
| SNOMED CT | 93996004 | Primary malignant neoplasm of sacrum |
| SNOMED CT | 93997008 | Primary malignant neoplasm of scapula |
| SNOMED CT | 93998003 | Primary malignant neoplasm of sclera of eye |
| SNOMED CT | 94001007 | Primary malignant neoplasm of second cuneiform bone of foot |
| SNOMED CT | 94002000 | Primary malignant neoplasm of septum of nose |
| SNOMED CT | 94003005 | Primary malignant neoplasm of short bone of lower limb |

Continued on next page

**TableS24 – continued from previous page**

| CodeSystem | ConceptCode | ConceptName |
|---|---|---|
| SNOMED CT | 94006002 | Primary malignant neoplasm of sigmoid colon |
| SNOMED CT | 94010004 | Primary malignant neoplasm of skin of axilla |
| SNOMED CT | 94014008 | Primary malignant neoplasm of skin of cheek |
| SNOMED CT | 94015009 | Primary malignant neoplasm of skin of chest |
| SNOMED CT | 94020009 | Primary malignant neoplasm of skin of eyebrow |
| SNOMED CT | 94021008 | Primary malignant neoplasm of skin of eyelid |
| SNOMED CT | 94049001 | Primary malignant neoplasm of soft palate |
| SNOMED CT | 94051002 | Primary malignant neoplasm of soft tissues of axilla |
| SNOMED CT | 94062002 | Primary malignant neoplasm of soft tissues of thorax |
| SNOMED CT | 94066004 | Primary malignant neoplasm of sphenoid bone |
| SNOMED CT | 94067008 | Primary malignant neoplasm of sphenoidal sinus |
| SNOMED CT | 94068003 | Primary malignant neoplasm of spinal cord |
| SNOMED CT | 94069006 | Primary malignant neoplasm of spinal meninges |
| SNOMED CT | 94071006 | Primary malignant neoplasm of spleen |
| SNOMED CT | 94072004 | Primary malignant neoplasm of splenic flexure of colon |
| SNOMED CT | 94073009 | Primary malignant neoplasm of sternum |
| SNOMED CT | 94075002 | Primary malignant neoplasm of subglottis |
| SNOMED CT | 94076001 | Primary malignant neoplasm of sublingual gland |
| SNOMED CT | 94077005 | Primary malignant neoplasm of submaxillary gland |
| SNOMED CT | 94078000 | Primary malignant neoplasm of superior wall of nasopharynx |
| SNOMED CT | 94080006 | Primary malignant neoplasm of supraglottis |
| SNOMED CT | 94083008 | Primary malignant neoplasm of talus |
| SNOMED CT | 94084002 | Primary malignant neoplasm of tarsal bone |
| SNOMED CT | 94085001 | Primary malignant neoplasm of temporal bone |
| SNOMED CT | 94086000 | Primary malignant neoplasm of temporal lobe |
| SNOMED CT | 94087009 | Primary malignant neoplasm of testis |
| SNOMED CT | 94089007 | Primary malignant neoplasm of the mesocolon |
| SNOMED CT | 94090003 | Primary malignant neoplasm of omentum |
| SNOMED CT | 94094007 | Primary malignant neoplasm of third cuneiform bone of foot |
| SNOMED CT | 94096009 | Primary malignant neoplasm of thymus |
| SNOMED CT | 94098005 | Primary malignant neoplasm of thyroid gland |
| SNOMED CT | 94099002 | Primary malignant neoplasm of tibia |
| SNOMED CT | 94101009 | Primary malignant neoplasm of tongue |
| SNOMED CT | 94102002 | Primary malignant neoplasm of tonsillar fossa |
| SNOMED CT | 94103007 | Primary malignant neoplasm of tonsillar pillar |
| SNOMED CT | 94104001 | Primary malignant neoplasm of trachea |
| SNOMED CT | 94105000 | Primary malignant neoplasm of transverse colon |
| SNOMED CT | 94107008 | Primary malignant neoplasm of trapezoid bone |
| SNOMED CT | 94109006 | Primary malignant neoplasm of trigone of urinary bladder |
| SNOMED CT | 94112009 | Primary malignant neoplasm of ulna |
| SNOMED CT | 94113004 | Primary malignant neoplasm of undescended testis |
| SNOMED CT | 94115006 | Primary malignant neoplasm of upper inner quadrant of female breast |
| SNOMED CT | 94117003 | Primary malignant neoplasm of upper outer quadrant of female breast |
| SNOMED CT | 94118008 | Primary malignant neoplasm of upper respiratory tract |
| SNOMED CT | 94120006 | Primary malignant neoplasm of urachus |

Continued on next page

**TableS24 – continued from previous page**

| CodeSystem | ConceptCode | ConceptName |
|---|---|---|
| SNOMED CT | 94121005 | Primary malignant neoplasm of ureter |
| SNOMED CT | 94122003 | Primary malignant neoplasm of ureteric orifice of urinary bladder |
| SNOMED CT | 94123008 | Primary malignant neoplasm of urethra |
| SNOMED CT | 94124002 | Primary malignant neoplasm of urinary bladder neck |
| SNOMED CT | 94125001 | Primary malignant neoplasm of urinary system |
| SNOMED CT | 94128004 | Primary malignant neoplasm of uveal tract of eye |
| SNOMED CT | 94129007 | Primary malignant neoplasm of uvula |
| SNOMED CT | 94132005 | Primary malignant neoplasm of vallecula |
| SNOMED CT | 94133000 | Primary malignant neoplasm of vas deferens |
| SNOMED CT | 94134006 | Primary malignant neoplasm of ventral surface of tongue |
| SNOMED CT | 94135007 | Primary malignant neoplasm of vermilion border of lip |
| SNOMED CT | 94138009 | Primary malignant neoplasm of vestibule of mouth |
| SNOMED CT | 94139001 | Primary malignant neoplasm of vestibule of nose |
| SNOMED CT | 94140004 | Primary malignant neoplasm of visceral pleura |
| SNOMED CT | 94142007 | Primary malignant neoplasm of vomer |
| SNOMED CT | 94143002 | Primary malignant neoplasm of vulva |
| SNOMED CT | 94145009 | Primary malignant neoplasm of zygomatic bone |
| SNOMED CT | 94148006 | Megakaryocytic leukemia in remission |
| SNOMED CT | 94179005 | Secondary malignant neoplasm of ascending colon |
| SNOMED CT | 94260004 | Secondary malignant neoplasm of colon |
| SNOMED CT | 94271003 | Secondary malignant neoplasm of descending colon |
| SNOMED CT | 94328005 | Secondary malignant neoplasm of hepatic flexure of colon |
| SNOMED CT | 94509004 | Secondary malignant neoplasm of rectosigmoid junction |
| SNOMED CT | 94538001 | Secondary malignant neoplasm of sigmoid colon |
| SNOMED CT | 94604000 | Secondary malignant neoplasm of splenic flexure of colon |
| SNOMED CT | 94643001 | Secondary malignant neoplasm of transverse colon |
| SNOMED CT | 94686001 | Mixed cell type lymphosarcoma of intra-abdominal lymph nodes |
| SNOMED CT | 94687005 | Mixed cell type lymphosarcoma of intrapelvic lymph nodes |
| SNOMED CT | 94688000 | Mixed cell type lymphosarcoma of intrathoracic lymph nodes |
| SNOMED CT | 94690004 | Mixed cell type lymphosarcoma of lymph nodes of head, face, and neck |
| SNOMED CT | 94707004 | Mycosis fungoides of intra-abdominal lymph nodes |
| SNOMED CT | 94708009 | Mycosis fungoides of intrapelvic lymph nodes |
| SNOMED CT | 94709001 | Mycosis fungoides of intrathoracic lymph nodes |
| SNOMED CT | 94710006 | Mycosis fungoides of lymph nodes of axilla AND/OR upper limb |
| SNOMED CT | 94711005 | Mycosis fungoides of lymph nodes of head, face AND/OR neck |
| SNOMED CT | 94714002 | Mycosis fungoides of spleen |
| SNOMED CT | 94715001 | Mycosis fungoides of extranodal AND/OR solid organ site |
| SNOMED CT | 94716000 | Myeloid leukemia in remission |
| SNOMED CT | 94718004 | Myeloid sarcoma in remission |
| SNOMED CT | 94726007 | Neoplasm of uncertain behavior of acromion |
| SNOMED CT | 94745003 | Neoplasm of uncertain behavior of areola of female breast |
| SNOMED CT | 94750009 | Neoplasm of uncertain behavior of axillary tail of female breast |
| SNOMED CT | 95186006 | Nodular lymphoma of intra-abdominal lymph nodes |
| SNOMED CT | 95187002 | Nodular lymphoma of intrapelvic lymph nodes |
| SNOMED CT | 95188007 | Nodular lymphoma of intrathoracic lymph nodes |
| SNOMED CT | 95192000 | Nodular lymphoma of lymph nodes of multiple sites |

Continued on next page

**TableS24 – continued from previous page**

| CodeSystem | ConceptCode | ConceptName |
|---|---|---|
| SNOMED CT | 95193005 | Nodular lymphoma of spleen |
| SNOMED CT | 95194004 | Nodular lymphoma of extranodal AND/OR solid organ site |
| SNOMED CT | 95209008 | Plasma cell leukemia in remission |
| SNOMED CT | 95210003 | Plasma cell leukemia |
| SNOMED CT | 95224004 | Reticulosarcoma of intra-abdominal lymph nodes |
| SNOMED CT | 95225003 | Reticulosarcoma of intrapelvic lymph nodes |
| SNOMED CT | 95226002 | Reticulosarcoma of intrathoracic lymph nodes |
| SNOMED CT | 95230004 | Reticulosarcoma of lymph nodes of multiple sites |
| SNOMED CT | 95231000 | Reticulosarcoma of spleen |
| SNOMED CT | 95260009 | Sézary's disease of lymph nodes of head, face AND/OR neck |
| SNOMED CT | 95263006 | Sézary's disease of spleen |
| SNOMED CT | 95264000 | Sézary's disease of extranodal AND/OR solid organ site |
| SNOMED CT | 95413004 | Nodular tenosynovitis |
| SNOMED CT | 109267002 | Overlapping malignant melanoma of skin |
| SNOMED CT | 109348004 | Overlapping malignant neoplasm of bone and articular cartilage of limb |
| SNOMED CT | 109351006 | Lipoma of skin and subcutaneous tissue of limb |
| SNOMED CT | 109366009 | Overlapping malignant neoplasm of accessory sinuses |
| SNOMED CT | 109367000 | Overlapping malignant neoplasm of nasopharynx |
| SNOMED CT | 109368005 | Malignant neoplasm of overlapping sites of hypopharynx |
| SNOMED CT | 109369002 | Malignant neoplasm of overlapping sites of larynx |
| SNOMED CT | 109370001 | Primary malignant neoplasm of laryngeal cartilage |
| SNOMED CT | 109371002 | Malignant neoplasm of overlapping sites of bronchus and lung |
| SNOMED CT | 109374005 | Overlapping malignant neoplasm of mediastinum and pleura |
| SNOMED CT | 109383000 | Malignant mesothelioma of pericardium |
| SNOMED CT | 109384006 | Malignant neoplasm of overlapping sites of heart mediastinum and pleura |
| SNOMED CT | 109822001 | Overlapping malignant neoplasm of lip |
| SNOMED CT | 109823006 | Malignant neoplasm of overlapping sites of tongue |
| SNOMED CT | 109824000 | Malignant neoplasm of overlapping sites of major salivary gland |
| SNOMED CT | 109828002 | Primary malignant neoplasm of salivary gland duct |
| SNOMED CT | 109830000 | Malignant neoplasm of overlapping sites of floor of mouth |
| SNOMED CT | 109831001 | Malignant neoplasm of overlapping sites of palate |
| SNOMED CT | 109832008 | Overlapping malignant neoplasm of oropharynx |
| SNOMED CT | 109835005 | Malignant neoplasm of overlapping sites of esophagus |
| SNOMED CT | 109836006 | Malignant neoplasm of overlapping sites of stomach |
| SNOMED CT | 109838007 | Malignant neoplasm of overlapping sites of colon |
| SNOMED CT | 109839004 | Overlapping malignant neoplasm of rectum, anus and anal canal |
| SNOMED CT | 109840002 | Primary malignant neoplasm of cloacogenic zone |
| SNOMED CT | 109851002 | Overlapping malignant neoplasm of retroperitoneum and peritoneum |
| SNOMED CT | 109853004 | Mesothelioma of peritoneum |
| SNOMED CT | 109854005 | Mesothelioma of parietal peritoneum |
| SNOMED CT | 109855006 | Mesothelioma of pelvic peritoneum |
| SNOMED CT | 109856007 | Mesothelioma of mesentery |
| SNOMED CT | 109857003 | Mesothelioma of mesocolon |
| SNOMED CT | 109858008 | Mesothelioma of omentum |

Continued on next page

**TableS24 – continued from previous page**

| CodeSystem | ConceptCode | ConceptName |
|---|---|---|
| SNOMED CT | 109870007 | Overlapping malignant neoplasm of urinary system |
| SNOMED CT | 109874003 | Malignant neoplasm of overlapping sites of male genital organs |
| SNOMED CT | 109875002 | Malignant neoplasm of overlapping sites of penis |
| SNOMED CT | 109876001 | Primary malignant neoplasm of descended testis |
| SNOMED CT | 109878000 | Overlapping malignant neoplasm of female genital organs |
| SNOMED CT | 109879008 | Overlapping malignant neoplasm of body of uterus |
| SNOMED CT | 109880006 | Overlapping malignant neoplasm of uterine cervix |
| SNOMED CT | 109882003 | Primary malignant neoplasm of fundus uteri |
| SNOMED CT | 109886000 | Overlapping malignant neoplasm of female breast |
| SNOMED CT | 109911004 | Overlapping malignant neoplasm of brain and other parts of the central nervous system |
| SNOMED CT | 109912006 | Malignant neoplasm of overlapping sites of brain |
| SNOMED CT | 109915008 | Primary malignant neoplasm of meninges |
| SNOMED CT | 109941002 | Primary malignant neoplasm of peripheral nerves of thorax |
| SNOMED CT | 109948008 | Malignant neoplasm of overlapping sites of eye and adnexa |
| SNOMED CT | 109951001 | Overlapping malignant neoplasm of multiple endocrine glands |
| SNOMED CT | 109975001 | T-zone lymphoma (clinical) |
| SNOMED CT | 109976000 | Lymphoepithelioid lymphoma (clinical) |
| SNOMED CT | 109978004 | T-cell lymphoma (clinical) |
| SNOMED CT | 109980005 | Malignant immunoproliferative disease (clinical) |
| SNOMED CT | 109988003 | Histiocytic sarcoma (clinical) |
| SNOMED CT | 109994006 | Essential thrombocythemia |
| SNOMED CT | 109995007 | Myelodysplastic syndrome (clinical) |
| SNOMED CT | 109996008 | Refractory anemia (clinical) |
| SNOMED CT | 109998009 | Refractory anemia with ringed sideroblasts associated with marked thrombocytosis |
| SNOMED CT | 110000005 | Refractory anemia with excess blasts in transformation (clinical) |
| SNOMED CT | 110002002 | Mast cell leukemia (clinical) |
| SNOMED CT | 110006004 | Prolymphocytic leukemia (clinical) |
| SNOMED CT | 110013004 | Overlapping malignant neoplasm of tonsil |
| SNOMED CT | 118607005 | Lymphocyte-rich classical Hodgkin lymphoma |
| SNOMED CT | 118611004 | Sézary's disease (clinical) |
| SNOMED CT | 118612006 | Malignant histiocytosis (clinical) |
| SNOMED CT | 118615008 | Malignant mast cell tumor (clinical) |
| SNOMED CT | 118618005 | Mycosis fungoides (clinical) |
| SNOMED CT | 123842006 | Endocervical adenocarcinoma |
| SNOMED CT | 126948004 | Cerebellopontine angle meningioma |
| SNOMED CT | 127070008 | Malignant histiocytic disorder |
| SNOMED CT | 127220001 | Malignant lymphoma of lymph nodes |
| SNOMED CT | 127225006 | Chronic myelomonocytic leukemia |
| SNOMED CT | 134312002 | Odontogenic ghost cell carcinoma |
| SNOMED CT | 187601000 | Malignant neoplasm of upper lip, lipstick area |
| SNOMED CT | 187604008 | Malignant neoplasm of lower lip, external |
| SNOMED CT | 187606005 | Malignant tumor of upper labial mucosa |
| SNOMED CT | 187608006 | Malignant tumor of frenum of upper lip |
| SNOMED CT | 187613005 | Malignant neoplasm of lower lip, buccal aspect |
| SNOMED CT | 187614004 | Malignant tumor of frenum of lower lip |

Continued on next page

**TableS24 – continued from previous page**

| CodeSystem | ConceptCode | ConceptName |
|---|---|---|
| SNOMED CT | 187622006 | Malignant tumor of labial mucosa |
| SNOMED CT | 187631006 | Malignant neoplasm of base of tongue dorsal surface |
| SNOMED CT | 187634003 | Malignant tumor of anterior two-thirds of tongue - dorsal surface |
| SNOMED CT | 187635002 | Malignant neoplasm of midline of tongue |
| SNOMED CT | 187637005 | Malignant neoplasm of tongue, tip and lateral border |
| SNOMED CT | 187640005 | Malignant tumor of anterior two-thirds of tongue - ventral surface |
| SNOMED CT | 187641009 | Malignant tumor of frenum linguae |
| SNOMED CT | 187644001 | Malignant tumor of junctional zone of tongue |
| SNOMED CT | 187652003 | Malignant tumor of anterior floor of mouth |
| SNOMED CT | 187653008 | Malignant tumor of lateral floor of mouth |
| SNOMED CT | 187658004 | Malignant tumor of vestibule of mouth |
| SNOMED CT | 187659007 | Malignant tumor of upper buccal sulcus |
| SNOMED CT | 187660002 | Malignant tumor of lower buccal sulcus |
| SNOMED CT | 187661003 | Malignant tumor of upper labial sulcus |
| SNOMED CT | 187662005 | Malignant tumor of lower labial sulcus |
| SNOMED CT | 187666008 | Malignant neoplasm of junction of hard and soft palate |
| SNOMED CT | 187675005 | Malignant tumor of tonsillar pillar |
| SNOMED CT | 187681002 | Malignant neoplasm of anterior epiglottis |
| SNOMED CT | 187682009 | Malignant neoplasm of epiglottis, free border |
| SNOMED CT | 187683004 | Malignant neoplasm of glossoepiglottic fold |
| SNOMED CT | 187685006 | Malignant neoplasm of junctional region of epiglottis |
| SNOMED CT | 187688008 | Malignant tumor of posterior wall of oropharynx |
| SNOMED CT | 187692001 | Malignant tumor of nasopharynx |
| SNOMED CT | 187693006 | Malignant tumor of posterior wall of nasopharynx |
| SNOMED CT | 187694000 | Malignant tumor of adenoid |
| SNOMED CT | 187697007 | Malignant tumor of pharyngeal recess |
| SNOMED CT | 187698002 | Malignant tumor of opening of auditory tube |
| SNOMED CT | 187700006 | Malignant tumor of anterior wall of nasopharynx |
| SNOMED CT | 187701005 | Malignant neoplasm of floor of nasopharynx |
| SNOMED CT | 187708004 | Malignant tumor aryepiglottic fold - hypopharyngeal aspect |
| SNOMED CT | 187709007 | Malignant neoplasm of posterior pharynx |
| SNOMED CT | 187722004 | Malignant tumor of cervical part of esophagus |
| SNOMED CT | 187723009 | Malignant tumor of thoracic part of esophagus |
| SNOMED CT | 187724003 | Malignant tumor of abdominal part of esophagus |
| SNOMED CT | 187725002 | Malignant tumor of upper third of esophagus |
| SNOMED CT | 187726001 | Malignant tumor of middle third of esophagus |
| SNOMED CT | 187727005 | Malignant tumor of lower third of esophagus |
| SNOMED CT | 187732006 | Malignant tumor of cardia |
| SNOMED CT | 187733001 | Malignant neoplasm of cardiac orifice of stomach |
| SNOMED CT | 187736009 | Malignant tumor of pylorus |
| SNOMED CT | 187738005 | Malignant neoplasm of pyloric canal of stomach |
| SNOMED CT | 187740000 | Malignant tumor of pyloric antrum |
| SNOMED CT | 187741001 | Malignant tumor of fundus of stomach |
| SNOMED CT | 187742008 | Malignant tumor of body of stomach |
| SNOMED CT | 187757001 | Malignant neoplasm, overlapping lesion of colon |
| SNOMED CT | 187786003 | Malignant neoplasm of sphincter of Oddi |

Continued on next page

**TableS24 – continued from previous page**

| CodeSystem | ConceptCode | ConceptName |
|---|---|---|
| SNOMED CT | 187794005 | Malignant tumor of Islets of Langerhans |
| SNOMED CT | 187801002 | Malignant tumor of peritoneum and retroperitoneum |
| SNOMED CT | 187803004 | Malignant neoplasm of perinephric tissue |
| SNOMED CT | 187809000 | Malignant neoplasm of mesocolon |
| SNOMED CT | 187810005 | Malignant neoplasm of mesocecum |
| SNOMED CT | 187811009 | Malignant neoplasm of mesorectum |
| SNOMED CT | 187814001 | Malignant neoplasm of the pouch of Douglas |
| SNOMED CT | 187822008 | Fibrosarcoma of spleen |
| SNOMED CT | 187829004 | Malignant neoplasm of cartilage of nose |
| SNOMED CT | 187830009 | Malignant neoplasm of nasal conchae |
| SNOMED CT | 187831008 | Malignant tumor of nasal vestibule |
| SNOMED CT | 187833006 | Malignant neoplasm of auditory tube, middle ear and mastoid air cells |
| SNOMED CT | 187834000 | Malignant tumor of Eustachian tube |
| SNOMED CT | 187835004 | Malignant tumor of tympanic cavity |
| SNOMED CT | 187836003 | Malignant tumor of tympanic antrum |
| SNOMED CT | 187841006 | Malignant tumor of glottis |
| SNOMED CT | 187842004 | Malignant tumor of supraglottis |
| SNOMED CT | 187843009 | Malignant neoplasm of arytenoid cartilage |
| SNOMED CT | 187844003 | Malignant neoplasm of cricoid cartilage |
| SNOMED CT | 187845002 | Malignant neoplasm of cuneiform cartilage |
| SNOMED CT | 187846001 | Malignant neoplasm of thyroid cartilage |
| SNOMED CT | 187853005 | Malignant neoplasm of cartilage of trachea |
| SNOMED CT | 187854004 | Malignant neoplasm of mucosa of trachea |
| SNOMED CT | 187857006 | Malignant neoplasm of carina of bronchus |
| SNOMED CT | 187861000 | Malignant neoplasm of upper lobe bronchus |
| SNOMED CT | 187862007 | Malignant neoplasm of upper lobe of lung |
| SNOMED CT | 187865009 | Malignant neoplasm of middle lobe bronchus |
| SNOMED CT | 187866005 | Malignant neoplasm of middle lobe of lung |
| SNOMED CT | 187869003 | Malignant neoplasm of lower lobe bronchus |
| SNOMED CT | 187870002 | Malignant neoplasm of lower lobe of lung |
| SNOMED CT | 187900002 | Malignant neoplasm of bones of skull and face |
| SNOMED CT | 187903000 | Malignant neoplasm of malar bone |
| SNOMED CT | 187906008 | Malignant neoplasm of orbital bone |
| SNOMED CT | 187916000 | Malignant neoplasm of cervical vertebra |
| SNOMED CT | 187917009 | Malignant neoplasm of thoracic vertebra |
| SNOMED CT | 187918004 | Malignant neoplasm of lumbar vertebra |
| SNOMED CT | 187920001 | Malignant neoplasm of ribs and/or sternum and/or clavicle |
| SNOMED CT | 187925006 | Malignant neoplasm of costal cartilage |
| SNOMED CT | 187926007 | Malignant neoplasm of costovertebral joint |
| SNOMED CT | 187927003 | Malignant neoplasm of xiphoid process |
| SNOMED CT | 187929000 | Malignant neoplasm of scapula and long bones of upper arm |
| SNOMED CT | 187932002 | Malignant neoplasm of humerus |
| SNOMED CT | 187937008 | Malignant neoplasm of carpal bone - scaphoid |
| SNOMED CT | 187938003 | Malignant neoplasm of carpal bone - lunate |
| SNOMED CT | 187939006 | Malignant neoplasm of carpal bone - triquetrum |
| SNOMED CT | 187940008 | Malignant neoplasm of carpal bone - pisiform |

Continued on next page

**TableS24 – continued from previous page**

| CodeSystem | ConceptCode | ConceptName |
|---|---|---|
| SNOMED CT | 187941007 | Malignant neoplasm of carpal bone - trapezium |
| SNOMED CT | 187942000 | Malignant neoplasm of carpal bone - trapezoid |
| SNOMED CT | 187943005 | Malignant neoplasm of carpal bone - capitate |
| SNOMED CT | 187944004 | Malignant neoplasm of carpal bone - hamate |
| SNOMED CT | 187945003 | Malignant neoplasm of first metacarpal bone |
| SNOMED CT | 187946002 | Malignant neoplasm of second metacarpal bone |
| SNOMED CT | 187947006 | Malignant neoplasm of third metacarpal bone |
| SNOMED CT | 187948001 | Malignant neoplasm of fourth metacarpal bone |
| SNOMED CT | 187949009 | Malignant neoplasm of fifth metacarpal bone |
| SNOMED CT | 187950009 | Malignant neoplasm of phalanges of hand |
| SNOMED CT | 187952001 | Malignant neoplasm of pelvic bones, sacrum and coccyx |
| SNOMED CT | 187956003 | Malignant neoplasm of sacral vertebra |
| SNOMED CT | 187957007 | Malignant neoplasm of coccygeal vertebra |
| SNOMED CT | 187967002 | Malignant neoplasm of calcaneum |
| SNOMED CT | 187968007 | Malignant neoplasm of medial cuneiform |
| SNOMED CT | 187969004 | Malignant neoplasm of intermediate cuneiform |
| SNOMED CT | 187970003 | Malignant neoplasm of lateral cuneiform |
| SNOMED CT | 187972006 | Malignant neoplasm of navicular |
| SNOMED CT | 187973001 | Malignant neoplasm of first metatarsal bone |
| SNOMED CT | 187974007 | Malignant neoplasm of second metatarsal bone |
| SNOMED CT | 187975008 | Malignant neoplasm of third metatarsal bone |
| SNOMED CT | 187976009 | Malignant neoplasm of fourth metatarsal bone |
| SNOMED CT | 187977000 | Malignant neoplasm of fifth metatarsal bone |
| SNOMED CT | 187978005 | Malignant neoplasm of phalanges of foot |
| SNOMED CT | 187988006 | Malignant neoplasm of tarsus of eyelid |
| SNOMED CT | 188010006 | Malignant neoplasm of connective and soft tissue of axilla |
| SNOMED CT | 188013008 | Malignant neoplasm of connective and soft tissues of thoracic spine |
| SNOMED CT | 188049009 | Malignant melanoma of axilla |
| SNOMED CT | 188030005 | Malignant melanoma of lip |
| SNOMED CT | 188033007 | Malignant melanoma of auricle (ear) |
| SNOMED CT | 188071004 | Malignant melanoma of popliteal fossa area |
| SNOMED CT | 188072006 | Malignant melanoma of lower leg |
| SNOMED CT | 188095002 | Malignant neoplasm of skin of cheek, external |
| SNOMED CT | 188107002 | Malignant neoplasm of skin of axillary fold |
| SNOMED CT | 188147009 | Malignant neoplasm of nipple and areola of female breast |
| SNOMED CT | 188151006 | Malignant neoplasm of central part of female breast |
| SNOMED CT | 188152004 | Malignant neoplasm of upper-inner quadrant of female breast |
| SNOMED CT | 188153009 | Malignant neoplasm of lower-inner quadrant of female breast |
| SNOMED CT | 188154003 | Malignant neoplasm of upper-outer quadrant of female breast |
| SNOMED CT | 188155002 | Malignant neoplasm of lower-outer quadrant of female breast |
| SNOMED CT | 188156001 | Malignant neoplasm of axillary tail of female breast |
| SNOMED CT | 188159008 | Malignant neoplasm of ectopic site of female breast |
| SNOMED CT | 188176007 | Malignant neoplasm of endocervical canal |
| SNOMED CT | 188177003 | Malignant neoplasm of endocervical gland |
| SNOMED CT | 188180002 | Primary malignant neoplasm of overlapping sites of cervix uteri |
| SNOMED CT | 188183000 | Malignant neoplasm of cervical stump |

Continued on next page

**TableS24 – continued from previous page**

| CodeSystem | ConceptCode | ConceptName |
|---|---|---|
| SNOMED CT | 188184006 | Malignant neoplasm of squamocolumnar junction of cervix |
| SNOMED CT | 188189001 | Malignant neoplasm of corpus uteri, excluding isthmus |
| SNOMED CT | 188190005 | Malignant neoplasm of cornu of corpus uteri |
| SNOMED CT | 188191009 | Malignant neoplasm of fundus of corpus uteri |
| SNOMED CT | 188192002 | Malignant neoplasm of endometrium of corpus uteri |
| SNOMED CT | 188193007 | Malignant neoplasm of myometrium of corpus uteri |
| SNOMED CT | 188195000 | Malignant neoplasm of isthmus of uterine body |
| SNOMED CT | 188204000 | Malignant neoplasm of round ligament |
| SNOMED CT | 188209005 | Malignant neoplasm of vaginal vault |
| SNOMED CT | 188219004 | Malignant tumor of undescended testis |
| SNOMED CT | 188220005 | Malignant tumor of ectopic testis |
| SNOMED CT | 188230001 | Malignant tumor of body of penis |
| SNOMED CT | 188234005 | Malignant tumor of seminal vesicle |
| SNOMED CT | 188235006 | Malignant tumor of tunica vaginalis |
| SNOMED CT | 188239000 | Malignant tumor of trigone of urinary bladder |
| SNOMED CT | 188240003 | Malignant tumor of vault of bladder |
| SNOMED CT | 188241004 | Malignant neoplasm of lateral wall of urinary bladder |
| SNOMED CT | 188242006 | Malignant neoplasm of anterior wall of urinary bladder |
| SNOMED CT | 188243001 | Malignant neoplasm of posterior wall of urinary bladder |
| SNOMED CT | 188244007 | Malignant tumor of bladder neck |
| SNOMED CT | 188245008 | Malignant tumor of ureteric orifice |
| SNOMED CT | 188247000 | Malignant neoplasm of overlapping sites of bladder |
| SNOMED CT | 188250002 | Malignant tumor of kidney parenchyma |
| SNOMED CT | 188252005 | Malignant tumor of renal calyx |
| SNOMED CT | 188253000 | Malignant tumor of pelviureteric junction |
| SNOMED CT | 188256008 | Malignant neoplasm of overlapping sites of urinary organs |
| SNOMED CT | 188261005 | Malignant neoplasm of eyeball excluding conjunctiva, cornea, retina and choroid |
| SNOMED CT | 188263008 | Malignant tumor of ciliary body |
| SNOMED CT | 188264002 | Malignant tumor of iris |
| SNOMED CT | 188265001 | Malignant neoplasm of crystalline lens |
| SNOMED CT | 188266000 | Malignant neoplasm of sclera |
| SNOMED CT | 188268004 | Malignant neoplasm of connective tissue of orbit |
| SNOMED CT | 188269007 | Malignant neoplasm of extraocular muscle of orbit |
| SNOMED CT | 188272000 | Malignant tumor of lacrimal gland |
| SNOMED CT | 188273005 | Malignant neoplasm of lacrimal sac |
| SNOMED CT | 188274004 | Malignant neoplasm of nasolacrimal duct |
| SNOMED CT | 188280007 | Malignant neoplasm of cerebrum (excluding lobes and ventricles) |
| SNOMED CT | 188281006 | Malignant neoplasm of basal ganglia |
| SNOMED CT | 188282004 | Malignant neoplasm of cerebral cortex |
| SNOMED CT | 188283009 | Malignant neoplasm of corpus striatum |
| SNOMED CT | 188285002 | Malignant neoplasm of globus pallidus |
| SNOMED CT | 188286001 | Malignant tumor of hypothalamus |
| SNOMED CT | 188287005 | Malignant neoplasm of thalamus |
| SNOMED CT | 188289008 | Malignant neoplasm of hippocampus |
| SNOMED CT | 188290004 | Malignant neoplasm of uncus |
| SNOMED CT | 188292007 | Malignant tumor of choroid plexus |

Continued on next page

**TableS24 – continued from previous page**

| CodeSystem | ConceptCode | ConceptName |
|---|---|---|
| SNOMED CT | 188293002 | Malignant neoplasm of floor of cerebral ventricle |
| SNOMED CT | 188295009 | Malignant neoplasm of cerebral peduncle |
| SNOMED CT | 188296005 | Malignant neoplasm of medulla oblongata |
| SNOMED CT | 188297001 | Malignant neoplasm of midbrain |
| SNOMED CT | 188298006 | Malignant neoplasm of pons |
| SNOMED CT | 188301005 | Malignant neoplasm of corpus callosum |
| SNOMED CT | 188302003 | Malignant neoplasm of tapetum |
| SNOMED CT | 188308004 | Malignant neoplasm of olfactory bulb |
| SNOMED CT | 188312005 | Malignant neoplasm of cerebral dura mater |
| SNOMED CT | 188313000 | Malignant neoplasm of cerebral arachnoid mater |
| SNOMED CT | 188315007 | Malignant neoplasm of cerebral pia mater |
| SNOMED CT | 188317004 | Malignant neoplasm of spinal dura mater |
| SNOMED CT | 188318009 | Malignant neoplasm of spinal arachnoid mater |
| SNOMED CT | 188319001 | Malignant neoplasm of spinal pia mater |
| SNOMED CT | 188325002 | Malignant neoplasm of peripheral nerve of thorax |
| SNOMED CT | 188339002 | Malignant neoplasm of pituitary gland and craniopharyngeal duct |
| SNOMED CT | 188340000 | Malignant tumor of craniopharyngeal duct |
| SNOMED CT | 188361007 | Malignant neoplasm of thorax |
| SNOMED CT | 188487008 | Lymphosarcoma and reticulosarcoma |
| SNOMED CT | 188489006 | Reticulosarcoma of lymph nodes of head, face and neck |
| SNOMED CT | 188492005 | Reticulosarcoma of lymph nodes of axilla and upper limb |
| SNOMED CT | 188493000 | Reticulosarcoma of lymph nodes of inguinal region and lower limb |
| SNOMED CT | 188498009 | Lymphosarcoma |
| SNOMED CT | 188500005 | Lymphosarcoma of lymph nodes of head, face and neck |
| SNOMED CT | 188501009 | Lymphosarcoma of intrathoracic lymph nodes |
| SNOMED CT | 188502002 | Lymphosarcoma of intra-abdominal lymph nodes |
| SNOMED CT | 188503007 | Lymphosarcoma of lymph nodes of axilla and upper limb |
| SNOMED CT | 188504001 | Lymphosarcoma of lymph nodes of inguinal region and lower limb |
| SNOMED CT | 188505000 | Lymphosarcoma of intrapelvic lymph nodes |
| SNOMED CT | 188506004 | Lymphosarcoma of spleen |
| SNOMED CT | 188507008 | Lymphosarcoma of lymph nodes of multiple sites |
| SNOMED CT | 188609000 | Nodular lymphoma of lymph nodes of head, face and neck |
| SNOMED CT | 188612002 | Nodular lymphoma of lymph nodes of axilla and upper limb |
| SNOMED CT | 188613007 | Nodular lymphoma of lymph nodes of inguinal region and lower limb |
| SNOMED CT | 188627002 | Mycosis fungoides of lymph nodes of multiple sites |
| SNOMED CT | 188631008 | Sézary's disease of intrathoracic lymph nodes |
| SNOMED CT | 188633006 | Sézary's disease of lymph nodes of axilla AND/OR upper limb |
| SNOMED CT | 188634000 | Sézary's disease of lymph nodes of inguinal region and lower limb |
| SNOMED CT | 188635004 | Sézary's disease of intrapelvic lymph nodes |
| SNOMED CT | 188637007 | Sézary's disease of lymph nodes of multiple sites |
| SNOMED CT | 188641006 | Malignant histiocytosis of lymph nodes of axilla and upper limb |
| SNOMED CT | 188642004 | Malignant histiocytosis of lymph nodes of inguinal region and lower limb |

Continued on next page

**TableS24 – continued from previous page**

| CodeSystem | ConceptCode | ConceptName |
|---|---|---|
| SNOMED CT | 188645002 | Leukemic reticuloendotheliosis of lymph nodes of head, face and neck |
| SNOMED CT | 188648000 | Leukemic reticuloendotheliosis of lymph nodes of axilla and upper limb |
| SNOMED CT | 188649008 | Leukemic reticuloendotheliosis of lymph nodes of inguinal region and lower limb |
| SNOMED CT | 188662007 | Mast cell malignancy of lymph nodes of head, face and neck |
| SNOMED CT | 188665009 | Mast cell malignancy of lymph nodes of axilla and upper limb |
| SNOMED CT | 188666005 | Mast cell malignancy of lymph nodes of inguinal region and lower limb |
| SNOMED CT | 188668006 | Mast cell malignancy of spleen |
| SNOMED CT | 188669003 | Mast cell malignancy of lymph nodes of multiple sites |
| SNOMED CT | 188674006 | Diffuse malignant lymphoma - small non-cleaved cell |
| SNOMED CT | 188675007 | Malignant lymphoma - small cleaved cell |
| SNOMED CT | 188676008 | Malignant lymphoma - mixed small and large cell |
| SNOMED CT | 188718006 | Extramedullary plasmacytoma |
| SNOMED CT | 188725004 | Lymphoid leukemia |
| SNOMED CT | 188726003 | Subacute lymphoid leukemia |
| SNOMED CT | 188728002 | Aleukemic lymphoid leukemia |
| SNOMED CT | 188729005 | Adult T-cell leukemia |
| SNOMED CT | 188732008 | Myeloid leukemia |
| SNOMED CT | 188733003 | Chronic eosinophilic leukemia |
| SNOMED CT | 188734009 | Chronic neutrophilic leukemia |
| SNOMED CT | 188736006 | Subacute myeloid leukemia |
| SNOMED CT | 188737002 | Chloroma |
| SNOMED CT | 188741003 | Aleukemic myeloid leukemia |
| SNOMED CT | 188744006 | Monocytic leukemia |
| SNOMED CT | 188745007 | Chronic monocytic leukemia |
| SNOMED CT | 188746008 | Subacute monocytic leukemia |
| SNOMED CT | 188748009 | Aleukemic monocytic leukemia |
| SNOMED CT | 188754005 | Megakaryocytic leukemia |
| SNOMED CT | 188768003 | Myelomonocytic leukemia |
| SNOMED CT | 188770007 | Subacute myelomonocytic leukemia |
| SNOMED CT | 189164002 | Cerebral meningioma |
| SNOMED CT | 189397006 | Neoplasm of uncertain behavior of sphincter of Oddi |
| SNOMED CT | 189406008 | Neoplasm of uncertain behavior of anal canal and sphincter |
| SNOMED CT | 189509003 | Refractory anemia without sideroblasts, so stated |
| SNOMED CT | 190030009 | Compound leukemias |
| SNOMED CT | 230156002 | Malignant meningitis |
| SNOMED CT | 231829006 | Malignant tumor of eyelid |
| SNOMED CT | 231831002 | Squamous cell carcinoma of eyelid |
| SNOMED CT | 231832009 | Basal cell carcinoma of eyelid |
| SNOMED CT | 231833004 | Meibomian gland carcinoma |
| SNOMED CT | 231834005 | Malignant melanoma of eyelid |
| SNOMED CT | 232075002 | Lymphoma of retina |
| SNOMED CT | 233854003 | Atrial myxoma |
| SNOMED CT | 233855002 | Familial atrial myxoma |

Continued on next page

**TableS24 – continued from previous page**

| CodeSystem | ConceptCode | ConceptName |
|---|---|---|
| SNOMED CT | 235686008 | Fundic gland polyposis of stomach |
| SNOMED CT | 236512004 | Leukemic infiltrate of kidney |
| SNOMED CT | 236513009 | Lymphoma of kidney |
| SNOMED CT | 237719001 | Pituitary adenoma with extrasellar extension |
| SNOMED CT | 237795006 | Virilizing ovarian tumor |
| SNOMED CT | 240163000 | Malignant neoplasm of nasopharyngeal wall |
| SNOMED CT | 241861008 | Metastatic malignant neoplasm to nasopharynx |
| SNOMED CT | 252991009 | Cervical intraepithelial neoplasia grade III with severe dysplasia |
| SNOMED CT | 253003009 | Carcinoid bronchial adenoma |
| SNOMED CT | 254389005 | Carcinoma of vermilion border of upper lip |
| SNOMED CT | 254390001 | Carcinoma of vermilion border of lower lip |
| SNOMED CT | 254393004 | Carcinoma of frenum of lip |
| SNOMED CT | 254398008 | Carcinoma of frenum of upper lip |
| SNOMED CT | 254402004 | Carcinoma of frenum of lower lip |
| SNOMED CT | 254404003 | Carcinoma of commissure of lip |
| SNOMED CT | 254408000 | Malignant tumor of anterior two-thirds of tongue - lateral margin |
| SNOMED CT | 254412006 | Malignant tumor of tip of tongue |
| SNOMED CT | 254417000 | Carcinoma of frenum linguae |
| SNOMED CT | 254423005 | Carcinoma of lingual tonsil |
| SNOMED CT | 254424004 | Carcinoma of upper gum |
| SNOMED CT | 254425003 | Carcinoma of lower gum |
| SNOMED CT | 254427006 | Carcinoma of anterior part of floor of mouth |
| SNOMED CT | 254431000 | Carcinoma of lateral part of floor of mouth |
| SNOMED CT | 254434008 | Carcinoma of hard palate |
| SNOMED CT | 254435009 | Carcinoma of soft palate |
| SNOMED CT | 254437001 | Squamous cell carcinoma of buccal mucosa |
| SNOMED CT | 254441002 | Carcinoma of upper buccal sulcus |
| SNOMED CT | 254445006 | Carcinoma of lower buccal sulcus |
| SNOMED CT | 254450000 | Carcinoma of upper labial sulcus |
| SNOMED CT | 254454009 | Carcinoma of lower labial sulcus |
| SNOMED CT | 254457002 | Carcinoma of retromolar area |
| SNOMED CT | 254459004 | Malignant tumor of anterior pillar of fauces |
| SNOMED CT | 254462001 | Carcinoma of parotid gland |
| SNOMED CT | 254465004 | Carcinoma of submandibular gland |
| SNOMED CT | 254466003 | Carcinoma of sublingual gland |
| SNOMED CT | 254478004 | Malignant tumor of inferior turbinate |
| SNOMED CT | 254481009 | Malignant tumor of middle turbinate |
| SNOMED CT | 254484001 | Malignant tumor of posterior margin of nasal septum and choanae |
| SNOMED CT | 254501009 | Tumor of inferior surface of soft palate |
| SNOMED CT | 254503007 | Malignant tumor of inferior surface of soft palate |
| SNOMED CT | 254509006 | Malignant tumor of anterior commissure |
| SNOMED CT | 254513004 | Malignant tumor of posterior commissure |
| SNOMED CT | 254517003 | Malignant tumor of suprahyoid epiglottis |
| SNOMED CT | 254520006 | Malignant tumor of infrahyoid epiglottis |
| SNOMED CT | 254526000 | Malignant tumor of laryngeal ventricle |
| SNOMED CT | 254530002 | Malignant tumor of parapharyngeal space |

Continued on next page

**TableS24 – continued from previous page**

| CodeSystem | ConceptCode | ConceptName |
|---|---|---|
| SNOMED CT | 254535007 | Carcinoma of cervical part of esophagus |
| SNOMED CT | 254539001 | Carcinoma of thoracic part of esophagus |
| SNOMED CT | 254543002 | Carcinoma of abdominal part of esophagus |
| SNOMED CT | 254547001 | Carcinoma of upper third of esophagus |
| SNOMED CT | 254549003 | Carcinoma of middle third of esophagus |
| SNOMED CT | 254551004 | Carcinoma of lower third of esophagus |
| SNOMED CT | 254553001 | Carcinoma of cardia |
| SNOMED CT | 254555008 | Carcinoma of fundus of stomach |
| SNOMED CT | 254557000 | Carcinoma of body of stomach |
| SNOMED CT | 254559002 | Carcinoma of pyloric antrum |
| SNOMED CT | 254561006 | Carcinoma of pylorus |
| SNOMED CT | 254563009 | Carcinoma of lesser curve of stomach |
| SNOMED CT | 254567005 | Carcinoma of greater curve of stomach |
| SNOMED CT | 254570009 | Carcinoma of duodenum |
| SNOMED CT | 254574000 | Diffuse leiomyomatosis of esophagus |
| SNOMED CT | 254586002 | Malignant tumor of anorectal junction |
| SNOMED CT | 254609000 | Carcinoma of ampulla of Vater |
| SNOMED CT | 254611009 | Malignant tumor of endocrine pancreas |
| SNOMED CT | 254612002 | Carcinoma of endocrine pancreas |
| SNOMED CT | 254613007 | Carcinoid tumor of pancreas |
| SNOMED CT | 254615000 | Endocrine pancreatic adenoma |
| SNOMED CT | 254619006 | Adenoid cystic carcinoma of trachea |
| SNOMED CT | 254620000 | Squamous cell carcinoma of trachea |
| SNOMED CT | 254621001 | Laryngotracheal papillomatosis |
| SNOMED CT | 254622008 | Squamous cell carcinoma of bronchus |
| SNOMED CT | 254625005 | Malignant tumor of lung parenchyma |
| SNOMED CT | 254626006 | Adenocarcinoma of lung |
| SNOMED CT | 254627002 | Carcinoid tumor of lung |
| SNOMED CT | 254629004 | Large cell carcinoma of lung |
| SNOMED CT | 254631008 | Giant cell carcinoma of lung |
| SNOMED CT | 254632001 | Small cell carcinoma of lung |
| SNOMED CT | 254633006 | Oat cell carcinoma of lung |
| SNOMED CT | 254634000 | Squamous cell carcinoma of lung |
| SNOMED CT | 254635004 | Epithelioid hemangioendothelioma of lung |
| SNOMED CT | 254637007 | Non-small cell lung cancer |
| SNOMED CT | 254638002 | Pancoast tumor |
| SNOMED CT | 254641006 | Histiocytoma of lung |
| SNOMED CT | 254643009 | Intrapulmonary teratoma |
| SNOMED CT | 254644003 | Hamartoma of lung |
| SNOMED CT | 254645002 | Malignant mesothelioma of pleura |
| SNOMED CT | 254730000 | Superficial spreading malignant melanoma of skin |
| SNOMED CT | 254731001 | Nodular malignant melanoma of skin |
| SNOMED CT | 254732008 | Acral lentiginous malignant melanoma of skin |
| SNOMED CT | 254733003 | Malignant melanoma arising in intradermal nevus |
| SNOMED CT | 254734009 | Malignant melanoma arising in congenital nevus |
| SNOMED CT | 254792006 | Proliferating angioendotheliomatosis |
| SNOMED CT | 254849005 | Malignant epithelial tumor of ovary |

Continued on next page

**TableS24 – continued from previous page**

| CodeSystem | ConceptCode | ConceptName |
|---|---|---|
| SNOMED CT | 254850005 | Serous papillary cystadenocarcinoma ovary |
| SNOMED CT | 254851009 | Mucinous cystadenocarcinoma of ovary |
| SNOMED CT | 254852002 | Endometrioid carcinoma ovary |
| SNOMED CT | 254856004 | Undifferentiated carcinoma of ovary |
| SNOMED CT | 254860001 | Malignant sex cord tumor of ovary |
| SNOMED CT | 254863004 | Granulosa cell tumor of ovary |
| SNOMED CT | 254864005 | Theca cell tumor of ovary |
| SNOMED CT | 254866007 | Sertoli-Leydig cell tumor of ovary |
| SNOMED CT | 254867003 | Gynandroblastoma of ovary |
| SNOMED CT | 254868008 | Hilus cell tumor of ovary |
| SNOMED CT | 254869000 | Malignant germ cell tumor of ovary |
| SNOMED CT | 254870004 | Choriocarcinoma of ovary |
| SNOMED CT | 254872007 | Embryonal carcinoma of ovary |
| SNOMED CT | 254874008 | Dysgerminoma of ovary |
| SNOMED CT | 254876005 | Endodermal sinus tumor of ovary |
| SNOMED CT | 254877001 | Sarcoma of uterus |
| SNOMED CT | 254878006 | Endometrial carcinoma |
| SNOMED CT | 254883003 | Intravenous leiomyomatosis |
| SNOMED CT | 254886006 | Squamous cell carcinoma of cervix |
| SNOMED CT | 254887002 | Adenocarcinoma of cervix |
| SNOMED CT | 254888007 | Adenosquamous carcinoma of cervix |
| SNOMED CT | 254889004 | Carcinoma of cervix stage 0 |
| SNOMED CT | 254893005 | Carcinoma of vagina |
| SNOMED CT | 254894004 | Vaginal intraepithelial neoplasia |
| SNOMED CT | 254900004 | Carcinoma of prostate |
| SNOMED CT | 254901000 | Prostatic intraepithelial neoplasia |
| SNOMED CT | 254904008 | Carcinoma of glans penis |
| SNOMED CT | 254908006 | Malignant tumor of skin of penis |
| SNOMED CT | 254909003 | Carcinoma of foreskin |
| SNOMED CT | 254912000 | Regressed malignant testicular tumor |
| SNOMED CT | 254915003 | Clear cell carcinoma of kidney |
| SNOMED CT | 254916002 | Cystadenocarcinoma of kidney |
| SNOMED CT | 254917006 | Papillary cystadenocarcinoma of kidney |
| SNOMED CT | 254918001 | Sarcoma of kidney |
| SNOMED CT | 254919009 | Cortical adenoma of kidney |
| SNOMED CT | 254920003 | Cystadenoma of kidney |
| SNOMED CT | 254921004 | Angiomyolipoma of kidney |
| SNOMED CT | 254922006 | Oncocytoma of kidney |
| SNOMED CT | 254923001 | Hemangiopericytoma of kidney |
| SNOMED CT | 254934003 | Malignant tumor of urethral stump |
| SNOMED CT | 254936001 | Glial tumor of brain |
| SNOMED CT | 254938000 | Astrocytoma of brain |
| SNOMED CT | 254940005 | Oligodendroglioma of brain |
| SNOMED CT | 254941009 | Mixed glial tumor of brain |
| SNOMED CT | 254945000 | Embryonal tumor of brain |
| SNOMED CT | 254946004 | Glial tumor of spinal cord |
| SNOMED CT | 254947008 | Glioma of spinal cord |

Continued on next page

**TableS24 – continued from previous page**

| CodeSystem | ConceptCode | ConceptName |
|---|---|---|
| SNOMED CT | 254948003 | Astrocytoma of spinal cord |
| SNOMED CT | 254950006 | Oligodendroglioma of spinal cord |
| SNOMED CT | 254951005 | Mixed glial tumor of spinal cord |
| SNOMED CT | 254954002 | Embryonal tumor of spinal cord |
| SNOMED CT | 254955001 | Pituitary carcinoma |
| SNOMED CT | 254956000 | Pituitary adenoma |
| SNOMED CT | 254957009 | Somatotroph adenoma |
| SNOMED CT | 254958004 | Corticotroph adenoma |
| SNOMED CT | 254959007 | Thyrotroph adenoma |
| SNOMED CT | 254960002 | Gonadotroph adenoma |
| SNOMED CT | 254961003 | Mixed-functioning pituitary adenoma |
| SNOMED CT | 254962005 | Functionless pituitary adenoma |
| SNOMED CT | 254963000 | Pituitary microadenoma |
| SNOMED CT | 254964006 | Pituitary mesoadenoma |
| SNOMED CT | 254965007 | Pituitary macroadenoma |
| SNOMED CT | 254966008 | Suprasellar extension of pituitary adenoma |
| SNOMED CT | 254968009 | Tumor of hypothalamus |
| SNOMED CT | 254969001 | Malignant tumor of olfactory tract |
| SNOMED CT | 254972008 | Malignant tumor of optic nerve and sheath |
| SNOMED CT | 254973003 | Malignant astrocytoma of optic nerve |
| SNOMED CT | 254974009 | Malignant tumor of optic nerve sheath |
| SNOMED CT | 254975005 | Malignant meningioma of optic nerve sheath |
| SNOMED CT | 254976006 | Optic nerve glioma |
| SNOMED CT | 254978007 | Meningioma of optic nerve sheath |
| SNOMED CT | 254979004 | Melanocytoma of optic nerve head |
| SNOMED CT | 254987003 | Adenoid cystic carcinoma of lacrimal gland |
| SNOMED CT | 254988008 | Adenocarcinoma of lacrimal gland |
| SNOMED CT | 254989000 | Carcinoma ex pleomorphic adenoma of lacrimal gland |
| SNOMED CT | 254990009 | Mucoepidermoid tumor of lacrimal gland |
| SNOMED CT | 254993006 | Liposarcoma of orbit |
| SNOMED CT | 254994000 | Rhabdomyosarcoma of orbit |
| SNOMED CT | 254995004 | Malignant hemangiopericytoma of orbit |
| SNOMED CT | 254996003 | Malignant fibrous histiocytoma of orbit |
| SNOMED CT | 254997007 | Capillary hemangioma of orbit |
| SNOMED CT | 254998002 | Cavernous hemangioma of orbit |
| SNOMED CT | 254999005 | Hemangiopericytoma of orbit |
| SNOMED CT | 255000005 | Neurofibroma of orbit |
| SNOMED CT | 255003007 | Squamous cell carcinoma of conjunctiva |
| SNOMED CT | 255004001 | Malignant melanoma of conjunctiva |
| SNOMED CT | 255008003 | Squamous cell carcinoma of cornea |
| SNOMED CT | 255012009 | Malignant melanoma of iris |
| SNOMED CT | 255015006 | Malignant melanoma of ciliary body |
| SNOMED CT | 255016007 | Adenocarcinoma of pigmented epithelium of ciliary body |
| SNOMED CT | 255017003 | Adenocarcinoma of non-pigmented epithelium of ciliary body |
| SNOMED CT | 255021005 | Malignant melanoma of choroid |
| SNOMED CT | 255028004 | Follicular thyroid carcinoma |
| SNOMED CT | 255029007 | Papillary thyroid carcinoma |

Continued on next page

**TableS24 – continued from previous page**

| CodeSystem | ConceptCode | ConceptName |
|---|---|---|
| SNOMED CT | 255030002 | Mixed follicular and papillary thyroid carcinoma |
| SNOMED CT | 255031003 | Anaplastic thyroid carcinoma |
| SNOMED CT | 255032005 | Medullary thyroid carcinoma |
| SNOMED CT | 255034006 | Thyroid follicular adenoma |
| SNOMED CT | 255035007 | Adrenal carcinoma |
| SNOMED CT | 255037004 | Parathyroid carcinoma |
| SNOMED CT | 255067005 | Sarcoma of bone and connective tissue |
| SNOMED CT | 255070009 | Overlapping malignant neoplasm of oral cavity and lip and salivary gland |
| SNOMED CT | 255071008 | Squamous cell carcinoma of lip |
| SNOMED CT | 255072001 | Malignant tumor of salivary gland |
| SNOMED CT | 255074000 | Malignant tumor of nasal cavity and nasopharynx |
| SNOMED CT | 255075004 | Malignant tumor of lateral nasal wall |
| SNOMED CT | 255078002 | Malignant tumor of esophagus, stomach and duodenum |
| SNOMED CT | 255084004 | Squamous cell carcinoma of anal margin |
| SNOMED CT | 255090000 | Malignant neoplasm of carpal bones |
| SNOMED CT | 255091001 | Malignant neoplasm of metacarpal bones |
| SNOMED CT | 255108000 | Carcinoma of bladder |
| SNOMED CT | 255109008 | Transitional cell carcinoma of bladder |
| SNOMED CT | 255110003 | Adenocarcinoma of bladder |
| SNOMED CT | 255111004 | Squamous cell carcinoma of bladder |
| SNOMED CT | 255112006 | Malignant tumor of pituitary and hypothalamus |
| SNOMED CT | 255119002 | Lymphangitis carcinomatosa |
| SNOMED CT | 255123005 | Metastasis to nervous system and eye |
| SNOMED CT | 255191003 | Localized malignant reticulohistiocytoma |
| SNOMED CT | 223003 | Tumor of body of uterus affecting pregnancy |
| SNOMED CT | 308006 | Pearly penile papules |
| SNOMED CT | 4135001 | 11p partial monosomy syndrome |
| SNOMED CT | 6331000 | Angiokeratoma of Fordyce |
| SNOMED CT | 7021009 | Gastrointestinal eosinophilic granuloma |
| SNOMED CT | 8090002 | Eosinophilic granuloma of oral mucosa |
| SNOMED CT | 13048006 | Orbital lymphoma |
| SNOMED CT | 22288000 | Tumor of cervix affecting pregnancy |
| SNOMED CT | 28122003 | Pulmonary eosinophilic granuloma |
| SNOMED CT | 30664006 | Multiple endocrine neoplasia, type 1 |
| SNOMED CT | 31047003 | Lymphomatoid papulosis |
| SNOMED CT | 31383003 | Tumor of vagina affecting pregnancy |
| SNOMED CT | 46724008 | Polyglandular activity in multiple endocrine adenomatosis |
| SNOMED CT | 58961005 | Lethal midline granuloma |
| SNOMED CT | 61530001 | Multiple endocrine neoplasia, type 3 |
| SNOMED CT | 61808009 | Multiple endocrine neoplasia, type 2 |
| SNOMED CT | 64862009 | Congenital rhabdomyoma of heart |
| SNOMED CT | 67944007 | Lhermitte-Duclos disease |
| SNOMED CT | 722795004 | Meningeal leukemia |
| SNOMED CT | 722953004 | B-cell lymphoma unclassifiable with features intermediate between Burkitt lymphoma and diffuse large B-cell lymphoma |

Continued on next page

**TableS24 – continued from previous page**

| CodeSystem | ConceptCode | ConceptName |
|---|---|---|
| SNOMED CT | 722954005 | B-cell lymphoma unclassifiable with features intermediate between classical Hodgkin lymphoma and diffuse large B-cell lymphoma |
| SNOMED CT | 723889003 | B lymphoblastic leukemia lymphoma with t(9:22) (q34;q11.2); BCR-ABL 1 |
| SNOMED CT | 724644005 | Myeloid leukemia co-occurrent with Down syndrome |
| SNOMED CT | 724645006 | T-cell histiocyte rich large B-cell lymphoma |
| SNOMED CT | 724647003 | Diffuse large B-cell lymphoma co-occurrent with chronic inflammation caused by Epstein-Barr virus |
| SNOMED CT | 724648008 | Plasmablastic lymphoma |
| SNOMED CT | 724649000 | Langerhans cell sarcoma |
| SNOMED CT | 724650000 | Primary follicular dendritic cell sarcoma |
| SNOMED CT | 725390002 | Acute myeloid leukemia with t(8;16)(p11;p13) translocation |
| SNOMED CT | 725437002 | Chronic lymphocytic leukemia genetic mutation variant |
| SNOMED CT | 726721002 | Nodal marginal zone B-cell lymphoma |
| SNOMED CT | 733598001 | Acute myeloid leukemia with t(6;9)(p23;q34) translocation |
| SNOMED CT | 734066005 | Diffuse large B-cell lymphoma of central nervous system |
| SNOMED CT | 734522002 | Acute myeloid leukemia with FMS-like tyrosine kinase-3 mutation |
| SNOMED CT | 735332000 | Primary cutaneous diffuse large cell B-cell lymphoma leg type |
| SNOMED CT | 736322001 | Pediatric follicular lymphoma |
| SNOMED CT | 737058005 | Microsatellite instability-high colorectal cancer |
| SNOMED CT | 738527001 | Myeloid and/or lymphoid neoplasm associated with platelet derived growth factor receptor alpha rearrangement |
| SNOMED CT | 762315004 | Therapy related acute myeloid leukemia due to and following administration of antineoplastic agent |
| SNOMED CT | 762690000 | Classical Hodgkin lymphoma |
| SNOMED CT | 763309005 | Acute myeloid leukemia with NPM1 somatic mutation |
| SNOMED CT | 763477007 | Primary lymphoma of conjunctiva |
| SNOMED CT | 763666008 | Splenic marginal zone B-cell lymphoma |
| SNOMED CT | 763719001 | Hydroa vacciniforme-like lymphoma |
| SNOMED CT | 763796007 | Megakaryoblastic acute myeloid leukemia with t(1;22)(p13;q13) |
| SNOMED CT | 763884007 | Splenic diffuse red pulp small B-cell lymphoma |
| SNOMED CT | 764855007 | Acute myeloid leukemia with CEBPA somatic mutations |
| SNOMED CT | 764940002 | Inherited acute myeloid leukemia |
| SNOMED CT | 765136002 | Primary cutaneous CD8 positive aggressive epidermotropic cytotoxic T-cell lymphoma |
| SNOMED CT | 765328000 | Classic mycosis fungoides |
| SNOMED CT | 766045006 | Acute myeloid leukemia and myelodysplastic syndrome related to alkylating agent |
| SNOMED CT | 766046007 | Acute myeloid leukemia and myelodysplastic syndrome related to topoisomerase type 2 inhibitor |
| SNOMED CT | 766048008 | Acute myeloid leukemia and myelodysplastic syndrome related to radiation |
| SNOMED CT | 766935007 | Primary bone lymphoma |
| SNOMED CT | 766981007 | Squamous cell carcinoma of colon |
| SNOMED CT | 770402000 | Aleukemic mast cell leukemia |

Continued on next page

**TableS24 – continued from previous page**

| CodeSystem | ConceptCode | ConceptName |
|---|---|---|
| SNOMED CT | 773537001 | Differentiation syndrome due to and following chemotherapy co-occurrent with acute promyelocytic leukemia |
| SNOMED CT | 773995001 | Primary cutaneous anaplastic large cell lymphoma |
| SNOMED CT | 780844005 | Acute myeloid leukemia with inv(3)(q21q26.2) or t(3;3)(q21;q26.2); RPN1-EVI1 |
| SNOMED CT | 783541009 | Breast implant-associated anaplastic large-cell lymphoma |
| SNOMED CT | 783565007 | Indolent T-cell lymphoproliferative disorder of gastrointestinal tract |
| SNOMED CT | 783615009 | Erythropoietic uroporphyria associated with myeloid malignancy |
| SNOMED CT | 788874003 | B-cell prolymphocytic leukemia in remission |
| SNOMED CT | 788972003 | Juvenile myelomonocytic leukemia in remission |
| SNOMED CT | 830057003 | Relapsing classical Hodgkin lymphoma |
| SNOMED CT | 836486002 | Lymphomatous infiltrate of kidney |
| SNOMED CT | 838340006 | B lymphoblastic leukemia lymphoma with t(5;14)(q31;q32); IL3-IGH |
| SNOMED CT | 838341005 | B lymphoblastic leukemia lymphoma with t(v;11q23); MLL rearranged |
| SNOMED CT | 838342003 | B lymphoblastic leukemia lymphoma with t(12;21) (p13;q22); TEL/AML1 (ETV6-RUNX1) |
| SNOMED CT | 838343008 | B lymphoblastic leukemia lymphoma with t(1;19)(Q23;P13.3); E2A-PBX1 (TCF3/PBX1) |
| SNOMED CT | 838344002 | B lymphoblastic leukemia lymphoma with hypodiploidy |
| SNOMED CT | 838346000 | B lymphoblastic leukemia lymphoma with hyperdiploidy |
| SNOMED CT | 838355002 | Acute myeloid leukemia with inv(16)(p13.1q22) or t(16;16)(p13.1;q22) CBFB-MYH11 |
| SNOMED CT | 840423002 | Diffuse large B-cell lymphoma of small intestine |
| SNOMED CT | 840424008 | Diffuse large B-cell lymphoma of stomach |
| SNOMED CT | 866098005 | Large B-cell lymphoma arising in HHV8-associated multicentric Castleman disease |
| SNOMED CT | 1701000119104 | Primary adenocarcinoma of colon |
| SNOMED CT | 12281000132104 | Relapsing acute myeloid leukemia |
| SNOMED CT | 12291000132102 | Refractory acute myeloid leukemia |
| SNOMED CT | 12301000132103 | Acute lymphoid leukemia relapse |
| SNOMED CT | 12311000132101 | Refractory acute lymphoid leukemia |
| SNOMED CT | 61291000119103 | Disorder of central nervous system co-occurrent and due to acute lymphoid leukemia in remission |
| SNOMED CT | 61301000119102 | Disorder of central nervous system co-occurrent and due to acute lymphoid leukemia |
| SNOMED CT | 61311000119104 | Acute myeloid leukemia, minimal differentiation, FAB M0 in remission |
| SNOMED CT | 61321000119106 | Acute myeloid leukemia without maturation, FAB M1 in remission |
| SNOMED CT | 68261000119102 | Diffuse non-Hodgkin's lymphoma Lugano stage I |
| SNOMED CT | 68271000119108 | Diffuse non-Hodgkin's lymphoma Lugano stage II |
| SNOMED CT | 68291000119109 | Diffuse non-Hodgkin's lymphoma Lugano stage IV |
| SNOMED CT | 84811000119107 | Follicular dendritic sarcoma of intraabdominal lymph node |
| SNOMED CT | 84831000119102 | Follicular dendritic sarcoma of lymph nodes of multiple sites |

Continued on next page

**TableS24 – continued from previous page**

| CodeSystem | ConceptCode | ConceptName |
|---|---|---|
| SNOMED CT | 96281000119107 | Overlapping malignant neoplasm of colon and rectum |
| SNOMED CT | 96981000119102 | Malignant neoplasm of rectosigmoid junction metastatic to brain |
| SNOMED CT | 116691000119101 | Marginal zone lymphoma of spleen |
| SNOMED CT | 116711000119103 | Marginal zone lymphoma of inguinal lymph node |
| SNOMED CT | 116741000119104 | Marginal zone lymphoma of thoracic lymph node |
| SNOMED CT | 116811000119106 | Non-Hodgkin lymphoma of central nervous system metastatic to lymph node of lower limb |
| SNOMED CT | 116821000119104 | Non-Hodgkin lymphoma of central nervous system metastatic to lymph node of upper limb |
| SNOMED CT | 116841000119105 | Marginal zone lymphoma of lymph nodes of multiple sites |
| SNOMED CT | 116871000119103 | Mantle cell lymphoma of lymph nodes of multiple sites |
| SNOMED CT | 117061000119101 | Peripheral T-cell lymphoma of spleen |
| SNOMED CT | 117091000119108 | Peripheral T-cell lymphoma of axillary lymph node |
| SNOMED CT | 117111000119100 | Peripheral T-cell lymphoma of thoracic lymph node |
| SNOMED CT | 117211000119105 | Peripheral T-cell lymphoma of lymph nodes of multiple sites |
| SNOMED CT | 122881000119107 | Chronic monocytic leukemia in relapse |
| SNOMED CT | 122901000119109 | Myeloid leukemia in relapse |
| SNOMED CT | 122951000119108 | Lymphoid leukemia in relapse |
| SNOMED CT | 122961000119105 | Chronic lymphoid leukemia in relapse |
| SNOMED CT | 122981000119101 | Plasma cell leukemia in relapse |
| SNOMED CT | 123781000119107 | Chronic leukemia in relapse |
| SNOMED CT | 127961000119108 | Aleukemic myeloid leukemia in relapse |
| SNOMED CT | 130381000119103 | Primary malignant neuroendocrine neoplasm of ascending colon |
| SNOMED CT | 133751000119102 | Lymphoma of colon |
| SNOMED CT | 184881000119106 | Primary adenocarcinoma of rectosigmoid junction |
| SNOMED CT | 350951000119101 | B-cell lymphoma of intra-abdominal lymph nodes |
| SNOMED CT | 351211000119104 | B-cell lymphoma of lymph nodes of multiple sites |
| SNOMED CT | 352251000119109 | Small lymphocytic B-cell lymphoma of lymph nodes of multiple sites |
| SNOMED CT | 352411000119109 | Small lymphocytic B-cell lymphoma of intra-abdominal lymph nodes |
| SNOMED CT | 352791000119108 | Non-Hodgkin's lymphoma of lymph nodes of multiple sites |
| SNOMED CT | 354851000119101 | Follicular non-Hodgkin's lymphoma of lymph nodes of multiple sites |
| SNOMED CT | 450521000124109 | Marginal zone lymphoma of axillary lymph node |
| SNOMED CT | 456911000124108 | Diffuse non-Hodgkin's lymphoma Lugano stage III |
| SNOMED CT | 461311000124107 | Refractory Hodgkin's lymphoma |
| SNOMED CT | 681601000119101 | Primary adenocarcinoma of ascending colon |
| SNOMED CT | 53132006 | Excessive gastrin secretion |
| SNOMED CT | 92516002 | Burkitt's tumor of unspecified, extranodal, or solid organ site |
| SNOMED CT | 93451002 | Erythroleukaemia, FAB M6 |
| SNOMED CT | 93487009 | Hodgkin disease, lymphocytic depletion of lymph nodes of axilla AND/OR upper limb |
| SNOMED CT | 93488004 | Hodgkin disease, lymphocytic depletion of lymph nodes of head, face AND/OR neck |
| SNOMED CT | 93489007 | Hodgkin disease, lymphocytic depletion of lymph nodes of inguinal region AND/OR lower limb |

Continued on next page

**TableS24 – continued from previous page**

| CodeSystem | ConceptCode | ConceptName |
|---|---|---|
| SNOMED CT | 93492006 | Hodgkin disease, lymphocytic depletion of extranodal AND/OR solid organ site |
| SNOMED CT | 93493001 | Hodgkin's paragranuloma of intra-abdominal lymph nodes |
| SNOMED CT | 93494007 | Hodgkin disease, lymphocytic-histiocytic predominance of intrapelvic lymph nodes |
| SNOMED CT | 93495008 | Hodgkin disease, lymphocytic-histiocytic predominance of intrathoracic lymph nodes |
| SNOMED CT | 93496009 | Hodgkin disease, lymphocytic-histiocytic predominance of lymph nodes of axilla AND/OR upper limb |
| SNOMED CT | 93497000 | Hodgkin disease, lymphocytic-histiocytic predominance of lymph nodes of head, face AND/OR neck |
| SNOMED CT | 93498005 | Hodgkin disease, lymphocytic-histiocytic predominance of lymph nodes of inguinal region AND/OR lower limb |
| SNOMED CT | 93500006 | Hodgkin paragranuloma of spleen |
| SNOMED CT | 93501005 | Hodgkin disease, lymphocytic-histiocytic predominance of extranodal AND/OR solid organ site |
| SNOMED CT | 93505001 | Hodgkin disease, mixed cellularity of lymph nodes of axilla AND/OR upper limb |
| SNOMED CT | 93506000 | Hodgkin disease, mixed cellularity of lymph nodes of head, face AND/OR neck |
| SNOMED CT | 93507009 | Hodgkin disease, mixed cellularity of lymph nodes of inguinal region AND/OR lower limb |
| SNOMED CT | 93509007 | Hodgkin disease, mixed cellularity of spleen |
| SNOMED CT | 93510002 | Hodgkin disease, mixed cellularity of extranodal AND/OR solid organ site |
| SNOMED CT | 93514006 | Hodgkin disease, nodular sclerosis of lymph nodes of axilla AND/OR upper limb |
| SNOMED CT | 93515007 | Hodgkin disease, nodular sclerosis of lymph nodes of head, face AND/OR neck |
| SNOMED CT | 93516008 | Hodgkin disease, nodular sclerosis of lymph nodes of inguinal region AND/OR lower limb |
| SNOMED CT | 93518009 | Hodgkin disease, nodular sclerosis of spleen |
| SNOMED CT | 93519001 | Hodgkin disease, nodular sclerosis of extranodal AND/OR solid organ site |
| SNOMED CT | 93520007 | Hodgkin disease of intra-abdominal lymph nodes |
| SNOMED CT | 93521006 | Hodgkin disease of intrapelvic lymph nodes |
| SNOMED CT | 93522004 | Hodgkin disease of intrathoracic lymph nodes |
| SNOMED CT | 93523009 | Hodgkin disease of lymph nodes of axilla AND/OR upper limb |
| SNOMED CT | 93524003 | Hodgkin disease of lymph nodes of head, face AND/OR neck |
| SNOMED CT | 93525002 | Hodgkin disease of lymph nodes of inguinal region AND/OR lower limb |
| SNOMED CT | 93526001 | Hodgkin disease of lymph nodes of multiple sites |
| SNOMED CT | 93527005 | Hodgkin disease of spleen |
| SNOMED CT | 93528000 | Hodgkin disease of extranodal AND/OR solid organ site |
| SNOMED CT | 93530003 | Hodgkin granuloma of intrapelvic lymph nodes |
| SNOMED CT | 93531004 | Hodgkin granuloma of intrathoracic lymph nodes |

Continued on next page

**TableS24 – continued from previous page**

| CodeSystem | ConceptCode | ConceptName |
|---|---|---|
| SNOMED CT | 93532006 | Hodgkin granuloma of lymph nodes of axilla AND/OR upper limb |
| SNOMED CT | 93533001 | Hodgkin granuloma of lymph nodes of head, face AND/OR neck |
| SNOMED CT | 93534007 | Hodgkin granuloma of lymph nodes of inguinal region AND/OR lower limb |
| SNOMED CT | 93536009 | Hodgkin granuloma of spleen |
| SNOMED CT | 93537000 | Hodgkin granuloma of extranodal AND/OR solid organ site |
| SNOMED CT | 93541001 | Hodgkin paragranuloma of lymph nodes of axilla AND/OR upper limb |
| SNOMED CT | 93542008 | Hodgkin paragranuloma of lymph nodes of head, face AND/OR neck |
| SNOMED CT | 93543003 | Hodgkin paragranuloma of lymph nodes of inguinal region AND/OR lower limb |
| SNOMED CT | 93546006 | Hodgkin paragranuloma of extranodal AND/OR solid organ site |
| SNOMED CT | 93547002 | Hodgkin sarcoma of intra-abdominal lymph nodes |
| SNOMED CT | 93548007 | Hodgkin sarcoma of intrapelvic lymph nodes |
| SNOMED CT | 93549004 | Hodgkin sarcoma of intrathoracic lymph nodes |
| SNOMED CT | 93550004 | Hodgkin sarcoma of lymph nodes of axilla AND/OR upper limb |
| SNOMED CT | 93551000 | Hodgkin sarcoma of lymph nodes of head, face AND/OR neck |
| SNOMED CT | 93552007 | Hodgkin sarcoma of lymph nodes of inguinal region AND/OR lower limb |
| SNOMED CT | 93554008 | Hodgkin sarcoma of spleen |
| SNOMED CT | 93555009 | Hodgkin sarcoma of extranodal AND/OR solid organ site |
| SNOMED CT | 94144008 | Primary malignant neoplasm of Waldeyer ring |
| SNOMED CT | 94719007 | Granulocytic sarcoma |
| SNOMED CT | 109350007 | Lipoma of trunk |
| SNOMED CT | 109388009 | Kaposi sarcoma of palate |
| SNOMED CT | 109390005 | Kaposi sarcoma of lung |
| SNOMED CT | 109962001 | Nonfollicular lymphoma |
| SNOMED CT | 109964000 | Diffuse non-Hodgkin lymphoma, undifferentiated |
| SNOMED CT | 109965004 | Lymphoblastoma |
| SNOMED CT | 109966003 | Malignant lymphoma - immunoblastic |
| SNOMED CT | 109967007 | Diffuse non-Hodgkin lymphoma, small cleaved cell |
| SNOMED CT | 109968002 | Diffuse non-Hodgkin lymphoma, small cell |
| SNOMED CT | 109969005 | DLBCL - diffuse large B cell lymphoma |
| SNOMED CT | 109970006 | Follicular lymphoma, grade 1 |
| SNOMED CT | 109971005 | Follicular lymphoma grade 2 |
| SNOMED CT | 109972003 | Follicular lymphoma grade 3 |
| SNOMED CT | 109977009 | Peripheral T-cell lymphoma pleomorphic small cell |
| SNOMED CT | 109979007 | B-cell lymphoma |
| SNOMED CT | 110004001 | Acute promyelocytic leukemia (clinical) |
| SNOMED CT | 110005000 | Acute myelomonocytic leukemia (clinical) |
| SNOMED CT | 110007008 | Adult T-cell leukemia |
| SNOMED CT | 118599009 | Lymphoma, Hodgkins |
| SNOMED CT | 118600007 | Malignant lymphoma (clinical) |
| SNOMED CT | 118601006 | Non-Hodgkin lymphoma |
| SNOMED CT | 118602004 | Hodgkin granuloma |

Continued on next page

**TableS24 – continued from previous page**

| CodeSystem | ConceptCode | ConceptName |
|---|---|---|
| SNOMED CT | 118605002 | Hodgkin lymphoma, nodular lymphocyte predominance |
| SNOMED CT | 118606001 | Hodgkin sarcoma |
| SNOMED CT | 118608000 | Hodgkin disease, nodular sclerosis |
| SNOMED CT | 118609008 | Hodgkin mixed cellularity lymphoma |
| SNOMED CT | 118610003 | Hodgkin disease, lymphocytic depletion |
| SNOMED CT | 118613001 | Leukemic reticuloendotheliosis |
| SNOMED CT | 118614007 | Acute infancy reticulosis |
| SNOMED CT | 118617000 | Burkitt lymphoma |
| SNOMED CT | 128875000 | Primary cutaneous CD30 antigen positive large T-cell lymphoma |
| SNOMED CT | 187716008 | Malignant tumour of Waldeyer ring |
| SNOMED CT | 187734007 | Malignant neoplasm of gastroesophageal junction of stomach |
| SNOMED CT | 188511002 | Burkitt tumor of intrathoracic lymph nodes |
| SNOMED CT | 188512009 | Burkitt tumor of intra-abdominal lymph nodes |
| SNOMED CT | 188513004 | Burkitt lymphoma of lymph nodes of axilla and upper limb |
| SNOMED CT | 188514005 | Burkitt lymphoma of lymph nodes of inguinal region and lower limb |
| SNOMED CT | 188515006 | Burkitt tumor of intrapelvic lymph nodes |
| SNOMED CT | 188516007 | Burkitt tumor of spleen |
| SNOMED CT | 188517003 | Burkitt tumor of lymph nodes of multiple sites |
| SNOMED CT | 188524002 | Hodgkin paragranuloma of intrathoracic lymph nodes |
| SNOMED CT | 188529007 | Hodgkin paragranuloma of intrapelvic lymph nodes |
| SNOMED CT | 188531003 | Hodgkin paragranuloma of lymph nodes of multiple sites |
| SNOMED CT | 188536008 | Hodgkin granuloma of intra-abdominal lymph nodes |
| SNOMED CT | 188537004 | Hodgkin granuloma of lymph nodes of axilla and upper limb |
| SNOMED CT | 188538009 | Hodgkin granuloma of lymph nodes of inguinal region and lower limb |
| SNOMED CT | 188541000 | Hodgkin granuloma of lymph nodes of multiple sites |
| SNOMED CT | 188547001 | Hodgkin sarcoma of lymph nodes of axilla and upper limb |
| SNOMED CT | 188548006 | Hodgkin sarcoma of lymph nodes of inguinal region and lower limb |
| SNOMED CT | 188551004 | Hodgkin sarcoma of lymph nodes of multiple sites |
| SNOMED CT | 188554007 | Hodgkin disease, lymphocytic-histiocytic predominance of lymph nodes of head, face and neck |
| SNOMED CT | 188558005 | Hodgkin disease, lymphocytic-histiocytic predominance of lymph nodes of axilla and upper limb |
| SNOMED CT | 188559002 | Hodgkin disease, lymphocytic-histiocytic predominance of lymph nodes of inguinal region and lower limb |
| SNOMED CT | 188562004 | Hodgkin disease, lymphocytic-histiocytic predominance of lymph nodes of multiple sites |
| SNOMED CT | 188565002 | Hodgkin disease, nodular sclerosis of lymph nodes of head, face and neck |
| SNOMED CT | 188566001 | Hodgkin disease, nodular sclerosis of intrathoracic lymph nodes |
| SNOMED CT | 188567005 | Hodgkin disease, nodular sclerosis of intra-abdominal lymph nodes |
| SNOMED CT | 188568000 | Hodgkin disease, nodular sclerosis of lymph nodes of axilla and upper limb |

Continued on next page

**TableS24 – continued from previous page**

| CodeSystem | ConceptCode | ConceptName |
|---|---|---|
| SNOMED CT | 188569008 | Hodgkin disease, nodular sclerosis of lymph nodes of inguinal region and lower limb |
| SNOMED CT | 188570009 | Hodgkin disease, nodular sclerosis of intrapelvic lymph nodes |
| SNOMED CT | 188572001 | Hodgkin disease, nodular sclerosis of lymph nodes of multiple sites |
| SNOMED CT | 188575004 | Hodgkin disease, mixed cellularity of lymph nodes of head, face and neck |
| SNOMED CT | 188576003 | Hodgkin disease, mixed cellularity of intrathoracic lymph nodes |
| SNOMED CT | 188577007 | Hodgkin disease, mixed cellularity of intra-abdominal lymph nodes |
| SNOMED CT | 188578002 | Hodgkin disease, mixed cellularity of lymph nodes of axilla and upper limb |
| SNOMED CT | 188579005 | Hodgkin disease, mixed cellularity of lymph nodes of inguinal region and lower limb |
| SNOMED CT | 188580008 | Hodgkin disease, mixed cellularity of intrapelvic lymph nodes |
| SNOMED CT | 188582000 | Hodgkin disease, mixed cellularity of lymph nodes of multiple sites |
| SNOMED CT | 188585003 | Hodgkin disease, lymphocytic depletion of lymph nodes of head, face and neck |
| SNOMED CT | 188586002 | Hodgkin disease, lymphocytic depletion of intrathoracic lymph nodes |
| SNOMED CT | 188587006 | Hodgkin disease, lymphocytic depletion of intra-abdominal lymph nodes |
| SNOMED CT | 188589009 | Hodgkin disease, lymphocytic depletion of lymph nodes of axilla and upper limb |
| SNOMED CT | 188590000 | Hodgkin disease, lymphocytic depletion of lymph nodes of inguinal region and lower limb |
| SNOMED CT | 188591001 | Hodgkin disease, lymphocytic depletion of intrapelvic lymph nodes |
| SNOMED CT | 188592008 | Hodgkin disease, lymphocytic depletion of spleen |
| SNOMED CT | 188593003 | Hodgkin disease, lymphocytic depletion of lymph nodes of multiple sites |
| SNOMED CT | 188632001 | Lymphoma intra abdominal, Sezarys |
| SNOMED CT | 188672005 | Follicular non-Hodgkin mixed small cleaved and large cell lymphoma |
| SNOMED CT | 188679001 | Diffuse non-Hodgkin lymphoma undifferentiated (diffuse) |
| SNOMED CT | 231830001 | Bowen disease of eyelid |
| SNOMED CT | 231835006 | Kaposi sarcoma of eyelid |
| SNOMED CT | 240531002 | African Burkitt lymphoma |
| SNOMED CT | 255114007 | Kaposi sarcoma of conjunctiva |
| SNOMED CT | 255115008 | Kaposi sarcoma of cornea |
| SNOMED CT | 255121007 | Peritoneal carcinomatosis |
| SNOMED CT | 276876007 | Carcinoma of Bartholin gland |
| SNOMED CT | 277550009 | Richter syndrome |
| SNOMED CT | 277609007 | Hodgkin disease, lymphocytic predominance - diffuse |
| SNOMED CT | 277610002 | Hodgkin disease, nodular sclerosis - lymphocytic predominance |
| SNOMED CT | 277611003 | Hodgkin disease, nodular sclerosis - mixed cellularity |

Continued on next page

**TableS24 – continued from previous page**

| CodeSystem | ConceptCode | ConceptName |
|---|---|---|
| SNOMED CT | 277612005 | Hodgkin disease, nodular sclerosis - lymphocytic depletion |
| SNOMED CT | 277654008 | Enteropathy-type T-cell lymphoma |
| SNOMED CT | 307608006 | Ewing sarcoma of bone |
| SNOMED CT | 307633009 | Hodgkin disease, lymphocytic depletion, diffuse fibrosis |
| SNOMED CT | 307634003 | Hodgkin disease, lymphocytic depletion, reticular type |
| SNOMED CT | 307635002 | Hodgkin disease, nodular sclerosis - cellular phase |
| SNOMED CT | 308121000 | Follicular non-Hodgkin lymphoma |
| SNOMED CT | 315058005 | Lynch syndrome |
| SNOMED CT | 359631009 | M0 - Acute myeloblastic leukemia - undifferentiated |
| SNOMED CT | 359640008 | Acute myelogenous leukemia without maturation |
| SNOMED CT | 359648001 | M2 - Acute myeloblastic leukemia with maturation |
| SNOMED CT | 363510005 | CA - Cancer of large bowel |
| SNOMED CT | 372131006 | Ca upper limb bones/scapula |
| SNOMED CT | 402881008 | Cutaneous B-cell lymphoma |
| SNOMED CT | 404106004 | Lymphomatoid papulosis with Hodgkin disease |
| SNOMED CT | 404122003 | Leukaemic infiltration of skin (chronic T-cell lymphocytic leukaemia) |
| SNOMED CT | 404123008 | Leukaemic infiltration of skin (T-cell prolymphocytic leukaemia) |
| SNOMED CT | 404124002 | Leukaemic infiltration of skin (T-cell lymphoblastic leukaemia) |
| SNOMED CT | 404135007 | Angiocentric lymphoma involving skin |
| SNOMED CT | 404136008 | Aggressive natural killer-cell leukemia involving skin |
| SNOMED CT | 404143002 | Crosti's lymphoma |
| SNOMED CT | 404157000 | Specific skin infiltration in Hodgkin disease |
| SNOMED CT | 414166008 | Extranodal natural killer/T-cell lymphoma, nasal type |
| SNOMED CT | 414780005 | Mucosa-associated lymphoid tissue lymphoma of orbit |
| SNOMED CT | 420302007 | Reticulosarcoma with AIDS (acquired immunodeficiency syndrome) |
| SNOMED CT | 421283008 | Primary lymphoma of brain with AIDS (acquired immunodeficiency syndrome) |
| SNOMED CT | 424779008 | Primary Kaposi sarcoma of oral cavity |
| SNOMED CT | 445269007 | Extranodal marginal zone B-cell lymphoma of mucosa-associated lymphoid tissue |
| SNOMED CT | 733627006 | Primary cutaneous gamma-delta T-cell lymphoma |
| SNOMED CT | 271467005 | Malignant neoplasm of bone, connective tissue, skin and breast |
| SNOMED CT | 271479005 | Benign neoplasm of pituitary gland and craniopharyngeal duct |
| SNOMED CT | 276822007 | Malignant melanoma of rectum |
| SNOMED CT | 277579002 | Light chain myeloma |
| SNOMED CT | 277580004 | Non-secretory myeloma |
| SNOMED CT | 278050001 | Sarcoma of breast |
| SNOMED CT | 278054005 | Infiltrating lobular carcinoma of breast |
| SNOMED CT | 280960002 | Benign tumor of lacrimal drainage structure |
| SNOMED CT | 285610008 | Metastasis to large intestine of unknown primary |
| SNOMED CT | 285612000 | Metastasis to rectum of unknown primary |
| SNOMED CT | 285638000 | Metastasis to vagina of unknown primary |
| SNOMED CT | 286893002 | Carcinoma of breast - upper, inner quadrant |
| SNOMED CT | 286894008 | Carcinoma of breast - lower, inner quadrant |
| SNOMED CT | 286895009 | Carcinoma of breast - upper, outer quadrant |

Continued on next page

**TableS24 – continued from previous page**

| CodeSystem | ConceptCode | ConceptName |
|---|---|---|
| SNOMED CT | 286896005 | Carcinoma breast - lower, outer quadrant |
| SNOMED CT | 286897001 | Carcinoma of breast - axillary tail |
| SNOMED CT | 307591004 | Malignant mastocytosis |
| SNOMED CT | 311779007 | Malignant neoplasm of skin of scapular region |
| SNOMED CT | 313250007 | Malignant neoplasm of lower eyelid |
| SNOMED CT | 313427003 | Lambda light chain myeloma |
| SNOMED CT | 314955001 | Local recurrence of malignant tumor of breast |
| SNOMED CT | 314966008 | Local recurrence of malignant tumor of rectum |
| SNOMED CT | 314987003 | Metastasis from malignant melanoma of skin |
| SNOMED CT | 359780007 | Metastatic malignant neoplasm to lateral axillary lymph nodes |
| SNOMED CT | 363350007 | Malignant tumor of cecum |
| SNOMED CT | 363351006 | Malignant tumor of rectum |
| SNOMED CT | 363411007 | Malignant tumor of appendix |
| SNOMED CT | 363450006 | Malignant tumor of foreskin |
| SNOMED CT | 369448007 | Malignant tumor involving rectum by direct extension from endometrium |
| SNOMED CT | 369449004 | Malignant tumor involving rectum by direct extension from fallopian tube |
| SNOMED CT | 369450004 | Malignant tumor involving rectum by direct extension from ovary |
| SNOMED CT | 369451000 | Malignant tumor involving rectum by direct extension from prostate |
| SNOMED CT | 369452007 | Malignant tumor involving rectum by direct extension from uterine cervix |
| SNOMED CT | 369453002 | Malignant tumor involving rectum by direct extension from uterus |
| SNOMED CT | 369454008 | Malignant tumor involving rectum by direct extension from vagina |
| SNOMED CT | 369455009 | Malignant tumor involving rectum by separate metastasis from endometrium |
| SNOMED CT | 369456005 | Malignant tumor involving rectum by separate metastasis from fallopian tube |
| SNOMED CT | 369457001 | Malignant tumor involving rectum by separate metastasis from ovary |
| SNOMED CT | 369458006 | Malignant tumor involving rectum by separate metastasis from prostate |
| SNOMED CT | 369459003 | Malignant tumor involving rectum by separate metastasis from uterine cervix |
| SNOMED CT | 369460008 | Malignant tumor involving rectum by separate metastasis from uterus |
| SNOMED CT | 369461007 | Malignant tumor involving rectum by separate metastasis from vagina |
| SNOMED CT | 369473007 | Malignant tumor involving bladder by direct extension from uterine cervix |
| SNOMED CT | 369474001 | Malignant tumor involving bladder by direct extension from uterus |
| SNOMED CT | 369488002 | Secondary malignant neoplasm of seminal vesicle |

Continued on next page

**TableS24 – continued from previous page**

| CodeSystem | ConceptCode | ConceptName |
|---|---|---|
| SNOMED CT | 369514009 | Secondary malignant neoplasm of left fallopian tube |
| SNOMED CT | 369521009 | Secondary malignant neoplasm of right fallopian tube |
| SNOMED CT | 369523007 | Secondary malignant neoplasm of left ovary |
| SNOMED CT | 369530001 | Secondary malignant neoplasm of right ovary |
| SNOMED CT | 369535006 | Secondary neoplasm of left broad ligament |
| SNOMED CT | 369536007 | Secondary neoplasm of right broad ligament |
| SNOMED CT | 369542006 | Malignant tumor involving left fallopian tube by separate metastasis from endometrium |
| SNOMED CT | 369543001 | Malignant tumor involving left fallopian tube by separate metastasis from ovary |
| SNOMED CT | 369544007 | Malignant tumor involving left fallopian tube by separate metastasis from right fallopian tube |
| SNOMED CT | 369545008 | Malignant tumor involving left fallopian tube by separate metastasis from uterus |
| SNOMED CT | 369546009 | Malignant tumor involving left fallopian tube by separate metastasis from vagina |
| SNOMED CT | 369553000 | Malignant tumor involving right fallopian tube by separate metastasis from endometrium |
| SNOMED CT | 369554006 | Malignant tumor involving right fallopian tube by separate metastasis from left fallopian tube |
| SNOMED CT | 369555007 | Malignant tumor involving right fallopian tube by separate metastasis from ovary |
| SNOMED CT | 369556008 | Malignant tumor involving right fallopian tube by separate metastasis from uterine cervix |
| SNOMED CT | 369557004 | Malignant tumor involving right fallopian tube by separate metastasis from uterus |
| SNOMED CT | 369558009 | Malignant tumor involving right fallopian tube by separate metastasis from vagina |
| SNOMED CT | 369560006 | Malignant tumor involving left ovary by separate metastasis from endometrium |
| SNOMED CT | 369561005 | Malignant tumor involving left ovary by separate metastasis from fallopian tube |
| SNOMED CT | 369562003 | Malignant tumor involving left ovary by separate metastasis from right ovary |
| SNOMED CT | 369563008 | Malignant tumor involving left ovary by separate metastasis from uterine cervix |
| SNOMED CT | 369564002 | Malignant tumor involving left ovary by separate metastasis from vagina |
| SNOMED CT | 369565001 | Malignant tumor involving left ovary by separate metastasis uterus |
| SNOMED CT | 369568004 | Malignant tumor involving right ovary by separate metastasis from endometrium |
| SNOMED CT | 369569007 | Malignant tumor involving right ovary by separate metastasis from fallopian tube |
| SNOMED CT | 369570008 | Malignant tumor involving right ovary by separate metastasis from left ovary |

Continued on next page

**TableS24 – continued from previous page**

| CodeSystem | ConceptCode | ConceptName |
|---|---|---|
| SNOMED CT | 369571007 | Malignant tumor involving right ovary by separate metastasis from uterine cervix |
| SNOMED CT | 369572000 | Malignant tumor involving right ovary by separate metastasis from uterus |
| SNOMED CT | 369573005 | Malignant tumor involving right ovary by separate metastasis from vagina |
| SNOMED CT | 369581006 | Malignant tumor involving vagina by separate metastasis from bladder |
| SNOMED CT | 369582004 | Malignant tumor involving vagina by separate metastasis from endometrium |
| SNOMED CT | 369583009 | Malignant tumor involving vagina by separate metastasis from fallopian tube |
| SNOMED CT | 369584003 | Malignant tumor involving vagina by separate metastasis from ovary |
| SNOMED CT | 369585002 | Malignant tumor involving vagina by separate metastasis from uterine cervix |
| SNOMED CT | 369586001 | Malignant tumor involving vagina by separate metastasis from uterus |
| SNOMED CT | 369588000 | Malignant tumor involving vulva by separate metastasis from endometrium |
| SNOMED CT | 369589008 | Malignant tumor involving vulva by separate metastasis from fallopian tube |
| SNOMED CT | 369590004 | Malignant tumor involving vulva by separate metastasis from ovary |
| SNOMED CT | 369591000 | Malignant tumor involving vulva by separate metastasis from uterine cervix |
| SNOMED CT | 369592007 | Malignant tumor involving vulva by separate metastasis from uterus |
| SNOMED CT | 369593002 | Malignant tumor involving vulva by separate metastasis from vagina |
| SNOMED CT | 369610009 | Malignant tumor involving left fallopian tube by separate metastasis from uterine cervix |
| SNOMED CT | 371977004 | Primary malignant neoplasm of cecum |
| SNOMED CT | 372023003 | Primary malignant neoplasm of upper third of esophagus |
| SNOMED CT | 372092003 | Primary malignant neoplasm of axillary tail of breast |
| SNOMED CT | 372093008 | Secondary malignant neoplasm of axillary tail of breast |
| SNOMED CT | 372094002 | Malignant neoplasm of axillary tail of breast |
| SNOMED CT | 372095001 | Malignant neoplasm of male breast |
| SNOMED CT | 372096000 | Carcinoma of male breast |
| SNOMED CT | 372112000 | Primary malignant neoplasm of middle lobe, bronchus or lung |
| SNOMED CT | 372113005 | Carcinoma of middle lobe, bronchus or lung |
| SNOMED CT | 372135002 | Primary malignant neoplasm of upper lobe, bronchus or lung |
| SNOMED CT | 372136001 | Carcinoma of upper lobe, bronchus or lung |
| SNOMED CT | 372137005 | Primary malignant neoplasm of breast |
| SNOMED CT | 373080008 | Malignant neoplasm of breast lower inner quadrant |
| SNOMED CT | 373081007 | Malignant neoplasm of breast lower outer quadrant |
| SNOMED CT | 373082000 | Malignant neoplasm of breast upper inner quadrant |

Continued on next page

**TableS24 – continued from previous page**

| CodeSystem | ConceptCode | ConceptName |
|---|---|---|
| SNOMED CT | 373083005 | Malignant neoplasm of breast upper outer quadrant |
| SNOMED CT | 373088001 | Primary malignant neoplasm of breast upper outer quadrant |
| SNOMED CT | 373089009 | Primary malignant neoplasm of breast upper inner quadrant |
| SNOMED CT | 373090000 | Primary malignant neoplasm of breast lower inner quadrant |
| SNOMED CT | 373091001 | Primary malignant neoplasm of breast lower outer quadrant |
| SNOMED CT | 385427008 | Osteochondroma of elbow |
| SNOMED CT | 385428003 | Osteochondroma of femur |
| SNOMED CT | 385429006 | Osteochondroma of foot |
| SNOMED CT | 385430001 | Osteochondroma of scapula |
| SNOMED CT | 385431002 | Osteochondroma of tibia |
| SNOMED CT | 388872005 | Secondary optic nerve sheath meningioma |
| SNOMED CT | 389272007 | Carpotarsal osteochondromatosis |
| SNOMED CT | 402517006 | Basal cell carcinoma of upper back |
| SNOMED CT | 402841001 | Angiokeratoma of vulva |
| SNOMED CT | 403274000 | Hypomelanosis surrounding malignant melanoma |
| SNOMED CT | 403458008 | Localized skin involvement by breast carcinoma |
| SNOMED CT | 403918002 | Basal cell carcinoma of glabella |
| SNOMED CT | 403946000 | Paget's disease of nipple |
| SNOMED CT | 404145009 | Primary cutaneous anaplastic large cell B-cell lymphoma |
| SNOMED CT | 406103009 | Squamous cell carcinoma in situ of uterine cervix |
| SNOMED CT | 408643008 | Infiltrating duct carcinoma of breast |
| SNOMED CT | 408645001 | Adenocarcinoma of large intestine |
| SNOMED CT | 413445002 | Adenocarcinoma of appendix |
| SNOMED CT | 413446001 | Adenocarcinoma of cecum |
| SNOMED CT | 413587002 | Smoldering myeloma |
| SNOMED CT | 414553000 | Kappa light chain myeloma |
| SNOMED CT | 415110002 | Plasma cell myeloma/plasmacytoma |
| SNOMED CT | 416769008 | Malignant teratoma of testis |
| SNOMED CT | 416921001 | Carcinoma in situ of nasolacrimal duct |
| SNOMED CT | 417181009 | Hormone receptor positive malignant neoplasm of breast |
| SNOMED CT | 417234002 | Benign sacral teratoma |
| SNOMED CT | 418529003 | Secondary malignant neoplasm of lacrimal drainage structure |
| SNOMED CT | 422607004 | Carcinoma in situ of lacrimal drainage system |
| SNOMED CT | 422893006 | Carcinoma in situ of lacrimal gland duct |
| SNOMED CT | 423318000 | Adenoid cystic carcinoma of oropharynx |
| SNOMED CT | 423384009 | Secondary malignant neoplasm of lacrimal gland duct |
| SNOMED CT | 423535002 | Basal cell carcinoma of chest wall |
| SNOMED CT | 423793008 | Mucoepidermoid carcinoma of parotid gland |
| SNOMED CT | 425657001 | Osteosclerotic myeloma |
| SNOMED CT | 427685000 | HER2-positive carcinoma of breast |
| SNOMED CT | 431396003 | Human epidermal growth factor 2 negative carcinoma of breast |
| SNOMED CT | 440422002 | Asymptomatic multiple myeloma |
| SNOMED CT | 441313008 | Indolent multiple myeloma |
| SNOMED CT | 443333004 | Medulloblastoma |
| SNOMED CT | 444374006 | Type C thymoma |
| SNOMED CT | 444596001 | Malignant thymoma |
| SNOMED CT | 444604002 | Mixed ductal and lobular carcinoma of breast |

Continued on next page

**TableS24 – continued from previous page**

| CodeSystem | ConceptCode | ConceptName |
|---|---|---|
| SNOMED CT | 444712000 | Mucinous carcinoma of breast |
| SNOMED CT | 445105005 | Blastic plasmacytoid dendritic cell neoplasm |
| SNOMED CT | 446939001 | Chordoma of clivus |
| SNOMED CT | 446945009 | Invasive pituitary adenoma |
| SNOMED CT | 447729009 | Ameloblastoma of mandible |
| SNOMED CT | 447730004 | Chordoma of sacrum |
| SNOMED CT | 447792005 | Chondrosarcoma of bone |
| SNOMED CT | 448135004 | Benign teratoma of pineal region |
| SNOMED CT | 448257000 | Sarcoma of male breast |
| SNOMED CT | 448296006 | Dermatofibrosarcoma protuberans of skin of chest |
| SNOMED CT | 448314007 | Carcinoma of spinal cord |
| SNOMED CT | 448563005 | Functionless pituitary neoplasm |
| SNOMED CT | 448992002 | Carcinoma of appendix |
| SNOMED CT | 448994001 | Carcinoma of upper rectum |
| SNOMED CT | 449627008 | Malignant neoplasm of long bone of lower limb |
| SNOMED CT | 449799008 | Subependymal giant cell astrocytoma |
| SNOMED CT | 449803009 | Primary malignant neoplasm of dome of urinary bladder |
| SNOMED CT | 608817003 | Pituicytoma |
| SNOMED CT | 697993003 | Undifferentiated carcinoma of nasal sinus |
| SNOMED CT | 698011002 | Keratinizing squamous cell carcinoma of nasopharynx |
| SNOMED CT | 698040004 | Malignant melanoma of nasal cavity |
| SNOMED CT | 698041000 | Malignant melanoma of vestibule of mouth |
| SNOMED CT | 698042007 | Malignant melanoma of tongue |
| SNOMED CT | 698043002 | Malignant melanoma of floor of mouth |
| SNOMED CT | 698044008 | Malignant melanoma of palatine arch |
| SNOMED CT | 698045009 | Malignant melanoma of buccal mucosa |
| SNOMED CT | 698048006 | Undifferentiated nonkeratinizing carcinoma of nasopharynx |
| SNOMED CT | 698200003 | Large cell Ewing sarcoma of bone |
| SNOMED CT | 698285005 | Malignant melanoma of ethmoid sinus |
| SNOMED CT | 698286006 | Malignant melanoma of palate |
| SNOMED CT | 698287002 | Malignant melanoma of gum |
| SNOMED CT | 698288007 | Malignant melanoma of maxillary sinus |
| SNOMED CT | 698770000 | Multicystic intraosseous ameloblastoma |
| SNOMED CT | 699317002 | Paratesticular malignant neoplasm |
| SNOMED CT | 699318007 | Supratentorial primitive neuroectodermal tumor |
| SNOMED CT | 699331002 | Granular cell tumor of neurohypophysis |
| SNOMED CT | 699354006 | Sarcoma of orbit |
| SNOMED CT | 699355007 | Leiomyosarcoma of orbit |
| SNOMED CT | 699356008 | Endometrial stromal sarcoma |
| SNOMED CT | 699357004 | Low grade endometrial stromal sarcoma |
| SNOMED CT | 699358009 | High grade endometrial stromal sarcoma |
| SNOMED CT | 699704002 | Classic medulloblastoma |
| SNOMED CT | 700057001 | Emberger syndrome |
| SNOMED CT | 700488005 | Malignant sex cord tumor of testis |
| SNOMED CT | 702346005 | Potocki-Shaffer syndrome |
| SNOMED CT | 702368000 | Carcinosarcoma of ovary |
| SNOMED CT | 702369008 | Carcinosarcoma of uterus |

Continued on next page

**TableS24 – continued from previous page**

| CodeSystem | ConceptCode | ConceptName |
|---|---|---|
| SNOMED CT | 702375004 | Familial isolated pituitary adenoma |
| SNOMED CT | 702391001 | Renal cell carcinoma |
| SNOMED CT | 702405001 | Malignant granulosa cell tumor of testis |
| SNOMED CT | 702467006 | Malignant neoplasm of augmented bladder |
| SNOMED CT | 703228009 | Non-small cell lung cancer with mutation in epidermal growth factor receptor |
| SNOMED CT | 703230006 | Non-small cell lung cancer without mutation in epidermal growth factor receptor |
| SNOMED CT | 703429003 | Malignant optic glioma of adulthood |
| SNOMED CT | 706970001 | Triple-negative breast cancer |
| SNOMED CT | 707337006 | Primary adenocarcinoma of accessory sinus |
| SNOMED CT | 707339009 | Primary adenocarcinoma of maxillary sinus |
| SNOMED CT | 707342003 | Primary adenocarcinoma of ethmoidal sinus |
| SNOMED CT | 707343008 | Primary adenocarcinoma of frontal sinus |
| SNOMED CT | 707344002 | Primary adenocarcinoma of sphenoidal sinus |
| SNOMED CT | 707345001 | Primary carcinoma of accessory sinus |
| SNOMED CT | 707346000 | Primary carcinoma of ethmoidal sinus |
| SNOMED CT | 707347009 | Primary carcinoma of maxillary sinus |
| SNOMED CT | 707348004 | Primary carcinoma of sphenoidal sinus |
| SNOMED CT | 707349007 | Primary carcinoma of frontal sinus |
| SNOMED CT | 707350007 | Malignant melanoma of accessory sinus |
| SNOMED CT | 707353009 | Primary squamous cell carcinoma of accessory sinus |
| SNOMED CT | 707354003 | Primary squamous cell carcinoma of maxillary sinus |
| SNOMED CT | 707355002 | Primary squamous cell carcinoma of sphenoidal sinus |
| SNOMED CT | 707356001 | Primary squamous cell carcinoma of frontal sinus |
| SNOMED CT | 707357005 | Primary squamous cell carcinoma of laryngeal cartilage |
| SNOMED CT | 707358000 | Primary squamous cell carcinoma of larynx |
| SNOMED CT | 707359008 | Primary squamous cell carcinoma of ethmoidal sinus |
| SNOMED CT | 707360003 | Primary lymphoepithelial carcinoma of larynx |
| SNOMED CT | 707361004 | Malignant melanoma of frontal sinus |
| SNOMED CT | 707362006 | Malignant melanoma of sphenoidal sinus |
| SNOMED CT | 707377003 | Primary signet ring cell carcinoma of trachea |
| SNOMED CT | 707378008 | Primary myoepithelial carcinoma of trachea |
| SNOMED CT | 707379000 | Primary mucoepidermoid carcinoma of trachea |
| SNOMED CT | 707380002 | Primary salivary gland type carcinoma of trachea |
| SNOMED CT | 707381003 | Solitary hamartoma of lung |
| SNOMED CT | 707382005 | Multiple hamartoma of lung |
| SNOMED CT | 707383000 | Primary mucinous cystadenocarcinoma of trachea |
| SNOMED CT | 707384006 | Primary solid carcinoma of trachea |
| SNOMED CT | 707385007 | Primary undifferentiated carcinoma of trachea |
| SNOMED CT | 707386008 | Primary acinar cell carcinoma of trachea |
| SNOMED CT | 707388009 | Primary squamous cell carcinoma of trachea |
| SNOMED CT | 707389001 | Primary clear cell squamous cell carcinoma of trachea |
| SNOMED CT | 707390005 | Primary basaloid squamous cell carcinoma of trachea |
| SNOMED CT | 707391009 | Primary papillary squamous cell carcinoma of trachea |
| SNOMED CT | 707392002 | Primary giant cell carcinoma of trachea |
| SNOMED CT | 707393007 | Primary adenosquamous carcinoma of trachea |

Continued on next page

**TableS24 – continued from previous page**

| CodeSystem | ConceptCode | ConceptName |
|---|---|---|
| SNOMED CT | 707394001 | Primary spindle cell carcinoma of trachea |
| SNOMED CT | 707395000 | Primary adenocarcinoma of hypopharynx |
| SNOMED CT | 707396004 | Primary oxyphilic adenocarcinoma of oropharynx |
| SNOMED CT | 707397008 | Primary basal cell adenocarcinoma of oropharynx |
| SNOMED CT | 707398003 | Primary polymorphous low grade adenocarcinoma of oropharynx |
| SNOMED CT | 707399006 | Primary papillary adenocarcinoma of oropharynx |
| SNOMED CT | 707400004 | Primary mucinous adenocarcinoma of oropharynx |
| SNOMED CT | 707401000 | Primary clear cell adenocarcinoma of oropharynx |
| SNOMED CT | 707402007 | Primary adenocarcinoma of oropharynx |
| SNOMED CT | 707403002 | Primary fetal adenocarcinoma of lung |
| SNOMED CT | 707404008 | Primary mixed subtype adenocarcinoma of lung |
| SNOMED CT | 707405009 | Primary adenosquamous carcinoma of lung |
| SNOMED CT | 707406005 | Primary mucoepidermoid carcinoma of hypopharynx |
| SNOMED CT | 707407001 | Primary signet ring cell carcinoma of lung |
| SNOMED CT | 707408006 | Primary small cell non-keratinizing squamous cell carcinoma of lung |
| SNOMED CT | 707409003 | Primary acinar cell carcinoma of lung |
| SNOMED CT | 707410008 | Primary solid carcinoma of lung |
| SNOMED CT | 707411007 | Primary papillary adenocarcinoma of lung |
| SNOMED CT | 707421004 | Primary undifferentiated carcinoma of larynx |
| SNOMED CT | 707422006 | Primary spindle cell squamous cell carcinoma of larynx |
| SNOMED CT | 707423001 | Primary basaloid carcinoma of larynx |
| SNOMED CT | 707424007 | Primary adenosquamous cell carcinoma of larynx |
| SNOMED CT | 707425008 | Primary adenoid squamous cell carcinoma of larynx |
| SNOMED CT | 707426009 | Primary papillary squamous cell carcinoma of larynx |
| SNOMED CT | 707427000 | Primary verrucous carcinoma of larynx |
| SNOMED CT | 707429002 | Overlapping squamous cell carcinoma of larynx |
| SNOMED CT | 707430007 | Overlapping squamous cell carcinoma of laryngeal cartilage |
| SNOMED CT | 707451005 | Primary adenocarcinoma of lung |
| SNOMED CT | 707452003 | Primary mucinous adenocarcinoma of lung |
| SNOMED CT | 707453008 | Primary clear cell squamous cell carcinoma of lung |
| SNOMED CT | 707454002 | Primary basaloid squamous cell carcinoma of lung |
| SNOMED CT | 707455001 | Primary papillary squamous cell carcinoma of lung |
| SNOMED CT | 707456000 | Primary undifferentiated carcinoma of lung |
| SNOMED CT | 707457009 | Primary spindle cell carcinoma of lung |
| SNOMED CT | 707458004 | Primary pleomorphic carcinoma of lung |
| SNOMED CT | 707460002 | Primary pseudosarcomatous carcinoma of lung |
| SNOMED CT | 707464006 | Primary myoepithelial carcinoma of lung |
| SNOMED CT | 707465007 | Primary mucoepidermoid carcinoma of lung |
| SNOMED CT | 707466008 | Primary adenoid cystic carcinoma of lung |
| SNOMED CT | 707467004 | Primary salivary gland type carcinoma of lung |
| SNOMED CT | 707468009 | Primary mixed mucinous and non-mucinous bronchiolo-alveolar carcinoma of lung |
| SNOMED CT | 707469001 | Primary non-mucinous bronchiolo-alveolar carcinoma of lung |
| SNOMED CT | 707470000 | Primary mucinous bronchiolo-alveolar carcinoma of lung |
| SNOMED CT | 707471001 | Primary clear cell adenocarcinoma of trachea |
| SNOMED CT | 707472008 | Primary papillary adenocarcinoma of trachea |

Continued on next page

**TableS24 – continued from previous page**

| CodeSystem | ConceptCode | ConceptName |
|---|---|---|
| SNOMED CT | 707473003 | Primary mucinous adenocarcinoma of trachea |
| SNOMED CT | 707475005 | Primary adenocarcinoma of trachea |
| SNOMED CT | 707479004 | Primary adenocarcinoma of subglottis |
| SNOMED CT | 707481002 | Primary basaloid squamous cell carcinoma of hypopharynx |
| SNOMED CT | 707482009 | Primary papillary squamous cell carcinoma of hypopharynx |
| SNOMED CT | 707483004 | Primary undifferentiated carcinoma of hypopharynx |
| SNOMED CT | 707484005 | Primary adenoid squamous cell carcinoma of hypopharynx |
| SNOMED CT | 707485006 | Primary adenosquamous carcinoma of hypopharynx |
| SNOMED CT | 707486007 | Primary basaloid carcinoma of hypopharynx |
| SNOMED CT | 707487003 | Primary giant cell carcinoma of hypopharynx |
| SNOMED CT | 707489000 | Primary spindle cell squamous cell carcinoma of hypopharynx |
| SNOMED CT | 707490009 | Primary verrucous carcinoma of hypopharynx |
| SNOMED CT | 707491008 | Primary lymphoepithelial carcinoma of hypopharynx |
| SNOMED CT | 707492001 | Primary squamous cell carcinoma of hypopharynx |
| SNOMED CT | 707493006 | Primary lymphoepithelial carcinoma of trachea |
| SNOMED CT | 707494000 | Primary verrucous carcinoma of trachea |
| SNOMED CT | 707495004 | Primary squamous cell adenoid carcinoma of trachea |
| SNOMED CT | 707528007 | Primary squamous cell carcinoma of nasopharynx |
| SNOMED CT | 707529004 | Overlapping squamous cell carcinoma of oropharynx |
| SNOMED CT | 707530009 | Cystic hamartoma of lung and kidney |
| SNOMED CT | 707532001 | Primary squamous cell carcinoma of posterior wall of oropharynx |
| SNOMED CT | 707535004 | Primary squamous cell carcinoma of lateral wall of oropharynx |
| SNOMED CT | 707537007 | Primary squamous cell carcinoma of anterior surface of epiglottis |
| SNOMED CT | 707538002 | Primary squamous cell carcinoma of vallecula |
| SNOMED CT | 707539005 | Primary adenoid cystic carcinoma of hypopharynx |
| SNOMED CT | 707575007 | Primary squamous cell carcinoma of supraglottis |
| SNOMED CT | 707576008 | Primary squamous cell carcinoma of subglottis |
| SNOMED CT | 707579001 | Primary basaloid squamous cell carcinoma of oropharynx |
| SNOMED CT | 707580003 | Primary basaloid carcinoma of oropharynx |
| SNOMED CT | 707581004 | Primary papillary squamous cell carcinoma of oropharynx |
| SNOMED CT | 707582006 | Primary spindle cell squamous cell carcinoma of oropharynx |
| SNOMED CT | 707583001 | Primary adenosquamous carcinoma of oropharynx |
| SNOMED CT | 707584007 | Primary lymphoepithelial carcinoma of oropharynx |
| SNOMED CT | 707585008 | Primary squamous cell carcinoma of oropharynx |
| SNOMED CT | 707586009 | Primary myoepithelial carcinoma of oropharynx |
| SNOMED CT | 707587000 | Primary carcinoma ex pleomorphic adenoma of oropharynx |
| SNOMED CT | 707588005 | Primary epithelial-myoepithelial carcinoma of oropharynx |
| SNOMED CT | 707589002 | Primary cystadenocarcinoma of oropharynx |
| SNOMED CT | 707590006 | Primary acinar cell carcinoma of oropharynx |
| SNOMED CT | 707591005 | Primary mucoepidermoid carcinoma of oropharynx |
| SNOMED CT | 707592003 | Primary infiltrating duct carcinoma of oropharynx |
| SNOMED CT | 707593008 | Primary salivary gland-type tumor of oropharynx |
| SNOMED CT | 707595001 | Primary mucinous cystadenocarcinoma of lung |
| SNOMED CT | 707596000 | Primary carcinosarcoma of lung |
| SNOMED CT | 707627009 | Primary salivary gland type carcinoma of hypopharynx |
| SNOMED CT | 707628004 | Overlapping squamous cell carcinoma of hypopharynx |
| SNOMED CT | 707660007 | Primary giant cell carcinoma of larynx |

Continued on next page

**TableS24 – continued from previous page**

| CodeSystem | ConceptCode | ConceptName |
|---|---|---|
| SNOMED CT | 707662004 | Primary basaloid squamous cell carcinoma of larynx |
| SNOMED CT | 707664003 | Primary squamous cell carcinoma of glottis |
| SNOMED CT | 707670009 | Pleuropulmonary blastoma |
| SNOMED CT | 707671008 | Pleuropulmonary blastoma type I |
| SNOMED CT | 707672001 | Pleuropulmonary blastoma type II |
| SNOMED CT | 707673006 | Pleuropulmonary blastoma type III |
| SNOMED CT | 707674000 | Primary malignant epithelial neoplasm of trachea |
| SNOMED CT | 707686002 | Primary squamous cell carcinoma of posterior wall of hypopharynx |
| SNOMED CT | 707697002 | Primary squamous cell carcinoma of hypopharyngeal aspect of aryepiglottic fold |
| SNOMED CT | 707703001 | Primary squamous cell carcinoma of postcricoid region |
| SNOMED CT | 707704007 | Primary squamous cell carcinoma of pyriform sinus |
| SNOMED CT | 707705008 | Nonkeratinizing carcinoma of the nasopharynx |
| SNOMED CT | 708504008 | Periosteal osteosarcoma of jaw |
| SNOMED CT | 708921005 | Carcinoma of central portion of breast |
| SNOMED CT | 708971008 | Diffuse sclerosing papillary thyroid carcinoma |
| SNOMED CT | 709031009 | Malignant neoplasm of superior wall of nasopharynx |
| SNOMED CT | 709830006 | Malignant carcinoid tumor of stomach |
| SNOMED CT | 710193006 | Benign odontogenic tumor of lower jaw |
| SNOMED CT | 710194000 | Benign odontogenic tumor of upper jaw |
| SNOMED CT | 710195004 | Malignant odontogenic neoplasm of lower jaw |
| SNOMED CT | 710196003 | Malignant odontogenic tumor of upper jaw |
| SNOMED CT | 711414003 | Primary clear cell adenocarcinoma of lung |
| SNOMED CT | 712525007 | Malignant neoplasm of short bone of lower limb |
| SNOMED CT | 712750007 | Malignant neoplasm of chest wall |
| SNOMED CT | 712849003 | Prostate cancer metastatic to bone |
| SNOMED CT | 713038003 | Overlapping primary malignant neoplasm of bone and articular cartilage of upper limb |
| SNOMED CT | 713290004 | Malignant ameloblastoma of mandible |
| SNOMED CT | 713293002 | Malignant germ cell neoplasm of mediastinum |
| SNOMED CT | 713327005 | Malignant meningioma of meninges of brain |
| SNOMED CT | 713573006 | Malignant carcinoid tumor of rectum |
| SNOMED CT | 713574000 | Malignant carcinoid tumor of kidney |
| SNOMED CT | 713609000 | Invasive carcinoma of breast |
| SNOMED CT | 713646001 | Malignant germ cell tumor of testis |
| SNOMED CT | 715215007 | WAGR syndrome |
| SNOMED CT | 715412008 | Familial prostate cancer |
| SNOMED CT | 715634002 | Florid cemento-osseous dysplasia |
| SNOMED CT | 715904005 | Pineal parenchymal tumor of intermediate differentiation |
| SNOMED CT | 715907003 | Multiple endocrine neoplasia type 4 |
| SNOMED CT | 716586009 | Epstein-Barr virus associated gastric carcinoma |
| SNOMED CT | 716588005 | Primary non-gestational choriocarcinoma of ovary |
| SNOMED CT | 716593008 | Carcinoma of salivary gland type of breast |
| SNOMED CT | 716649003 | Extraovarian primary peritoneal carcinoma |
| SNOMED CT | 716653001 | Neuroendocrine carcinoma of thymus |
| SNOMED CT | 716655008 | Aggressive systemic mastocytosis |

Continued on next page

**TableS24 – continued from previous page**

| CodeSystem | ConceptCode | ConceptName |
|---|---|---|
| SNOMED CT | 716657000 | Familial papillary thyroid carcinoma with renal papillary neoplasia syndrome |
| SNOMED CT | 716742001 | Multiple osteochondroma |
| SNOMED CT | 716855006 | Theca steroid producing cell malignant neoplasm of ovary |
| SNOMED CT | 716859000 | Hereditary diffuse carcinoma of stomach |
| SNOMED CT | 717003001 | Hereditary cavernous hemangioma of brain |
| SNOMED CT | 717734005 | Papillary thyroid carcinoma with renal papillary neoplasia |
| SNOMED CT | 717735006 | Renal cell carcinoma of kidney except renal pelvis |
| SNOMED CT | 717736007 | Familial renal cell carcinoma |
| SNOMED CT | 717916003 | Neuroendocrine carcinoma of appendix |
| SNOMED CT | 717921000 | Poorly-differentiated neuroendocrine carcinoma of thymus |
| SNOMED CT | 717922007 | Well-differentiated neuroendocrine carcinoma of thymus |
| SNOMED CT | 717968005 | Melanoma and neural system tumor syndrome |
| SNOMED CT | 718220008 | Hereditary breast and ovarian cancer syndrome |
| SNOMED CT | 718604008 | Small cell neuroendocrine carcinoma of bladder |
| SNOMED CT | 721188000 | Multiple endocrine neoplasia type 2A |
| SNOMED CT | 721311006 | Systemic Epstein-Barr virus positive T-cell lymphoproliferative disease of childhood |
| SNOMED CT | 721546004 | Primary adenocarcinoma of ciliary epithelium |
| SNOMED CT | 721547008 | Primary adenocarcinoma of epithelium of iris |
| SNOMED CT | 721548003 | Primary malignant neoplasm of lacrimal apparatus |
| SNOMED CT | 721549006 | Primary adenocarcinoma of lacrimal apparatus |
| SNOMED CT | 721556000 | Primary adenocarcinoma of palate |
| SNOMED CT | 721557009 | Primary adenocarcinoma of parotid gland |
| SNOMED CT | 721560002 | Primary adenocarcinoma of nasal cavity |
| SNOMED CT | 721561003 | Primary adenocarcinoma of middle ear |
| SNOMED CT | 721562005 | Primary adenocarcinoma of overlapping sites of retroperitoneum, peritoneum and omentum |
| SNOMED CT | 721563000 | Primary malignant melanoma of vagina |
| SNOMED CT | 721568009 | Primary adenocarcinoma of parametrium |
| SNOMED CT | 721569001 | Primary adenocarcinoma of uterine ligament |
| SNOMED CT | 721571001 | Benign endometrial stromal neoplasm |
| SNOMED CT | 721574009 | Primary rhabdomyosarcoma of male genital organ |
| SNOMED CT | 721575005 | Primary angiosarcoma of heart |
| SNOMED CT | 721576006 | Primary angiosarcoma of breast |
| SNOMED CT | 721578007 | Primary liposarcoma of male genital organ |
| SNOMED CT | 721580001 | Primary myosarcoma of omentum |
| SNOMED CT | 721602000 | Primary embryonal carcinoma of testis |
| SNOMED CT | 721603005 | Primary choriocarcinoma of testis |
| SNOMED CT | 721604004 | Primary squamous cell carcinoma of overlapping sites of male genital organ |
| SNOMED CT | 721605003 | Primary squamous cell carcinoma of overlapping sites of accessory sinuses |
| SNOMED CT | 721606002 | Primary adenocarcinoma of overlapping sites of accessory sinuses |
| SNOMED CT | 721607006 | Primary undifferentiated carcinoma of oropharynx |
| SNOMED CT | 721617001 | Primary adenocarcinoma of lower third of esophagus due to Barrett esophagus |

Continued on next page

**TableS24 – continued from previous page**

| CodeSystem | ConceptCode | ConceptName |
|---|---|---|
| SNOMED CT | 721618006 | Primary squamous cell carcinoma of upper third of esophagus |
| SNOMED CT | 721619003 | Primary squamous cell carcinoma of middle third of esophagus |
| SNOMED CT | 721620009 | Primary squamous cell carcinoma of lower third of esophagus |
| SNOMED CT | 721621008 | Primary squamous cell carcinoma of overlapping sites of esophagus |
| SNOMED CT | 721622001 | Primary adenocarcinoma of upper third of esophagus |
| SNOMED CT | 721623006 | Primary adenocarcinoma of middle third of esophagus |
| SNOMED CT | 721624000 | Primary adenocarcinoma of overlapping sites of esophagus |
| SNOMED CT | 721625004 | Primary neuroendocrine carcinoma of esophagus |
| SNOMED CT | 721626003 | Primary malignant neuroendocrine neoplasm of esophagus |
| SNOMED CT | 721627007 | Malignant melanoma of esophagus |
| SNOMED CT | 721628002 | Primary adenocarcinoma of esophagogastric junction |
| SNOMED CT | 721629005 | Linitis plastica of stomach |
| SNOMED CT | 721630000 | Primary adenocarcinoma of cardia of stomach |
| SNOMED CT | 721632008 | Primary adenocarcinoma of pyloric antrum of stomach |
| SNOMED CT | 721633003 | Primary adenocarcinoma of overlapping sites of stomach |
| SNOMED CT | 721634009 | Primary malignant neuroendocrine neoplasm of stomach |
| SNOMED CT | 721635005 | Primary malignant neuroendocrine neoplasm of cardia of stomach |
| SNOMED CT | 721636006 | Primary malignant neuroendocrine neoplasm of body of stomach |
| SNOMED CT | 721637002 | Primary malignant neuroendocrine neoplasm of pyloric antrum of stomach |
| SNOMED CT | 721638007 | Primary neuroendocrine carcinoma of stomach |
| SNOMED CT | 721639004 | Primary neuroendocrine carcinoma of cardia of stomach |
| SNOMED CT | 721640002 | Primary neuroendocrine carcinoma of body of stomach |
| SNOMED CT | 721641003 | Primary neuroendocrine carcinoma of pyloric antrum of stomach |
| SNOMED CT | 721642005 | Primary neuroendocrine carcinoma of overlapping sites of stomach |
| SNOMED CT | 721643000 | Primary malignant mesenchymal neoplasm of stomach |
| SNOMED CT | 721644006 | Primary malignant neuroendocrine neoplasm of duodenum |
| SNOMED CT | 721645007 | Primary neuroendocrine carcinoma of duodenum |
| SNOMED CT | 721672004 | Primary mucinous adenocarcinoma of appendix |
| SNOMED CT | 721673009 | Primary malignant neuroendocrine neoplasm of appendix |
| SNOMED CT | 721697000 | Primary neuroendocrine carcinoma of colon |
| SNOMED CT | 721698005 | Primary malignant neuroendocrine neoplasm of colon |
| SNOMED CT | 721700001 | Primary malignant neuroendocrine neoplasm of rectum |
| SNOMED CT | 721701002 | Primary neuroendocrine carcinoma of rectum |
| SNOMED CT | 721709000 | Primary cloacogenic carcinoma of anal canal |
| SNOMED CT | 721718003 | Primary adenocarcinoma of ampulla of Vater |
| SNOMED CT | 721725005 | Primary adenocarcinoma of peritoneum |
| SNOMED CT | 68979007 | Heavy chain disease |
| SNOMED CT | 82546001 | Reactive immunoproliferative disease |
| SNOMED CT | 92072003 | Benign neoplasm of craniopharyngeal duct |
| SNOMED CT | 92169007 | Benign neoplasm of lacrimal gland |
| SNOMED CT | 92296004 | Benign neoplasm of pituitary gland |
| SNOMED CT | 92564006 | Carcinoma in situ of uterine cervix |
| SNOMED CT | 92573003 | Carcinoma in situ of craniopharyngeal duct |

Continued on next page

**TableS24 – continued from previous page**

| CodeSystem | ConceptCode | ConceptName |
|---|---|---|
| SNOMED CT | 92580001 | Carcinoma in situ of endocervix |
| SNOMED CT | 92588008 | Carcinoma in situ of exocervix |
| SNOMED CT | 92628002 | Carcinoma in situ of lacrimal gland |
| SNOMED CT | 92683008 | Carcinoma in situ of pituitary gland |
| SNOMED CT | 92791005 | Carcinoma in situ of vagina |
| SNOMED CT | 93675008 | Primary malignant neoplasm of anterior wall of urinary bladder |
| SNOMED CT | 93679002 | Primary malignant neoplasm of appendix |
| SNOMED CT | 93681000 | Primary malignant neoplasm of areola of male breast |
| SNOMED CT | 93683002 | Primary malignant neoplasm of ascending colon |
| SNOMED CT | 93751003 | Primary malignant neoplasm of cervical vertebral column |
| SNOMED CT | 93777006 | Primary malignant neoplasm of ectopic male breast tissue |
| SNOMED CT | 93854002 | Primary malignant neoplasm of large intestine |
| SNOMED CT | 93863000 | Primary malignant neoplasm of lateral wall of urinary bladder |
| SNOMED CT | 93878007 | Primary malignant neoplasm of lumbar vertebral column |
| SNOMED CT | 93884005 | Primary malignant neoplasm of male breast |
| SNOMED CT | 93914000 | Primary malignant neoplasm of myocardium |
| SNOMED CT | 93925009 | Primary malignant neoplasm of nipple of male breast |
| SNOMED CT | 93972009 | Primary malignant neoplasm of posterior wall of urinary bladder |
| SNOMED CT | 93984006 | Primary malignant neoplasm of rectum |
| SNOMED CT | 94004004 | Primary malignant neoplasm of short bone of upper limb |
| SNOMED CT | 94012007 | Primary malignant neoplasm of skin of breast |
| SNOMED CT | 94016005 | Primary malignant neoplasm of skin of chin |
| SNOMED CT | 94032008 | Primary malignant neoplasm of skin of lip |
| SNOMED CT | 94152006 | Secondary malignant neoplasm of abdominal esophagus |
| SNOMED CT | 94155008 | Secondary malignant neoplasm of accessory sinus |
| SNOMED CT | 94157000 | Secondary malignant neoplasm of acromion |
| SNOMED CT | 94158005 | Secondary malignant neoplasm of adenoid |
| SNOMED CT | 94160007 | Secondary malignant neoplasm of adrenal cortex |
| SNOMED CT | 94161006 | Secondary malignant neoplasm of adrenal gland |
| SNOMED CT | 94162004 | Secondary malignant neoplasm of adrenal medulla |
| SNOMED CT | 94163009 | Secondary malignant neoplasm of alveolar ridge mucosa |
| SNOMED CT | 94164003 | Secondary malignant neoplasm of ampulla of Vater |
| SNOMED CT | 94165002 | Secondary malignant neoplasm of anal canal |
| SNOMED CT | 94166001 | Secondary malignant neoplasm of anterior aspect of epiglottis |
| SNOMED CT | 94167005 | Secondary malignant neoplasm of anterior mediastinum |
| SNOMED CT | 94168000 | Secondary malignant neoplasm of anterior portion of floor of mouth |
| SNOMED CT | 94169008 | Secondary malignant neoplasm of anterior two-thirds of tongue |
| SNOMED CT | 94170009 | Secondary malignant neoplasm of anterior wall of nasopharynx |
| SNOMED CT | 94171008 | Secondary malignant neoplasm of anterior wall of urinary bladder |
| SNOMED CT | 94172001 | Secondary malignant neoplasm of anus |
| SNOMED CT | 94175004 | Secondary malignant neoplasm of appendix |
| SNOMED CT | 94176003 | Secondary malignant neoplasm of areola of female breast |
| SNOMED CT | 94177007 | Secondary malignant neoplasm of areola of male breast |
| SNOMED CT | 94180008 | Secondary malignant neoplasm of axilla |
| SNOMED CT | 94181007 | Secondary malignant neoplasm of axillary lymph nodes |

Continued on next page

**TableS24 – continued from previous page**

| CodeSystem | ConceptCode | ConceptName |
|---|---|---|
| SNOMED CT | 94182000 | Secondary malignant neoplasm of axillary tail of female breast |
| SNOMED CT | 94184004 | Secondary malignant neoplasm of base of tongue |
| SNOMED CT | 94186002 | Secondary malignant neoplasm of bladder |
| SNOMED CT | 94189009 | Secondary malignant neoplasm of blood vessel of axilla |
| SNOMED CT | 94207003 | Secondary malignant neoplasm of blood vessel of thorax |
| SNOMED CT | 94213007 | Secondary malignant neoplasm of body of penis |
| SNOMED CT | 94214001 | Secondary malignant neoplasm of body of stomach |
| SNOMED CT | 94215000 | Secondary malignant neoplasm of body of uterus |
| SNOMED CT | 94218003 | Secondary malignant neoplasm of bone of face |
| SNOMED CT | 94219006 | Secondary malignant neoplasm of bone of lower limb |
| SNOMED CT | 94220000 | Secondary malignant neoplasm of bone of skull |
| SNOMED CT | 94221001 | Secondary malignant neoplasm of bone of upper limb |
| SNOMED CT | 94222008 | Secondary malignant neoplasm of bone |
| SNOMED CT | 94224009 | Secondary malignant neoplasm of brain stem |
| SNOMED CT | 94225005 | Secondary malignant neoplasm of brain |
| SNOMED CT | 94226006 | Secondary malignant neoplasm of broad ligament |
| SNOMED CT | 94227002 | Secondary malignant neoplasm of bronchopulmonary lymph nodes |
| SNOMED CT | 94228007 | Secondary malignant neoplasm of bronchus of left lower lobe |
| SNOMED CT | 94229004 | Secondary malignant neoplasm of bronchus of left upper lobe |
| SNOMED CT | 94230009 | Secondary malignant neoplasm of bronchus of right lower lobe |
| SNOMED CT | 94231008 | Secondary malignant neoplasm of bronchus of right middle lobe |
| SNOMED CT | 94232001 | Secondary malignant neoplasm of bronchus of right upper lobe |
| SNOMED CT | 94233006 | Secondary malignant neoplasm of bronchus |
| SNOMED CT | 94234000 | Secondary malignant neoplasm of buccal mucosa |
| SNOMED CT | 94235004 | Secondary malignant neoplasm of cecum |
| SNOMED CT | 94236003 | Secondary malignant neoplasm of calcaneus |
| SNOMED CT | 94237007 | Secondary malignant neoplasm of cardia of stomach |
| SNOMED CT | 94238002 | Secondary malignant neoplasm of carina |
| SNOMED CT | 94240007 | Secondary malignant neoplasm of carpal bone |
| SNOMED CT | 94241006 | Secondary malignant neoplasm of cartilage of nose |
| SNOMED CT | 94243009 | Secondary malignant neoplasm of central nervous system |
| SNOMED CT | 94244003 | Secondary malignant neoplasm of central portion of female breast |
| SNOMED CT | 94245002 | Secondary malignant neoplasm of cerebellum |
| SNOMED CT | 94246001 | Secondary malignant neoplasm of cerebral meninges |
| SNOMED CT | 94247005 | Secondary malignant neoplasm of cerebral ventricle |
| SNOMED CT | 94248000 | Secondary malignant neoplasm of cerebrum |
| SNOMED CT | 94249008 | Secondary malignant neoplasm of cervical esophagus |
| SNOMED CT | 94250008 | Secondary malignant neoplasm of cervical vertebral column |
| SNOMED CT | 94253005 | Secondary malignant neoplasm of chest wall |
| SNOMED CT | 94254004 | Secondary malignant neoplasm of choroid |
| SNOMED CT | 94255003 | Secondary malignant neoplasm of ciliary body |
| SNOMED CT | 94256002 | Secondary malignant neoplasm of clavicle |
| SNOMED CT | 94257006 | Secondary malignant neoplasm of clitoris |
| SNOMED CT | 94259009 | Secondary malignant neoplasm of coccyx |
| SNOMED CT | 94261000 | Secondary malignant neoplasm of commissure of lip |

Continued on next page

**TableS24 – continued from previous page**

| CodeSystem | ConceptCode | ConceptName |
|---|---|---|
| SNOMED CT | 94263002 | Secondary malignant neoplasm of conjunctiva |
| SNOMED CT | 94265009 | Secondary malignant neoplasm of cornea |
| SNOMED CT | 94267001 | Secondary malignant neoplasm of craniopharyngeal duct |
| SNOMED CT | 94269003 | Secondary malignant neoplasm of cuboid |
| SNOMED CT | 94272005 | Secondary malignant neoplasm of diaphragm |
| SNOMED CT | 94273000 | Secondary malignant neoplasm of dorsal surface of tongue |
| SNOMED CT | 94274006 | Secondary malignant neoplasm of thoracic vertebral column |
| SNOMED CT | 94275007 | Secondary malignant neoplasm of duodenum |
| SNOMED CT | 94276008 | Secondary malignant neoplasm of ectopic female breast tissue |
| SNOMED CT | 94277004 | Secondary malignant neoplasm of ectopic male breast tissue |
| SNOMED CT | 94278009 | Secondary malignant neoplasm of endocardium |
| SNOMED CT | 94279001 | Secondary malignant neoplasm of endocervix |
| SNOMED CT | 94280003 | Secondary malignant neoplasm of endocrine gland |
| SNOMED CT | 94281004 | Secondary malignant neoplasm of endometrium |
| SNOMED CT | 94282006 | Secondary malignant neoplasm of epicardium |
| SNOMED CT | 94283001 | Secondary malignant neoplasm of epididymis |
| SNOMED CT | 94284007 | Secondary malignant neoplasm of epiglottis |
| SNOMED CT | 94286009 | Secondary malignant neoplasm of esophagus |
| SNOMED CT | 94287000 | Secondary malignant neoplasm of ethmoid bone |
| SNOMED CT | 94288005 | Secondary malignant neoplasm of ethmoidal sinus |
| SNOMED CT | 94289002 | Secondary malignant neoplasm of eustachian tube |
| SNOMED CT | 94290006 | Secondary malignant neoplasm of exocervix |
| SNOMED CT | 94292003 | Secondary malignant neoplasm of eye |
| SNOMED CT | 94295001 | Secondary malignant neoplasm of fallopian tube |
| SNOMED CT | 94296000 | Secondary malignant neoplasm of false vocal cord |
| SNOMED CT | 94297009 | Secondary malignant neoplasm of female breast |
| SNOMED CT | 94298004 | Secondary malignant neoplasm of female genital organ |
| SNOMED CT | 94300004 | Secondary malignant neoplasm of femur |
| SNOMED CT | 94301000 | Secondary malignant neoplasm of fibula |
| SNOMED CT | 94302007 | Secondary malignant neoplasm of first cuneiform bone of foot |
| SNOMED CT | 94304008 | Secondary malignant neoplasm of floor of mouth |
| SNOMED CT | 94307001 | Secondary malignant neoplasm of prepuce |
| SNOMED CT | 94308006 | Secondary malignant neoplasm of frontal bone |
| SNOMED CT | 94309003 | Secondary malignant neoplasm of frontal lobe |
| SNOMED CT | 94310008 | Secondary malignant neoplasm of frontal sinus |
| SNOMED CT | 94311007 | Secondary malignant neoplasm of fundus of stomach |
| SNOMED CT | 94314004 | Secondary malignant neoplasm of gingival mucosa |
| SNOMED CT | 94315003 | Secondary malignant neoplasm of glans penis |
| SNOMED CT | 94318001 | Secondary malignant neoplasm of glottis |
| SNOMED CT | 94320003 | Secondary malignant neoplasm of greater curvature of stomach |
| SNOMED CT | 94321004 | Secondary malignant neoplasm of gum |
| SNOMED CT | 94322006 | Secondary malignant neoplasm of hamate bone |
| SNOMED CT | 94324007 | Secondary malignant neoplasm of hard palate |
| SNOMED CT | 94327000 | Secondary malignant neoplasm of heart |
| SNOMED CT | 94329002 | Secondary malignant neoplasm of hilus of lung |
| SNOMED CT | 94332004 | Secondary malignant neoplasm of hypopharyngeal aspect of aryepiglottic fold |

Continued on next page

**TableS24 – continued from previous page**

| CodeSystem | ConceptCode | ConceptName |
|---|---|---|
| SNOMED CT | 94333009 | Secondary malignant neoplasm of hypopharyngeal aspect of interarytenoid fold |
| SNOMED CT | 94334003 | Secondary malignant neoplasm of hypopharynx |
| SNOMED CT | 94337005 | Secondary malignant neoplasm of ilium |
| SNOMED CT | 94338000 | Secondary malignant neoplasm of infraclavicular lymph nodes |
| SNOMED CT | 94341009 | Secondary malignant neoplasm of inner aspect of lip |
| SNOMED CT | 94342002 | Secondary malignant neoplasm of inner aspect of lower lip |
| SNOMED CT | 94343007 | Secondary malignant neoplasm of inner aspect of upper lip |
| SNOMED CT | 94344001 | Secondary malignant neoplasm of intercostal lymph nodes |
| SNOMED CT | 94351005 | Secondary malignant neoplasm of intrathoracic lymph nodes |
| SNOMED CT | 94352003 | Secondary malignant neoplasm of intrathoracic organs |
| SNOMED CT | 94353008 | Secondary malignant neoplasm of ischium |
| SNOMED CT | 94354002 | Secondary malignant neoplasm of islets of Langerhans |
| SNOMED CT | 94355001 | Secondary malignant neoplasm of isthmus of uterus |
| SNOMED CT | 94358004 | Secondary malignant neoplasm of junctional region of epiglottis |
| SNOMED CT | 94359007 | Secondary malignant neoplasm of junctional zone of tongue |
| SNOMED CT | 94360002 | Secondary malignant neoplasm of kidney |
| SNOMED CT | 94361003 | Secondary malignant neoplasm of labia majora |
| SNOMED CT | 94362005 | Secondary malignant neoplasm of labia minora |
| SNOMED CT | 94364006 | Secondary malignant neoplasm of lacrimal gland |
| SNOMED CT | 94366008 | Secondary malignant neoplasm of laryngeal aspect of aryepiglottic fold |
| SNOMED CT | 94367004 | Secondary malignant neoplasm of laryngeal aspect of interarytenoid fold |
| SNOMED CT | 94368009 | Secondary malignant neoplasm of laryngeal commissure |
| SNOMED CT | 94369001 | Secondary malignant neoplasm of laryngeal surface of epiglottis |
| SNOMED CT | 94370000 | Secondary malignant neoplasm of larynx |
| SNOMED CT | 94371001 | Secondary malignant neoplasm of lateral portion of floor of mouth |
| SNOMED CT | 94372008 | Secondary malignant neoplasm of lateral wall of nasopharynx |
| SNOMED CT | 94373003 | Secondary malignant neoplasm of lateral wall of oropharynx |
| SNOMED CT | 94374009 | Secondary malignant neoplasm of lateral wall of urinary bladder |
| SNOMED CT | 94375005 | Secondary malignant neoplasm of left lower lobe of lung |
| SNOMED CT | 94376006 | Secondary malignant neoplasm of left upper lobe of lung |
| SNOMED CT | 94378007 | Secondary malignant neoplasm of lesser curvature of stomach |
| SNOMED CT | 94379004 | Secondary malignant neoplasm of lingual tonsil |
| SNOMED CT | 94380001 | Secondary malignant neoplasm of lip |
| SNOMED CT | 94382009 | Secondary malignant neoplasm of long bone of lower limb |
| SNOMED CT | 94383004 | Secondary malignant neoplasm of long bone of upper limb |
| SNOMED CT | 94384005 | Secondary malignant neoplasm of lower gum |
| SNOMED CT | 94385006 | Secondary malignant neoplasm of lower inner quadrant of female breast |
| SNOMED CT | 94387003 | Secondary malignant neoplasm of lower outer quadrant of female breast |
| SNOMED CT | 94388008 | Secondary malignant neoplasm of lower third of esophagus |
| SNOMED CT | 94389000 | Secondary malignant neoplasm of lumbar vertebral column |
| SNOMED CT | 94390009 | Secondary malignant neoplasm of lunate bone |

Continued on next page

**TableS24 – continued from previous page**

| CodeSystem | ConceptCode | ConceptName |
|---|---|---|
| SNOMED CT | 94391008 | Secondary malignant neoplasm of lung |
| SNOMED CT | 94393006 | Secondary malignant neoplasm of lymph nodes of face |
| SNOMED CT | 94394000 | Secondary malignant neoplasm of lymph nodes of head |
| SNOMED CT | 94397007 | Secondary malignant neoplasm of lymph nodes of neck |
| SNOMED CT | 94399005 | Secondary malignant neoplasm of main bronchus |
| SNOMED CT | 94400003 | Secondary malignant neoplasm of major salivary gland |
| SNOMED CT | 94401004 | Secondary malignant neoplasm of male breast |
| SNOMED CT | 94402006 | Secondary malignant neoplasm of male genital organ |
| SNOMED CT | 94403001 | Secondary malignant neoplasm of mandible |
| SNOMED CT | 94404007 | Secondary malignant neoplasm of mastoid air cells |
| SNOMED CT | 94405008 | Secondary malignant neoplasm of maxilla |
| SNOMED CT | 94406009 | Secondary malignant neoplasm of maxillary sinus |
| SNOMED CT | 94408005 | Secondary malignant neoplasm of mediastinal lymph nodes |
| SNOMED CT | 94409002 | Secondary malignant neoplasm of mediastinum |
| SNOMED CT | 94411006 | Secondary malignant neoplasm of metacarpal bone |
| SNOMED CT | 94412004 | Secondary malignant neoplasm of metatarsal bone |
| SNOMED CT | 94413009 | Secondary malignant neoplasm of middle ear |
| SNOMED CT | 94414003 | Secondary malignant neoplasm of middle third of esophagus |
| SNOMED CT | 94415002 | Secondary malignant neoplasm of minor salivary gland |
| SNOMED CT | 94417005 | Secondary malignant neoplasm of multiple endocrine glands |
| SNOMED CT | 94429001 | Secondary malignant neoplasm of muscle of thorax |
| SNOMED CT | 94433008 | Secondary malignant neoplasm of myocardium |
| SNOMED CT | 94434002 | Secondary malignant neoplasm of myometrium |
| SNOMED CT | 94435001 | Secondary malignant neoplasm of nasal bone |
| SNOMED CT | 94436000 | Secondary malignant neoplasm of nasal cavity |
| SNOMED CT | 94437009 | Secondary malignant neoplasm of nasal concha |
| SNOMED CT | 94439007 | Secondary malignant neoplasm of navicular bone of foot |
| SNOMED CT | 94440009 | Secondary malignant neoplasm of scaphoid bone |
| SNOMED CT | 94443006 | Secondary malignant neoplasm of nipple of female breast |
| SNOMED CT | 94444000 | Secondary malignant neoplasm of nipple of male breast |
| SNOMED CT | 94447007 | Secondary malignant neoplasm of occipital bone |
| SNOMED CT | 94448002 | Secondary malignant neoplasm of occipital lobe |
| SNOMED CT | 94449005 | Secondary malignant neoplasm of occipital lymph nodes |
| SNOMED CT | 94452002 | Secondary malignant neoplasm of optic nerve |
| SNOMED CT | 94453007 | Secondary malignant neoplasm of orbit |
| SNOMED CT | 94454001 | Secondary malignant neoplasm of oropharynx |
| SNOMED CT | 94455000 | Secondary malignant neoplasm of ovary |
| SNOMED CT | 94456004 | Secondary malignant neoplasm of palate |
| SNOMED CT | 94457008 | Secondary malignant neoplasm of palatine bone |
| SNOMED CT | 94458003 | Secondary malignant neoplasm of tonsil |
| SNOMED CT | 94463004 | Secondary malignant neoplasm of paramammary lymph nodes |
| SNOMED CT | 94464005 | Secondary malignant neoplasm of parametrial lymph nodes |
| SNOMED CT | 94465006 | Secondary malignant neoplasm of parametrium |
| SNOMED CT | 94467003 | Secondary malignant neoplasm of parathyroid gland |
| SNOMED CT | 94468008 | Secondary malignant neoplasm of paraurethral glands |
| SNOMED CT | 94470004 | Secondary malignant neoplasm of parietal bone |
| SNOMED CT | 94471000 | Secondary malignant neoplasm of parietal lobe |

Continued on next page

**TableS24 – continued from previous page**

| CodeSystem | ConceptCode | ConceptName |
|---|---|---|
| SNOMED CT | 94472007 | Secondary malignant neoplasm of parietal peritoneum |
| SNOMED CT | 94473002 | Secondary malignant neoplasm of parietal pleura |
| SNOMED CT | 94474008 | Secondary malignant neoplasm of parotid gland |
| SNOMED CT | 94475009 | Secondary malignant neoplasm of parotid lymph nodes |
| SNOMED CT | 94476005 | Secondary malignant neoplasm of patella |
| SNOMED CT | 94477001 | Secondary malignant neoplasm of pectoral axillary lymph nodes |
| SNOMED CT | 94478006 | Secondary malignant neoplasm of pelvic bone |
| SNOMED CT | 94479003 | Secondary malignant neoplasm of pelvic peritoneum |
| SNOMED CT | 94481001 | Secondary malignant neoplasm of penis |
| SNOMED CT | 94484009 | Secondary malignant neoplasm of pericardium |
| SNOMED CT | 94485005 | Secondary malignant neoplasm of perirenal tissue |
| SNOMED CT | 94486006 | Secondary malignant neoplasm of phalanx of foot |
| SNOMED CT | 94487002 | Secondary malignant neoplasm of phalanx of hand |
| SNOMED CT | 94488007 | Secondary malignant neoplasm of pharynx |
| SNOMED CT | 94489004 | Secondary malignant neoplasm of pineal gland |
| SNOMED CT | 94490008 | Secondary malignant neoplasm of pisiform bone of hand |
| SNOMED CT | 94491007 | Secondary malignant neoplasm of pituitary gland |
| SNOMED CT | 94493005 | Secondary malignant neoplasm of pleura |
| SNOMED CT | 94495003 | Secondary malignant neoplasm of postcricoid region |
| SNOMED CT | 94496002 | Secondary malignant neoplasm of posterior hypopharyngeal wall |
| SNOMED CT | 94497006 | Secondary malignant neoplasm of posterior mediastinum |
| SNOMED CT | 94498001 | Secondary malignant neoplasm of posterior wall of nasopharynx |
| SNOMED CT | 94499009 | Secondary malignant neoplasm of posterior wall of oropharynx |
| SNOMED CT | 94500000 | Secondary malignant neoplasm of posterior wall of urinary bladder |
| SNOMED CT | 94501001 | Secondary malignant neoplasm of preauricular lymph nodes |
| SNOMED CT | 94503003 | Secondary malignant neoplasm of prostate |
| SNOMED CT | 94504009 | Secondary malignant neoplasm of pubis |
| SNOMED CT | 94505005 | Secondary malignant neoplasm of pyloric antrum |
| SNOMED CT | 94506006 | Secondary malignant neoplasm of pylorus |
| SNOMED CT | 94507002 | Secondary malignant neoplasm of pyriform sinus |
| SNOMED CT | 94508007 | Secondary malignant neoplasm of radius |
| SNOMED CT | 94510009 | Secondary malignant neoplasm of rectouterine pouch |
| SNOMED CT | 94511008 | Secondary malignant neoplasm of rectovaginal septum |
| SNOMED CT | 94512001 | Secondary malignant neoplasm of rectovesical septum |
| SNOMED CT | 94513006 | Secondary malignant neoplasm of rectum |
| SNOMED CT | 94514000 | Secondary malignant neoplasm of renal pelvis |
| SNOMED CT | 94516003 | Secondary malignant neoplasm of retina |
| SNOMED CT | 94518002 | Secondary malignant neoplasm of retromolar area |
| SNOMED CT | 94520004 | Secondary malignant neoplasm of retropharyngeal lymph nodes |
| SNOMED CT | 94521000 | Secondary malignant neoplasm of rib |
| SNOMED CT | 94522007 | Secondary malignant neoplasm of right lower lobe of lung |
| SNOMED CT | 94523002 | Secondary malignant neoplasm of right middle lobe of lung |
| SNOMED CT | 94524008 | Secondary malignant neoplasm of right upper lobe of lung |
| SNOMED CT | 94525009 | Secondary malignant neoplasm of round ligament of uterus |
| SNOMED CT | 94527001 | Secondary malignant neoplasm of sacrum |
| SNOMED CT | 94528006 | Secondary malignant neoplasm of scalene lymph nodes |

Continued on next page

**TableS24 – continued from previous page**

| CodeSystem | ConceptCode | ConceptName |
|---|---|---|
| SNOMED CT | 94529003 | Secondary malignant neoplasm of scapula |
| SNOMED CT | 94530008 | Secondary malignant neoplasm of sclera |
| SNOMED CT | 94531007 | Secondary malignant neoplasm of scrotum |
| SNOMED CT | 94533005 | Secondary malignant neoplasm of second cuneiform bone of foot |
| SNOMED CT | 94534004 | Secondary malignant neoplasm of septum of nose |
| SNOMED CT | 94535003 | Secondary malignant neoplasm of short bone of lower limb |
| SNOMED CT | 94536002 | Secondary malignant neoplasm of short bone of upper limb |
| SNOMED CT | 94542003 | Secondary malignant neoplasm of skin of axilla |
| SNOMED CT | 94544002 | Secondary malignant neoplasm of skin of breast |
| SNOMED CT | 94546000 | Secondary malignant neoplasm of skin of cheek |
| SNOMED CT | 94547009 | Secondary malignant neoplasm of skin of chest |
| SNOMED CT | 94553009 | Secondary malignant neoplasm of skin of eyelid |
| SNOMED CT | 94564007 | Secondary malignant neoplasm of skin of lip |
| SNOMED CT | 94581003 | Secondary malignant neoplasm of soft palate |
| SNOMED CT | 94583000 | Secondary malignant neoplasm of soft tissues of axilla |
| SNOMED CT | 94594001 | Secondary malignant neoplasm of soft tissues of thorax |
| SNOMED CT | 94597008 | Secondary malignant neoplasm of spermatic cord |
| SNOMED CT | 94598003 | Secondary malignant neoplasm of sphenoid bone |
| SNOMED CT | 94599006 | Secondary malignant neoplasm of sphenoidal sinus |
| SNOMED CT | 94600009 | Secondary malignant neoplasm of spinal cord |
| SNOMED CT | 94601008 | Secondary malignant neoplasm of spinal meninges |
| SNOMED CT | 94602001 | Secondary malignant neoplasm of vertebral column |
| SNOMED CT | 94605004 | Secondary malignant neoplasm of sternum |
| SNOMED CT | 94606003 | Secondary malignant neoplasm of stomach |
| SNOMED CT | 94607007 | Secondary malignant neoplasm of subglottis |
| SNOMED CT | 94608002 | Secondary malignant neoplasm of sublingual gland |
| SNOMED CT | 94609005 | Secondary malignant neoplasm of submandibular lymph nodes |
| SNOMED CT | 94611001 | Secondary malignant neoplasm of submental lymph nodes |
| SNOMED CT | 94613003 | Secondary malignant neoplasm of superior wall of nasopharynx |
| SNOMED CT | 94614009 | Secondary malignant neoplasm of supraclavicular lymph nodes |
| SNOMED CT | 94616006 | Secondary malignant neoplasm of supraglottis |
| SNOMED CT | 94619004 | Secondary malignant neoplasm of talus |
| SNOMED CT | 94620005 | Secondary malignant neoplasm of tarsal bone |
| SNOMED CT | 94621009 | Secondary malignant neoplasm of temporal bone |
| SNOMED CT | 94622002 | Secondary malignant neoplasm of temporal lobe |
| SNOMED CT | 94623007 | Secondary malignant neoplasm of testis |
| SNOMED CT | 94624001 | Secondary malignant neoplasm of the mesentery |
| SNOMED CT | 94625000 | Secondary malignant neoplasm of the mesocolon |
| SNOMED CT | 94626004 | Secondary malignant neoplasm of omentum |
| SNOMED CT | 94627008 | Secondary malignant neoplasm of peritoneum |
| SNOMED CT | 94630001 | Secondary malignant neoplasm of third cuneiform bone of foot |
| SNOMED CT | 94631002 | Secondary malignant neoplasm of thoracic esophagus |
| SNOMED CT | 94632009 | Secondary malignant neoplasm of thymus |
| SNOMED CT | 94634005 | Secondary malignant neoplasm of thyroid gland |
| SNOMED CT | 94635006 | Secondary malignant neoplasm of tibia |
| SNOMED CT | 94637003 | Secondary malignant neoplasm of tip and lateral border of tongue |

Continued on next page

**TableS24 – continued from previous page**

| CodeSystem | ConceptCode | ConceptName |
|---|---|---|
| SNOMED CT | 94638008 | Secondary malignant neoplasm of tongue |
| SNOMED CT | 94639000 | Secondary malignant neoplasm of tonsillar fossa |
| SNOMED CT | 94640003 | Secondary malignant neoplasm of tonsillar pillar |
| SNOMED CT | 94641004 | Secondary malignant neoplasm of trachea |
| SNOMED CT | 94642006 | Secondary malignant neoplasm of tracheobronchial lymph nodes |
| SNOMED CT | 94644007 | Secondary malignant neoplasm of trapezium |
| SNOMED CT | 94645008 | Secondary malignant neoplasm of trapezoid bone |
| SNOMED CT | 94647000 | Secondary malignant neoplasm of trigone of urinary bladder |
| SNOMED CT | 94650002 | Secondary malignant neoplasm of ulna |
| SNOMED CT | 94651003 | Secondary malignant neoplasm of undescended testis |
| SNOMED CT | 94652005 | Secondary malignant neoplasm of upper gum |
| SNOMED CT | 94653000 | Secondary malignant neoplasm of upper inner quadrant of female breast |
| SNOMED CT | 94655007 | Secondary malignant neoplasm of upper outer quadrant of female breast |
| SNOMED CT | 94656008 | Secondary malignant neoplasm of upper respiratory tract |
| SNOMED CT | 94657004 | Secondary malignant neoplasm of upper third of esophagus |
| SNOMED CT | 94658009 | Secondary malignant neoplasm of urachus |
| SNOMED CT | 94659001 | Secondary malignant neoplasm of ureter |
| SNOMED CT | 94660006 | Secondary malignant neoplasm of ureteric orifice of urinary bladder |
| SNOMED CT | 94661005 | Secondary malignant neoplasm of urethra |
| SNOMED CT | 94662003 | Secondary malignant neoplasm of urinary bladder neck |
| SNOMED CT | 94663008 | Secondary malignant neoplasm of urinary system |
| SNOMED CT | 94665001 | Secondary malignant neoplasm of uterus |
| SNOMED CT | 94666000 | Secondary malignant neoplasm of uveal tract |
| SNOMED CT | 94667009 | Secondary malignant neoplasm of uvula |
| SNOMED CT | 94668004 | Secondary malignant neoplasm of vagina |
| SNOMED CT | 94670008 | Secondary malignant neoplasm of vallecula |
| SNOMED CT | 94671007 | Secondary malignant neoplasm of vas deferens |
| SNOMED CT | 94672000 | Secondary malignant neoplasm of ventral surface of tongue |
| SNOMED CT | 94673005 | Secondary malignant neoplasm of vermilion border of lip |
| SNOMED CT | 94674004 | Secondary malignant neoplasm of vermilion border of lower lip |
| SNOMED CT | 94675003 | Secondary malignant neoplasm of vermilion border of upper lip |
| SNOMED CT | 94676002 | Secondary malignant neoplasm of vestibule of mouth |
| SNOMED CT | 94677006 | Secondary malignant neoplasm of vestibule of nose |
| SNOMED CT | 94678001 | Secondary malignant neoplasm of visceral pleura |
| SNOMED CT | 94679009 | Secondary malignant neoplasm of vocal cord |
| SNOMED CT | 94680007 | Secondary malignant neoplasm of vomer |
| SNOMED CT | 94683009 | Secondary malignant neoplasm of zygomatic bone |
| SNOMED CT | 94704006 | Multiple myeloma in remission |
| SNOMED CT | 94712003 | Mycosis fungoides of lymph nodes of inguinal region AND/OR lower limb |
| SNOMED CT | 94758002 | Neoplasm of uncertain behavior of body of stomach |
| SNOMED CT | 94777000 | Neoplasm of uncertain behavior of cardia of stomach |
| SNOMED CT | 94808000 | Neoplasm of uncertain behavior of craniopharyngeal duct |
| SNOMED CT | 94849000 | Neoplasm of uncertain behavior of fundus of stomach |

Continued on next page

**TableS24 – continued from previous page**

| CodeSystem | ConceptCode | ConceptName |
|---|---|---|
| SNOMED CT | 94858007 | Neoplasm of uncertain behavior of greater curvature of stomach |
| SNOMED CT | 94893000 | Neoplasm of uncertain behavior of lacrimal gland |
| SNOMED CT | 94907009 | Neoplasm of uncertain behavior of lesser curvature of stomach |
| SNOMED CT | 95004009 | Neoplasm of uncertain behavior of pituitary gland |
| SNOMED CT | 95016003 | Neoplasm of uncertain behavior of pyloric antrum |
| SNOMED CT | 95017007 | Neoplasm of uncertain behavior of pylorus |
| SNOMED CT | 95261008 | Sézary's disease of lymph nodes of inguinal region AND/OR lower limb |
| SNOMED CT | 109347009 | Overlapping malignant neoplasm of bone and articular cartilage |
| SNOMED CT | 109833003 | Overlapping malignant neoplasm of lip, oral cavity and pharynx |
| SNOMED CT | 109885001 | Overlapping malignant neoplasm of vulva |
| SNOMED CT | 109887009 | Overlapping malignant neoplasm of male breast |
| SNOMED CT | 109982002 | Alpha heavy chain disease (clinical) |
| SNOMED CT | 109984001 | Gamma heavy chain disease (clinical) |
| SNOMED CT | 109985000 | Immunoproliferative small intestinal disease (clinical) |
| SNOMED CT | 109989006 | Multiple myeloma |
| SNOMED CT | 109991003 | Acute panmyelosis with myelofibrosis |
| SNOMED CT | 123313007 | Alpha heavy chain disease, enteric form |
| SNOMED CT | 123845008 | Adenocarcinoma of endometrium |
| SNOMED CT | 127004000 | Neoplasm of lacrimal gland |
| SNOMED CT | 127024001 | Neoplasm of pituitary gland |
| SNOMED CT | 127025000 | Neoplasm of craniopharyngeal duct |
| SNOMED CT | 127218004 | Reactive follicular hyperplasia in the elderly |
| SNOMED CT | 134209002 | Prolactinoma |
| SNOMED CT | 134421000 | Pathologic fracture of bone at site of metastatic neoplasm |
| SNOMED CT | 187633009 | Malignant neoplasm of dorsal surface of tongue |
| SNOMED CT | 187760008 | Malignant neoplasm of rectum, rectosigmoid junction and anus |
| SNOMED CT | 187828007 | Malignant neoplasm of nasal cavities, middle ear and accessory sinuses |
| SNOMED CT | 187864008 | Malignant neoplasm of middle lobe, bronchus or lung |
| SNOMED CT | 187881004 | Malignant neoplasm of thymus, heart and mediastinum |
| SNOMED CT | 187989003 | Malignant neoplasm soft tissues of cervical spine |
| SNOMED CT | 188009001 | Malignant neoplasm of connective and soft tissue of thorax |
| SNOMED CT | 188042000 | Malignant melanoma of temple |
| SNOMED CT | 188050009 | Malignant melanoma of breast |
| SNOMED CT | 188051008 | Malignant melanoma of buttock |
| SNOMED CT | 188068007 | Malignant melanoma of hip |
| SNOMED CT | 188069004 | Malignant melanoma of thigh |
| SNOMED CT | 188070003 | Malignant melanoma of knee |
| SNOMED CT | 188073001 | Malignant melanoma of ankle |
| SNOMED CT | 188074007 | Malignant melanoma of heel |
| SNOMED CT | 188075008 | Malignant melanoma of foot |
| SNOMED CT | 188076009 | Malignant melanoma of toe |
| SNOMED CT | 188077000 | Malignant melanoma of great toe |
| SNOMED CT | 188157005 | Malignant neoplasm of overlapping sites of breast |
| SNOMED CT | 188163001 | Malignant neoplasm of nipple and areola of male breast |
| SNOMED CT | 188168005 | Malignant neoplasm of ectopic site of male breast |

Continued on next page

**TableS24 – continued from previous page**

| CodeSystem | ConceptCode | ConceptName |
|---|---|---|
| SNOMED CT | 188445006 | Secondary malignant neoplasm of retroperitoneum and peritoneum |
| SNOMED CT | 188462001 | Secondary malignant neoplasm of brain and spinal cord |
| SNOMED CT | 188469005 | Secondary malignant neoplasm of cervix uteri |
| SNOMED CT | 188471005 | Secondary malignant neoplasm of epididymis and vas deferens |
| SNOMED CT | 189125000 | Skin tag in vagina |
| SNOMED CT | 189155007 | Benign neoplasm lacrimal sac |
| SNOMED CT | 189156008 | Benign neoplasm nasolacrimal duct |
| SNOMED CT | 189179009 | Craniopharyngioma |
| SNOMED CT | 189815007 | Pulmonary blastoma |
| SNOMED CT | 190818004 | Waldenström macroglobulinemia |
| SNOMED CT | 770559003 | Leiomyosarcoma of corpus uteri |
| SNOMED CT | 231828003 | Capillary hemangioma of eyelid |
| SNOMED CT | 232457008 | Laryngeal papillomatosis |
| SNOMED CT | 233940007 | Pulmonary tumor embolism |
| SNOMED CT | 237721006 | Tumor of pituitary and suprasellar region |
| SNOMED CT | 242862004 | Secondary malignant neoplasm of nasopharyngeal wall |
| SNOMED CT | 253010003 | Microprolactinoma |
| SNOMED CT | 253011004 | Macroprolactinoma |
| SNOMED CT | 254436005 | Carcinoma of uvula |
| SNOMED CT | 254504001 | Benign tumor of inferior surface of soft palate |
| SNOMED CT | 254582000 | Adenocarcinoma of rectum |
| SNOMED CT | 254837009 | Malignant tumor of breast |
| SNOMED CT | 254838004 | Carcinoma of breast |
| SNOMED CT | 254839007 | Scirrhous carcinoma of breast |
| SNOMED CT | 254840009 | Inflammatory carcinoma of breast |
| SNOMED CT | 254841008 | Cancer en cuirasse |
| SNOMED CT | 254843006 | Familial cancer of breast |
| SNOMED CT | 254844000 | Malignant phyllodes tumor of breast |
| SNOMED CT | 254939008 | Ependymoma of brain |
| SNOMED CT | 254949006 | Ependymoma of spinal cord |
| SNOMED CT | 254992001 | Pleomorphic adenoma of lacrimal gland |
| SNOMED CT | 255068000 | Carcinoma of bone, connective tissue, skin and breast |
| SNOMED CT | 255081007 | Carcinoma of cecum |
| SNOMED CT | 255101006 | Sézary disease of skin |
| SNOMED CT | 255102004 | Angioendotheliomatosis |
| SNOMED CT | 255107005 | Seminoma of testis |
| SNOMED CT | 255200003 | Benign tumor of hypothalamus |
| SNOMED CT | 1229988003 | Myeloid and/or lymphoid neoplasm with PCM1-JAK2 |
| SNOMED CT | 20224008 | Delta heavy chain disease |
| SNOMED CT | 25050002 | Alpha heavy chain disease, respiratory form |
| SNOMED CT | 39795003 | Hand-Schüller-Christian disease |
| SNOMED CT | 60620005 | Epsilon heavy chain disease |
| SNOMED CT | 61493004 | Mu heavy chain disease |
| SNOMED CT | 1217301006 | Precursor cell lymphoblastic lymphoma |
| SNOMED CT | 722103009 | Hormone sensitive prostate cancer |
| SNOMED CT | 722425009 | Reactive oxygen species 1 positive non-small cell lung cancer |

Continued on next page

**TableS24 – continued from previous page**

| CodeSystem | ConceptCode | ConceptName |
|---|---|---|
| SNOMED CT | 722445001 | Primary giant cell sarcoma of peritoneum |
| SNOMED CT | 722509001 | Primary rhabdomyosarcoma of oral cavity |
| SNOMED CT | 722510006 | Primary rhabdomyosarcoma of pharynx |
| SNOMED CT | 722512003 | Primary rhabdomyosarcoma of intrathoracic organ |
| SNOMED CT | 722514002 | Primary leiomyosarcoma of peritoneum |
| SNOMED CT | 722516000 | Primary liposarcoma of peritoneum |
| SNOMED CT | 722519007 | Primary sarcoma of peritoneum |
| SNOMED CT | 722524005 | Primary invasive pleomorphic lobular carcinoma of breast |
| SNOMED CT | 722527003 | Primary malignant neuroendocrine neoplasm of bronchus |
| SNOMED CT | 722528008 | Primary malignant neuroendocrine neoplasm of lung |
| SNOMED CT | 722529000 | Primary malignant epithelial neoplasm of nasopharynx |
| SNOMED CT | 722530005 | Primary squamous cell carcinoma of pharyngeal tonsil |
| SNOMED CT | 722533007 | Primary malignant neoplasm of esophagogastric junction |
| SNOMED CT | 722542000 | Primary squamous cell carcinoma of anal canal |
| SNOMED CT | 722543005 | Primary malignant melanoma of anal canal |
| SNOMED CT | 722665003 | Primary malignant melanoma of cornea |
| SNOMED CT | 722666002 | Primary squamous cell carcinoma of lacrimal apparatus |
| SNOMED CT | 722670005 | Primary thymic carcinoma |
| SNOMED CT | 722671009 | Metastatic malignant neoplasm of meninges |
| SNOMED CT | 722672002 | Primary squamous cell carcinoma of base of tongue |
| SNOMED CT | 722673007 | Primary squamous cell carcinoma of lingual tonsil |
| SNOMED CT | 722674001 | Primary squamous cell carcinoma of parotid gland |
| SNOMED CT | 722676004 | Primary squamous cell carcinoma of middle ear |
| SNOMED CT | 722677008 | Primary mesothelioma of overlapping sites of retroperitoneum, peritoneum and omentum |
| SNOMED CT | 722678003 | Primary squamous cell carcinoma of vagina |
| SNOMED CT | 722679006 | Primary mucinous adenocarcinoma of endometrium |
| SNOMED CT | 722680009 | Primary serous adenocarcinoma of endometrium |
| SNOMED CT | 722681008 | Primary mixed adenocarcinoma of endometrium |
| SNOMED CT | 722682001 | Primary small cell carcinoma of endometrium |
| SNOMED CT | 722683006 | Primary neuroendocrine carcinoma of cervix uteri |
| SNOMED CT | 722684000 | Primary low grade serous adenocarcinoma of ovary |
| SNOMED CT | 722685004 | Primary high grade serous adenocarcinoma of ovary |
| SNOMED CT | 722714004 | Primary mucoepidermoid carcinoma of lacrimal apparatus |
| SNOMED CT | 722718001 | Primary malignant meningioma |
| SNOMED CT | 722828003 | Primary synovial sarcoma of intrathoracic organ |
| SNOMED CT | 722832009 | Primary solid papillary carcinoma with invasion of breast |
| SNOMED CT | 723077004 | Primary myosarcoma of uterus |
| SNOMED CT | 723182009 | Primary squamous cell carcinoma of nasal cavity |
| SNOMED CT | 723265000 | Primary squamous cell carcinoma of anus |
| SNOMED CT | 723281005 | Primary malignant melanoma of anus |
| SNOMED CT | 723301009 | Squamous non-small cell lung cancer |
| SNOMED CT | 723843005 | Primary chondrosarcoma of bone of limb |
| SNOMED CT | 723846002 | Primary chondrosarcoma of bone of rib |
| SNOMED CT | 723847006 | Primary chondrosarcoma of articular cartilage of rib |
| SNOMED CT | 723848001 | Primary osteosarcoma of bone of jaw |
| SNOMED CT | 723849009 | Primary osteosarcoma of articular cartilage of jaw |

Continued on next page

**TableS24 – continued from previous page**

| CodeSystem | ConceptCode | ConceptName |
|---|---|---|
| SNOMED CT | 723850009 | Primary osteosarcoma of bone of limb |
| SNOMED CT | 723853006 | Primary Ewing sarcoma of bone of limb |
| SNOMED CT | 723856003 | Primary Ewing sarcoma of articular cartilage of rib |
| SNOMED CT | 723879002 | Primary adenocarcinoma of overlapping sites of urinary organ |
| SNOMED CT | 723887001 | Primary Ewing sarcoma of bone of rib |
| SNOMED CT | 724056005 | Malignant neoplasm of lower lobe of right lung |
| SNOMED CT | 724058006 | Malignant neoplasm of upper lobe of left lung |
| SNOMED CT | 724059003 | Malignant neoplasm of lower lobe of left lung |
| SNOMED CT | 724060008 | Malignant neoplasm of right upper lobe of lung |
| SNOMED CT | 724467001 | Primary squamous cell carcinoma of overlapping sites of urinary organs |
| SNOMED CT | 724468006 | Primary urothelial carcinoma of overlapping sites of urinary organs |
| SNOMED CT | 724552002 | Primary poorly differentiated carcinoma of thyroid gland |
| SNOMED CT | 724553007 | Primary undifferentiated carcinoma of thyroid gland |
| SNOMED CT | 724554001 | Primary malignant epithelial neoplasm of endocrine gland |
| SNOMED CT | 726019003 | Familial malignant melanoma of skin |
| SNOMED CT | 726652005 | Malignant carcinoid tumor of thymus |
| SNOMED CT | 726653000 | Malignant carcinoid tumor of bronchus |
| SNOMED CT | 726654006 | Malignant carcinoid tumor of colon |
| SNOMED CT | 732201008 | Endometrial carcinosarcoma |
| SNOMED CT | 733064004 | Osteosarcoma, limb anomalies, erythroid macrocytosis syndrome |
| SNOMED CT | 733134009 | Primary squamous cell carcinoma of paraurethral gland |
| SNOMED CT | 733135005 | Primary urothelial carcinoma of paraurethral gland |
| SNOMED CT | 733136006 | Primary adenocarcinoma of urethra |
| SNOMED CT | 733144006 | Malignant epithelial neoplasm of bronchus |
| SNOMED CT | 733162002 | Primary malignant neuroendocrine neoplasm of anus |
| SNOMED CT | 733163007 | Primary malignant neuroendocrine neoplasm of anal canal |
| SNOMED CT | 733343005 | Primary squamous cell carcinoma of oral cavity |
| SNOMED CT | 733344004 | Primary squamous cell carcinoma of lip |
| SNOMED CT | 733345003 | Primary squamous cell carcinoma of pharynx |
| SNOMED CT | 733351008 | Primary malignant neuroendocrine neoplasm of ampulla of Vater |
| SNOMED CT | 733357007 | Primary squamous cell carcinoma of intrathoracic organ |
| SNOMED CT | 733359005 | Primary squamous cell carcinoma of endometrium |
| SNOMED CT | 733360000 | Primary undifferentiated carcinoma of endometrium |
| SNOMED CT | 733361001 | Primary mucinous adenocarcinoma of ovary |
| SNOMED CT | 733362008 | Primary adenocarcinoma of paraurethral gland |
| SNOMED CT | 733470002 | Collecting duct carcinoma of kidney |
| SNOMED CT | 733471003 | Chromophobe renal cell carcinoma |
| SNOMED CT | 733491005 | Carney complex |
| SNOMED CT | 733603009 | Tubulocystic renal cell carcinoma |
| SNOMED CT | 733608000 | Papillary renal cell carcinoma |
| SNOMED CT | 733834006 | Invasive carcinoma of uterine cervix co-occurrent with human immunodeficiency virus infection |
| SNOMED CT | 733926004 | Ganglioneuroblastoma of central nervous system |
| SNOMED CT | 734015000 | Clear cell papillary renal cell carcinoma |
| SNOMED CT | 734099007 | Neuroblastoma of central nervous system |

Continued on next page

**TableS24 – continued from previous page**

| CodeSystem | ConceptCode | ConceptName |
|---|---|---|
| SNOMED CT | 735082004 | Fordyce angiokeratoma of scrotum |
| SNOMED CT | 735450006 | Primary malignant neuroepitheliomatous neoplasm of nasal cavity |
| SNOMED CT | 735679005 | Primary chondrosarcoma of bone |
| SNOMED CT | 735757008 | Primary ganglioneuroblastoma of brain |
| SNOMED CT | 735916009 | Primary malignant neuroepithelial neoplasm of retina |
| SNOMED CT | 735918005 | Primary malignant neoplasm of lacrimal caruncle |
| SNOMED CT | 735919002 | Primary malignant neuroepithelial neoplasm of iris |
| SNOMED CT | 735920008 | Primary malignant neuroepithelial neoplasm of ciliary body |
| SNOMED CT | 735921007 | Primary malignant neuroepithelial neoplasm of orbit |
| SNOMED CT | 737267003 | Hemolymphangioma of conjunctiva |
| SNOMED CT | 737308008 | Primary adenocarcinoma of sublingual gland |
| SNOMED CT | 737309000 | Primary adenocarcinoma of submandibular gland |
| SNOMED CT | 737310005 | Primary squamous cell carcinoma of submandibular gland |
| SNOMED CT | 737311009 | Primary squamous cell carcinoma of sublingual gland |
| SNOMED CT | 738770003 | Anaplastic lymphoma kinase positive anaplastic large cell lymphoma |
| SNOMED CT | 739301006 | Osteoporosis co-occurrent and due to multiple myeloma |
| SNOMED CT | 762457009 | Astroblastoma of brain |
| SNOMED CT | 762458004 | Primary endometrioid carcinoma of endometrium of body of uterus |
| SNOMED CT | 763063001 | Adenoid basal carcinoma of cervix uteri |
| SNOMED CT | 763064007 | Adenoid cystic carcinoma of cervix uteri |
| SNOMED CT | 763131005 | Clear cell adenocarcinoma of ovary |
| SNOMED CT | 763408003 | Rhabdomyosarcoma of cervix uteri |
| SNOMED CT | 763409006 | Rhabdomyosarcoma of corpus uteri |
| SNOMED CT | 763479005 | Metaplastic carcinoma of breast |
| SNOMED CT | 763771009 | Leiomyosarcoma of cervix uteri |
| SNOMED CT | 764694005 | MiT family translocation renal cell carcinoma |
| SNOMED CT | 764737005 | Squamous cell carcinoma of corpus uteri |
| SNOMED CT | 764845008 | Adenocarcinoma of anal canal |
| SNOMED CT | 764846009 | Adenocarcinoma of penis |
| SNOMED CT | 764847000 | Adenosarcoma of cervix uteri |
| SNOMED CT | 764856008 | Acquired cystic disease associated renal cell carcinoma |
| SNOMED CT | 764951002 | Carcinosarcoma of cervix uteri |
| SNOMED CT | 764952009 | Carcinosarcoma of corpus uteri |
| SNOMED CT | 764961009 | Hereditary clear cell renal cell carcinoma |
| SNOMED CT | 764990003 | Mucinous tubular and spindle cell renal carcinoma |
| SNOMED CT | 765095002 | Renal medullary carcinoma |
| SNOMED CT | 765190005 | Fibrothecoma of ovary |
| SNOMED CT | 765740002 | Adenosarcoma of corpus uteri |
| SNOMED CT | 766247009 | Primitive neuroectodermal tumor of corpus uteri |
| SNOMED CT | 766248004 | Primitive neuroectodermal tumor of cervix uteri |
| SNOMED CT | 766757006 | Undifferentiated carcinoma of stomach |
| SNOMED CT | 766758001 | Undifferentiated carcinoma of corpus uteri |
| SNOMED CT | 766759009 | Vulvovaginal rhabdomyosarcoma |
| SNOMED CT | 766881008 | Carney complex, trismus, pseudocamptodactyly syndrome |

Continued on next page

**TableS24 – continued from previous page**

| CodeSystem | ConceptCode | ConceptName |
|---|---|---|
| SNOMED CT | 766930002 | Glassy cell carcinoma of cervix uteri |
| SNOMED CT | 766979005 | Squamous cell carcinoma of rectum |
| SNOMED CT | 766980008 | Squamous cell carcinoma of stomach |
| SNOMED CT | 767444009 | Germline BRCA-mutated, HER2-negative metastatic breast cancer |
| SNOMED CT | 767448007 | Pineoblastoma |
| SNOMED CT | 768470007 | Aldosterone-producing adenoma |
| SNOMED CT | 770601003 | Small cell carcinoma of ovary |
| SNOMED CT | 770628008 | Diffuse leptomeningeal melanocytosis |
| SNOMED CT | 770686005 | Malignant germ cell neoplasm of vagina |
| SNOMED CT | 771080008 | Hereditary site-specific ovarian cancer syndrome |
| SNOMED CT | 772992009 | Primary differentiated carcinoma of thyroid gland |
| SNOMED CT | 773283006 | Malignant germ cell neoplasm of cervix uteri |
| SNOMED CT | 773284000 | Malignant germ cell neoplasm of corpus uteri |
| SNOMED CT | 773624006 | Primary ameloblastic carcinoma |
| SNOMED CT | 773774000 | High-grade neuroendocrine carcinoma of corpus uteri |
| SNOMED CT | 773775004 | High-grade neuroendocrine carcinoma of cervix uteri |
| SNOMED CT | 778046002 | Somatomammotropinoma |
| SNOMED CT | 778066006 | Carcinofibroma of corpus uteri |
| SNOMED CT | 781076008 | Metastatic colorectal cancer |
| SNOMED CT | 781382000 | Malignant neoplasm of colon and/or rectum |
| SNOMED CT | 782722002 | Global developmental delay, lung cysts, overgrowth, Wilms tumor syndrome |
| SNOMED CT | 783006001 | Low-grade neuroendocrine neoplasm of corpus uteri |
| SNOMED CT | 783008000 | Pituitary dermoid and epidermoid cysts |
| SNOMED CT | 783056006 | Paratesticular adenocarcinoma |
| SNOMED CT | 783183009 | Salivary gland type carcinoma of esophagus |
| SNOMED CT | 783704005 | Undifferentiated carcinoma of esophagus |
| SNOMED CT | 785807007 | Transitional cell carcinoma of corpus uteri |
| SNOMED CT | 786038001 | Familial nonmedullary thyroid carcinoma |
| SNOMED CT | 788756004 | Spindle cell oncocytoma of posterior pituitary gland |
| SNOMED CT | 788757008 | Pituicytoma of posterior pituitary gland |
| SNOMED CT | 788758003 | Sellar ependymoma of posterior pituitary gland |
| SNOMED CT | 789689004 | Malignant lymphomatoid granulomatosis grade 3 |
| SNOMED CT | 789690008 | Malignant lymphomatoid granulomatosis grade 3 of lung |
| SNOMED CT | 789721005 | Malignant middle ear paraganglioma |
| SNOMED CT | 816205008 | Malignant pituitary blastoma |
| SNOMED CT | 818967003 | Medulloepithelioma of central nervous system |
| SNOMED CT | 822969007 | Acinar cell cystadenocarcinoma of lung |
| SNOMED CT | 822970008 | Acinar cell cystadenocarcinoma of prostate |
| SNOMED CT | 823017009 | Infiltrating duct carcinoma of prostate |
| SNOMED CT | 827162007 | Malignant immature teratoma of ovary |
| SNOMED CT | 830032008 | Dermoid cyst of occipital lobe |
| SNOMED CT | 830033003 | Dermoid cyst of brain |
| SNOMED CT | 830055006 | Anaplastic lymphoma kinase fusion oncogene negative non-small cell lung cancer |
| SNOMED CT | 830060005 | Reactive oxygen species 1 negative non-small cell lung cancer |

Continued on next page

**TableS24 – continued from previous page**

| CodeSystem | ConceptCode | ConceptName |
|---|---|---|
| SNOMED CT | 830151004 | Anaplastic lymphoma kinase fusion oncogene positive non-small cell lung cancer |
| SNOMED CT | 830196006 | Chordoma of cervical spine |
| SNOMED CT | 833285007 | Chordoma of lumbar spine |
| SNOMED CT | 833295000 | Chordoma of thoracic spine |
| SNOMED CT | 836274002 | Carcinomatosis of peritoneum |
| SNOMED CT | 840573001 | Carcinoma of epididymis and spermatic cord |
| SNOMED CT | 860830009 | Gastrointestinal stromal neoplasm of overlapping sites of stomach |
| SNOMED CT | 865954003 | Adenocarcinoma of breast |
| SNOMED CT | 866050001 | Mixed germ cell neoplasm of central nervous system |
| SNOMED CT | 866072004 | Malignant melanoma of orbit |
| SNOMED CT | 866073009 | Malignant melanoma of overlapping sites of eye and adnexa |
| SNOMED CT | 866079008 | Malignant melanoma of lacrimal gland |
| SNOMED CT | 866080006 | Malignant germ cell neoplasm of heart |
| SNOMED CT | 866082003 | Malignant melanoma of lacrimal drainage system |
| SNOMED CT | 866087009 | Pathologic fracture of vertebra at site of metastatic neoplasm |
| SNOMED CT | 866176003 | Primary clear cell peritoneal carcinoma |
| SNOMED CT | 870327007 | Papillary squamous cell carcinoma of nasopharynx |
| SNOMED CT | 870318006 | Osteosarcoma of bone of head |
| SNOMED CT | 870355006 | Neuroendocrine carcinoma of anus |
| SNOMED CT | 1661000119106 | Metastasis to lung from adenocarcinoma |
| SNOMED CT | 7391000119103 | Primary adenoid cystic carcinoma of nasopharynx |
| SNOMED CT | 11471000224106 | Diffuse intrinsic pontine glioma |
| SNOMED CT | 18121000119104 | Primary squamous cell carcinoma of palatine tonsil |
| SNOMED CT | 18831000119107 | Benign multiple endocrine neoplasia type 2a |
| SNOMED CT | 33581000119109 | Primary malignant neoplasm of extradural spinal cord |
| SNOMED CT | 45221000119105 | Primary invasive malignant neoplasm of female breast |
| SNOMED CT | 61471000119100 | Basal cell carcinoma of naris |
| SNOMED CT | 67771000119102 | Grade 2 astrocytoma of brain |
| SNOMED CT | 67811000119102 | Primary small cell malignant neoplasm of lung, TNM stage 1 |
| SNOMED CT | 67821000119109 | Primary small cell malignant neoplasm of lung, TNM stage 2 |
| SNOMED CT | 67831000119107 | Primary small cell malignant neoplasm of lung, TNM stage 3 |
| SNOMED CT | 67841000119103 | Primary small cell malignant neoplasm of lung, TNM stage 4 |
| SNOMED CT | 71111000119109 | Recurrent primary malignant neoplasm of vulva |
| SNOMED CT | 74411000119100 | Prostatic intraepithelial neoplasia and adjacent atypia |
| SNOMED CT | 78411000119107 | Ewing sarcoma of bone of pelvis |
| SNOMED CT | 82501000119102 | Anaplastic astrocytoma of central nervous system |
| SNOMED CT | 82761000119108 | Osteosarcoma of bone of upper limb |
| SNOMED CT | 87091000119101 | Malignant glioma of cerebrum |
| SNOMED CT | 87101000119106 | Primary malignant glioma of brain |
| SNOMED CT | 87111000119109 | Malignant glioma of hypothalamus |
| SNOMED CT | 87121000119102 | Malignant glioma of cerebellum |
| SNOMED CT | 87131000119104 | Primary extramedullary malignant tumor of spinal cord |
| SNOMED CT | 87151000119105 | Malignant glioma of central nervous system |
| SNOMED CT | 90811000119100 | Low grade malignant glioma of brain |
| SNOMED CT | 90831000119105 | Grade 4 malignant glioma of brain |

Continued on next page

**TableS24 – continued from previous page**

| CodeSystem | ConceptCode | ConceptName |
|---|---|---|
| SNOMED CT | 91031000119108 | Primary chondrosarcoma of bone of upper limb |
| SNOMED CT | 91041000119104 | Ewing sarcoma of bone structure of upper limb |
| SNOMED CT | 91061000119100 | Primary chondrosarcoma of bone of pelvis |
| SNOMED CT | 91081000119109 | Primary chondrosarcoma of bone of lower limb |
| SNOMED CT | 91131000119109 | Primary adenocarcinoma of vulva |
| SNOMED CT | 91141000119100 | Primary spindle cell carcinoma of urinary bladder |
| SNOMED CT | 91151000119103 | Primary squamous cell carcinoma of chest wall |
| SNOMED CT | 91161000119101 | Primary adenocarcinoma of chest wall |
| SNOMED CT | 91171000119107 | Primary undifferentiated large cell malignancy of chest wall |
| SNOMED CT | 91181000119105 | Squamous cell carcinoma metastatic to pleura |
| SNOMED CT | 91281000119103 | Secondary adenocarcinoma of bone |
| SNOMED CT | 94281000119101 | Malignant multiple endocrine neoplasia type 2a |
| SNOMED CT | 96291000119105 | Primary malignant inflammatory neoplasm of female breast |
| SNOMED CT | 96901000119105 | Prostate cancer metastatic to eye |
| SNOMED CT | 98981000119103 | Primary malignant neoplasm of ileocecal valve |
| SNOMED CT | 99101000119101 | Primary adenocarcinoma of fallopian tube |
| SNOMED CT | 99121000119105 | Primary adenocarcinoma of vagina |
| SNOMED CT | 99131000119108 | Astrocytoma of cerebrum |
| SNOMED CT | 100721000119109 | High grade astrocytoma of brain |
| SNOMED CT | 100731000119107 | Low grade astrocytoma of brain |
| SNOMED CT | 101371000119101 | Adamantinoma of long bone of lower limb |
| SNOMED CT | 101571000119107 | Ependymoma of cerebrum |
| SNOMED CT | 104981000119104 | Oligodendroglioma of cerebrum |
| SNOMED CT | 105111000119109 | Primary squamous cell carcinoma of thymus |
| SNOMED CT | 105121000119102 | Squamous cell carcinoma of vagina |
| SNOMED CT | 107561000119107 | Ependymoma of brain stem |
| SNOMED CT | 107571000119101 | Oligodendroglioma of brain stem |
| SNOMED CT | 107581000119103 | Astrocytoma of brain stem |
| SNOMED CT | 107591000119100 | Primary squamous cell carcinoma of urethra |
| SNOMED CT | 107601000119107 | Primary transitional cell carcinoma of urethra |
| SNOMED CT | 107691000119101 | Non-seminomatous germ cell neoplasm of testis |
| SNOMED CT | 107751000119102 | Primary malignant mixed Mullerian neoplasm of endometrium |
| SNOMED CT | 107771000119106 | Primary malignant clear cell neoplasm of endometrium |
| SNOMED CT | 107791000119107 | Primary adenosquamous carcinoma of endometrium |
| SNOMED CT | 108101000119101 | Malignant melanoma metastatic to kidney |
| SNOMED CT | 108131000119108 | Small cell carcinoma metastatic to kidney |
| SNOMED CT | 108201000119105 | Secondary undifferentiated large cell malignant neoplasm of kidney |
| SNOMED CT | 108211000119108 | Secondary adenocarcinoma of kidney |
| SNOMED CT | 113071000119101 | Chondromyxoid fibroma of long bone of lower limb |
| SNOMED CT | 113081000119103 | Chondromyxoid fibroma of long bone of upper limb |
| SNOMED CT | 113121000119101 | Osteochondroma of fibula |
| SNOMED CT | 113131000119103 | Osteochondroma of humerus |
| SNOMED CT | 113141000119107 | Osteochondroma of pelvis |
| SNOMED CT | 123241000119109 | Benign carcinoid tumor of stomach |
| SNOMED CT | 123701000119104 | Malignant carcinoid tumor of descending colon |
| SNOMED CT | 128041000119107 | Primary adenocarcinoma of distal third of esophagus |

Continued on next page

**TableS24 – continued from previous page**

| CodeSystem | ConceptCode | ConceptName |
|---|---|---|
| SNOMED CT | 130351000119105 | Primary malignant neuroendocrine neoplasm of large intestine |
| SNOMED CT | 130371000119101 | Primary malignant neuroendocrine neoplasm of cecum |
| SNOMED CT | 130891000119106 | Chordoma of coccyx |
| SNOMED CT | 130911000119108 | Chordoma of pelvis |
| SNOMED CT | 135091000119106 | Secondary neuroendocrine carcinoma of peritoneum |
| SNOMED CT | 147101000119108 | Primary malignant astrocytoma of central nervous system |
| SNOMED CT | 147131000119101 | Glioblastoma multiforme of central nervous system |
| SNOMED CT | 184741000119107 | Primary adenocarcinoma of anus |
| SNOMED CT | 184781000119102 | Primary adenocarcinoma of cervix uteri |
| SNOMED CT | 184801000119103 | Primary adenocarcinoma of lacrimal gland |
| SNOMED CT | 184861000119102 | Primary adenocarcinoma of nasopharynx |
| SNOMED CT | 208041000119100 | Primary adenocarcinoma of endocervix |
| SNOMED CT | 226521000119108 | Primary malignant neoplasm of nasopharynx |
| SNOMED CT | 234941000119100 | Malignant glioma of eye |
| SNOMED CT | 236811000119101 | Paget disease of anal canal |
| SNOMED CT | 242951000119108 | Primary squamous cell carcinoma of vermilion border of lip |
| SNOMED CT | 350861000119106 | Primary malignant neoplasm of right adrenal gland |
| SNOMED CT | 350921000119109 | Primary malignant neoplasm of left adrenal gland |
| SNOMED CT | 351171000119101 | Benign carcinoid of lung |
| SNOMED CT | 351961000119109 | Malignant melanoma of skin of right upper limb |
| SNOMED CT | 352001000119100 | Malignant melanoma of skin of right lower limb |
| SNOMED CT | 352201000119105 | Malignant melanoma of skin of left upper limb |
| SNOMED CT | 353421000119109 | Primary malignant neoplasm of axillary tail of left female breast |
| SNOMED CT | 353431000119107 | Primary malignant neoplasm of female left breast |
| SNOMED CT | 353441000119103 | Primary malignant neoplasm of central portion of female left breast |
| SNOMED CT | 353501000119104 | Primary malignant neoplasm of axillary tail of right female breast |
| SNOMED CT | 353521000119108 | Primary malignant neoplasm of central portion of female right breast |
| SNOMED CT | 353561000119103 | Secondary malignant neoplasm of right lung |
| SNOMED CT | 353661000119102 | Overlapping primary malignant neoplasm of bone and articular cartilage of lower limb |
| SNOMED CT | 353671000119108 | Overlapping primary malignant neoplasm of bone and articular cartilage of right upper limb |
| SNOMED CT | 353741000119106 | Secondary malignant neoplasm of left lung |
| SNOMED CT | 354351000119105 | Primary malignant neoplasm of left kidney |
| SNOMED CT | 354361000119107 | Primary malignant neoplasm of right kidney |
| SNOMED CT | 354491000119109 | Primary malignant neoplasm of left male breast |
| SNOMED CT | 354591000119108 | Primary malignant neoplasm of right male breast |
| SNOMED CT | 354621000119105 | Overlapping primary malignant neoplasm of bone and articular cartilage of left upper limb |
| SNOMED CT | 354641000119104 | Primary malignant neoplasm of bone of left upper limb |
| SNOMED CT | 354651000119102 | Primary malignant neoplasm of bone of left lower limb |
| SNOMED CT | 354671000119106 | Primary malignant neoplasm of bone of right upper limb |
| SNOMED CT | 354681000119109 | Primary malignant neoplasm of bone of right lower limb |
| SNOMED CT | 354701000119107 | Primary malignant neoplasm of left lung |

Continued on next page

**TableS24 – continued from previous page**

| CodeSystem | ConceptCode | ConceptName |
|---|---|---|
| SNOMED CT | 354741000119109 | Primary malignant neoplasm of right lung |
| SNOMED CT | 431911000124102 | Gnathic osteosarcoma |
| SNOMED CT | 432131000124100 | Malignant mesothelioma of the tunica vaginalis |
| SNOMED CT | 446531000124106 | Meningioma of cerebellum |
| SNOMED CT | 452291000124109 | Relapse multiple myeloma |
| SNOMED CT | 458321000124102 | Metastatic urothelial carcinoma |
| SNOMED CT | 458581000124106 | Metastatic HER2 positive gastroesophageal junction cancer |
| SNOMED CT | 459381000124106 | Metastatic castration-resistant prostate cancer |
| SNOMED CT | 459391000124109 | Metastatic human epidermal growth factor 2 positive carcinoma of breast |
| SNOMED CT | 459411000124109 | Metastatic collecting duct carcinoma |
| SNOMED CT | 459421000124101 | Metastatic penile cancer |
| SNOMED CT | 461351000124108 | Breast implant-associated anaplastic large cell lymphoma |
| SNOMED CT | 461371000124103 | Recurrent ovarian cancer |
| SNOMED CT | 681631000119108 | Primary adenocarcinoma of body of stomach |
| SNOMED CT | 681651000119102 | Primary adenocarcinoma of descending colon |
| SNOMED CT | 682701000119104 | Benign neoplasm of left lacrimal gland |
| SNOMED CT | 682761000119103 | Benign neoplasm of right lacrimal gland |
| SNOMED CT | 683991000119103 | Extensive stage primary small cell carcinoma of lung |
| SNOMED CT | 684911000119105 | Primary glioblastoma multiforme of frontal lobe |
| SNOMED CT | 1078881000119102 | Primary adenocarcinoma of lower lobe of left lung |
| SNOMED CT | 1078901000119100 | Primary adenocarcinoma of upper lobe of left lung |
| SNOMED CT | 1078931000119107 | Primary adenocarcinoma of lower lobe of right lung |
| SNOMED CT | 1078961000119104 | Primary adenocarcinoma of upper lobe of right lung |
| SNOMED CT | 1078991000119106 | Angiomyolipoma of left kidney |
| SNOMED CT | 1079001000119106 | Angiomyolipoma of right kidney |
| SNOMED CT | 1079111000119108 | Primary basal cell carcinoma of left eyelid |
| SNOMED CT | 1079171000119100 | Primary basal cell carcinoma of right eyelid |
| SNOMED CT | 1080091000119100 | Infiltrating ductal carcinoma of axillary tail of left female breast |
| SNOMED CT | 1080101000119105 | Infiltrating duct carcinoma of left female breast |
| SNOMED CT | 1080111000119108 | Infiltrating ductal carcinoma of central portion of left female breast |
| SNOMED CT | 1080121000119101 | Infiltrating ductal carcinoma of lower inner quadrant of left female breast |
| SNOMED CT | 1080131000119103 | Infiltrating ductal carcinoma of lower outer quadrant of left female breast |
| SNOMED CT | 1080151000119109 | Infiltrating ductal carcinoma of upper inner quadrant of left female breast |
| SNOMED CT | 1080161000119106 | Infiltrating ductal carcinoma of upper outer quadrant of left female breast |
| SNOMED CT | 1080171000119100 | Infiltrating ductal carcinoma of axillary tail of right female breast |
| SNOMED CT | 1080181000119102 | Invasive ductal carcinoma of right female breast |
| SNOMED CT | 1080191000119104 | Infiltrating ductal carcinoma of central portion of right female breast |
| SNOMED CT | 1080201000119101 | Infiltrating ductal carcinoma of lower inner quadrant of right female breast |

Continued on next page

**TableS24 – continued from previous page**

| CodeSystem | ConceptCode | ConceptName |
|---|---|---|
| SNOMED CT | 1080211000119103 | Infiltrating ductal carcinoma of lower outer quadrant of right female breast |
| SNOMED CT | 1080231000119108 | Infiltrating ductal carcinoma of upper inner quadrant of right female breast |
| SNOMED CT | 1080241000119104 | Infiltrating ductal carcinoma of upper outer quadrant of right female breast |
| SNOMED CT | 1080261000119100 | Infiltrating lobular carcinoma of left female breast |
| SNOMED CT | 1080341000119105 | Infiltrating lobular carcinoma of right female breast |
| SNOMED CT | 1080941000119109 | Malignant melanoma of left choroid |
| SNOMED CT | 1080981000119104 | Malignant melanoma of right choroid |
| SNOMED CT | 1081021000119109 | Malignant melanoma of skin of left lower limb |
| SNOMED CT | 1081231000119103 | Oncocytoma of left kidney |
| SNOMED CT | 1081241000119107 | Oncocytoma of right kidney |
| SNOMED CT | 1081551000119106 | Recurrent primary malignant neoplasm of left female breast |
| SNOMED CT | 1081561000119108 | Recurrent primary malignant neoplasm of right female breast |
| SNOMED CT | 1081571000119102 | Clear cell carcinoma of left kidney |
| SNOMED CT | 1081581000119104 | Clear cell carcinoma of right kidney |
| SNOMED CT | 1081781000119105 | Primary seminoma of left testis |
| SNOMED CT | 1081811000119107 | Primary seminoma of right testis |
| SNOMED CT | 1082281000119105 | Transitional cell carcinoma of left renal pelvis |
| SNOMED CT | 1082291000119108 | Transitional cell carcinoma of left ureter |
| SNOMED CT | 1082301000119109 | Transitional cell carcinoma of right renal pelvis |
| SNOMED CT | 1082311000119107 | Transitional cell carcinoma of right ureter |
| SNOMED CT | 1082321000119100 | Nephroblastoma of left kidney |
| SNOMED CT | 1082331000119102 | Nephroblastoma of right kidney |
| SNOMED CT | 1082701000112100 | Locally advanced breast cancer |
| SNOMED CT | 1082901000112103 | Primary malignant neoplasm of breast with axillary lymph node invasion |
| SNOMED CT | 1098981000119101 | Recurrent malignant neoplasm of prostate |
| SNOMED CT | 10708511000119108 | Primary malignant neoplasm of uterus |
| SNOMED CT | 10737861000119101 | Malignant germ cell neoplasm of right ovary |
| SNOMED CT | 10737911000119105 | Malignant germ cell neoplasm of left ovary |
| SNOMED CT | 12235561000119105 | Large cell carcinoma of left lung |
| SNOMED CT | 12235601000119105 | Large cell carcinoma of right lung |
| SNOMED CT | 12240951000119107 | Squamous cell carcinoma of left lung |
| SNOMED CT | 12240991000119102 | Squamous cell carcinoma of right lung |
| SNOMED CT | 12246561000119101 | Secondary malignant neoplasm of bilateral adrenal glands |
| SNOMED CT | 12246601000119101 | Secondary malignant neoplasm of bilateral lungs |
| SNOMED CT | 13351431000119102 | Secondary malignant neoplasm of lymph nodes of neck from thyroid |
| SNOMED CT | 15635721000119108 | Primary malignant neoplasm of both ovaries |
| SNOMED CT | 15635761000119103 | Infiltrating duct carcinoma of bilateral female breasts |
| SNOMED CT | 15635801000119106 | Primary malignant neoplasm of bilateral female breasts |
| SNOMED CT | 15636991000119103 | Renal cell carcinoma of bilateral kidneys |
| SNOMED CT | 15638291000119105 | Angiomyolipoma of bilateral kidneys |
| SNOMED CT | 15930861000119100 | Carcinosarcoma of left ovary |
| SNOMED CT | 15930901000119106 | Carcinosarcoma of right ovary |

Continued on next page

**TableS24 – continued from previous page**

| CodeSystem | ConceptCode | ConceptName |
|---|---|---|
| SNOMED CT | 15930941000119108 | Carcinosarcoma of bilateral ovaries |
| SNOMED CT | 15956181000119102 | Secondary adenocarcinoma of bilateral lungs |
| SNOMED CT | 15956341000119105 | Adenocarcinoma of left lung |
| SNOMED CT | 15956381000119100 | Adenocarcinoma of right lung |
| SNOMED CT | 15959061000119101 | Transitional cell carcinoma of left kidney |
| SNOMED CT | 15959101000119103 | Transitional cell carcinoma of right kidney |
| SNOMED CT | 15978071000119100 | Capillary hemangioma of right orbit region |
| SNOMED CT | 15978151000119108 | Capillary hemangioma of left orbit region |
| SNOMED CT | 16091171000119106 | Angiosarcoma of skin of cheek |
| SNOMED CT | 19090001000004101 | Metastatic neoplasm of left basal ganglion |
| SNOMED CT | 92511007 | Burkitt's tumor of lymph nodes of axilla and upper limb |
| SNOMED CT | 92512000 | Burkitt's tumor of lymph nodes of head, face, and neck |
| SNOMED CT | 92513005 | Burkitt's tumor of lymph nodes of inguinal region and lower limb |
| SNOMED CT | 94365007 | Cancer metastatic to large intestine |
| SNOMED CT | 94682004 | Secondary malignant neoplasm of Waldeyer ring |
| SNOMED CT | 254898001 | Paget disease vulvar cancer |
| SNOMED CT | 285420006 | Immunoglobulin A myeloma |
| SNOMED CT | 285421005 | Immunoglobulin G myeloma |
| SNOMED CT | 285422003 | Immunoglobulin D myeloma |
| SNOMED CT | 402911002 | Buschke-Löwenstein tumor of penis |
| SNOMED CT | 402912009 | Vulval verrucous carcinoma of Buschke-Lowenstein |
| SNOMED CT | 423829008 | Invasive vulval Paget disease |
| SNOMED CT | 123661000119106 | Malignant carcinoid tumour of lung |
| SNOMED CT | 123721000119108 | Malignant carcinoid tumour of ascending colon |
| SNOMED CT | 123731000119106 | Malignant carcinoid tumour of caecum |
| SNOMED CT | 123741000119102 | Malignant carcinoid tumour of large intestine |
| SNOMED CT | 123761000119103 | Malignant carcinoid tumour of duodenum |
| SNOMED CT | 143401000119106 | Malignant carcinoid tumour of pancreas |
| SNOMED CT | 192681000119104 | Malignant carcinoid tumour of appendix |
| SNOMED CT | 353511000119101 | Primary malignant neoplasm of female right breast |
| SNOMED CT | 1010603001 | Primary mucoepidermoid carcinoma of nasopharynx |
| SNOMED CT | 1010607000 | Primary lymphoepithelial carcinoma of nasopharynx |
| SNOMED CT | 1010624006 | Primary salivary gland type neoplasm of nasopharynx |
| SNOMED CT | 1010631005 | Primary malignant melanoma of overlapping sites of accessory sinuses |
| SNOMED CT | 1010633008 | Primary papillary adenocarcinoma of nasopharynx |
| SNOMED CT | 1010656004 | Adenoid squamous cell carcinoma of nasopharynx |
| SNOMED CT | 1010669000 | Primary squamous cell carcinoma of superior wall of nasopharynx |
| SNOMED CT | 1010674008 | Basaloid squamous cell carcinoma of nasopharynx |
| SNOMED CT | 1010675009 | Squamous cell carcinoma of posterior wall of nasopharynx |
| SNOMED CT | 1010677001 | Primary squamous cell carcinomas of anterior wall of nasopharynx |
| SNOMED CT | 1010679003 | Primary squamous cell carcinoma of lateral wall of nasopharynx |
| SNOMED CT | 1010681001 | Primary squamous cell carcinoma of overlapping sites of nasopharynx |
| SNOMED CT | 105041000119109 | Secondary squamous cell carcinoma of lung |

Continued on next page

**TableS24 – continued from previous page**

| CodeSystem | ConceptCode | ConceptName |
|---|---|---|
| SNOMED CT | 105051000119106 | Secondary undifferentiated large cell carcinoma of lung |
| SNOMED CT | 143291000119104 | Non-Hodgkin's lymphoma in remission |
| SNOMED CT | 15629411000119106 | Subependymoma of brain |
| SNOMED CT | 15957501000119101 | Bilateral oncocytoma of kidneys |
| SNOMED CT | 16903531000119107 | Atypical meningioma of cerebral meninges |
| SNOMED CT | 25521000119106 | Neoplasm of uncertain behavior of hypothalamus |
| SNOMED CT | 71101000119106 | Recurrent primary malignant neoplasm of vagina |
| SNOMED CT | 878805009 | Renal granular cell carcinoma |
| SNOMED CT | 878807001 | HER2-positive gastric cancer |
| SNOMED CT | 878808006 | Nongerminomatous germ cell tumor of central nervous system |
| SNOMED CT | 878857009 | Malignant lymphomatoid granulomatosis grade 3 |
| SNOMED CT | 878858004 | Malignant lymphomatoid granulomatosis grade 3 of lung |
| SNOMED CT | 879818005 | Fetal sacral teratoma |
| SNOMED CT | 879991002 | Gastrinoma of pancreas |
| SNOMED CT | 879992009 | Gastrinoma of duodenum |
| SNOMED CT | 880078001 | 11p15 deletion syndrome |
| SNOMED CT | 890528009 | Cancer of left lung |
| SNOMED CT | 890529001 | Cancer of right lung |
| SNOMED CT | 890534002 | Bilateral primary malignant neoplasm of lungs |
| SNOMED CT | 895111002 | Chordoma of vertebral column |
| SNOMED CT | 895345000 | Bilateral primary malignant neoplasm of kidneys |
| SNOMED CT | 895354002 | Primary malignant neoplasm of left ureter |
| SNOMED CT | 895355001 | Primary malignant neoplasm of right ureter |
| SNOMED CT | 895356000 | Bilateral primary malignant neoplasm of ureters |
| SNOMED CT | 895358004 | Primary malignant neoplasm of left testis |
| SNOMED CT | 895359007 | Primary malignant neoplasm of right testis |
| SNOMED CT | 895361003 | Bilateral primary malignant neoplasm of testes |
| SNOMED CT | 898197001 | Adult T-cell leukemia/lymphoma in remission |
| SNOMED CT | 898198006 | T-cell prolymphocytic leukemia in remission |
| SNOMED CT | 10737361000119109 | Primary adenocarcinoma of right fallopian tube |
| SNOMED CT | 10737401000119100 | Primary adenocarcinoma of left fallopian tube |
| SNOMED CT | 10737521000119108 | Right ovarian primary mucinous cystadenocarcinoma |
| SNOMED CT | 10737561000119103 | Left ovarian primary mucinous cystadenocarcinoma |
| SNOMED CT | 10737601000119103 | Primary serous papillary cystadenocarcinoma of right ovary |
| SNOMED CT | 10737641000119101 | Primary serous papillary cystadenocarcinoma of left ovary |
| SNOMED CT | 10737781000119101 | Right ovarian primary endometrioid carcinoma |
| SNOMED CT | 10737821000119106 | Left ovarian primary endometrioid carcinoma |
| SNOMED CT | 1078891000119104 | Primary adenocarcinoma of left main bronchus |
| SNOMED CT | 1078941000119103 | Primary adenocarcinoma of right main bronchus |
| SNOMED CT | 1078951000119101 | Primary adenocarcinoma of middle lobe of right lung |
| SNOMED CT | 1078971000119105 | Left lacrimal adenoid cystic carcinoma |
| SNOMED CT | 1078981000119108 | Right lacrimal adenoid cystic carcinoma |
| SNOMED CT | 1079531000119106 | Secondary malignant neoplasm of left kidney |
| SNOMED CT | 1079541000119102 | Secondary malignant neoplasm of right kidney |
| SNOMED CT | 1079551000119100 | Primary chondrosarcoma of bone of left upper limb |
| SNOMED CT | 1079561000119103 | Primary chondrosarcoma of bone of left foot |
| SNOMED CT | 1079571000119109 | Primary chondrosarcoma of bone of left hand |

Continued on next page

**TableS24 – continued from previous page**

| CodeSystem | ConceptCode | ConceptName |
|---|---|---|
| SNOMED CT | 1079581000119107 | Primary chondrosarcoma of bone of left lower limb |
| SNOMED CT | 1079591000119105 | Primary chondrosarcoma of left scapula |
| SNOMED CT | 1079601000119103 | Primary chondrosarcoma of bone of right upper limb |
| SNOMED CT | 1079611000119100 | Primary chondrosarcoma of bone of right foot |
| SNOMED CT | 1079621000119107 | Primary chondrosarcoma of bone of right hand |
| SNOMED CT | 1079631000119105 | Primary chondrosarcoma of bone of right lower limb |
| SNOMED CT | 1079641000119101 | Primary chondrosarcoma of right scapula |
| SNOMED CT | 1079991000119101 | Primary Ewing sarcoma of bone of left upper limb |
| SNOMED CT | 1080001000119106 | Primary Ewing sarcoma of bone of left foot |
| SNOMED CT | 1080021000119102 | Primary Ewing sarcoma of bone of left lower limb |
| SNOMED CT | 1080041000119108 | Primary Ewing sarcoma of bone of right upper limb |
| SNOMED CT | 1080051000119105 | Primary Ewing sarcoma of bone of right foot |
| SNOMED CT | 1080071000119101 | Primary Ewing sarcoma of bone of right lower limb |
| SNOMED CT | 1080451000119101 | Primary large cell carcinoma of lower lobe of left lung |
| SNOMED CT | 1080471000119105 | Primary large cell carcinoma of upper lobe of left lung |
| SNOMED CT | 1080501000119104 | Primary large cell carcinoma of lower lobe of right lung |
| SNOMED CT | 1080531000119106 | Primary large cell carcinoma of upper lobe of right lung |
| SNOMED CT | 1080561000119103 | Primary leiomyosarcoma of left kidney |
| SNOMED CT | 1080611000119100 | Primary leiomyosarcoma of right kidney |
| SNOMED CT | 1081271000119100 | Primary osteosarcoma of bone of left upper limb |
| SNOMED CT | 1081281000119102 | Primary osteosarcoma of bone of left foot |
| SNOMED CT | 1081291000119104 | Primary osteosarcoma of bone of left hand |
| SNOMED CT | 1081301000119103 | Primary osteosarcoma of bone of left lower limb |
| SNOMED CT | 1081311000119100 | Primary osteosarcoma of left scapula |
| SNOMED CT | 1081321000119107 | Primary osteosarcoma of bone of right upper limb |
| SNOMED CT | 1081331000119105 | Primary osteosarcoma of bone of right foot |
| SNOMED CT | 1081341000119101 | Primary osteosarcoma of bone of right hand |
| SNOMED CT | 1081351000119104 | Primary osteosarcoma of bone of right lower limb |
| SNOMED CT | 1081361000119102 | Primary osteosarcoma of right scapula |
| SNOMED CT | 1081591000119101 | Left primary retinoblastoma |
| SNOMED CT | 1081601000119108 | Right primary retinoblastoma |
| SNOMED CT | 1081841000119106 | Primary small cell carcinoma of lower lobe of left lung |
| SNOMED CT | 1081851000119108 | Primary small cell carcinoma of left main bronchus |
| SNOMED CT | 1081861000119105 | Primary small cell carcinoma of upper lobe of left lung |
| SNOMED CT | 1081891000119103 | Primary small cell carcinoma of lower lobe of right lung |
| SNOMED CT | 1081901000119104 | Primary small cell carcinoma of right main bronchus |
| SNOMED CT | 1081911000119101 | Primary small cell carcinoma of middle lobe of right lung |
| SNOMED CT | 1081921000119108 | Primary small cell carcinoma of upper lobe of right lung |
| SNOMED CT | 1082061000119108 | Primary squamous cell carcinoma of conjunctiva of left eye |
| SNOMED CT | 1082111000119109 | Primary squamous cell carcinoma of lower lobe of left lung |
| SNOMED CT | 1082121000119102 | Primary squamous cell carcinoma of left main bronchus |
| SNOMED CT | 1082141000119108 | Primary squamous cell carcinoma of upper lobe of left lung |
| SNOMED CT | 1082181000119103 | Primary squamous cell carcinoma of conjunctiva of right eye |
| SNOMED CT | 1082231000119109 | Primary squamous cell carcinoma of lower lobe of right lung |
| SNOMED CT | 1082241000119100 | Primary squamous cell carcinoma of right main bronchus |
| SNOMED CT | 1082251000119103 | Primary squamous cell carcinoma of middle lobe of right lung |
| SNOMED CT | 1082271000119107 | Primary squamous cell carcinoma of upper lobe of right lung |

Continued on next page

**TableS24 – continued from previous page**

| CodeSystem | ConceptCode | ConceptName |
|---|---|---|
| SNOMED CT | 10976631000119101 | Leiomyosarcoma of skin of chest |
| SNOMED CT | 10976711000119104 | Primary primitive neuroectodermal neoplasm of central nervous system |
| SNOMED CT | 1099571000119100 | Primary malignant neoplasm of left lacrimal sac |
| SNOMED CT | 1099581000119102 | Primary malignant neoplasm of right lacrimal sac |
| SNOMED CT | 1137644008 | Leiomyosarcoma of fundus of stomach |
| SNOMED CT | 1141621000 | Carcinoma of pyriform fossa |
| SNOMED CT | 1141627001 | Non-small cell lung carcinoma with NRG1 fusion |
| SNOMED CT | 1144252005 | Malignant neoplasm of lateral border of tongue |
| SNOMED CT | 1144303008 | Leiomyosarcoma of pylorus of stomach |
| SNOMED CT | 1144304002 | Leiomyosarcoma of pyloric antrum of stomach |
| SNOMED CT | 1144305001 | Leiomyosarcoma of gastric corpus structure |
| SNOMED CT | 1144306000 | Leiomyosarcoma of lesser curvature of stomach |
| SNOMED CT | 1144307009 | Leiomyosarcoma of greater curvature of stomach |
| SNOMED CT | 1144764001 | Malignant tumor of esophagus with NRG1 fusion |
| SNOMED CT | 1144922006 | Thymoma type AB |
| SNOMED CT | 1144924007 | Thymoma type B |
| SNOMED CT | 1144928005 | Thymoma type A |
| SNOMED CT | 1144945000 | Adnexal carcinoma of vulva |
| SNOMED CT | 1145245004 | Carcinoma of meibomian gland |
| SNOMED CT | 1148845007 | Follicular lymphoma grade 3b |
| SNOMED CT | 1148847004 | Follicular non-Hodgkin's lymphoma in situ |
| SNOMED CT | 1148851002 | Follicular lymphoma grade 3a |
| SNOMED CT | 1148852009 | Malignant cystic nephroma |
| SNOMED CT | 1148901006 | Squamous cell carcinoma of male genital |
| SNOMED CT | 1148904003 | Spermatocytic seminoma of testis |
| SNOMED CT | 1148906001 | Acute myeloid leukemia with t(8;21)(q22;q22) RUNX1-RUNX1T1 |
| SNOMED CT | 1148931000 | Cystic renal cell carcinoma of kidney |
| SNOMED CT | 1153336009 | Pediatric nodal marginal zone B cell lymphoma |
| SNOMED CT | 1153345005 | Myelodysplastic syndrome with single lineage dysplasia |
| SNOMED CT | 1153346006 | Primary non-Hodgkin malignant lymphoma of uveal tract |
| SNOMED CT | 1153347002 | Primary non-Hodgkin malignant lymphoma of vitreoretinal tract |
| SNOMED CT | 1153348007 | Primary effusion lymphoma due to human immune deficiency virus infection |
| SNOMED CT | 1153354008 | Malignant lymphoma of duodenum |
| SNOMED CT | 1153355009 | Malignant lymphoma of esophagus |
| SNOMED CT | 1153356005 | Lymphoma of anus |
| SNOMED CT | 1153357001 | Lymphoma of appendix |
| SNOMED CT | 1153377008 | Mixed phenotype acute leukemia with t(9;22) (q34;q11.2); BCR-ABL1 |
| SNOMED CT | 1153379006 | Mixed phenotype acute leukemia with T-cell and myeloid lineage |
| SNOMED CT | 1153381008 | Mixed phenotype acute leukemia with myeloid and B-cell lymphoid phenotypes |
| SNOMED CT | 1153382001 | Lymphoma of lacrimal gland |
| SNOMED CT | 1153383006 | Lymphoma of liver |
| SNOMED CT | 1153386003 | Gastrointestinal stromal neoplasm of cardia of stomach |

Continued on next page

**TableS24 – continued from previous page**

| CodeSystem | ConceptCode | ConceptName |
|---|---|---|
| SNOMED CT | 1153388002 | Gastrointestinal stromal neoplasm of body of stomach |
| SNOMED CT | 1153404003 | Gastrointestinal stromal neoplasm of pyloric antrum of stomach |
| SNOMED CT | 1153421007 | Chondrosarcoma of sternum |
| SNOMED CT | 1153422000 | Chondrosarcoma of mandible |
| SNOMED CT | 1153423005 | Chondrosarcoma of rib |
| SNOMED CT | 1153424004 | Chondrosarcoma of bone of pelvic wall |
| SNOMED CT | 1153425003 | Chondrosarcoma of clavicle |
| SNOMED CT | 1153426002 | Chondrosarcoma of skull |
| SNOMED CT | 1153427006 | Chondrosarcoma of vertebral column |
| SNOMED CT | 1155991005 | Secondary malignant neoplasm of leptomeninges |
| SNOMED CT | 1156403002 | Composite Hodgkin and non-Hodgkin lymphoma |
| SNOMED CT | 1156406005 | Anaplastic oligodendroglioma of central nervous system |
| SNOMED CT | 1156407001 | Myxopapillary ependymoma of spinal cord |
| SNOMED CT | 1156409003 | Anaplastic ependymoma of central nervous system |
| SNOMED CT | 1156410008 | Anaplastic oligoastrocytoma of central nervous system |
| SNOMED CT | 1156413005 | Gliomatosis cerebri |
| SNOMED CT | 1156414004 | Giant cell glioblastoma of brain |
| SNOMED CT | 1156415003 | Protoplasmic astrocytoma of brain |
| SNOMED CT | 1156416002 | Gemistocytic astrocytoma of brain |
| SNOMED CT | 1156417006 | Fibrillary astrocytoma of brain |
| SNOMED CT | 1156455001 | Pleomorphic xanthoastrocytoma of brain |
| SNOMED CT | 1156456000 | Pilomyxoid astrocytoma of brain |
| SNOMED CT | 1156459007 | Anaplastic ganglioglioma of central nervous system |
| SNOMED CT | 1156460002 | Desmoplastic medulloblastoma of brain |
| SNOMED CT | 1156461003 | Gliosarcoma of brain |
| SNOMED CT | 1156469001 | Large cell medulloblastoma of brain |
| SNOMED CT | 1156471001 | Choroid plexus carcinoma |
| SNOMED CT | 1156472008 | Papillary tumor of pineal region |
| SNOMED CT | 1156495004 | Dendritic cell sarcoma |
| SNOMED CT | 1156783003 | Leiomyosarcoma of rectum |
| SNOMED CT | 1156785005 | Leiomyosarcoma of duodenum |
| SNOMED CT | 1156786006 | Leiomyosarcoma of esophagus |
| SNOMED CT | 1156788007 | Leiomyosarcoma of colon |
| SNOMED CT | 1156795003 | Kaposi sarcoma of colon |
| SNOMED CT | 1156797006 | Kaposi sarcoma of rectum |
| SNOMED CT | 1156804004 | Peripheral neuroectodermal neoplasm of corpus uteri |
| SNOMED CT | 1156807006 | Peripheral neuroectodermal neoplasm of cervix uteri |
| SNOMED CT | 1156808001 | Malignant mixed Müllerian neoplasm of corpus uteri |
| SNOMED CT | 1156809009 | Malignant mesenchymal neoplasm of duodenum |
| SNOMED CT | 1156810004 | Malignant mesenchymal neoplasm of esophagus |
| SNOMED CT | 1156811000 | Malignant mesenchymal neoplasm of anus |
| SNOMED CT | 1156812007 | Malignant mesenchymal neoplasm of appendix |
| SNOMED CT | 1156833003 | Germinoma of central nervous system |
| SNOMED CT | 1157060001 | Diffuse astrocytoma of brain |
| SNOMED CT | 1157061002 | Gliosarcoma of central nervous system |
| SNOMED CT | 1157062009 | Giant cell glioblastoma of central nervous system |
| SNOMED CT | 1157064005 | Gemistocytic astrocytoma of central nervous system |

Continued on next page

**TableS24 – continued from previous page**

| CodeSystem | ConceptCode | ConceptName |
|---|---|---|
| SNOMED CT | 1157067003 | Malignant neoplasm of vertebra |
| SNOMED CT | 1157068008 | Fibrillary astrocytoma of central nervous system |
| SNOMED CT | 1157072007 | Gliosarcoma of spinal cord |
| SNOMED CT | 1157141006 | Astroblastoma of central nervous system |
| SNOMED CT | 1157157006 | Acute myeloid leukemia with 11q23 abnormality |
| SNOMED CT | 1157162007 | Intravascular large B-cell lymphoma |
| SNOMED CT | 1157249003 | Invasive benign pituitary adenoma |
| SNOMED CT | 12241031000119108 | Secondary malignant neoplasm of left breast |
| SNOMED CT | 12241071000119106 | Secondary malignant neoplasm of right breast |
| SNOMED CT | 12246481000119106 | Bilateral secondary malignant neoplasm of kidneys |
| SNOMED CT | 12246641000119104 | Bilateral secondary malignant neoplasm of breasts |
| SNOMED CT | 130941000119107 | Primary chordoma of bone of skull |
| SNOMED CT | 145501000119108 | Secondary malignant neoplasm of breast |
| SNOMED CT | 15930821000119105 | Bilateral secondary malignant neoplasm of ovaries |
| SNOMED CT | 15931021000119108 | Left ovarian primary sarcoma |
| SNOMED CT | 15931061000119103 | Right ovarian primary sarcoma |
| SNOMED CT | 15949941000119101 | Primary squamous cell carcinoma of skin of left breast |
| SNOMED CT | 15950021000119100 | Primary squamous cell carcinoma of skin of right breast |
| SNOMED CT | 15950061000119105 | Primary malignant neoplasm of skin of right breast |
| SNOMED CT | 15950101000119108 | Primary basal cell carcinoma of skin of right breast |
| SNOMED CT | 15950141000119105 | Primary basal cell carcinoma of skin of left breast |
| SNOMED CT | 15950221000119108 | Primary malignant neoplasm of skin of left breast |
| SNOMED CT | 15951021000119107 | Bilateral malignant melanoma of skin of lower limbs |
| SNOMED CT | 15951061000119102 | Bilateral malignant melanoma of skin of upper limbs |
| SNOMED CT | 15955101000119101 | Primary malignant neoplasm of right greater vestibular gland |
| SNOMED CT | 15955141000119104 | Primary malignant neoplasm of left greater vestibular gland |
| SNOMED CT | 15955901000119104 | Primary small cell carcinoma of right lung |
| SNOMED CT | 15955941000119102 | Primary small cell carcinoma of left lung |
| SNOMED CT | 15956221000119105 | Secondary adenocarcinoma of right lung |
| SNOMED CT | 15956261000119100 | Secondary adenocarcinoma of left lung |
| SNOMED CT | 15956661000119102 | Secondary small cell carcinoma of right lung |
| SNOMED CT | 15956701000119109 | Secondary small cell carcinoma of left lung |
| SNOMED CT | 15956741000119106 | Bilateral secondary small cell carcinoma of lungs |
| SNOMED CT | 15957141000119109 | Bilateral secondary squamous cell carcinoma of lungs |
| SNOMED CT | 15957181000119104 | Secondary squamous cell carcinoma of right lung |
| SNOMED CT | 15957221000119107 | Secondary squamous cell carcinoma of left lung |
| SNOMED CT | 15957261000119102 | Bilateral Kaposi sarcoma of lungs |
| SNOMED CT | 15958021000119103 | Primary sarcoma of left kidney |
| SNOMED CT | 15958101000119106 | Primary sarcoma of right kidney |
| SNOMED CT | 15958181000119103 | Bilateral secondary adenocarcinoma of kidneys |
| SNOMED CT | 15958221000119106 | Secondary adenocarcinoma of left kidney |
| SNOMED CT | 15958261000119101 | Secondary adenocarcinoma of right kidney |
| SNOMED CT | 15958301000119109 | Secondary squamous cell carcinoma of right kidney |
| SNOMED CT | 15958381000119101 | Secondary squamous cell carcinoma of left kidney |
| SNOMED CT | 15958661000119103 | Secondary malignant melanoma of left kidney |
| SNOMED CT | 15958701000119105 | Bilateral secondary malignant melanoma of kidneys |
| SNOMED CT | 15958741000119107 | Secondary malignant melanoma of right kidney |

Continued on next page

**TableS24 – continued from previous page**

| CodeSystem | ConceptCode | ConceptName |
|---|---|---|
| SNOMED CT | 15977551000119100 | Bilateral secondary malignant neoplasm of eyes |
| SNOMED CT | 15977591000119105 | Secondary malignant neoplasm of right eye |
| SNOMED CT | 15977631000119105 | Secondary malignant neoplasm of left eye |
| SNOMED CT | 16091411000119109 | Primary angiosarcoma of skin of lip |
| SNOMED CT | 16218761000119102 | Primary squamous cell carcinoma of skin of lip |
| SNOMED CT | 16527441000119105 | Primary non-small cell carcinoma of lower lobe of left lung |
| SNOMED CT | 16527481000119100 | Primary non-small cell carcinoma of middle lobe of right lung |
| SNOMED CT | 16527521000119100 | Primary non-small cell carcinoma of upper lobe of left lung |
| SNOMED CT | 16527561000119105 | Primary non-small cell carcinoma of lower lobe of right lung |
| SNOMED CT | 16527601000119105 | Primary non-small cell carcinoma of right lung |
| SNOMED CT | 16527641000119107 | Primary non-small cell carcinoma of left lung |
| SNOMED CT | 16527721000119108 | Primary non-small cell carcinoma of upper lobe of right lung |
| SNOMED CT | 16636051000119105 | Primary malignant gastrointestinal stromal neoplasm of colon |
| SNOMED CT | 16636101000119105 | Primary malignant gastrointestinal stromal neoplasm of rectum |
| SNOMED CT | 16774671000119108 | Primary basal cell carcinoma of skin of labium majus |
| SNOMED CT | 16775311000119107 | Mycosis fungoides of facial lymph node |
| SNOMED CT | 16845301000119108 | Primary glioblastoma multiforme of cerebellum |
| SNOMED CT | 16845341000119105 | Primary glioblastoma multiforme of brainstem |
| SNOMED CT | 16845381000119100 | Primary chondrosarcoma of sternum |
| SNOMED CT | 16899941000119102 | Primary squamous cell carcinoma of skin of anus |
| SNOMED CT | 16905141000119107 | Primary malignant neoplasm of skin of right upper eyelid |
| SNOMED CT | 16905341000119105 | Primary malignant neoplasm of skin of right lower eyelid |
| SNOMED CT | 16905381000119100 | Primary basal cell carcinoma of left upper eyelid |
| SNOMED CT | 16905421000119109 | Primary basal cell carcinoma of right upper eyelid |
| SNOMED CT | 16905461000119104 | Primary basal cell carcinoma of right lower eyelid |
| SNOMED CT | 16905501000119104 | Primary basal cell carcinoma of left lower eyelid |
| SNOMED CT | 16905541000119102 | Primary malignant neoplasm of skin of left lower eyelid |
| SNOMED CT | 16905581000119107 | Primary malignant neoplasm of skin of left upper eyelid |
| SNOMED CT | 16906481000119103 | Primary squamous cell carcinoma of right upper eyelid |
| SNOMED CT | 16906521000119103 | Primary squamous cell carcinoma of right lower eyelid |
| SNOMED CT | 16906561000119108 | Primary squamous cell carcinoma of left upper eyelid |
| SNOMED CT | 16906601000119108 | Primary squamous cell carcinoma of left lower eyelid |
| SNOMED CT | 16908281000119101 | Squamous cell carcinoma in situ of skin of right lower eyelid |
| SNOMED CT | 16908321000119106 | Squamous cell carcinoma in situ of skin of left lower eyelid |
| SNOMED CT | 243971001000119103 | Malignant melanoma of skin of right ankle |
| SNOMED CT | 249389151000119103 | Malignant melanoma of skin of right wrist |
| SNOMED CT | 350681000119101 | Primary malignant neoplasm of adrenal medulla of right adrenal gland |
| SNOMED CT | 350801000119105 | Primary malignant neoplasm of left adrenal cortex |
| SNOMED CT | 350851000119109 | Primary malignant neoplasm of adrenal medulla of left adrenal gland |
| SNOMED CT | 350881000119102 | Primary malignant neoplasm of right adrenal cortex |
| SNOMED CT | 351251000119103 | Acute myelofibrosis in remission |
| SNOMED CT | 352331000119105 | Primary malignant neoplasm of left spermatic cord |
| SNOMED CT | 352341000119101 | Primary malignant neoplasm of right spermatic cord |
| SNOMED CT | 352911000119101 | Primary malignant neoplasm of left optic nerve |
| SNOMED CT | 352921000119108 | Primary malignant neoplasm of right optic nerve |

Continued on next page

**TableS24 – continued from previous page**

| CodeSystem | ConceptCode | ConceptName |
|---|---|---|
| SNOMED CT | 352961000119103 | Primary malignant neoplasm of right renal pelvis |
| SNOMED CT | 353061000119107 | Right retinal primary malignant neoplasm |
| SNOMED CT | 353081000119103 | Left retinal primary malignant neoplasm |
| SNOMED CT | 353111000119108 | Primary malignant neoplasm of left renal pelvis |
| SNOMED CT | 353571000119109 | Secondary malignant neoplasm of right adrenal gland |
| SNOMED CT | 353721000119100 | Secondary malignant neoplasm of left adrenal gland |
| SNOMED CT | 353761000119105 | Left corneal primary malignant neoplasm |
| SNOMED CT | 353791000119103 | Right conjunctival primary malignant neoplasm |
| SNOMED CT | 353801000119102 | Right choroidal primary malignant neoplasm |
| SNOMED CT | 353811000119104 | Right corneal primary malignant neoplasm |
| SNOMED CT | 353841000119100 | Left conjunctival primary malignant neoplasm |
| SNOMED CT | 354301000119106 | Left choroidal primary malignant neoplasm |
| SNOMED CT | 354391000119100 | Kaposi sarcoma of left lung |
| SNOMED CT | 354461000119102 | Kaposi sarcoma of right lung |
| SNOMED CT | 354721000119103 | Primary malignant neoplasm of left main bronchus |
| SNOMED CT | 354731000119100 | Primary malignant neoplasm of right main bronchus |
| SNOMED CT | 400375351000119102 | Malignant melanoma of skin of left ankle |
| SNOMED CT | 681591000119108 | Primary adenocarcinoma of anterior wall of urinary bladder |
| SNOMED CT | 681611000119103 | Primary adenocarcinoma of neck of urinary bladder |
| SNOMED CT | 681661000119100 | Primary adenocarcinoma of dome of urinary bladder |
| SNOMED CT | 681681000119109 | Primary adenocarcinoma of fundus of stomach |
| SNOMED CT | 681701000119107 | Primary adenocarcinoma of greater curvature of stomach |
| SNOMED CT | 681781000119104 | Primary adenocarcinoma of lesser curvature of stomach |
| SNOMED CT | 681941000119107 | Primary adenocarcinoma of pylorus |
| SNOMED CT | 681991000119104 | Primary adenocarcinoma of trigone of urinary bladder |
| SNOMED CT | 682031000119109 | Primary adenocarcinoma of urachus |
| SNOMED CT | 682041000119100 | Primary adenocarcinoma of ureteral orifice |
| SNOMED CT | 682221000119103 | Primary anaplastic astrocytoma of cerebrum |
| SNOMED CT | 682231000119100 | Primary anaplastic astrocytoma of frontal lobe |
| SNOMED CT | 682241000119109 | Primary anaplastic astrocytoma of occipital lobe |
| SNOMED CT | 682251000119106 | Primary anaplastic astrocytoma of parietal lobe |
| SNOMED CT | 682261000119108 | Primary anaplastic astrocytoma of temporal lobe |
| SNOMED CT | 682291000119101 | Primary angiosarcoma of axillary region |
| SNOMED CT | 682411000119106 | Primary angiosarcoma of thorax |
| SNOMED CT | 682451000119107 | Primary astrocytoma of frontal lobe |
| SNOMED CT | 682471000119103 | Primary astrocytoma of parietal lobe |
| SNOMED CT | 682481000119100 | Primary astrocytoma of temporal lobe |
| SNOMED CT | 683031000119101 | Primary carcinoma ex pleomorphic adenoma of salivary gland |
| SNOMED CT | 683041000119105 | Primary carcinoma ex pleomorphic adenoma of submandibular gland |
| SNOMED CT | 683121000119103 | Primary chondrosarcoma of mandible |
| SNOMED CT | 683131000119100 | Primary chondrosarcoma of vertebral column |
| SNOMED CT | 683761000119107 | Primary ependymoma of brain ventricle |
| SNOMED CT | 683791000119100 | Primary ependymoma of parietal lobe |
| SNOMED CT | 683981000119101 | Primary Ewing sarcoma of bone of spine |
| SNOMED CT | 684901000119107 | Primary glioblastoma multiforme of cerebrum |
| SNOMED CT | 684921000119103 | Primary glioblastoma multiforme of occipital lobe |

Continued on next page

**TableS24 – continued from previous page**

| CodeSystem | ConceptCode | ConceptName |
|---|---|---|
| SNOMED CT | 684931000119100 | Primary glioblastoma multiforme of parietal lobe |
| SNOMED CT | 684941000119109 | Primary glioblastoma multiforme of temporal lobe |
| SNOMED CT | 685721000119101 | Primary leiomyosarcoma of thorax |
| SNOMED CT | 685851000119104 | Primary liposarcoma of soft tissue of axilla |
| SNOMED CT | 685971000119103 | Primary liposarcoma of soft tissue of thorax |
| SNOMED CT | 686431000119100 | Primary malignant glioma of frontal lobe |
| SNOMED CT | 686441000119109 | Primary malignant glioma of occipital lobe |
| SNOMED CT | 686451000119106 | Primary malignant glioma of parietal lobe |
| SNOMED CT | 686461000119108 | Primary malignant glioma of temporal lobe |
| SNOMED CT | 686591000119102 | Primary malignant pheochromocytoma of left adrenal gland |
| SNOMED CT | 686601000119109 | Primary malignant pheochromocytoma of right adrenal gland |
| SNOMED CT | 686981000119108 | Mycosis fungoides of inguinal lymph node |
| SNOMED CT | 686991000119106 | Mycosis fungoides of lower limb lymph node |
| SNOMED CT | 687331000119109 | Primary oligodendroglioma of frontal lobe |
| SNOMED CT | 687351000119103 | Primary oligodendroglioma of parietal lobe |
| SNOMED CT | 687361000119101 | Primary oligodendroglioma of temporal lobe |
| SNOMED CT | 687401000119105 | Primary osteosarcoma of mandible |
| SNOMED CT | 687421000119101 | Primary osteosarcoma of vertebral column |
| SNOMED CT | 687911000119100 | Primary rhabdomyosarcoma of thorax |
| SNOMED CT | 687961000119102 | Primary sarcoma of axilla |
| SNOMED CT | 688081000119108 | Primary sarcoma of thorax |
| SNOMED CT | 688671000119103 | Primary squamous cell carcinoma of anterior floor of mouth |
| SNOMED CT | 688681000119100 | Primary squamous cell carcinoma of body of tongue |
| SNOMED CT | 688691000119102 | Primary squamous cell carcinoma of anterior wall of urinary bladder |
| SNOMED CT | 688711000119104 | Primary squamous cell carcinoma of aryepiglottic fold |
| SNOMED CT | 688731000119109 | Primary squamous cell carcinoma of neck of urinary bladder |
| SNOMED CT | 688751000119103 | Primary squamous cell carcinoma of border of tongue |
| SNOMED CT | 688781000119105 | Primary squamous cell carcinoma of commissure of lip |
| SNOMED CT | 688801000119109 | Primary squamous cell carcinoma of dome of urinary bladder |
| SNOMED CT | 688811000119107 | Primary squamous cell carcinoma of dorsal surface of tongue |
| SNOMED CT | 688861000119105 | Primary squamous cell carcinoma of hard palate |
| SNOMED CT | 688891000119103 | Primary squamous cell carcinoma of mucous membrane of lip |
| SNOMED CT | 688901000119104 | Primary squamous cell carcinoma of mucous membrane of lower lip |
| SNOMED CT | 688911000119101 | Primary squamous cell carcinoma of mucous membrane of upper lip |
| SNOMED CT | 688931000119106 | Primary squamous cell carcinoma of lateral floor of mouth |
| SNOMED CT | 688941000119102 | Primary squamous cell carcinoma of lateral wall of urinary bladder |
| SNOMED CT | 688981000119107 | Primary squamous cell carcinoma of lower gum |
| SNOMED CT | 689231000119107 | Primary squamous cell carcinoma of posterior wall of urinary bladder |
| SNOMED CT | 689251000119101 | Primary squamous cell carcinoma of posterior wall of nasopharynx |
| SNOMED CT | 689281000119108 | Primary squamous cell carcinoma of retromolar area |
| SNOMED CT | 689311000119105 | Primary squamous cell carcinoma of soft palate |

Continued on next page

**TableS24 – continued from previous page**

| CodeSystem | ConceptCode | ConceptName |
|---|---|---|
| SNOMED CT | 689361000119108 | Primary squamous cell carcinoma of tonsillar fossa |
| SNOMED CT | 689371000119102 | Primary squamous cell carcinoma of tonsillar pillar |
| SNOMED CT | 689401000119104 | Primary squamous cell carcinoma of upper gum |
| SNOMED CT | 689441000119102 | Primary squamous cell carcinoma of ureteral orifice |
| SNOMED CT | 689451000119100 | Primary squamous cell carcinoma of uvula |
| SNOMED CT | 689471000119109 | Primary squamous cell carcinoma of inferior surface of tongue |
| SNOMED CT | 689481000119107 | Primary squamous cell carcinoma of vermilion border of lower lip |
| SNOMED CT | 689491000119105 | Primary squamous cell carcinoma of vermilion border of upper lip |
| SNOMED CT | 689501000119103 | Primary squamous cell carcinoma of vestibule of mouth |
| SNOMED CT | 689771000119103 | Primary transitional cell carcinoma of anterior wall of urinary bladder |
| SNOMED CT | 689781000119100 | Primary transitional cell carcinoma of neck of urinary bladder |
| SNOMED CT | 689791000119102 | Primary transitional cell carcinoma of dome of urinary bladder |
| SNOMED CT | 689801000119101 | Primary transitional cell carcinoma of lateral wall of urinary bladder |
| SNOMED CT | 689821000119105 | Primary transitional cell carcinoma of posterior wall of urinary bladder |
| SNOMED CT | 689831000119108 | Primary transitional cell carcinoma of trigone of urinary bladder |
| SNOMED CT | 689851000119102 | Primary transitional cell carcinoma of ureteral orifice |
| SNOMED CT | 690121000119109 | Primary basal cell carcinoma of skin of scrotum |
| SNOMED CT | 690141000119103 | Primary basal cell carcinoma of skin of male genitalia |
| SNOMED CT | 763101411000119100 | Primary renal cell carcinoma of left kidney |
| SNOMED CT | 808558991000119105 | Malignant melanoma of skin of left forearm |
| SNOMED CT | 84821000119100 | Primary follicular dendritic cell sarcoma of spleen |
| SNOMED CT | 87141000119108 | Primary malignant neoplasm of intramedullary spinal cord |
| SNOMED CT | 894171571000119107 | Primary renal cell carcinoma of right kidney |
| SNOMED CT | 91191000119108 | Secondary adenocarcinoma of pleura |
| SNOMED CT | 91211000119109 | Secondary small cell carcinoma of pleura |
| SNOMED CT | 91221000119102 | Secondary malignant melanoma of pleura |
| SNOMED CT | 91271000119101 | Secondary squamous cell carcinoma of bone |
| SNOMED CT | 939595491000119108 | Malignant melanoma of skin of right forearm |
| SNOMED CT | 985355341000119101 | Malignant melanoma of skin of left wrist |
| SNOMED CT | 1162280003 | Rhabdomyosarcoma of isthmus uteri |
| SNOMED CT | 1162282006 | Rhabdomyosarcoma of myometrium of corpus uteri |
| SNOMED CT | 1217383007 | RELA fusion-positive supratentorial ependymoma |
| SNOMED CT | 1197323000 | Metastatic carcinoma of thoracic lymph node |
| SNOMED CT | 1197324006 | Metastatic malignant melanoma of skin |
| SNOMED CT | 1197293007 | Metastatic squamous cell carcinoma of lymph nodes of head and neck |
| SNOMED CT | 1208457007 | Small cell neuroendocrine carcinoma of prostate |
| SNOMED CT | 1196815001 | Benign neoplasm of lacrimal apparatus |
| SNOMED CT | 1208741006 | SMARCA4-deficient undifferentiated neoplasm of thorax |
| SNOMED CT | 1197285007 | Metastatic squamous cell carcinoma of thoracic lymph node |
| SNOMED CT | 1197344000 | T-cell lymphoma of small intestine |
| SNOMED CT | 1201780008 | Chondrosarcoma of gingiva |

Continued on next page

**TableS24 – continued from previous page**

| CodeSystem | ConceptCode | ConceptName |
|---|---|---|
| SNOMED CT | 1208461001 | Plasmacytoid urothelial carcinoma of urinary bladder |
| SNOMED CT | 1197262006 | Pleomorphic adenoma of lacrimal system |
| SNOMED CT | 353211000119103 | Extramedullary plasmacytoma in remission |
| SNOMED CT | 352581000119104 | Primary malignant neoplasm of right descended testis |
| SNOMED CT | 1177173001 | Congenital progressive bone marrow failure, B-cell immunodeficiency, skeletal dysplasia syndrome |
| SNOMED CT | 1162531007 | Acute myeloid leukemia in complete remission |
| SNOMED CT | 1231381006 | Clear cell urothelial carcinoma of urinary system |
| SNOMED CT | 1197359006 | Familial colorectal cancer type X |
| SNOMED CT | 1196879002 | Familial ovarian cancer |
| SNOMED CT | 1217010007 | HER2-expressing (human epidermal growth factor 2-expressing) colorectal malignant neoplasm |
| SNOMED CT | 1217490003 | Kaposi sarcoma of gingiva |
| SNOMED CT | 1222581000 | Lipid-rich urothelial carcinoma of urinary system |
| SNOMED CT | 1196849009 | Lymphangioma of conjunctiva |
| SNOMED CT | 145298771000119104 | Malignant carcinoid tumor of left lung |
| SNOMED CT | 793197981000119106 | Malignant carcinoid tumor of right lung |
| SNOMED CT | 123691000119104 | Malignant carcinoid tumor of sigmoid colon |
| SNOMED CT | 1197294001 | Malignant melanoma arising in melanocytic nevus |
| SNOMED CT | 1197334002 | Malignant melanoma of uveal tract |
| SNOMED CT | 1197313009 | Metastatic adenocarcinoma of lymph node of head and neck |
| SNOMED CT | 1197328009 | Metastatic adenocarcinoma of thoracic lymph node |
| SNOMED CT | 1197326008 | Metastatic carcinoma of lymph node of head and neck |
| SNOMED CT | 1197267000 | Oxyphilic adenoma of lacrimal system |
| SNOMED CT | 1196856003 | Primary biphasic malignant mesothelioma of pleura |
| SNOMED CT | 1208483002 | Primary choriocarcinoma of central nervous system |
| SNOMED CT | 1196841007 | Primary desmoplastic mesothelioma of pleura |
| SNOMED CT | 1196839006 | Primary embryonal carcinoma of brain |
| SNOMED CT | 1196883002 | Primary epithelioid malignant mesothelioma of pleura |
| SNOMED CT | 1196844004 | Primary malignant astrocytoma of brain |
| SNOMED CT | 1196898000 | Primary malignant atypical teratoid rhabdoid neoplasm of brain |
| SNOMED CT | 1231267007 | Primary malignant ependymoma of optic nerve |
| SNOMED CT | 1197298003 | Primary malignant germ cell neoplasm of pineal gland |
| SNOMED CT | 1197296004 | Primary malignant germ cell neoplasm of pleura |
| SNOMED CT | 1197332003 | Primary malignant melanoma of central nervous system |
| SNOMED CT | 1197354001 | Primary malignant mesenchymal neoplasm of colon |
| SNOMED CT | 1197355000 | Primary malignant mesenchymal neoplasm of rectum |
| SNOMED CT | 1197266009 | Primary papillary carcinoma of body of uterus |
| SNOMED CT | 1197268005 | Primary papillary carcinoma of cervix uteri |
| SNOMED CT | 1197263001 | Primary poorly differentiated endocrine carcinoma of cervix uteri |
| SNOMED CT | 1197260003 | Primary poorly differentiated endocrine carcinoma of corpus uteri |
| SNOMED CT | 1197258000 | Primary salivary gland type carcinoma of palate |
| SNOMED CT | 1197259008 | Primary sarcomatoid malignant mesothelioma of pleura |
| SNOMED CT | 1197275006 | Primary squamous cell carcinoma of fallopian tube |
| SNOMED CT | 1229871006 | Primary squamous cell carcinoma of nasal cavity and paranasal sinus |

Continued on next page

**TableS24 – continued from previous page**

| CodeSystem | ConceptCode | ConceptName |
|---|---|---|
| SNOMED CT | 16899901000119104 | Primary squamous cell carcinoma of skin of chest |
| SNOMED CT | 1197276007 | Primary squamous cell carcinoma of Waldeyer ring |
| SNOMED CT | 1197343006 | Primary teratoma of brain |
| SNOMED CT | 1217020002 | Sarcomatoid urothelial carcinoma of ureter |
| SNOMED CT | 1208460000 | Sarcomatoid urothelial carcinoma of urinary bladder |
| SNOMED CT | 1230318000 | Serous carcinoma of body of uterus |
| SNOMED CT | 1208451008 | Transitional cell carcinoma of upper urinary tract |
| SNOMED CT | 1197280002 | Yolk sac tumor of central nervous system |
| SNOMED CT | 1196900003 | B cell lymphoma of small intestine |
| SNOMED CT | 1196876009 | Primary acinar cell carcinoma of parotid gland |
| SNOMED CT | 1196911006 | Primary adenocarcinoma of iris neuroepithelium |
| SNOMED CT | 682461000119109 | Primary astrocytoma of occipital lobe |
| SNOMED CT | 681921000119101 | Primary adenocarcinoma of posterior wall of urinary bladder |
| SNOMED CT | 1179762006 | Malignant neoplasm of middle lobe of right lung |
| SNOMED CT | 682441000119105 | Primary astrocytoma of cerebral ventricle |
| SNOMED CT | 1197350005 | Primary malignant Müllerian mixed neoplasm of cervix uteri |
| SNOMED CT | 352551000119106 | Primary malignant neoplasm of left descended testis |
| SNOMED CT | 1196860000 | Primary anaplastic large cell medulloblastoma of brain |
| SNOMED CT | 1163357003 | Malignant neoplasm of oral cavity and lip and salivary gland |
| SNOMED CT | 1186615002 | Primary Kaposi sarcoma of palate |
| SNOMED CT | 1163568002 | Primary malignant neoplasm of overlapping sites of colon |
| SNOMED CT | 1179375008 | Adenocarcinoma of parathyroid gland |
| SNOMED CT | 1179377000 | Adenocarcinoma of pituitary gland |
| SNOMED CT | 1187123005 | Mixed phenotype acute leukemia |
| SNOMED CT | 1172704005 | High grade B-cell lymphoma with MYC and BCL2 and/or BCL6 rearrangements |
| SNOMED CT | 1162264008 | Malignant lymphoma of uveal tract |
| SNOMED CT | 1162354002 | Osteosarcoma of articular cartilage of rib |
| SNOMED CT | 1162353008 | Osteosarcoma of articular cartilage of vertebra |
| SNOMED CT | 1162349006 | Osteosarcoma of bone of clavicle |
| SNOMED CT | 1162351005 | Osteosarcoma of bone of rib |
| SNOMED CT | 1162348003 | Osteosarcoma of bone of vertebra |
| SNOMED CT | 1162346004 | Osteosarcoma of sternum |
| SNOMED CT | 1162284007 | Rhabdomyosarcoma of endometrium of corpus uteri |
| SNOMED CT | 1162286009 | Rhabdomyosarcoma of fundus uteri |
| SNOMED CT | 1162311007 | Sarcoma of corpus uteri |
| SNOMED CT | 1187464007 | Clear cell sarcoma of kidney |

#### 5.4 Cerebrovascular disease, stroke, or TIA

Table S25: Concept codes used to identify if a patient was diagnosed with cerebrovascular disease, stroke, or TIA.

| CodeSystem | ConceptCode | ConceptName |
|---|---|---|
| ICD10CM | G45.0 | Vertebro-basilar artery syndrome |

Continued on next page

**TableS25 – continued from previous page**

| CodeSystem | ConceptCode | ConceptName |
|---|---|---|
| ICD10CM | G45.1 | Carotid artery syndrome (hemispheric) |
| ICD10CM | G45.2 | Multiple and bilateral precerebral artery syndromes |
| ICD10CM | G45.8 | Other transient cerebral ischemic attacks and related syndromes |
| ICD10CM | G45.9 | Transient cerebral ischemic attack, unspecified |
| ICD10CM | G46.0 | Middle cerebral artery syndrome |
| ICD10CM | G46.1 | Anterior cerebral artery syndrome |
| ICD10CM | G46.2 | Posterior cerebral artery syndrome |
| ICD10CM | G46.3 | Brain stem stroke syndrome |
| ICD10CM | G46.4 | Cerebellar stroke syndrome |
| ICD10CM | G46.5 | Pure motor lacunar syndrome |
| ICD10CM | G46.6 | Pure sensory lacunar syndrome |
| ICD10CM | G46.7 | Other lacunar syndromes |
| ICD10CM | G46.8 | Other vascular syndromes of brain in cerebrovascular diseases |
| ICD10CM | I63.00 | Cerebral infarction due to thrombosis of unspecified precerebral artery |
| ICD10CM | I63.011 | Cerebral infarction due to thrombosis of right vertebral artery |
| ICD10CM | I63.012 | Cerebral infarction due to thrombosis of left vertebral artery |
| ICD10CM | I63.013 | Cerebral infarction due to thrombosis of bilateral vertebral arteries |
| ICD10CM | I63.019 | Cerebral infarction due to thrombosis of unspecified vertebral artery |
| ICD10CM | I63.02 | Cerebral infarction due to thrombosis of basilar artery |
| ICD10CM | I63.031 | Cerebral infarction due to thrombosis of right carotid artery |
| ICD10CM | I63.032 | Cerebral infarction due to thrombosis of left carotid artery |
| ICD10CM | I63.033 | Cerebral infarction due to thrombosis of bilateral carotid arteries |
| ICD10CM | I63.039 | Cerebral infarction due to thrombosis of unspecified carotid artery |
| ICD10CM | I63.09 | Cerebral infarction due to thrombosis of other precerebral artery |
| ICD10CM | I63.10 | Cerebral infarction due to embolism of unspecified precerebral artery |
| ICD10CM | I63.111 | Cerebral infarction due to embolism of right vertebral artery |
| ICD10CM | I63.112 | Cerebral infarction due to embolism of left vertebral artery |
| ICD10CM | I63.113 | Cerebral infarction due to embolism of bilateral vertebral arteries |
| ICD10CM | I63.119 | Cerebral infarction due to embolism of unspecified vertebral artery |
| ICD10CM | I63.12 | Cerebral infarction due to embolism of basilar artery |
| ICD10CM | I63.131 | Cerebral infarction due to embolism of right carotid artery |
| ICD10CM | I63.132 | Cerebral infarction due to embolism of left carotid artery |
| ICD10CM | I63.133 | Cerebral infarction due to embolism of bilateral carotid arteries |
| ICD10CM | I63.139 | Cerebral infarction due to embolism of unspecified carotid artery |
| ICD10CM | I63.19 | Cerebral infarction due to embolism of other precerebral artery |
| ICD10CM | I63.20 | Cerebral infarction due to unspecified occlusion or stenosis of unspecified precerebral arteries |
| ICD10CM | I63.211 | Cerebral infarction due to unspecified occlusion or stenosis of right vertebral artery |
| ICD10CM | I63.212 | Cerebral infarction due to unspecified occlusion or stenosis of left vertebral artery |

Continued on next page

**TableS25 – continued from previous page**

| CodeSystem | ConceptCode | ConceptName |
|---|---|---|
| ICD10CM | I63.213 | Cerebral infarction due to unspecified occlusion or stenosis of bilateral vertebral arteries |
| ICD10CM | I63.219 | Cerebral infarction due to unspecified occlusion or stenosis of unspecified vertebral artery |
| ICD10CM | I63.22 | Cerebral infarction due to unspecified occlusion or stenosis of basilar artery |
| ICD10CM | I63.231 | Cerebral infarction due to unspecified occlusion or stenosis of right carotid arteries |
| ICD10CM | I63.232 | Cerebral infarction due to unspecified occlusion or stenosis of left carotid arteries |
| ICD10CM | I63.233 | Cerebral infarction due to unspecified occlusion or stenosis of bilateral carotid arteries |
| ICD10CM | I63.239 | Cerebral infarction due to unspecified occlusion or stenosis of unspecified carotid artery |
| ICD10CM | I63.29 | Cerebral infarction due to unspecified occlusion or stenosis of other precerebral arteries |
| ICD10CM | I63.30 | Cerebral infarction due to thrombosis of unspecified cerebral artery |
| ICD10CM | I63.311 | Cerebral infarction due to thrombosis of right middle cerebral artery |
| ICD10CM | I63.312 | Cerebral infarction due to thrombosis of left middle cerebral artery |
| ICD10CM | I63.313 | Cerebral infarction due to thrombosis of bilateral middle cerebral arteries |
| ICD10CM | I63.319 | Cerebral infarction due to thrombosis of unspecified middle cerebral artery |
| ICD10CM | I63.321 | Cerebral infarction due to thrombosis of right anterior cerebral artery |
| ICD10CM | I63.322 | Cerebral infarction due to thrombosis of left anterior cerebral artery |
| ICD10CM | I63.323 | Cerebral infarction due to thrombosis of bilateral anterior cerebral arteries |
| ICD10CM | I63.329 | Cerebral infarction due to thrombosis of unspecified anterior cerebral artery |
| ICD10CM | I63.331 | Cerebral infarction due to thrombosis of right posterior cerebral artery |
| ICD10CM | I63.332 | Cerebral infarction due to thrombosis of left posterior cerebral artery |
| ICD10CM | I63.333 | Cerebral infarction due to thrombosis of bilateral posterior cerebral arteries |
| ICD10CM | I63.339 | Cerebral infarction due to thrombosis of unspecified posterior cerebral artery |
| ICD10CM | I63.341 | Cerebral infarction due to thrombosis of right cerebellar artery |
| ICD10CM | I63.342 | Cerebral infarction due to thrombosis of left cerebellar artery |
| ICD10CM | I63.343 | Cerebral infarction due to thrombosis of bilateral cerebellar arteries |

Continued on next page

**TableS25 – continued from previous page**

| CodeSystem | ConceptCode | ConceptName |
|---|---|---|
| ICD10CM | I63.349 | Cerebral infarction due to thrombosis of unspecified cerebellar artery |
| ICD10CM | I63.39 | Cerebral infarction due to thrombosis of other cerebral artery |
| ICD10CM | I63.40 | Cerebral infarction due to embolism of unspecified cerebral artery |
| ICD10CM | I63.411 | Cerebral infarction due to embolism of right middle cerebral artery |
| ICD10CM | I63.412 | Cerebral infarction due to embolism of left middle cerebral artery |
| ICD10CM | I63.413 | Cerebral infarction due to embolism of bilateral middle cerebral arteries |
| ICD10CM | I63.419 | Cerebral infarction due to embolism of unspecified middle cerebral artery |
| ICD10CM | I63.421 | Cerebral infarction due to embolism of right anterior cerebral artery |
| ICD10CM | I63.422 | Cerebral infarction due to embolism of left anterior cerebral artery |
| ICD10CM | I63.423 | Cerebral infarction due to embolism of bilateral anterior cerebral arteries |
| ICD10CM | I63.429 | Cerebral infarction due to embolism of unspecified anterior cerebral artery |
| ICD10CM | I63.431 | Cerebral infarction due to embolism of right posterior cerebral artery |
| ICD10CM | I63.432 | Cerebral infarction due to embolism of left posterior cerebral artery |
| ICD10CM | I63.433 | Cerebral infarction due to embolism of bilateral posterior cerebral arteries |
| ICD10CM | I63.439 | Cerebral infarction due to embolism of unspecified posterior cerebral artery |
| ICD10CM | I63.441 | Cerebral infarction due to embolism of right cerebellar artery |
| ICD10CM | I63.442 | Cerebral infarction due to embolism of left cerebellar artery |
| ICD10CM | I63.443 | Cerebral infarction due to embolism of bilateral cerebellar arteries |
| ICD10CM | I63.449 | Cerebral infarction due to embolism of unspecified cerebellar artery |
| ICD10CM | I63.49 | Cerebral infarction due to embolism of other cerebral artery |
| ICD10CM | I63.50 | Cerebral infarction due to unspecified occlusion or stenosis of unspecified cerebral artery |
| ICD10CM | I63.511 | Cerebral infarction due to unspecified occlusion or stenosis of right middle cerebral artery |
| ICD10CM | I63.512 | Cerebral infarction due to unspecified occlusion or stenosis of left middle cerebral artery |
| ICD10CM | I63.513 | Cerebral infarction due to unspecified occlusion or stenosis of bilateral middle cerebral arteries |
| ICD10CM | I63.519 | Cerebral infarction due to unspecified occlusion or stenosis of unspecified middle cerebral artery |
| ICD10CM | I63.521 | Cerebral infarction due to unspecified occlusion or stenosis of right anterior cerebral artery |

Continued on next page

**TableS25 – continued from previous page**

| CodeSystem | ConceptCode | ConceptName |
|---|---|---|
| ICD10CM | I63.522 | Cerebral infarction due to unspecified occlusion or stenosis of left anterior cerebral artery |
| ICD10CM | I63.523 | Cerebral infarction due to unspecified occlusion or stenosis of bilateral anterior cerebral arteries |
| ICD10CM | I63.529 | Cerebral infarction due to unspecified occlusion or stenosis of unspecified anterior cerebral artery |
| ICD10CM | I63.531 | Cerebral infarction due to unspecified occlusion or stenosis of right posterior cerebral artery |
| ICD10CM | I63.532 | Cerebral infarction due to unspecified occlusion or stenosis of left posterior cerebral artery |
| ICD10CM | I63.533 | Cerebral infarction due to unspecified occlusion or stenosis of bilateral posterior cerebral arteries |
| ICD10CM | I63.539 | Cerebral infarction due to unspecified occlusion or stenosis of unspecified posterior cerebral artery |
| ICD10CM | I63.541 | Cerebral infarction due to unspecified occlusion or stenosis of right cerebellar artery |
| ICD10CM | I63.542 | Cerebral infarction due to unspecified occlusion or stenosis of left cerebellar artery |
| ICD10CM | I63.543 | Cerebral infarction due to unspecified occlusion or stenosis of bilateral cerebellar arteries |
| ICD10CM | I63.549 | Cerebral infarction due to unspecified occlusion or stenosis of unspecified cerebellar artery |
| ICD10CM | I63.59 | Cerebral infarction due to unspecified occlusion or stenosis of other cerebral artery |
| ICD10CM | I63.6 | Cerebral infarction due to cerebral venous thrombosis, nonpyogenic |
| ICD10CM | I63.81 | Other cerebral infarction due to occlusion or stenosis of small artery |
| ICD10CM | I63.89 | Other cerebral infarction |
| ICD10CM | I63.9 | Cerebral infarction, unspecified |
| ICD10CM | I69.00 | Unspecified sequelae of nontraumatic subarachnoid hemorrhage |
| ICD10CM | I69.010 | Attention and concentration deficit following nontraumatic subarachnoid hemorrhage |
| ICD10CM | I69.011 | Memory deficit following nontraumatic subarachnoid hemorrhage |
| ICD10CM | I69.012 | Visuospatial deficit and spatial neglect following nontraumatic subarachnoid hemorrhage |
| ICD10CM | I69.013 | Psychomotor deficit following nontraumatic subarachnoid hemorrhage |
| ICD10CM | I69.014 | Frontal lobe and executive function deficit following nontraumatic subarachnoid hemorrhage |
| ICD10CM | I69.015 | Cognitive social or emotional deficit following nontraumatic subarachnoid hemorrhage |
| ICD10CM | I69.018 | Other symptoms and signs involving cognitive functions following nontraumatic subarachnoid hemorrhage |
| ICD10CM | I69.019 | Unspecified symptoms and signs involving cognitive functions following nontraumatic subarachnoid hemorrhage |

Continued on next page

**TableS25 – continued from previous page**

| CodeSystem | ConceptCode | ConceptName |
|---|---|---|
| ICD10CM | I69.020 | Aphasia following nontraumatic subarachnoid hemorrhage |
| ICD10CM | I69.021 | Dysphasia following nontraumatic subarachnoid hemorrhage |
| ICD10CM | I69.022 | Dysarthria following nontraumatic subarachnoid hemorrhage |
| ICD10CM | I69.023 | Fluency disorder following nontraumatic subarachnoid hemorrhage |
| ICD10CM | I69.028 | Other speech and language deficits following nontraumatic subarachnoid hemorrhage |
| ICD10CM | I69.031 | Monoplegia of upper limb following nontraumatic subarachnoid hemorrhage affecting right dominant side |
| ICD10CM | I69.032 | Monoplegia of upper limb following nontraumatic subarachnoid hemorrhage affecting left dominant side |
| ICD10CM | I69.033 | Monoplegia of upper limb following nontraumatic subarachnoid hemorrhage affecting right non-dominant side |
| ICD10CM | I69.034 | Monoplegia of upper limb following nontraumatic subarachnoid hemorrhage affecting left non-dominant side |
| ICD10CM | I69.039 | Monoplegia of upper limb following nontraumatic subarachnoid hemorrhage affecting unspecified side |
| ICD10CM | I69.041 | Monoplegia of lower limb following nontraumatic subarachnoid hemorrhage affecting right dominant side |
| ICD10CM | I69.042 | Monoplegia of lower limb following nontraumatic subarachnoid hemorrhage affecting left dominant side |
| ICD10CM | I69.043 | Monoplegia of lower limb following nontraumatic subarachnoid hemorrhage affecting right non-dominant side |
| ICD10CM | I69.044 | Monoplegia of lower limb following nontraumatic subarachnoid hemorrhage affecting left non-dominant side |
| ICD10CM | I69.049 | Monoplegia of lower limb following nontraumatic subarachnoid hemorrhage affecting unspecified side |
| ICD10CM | I69.051 | Hemiplegia and hemiparesis following nontraumatic subarachnoid hemorrhage affecting right dominant side |
| ICD10CM | I69.052 | Hemiplegia and hemiparesis following nontraumatic subarachnoid hemorrhage affecting left dominant side |
| ICD10CM | I69.053 | Hemiplegia and hemiparesis following nontraumatic subarachnoid hemorrhage affecting right non-dominant side |
| ICD10CM | I69.054 | Hemiplegia and hemiparesis following nontraumatic subarachnoid hemorrhage affecting left non-dominant side |
| ICD10CM | I69.059 | Hemiplegia and hemiparesis following nontraumatic subarachnoid hemorrhage affecting unspecified side |
| ICD10CM | I69.061 | Other paralytic syndrome following nontraumatic subarachnoid hemorrhage affecting right dominant side |
| ICD10CM | I69.323 | Fluency disorder following cerebral infarction |
| ICD10CM | I69.062 | Other paralytic syndrome following nontraumatic subarachnoid hemorrhage affecting left dominant side |
| ICD10CM | I69.063 | Other paralytic syndrome following nontraumatic subarachnoid hemorrhage affecting right non-dominant side |
| ICD10CM | I69.064 | Other paralytic syndrome following nontraumatic subarachnoid hemorrhage affecting left non-dominant side |

Continued on next page

**TableS25 – continued from previous page**

| CodeSystem | ConceptCode | ConceptName |
|---|---|---|
| ICD10CM | I69.065 | Other paralytic syndrome following nontraumatic subarachnoid hemorrhage, bilateral |
| ICD10CM | I69.069 | Other paralytic syndrome following nontraumatic subarachnoid hemorrhage affecting unspecified side |
| ICD10CM | I69.090 | Apraxia following nontraumatic subarachnoid hemorrhage |
| ICD10CM | I69.091 | Dysphagia following nontraumatic subarachnoid hemorrhage |
| ICD10CM | I69.092 | Facial weakness following nontraumatic subarachnoid hemorrhage |
| ICD10CM | I69.093 | Ataxia following nontraumatic subarachnoid hemorrhage |
| ICD10CM | I69.098 | Other sequelae following nontraumatic subarachnoid hemorrhage |
| ICD10CM | I69.10 | Unspecified sequelae of nontraumatic intracerebral hemorrhage |
| ICD10CM | I69.110 | Attention and concentration deficit following nontraumatic intracerebral hemorrhage |
| ICD10CM | I69.111 | Memory deficit following nontraumatic intracerebral hemorrhage |
| ICD10CM | I69.112 | Visuospatial deficit and spatial neglect following nontraumatic intracerebral hemorrhage |
| ICD10CM | I69.113 | Psychomotor deficit following nontraumatic intracerebral hemorrhage |
| ICD10CM | I69.114 | Frontal lobe and executive function deficit following nontraumatic intracerebral hemorrhage |
| ICD10CM | I69.115 | Cognitive social or emotional deficit following nontraumatic intracerebral hemorrhage |
| ICD10CM | I69.118 | Other symptoms and signs involving cognitive functions following nontraumatic intracerebral hemorrhage |
| ICD10CM | I69.119 | Unspecified symptoms and signs involving cognitive functions following nontraumatic intracerebral hemorrhage |
| ICD10CM | I69.120 | Aphasia following nontraumatic intracerebral hemorrhage |
| ICD10CM | I69.121 | Dysphasia following nontraumatic intracerebral hemorrhage |
| ICD10CM | I69.122 | Dysarthria following nontraumatic intracerebral hemorrhage |
| ICD10CM | I69.123 | Fluency disorder following nontraumatic intracerebral hemorrhage |
| ICD10CM | I69.128 | Other speech and language deficits following nontraumatic intracerebral hemorrhage |
| ICD10CM | I69.131 | Monoplegia of upper limb following nontraumatic intracerebral hemorrhage affecting right dominant side |
| ICD10CM | I69.132 | Monoplegia of upper limb following nontraumatic intracerebral hemorrhage affecting left dominant side |
| ICD10CM | I69.133 | Monoplegia of upper limb following nontraumatic intracerebral hemorrhage affecting right non-dominant side |
| ICD10CM | I69.134 | Monoplegia of upper limb following nontraumatic intracerebral hemorrhage affecting left non-dominant side |
| ICD10CM | I69.139 | Monoplegia of upper limb following nontraumatic intracerebral hemorrhage affecting unspecified side |
| ICD10CM | I69.141 | Monoplegia of lower limb following nontraumatic intracerebral hemorrhage affecting right dominant side |
| ICD10CM | I69.142 | Monoplegia of lower limb following nontraumatic intracerebral hemorrhage affecting left dominant side |

Continued on next page

**TableS25 – continued from previous page**

| CodeSystem | ConceptCode | ConceptName |
|---|---|---|
| ICD10CM | I69.143 | Monoplegia of lower limb following nontraumatic intracerebral hemorrhage affecting right non-dominant side |
| ICD10CM | I69.144 | Monoplegia of lower limb following nontraumatic intracerebral hemorrhage affecting left non-dominant side |
| ICD10CM | I69.149 | Monoplegia of lower limb following nontraumatic intracerebral hemorrhage affecting unspecified side |
| ICD10CM | I69.151 | Hemiplegia and hemiparesis following nontraumatic intracerebral hemorrhage affecting right dominant side |
| ICD10CM | I69.152 | Hemiplegia and hemiparesis following nontraumatic intracerebral hemorrhage affecting left dominant side |
| ICD10CM | I69.153 | Hemiplegia and hemiparesis following nontraumatic intracerebral hemorrhage affecting right non-dominant side |
| ICD10CM | I69.154 | Hemiplegia and hemiparesis following nontraumatic intracerebral hemorrhage affecting left non-dominant side |
| ICD10CM | I69.159 | Hemiplegia and hemiparesis following nontraumatic intracerebral hemorrhage affecting unspecified side |
| ICD10CM | I69.161 | Other paralytic syndrome following nontraumatic intracerebral hemorrhage affecting right dominant side |
| ICD10CM | I69.162 | Other paralytic syndrome following nontraumatic intracerebral hemorrhage affecting left dominant side |
| ICD10CM | I69.163 | Other paralytic syndrome following nontraumatic intracerebral hemorrhage affecting right non-dominant side |
| ICD10CM | I69.164 | Other paralytic syndrome following nontraumatic intracerebral hemorrhage affecting left non-dominant side |
| ICD10CM | I69.165 | Other paralytic syndrome following nontraumatic intracerebral hemorrhage, bilateral |
| ICD10CM | I69.169 | Other paralytic syndrome following nontraumatic intracerebral hemorrhage affecting unspecified side |
| ICD10CM | I69.190 | Apraxia following nontraumatic intracerebral hemorrhage |
| ICD10CM | I69.191 | Dysphagia following nontraumatic intracerebral hemorrhage |
| ICD10CM | I69.192 | Facial weakness following nontraumatic intracerebral hemorrhage |
| ICD10CM | I69.193 | Ataxia following nontraumatic intracerebral hemorrhage |
| ICD10CM | I69.198 | Other sequelae of nontraumatic intracerebral hemorrhage |
| ICD10CM | I69.20 | Unspecified sequelae of other nontraumatic intracranial hemorrhage |
| ICD10CM | I69.210 | Attention and concentration deficit following other nontraumatic intracranial hemorrhage |
| ICD10CM | I69.211 | Memory deficit following other nontraumatic intracranial hemorrhage |
| ICD10CM | I69.212 | Visuospatial deficit and spatial neglect following other nontraumatic intracranial hemorrhage |
| ICD10CM | I69.213 | Psychomotor deficit following other nontraumatic intracranial hemorrhage |
| ICD10CM | I69.214 | Frontal lobe and executive function deficit following other nontraumatic intracranial hemorrhage |

Continued on next page

**TableS25 – continued from previous page**

| CodeSystem | ConceptCode | ConceptName |
|---|---|---|
| ICD10CM | I69.215 | Cognitive social or emotional deficit following other nontraumatic intracranial hemorrhage |
| ICD10CM | I69.218 | Other symptoms and signs involving cognitive functions following other nontraumatic intracranial hemorrhage |
| ICD10CM | I69.328 | Other speech and language deficits following cerebral infarction |
| ICD10CM | I69.219 | Unspecified symptoms and signs involving cognitive functions following other nontraumatic intracranial hemorrhage |
| ICD10CM | I69.220 | Aphasia following other nontraumatic intracranial hemorrhage |
| ICD10CM | I69.221 | Dysphasia following other nontraumatic intracranial hemorrhage |
| ICD10CM | I69.222 | Dysarthria following other nontraumatic intracranial hemorrhage |
| ICD10CM | I69.223 | Fluency disorder following other nontraumatic intracranial hemorrhage |
| ICD10CM | I69.228 | Other speech and language deficits following other nontraumatic intracranial hemorrhage |
| ICD10CM | I69.231 | Monoplegia of upper limb following other nontraumatic intracranial hemorrhage affecting right dominant side |
| ICD10CM | I69.232 | Monoplegia of upper limb following other nontraumatic intracranial hemorrhage affecting left dominant side |
| ICD10CM | I69.233 | Monoplegia of upper limb following other nontraumatic intracranial hemorrhage affecting right non-dominant side |
| ICD10CM | I69.234 | Monoplegia of upper limb following other nontraumatic intracranial hemorrhage affecting left non-dominant side |
| ICD10CM | I69.239 | Monoplegia of upper limb following other nontraumatic intracranial hemorrhage affecting unspecified side |
| ICD10CM | I69.241 | Monoplegia of lower limb following other nontraumatic intracranial hemorrhage affecting right dominant side |
| ICD10CM | I69.242 | Monoplegia of lower limb following other nontraumatic intracranial hemorrhage affecting left dominant side |
| ICD10CM | I69.243 | Monoplegia of lower limb following other nontraumatic intracranial hemorrhage affecting right non-dominant side |
| ICD10CM | I69.244 | Monoplegia of lower limb following other nontraumatic intracranial hemorrhage affecting left non-dominant side |
| ICD10CM | I69.249 | Monoplegia of lower limb following other nontraumatic intracranial hemorrhage affecting unspecified side |
| ICD10CM | I69.251 | Hemiplegia and hemiparesis following other nontraumatic intracranial hemorrhage affecting right dominant side |
| ICD10CM | I69.252 | Hemiplegia and hemiparesis following other nontraumatic intracranial hemorrhage affecting left dominant side |
| ICD10CM | I69.253 | Hemiplegia and hemiparesis following other nontraumatic intracranial hemorrhage affecting right non-dominant side |
| ICD10CM | I69.254 | Hemiplegia and hemiparesis following other nontraumatic intracranial hemorrhage affecting left non-dominant side |
| ICD10CM | I69.259 | Hemiplegia and hemiparesis following other nontraumatic intracranial hemorrhage affecting unspecified side |
| ICD10CM | I69.261 | Other paralytic syndrome following other nontraumatic intracranial hemorrhage affecting right dominant side |

Continued on next page

**TableS25 – continued from previous page**

| CodeSystem | ConceptCode | ConceptName |
|---|---|---|
| ICD10CM | I69.262 | Other paralytic syndrome following other nontraumatic intracranial hemorrhage affecting left dominant side |
| ICD10CM | I69.263 | Other paralytic syndrome following other nontraumatic intracranial hemorrhage affecting right non-dominant side |
| ICD10CM | I69.264 | Other paralytic syndrome following other nontraumatic intracranial hemorrhage affecting left non-dominant side |
| ICD10CM | I69.265 | Other paralytic syndrome following other nontraumatic intracranial hemorrhage, bilateral |
| ICD10CM | I69.269 | Other paralytic syndrome following other nontraumatic intracranial hemorrhage affecting unspecified side |
| ICD10CM | I69.290 | Apraxia following other nontraumatic intracranial hemorrhage |
| ICD10CM | I69.291 | Dysphagia following other nontraumatic intracranial hemorrhage |
| ICD10CM | I69.292 | Facial weakness following other nontraumatic intracranial hemorrhage |
| ICD10CM | I69.293 | Ataxia following other nontraumatic intracranial hemorrhage |
| ICD10CM | I69.298 | Other sequelae of other nontraumatic intracranial hemorrhage |
| ICD10CM | I69.30 | Unspecified sequelae of cerebral infarction |
| ICD10CM | I69.310 | Attention and concentration deficit following cerebral infarction |
| ICD10CM | I69.311 | Memory deficit following cerebral infarction |
| ICD10CM | I69.312 | Visuospatial deficit and spatial neglect following cerebral infarction |
| ICD10CM | I69.313 | Psychomotor deficit following cerebral infarction |
| ICD10CM | I69.314 | Frontal lobe and executive function deficit following cerebral infarction |
| ICD10CM | I69.315 | Cognitive social or emotional deficit following cerebral infarction |
| ICD10CM | I69.318 | Other symptoms and signs involving cognitive functions following cerebral infarction |
| ICD10CM | I69.319 | Unspecified symptoms and signs involving cognitive functions following cerebral infarction |
| ICD10CM | I69.320 | Aphasia following cerebral infarction |
| ICD10CM | I69.321 | Dysphasia following cerebral infarction |
| ICD10CM | I69.322 | Dysarthria following cerebral infarction |
| ICD10CM | I69.331 | Monoplegia of upper limb following cerebral infarction affecting right dominant side |
| ICD10CM | I69.332 | Monoplegia of upper limb following cerebral infarction affecting left dominant side |
| ICD10CM | I69.333 | Monoplegia of upper limb following cerebral infarction affecting right non-dominant side |
| ICD10CM | I69.334 | Monoplegia of upper limb following cerebral infarction affecting left non-dominant side |
| ICD10CM | I69.339 | Monoplegia of upper limb following cerebral infarction affecting unspecified side |
| ICD10CM | I69.341 | Monoplegia of lower limb following cerebral infarction affecting right dominant side |
| ICD10CM | I69.342 | Monoplegia of lower limb following cerebral infarction affecting left dominant side |

Continued on next page

**TableS25 – continued from previous page**

| CodeSystem | ConceptCode | ConceptName |
|---|---|---|
| ICD10CM | I69.343 | Monoplegia of lower limb following cerebral infarction affecting right non-dominant side |
| ICD10CM | I69.344 | Monoplegia of lower limb following cerebral infarction affecting left non-dominant side |
| ICD10CM | I69.349 | Monoplegia of lower limb following cerebral infarction affecting unspecified side |
| ICD10CM | I69.351 | Hemiplegia and hemiparesis following cerebral infarction affecting right dominant side |
| ICD10CM | I69.352 | Hemiplegia and hemiparesis following cerebral infarction affecting left dominant side |
| ICD10CM | I69.353 | Hemiplegia and hemiparesis following cerebral infarction affecting right non-dominant side |
| ICD10CM | I69.354 | Hemiplegia and hemiparesis following cerebral infarction affecting left non-dominant side |
| ICD10CM | I69.359 | Hemiplegia and hemiparesis following cerebral infarction affecting unspecified side |
| ICD10CM | I69.361 | Other paralytic syndrome following cerebral infarction affecting right dominant side |
| ICD10CM | I69.362 | Other paralytic syndrome following cerebral infarction affecting left dominant side |
| ICD10CM | I69.363 | Other paralytic syndrome following cerebral infarction affecting right non-dominant side |
| ICD10CM | I69.364 | Other paralytic syndrome following cerebral infarction affecting left non-dominant side |
| ICD10CM | I69.365 | Other paralytic syndrome following cerebral infarction, bilateral |
| ICD10CM | I69.369 | Other paralytic syndrome following cerebral infarction affecting unspecified side |
| ICD10CM | I69.390 | Apraxia following cerebral infarction |
| ICD10CM | I69.391 | Dysphagia following cerebral infarction |
| ICD10CM | I69.392 | Facial weakness following cerebral infarction |
| ICD10CM | I69.393 | Ataxia following cerebral infarction |
| ICD10CM | I69.398 | Other sequelae of cerebral infarction |
| ICD10CM | I69.80 | Unspecified sequelae of other cerebrovascular disease |
| ICD10CM | I69.810 | Attention and concentration deficit following other cerebrovascular disease |
| ICD10CM | I69.811 | Memory deficit following other cerebrovascular disease |
| ICD10CM | I69.812 | Visuospatial deficit and spatial neglect following other cerebrovascular disease |
| ICD10CM | I69.813 | Psychomotor deficit following other cerebrovascular disease |
| ICD10CM | I69.814 | Frontal lobe and executive function deficit following other cerebrovascular disease |
| ICD10CM | I69.815 | Cognitive social or emotional deficit following other cerebrovascular disease |
| ICD10CM | I69.818 | Other symptoms and signs involving cognitive functions following other cerebrovascular disease |
| ICD10CM | I69.819 | Unspecified symptoms and signs involving cognitive functions following other cerebrovascular disease |

Continued on next page

**TableS25 – continued from previous page**

| CodeSystem | ConceptCode | ConceptName |
|---|---|---|
| ICD10CM | I69.820 | Aphasia following other cerebrovascular disease |
| ICD10CM | I69.821 | Dysphasia following other cerebrovascular disease |
| ICD10CM | I69.822 | Dysarthria following other cerebrovascular disease |
| ICD10CM | I69.831 | Monoplegia of upper limb following other cerebrovascular disease affecting right dominant side |
| ICD10CM | I69.832 | Monoplegia of upper limb following other cerebrovascular disease affecting left dominant side |
| ICD10CM | I69.833 | Monoplegia of upper limb following other cerebrovascular disease affecting right non-dominant side |
| ICD10CM | I69.834 | Monoplegia of upper limb following other cerebrovascular disease affecting left non-dominant side |
| ICD10CM | I69.839 | Monoplegia of upper limb following other cerebrovascular disease affecting unspecified side |
| ICD10CM | I69.841 | Monoplegia of lower limb following other cerebrovascular disease affecting right dominant side |
| ICD10CM | I69.842 | Monoplegia of lower limb following other cerebrovascular disease affecting left dominant side |
| ICD10CM | I69.843 | Monoplegia of lower limb following other cerebrovascular disease affecting right non-dominant side |
| ICD10CM | I69.844 | Monoplegia of lower limb following other cerebrovascular disease affecting left non-dominant side |
| ICD10CM | I69.849 | Monoplegia of lower limb following other cerebrovascular disease affecting unspecified side |
| ICD10CM | I69.863 | Other paralytic syndrome following other cerebrovascular disease affecting right non-dominant side |
| ICD10CM | I69.864 | Other paralytic syndrome following other cerebrovascular disease affecting left non-dominant side |
| ICD10CM | I69.865 | Other paralytic syndrome following other cerebrovascular disease, bilateral |
| ICD10CM | I69.869 | Other paralytic syndrome following other cerebrovascular disease affecting unspecified side |
| ICD10CM | I69.890 | Apraxia following other cerebrovascular disease |
| ICD10CM | I69.892 | Facial weakness following other cerebrovascular disease |
| ICD10CM | I69.893 | Ataxia following other cerebrovascular disease |
| ICD10CM | I69.898 | Other sequelae of other cerebrovascular disease |
| ICD10CM | I69.90 | Unspecified sequelae of unspecified cerebrovascular disease |
| ICD10CM | I69.910 | Attention and concentration deficit following unspecified cerebrovascular disease |
| ICD10CM | I69.911 | Memory deficit following unspecified cerebrovascular disease |
| ICD10CM | I69.912 | Visuospatial deficit and spatial neglect following unspecified cerebrovascular disease |
| ICD10CM | I69.913 | Psychomotor deficit following unspecified cerebrovascular disease |
| ICD10CM | I69.914 | Frontal lobe and executive function deficit following unspecified cerebrovascular disease |
| ICD10CM | I69.915 | Cognitive social or emotional deficit following unspecified cerebrovascular disease |

Continued on next page

**TableS25 – continued from previous page**

| CodeSystem | ConceptCode | ConceptName |
|---|---|---|
| ICD10CM | I69.918 | Other symptoms and signs involving cognitive functions following unspecified cerebrovascular disease |
| ICD10CM | I69.919 | Unspecified symptoms and signs involving cognitive functions following unspecified cerebrovascular disease |
| ICD10CM | I69.920 | Aphasia following unspecified cerebrovascular disease |
| ICD10CM | I69.921 | Dysphasia following unspecified cerebrovascular disease |
| ICD10CM | I69.922 | Dysarthria following unspecified cerebrovascular disease |
| ICD10CM | I69.923 | Fluency disorder following unspecified cerebrovascular disease |
| ICD10CM | I69.928 | Other speech and language deficits following unspecified cerebrovascular disease |
| ICD10CM | I69.931 | Monoplegia of upper limb following unspecified cerebrovascular disease affecting right dominant side |
| ICD10CM | I69.932 | Monoplegia of upper limb following unspecified cerebrovascular disease affecting left dominant side |
| ICD10CM | I69.933 | Monoplegia of upper limb following unspecified cerebrovascular disease affecting right non-dominant side |
| ICD10CM | I69.934 | Monoplegia of upper limb following unspecified cerebrovascular disease affecting left non-dominant side |
| ICD10CM | I69.939 | Monoplegia of upper limb following unspecified cerebrovascular disease affecting unspecified side |
| ICD10CM | I69.941 | Monoplegia of lower limb following unspecified cerebrovascular disease affecting right dominant side |
| ICD10CM | I69.942 | Monoplegia of lower limb following unspecified cerebrovascular disease affecting left dominant side |
| ICD10CM | I69.943 | Monoplegia of lower limb following unspecified cerebrovascular disease affecting right non-dominant side |
| ICD10CM | I69.944 | Monoplegia of lower limb following unspecified cerebrovascular disease affecting left non-dominant side |
| ICD10CM | I69.949 | Monoplegia of lower limb following unspecified cerebrovascular disease affecting unspecified side |
| ICD10CM | I69.990 | Apraxia following unspecified cerebrovascular disease |
| ICD10CM | I69.991 | Dysphagia following unspecified cerebrovascular disease |
| ICD10CM | I69.992 | Facial weakness following unspecified cerebrovascular disease |
| ICD10CM | I69.993 | Ataxia following unspecified cerebrovascular disease |
| ICD10CM | I69.998 | Other sequelae following unspecified cerebrovascular disease |
| ICD10CM | Z86.73 | Personal history of transient ischemic attack (TIA), and cerebral infarction without residual deficits |
| ICD9CM | 433.00 | Occlusion and stenosis of basilar artery without mention of cerebral infarction |
| ICD9CM | 433.01 | Occlusion and stenosis of basilar artery with cerebral infarction |
| ICD9CM | 433.10 | Occlusion and stenosis of carotid artery without mention of cerebral infarction |
| ICD9CM | 433.11 | Occlusion and stenosis of carotid artery with cerebral infarction |
| ICD9CM | 433.20 | Occlusion and stenosis of vertebral artery without mention of cerebral infarction |
| ICD9CM | 433.21 | Occlusion and stenosis of vertebral artery with cerebral infarction |

Continued on next page

**TableS25 – continued from previous page**

| CodeSystem | ConceptCode | ConceptName |
|---|---|---|
| ICD9CM | 435.8 | Other specified transient cerebral ischemias |
| ICD9CM | 433.30 | Occlusion and stenosis of multiple and bilateral precerebral arteries without mention of cerebral infarction |
| ICD9CM | 433.31 | Occlusion and stenosis of multiple and bilateral precerebral arteries with cerebral infarction |
| ICD9CM | 433.80 | Occlusion and stenosis of other specified precerebral artery without mention of cerebral infarction |
| ICD9CM | 433.81 | Occlusion and stenosis of other specified precerebral artery with cerebral infarction |
| ICD9CM | 433.90 | Occlusion and stenosis of unspecified precerebral artery without mention of cerebral infarction |
| ICD9CM | 433.91 | Occlusion and stenosis of unspecified precerebral artery with cerebral infarction |
| ICD9CM | 434.00 | Cerebral thrombosis without mention of cerebral infarction |
| ICD9CM | 434.01 | Cerebral thrombosis with cerebral infarction |
| ICD9CM | 434.10 | Cerebral embolism without mention of cerebral infarction |
| ICD9CM | 434.11 | Cerebral embolism with cerebral infarction |
| ICD9CM | 434.90 | Cerebral artery occlusion, unspecified without mention of cerebral infarction |
| ICD9CM | 434.91 | Cerebral artery occlusion, unspecified with cerebral infarction |
| ICD9CM | 435.0 | Basilar artery syndrome |
| ICD9CM | 435.1 | Vertebral artery syndrome |
| ICD9CM | 435.3 | Vertebrobasilar artery syndrome |
| ICD9CM | 435.9 | Unspecified transient cerebral ischemia |
| ICD9CM | 436 | Acute, but ill-defined, cerebrovascular disease |
| ICD9CM | 437.1 | Other generalized ischemic cerebrovascular disease |
| ICD9CM | 438.0 | Late effects of cerebrovascular disease, cognitive deficits |
| ICD9CM | 438.10 | Late effects of cerebrovascular disease, speech and language deficit, unspecified |
| ICD9CM | 438.11 | Late effects of cerebrovascular disease, aphasia |
| ICD9CM | 438.12 | Late effects of cerebrovascular disease, dysphasia |
| ICD9CM | 438.13 | Late effects of cerebrovascular disease, dysarthria |
| ICD9CM | 438.14 | Late effects of cerebrovascular disease, fluency disorder |
| ICD9CM | 438.19 | Late effects of cerebrovascular disease, other speech and language deficits |
| ICD9CM | 438.20 | Late effects of cerebrovascular disease, hemiplegia affecting unspecified side |
| ICD9CM | 438.21 | Late effects of cerebrovascular disease, hemiplegia affecting dominant side |
| ICD9CM | 438.22 | Late effects of cerebrovascular disease, hemiplegia affecting nondominant side |
| ICD9CM | 438.30 | Late effects of cerebrovascular disease, monoplegia of upper limb affecting unspecified side |
| ICD9CM | 438.31 | Late effects of cerebrovascular disease, monoplegia of upper limb affecting dominant side |
| ICD9CM | 438.32 | Late effects of cerebrovascular disease, monoplegia of upper limb affecting nondominant side |

Continued on next page

**TableS25 – continued from previous page**

| CodeSystem | ConceptCode | ConceptName |
|---|---|---|
| ICD9CM | 438.40 | Late effects of cerebrovascular disease, monoplegia of lower limb affecting unspecified side |
| ICD9CM | 438.41 | Late effects of cerebrovascular disease, monoplegia of lower limb affecting dominant side |
| ICD9CM | 438.42 | Late effects of cerebrovascular disease, monoplegia of lower limb affecting nondominant side |
| ICD9CM | 438.50 | Late effects of cerebrovascular disease, other paralytic syndrome affecting unspecified side |
| ICD9CM | 438.51 | Late effects of cerebrovascular disease, other paralytic syndrome affecting dominant side |
| ICD9CM | 438.52 | Late effects of cerebrovascular disease, other paralytic syndrome affecting nondominant side |
| ICD9CM | 438.53 | Late effects of cerebrovascular disease, other paralytic syndrome, bilateral |
| ICD9CM | 438.6 | Late effects of cerebrovascular disease, alterations of sensations |
| ICD9CM | 438.7 | Late effects of cerebrovascular disease, disturbances of vision |
| ICD9CM | 438.81 | Other late effects of cerebrovascular disease, apraxia |
| ICD9CM | 438.82 | Other late effects of cerebrovascular disease, dysphagia |
| ICD9CM | 438.83 | Other late effects of cerebrovascular disease, facial weakness |
| ICD9CM | 438.84 | Other late effects of cerebrovascular disease, ataxia |
| ICD9CM | 438.85 | Other late effects of cerebrovascular disease, vertigo |
| ICD9CM | 438.89 | Other late effects of cerebrovascular disease |
| ICD9CM | 438.9 | Unspecified late effects of cerebrovascular disease |
| ICD9CM | V12.54 | Personal history of transient ischemic attack (TIA), and cerebral infarction without residual deficits |
| SNOMED CT | 266257000 | Transient cerebral ischemia |
| SNOMED CT | 275434003 | Stroke in the puerperium |
| SNOMED CT | 275526006 | History of cerebrovascular accident |
| SNOMED CT | 276219001 | Occipital cerebral infarction |
| SNOMED CT | 281240008 | Extension of cerebrovascular accident |
| SNOMED CT | 302909007 | Diffuse cerebrovascular disease |
| SNOMED CT | 307766002 | Left sided cerebral infarction |
| SNOMED CT | 307767006 | Right sided cerebral infarction |
| SNOMED CT | 371040005 | Thrombotic stroke |
| SNOMED CT | 371041009 | Embolic stroke |
| SNOMED CT | 373606000 | Occlusive stroke |
| SNOMED CT | 390936003 | Cerebral autosomal dominant arteriopathy with subcortical infarcts and leukoencephalopathy |
| SNOMED CT | 413102000 | Infarction of basal ganglia |
| SNOMED CT | 413758000 | Cardioembolic stroke |
| SNOMED CT | 422504002 | Ischemic stroke |
| SNOMED CT | 425642008 | Monoplegia of dominant lower limb as a late effect of cerebrovascular accident |
| SNOMED CT | 425882004 | Paralytic syndrome as late effect of stroke |
| SNOMED CT | 426033005 | Dysphagia as a late effect of cerebrovascular accident |
| SNOMED CT | 426788002 | Vertigo as late effect of stroke |
| SNOMED CT | 426983002 | Infarction of medulla oblongata |

Continued on next page

**TableS25 – continued from previous page**

| CodeSystem | ConceptCode | ConceptName |
|---|---|---|
| SNOMED CT | 427065003 | Monoplegia of dominant upper limb as a late effect of cerebrovascular accident |
| SNOMED CT | 427296003 | Thalamic stroke |
| SNOMED CT | 427432001 | Paralytic syndrome as late effect of thalamic stroke |
| SNOMED CT | 428668000 | Apraxia due to cerebrovascular accident |
| SNOMED CT | 430947007 | Paralytic syndrome of nondominant side as late effect of stroke |
| SNOMED CT | 430959006 | Paralytic syndrome of dominant side as late effect of stroke |
| SNOMED CT | 432504007 | Cerebral infarction |
| SNOMED CT | 433183000 | Neurogenic bladder as late effect of cerebrovascular accident |
| SNOMED CT | 441526008 | Infarct of cerebrum due to iatrogenic cerebrovascular accident |
| SNOMED CT | 441529001 | Dysphasia as late effect of cerebrovascular disease |
| SNOMED CT | 441630004 | Aphasia as late effect of cerebrovascular disease |
| SNOMED CT | 441735003 | Sensory disorder as a late effect of cerebrovascular disease |
| SNOMED CT | 441759008 | Abnormal vision as a late effect of cerebrovascular disease |
| SNOMED CT | 441887006 | Monoplegia of lower limb as late effect of cerebrovascular disease |
| SNOMED CT | 441894009 | Monoplegia of nondominant lower limb as a late effect of cerebrovascular accident |
| SNOMED CT | 441960006 | Speech and language deficit as late effect of cerebrovascular accident |
| SNOMED CT | 441991000 | Hemiparesis as late effect of cerebrovascular accident |
| SNOMED CT | 442024001 | Hemiplegia as late effect of cerebrovascular disease |
| SNOMED CT | 442097001 | Monoplegia of upper limb as late effect of cerebrovascular disease |
| SNOMED CT | 442181008 | Monoplegia of nondominant upper limb as a late effect of cerebrovascular accident |
| SNOMED CT | 442212003 | Residual cognitive deficit as late effect of cerebrovascular accident |
| SNOMED CT | 442617003 | Aphasia as late effect of cerebrovascular accident |
| SNOMED CT | 442668000 | Hemiplegia of nondominant side as late effect of cerebrovascular disease |
| SNOMED CT | 442676003 | Hemiplegia of dominant side as late effect of cerebrovascular disease |
| SNOMED CT | 442733008 | Hemiplegia as late effect of cerebrovascular accident |
| SNOMED CT | 443929000 | Small vessel cerebrovascular disease |
| SNOMED CT | 703163006 | Secondary cerebrovascular disease |
| SNOMED CT | 705128004 | Cerebral infarction due to embolism of middle cerebral artery |
| SNOMED CT | 705130002 | Cerebral infarction due to thrombosis of middle cerebral artery |
| SNOMED CT | 95457000 | Brain stem infarction |
| SNOMED CT | 111297002 | Nonparalytic stroke |
| SNOMED CT | 111298007 | Chronic cerebral ischemia |
| SNOMED CT | 116288000 | Paralytic stroke |
| SNOMED CT | 161511000 | History of transient ischemic attack |
| SNOMED CT | 192813004 | Cerebral degeneration due to cerebrovascular disease |
| SNOMED CT | 195185009 | Cerebral infarct due to thrombosis of precerebral arteries |
| SNOMED CT | 195186005 | Cerebral infarction due to embolism of precerebral arteries |
| SNOMED CT | 195189003 | Cerebral infarction due to thrombosis of cerebral arteries |
| SNOMED CT | 195190007 | Cerebral infarction due to embolism of cerebral arteries |
| SNOMED CT | 195212005 | Brainstem stroke syndrome |

Continued on next page

**TableS25 – continued from previous page**

| CodeSystem | ConceptCode | ConceptName |
|---|---|---|
| SNOMED CT | 195213000 | Cerebellar stroke syndrome |
| SNOMED CT | 195230003 | Cerebral infarction due to cerebral venous thrombosis, non-pyogenic |
| SNOMED CT | 195239002 | Late effects of cerebrovascular disease |
| SNOMED CT | 195243003 | Sequelae of cerebral infarction |
| SNOMED CT | 230690007 | Cerebrovascular accident |
| SNOMED CT | 230693009 | Anterior cerebral circulation infarction |
| SNOMED CT | 230694003 | Total anterior cerebral circulation infarction |
| SNOMED CT | 230695002 | Partial anterior cerebral circulation infarction |
| SNOMED CT | 230696001 | Posterior cerebral circulation infarction |
| SNOMED CT | 230698000 | Lacunar infarction |
| SNOMED CT | 230707007 | Anterior cerebral circulation hemorrhagic infarction |
| SNOMED CT | 230708002 | Posterior cerebral circulation hemorrhagic infarction |
| SNOMED CT | 230713003 | Stroke of uncertain pathology |
| SNOMED CT | 230714009 | Anterior circulation stroke of uncertain pathology |
| SNOMED CT | 230715005 | Posterior circulation stroke of uncertain pathology |
| SNOMED CT | 230738008 | Asymptomatic cerebrovascular disease |
| SNOMED CT | 20059004 | Cerebral artery occlusion |
| SNOMED CT | 25133001 | Completed stroke |
| SNOMED CT | 57981008 | Progressing stroke |
| SNOMED CT | 62914000 | Cerebrovascular disease |
| SNOMED CT | 361000119103 | Paralytic syndrome on one side of the body as late effect of cerebrovascular accident |
| SNOMED CT | 5571000124103 | Cerebrovascular accident with intracranial hemorrhage |
| SNOMED CT | 9901000119100 | Occlusion of cerebral artery with stroke |
| SNOMED CT | 18761000119108 | Monoplegia of arm dominant side as sequela of cerebrovascular disease |
| SNOMED CT | 23671000119107 | Sequela of ischemic cerebral infarction |
| SNOMED CT | 26021000119107 | Vertigo as sequela of cerebrovascular disease |
| SNOMED CT | 29941000119105 | Ataxia as sequela of cerebrovascular disease |
| SNOMED CT | 33301000119105 | Sequela of cardioembolic stroke |
| SNOMED CT | 33331000119103 | Sequela of lacunar stroke |
| SNOMED CT | 34191000119104 | Cerebral infarction due to vertebral artery occlusion |
| SNOMED CT | 40161000119102 | Weakness of face muscles as sequela of stroke |
| SNOMED CT | 46421000119102 | Behavior disorder as sequela of cerebral infarction |
| SNOMED CT | 48601000119107 | Hemiplegia and/or hemiparesis following stroke |
| SNOMED CT | 87551000119101 | Visual disturbance as sequela of cerebrovascular disease |
| SNOMED CT | 91601000119109 | Sequela of thrombotic stroke |
| SNOMED CT | 92341000119107 | Weakness of extremities as sequela of stroke |
| SNOMED CT | 99451000119105 | Cerebral infarction due to stenosis of carotid artery |
| SNOMED CT | 102831000119104 | Paralytic syndrome of both lower limbs as sequela of stroke |
| SNOMED CT | 103761000119107 | Paralytic syndrome of all four limbs as sequela of stroke |
| SNOMED CT | 108691000119102 | Spasticity as sequela of stroke |
| SNOMED CT | 133981000119106 | Dysarthria as late effects of cerebrovascular disease |
| SNOMED CT | 133991000119109 | Fluency disorder as sequela of cerebrovascular disease |
| SNOMED CT | 134771000119108 | Alteration of sensation as late effect of stroke |
| SNOMED CT | 137991000119103 | Seizure disorder as sequela of stroke |

Continued on next page

**TableS25 – continued from previous page**

| CodeSystem | ConceptCode | ConceptName |
|---|---|---|
| SNOMED CT | 140281000119108 | Hemiparesis as late effect of cerebrovascular disease |
| SNOMED CT | 140911000119109 | Ischemic stroke with coma |
| SNOMED CT | 140921000119102 | Ischemic stroke without coma |
| SNOMED CT | 145741000119101 | Apraxia as late effect of cerebrovascular disease |
| SNOMED CT | 148871000119109 | Weakness as a late effect of stroke |
| SNOMED CT | 149821000119103 | Cerebral infarction due to carotid artery occlusion |
| SNOMED CT | 433891000124100 | Cerebral infarction due to cerebral artery occlusion |
| SNOMED CT | 433911000124103 | Cerebral infarction due to posterior cerebral artery occlusion |
| SNOMED CT | 433931000124109 | Cerebral infarction due to internal carotid artery occlusion |
| SNOMED CT | 433941000124104 | Occlusion and stenosis of middle cerebral artery with infarction |
| SNOMED CT | 433951000124102 | Occlusion and stenosis of posterior cerebral artery with infarction |
| SNOMED CT | 433961000124100 | Cerebral infarction due to cerebral venous thrombosis |
| SNOMED CT | 433971000124107 | Cerebral venous thrombosis of cortical vein with infarction |
| SNOMED CT | 434141000124103 | Chronic cerebrovascular accident |
| SNOMED CT | 434151000124101 | Cerebral infarction due to anterior cerebral artery occlusion |
| SNOMED CT | 434961000124102 | Cerebral infarction due to middle cerebral artery occlusion |
| SNOMED CT | 434991000124105 | Cerebral infarction due to basilar artery stenosis |
| SNOMED CT | 230706003 | Hemorrhagic stroke |
| SNOMED CT | 262940009 | Transection of vertebral artery |
| SNOMED CT | 262942001 | Transection of carotid artery |
| SNOMED CT | 266253001 | Precerebral arterial occlusion |
| SNOMED CT | 266254007 | Carotid artery occlusion |
| SNOMED CT | 275363001 | Rupture of superficial cerebral vein |
| SNOMED CT | 276220007 | Foville syndrome |
| SNOMED CT | 276222004 | Top of basilar syndrome |
| SNOMED CT | 276594006 | Perinatal rupture of superficial cerebral vein |
| SNOMED CT | 276706004 | Perinatal cerebral ischemia |
| SNOMED CT | 277196008 | Berry aneurysm |
| SNOMED CT | 277299009 | Ruptured cerebral arteriovenous malformation |
| SNOMED CT | 277315000 | Ruptured aneurysm of anterior cerebral artery |
| SNOMED CT | 277316004 | Ruptured aneurysm of middle cerebral artery |
| SNOMED CT | 277319006 | Ruptured aneurysm of posterior cerebral artery |
| SNOMED CT | 277320000 | Ruptured aneurysm of anterior communicating artery |
| SNOMED CT | 277322008 | Ruptured aneurysm of posterior communicating artery |
| SNOMED CT | 277324009 | Ruptured aneurysm of basilar artery |
| SNOMED CT | 277325005 | Ruptured aneurysm of posterior inferior cerebellar artery |
| SNOMED CT | 277328007 | Ruptured internal carotid-anterior communicating artery zone aneurysm |
| SNOMED CT | 277329004 | Ruptured internal carotid-posterior communicating artery zone aneurysm |
| SNOMED CT | 277330009 | Ruptured internal carotid bifurcation aneurysm |
| SNOMED CT | 286742002 | Impending cerebrovascular accident |
| SNOMED CT | 287731003 | Cerebral ischemia |
| SNOMED CT | 288723005 | Acute ill-defined cerebrovascular disease |
| SNOMED CT | 297138001 | Embolus of circle of Willis |
| SNOMED CT | 297157005 | Thrombus of intracranial vein |
| SNOMED CT | 297176007 | Vertebral artery aneurysm |

Continued on next page

**TableS25 – continued from previous page**

| CodeSystem | ConceptCode | ConceptName |
|---|---|---|
| SNOMED CT | 300920004 | Carotid atherosclerosis |
| SNOMED CT | 302213007 | Carotid cavernous fistula |
| SNOMED CT | 302878004 | Intracranial septic thrombophlebitis |
| SNOMED CT | 302879007 | Septic thrombophlebitis of cavernous sinus |
| SNOMED CT | 302880005 | Septic thrombophlebitis of sagittal sinus |
| SNOMED CT | 302881009 | Septic thrombophlebitis of lateral sinus |
| SNOMED CT | 302902003 | Infarction of optic chiasm |
| SNOMED CT | 302904002 | Infarction of visual cortex |
| SNOMED CT | 307362003 | Intracranial venous septic embolism |
| SNOMED CT | 307363008 | Multiple lacunar infarcts |
| SNOMED CT | 308067002 | H/O: Stroke in last year |
| SNOMED CT | 312586003 | Intracranial thrombophlebitis |
| SNOMED CT | 371121002 | Neonatal stroke |
| SNOMED CT | 371158002 | Disorder of basilar artery |
| SNOMED CT | 371160000 | Disorder of carotid artery |
| SNOMED CT | 384993003 | Periventricular hemorrhagic venous infarct |
| SNOMED CT | 403775003 | Hereditary neurocutaneous angiomata |
| SNOMED CT | 408664007 | Pontine artery occlusion |
| SNOMED CT | 408665008 | Pontine artery thrombosis |
| SNOMED CT | 416792008 | Vein of Galen malformation |
| SNOMED CT | 423462008 | Dural carotid cavernous fistula |
| SNOMED CT | 425420004 | Thrombosis of internal carotid artery |
| SNOMED CT | 425932008 | Thrombosis of posterior communicating artery |
| SNOMED CT | 426107000 | Acute lacunar infarction |
| SNOMED CT | 426651005 | Bilateral carotid artery occlusion |
| SNOMED CT | 426814001 | Transient cerebral ischemia due to atrial fibrillation |
| SNOMED CT | 427020007 | Cerebral vasculitis |
| SNOMED CT | 428089008 | Venous hemangioma of brain |
| SNOMED CT | 429235008 | History of cardioembolic stroke |
| SNOMED CT | 429993008 | History of cerebrovascular accident without residual deficits |
| SNOMED CT | 440140008 | History of cerebrovascular accident with residual deficit |
| SNOMED CT | 444172003 | Recurrent transient cerebral ischemic attack |
| SNOMED CT | 444657001 | Superior cerebellar artery syndrome |
| SNOMED CT | 444869007 | Cavernous hemangioma of brain |
| SNOMED CT | 445109004 | Isolation of left common carotid artery |
| SNOMED CT | 445349004 | Isolation of right common carotid artery |
| SNOMED CT | 446712002 | Thromboembolus of precerebral artery |
| SNOMED CT | 449903008 | Type I arteriovenous malformation of spinal cord |
| SNOMED CT | 460307002 | Systemic to pulmonary collateral artery from right carotid artery |
| SNOMED CT | 460312001 | Systemic to pulmonary collateral artery from left carotid artery |
| SNOMED CT | 460880006 | Arteriovenous fistula of great cerebral vein of Galen |
| SNOMED CT | 460890003 | Anomalous common origin of brachiocephalic artery and left common carotid artery |
| SNOMED CT | 460899002 | Anomalous origin of left common carotid artery from brachiocephalic artery |
| SNOMED CT | 461326001 | Anomalous separate origins of internal carotid arteries and external carotid arteries from single aortic arch |

Continued on next page

**TableS25 – continued from previous page**

| CodeSystem | ConceptCode | ConceptName |
|---|---|---|
| SNOMED CT | 472746006 | Cerebrovascular disorder due to paradoxical embolus |
| SNOMED CT | 698363002 | Postoperative thromboembolism of precerebral artery |
| SNOMED CT | 698627005 | Postoperative phlebitis and thrombophlebitis of intracranial sinuses |
| SNOMED CT | 699429007 | History of cerebrovascular accident in last eight weeks |
| SNOMED CT | 699706000 | Embolism of middle cerebral artery |
| SNOMED CT | 700467001 | Reversible cerebral vasoconstriction syndrome |
| SNOMED CT | 702374000 | Neonatal noninfectious cerebral venous sinus thrombosis |
| SNOMED CT | 702575003 | Retinocochleocerebral vasculopathy |
| SNOMED CT | 703166003 | Dural arteriovenous fistula |
| SNOMED CT | 703176000 | Ruptured aneurysm of vertebral artery |
| SNOMED CT | 703180005 | Asymptomatic occlusion of extracranial carotid artery |
| SNOMED CT | 703184001 | Asymptomatic occlusion of intracranial carotid artery |
| SNOMED CT | 703193000 | Congenital malformation of dural sinus |
| SNOMED CT | 703205008 | Asymptomatic occlusion of posterior cerebral artery |
| SNOMED CT | 703206009 | Asymptomatic occlusion of basilar artery |
| SNOMED CT | 703207000 | Asymptomatic occlusion of anterior cerebral artery |
| SNOMED CT | 703208005 | Asymptomatic occlusion of middle cerebral artery |
| SNOMED CT | 703218000 | Cerebral vasoconstriction syndrome |
| SNOMED CT | 703219008 | Cerebral autosomal recessive arteriopathy with subcortical infarcts and leucoencephalopathy |
| SNOMED CT | 703221003 | Congenital intracranial vascular malformation |
| SNOMED CT | 703226008 | Familial cerebral saccular aneurysm |
| SNOMED CT | 703266007 | Cerebrofacial arteriovenous metameric syndrome |
| SNOMED CT | 703267003 | Cerebrofacial arteriovenous metameric syndrome type 1 |
| SNOMED CT | 703268008 | Cerebrofacial arteriovenous metameric syndrome type 3 |
| SNOMED CT | 703311009 | Cerebral arteritis due to infectious disease |
| SNOMED CT | 703312002 | Primary cerebral arteritis |
| SNOMED CT | 703313007 | Cerebral amyloid angiopathy associated with systemic amyloidosis |
| SNOMED CT | 705066004 | Dissection of internal carotid artery |
| SNOMED CT | 705129007 | Thrombosis of middle cerebral artery |
| SNOMED CT | 710575003 | Transient ischemic attack due to embolism |
| SNOMED CT | 713035000 | Dissection of precerebral artery |
| SNOMED CT | 713081000 | Dissection of cerebral artery |
| SNOMED CT | 713265001 | Nontraumatic ruptured cerebral aneurysm |
| SNOMED CT | 716051003 | CVA (cerebrovascular accident) during surgery |
| SNOMED CT | 716745004 | Sneddon syndrome |
| SNOMED CT | 717003001 | Hereditary cavernous hemangioma of brain |
| SNOMED CT | 718551002 | Moyamoya disease with early onset achalasia |
| SNOMED CT | 720626009 | Dissection of carotid artery |
| SNOMED CT | 720809000 | Dissection of external carotid artery |
| SNOMED CT | 721328009 | Injury of intracranial vessel of head |
| SNOMED CT | 721411005 | Minor laceration of carotid artery |
| SNOMED CT | 721412003 | Minor laceration of vertebral artery |
| SNOMED CT | 721979005 | Lymphedema and cerebral arteriovenous anomaly syndrome |
| SNOMED CT | 722004001 | Agenesis of internal carotid artery |

Continued on next page

**TableS25 – continued from previous page**

| CodeSystem | ConceptCode | ConceptName |
|---|---|---|
| SNOMED CT | 69116000 | Moyamoya disease |
| SNOMED CT | 69798007 | Carotid artery obstruction |
| SNOMED CT | 70607008 | Thrombosis of superior sagittal sinus |
| SNOMED CT | 70936005 | Multi-infarct dementia, uncomplicated |
| SNOMED CT | 71444005 | Cerebral thrombosis |
| SNOMED CT | 73173006 | Spasm of cerebral arteries |
| SNOMED CT | 73192008 | Multiple AND bilateral precerebral artery stenosis |
| SNOMED CT | 73390009 | Endophlebitis of cavernous venous sinus |
| SNOMED CT | 75138007 | Endophlebitis of superior sagittal sinus |
| SNOMED CT | 75543006 | Cerebral embolism |
| SNOMED CT | 76402003 | Carotid artery insufficiency syndrome |
| SNOMED CT | 78569004 | Posterior inferior cerebellar artery syndrome |
| SNOMED CT | 80606009 | Carotid artery embolism |
| SNOMED CT | 80758005 | Embolism of lateral venous sinus |
| SNOMED CT | 80820004 | Injury of internal carotid artery |
| SNOMED CT | 80901002 | Endophlebitis of torcular Herophili |
| SNOMED CT | 84216001 | Cerebral venous thrombosis of pregnancy AND/OR puerperium |
| SNOMED CT | 86003009 | Carotid artery thrombosis |
| SNOMED CT | 87937009 | Endophlebitis of intracranial venous sinus |
| SNOMED CT | 88032003 | Amaurosis fugax |
| SNOMED CT | 88174006 | Basilar artery thrombosis |
| SNOMED CT | 88755007 | Phlebitis of lateral venous sinus |
| SNOMED CT | 88922007 | Thrombosis of basilar sinus |
| SNOMED CT | 89142007 | Progressive intracranial arterial occlusion |
| SNOMED CT | 89980009 | Thrombosis of cavernous venous sinus |
| SNOMED CT | 90099008 | Subcortical leukoencephalopathy |
| SNOMED CT | 90520006 | Vertebral artery stenosis |
| SNOMED CT | 92962004 | Congenital absence of carotid artery |
| SNOMED CT | 92997002 | Congenital anomaly of carotid artery |
| SNOMED CT | 93054001 | Congenital dilatation of carotid artery |
| SNOMED CT | 93312006 | Congenital malposition of carotid artery |
| SNOMED CT | 93396008 | Congenital stenosis of carotid artery |
| SNOMED CT | 93468003 | Hemangioma of intracranial structure |
| SNOMED CT | 95235009 | Retroesophageal carotid artery |
| SNOMED CT | 95455008 | Thrombosis of cerebral veins |
| SNOMED CT | 95456009 | Brain stem ischemia |
| SNOMED CT | 95458005 | Cerebellar artery occlusion |
| SNOMED CT | 95459002 | Cerebellar artery thrombosis |
| SNOMED CT | 95460007 | Cerebellar infarction |
| SNOMED CT | 95461006 | Thrombophlebitis of cerebral vein |
| SNOMED CT | 95644001 | Systemic lupus erythematosus encephalitis |
| SNOMED CT | 95830009 | Pituitary infarction |
| SNOMED CT | 111028009 | Arteriopathic granular atrophy of cerebral cortex |
| SNOMED CT | 111296006 | Basilar artery embolism |
| SNOMED CT | 111299004 | Atheroma of cerebral arteries |
| SNOMED CT | 128218002 | Disorder of intracranial venous sinus |
| SNOMED CT | 128608001 | Cerebral arterial aneurysm |

Continued on next page

**TableS25 – continued from previous page**

| CodeSystem | ConceptCode | ConceptName |
|---|---|---|
| SNOMED CT | 128609009 | Intracranial aneurysm |
| SNOMED CT | 186317009 | Listerial cerebral arteritis |
| SNOMED CT | 186893003 | Rupture of syphilitic cerebral aneurysm |
| SNOMED CT | 192753009 | Phlebitis and thrombophlebitis of intracranial sinuses |
| SNOMED CT | 192755002 | Embolism superior longitudinal sinus |
| SNOMED CT | 192759008 | Cerebral venous sinus thrombosis |
| SNOMED CT | 192760003 | Thrombosis of superior longitudinal sinus |
| SNOMED CT | 192761004 | Thrombosis transverse sinus |
| SNOMED CT | 192764007 | Phlebitis cavernous sinus |
| SNOMED CT | 192765008 | Phlebitis of superior longitudinal sinus |
| SNOMED CT | 192769002 | Thrombophlebitis of central nervous system venous sinuses |
| SNOMED CT | 192770001 | Thrombophlebitis of cavernous sinus |
| SNOMED CT | 192771002 | Thrombophlebitis of superior longitudinal venous sinus |
| SNOMED CT | 192772009 | Thrombophlebitis lateral venous sinus |
| SNOMED CT | 195154000 | Ruptured berry aneurysm |
| SNOMED CT | 195155004 | Subarachnoid hemorrhage from carotid siphon and bifurcation |
| SNOMED CT | 195160000 | Subarachnoid hemorrhage from vertebral artery |
| SNOMED CT | 195180004 | Basilar artery occlusion |
| SNOMED CT | 195182007 | Vertebral artery occlusion |
| SNOMED CT | 195183002 | Multiple and bilateral precerebral arterial occlusion |
| SNOMED CT | 195200006 | Carotid artery syndrome hemispheric |
| SNOMED CT | 195205001 | Impending cerebral ischemia |
| SNOMED CT | 195206000 | Intermittent cerebral ischemia |
| SNOMED CT | 195209007 | Middle cerebral artery syndrome |
| SNOMED CT | 195210002 | Anterior cerebral artery syndrome |
| SNOMED CT | 195211003 | Posterior cerebral artery syndrome |
| SNOMED CT | 195216008 | Left sided cerebral hemisphere cerebrovascular accident |
| SNOMED CT | 195217004 | Right sided cerebral hemisphere cerebrovascular accident |
| SNOMED CT | 195229008 | Non-pyogenic venous sinus thrombosis |
| SNOMED CT | 195232006 | Occlusion and stenosis of middle cerebral artery |
| SNOMED CT | 195233001 | Occlusion and stenosis of anterior cerebral artery |
| SNOMED CT | 195234007 | Occlusion and stenosis of posterior cerebral artery |
| SNOMED CT | 195235008 | Occlusion and stenosis of cerebellar arteries |
| SNOMED CT | 195236009 | Occlusion and stenosis of multiple and bilateral cerebral arteries |
| SNOMED CT | 195373009 | Anterior spinal and vertebral artery compression syndromes |
| SNOMED CT | 200258006 | Obstetric cerebral venous thrombosis |
| SNOMED CT | 200259003 | Cerebral venous thrombosis in pregnancy |
| SNOMED CT | 200260008 | Cerebral venous thrombosis in puerperium |
| SNOMED CT | 200330000 | Puerperal cerebrovascular disorder - delivered |
| SNOMED CT | 200331001 | Puerperal cerebrovascular disorder - delivered with postnatal complication |
| SNOMED CT | 200332008 | Puerperal cerebrovascular disorder with antenatal complication |
| SNOMED CT | 200333003 | Puerperal cerebrovascular disorder with postnatal complication |
| SNOMED CT | 204493007 | Arteriovenous malformation of precerebral vessels |
| SNOMED CT | 204497008 | Cerebrovascular system anomalies |
| SNOMED CT | 204501003 | Congenital stricture of cerebral artery |
| SNOMED CT | 206576006 | Neonatal cerebral ischemia |

Continued on next page

**TableS25 – continued from previous page**

| CodeSystem | ConceptCode | ConceptName |
|---|---|---|
| SNOMED CT | 213044006 | Mechanical complication of carotid artery bypass |
| SNOMED CT | 230220006 | Intracranial septic embolism |
| SNOMED CT | 230221005 | Intracranial arterial septic embolism |
| SNOMED CT | 230222003 | Septic thrombophlebitis of straight sinus |
| SNOMED CT | 230223008 | Septic thrombophlebitis of sigmoid sinus |
| SNOMED CT | 230224002 | Septic thrombophlebitis of cortical vein |
| SNOMED CT | 230225001 | Septic thrombophlebitis of great cerebral vein |
| SNOMED CT | 230285003 | Vascular dementia of acute onset |
| SNOMED CT | 230287006 | Mixed cortical and subcortical vascular dementia |
| SNOMED CT | 230482003 | Carotidynia |
| SNOMED CT | 230518009 | Infarction of optic tract |
| SNOMED CT | 230523009 | Infarction of optic radiation |
| SNOMED CT | 230691006 | CVA - cerebrovascular accident due to cerebral artery occlusion |
| SNOMED CT | 230692004 | Infarction - precerebral |
| SNOMED CT | 230699008 | Pure motor lacunar infarction |
| SNOMED CT | 230700009 | Pure sensory lacunar infarction |
| SNOMED CT | 230701008 | Pure sensorimotor lacunar infarction |
| SNOMED CT | 230702001 | Lacunar ataxic hemiparesis |
| SNOMED CT | 230703006 | Dysarthria-clumsy hand syndrome |
| SNOMED CT | 230716006 | Carotid territory transient ischemic attack |
| SNOMED CT | 230717002 | Vertebrobasilar territory transient ischemic attack |
| SNOMED CT | 230720005 | Cerebral venous thrombosis of straight sinus |
| SNOMED CT | 230721009 | Cerebral venous thrombosis of sigmoid sinus |
| SNOMED CT | 230722002 | Cerebral venous thrombosis of cortical vein |
| SNOMED CT | 230723007 | Cerebral venous thrombosis of great cerebral vein |
| SNOMED CT | 230724001 | Cerebral amyloid angiopathy |
| SNOMED CT | 230725000 | Sporadic cerebral amyloid angiopathy |
| SNOMED CT | 230730001 | Dissection of vertebral artery |
| SNOMED CT | 230731002 | Cerebral arteritis in systemic vasculitis |
| SNOMED CT | 230732009 | Cerebral arteritis in giant cell arteritis |
| SNOMED CT | 230735006 | Syphilitic cerebral arteritis |
| SNOMED CT | 230769007 | Periventricular leucomalacia |
| SNOMED CT | 232036006 | Cilioretinal artery occlusion |
| SNOMED CT | 233964008 | Internal carotid artery stenosis |
| SNOMED CT | 233983001 | Ruptured cerebral aneurysm |
| SNOMED CT | 233988005 | Carotid artery aneurysm |
| SNOMED CT | 234005004 | Vertebral artery rupture |
| SNOMED CT | 234006003 | Carotid artery rupture |
| SNOMED CT | 234142008 | Cerebral arteriovenous malformation |
| SNOMED CT | 234149004 | Congenital arteriovenous fistula of brain |
| SNOMED CT | 237701005 | Pituitary apoplexy |
| SNOMED CT | 237867001 | Hereditary cerebrovascular amyloidosis |
| SNOMED CT | 253194008 | Aneurysm of the vein of Galen |
| SNOMED CT | 253699002 | Isolation of common carotid artery |
| SNOMED CT | 262717000 | Injury of vertebral artery |
| SNOMED CT | 590005 | Congenital aneurysm of anterior communicating artery |
| SNOMED CT | 1055001 | Stenosis of precerebral artery |

Continued on next page

**TableS25 – continued from previous page**

| CodeSystem | ConceptCode | ConceptName |
|---|---|---|
| SNOMED CT | 2495006 | Congenital cerebral arteriovenous aneurysm |
| SNOMED CT | 3681008 | Thrombophlebitis of torcular Herophili |
| SNOMED CT | 4262001 | Phlebitis of superior sagittal sinus |
| SNOMED CT | 6594005 | Cerebrovascular disorder in the puerperium |
| SNOMED CT | 6715005 | Injury of external carotid artery |
| SNOMED CT | 6729006 | Cerebrofacial arteriovenous metameric syndrome type 2 |
| SNOMED CT | 6956001 | Injury of carotid artery |
| SNOMED CT | 8166000 | Thrombophlebitis of basilar sinus |
| SNOMED CT | 8269002 | Cerebrospinal angiopathy |
| SNOMED CT | 10349009 | Multi-infarct dementia with delirium |
| SNOMED CT | 10878002 | Aneurysm of common carotid artery |
| SNOMED CT | 12853006 | Embolism of torcular Herophili |
| SNOMED CT | 14070001 | Multi-infarct dementia with depression |
| SNOMED CT | 14246007 | Embolism of intracranial venous sinus |
| SNOMED CT | 14309005 | Anterior choroidal artery syndrome |
| SNOMED CT | 14977000 | Multiple AND bilateral precerebral artery thrombosis |
| SNOMED CT | 15705007 | Phlebitis of basilar sinus |
| SNOMED CT | 15742000 | Thrombosis of inferior sagittal sinus |
| SNOMED CT | 16061002 | Endophlebitis of lateral venous sinus |
| SNOMED CT | 16418006 | Embolism of basilar sinus |
| SNOMED CT | 18058007 | Phlebitis of intracranial venous sinus |
| SNOMED CT | 18322005 | Thrombosis of torcular Herophili |
| SNOMED CT | 19474002 | Injury of common carotid artery |
| SNOMED CT | 21258007 | Thrombosis of lateral venous sinus |
| SNOMED CT | 23808003 | Rolandic vein occlusion syndrome |
| SNOMED CT | 23819000 | Embolism of cavernous venous sinus |
| SNOMED CT | 24624008 | Aneurysm of internal carotid artery |
| SNOMED CT | 25772007 | Multi-infarct dementia with delusions |
| SNOMED CT | 26954004 | Thrombophlebitis of superior sagittal sinus |
| SNOMED CT | 28366008 | Cerebral arteritis |
| SNOMED CT | 28790007 | Obstruction of precerebral artery |
| SNOMED CT | 29322000 | Acute cerebrovascular insufficiency |
| SNOMED CT | 30400005 | Middle meningeal hemorrhage following injury |
| SNOMED CT | 32112006 | Phlebitis of inferior sagittal sinus |
| SNOMED CT | 34781003 | Vertebral artery syndrome |
| SNOMED CT | 35386004 | Cavernous sinus syndrome |
| SNOMED CT | 37943007 | Multiple AND bilateral precerebral artery embolism |
| SNOMED CT | 38742007 | Central retinal artery occlusion |
| SNOMED CT | 40276003 | Embolism of precerebral artery |
| SNOMED CT | 40450001 | Embolism of superior sagittal sinus |
| SNOMED CT | 42970005 | Nonpyogenic thrombosis of intracranial venous sinus |
| SNOMED CT | 43658003 | Vertebral artery obstruction |
| SNOMED CT | 48248005 | Thrombophlebitis of inferior sagittal sinus |
| SNOMED CT | 48601002 | Thrombosis of precerebral artery |
| SNOMED CT | 50751005 | Sinus pericranii |
| SNOMED CT | 51723007 | Carotid sinus syncope |
| SNOMED CT | 54265003 | Congenital anomaly of cerebral artery |

Continued on next page

**TableS25 – continued from previous page**

| CodeSystem | ConceptCode | ConceptName |
|---|---|---|
| SNOMED CT | 54519002 | Basilar artery stenosis |
| SNOMED CT | 55382008 | Cerebral atherosclerosis |
| SNOMED CT | 55734000 | Endophlebitis of basilar sinus |
| SNOMED CT | 56267009 | Multi-infarct dementia |
| SNOMED CT | 56384000 | Embolism of inferior sagittal sinus |
| SNOMED CT | 60706008 | Phlebitis of torcular Herophili |
| SNOMED CT | 61091005 | Aneurysm of external carotid artery |
| SNOMED CT | 61687004 | Endophlebitis of inferior sagittal sinus |
| SNOMED CT | 62702001 | Cerebral vein occlusion |
| SNOMED CT | 63795001 | Thrombosis of intracranial venous sinus of pregnancy AND/OR puerperium |
| SNOMED CT | 64009001 | Basilar artery syndrome |
| SNOMED CT | 64586002 | Carotid artery stenosis |
| SNOMED CT | 64775002 | Vertebral artery thrombosis |
| SNOMED CT | 65084004 | Vertebral artery embolism |
| SNOMED CT | 65312002 | Cerebral arteriosclerosis |
| SNOMED CT | 65587001 | Congenital anomaly of cerebrovascular system |
| SNOMED CT | 67992007 | Multiple AND bilateral precerebral artery obstruction |
| SNOMED CT | 722642008 | Major laceration of carotid artery |
| SNOMED CT | 722643003 | Major laceration of vertebral artery |
| SNOMED CT | 722929005 | Perinatal arterial ischemic stroke of fetus and/or neonate |
| SNOMED CT | 722930000 | Neonatal thrombosis of cerebral venous sinus |
| SNOMED CT | 723082006 | Silent cerebral infarct |
| SNOMED CT | 723083001 | Late effects of cerebral ischemic stroke |
| SNOMED CT | 723857007 | Silent microhemorrhage of brain |
| SNOMED CT | 724097003 | Moyamoya angiopathy, short stature, facial dysmorphism, hypergonadotropic hypogonadism syndrome |
| SNOMED CT | 724357007 | Hereditary cerebral hemorrhage with amyloidosis |
| SNOMED CT | 724424009 | Cerebral ischemic stroke due to small artery occlusion |
| SNOMED CT | 724425005 | Cerebral ischemic stroke due to intracranial large artery atherosclerosis |
| SNOMED CT | 724426006 | Cerebral ischemic stroke due to extracranial large artery atherosclerosis |
| SNOMED CT | 724427002 | Asymptomatic stenosis of intracranial artery |
| SNOMED CT | 724428007 | Asymptomatic stenosis of extracranial artery |
| SNOMED CT | 724429004 | Stroke co-occurrent with migraine |
| SNOMED CT | 724993002 | Cerebral ischemic stroke due to occlusion of extracranial large artery |
| SNOMED CT | 724994008 | Cerebral ischemic stroke due to stenosis of extracranial large artery |
| SNOMED CT | 725132001 | Ischemic stroke without residual deficits |
| SNOMED CT | 733199002 | Multifocal cerebral infarction due to and following procedure on cardiovascular system |
| SNOMED CT | 734326000 | Stenosis of left vertebral artery |
| SNOMED CT | 734327009 | Stenosis of right vertebral artery |
| SNOMED CT | 734374000 | Thrombosis of left carotid artery |
| SNOMED CT | 734382000 | Thrombosis of right carotid artery |

Continued on next page

**TableS25 – continued from previous page**

| CodeSystem | ConceptCode | ConceptName |
|---|---|---|
| SNOMED CT | 734383005 | Thrombosis of left middle cerebral artery |
| SNOMED CT | 734384004 | Thrombosis of right middle cerebral artery |
| SNOMED CT | 734396006 | Spontaneous rupture of left posterior communicating artery |
| SNOMED CT | 734397002 | Spontaneous rupture of right posterior communicating artery |
| SNOMED CT | 734879002 | Ruptured aneurysm of right posterior communicating artery |
| SNOMED CT | 734880004 | Ruptured aneurysm of left posterior communicating artery |
| SNOMED CT | 734959006 | Embolus of left cerebellar artery |
| SNOMED CT | 734960001 | Embolus of right cerebellar artery |
| SNOMED CT | 734961002 | Embolus of left posterior cerebral artery |
| SNOMED CT | 734963004 | Embolus of right posterior cerebral artery |
| SNOMED CT | 734964005 | Embolus of left middle cerebral artery |
| SNOMED CT | 734965006 | Embolus of right middle cerebral artery |
| SNOMED CT | 735114006 | Occlusion of right pontine artery |
| SNOMED CT | 735115007 | Occlusion of left pontine artery |
| SNOMED CT | 735131004 | Occlusion of left cerebellar artery |
| SNOMED CT | 735132006 | Occlusion of right cerebellar artery |
| SNOMED CT | 737159004 | Aneurysm of basilar artery |
| SNOMED CT | 737160009 | Dissection of basilar artery |
| SNOMED CT | 762629007 | Occlusion of right middle cerebral artery by embolus |
| SNOMED CT | 762630002 | Occlusion of left middle cerebral artery by embolus |
| SNOMED CT | 762632005 | Occlusion of left cerebellar artery by embolus |
| SNOMED CT | 762633000 | Occlusion of right cerebellar artery by embolus |
| SNOMED CT | 762648006 | Stenosis of right cerebellar artery |
| SNOMED CT | 762649003 | Stenosis of left cerebellar artery |
| SNOMED CT | 762651004 | Occlusion of right posterior cerebral artery by embolus |
| SNOMED CT | 762652006 | Occlusion of left posterior cerebral artery by embolus |
| SNOMED CT | 771476007 | Autosomal recessive leukoencephalopathy, ischemic stroke, retinitis pigmentosa syndrome |
| SNOMED CT | 783161005 | ABri amyloidosis |
| SNOMED CT | 783258000 | ADan amyloidosis |
| SNOMED CT | 783413008 | Multiple aneurysms of cerebral artery |
| SNOMED CT | 783415001 | Aneurysm of internal carotid bifurcation |
| SNOMED CT | 783416000 | Aneurysm of anterior cerebral artery |
| SNOMED CT | 783417009 | Aneurysm of posterior inferior cerebellar artery |
| SNOMED CT | 783418004 | Aneurysm of anterior communicating artery |
| SNOMED CT | 783419007 | Aneurysm of posterior cerebral artery |
| SNOMED CT | 783420001 | Aneurysm of middle cerebral artery |
| SNOMED CT | 783421002 | Aneurysm of posterior communicating artery |
| SNOMED CT | 783422009 | Aneurysm of internal carotid-anterior communicating artery zone |
| SNOMED CT | 783423004 | Aneurysm of internal carotid-posterior communicating artery zone |
| SNOMED CT | 783629005 | Congenital aneurysm of cerebral artery |
| SNOMED CT | 783630000 | Congenital aneurysm of precerebral artery |
| SNOMED CT | 783707003 | Cerebral aneurysm due to dissection of cerebral artery |
| SNOMED CT | 783716004 | Acquired aneurysm of cerebral artery |
| SNOMED CT | 783731008 | Fibromuscular dysplasia of wall of carotid artery |

Continued on next page

**TableS25 – continued from previous page**

| CodeSystem | ConceptCode | ConceptName |
|---|---|---|
| SNOMED CT | 783733006 | Bilateral fibromuscular dysplasia of wall of carotid arteries |
| SNOMED CT | 787044009 | Bilateral stenosis of carotid arteries |
| SNOMED CT | 788454002 | Bilateral stenosis of vertebral arteries |
| SNOMED CT | 788455001 | Bilateral occlusion of pontine arteries |
| SNOMED CT | 788880006 | Cerebral ischemic stroke due to dissection of artery |
| SNOMED CT | 788881005 | Cerebral ischemic stroke due to aortic arch embolism |
| SNOMED CT | 788882003 | Cerebral ischemic stroke due to global hypoperfusion with watershed infarct |
| SNOMED CT | 788883008 | Cerebral ischemic stroke due to hypercoagulable state |
| SNOMED CT | 788884002 | Cerebral ischemic stroke due to subarachnoid hemorrhage |
| SNOMED CT | 838275008 | Stenosis of cerebral artery |
| SNOMED CT | 838308007 | Fibromuscular dysplasia of wall of intracranial artery |
| SNOMED CT | 838309004 | Cerebrovascular abnormality due to Takayasu disease |
| SNOMED CT | 840419005 | Dissection of extracranial carotid artery |
| SNOMED CT | 840420004 | Dissection of extracranial vertebral artery |
| SNOMED CT | 840422007 | Dissection of anterior cerebral artery |
| SNOMED CT | 840434004 | Dissection of posterior cerebral artery |
| SNOMED CT | 840436002 | Dissection of middle cerebral artery |
| SNOMED CT | 840437006 | Dissection of multiple cerebral arteries |
| SNOMED CT | 840438001 | Dissection of intracranial vertebral artery |
| SNOMED CT | 840439009 | Dissection of intracranial carotid artery |
| SNOMED CT | 840441005 | Dissection of intracranial artery |
| SNOMED CT | 860820000 | Hypoplasia of cranial sinus |
| SNOMED CT | 870544005 | Occlusion of distal basilar artery |
| SNOMED CT | 870566003 | Occlusion of anterior choroidal artery |
| SNOMED CT | 870579007 | Occlusion of branch of basilar artery |
| SNOMED CT | 870637009 | Cervical artery dissection |
| SNOMED CT | 871637001 | Thrombosis of multiple cerebral veins |
| SNOMED CT | 21000119103 | Asymptomatic carotid artery stenosis |
| SNOMED CT | 1131000119105 | Sequela of cerebrovascular accident |
| SNOMED CT | 5431000124100 | Thrombosis of anterior cerebral artery |
| SNOMED CT | 5451000124107 | Thrombosis of superior cerebellar artery |
| SNOMED CT | 5461000124109 | Thrombosis of anterior inferior cerebellar artery |
| SNOMED CT | 5471000124102 | Ruptured aneurysm of intracranial artery |
| SNOMED CT | 5481000124104 | Thrombophlebitis of straight sinus |
| SNOMED CT | 5491000124101 | Thrombophlebitis of transverse sinus |
| SNOMED CT | 5501000124109 | Thrombophlebitis of internal cerebral vein |
| SNOMED CT | 5511000124107 | Thrombophlebitis of sigmoid sinus |
| SNOMED CT | 5531000124101 | Thrombophlebitis of basal vein of Rosenthal |
| SNOMED CT | 5581000124100 | Thrombosis of posterior cerebral artery |
| SNOMED CT | 5591000124102 | Thrombosis of superior anastomotic vein |
| SNOMED CT | 5601000124105 | Thrombosis of posterior inferior cerebellar artery |
| SNOMED CT | 5611000124108 | Thrombosis of basal vein |
| SNOMED CT | 5651000124109 | Pseudoaneurysm of vertebral artery |
| SNOMED CT | 7931000119101 | Anterior choroidal artery thrombosis |
| SNOMED CT | 9611000119107 | Symptomatic carotid artery stenosis |
| SNOMED CT | 9631000119102 | Carotid sinus hypersensitivity |

Continued on next page

**TableS25 – continued from previous page**

| CodeSystem | ConceptCode | ConceptName |
|---|---|---|
| SNOMED CT | 18751000119106 | Monoplegia of leg dominant side as sequela of cerebrovascular disease |
| SNOMED CT | 34181000119102 | Cerebral infarction due to occlusion of basilar artery |
| SNOMED CT | 79341000119107 | Mixed dementia |
| SNOMED CT | 90921000119104 | History of cerebrovascular accident greater than eight weeks in the past |
| SNOMED CT | 97531000119106 | History of parietal cerebrovascular accident |
| SNOMED CT | 99051000119101 | History of lacunar cerebrovascular accident |
| SNOMED CT | 106021000119105 | Multi-infarct dementia due to atherosclerosis |
| SNOMED CT | 106241000119108 | Bilateral paralytic syndrome as late effect of stroke |
| SNOMED CT | 118951000119103 | History of thrombotic stroke without residual deficits |
| SNOMED CT | 118961000119101 | History of hemorrhagic cerebrovascular accident without residual deficits |
| SNOMED CT | 118971000119107 | History of embolic stroke without deficits |
| SNOMED CT | 125081000119106 | Cerebral infarction due to occlusion of precerebral artery |
| SNOMED CT | 126011000119107 | Acquired caroticocavernous sinus fistula |
| SNOMED CT | 128171000119104 | Spontaneous caroticocavernous sinus fistula |
| SNOMED CT | 140221000119109 | History of transient ischemic attack due to embolism |
| SNOMED CT | 140251000119101 | Nonatherosclerotic cerbrovascular accident |
| SNOMED CT | 140701000119108 | History of hemorrhagic stroke with hemiparesis |
| SNOMED CT | 140711000119106 | History of hemorrhagic stroke with hemiplegia |
| SNOMED CT | 141281000119101 | History of ischemic stroke without residual deficits |
| SNOMED CT | 141811000119106 | History of hemorrhagic cerebrovascular accident with residual deficit |
| SNOMED CT | 141821000119104 | History of ischemic cerebrovascular accident with residual deficit |
| SNOMED CT | 141831000119101 | History of embolic stroke with deficits |
| SNOMED CT | 142031000119104 | Visual field defect due to and following cerebrovascular accident |
| SNOMED CT | 151161000119102 | Bilateral cerebral infarction due to precererbral arterial occlusion |
| SNOMED CT | 186831000119104 | Apraxia as sequela of cerebrovascular accident |
| SNOMED CT | 284811000119102 | Aneurysm of extracranial portion of internal carotid artery |
| SNOMED CT | 284821000119109 | Aneurysm of intracranial portion of internal carotid artery |
| SNOMED CT | 284871000119105 | Atherosclerosis of left carotid artery |
| SNOMED CT | 284881000119108 | Atherosclerosis of right carotid artery |
| SNOMED CT | 285161000119105 | Left carotid artery occlusion |
| SNOMED CT | 285171000119104 | Right carotid artery occlusion |
| SNOMED CT | 285191000119103 | Left carotid artery stenosis |
| SNOMED CT | 285201000119100 | Right carotid artery stenosis |
| SNOMED CT | 290581000119101 | Ataxia as sequela of cerebrovascular accident |
| SNOMED CT | 290621000119101 | Cognitive deficit due to and following cerebrovascular disease |
| SNOMED CT | 290631000119103 | Dysarthria due to and following cerebrovascular accident |
| SNOMED CT | 290791000119105 | Fluency disorder as sequela of cerebrovascular accident |
| SNOMED CT | 290891000119100 | Monoplegia of left dominant lower limb due to and following cerebrovascular accident |
| SNOMED CT | 290901000119101 | Monoplegia of left nondominant lower limb due to and following cerebrovascular accident |
| SNOMED CT | 290911000119103 | Monoplegia of right dominant lower limb due to and following cerebrovascular accident |

Continued on next page

**TableS25 – continued from previous page**

| CodeSystem | ConceptCode | ConceptName |
|---|---|---|
| SNOMED CT | 290921000119105 | Monoplegia of right nondominant lower limb due to and following cerebrovascular accident |
| SNOMED CT | 290931000119108 | Monoplegia of lower limb as sequela of cerebrovascular accident |
| SNOMED CT | 291081000119100 | Monoplegia of left dominant upper limb due to and following cerebrovascular accident |
| SNOMED CT | 291091000119102 | Monoplegia of left nondominant upper limb as sequela of cerebrovascular accident |
| SNOMED CT | 291101000119107 | Monoplegia of right dominant upper limb due to and following cerebrovascular accident |
| SNOMED CT | 291111000119105 | Monoplegia of right nondominant arm as late effect of stroke |
| SNOMED CT | 291121000119103 | Monoplegia of arm as late effect of stroke |
| SNOMED CT | 291351000119109 | Spontaneous hemorrhage of subarachnoid space from basilar artery |
| SNOMED CT | 291371000119100 | Spontaneous hemorrhage of subarachnoid space from intracranial artery |
| SNOMED CT | 291401000119102 | Spontaneous hemorrhage of subarachnoid space from left middle cerebral artery |
| SNOMED CT | 291411000119104 | Spontaneous hemorrhage of subarachnoid space from left posterior communicating artery |
| SNOMED CT | 291471000119107 | Non-traumatic hemorrhage of subarachnoid space from right middle cerebral artery |
| SNOMED CT | 291481000119105 | Spontaneous hemorrhage of subarachnoid space from right posterior communicating artery |
| SNOMED CT | 291751000119105 | Fluency disorder due to and following non-traumatic intracerebral hemorrhage |
| SNOMED CT | 292621000119100 | Occlusion of right vertebral artery |
| SNOMED CT | 292631000119102 | Occlusion of left vertebral artery |
| SNOMED CT | 292661000119105 | Cerebrovascular accident due to stenosis of right vertebral artery |
| SNOMED CT | 292671000119104 | Cerebrovascular accident due to stenosis of left vertebral artery |
| SNOMED CT | 292681000119101 | Cerebrovascular accident due to right vertebral artery occlusion |
| SNOMED CT | 292691000119103 | Cerebrovascular accident due to occlusion of left vertebral artery |
| SNOMED CT | 292851000119109 | Lacunar ataxic hemiparesis of right dominant side |
| SNOMED CT | 292861000119106 | Lacunar ataxic hemiparesis of left dominant side |
| SNOMED CT | 293811000119100 | Cerebral infarction due to vertebral artery stenosis |
| SNOMED CT | 293831000119105 | Cerebral infarction due to stenosis of precerebral artery |
| SNOMED CT | 329361000119107 | Cerebrovascular accident due to occlusion of right middle cerebral artery by embolus |
| SNOMED CT | 329371000119101 | Cerebrovascular accident due to occlusion of left middle cerebral artery by embolus |
| SNOMED CT | 329391000119100 | Right anterior cerebral artery embolism with stroke |
| SNOMED CT | 329401000119103 | Left anterior cerebral artery embolism with stroke |
| SNOMED CT | 329421000119107 | Cerebrovascular accident due to occlusion of right posterior cerebral artery by embolus |
| SNOMED CT | 329431000119105 | Cerebrovascular accident due to occlusion of left posterior cerebral artery by embolus |
| SNOMED CT | 329451000119104 | Cerebrovascular accident due to occlusion of right cerebellar artery by embolus |

Continued on next page

**TableS25 – continued from previous page**

| CodeSystem | ConceptCode | ConceptName |
|---|---|---|
| SNOMED CT | 329461000119102 | Cerebrovascular accident due to occlusion of left cerebellar artery by embolus |
| SNOMED CT | 329481000119106 | Occlusion of right middle cerebral artery |
| SNOMED CT | 329491000119109 | Occlusion of left middle cerebral artery |
| SNOMED CT | 329521000119106 | Occlusion of right anterior cerebral artery |
| SNOMED CT | 329531000119109 | Occlusion of left anterior cerebral artery |
| SNOMED CT | 329561000119101 | Occlusion of right posterior cerebral artery |
| SNOMED CT | 329571000119107 | Occlusion of left posterior cerebral artery |
| SNOMED CT | 329581000119105 | Occlusion of bilateral posterior cerebral arteries |
| SNOMED CT | 329621000119105 | Cerebrovascular accident due to thrombus of right vertebral artery |
| SNOMED CT | 329631000119108 | Left vertebral artery thrombosis with stroke |
| SNOMED CT | 329641000119104 | Cerebrovascular accident due to thrombus of basilar artery |
| SNOMED CT | 329651000119102 | Cerebrovascular accident due to thrombus of right carotid artery |
| SNOMED CT | 329671000119106 | Traumatic subarachnoid hemorrhage with loss of consciousness |
| SNOMED CT | 330111000119103 | Ruptured acquired aneurysm of cerebral artery |
| SNOMED CT | 330411000119109 | Lacunar ataxic hemiparesis of left nondominant side |
| SNOMED CT | 330421000119102 | Lacunar ataxic hemiparesis of right nondominant side |
| SNOMED CT | 330791000119108 | Cerebrovascular accident due to thrombus of left carotid artery |
| SNOMED CT | 336191000119105 | Occlusion of right central retinal artery |
| SNOMED CT | 341801000119101 | Occlusion of left central retinal artery |
| SNOMED CT | 347011000119102 | Bilateral occlusion of central retinal arteries |
| SNOMED CT | 429221000124104 | Dissecting aneurysm of cerebral artery |
| SNOMED CT | 429231000124101 | Dissection of middle cerebral artery |
| SNOMED CT | 429251000124108 | Dissection of anterior cerebral artery |
| SNOMED CT | 429811000124106 | Anterior cerebral artery embolism |
| SNOMED CT | 429831000124100 | Cerebellar artery embolism |
| SNOMED CT | 429841000124105 | Superior cerebellar artery embolism |
| SNOMED CT | 429851000124107 | Anterior inferior cerebellar artery embolism |
| SNOMED CT | 429861000124109 | Posterior inferior cerebellar artery embolism |
| SNOMED CT | 430721000124101 | Carotid artery occlusion without infarction |
| SNOMED CT | 430731000124103 | Basilar artery occlusion without infarction |
| SNOMED CT | 430781000124102 | Occlusion and stenosis of middle cerebral artery without infarction |
| SNOMED CT | 430831000124106 | Occlusion and stenosis of posterior cerebral artery without infarction |
| SNOMED CT | 430841000124101 | Occlusion and stenosis of anterior cerebral artery without infarction |
| SNOMED CT | 430851000124104 | Multiple and bilateral precerebral artery stenosis with infarction |
| SNOMED CT | 430861000124102 | Multiple and bilateral precerebral artery stenosis without infarction |
| SNOMED CT | 431421000124103 | Arteriosclerosis of carotid artery |
| SNOMED CT | 432051000124108 | History of embolic stroke without residual deficits |
| SNOMED CT | 432181000124104 | History of hemorrhagic stroke with residual hemiplegia |
| SNOMED CT | 432191000124101 | History of hemorrhagic stroke with residual hemiparesis |
| SNOMED CT | 432261000124102 | Cerebral venous thrombosis of cortical vein without infarction |
| SNOMED CT | 432481000124106 | Thrombosis of cerebral medullary veins |

Continued on next page

**TableS25 – continued from previous page**

| CodeSystem | ConceptCode | ConceptName |
|---|---|---|
| SNOMED CT | 433821000124102 | Occlusion of internal carotid artery |
| SNOMED CT | 434191000124107 | Post-radiotherapy cerebral vasculopathy |
| SNOMED CT | 434821000124104 | Vertebral artery occlusion with infarction |
| SNOMED CT | 434831000124101 | Vertebral artery stenosis with infarction |
| SNOMED CT | 434881000124100 | Multiple and bilateral precerebral arterial occlusion with infarction |
| SNOMED CT | 434891000124102 | Internal carotid artery stenosis with infarction |
| SNOMED CT | 434951000124104 | Middle cerebral artery occlusion |
| SNOMED CT | 435271000124103 | Vertebral artery occlusion without infarction |
| SNOMED CT | 435281000124100 | Vertebral artery stenosis without infarction |
| SNOMED CT | 436021000124100 | Carotid artery stenosis without infarction |
| SNOMED CT | 436031000124102 | Anterior inferior cerebellar artery occlusion with infarction |
| SNOMED CT | 436041000124107 | Posterior inferior cerebellar artery occlusion with infarction |
| SNOMED CT | 436591000124109 | Thrombophlebitis of superior anastomotic vein |
| SNOMED CT | 436781000124104 | Intracranial artery occlusion with infarction |
| SNOMED CT | 436791000124101 | Anterior choroidal artery occlusion with infarction |
| SNOMED CT | 437911000124106 | Recurrent artery of Huebner occlusion with infarction |
| SNOMED CT | 454031000124106 | Occlusion of superior cerebellar artery |
| SNOMED CT | 454041000124101 | Cerebellar infarction due to occlusion of superior cerebellar artery |
| SNOMED CT | 455011000124104 | Occlusion of anterior cerebral artery |
| SNOMED CT | 455031000124105 | Occlusion of right posterior communicating artery |
| SNOMED CT | 455041000124100 | Occlusion of left posterior communicating artery |
| SNOMED CT | 455791000124106 | Occlusive embolus of left vertebral artery |
| SNOMED CT | 455801000124107 | Occlusive embolus of right vertebral artery |
| SNOMED CT | 455811000124105 | Occlusive embolus of basilar artery |
| SNOMED CT | 455821000124102 | Occlusive embolus of left carotid artery |
| SNOMED CT | 455831000124104 | Occlusive embolus of right carotid artery |
| SNOMED CT | 455841000124109 | Occlusive embolus of left anterior cerebral |
| SNOMED CT | 455851000124106 | Occlusive embolus of right anterior cerebral |
| SNOMED CT | 455901000124101 | Occlusion of cerebral vein by nonpyogenic thrombus |
| SNOMED CT | 455911000124103 | Occlusive thrombus of left cerebellar artery |
| SNOMED CT | 455921000124106 | Occlusive thrombus of left vertebral artery |
| SNOMED CT | 455931000124109 | Occlusive thrombus of left posterior cerebral artery |
| SNOMED CT | 457551000124104 | Acute stroke |
| SNOMED CT | 461431000124108 | Vertebral artery atherosclerosis |
| SNOMED CT | 672371000119103 | Monoplegia of nondominant lower limb due to and following ischemic cerebrovascular accident |
| SNOMED CT | 672381000119100 | Monoplegia of dominant lower limb due to and following ischemic cerebrovascular accident |
| SNOMED CT | 672421000119109 | Monoplegia of dominant upper limb due to and following ischemic cerebrovascular accident |
| SNOMED CT | 672451000119101 | Hemiplegia of nondominant side due to and following hemorrhagic cerebrovascular accident |
| SNOMED CT | 672471000119105 | Hemiplegia of dominant side due to and following hemorrhagic cerebrovascular accident |

Continued on next page

**TableS25 – continued from previous page**

| CodeSystem | ConceptCode | ConceptName |
|---|---|---|
| SNOMED CT | 672481000119108 | Fluency disorder due to and following ischemic cerebrovascular accident |
| SNOMED CT | 672491000119106 | Fluency disorder due to and following hemorrhagic cerebrovascular accident |
| SNOMED CT | 672501000119104 | Dysarthria due to and following ischemic cerebrovascular accident |
| SNOMED CT | 672511000119101 | Dysarthria due to and following hemorrhagic cerebrovascular accident |
| SNOMED CT | 672521000119108 | Dysphasia due to and following ischemic cerebrovascular accident |
| SNOMED CT | 672531000119106 | Dysphasia due to and following hemorrhagic cerebrovascular accident |
| SNOMED CT | 672541000119102 | Aphasia due to and following ischemic cerebrovascular accident |
| SNOMED CT | 672551000119100 | Aphasia due to and following hemorrhagic cerebrovascular accident |
| SNOMED CT | 672561000119103 | Cognitive deficit due to and following ischemic cerebrovascular accident |
| SNOMED CT | 672571000119109 | Cognitive deficit due to and following hemorrhagic cerebrovascular accident |
| SNOMED CT | 674091000119108 | Vertigo as sequela of ischemic cerebrovascular accident |
| SNOMED CT | 674101000119103 | Vertigo due to and following hemorrhagic cerebrovascular accident |
| SNOMED CT | 674111000119100 | Ataxia due to and following ischemic cerebrovascular accident |
| SNOMED CT | 674121000119107 | Ataxia as sequela of hemorrhagic cerebrovascular accident |
| SNOMED CT | 674131000119105 | Monoplegia of lower limb due to and following ischemic cerebrovascular accident |
| SNOMED CT | 674161000119102 | Monoplegia of arm as late effect of ischemic stroke |
| SNOMED CT | 674351000119101 | Apraxia due to and following hemorrhagic cerebrovascular accident |
| SNOMED CT | 674361000119104 | Apraxia as sequela of ischemic cerebrovascular accident |
| SNOMED CT | 674381000119108 | Weakness of facial muscle as sequela of ischemic cerebrovascular accident |
| SNOMED CT | 674391000119106 | Speech and language deficit as sequela of hemorrhagic cerebrovascular accident |
| SNOMED CT | 674401000119108 | Speech and language deficit as sequela of ischemic cerebrovascular accident |
| SNOMED CT | 690051000119100 | History of cerebellar stroke |
| SNOMED CT | 690171000119105 | Weakness of facial muscle as sequela of embolic cerebrovascular accident |
| SNOMED CT | 690181000119108 | Apraxia due to and following embolic cerebrovascular accident |
| SNOMED CT | 690191000119106 | Vertigo due to and following embolic cerebrovascular accident |
| SNOMED CT | 690201000119109 | Ataxia as sequela of embolic cerebrovascular accident |
| SNOMED CT | 690231000119102 | Monoplegia of lower limb due to and following embolic cerebrovascular accident |
| SNOMED CT | 690241000119106 | Monoplegia of nondominant upper limb due to and following embolic cerebrovascular accident |
| SNOMED CT | 690251000119108 | Monoplegia of dominant upper limb due to and following embolic cerebrovascular accident |

Continued on next page

**TableS25 – continued from previous page**

| CodeSystem | ConceptCode | ConceptName |
|---|---|---|
| SNOMED CT | 690261000119105 | Monoplegia of upper limb due to and following embolic cerebrovascular accident |
| SNOMED CT | 690271000119104 | Hemiplegia of nondominant side as sequela of embolic cerebrovascular accident |
| SNOMED CT | 690281000119101 | Hemiplegia of dominant side due to and following embolic cerebrovascular accident |
| SNOMED CT | 690311000119104 | Dysarthria due to and following embolic cerebrovascular accident |
| SNOMED CT | 690321000119106 | Aphasia due to and following embolic cerebrovascular accident |
| SNOMED CT | 690331000119109 | Speech and language deficit as sequela of embolic cerebrovascular accident |
| SNOMED CT | 690341000119100 | Cognitive deficit due to and following embolic cerebrovascular accident |
| SNOMED CT | 690351000119103 | Dysphasia due to and following embolic cerebrovascular accident |
| SNOMED CT | 12237911000119109 | Amaurosis fugax of left eye |
| SNOMED CT | 12237951000119105 | Amaurosis fugax of right eye |
| SNOMED CT | 12242711000119109 | Weakness of left facial muscle as sequela of cerebrovascular accident |
| SNOMED CT | 12242751000119105 | Weakness of right facial muscle as sequela of cerebrovascular accident |
| SNOMED CT | 12367511000119101 | Paraplegia as sequela of cerebrovascular accident |
| SNOMED CT | 12720001000004100 | Stenosis of cavernous portion of right internal carotid artery |
| SNOMED CT | 13016361000119101 | History of amaurosis fugax |
| SNOMED CT | 15648201000119100 | Aneurysm of intracranial portion of right internal carotid artery |
| SNOMED CT | 15648241000119103 | Aneurysm of intracranial portion of left internal carotid artery |
| SNOMED CT | 15648281000119108 | Aneurysm of extracranial portion of right internal carotid artery |
| SNOMED CT | 15648321000119103 | Aneurysm of extracranial portion of left internal carotid artery |
| SNOMED CT | 15648361000119108 | Aneurysm of right internal carotid artery |
| SNOMED CT | 15648401000119104 | Aneurysm of left internal carotid artery |
| SNOMED CT | 15648441000119102 | Aneurysm of right common carotid artery |
| SNOMED CT | 15648481000119107 | Aneurysm of left common carotid artery |
| SNOMED CT | 15648521000119107 | Aneurysm of right carotid artery |
| SNOMED CT | 15648561000119102 | Aneurysm of left carotid artery |
| SNOMED CT | 15707841000119101 | Thrombosis of right internal carotid artery |
| SNOMED CT | 15707881000119106 | Thrombosis of left internal carotid artery |
| SNOMED CT | 15707921000119104 | Thrombosis of bilateral internal carotid arteries |
| SNOMED CT | 15708001000119106 | Dissection of left carotid artery |
| SNOMED CT | 15708041000119108 | Dissection of right carotid artery |
| SNOMED CT | 15710681000119105 | Dissection of right vertebral artery |
| SNOMED CT | 15710721000119104 | Dissection of left vertebral artery |
| SNOMED CT | 15978431000119106 | Thrombosis of right vertebral artery |
| SNOMED CT | 15978471000119109 | Thrombosis of left vertebral artery |
| SNOMED CT | 15978631000119109 | Bilateral vertebral artery occlusion |
| SNOMED CT | 15982191000119106 | Weakness of right facial muscle due to and following non-traumatic intracerebral hemorrhage |
| SNOMED CT | 15982271000119104 | Weakness of right facial muscle as sequela of cerebrovascular disease |

Continued on next page

**TableS25 – continued from previous page**

| CodeSystem | ConceptCode | ConceptName |
|---|---|---|
| SNOMED CT | 15982311000119104 | Weakness of left facial muscle as sequela of cerebrovascular disease |
| SNOMED CT | 15982431000119103 | Weakness of left facial muscle due to and following non-traumatic intracerebral hemorrhage |
| SNOMED CT | 15988351000119101 | Acquired right carotid cavernous fistula |
| SNOMED CT | 15988391000119106 | Acquired left carotid cavernous fistula |
| SNOMED CT | 16000351000119109 | Cerebrovascular accident due to occlusion of left posterior cerebral artery |
| SNOMED CT | 16000391000119104 | Cerebrovascular accident due to occlusion of right posterior cerebral artery |
| SNOMED CT | 16000431000119109 | Cerebrovascular accident due to occlusion of right middle cerebral artery |
| SNOMED CT | 16000471000119107 | Cerebrovascular accident due to occlusion of left anterior cerebral artery |
| SNOMED CT | 16000511000119103 | Cerebrovascular accident due to occlusion of left middle cerebral artery |
| SNOMED CT | 16000551000119102 | Cerebrovascular accident due to occlusion of right anterior cerebral artery |
| SNOMED CT | 16001671000119100 | Fibromuscular hyperplasia of left carotid artery |
| SNOMED CT | 16001711000119101 | Fibromuscular hyperplasia of right carotid artery |
| SNOMED CT | 16002031000119102 | Cerebrovascular accident due to thrombus of right middle cerebral artery |
| SNOMED CT | 16002071000119104 | Cerebrovascular accident due to thrombus of right cerebellar artery |
| SNOMED CT | 16002111000119106 | Cerebrovascular accident due to thrombus of left middle cerebral artery |
| SNOMED CT | 16002151000119107 | Nonpyogenic cerebral venous thrombosis with stroke |
| SNOMED CT | 16002191000119102 | Left cerebellar artery thrombosis with stroke |
| SNOMED CT | 16002231000119106 | Cerebrovascular accident due to thrombus of right posterior cerebral artery |
| SNOMED CT | 16002271000119109 | Left posterior cerebral artery thrombosis with stroke |
| SNOMED CT | 16002351000119105 | Right vertebral artery embolism with stroke |
| SNOMED CT | 16002391000119100 | Left vertebral artery embolism with stroke |
| SNOMED CT | 16002431000119105 | Right carotid artery embolism with stroke |
| SNOMED CT | 16002471000119108 | Left carotid artery embolism with stroke |
| SNOMED CT | 16002511000119104 | Basilar artery embolism with stroke |
| SNOMED CT | 16002591000119108 | Pain of left hand due to and following cerebrovascular accident |
| SNOMED CT | 16002631000119108 | Pain of right hand due to and following cerebrovascular accident |
| SNOMED CT | 16023911000119108 | Cerebrovascular accident due to right carotid artery occlusion |
| SNOMED CT | 16023991000119104 | Cerebrovascular accident due to occlusion of left pontine artery |
| SNOMED CT | 16024031000119100 | Cerebrovascular accident due to occlusion of right pontine artery |
| SNOMED CT | 16024111000119109 | Cerebrovascular accident due to occlusion of left carotid artery |
| SNOMED CT | 16024151000119105 | Cerebrovascular accident due to occlusion of left cerebellar artery |
| SNOMED CT | 16024191000119100 | Cerebrovascular accident due to occlusion of left posterior communicating artery |
| SNOMED CT | 16024231000119109 | Cerebrovascular accident due to occlusion of right posterior communicating artery |

Continued on next page

**TableS25 – continued from previous page**

| CodeSystem | ConceptCode | ConceptName |
|---|---|---|
| SNOMED CT | 16024271000119107 | Cerebrovascular accident due to occlusion of right cerebellar artery |
| SNOMED CT | 16026951000119102 | Cerebrovascular accident due to right carotid artery stenosis |
| SNOMED CT | 16218291000119100 | Acute cerebral ischemia |
| SNOMED CT | 16260551000119106 | Dysphasia due to and following cerebrovascular accident |
| SNOMED CT | 16279401000119108 | Occlusion of right cilioretinal artery |
| SNOMED CT | 16279441000119105 | Occlusion of left cilioretinal artery |
| SNOMED CT | 16371781000119100 | Cerebellar stroke |
| SNOMED CT | 16476641000119100 | Acquired arteriovenous fistula of dura of cerebrum |
| SNOMED CT | 16644681000119102 | Cerebrovascular accident due to occlusion of bilateral pontine arteries |
| SNOMED CT | 16661931000119102 | Cerebrovascular accident due to stenosis of bilateral vertebral arteries |
| SNOMED CT | 16661971000119104 | Cerebrovascular accident due to stenosis of bilateral carotid arteries |
| SNOMED CT | 16662331000119106 | Aneurysm of right vertebral artery |
| SNOMED CT | 16662371000119109 | Aneurysm of left vertebral artery |
| SNOMED CT | 16703551000119107 | Memory deficit due to and following cerebrovascular disease |
| SNOMED CT | 16703661000119105 | Memory deficit due to and following cerebrovascular accident |
| SNOMED CT | 16703761000119102 | Memory deficit due to and following ischemic cerebrovascular accident |
| SNOMED CT | 16703821000119101 | Memory deficit due to and following hemorrhagic cerebrovascular accident |
| SNOMED CT | 16709811000119106 | Spontaneous hemorrhage of subarachnoid space from anterior communicating artery |
| SNOMED CT | 16896851000119101 | History of embolic cerebrovascular accident |
| SNOMED CT | 16896891000119106 | History of cerebrovascular accident due to ischemia |
| SNOMED CT | 21290001000004104 | Stenosis of middle cerebral artery |
| SNOMED CT | 27820001000004104 | Stenosis of intracranial vessel |
| SNOMED CT | 24654003 | Weber syndrome |
| SNOMED CT | 36179005 | Reversible ischemic neurologic deficit syndrome |
| SNOMED CT | 39925003 | Mitochondrial myopathy, encephalopathy, lactic acidosis and stroke-like episodes |
| SNOMED CT | 45639009 | Hereditary cystatin C amyloid angiopathy |
| SNOMED CT | 56453003 | Dutch type amyloidosis |
| SNOMED CT | 87555007 | Rubrospinal cerebellar peduncle syndrome |
| SNOMED CT | 110997000 | Idiopathic nonarteriosclerotic cerebrovascular calcification |
| SNOMED CT | 254775002 | Bregeat syndrome |
| SNOMED CT | 276221006 | Gubler syndrome |
| SNOMED CT | 724779000 | White matter disorder co-occurrent and due to cerebral autosomal dominant arteriopathy with subcortical infarcts and leucoencephalopathy |
| SNOMED CT | 778060000 | COL4A1-related familial vascular leucoencephalopathy |
| SNOMED CT | 783787000 | Retinal vasculopathy with cerebral leucoencephalopathy and systemic manifestations |
| SNOMED CT | 284861000119104 | Atherosclerosis of bilateral carotid arteries |
| SNOMED CT | 429241000124106 | Dissection of posterior cerebral artery |

Continued on next page

**TableS25 – continued from previous page**

| CodeSystem | ConceptCode | ConceptName |
|---|---|---|
| SNOMED CT | 429821000124103 | Embolism of posterior cerebral artery |
| SNOMED CT | 672441000119103 | Hemiplegia of nondominant side due to and following ischaemic cerebrovascular accident |
| SNOMED CT | 672461000119104 | Hemiplegia of dominant side due to and following ischaemic cerebrovascular accident |
| SNOMED CT | 15707961000119109 | Dissection of bilateral carotid arteries |
| SNOMED CT | 15710641000119100 | Dissection of bilateral vertebral arteries |
| SNOMED CT | 16026991000119107 | Cerebrovascular accident due to stenosis of left carotid artery |
| SNOMED CT | 1078223005 | Thrombosis of cerebral venous sinus due to and following surgical procedure |
| SNOMED CT | 16520041000119104 | Congenital fenestration of basilar artery |
| SNOMED CT | 16528221000119104 | Congenital duplication of anterior communicating artery |
| SNOMED CT | 16703711000119100 | Memory deficit due to and following embolic cerebrovascular accident |
| SNOMED CT | 16891111000119104 | Idiopathic ischemic stroke |
| SNOMED CT | 329501000119102 | Bilateral occlusion of middle cerebral arteries |
| SNOMED CT | 329541000119100 | Bilateral occlusion of anterior cerebral arteries |
| SNOMED CT | 595899961000119100 | Cerebrovascular accident of basal ganglia |
| SNOMED CT | 652287331000119104 | Cerebrovascular accident of brainstem |
| SNOMED CT | 881694631000119107 | Cerebrovascular accident of medulla oblongata |
| SNOMED CT | 915141931000119109 | Cerebrovascular accident due to embolism of basilar artery |
| SNOMED CT | 107557061000119108 | Cerebrovascular accident due to embolism of bilateral anterior cerebral arteries |
| SNOMED CT | 1142056004 | Neonatal ischemic stroke |
| SNOMED CT | 1153543002 | Occlusion of anterior cerebral artery |
| SNOMED CT | 1153544008 | Occlusion of right anterior cerebral artery |
| SNOMED CT | 1153545009 | Occlusion of left anterior cerebral artery |
| SNOMED CT | 1153546005 | Bilateral occlusion of posterior cerebral arteries |
| SNOMED CT | 1153607003 | Occlusion of right posterior communicating artery |
| SNOMED CT | 1153608008 | Occlusion of left posterior communicating artery |
| SNOMED CT | 1153611009 | Embolism of left anterior cerebral artery |
| SNOMED CT | 1153612002 | Embolism of right anterior cerebral artery |
| SNOMED CT | 1153630009 | Embolism of left carotid artery |
| SNOMED CT | 1153631008 | Embolism of right carotid artery |
| SNOMED CT | 1153632001 | Bilateral embolism of middle cerebral arteries |
| SNOMED CT | 1153633006 | Bilateral embolism of posterior cerebral arteries |
| SNOMED CT | 1153634000 | Bilateral embolism of anterior cerebral arteries |
| SNOMED CT | 1153638002 | Occlusion of bilateral cerebellar arteries |
| SNOMED CT | 1155688007 | Bilateral embolism of carotid arteries |
| SNOMED CT | 1155689004 | Bilateral embolism of cerebellar arteries |
| SNOMED CT | 1155697006 | Embolism of left vertebral artery |
| SNOMED CT | 1155698001 | Embolism of right vertebral artery |
| SNOMED CT | 1155699009 | Bilateral embolism of vertebral arteries |
| SNOMED CT | 1156016006 | Left posterior cerebral artery thrombosis |
| SNOMED CT | 1156017002 | Right posterior cerebral artery thrombosis |
| SNOMED CT | 1156018007 | Left cerebellar artery thrombosis |
| SNOMED CT | 1156019004 | Right cerebellar artery thrombosis |

Continued on next page

**TableS25 – continued from previous page**

| CodeSystem | ConceptCode | ConceptName |
|---|---|---|
| SNOMED CT | 1156027008 | Cerebral venous sinus thrombosis in pregnancy |
| SNOMED CT | 1156029006 | Cerebral venous sinus thrombosis in puerperium |
| SNOMED CT | 117776611000119102 | Cerebrovascular accident due to occlusion of left posterior communicating artery |
| SNOMED CT | 11842891000119102 | Right carotid artery injury |
| SNOMED CT | 11842931000119105 | Left carotid artery injury |
| SNOMED CT | 11844791000119100 | Right vertebral artery injury |
| SNOMED CT | 11844831000119106 | Left vertebral artery injury |
| SNOMED CT | 152148641000119104 | Cerebrovascular accident due to embolism of bilateral carotid arteries |
| SNOMED CT | 168747591000119109 | Cerebrovascular accident due to embolism of bilateral posterior cerebral arteries |
| SNOMED CT | 182960891000119101 | Cerebrovascular accident due to occlusion of left anterior cerebral artery |
| SNOMED CT | 188174841000119103 | Cerebrovascular accident due to occlusion of bilateral middle cerebral arteries |
| SNOMED CT | 239965291000119107 | Cerebrovascular accident due to occlusion of basilar artery |
| SNOMED CT | 251770561000119107 | Cerebrovascular accident due to embolism of left anterior cerebral artery |
| SNOMED CT | 346674811000119104 | Cerebrovascular accident due to occlusion of bilateral cerebellar arteries |
| SNOMED CT | 384430101000119103 | Cerebrovascular accident due to embolism of right carotid artery |
| SNOMED CT | 38595071000119104 | Cerebrovascular accident due to thrombosis of right posterior cerebral artery |
| SNOMED CT | 496369931000119104 | Cerebrovascular accident due to thrombosis of right cerebellar artery |
| SNOMED CT | 511452481000119102 | Cerebrovascular accident due to thrombosis of right vertebral artery |
| SNOMED CT | 517253051000119105 | Cerebrovascular accident due to embolism of left vertebral artery |
| SNOMED CT | 58173271000119101 | Cerebrovascular accident due to embolism of bilateral cerebellar arteries |
| SNOMED CT | 655081461000119101 | Cerebrovascular accident due to occlusion of right anterior cerebral artery |
| SNOMED CT | 720261501000119107 | Cerebrovascular accident due to occlusion of bilateral anterior cerebral arteries |
| SNOMED CT | 732330391000119107 | Cerebrovascular accident due to embolism of bilateral vertebral arteries |
| SNOMED CT | 759950981000119101 | Cerebrovascular accident due to thrombosis of left vertebral artery |
| SNOMED CT | 769023031000119104 | Cerebrovascular accident of thalamus |
| SNOMED CT | 806161651000119106 | Cerebrovascular accident due to embolism of right vertebral artery |
| SNOMED CT | 849488701000119104 | Cerebrovascular accident due to embolism of right anterior cerebral artery |
| SNOMED CT | 849579281000119106 | Cerebrovascular accident due to occlusion of right posterior communicating artery |

Continued on next page

**TableS25 – continued from previous page**

| CodeSystem | ConceptCode | ConceptName |
|---|---|---|
| SNOMED CT | 851365731000119106 | Cerebrovascular accident due to thrombosis of left posterior cerebral artery |
| SNOMED CT | 859422751000119101 | Cerebrovascular accident due to embolism of left carotid artery |
| SNOMED CT | 86553761000119103 | Cerebrovascular accident due to occlusion of bilateral posterior cerebral arteries |
| SNOMED CT | 898941951000119108 | Cerebrovascular accident due to occlusion of bilateral vertebral arteries |
| SNOMED CT | 939885431000119109 | Cerebrovascular accident due to embolism of bilateral middle cerebral arteries |
| SNOMED CT | 957319791000119104 | Cerebrovascular accident due to thrombosis of left cerebellar artery |
| Truveta | 1215926 | Cerebrovascular accident (CVA) associated with severe acute respiratory syndrome coronavirus 2 (SARS-CoV-2) infection |
| SNOMED CT | 1186724002 | HTRA1-related autosomal dominant cerebral small vessel disease |
| SNOMED CT | 1163536005 | Aspergillus mycotic cerebral aneurysm |
| SNOMED CT | 1204162009 | Asymptomatic stenosis of anterior cerebral artery |
| SNOMED CT | 1204164005 | Asymptomatic stenosis of basilar artery |
| SNOMED CT | 1204158003 | Asymptomatic stenosis of extracranial carotid artery |
| SNOMED CT | 1204188007 | Asymptomatic occlusion of extracranial vertebral artery |
| SNOMED CT | 1204160001 | Asymptomatic stenosis of extracranial vertebral artery |
| SNOMED CT | 1204156004 | Asymptomatic stenosis of intracranial carotid artery |
| SNOMED CT | 1204147007 | Asymptomatic stenosis of middle cerebral artery |
| SNOMED CT | 1204148002 | Asymptomatic stenosis of posterior cerebral artery |
| SNOMED CT | 1204128009 | Cholesterol atheroembolism of cerebrum |
| SNOMED CT | 1197429000 | Cathepsin A-related arteriopathy, strokes, leukoencephalopathy |
| SNOMED CT | 1204202004 | Infective occlusion of cerebral artery |
| SNOMED CT | 1231168008 | Malignant middle cerebral artery syndrome |
| SNOMED CT | 1197363004 | Pediatric arterial ischemic stroke |
| SNOMED CT | 1208841001 | Secondary moyamoya disease |
| SNOMED CT | 1208871009 | Transient ischemic attack co-occurrent with subarachnoid hemorrhage |
| SNOMED CT | 293671000119109 | Behavioral disturbance due to multi-infarct dementia |
| SNOMED CT | 1187538003 | Cerebral secondary systemic amyloid angiopathy |
| SNOMED CT | 1187126002 | ITM2B-related amyloidosis |
| SNOMED CT | 1186710001 | Leukoencephalopathy with calcifications and cysts |
| SNOMED CT | 1177015003 | Tuberculous cerebral arteritis |
| SNOMED CT | 1173997008 | Pontine autosomal dominant microangiopathy with leukoencephalopathy |
| SNOMED CT | 1163482004 | Hemorrhagic cerebral infarction caused by Aspergillus |

#### 5.5 Chronic kidney disease

Table S26: Concept codes used to identify if a patient was diagnosed with CKD.

| CodeSystem | ConceptCode | ConceptName |
|---|---|---|
| ICD10CM | D63.1 | Anemia in chronic kidney disease |
| ICD10CM | E08.22 | Diabetes mellitus due to underlying condition with diabetic chronic kidney disease |
| ICD10CM | E09.22 | Drug or chemical induced diabetes mellitus with diabetic chronic kidney disease |
| ICD10CM | E13.22 | Other specified diabetes mellitus with diabetic chronic kidney disease |
| ICD10CM | I12 | Hypertensive chronic kidney disease |
| ICD10CM | I12.0 | Hypertensive chronic kidney disease with stage 5 chronic kidney disease or end stage renal disease |
| ICD10CM | I12.9 | Hypertensive chronic kidney disease with stage 1 through stage 4 chronic kidney disease, or unspecified chronic kidney disease |
| ICD10CM | I13 | Hypertensive heart and chronic kidney disease |
| ICD10CM | I13.0 | Hypertensive heart and chronic kidney disease with heart failure and stage 1 through stage 4 chronic kidney disease, or unspecified chronic kidney disease |
| ICD10CM | I13.1 | Hypertensive heart and chronic kidney disease without heart failure |
| ICD10CM | I13.10 | Hypertensive heart and chronic kidney disease without heart failure, with stage 1 through stage 4 chronic kidney disease, or unspecified chronic kidney disease |
| ICD10CM | I13.11 | Hypertensive heart and chronic kidney disease without heart failure, with stage 5 chronic kidney disease, or end stage renal disease |
| ICD10CM | I13.2 | Hypertensive heart and chronic kidney disease with heart failure and with stage 5 chronic kidney disease, or end stage renal disease |
| ICD10CM | N18 | Chronic kidney disease (CKD) |
| ICD10CM | O10.211 | Pre-existing hypertensive chronic kidney disease complicating pregnancy, first trimester |
| ICD10CM | O10.212 | Pre-existing hypertensive chronic kidney disease complicating pregnancy, second trimester |
| ICD10CM | O10.213 | Pre-existing hypertensive chronic kidney disease complicating pregnancy, third trimester |
| ICD10CM | O10.219 | Pre-existing hypertensive chronic kidney disease complicating pregnancy, unspecified trimester |
| ICD10CM | O10.22 | Pre-existing hypertensive chronic kidney disease complicating childbirth |
| ICD10CM | O10.31 | Pre-existing hypertensive heart and chronic kidney disease complicating pregnancy |
| ICD10CM | O10.311 | Pre-existing hypertensive heart and chronic kidney disease complicating pregnancy, first trimester |
| ICD10CM | O10.312 | Pre-existing hypertensive heart and chronic kidney disease complicating pregnancy, second trimester |

Continued on next page

**TableS26 – continued from previous page**

| CodeSystem | ConceptCode | ConceptName |
|---|---|---|
| ICD10CM | O10.313 | Pre-existing hypertensive heart and chronic kidney disease complicating pregnancy, third trimester |
| ICD10CM | O10.319 | Pre-existing hypertensive heart and chronic kidney disease complicating pregnancy, unspecified trimester |
| ICD10CM | O10.32 | Pre-existing hypertensive heart and chronic kidney disease complicating childbirth |
| ICD10CM | O10.33 | Pre-existing hypertensive heart and chronic kidney disease complicating the puerperium |
| ICD10CM | Q61.2 | Polycystic kidney, adult type |
| ICD10CM | Q61.3 | Polycystic kidney, unspecified |
| ICD10CM | Q61.8 | Other cystic kidney diseases |
| ICD10CM | Z94.0 | Kidney transplant status |
| ICD10CM | N18.1 | Chronic kidney disease, stage 1 |
| ICD10CM | N18.2 | Chronic kidney disease, stage 2 (mild) |
| ICD10CM | N18.3 | Chronic kidney disease, stage 3 (moderate) |
| ICD10CM | N18.4 | Chronic kidney disease, stage 4 (severe) |
| ICD10CM | N18.5 | Chronic kidney disease, stage 5 |
| ICD10CM | N18.6 | End stage renal disease |
| ICD10CM | N18.9 | Chronic kidney disease, unspecified |
| ICD10CM | N18.30 | Chronic kidney disease, stage 3 unspecified |
| ICD10CM | N18.31 | Chronic kidney disease, stage 3a |
| ICD10CM | N18.32 | Chronic kidney disease, stage 3b |
| SNOMED CT | 709044004 | Chronic kidney disease |
| SNOMED CT | 425369003 | Chronic progressive renal failure |
| SNOMED CT | 431855005 | Chronic kidney disease stage 1 |
| SNOMED CT | 431856006 | Chronic kidney disease stage 2 |
| SNOMED CT | 431857002 | Chronic kidney disease stage 4 |
| SNOMED CT | 433144002 | Chronic kidney disease stage 3 |
| SNOMED CT | 433146000 | Chronic kidney disease stage 5 |
| SNOMED CT | 700378005 | Chronic kidney disease stage 3A |
| SNOMED CT | 700379002 | Chronic kidney disease stage 3B |
| SNOMED CT | 704667004 | Hypertension concurrent and due to end stage renal disease on dialysis |
| SNOMED CT | 707324008 | Anemia in end stage renal disease |
| SNOMED CT | 708975004 | Mixed renal osteodystrophy |
| SNOMED CT | 712487000 | End stage renal disease due to benign hypertension |
| SNOMED CT | 713313000 | Chronic kidney disease mineral and bone disorder |
| SNOMED CT | 714152005 | Chronic kidney disease stage 5 on dialysis |
| SNOMED CT | 714153000 | Chronic kidney disease stage 5 with transplant |
| SNOMED CT | 90688005 | Chronic renal failure |
| SNOMED CT | 236433006 | Acute-on-chronic renal failure |
| SNOMED CT | 236434000 | End stage renal failure untreated by renal replacement therapy |
| SNOMED CT | 236435004 | End stage renal failure on dialysis |
| SNOMED CT | 236436003 | End stage renal failure with renal transplant |
| SNOMED CT | 236552002 | Adynamic bone disease |
| SNOMED CT | 16726004 | Renal osteodystrophy |
| SNOMED CT | 46177005 | End-stage renal disease |

Continued on next page

**TableS26 – continued from previous page**

| CodeSystem | ConceptCode | ConceptName |
|---|---|---|
| SNOMED CT | 49708008 | Anemia of chronic renal failure |
| SNOMED CT | 57557005 | Chronic milk alkali syndrome |
| SNOMED CT | 722098007 | Chronic kidney disease following donor nephrectomy |
| SNOMED CT | 722149000 | Chronic kidney disease due to and following excision of neoplasm of kidney |
| SNOMED CT | 722150000 | Chronic kidney disease due to systemic infection |
| SNOMED CT | 722467000 | Chronic kidney disease due to traumatic loss of kidney |
| SNOMED CT | 723190009 | Chronic renal insufficiency |
| SNOMED CT | 776416004 | Hyperuricemia, pulmonary hypertension, renal failure, alkalosis syndrome |
| SNOMED CT | 711000119100 | Chronic kidney disease stage 5 due to type 2 diabetes mellitus |
| SNOMED CT | 721000119107 | Chronic kidney disease stage 4 due to type 2 diabetes mellitus |
| SNOMED CT | 731000119105 | Chronic kidney disease stage 3 due to type 2 diabetes mellitus |
| SNOMED CT | 741000119101 | Chronic kidney disease stage 2 due to type 2 diabetes mellitus |
| SNOMED CT | 751000119104 | Chronic kidney disease stage 1 due to type 2 diabetes mellitus |
| SNOMED CT | 771000119108 | Chronic kidney disease due to type 2 diabetes mellitus |
| SNOMED CT | 1801000119106 | Anemia, pre-end stage renal disease on erythropoietin protocol |
| SNOMED CT | 8501000119104 | Hypertensive heart and chronic kidney disease |
| SNOMED CT | 71421000119105 | Hypertension in chronic kidney disease due to type 2 diabetes mellitus |
| SNOMED CT | 71701000119105 | Hypertension in chronic kidney disease due to type 1 diabetes mellitus |
| SNOMED CT | 90721000119101 | Chronic kidney disease stage 1 due to type 1 diabetes mellitus |
| SNOMED CT | 90731000119103 | Chronic kidney disease stage 2 due to type 1 diabetes mellitus |
| SNOMED CT | 90741000119107 | Chronic kidney disease stage 3 due to type 1 diabetes mellitus |
| SNOMED CT | 90751000119109 | Chronic kidney disease stage 4 due to type 1 diabetes mellitus |
| SNOMED CT | 90761000119106 | Chronic kidney disease stage 5 due to type 1 diabetes mellitus |
| SNOMED CT | 90771000119100 | End stage renal disease on dialysis due to type 1 diabetes mellitus |
| SNOMED CT | 90791000119104 | End stage renal disease on dialysis due to type 2 diabetes mellitus |
| SNOMED CT | 96441000119101 | Chronic kidney disease due to type 1 diabetes mellitus |
| SNOMED CT | 96701000119107 | Hypertensive heart AND chronic kidney disease on dialysis |
| SNOMED CT | 96711000119105 | Hypertensive heart AND chronic kidney disease stage 5 |
| SNOMED CT | 96721000119103 | Hypertensive heart AND chronic kidney disease stage 4 |
| SNOMED CT | 96731000119100 | Hypertensive heart AND chronic kidney disease stage 3 |
| SNOMED CT | 96741000119109 | Hypertensive heart AND chronic kidney disease stage 2 |
| SNOMED CT | 96751000119106 | Hypertensive heart AND chronic kidney disease stage 1 |
| SNOMED CT | 104931000119100 | Chronic kidney disease due to hypertension |
| SNOMED CT | 111411000119103 | End stage renal disease due to hypertension |
| SNOMED CT | 117681000119102 | Chronic kidney disease stage 1 due to hypertension |
| SNOMED CT | 118781000119108 | Pre-existing hypertensive chronic kidney disease in mother complicating pregnancy |
| SNOMED CT | 120261000119101 | Benign hypertensive heart disease and chronic renal disease stage 5 |
| SNOMED CT | 127991000119101 | Hypertension concurrent and due to end stage renal disease on dialysis due to type 2 diabetes mellitus |
| SNOMED CT | 128001000119105 | Hypertension concurrent and due to end stage renal disease on dialysis due to type 1 diabetes mellitus |

Continued on next page

**TableS26 – continued from previous page**

| CodeSystem | ConceptCode | ConceptName |
|---|---|---|
| SNOMED CT | 129151000119102 | Chronic kidney disease stage 4 due to hypertension |
| SNOMED CT | 129161000119100 | Chronic kidney disease stage 5 due to hypertension |
| SNOMED CT | 129171000119106 | Chronic kidney disease stage 3 due to hypertension |
| SNOMED CT | 129181000119109 | Chronic kidney disease stage 2 due to hypertension |
| SNOMED CT | 140101000119109 | Hypertension in chronic kidney disease stage 5 due to type 2 diabetes mellitus |
| SNOMED CT | 140111000119107 | Hypertension in chronic kidney disease stage 4 due to type 2 diabetes mellitus |
| SNOMED CT | 140121000119100 | Hypertension in chronic kidney disease stage 3 due to type 2 diabetes mellitus |
| SNOMED CT | 140131000119102 | Hypertension in chronic kidney disease stage 2 due to type 2 diabetes mellitus |
| SNOMED CT | 153851000119106 | Malignant hypertensive chronic kidney disease stage 5 |
| SNOMED CT | 153891000119101 | End stage renal disease on dialysis due to hypertension |
| SNOMED CT | 284961000119106 | Chronic kidney disease due to benign hypertension |
| SNOMED CT | 284971000119100 | Chronic kidney disease stage 1 due to benign hypertension |
| SNOMED CT | 284981000119102 | Chronic kidney disease stage 2 due to benign hypertension |
| SNOMED CT | 284991000119104 | Chronic kidney disease stage 3 due to benign hypertension |
| SNOMED CT | 285001000119105 | Chronic kidney disease stage 4 due to benign hypertension |
| SNOMED CT | 285011000119108 | Chronic kidney disease stage 5 due to benign hypertension |
| SNOMED CT | 285041000119107 | Benign hypertensive heart disease and chronic renal disease stage 1 |
| SNOMED CT | 285061000119106 | Benign hypertensive heart disease and chronic renal disease stage 2 |
| SNOMED CT | 285081000119102 | Benign hypertensive heart disease and chronic renal disease stage 3 |
| SNOMED CT | 285101000119109 | Benign hypertensive heart disease and chronic renal disease stage 4 |
| SNOMED CT | 285831000119108 | Malignant hypertensive chronic kidney disease |
| SNOMED CT | 285841000119104 | Malignant hypertensive end stage renal disease |
| SNOMED CT | 285851000119102 | Malignant hypertensive chronic kidney disease stage 1 |
| SNOMED CT | 285861000119100 | Malignant hypertensive chronic kidney disease stage 2 |
| SNOMED CT | 285871000119106 | Malignant hypertensive chronic kidney disease stage 3 |
| SNOMED CT | 285881000119109 | Malignant hypertensive chronic kidney disease stage 4 |
| SNOMED CT | 285911000119109 | Malignant hypertensive heart disease and chronic renal disease stage 3 |
| SNOMED CT | 285921000119102 | Malignant hypertensive heart disease and chronic renal disease stage 4 |
| SNOMED CT | 286371000119107 | Malignant hypertensive end stage renal disease on dialysis |
| SNOMED CT | 434431000124103 | Hypertensive end stage renal disease |
| SNOMED CT | 449631000124102 | Benign hypertensive heart disease and chronic renal disease |
| SNOMED CT | 691421000119108 | Anemia co-occurrent and due to chronic kidney disease stage 3 |
| SNOMED CT | 10757401000119104 | Pre-existing hypertensive heart and chronic kidney disease in mother complicating childbirth |
| SNOMED CT | 10757481000119107 | Pre-existing hypertensive heart and chronic kidney disease in mother complicating pregnancy |

Continued on next page

**TableS26 – continued from previous page**

| CodeSystem | ConceptCode | ConceptName |
|---|---|---|
| SNOMED CT | 368421000119108 | Chronic kidney disease stage 1 due to drug induced diabetes mellitus |
| SNOMED CT | 368431000119106 | Chronic kidney disease stage 2 due to drug induced diabetes mellitus |
| SNOMED CT | 368441000119102 | Chronic kidney disease stage 3 due to drug induced diabetes mellitus |
| SNOMED CT | 368451000119100 | Chronic kidney disease stage 4 due to drug induced diabetes mellitus |
| SNOMED CT | 368461000119103 | Chronic kidney disease stage 5 due to drug induced diabetes mellitus |
| SNOMED CT | 368471000119109 | End stage renal disease on dialysis due to drug induced diabetes mellitus |
| SNOMED CT | 897308007 | Chronic kidney disease with osteoporosis |
| SNOMED CT | 897310009 | Renal osteodystrophy with high bone turnover |
| SNOMED CT | 897311008 | Renal osteodystrophy with normal bone turnover |
| SNOMED CT | 897312001 | Renal osteodystrophy with low bone turnover |
| SNOMED CT | 1217070004 | Renal osteodystrophy due to hyperparathyroidism |
| SNOMED CT | 1208934006 | Sagliker syndrome |

#### 5.6 Chronic obstructive pulmonary disease

Table S27: Concept codes used to identify if a patient was diagnosed with COPD.

| CodeSystem | ConceptCode | ConceptName |
|---|---|---|
| ICD10CM | J44.0 | Chronic obstructive pulmonary disease with (acute) lower respiratory infection |
| ICD10CM | J44.1 | Chronic obstructive pulmonary disease with (acute) exacerbation |
| ICD10CM | J44.9 | Chronic obstructive pulmonary disease, unspecified |
| ICD9CM | 493.20 | Chronic obstructive asthma, unspecified |
| ICD9CM | 493.21 | Chronic obstructive asthma with status asthmaticus |
| ICD9CM | 493.22 | Chronic obstructive asthma with (acute) exacerbation |
| ICD9CM | 496 | Chronic airway obstruction, not elsewhere classified |
| SNOMED CT | 285381006 | Acute infective exacerbation of chronic obstructive airways disease |
| SNOMED CT | 313296004 | Mild chronic obstructive pulmonary disease |
| SNOMED CT | 313297008 | Moderate chronic obstructive pulmonary disease |
| SNOMED CT | 313299006 | Severe chronic obstructive pulmonary disease |
| SNOMED CT | 135836000 | End stage chronic obstructive airways disease |
| SNOMED CT | 195951007 | Acute exacerbation of chronic obstructive airways disease |
| SNOMED CT | 196001008 | Chronic obstructive pulmonary disease with acute lower respiratory infection |
| SNOMED CT | 13645005 | Chronic obstructive lung disease |
| SNOMED CT | 266355005 | Bullous emphysema with collapse |

Continued on next page

**TableS27 – continued from previous page**

| CodeSystem | ConceptCode | ConceptName |
|---|---|---|
| SNOMED CT | 266356006 | Atrophic (senile) emphysema |
| SNOMED CT | 708030004 | Pulmonary emphysema co-occurrent with fibrosis of lung |
| SNOMED CT | 70756004 | Bronchial atresia with segmental pulmonary emphysema |
| SNOMED CT | 77690003 | Interstitial emphysema of lung |
| SNOMED CT | 86680006 | Ruptured emphysematous bleb of lung |
| SNOMED CT | 87433001 | Pulmonary emphysema |
| SNOMED CT | 195957006 | Chronic bullous emphysema |
| SNOMED CT | 195958001 | Segmental bullous emphysema |
| SNOMED CT | 195959009 | Zonal bullous emphysema |
| SNOMED CT | 196026004 | Chronic emphysema due to chemical fumes |
| SNOMED CT | 233675009 | Toxic emphysema |
| SNOMED CT | 233677001 | Scar emphysema |
| SNOMED CT | 4981000 | Panacinar emphysema |
| SNOMED CT | 16003001 | Giant bullous emphysema |
| SNOMED CT | 16846004 | Obstructive emphysema |
| SNOMED CT | 23958009 | Vanishing lung |
| SNOMED CT | 31898008 | Paraseptal emphysema |
| SNOMED CT | 33325001 | Compensatory emphysema |
| SNOMED CT | 47895001 | Congenital emphysema |
| SNOMED CT | 47938003 | Chronic obliterative bronchiolitis |
| SNOMED CT | 57686001 | Emphysematous bleb of lung |
| SNOMED CT | 60805002 | Hemolytic anemia with emphysema AND cutis laxa |
| SNOMED CT | 66987001 | Congenital lobar emphysema |
| SNOMED CT | 68328006 | Centriacinar emphysema |
| SNOMED CT | 762618008 | Bronchiolitis obliterans syndrome due to and after lung transplantation |
| SNOMED CT | 836477007 | Chronic emphysema due to vapor |
| SNOMED CT | 840350008 | Chronic obliterative bronchiolitis due to chemical fumes |
| SNOMED CT | 840351007 | Chronic obliterative bronchiolitis due to vapor |
| SNOMED CT | 1751000119100 | Acute exacerbation of chronic obstructive airways disease with asthma |
| SNOMED CT | 106001000119101 | Chronic obstructive lung disease co-occurrent with acute bronchitis |
| SNOMED CT | 10692761000119107 | Asthma-chronic obstructive pulmonary disease overlap syndrome |
| SNOMED CT | 233674008 | Pulmonary emphysema in alpha-1 primary immunodeficiency deficiency |
| SNOMED CT | 1010333003 | Emphysema of left lung |
| SNOMED CT | 1010334009 | Emphysema of right lung |
| SNOMED CT | 1177120001 | Bronchiolitis obliterans syndrome due to and following allogeneic stem cell transplant |

86 ## 5.7 Dementia

Table S28: Concept codes used to identify if a patient was diagnosed with dementia.

| CodeSystem | ConceptCode | ConceptName |
|---|---|---|
| RxNorm | 1100184 | donepezil hydrochloride 23 MG Oral Tablet |
| RxNorm | 1308569 | 24 HR rivastigmine 0.554 MG/HR Transdermal System |
| RxNorm | 1599803 | 24 HR donepezil hydrochloride 10 MG / memantine hydrochloride 28 MG Extended Release Oral Capsule |
| RxNorm | 1599805 | 24 HR donepezil hydrochloride 10 MG / memantine hydrochloride 14 MG Extended Release Oral Capsule |
| RxNorm | 1805420 | 24 HR donepezil hydrochloride 10 MG / memantine hydrochloride 21 MG Extended Release Oral Capsule |
| RxNorm | 1805425 | 24 HR donepezil hydrochloride 10 MG / memantine hydrochloride 7 MG Extended Release Oral Capsule |
| RxNorm | 310436 | galantamine 4 MG Oral Tablet |
| RxNorm | 310437 | galantamine 8 MG Oral Tablet |
| RxNorm | 312835 | rivastigmine 3 MG Oral Capsule |
| RxNorm | 312836 | rivastigmine 6 MG Oral Capsule |
| RxNorm | 314214 | rivastigmine 1.5 MG Oral Capsule |
| RxNorm | 314215 | rivastigmine 4.5 MG Oral Capsule |
| RxNorm | 579148 | galantamine 12 MG Oral Tablet |
| RxNorm | 725021 | 24 HR rivastigmine 0.192 MG/HR Transdermal System |
| RxNorm | 725023 | 24 HR rivastigmine 0.396 MG/HR Transdermal System |
| RxNorm | 860695 | 24 HR galantamine hydrobromide 16 MG Extended Release Oral Capsule |
| RxNorm | 860707 | 24 HR galantamine hydrobromide 24 MG Extended Release Oral Capsule |
| RxNorm | 860715 | 24 HR galantamine hydrobromide 8 MG Extended Release Oral Capsule |
| RxNorm | 860901 | galantamine hydrobromide 4 MG/ML Oral Solution |
| RxNorm | 996740 | memantine hydrochloride 2 MG/ML Oral Solution |
| RxNorm | 997220 | donepezil hydrochloride 10 MG Disintegrating Oral Tablet |
| RxNorm | 997223 | donepezil hydrochloride 10 MG Oral Tablet |
| RxNorm | 997226 | donepezil hydrochloride 5 MG Disintegrating Oral Tablet |
| RxNorm | 997229 | donepezil hydrochloride 5 MG Oral Tablet |
| RxNorm | 996561 | memantine hydrochloride 10 MG Oral Tablet |
| RxNorm | 996571 | memantine hydrochloride 5 MG Oral Tablet |
| RxNorm | 996594 | 24 HR memantine hydrochloride 14 MG Extended Release Oral Capsule |
| RxNorm | 996603 | 24 HR memantine hydrochloride 21 MG Extended Release Oral Capsule |
| RxNorm | 996609 | 24 HR memantine hydrochloride 28 MG Extended Release Oral Capsule |
| RxNorm | 996615 | 24 HR memantine hydrochloride 7 MG Extended Release Oral Capsule |
| RxNorm | 1100187 | donepezil hydrochloride 23 MG Oral Tablet [Aricept] |
| RxNorm | 1308571 | 24 HR rivastigmine 0.554 MG/HR Transdermal System [Exelon] |
| RxNorm | 1602588 | 24 HR donepezil hydrochloride 10 MG / memantine hydrochloride 14 MG Extended Release Oral Capsule [Namzaric] |

Continued on next page

**TableS28 – continued from previous page**

| CodeSystem | ConceptCode | ConceptName |
|---|---|---|
| RxNorm | 1602594 | 24 HR donepezil hydrochloride 10 MG / memantine hydrochloride 28 MG Extended Release Oral Capsule [Namzaric] |
| RxNorm | 1805422 | 24 HR donepezil hydrochloride 10 MG / memantine hydrochloride 21 MG Extended Release Oral Capsule [Namzaric] |
| RxNorm | 1805427 | 24 HR donepezil hydrochloride 10 MG / memantine hydrochloride 7 MG Extended Release Oral Capsule [Namzaric] |
| RxNorm | 1858970 | {7 (24 HR donepezil hydrochloride 10 MG / memantine hydrochloride 14 MG Extended Release Oral Capsule) / 7 (24 HR donepezil hydrochloride 10 MG / memantine hydrochloride 21 MG Extended Release Oral Capsule) / 7 (24 HR donepezil hydrochloride 10 MG / memantine hydrochloride 28 MG Extended Release Oral Capsule) / 7 (24 HR donepezil hydrochloride 10 MG / memantine hydrochloride 7 MG Extended Release Oral Capsule) } Pack |
| RxNorm | 1858971 | {7 (24 HR donepezil hydrochloride 10 MG / memantine hydrochloride 14 MG Extended Release Oral Capsule [Namzaric]) / 7 (24 HR donepezil hydrochloride 10 MG / memantine hydrochloride 21 MG Extended Release Oral Capsule [Namzaric]) / 7 (24 HR donepezil hydrochloride 10 MG / memantine hydrochloride 28 MG Extended Release Oral Capsule [Namzaric]) / 7 (24 HR donepezil hydrochloride 10 MG / memantine hydrochloride 7 MG Extended Release Oral Capsule [Namzaric]) } Pack [Namzaric Titration Pack] |
| RxNorm | 226665 | rivastigmine 1.5 MG Oral Capsule [Exelon] |
| RxNorm | 226666 | rivastigmine 3 MG Oral Capsule [Exelon] |
| RxNorm | 226667 | rivastigmine 4.5 MG Oral Capsule [Exelon] |
| RxNorm | 226668 | rivastigmine 6 MG Oral Capsule [Exelon] |
| RxNorm | 602734 | galantamine 12 MG Oral Tablet [Razadyne] |
| RxNorm | 602736 | galantamine 4 MG Oral Tablet [Razadyne] |
| RxNorm | 602737 | galantamine 8 MG Oral Tablet [Razadyne] |
| RxNorm | 725105 | 24 HR rivastigmine 0.396 MG/HR Transdermal System [Exelon] |
| RxNorm | 751302 | 24 HR rivastigmine 0.192 MG/HR Transdermal System [Exelon] |
| RxNorm | 860697 | 24 HR galantamine hydrobromide 16 MG Extended Release Oral Capsule [Razadyne] |
| RxNorm | 860709 | 24 HR galantamine hydrobromide 24 MG Extended Release Oral Capsule [Razadyne] |
| RxNorm | 860717 | 24 HR galantamine hydrobromide 8 MG Extended Release Oral Capsule [Razadyne] |
| RxNorm | 860903 | galantamine hydrobromide 4 MG/ML Oral Solution [Razadyne] |
| RxNorm | 996742 | memantine hydrochloride 2 MG/ML Oral Solution [Namenda] |
| RxNorm | 997222 | donepezil hydrochloride 10 MG Disintegrating Oral Tablet [Aricept] |
| RxNorm | 997224 | donepezil hydrochloride 10 MG Oral Tablet [Aricept] |
| RxNorm | 997228 | donepezil hydrochloride 5 MG Disintegrating Oral Tablet [Aricept] |
| RxNorm | 997230 | donepezil hydrochloride 5 MG Oral Tablet [Aricept] |
| RxNorm | 996563 | memantine hydrochloride 10 MG Oral Tablet [Namenda] |

Continued on next page

**TableS28 – continued from previous page**

| CodeSystem | ConceptCode | ConceptName |
|---|---|---|
| RxNorm | 996572 | {21 (memantine hydrochloride 10 MG Oral Tablet) / 28 (memantine hydrochloride 5 MG Oral Tablet) } Pack |
| RxNorm | 996574 | memantine hydrochloride 5 MG Oral Tablet [Namenda] |
| RxNorm | 996597 | 24 HR memantine hydrochloride 14 MG Extended Release Oral Capsule [Namenda] |
| RxNorm | 996605 | 24 HR memantine hydrochloride 21 MG Extended Release Oral Capsule [Namenda] |
| RxNorm | 996611 | 24 HR memantine hydrochloride 28 MG Extended Release Oral Capsule [Namenda] |
| RxNorm | 996617 | 24 HR memantine hydrochloride 7 MG Extended Release Oral Capsule [Namenda] |
| RxNorm | 996624 | {7 (24 HR memantine hydrochloride 14 MG Extended Release Oral Capsule) / 7 (24 HR memantine hydrochloride 21 MG Extended Release Oral Capsule) / 7 (24 HR memantine hydrochloride 28 MG Extended Release Oral Capsule) / 7 (24 HR memantine hydrochloride 7 MG Extended Release Oral Capsule) } Pack |
| RxNorm | 996633 | {7 (24 HR memantine hydrochloride 14 MG Extended Release Oral Capsule [Namenda]) / 7 (24 HR memantine hydrochloride 21 MG Extended Release Oral Capsule [Namenda]) / 7 (24 HR memantine hydrochloride 28 MG Extended Release Oral Capsule [Namenda]) / 7 (24 HR memantine hydrochloride 7 MG Extended Release Oral Capsule [Namenda]) } Pack [Namenda 28 Titration Pack] |
| RxNorm | 996634 | {21 (memantine hydrochloride 10 MG Oral Tablet [Namenda]) / 28 (memantine hydrochloride 5 MG Oral Tablet [Namenda]) } Pack [Namenda 49 Titration Pack] |
| RxNorm | 284556 | Galantamine 4 MG Oral Tablet [Reminyl] |
| RxNorm | 284557 | Galantamine 8 MG Oral Tablet [Reminyl] |
| RxNorm | 579150 | Galantamine 12 MG Oral Tablet [Reminyl] |
| ICD10CM | A52.17 | General paresis |
| ICD10CM | F01.50 | Vascular dementia without behavioral disturbance |
| ICD10CM | F01.51 | Vascular dementia with behavioral disturbance |
| ICD10CM | F02.80 | Dementia in other diseases classified elsewhere without behavioral disturbance |
| ICD10CM | F02.81 | Dementia in other diseases classified elsewhere with behavioral disturbance |
| ICD10CM | F03.90 | Unspecified dementia without behavioral disturbance |
| ICD10CM | F03.91 | Unspecified dementia with behavioral disturbance |
| ICD10CM | F05 | Delirium due to known physiological condition |
| ICD10CM | F06.8 | Other specified mental disorders due to known physiological condition |
| ICD10CM | G30.0 | Alzheimer's disease with early onset |
| ICD10CM | G30.1 | Alzheimer's disease with late onset |
| ICD10CM | G30.8 | Other Alzheimer's disease |
| ICD10CM | G30.9 | Alzheimer's disease, unspecified |
| ICD10CM | G31.01 | Pick's disease |

Continued on next page

**TableS28 – continued from previous page**

| CodeSystem | ConceptCode | ConceptName |
|---|---|---|
| ICD10CM | G31.09 | Other frontotemporal dementia |
| ICD10CM | G31.83 | Dementia with Lewy bodies |
| ICD9CM | 290.0 | Senile dementia, uncomplicated |
| ICD9CM | 290.10 | Presenile dementia, uncomplicated |
| ICD9CM | 290.11 | Presenile dementia with delirium |
| ICD9CM | 290.12 | Presenile dementia with delusional features |
| ICD9CM | 290.13 | Presenile dementia with depressive features |
| ICD9CM | 290.20 | Senile dementia with delusional features |
| ICD9CM | 290.21 | Senile dementia with depressive features |
| ICD9CM | 290.3 | Senile dementia with delirium |
| ICD9CM | 290.40 | Vascular dementia, uncomplicated |
| ICD9CM | 290.41 | Vascular dementia, with delirium |
| ICD9CM | 290.42 | Vascular dementia, with delusions |
| ICD9CM | 290.43 | Vascular dementia, with depressed mood |
| ICD9CM | 290.8 | Other specified senile psychotic conditions |
| ICD9CM | 290.9 | Unspecified senile psychotic condition |
| ICD9CM | 294.10 | Dementia in conditions classified elsewhere without behavioral disturbance |
| ICD9CM | 294.11 | Dementia in conditions classified elsewhere with behavioral disturbance |
| ICD9CM | 294.20 | Dementia, unspecified, without behavioral disturbance |
| ICD9CM | 294.21 | Dementia, unspecified, with behavioral disturbance |
| ICD9CM | 294.8 | Other persistent mental disorders due to conditions classified elsewhere |
| ICD9CM | 331.0 | Alzheimer's disease |
| ICD9CM | 331.11 | Pick's disease |
| ICD9CM | 331.19 | Other frontotemporal dementia |
| ICD9CM | 331.82 | Dementia with lewy bodies |
| ICD9CM | 94.1 | Psychiatric interviews, consultations, and evaluations |
| SNOMED CT | 278857002 | Dementia of frontal lobe type |
| SNOMED CT | 279982005 | Cerebral degeneration presenting primarily with dementia |
| SNOMED CT | 371024007 | Senile dementia with delusion |
| SNOMED CT | 371026009 | Senile dementia with psychosis |
| SNOMED CT | 429998004 | Vascular dementia |
| SNOMED CT | 70936005 | Multi-infarct dementia, uncomplicated |
| SNOMED CT | 82959004 | Dementia paralytica juvenilis |
| SNOMED CT | 90099008 | Subcortical leukoencephalopathy |
| SNOMED CT | 111480006 | Psychoactive substance-induced organic dementia |
| SNOMED CT | 191449005 | Uncomplicated senile dementia |
| SNOMED CT | 191451009 | Uncomplicated presenile dementia |
| SNOMED CT | 191452002 | Presenile dementia with delirium |
| SNOMED CT | 191454001 | Presenile dementia with paranoia |
| SNOMED CT | 191455000 | Presenile dementia with depression |
| SNOMED CT | 191457008 | Senile dementia with depressive or paranoid features |
| SNOMED CT | 191458003 | Senile dementia with paranoia |
| SNOMED CT | 191459006 | Senile dementia with depression |
| SNOMED CT | 191461002 | Senile dementia with delirium |

Continued on next page

**TableS28 – continued from previous page**

| CodeSystem | ConceptCode | ConceptName |
|---|---|---|
| SNOMED CT | 191463004 | Uncomplicated arteriosclerotic dementia |
| SNOMED CT | 191464005 | Arteriosclerotic dementia with delirium |
| SNOMED CT | 191465006 | Arteriosclerotic dementia with paranoia |
| SNOMED CT | 191466007 | Arteriosclerotic dementia with depression |
| SNOMED CT | 191493005 | Drug-induced dementia |
| SNOMED CT | 230270009 | Frontotemporal dementia |
| SNOMED CT | 230283005 | Punch drunk syndrome |
| SNOMED CT | 230286002 | Subcortical vascular dementia |
| SNOMED CT | 230287006 | Mixed cortical and subcortical vascular dementia |
| SNOMED CT | 230288001 | Semantic dementia |
| SNOMED CT | 281004 | Dementia associated with alcoholism |
| SNOMED CT | 10349009 | Multi-infarct dementia with delirium |
| SNOMED CT | 12348006 | Presenile dementia |
| SNOMED CT | 14070001 | Multi-infarct dementia with depression |
| SNOMED CT | 15662003 | Senile dementia |
| SNOMED CT | 25772007 | Multi-infarct dementia with delusions |
| SNOMED CT | 32875003 | Inhalant-induced persisting dementia |
| SNOMED CT | 51928006 | General paresis - neurosyphilis |
| SNOMED CT | 52448006 | Dementia |
| SNOMED CT | 54502004 | Primary degenerative dementia of the Alzheimer type, presenile onset, with delusions |
| SNOMED CT | 56267009 | Multi-infarct dementia |
| SNOMED CT | 59651006 | Sedative, hypnotic AND/OR anxiolytic-induced persisting dementia |
| SNOMED CT | 65096006 | Primary degenerative dementia of the Alzheimer type, presenile onset, with delirium |
| SNOMED CT | 4817008 | Dementia Alzheimers type, late onset with delirium |
| SNOMED CT | 6475002 | Dementia in Alzheimer's disease - type 2 |
| SNOMED CT | 10532003 | Alzheimers dementia, early onset, with depressed mood |
| SNOMED CT | 26852004 | Alzheimers dementia, late onset, with depressive mood |
| SNOMED CT | 55009008 | Alzheimers dementia, late onset, with delusions |
| SNOMED CT | 66108005 | Dementia in Alzheimer's disease - type 1 |
| SNOMED CT | 312991009 | Senile Lewy body dementia |
| SNOMED CT | 416780008 | Dementia in Alzheimer's disease with early onset |
| SNOMED CT | 420614009 | Organic dementia with AIDS (acquired immunodeficiency syndrome) |
| SNOMED CT | 421023003 | Presenile dementia with AIDS (acquired immunodeficiency syndrome) |
| SNOMED CT | 421529006 | Dementia with AIDS (acquired immunodeficiency syndrome) |
| SNOMED CT | 425390006 | Dementia in Parkinsons disease |
| SNOMED CT | 442344002 | Dementia due to Huntingtons disease |
| SNOMED CT | 268612007 | Senile and presenile organic psychotic conditions |
| SNOMED CT | 429458009 | Dementia due to Creutzfeldt Jakob disease |
| SNOMED CT | 698624003 | Dementia associated with cerebral lipidosis |
| SNOMED CT | 698625002 | Dementia associated with normal pressure hydrocephalus |
| SNOMED CT | 698626001 | Dementia associated with multiple sclerosis |
| SNOMED CT | 698687007 | Post-traumatic dementia with behavioral change |

Continued on next page

**TableS28 – continued from previous page**

| CodeSystem | ConceptCode | ConceptName |
|---|---|---|
| SNOMED CT | 698725008 | Dementia associated with neurosyphilis |
| SNOMED CT | 698726009 | Dementia associated with viral encephalitis |
| SNOMED CT | 698781002 | Dementia associated with cerebral anoxia |
| SNOMED CT | 698948009 | Vascular dementia in remission |
| SNOMED CT | 698949001 | Dementia in remission |
| SNOMED CT | 698954005 | Primary degenerative dementia of the Alzheimer type, senile onset in remission |
| SNOMED CT | 698955006 | Primary degenerative dementia of the Alzheimer type, presenile onset in remission |
| SNOMED CT | 702393003 | CHMP2B-related frontotemporal dementia |
| SNOMED CT | 713488003 | Presenile dementia co-occurrent with human immunodeficiency virus infection |
| SNOMED CT | 713844000 | Dementia co-occurrent with human immunodeficiency virus infection |
| SNOMED CT | 715737004 | Parkinsonism with dementia of Guadeloupe |
| SNOMED CT | 716667005 | Right temporal lobar atrophy |
| SNOMED CT | 716994006 | Behavioral variant of frontotemporal dementia |
| SNOMED CT | 191519005 | Dementia associated with another disease |
| SNOMED CT | 230274000 | Frontal lobe degeneration with motor neurone disease |
| SNOMED CT | 230281007 | Argyrophilic grain disease |
| SNOMED CT | 230282000 | Traumatic encephalopathy |
| SNOMED CT | 230285003 | Vascular dementia of acute onset |
| SNOMED CT | 230289009 | Patchy dementia |
| SNOMED CT | 9345005 | Dialysis dementia |
| SNOMED CT | 40425004 | Postconcussion syndrome |
| SNOMED CT | 42769004 | Diffuse Lewy body disease with spongiform cortical change |
| SNOMED CT | 722600006 | Non-amnestic Alzheimer disease |
| SNOMED CT | 722977005 | Dementia co-occurrent and due to neurocysticercosis |
| SNOMED CT | 722978000 | Dementia caused by toxin |
| SNOMED CT | 722979008 | Dementia due to metabolic abnormality |
| SNOMED CT | 722980006 | Dementia due to chromosomal anomaly |
| SNOMED CT | 723123001 | Ischemic vascular dementia |
| SNOMED CT | 723390000 | Rapidly progressive dementia |
| SNOMED CT | 724776007 | Dementia due to disorder of central nervous system |
| SNOMED CT | 724777003 | Dementia due to infectious disease |
| SNOMED CT | 724992007 | Epilepsy co-occurrent and due to dementia |
| SNOMED CT | 725898002 | Delirium co-occurrent with dementia |
| SNOMED CT | 733184002 | Dementia caused by heavy metal exposure |
| SNOMED CT | 733185001 | Dementia following injury caused by exposure to ionizing radiation |
| SNOMED CT | 733190003 | Dementia due to primary malignant neoplasm of brain |
| SNOMED CT | 733191004 | Dementia due to chronic subdural hematoma |
| SNOMED CT | 733192006 | Dementia due to herpes encephalitis |
| SNOMED CT | 733194007 | Dementia with Down syndrome |
| SNOMED CT | 762350007 | Dementia due to prion disease |
| SNOMED CT | 762351006 | Dementia due to and following injury of head |
| SNOMED CT | 762707000 | Subcortical dementia |

Continued on next page

**TableS28 – continued from previous page**

| CodeSystem | ConceptCode | ConceptName |
|---|---|---|
| SNOMED CT | 774069007 | PRKAR1B-related neurodegenerative dementia with intermediate filaments |
| SNOMED CT | 783161005 | ABri amyloidosis |
| SNOMED CT | 783258000 | ADan amyloidosis |
| SNOMED CT | 788898005 | Dementia caused by volatile inhalant |
| SNOMED CT | 788899002 | Dementia due to pellagra |
| SNOMED CT | 833326008 | Cortical vascular dementia |
| SNOMED CT | 838276009 | Amyotrophic lateral sclerosis, parkinsonism, dementia complex |
| SNOMED CT | 840464007 | Dementia due to carbon monoxide poisoning |
| SNOMED CT | 840465008 | Dementia due to iron deficiency |
| SNOMED CT | 1581000119101 | Dementia of the Alzheimer type with behavioral disturbance |
| SNOMED CT | 1591000119103 | Dementia with behavioral disturbance |
| SNOMED CT | 2421000119107 | Hallucinations co-occurrent and due to late onset dementia |
| SNOMED CT | 22381000119105 | Primary degenerative dementia |
| SNOMED CT | 31081000119101 | Presenile dementia with delusions |
| SNOMED CT | 79341000119107 | Mixed dementia |
| SNOMED CT | 82351000119105 | Altered behavior due to Pick's disease |
| SNOMED CT | 82371000119101 | Dementia due to multiple sclerosis with altered behavior |
| SNOMED CT | 82381000119103 | Epileptic dementia with behavioral disturbance |
| SNOMED CT | 97751000119108 | Altered behavior in Alzheimer's disease |
| SNOMED CT | 101421000119107 | Dementia due to Parkinson's disease |
| SNOMED CT | 105421000119105 | Early onset Alzheimer's disease with behavioral disturbance |
| SNOMED CT | 106021000119105 | Multi-infarct dementia due to atherosclerosis |
| SNOMED CT | 130121000119104 | Dementia due to Rett's syndrome |
| SNOMED CT | 135811000119107 | Lewy body dementia with behavioral disturbance |
| SNOMED CT | 141991000119109 | Delusions in Alzheimer's disease |
| SNOMED CT | 142001000119106 | Depressed mood in Alzheimer's disease |
| SNOMED CT | 142011000119109 | Alzheimer's disease co-occurrent with delirium |
| SNOMED CT | 288631000119104 | Vascular dementia with behavioral disturbance |
| SNOMED CT | 428051000124108 | Mild dementia |
| SNOMED CT | 428351000124105 | Severe dementia |
| SNOMED CT | 430771000124100 | Moderate dementia |
| SNOMED CT | 16219201000119101 | Behavioural disturbance co-occurrent and due to late onset Alzheimer dementia |
| SNOMED CT | 16276361000119109 | Vascular dementia without behavioral disturbance |
| SNOMED CT | 26929004 | Alzheimer dementia |
| SNOMED CT | 58756001 | Huntington disease |
| SNOMED CT | 80098002 | SDLT - Senile dementia of the Lewy body type |
| SNOMED CT | 230265002 | Familial Alzheimer disease of early onset |
| SNOMED CT | 230266001 | Non-familial Alzheimer disease of early onset |
| SNOMED CT | 230267005 | Familial Alzheimer disease of late onset |
| SNOMED CT | 230268000 | Non-familial Alzheimer disease of late onset |
| SNOMED CT | 230269008 | Focal Alzheimer disease |
| SNOMED CT | 230271008 | Pick disease with Pick bodies |
| SNOMED CT | 230272001 | Pick disease with Pick cells and no Pick bodies |
| SNOMED CT | 230280008 | Progressive aphasia in Alzheimer disease |
| SNOMED CT | 230299004 | Juvenile onset Huntington disease |

Continued on next page

**TableS28 – continued from previous page**

| CodeSystem | ConceptCode | ConceptName |
|---|---|---|
| SNOMED CT | 230300007 | Late onset Huntington disease |
| SNOMED CT | 230301006 | Akinetic-rigid form of Huntington disease |
| SNOMED CT | 416975007 | Dementia of the Alzheimers type, late onset |
| SNOMED CT | 733193001 | Dementia with progressive multifocal leucoencephalopathy |
| SNOMED CT | 21921000119103 | Dementia co-occurrent and due to Pick's disease |
| SNOMED CT | 82361000119107 | Altered behavior in dementia due to Huntington chorea |
| SNOMED CT | 429161000124103 | Primary degenerative dementia of the Alzheimer type, senile onset, with behavioral disturbance |
| SNOMED CT | 1148924004 | Dementia due to deficiency of folic acid |
| SNOMED CT | 1156789004 | Autosomal dominant Alzheimer disease due to mutation of amyloid precursor protein |
| SNOMED CT | 1156798001 | Autosomal dominant Alzheimer disease due to mutation of presenilin 2 |
| SNOMED CT | 1156800008 | Autosomal dominant Alzheimer disease due to mutation of presenilin 1 |
| SNOMED CT | 1186724002 | HTRA1-related autosomal dominant cerebral small vessel disease |
| SNOMED CT | 1187004001 | Chronic traumatic encephalopathy |
| SNOMED CT | 293671000119109 | Behavioral disturbance due to multi-infarct dementia |
| SNOMED CT | 1187126002 | ITM2B-related amyloidosis |
| SNOMED CT | 1186887004 | Dementia caused by manganese |
| SNOMED CT | 1186880002 | Dementia due to cobalamin deficiency |
| SNOMED CT | 1186879000 | Dementia due to thiamine deficiency |
| SNOMED CT | 1186877003 | Dementia due to vitamin E deficiency |
| SNOMED CT | 1186881003 | Dementia due to niacin deficiency |
| SNOMED CT | 1186883000 | Dementia due to nutritional deficiency |

#### 5.8 Depression

Table S29: Concept codes used to identify if a patient was diagnosed with depression.

| CodeSystem | ConceptCode | ConceptName |
|---|---|---|
| RxNorm | 1000048 | doxepin hydrochloride 10 MG Oral Capsule |
| RxNorm | 1000054 | doxepin hydrochloride 10 MG/ML Oral Solution |
| RxNorm | 1000058 | doxepin hydrochloride 100 MG Oral Capsule |
| RxNorm | 1000064 | doxepin hydrochloride 150 MG Oral Capsule |
| RxNorm | 1000070 | doxepin hydrochloride 25 MG Oral Capsule |
| RxNorm | 1000076 | doxepin hydrochloride 50 MG Oral Capsule |
| RxNorm | 1000097 | doxepin hydrochloride 75 MG Oral Capsule |
| RxNorm | 103968 | lamotrigine 100 MG Disintegrating Oral Tablet |
| RxNorm | 1086772 | vilazodone hydrochloride 10 MG Oral Tablet |
| RxNorm | 1086778 | vilazodone hydrochloride 20 MG Oral Tablet |
| RxNorm | 1086784 | vilazodone hydrochloride 40 MG Oral Tablet |

Continued on next page

**TableS29 – continued from previous page**

| CodeSystem | ConceptCode | ConceptName |
|---|---|---|
| RxNorm | 1086789 | {7 (vilazodone hydrochloride 10 MG Oral Tablet) / 7 (vilazodone hydrochloride 20 MG Oral Tablet) / 16 (vilazodone hydrochloride 40 MG Oral Tablet) } Pack |
| RxNorm | 1098608 | 24 HR lamotrigine 300 MG Extended Release Oral Tablet |
| RxNorm | 1098649 | nefazodone hydrochloride 100 MG Oral Tablet |
| RxNorm | 1098666 | nefazodone hydrochloride 150 MG Oral Tablet |
| RxNorm | 1098670 | nefazodone hydrochloride 200 MG Oral Tablet |
| RxNorm | 1098674 | nefazodone hydrochloride 250 MG Oral Tablet |
| RxNorm | 1098678 | nefazodone hydrochloride 50 MG Oral Tablet |
| RxNorm | 1099288 | desipramine hydrochloride 10 MG Oral Tablet |
| RxNorm | 1099292 | desipramine hydrochloride 100 MG Oral Tablet |
| RxNorm | 1099296 | desipramine hydrochloride 150 MG Oral Tablet |
| RxNorm | 1099300 | desipramine hydrochloride 25 MG Oral Tablet |
| RxNorm | 1099304 | desipramine hydrochloride 50 MG Oral Tablet |
| RxNorm | 1099316 | desipramine hydrochloride 75 MG Oral Tablet |
| RxNorm | 1146690 | 24 HR lamotrigine 250 MG Extended Release Oral Tablet |
| RxNorm | 1190110 | fluoxetine 60 MG Oral Tablet |
| RxNorm | 1232585 | 24 HR bupropion hydrochloride 450 MG Extended Release Oral Tablet |
| RxNorm | 1298857 | maprotiline hydrochloride 25 MG Oral Tablet |
| RxNorm | 1298861 | maprotiline hydrochloride 50 MG Oral Tablet |
| RxNorm | 1298870 | maprotiline hydrochloride 75 MG Oral Tablet |
| RxNorm | 1738483 | paroxetine hydrochloride 10 MG Oral Tablet |
| RxNorm | 1738495 | paroxetine hydrochloride 20 MG Oral Tablet |
| RxNorm | 1738503 | paroxetine hydrochloride 30 MG Oral Tablet |
| RxNorm | 1738511 | paroxetine hydrochloride 40 MG Oral Tablet |
| RxNorm | 1738515 | paroxetine mesylate 10 MG Oral Tablet |
| RxNorm | 1738519 | paroxetine mesylate 20 MG Oral Tablet |
| RxNorm | 1738523 | paroxetine mesylate 30 MG Oral Tablet |
| RxNorm | 1738527 | paroxetine mesylate 40 MG Oral Tablet |
| RxNorm | 1738803 | 24 HR paroxetine hydrochloride 12.5 MG Extended Release Oral Tablet |
| RxNorm | 1738805 | 24 HR paroxetine hydrochloride 25 MG Extended Release Oral Tablet |
| RxNorm | 1738807 | 24 HR paroxetine hydrochloride 37.5 MG Extended Release Oral Tablet |
| RxNorm | 198045 | nortriptyline 10 MG Oral Capsule |
| RxNorm | 198046 | nortriptyline 50 MG Oral Capsule |
| RxNorm | 198047 | nortriptyline 75 MG Oral Capsule |
| RxNorm | 197363 | amoxapine 100 MG Oral Tablet |
| RxNorm | 197364 | amoxapine 150 MG Oral Tablet |
| RxNorm | 197365 | amoxapine 25 MG Oral Tablet |
| RxNorm | 197366 | amoxapine 50 MG Oral Tablet |
| RxNorm | 198427 | lamotrigine 100 MG Oral Tablet |
| RxNorm | 198428 | lamotrigine 150 MG Oral Tablet |
| RxNorm | 198429 | lamotrigine 200 MG Oral Tablet |
| RxNorm | 198430 | lamotrigine 25 MG Disintegrating Oral Tablet |

Continued on next page

**TableS29 – continued from previous page**

| CodeSystem | ConceptCode | ConceptName |
|---|---|---|
| RxNorm | 200371 | citalopram 20 MG Oral Tablet |
| RxNorm | 248642 | fluoxetine 20 MG Oral Tablet |
| RxNorm | 252478 | lamotrigine 50 MG Disintegrating Oral Tablet |
| RxNorm | 252479 | lamotrigine 200 MG Disintegrating Oral Tablet |
| RxNorm | 310384 | fluoxetine 10 MG Oral Capsule |
| RxNorm | 310385 | fluoxetine 20 MG Oral Capsule |
| RxNorm | 310386 | fluoxetine 4 MG/ML Oral Solution |
| RxNorm | 283406 | mirtazapine 15 MG Disintegrating Oral Tablet |
| RxNorm | 283407 | mirtazapine 30 MG Disintegrating Oral Tablet |
| RxNorm | 309313 | citalopram 2 MG/ML Oral Solution |
| RxNorm | 309314 | citalopram 40 MG Oral Tablet |
| RxNorm | 283485 | mirtazapine 45 MG Disintegrating Oral Tablet |
| RxNorm | 282401 | lamotrigine 25 MG Oral Tablet |
| RxNorm | 313447 | tranylcypromine 10 MG Oral Tablet |
| RxNorm | 283672 | citalopram 10 MG Oral Tablet |
| RxNorm | 311725 | mirtazapine 15 MG Oral Tablet |
| RxNorm | 311726 | mirtazapine 45 MG Oral Tablet |
| RxNorm | 312938 | sertraline 100 MG Oral Tablet |
| RxNorm | 312940 | sertraline 25 MG Oral Tablet |
| RxNorm | 312941 | sertraline 50 MG Oral Tablet |
| RxNorm | 313496 | trimipramine 100 MG Oral Capsule |
| RxNorm | 313498 | trimipramine 25 MG Oral Capsule |
| RxNorm | 313499 | trimipramine 50 MG Oral Capsule |
| RxNorm | 313580 | venlafaxine 100 MG Oral Tablet |
| RxNorm | 313581 | 24 HR venlafaxine 150 MG Extended Release Oral Capsule |
| RxNorm | 313582 | venlafaxine 25 MG Oral Tablet |
| RxNorm | 313583 | 24 HR venlafaxine 37.5 MG Extended Release Oral Capsule |
| RxNorm | 313584 | venlafaxine 37.5 MG Oral Tablet |
| RxNorm | 313585 | 24 HR venlafaxine 75 MG Extended Release Oral Capsule |
| RxNorm | 313586 | venlafaxine 75 MG Oral Tablet |
| RxNorm | 312036 | nortriptyline 2 MG/ML Oral Solution |
| RxNorm | 314277 | venlafaxine 50 MG Oral Tablet |
| RxNorm | 312242 | paroxetine hydrochloride 2 MG/ML Oral Suspension |
| RxNorm | 313989 | fluoxetine 40 MG Oral Capsule |
| RxNorm | 313990 | fluoxetine 10 MG Oral Tablet |
| RxNorm | 313995 | fluoxetine 90 MG Delayed Release Oral Capsule |
| RxNorm | 312347 | phenelzine 15 MG Oral Tablet |
| RxNorm | 314111 | mirtazapine 30 MG Oral Tablet |
| RxNorm | 317136 | nortriptyline 25 MG Oral Capsule |
| RxNorm | 311264 | lamotrigine 25 MG Chewable Tablet |
| RxNorm | 311265 | lamotrigine 5 MG Chewable Tablet |
| RxNorm | 349010 | lamotrigine 2 MG Chewable Tablet |
| RxNorm | 349332 | escitalopram 10 MG Oral Tablet |
| RxNorm | 351249 | escitalopram 5 MG Oral Tablet |
| RxNorm | 351250 | escitalopram 20 MG Oral Tablet |
| RxNorm | 351285 | escitalopram 1 MG/ML Oral Solution |
| RxNorm | 410503 | 5-hydroxytryptophan 100 MG Oral Capsule |

Continued on next page

**TableS29 – continued from previous page**

| CodeSystem | ConceptCode | ConceptName |
|---|---|---|
| RxNorm | 476809 | mirtazapine 7.5 MG Oral Tablet |
| RxNorm | 485514 | 5-hydroxytryptophan 50 MG Oral Capsule |
| RxNorm | 596926 | duloxetine 20 MG Delayed Release Oral Capsule |
| RxNorm | 596930 | duloxetine 30 MG Delayed Release Oral Capsule |
| RxNorm | 596934 | duloxetine 60 MG Delayed Release Oral Capsule |
| RxNorm | 616402 | duloxetine 40 MG Delayed Release Oral Capsule |
| RxNorm | 751563 | {7 (lamotrigine 100 MG Oral Tablet) / 42 (lamotrigine 25 MG Oral Tablet) } Pack |
| RxNorm | 753451 | {14 (lamotrigine 100 MG Oral Tablet) / 84 (lamotrigine 25 MG Oral Tablet) } Pack |
| RxNorm | 790264 | 24 HR desvenlafaxine 100 MG Extended Release Oral Tablet |
| RxNorm | 790288 | 24 HR desvenlafaxine 50 MG Extended Release Oral Tablet |
| RxNorm | 751139 | {35 (lamotrigine 25 MG Oral Tablet) } Pack |
| RxNorm | 808744 | 24 HR venlafaxine 150 MG Extended Release Oral Tablet |
| RxNorm | 808748 | 24 HR venlafaxine 225 MG Extended Release Oral Tablet |
| RxNorm | 808751 | 24 HR venlafaxine 37.5 MG Extended Release Oral Tablet |
| RxNorm | 808753 | 24 HR venlafaxine 75 MG Extended Release Oral Tablet |
| RxNorm | 851750 | {7 (lamotrigine 100 MG Disintegrating Oral Tablet) / 14 (lamotrigine 25 MG Disintegrating Oral Tablet) / 14 (lamotrigine 50 MG Disintegrating Oral Tablet) } Pack |
| RxNorm | 851752 | {14 (lamotrigine 100 MG Disintegrating Oral Tablet) / 42 (lamotrigine 50 MG Disintegrating Oral Tablet) } Pack |
| RxNorm | 850087 | 24 HR lamotrigine 100 MG Extended Release Oral Tablet |
| RxNorm | 850091 | 24 HR lamotrigine 50 MG Extended Release Oral Tablet |
| RxNorm | 851748 | {21 (lamotrigine 25 MG Disintegrating Oral Tablet) / 7 (lamotrigine 50 MG Disintegrating Oral Tablet) } Pack |
| RxNorm | 857297 | clomipramine hydrochloride 25 MG Oral Capsule |
| RxNorm | 857301 | clomipramine hydrochloride 50 MG Oral Capsule |
| RxNorm | 857305 | clomipramine hydrochloride 75 MG Oral Capsule |
| RxNorm | 856364 | trazodone hydrochloride 150 MG Oral Tablet |
| RxNorm | 856369 | trazodone hydrochloride 300 MG Oral Tablet |
| RxNorm | 856373 | trazodone hydrochloride 100 MG Oral Tablet |
| RxNorm | 856377 | trazodone hydrochloride 50 MG Oral Tablet |
| RxNorm | 835564 | imipramine hydrochloride 25 MG Oral Tablet |
| RxNorm | 835568 | imipramine hydrochloride 50 MG Oral Tablet |
| RxNorm | 835572 | imipramine pamoate 75 MG Oral Capsule |
| RxNorm | 835577 | imipramine pamoate 150 MG Oral Capsule |
| RxNorm | 835589 | imipramine pamoate 125 MG Oral Capsule |
| RxNorm | 835591 | imipramine pamoate 100 MG Oral Capsule |
| RxNorm | 835593 | imipramine hydrochloride 10 MG Oral Tablet |
| RxNorm | 856706 | amitriptyline hydrochloride 10 MG / perphenazine 2 MG Oral Tablet |
| RxNorm | 856720 | amitriptyline hydrochloride 10 MG / perphenazine 4 MG Oral Tablet |
| RxNorm | 856762 | amitriptyline hydrochloride 100 MG Oral Tablet |
| RxNorm | 856769 | amitriptyline hydrochloride 12.5 MG / chlordiazepoxide 5 MG Oral Tablet |

Continued on next page

**TableS29 – continued from previous page**

| CodeSystem | ConceptCode | ConceptName |
|---|---|---|
| RxNorm | 856773 | amitriptyline hydrochloride 150 MG Oral Tablet |
| RxNorm | 856783 | amitriptyline hydrochloride 10 MG Oral Tablet |
| RxNorm | 856792 | amitriptyline hydrochloride 25 MG / chlordiazepoxide 10 MG Oral Tablet |
| RxNorm | 856797 | amitriptyline hydrochloride 25 MG / perphenazine 2 MG Oral Tablet |
| RxNorm | 856825 | amitriptyline hydrochloride 25 MG / perphenazine 4 MG Oral Tablet |
| RxNorm | 856834 | amitriptyline hydrochloride 25 MG Oral Tablet |
| RxNorm | 856840 | amitriptyline hydrochloride 50 MG / perphenazine 4 MG Oral Tablet |
| RxNorm | 856845 | amitriptyline hydrochloride 50 MG Oral Tablet |
| RxNorm | 856853 | amitriptyline hydrochloride 75 MG Oral Tablet |
| RxNorm | 859186 | selegiline hydrochloride 5 MG Oral Capsule |
| RxNorm | 859190 | selegiline hydrochloride 1.25 MG Disintegrating Oral Tablet |
| RxNorm | 859193 | selegiline hydrochloride 5 MG Oral Tablet |
| RxNorm | 861064 | sertraline 20 MG/ML Oral Solution |
| RxNorm | 865206 | 24 HR selegiline 0.25 MG/HR Transdermal System |
| RxNorm | 865210 | 24 HR selegiline 0.375 MG/HR Transdermal System |
| RxNorm | 865214 | 24 HR selegiline 0.5 MG/HR Transdermal System |
| RxNorm | 900156 | 24 HR lamotrigine 200 MG Extended Release Oral Tablet |
| RxNorm | 900164 | 24 HR lamotrigine 25 MG Extended Release Oral Tablet |
| RxNorm | 900865 | {14 (24 HR lamotrigine 100 MG Extended Release Oral Tablet) / 7 (24 HR lamotrigine 200 MG Extended Release Oral Tablet) / 14 (24 HR lamotrigine 50 MG Extended Release Oral Tablet) } Pack |
| RxNorm | 900890 | {7 (24 HR lamotrigine 100 MG Extended Release Oral Tablet) / 14 (24 HR lamotrigine 25 MG Extended Release Oral Tablet) / 14 (24 HR lamotrigine 50 MG Extended Release Oral Tablet) } Pack |
| RxNorm | 905168 | protriptyline hydrochloride 10 MG Oral Tablet |
| RxNorm | 905172 | protriptyline hydrochloride 5 MG Oral Tablet |
| RxNorm | 900983 | {21 (24 HR lamotrigine 25 MG Extended Release Oral Tablet) / 7 (24 HR lamotrigine 50 MG Extended Release Oral Tablet) } Pack |
| RxNorm | 903873 | 24 HR fluvoxamine maleate 100 MG Extended Release Oral Capsule |
| RxNorm | 903879 | 24 HR fluvoxamine maleate 150 MG Extended Release Oral Capsule |
| RxNorm | 903884 | fluvoxamine maleate 100 MG Oral Tablet |
| RxNorm | 903887 | fluvoxamine maleate 25 MG Oral Tablet |
| RxNorm | 903891 | fluvoxamine maleate 50 MG Oral Tablet |
| RxNorm | 966787 | doxepin 3 MG Oral Tablet |
| RxNorm | 966793 | doxepin 6 MG Oral Tablet |
| RxNorm | 898697 | 24 HR trazodone hydrochloride 150 MG Extended Release Oral Tablet |

Continued on next page

**TableS29 – continued from previous page**

| CodeSystem | ConceptCode | ConceptName |
|---|---|---|
| RxNorm | 898704 | 24 HR trazodone hydrochloride 300 MG Extended Release Oral Tablet |
| RxNorm | 993503 | 12 HR bupropion hydrochloride 100 MG Extended Release Oral Tablet |
| RxNorm | 993518 | 12 HR bupropion hydrochloride 150 MG Extended Release Oral Tablet |
| RxNorm | 993536 | 12 HR bupropion hydrochloride 200 MG Extended Release Oral Tablet |
| RxNorm | 993541 | 24 HR bupropion hydrochloride 150 MG Extended Release Oral Tablet |
| RxNorm | 993550 | 24 HR bupropion hydrobromide 174 MG Extended Release Oral Tablet |
| RxNorm | 993557 | 24 HR bupropion hydrochloride 300 MG Extended Release Oral Tablet |
| RxNorm | 993567 | 24 HR bupropion hydrobromide 348 MG Extended Release Oral Tablet |
| RxNorm | 993681 | 24 HR bupropion hydrobromide 522 MG Extended Release Oral Tablet |
| RxNorm | 993687 | bupropion hydrochloride 100 MG Oral Tablet |
| RxNorm | 993691 | bupropion hydrochloride 75 MG Oral Tablet |
| RxNorm | 1000050 | doxepin hydrochloride 10 MG Oral Capsule [Adapin] |
| RxNorm | 1000060 | doxepin hydrochloride 100 MG Oral Capsule [Adapin] |
| RxNorm | 1000052 | doxepin hydrochloride 10 MG Oral Capsule [Sinequan] |
| RxNorm | 1000056 | doxepin hydrochloride 10 MG/ML Oral Solution [Sinequan] |
| RxNorm | 1000062 | doxepin hydrochloride 100 MG Oral Capsule [Sinequan] |
| RxNorm | 1000066 | doxepin hydrochloride 150 MG Oral Capsule [Adapin] |
| RxNorm | 1000068 | doxepin hydrochloride 150 MG Oral Capsule [Sinequan] |
| RxNorm | 1000072 | doxepin hydrochloride 25 MG Oral Capsule [Adapin] |
| RxNorm | 1000074 | doxepin hydrochloride 25 MG Oral Capsule [Sinequan] |
| RxNorm | 1000078 | doxepin hydrochloride 50 MG Oral Capsule [Adapin] |
| RxNorm | 1000081 | doxepin hydrochloride 50 MG Oral Capsule [Sinequan] |
| RxNorm | 1000099 | doxepin hydrochloride 75 MG Oral Capsule [Adapin] |
| RxNorm | 1000101 | doxepin hydrochloride 75 MG Oral Capsule [Sinequan] |
| RxNorm | 102550 | imipramine hydrochloride 10 MG Oral Tablet [Pramimil] |
| RxNorm | 102551 | imipramine hydrochloride 25 MG Oral Tablet [Pramimil] |
| RxNorm | 108782 | lamotrigine 200 MG Oral Tablet [Lamictal] |
| RxNorm | 105018 | lamotrigine 100 MG Oral Tablet [Lamictal] |
| RxNorm | 105019 | lamotrigine 25 MG Oral Tablet [Lamictal] |
| RxNorm | 108886 | citalopram 20 MG Oral Tablet [Cipramil] |
| RxNorm | 1086776 | vilazodone hydrochloride 10 MG Oral Tablet [Viibryd] |
| RxNorm | 1086780 | vilazodone hydrochloride 20 MG Oral Tablet [Viibryd] |
| RxNorm | 1086786 | vilazodone hydrochloride 40 MG Oral Tablet [Viibryd] |
| RxNorm | 1086790 | {7 (vilazodone hydrochloride 10 MG Oral Tablet [Viibryd]) / 7 (vilazodone hydrochloride 20 MG Oral Tablet [Viibryd]) / 16 (vilazodone hydrochloride 40 MG Oral Tablet [Viibryd]) } Pack [Viibryd 10/20/40 30 Day Pack] |

Continued on next page

**TableS29 – continued from previous page**

| CodeSystem | ConceptCode | ConceptName |
|---|---|---|
| RxNorm | 1098610 | 24 HR lamotrigine 300 MG Extended Release Oral Tablet [Lamictal] |
| RxNorm | 1099398 | desipramine hydrochloride 25 MG Oral Tablet [Pertofrane] |
| RxNorm | 1098651 | nefazodone hydrochloride 100 MG Oral Tablet [Serzone] |
| RxNorm | 1098668 | nefazodone hydrochloride 150 MG Oral Tablet [Serzone] |
| RxNorm | 1098672 | nefazodone hydrochloride 200 MG Oral Tablet [Serzone] |
| RxNorm | 1098676 | nefazodone hydrochloride 250 MG Oral Tablet [Serzone] |
| RxNorm | 1098680 | nefazodone hydrochloride 50 MG Oral Tablet [Serzone] |
| RxNorm | 1098681 | {14 (nefazodone hydrochloride 100 MG Oral Tablet) / 14 (nefazodone hydrochloride 200 MG Oral Tablet) / 14 (nefazodone hydrochloride 50 MG Oral Tablet) } Pack |
| RxNorm | 1098682 | {14 (nefazodone hydrochloride 100 MG Oral Tablet [Serzone]) / 14 (nefazodone hydrochloride 200 MG Oral Tablet [Serzone]) / 14 (nefazodone hydrochloride 50 MG Oral Tablet [Serzone]) } Pack [Serzone Starter Pack] |
| RxNorm | 1099645 | {7 (vilazodone hydrochloride 10 MG Oral Tablet) / 7 (vilazodone hydrochloride 20 MG Oral Tablet) } Pack |
| RxNorm | 1099646 | {7 (vilazodone hydrochloride 10 MG Oral Tablet [Viibryd]) / 7 (vilazodone hydrochloride 20 MG Oral Tablet [Viibryd]) } Pack [Viibryd 14] |
| RxNorm | 1099290 | desipramine hydrochloride 10 MG Oral Tablet [Norpramin] |
| RxNorm | 1099294 | desipramine hydrochloride 100 MG Oral Tablet [Norpramin] |
| RxNorm | 1099298 | desipramine hydrochloride 150 MG Oral Tablet [Norpramin] |
| RxNorm | 1099302 | desipramine hydrochloride 25 MG Oral Tablet [Norpramin] |
| RxNorm | 1099306 | desipramine hydrochloride 50 MG Oral Tablet [Norpramin] |
| RxNorm | 1099318 | desipramine hydrochloride 75 MG Oral Tablet [Norpramin] |
| RxNorm | 104824 | trazodone hydrochloride 150 MG Oral Tablet [Molipaxin] |
| RxNorm | 104836 | phenelzine 15 MG Oral Tablet [Nardil] |
| RxNorm | 104838 | tranylcypromine 10 MG Oral Tablet [Parnate] |
| RxNorm | 1100363 | nortriptyline 10 MG Oral Capsule [Aventyl] |
| RxNorm | 1100366 | nortriptyline 2 MG/ML Oral Solution [Aventyl] |
| RxNorm | 1100384 | nortriptyline 25 MG Oral Capsule [Aventyl] |
| RxNorm | 104849 | fluoxetine 20 MG Oral Capsule [Prozac] |
| RxNorm | 104850 | fluoxetine 4 MG/ML Oral Solution [Prozac] |
| RxNorm | 106834 | trimipramine 50 MG Oral Capsule [Surmontil] |
| RxNorm | 1146692 | 24 HR lamotrigine 250 MG Extended Release Oral Tablet [Lamictal] |
| RxNorm | 1232591 | 24 HR bupropion hydrochloride 450 MG Extended Release Oral Tablet [Forfivo] |
| RxNorm | 1298859 | maprotiline hydrochloride 25 MG Oral Tablet [Ludiomil] |
| RxNorm | 1298863 | maprotiline hydrochloride 50 MG Oral Tablet [Ludiomil] |
| RxNorm | 1298872 | maprotiline hydrochloride 75 MG Oral Tablet [Ludiomil] |
| RxNorm | 141945 | protriptyline hydrochloride 5 MG Oral Tablet [Concordin] |
| RxNorm | 141946 | protriptyline hydrochloride 10 MG Oral Tablet [Concordin] |
| RxNorm | 1435480 | 24 HR desvenlafaxine 100 MG Extended Release Oral Tablet [Khedezla] |

Continued on next page

**TableS29 – continued from previous page**

| CodeSystem | ConceptCode | ConceptName |
|---|---|---|
| RxNorm | 1435483 | 24 HR desvenlafaxine 50 MG Extended Release Oral Tablet [Khedezla] |
| RxNorm | 151256 | selegiline hydrochloride 5 MG Oral Tablet [Vivapryl] |
| RxNorm | 151281 | selegiline hydrochloride 5 MG Oral Tablet [Stilline] |
| RxNorm | 153503 | citalopram 10 MG Oral Tablet [Cipramil] |
| RxNorm | 152723 | selegiline hydrochloride 5 MG Oral Tablet [Centrapryl] |
| RxNorm | 1653469 | {7 (vilazodone hydrochloride 10 MG Oral Tablet) / 23 (vilazodone hydrochloride 20 MG Oral Tablet) } Pack |
| RxNorm | 1653470 | {7 (vilazodone hydrochloride 10 MG Oral Tablet [Viibryd]) / 23 (vilazodone hydrochloride 20 MG Oral Tablet [Viibryd]) } Pack [Viibryd Starter Pack 10/20 30 Day Pack] |
| RxNorm | 1652069 | duloxetine 40 MG Delayed Release Oral Capsule [Irenka] |
| RxNorm | 1738804 | 24 HR paroxetine hydrochloride 12.5 MG Extended Release Oral Tablet [Paxil] |
| RxNorm | 1738806 | 24 HR paroxetine hydrochloride 25 MG Extended Release Oral Tablet [Paxil] |
| RxNorm | 1738808 | 24 HR paroxetine hydrochloride 37.5 MG Extended Release Oral Tablet [Paxil] |
| RxNorm | 2001506 | lamotrigine 150 MG Oral Tablet [Subvenite] |
| RxNorm | 2001508 | lamotrigine 25 MG Oral Tablet [Subvenite] |
| RxNorm | 201239 | lamotrigine 5 MG Chewable Tablet [Lamictal] |
| RxNorm | 201240 | lamotrigine 25 MG Chewable Tablet [Lamictal] |
| RxNorm | 2003796 | lamotrigine 100 MG Oral Tablet [Subvenite] |
| RxNorm | 2003797 | {14 (lamotrigine 100 MG Oral Tablet [Subvenite]) / 84 (lamotrigine 25 MG Oral Tablet [Subvenite]) } Pack [Subvenite Green (for Patients Taking Carbamazepine, Phenytoin, Phenobarbital, or Primidone and Not Taking Valproate)] |
| RxNorm | 2003798 | {7 (lamotrigine 100 MG Oral Tablet [Subvenite]) / 42 (lamotrigine 25 MG Oral Tablet [Subvenite]) } Pack [Subvenite Orange (for Patients Not Taking Carbamazepine, Phenytoin, Phenobarbital, Primidone, or Valproate)] |
| RxNorm | 2003799 | {35 (lamotrigine 25 MG Oral Tablet [Subvenite]) } Pack [Subvenite Blue (for Patients Taking Valproate)] |
| RxNorm | 2003801 | lamotrigine 200 MG Oral Tablet [Subvenite] |
| RxNorm | 201692 | amitriptyline hydrochloride 10 MG Oral Tablet [Domical] |
| RxNorm | 201694 | amitriptyline hydrochloride 25 MG Oral Tablet [Domical] |
| RxNorm | 201696 | amitriptyline hydrochloride 50 MG Oral Tablet [Domical] |
| RxNorm | 205535 | fluoxetine 10 MG Oral Capsule [Prozac] |
| RxNorm | 207350 | paroxetine hydrochloride 30 MG Oral Tablet [Paxil] |
| RxNorm | 207349 | paroxetine hydrochloride 20 MG Oral Tablet [Paxil] |
| RxNorm | 206201 | lamotrigine 150 MG Oral Tablet [Lamictal] |
| RxNorm | 205696 | amoxapine 100 MG Oral Tablet [Asendin] |
| RxNorm | 205703 | amoxapine 150 MG Oral Tablet [Asendin] |
| RxNorm | 205706 | amoxapine 25 MG Oral Tablet [Asendin] |
| RxNorm | 205709 | amoxapine 50 MG Oral Tablet [Asendin] |
| RxNorm | 209329 | nortriptyline 10 MG Oral Capsule [Pamelor] |
| RxNorm | 209339 | nortriptyline 25 MG Oral Capsule [Pamelor] |

Continued on next page

**TableS29 – continued from previous page**

| CodeSystem | ConceptCode | ConceptName |
|---|---|---|
| RxNorm | 209350 | nortriptyline 50 MG Oral Capsule [Pamelor] |
| RxNorm | 208645 | trimipramine 100 MG Oral Capsule [Surmontil] |
| RxNorm | 208646 | trimipramine 25 MG Oral Capsule [Surmontil] |
| RxNorm | 209391 | nortriptyline 75 MG Oral Capsule [Pamelor] |
| RxNorm | 208848 | venlafaxine 100 MG Oral Tablet [Effexor] |
| RxNorm | 208849 | venlafaxine 25 MG Oral Tablet [Effexor] |
| RxNorm | 208850 | venlafaxine 37.5 MG Oral Tablet [Effexor] |
| RxNorm | 208851 | venlafaxine 50 MG Oral Tablet [Effexor] |
| RxNorm | 208853 | venlafaxine 75 MG Oral Tablet [Effexor] |
| RxNorm | 208149 | sertraline 100 MG Oral Tablet [Zoloft] |
| RxNorm | 208161 | sertraline 50 MG Oral Tablet [Zoloft] |
| RxNorm | 209464 | nortriptyline 2 MG/ML Oral Solution [Pamelor] |
| RxNorm | 211322 | mirtazapine 15 MG Oral Tablet [Remeron] |
| RxNorm | 211323 | mirtazapine 30 MG Oral Tablet [Remeron] |
| RxNorm | 211699 | paroxetine hydrochloride 10 MG Oral Tablet [Paxil] |
| RxNorm | 211700 | paroxetine hydrochloride 40 MG Oral Tablet [Paxil] |
| RxNorm | 212233 | sertraline 25 MG Oral Tablet [Zoloft] |
| RxNorm | 213291 | paroxetine hydrochloride 2 MG/ML Oral Suspension [Paxil] |
| RxNorm | 213344 | citalopram 20 MG Oral Tablet [Celexa] |
| RxNorm | 213345 | citalopram 40 MG Oral Tablet [Celexa] |
| RxNorm | 226376 | citalopram 40 MG Oral Tablet [Cipramil] |
| RxNorm | 261135 | mirtazapine 45 MG Oral Tablet [Remeron] |
| RxNorm | 284591 | citalopram 10 MG Oral Tablet [Celexa] |
| RxNorm | 261282 | fluoxetine 10 MG Oral Tablet [Prozac] |
| RxNorm | 261287 | fluoxetine 40 MG Oral Capsule [Prozac] |
| RxNorm | 261342 | citalopram 2 MG/ML Oral Solution [Celexa] |
| RxNorm | 352004 | fluoxetine 20 MG Oral Tablet [Rapiflux] |
| RxNorm | 352272 | escitalopram 10 MG Oral Tablet [Lexapro] |
| RxNorm | 352273 | escitalopram 20 MG Oral Tablet [Lexapro] |
| RxNorm | 404408 | escitalopram 5 MG Oral Tablet [Lexapro] |
| RxNorm | 404420 | escitalopram 1 MG/ML Oral Solution [Lexapro] |
| RxNorm | 540848 | citalopram 20 MG Oral Tablet [Citalopraam] |
| RxNorm | 541660 | paroxetine mesylate 10 MG Oral Tablet [Pexeva] |
| RxNorm | 541662 | paroxetine mesylate 20 MG Oral Tablet [Pexeva] |
| RxNorm | 541664 | paroxetine mesylate 30 MG Oral Tablet [Pexeva] |
| RxNorm | 541666 | paroxetine mesylate 40 MG Oral Tablet [Pexeva] |
| RxNorm | 542426 | lamotrigine 2 MG Chewable Tablet [Lamictal] |
| RxNorm | 583239 | nortriptyline 25 MG Oral Capsule [Notriptyline] |
| RxNorm | 598032 | fluoxetine 90 MG Delayed Release Oral Capsule [Prozac] |
| RxNorm | 596928 | duloxetine 20 MG Delayed Release Oral Capsule [Cymbalta] |
| RxNorm | 596932 | duloxetine 30 MG Delayed Release Oral Capsule [Cymbalta] |
| RxNorm | 615186 | duloxetine 60 MG Delayed Release Oral Capsule [Cymbalta] |
| RxNorm | 647325 | fluoxetine 10 MG Oral Capsule [Sarafem] |
| RxNorm | 647442 | fluoxetine 20 MG Oral Capsule [Sarafem] |
| RxNorm | 647555 | fluoxetine 10 MG Oral Tablet [Sarafem] |
| RxNorm | 647556 | fluoxetine 20 MG Oral Tablet [Sarafem] |
| RxNorm | 752869 | mirtazapine 15 MG Disintegrating Oral Tablet [Remeron] |

Continued on next page

**TableS29 – continued from previous page**

| CodeSystem | ConceptCode | ConceptName |
|---|---|---|
| RxNorm | 752870 | mirtazapine 30 MG Disintegrating Oral Tablet [Remeron] |
| RxNorm | 752871 | mirtazapine 45 MG Disintegrating Oral Tablet [Remeron] |
| RxNorm | 729929 | 24 HR venlafaxine 75 MG Extended Release Oral Capsule [Effexor] |
| RxNorm | 729931 | 24 HR venlafaxine 150 MG Extended Release Oral Capsule [Effexor] |
| RxNorm | 729932 | 24 HR venlafaxine 37.5 MG Extended Release Oral Capsule [Effexor] |
| RxNorm | 763188 | {7 (24 HR venlafaxine 37.5 MG Extended Release Oral Capsule) / 7 (24 HR venlafaxine 75 MG Extended Release Oral Capsule) } Pack |
| RxNorm | 763189 | {7 (24 HR venlafaxine 37.5 MG Extended Release Oral Capsule [Effexor]) / 7 (24 HR venlafaxine 75 MG Extended Release Oral Capsule [Effexor]) } Pack [Effexor XR Starter Kit] |
| RxNorm | 790267 | 24 HR desvenlafaxine succinate 100 MG Extended Release Oral Tablet [Pristiq] |
| RxNorm | 790290 | 24 HR desvenlafaxine succinate 50 MG Extended Release Oral Tablet [Pristiq] |
| RxNorm | 799026 | fluoxetine 10 MG Oral Capsule [Selfemra] |
| RxNorm | 799059 | fluoxetine 20 MG Oral Capsule [Selfemra] |
| RxNorm | 795772 | {35 (lamotrigine 25 MG Oral Tablet [Lamictal]) } Pack [Lamictal Blue (For Patients Taking Valproate)] |
| RxNorm | 795774 | {7 (lamotrigine 100 MG Oral Tablet [Lamictal]) / 42 (lamotrigine 25 MG Oral Tablet [Lamictal]) } Pack [Lamictal Orange (For Patients Not Taking Carbamazepine, Phenytoin, Phenobarbital, Primidone, or Rifampin and Not Taking Valproate)] |
| RxNorm | 795778 | {14 (lamotrigine 100 MG Oral Tablet [Lamictal]) / 84 (lamotrigine 25 MG Oral Tablet [Lamictal]) } Pack [Lamictal Green (For Patients Taking Carbamazepine, Phenytoin, Phenobarbital, Primidone, or Rifampin and Not Taking Valproate)] |
| RxNorm | 751140 | {5 (lamotrigine 100 MG Oral Tablet) / 42 (lamotrigine 25 MG Oral Tablet) } Pack |
| RxNorm | 851751 | {7 (lamotrigine 100 MG Disintegrating Oral Tablet [Lamictal]) / 14 (lamotrigine 25 MG Disintegrating Oral Tablet [Lamictal]) / 14 (lamotrigine 50 MG Disintegrating Oral Tablet [Lamictal]) } Pack [Lamictal ODT Orange Patient Titration Kit (For Patients Not Taking Enzyme-Inducing Drugs or Valproate)] |
| RxNorm | 851753 | {14 (lamotrigine 100 MG Disintegrating Oral Tablet [Lamictal]) / 42 (lamotrigine 50 MG Disintegrating Oral Tablet [Lamictal]) } Pack [Lamictal ODT Green Titration Kit (for Patients Taking Enzyme-Inducing Drugs and Not Taking Valproate)] |
| RxNorm | 849049 | lamotrigine 100 MG Disintegrating Oral Tablet [Lamictal] |
| RxNorm | 849050 | lamotrigine 200 MG Disintegrating Oral Tablet [Lamictal] |
| RxNorm | 849051 | lamotrigine 25 MG Disintegrating Oral Tablet [Lamictal] |
| RxNorm | 849052 | lamotrigine 50 MG Disintegrating Oral Tablet [Lamictal] |
| RxNorm | 834025 | {10 (lamotrigine 25 MG Oral Tablet) } Pack |

Continued on next page

**TableS29 – continued from previous page**

| CodeSystem | ConceptCode | ConceptName |
|---|---|---|
| RxNorm | 850097 | {14 (24 HR lamotrigine 100 MG Extended Release Oral Tablet) / 7 (24 HR lamotrigine 200 MG Extended Release Oral Capsule) / 14 (24 HR lamotrigine 50 MG Extended Release Oral Tablet) } Pack |
| RxNorm | 851749 | {21 (lamotrigine 25 MG Disintegrating Oral Tablet [Lamictal]) / 7 (lamotrigine 50 MG Disintegrating Oral Tablet [Lamictal]) } Pack [Lamictal ODT Blue Patient Titration Kit (for Patients Taking Valproate)] |
| RxNorm | 857299 | clomipramine hydrochloride 25 MG Oral Capsule [Anafranil] |
| RxNorm | 857303 | clomipramine hydrochloride 50 MG Oral Capsule [Anafranil] |
| RxNorm | 857307 | clomipramine hydrochloride 75 MG Oral Capsule [Anafranil] |
| RxNorm | 856366 | trazodone hydrochloride 150 MG Oral Tablet [Desyrel] |
| RxNorm | 856371 | trazodone hydrochloride 300 MG Oral Tablet [Desyrel] |
| RxNorm | 856375 | trazodone hydrochloride 100 MG Oral Tablet [Desyrel] |
| RxNorm | 856379 | trazodone hydrochloride 50 MG Oral Tablet [Desyrel] |
| RxNorm | 835566 | imipramine hydrochloride 25 MG Oral Tablet [Tofranil] |
| RxNorm | 835570 | imipramine hydrochloride 50 MG Oral Tablet [Tofranil] |
| RxNorm | 835574 | imipramine pamoate 75 MG Oral Capsule [Tofranil-PM] |
| RxNorm | 835579 | imipramine pamoate 150 MG Oral Capsule [Tofranil-PM] |
| RxNorm | 835596 | imipramine hydrochloride 10 MG Oral Tablet [Tofranil] |
| RxNorm | 835598 | imipramine pamoate 100 MG Oral Capsule [Tofranil-PM] |
| RxNorm | 835600 | imipramine pamoate 125 MG Oral Capsule [Tofranil-PM] |
| RxNorm | 856708 | amitriptyline hydrochloride 10 MG / perphenazine 2 MG Oral Tablet [Duo-Vil] |
| RxNorm | 856710 | amitriptyline hydrochloride 10 MG / perphenazine 2 MG Oral Tablet [Etrafon] |
| RxNorm | 856717 | amitriptyline hydrochloride 10 MG / perphenazine 2 MG Oral Tablet [Triavil] |
| RxNorm | 856722 | amitriptyline hydrochloride 10 MG / perphenazine 4 MG Oral Tablet [Duo-Vil] |
| RxNorm | 856755 | amitriptyline hydrochloride 10 MG / perphenazine 4 MG Oral Tablet [Etrafon] |
| RxNorm | 856760 | amitriptyline hydrochloride 10 MG / perphenazine 4 MG Oral Tablet [Triavil] |
| RxNorm | 856764 | amitriptyline hydrochloride 100 MG Oral Tablet [Elavil] |
| RxNorm | 856766 | amitriptyline hydrochloride 100 MG Oral Tablet [Endep] |
| RxNorm | 856775 | amitriptyline hydrochloride 150 MG Oral Tablet [Elavil] |
| RxNorm | 856777 | amitriptyline hydrochloride 150 MG Oral Tablet [Endep] |
| RxNorm | 856781 | amitriptyline hydrochloride 12.5 MG / chlordiazepoxide 5 MG Oral Tablet [Limbitrol] |
| RxNorm | 856785 | amitriptyline hydrochloride 10 MG Oral Tablet [Endep] |
| RxNorm | 856796 | amitriptyline hydrochloride 25 MG / chlordiazepoxide 10 MG Oral Tablet [Limbitrol] |
| RxNorm | 856799 | amitriptyline hydrochloride 25 MG / perphenazine 2 MG Oral Tablet [Duo-Vil] |
| RxNorm | 856806 | amitriptyline hydrochloride 25 MG / perphenazine 2 MG Oral Tablet [Etrafon] |

Continued on next page

**TableS29 – continued from previous page**

| CodeSystem | ConceptCode | ConceptName |
|---|---|---|
| RxNorm | 856810 | amitriptyline hydrochloride 25 MG / perphenazine 2 MG Oral Tablet [Triavil] |
| RxNorm | 856812 | amitriptyline hydrochloride 10 MG Oral Tablet [Elavil] |
| RxNorm | 856829 | amitriptyline hydrochloride 25 MG / perphenazine 4 MG Oral Tablet [Etrafon] |
| RxNorm | 856833 | amitriptyline hydrochloride 25 MG / perphenazine 4 MG Oral Tablet [Triavil] |
| RxNorm | 856836 | amitriptyline hydrochloride 25 MG Oral Tablet [Elavil] |
| RxNorm | 856838 | amitriptyline hydrochloride 25 MG Oral Tablet [Endep] |
| RxNorm | 856844 | amitriptyline hydrochloride 50 MG / perphenazine 4 MG Oral Tablet [Triavil] |
| RxNorm | 856847 | amitriptyline hydrochloride 50 MG Oral Tablet [Elavil] |
| RxNorm | 856849 | amitriptyline hydrochloride 50 MG Oral Tablet [Endep] |
| RxNorm | 856851 | amitriptyline hydrochloride 50 MG Oral Tablet [Vanatrip] |
| RxNorm | 856855 | amitriptyline hydrochloride 75 MG Oral Tablet [Elavil] |
| RxNorm | 856857 | amitriptyline hydrochloride 75 MG Oral Tablet [Endep] |
| RxNorm | 859188 | selegiline hydrochloride 5 MG Oral Capsule [Eldepryl] |
| RxNorm | 859192 | selegiline hydrochloride 1.25 MG Disintegrating Oral Tablet [Zelapar] |
| RxNorm | 859195 | selegiline hydrochloride 5 MG Oral Tablet [Atapryl] |
| RxNorm | 859200 | selegiline hydrochloride 5 MG Oral Tablet [Carbex] |
| RxNorm | 861066 | sertraline 20 MG/ML Oral Solution [Zoloft] |
| RxNorm | 897065 | selegiline hydrochloride 5 MG Oral Tablet [Anipryl] |
| RxNorm | 865208 | 24 HR selegiline 0.25 MG/HR Transdermal System [Emsam] |
| RxNorm | 865212 | 24 HR selegiline 0.375 MG/HR Transdermal System [Emsam] |
| RxNorm | 865216 | 24 HR selegiline 0.5 MG/HR Transdermal System [Emsam] |
| RxNorm | 865217 | selegiline hydrochloride 5 MG Oral Tablet [Eldepryl] |
| RxNorm | 900145 | 24 HR lamotrigine 100 MG Extended Release Oral Tablet [Lamictal] |
| RxNorm | 898706 | 24 HR trazodone hydrochloride 300 MG Extended Release Oral Tablet [Oleptro] |
| RxNorm | 900157 | 24 HR lamotrigine 200 MG Extended Release Oral Tablet [Lamictal] |
| RxNorm | 900165 | 24 HR lamotrigine 25 MG Extended Release Oral Tablet [Lamictal] |
| RxNorm | 900169 | 24 HR lamotrigine 50 MG Extended Release Oral Tablet [Lamictal] |
| RxNorm | 900866 | {14 (24 HR lamotrigine 100 MG Extended Release Oral Tablet [Lamictal]) / 7 (24 HR lamotrigine 200 MG Extended Release Oral Tablet [Lamictal]) / 14 (24 HR lamotrigine 50 MG Extended Release Oral Tablet [Lamictal]) } Pack [Lamictal XR Green Patient Titration Kit (for Patients Taking Carbamazepine, Phenytoin, Phenobarbital, or Primidone, and Not Taking Valproate)] |

Continued on next page

**TableS29 – continued from previous page**

| CodeSystem | ConceptCode | ConceptName |
|---|---|---|
| RxNorm | 900891 | {7 (24 HR lamotrigine 100 MG Extended Release Oral Tablet [Lamictal]) / 14 (24 HR lamotrigine 25 MG Extended Release Oral Tablet [Lamictal]) / 14 (24 HR lamotrigine 50 MG Extended Release Oral Tablet [Lamictal]) } Pack [Lamictal XR Orange Patient Titration Kit (for Patients Not Taking Carbamazepine, Phenytoin, Phenobarbital, Primidone, or Valproate)] |
| RxNorm | 905170 | protriptyline hydrochloride 10 MG Oral Tablet [Vivactil] |
| RxNorm | 905174 | protriptyline hydrochloride 5 MG Oral Tablet [Vivactil] |
| RxNorm | 900984 | {21 (24 HR lamotrigine 25 MG Extended Release Oral Tablet [Lamictal]) / 7 (24 HR lamotrigine 50 MG Extended Release Oral Tablet [Lamictal]) } Pack [Lamictal XR Blue Patient Titration Kit (for Patients Taking Valproate)] |
| RxNorm | 903875 | 24 HR fluvoxamine maleate 100 MG Extended Release Oral Capsule [Luvox] |
| RxNorm | 903881 | 24 HR fluvoxamine maleate 150 MG Extended Release Oral Capsule [Luvox] |
| RxNorm | 903885 | fluvoxamine maleate 100 MG Oral Tablet [Luvox] |
| RxNorm | 903889 | fluvoxamine maleate 25 MG Oral Tablet [Luvox] |
| RxNorm | 903893 | fluvoxamine maleate 50 MG Oral Tablet [Luvox] |
| RxNorm | 966791 | doxepin 3 MG Oral Tablet [Silenor] |
| RxNorm | 966795 | doxepin 6 MG Oral Tablet [Silenor] |
| RxNorm | 898701 | 24 HR trazodone hydrochloride 150 MG Extended Release Oral Tablet [Oleptro] |
| RxNorm | 993505 | 12 HR bupropion hydrochloride 100 MG Extended Release Oral Tablet [Budeprion] |
| RxNorm | 993511 | 12 HR bupropion hydrochloride 100 MG Extended Release Oral Tablet [Wellbutrin] |
| RxNorm | 993520 | 12 HR bupropion hydrochloride 150 MG Extended Release Oral Tablet [Budeprion] |
| RxNorm | 993524 | Smoking Cessation 12 HR bupropion hydrochloride 150 MG Extended Release Oral Tablet [Buproban] |
| RxNorm | 993528 | 12 HR bupropion hydrochloride 150 MG Extended Release Oral Tablet [Wellbutrin] |
| RxNorm | 993537 | 12 HR bupropion hydrochloride 200 MG Extended Release Oral Tablet [Wellbutrin] |
| RxNorm | 993542 | 24 HR bupropion hydrochloride 150 MG Extended Release Oral Tablet [Budeprion] |
| RxNorm | 993545 | 24 HR bupropion hydrochloride 150 MG Extended Release Oral Tablet [Wellbutrin] |
| RxNorm | 993552 | 24 HR bupropion hydrobromide 174 MG Extended Release Oral Tablet [Aplenzin] |
| RxNorm | 993559 | 24 HR bupropion hydrochloride 300 MG Extended Release Oral Tablet [Budeprion] |
| RxNorm | 993564 | 24 HR bupropion hydrochloride 300 MG Extended Release Oral Tablet [Wellbutrin] |
| RxNorm | 993569 | 24 HR bupropion hydrobromide 348 MG Extended Release Oral Tablet [Aplenzin] |

Continued on next page

**TableS29 – continued from previous page**

| CodeSystem | ConceptCode | ConceptName |
|---|---|---|
| RxNorm | 993683 | 24 HR bupropion hydrobromide 522 MG Extended Release Oral Tablet [Aplenzin] |
| RxNorm | 993688 | bupropion hydrochloride 100 MG Oral Tablet [Wellbutrin] |
| RxNorm | 993693 | bupropion hydrochloride 75 MG Oral Tablet [Wellbutrin] |
| RxNorm | 993954 | Smoking Cessation 12 HR bupropion hydrochloride 150 MG Extended Release Oral Tablet [Zyban] |
| RxNorm | 351962 | Fluoxetine 90 MG Enteric Coated Capsule [Prozac Weekly] |
| RxNorm | 209463 | Nortriptyline 2 MG/ML Oral Solution [Aventyl HCl] |
| RxNorm | 209328 | Nortriptyline 10 MG Oral Capsule [Aventyl HCl] |
| RxNorm | 209338 | Nortriptyline 25 MG Oral Capsule [Aventyl HCl] |
| RxNorm | 284535 | Mirtazapine 15 MG Disintegrating Tablet [Remeron SolTab] |
| RxNorm | 284536 | Mirtazapine 30 MG Disintegrating Tablet [Remeron SolTab] |
| RxNorm | 213095 | venlafaxine 37.5 MG 24 Hour Extended Release Capsule [Effexor XR] |
| RxNorm | 351990 | Mirtazapine 45 MG Disintegrating Tablet [Remeron SolTab] |
| RxNorm | 213099 | venlafaxine 75 MG 24 Hour Extended Release Capsule [Effexor XR] |
| RxNorm | 544236 | Mirtazapine 45 MG Oral Tablet [Mirtazdine] |
| RxNorm | 213094 | venlafaxine 150 MG 24 Hour Extended Release Capsule [Effexor XR] |
| RxNorm | 542424 | lamotrigine 2 MG Chewable Tablet [Itmd Lamictal Cd] |
| RxNorm | 629007 | Fluoxetine 20 MG Oral Tablet [Prozac] |
| RxNorm | 753538 | {14 (lamotrigine 100 MG Oral Tablet) / 84 (lamotrigine 25 MG Oral Tablet) } Pack [Lamictal Green (For Patients Taking Carbamazepine, Phenytoin, Primidone, Phenobarbital, or Rifampin and Not Taking Valproate)] |
| RxNorm | 753537 | {7 (lamotrigine 100 MG Oral Tablet) / 42 (lamotrigine 25 MG Oral Tablet) } Pack [Lamictal Orange (For Patients Not Taking Carbamazepine, Phenytoin, Phenobarbital, Primidone, or Rifampin and Not Taking Valproate)] |
| RxNorm | 753448 | {35 (lamotrigine 25 MG Oral Tablet) } Pack [Lamictal Blue (For Patients Taking Valproate)] |
| ICD10CM | F01.51 | Vascular dementia with behavioral disturbance |
| ICD10CM | F32.0 | Major depressive disorder, single episode, mild |
| ICD10CM | F32.1 | Major depressive disorder, single episode, moderate |
| ICD10CM | F32.2 | Major depressive disorder, single episode, severe without psychotic features |
| ICD10CM | F32.3 | Major depressive disorder, single episode, severe with psychotic features |
| ICD10CM | F32.4 | Major depressive disorder, single episode, in partial remission |
| ICD10CM | F32.5 | Major depressive disorder, single episode, in full remission |
| ICD10CM | F32.89 | Other specified depressive episodes |
| ICD10CM | F32.9 | Major depressive disorder, single episode, unspecified |
| ICD10CM | F33.0 | Major depressive disorder, recurrent, mild |
| ICD10CM | F33.1 | Major depressive disorder, recurrent, moderate |
| ICD10CM | F33.2 | Major depressive disorder, recurrent severe without psychotic features |

Continued on next page

**TableS29 – continued from previous page**

| CodeSystem | ConceptCode | ConceptName |
|---|---|---|
| ICD10CM | F33.3 | Major depressive disorder, recurrent, severe with psychotic symptoms |
| ICD10CM | F33.40 | Major depressive disorder, recurrent, in remission, unspecified |
| ICD10CM | F33.41 | Major depressive disorder, recurrent, in partial remission |
| ICD10CM | F33.42 | Major depressive disorder, recurrent, in full remission |
| ICD10CM | F33.8 | Other recurrent depressive disorders |
| ICD10CM | F33.9 | Major depressive disorder, recurrent, unspecified |
| ICD10CM | F34.1 | Dysthymic disorder |
| ICD10CM | F34.81 | Disruptive mood dysregulation disorder |
| ICD10CM | F34.89 | Other specified persistent mood disorders |
| ICD10CM | F43.21 | Adjustment disorder with depressed mood |
| ICD10CM | F43.23 | Adjustment disorder with mixed anxiety and depressed mood |
| ICD10CM | F53.0 | Postpartum depression |
| ICD10CM | F53.1 | Puerperal psychosis |
| ICD9CM | 290.13 | Presenile dementia with depressive features |
| ICD9CM | 290.21 | Senile dementia with depressive features |
| ICD9CM | 290.43 | Vascular dementia, with depressed mood |
| ICD9CM | 296.20 | Major depressive affective disorder, single episode, unspecified |
| ICD9CM | 296.21 | Major depressive affective disorder, single episode, mild |
| ICD9CM | 296.22 | Major depressive affective disorder, single episode, moderate |
| ICD9CM | 296.23 | Major depressive affective disorder, single episode, severe, without mention of psychotic behavior |
| ICD9CM | 296.24 | Major depressive affective disorder, single episode, severe, specified as with psychotic behavior |
| ICD9CM | 296.25 | Major depressive affective disorder, single episode, in partial or unspecified remission |
| ICD9CM | 296.26 | Major depressive affective disorder, single episode, in full remission |
| ICD9CM | 296.30 | Major depressive affective disorder, recurrent episode, unspecified |
| ICD9CM | 296.31 | Major depressive affective disorder, recurrent episode, mild |
| ICD9CM | 296.32 | Major depressive affective disorder, recurrent episode, moderate |
| ICD9CM | 296.33 | Major depressive affective disorder, recurrent episode, severe, without mention of psychotic behavior |
| ICD9CM | 296.34 | Major depressive affective disorder, recurrent episode, severe, specified as with psychotic behavior |
| ICD9CM | 296.35 | Major depressive affective disorder, recurrent episode, in partial or unspecified remission |
| ICD9CM | 296.36 | Major depressive affective disorder, recurrent episode, in full remission |
| ICD9CM | 296.82 | Atypical depressive disorder |
| ICD9CM | 298.0 | Depressive type psychosis |
| ICD9CM | 300.4 | Dysthymic disorder |
| ICD9CM | 301.12 | Chronic depressive personality disorder |
| ICD9CM | 309.0 | Adjustment disorder with depressed mood |
| ICD9CM | 309.1 | Prolonged depressive reaction |
| ICD9CM | 309.28 | Adjustment disorder with mixed anxiety and depressed mood |

Continued on next page

**TableS29 – continued from previous page**

| CodeSystem | ConceptCode | ConceptName |
|---|---|---|
| ICD9CM | 311 | Depressive disorder, not elsewhere classified |
| SNOMED CT | 268621008 | Recurrent major depressive episodes |
| SNOMED CT | 319768000 | Recurrent major depressive disorder with melancholic features |
| SNOMED CT | 320751009 | Major depression, melancholic type |
| SNOMED CT | 370143000 | Major depressive disorder |
| SNOMED CT | 430852001 | Severe major depression, single episode, with psychotic features |
| SNOMED CT | 442057004 | Chronic depressive personality disorder |
| SNOMED CT | 69392006 | Major depressive disorder, single episode with catatonic features |
| SNOMED CT | 71336009 | Recurrent major depressive disorder with postpartum onset |
| SNOMED CT | 73867007 | Severe major depression with psychotic features |
| SNOMED CT | 75084000 | Severe major depression without psychotic features |
| SNOMED CT | 75837004 | Mood disorder with depressive features due to general medical condition |
| SNOMED CT | 76441001 | Severe major depression, single episode, without psychotic features |
| SNOMED CT | 77486005 | Mood disorder with major depressive-like episode due to general medical condition |
| SNOMED CT | 77911002 | Severe major depression, single episode, with psychotic features, mood-congruent |
| SNOMED CT | 78667006 | Dysthymia |
| SNOMED CT | 79298009 | Mild major depression, single episode |
| SNOMED CT | 81319007 | Severe bipolar II disorder, most recent episode major depressive without psychotic features |
| SNOMED CT | 83176005 | Primary dysthymia |
| SNOMED CT | 84760002 | Schizoaffective disorder, depressive type |
| SNOMED CT | 85080004 | Secondary dysthymia |
| SNOMED CT | 87512008 | Mild major depression |
| SNOMED CT | 191604000 | Single major depressive episode, severe, with psychosis |
| SNOMED CT | 191610000 | Recurrent major depressive episodes, mild |
| SNOMED CT | 191611001 | Recurrent major depressive episodes, moderate |
| SNOMED CT | 191613003 | Recurrent major depressive episodes, severe, with psychosis |
| SNOMED CT | 191616006 | Recurrent depression |
| SNOMED CT | 191659001 | Atypical depressive disorder |
| SNOMED CT | 192080009 | Chronic depression |
| SNOMED CT | 231504006 | Mixed anxiety and depressive disorder |
| SNOMED CT | 231542000 | Depressive conduct disorder |
| SNOMED CT | 832007 | Moderate major depression |
| SNOMED CT | 2506003 | Early onset dysthymia |
| SNOMED CT | 2618002 | Chronic recurrent major depressive disorder |
| SNOMED CT | 3109008 | Secondary dysthymia early onset |
| SNOMED CT | 14183003 | Chronic major depressive disorder, single episode |
| SNOMED CT | 15193003 | Severe recurrent major depression with psychotic features, mood-incongruent |
| SNOMED CT | 15639000 | Moderate major depression, single episode |
| SNOMED CT | 18818009 | Moderate recurrent major depression |
| SNOMED CT | 19527009 | Single episode of major depression in full remission |
| SNOMED CT | 19694002 | Late onset dysthymia |

Continued on next page

**TableS29 – continued from previous page**

| CodeSystem | ConceptCode | ConceptName |
|---|---|---|
| SNOMED CT | 20250007 | Severe major depression, single episode, with psychotic features, mood-incongruent |
| SNOMED CT | 25922000 | Major depressive disorder, single episode with postpartum onset |
| SNOMED CT | 28475009 | Severe recurrent major depression with psychotic features |
| SNOMED CT | 33078009 | Severe recurrent major depression with psychotic features, mood-congruent |
| SNOMED CT | 36170009 | Secondary dysthymia late onset |
| SNOMED CT | 36474008 | Severe recurrent major depression without psychotic features |
| SNOMED CT | 36923009 | Major depression, single episode |
| SNOMED CT | 38451003 | Primary dysthymia early onset |
| SNOMED CT | 38694004 | Recurrent major depressive disorder with atypical features |
| SNOMED CT | 39809009 | Recurrent major depressive disorder with catatonic features |
| SNOMED CT | 40379007 | Mild recurrent major depression |
| SNOMED CT | 40568001 | Recurrent brief depressive disorder |
| SNOMED CT | 42925002 | Major depressive disorder, single episode with atypical features |
| SNOMED CT | 48589009 | Minor depressive disorder |
| SNOMED CT | 63778009 | Major depressive disorder, single episode with melancholic features |
| SNOMED CT | 66344007 | Recurrent major depression |
| SNOMED CT | 67711008 | Primary dysthymia late onset |
| SNOMED CT | 133121000119109 | Severe seasonal affective disorder |
| SNOMED CT | 10811121000119102 | Major depressive disorder in mother complicating childbirth |
| SNOMED CT | 35489007 | Depressive episode |
| SNOMED CT | 274948002 | Endogenous depression - recurrent |
| SNOMED CT | 279225001 | Maternity blues |
| SNOMED CT | 300706003 | Endogenous depression |
| SNOMED CT | 310495003 | Mild depression |
| SNOMED CT | 310496002 | Moderate depression |
| SNOMED CT | 310497006 | Severe depression |
| SNOMED CT | 321717001 | Involutional depression |
| SNOMED CT | 426578000 | Premenstrual dysphoric disorder in remission |
| SNOMED CT | 450714000 | Severe major depression |
| SNOMED CT | 698957003 | Depressive disorder in remission |
| SNOMED CT | 712823008 | Acute depression |
| SNOMED CT | 718636001 | Minimal depression |
| SNOMED CT | 719592004 | Moderately severe major depression |
| SNOMED CT | 719593009 | Moderately severe depression |
| SNOMED CT | 720451004 | Minimal recurrent major depression |
| SNOMED CT | 720452006 | Moderately severe recurrent major depression |
| SNOMED CT | 720453001 | Moderately severe major depression single episode |
| SNOMED CT | 720454007 | Minimal major depression single episode |
| SNOMED CT | 720455008 | Minimal major depression |
| SNOMED CT | 70747007 | Major depression single episode, in partial remission |
| SNOMED CT | 79842004 | Stuporous depression |
| SNOMED CT | 82218004 | Postoperative depression |
| SNOMED CT | 83458005 | Agitated depression |
| SNOMED CT | 84788008 | Menopausal depression |

Continued on next page

**TableS29 – continued from previous page**

| CodeSystem | ConceptCode | ConceptName |
|---|---|---|
| SNOMED CT | 87414006 | Reactive depression (situational) |
| SNOMED CT | 87842000 | Generalized neuromuscular exhaustion syndrome |
| SNOMED CT | 162722001 | O/E - depressed |
| SNOMED CT | 191495003 | Depressive disorder caused by drug |
| SNOMED CT | 191627008 | Bipolar affective disorder, current episode depression |
| SNOMED CT | 191629006 | Bipolar affective disorder, currently depressed, mild |
| SNOMED CT | 191630001 | Bipolar affective disorder, currently depressed, moderate |
| SNOMED CT | 191634005 | Bipolar affective disorder, currently depressed, in full remission |
| SNOMED CT | 191676002 | Reactive depressive psychosis |
| SNOMED CT | 192079006 | Postviral depression |
| SNOMED CT | 231485007 | Post-schizophrenic depression |
| SNOMED CT | 231499006 | Endogenous depression first episode |
| SNOMED CT | 231500002 | Masked depression |
| SNOMED CT | 237349002 | Mild postnatal depression |
| SNOMED CT | 237350002 | Severe postnatal depression |
| SNOMED CT | 247803002 | Seasonal affective disorder |
| SNOMED CT | 596004 | Premenstrual dysphoric disorder |
| SNOMED CT | 14070001 | Multi-infarct dementia with depression |
| SNOMED CT | 29929003 | Bipolar I disorder, most recent episode depressed with atypical features |
| SNOMED CT | 30605009 | Major depression in partial remission |
| SNOMED CT | 33135002 | Recurrent major depression in partial remission |
| SNOMED CT | 33736005 | Severe major depression with psychotic features, mood-congruent |
| SNOMED CT | 42810003 | Major depression in remission |
| SNOMED CT | 46244001 | Recurrent major depression in full remission |
| SNOMED CT | 58703003 | Postpartum depression |
| SNOMED CT | 60099002 | Severe major depression with psychotic features, mood-incongruent |
| SNOMED CT | 63412003 | Major depression in full remission |
| SNOMED CT | 68019004 | Recurrent major depression in remission |
| SNOMED CT | 723928009 | Mood disorder with depressive symptoms caused by alcohol |
| SNOMED CT | 723930006 | Mood disorder with mixed manic and depressive symptoms caused by alcohol |
| SNOMED CT | 724676000 | Mood disorder with depressive symptoms caused by sedative |
| SNOMED CT | 724677009 | Mood disorder with depressive symptoms caused by hypnotic |
| SNOMED CT | 724678004 | Mood disorder with depressive symptoms caused by anxiolytic |
| SNOMED CT | 724690002 | Mood disorder with depressive symptoms caused by cocaine |
| SNOMED CT | 726772006 | Major depression with psychotic features |
| SNOMED CT | 762321000 | Mood disorder with depressive symptoms caused by opioid |
| SNOMED CT | 762329003 | Mood disorder with depressive symptoms caused by stimulant |
| SNOMED CT | 762336002 | Mood disorder with depressive symptoms caused by hallucinogen |
| SNOMED CT | 762339009 | Mood disorder with depressive symptoms caused by volatile inhalant |
| SNOMED CT | 762345001 | Mood disorder with depressive symptoms caused by dissociative drug |

Continued on next page

**TableS29 – continued from previous page**

| CodeSystem | ConceptCode | ConceptName |
|---|---|---|
| SNOMED CT | 762512002 | Mood disorder with depressive symptoms caused by synthetic cathinone |
| SNOMED CT | 765176007 | Psychosis and severe depression co-occurrent and due to bipolar affective disorder |
| SNOMED CT | 788120007 | Antenatal depression |
| SNOMED CT | 838530009 | Mood disorder with depressive symptoms caused by amphetamine and/or amphetamine derivative |
| SNOMED CT | 251000119105 | Severe major depression, single episode |
| SNOMED CT | 281000119103 | Severe recurrent major depression |
| SNOMED CT | 10211000132109 | Perinatal depression |
| SNOMED CT | 94631000119100 | Depressive disorder in mother complicating pregnancy |
| SNOMED CT | 104851000119103 | Postpartum major depression in remission |
| SNOMED CT | 288751000119101 | Reactive depressive psychosis, single episode |
| SNOMED CT | 10811161000119107 | Major depressive disorder in mother complicating pregnancy |
| SNOMED CT | 10835871000119104 | Depressive disorder in mother complicating childbirth |
| SNOMED CT | 16238181000119101 | Depressive disorder caused by amphetamine |
| SNOMED CT | 16238221000119109 | Depressive disorder caused by metamfetamine |
| SNOMED CT | 16264621000119109 | Recurrent mild major depressive disorder co-occurrent with anxiety |
| SNOMED CT | 16264821000119108 | Recurrent severe major depressive disorder co-occurrent with anxiety |
| SNOMED CT | 16264901000119109 | Recurrent moderate major depressive disorder co-occurrent with anxiety |
| SNOMED CT | 16265061000119105 | Recurrent major depressive disorder co-occurrent with anxiety in full remission |
| SNOMED CT | 16265301000119106 | Recurrent major depressive disorder in partial remission co-occurrent with anxiety |
| SNOMED CT | 16265951000119109 | Mild major depressive disorder co-occurrent with anxiety single episode |
| SNOMED CT | 16266831000119100 | Moderate major depressive disorder co-occurrent with anxiety single episode |
| SNOMED CT | 16266991000119108 | Severe major depressive disorder co-occurrent with anxiety single episode |
| SNOMED CT | 1153570009 | Treatment resistant depression |
| SNOMED CT | 1153575004 | Persistent depressive disorder |

#### 5.9 Diabetes

Table S30: Concept codes used to identify if a patient was diagnosed with diabetes.

| CodeSystem | ConceptCode | ConceptName |
|---|---|---|
| ICD10CM | E11.610 | Type 2 diabetes mellitus with diabetic neuropathic arthropathy |
| ICD10CM | E11.00 | Type 2 diabetes mellitus with hyperosmolarity without nonketotic hyperglycemic-hyperosmolar coma (NKHHC) |

Continued on next page

**TableS30 – continued from previous page**

| CodeSystem | ConceptCode | ConceptName |
|---|---|---|
| ICD10CM | E11.01 | Type 2 diabetes mellitus with hyperosmolarity with coma |
| ICD10CM | E11.10 | Type 2 diabetes mellitus with ketoacidosis without coma |
| ICD10CM | E11.11 | Type 2 diabetes mellitus with ketoacidosis with coma |
| ICD10CM | E11.21 | Type 2 diabetes mellitus with diabetic nephropathy |
| ICD10CM | E11.22 | Type 2 diabetes mellitus with diabetic chronic kidney disease |
| ICD10CM | E11.29 | Type 2 diabetes mellitus with other diabetic kidney complication |
| ICD10CM | E11.311 | Type 2 diabetes mellitus with unspecified diabetic retinopathy with macular edema |
| ICD10CM | E11.319 | Type 2 diabetes mellitus with unspecified diabetic retinopathy without macular edema |
| ICD10CM | E11.321 | Type 2 diabetes mellitus with mild nonproliferative diabetic retinopathy with macular edema |
| ICD10CM | E11.3211 | Type 2 diabetes mellitus with mild nonproliferative diabetic retinopathy with macular edema, right eye |
| ICD10CM | E11.3212 | Type 2 diabetes mellitus with mild nonproliferative diabetic retinopathy with macular edema, left eye |
| ICD10CM | E11.3213 | Type 2 diabetes mellitus with mild nonproliferative diabetic retinopathy with macular edema, bilateral |
| ICD10CM | E11.3219 | Type 2 diabetes mellitus with mild nonproliferative diabetic retinopathy with macular edema, unspecified eye |
| ICD10CM | E11.329 | Type 2 diabetes mellitus with mild nonproliferative diabetic retinopathy without macular edema |
| ICD10CM | E11.3291 | Type 2 diabetes mellitus with mild nonproliferative diabetic retinopathy without macular edema, right eye |
| ICD10CM | E11.3292 | Type 2 diabetes mellitus with mild nonproliferative diabetic retinopathy without macular edema, left eye |
| ICD10CM | E11.3293 | Type 2 diabetes mellitus with mild nonproliferative diabetic retinopathy without macular edema, bilateral |
| ICD10CM | E11.3299 | Type 2 diabetes mellitus with mild nonproliferative diabetic retinopathy without macular edema, unspecified eye |
| ICD10CM | E11.331 | Type 2 diabetes mellitus with moderate nonproliferative diabetic retinopathy with macular edema |
| ICD10CM | E11.3311 | Type 2 diabetes mellitus with moderate nonproliferative diabetic retinopathy with macular edema, right eye |
| ICD10CM | E11.3312 | Type 2 diabetes mellitus with moderate nonproliferative diabetic retinopathy with macular edema, left eye |
| ICD10CM | E11.3313 | Type 2 diabetes mellitus with moderate nonproliferative diabetic retinopathy with macular edema, bilateral |
| ICD10CM | E11.3319 | Type 2 diabetes mellitus with moderate nonproliferative diabetic retinopathy with macular edema, unspecified eye |
| ICD10CM | E11.339 | Type 2 diabetes mellitus with moderate nonproliferative diabetic retinopathy without macular edema |
| ICD10CM | E11.3391 | Type 2 diabetes mellitus with moderate nonproliferative diabetic retinopathy without macular edema, right eye |
| ICD10CM | E11.3392 | Type 2 diabetes mellitus with moderate nonproliferative diabetic retinopathy without macular edema, left eye |

Continued on next page

**TableS30 – continued from previous page**

| CodeSystem | ConceptCode | ConceptName |
|---|---|---|
| ICD10CM | E11.3393 | Type 2 diabetes mellitus with moderate nonproliferative diabetic retinopathy without macular edema, bilateral |
| ICD10CM | E11.3399 | Type 2 diabetes mellitus with moderate nonproliferative diabetic retinopathy without macular edema, unspecified eye |
| ICD10CM | E11.341 | Type 2 diabetes mellitus with severe nonproliferative diabetic retinopathy with macular edema |
| ICD10CM | E11.3411 | Type 2 diabetes mellitus with severe nonproliferative diabetic retinopathy with macular edema, right eye |
| ICD10CM | E11.3412 | Type 2 diabetes mellitus with severe nonproliferative diabetic retinopathy with macular edema, left eye |
| ICD10CM | E11.3413 | Type 2 diabetes mellitus with severe nonproliferative diabetic retinopathy with macular edema, bilateral |
| ICD10CM | E11.3419 | Type 2 diabetes mellitus with severe nonproliferative diabetic retinopathy with macular edema, unspecified eye |
| ICD10CM | E11.349 | Type 2 diabetes mellitus with severe nonproliferative diabetic retinopathy without macular edema |
| ICD10CM | E11.3491 | Type 2 diabetes mellitus with severe nonproliferative diabetic retinopathy without macular edema, right eye |
| ICD10CM | E11.3492 | Type 2 diabetes mellitus with severe nonproliferative diabetic retinopathy without macular edema, left eye |
| ICD10CM | E11.3493 | Type 2 diabetes mellitus with severe nonproliferative diabetic retinopathy without macular edema, bilateral |
| ICD10CM | E11.3499 | Type 2 diabetes mellitus with severe nonproliferative diabetic retinopathy without macular edema, unspecified eye |
| ICD10CM | E11.351 | Type 2 diabetes mellitus with proliferative diabetic retinopathy with macular edema |
| ICD10CM | E11.3511 | Type 2 diabetes mellitus with proliferative diabetic retinopathy with macular edema, right eye |
| ICD10CM | E11.3512 | Type 2 diabetes mellitus with proliferative diabetic retinopathy with macular edema, left eye |
| ICD10CM | E11.3513 | Type 2 diabetes mellitus with proliferative diabetic retinopathy with macular edema, bilateral |
| ICD10CM | E11.3519 | Type 2 diabetes mellitus with proliferative diabetic retinopathy with macular edema, unspecified eye |
| ICD10CM | E11.3521 | Type 2 diabetes mellitus with proliferative diabetic retinopathy with traction retinal detachment involving the macula, right eye |
| ICD10CM | E11.3522 | Type 2 diabetes mellitus with proliferative diabetic retinopathy with traction retinal detachment involving the macula, left eye |
| ICD10CM | E11.3523 | Type 2 diabetes mellitus with proliferative diabetic retinopathy with traction retinal detachment involving the macula, bilateral |
| ICD10CM | E11.3529 | Type 2 diabetes mellitus with proliferative diabetic retinopathy with traction retinal detachment involving the macula, unspecified eye |
| ICD10CM | E11.3531 | Type 2 diabetes mellitus with proliferative diabetic retinopathy with traction retinal detachment not involving the macula, right eye |

Continued on next page

**TableS30 – continued from previous page**

| CodeSystem | ConceptCode | ConceptName |
|---|---|---|
| ICD10CM | E11.3532 | Type 2 diabetes mellitus with proliferative diabetic retinopathy with traction retinal detachment not involving the macula, left eye |
| ICD10CM | E11.3533 | Type 2 diabetes mellitus with proliferative diabetic retinopathy with traction retinal detachment not involving the macula, bilateral |
| ICD10CM | E11.3539 | Type 2 diabetes mellitus with proliferative diabetic retinopathy with traction retinal detachment not involving the macula, unspecified eye |
| ICD10CM | E11.3541 | Type 2 diabetes mellitus with proliferative diabetic retinopathy with combined traction retinal detachment and rhegmatogenous retinal detachment, right eye |
| ICD10CM | E11.3542 | Type 2 diabetes mellitus with proliferative diabetic retinopathy with combined traction retinal detachment and rhegmatogenous retinal detachment, left eye |
| ICD10CM | E11.3543 | Type 2 diabetes mellitus with proliferative diabetic retinopathy with combined traction retinal detachment and rhegmatogenous retinal detachment, bilateral |
| ICD10CM | E11.3549 | Type 2 diabetes mellitus with proliferative diabetic retinopathy with combined traction retinal detachment and rhegmatogenous retinal detachment, unspecified eye |
| ICD10CM | E11.3551 | Type 2 diabetes mellitus with stable proliferative diabetic retinopathy, right eye |
| ICD10CM | E11.3552 | Type 2 diabetes mellitus with stable proliferative diabetic retinopathy, left eye |
| ICD10CM | E11.3553 | Type 2 diabetes mellitus with stable proliferative diabetic retinopathy, bilateral |
| ICD10CM | E11.3559 | Type 2 diabetes mellitus with stable proliferative diabetic retinopathy, unspecified eye |
| ICD10CM | E11.359 | Type 2 diabetes mellitus with proliferative diabetic retinopathy without macular edema |
| ICD10CM | E11.3591 | Type 2 diabetes mellitus with proliferative diabetic retinopathy without macular edema, right eye |
| ICD10CM | E11.3592 | Type 2 diabetes mellitus with proliferative diabetic retinopathy without macular edema, left eye |
| ICD10CM | E11.3593 | Type 2 diabetes mellitus with proliferative diabetic retinopathy without macular edema, bilateral |
| ICD10CM | E11.3599 | Type 2 diabetes mellitus with proliferative diabetic retinopathy without macular edema, unspecified eye |
| ICD10CM | E11.36 | Type 2 diabetes mellitus with diabetic cataract |
| ICD10CM | E11.37X1 | Type 2 diabetes mellitus with diabetic macular edema, resolved following treatment, right eye |
| ICD10CM | E11.37X2 | Type 2 diabetes mellitus with diabetic macular edema, resolved following treatment, left eye |
| ICD10CM | E11.37X3 | Type 2 diabetes mellitus with diabetic macular edema, resolved following treatment, bilateral |

Continued on next page

**TableS30 – continued from previous page**

| CodeSystem | ConceptCode | ConceptName |
|---|---|---|
| ICD10CM | E11.37X9 | Type 2 diabetes mellitus with diabetic macular edema, resolved following treatment, unspecified eye |
| ICD10CM | E11.39 | Type 2 diabetes mellitus with other diabetic ophthalmic complication |
| ICD10CM | E11.40 | Type 2 diabetes mellitus with diabetic neuropathy, unspecified |
| ICD10CM | E11.41 | Type 2 diabetes mellitus with diabetic mononeuropathy |
| ICD10CM | E11.42 | Type 2 diabetes mellitus with diabetic polyneuropathy |
| ICD10CM | E11.43 | Type 2 diabetes mellitus with diabetic autonomic (poly)neuropathy |
| ICD10CM | E11.44 | Type 2 diabetes mellitus with diabetic amyotrophy |
| ICD10CM | E11.49 | Type 2 diabetes mellitus with other diabetic neurological complication |
| ICD10CM | E11.51 | Type 2 diabetes mellitus with diabetic peripheral angiopathy without gangrene |
| ICD10CM | E11.52 | Type 2 diabetes mellitus with diabetic peripheral angiopathy with gangrene |
| ICD10CM | E11.59 | Type 2 diabetes mellitus with other circulatory complications |
| ICD10CM | E11.618 | Type 2 diabetes mellitus with other diabetic arthropathy |
| ICD10CM | E11.620 | Type 2 diabetes mellitus with diabetic dermatitis |
| ICD10CM | E11.621 | Type 2 diabetes mellitus with foot ulcer |
| ICD10CM | E11.622 | Type 2 diabetes mellitus with other skin ulcer |
| ICD10CM | E11.628 | Type 2 diabetes mellitus with other skin complications |
| ICD10CM | E11.630 | Type 2 diabetes mellitus with periodontal disease |
| ICD10CM | E11.638 | Type 2 diabetes mellitus with other oral complications |
| ICD10CM | E11.641 | Type 2 diabetes mellitus with hypoglycemia with coma |
| ICD10CM | E11.649 | Type 2 diabetes mellitus with hypoglycemia without coma |
| ICD10CM | E11.65 | Type 2 diabetes mellitus with hyperglycemia |
| ICD10CM | E11.69 | Type 2 diabetes mellitus with other specified complication |
| ICD10CM | E11.8 | Type 2 diabetes mellitus with unspecified complications |
| ICD10CM | E11.9 | Type 2 diabetes mellitus without complications |
| ICD10CM | O24.111 | Pre-existing type 2 diabetes mellitus, in pregnancy, first trimester |
| ICD10CM | O24.112 | Pre-existing type 2 diabetes mellitus, in pregnancy, second trimester |
| ICD10CM | O24.113 | Pre-existing type 2 diabetes mellitus, in pregnancy, third trimester |
| ICD10CM | O24.119 | Pre-existing type 2 diabetes mellitus, in pregnancy, unspecified trimester |
| ICD10CM | O24.12 | Pre-existing type 2 diabetes mellitus, in childbirth |
| ICD10CM | O24.13 | Pre-existing type 2 diabetes mellitus, in the puerperium |
| SNOMED CT | 313436004 | Type 2 diabetes mellitus without complication |
| SNOMED CT | 314902007 | Type 2 diabetes mellitus with peripheral angiopathy |
| SNOMED CT | 314903002 | Arthropathy due to type 2 diabetes mellitus |
| SNOMED CT | 359642000 | Type 2 diabetes mellitus in nonobese |
| SNOMED CT | 420279001 | Renal disorder due to type 2 diabetes mellitus |
| SNOMED CT | 420436000 | Mononeuropathy due to type 2 diabetes mellitus |
| SNOMED CT | 421326000 | Disorder of nervous system due to type 2 diabetes mellitus |

Continued on next page

**TableS30 – continued from previous page**

| CodeSystem | ConceptCode | ConceptName |
|---|---|---|
| SNOMED CT | 421779007 | Exudative maculopathy due to type 2 diabetes mellitus |
| SNOMED CT | 421847006 | Ketoacidotic coma due to type 2 diabetes mellitus |
| SNOMED CT | 422034002 | Retinopathy due to type 2 diabetes mellitus |
| SNOMED CT | 422099009 | Disorder of eye due to type 2 diabetes mellitus |
| SNOMED CT | 422166005 | Peripheral circulatory disorder due to type 2 diabetes mellitus |
| SNOMED CT | 427027005 | Lumbosacral radiculoplexus neuropathy due to type 2 diabetes mellitus |
| SNOMED CT | 428007007 | Erectile dysfunction due to type 2 diabetes mellitus |
| SNOMED CT | 609567009 | Pre-existing type 2 diabetes mellitus in pregnancy |
| SNOMED CT | 712883005 | Autonomic neuropathy due to type 2 diabetes mellitus |
| SNOMED CT | 713703005 | Gastroparesis due to type 2 diabetes mellitus |
| SNOMED CT | 713706002 | Polyneuropathy due to type 2 diabetes mellitus |
| SNOMED CT | 719216001 | Hypoglycemic coma due to type 2 diabetes mellitus |
| SNOMED CT | 81531005 | Type 2 diabetes mellitus in obese |
| SNOMED CT | 190331003 | Hyperosmolar coma due to type 2 diabetes mellitus |
| SNOMED CT | 190389009 | Type 2 diabetes mellitus with ulcer |
| SNOMED CT | 199230006 | Pre-existing type 2 diabetes mellitus |
| SNOMED CT | 237599002 | Insulin treated type 2 diabetes mellitus |
| SNOMED CT | 237604008 | Maturity onset diabetes of the young, type 2 |
| SNOMED CT | 9859006 | Acanthosis nigricans due to type 2 diabetes mellitus |
| SNOMED CT | 44054006 | Type 2 diabetes mellitus |
| SNOMED CT | 1481000119100 | Diabetes mellitus type 2 without retinopathy |
| SNOMED CT | 1501000119109 | Proliferative retinopathy due to type 2 diabetes mellitus |
| SNOMED CT | 1511000119107 | Peripheral neuropathy due to type 2 diabetes mellitus |
| SNOMED CT | 1551000119108 | Nonproliferative retinopathy due to type 2 diabetes mellitus |
| SNOMED CT | 28331000119107 | Retinal edema due to type 2 diabetes mellitus |
| SNOMED CT | 41911000119107 | Glaucoma due to type 2 diabetes mellitus |
| SNOMED CT | 60951000119105 | Blindness due to type 2 diabetes mellitus |
| SNOMED CT | 71441000119104 | Nephrotic syndrome due to type 2 diabetes mellitus |
| SNOMED CT | 82541000119100 | Traction detachment of retina due to type 2 diabetes mellitus |
| SNOMED CT | 82551000119103 | Rubeosis iridis due to type 2 diabetes mellitus |
| SNOMED CT | 97331000119101 | Macular edema and retinopathy due to type 2 diabetes mellitus |
| SNOMED CT | 104961000119108 | Ischemia of retina due to type 2 diabetes mellitus |
| SNOMED CT | 110181000119105 | Peripheral sensory neuropathy due to type 2 diabetes mellitus |
| SNOMED CT | 138911000119106 | Mild nonproliferative retinopathy due to type 2 diabetes mellitus |
| SNOMED CT | 138921000119104 | Moderate nonproliferative retinopathy due to type 2 diabetes mellitus |
| SNOMED CT | 138941000119105 | Severe nonproliferative retinopathy due to diabetes mellitus type 2 |
| SNOMED CT | 368101000119109 | Diabetes mellitus type 2 with periodontal disease |
| SNOMED CT | 368581000119106 | Neuropathy due to type 2 diabetes mellitus |
| SNOMED CT | 87921000119104 | Cranial nerve palsy with type II diabetes mellitus |
| SNOMED CT | 420715001 | Persistent microalbuminuria due to type 2 diabetes mellitus |
| SNOMED CT | 421986006 | Persistent proteinuria due to type 2 diabetes mellitus |
| SNOMED CT | 703138006 | Type II diabetes mellitus in remission |
| SNOMED CT | 237627000 | Pregnancy and type 2 diabetes mellitus |
| SNOMED CT | 711000119100 | Chronic kidney disease stage 5 due to type 2 diabetes mellitus |

Continued on next page

**TableS30 – continued from previous page**

| CodeSystem | ConceptCode | ConceptName |
|---|---|---|
| SNOMED CT | 721000119107 | Chronic kidney disease stage 4 due to type 2 diabetes mellitus |
| SNOMED CT | 731000119105 | Chronic kidney disease stage 3 due to type 2 diabetes mellitus |
| SNOMED CT | 741000119101 | Chronic kidney disease stage 2 due to type 2 diabetes mellitus |
| SNOMED CT | 751000119104 | Chronic kidney disease stage 1 due to type 2 diabetes mellitus |
| SNOMED CT | 771000119108 | Chronic kidney disease due to type 2 diabetes mellitus |
| SNOMED CT | 781000119106 | Neuropathic arthropathy due to type 2 diabetes mellitus |
| SNOMED CT | 1491000119102 | Vitreous hemorrhage due to type 2 diabetes mellitus |
| SNOMED CT | 71421000119105 | Hypertension in chronic kidney disease due to type 2 diabetes mellitus |
| SNOMED CT | 90781000119102 | Microalbuminuria due to type 2 diabetes mellitus |
| SNOMED CT | 90791000119104 | End stage renal disease on dialysis due to type 2 diabetes mellitus |
| SNOMED CT | 97341000119105 | Proliferative retinopathy with retinal edema due to type 2 diabetes mellitus |
| SNOMED CT | 127991000119101 | Hypertension concurrent and due to end stage renal disease on dialysis due to type 2 diabetes mellitus |
| SNOMED CT | 140101000119109 | Hypertension in chronic kidney disease stage 5 due to type 2 diabetes mellitus |
| SNOMED CT | 140111000119107 | Hypertension in chronic kidney disease stage 4 due to type 2 diabetes mellitus |
| SNOMED CT | 140121000119100 | Hypertension in chronic kidney disease stage 3 due to type 2 diabetes mellitus |
| SNOMED CT | 140131000119102 | Hypertension in chronic kidney disease stage 2 due to type 2 diabetes mellitus |
| SNOMED CT | 157141000119108 | Proteinuria due to type 2 diabetes mellitus |
| SNOMED CT | 368591000119109 | Cheiroarthropathy due to diabetes mellitus type 2 |
| SNOMED CT | 420756003 | Cataract of eye due to diabetes mellitus type 2 |
| SNOMED CT | 769220000 | Macular edema co-occurrent and due to type 2 diabetes mellitus |
| SNOMED CT | 1003605004 | Glaucoma suspect due to diabetes mellitus type 2 |
| SNOMED CT | 16745051000119107 | Nonproliferative retinopathy of right eye due to diabetes mellitus type 2 |
| SNOMED CT | 16745131000119100 | Nonproliferative retinopathy of left eye due to diabetes mellitus type 2 |
| SNOMED CT | 16745531000119108 | Mild nonproliferative retinopathy of left eye due to diabetes mellitus type 2 |
| SNOMED CT | 16745651000119101 | Mild nonproliferative retinopathy of right eye due to diabetes mellitus type 2 |
| SNOMED CT | 16745931000119102 | Bilateral mild nonproliferative retinopathy due to diabetes mellitus type 2 |
| SNOMED CT | 16746131000119105 | Moderate nonproliferative retinopathy of left eye due to diabetes mellitus type 2 |
| SNOMED CT | 16746261000119108 | Moderate nonproliferative retinopathy of right eye due to diabetes mellitus type 2 |
| SNOMED CT | 16746341000119103 | Bilateral moderate nonproliferative retinopathy due to diabetes mellitus type 2 |
| SNOMED CT | 16746901000119101 | Severe nonproliferative retinopathy of left eye due to diabetes mellitus type 2 |

Continued on next page

**TableS30 – continued from previous page**

| CodeSystem | ConceptCode | ConceptName |
|---|---|---|
| SNOMED CT | 16747021000119109 | Bilateral severe nonproliferative retinopathy of eyes due to diabetes mellitus type 2 |
| SNOMED CT | 16747141000119104 | Severe nonproliferative retinopathy of right eye due to diabetes mellitus type 2 |
| SNOMED CT | 16747661000119109 | Bilateral nonproliferative retinopathy due to diabetes mellitus type 2 |
| SNOMED CT | 16747901000119104 | Proliferative retinopathy of left eye due to diabetes mellitus type 2 |
| SNOMED CT | 16748141000119100 | Bilateral proliferative retinopathy due to diabetes mellitus type 2 |
| SNOMED CT | 16749661000119102 | Proliferative retinopathy of right eye due to diabetes mellitus type 2 |
| SNOMED CT | 1196922005 | Neovascular glaucoma due to diabetes mellitus type 2 |
| SNOMED CT | 1217044000 | Ketosis-resistant diabetes mellitus |

#### 5.10 Immunocompromised

Table S31: Concept codes used to identify if a patient was diagnosed with immunocompromised.

| CodeSystem | ConceptCode | ConceptName |
|---|---|---|
| ICD10CM | C01 | Malignant neoplasm of base of tongue |
| ICD10CM | C02 | Malignant neoplasm of other and unspecified parts of tongue |
| ICD10CM | C03 | Malignant neoplasm of gum |
| ICD10CM | C04 | Malignant neoplasm of floor of mouth |
| ICD10CM | C05 | Malignant neoplasm of palate |
| ICD10CM | C06 | Malignant neoplasm of other and unspecified parts of mouth |
| ICD10CM | C07 | Malignant neoplasm of parotid gland |
| ICD10CM | C08 | Malignant neoplasm of other and unspecified major salivary glands |
| ICD10CM | C09 | Malignant neoplasm of tonsil |
| ICD10CM | C10 | Malignant neoplasm of oropharynx |
| ICD10CM | C11 | Malignant neoplasm of nasopharynx |
| ICD10CM | C12 | Malignant neoplasm of pyriform sinus |
| ICD10CM | C13 | Malignant neoplasm of hypopharynx |
| ICD10CM | C14 | Malignant neoplasm of other and ill-defined sites in the lip, oral cavity and pharynx |
| ICD10CM | C15 | Malignant neoplasm of esophagus |
| ICD10CM | C16 | Malignant neoplasm of stomach |
| ICD10CM | C17 | Malignant neoplasm of small intestine |
| ICD10CM | C18 | Malignant neoplasm of colon |
| ICD10CM | C19 | Malignant neoplasm of rectosigmoid junction |
| ICD10CM | C20 | Malignant neoplasm of rectum |
| ICD10CM | C21 | Malignant neoplasm of anus and anal canal |
| ICD10CM | C22 | Malignant neoplasm of liver and intrahepatic bile ducts |

Continued on next page

**TableS31 – continued from previous page**

| CodeSystem | ConceptCode | ConceptName |
|---|---|---|
| ICD10CM | C23 | Malignant neoplasm of gallbladder |
| ICD10CM | C24 | Malignant neoplasm of other and unspecified parts of biliary tract |
| ICD10CM | C25 | Malignant neoplasm of pancreas |
| ICD10CM | C26 | Malignant neoplasm of other and ill-defined digestive organs |
| ICD10CM | C30 | Malignant neoplasm of nasal cavity and middle ear |
| ICD10CM | C31 | Malignant neoplasm of accessory sinuses |
| ICD10CM | C32 | Malignant neoplasm of larynx |
| ICD10CM | C33 | Malignant neoplasm of trachea |
| ICD10CM | C34 | Malignant neoplasm of bronchus and lung |
| ICD10CM | C37 | Malignant neoplasm of thymus |
| ICD10CM | C38 | Malignant neoplasm of heart, mediastinum and pleura |
| ICD10CM | C39 | Malignant neoplasm of other and ill-defined sites in the respiratory system and intrathoracic organs |
| ICD10CM | C40 | Malignant neoplasm of bone and articular cartilage of limbs |
| ICD10CM | C41 | Malignant neoplasm of bone and articular cartilage of other and unspecified sites |
| ICD10CM | C43 | Malignant melanoma of skin |
| ICD10CM | C44 | Other and unspecified malignant neoplasm of skin |
| ICD10CM | C45 | Mesothelioma |
| ICD10CM | C46 | Kaposi's sarcoma |
| ICD10CM | C47 | Malignant neoplasm of peripheral nerves and autonomic nervous system |
| ICD10CM | C48 | Malignant neoplasm of retroperitoneum and peritoneum |
| ICD10CM | C49 | Malignant neoplasm of other connective and soft tissue |
| ICD10CM | C50 | Malignant neoplasm of breast |
| ICD10CM | C51 | Malignant neoplasm of vulva |
| ICD10CM | C52 | Malignant neoplasm of vagina |
| ICD10CM | C53 | Malignant neoplasm of cervix uteri |
| ICD10CM | C54 | Malignant neoplasm of corpus uteri |
| ICD10CM | C55 | Malignant neoplasm of uterus, part unspecified |
| ICD10CM | C56 | Malignant neoplasm of ovary |
| ICD10CM | C57 | Malignant neoplasm of other and unspecified female genital organs |
| ICD10CM | C58 | Malignant neoplasm of placenta |
| ICD10CM | C60 | Malignant neoplasm of penis |
| ICD10CM | C61 | Malignant neoplasm of prostate |
| ICD10CM | C62 | Malignant neoplasm of testis |
| ICD10CM | C63 | Malignant neoplasm of other and unspecified male genital organs |
| ICD10CM | C64 | Malignant neoplasm of kidney, except renal pelvis |
| ICD10CM | C65 | Malignant neoplasm of renal pelvis |
| ICD10CM | C66 | Malignant neoplasm of ureter |
| ICD10CM | C67 | Malignant neoplasm of bladder |
| ICD10CM | C68 | Malignant neoplasm of other and unspecified urinary organs |
| ICD10CM | C69 | Malignant neoplasm of eye and adnexa |
| ICD10CM | C70 | Malignant neoplasm of meninges |
| ICD10CM | C71 | Malignant neoplasm of brain |

Continued on next page

**TableS31 – continued from previous page**

| CodeSystem | ConceptCode | ConceptName |
|---|---|---|
| ICD10CM | C72 | Malignant neoplasm of spinal cord, cranial nerves and other parts of central nervous system |
| ICD10CM | C73 | Malignant neoplasm of thyroid gland |
| ICD10CM | C74 | Malignant neoplasm of adrenal gland |
| ICD10CM | C75 | Malignant neoplasm of other endocrine glands and related structures |
| ICD10CM | C76 | Malignant neoplasm of other and ill-defined sites |
| ICD10CM | C77 | Secondary and unspecified malignant neoplasm of lymph nodes |
| ICD10CM | C78 | Secondary malignant neoplasm of respiratory and digestive organs |
| ICD10CM | C79 | Secondary malignant neoplasm of other and unspecified sites |
| ICD10CM | C7A | Malignant neuroendocrine tumors |
| ICD10CM | C7B | Secondary neuroendocrine tumors |
| ICD10CM | C80 | Malignant neoplasm without specification of site |
| ICD10CM | C81 | Hodgkin lymphoma |
| ICD10CM | C82 | Follicular lymphoma |
| ICD10CM | C83 | Non-follicular lymphoma |
| ICD10CM | C84 | Mature T/NK-cell lymphomas |
| ICD10CM | C85 | Other specified and unspecified types of non-Hodgkin lymphoma |
| ICD10CM | C86 | Other specified types of T/NK-cell lymphoma |
| ICD10CM | C88 | Malignant immunoproliferative diseases and certain other B-cell lymphomas |
| ICD10CM | C90 | Multiple myeloma and malignant plasma cell neoplasms |
| ICD10CM | C91 | Lymphoid leukemia |
| ICD10CM | C92 | Myeloid leukemia |
| ICD10CM | C93 | Monocytic leukemia |
| ICD10CM | C94 | Other leukemias of specified cell type |
| ICD10CM | C95 | Leukemia of unspecified cell type |
| ICD10CM | C96 | Other and unspecified malignant neoplasms of lymphoid, hematopoietic and related tissue |
| ICD10CM | D27.9 | Benign neoplasm of unspecified ovary |
| ICD10CM | D3A | Benign neuroendocrine tumors |
| ICD10CM | D46 | Myelodysplastic syndromes |
| ICD10CM | D47.Z1 | Post-transplant lymphoproliferative disorder (PTLD) |
| ICD10CM | D61.0 | Constitutional aplastic anemia |
| ICD10CM | D61.09 | Other constitutional aplastic anemia |
| ICD10CM | D61.2 | Aplastic anemia due to other external agents |
| ICD10CM | D61.9 | Aplastic anemia, unspecified |
| ICD10CM | D70.0 | Congenital agranulocytosis |
| ICD10CM | D71 | Functional disorders of polymorphonuclear neutrophils |
| ICD10CM | D72.89 | Other specified disorders of white blood cells |
| ICD10CM | D80 | Immunodeficiency with predominantly antibody defects |
| ICD10CM | D81 | Combined immunodeficiencies |
| ICD10CM | D82 | Immunodeficiency associated with other major defects |
| ICD10CM | D83 | Common variable immunodeficiency |
| ICD10CM | D84 | Other immunodeficiencies |

Continued on next page

**TableS31 – continued from previous page**

| CodeSystem | ConceptCode | ConceptName |
|---|---|---|
| ICD10CM | D89 | Other disorders involving the immune mechanism, not elsewhere classified |
| ICD10CM | G35 | Multiple sclerosis |
| ICD10CM | J67.9 | Hypersensitivity pneumonitis due to unspecified organic dust |
| ICD10CM | K70.3 | Alcoholic cirrhosis of liver |
| ICD10CM | K70.4 | Alcoholic hepatic failure |
| ICD10CM | K72 | Hepatic failure, not elsewhere classified |
| ICD10CM | K74.3 | Primary biliary cirrhosis |
| ICD10CM | K74.4 | Secondary biliary cirrhosis |
| ICD10CM | K74.5 | Biliary cirrhosis, unspecified |
| ICD10CM | K74.6 | Other and unspecified cirrhosis of liver |
| ICD10CM | L40.54 | Psoriatic juvenile arthropathy |
| ICD10CM | L40.59 | Other psoriatic arthropathy |
| ICD10CM | L93.0 | Discoid lupus erythematosus |
| ICD10CM | L93.2 | Other local lupus erythematosus |
| ICD10CM | L94 | Other localized connective tissue disorders |
| ICD10CM | M05 | Rheumatoid arthritis with rheumatoid factor |
| ICD10CM | M06 | Other rheumatoid arthritis |
| ICD10CM | M07 | Enteropathic arthropathies |
| ICD10CM | M08 | Juvenile arthritis |
| ICD10CM | M30 | Polyarteritis nodosa and related conditions |
| ICD10CM | M31.3 | Wegener's granulomatosis |
| ICD10CM | M31.5 | Giant cell arteritis with polymyalgia rheumatica |
| ICD10CM | M32 | Systemic lupus erythematosus (SLE) |
| ICD10CM | M33 | Dermatopolymyositis |
| ICD10CM | M34 | Systemic sclerosis [scleroderma] |
| ICD10CM | M35.3 | Polymyalgia rheumatica |
| ICD10CM | M35.8 | Other specified systemic involvement of connective tissue |
| ICD10CM | M35.9 | Systemic involvement of connective tissue, unspecified |
| ICD10CM | M46 | Other inflammatory spondylopathies |
| ICD10CM | T78.40 | Allergy, unspecified |
| ICD10CM | T86 | Complications of transplanted organs and tissue |
| ICD10CM | Z48.2 | Encounter for aftercare following organ transplant |
| ICD10CM | Z51.0 | Encounter for antineoplastic radiation therapy |
| ICD10CM | Z51.1 | Encounter for antineoplastic chemotherapy and immunotherapy |
| ICD10CM | Z94 | Transplanted organ and tissue status |
| ICD10CM | Z98.85 | Transplanted organ removal status |
| ICD10CM | C02.0 | Malignant neoplasm of dorsal surface of tongue |
| ICD10CM | C02.1 | Malignant neoplasm of border of tongue |
| ICD10CM | C02.2 | Malignant neoplasm of ventral surface of tongue |
| ICD10CM | C02.3 | Malignant neoplasm of anterior two-thirds of tongue, part unspecified |
| ICD10CM | C02.4 | Malignant neoplasm of lingual tonsil |
| ICD10CM | C02.8 | Malignant neoplasm of overlapping sites of tongue |
| ICD10CM | C02.9 | Malignant neoplasm of tongue, unspecified |
| ICD10CM | C03.0 | Malignant neoplasm of upper gum |
| ICD10CM | C03.1 | Malignant neoplasm of lower gum |

Continued on next page

**TableS31 – continued from previous page**

| CodeSystem | ConceptCode | ConceptName |
|---|---|---|
| ICD10CM | C03.9 | Malignant neoplasm of gum, unspecified |
| ICD10CM | C04.0 | Malignant neoplasm of anterior floor of mouth |
| ICD10CM | C04.1 | Malignant neoplasm of lateral floor of mouth |
| ICD10CM | C04.8 | Malignant neoplasm of overlapping sites of floor of mouth |
| ICD10CM | C04.9 | Malignant neoplasm of floor of mouth, unspecified |
| ICD10CM | C05.0 | Malignant neoplasm of hard palate |
| ICD10CM | C05.1 | Malignant neoplasm of soft palate |
| ICD10CM | C05.2 | Malignant neoplasm of uvula |
| ICD10CM | C05.8 | Malignant neoplasm of overlapping sites of palate |
| ICD10CM | C05.9 | Malignant neoplasm of palate, unspecified |
| ICD10CM | C06.0 | Malignant neoplasm of cheek mucosa |
| ICD10CM | C06.1 | Malignant neoplasm of vestibule of mouth |
| ICD10CM | C06.2 | Malignant neoplasm of retromolar area |
| ICD10CM | C06.8 | Malignant neoplasm of overlapping sites of other and unspecified parts of mouth |
| ICD10CM | C06.80 | Malignant neoplasm of overlapping sites of unspecified parts of mouth |
| ICD10CM | C06.89 | Malignant neoplasm of overlapping sites of other parts of mouth |
| ICD10CM | C06.9 | Malignant neoplasm of mouth, unspecified |
| ICD10CM | C08.0 | Malignant neoplasm of submandibular gland |
| ICD10CM | C08.1 | Malignant neoplasm of sublingual gland |
| ICD10CM | C08.9 | Malignant neoplasm of major salivary gland, unspecified |
| ICD10CM | C09.0 | Malignant neoplasm of tonsillar fossa |
| ICD10CM | C09.1 | Malignant neoplasm of tonsillar pillar (anterior) (posterior) |
| ICD10CM | C09.8 | Malignant neoplasm of overlapping sites of tonsil |
| ICD10CM | C09.9 | Malignant neoplasm of tonsil, unspecified |
| ICD10CM | C10.0 | Malignant neoplasm of vallecula |
| ICD10CM | C10.1 | Malignant neoplasm of anterior surface of epiglottis |
| ICD10CM | C10.2 | Malignant neoplasm of lateral wall of oropharynx |
| ICD10CM | C10.3 | Malignant neoplasm of posterior wall of oropharynx |
| ICD10CM | C10.4 | Malignant neoplasm of branchial cleft |
| ICD10CM | C10.8 | Malignant neoplasm of overlapping sites of oropharynx |
| ICD10CM | C10.9 | Malignant neoplasm of oropharynx, unspecified |
| ICD10CM | C11.0 | Malignant neoplasm of superior wall of nasopharynx |
| ICD10CM | C11.1 | Malignant neoplasm of posterior wall of nasopharynx |
| ICD10CM | C11.2 | Malignant neoplasm of lateral wall of nasopharynx |
| ICD10CM | C11.3 | Malignant neoplasm of anterior wall of nasopharynx |
| ICD10CM | C11.8 | Malignant neoplasm of overlapping sites of nasopharynx |
| ICD10CM | C11.9 | Malignant neoplasm of nasopharynx, unspecified |
| ICD10CM | C13.0 | Malignant neoplasm of postcricoid region |
| ICD10CM | C13.1 | Malignant neoplasm of aryepiglottic fold, hypopharyngeal aspect |
| ICD10CM | C13.2 | Malignant neoplasm of posterior wall of hypopharynx |
| ICD10CM | C13.8 | Malignant neoplasm of overlapping sites of hypopharynx |
| ICD10CM | C13.9 | Malignant neoplasm of hypopharynx, unspecified |
| ICD10CM | C14.0 | Malignant neoplasm of pharynx, unspecified |
| ICD10CM | C14.2 | Malignant neoplasm of Waldeyer's ring |

Continued on next page

**TableS31 – continued from previous page**

| CodeSystem | ConceptCode | ConceptName |
|---|---|---|
| ICD10CM | C14.8 | Malignant neoplasm of overlapping sites of lip, oral cavity and pharynx |
| ICD10CM | C15.3 | Malignant neoplasm of upper third of esophagus |
| ICD10CM | C15.4 | Malignant neoplasm of middle third of esophagus |
| ICD10CM | C15.5 | Malignant neoplasm of lower third of esophagus |
| ICD10CM | C15.8 | Malignant neoplasm of overlapping sites of esophagus |
| ICD10CM | C15.9 | Malignant neoplasm of esophagus, unspecified |
| ICD10CM | C16.0 | Malignant neoplasm of cardia |
| ICD10CM | C16.1 | Malignant neoplasm of fundus of stomach |
| ICD10CM | C16.2 | Malignant neoplasm of body of stomach |
| ICD10CM | C16.3 | Malignant neoplasm of pyloric antrum |
| ICD10CM | C16.4 | Malignant neoplasm of pylorus |
| ICD10CM | C16.5 | Malignant neoplasm of lesser curvature of stomach, unspecified |
| ICD10CM | C16.6 | Malignant neoplasm of greater curvature of stomach, unspecified |
| ICD10CM | C16.8 | Malignant neoplasm of overlapping sites of stomach |
| ICD10CM | C16.9 | Malignant neoplasm of stomach, unspecified |
| ICD10CM | C17.0 | Malignant neoplasm of duodenum |
| ICD10CM | C17.1 | Malignant neoplasm of jejunum |
| ICD10CM | C17.2 | Malignant neoplasm of ileum |
| ICD10CM | C17.3 | Meckel's diverticulum, malignant |
| ICD10CM | C17.8 | Malignant neoplasm of overlapping sites of small intestine |
| ICD10CM | C17.9 | Malignant neoplasm of small intestine, unspecified |
| ICD10CM | C18.0 | Malignant neoplasm of cecum |
| ICD10CM | C18.1 | Malignant neoplasm of appendix |
| ICD10CM | C18.2 | Malignant neoplasm of ascending colon |
| ICD10CM | C18.3 | Malignant neoplasm of hepatic flexure |
| ICD10CM | C18.4 | Malignant neoplasm of transverse colon |
| ICD10CM | C18.5 | Malignant neoplasm of splenic flexure |
| ICD10CM | C18.6 | Malignant neoplasm of descending colon |
| ICD10CM | C18.7 | Malignant neoplasm of sigmoid colon |
| ICD10CM | C18.8 | Malignant neoplasm of overlapping sites of colon |
| ICD10CM | C18.9 | Malignant neoplasm of colon, unspecified |
| ICD10CM | C21.0 | Malignant neoplasm of anus, unspecified |
| ICD10CM | C21.1 | Malignant neoplasm of anal canal |
| ICD10CM | C21.2 | Malignant neoplasm of cloacogenic zone |
| ICD10CM | C21.8 | Malignant neoplasm of overlapping sites of rectum, anus and anal canal |
| ICD10CM | C22.0 | Liver cell carcinoma |
| ICD10CM | C22.1 | Intrahepatic bile duct carcinoma |
| ICD10CM | C22.2 | Hepatoblastoma |
| ICD10CM | C22.3 | Angiosarcoma of liver |
| ICD10CM | C22.4 | Other sarcomas of liver |
| ICD10CM | C22.7 | Other specified carcinomas of liver |
| ICD10CM | C22.8 | Malignant neoplasm of liver, primary, unspecified as to type |
| ICD10CM | C22.9 | Malignant neoplasm of liver, not specified as primary or secondary |
| ICD10CM | C24.0 | Malignant neoplasm of extrahepatic bile duct |

Continued on next page

**TableS31 – continued from previous page**

| CodeSystem | ConceptCode | ConceptName |
|---|---|---|
| ICD10CM | C24.1 | Malignant neoplasm of ampulla of Vater |
| ICD10CM | C24.8 | Malignant neoplasm of overlapping sites of biliary tract |
| ICD10CM | C24.9 | Malignant neoplasm of biliary tract, unspecified |
| ICD10CM | C25.0 | Malignant neoplasm of head of pancreas |
| ICD10CM | C25.1 | Malignant neoplasm of body of pancreas |
| ICD10CM | C25.2 | Malignant neoplasm of tail of pancreas |
| ICD10CM | C25.3 | Malignant neoplasm of pancreatic duct |
| ICD10CM | C25.4 | Malignant neoplasm of endocrine pancreas |
| ICD10CM | C25.7 | Malignant neoplasm of other parts of pancreas |
| ICD10CM | C25.8 | Malignant neoplasm of overlapping sites of pancreas |
| ICD10CM | C25.9 | Malignant neoplasm of pancreas, unspecified |
| ICD10CM | C26.0 | Malignant neoplasm of intestinal tract, part unspecified |
| ICD10CM | C26.1 | Malignant neoplasm of spleen |
| ICD10CM | C26.9 | Malignant neoplasm of ill-defined sites within the digestive system |
| ICD10CM | C30.0 | Malignant neoplasm of nasal cavity |
| ICD10CM | C30.1 | Malignant neoplasm of middle ear |
| ICD10CM | C31.0 | Malignant neoplasm of maxillary sinus |
| ICD10CM | C31.1 | Malignant neoplasm of ethmoidal sinus |
| ICD10CM | C31.2 | Malignant neoplasm of frontal sinus |
| ICD10CM | C31.3 | Malignant neoplasm of sphenoid sinus |
| ICD10CM | C31.8 | Malignant neoplasm of overlapping sites of accessory sinuses |
| ICD10CM | C31.9 | Malignant neoplasm of accessory sinus, unspecified |
| ICD10CM | C32.0 | Malignant neoplasm of glottis |
| ICD10CM | C32.1 | Malignant neoplasm of supraglottis |
| ICD10CM | C32.2 | Malignant neoplasm of subglottis |
| ICD10CM | C32.3 | Malignant neoplasm of laryngeal cartilage |
| ICD10CM | C32.8 | Malignant neoplasm of overlapping sites of larynx |
| ICD10CM | C32.9 | Malignant neoplasm of larynx, unspecified |
| ICD10CM | C34.0 | Malignant neoplasm of main bronchus |
| ICD10CM | C34.00 | Malignant neoplasm of unspecified main bronchus |
| ICD10CM | C34.01 | Malignant neoplasm of right main bronchus |
| ICD10CM | C34.02 | Malignant neoplasm of left main bronchus |
| ICD10CM | C34.1 | Malignant neoplasm of upper lobe, bronchus or lung |
| ICD10CM | C34.10 | Malignant neoplasm of upper lobe, unspecified bronchus or lung |
| ICD10CM | C34.11 | Malignant neoplasm of upper lobe, right bronchus or lung |
| ICD10CM | C34.12 | Malignant neoplasm of upper lobe, left bronchus or lung |
| ICD10CM | C34.2 | Malignant neoplasm of middle lobe, bronchus or lung |
| ICD10CM | C34.3 | Malignant neoplasm of lower lobe, bronchus or lung |
| ICD10CM | C34.30 | Malignant neoplasm of lower lobe, unspecified bronchus or lung |
| ICD10CM | C34.31 | Malignant neoplasm of lower lobe, right bronchus or lung |
| ICD10CM | C34.32 | Malignant neoplasm of lower lobe, left bronchus or lung |
| ICD10CM | C34.8 | Malignant neoplasm of overlapping sites of bronchus and lung |
| ICD10CM | C34.80 | Malignant neoplasm of overlapping sites of unspecified bronchus and lung |
| ICD10CM | C34.81 | Malignant neoplasm of overlapping sites of right bronchus and lung |

Continued on next page

**TableS31 – continued from previous page**

| CodeSystem | ConceptCode | ConceptName |
|---|---|---|
| ICD10CM | C34.82 | Malignant neoplasm of overlapping sites of left bronchus and lung |
| ICD10CM | C34.9 | Malignant neoplasm of unspecified part of bronchus or lung |
| ICD10CM | C34.90 | Malignant neoplasm of unspecified part of unspecified bronchus or lung |
| ICD10CM | C34.91 | Malignant neoplasm of unspecified part of right bronchus or lung |
| ICD10CM | C34.92 | Malignant neoplasm of unspecified part of left bronchus or lung |
| ICD10CM | C38.0 | Malignant neoplasm of heart |
| ICD10CM | C38.1 | Malignant neoplasm of anterior mediastinum |
| ICD10CM | C38.2 | Malignant neoplasm of posterior mediastinum |
| ICD10CM | C38.3 | Malignant neoplasm of mediastinum, part unspecified |
| ICD10CM | C38.4 | Malignant neoplasm of pleura |
| ICD10CM | C38.8 | Malignant neoplasm of overlapping sites of heart, mediastinum and pleura |
| ICD10CM | C39.0 | Malignant neoplasm of upper respiratory tract, part unspecified |
| ICD10CM | C39.9 | Malignant neoplasm of lower respiratory tract, part unspecified |
| ICD10CM | C40.0 | Malignant neoplasm of scapula and long bones of upper limb |
| ICD10CM | C40.00 | Malignant neoplasm of scapula and long bones of unspecified upper limb |
| ICD10CM | C40.01 | Malignant neoplasm of scapula and long bones of right upper limb |
| ICD10CM | C40.02 | Malignant neoplasm of scapula and long bones of left upper limb |
| ICD10CM | C40.1 | Malignant neoplasm of short bones of upper limb |
| ICD10CM | C40.10 | Malignant neoplasm of short bones of unspecified upper limb |
| ICD10CM | C40.11 | Malignant neoplasm of short bones of right upper limb |
| ICD10CM | C40.12 | Malignant neoplasm of short bones of left upper limb |
| ICD10CM | C40.2 | Malignant neoplasm of long bones of lower limb |
| ICD10CM | C40.20 | Malignant neoplasm of long bones of unspecified lower limb |
| ICD10CM | C40.21 | Malignant neoplasm of long bones of right lower limb |
| ICD10CM | C40.22 | Malignant neoplasm of long bones of left lower limb |
| ICD10CM | C40.3 | Malignant neoplasm of short bones of lower limb |
| ICD10CM | C40.30 | Malignant neoplasm of short bones of unspecified lower limb |
| ICD10CM | C40.31 | Malignant neoplasm of short bones of right lower limb |
| ICD10CM | C40.32 | Malignant neoplasm of short bones of left lower limb |
| ICD10CM | C40.8 | Malignant neoplasm of overlapping sites of bone and articular cartilage of limb |
| ICD10CM | C40.80 | Malignant neoplasm of overlapping sites of bone and articular cartilage of unspecified limb |
| ICD10CM | C40.81 | Malignant neoplasm of overlapping sites of bone and articular cartilage of right limb |
| ICD10CM | C40.82 | Malignant neoplasm of overlapping sites of bone and articular cartilage of left limb |
| ICD10CM | C40.9 | Malignant neoplasm of unspecified bones and articular cartilage of limb |
| ICD10CM | C40.90 | Malignant neoplasm of unspecified bones and articular cartilage of unspecified limb |

Continued on next page

**TableS31 – continued from previous page**

| CodeSystem | ConceptCode | ConceptName |
|---|---|---|
| ICD10CM | C40.91 | Malignant neoplasm of unspecified bones and articular cartilage of right limb |
| ICD10CM | C40.92 | Malignant neoplasm of unspecified bones and articular cartilage of left limb |
| ICD10CM | C41.0 | Malignant neoplasm of bones of skull and face |
| ICD10CM | C41.1 | Malignant neoplasm of mandible |
| ICD10CM | C41.2 | Malignant neoplasm of vertebral column |
| ICD10CM | C41.3 | Malignant neoplasm of ribs, sternum and clavicle |
| ICD10CM | C41.4 | Malignant neoplasm of pelvic bones, sacrum and coccyx |
| ICD10CM | C41.9 | Malignant neoplasm of bone and articular cartilage, unspecified |
| ICD10CM | C43.0 | Malignant melanoma of lip |
| ICD10CM | C43.1 | Malignant melanoma of eyelid, including canthus |
| ICD10CM | C43.10 | Malignant melanoma of unspecified eyelid, including canthus |
| ICD10CM | C43.11 | Malignant melanoma of right eyelid, including canthus |
| ICD10CM | C43.111 | Malignant melanoma of right upper eyelid, including canthus |
| ICD10CM | C43.112 | Malignant melanoma of right lower eyelid, including canthus |
| ICD10CM | C43.12 | Malignant melanoma of left eyelid, including canthus |
| ICD10CM | C43.121 | Malignant melanoma of left upper eyelid, including canthus |
| ICD10CM | C43.122 | Malignant melanoma of left lower eyelid, including canthus |
| ICD10CM | C43.2 | Malignant melanoma of ear and external auricular canal |
| ICD10CM | C43.20 | Malignant melanoma of unspecified ear and external auricular canal |
| ICD10CM | C43.21 | Malignant melanoma of right ear and external auricular canal |
| ICD10CM | C43.22 | Malignant melanoma of left ear and external auricular canal |
| ICD10CM | C43.3 | Malignant melanoma of other and unspecified parts of face |
| ICD10CM | C43.30 | Malignant melanoma of unspecified part of face |
| ICD10CM | C43.31 | Malignant melanoma of nose |
| ICD10CM | C43.39 | Malignant melanoma of other parts of face |
| ICD10CM | C43.4 | Malignant melanoma of scalp and neck |
| ICD10CM | C43.5 | Malignant melanoma of trunk |
| ICD10CM | C43.51 | Malignant melanoma of anal skin |
| ICD10CM | C43.52 | Malignant melanoma of skin of breast |
| ICD10CM | C43.59 | Malignant melanoma of other part of trunk |
| ICD10CM | C43.6 | Malignant melanoma of upper limb, including shoulder |
| ICD10CM | C43.60 | Malignant melanoma of unspecified upper limb, including shoulder |
| ICD10CM | C43.61 | Malignant melanoma of right upper limb, including shoulder |
| ICD10CM | C43.62 | Malignant melanoma of left upper limb, including shoulder |
| ICD10CM | C43.7 | Malignant melanoma of lower limb, including hip |
| ICD10CM | C43.70 | Malignant melanoma of unspecified lower limb, including hip |
| ICD10CM | C43.71 | Malignant melanoma of right lower limb, including hip |
| ICD10CM | C43.72 | Malignant melanoma of left lower limb, including hip |
| ICD10CM | C43.8 | Malignant melanoma of overlapping sites of skin |
| ICD10CM | C43.9 | Malignant melanoma of skin, unspecified |
| ICD10CM | C44.0 | Other and unspecified malignant neoplasm of skin of lip |
| ICD10CM | C44.00 | Unspecified malignant neoplasm of skin of lip |
| ICD10CM | C44.01 | Basal cell carcinoma of skin of lip |

Continued on next page

**TableS31 – continued from previous page**

| CodeSystem | ConceptCode | ConceptName |
|---|---|---|
| ICD10CM | C44.02 | Squamous cell carcinoma of skin of lip |
| ICD10CM | C44.09 | Other specified malignant neoplasm of skin of lip |
| ICD10CM | C44.1 | Other and unspecified malignant neoplasm of skin of eyelid, including canthus |
| ICD10CM | C44.10 | Unspecified malignant neoplasm of skin of eyelid, including canthus |
| ICD10CM | C44.101 | Unspecified malignant neoplasm of skin of unspecified eyelid, including canthus |
| ICD10CM | C44.102 | Unspecified malignant neoplasm of skin of right eyelid, including canthus |
| ICD10CM | C44.1021 | Unspecified malignant neoplasm of skin of right upper eyelid, including canthus |
| ICD10CM | C44.1022 | Unspecified malignant neoplasm of skin of right lower eyelid, including canthus |
| ICD10CM | C44.109 | Unspecified malignant neoplasm of skin of left eyelid, including canthus |
| ICD10CM | C44.1091 | Unspecified malignant neoplasm of skin of left upper eyelid, including canthus |
| ICD10CM | C44.1092 | Unspecified malignant neoplasm of skin of left lower eyelid, including canthus |
| ICD10CM | C44.11 | Basal cell carcinoma of skin of eyelid, including canthus |
| ICD10CM | C44.111 | Basal cell carcinoma of skin of unspecified eyelid, including canthus |
| ICD10CM | C44.112 | Basal cell carcinoma of skin of right eyelid, including canthus |
| ICD10CM | C44.1121 | Basal cell carcinoma of skin of right upper eyelid, including canthus |
| ICD10CM | C44.1122 | Basal cell carcinoma of skin of right lower eyelid, including canthus |
| ICD10CM | C44.119 | Basal cell carcinoma of skin of left eyelid, including canthus |
| ICD10CM | C44.1191 | Basal cell carcinoma of skin of left upper eyelid, including canthus |
| ICD10CM | C44.1192 | Basal cell carcinoma of skin of left lower eyelid, including canthus |
| ICD10CM | C44.12 | Squamous cell carcinoma of skin of eyelid, including canthus |
| ICD10CM | C44.121 | Squamous cell carcinoma of skin of unspecified eyelid, including canthus |
| ICD10CM | C44.122 | Squamous cell carcinoma of skin of right eyelid, including canthus |
| ICD10CM | C44.1221 | Squamous cell carcinoma of skin of right upper eyelid, including canthus |
| ICD10CM | C44.1222 | Squamous cell carcinoma of skin of right lower eyelid, including canthus |
| ICD10CM | C44.129 | Squamous cell carcinoma of skin of left eyelid, including canthus |
| ICD10CM | C44.1291 | Squamous cell carcinoma of skin of left upper eyelid, including canthus |
| ICD10CM | C44.1292 | Squamous cell carcinoma of skin of left lower eyelid, including canthus |
| ICD10CM | C44.13 | Sebaceous cell carcinoma of skin of eyelid, including canthus |

Continued on next page

**TableS31 – continued from previous page**

| CodeSystem | ConceptCode | ConceptName |
|---|---|---|
| ICD10CM | C44.131 | Sebaceous cell carcinoma of skin of unspecified eyelid, including canthus |
| ICD10CM | C44.132 | Sebaceous cell carcinoma of skin of right eyelid, including canthus |
| ICD10CM | C44.1321 | Sebaceous cell carcinoma of skin of right upper eyelid, including canthus |
| ICD10CM | C44.1322 | Sebaceous cell carcinoma of skin of right lower eyelid, including canthus |
| ICD10CM | C44.139 | Sebaceous cell carcinoma of skin of left eyelid, including canthus |
| ICD10CM | C44.1391 | Sebaceous cell carcinoma of skin of left upper eyelid, including canthus |
| ICD10CM | C44.1392 | Sebaceous cell carcinoma of skin of left lower eyelid, including canthus |
| ICD10CM | C44.19 | Other specified malignant neoplasm of skin of eyelid, including canthus |
| ICD10CM | C44.191 | Other specified malignant neoplasm of skin of unspecified eyelid, including canthus |
| ICD10CM | C44.192 | Other specified malignant neoplasm of skin of right eyelid, including canthus |
| ICD10CM | C44.1921 | Other specified malignant neoplasm of skin of right upper eyelid, including canthus |
| ICD10CM | C44.1922 | Other specified malignant neoplasm of skin of right lower eyelid, including canthus |
| ICD10CM | C44.199 | Other specified malignant neoplasm of skin of left eyelid, including canthus |
| ICD10CM | C44.1991 | Other specified malignant neoplasm of skin of left upper eyelid, including canthus |
| ICD10CM | C44.1992 | Other specified malignant neoplasm of skin of left lower eyelid, including canthus |
| ICD10CM | C44.2 | Other and unspecified malignant neoplasm of skin of ear and external auricular canal |
| ICD10CM | C44.20 | Unspecified malignant neoplasm of skin of ear and external auricular canal |
| ICD10CM | C44.201 | Unspecified malignant neoplasm of skin of unspecified ear and external auricular canal |
| ICD10CM | C44.202 | Unspecified malignant neoplasm of skin of right ear and external auricular canal |
| ICD10CM | C44.209 | Unspecified malignant neoplasm of skin of left ear and external auricular canal |
| ICD10CM | C44.21 | Basal cell carcinoma of skin of ear and external auricular canal |
| ICD10CM | C44.211 | Basal cell carcinoma of skin of unspecified ear and external auricular canal |
| ICD10CM | C44.212 | Basal cell carcinoma of skin of right ear and external auricular canal |
| ICD10CM | C44.219 | Basal cell carcinoma of skin of left ear and external auricular canal |
| ICD10CM | C44.22 | Squamous cell carcinoma of skin of ear and external auricular canal |

Continued on next page

**TableS31 – continued from previous page**

| CodeSystem | ConceptCode | ConceptName |
|---|---|---|
| ICD10CM | C44.221 | Squamous cell carcinoma of skin of unspecified ear and external auricular canal |
| ICD10CM | C44.222 | Squamous cell carcinoma of skin of right ear and external auricular canal |
| ICD10CM | C44.229 | Squamous cell carcinoma of skin of left ear and external auricular canal |
| ICD10CM | C44.29 | Other specified malignant neoplasm of skin of ear and external auricular canal |
| ICD10CM | C44.291 | Other specified malignant neoplasm of skin of unspecified ear and external auricular canal |
| ICD10CM | C44.292 | Other specified malignant neoplasm of skin of right ear and external auricular canal |
| ICD10CM | C44.299 | Other specified malignant neoplasm of skin of left ear and external auricular canal |
| ICD10CM | C44.3 | Other and unspecified malignant neoplasm of skin of other and unspecified parts of face |
| ICD10CM | C44.30 | Unspecified malignant neoplasm of skin of other and unspecified parts of face |
| ICD10CM | C44.300 | Unspecified malignant neoplasm of skin of unspecified part of face |
| ICD10CM | C44.301 | Unspecified malignant neoplasm of skin of nose |
| ICD10CM | C44.309 | Unspecified malignant neoplasm of skin of other parts of face |
| ICD10CM | C44.31 | Basal cell carcinoma of skin of other and unspecified parts of face |
| ICD10CM | C44.310 | Basal cell carcinoma of skin of unspecified parts of face |
| ICD10CM | C44.311 | Basal cell carcinoma of skin of nose |
| ICD10CM | C44.319 | Basal cell carcinoma of skin of other parts of face |
| ICD10CM | C44.32 | Squamous cell carcinoma of skin of other and unspecified parts of face |
| ICD10CM | C44.320 | Squamous cell carcinoma of skin of unspecified parts of face |
| ICD10CM | C44.321 | Squamous cell carcinoma of skin of nose |
| ICD10CM | C44.329 | Squamous cell carcinoma of skin of other parts of face |
| ICD10CM | C44.39 | Other specified malignant neoplasm of skin of other and unspecified parts of face |
| ICD10CM | C44.390 | Other specified malignant neoplasm of skin of unspecified parts of face |
| ICD10CM | C44.391 | Other specified malignant neoplasm of skin of nose |
| ICD10CM | C44.399 | Other specified malignant neoplasm of skin of other parts of face |
| ICD10CM | C44.4 | Other and unspecified malignant neoplasm of skin of scalp and neck |
| ICD10CM | C44.40 | Unspecified malignant neoplasm of skin of scalp and neck |
| ICD10CM | C44.41 | Basal cell carcinoma of skin of scalp and neck |
| ICD10CM | C44.42 | Squamous cell carcinoma of skin of scalp and neck |
| ICD10CM | C44.49 | Other specified malignant neoplasm of skin of scalp and neck |
| ICD10CM | C44.5 | Other and unspecified malignant neoplasm of skin of trunk |
| ICD10CM | C44.50 | Unspecified malignant neoplasm of skin of trunk |
| ICD10CM | C44.500 | Unspecified malignant neoplasm of anal skin |
| ICD10CM | C44.501 | Unspecified malignant neoplasm of skin of breast |

Continued on next page

**TableS31 – continued from previous page**

| CodeSystem | ConceptCode | ConceptName |
|---|---|---|
| ICD10CM | C44.509 | Unspecified malignant neoplasm of skin of other part of trunk |
| ICD10CM | C44.51 | Basal cell carcinoma of skin of trunk |
| ICD10CM | C44.510 | Basal cell carcinoma of anal skin |
| ICD10CM | C44.511 | Basal cell carcinoma of skin of breast |
| ICD10CM | C44.519 | Basal cell carcinoma of skin of other part of trunk |
| ICD10CM | C44.52 | Squamous cell carcinoma of skin of trunk |
| ICD10CM | C44.520 | Squamous cell carcinoma of anal skin |
| ICD10CM | C44.521 | Squamous cell carcinoma of skin of breast |
| ICD10CM | C44.529 | Squamous cell carcinoma of skin of other part of trunk |
| ICD10CM | C44.59 | Other specified malignant neoplasm of skin of trunk |
| ICD10CM | C44.590 | Other specified malignant neoplasm of anal skin |
| ICD10CM | C44.591 | Other specified malignant neoplasm of skin of breast |
| ICD10CM | C44.599 | Other specified malignant neoplasm of skin of other part of trunk |
| ICD10CM | C44.6 | Other and unspecified malignant neoplasm of skin of upper limb, including shoulder |
| ICD10CM | C44.60 | Unspecified malignant neoplasm of skin of upper limb, including shoulder |
| ICD10CM | C44.601 | Unspecified malignant neoplasm of skin of unspecified upper limb, including shoulder |
| ICD10CM | C44.602 | Unspecified malignant neoplasm of skin of right upper limb, including shoulder |
| ICD10CM | C44.609 | Unspecified malignant neoplasm of skin of left upper limb, including shoulder |
| ICD10CM | C44.61 | Basal cell carcinoma of skin of upper limb, including shoulder |
| ICD10CM | C44.611 | Basal cell carcinoma of skin of unspecified upper limb, including shoulder |
| ICD10CM | C44.612 | Basal cell carcinoma of skin of right upper limb, including shoulder |
| ICD10CM | C44.619 | Basal cell carcinoma of skin of left upper limb, including shoulder |
| ICD10CM | C44.62 | Squamous cell carcinoma of skin of upper limb, including shoulder |
| ICD10CM | C44.621 | Squamous cell carcinoma of skin of unspecified upper limb, including shoulder |
| ICD10CM | C44.622 | Squamous cell carcinoma of skin of right upper limb, including shoulder |
| ICD10CM | C44.629 | Squamous cell carcinoma of skin of left upper limb, including shoulder |
| ICD10CM | C44.69 | Other specified malignant neoplasm of skin of upper limb, including shoulder |
| ICD10CM | C44.691 | Other specified malignant neoplasm of skin of unspecified upper limb, including shoulder |
| ICD10CM | C44.692 | Other specified malignant neoplasm of skin of right upper limb, including shoulder |
| ICD10CM | C44.699 | Other specified malignant neoplasm of skin of left upper limb, including shoulder |
| ICD10CM | C44.7 | Other and unspecified malignant neoplasm of skin of lower limb, including hip |

Continued on next page

**TableS31 – continued from previous page**

| CodeSystem | ConceptCode | ConceptName |
|---|---|---|
| ICD10CM | C44.70 | Unspecified malignant neoplasm of skin of lower limb, including hip |
| ICD10CM | C44.701 | Unspecified malignant neoplasm of skin of unspecified lower limb, including hip |
| ICD10CM | C44.702 | Unspecified malignant neoplasm of skin of right lower limb, including hip |
| ICD10CM | C44.709 | Unspecified malignant neoplasm of skin of left lower limb, including hip |
| ICD10CM | C44.71 | Basal cell carcinoma of skin of lower limb, including hip |
| ICD10CM | C44.711 | Basal cell carcinoma of skin of unspecified lower limb, including hip |
| ICD10CM | C44.712 | Basal cell carcinoma of skin of right lower limb, including hip |
| ICD10CM | C44.719 | Basal cell carcinoma of skin of left lower limb, including hip |
| ICD10CM | C44.72 | Squamous cell carcinoma of skin of lower limb, including hip |
| ICD10CM | C44.721 | Squamous cell carcinoma of skin of unspecified lower limb, including hip |
| ICD10CM | C44.722 | Squamous cell carcinoma of skin of right lower limb, including hip |
| ICD10CM | C44.729 | Squamous cell carcinoma of skin of left lower limb, including hip |
| ICD10CM | C44.79 | Other specified malignant neoplasm of skin of lower limb, including hip |
| ICD10CM | C44.791 | Other specified malignant neoplasm of skin of unspecified lower limb, including hip |
| ICD10CM | C44.792 | Other specified malignant neoplasm of skin of right lower limb, including hip |
| ICD10CM | C44.799 | Other specified malignant neoplasm of skin of left lower limb, including hip |
| ICD10CM | C44.8 | Other and unspecified malignant neoplasm of overlapping sites of skin |
| ICD10CM | C44.80 | Unspecified malignant neoplasm of overlapping sites of skin |
| ICD10CM | C44.81 | Basal cell carcinoma of overlapping sites of skin |
| ICD10CM | C44.82 | Squamous cell carcinoma of overlapping sites of skin |
| ICD10CM | C44.89 | Other specified malignant neoplasm of overlapping sites of skin |
| ICD10CM | C44.9 | Other and unspecified malignant neoplasm of skin, unspecified |
| ICD10CM | C44.90 | Unspecified malignant neoplasm of skin, unspecified |
| ICD10CM | C44.91 | Basal cell carcinoma of skin, unspecified |
| ICD10CM | C44.92 | Squamous cell carcinoma of skin, unspecified |
| ICD10CM | C44.99 | Other specified malignant neoplasm of skin, unspecified |
| ICD10CM | C45.0 | Mesothelioma of pleura |
| ICD10CM | C45.1 | Mesothelioma of peritoneum |
| ICD10CM | C45.2 | Mesothelioma of pericardium |
| ICD10CM | C45.7 | Mesothelioma of other sites |
| ICD10CM | C45.9 | Mesothelioma, unspecified |
| ICD10CM | C46.0 | Kaposi's sarcoma of skin |
| ICD10CM | C46.1 | Kaposi's sarcoma of soft tissue |
| ICD10CM | C46.2 | Kaposi's sarcoma of palate |
| ICD10CM | C46.3 | Kaposi's sarcoma of lymph nodes |

Continued on next page

**TableS31 – continued from previous page**

| CodeSystem | ConceptCode | ConceptName |
|---|---|---|
| ICD10CM | C46.4 | Kaposi's sarcoma of gastrointestinal sites |
| ICD10CM | C46.5 | Kaposi's sarcoma of lung |
| ICD10CM | C46.50 | Kaposi's sarcoma of unspecified lung |
| ICD10CM | C46.51 | Kaposi's sarcoma of right lung |
| ICD10CM | C46.52 | Kaposi's sarcoma of left lung |
| ICD10CM | C46.7 | Kaposi's sarcoma of other sites |
| ICD10CM | C46.9 | Kaposi's sarcoma, unspecified |
| ICD10CM | C47.0 | Malignant neoplasm of peripheral nerves of head, face and neck |
| ICD10CM | C47.1 | Malignant neoplasm of peripheral nerves of upper limb, including shoulder |
| ICD10CM | C47.10 | Malignant neoplasm of peripheral nerves of unspecified upper limb, including shoulder |
| ICD10CM | C47.11 | Malignant neoplasm of peripheral nerves of right upper limb, including shoulder |
| ICD10CM | C47.12 | Malignant neoplasm of peripheral nerves of left upper limb, including shoulder |
| ICD10CM | C47.2 | Malignant neoplasm of peripheral nerves of lower limb, including hip |
| ICD10CM | C47.20 | Malignant neoplasm of peripheral nerves of unspecified lower limb, including hip |
| ICD10CM | C47.21 | Malignant neoplasm of peripheral nerves of right lower limb, including hip |
| ICD10CM | C47.22 | Malignant neoplasm of peripheral nerves of left lower limb, including hip |
| ICD10CM | C47.3 | Malignant neoplasm of peripheral nerves of thorax |
| ICD10CM | C47.4 | Malignant neoplasm of peripheral nerves of abdomen |
| ICD10CM | C47.5 | Malignant neoplasm of peripheral nerves of pelvis |
| ICD10CM | C47.6 | Malignant neoplasm of peripheral nerves of trunk, unspecified |
| ICD10CM | C47.8 | Malignant neoplasm of overlapping sites of peripheral nerves and autonomic nervous system |
| ICD10CM | C47.9 | Malignant neoplasm of peripheral nerves and autonomic nervous system, unspecified |
| ICD10CM | C48.0 | Malignant neoplasm of retroperitoneum |
| ICD10CM | C48.1 | Malignant neoplasm of specified parts of peritoneum |
| ICD10CM | C48.2 | Malignant neoplasm of peritoneum, unspecified |
| ICD10CM | C48.8 | Malignant neoplasm of overlapping sites of retroperitoneum and peritoneum |
| ICD10CM | C49.0 | Malignant neoplasm of connective and soft tissue of head, face and neck |
| ICD10CM | C49.1 | Malignant neoplasm of connective and soft tissue of upper limb, including shoulder |
| ICD10CM | C49.10 | Malignant neoplasm of connective and soft tissue of unspecified upper limb, including shoulder |
| ICD10CM | C49.11 | Malignant neoplasm of connective and soft tissue of right upper limb, including shoulder |
| ICD10CM | C49.12 | Malignant neoplasm of connective and soft tissue of left upper limb, including shoulder |

Continued on next page

**TableS31 – continued from previous page**

| CodeSystem | ConceptCode | ConceptName |
|---|---|---|
| ICD10CM | C49.2 | Malignant neoplasm of connective and soft tissue of lower limb, including hip |
| ICD10CM | C49.20 | Malignant neoplasm of connective and soft tissue of unspecified lower limb, including hip |
| ICD10CM | C49.21 | Malignant neoplasm of connective and soft tissue of right lower limb, including hip |
| ICD10CM | C49.22 | Malignant neoplasm of connective and soft tissue of left lower limb, including hip |
| ICD10CM | C49.3 | Malignant neoplasm of connective and soft tissue of thorax |
| ICD10CM | C49.4 | Malignant neoplasm of connective and soft tissue of abdomen |
| ICD10CM | C49.5 | Malignant neoplasm of connective and soft tissue of pelvis |
| ICD10CM | C49.6 | Malignant neoplasm of connective and soft tissue of trunk, unspecified |
| ICD10CM | C49.8 | Malignant neoplasm of overlapping sites of connective and soft tissue |
| ICD10CM | C49.9 | Malignant neoplasm of connective and soft tissue, unspecified |
| ICD10CM | C49.A | Gastrointestinal stromal tumor |
| ICD10CM | C49.A0 | Gastrointestinal stromal tumor, unspecified site |
| ICD10CM | C49.A1 | Gastrointestinal stromal tumor of esophagus |
| ICD10CM | C49.A2 | Gastrointestinal stromal tumor of stomach |
| ICD10CM | C49.A3 | Gastrointestinal stromal tumor of small intestine |
| ICD10CM | C49.A4 | Gastrointestinal stromal tumor of large intestine |
| ICD10CM | C49.A5 | Gastrointestinal stromal tumor of rectum |
| ICD10CM | C49.A9 | Gastrointestinal stromal tumor of other sites |
| ICD10CM | C50.0 | Malignant neoplasm of nipple and areola |
| ICD10CM | C50.01 | Malignant neoplasm of nipple and areola, female |
| ICD10CM | C50.011 | Malignant neoplasm of nipple and areola, right female breast |
| ICD10CM | C50.012 | Malignant neoplasm of nipple and areola, left female breast |
| ICD10CM | C50.019 | Malignant neoplasm of nipple and areola, unspecified female breast |
| ICD10CM | C50.02 | Malignant neoplasm of nipple and areola, male |
| ICD10CM | C50.021 | Malignant neoplasm of nipple and areola, right male breast |
| ICD10CM | C50.022 | Malignant neoplasm of nipple and areola, left male breast |
| ICD10CM | C50.029 | Malignant neoplasm of nipple and areola, unspecified male breast |
| ICD10CM | C50.1 | Malignant neoplasm of central portion of breast |
| ICD10CM | C50.11 | Malignant neoplasm of central portion of breast, female |
| ICD10CM | C50.111 | Malignant neoplasm of central portion of right female breast |
| ICD10CM | C50.112 | Malignant neoplasm of central portion of left female breast |
| ICD10CM | C50.119 | Malignant neoplasm of central portion of unspecified female breast |
| ICD10CM | C50.12 | Malignant neoplasm of central portion of breast, male |
| ICD10CM | C50.121 | Malignant neoplasm of central portion of right male breast |
| ICD10CM | C50.122 | Malignant neoplasm of central portion of left male breast |
| ICD10CM | C50.129 | Malignant neoplasm of central portion of unspecified male breast |
| ICD10CM | C50.2 | Malignant neoplasm of upper-inner quadrant of breast |
| ICD10CM | C50.21 | Malignant neoplasm of upper-inner quadrant of breast, female |

Continued on next page

**TableS31 – continued from previous page**

| CodeSystem | ConceptCode | ConceptName |
|---|---|---|
| ICD10CM | C50.211 | Malignant neoplasm of upper-inner quadrant of right female breast |
| ICD10CM | C50.212 | Malignant neoplasm of upper-inner quadrant of left female breast |
| ICD10CM | C50.219 | Malignant neoplasm of upper-inner quadrant of unspecified female breast |
| ICD10CM | C50.22 | Malignant neoplasm of upper-inner quadrant of breast, male |
| ICD10CM | C50.221 | Malignant neoplasm of upper-inner quadrant of right male breast |
| ICD10CM | C50.222 | Malignant neoplasm of upper-inner quadrant of left male breast |
| ICD10CM | C50.229 | Malignant neoplasm of upper-inner quadrant of unspecified male breast |
| ICD10CM | C50.3 | Malignant neoplasm of lower-inner quadrant of breast |
| ICD10CM | C50.31 | Malignant neoplasm of lower-inner quadrant of breast, female |
| ICD10CM | C50.311 | Malignant neoplasm of lower-inner quadrant of right female breast |
| ICD10CM | C50.312 | Malignant neoplasm of lower-inner quadrant of left female breast |
| ICD10CM | C50.319 | Malignant neoplasm of lower-inner quadrant of unspecified female breast |
| ICD10CM | C50.32 | Malignant neoplasm of lower-inner quadrant of breast, male |
| ICD10CM | C50.321 | Malignant neoplasm of lower-inner quadrant of right male breast |
| ICD10CM | C50.322 | Malignant neoplasm of lower-inner quadrant of left male breast |
| ICD10CM | C50.329 | Malignant neoplasm of lower-inner quadrant of unspecified male breast |
| ICD10CM | C50.4 | Malignant neoplasm of upper-outer quadrant of breast |
| ICD10CM | C50.41 | Malignant neoplasm of upper-outer quadrant of breast, female |
| ICD10CM | C50.411 | Malignant neoplasm of upper-outer quadrant of right female breast |
| ICD10CM | C50.412 | Malignant neoplasm of upper-outer quadrant of left female breast |
| ICD10CM | C50.419 | Malignant neoplasm of upper-outer quadrant of unspecified female breast |
| ICD10CM | C50.42 | Malignant neoplasm of upper-outer quadrant of breast, male |
| ICD10CM | C50.421 | Malignant neoplasm of upper-outer quadrant of right male breast |
| ICD10CM | C50.422 | Malignant neoplasm of upper-outer quadrant of left male breast |
| ICD10CM | C50.429 | Malignant neoplasm of upper-outer quadrant of unspecified male breast |
| ICD10CM | C50.5 | Malignant neoplasm of lower-outer quadrant of breast |
| ICD10CM | C50.51 | Malignant neoplasm of lower-outer quadrant of breast, female |
| ICD10CM | C50.511 | Malignant neoplasm of lower-outer quadrant of right female breast |
| ICD10CM | C50.512 | Malignant neoplasm of lower-outer quadrant of left female breast |
| ICD10CM | C50.519 | Malignant neoplasm of lower-outer quadrant of unspecified female breast |
| ICD10CM | C50.52 | Malignant neoplasm of lower-outer quadrant of breast, male |
| ICD10CM | C50.521 | Malignant neoplasm of lower-outer quadrant of right male breast |
| ICD10CM | C50.522 | Malignant neoplasm of lower-outer quadrant of left male breast |
| ICD10CM | C50.529 | Malignant neoplasm of lower-outer quadrant of unspecified male breast |
| ICD10CM | C50.6 | Malignant neoplasm of axillary tail of breast |

Continued on next page

**TableS31 – continued from previous page**

| CodeSystem | ConceptCode | ConceptName |
|---|---|---|
| ICD10CM | C50.61 | Malignant neoplasm of axillary tail of breast, female |
| ICD10CM | C50.611 | Malignant neoplasm of axillary tail of right female breast |
| ICD10CM | C50.612 | Malignant neoplasm of axillary tail of left female breast |
| ICD10CM | C50.619 | Malignant neoplasm of axillary tail of unspecified female breast |
| ICD10CM | C50.62 | Malignant neoplasm of axillary tail of breast, male |
| ICD10CM | C50.621 | Malignant neoplasm of axillary tail of right male breast |
| ICD10CM | C50.622 | Malignant neoplasm of axillary tail of left male breast |
| ICD10CM | C50.629 | Malignant neoplasm of axillary tail of unspecified male breast |
| ICD10CM | C50.8 | Malignant neoplasm of overlapping sites of breast |
| ICD10CM | C50.81 | Malignant neoplasm of overlapping sites of breast, female |
| ICD10CM | C50.811 | Malignant neoplasm of overlapping sites of right female breast |
| ICD10CM | C50.812 | Malignant neoplasm of overlapping sites of left female breast |
| ICD10CM | C50.819 | Malignant neoplasm of overlapping sites of unspecified female breast |
| ICD10CM | C50.82 | Malignant neoplasm of overlapping sites of breast, male |
| ICD10CM | C50.821 | Malignant neoplasm of overlapping sites of right male breast |
| ICD10CM | C50.822 | Malignant neoplasm of overlapping sites of left male breast |
| ICD10CM | C50.829 | Malignant neoplasm of overlapping sites of unspecified male breast |
| ICD10CM | C50.9 | Malignant neoplasm of breast of unspecified site |
| ICD10CM | C50.91 | Malignant neoplasm of breast of unspecified site, female |
| ICD10CM | C50.911 | Malignant neoplasm of unspecified site of right female breast |
| ICD10CM | C50.912 | Malignant neoplasm of unspecified site of left female breast |
| ICD10CM | C50.919 | Malignant neoplasm of unspecified site of unspecified female breast |
| ICD10CM | C50.92 | Malignant neoplasm of breast of unspecified site, male |
| ICD10CM | C50.921 | Malignant neoplasm of unspecified site of right male breast |
| ICD10CM | C50.922 | Malignant neoplasm of unspecified site of left male breast |
| ICD10CM | C50.929 | Malignant neoplasm of unspecified site of unspecified male breast |
| ICD10CM | C51.0 | Malignant neoplasm of labium majus |
| ICD10CM | C51.1 | Malignant neoplasm of labium minus |
| ICD10CM | C51.2 | Malignant neoplasm of clitoris |
| ICD10CM | C51.8 | Malignant neoplasm of overlapping sites of vulva |
| ICD10CM | C51.9 | Malignant neoplasm of vulva, unspecified |
| ICD10CM | C53.0 | Malignant neoplasm of endocervix |
| ICD10CM | C53.1 | Malignant neoplasm of exocervix |
| ICD10CM | C53.8 | Malignant neoplasm of overlapping sites of cervix uteri |
| ICD10CM | C53.9 | Malignant neoplasm of cervix uteri, unspecified |
| ICD10CM | C54.0 | Malignant neoplasm of isthmus uteri |
| ICD10CM | C54.1 | Malignant neoplasm of endometrium |
| ICD10CM | C54.2 | Malignant neoplasm of myometrium |
| ICD10CM | C54.3 | Malignant neoplasm of fundus uteri |
| ICD10CM | C54.8 | Malignant neoplasm of overlapping sites of corpus uteri |
| ICD10CM | C54.9 | Malignant neoplasm of corpus uteri, unspecified |
| ICD10CM | C56.1 | Malignant neoplasm of right ovary |
| ICD10CM | C56.2 | Malignant neoplasm of left ovary |
| ICD10CM | C56.9 | Malignant neoplasm of unspecified ovary |

Continued on next page

**TableS31 – continued from previous page**

| CodeSystem | ConceptCode | ConceptName |
|---|---|---|
| ICD10CM | C57.0 | Malignant neoplasm of fallopian tube |
| ICD10CM | C57.00 | Malignant neoplasm of unspecified fallopian tube |
| ICD10CM | C57.01 | Malignant neoplasm of right fallopian tube |
| ICD10CM | C57.02 | Malignant neoplasm of left fallopian tube |
| ICD10CM | C57.1 | Malignant neoplasm of broad ligament |
| ICD10CM | C57.10 | Malignant neoplasm of unspecified broad ligament |
| ICD10CM | C57.11 | Malignant neoplasm of right broad ligament |
| ICD10CM | C57.12 | Malignant neoplasm of left broad ligament |
| ICD10CM | C57.2 | Malignant neoplasm of round ligament |
| ICD10CM | C57.20 | Malignant neoplasm of unspecified round ligament |
| ICD10CM | C57.21 | Malignant neoplasm of right round ligament |
| ICD10CM | C57.22 | Malignant neoplasm of left round ligament |
| ICD10CM | C57.3 | Malignant neoplasm of parametrium |
| ICD10CM | C57.4 | Malignant neoplasm of uterine adnexa, unspecified |
| ICD10CM | C57.7 | Malignant neoplasm of other specified female genital organs |
| ICD10CM | C57.8 | Malignant neoplasm of overlapping sites of female genital organs |
| ICD10CM | C57.9 | Malignant neoplasm of female genital organ, unspecified |
| ICD10CM | C60.0 | Malignant neoplasm of prepuce |
| ICD10CM | C60.1 | Malignant neoplasm of glans penis |
| ICD10CM | C60.2 | Malignant neoplasm of body of penis |
| ICD10CM | C60.8 | Malignant neoplasm of overlapping sites of penis |
| ICD10CM | C60.9 | Malignant neoplasm of penis, unspecified |
| ICD10CM | C62.0 | Malignant neoplasm of undescended testis |
| ICD10CM | C62.00 | Malignant neoplasm of unspecified undescended testis |
| ICD10CM | C62.01 | Malignant neoplasm of undescended right testis |
| ICD10CM | C62.02 | Malignant neoplasm of undescended left testis |
| ICD10CM | C62.1 | Malignant neoplasm of descended testis |
| ICD10CM | C62.10 | Malignant neoplasm of unspecified descended testis |
| ICD10CM | C62.11 | Malignant neoplasm of descended right testis |
| ICD10CM | C62.12 | Malignant neoplasm of descended left testis |
| ICD10CM | C62.9 | Malignant neoplasm of testis, unspecified whether descended or undescended |
| ICD10CM | C62.90 | Malignant neoplasm of unspecified testis, unspecified whether descended or undescended |
| ICD10CM | C62.91 | Malignant neoplasm of right testis, unspecified whether descended or undescended |
| ICD10CM | C62.92 | Malignant neoplasm of left testis, unspecified whether descended or undescended |
| ICD10CM | C63.0 | Malignant neoplasm of epididymis |
| ICD10CM | C63.00 | Malignant neoplasm of unspecified epididymis |
| ICD10CM | C63.01 | Malignant neoplasm of right epididymis |
| ICD10CM | C63.02 | Malignant neoplasm of left epididymis |
| ICD10CM | C63.1 | Malignant neoplasm of spermatic cord |
| ICD10CM | C63.10 | Malignant neoplasm of unspecified spermatic cord |
| ICD10CM | C63.11 | Malignant neoplasm of right spermatic cord |
| ICD10CM | C63.12 | Malignant neoplasm of left spermatic cord |
| ICD10CM | C63.2 | Malignant neoplasm of scrotum |

Continued on next page

**TableS31 – continued from previous page**

| CodeSystem | ConceptCode | ConceptName |
|---|---|---|
| ICD10CM | C63.7 | Malignant neoplasm of other specified male genital organs |
| ICD10CM | C63.8 | Malignant neoplasm of overlapping sites of male genital organs |
| ICD10CM | C63.9 | Malignant neoplasm of male genital organ, unspecified |
| ICD10CM | C64.1 | Malignant neoplasm of right kidney, except renal pelvis |
| ICD10CM | C64.2 | Malignant neoplasm of left kidney, except renal pelvis |
| ICD10CM | C64.9 | Malignant neoplasm of unspecified kidney, except renal pelvis |
| ICD10CM | C65.1 | Malignant neoplasm of right renal pelvis |
| ICD10CM | C65.2 | Malignant neoplasm of left renal pelvis |
| ICD10CM | C65.9 | Malignant neoplasm of unspecified renal pelvis |
| ICD10CM | C66.1 | Malignant neoplasm of right ureter |
| ICD10CM | C66.2 | Malignant neoplasm of left ureter |
| ICD10CM | C66.9 | Malignant neoplasm of unspecified ureter |
| ICD10CM | C67.0 | Malignant neoplasm of trigone of bladder |
| ICD10CM | C67.1 | Malignant neoplasm of dome of bladder |
| ICD10CM | C67.2 | Malignant neoplasm of lateral wall of bladder |
| ICD10CM | C67.3 | Malignant neoplasm of anterior wall of bladder |
| ICD10CM | C67.4 | Malignant neoplasm of posterior wall of bladder |
| ICD10CM | C67.5 | Malignant neoplasm of bladder neck |
| ICD10CM | C67.6 | Malignant neoplasm of ureteric orifice |
| ICD10CM | C67.7 | Malignant neoplasm of urachus |
| ICD10CM | C67.8 | Malignant neoplasm of overlapping sites of bladder |
| ICD10CM | C67.9 | Malignant neoplasm of bladder, unspecified |
| ICD10CM | C68.0 | Malignant neoplasm of urethra |
| ICD10CM | C68.1 | Malignant neoplasm of paraurethral glands |
| ICD10CM | C68.8 | Malignant neoplasm of overlapping sites of urinary organs |
| ICD10CM | C68.9 | Malignant neoplasm of urinary organ, unspecified |
| ICD10CM | C69.0 | Malignant neoplasm of conjunctiva |
| ICD10CM | C69.00 | Malignant neoplasm of unspecified conjunctiva |
| ICD10CM | C69.01 | Malignant neoplasm of right conjunctiva |
| ICD10CM | C69.02 | Malignant neoplasm of left conjunctiva |
| ICD10CM | C69.1 | Malignant neoplasm of cornea |
| ICD10CM | C69.10 | Malignant neoplasm of unspecified cornea |
| ICD10CM | C69.11 | Malignant neoplasm of right cornea |
| ICD10CM | C69.12 | Malignant neoplasm of left cornea |
| ICD10CM | C69.2 | Malignant neoplasm of retina |
| ICD10CM | C69.20 | Malignant neoplasm of unspecified retina |
| ICD10CM | C69.21 | Malignant neoplasm of right retina |
| ICD10CM | C69.22 | Malignant neoplasm of left retina |
| ICD10CM | C69.3 | Malignant neoplasm of choroid |
| ICD10CM | C69.30 | Malignant neoplasm of unspecified choroid |
| ICD10CM | C69.31 | Malignant neoplasm of right choroid |
| ICD10CM | C69.32 | Malignant neoplasm of left choroid |
| ICD10CM | C69.4 | Malignant neoplasm of ciliary body |
| ICD10CM | C69.40 | Malignant neoplasm of unspecified ciliary body |
| ICD10CM | C69.41 | Malignant neoplasm of right ciliary body |
| ICD10CM | C69.42 | Malignant neoplasm of left ciliary body |
| ICD10CM | C69.5 | Malignant neoplasm of lacrimal gland and duct |

Continued on next page

**TableS31 – continued from previous page**

| CodeSystem | ConceptCode | ConceptName |
|---|---|---|
| ICD10CM | C69.50 | Malignant neoplasm of unspecified lacrimal gland and duct |
| ICD10CM | C69.51 | Malignant neoplasm of right lacrimal gland and duct |
| ICD10CM | C69.52 | Malignant neoplasm of left lacrimal gland and duct |
| ICD10CM | C69.6 | Malignant neoplasm of orbit |
| ICD10CM | C69.60 | Malignant neoplasm of unspecified orbit |
| ICD10CM | C69.61 | Malignant neoplasm of right orbit |
| ICD10CM | C69.62 | Malignant neoplasm of left orbit |
| ICD10CM | C69.8 | Malignant neoplasm of overlapping sites of eye and adnexa |
| ICD10CM | C69.80 | Malignant neoplasm of overlapping sites of unspecified eye and adnexa |
| ICD10CM | C69.81 | Malignant neoplasm of overlapping sites of right eye and adnexa |
| ICD10CM | C69.82 | Malignant neoplasm of overlapping sites of left eye and adnexa |
| ICD10CM | C69.9 | Malignant neoplasm of unspecified site of eye |
| ICD10CM | C69.90 | Malignant neoplasm of unspecified site of unspecified eye |
| ICD10CM | C69.91 | Malignant neoplasm of unspecified site of right eye |
| ICD10CM | C69.92 | Malignant neoplasm of unspecified site of left eye |
| ICD10CM | C70.0 | Malignant neoplasm of cerebral meninges |
| ICD10CM | C70.1 | Malignant neoplasm of spinal meninges |
| ICD10CM | C70.9 | Malignant neoplasm of meninges, unspecified |
| ICD10CM | C71.0 | Malignant neoplasm of cerebrum, except lobes and ventricles |
| ICD10CM | C71.1 | Malignant neoplasm of frontal lobe |
| ICD10CM | C71.2 | Malignant neoplasm of temporal lobe |
| ICD10CM | C71.3 | Malignant neoplasm of parietal lobe |
| ICD10CM | C71.4 | Malignant neoplasm of occipital lobe |
| ICD10CM | C71.5 | Malignant neoplasm of cerebral ventricle |
| ICD10CM | C71.6 | Malignant neoplasm of cerebellum |
| ICD10CM | C71.7 | Malignant neoplasm of brain stem |
| ICD10CM | C71.8 | Malignant neoplasm of overlapping sites of brain |
| ICD10CM | C71.9 | Malignant neoplasm of brain, unspecified |
| ICD10CM | C72.0 | Malignant neoplasm of spinal cord |
| ICD10CM | C72.1 | Malignant neoplasm of cauda equina |
| ICD10CM | C72.2 | Malignant neoplasm of olfactory nerve |
| ICD10CM | C72.20 | Malignant neoplasm of unspecified olfactory nerve |
| ICD10CM | C72.21 | Malignant neoplasm of right olfactory nerve |
| ICD10CM | C72.22 | Malignant neoplasm of left olfactory nerve |
| ICD10CM | C72.3 | Malignant neoplasm of optic nerve |
| ICD10CM | C72.30 | Malignant neoplasm of unspecified optic nerve |
| ICD10CM | C72.31 | Malignant neoplasm of right optic nerve |
| ICD10CM | C72.32 | Malignant neoplasm of left optic nerve |
| ICD10CM | C72.4 | Malignant neoplasm of acoustic nerve |
| ICD10CM | C72.40 | Malignant neoplasm of unspecified acoustic nerve |
| ICD10CM | C72.41 | Malignant neoplasm of right acoustic nerve |
| ICD10CM | C72.42 | Malignant neoplasm of left acoustic nerve |
| ICD10CM | C72.5 | Malignant neoplasm of other and unspecified cranial nerves |
| ICD10CM | C72.50 | Malignant neoplasm of unspecified cranial nerve |
| ICD10CM | C72.59 | Malignant neoplasm of other cranial nerves |
| ICD10CM | C72.9 | Malignant neoplasm of central nervous system, unspecified |

Continued on next page

**TableS31 – continued from previous page**

| CodeSystem | ConceptCode | ConceptName |
|---|---|---|
| ICD10CM | C74.0 | Malignant neoplasm of cortex of adrenal gland |
| ICD10CM | C74.00 | Malignant neoplasm of cortex of unspecified adrenal gland |
| ICD10CM | C74.01 | Malignant neoplasm of cortex of right adrenal gland |
| ICD10CM | C74.02 | Malignant neoplasm of cortex of left adrenal gland |
| ICD10CM | C74.1 | Malignant neoplasm of medulla of adrenal gland |
| ICD10CM | C74.10 | Malignant neoplasm of medulla of unspecified adrenal gland |
| ICD10CM | C74.11 | Malignant neoplasm of medulla of right adrenal gland |
| ICD10CM | C74.12 | Malignant neoplasm of medulla of left adrenal gland |
| ICD10CM | C74.9 | Malignant neoplasm of unspecified part of adrenal gland |
| ICD10CM | C74.90 | Malignant neoplasm of unspecified part of unspecified adrenal gland |
| ICD10CM | C74.91 | Malignant neoplasm of unspecified part of right adrenal gland |
| ICD10CM | C74.92 | Malignant neoplasm of unspecified part of left adrenal gland |
| ICD10CM | C75.0 | Malignant neoplasm of parathyroid gland |
| ICD10CM | C75.1 | Malignant neoplasm of pituitary gland |
| ICD10CM | C75.2 | Malignant neoplasm of craniopharyngeal duct |
| ICD10CM | C75.3 | Malignant neoplasm of pineal gland |
| ICD10CM | C75.4 | Malignant neoplasm of carotid body |
| ICD10CM | C75.5 | Malignant neoplasm of aortic body and other paraganglia |
| ICD10CM | C75.8 | Malignant neoplasm with pluriglandular involvement, unspecified |
| ICD10CM | C75.9 | Malignant neoplasm of endocrine gland, unspecified |
| ICD10CM | C76.0 | Malignant neoplasm of head, face and neck |
| ICD10CM | C76.1 | Malignant neoplasm of thorax |
| ICD10CM | C76.2 | Malignant neoplasm of abdomen |
| ICD10CM | C76.3 | Malignant neoplasm of pelvis |
| ICD10CM | C76.4 | Malignant neoplasm of upper limb |
| ICD10CM | C76.40 | Malignant neoplasm of unspecified upper limb |
| ICD10CM | C76.41 | Malignant neoplasm of right upper limb |
| ICD10CM | C76.42 | Malignant neoplasm of left upper limb |
| ICD10CM | C76.5 | Malignant neoplasm of lower limb |
| ICD10CM | C76.50 | Malignant neoplasm of unspecified lower limb |
| ICD10CM | C76.51 | Malignant neoplasm of right lower limb |
| ICD10CM | C76.52 | Malignant neoplasm of left lower limb |
| ICD10CM | C76.8 | Malignant neoplasm of other specified ill-defined sites |
| ICD10CM | C77.0 | Secondary and unspecified malignant neoplasm of lymph nodes of head, face and neck |
| ICD10CM | C77.1 | Secondary and unspecified malignant neoplasm of intrathoracic lymph nodes |
| ICD10CM | C77.2 | Secondary and unspecified malignant neoplasm of intra-abdominal lymph nodes |
| ICD10CM | C77.3 | Secondary and unspecified malignant neoplasm of axilla and upper limb lymph nodes |
| ICD10CM | C77.4 | Secondary and unspecified malignant neoplasm of inguinal and lower limb lymph nodes |
| ICD10CM | C77.5 | Secondary and unspecified malignant neoplasm of intrapelvic lymph nodes |

Continued on next page

**TableS31 – continued from previous page**

| CodeSystem | ConceptCode | ConceptName |
|---|---|---|
| ICD10CM | C77.8 | Secondary and unspecified malignant neoplasm of lymph nodes of multiple regions |
| ICD10CM | C77.9 | Secondary and unspecified malignant neoplasm of lymph node, unspecified |
| ICD10CM | C78.0 | Secondary malignant neoplasm of lung |
| ICD10CM | C78.00 | Secondary malignant neoplasm of unspecified lung |
| ICD10CM | C78.01 | Secondary malignant neoplasm of right lung |
| ICD10CM | C78.02 | Secondary malignant neoplasm of left lung |
| ICD10CM | C78.1 | Secondary malignant neoplasm of mediastinum |
| ICD10CM | C78.2 | Secondary malignant neoplasm of pleura |
| ICD10CM | C78.3 | Secondary malignant neoplasm of other and unspecified respiratory organs |
| ICD10CM | C78.30 | Secondary malignant neoplasm of unspecified respiratory organ |
| ICD10CM | C78.39 | Secondary malignant neoplasm of other respiratory organs |
| ICD10CM | C78.4 | Secondary malignant neoplasm of small intestine |
| ICD10CM | C78.5 | Secondary malignant neoplasm of large intestine and rectum |
| ICD10CM | C78.6 | Secondary malignant neoplasm of retroperitoneum and peritoneum |
| ICD10CM | C78.7 | Secondary malignant neoplasm of liver and intrahepatic bile duct |
| ICD10CM | C78.8 | Secondary malignant neoplasm of other and unspecified digestive organs |
| ICD10CM | C78.80 | Secondary malignant neoplasm of unspecified digestive organ |
| ICD10CM | C78.89 | Secondary malignant neoplasm of other digestive organs |
| ICD10CM | C79.0 | Secondary malignant neoplasm of kidney and renal pelvis |
| ICD10CM | C79.00 | Secondary malignant neoplasm of unspecified kidney and renal pelvis |
| ICD10CM | C79.01 | Secondary malignant neoplasm of right kidney and renal pelvis |
| ICD10CM | C79.02 | Secondary malignant neoplasm of left kidney and renal pelvis |
| ICD10CM | C79.1 | Secondary malignant neoplasm of bladder and other and unspecified urinary organs |
| ICD10CM | C79.10 | Secondary malignant neoplasm of unspecified urinary organs |
| ICD10CM | C79.11 | Secondary malignant neoplasm of bladder |
| ICD10CM | C79.19 | Secondary malignant neoplasm of other urinary organs |
| ICD10CM | C79.2 | Secondary malignant neoplasm of skin |
| ICD10CM | C79.3 | Secondary malignant neoplasm of brain and cerebral meninges |
| ICD10CM | C79.31 | Secondary malignant neoplasm of brain |
| ICD10CM | C79.32 | Secondary malignant neoplasm of cerebral meninges |
| ICD10CM | C79.4 | Secondary malignant neoplasm of other and unspecified parts of nervous system |
| ICD10CM | C79.40 | Secondary malignant neoplasm of unspecified part of nervous system |
| ICD10CM | C79.49 | Secondary malignant neoplasm of other parts of nervous system |
| ICD10CM | C79.5 | Secondary malignant neoplasm of bone and bone marrow |
| ICD10CM | C79.51 | Secondary malignant neoplasm of bone |
| ICD10CM | C79.52 | Secondary malignant neoplasm of bone marrow |
| ICD10CM | C79.6 | Secondary malignant neoplasm of ovary |
| ICD10CM | C79.60 | Secondary malignant neoplasm of unspecified ovary |

Continued on next page

**TableS31 – continued from previous page**

| CodeSystem | ConceptCode | ConceptName |
|---|---|---|
| ICD10CM | C79.61 | Secondary malignant neoplasm of right ovary |
| ICD10CM | C79.62 | Secondary malignant neoplasm of left ovary |
| ICD10CM | C79.7 | Secondary malignant neoplasm of adrenal gland |
| ICD10CM | C79.70 | Secondary malignant neoplasm of unspecified adrenal gland |
| ICD10CM | C79.71 | Secondary malignant neoplasm of right adrenal gland |
| ICD10CM | C79.72 | Secondary malignant neoplasm of left adrenal gland |
| ICD10CM | C79.8 | Secondary malignant neoplasm of other specified sites |
| ICD10CM | C79.81 | Secondary malignant neoplasm of breast |
| ICD10CM | C79.82 | Secondary malignant neoplasm of genital organs |
| ICD10CM | C79.89 | Secondary malignant neoplasm of other specified sites |
| ICD10CM | C79.9 | Secondary malignant neoplasm of unspecified site |
| ICD10CM | C7A.0 | Malignant carcinoid tumors |
| ICD10CM | C7A.00 | Malignant carcinoid tumor of unspecified site |
| ICD10CM | C7A.01 | Malignant carcinoid tumors of the small intestine |
| ICD10CM | C7A.010 | Malignant carcinoid tumor of the duodenum |
| ICD10CM | C7A.011 | Malignant carcinoid tumor of the jejunum |
| ICD10CM | C7A.012 | Malignant carcinoid tumor of the ileum |
| ICD10CM | C7A.019 | Malignant carcinoid tumor of the small intestine, unspecified portion |
| ICD10CM | C7A.02 | Malignant carcinoid tumors of the appendix, large intestine, and rectum |
| ICD10CM | C7A.020 | Malignant carcinoid tumor of the appendix |
| ICD10CM | C7A.021 | Malignant carcinoid tumor of the cecum |
| ICD10CM | C7A.022 | Malignant carcinoid tumor of the ascending colon |
| ICD10CM | C7A.023 | Malignant carcinoid tumor of the transverse colon |
| ICD10CM | C7A.024 | Malignant carcinoid tumor of the descending colon |
| ICD10CM | C7A.025 | Malignant carcinoid tumor of the sigmoid colon |
| ICD10CM | C7A.026 | Malignant carcinoid tumor of the rectum |
| ICD10CM | C7A.029 | Malignant carcinoid tumor of the large intestine, unspecified portion |
| ICD10CM | C7A.09 | Malignant carcinoid tumors of other sites |
| ICD10CM | C7A.090 | Malignant carcinoid tumor of the bronchus and lung |
| ICD10CM | C7A.091 | Malignant carcinoid tumor of the thymus |
| ICD10CM | C7A.092 | Malignant carcinoid tumor of the stomach |
| ICD10CM | C7A.093 | Malignant carcinoid tumor of the kidney |
| ICD10CM | C7A.094 | Malignant carcinoid tumor of the foregut, unspecified |
| ICD10CM | C7A.095 | Malignant carcinoid tumor of the midgut, unspecified |
| ICD10CM | C7A.096 | Malignant carcinoid tumor of the hindgut, unspecified |
| ICD10CM | C7A.098 | Malignant carcinoid tumors of other sites |
| ICD10CM | C7A.1 | Malignant poorly differentiated neuroendocrine tumors |
| ICD10CM | C7A.8 | Other malignant neuroendocrine tumors |
| ICD10CM | C7B.0 | Secondary carcinoid tumors |
| ICD10CM | C7B.00 | Secondary carcinoid tumors, unspecified site |
| ICD10CM | C7B.01 | Secondary carcinoid tumors of distant lymph nodes |
| ICD10CM | C7B.02 | Secondary carcinoid tumors of liver |
| ICD10CM | C7B.03 | Secondary carcinoid tumors of bone |
| ICD10CM | C7B.04 | Secondary carcinoid tumors of peritoneum |

Continued on next page

**TableS31 – continued from previous page**

| CodeSystem | ConceptCode | ConceptName |
|---|---|---|
| ICD10CM | C7B.09 | Secondary carcinoid tumors of other sites |
| ICD10CM | C7B.1 | Secondary Merkel cell carcinoma |
| ICD10CM | C7B.8 | Other secondary neuroendocrine tumors |
| ICD10CM | C80.0 | Disseminated malignant neoplasm, unspecified |
| ICD10CM | C80.1 | Malignant (primary) neoplasm, unspecified |
| ICD10CM | C80.2 | Malignant neoplasm associated with transplanted organ |
| ICD10CM | C81.0 | Nodular lymphocyte predominant Hodgkin lymphoma |
| ICD10CM | C81.00 | Nodular lymphocyte predominant Hodgkin lymphoma, unspecified site |
| ICD10CM | C81.01 | Nodular lymphocyte predominant Hodgkin lymphoma, lymph nodes of head, face, and neck |
| ICD10CM | C81.02 | Nodular lymphocyte predominant Hodgkin lymphoma, intrathoracic lymph nodes |
| ICD10CM | C81.03 | Nodular lymphocyte predominant Hodgkin lymphoma, intra-abdominal lymph nodes |
| ICD10CM | C81.04 | Nodular lymphocyte predominant Hodgkin lymphoma, lymph nodes of axilla and upper limb |
| ICD10CM | C81.05 | Nodular lymphocyte predominant Hodgkin lymphoma, lymph nodes of inguinal region and lower limb |
| ICD10CM | C81.06 | Nodular lymphocyte predominant Hodgkin lymphoma, intrapelvic lymph nodes |
| ICD10CM | C81.07 | Nodular lymphocyte predominant Hodgkin lymphoma, spleen |
| ICD10CM | C81.08 | Nodular lymphocyte predominant Hodgkin lymphoma, lymph nodes of multiple sites |
| ICD10CM | C81.09 | Nodular lymphocyte predominant Hodgkin lymphoma, extranodal and solid organ sites |
| ICD10CM | C81.1 | Nodular sclerosis Hodgkin lymphoma |
| ICD10CM | C81.10 | Nodular sclerosis Hodgkin lymphoma, unspecified site |
| ICD10CM | C81.11 | Nodular sclerosis Hodgkin lymphoma, lymph nodes of head, face, and neck |
| ICD10CM | C81.12 | Nodular sclerosis Hodgkin lymphoma, intrathoracic lymph nodes |
| ICD10CM | C81.13 | Nodular sclerosis Hodgkin lymphoma, intra-abdominal lymph nodes |
| ICD10CM | C81.14 | Nodular sclerosis Hodgkin lymphoma, lymph nodes of axilla and upper limb |
| ICD10CM | C81.15 | Nodular sclerosis Hodgkin lymphoma, lymph nodes of inguinal region and lower limb |
| ICD10CM | C81.16 | Nodular sclerosis Hodgkin lymphoma, intrapelvic lymph nodes |
| ICD10CM | C81.17 | Nodular sclerosis Hodgkin lymphoma, spleen |
| ICD10CM | C81.18 | Nodular sclerosis Hodgkin lymphoma, lymph nodes of multiple sites |
| ICD10CM | C81.19 | Nodular sclerosis Hodgkin lymphoma, extranodal and solid organ sites |
| ICD10CM | C81.2 | Mixed cellularity Hodgkin lymphoma |
| ICD10CM | C81.20 | Mixed cellularity Hodgkin lymphoma, unspecified site |
| ICD10CM | C81.21 | Mixed cellularity Hodgkin lymphoma, lymph nodes of head, face, and neck |

Continued on next page

**TableS31 – continued from previous page**

| CodeSystem | ConceptCode | ConceptName |
|---|---|---|
| ICD10CM | C81.22 | Mixed cellularity Hodgkin lymphoma, intrathoracic lymph nodes |
| ICD10CM | C81.23 | Mixed cellularity Hodgkin lymphoma, intra-abdominal lymph nodes |
| ICD10CM | C81.24 | Mixed cellularity Hodgkin lymphoma, lymph nodes of axilla and upper limb |
| ICD10CM | C81.25 | Mixed cellularity Hodgkin lymphoma, lymph nodes of inguinal region and lower limb |
| ICD10CM | C81.26 | Mixed cellularity Hodgkin lymphoma, intrapelvic lymph nodes |
| ICD10CM | C81.27 | Mixed cellularity Hodgkin lymphoma, spleen |
| ICD10CM | C81.28 | Mixed cellularity Hodgkin lymphoma, lymph nodes of multiple sites |
| ICD10CM | C81.29 | Mixed cellularity Hodgkin lymphoma, extranodal and solid organ sites |
| ICD10CM | C81.3 | Lymphocyte depleted Hodgkin lymphoma |
| ICD10CM | C81.30 | Lymphocyte depleted Hodgkin lymphoma, unspecified site |
| ICD10CM | C81.31 | Lymphocyte depleted Hodgkin lymphoma, lymph nodes of head, face, and neck |
| ICD10CM | C81.32 | Lymphocyte depleted Hodgkin lymphoma, intrathoracic lymph nodes |
| ICD10CM | C81.33 | Lymphocyte depleted Hodgkin lymphoma, intra-abdominal lymph nodes |
| ICD10CM | C81.34 | Lymphocyte depleted Hodgkin lymphoma, lymph nodes of axilla and upper limb |
| ICD10CM | C81.35 | Lymphocyte depleted Hodgkin lymphoma, lymph nodes of inguinal region and lower limb |
| ICD10CM | C81.36 | Lymphocyte depleted Hodgkin lymphoma, intrapelvic lymph nodes |
| ICD10CM | C81.37 | Lymphocyte depleted Hodgkin lymphoma, spleen |
| ICD10CM | C81.38 | Lymphocyte depleted Hodgkin lymphoma, lymph nodes of multiple sites |
| ICD10CM | C81.39 | Lymphocyte depleted Hodgkin lymphoma, extranodal and solid organ sites |
| ICD10CM | C81.4 | Lymphocyte-rich Hodgkin lymphoma |
| ICD10CM | C81.40 | Lymphocyte-rich Hodgkin lymphoma, unspecified site |
| ICD10CM | C81.41 | Lymphocyte-rich Hodgkin lymphoma, lymph nodes of head, face, and neck |
| ICD10CM | C81.42 | Lymphocyte-rich Hodgkin lymphoma, intrathoracic lymph nodes |
| ICD10CM | C81.43 | Lymphocyte-rich Hodgkin lymphoma, intra-abdominal lymph nodes |
| ICD10CM | C81.44 | Lymphocyte-rich Hodgkin lymphoma, lymph nodes of axilla and upper limb |
| ICD10CM | C81.45 | Lymphocyte-rich Hodgkin lymphoma, lymph nodes of inguinal region and lower limb |
| ICD10CM | C81.46 | Lymphocyte-rich Hodgkin lymphoma, intrapelvic lymph nodes |
| ICD10CM | C81.47 | Lymphocyte-rich Hodgkin lymphoma, spleen |

Continued on next page

**TableS31 – continued from previous page**

| CodeSystem | ConceptCode | ConceptName |
|---|---|---|
| ICD10CM | C81.48 | Lymphocyte-rich Hodgkin lymphoma, lymph nodes of multiple sites |
| ICD10CM | C81.49 | Lymphocyte-rich Hodgkin lymphoma, extranodal and solid organ sites |
| ICD10CM | C81.7 | Other Hodgkin lymphoma |
| ICD10CM | C81.70 | Other Hodgkin lymphoma, unspecified site |
| ICD10CM | C81.71 | Other Hodgkin lymphoma, lymph nodes of head, face, and neck |
| ICD10CM | C81.72 | Other Hodgkin lymphoma, intrathoracic lymph nodes |
| ICD10CM | C81.73 | Other Hodgkin lymphoma, intra-abdominal lymph nodes |
| ICD10CM | C81.74 | Other Hodgkin lymphoma, lymph nodes of axilla and upper limb |
| ICD10CM | C81.75 | Other Hodgkin lymphoma, lymph nodes of inguinal region and lower limb |
| ICD10CM | C81.76 | Other Hodgkin lymphoma, intrapelvic lymph nodes |
| ICD10CM | C81.77 | Other Hodgkin lymphoma, spleen |
| ICD10CM | C81.78 | Other Hodgkin lymphoma, lymph nodes of multiple sites |
| ICD10CM | C81.79 | Other Hodgkin lymphoma, extranodal and solid organ sites |
| ICD10CM | C81.9 | Hodgkin lymphoma, unspecified |
| ICD10CM | C81.90 | Hodgkin lymphoma, unspecified, unspecified site |
| ICD10CM | C81.91 | Hodgkin lymphoma, unspecified, lymph nodes of head, face, and neck |
| ICD10CM | C81.92 | Hodgkin lymphoma, unspecified, intrathoracic lymph nodes |
| ICD10CM | C81.93 | Hodgkin lymphoma, unspecified, intra-abdominal lymph nodes |
| ICD10CM | C81.94 | Hodgkin lymphoma, unspecified, lymph nodes of axilla and upper limb |
| ICD10CM | C81.95 | Hodgkin lymphoma, unspecified, lymph nodes of inguinal region and lower limb |
| ICD10CM | C81.96 | Hodgkin lymphoma, unspecified, intrapelvic lymph nodes |
| ICD10CM | C81.97 | Hodgkin lymphoma, unspecified, spleen |
| ICD10CM | C81.98 | Hodgkin lymphoma, unspecified, lymph nodes of multiple sites |
| ICD10CM | C81.99 | Hodgkin lymphoma, unspecified, extranodal and solid organ sites |
| ICD10CM | C82.0 | Follicular lymphoma grade I |
| ICD10CM | C82.00 | Follicular lymphoma grade I, unspecified site |
| ICD10CM | C82.01 | Follicular lymphoma grade I, lymph nodes of head, face, and neck |
| ICD10CM | C82.02 | Follicular lymphoma grade I, intrathoracic lymph nodes |
| ICD10CM | C82.03 | Follicular lymphoma grade I, intra-abdominal lymph nodes |
| ICD10CM | C82.04 | Follicular lymphoma grade I, lymph nodes of axilla and upper limb |
| ICD10CM | C82.05 | Follicular lymphoma grade I, lymph nodes of inguinal region and lower limb |
| ICD10CM | C82.06 | Follicular lymphoma grade I, intrapelvic lymph nodes |
| ICD10CM | C82.07 | Follicular lymphoma grade I, spleen |
| ICD10CM | C82.08 | Follicular lymphoma grade I, lymph nodes of multiple sites |
| ICD10CM | C82.09 | Follicular lymphoma grade I, extranodal and solid organ sites |
| ICD10CM | C82.1 | Follicular lymphoma grade II |
| ICD10CM | C82.10 | Follicular lymphoma grade II, unspecified site |

Continued on next page

**TableS31 – continued from previous page**

| CodeSystem | ConceptCode | ConceptName |
|---|---|---|
| ICD10CM | C82.11 | Follicular lymphoma grade II, lymph nodes of head, face, and neck |
| ICD10CM | C82.12 | Follicular lymphoma grade II, intrathoracic lymph nodes |
| ICD10CM | C82.13 | Follicular lymphoma grade II, intra-abdominal lymph nodes |
| ICD10CM | C82.14 | Follicular lymphoma grade II, lymph nodes of axilla and upper limb |
| ICD10CM | C82.15 | Follicular lymphoma grade II, lymph nodes of inguinal region and lower limb |
| ICD10CM | C82.16 | Follicular lymphoma grade II, intrapelvic lymph nodes |
| ICD10CM | C82.17 | Follicular lymphoma grade II, spleen |
| ICD10CM | C82.18 | Follicular lymphoma grade II, lymph nodes of multiple sites |
| ICD10CM | C82.19 | Follicular lymphoma grade II, extranodal and solid organ sites |
| ICD10CM | C82.2 | Follicular lymphoma grade III, unspecified |
| ICD10CM | C82.20 | Follicular lymphoma grade III, unspecified, unspecified site |
| ICD10CM | C82.21 | Follicular lymphoma grade III, unspecified, lymph nodes of head, face, and neck |
| ICD10CM | C82.22 | Follicular lymphoma grade III, unspecified, intrathoracic lymph nodes |
| ICD10CM | C82.23 | Follicular lymphoma grade III, unspecified, intra-abdominal lymph nodes |
| ICD10CM | C82.24 | Follicular lymphoma grade III, unspecified, lymph nodes of axilla and upper limb |
| ICD10CM | C82.25 | Follicular lymphoma grade III, unspecified, lymph nodes of inguinal region and lower limb |
| ICD10CM | C82.26 | Follicular lymphoma grade III, unspecified, intrapelvic lymph nodes |
| ICD10CM | C82.27 | Follicular lymphoma grade III, unspecified, spleen |
| ICD10CM | C82.28 | Follicular lymphoma grade III, unspecified, lymph nodes of multiple sites |
| ICD10CM | C82.29 | Follicular lymphoma grade III, unspecified, extranodal and solid organ sites |
| ICD10CM | C82.3 | Follicular lymphoma grade IIIa |
| ICD10CM | C82.30 | Follicular lymphoma grade IIIa, unspecified site |
| ICD10CM | C82.31 | Follicular lymphoma grade IIIa, lymph nodes of head, face, and neck |
| ICD10CM | C82.32 | Follicular lymphoma grade IIIa, intrathoracic lymph nodes |
| ICD10CM | C82.33 | Follicular lymphoma grade IIIa, intra-abdominal lymph nodes |
| ICD10CM | C82.34 | Follicular lymphoma grade IIIa, lymph nodes of axilla and upper limb |
| ICD10CM | C82.35 | Follicular lymphoma grade IIIa, lymph nodes of inguinal region and lower limb |
| ICD10CM | C82.36 | Follicular lymphoma grade IIIa, intrapelvic lymph nodes |
| ICD10CM | C82.37 | Follicular lymphoma grade IIIa, spleen |
| ICD10CM | C82.38 | Follicular lymphoma grade IIIa, lymph nodes of multiple sites |
| ICD10CM | C82.39 | Follicular lymphoma grade IIIa, extranodal and solid organ sites |
| ICD10CM | C82.4 | Follicular lymphoma grade IIIb |
| ICD10CM | C82.40 | Follicular lymphoma grade IIIb, unspecified site |

Continued on next page

**TableS31 – continued from previous page**

| CodeSystem | ConceptCode | ConceptName |
|---|---|---|
| ICD10CM | C82.41 | Follicular lymphoma grade IIIb, lymph nodes of head, face, and neck |
| ICD10CM | C82.42 | Follicular lymphoma grade IIIb, intrathoracic lymph nodes |
| ICD10CM | C82.43 | Follicular lymphoma grade IIIb, intra-abdominal lymph nodes |
| ICD10CM | C82.44 | Follicular lymphoma grade IIIb, lymph nodes of axilla and upper limb |
| ICD10CM | C82.45 | Follicular lymphoma grade IIIb, lymph nodes of inguinal region and lower limb |
| ICD10CM | C82.46 | Follicular lymphoma grade IIIb, intrapelvic lymph nodes |
| ICD10CM | C82.47 | Follicular lymphoma grade IIIb, spleen |
| ICD10CM | C82.48 | Follicular lymphoma grade IIIb, lymph nodes of multiple sites |
| ICD10CM | C82.49 | Follicular lymphoma grade IIIb, extranodal and solid organ sites |
| ICD10CM | C82.5 | Diffuse follicle center lymphoma |
| ICD10CM | C82.50 | Diffuse follicle center lymphoma, unspecified site |
| ICD10CM | C82.51 | Diffuse follicle center lymphoma, lymph nodes of head, face, and neck |
| ICD10CM | C82.52 | Diffuse follicle center lymphoma, intrathoracic lymph nodes |
| ICD10CM | C82.53 | Diffuse follicle center lymphoma, intra-abdominal lymph nodes |
| ICD10CM | C82.54 | Diffuse follicle center lymphoma, lymph nodes of axilla and upper limb |
| ICD10CM | C82.55 | Diffuse follicle center lymphoma, lymph nodes of inguinal region and lower limb |
| ICD10CM | C82.56 | Diffuse follicle center lymphoma, intrapelvic lymph nodes |
| ICD10CM | C82.57 | Diffuse follicle center lymphoma, spleen |
| ICD10CM | C82.58 | Diffuse follicle center lymphoma, lymph nodes of multiple sites |
| ICD10CM | C82.59 | Diffuse follicle center lymphoma, extranodal and solid organ sites |
| ICD10CM | C82.6 | Cutaneous follicle center lymphoma |
| ICD10CM | C82.60 | Cutaneous follicle center lymphoma, unspecified site |
| ICD10CM | C82.61 | Cutaneous follicle center lymphoma, lymph nodes of head, face, and neck |
| ICD10CM | C82.62 | Cutaneous follicle center lymphoma, intrathoracic lymph nodes |
| ICD10CM | C82.63 | Cutaneous follicle center lymphoma, intra-abdominal lymph nodes |
| ICD10CM | C82.64 | Cutaneous follicle center lymphoma, lymph nodes of axilla and upper limb |
| ICD10CM | C82.65 | Cutaneous follicle center lymphoma, lymph nodes of inguinal region and lower limb |
| ICD10CM | C82.66 | Cutaneous follicle center lymphoma, intrapelvic lymph nodes |
| ICD10CM | C82.67 | Cutaneous follicle center lymphoma, spleen |
| ICD10CM | C82.68 | Cutaneous follicle center lymphoma, lymph nodes of multiple sites |
| ICD10CM | C82.69 | Cutaneous follicle center lymphoma, extranodal and solid organ sites |
| ICD10CM | C82.8 | Other types of follicular lymphoma |
| ICD10CM | C82.80 | Other types of follicular lymphoma, unspecified site |
| ICD10CM | C82.81 | Other types of follicular lymphoma, lymph nodes of head, face, and neck |

Continued on next page

**TableS31 – continued from previous page**

| CodeSystem | ConceptCode | ConceptName |
|---|---|---|
| ICD10CM | C82.82 | Other types of follicular lymphoma, intrathoracic lymph nodes |
| ICD10CM | C82.83 | Other types of follicular lymphoma, intra-abdominal lymph nodes |
| ICD10CM | C82.84 | Other types of follicular lymphoma, lymph nodes of axilla and upper limb |
| ICD10CM | C82.85 | Other types of follicular lymphoma, lymph nodes of inguinal region and lower limb |
| ICD10CM | C82.86 | Other types of follicular lymphoma, intrapelvic lymph nodes |
| ICD10CM | C82.87 | Other types of follicular lymphoma, spleen |
| ICD10CM | C82.88 | Other types of follicular lymphoma, lymph nodes of multiple sites |
| ICD10CM | C82.89 | Other types of follicular lymphoma, extranodal and solid organ sites |
| ICD10CM | C82.9 | Follicular lymphoma, unspecified |
| ICD10CM | C82.90 | Follicular lymphoma, unspecified, unspecified site |
| ICD10CM | C82.91 | Follicular lymphoma, unspecified, lymph nodes of head, face, and neck |
| ICD10CM | C82.92 | Follicular lymphoma, unspecified, intrathoracic lymph nodes |
| ICD10CM | C82.93 | Follicular lymphoma, unspecified, intra-abdominal lymph nodes |
| ICD10CM | C82.94 | Follicular lymphoma, unspecified, lymph nodes of axilla and upper limb |
| ICD10CM | C82.95 | Follicular lymphoma, unspecified, lymph nodes of inguinal region and lower limb |
| ICD10CM | C82.96 | Follicular lymphoma, unspecified, intrapelvic lymph nodes |
| ICD10CM | C82.97 | Follicular lymphoma, unspecified, spleen |
| ICD10CM | C82.98 | Follicular lymphoma, unspecified, lymph nodes of multiple sites |
| ICD10CM | C82.99 | Follicular lymphoma, unspecified, extranodal and solid organ sites |
| ICD10CM | C83.0 | Small cell B-cell lymphoma |
| ICD10CM | C83.00 | Small cell B-cell lymphoma, unspecified site |
| ICD10CM | C83.01 | Small cell B-cell lymphoma, lymph nodes of head, face, and neck |
| ICD10CM | C83.02 | Small cell B-cell lymphoma, intrathoracic lymph nodes |
| ICD10CM | C83.03 | Small cell B-cell lymphoma, intra-abdominal lymph nodes |
| ICD10CM | C83.04 | Small cell B-cell lymphoma, lymph nodes of axilla and upper limb |
| ICD10CM | C83.05 | Small cell B-cell lymphoma, lymph nodes of inguinal region and lower limb |
| ICD10CM | C83.06 | Small cell B-cell lymphoma, intrapelvic lymph nodes |
| ICD10CM | C83.07 | Small cell B-cell lymphoma, spleen |
| ICD10CM | C83.08 | Small cell B-cell lymphoma, lymph nodes of multiple sites |
| ICD10CM | C83.09 | Small cell B-cell lymphoma, extranodal and solid organ sites |
| ICD10CM | C83.1 | Mantle cell lymphoma |
| ICD10CM | C83.10 | Mantle cell lymphoma, unspecified site |
| ICD10CM | C83.11 | Mantle cell lymphoma, lymph nodes of head, face, and neck |
| ICD10CM | C83.12 | Mantle cell lymphoma, intrathoracic lymph nodes |
| ICD10CM | C83.13 | Mantle cell lymphoma, intra-abdominal lymph nodes |
| ICD10CM | C83.14 | Mantle cell lymphoma, lymph nodes of axilla and upper limb |

Continued on next page

**TableS31 – continued from previous page**

| CodeSystem | ConceptCode | ConceptName |
|---|---|---|
| ICD10CM | C83.15 | Mantle cell lymphoma, lymph nodes of inguinal region and lower limb |
| ICD10CM | C83.16 | Mantle cell lymphoma, intrapelvic lymph nodes |
| ICD10CM | C83.17 | Mantle cell lymphoma, spleen |
| ICD10CM | C83.18 | Mantle cell lymphoma, lymph nodes of multiple sites |
| ICD10CM | C83.19 | Mantle cell lymphoma, extranodal and solid organ sites |
| ICD10CM | C83.3 | Diffuse large B-cell lymphoma |
| ICD10CM | C83.30 | Diffuse large B-cell lymphoma, unspecified site |
| ICD10CM | C83.31 | Diffuse large B-cell lymphoma, lymph nodes of head, face, and neck |
| ICD10CM | C83.32 | Diffuse large B-cell lymphoma, intrathoracic lymph nodes |
| ICD10CM | C83.33 | Diffuse large B-cell lymphoma, intra-abdominal lymph nodes |
| ICD10CM | C83.34 | Diffuse large B-cell lymphoma, lymph nodes of axilla and upper limb |
| ICD10CM | C83.35 | Diffuse large B-cell lymphoma, lymph nodes of inguinal region and lower limb |
| ICD10CM | C83.36 | Diffuse large B-cell lymphoma, intrapelvic lymph nodes |
| ICD10CM | C83.37 | Diffuse large B-cell lymphoma, spleen |
| ICD10CM | C83.38 | Diffuse large B-cell lymphoma, lymph nodes of multiple sites |
| ICD10CM | C83.39 | Diffuse large B-cell lymphoma, extranodal and solid organ sites |
| ICD10CM | C83.5 | Lymphoblastic (diffuse) lymphoma |
| ICD10CM | C83.50 | Lymphoblastic (diffuse) lymphoma, unspecified site |
| ICD10CM | C83.51 | Lymphoblastic (diffuse) lymphoma, lymph nodes of head, face, and neck |
| ICD10CM | C83.52 | Lymphoblastic (diffuse) lymphoma, intrathoracic lymph nodes |
| ICD10CM | C83.53 | Lymphoblastic (diffuse) lymphoma, intra-abdominal lymph nodes |
| ICD10CM | C83.54 | Lymphoblastic (diffuse) lymphoma, lymph nodes of axilla and upper limb |
| ICD10CM | C83.55 | Lymphoblastic (diffuse) lymphoma, lymph nodes of inguinal region and lower limb |
| ICD10CM | C83.56 | Lymphoblastic (diffuse) lymphoma, intrapelvic lymph nodes |
| ICD10CM | C83.57 | Lymphoblastic (diffuse) lymphoma, spleen |
| ICD10CM | C83.58 | Lymphoblastic (diffuse) lymphoma, lymph nodes of multiple sites |
| ICD10CM | C83.59 | Lymphoblastic (diffuse) lymphoma, extranodal and solid organ sites |
| ICD10CM | C83.7 | Burkitt lymphoma |
| ICD10CM | C83.70 | Burkitt lymphoma, unspecified site |
| ICD10CM | C83.71 | Burkitt lymphoma, lymph nodes of head, face, and neck |
| ICD10CM | C83.72 | Burkitt lymphoma, intrathoracic lymph nodes |
| ICD10CM | C83.73 | Burkitt lymphoma, intra-abdominal lymph nodes |
| ICD10CM | C83.74 | Burkitt lymphoma, lymph nodes of axilla and upper limb |
| ICD10CM | C83.75 | Burkitt lymphoma, lymph nodes of inguinal region and lower limb |
| ICD10CM | C83.76 | Burkitt lymphoma, intrapelvic lymph nodes |
| ICD10CM | C83.77 | Burkitt lymphoma, spleen |

Continued on next page

**TableS31 – continued from previous page**

| CodeSystem | ConceptCode | ConceptName |
|---|---|---|
| ICD10CM | C83.78 | Burkitt lymphoma, lymph nodes of multiple sites |
| ICD10CM | C83.79 | Burkitt lymphoma, extranodal and solid organ sites |
| ICD10CM | C83.8 | Other non-follicular lymphoma |
| ICD10CM | C83.80 | Other non-follicular lymphoma, unspecified site |
| ICD10CM | C83.81 | Other non-follicular lymphoma, lymph nodes of head, face, and neck |
| ICD10CM | C83.82 | Other non-follicular lymphoma, intrathoracic lymph nodes |
| ICD10CM | C83.83 | Other non-follicular lymphoma, intra-abdominal lymph nodes |
| ICD10CM | C83.84 | Other non-follicular lymphoma, lymph nodes of axilla and upper limb |
| ICD10CM | C83.85 | Other non-follicular lymphoma, lymph nodes of inguinal region and lower limb |
| ICD10CM | C83.86 | Other non-follicular lymphoma, intrapelvic lymph nodes |
| ICD10CM | C83.87 | Other non-follicular lymphoma, spleen |
| ICD10CM | C83.88 | Other non-follicular lymphoma, lymph nodes of multiple sites |
| ICD10CM | C83.89 | Other non-follicular lymphoma, extranodal and solid organ sites |
| ICD10CM | C83.9 | Non-follicular (diffuse) lymphoma, unspecified |
| ICD10CM | C83.90 | Non-follicular (diffuse) lymphoma, unspecified, unspecified site |
| ICD10CM | C83.91 | Non-follicular (diffuse) lymphoma, unspecified, lymph nodes of head, face, and neck |
| ICD10CM | C83.92 | Non-follicular (diffuse) lymphoma, unspecified, intrathoracic lymph nodes |
| ICD10CM | C83.93 | Non-follicular (diffuse) lymphoma, unspecified, intra-abdominal lymph nodes |
| ICD10CM | C83.94 | Non-follicular (diffuse) lymphoma, unspecified, lymph nodes of axilla and upper limb |
| ICD10CM | C83.95 | Non-follicular (diffuse) lymphoma, unspecified, lymph nodes of inguinal region and lower limb |
| ICD10CM | C83.96 | Non-follicular (diffuse) lymphoma, unspecified, intrapelvic lymph nodes |
| ICD10CM | C83.97 | Non-follicular (diffuse) lymphoma, unspecified, spleen |
| ICD10CM | C83.98 | Non-follicular (diffuse) lymphoma, unspecified, lymph nodes of multiple sites |
| ICD10CM | C83.99 | Non-follicular (diffuse) lymphoma, unspecified, extranodal and solid organ sites |
| ICD10CM | C84.0 | Mycosis fungoides |
| ICD10CM | C84.00 | Mycosis fungoides, unspecified site |
| ICD10CM | C84.01 | Mycosis fungoides, lymph nodes of head, face, and neck |
| ICD10CM | C84.02 | Mycosis fungoides, intrathoracic lymph nodes |
| ICD10CM | C84.03 | Mycosis fungoides, intra-abdominal lymph nodes |
| ICD10CM | C84.04 | Mycosis fungoides, lymph nodes of axilla and upper limb |
| ICD10CM | C84.05 | Mycosis fungoides, lymph nodes of inguinal region and lower limb |
| ICD10CM | C84.06 | Mycosis fungoides, intrapelvic lymph nodes |
| ICD10CM | C84.07 | Mycosis fungoides, spleen |
| ICD10CM | C84.08 | Mycosis fungoides, lymph nodes of multiple sites |
| ICD10CM | C84.09 | Mycosis fungoides, extranodal and solid organ sites |
| ICD10CM | C84.1 | Sezary disease |

Continued on next page

**TableS31 – continued from previous page**

| CodeSystem | ConceptCode | ConceptName |
|---|---|---|
| ICD10CM | C84.10 | Sezary disease, unspecified site |
| ICD10CM | C84.11 | Sezary disease, lymph nodes of head, face, and neck |
| ICD10CM | C84.12 | Sezary disease, intrathoracic lymph nodes |
| ICD10CM | C84.13 | Sezary disease, intra-abdominal lymph nodes |
| ICD10CM | C84.14 | Sezary disease, lymph nodes of axilla and upper limb |
| ICD10CM | C84.15 | Sezary disease, lymph nodes of inguinal region and lower limb |
| ICD10CM | C84.16 | Sezary disease, intrapelvic lymph nodes |
| ICD10CM | C84.17 | Sezary disease, spleen |
| ICD10CM | C84.18 | Sezary disease, lymph nodes of multiple sites |
| ICD10CM | C84.19 | Sezary disease, extranodal and solid organ sites |
| ICD10CM | C84.4 | Peripheral T-cell lymphoma, not classified |
| ICD10CM | C84.40 | Peripheral T-cell lymphoma, not classified, unspecified site |
| ICD10CM | C84.41 | Peripheral T-cell lymphoma, not classified, lymph nodes of head, face, and neck |
| ICD10CM | C84.42 | Peripheral T-cell lymphoma, not classified, intrathoracic lymph nodes |
| ICD10CM | C84.43 | Peripheral T-cell lymphoma, not classified, intra-abdominal lymph nodes |
| ICD10CM | C84.44 | Peripheral T-cell lymphoma, not classified, lymph nodes of axilla and upper limb |
| ICD10CM | C84.45 | Peripheral T-cell lymphoma, not classified, lymph nodes of inguinal region and lower limb |
| ICD10CM | C84.46 | Peripheral T-cell lymphoma, not classified, intrapelvic lymph nodes |
| ICD10CM | C84.47 | Peripheral T-cell lymphoma, not classified, spleen |
| ICD10CM | C84.48 | Peripheral T-cell lymphoma, not classified, lymph nodes of multiple sites |
| ICD10CM | C84.49 | Peripheral T-cell lymphoma, not classified, extranodal and solid organ sites |
| ICD10CM | C84.6 | Anaplastic large cell lymphoma, ALK-positive |
| ICD10CM | C84.60 | Anaplastic large cell lymphoma, ALK-positive, unspecified site |
| ICD10CM | C84.61 | Anaplastic large cell lymphoma, ALK-positive, lymph nodes of head, face, and neck |
| ICD10CM | C84.62 | Anaplastic large cell lymphoma, ALK-positive, intrathoracic lymph nodes |
| ICD10CM | C84.63 | Anaplastic large cell lymphoma, ALK-positive, intra-abdominal lymph nodes |
| ICD10CM | C84.64 | Anaplastic large cell lymphoma, ALK-positive, lymph nodes of axilla and upper limb |
| ICD10CM | C84.65 | Anaplastic large cell lymphoma, ALK-positive, lymph nodes of inguinal region and lower limb |
| ICD10CM | C84.66 | Anaplastic large cell lymphoma, ALK-positive, intrapelvic lymph nodes |
| ICD10CM | C84.67 | Anaplastic large cell lymphoma, ALK-positive, spleen |
| ICD10CM | C84.68 | Anaplastic large cell lymphoma, ALK-positive, lymph nodes of multiple sites |

Continued on next page

**TableS31 – continued from previous page**

| CodeSystem | ConceptCode | ConceptName |
|---|---|---|
| ICD10CM | C84.69 | Anaplastic large cell lymphoma, ALK-positive, extranodal and solid organ sites |
| ICD10CM | C84.7 | Anaplastic large cell lymphoma, ALK-negative |
| ICD10CM | C84.70 | Anaplastic large cell lymphoma, ALK-negative, unspecified site |
| ICD10CM | C84.71 | Anaplastic large cell lymphoma, ALK-negative, lymph nodes of head, face, and neck |
| ICD10CM | C84.72 | Anaplastic large cell lymphoma, ALK-negative, intrathoracic lymph nodes |
| ICD10CM | C84.73 | Anaplastic large cell lymphoma, ALK-negative, intra-abdominal lymph nodes |
| ICD10CM | C84.74 | Anaplastic large cell lymphoma, ALK-negative, lymph nodes of axilla and upper limb |
| ICD10CM | C84.75 | Anaplastic large cell lymphoma, ALK-negative, lymph nodes of inguinal region and lower limb |
| ICD10CM | C84.76 | Anaplastic large cell lymphoma, ALK-negative, intrapelvic lymph nodes |
| ICD10CM | C84.77 | Anaplastic large cell lymphoma, ALK-negative, spleen |
| ICD10CM | C84.78 | Anaplastic large cell lymphoma, ALK-negative, lymph nodes of multiple sites |
| ICD10CM | C84.79 | Anaplastic large cell lymphoma, ALK-negative, extranodal and solid organ sites |
| ICD10CM | C84.9 | Mature T/NK-cell lymphomas, unspecified |
| ICD10CM | C84.90 | Mature T/NK-cell lymphomas, unspecified, unspecified site |
| ICD10CM | C84.91 | Mature T/NK-cell lymphomas, unspecified, lymph nodes of head, face, and neck |
| ICD10CM | C84.92 | Mature T/NK-cell lymphomas, unspecified, intrathoracic lymph nodes |
| ICD10CM | C84.93 | Mature T/NK-cell lymphomas, unspecified, intra-abdominal lymph nodes |
| ICD10CM | C84.94 | Mature T/NK-cell lymphomas, unspecified, lymph nodes of axilla and upper limb |
| ICD10CM | C84.95 | Mature T/NK-cell lymphomas, unspecified, lymph nodes of inguinal region and lower limb |
| ICD10CM | C84.96 | Mature T/NK-cell lymphomas, unspecified, intrapelvic lymph nodes |
| ICD10CM | C84.97 | Mature T/NK-cell lymphomas, unspecified, spleen |
| ICD10CM | C84.98 | Mature T/NK-cell lymphomas, unspecified, lymph nodes of multiple sites |
| ICD10CM | C84.99 | Mature T/NK-cell lymphomas, unspecified, extranodal and solid organ sites |
| ICD10CM | C84.A | Cutaneous T-cell lymphoma, unspecified |
| ICD10CM | C84.A0 | Cutaneous T-cell lymphoma, unspecified, unspecified site |
| ICD10CM | C84.A1 | Cutaneous T-cell lymphoma, unspecified lymph nodes of head, face, and neck |
| ICD10CM | C84.A2 | Cutaneous T-cell lymphoma, unspecified, intrathoracic lymph nodes |

Continued on next page

**TableS31 – continued from previous page**

| CodeSystem | ConceptCode | ConceptName |
|---|---|---|
| ICD10CM | C84.A3 | Cutaneous T-cell lymphoma, unspecified, intra-abdominal lymph nodes |
| ICD10CM | C84.A4 | Cutaneous T-cell lymphoma, unspecified, lymph nodes of axilla and upper limb |
| ICD10CM | C84.A5 | Cutaneous T-cell lymphoma, unspecified, lymph nodes of inguinal region and lower limb |
| ICD10CM | C84.A6 | Cutaneous T-cell lymphoma, unspecified, intrapelvic lymph nodes |
| ICD10CM | C84.A7 | Cutaneous T-cell lymphoma, unspecified, spleen |
| ICD10CM | C84.A8 | Cutaneous T-cell lymphoma, unspecified, lymph nodes of multiple sites |
| ICD10CM | C84.A9 | Cutaneous T-cell lymphoma, unspecified, extranodal and solid organ sites |
| ICD10CM | C84.Z | Other mature T/NK-cell lymphomas |
| ICD10CM | C84.Z0 | Other mature T/NK-cell lymphomas, unspecified site |
| ICD10CM | C84.Z1 | Other mature T/NK-cell lymphomas, lymph nodes of head, face, and neck |
| ICD10CM | C84.Z2 | Other mature T/NK-cell lymphomas, intrathoracic lymph nodes |
| ICD10CM | C84.Z3 | Other mature T/NK-cell lymphomas, intra-abdominal lymph nodes |
| ICD10CM | C84.Z4 | Other mature T/NK-cell lymphomas, lymph nodes of axilla and upper limb |
| ICD10CM | C84.Z5 | Other mature T/NK-cell lymphomas, lymph nodes of inguinal region and lower limb |
| ICD10CM | C84.Z6 | Other mature T/NK-cell lymphomas, intrapelvic lymph nodes |
| ICD10CM | C84.Z7 | Other mature T/NK-cell lymphomas, spleen |
| ICD10CM | C84.Z8 | Other mature T/NK-cell lymphomas, lymph nodes of multiple sites |
| ICD10CM | C84.Z9 | Other mature T/NK-cell lymphomas, extranodal and solid organ sites |
| ICD10CM | C85.1 | Unspecified B-cell lymphoma |
| ICD10CM | C85.10 | Unspecified B-cell lymphoma, unspecified site |
| ICD10CM | C85.11 | Unspecified B-cell lymphoma, lymph nodes of head, face, and neck |
| ICD10CM | C85.12 | Unspecified B-cell lymphoma, intrathoracic lymph nodes |
| ICD10CM | C85.13 | Unspecified B-cell lymphoma, intra-abdominal lymph nodes |
| ICD10CM | C85.14 | Unspecified B-cell lymphoma, lymph nodes of axilla and upper limb |
| ICD10CM | C85.15 | Unspecified B-cell lymphoma, lymph nodes of inguinal region and lower limb |
| ICD10CM | C85.16 | Unspecified B-cell lymphoma, intrapelvic lymph nodes |
| ICD10CM | C85.17 | Unspecified B-cell lymphoma, spleen |
| ICD10CM | C85.18 | Unspecified B-cell lymphoma, lymph nodes of multiple sites |
| ICD10CM | C85.19 | Unspecified B-cell lymphoma, extranodal and solid organ sites |
| ICD10CM | C85.2 | Mediastinal (thymic) large B-cell lymphoma |
| ICD10CM | C85.20 | Mediastinal (thymic) large B-cell lymphoma, unspecified site |

Continued on next page

**TableS31 – continued from previous page**

| CodeSystem | ConceptCode | ConceptName |
|---|---|---|
| ICD10CM | C85.21 | Mediastinal (thymic) large B-cell lymphoma, lymph nodes of head, face, and neck |
| ICD10CM | C85.22 | Mediastinal (thymic) large B-cell lymphoma, intrathoracic lymph nodes |
| ICD10CM | C85.23 | Mediastinal (thymic) large B-cell lymphoma, intra-abdominal lymph nodes |
| ICD10CM | C85.24 | Mediastinal (thymic) large B-cell lymphoma, lymph nodes of axilla and upper limb |
| ICD10CM | C85.25 | Mediastinal (thymic) large B-cell lymphoma, lymph nodes of inguinal region and lower limb |
| ICD10CM | C85.26 | Mediastinal (thymic) large B-cell lymphoma, intrapelvic lymph nodes |
| ICD10CM | C85.27 | Mediastinal (thymic) large B-cell lymphoma, spleen |
| ICD10CM | C85.28 | Mediastinal (thymic) large B-cell lymphoma, lymph nodes of multiple sites |
| ICD10CM | C85.29 | Mediastinal (thymic) large B-cell lymphoma, extranodal and solid organ sites |
| ICD10CM | C85.8 | Other specified types of non-Hodgkin lymphoma |
| ICD10CM | C85.80 | Other specified types of non-Hodgkin lymphoma, unspecified site |
| ICD10CM | C85.81 | Other specified types of non-Hodgkin lymphoma, lymph nodes of head, face, and neck |
| ICD10CM | C85.82 | Other specified types of non-Hodgkin lymphoma, intrathoracic lymph nodes |
| ICD10CM | C85.83 | Other specified types of non-Hodgkin lymphoma, intra-abdominal lymph nodes |
| ICD10CM | C85.84 | Other specified types of non-Hodgkin lymphoma, lymph nodes of axilla and upper limb |
| ICD10CM | C85.85 | Other specified types of non-Hodgkin lymphoma, lymph nodes of inguinal region and lower limb |
| ICD10CM | C85.86 | Other specified types of non-Hodgkin lymphoma, intrapelvic lymph nodes |
| ICD10CM | C85.87 | Other specified types of non-Hodgkin lymphoma, spleen |
| ICD10CM | C85.88 | Other specified types of non-Hodgkin lymphoma, lymph nodes of multiple sites |
| ICD10CM | C85.89 | Other specified types of non-Hodgkin lymphoma, extranodal and solid organ sites |
| ICD10CM | C85.9 | Non-Hodgkin lymphoma, unspecified |
| ICD10CM | C85.90 | Non-Hodgkin lymphoma, unspecified, unspecified site |
| ICD10CM | C85.91 | Non-Hodgkin lymphoma, unspecified, lymph nodes of head, face, and neck |
| ICD10CM | C85.92 | Non-Hodgkin lymphoma, unspecified, intrathoracic lymph nodes |
| ICD10CM | C85.93 | Non-Hodgkin lymphoma, unspecified, intra-abdominal lymph nodes |
| ICD10CM | C85.94 | Non-Hodgkin lymphoma, unspecified, lymph nodes of axilla and upper limb |
| ICD10CM | C85.95 | Non-Hodgkin lymphoma, unspecified, lymph nodes of inguinal region and lower limb |

Continued on next page

**TableS31 – continued from previous page**

| CodeSystem | ConceptCode | ConceptName |
|---|---|---|
| ICD10CM | C85.96 | Non-Hodgkin lymphoma, unspecified, intrapelvic lymph nodes |
| ICD10CM | C85.97 | Non-Hodgkin lymphoma, unspecified, spleen |
| ICD10CM | C85.98 | Non-Hodgkin lymphoma, unspecified, lymph nodes of multiple sites |
| ICD10CM | C85.99 | Non-Hodgkin lymphoma, unspecified, extranodal and solid organ sites |
| ICD10CM | C86.0 | Extranodal NK/T-cell lymphoma, nasal type |
| ICD10CM | C86.1 | Hepatosplenic T-cell lymphoma |
| ICD10CM | C86.2 | Enteropathy-type (intestinal) T-cell lymphoma |
| ICD10CM | C86.3 | Subcutaneous panniculitis-like T-cell lymphoma |
| ICD10CM | C86.4 | Blastic NK-cell lymphoma |
| ICD10CM | C86.5 | Angioimmunoblastic T-cell lymphoma |
| ICD10CM | C86.6 | Primary cutaneous CD30-positive T-cell proliferations |
| ICD10CM | C88.0 | Waldenstrom macroglobulinemia |
| ICD10CM | C88.2 | Heavy chain disease |
| ICD10CM | C88.3 | Immunoproliferative small intestinal disease |
| ICD10CM | C88.4 | Extranodal marginal zone B-cell lymphoma of mucosa-associated lymphoid tissue [MALT-lymphoma] |
| ICD10CM | C88.8 | Other malignant immunoproliferative diseases |
| ICD10CM | C88.9 | Malignant immunoproliferative disease, unspecified |
| ICD10CM | C90.0 | Multiple myeloma |
| ICD10CM | C90.00 | Multiple myeloma not having achieved remission |
| ICD10CM | C90.01 | Multiple myeloma in remission |
| ICD10CM | C90.02 | Multiple myeloma in relapse |
| ICD10CM | C90.1 | Plasma cell leukemia |
| ICD10CM | C90.10 | Plasma cell leukemia not having achieved remission |
| ICD10CM | C90.11 | Plasma cell leukemia in remission |
| ICD10CM | C90.12 | Plasma cell leukemia in relapse |
| ICD10CM | C90.2 | Extramedullary plasmacytoma |
| ICD10CM | C90.20 | Extramedullary plasmacytoma not having achieved remission |
| ICD10CM | C90.21 | Extramedullary plasmacytoma in remission |
| ICD10CM | C90.22 | Extramedullary plasmacytoma in relapse |
| ICD10CM | C90.3 | Solitary plasmacytoma |
| ICD10CM | C90.30 | Solitary plasmacytoma not having achieved remission |
| ICD10CM | C90.31 | Solitary plasmacytoma in remission |
| ICD10CM | C90.32 | Solitary plasmacytoma in relapse |
| ICD10CM | C91.0 | Acute lymphoblastic leukemia [ALL] |
| ICD10CM | C91.00 | Acute lymphoblastic leukemia not having achieved remission |
| ICD10CM | C91.01 | Acute lymphoblastic leukemia, in remission |
| ICD10CM | C91.02 | Acute lymphoblastic leukemia, in relapse |
| ICD10CM | C91.1 | Chronic lymphocytic leukemia of B-cell type |
| ICD10CM | C91.10 | Chronic lymphocytic leukemia of B-cell type not having achieved remission |
| ICD10CM | C91.11 | Chronic lymphocytic leukemia of B-cell type in remission |
| ICD10CM | C91.12 | Chronic lymphocytic leukemia of B-cell type in relapse |
| ICD10CM | C91.3 | Prolymphocytic leukemia of B-cell type |

Continued on next page

**TableS31 – continued from previous page**

| CodeSystem | ConceptCode | ConceptName |
|---|---|---|
| ICD10CM | C91.30 | Prolymphocytic leukemia of B-cell type not having achieved remission |
| ICD10CM | C91.31 | Prolymphocytic leukemia of B-cell type, in remission |
| ICD10CM | C91.32 | Prolymphocytic leukemia of B-cell type, in relapse |
| ICD10CM | C91.4 | Hairy cell leukemia |
| ICD10CM | C91.40 | Hairy cell leukemia not having achieved remission |
| ICD10CM | C91.41 | Hairy cell leukemia, in remission |
| ICD10CM | C91.42 | Hairy cell leukemia, in relapse |
| ICD10CM | C91.5 | Adult T-cell lymphoma/leukemia (HTLV-1-associated) |
| ICD10CM | C91.50 | Adult T-cell lymphoma/leukemia (HTLV-1-associated) not having achieved remission |
| ICD10CM | C91.51 | Adult T-cell lymphoma/leukemia (HTLV-1-associated), in remission |
| ICD10CM | C91.52 | Adult T-cell lymphoma/leukemia (HTLV-1-associated), in relapse |
| ICD10CM | C91.6 | Prolymphocytic leukemia of T-cell type |
| ICD10CM | C91.60 | Prolymphocytic leukemia of T-cell type not having achieved remission |
| ICD10CM | C91.61 | Prolymphocytic leukemia of T-cell type, in remission |
| ICD10CM | C91.62 | Prolymphocytic leukemia of T-cell type, in relapse |
| ICD10CM | C91.9 | Lymphoid leukemia, unspecified |
| ICD10CM | C91.90 | Lymphoid leukemia, unspecified not having achieved remission |
| ICD10CM | C91.91 | Lymphoid leukemia, unspecified, in remission |
| ICD10CM | C91.92 | Lymphoid leukemia, unspecified, in relapse |
| ICD10CM | C91.A | Mature B-cell leukemia Burkitt-type |
| ICD10CM | C91.A0 | Mature B-cell leukemia Burkitt-type not having achieved remission |
| ICD10CM | C91.A1 | Mature B-cell leukemia Burkitt-type, in remission |
| ICD10CM | C91.A2 | Mature B-cell leukemia Burkitt-type, in relapse |
| ICD10CM | C91.Z | Other lymphoid leukemia |
| ICD10CM | C91.Z0 | Other lymphoid leukemia not having achieved remission |
| ICD10CM | C91.Z1 | Other lymphoid leukemia, in remission |
| ICD10CM | C91.Z2 | Other lymphoid leukemia, in relapse |
| ICD10CM | C92.0 | Acute myeloblastic leukemia |
| ICD10CM | C92.00 | Acute myeloblastic leukemia, not having achieved remission |
| ICD10CM | C92.01 | Acute myeloblastic leukemia, in remission |
| ICD10CM | C92.02 | Acute myeloblastic leukemia, in relapse |
| ICD10CM | C92.1 | Chronic myeloid leukemia, BCR/ABL-positive |
| ICD10CM | C92.10 | Chronic myeloid leukemia, BCR/ABL-positive, not having achieved remission |
| ICD10CM | C92.11 | Chronic myeloid leukemia, BCR/ABL-positive, in remission |
| ICD10CM | C92.12 | Chronic myeloid leukemia, BCR/ABL-positive, in relapse |
| ICD10CM | C92.2 | Atypical chronic myeloid leukemia, BCR/ABL-negative |
| ICD10CM | C92.20 | Atypical chronic myeloid leukemia, BCR/ABL-negative, not having achieved remission |
| ICD10CM | C92.21 | Atypical chronic myeloid leukemia, BCR/ABL-negative, in remission |

Continued on next page

**TableS31 – continued from previous page**

| CodeSystem | ConceptCode | ConceptName |
|---|---|---|
| ICD10CM | C92.22 | Atypical chronic myeloid leukemia, BCR/ABL-negative, in relapse |
| ICD10CM | C92.3 | Myeloid sarcoma |
| ICD10CM | C92.30 | Myeloid sarcoma, not having achieved remission |
| ICD10CM | C92.31 | Myeloid sarcoma, in remission |
| ICD10CM | C92.32 | Myeloid sarcoma, in relapse |
| ICD10CM | C92.4 | Acute promyelocytic leukemia |
| ICD10CM | C92.40 | Acute promyelocytic leukemia, not having achieved remission |
| ICD10CM | C92.41 | Acute promyelocytic leukemia, in remission |
| ICD10CM | C92.42 | Acute promyelocytic leukemia, in relapse |
| ICD10CM | C92.5 | Acute myelomonocytic leukemia |
| ICD10CM | C92.50 | Acute myelomonocytic leukemia, not having achieved remission |
| ICD10CM | C92.51 | Acute myelomonocytic leukemia, in remission |
| ICD10CM | C92.52 | Acute myelomonocytic leukemia, in relapse |
| ICD10CM | C92.6 | Acute myeloid leukemia with 11q23-abnormality |
| ICD10CM | C92.60 | Acute myeloid leukemia with 11q23-abnormality not having achieved remission |
| ICD10CM | C92.61 | Acute myeloid leukemia with 11q23-abnormality in remission |
| ICD10CM | C92.62 | Acute myeloid leukemia with 11q23-abnormality in relapse |
| ICD10CM | C92.9 | Myeloid leukemia, unspecified |
| ICD10CM | C92.90 | Myeloid leukemia, unspecified, not having achieved remission |
| ICD10CM | C92.91 | Myeloid leukemia, unspecified in remission |
| ICD10CM | C92.92 | Myeloid leukemia, unspecified in relapse |
| ICD10CM | C92.A | Acute myeloid leukemia with multilineage dysplasia |
| ICD10CM | C92.A0 | Acute myeloid leukemia with multilineage dysplasia, not having achieved remission |
| ICD10CM | C92.A1 | Acute myeloid leukemia with multilineage dysplasia, in remission |
| ICD10CM | C92.A2 | Acute myeloid leukemia with multilineage dysplasia, in relapse |
| ICD10CM | C92.Z | Other myeloid leukemia |
| ICD10CM | C92.Z0 | Other myeloid leukemia not having achieved remission |
| ICD10CM | C92.Z1 | Other myeloid leukemia, in remission |
| ICD10CM | C92.Z2 | Other myeloid leukemia, in relapse |
| ICD10CM | C93.0 | Acute monoblastic/monocytic leukemia |
| ICD10CM | C93.00 | Acute monoblastic/monocytic leukemia, not having achieved remission |
| ICD10CM | C93.01 | Acute monoblastic/monocytic leukemia, in remission |
| ICD10CM | C93.02 | Acute monoblastic/monocytic leukemia, in relapse |
| ICD10CM | C93.1 | Chronic myelomonocytic leukemia |
| ICD10CM | C93.10 | Chronic myelomonocytic leukemia not having achieved remission |
| ICD10CM | C93.11 | Chronic myelomonocytic leukemia, in remission |
| ICD10CM | C93.12 | Chronic myelomonocytic leukemia, in relapse |
| ICD10CM | C93.3 | Juvenile myelomonocytic leukemia |
| ICD10CM | C93.30 | Juvenile myelomonocytic leukemia, not having achieved remission |
| ICD10CM | C93.31 | Juvenile myelomonocytic leukemia, in remission |
| ICD10CM | C93.32 | Juvenile myelomonocytic leukemia, in relapse |
| ICD10CM | C93.9 | Monocytic leukemia, unspecified |

Continued on next page

**TableS31 – continued from previous page**

| CodeSystem | ConceptCode | ConceptName |
|---|---|---|
| ICD10CM | C93.90 | Monocytic leukemia, unspecified, not having achieved remission |
| ICD10CM | C93.91 | Monocytic leukemia, unspecified in remission |
| ICD10CM | C93.92 | Monocytic leukemia, unspecified in relapse |
| ICD10CM | C93.Z | Other monocytic leukemia |
| ICD10CM | C93.Z0 | Other monocytic leukemia, not having achieved remission |
| ICD10CM | C93.Z1 | Other monocytic leukemia, in remission |
| ICD10CM | C93.Z2 | Other monocytic leukemia, in relapse |
| ICD10CM | C94.0 | Acute erythroid leukemia |
| ICD10CM | C94.00 | Acute erythroid leukemia, not having achieved remission |
| ICD10CM | C94.01 | Acute erythroid leukemia, in remission |
| ICD10CM | C94.02 | Acute erythroid leukemia, in relapse |
| ICD10CM | C94.2 | Acute megakaryoblastic leukemia |
| ICD10CM | C94.20 | Acute megakaryoblastic leukemia not having achieved remission |
| ICD10CM | C94.21 | Acute megakaryoblastic leukemia, in remission |
| ICD10CM | C94.22 | Acute megakaryoblastic leukemia, in relapse |
| ICD10CM | C94.3 | Mast cell leukemia |
| ICD10CM | C94.30 | Mast cell leukemia not having achieved remission |
| ICD10CM | C94.31 | Mast cell leukemia, in remission |
| ICD10CM | C94.32 | Mast cell leukemia, in relapse |
| ICD10CM | C94.4 | Acute panmyelosis with myelofibrosis |
| ICD10CM | C94.40 | Acute panmyelosis with myelofibrosis not having achieved remission |
| ICD10CM | C94.41 | Acute panmyelosis with myelofibrosis, in remission |
| ICD10CM | C94.42 | Acute panmyelosis with myelofibrosis, in relapse |
| ICD10CM | C94.6 | Myelodysplastic disease, not classified |
| ICD10CM | C94.8 | Other specified leukemias |
| ICD10CM | C94.80 | Other specified leukemias not having achieved remission |
| ICD10CM | C94.81 | Other specified leukemias, in remission |
| ICD10CM | C94.82 | Other specified leukemias, in relapse |
| ICD10CM | C95.0 | Acute leukemia of unspecified cell type |
| ICD10CM | C95.00 | Acute leukemia of unspecified cell type not having achieved remission |
| ICD10CM | C95.01 | Acute leukemia of unspecified cell type, in remission |
| ICD10CM | C95.02 | Acute leukemia of unspecified cell type, in relapse |
| ICD10CM | C95.1 | Chronic leukemia of unspecified cell type |
| ICD10CM | C95.10 | Chronic leukemia of unspecified cell type not having achieved remission |
| ICD10CM | C95.11 | Chronic leukemia of unspecified cell type, in remission |
| ICD10CM | C95.12 | Chronic leukemia of unspecified cell type, in relapse |
| ICD10CM | C95.9 | Leukemia, unspecified |
| ICD10CM | C95.90 | Leukemia, unspecified not having achieved remission |
| ICD10CM | C95.91 | Leukemia, unspecified, in remission |
| ICD10CM | C95.92 | Leukemia, unspecified, in relapse |
| ICD10CM | C96.0 | Multifocal and multisystemic (disseminated) Langerhans-cell histiocytosis |
| ICD10CM | C96.2 | Malignant mast cell neoplasm |
| ICD10CM | C96.20 | Malignant mast cell neoplasm, unspecified |

Continued on next page

**TableS31 – continued from previous page**

| CodeSystem | ConceptCode | ConceptName |
|---|---|---|
| ICD10CM | C96.21 | Aggressive systemic mastocytosis |
| ICD10CM | C96.22 | Mast cell sarcoma |
| ICD10CM | C96.29 | Other malignant mast cell neoplasm |
| ICD10CM | C96.4 | Sarcoma of dendritic cells (accessory cells) |
| ICD10CM | C96.5 | Multifocal and unisystemic Langerhans-cell histiocytosis |
| ICD10CM | C96.6 | Unifocal Langerhans-cell histiocytosis |
| ICD10CM | C96.9 | Malignant neoplasm of lymphoid, hematopoietic and related tissue, unspecified |
| ICD10CM | C96.A | Histiocytic sarcoma |
| ICD10CM | C96.Z | Other specified malignant neoplasms of lymphoid, hematopoietic and related tissue |
| ICD10CM | D3A.0 | Benign carcinoid tumors |
| ICD10CM | D3A.00 | Benign carcinoid tumor of unspecified site |
| ICD10CM | D3A.01 | Benign carcinoid tumors of the small intestine |
| ICD10CM | D3A.010 | Benign carcinoid tumor of the duodenum |
| ICD10CM | D3A.011 | Benign carcinoid tumor of the jejunum |
| ICD10CM | D3A.012 | Benign carcinoid tumor of the ileum |
| ICD10CM | D3A.019 | Benign carcinoid tumor of the small intestine, unspecified portion |
| ICD10CM | D3A.02 | Benign carcinoid tumors of the appendix, large intestine, and rectum |
| ICD10CM | D3A.020 | Benign carcinoid tumor of the appendix |
| ICD10CM | D3A.021 | Benign carcinoid tumor of the cecum |
| ICD10CM | D3A.022 | Benign carcinoid tumor of the ascending colon |
| ICD10CM | D3A.023 | Benign carcinoid tumor of the transverse colon |
| ICD10CM | D3A.024 | Benign carcinoid tumor of the descending colon |
| ICD10CM | D3A.025 | Benign carcinoid tumor of the sigmoid colon |
| ICD10CM | D3A.026 | Benign carcinoid tumor of the rectum |
| ICD10CM | D3A.029 | Benign carcinoid tumor of the large intestine, unspecified portion |
| ICD10CM | D3A.09 | Benign carcinoid tumors of other sites |
| ICD10CM | D3A.090 | Benign carcinoid tumor of the bronchus and lung |
| ICD10CM | D3A.091 | Benign carcinoid tumor of the thymus |
| ICD10CM | D3A.092 | Benign carcinoid tumor of the stomach |
| ICD10CM | D3A.093 | Benign carcinoid tumor of the kidney |
| ICD10CM | D3A.094 | Benign carcinoid tumor of the foregut, unspecified |
| ICD10CM | D3A.095 | Benign carcinoid tumor of the midgut, unspecified |
| ICD10CM | D3A.096 | Benign carcinoid tumor of the hindgut, unspecified |
| ICD10CM | D3A.098 | Benign carcinoid tumors of other sites |
| ICD10CM | D3A.8 | Other benign neuroendocrine tumors |
| ICD10CM | D46.0 | Refractory anemia without ring sideroblasts, so stated |
| ICD10CM | D46.1 | Refractory anemia with ring sideroblasts |
| ICD10CM | D46.2 | Refractory anemia with excess of blasts [RAEB] |
| ICD10CM | D46.20 | Refractory anemia with excess of blasts, unspecified |
| ICD10CM | D46.21 | Refractory anemia with excess of blasts 1 |
| ICD10CM | D46.22 | Refractory anemia with excess of blasts 2 |
| ICD10CM | D46.4 | Refractory anemia, unspecified |
| ICD10CM | D46.9 | Myelodysplastic syndrome, unspecified |

Continued on next page

**TableS31 – continued from previous page**

| CodeSystem | ConceptCode | ConceptName |
|---|---|---|
| ICD10CM | D46.A | Refractory cytopenia with multilineage dysplasia |
| ICD10CM | D46.B | Refractory cytopenia with multilineage dysplasia and ring sideroblasts |
| ICD10CM | D46.C | Myelodysplastic syndrome with isolated del(5q) chromosomal abnormality |
| ICD10CM | D46.Z | Other myelodysplastic syndromes |
| ICD10CM | D61.01 | Constitutional (pure) red blood cell aplasia |
| ICD10CM | D80.0 | Hereditary hypogammaglobulinemia |
| ICD10CM | D80.1 | Nonfamilial hypogammaglobulinemia |
| ICD10CM | D80.2 | Selective deficiency of immunoglobulin A [IgA] |
| ICD10CM | D80.3 | Selective deficiency of immunoglobulin G [IgG] subclasses |
| ICD10CM | D80.4 | Selective deficiency of immunoglobulin M [IgM] |
| ICD10CM | D80.5 | Immunodeficiency with increased immunoglobulin M [IgM] |
| ICD10CM | D80.6 | Antibody deficiency with near-normal immunoglobulins or with hyperimmunoglobulinemia |
| ICD10CM | D80.7 | Transient hypogammaglobulinemia of infancy |
| ICD10CM | D80.8 | Other immunodeficiencies with predominantly antibody defects |
| ICD10CM | D80.9 | Immunodeficiency with predominantly antibody defects, unspecified |
| ICD10CM | D81.0 | Severe combined immunodeficiency [SCID] with reticular dysgenesis |
| ICD10CM | D81.1 | Severe combined immunodeficiency [SCID] with low T- and B-cell numbers |
| ICD10CM | D81.2 | Severe combined immunodeficiency [SCID] with low or normal B-cell numbers |
| ICD10CM | D81.3 | Adenosine deaminase [ADA] deficiency |
| ICD10CM | D81.30 | Adenosine deaminase deficiency, unspecified |
| ICD10CM | D81.31 | Severe combined immunodeficiency due to adenosine deaminase deficiency |
| ICD10CM | D81.32 | Adenosine deaminase 2 deficiency |
| ICD10CM | D81.39 | Other adenosine deaminase deficiency |
| ICD10CM | D81.4 | Nezelof's syndrome |
| ICD10CM | D81.5 | Purine nucleoside phosphorylase [PNP] deficiency |
| ICD10CM | D81.6 | Major histocompatibility complex class I deficiency |
| ICD10CM | D81.7 | Major histocompatibility complex class II deficiency |
| ICD10CM | D81.8 | Other combined immunodeficiencies |
| ICD10CM | D81.81 | Biotin-dependent carboxylase deficiency |
| ICD10CM | D81.810 | Biotinidase deficiency |
| ICD10CM | D81.818 | Other biotin-dependent carboxylase deficiency |
| ICD10CM | D81.819 | Biotin-dependent carboxylase deficiency, unspecified |
| ICD10CM | D81.89 | Other combined immunodeficiencies |
| ICD10CM | D81.9 | Combined immunodeficiency, unspecified |
| ICD10CM | D82.0 | Wiskott-Aldrich syndrome |
| ICD10CM | D82.1 | Di George's syndrome |
| ICD10CM | D82.2 | Immunodeficiency with short-limbed stature |
| ICD10CM | D82.3 | Immunodeficiency following hereditary defective response to Epstein-Barr virus |

Continued on next page

**TableS31 – continued from previous page**

| CodeSystem | ConceptCode | ConceptName |
|---|---|---|
| ICD10CM | D82.4 | Hyperimmunoglobulin E [IgE] syndrome |
| ICD10CM | D82.8 | Immunodeficiency associated with other specified major defects |
| ICD10CM | D82.9 | Immunodeficiency associated with major defect, unspecified |
| ICD10CM | D89.4 | Mast cell activation syndrome and related disorders |
| ICD10CM | D83.0 | Common variable immunodeficiency with predominant abnormalities of B-cell numbers and function |
| ICD10CM | D83.1 | Common variable immunodeficiency with predominant immunoregulatory T-cell disorders |
| ICD10CM | D83.2 | Common variable immunodeficiency with autoantibodies to B- or T-cells |
| ICD10CM | D83.8 | Other common variable immunodeficiencies |
| ICD10CM | D83.9 | Common variable immunodeficiency, unspecified |
| ICD10CM | D84.0 | Lymphocyte function antigen-1 [LFA-1] defect |
| ICD10CM | D84.1 | Defects in the complement system |
| ICD10CM | D84.8 | Other specified immunodeficiencies |
| ICD10CM | D84.9 | Immunodeficiency, unspecified |
| ICD10CM | D89.0 | Polyclonal hypergammaglobulinemia |
| ICD10CM | D89.1 | Cryoglobulinemia |
| ICD10CM | D89.2 | Hypergammaglobulinemia, unspecified |
| ICD10CM | D89.3 | Immune reconstitution syndrome |
| ICD10CM | D89.40 | Mast cell activation, unspecified |
| ICD10CM | D89.41 | Monoclonal mast cell activation syndrome |
| ICD10CM | D89.42 | Idiopathic mast cell activation syndrome |
| ICD10CM | D89.43 | Secondary mast cell activation |
| ICD10CM | D89.49 | Other mast cell activation disorder |
| ICD10CM | D89.8 | Other specified disorders involving the immune mechanism, not elsewhere classified |
| ICD10CM | D89.81 | Graft-versus-host disease |
| ICD10CM | D89.810 | Acute graft-versus-host disease |
| ICD10CM | D89.811 | Chronic graft-versus-host disease |
| ICD10CM | D89.812 | Acute on chronic graft-versus-host disease |
| ICD10CM | D89.813 | Graft-versus-host disease, unspecified |
| ICD10CM | D89.82 | Autoimmune lymphoproliferative syndrome [ALPS] |
| ICD10CM | D89.89 | Other specified disorders involving the immune mechanism, not elsewhere classified |
| ICD10CM | D89.9 | Disorder involving the immune mechanism, unspecified |
| ICD10CM | K70.30 | Alcoholic cirrhosis of liver without ascites |
| ICD10CM | K70.31 | Alcoholic cirrhosis of liver with ascites |
| ICD10CM | K70.40 | Alcoholic hepatic failure without coma |
| ICD10CM | K70.41 | Alcoholic hepatic failure with coma |
| ICD10CM | K72.0 | Acute and subacute hepatic failure |
| ICD10CM | K72.00 | Acute and subacute hepatic failure without coma |
| ICD10CM | K72.01 | Acute and subacute hepatic failure with coma |
| ICD10CM | K72.1 | Chronic hepatic failure |
| ICD10CM | K72.10 | Chronic hepatic failure without coma |
| ICD10CM | K72.11 | Chronic hepatic failure with coma |
| ICD10CM | K72.9 | Hepatic failure, unspecified |

Continued on next page

**TableS31 – continued from previous page**

| CodeSystem | ConceptCode | ConceptName |
|---|---|---|
| ICD10CM | K72.90 | Hepatic failure, unspecified without coma |
| ICD10CM | K72.91 | Hepatic failure, unspecified with coma |
| ICD10CM | K74.60 | Unspecified cirrhosis of liver |
| ICD10CM | K74.69 | Other cirrhosis of liver |
| ICD10CM | L94.0 | Localized scleroderma [morphea] |
| ICD10CM | L94.1 | Linear scleroderma |
| ICD10CM | L94.2 | Calcinosis cutis |
| ICD10CM | L94.3 | Sclerodactyly |
| ICD10CM | L94.4 | Gottron's papules |
| ICD10CM | L94.5 | Poikiloderma vasculare atrophicans |
| ICD10CM | L94.6 | Ainhum |
| ICD10CM | L94.8 | Other specified localized connective tissue disorders |
| ICD10CM | L94.9 | Localized connective tissue disorder, unspecified |
| ICD10CM | M05.0 | Felty's syndrome |
| ICD10CM | M05.00 | Felty's syndrome, unspecified site |
| ICD10CM | M05.01 | Felty's syndrome, shoulder |
| ICD10CM | M05.011 | Felty's syndrome, right shoulder |
| ICD10CM | M05.012 | Felty's syndrome, left shoulder |
| ICD10CM | M05.019 | Felty's syndrome, unspecified shoulder |
| ICD10CM | M05.02 | Felty's syndrome, elbow |
| ICD10CM | M05.021 | Felty's syndrome, right elbow |
| ICD10CM | M05.022 | Felty's syndrome, left elbow |
| ICD10CM | M05.029 | Felty's syndrome, unspecified elbow |
| ICD10CM | M05.03 | Felty's syndrome, wrist |
| ICD10CM | M05.031 | Felty's syndrome, right wrist |
| ICD10CM | M05.032 | Felty's syndrome, left wrist |
| ICD10CM | M05.039 | Felty's syndrome, unspecified wrist |
| ICD10CM | M05.04 | Felty's syndrome, hand |
| ICD10CM | M05.041 | Felty's syndrome, right hand |
| ICD10CM | M05.042 | Felty's syndrome, left hand |
| ICD10CM | M05.049 | Felty's syndrome, unspecified hand |
| ICD10CM | M05.05 | Felty's syndrome, hip |
| ICD10CM | M05.051 | Felty's syndrome, right hip |
| ICD10CM | M05.052 | Felty's syndrome, left hip |
| ICD10CM | M05.059 | Felty's syndrome, unspecified hip |
| ICD10CM | M05.06 | Felty's syndrome, knee |
| ICD10CM | M05.061 | Felty's syndrome, right knee |
| ICD10CM | M05.062 | Felty's syndrome, left knee |
| ICD10CM | M05.069 | Felty's syndrome, unspecified knee |
| ICD10CM | M05.07 | Felty's syndrome, ankle and foot |
| ICD10CM | M05.071 | Felty's syndrome, right ankle and foot |
| ICD10CM | M05.072 | Felty's syndrome, left ankle and foot |
| ICD10CM | M05.079 | Felty's syndrome, unspecified ankle and foot |
| ICD10CM | M05.09 | Felty's syndrome, multiple sites |
| ICD10CM | M05.1 | Rheumatoid lung disease with rheumatoid arthritis |
| ICD10CM | M05.10 | Rheumatoid lung disease with rheumatoid arthritis of unspecified site |

Continued on next page

**TableS31 – continued from previous page**

| CodeSystem | ConceptCode | ConceptName |
|---|---|---|
| ICD10CM | M05.11 | Rheumatoid lung disease with rheumatoid arthritis of shoulder |
| ICD10CM | M05.111 | Rheumatoid lung disease with rheumatoid arthritis of right shoulder |
| ICD10CM | M05.112 | Rheumatoid lung disease with rheumatoid arthritis of left shoulder |
| ICD10CM | M05.119 | Rheumatoid lung disease with rheumatoid arthritis of unspecified shoulder |
| ICD10CM | M05.12 | Rheumatoid lung disease with rheumatoid arthritis of elbow |
| ICD10CM | M05.121 | Rheumatoid lung disease with rheumatoid arthritis of right elbow |
| ICD10CM | M05.122 | Rheumatoid lung disease with rheumatoid arthritis of left elbow |
| ICD10CM | M05.129 | Rheumatoid lung disease with rheumatoid arthritis of unspecified elbow |
| ICD10CM | M05.13 | Rheumatoid lung disease with rheumatoid arthritis of wrist |
| ICD10CM | M05.131 | Rheumatoid lung disease with rheumatoid arthritis of right wrist |
| ICD10CM | M05.132 | Rheumatoid lung disease with rheumatoid arthritis of left wrist |
| ICD10CM | M05.139 | Rheumatoid lung disease with rheumatoid arthritis of unspecified wrist |
| ICD10CM | M05.14 | Rheumatoid lung disease with rheumatoid arthritis of hand |
| ICD10CM | M05.141 | Rheumatoid lung disease with rheumatoid arthritis of right hand |
| ICD10CM | M05.142 | Rheumatoid lung disease with rheumatoid arthritis of left hand |
| ICD10CM | M05.149 | Rheumatoid lung disease with rheumatoid arthritis of unspecified hand |
| ICD10CM | M05.15 | Rheumatoid lung disease with rheumatoid arthritis of hip |
| ICD10CM | M05.151 | Rheumatoid lung disease with rheumatoid arthritis of right hip |
| ICD10CM | M05.152 | Rheumatoid lung disease with rheumatoid arthritis of left hip |
| ICD10CM | M05.159 | Rheumatoid lung disease with rheumatoid arthritis of unspecified hip |
| ICD10CM | M05.16 | Rheumatoid lung disease with rheumatoid arthritis of knee |
| ICD10CM | M05.161 | Rheumatoid lung disease with rheumatoid arthritis of right knee |
| ICD10CM | M05.162 | Rheumatoid lung disease with rheumatoid arthritis of left knee |
| ICD10CM | M05.169 | Rheumatoid lung disease with rheumatoid arthritis of unspecified knee |
| ICD10CM | M05.17 | Rheumatoid lung disease with rheumatoid arthritis of ankle and foot |
| ICD10CM | M05.171 | Rheumatoid lung disease with rheumatoid arthritis of right ankle and foot |
| ICD10CM | M05.172 | Rheumatoid lung disease with rheumatoid arthritis of left ankle and foot |
| ICD10CM | M05.179 | Rheumatoid lung disease with rheumatoid arthritis of unspecified ankle and foot |
| ICD10CM | M05.19 | Rheumatoid lung disease with rheumatoid arthritis of multiple sites |
| ICD10CM | M05.2 | Rheumatoid vasculitis with rheumatoid arthritis |
| ICD10CM | M05.20 | Rheumatoid vasculitis with rheumatoid arthritis of unspecified site |
| ICD10CM | M05.21 | Rheumatoid vasculitis with rheumatoid arthritis of shoulder |

Continued on next page

**TableS31 – continued from previous page**

| CodeSystem | ConceptCode | ConceptName |
|---|---|---|
| ICD10CM | M05.211 | Rheumatoid vasculitis with rheumatoid arthritis of right shoulder |
| ICD10CM | M05.212 | Rheumatoid vasculitis with rheumatoid arthritis of left shoulder |
| ICD10CM | M05.219 | Rheumatoid vasculitis with rheumatoid arthritis of unspecified shoulder |
| ICD10CM | M05.22 | Rheumatoid vasculitis with rheumatoid arthritis of elbow |
| ICD10CM | M05.221 | Rheumatoid vasculitis with rheumatoid arthritis of right elbow |
| ICD10CM | M05.222 | Rheumatoid vasculitis with rheumatoid arthritis of left elbow |
| ICD10CM | M05.229 | Rheumatoid vasculitis with rheumatoid arthritis of unspecified elbow |
| ICD10CM | M05.23 | Rheumatoid vasculitis with rheumatoid arthritis of wrist |
| ICD10CM | M05.231 | Rheumatoid vasculitis with rheumatoid arthritis of right wrist |
| ICD10CM | M05.232 | Rheumatoid vasculitis with rheumatoid arthritis of left wrist |
| ICD10CM | M05.239 | Rheumatoid vasculitis with rheumatoid arthritis of unspecified wrist |
| ICD10CM | M05.24 | Rheumatoid vasculitis with rheumatoid arthritis of hand |
| ICD10CM | M05.241 | Rheumatoid vasculitis with rheumatoid arthritis of right hand |
| ICD10CM | M05.242 | Rheumatoid vasculitis with rheumatoid arthritis of left hand |
| ICD10CM | M05.249 | Rheumatoid vasculitis with rheumatoid arthritis of unspecified hand |
| ICD10CM | M05.25 | Rheumatoid vasculitis with rheumatoid arthritis of hip |
| ICD10CM | M05.251 | Rheumatoid vasculitis with rheumatoid arthritis of right hip |
| ICD10CM | M05.252 | Rheumatoid vasculitis with rheumatoid arthritis of left hip |
| ICD10CM | M05.259 | Rheumatoid vasculitis with rheumatoid arthritis of unspecified hip |
| ICD10CM | M05.26 | Rheumatoid vasculitis with rheumatoid arthritis of knee |
| ICD10CM | M05.261 | Rheumatoid vasculitis with rheumatoid arthritis of right knee |
| ICD10CM | M05.262 | Rheumatoid vasculitis with rheumatoid arthritis of left knee |
| ICD10CM | M05.269 | Rheumatoid vasculitis with rheumatoid arthritis of unspecified knee |
| ICD10CM | M05.27 | Rheumatoid vasculitis with rheumatoid arthritis of ankle and foot |
| ICD10CM | M05.271 | Rheumatoid vasculitis with rheumatoid arthritis of right ankle and foot |
| ICD10CM | M05.272 | Rheumatoid vasculitis with rheumatoid arthritis of left ankle and foot |
| ICD10CM | M05.279 | Rheumatoid vasculitis with rheumatoid arthritis of unspecified ankle and foot |
| ICD10CM | M05.29 | Rheumatoid vasculitis with rheumatoid arthritis of multiple sites |
| ICD10CM | M05.3 | Rheumatoid heart disease with rheumatoid arthritis |
| ICD10CM | M05.30 | Rheumatoid heart disease with rheumatoid arthritis of unspecified site |
| ICD10CM | M05.31 | Rheumatoid heart disease with rheumatoid arthritis of shoulder |
| ICD10CM | M05.311 | Rheumatoid heart disease with rheumatoid arthritis of right shoulder |
| ICD10CM | M05.312 | Rheumatoid heart disease with rheumatoid arthritis of left shoulder |

Continued on next page

**TableS31 – continued from previous page**

| CodeSystem | ConceptCode | ConceptName |
|---|---|---|
| ICD10CM | M05.319 | Rheumatoid heart disease with rheumatoid arthritis of unspecified shoulder |
| ICD10CM | M05.32 | Rheumatoid heart disease with rheumatoid arthritis of elbow |
| ICD10CM | M05.321 | Rheumatoid heart disease with rheumatoid arthritis of right elbow |
| ICD10CM | M05.322 | Rheumatoid heart disease with rheumatoid arthritis of left elbow |
| ICD10CM | M05.329 | Rheumatoid heart disease with rheumatoid arthritis of unspecified elbow |
| ICD10CM | M05.33 | Rheumatoid heart disease with rheumatoid arthritis of wrist |
| ICD10CM | M05.331 | Rheumatoid heart disease with rheumatoid arthritis of right wrist |
| ICD10CM | M05.332 | Rheumatoid heart disease with rheumatoid arthritis of left wrist |
| ICD10CM | M05.339 | Rheumatoid heart disease with rheumatoid arthritis of unspecified wrist |
| ICD10CM | M05.34 | Rheumatoid heart disease with rheumatoid arthritis of hand |
| ICD10CM | M05.341 | Rheumatoid heart disease with rheumatoid arthritis of right hand |
| ICD10CM | M05.342 | Rheumatoid heart disease with rheumatoid arthritis of left hand |
| ICD10CM | M05.349 | Rheumatoid heart disease with rheumatoid arthritis of unspecified hand |
| ICD10CM | M05.35 | Rheumatoid heart disease with rheumatoid arthritis of hip |
| ICD10CM | M05.351 | Rheumatoid heart disease with rheumatoid arthritis of right hip |
| ICD10CM | M05.352 | Rheumatoid heart disease with rheumatoid arthritis of left hip |
| ICD10CM | M05.359 | Rheumatoid heart disease with rheumatoid arthritis of unspecified hip |
| ICD10CM | M05.36 | Rheumatoid heart disease with rheumatoid arthritis of knee |
| ICD10CM | M05.361 | Rheumatoid heart disease with rheumatoid arthritis of right knee |
| ICD10CM | M05.362 | Rheumatoid heart disease with rheumatoid arthritis of left knee |
| ICD10CM | M05.369 | Rheumatoid heart disease with rheumatoid arthritis of unspecified knee |
| ICD10CM | M05.37 | Rheumatoid heart disease with rheumatoid arthritis of ankle and foot |
| ICD10CM | M05.371 | Rheumatoid heart disease with rheumatoid arthritis of right ankle and foot |
| ICD10CM | M05.372 | Rheumatoid heart disease with rheumatoid arthritis of left ankle and foot |
| ICD10CM | M05.379 | Rheumatoid heart disease with rheumatoid arthritis of unspecified ankle and foot |
| ICD10CM | M05.39 | Rheumatoid heart disease with rheumatoid arthritis of multiple sites |
| ICD10CM | M05.4 | Rheumatoid myopathy with rheumatoid arthritis |
| ICD10CM | M05.40 | Rheumatoid myopathy with rheumatoid arthritis of unspecified site |
| ICD10CM | M05.41 | Rheumatoid myopathy with rheumatoid arthritis of shoulder |
| ICD10CM | M05.411 | Rheumatoid myopathy with rheumatoid arthritis of right shoulder |
| ICD10CM | M05.412 | Rheumatoid myopathy with rheumatoid arthritis of left shoulder |

Continued on next page

**TableS31 – continued from previous page**

| CodeSystem | ConceptCode | ConceptName |
|---|---|---|
| ICD10CM | M05.419 | Rheumatoid myopathy with rheumatoid arthritis of unspecified shoulder |
| ICD10CM | M05.42 | Rheumatoid myopathy with rheumatoid arthritis of elbow |
| ICD10CM | M05.421 | Rheumatoid myopathy with rheumatoid arthritis of right elbow |
| ICD10CM | M05.422 | Rheumatoid myopathy with rheumatoid arthritis of left elbow |
| ICD10CM | M05.429 | Rheumatoid myopathy with rheumatoid arthritis of unspecified elbow |
| ICD10CM | M05.43 | Rheumatoid myopathy with rheumatoid arthritis of wrist |
| ICD10CM | M05.431 | Rheumatoid myopathy with rheumatoid arthritis of right wrist |
| ICD10CM | M05.432 | Rheumatoid myopathy with rheumatoid arthritis of left wrist |
| ICD10CM | M05.439 | Rheumatoid myopathy with rheumatoid arthritis of unspecified wrist |
| ICD10CM | M05.44 | Rheumatoid myopathy with rheumatoid arthritis of hand |
| ICD10CM | M05.441 | Rheumatoid myopathy with rheumatoid arthritis of right hand |
| ICD10CM | M05.442 | Rheumatoid myopathy with rheumatoid arthritis of left hand |
| ICD10CM | M05.449 | Rheumatoid myopathy with rheumatoid arthritis of unspecified hand |
| ICD10CM | M05.45 | Rheumatoid myopathy with rheumatoid arthritis of hip |
| ICD10CM | M05.451 | Rheumatoid myopathy with rheumatoid arthritis of right hip |
| ICD10CM | M05.452 | Rheumatoid myopathy with rheumatoid arthritis of left hip |
| ICD10CM | M05.459 | Rheumatoid myopathy with rheumatoid arthritis of unspecified hip |
| ICD10CM | M05.46 | Rheumatoid myopathy with rheumatoid arthritis of knee |
| ICD10CM | M05.461 | Rheumatoid myopathy with rheumatoid arthritis of right knee |
| ICD10CM | M05.462 | Rheumatoid myopathy with rheumatoid arthritis of left knee |
| ICD10CM | M05.469 | Rheumatoid myopathy with rheumatoid arthritis of unspecified knee |
| ICD10CM | M05.47 | Rheumatoid myopathy with rheumatoid arthritis of ankle and foot |
| ICD10CM | M05.471 | Rheumatoid myopathy with rheumatoid arthritis of right ankle and foot |
| ICD10CM | M05.472 | Rheumatoid myopathy with rheumatoid arthritis of left ankle and foot |
| ICD10CM | M05.479 | Rheumatoid myopathy with rheumatoid arthritis of unspecified ankle and foot |
| ICD10CM | M05.49 | Rheumatoid myopathy with rheumatoid arthritis of multiple sites |
| ICD10CM | M05.5 | Rheumatoid polyneuropathy with rheumatoid arthritis |
| ICD10CM | M05.50 | Rheumatoid polyneuropathy with rheumatoid arthritis of unspecified site |
| ICD10CM | M05.51 | Rheumatoid polyneuropathy with rheumatoid arthritis of shoulder |
| ICD10CM | M05.511 | Rheumatoid polyneuropathy with rheumatoid arthritis of right shoulder |
| ICD10CM | M05.512 | Rheumatoid polyneuropathy with rheumatoid arthritis of left shoulder |

Continued on next page

**TableS31 – continued from previous page**

| CodeSystem | ConceptCode | ConceptName |
|---|---|---|
| ICD10CM | M05.519 | Rheumatoid polyneuropathy with rheumatoid arthritis of unspecified shoulder |
| ICD10CM | M05.52 | Rheumatoid polyneuropathy with rheumatoid arthritis of elbow |
| ICD10CM | M05.521 | Rheumatoid polyneuropathy with rheumatoid arthritis of right elbow |
| ICD10CM | M05.522 | Rheumatoid polyneuropathy with rheumatoid arthritis of left elbow |
| ICD10CM | M05.529 | Rheumatoid polyneuropathy with rheumatoid arthritis of unspecified elbow |
| ICD10CM | M05.53 | Rheumatoid polyneuropathy with rheumatoid arthritis of wrist |
| ICD10CM | M05.531 | Rheumatoid polyneuropathy with rheumatoid arthritis of right wrist |
| ICD10CM | M05.532 | Rheumatoid polyneuropathy with rheumatoid arthritis of left wrist |
| ICD10CM | M05.539 | Rheumatoid polyneuropathy with rheumatoid arthritis of unspecified wrist |
| ICD10CM | M05.54 | Rheumatoid polyneuropathy with rheumatoid arthritis of hand |
| ICD10CM | M05.541 | Rheumatoid polyneuropathy with rheumatoid arthritis of right hand |
| ICD10CM | M05.542 | Rheumatoid polyneuropathy with rheumatoid arthritis of left hand |
| ICD10CM | M05.549 | Rheumatoid polyneuropathy with rheumatoid arthritis of unspecified hand |
| ICD10CM | M05.55 | Rheumatoid polyneuropathy with rheumatoid arthritis of hip |
| ICD10CM | M05.551 | Rheumatoid polyneuropathy with rheumatoid arthritis of right hip |
| ICD10CM | M05.552 | Rheumatoid polyneuropathy with rheumatoid arthritis of left hip |
| ICD10CM | M05.559 | Rheumatoid polyneuropathy with rheumatoid arthritis of unspecified hip |
| ICD10CM | M05.56 | Rheumatoid polyneuropathy with rheumatoid arthritis of knee |
| ICD10CM | M05.561 | Rheumatoid polyneuropathy with rheumatoid arthritis of right knee |
| ICD10CM | M05.562 | Rheumatoid polyneuropathy with rheumatoid arthritis of left knee |
| ICD10CM | M05.569 | Rheumatoid polyneuropathy with rheumatoid arthritis of unspecified knee |
| ICD10CM | M05.57 | Rheumatoid polyneuropathy with rheumatoid arthritis of ankle and foot |
| ICD10CM | M05.571 | Rheumatoid polyneuropathy with rheumatoid arthritis of right ankle and foot |
| ICD10CM | M05.572 | Rheumatoid polyneuropathy with rheumatoid arthritis of left ankle and foot |
| ICD10CM | M05.579 | Rheumatoid polyneuropathy with rheumatoid arthritis of unspecified ankle and foot |
| ICD10CM | M05.59 | Rheumatoid polyneuropathy with rheumatoid arthritis of multiple sites |

Continued on next page

**TableS31 – continued from previous page**

| CodeSystem | ConceptCode | ConceptName |
|---|---|---|
| ICD10CM | M05.6 | Rheumatoid arthritis with involvement of other organs and systems |
| ICD10CM | M05.60 | Rheumatoid arthritis of unspecified site with involvement of other organs and systems |
| ICD10CM | M05.61 | Rheumatoid arthritis of shoulder with involvement of other organs and systems |
| ICD10CM | M05.611 | Rheumatoid arthritis of right shoulder with involvement of other organs and systems |
| ICD10CM | M05.612 | Rheumatoid arthritis of left shoulder with involvement of other organs and systems |
| ICD10CM | M05.619 | Rheumatoid arthritis of unspecified shoulder with involvement of other organs and systems |
| ICD10CM | M05.62 | Rheumatoid arthritis of elbow with involvement of other organs and systems |
| ICD10CM | M05.621 | Rheumatoid arthritis of right elbow with involvement of other organs and systems |
| ICD10CM | M05.622 | Rheumatoid arthritis of left elbow with involvement of other organs and systems |
| ICD10CM | M05.629 | Rheumatoid arthritis of unspecified elbow with involvement of other organs and systems |
| ICD10CM | M05.63 | Rheumatoid arthritis of wrist with involvement of other organs and systems |
| ICD10CM | M05.631 | Rheumatoid arthritis of right wrist with involvement of other organs and systems |
| ICD10CM | M05.632 | Rheumatoid arthritis of left wrist with involvement of other organs and systems |
| ICD10CM | M05.639 | Rheumatoid arthritis of unspecified wrist with involvement of other organs and systems |
| ICD10CM | M05.64 | Rheumatoid arthritis of hand with involvement of other organs and systems |
| ICD10CM | M05.641 | Rheumatoid arthritis of right hand with involvement of other organs and systems |
| ICD10CM | M05.642 | Rheumatoid arthritis of left hand with involvement of other organs and systems |
| ICD10CM | M05.649 | Rheumatoid arthritis of unspecified hand with involvement of other organs and systems |
| ICD10CM | M05.65 | Rheumatoid arthritis of hip with involvement of other organs and systems |
| ICD10CM | M05.651 | Rheumatoid arthritis of right hip with involvement of other organs and systems |
| ICD10CM | M05.652 | Rheumatoid arthritis of left hip with involvement of other organs and systems |
| ICD10CM | M05.659 | Rheumatoid arthritis of unspecified hip with involvement of other organs and systems |
| ICD10CM | M05.66 | Rheumatoid arthritis of knee with involvement of other organs and systems |

Continued on next page

**TableS31 – continued from previous page**

| CodeSystem | ConceptCode | ConceptName |
|---|---|---|
| ICD10CM | M05.661 | Rheumatoid arthritis of right knee with involvement of other organs and systems |
| ICD10CM | M05.662 | Rheumatoid arthritis of left knee with involvement of other organs and systems |
| ICD10CM | M05.669 | Rheumatoid arthritis of unspecified knee with involvement of other organs and systems |
| ICD10CM | M05.67 | Rheumatoid arthritis of ankle and foot with involvement of other organs and systems |
| ICD10CM | M05.671 | Rheumatoid arthritis of right ankle and foot with involvement of other organs and systems |
| ICD10CM | M05.672 | Rheumatoid arthritis of left ankle and foot with involvement of other organs and systems |
| ICD10CM | M05.679 | Rheumatoid arthritis of unspecified ankle and foot with involvement of other organs and systems |
| ICD10CM | M05.69 | Rheumatoid arthritis of multiple sites with involvement of other organs and systems |
| ICD10CM | M05.7 | Rheumatoid arthritis with rheumatoid factor without organ or systems involvement |
| ICD10CM | M05.70 | Rheumatoid arthritis with rheumatoid factor of unspecified site without organ or systems involvement |
| ICD10CM | M05.71 | Rheumatoid arthritis with rheumatoid factor of shoulder without organ or systems involvement |
| ICD10CM | M05.711 | Rheumatoid arthritis with rheumatoid factor of right shoulder without organ or systems involvement |
| ICD10CM | M05.712 | Rheumatoid arthritis with rheumatoid factor of left shoulder without organ or systems involvement |
| ICD10CM | M05.719 | Rheumatoid arthritis with rheumatoid factor of unspecified shoulder without organ or systems involvement |
| ICD10CM | M05.72 | Rheumatoid arthritis with rheumatoid factor of elbow without organ or systems involvement |
| ICD10CM | M05.721 | Rheumatoid arthritis with rheumatoid factor of right elbow without organ or systems involvement |
| ICD10CM | M05.722 | Rheumatoid arthritis with rheumatoid factor of left elbow without organ or systems involvement |
| ICD10CM | M05.729 | Rheumatoid arthritis with rheumatoid factor of unspecified elbow without organ or systems involvement |
| ICD10CM | M05.73 | Rheumatoid arthritis with rheumatoid factor of wrist without organ or systems involvement |
| ICD10CM | M05.731 | Rheumatoid arthritis with rheumatoid factor of right wrist without organ or systems involvement |
| ICD10CM | M05.732 | Rheumatoid arthritis with rheumatoid factor of left wrist without organ or systems involvement |
| ICD10CM | M05.739 | Rheumatoid arthritis with rheumatoid factor of unspecified wrist without organ or systems involvement |
| ICD10CM | M05.74 | Rheumatoid arthritis with rheumatoid factor of hand without organ or systems involvement |

Continued on next page

**TableS31 – continued from previous page**

| CodeSystem | ConceptCode | ConceptName |
|---|---|---|
| ICD10CM | M05.741 | Rheumatoid arthritis with rheumatoid factor of right hand without organ or systems involvement |
| ICD10CM | M05.742 | Rheumatoid arthritis with rheumatoid factor of left hand without organ or systems involvement |
| ICD10CM | M05.749 | Rheumatoid arthritis with rheumatoid factor of unspecified hand without organ or systems involvement |
| ICD10CM | M05.75 | Rheumatoid arthritis with rheumatoid factor of hip without organ or systems involvement |
| ICD10CM | M05.751 | Rheumatoid arthritis with rheumatoid factor of right hip without organ or systems involvement |
| ICD10CM | M05.752 | Rheumatoid arthritis with rheumatoid factor of left hip without organ or systems involvement |
| ICD10CM | M05.759 | Rheumatoid arthritis with rheumatoid factor of unspecified hip without organ or systems involvement |
| ICD10CM | M05.76 | Rheumatoid arthritis with rheumatoid factor of knee without organ or systems involvement |
| ICD10CM | M05.761 | Rheumatoid arthritis with rheumatoid factor of right knee without organ or systems involvement |
| ICD10CM | M05.762 | Rheumatoid arthritis with rheumatoid factor of left knee without organ or systems involvement |
| ICD10CM | M05.769 | Rheumatoid arthritis with rheumatoid factor of unspecified knee without organ or systems involvement |
| ICD10CM | M05.77 | Rheumatoid arthritis with rheumatoid factor of ankle and foot without organ or systems involvement |
| ICD10CM | M05.771 | Rheumatoid arthritis with rheumatoid factor of right ankle and foot without organ or systems involvement |
| ICD10CM | M05.772 | Rheumatoid arthritis with rheumatoid factor of left ankle and foot without organ or systems involvement |
| ICD10CM | M05.779 | Rheumatoid arthritis with rheumatoid factor of unspecified ankle and foot without organ or systems involvement |
| ICD10CM | M05.852 | Other rheumatoid arthritis with rheumatoid factor of left hip |
| ICD10CM | M05.79 | Rheumatoid arthritis with rheumatoid factor of multiple sites without organ or systems involvement |
| ICD10CM | M05.8 | Other rheumatoid arthritis with rheumatoid factor |
| ICD10CM | M05.80 | Other rheumatoid arthritis with rheumatoid factor of unspecified site |
| ICD10CM | M05.81 | Other rheumatoid arthritis with rheumatoid factor of shoulder |
| ICD10CM | M05.811 | Other rheumatoid arthritis with rheumatoid factor of right shoulder |
| ICD10CM | M05.812 | Other rheumatoid arthritis with rheumatoid factor of left shoulder |
| ICD10CM | M05.819 | Other rheumatoid arthritis with rheumatoid factor of unspecified shoulder |
| ICD10CM | M05.82 | Other rheumatoid arthritis with rheumatoid factor of elbow |
| ICD10CM | M05.821 | Other rheumatoid arthritis with rheumatoid factor of right elbow |
| ICD10CM | M05.822 | Other rheumatoid arthritis with rheumatoid factor of left elbow |

Continued on next page

**TableS31 – continued from previous page**

| CodeSystem | ConceptCode | ConceptName |
|---|---|---|
| ICD10CM | M05.829 | Other rheumatoid arthritis with rheumatoid factor of unspecified elbow |
| ICD10CM | M05.83 | Other rheumatoid arthritis with rheumatoid factor of wrist |
| ICD10CM | M05.831 | Other rheumatoid arthritis with rheumatoid factor of right wrist |
| ICD10CM | M05.832 | Other rheumatoid arthritis with rheumatoid factor of left wrist |
| ICD10CM | M05.839 | Other rheumatoid arthritis with rheumatoid factor of unspecified wrist |
| ICD10CM | M05.84 | Other rheumatoid arthritis with rheumatoid factor of hand |
| ICD10CM | M05.841 | Other rheumatoid arthritis with rheumatoid factor of right hand |
| ICD10CM | M05.842 | Other rheumatoid arthritis with rheumatoid factor of left hand |
| ICD10CM | M05.849 | Other rheumatoid arthritis with rheumatoid factor of unspecified hand |
| ICD10CM | M05.85 | Other rheumatoid arthritis with rheumatoid factor of hip |
| ICD10CM | M05.851 | Other rheumatoid arthritis with rheumatoid factor of right hip |
| ICD10CM | M05.859 | Other rheumatoid arthritis with rheumatoid factor of unspecified hip |
| ICD10CM | M05.86 | Other rheumatoid arthritis with rheumatoid factor of knee |
| ICD10CM | M05.861 | Other rheumatoid arthritis with rheumatoid factor of right knee |
| ICD10CM | M05.862 | Other rheumatoid arthritis with rheumatoid factor of left knee |
| ICD10CM | M05.869 | Other rheumatoid arthritis with rheumatoid factor of unspecified knee |
| ICD10CM | M05.87 | Other rheumatoid arthritis with rheumatoid factor of ankle and foot |
| ICD10CM | M05.871 | Other rheumatoid arthritis with rheumatoid factor of right ankle and foot |
| ICD10CM | M05.872 | Other rheumatoid arthritis with rheumatoid factor of left ankle and foot |
| ICD10CM | M05.879 | Other rheumatoid arthritis with rheumatoid factor of unspecified ankle and foot |
| ICD10CM | M05.89 | Other rheumatoid arthritis with rheumatoid factor of multiple sites |
| ICD10CM | M05.9 | Rheumatoid arthritis with rheumatoid factor, unspecified |
| ICD10CM | M06.0 | Rheumatoid arthritis without rheumatoid factor |
| ICD10CM | M06.00 | Rheumatoid arthritis without rheumatoid factor, unspecified site |
| ICD10CM | M06.01 | Rheumatoid arthritis without rheumatoid factor, shoulder |
| ICD10CM | M06.011 | Rheumatoid arthritis without rheumatoid factor, right shoulder |
| ICD10CM | M06.012 | Rheumatoid arthritis without rheumatoid factor, left shoulder |
| ICD10CM | M06.019 | Rheumatoid arthritis without rheumatoid factor, unspecified shoulder |
| ICD10CM | M06.02 | Rheumatoid arthritis without rheumatoid factor, elbow |
| ICD10CM | M06.021 | Rheumatoid arthritis without rheumatoid factor, right elbow |
| ICD10CM | M06.022 | Rheumatoid arthritis without rheumatoid factor, left elbow |
| ICD10CM | M06.029 | Rheumatoid arthritis without rheumatoid factor, unspecified elbow |
| ICD10CM | M06.03 | Rheumatoid arthritis without rheumatoid factor, wrist |
| ICD10CM | M06.031 | Rheumatoid arthritis without rheumatoid factor, right wrist |
| ICD10CM | M06.032 | Rheumatoid arthritis without rheumatoid factor, left wrist |

Continued on next page

**TableS31 – continued from previous page**

| CodeSystem | ConceptCode | ConceptName |
|---|---|---|
| ICD10CM | M06.039 | Rheumatoid arthritis without rheumatoid factor, unspecified wrist |
| ICD10CM | M06.04 | Rheumatoid arthritis without rheumatoid factor, hand |
| ICD10CM | M06.041 | Rheumatoid arthritis without rheumatoid factor, right hand |
| ICD10CM | M06.042 | Rheumatoid arthritis without rheumatoid factor, left hand |
| ICD10CM | M06.049 | Rheumatoid arthritis without rheumatoid factor, unspecified hand |
| ICD10CM | M06.05 | Rheumatoid arthritis without rheumatoid factor, hip |
| ICD10CM | M06.051 | Rheumatoid arthritis without rheumatoid factor, right hip |
| ICD10CM | M06.052 | Rheumatoid arthritis without rheumatoid factor, left hip |
| ICD10CM | M06.059 | Rheumatoid arthritis without rheumatoid factor, unspecified hip |
| ICD10CM | M06.06 | Rheumatoid arthritis without rheumatoid factor, knee |
| ICD10CM | M06.061 | Rheumatoid arthritis without rheumatoid factor, right knee |
| ICD10CM | M06.062 | Rheumatoid arthritis without rheumatoid factor, left knee |
| ICD10CM | M06.069 | Rheumatoid arthritis without rheumatoid factor, unspecified knee |
| ICD10CM | M06.07 | Rheumatoid arthritis without rheumatoid factor, ankle and foot |
| ICD10CM | M06.071 | Rheumatoid arthritis without rheumatoid factor, right ankle and foot |
| ICD10CM | M06.072 | Rheumatoid arthritis without rheumatoid factor, left ankle and foot |
| ICD10CM | M06.079 | Rheumatoid arthritis without rheumatoid factor, unspecified ankle and foot |
| ICD10CM | M06.08 | Rheumatoid arthritis without rheumatoid factor, vertebrae |
| ICD10CM | M06.09 | Rheumatoid arthritis without rheumatoid factor, multiple sites |
| ICD10CM | M06.1 | Adult-onset Still's disease |
| ICD10CM | M06.2 | Rheumatoid bursitis |
| ICD10CM | M06.20 | Rheumatoid bursitis, unspecified site |
| ICD10CM | M06.21 | Rheumatoid bursitis, shoulder |
| ICD10CM | M06.211 | Rheumatoid bursitis, right shoulder |
| ICD10CM | M06.212 | Rheumatoid bursitis, left shoulder |
| ICD10CM | M06.219 | Rheumatoid bursitis, unspecified shoulder |
| ICD10CM | M06.22 | Rheumatoid bursitis, elbow |
| ICD10CM | M06.221 | Rheumatoid bursitis, right elbow |
| ICD10CM | M06.222 | Rheumatoid bursitis, left elbow |
| ICD10CM | M06.229 | Rheumatoid bursitis, unspecified elbow |
| ICD10CM | M06.23 | Rheumatoid bursitis, wrist |
| ICD10CM | M06.231 | Rheumatoid bursitis, right wrist |
| ICD10CM | M06.232 | Rheumatoid bursitis, left wrist |
| ICD10CM | M06.239 | Rheumatoid bursitis, unspecified wrist |
| ICD10CM | M06.24 | Rheumatoid bursitis, hand |
| ICD10CM | M06.241 | Rheumatoid bursitis, right hand |
| ICD10CM | M06.242 | Rheumatoid bursitis, left hand |
| ICD10CM | M06.249 | Rheumatoid bursitis, unspecified hand |
| ICD10CM | M06.25 | Rheumatoid bursitis, hip |
| ICD10CM | M06.251 | Rheumatoid bursitis, right hip |
| ICD10CM | M06.252 | Rheumatoid bursitis, left hip |

Continued on next page

**TableS31 – continued from previous page**

| CodeSystem | ConceptCode | ConceptName |
|---|---|---|
| ICD10CM | M06.259 | Rheumatoid bursitis, unspecified hip |
| ICD10CM | M06.26 | Rheumatoid bursitis, knee |
| ICD10CM | M06.261 | Rheumatoid bursitis, right knee |
| ICD10CM | M06.262 | Rheumatoid bursitis, left knee |
| ICD10CM | M06.269 | Rheumatoid bursitis, unspecified knee |
| ICD10CM | M06.27 | Rheumatoid bursitis, ankle and foot |
| ICD10CM | M06.271 | Rheumatoid bursitis, right ankle and foot |
| ICD10CM | M06.272 | Rheumatoid bursitis, left ankle and foot |
| ICD10CM | M06.279 | Rheumatoid bursitis, unspecified ankle and foot |
| ICD10CM | M06.28 | Rheumatoid bursitis, vertebrae |
| ICD10CM | M06.29 | Rheumatoid bursitis, multiple sites |
| ICD10CM | M06.3 | Rheumatoid nodule |
| ICD10CM | M06.30 | Rheumatoid nodule, unspecified site |
| ICD10CM | M06.31 | Rheumatoid nodule, shoulder |
| ICD10CM | M06.311 | Rheumatoid nodule, right shoulder |
| ICD10CM | M06.312 | Rheumatoid nodule, left shoulder |
| ICD10CM | M06.319 | Rheumatoid nodule, unspecified shoulder |
| ICD10CM | M06.32 | Rheumatoid nodule, elbow |
| ICD10CM | M06.321 | Rheumatoid nodule, right elbow |
| ICD10CM | M06.322 | Rheumatoid nodule, left elbow |
| ICD10CM | M06.329 | Rheumatoid nodule, unspecified elbow |
| ICD10CM | M06.33 | Rheumatoid nodule, wrist |
| ICD10CM | M06.331 | Rheumatoid nodule, right wrist |
| ICD10CM | M06.332 | Rheumatoid nodule, left wrist |
| ICD10CM | M06.339 | Rheumatoid nodule, unspecified wrist |
| ICD10CM | M06.34 | Rheumatoid nodule, hand |
| ICD10CM | M06.341 | Rheumatoid nodule, right hand |
| ICD10CM | M06.342 | Rheumatoid nodule, left hand |
| ICD10CM | M06.349 | Rheumatoid nodule, unspecified hand |
| ICD10CM | M06.35 | Rheumatoid nodule, hip |
| ICD10CM | M06.351 | Rheumatoid nodule, right hip |
| ICD10CM | M06.352 | Rheumatoid nodule, left hip |
| ICD10CM | M06.359 | Rheumatoid nodule, unspecified hip |
| ICD10CM | M06.36 | Rheumatoid nodule, knee |
| ICD10CM | M06.361 | Rheumatoid nodule, right knee |
| ICD10CM | M06.362 | Rheumatoid nodule, left knee |
| ICD10CM | M06.369 | Rheumatoid nodule, unspecified knee |
| ICD10CM | M06.37 | Rheumatoid nodule, ankle and foot |
| ICD10CM | M06.371 | Rheumatoid nodule, right ankle and foot |
| ICD10CM | M06.372 | Rheumatoid nodule, left ankle and foot |
| ICD10CM | M06.379 | Rheumatoid nodule, unspecified ankle and foot |
| ICD10CM | M06.38 | Rheumatoid nodule, vertebrae |
| ICD10CM | M06.39 | Rheumatoid nodule, multiple sites |
| ICD10CM | M06.4 | Inflammatory polyarthropathy |
| ICD10CM | M06.8 | Other specified rheumatoid arthritis |
| ICD10CM | M06.80 | Other specified rheumatoid arthritis, unspecified site |
| ICD10CM | M06.81 | Other specified rheumatoid arthritis, shoulder |

Continued on next page

**TableS31 – continued from previous page**

| CodeSystem | ConceptCode | ConceptName |
|---|---|---|
| ICD10CM | M06.811 | Other specified rheumatoid arthritis, right shoulder |
| ICD10CM | M06.812 | Other specified rheumatoid arthritis, left shoulder |
| ICD10CM | M06.819 | Other specified rheumatoid arthritis, unspecified shoulder |
| ICD10CM | M06.82 | Other specified rheumatoid arthritis, elbow |
| ICD10CM | M06.821 | Other specified rheumatoid arthritis, right elbow |
| ICD10CM | M06.822 | Other specified rheumatoid arthritis, left elbow |
| ICD10CM | M06.829 | Other specified rheumatoid arthritis, unspecified elbow |
| ICD10CM | M06.83 | Other specified rheumatoid arthritis, wrist |
| ICD10CM | M06.831 | Other specified rheumatoid arthritis, right wrist |
| ICD10CM | M06.832 | Other specified rheumatoid arthritis, left wrist |
| ICD10CM | M06.839 | Other specified rheumatoid arthritis, unspecified wrist |
| ICD10CM | M06.84 | Other specified rheumatoid arthritis, hand |
| ICD10CM | M06.841 | Other specified rheumatoid arthritis, right hand |
| ICD10CM | M06.842 | Other specified rheumatoid arthritis, left hand |
| ICD10CM | M06.849 | Other specified rheumatoid arthritis, unspecified hand |
| ICD10CM | M06.85 | Other specified rheumatoid arthritis, hip |
| ICD10CM | M06.851 | Other specified rheumatoid arthritis, right hip |
| ICD10CM | M06.852 | Other specified rheumatoid arthritis, left hip |
| ICD10CM | M06.859 | Other specified rheumatoid arthritis, unspecified hip |
| ICD10CM | M06.86 | Other specified rheumatoid arthritis, knee |
| ICD10CM | M06.861 | Other specified rheumatoid arthritis, right knee |
| ICD10CM | M06.862 | Other specified rheumatoid arthritis, left knee |
| ICD10CM | M06.869 | Other specified rheumatoid arthritis, unspecified knee |
| ICD10CM | M06.87 | Other specified rheumatoid arthritis, ankle and foot |
| ICD10CM | M06.871 | Other specified rheumatoid arthritis, right ankle and foot |
| ICD10CM | M06.872 | Other specified rheumatoid arthritis, left ankle and foot |
| ICD10CM | M06.879 | Other specified rheumatoid arthritis, unspecified ankle and foot |
| ICD10CM | M06.88 | Other specified rheumatoid arthritis, vertebrae |
| ICD10CM | M06.89 | Other specified rheumatoid arthritis, multiple sites |
| ICD10CM | M06.9 | Rheumatoid arthritis, unspecified |
| ICD10CM | M07.6 | Enteropathic arthropathies |
| ICD10CM | M07.60 | Enteropathic arthropathies, unspecified site |
| ICD10CM | M07.61 | Enteropathic arthropathies, shoulder |
| ICD10CM | M07.611 | Enteropathic arthropathies, right shoulder |
| ICD10CM | M07.612 | Enteropathic arthropathies, left shoulder |
| ICD10CM | M07.619 | Enteropathic arthropathies, unspecified shoulder |
| ICD10CM | M07.62 | Enteropathic arthropathies, elbow |
| ICD10CM | M07.621 | Enteropathic arthropathies, right elbow |
| ICD10CM | M07.622 | Enteropathic arthropathies, left elbow |
| ICD10CM | M07.629 | Enteropathic arthropathies, unspecified elbow |
| ICD10CM | M07.63 | Enteropathic arthropathies, wrist |
| ICD10CM | M07.631 | Enteropathic arthropathies, right wrist |
| ICD10CM | M07.632 | Enteropathic arthropathies, left wrist |
| ICD10CM | M07.639 | Enteropathic arthropathies, unspecified wrist |
| ICD10CM | M07.64 | Enteropathic arthropathies, hand |
| ICD10CM | M07.641 | Enteropathic arthropathies, right hand |
| ICD10CM | M07.642 | Enteropathic arthropathies, left hand |

Continued on next page

**TableS31 – continued from previous page**

| CodeSystem | ConceptCode | ConceptName |
|---|---|---|
| ICD10CM | M07.649 | Enteropathic arthropathies, unspecified hand |
| ICD10CM | M07.65 | Enteropathic arthropathies, hip |
| ICD10CM | M07.651 | Enteropathic arthropathies, right hip |
| ICD10CM | M07.652 | Enteropathic arthropathies, left hip |
| ICD10CM | M07.659 | Enteropathic arthropathies, unspecified hip |
| ICD10CM | M07.66 | Enteropathic arthropathies, knee |
| ICD10CM | M07.661 | Enteropathic arthropathies, right knee |
| ICD10CM | M07.662 | Enteropathic arthropathies, left knee |
| ICD10CM | M07.669 | Enteropathic arthropathies, unspecified knee |
| ICD10CM | M07.67 | Enteropathic arthropathies, ankle and foot |
| ICD10CM | M07.671 | Enteropathic arthropathies, right ankle and foot |
| ICD10CM | M07.672 | Enteropathic arthropathies, left ankle and foot |
| ICD10CM | M07.679 | Enteropathic arthropathies, unspecified ankle and foot |
| ICD10CM | M07.68 | Enteropathic arthropathies, vertebrae |
| ICD10CM | M07.69 | Enteropathic arthropathies, multiple sites |
| ICD10CM | M08.0 | Unspecified juvenile rheumatoid arthritis |
| ICD10CM | M08.00 | Unspecified juvenile rheumatoid arthritis of unspecified site |
| ICD10CM | M08.01 | Unspecified juvenile rheumatoid arthritis, shoulder |
| ICD10CM | M08.011 | Unspecified juvenile rheumatoid arthritis, right shoulder |
| ICD10CM | M08.012 | Unspecified juvenile rheumatoid arthritis, left shoulder |
| ICD10CM | M08.019 | Unspecified juvenile rheumatoid arthritis, unspecified shoulder |
| ICD10CM | M08.02 | Unspecified juvenile rheumatoid arthritis of elbow |
| ICD10CM | M08.021 | Unspecified juvenile rheumatoid arthritis, right elbow |
| ICD10CM | M08.022 | Unspecified juvenile rheumatoid arthritis, left elbow |
| ICD10CM | M08.029 | Unspecified juvenile rheumatoid arthritis, unspecified elbow |
| ICD10CM | M08.03 | Unspecified juvenile rheumatoid arthritis, wrist |
| ICD10CM | M08.031 | Unspecified juvenile rheumatoid arthritis, right wrist |
| ICD10CM | M08.032 | Unspecified juvenile rheumatoid arthritis, left wrist |
| ICD10CM | M08.039 | Unspecified juvenile rheumatoid arthritis, unspecified wrist |
| ICD10CM | M08.04 | Unspecified juvenile rheumatoid arthritis, hand |
| ICD10CM | M08.041 | Unspecified juvenile rheumatoid arthritis, right hand |
| ICD10CM | M08.042 | Unspecified juvenile rheumatoid arthritis, left hand |
| ICD10CM | M08.049 | Unspecified juvenile rheumatoid arthritis, unspecified hand |
| ICD10CM | M08.05 | Unspecified juvenile rheumatoid arthritis, hip |
| ICD10CM | M08.051 | Unspecified juvenile rheumatoid arthritis, right hip |
| ICD10CM | M08.052 | Unspecified juvenile rheumatoid arthritis, left hip |
| ICD10CM | M08.059 | Unspecified juvenile rheumatoid arthritis, unspecified hip |
| ICD10CM | M08.06 | Unspecified juvenile rheumatoid arthritis, knee |
| ICD10CM | M08.061 | Unspecified juvenile rheumatoid arthritis, right knee |
| ICD10CM | M08.062 | Unspecified juvenile rheumatoid arthritis, left knee |
| ICD10CM | M08.069 | Unspecified juvenile rheumatoid arthritis, unspecified knee |
| ICD10CM | M08.07 | Unspecified juvenile rheumatoid arthritis, ankle and foot |
| ICD10CM | M08.071 | Unspecified juvenile rheumatoid arthritis, right ankle and foot |
| ICD10CM | M08.072 | Unspecified juvenile rheumatoid arthritis, left ankle and foot |
| ICD10CM | M08.079 | Unspecified juvenile rheumatoid arthritis, unspecified ankle and foot |
| ICD10CM | M08.08 | Unspecified juvenile rheumatoid arthritis, vertebrae |

Continued on next page

**TableS31 – continued from previous page**

| CodeSystem | ConceptCode | ConceptName |
|---|---|---|
| ICD10CM | M08.09 | Unspecified juvenile rheumatoid arthritis, multiple sites |
| ICD10CM | M08.1 | Juvenile ankylosing spondylitis |
| ICD10CM | M08.2 | Juvenile rheumatoid arthritis with systemic onset |
| ICD10CM | M08.20 | Juvenile rheumatoid arthritis with systemic onset, unspecified site |
| ICD10CM | M08.21 | Juvenile rheumatoid arthritis with systemic onset, shoulder |
| ICD10CM | M08.211 | Juvenile rheumatoid arthritis with systemic onset, right shoulder |
| ICD10CM | M08.212 | Juvenile rheumatoid arthritis with systemic onset, left shoulder |
| ICD10CM | M08.219 | Juvenile rheumatoid arthritis with systemic onset, unspecified shoulder |
| ICD10CM | M08.22 | Juvenile rheumatoid arthritis with systemic onset, elbow |
| ICD10CM | M08.221 | Juvenile rheumatoid arthritis with systemic onset, right elbow |
| ICD10CM | M08.222 | Juvenile rheumatoid arthritis with systemic onset, left elbow |
| ICD10CM | M08.229 | Juvenile rheumatoid arthritis with systemic onset, unspecified elbow |
| ICD10CM | M08.23 | Juvenile rheumatoid arthritis with systemic onset, wrist |
| ICD10CM | M08.231 | Juvenile rheumatoid arthritis with systemic onset, right wrist |
| ICD10CM | M08.232 | Juvenile rheumatoid arthritis with systemic onset, left wrist |
| ICD10CM | M08.239 | Juvenile rheumatoid arthritis with systemic onset, unspecified wrist |
| ICD10CM | M08.24 | Juvenile rheumatoid arthritis with systemic onset, hand |
| ICD10CM | M08.241 | Juvenile rheumatoid arthritis with systemic onset, right hand |
| ICD10CM | M08.242 | Juvenile rheumatoid arthritis with systemic onset, left hand |
| ICD10CM | M08.249 | Juvenile rheumatoid arthritis with systemic onset, unspecified hand |
| ICD10CM | M08.25 | Juvenile rheumatoid arthritis with systemic onset, hip |
| ICD10CM | M08.251 | Juvenile rheumatoid arthritis with systemic onset, right hip |
| ICD10CM | M08.252 | Juvenile rheumatoid arthritis with systemic onset, left hip |
| ICD10CM | M08.259 | Juvenile rheumatoid arthritis with systemic onset, unspecified hip |
| ICD10CM | M08.26 | Juvenile rheumatoid arthritis with systemic onset, knee |
| ICD10CM | M08.261 | Juvenile rheumatoid arthritis with systemic onset, right knee |
| ICD10CM | M08.262 | Juvenile rheumatoid arthritis with systemic onset, left knee |
| ICD10CM | M08.269 | Juvenile rheumatoid arthritis with systemic onset, unspecified knee |
| ICD10CM | M08.27 | Juvenile rheumatoid arthritis with systemic onset, ankle and foot |
| ICD10CM | M08.271 | Juvenile rheumatoid arthritis with systemic onset, right ankle and foot |
| ICD10CM | M08.272 | Juvenile rheumatoid arthritis with systemic onset, left ankle and foot |
| ICD10CM | M08.279 | Juvenile rheumatoid arthritis with systemic onset, unspecified ankle and foot |
| ICD10CM | M08.28 | Juvenile rheumatoid arthritis with systemic onset, vertebrae |
| ICD10CM | M08.29 | Juvenile rheumatoid arthritis with systemic onset, multiple sites |
| ICD10CM | M08.3 | Juvenile rheumatoid polyarthritis (seronegative) |
| ICD10CM | M08.4 | Pauciarticular juvenile rheumatoid arthritis |
| ICD10CM | M08.40 | Pauciarticular juvenile rheumatoid arthritis, unspecified site |

Continued on next page

**TableS31 – continued from previous page**

| CodeSystem | ConceptCode | ConceptName |
|---|---|---|
| ICD10CM | M08.41 | Pauciarticular juvenile rheumatoid arthritis, shoulder |
| ICD10CM | M08.411 | Pauciarticular juvenile rheumatoid arthritis, right shoulder |
| ICD10CM | M08.412 | Pauciarticular juvenile rheumatoid arthritis, left shoulder |
| ICD10CM | M08.419 | Pauciarticular juvenile rheumatoid arthritis, unspecified shoulder |
| ICD10CM | M08.42 | Pauciarticular juvenile rheumatoid arthritis, elbow |
| ICD10CM | M08.421 | Pauciarticular juvenile rheumatoid arthritis, right elbow |
| ICD10CM | M08.422 | Pauciarticular juvenile rheumatoid arthritis, left elbow |
| ICD10CM | M08.429 | Pauciarticular juvenile rheumatoid arthritis, unspecified elbow |
| ICD10CM | M08.43 | Pauciarticular juvenile rheumatoid arthritis, wrist |
| ICD10CM | M08.431 | Pauciarticular juvenile rheumatoid arthritis, right wrist |
| ICD10CM | M08.432 | Pauciarticular juvenile rheumatoid arthritis, left wrist |
| ICD10CM | M08.439 | Pauciarticular juvenile rheumatoid arthritis, unspecified wrist |
| ICD10CM | M08.44 | Pauciarticular juvenile rheumatoid arthritis, hand |
| ICD10CM | M08.441 | Pauciarticular juvenile rheumatoid arthritis, right hand |
| ICD10CM | M08.442 | Pauciarticular juvenile rheumatoid arthritis, left hand |
| ICD10CM | M08.449 | Pauciarticular juvenile rheumatoid arthritis, unspecified hand |
| ICD10CM | M08.45 | Pauciarticular juvenile rheumatoid arthritis, hip |
| ICD10CM | M08.451 | Pauciarticular juvenile rheumatoid arthritis, right hip |
| ICD10CM | M08.452 | Pauciarticular juvenile rheumatoid arthritis, left hip |
| ICD10CM | M08.459 | Pauciarticular juvenile rheumatoid arthritis, unspecified hip |
| ICD10CM | M08.46 | Pauciarticular juvenile rheumatoid arthritis, knee |
| ICD10CM | M08.461 | Pauciarticular juvenile rheumatoid arthritis, right knee |
| ICD10CM | M08.462 | Pauciarticular juvenile rheumatoid arthritis, left knee |
| ICD10CM | M08.469 | Pauciarticular juvenile rheumatoid arthritis, unspecified knee |
| ICD10CM | M08.47 | Pauciarticular juvenile rheumatoid arthritis, ankle and foot |
| ICD10CM | M08.471 | Pauciarticular juvenile rheumatoid arthritis, right ankle and foot |
| ICD10CM | M08.472 | Pauciarticular juvenile rheumatoid arthritis, left ankle and foot |
| ICD10CM | M08.479 | Pauciarticular juvenile rheumatoid arthritis, unspecified ankle and foot |
| ICD10CM | M08.48 | Pauciarticular juvenile rheumatoid arthritis, vertebrae |
| ICD10CM | M08.8 | Other juvenile arthritis |
| ICD10CM | M08.80 | Other juvenile arthritis, unspecified site |
| ICD10CM | M08.81 | Other juvenile arthritis, shoulder |
| ICD10CM | M08.811 | Other juvenile arthritis, right shoulder |
| ICD10CM | M08.812 | Other juvenile arthritis, left shoulder |
| ICD10CM | M08.819 | Other juvenile arthritis, unspecified shoulder |
| ICD10CM | M08.82 | Other juvenile arthritis, elbow |
| ICD10CM | M08.821 | Other juvenile arthritis, right elbow |
| ICD10CM | M08.822 | Other juvenile arthritis, left elbow |
| ICD10CM | M08.829 | Other juvenile arthritis, unspecified elbow |
| ICD10CM | M08.83 | Other juvenile arthritis, wrist |
| ICD10CM | M08.831 | Other juvenile arthritis, right wrist |
| ICD10CM | M08.832 | Other juvenile arthritis, left wrist |
| ICD10CM | M08.839 | Other juvenile arthritis, unspecified wrist |
| ICD10CM | M08.84 | Other juvenile arthritis, hand |
| ICD10CM | M08.841 | Other juvenile arthritis, right hand |

Continued on next page

**TableS31 – continued from previous page**

| CodeSystem | ConceptCode | ConceptName |
|---|---|---|
| ICD10CM | M08.842 | Other juvenile arthritis, left hand |
| ICD10CM | M08.849 | Other juvenile arthritis, unspecified hand |
| ICD10CM | M08.85 | Other juvenile arthritis, hip |
| ICD10CM | M08.851 | Other juvenile arthritis, right hip |
| ICD10CM | M08.852 | Other juvenile arthritis, left hip |
| ICD10CM | M08.859 | Other juvenile arthritis, unspecified hip |
| ICD10CM | M08.86 | Other juvenile arthritis, knee |
| ICD10CM | M08.861 | Other juvenile arthritis, right knee |
| ICD10CM | M08.862 | Other juvenile arthritis, left knee |
| ICD10CM | M08.869 | Other juvenile arthritis, unspecified knee |
| ICD10CM | M08.87 | Other juvenile arthritis, ankle and foot |
| ICD10CM | M08.871 | Other juvenile arthritis, right ankle and foot |
| ICD10CM | M08.872 | Other juvenile arthritis, left ankle and foot |
| ICD10CM | M08.879 | Other juvenile arthritis, unspecified ankle and foot |
| ICD10CM | M08.88 | Other juvenile arthritis, other specified site |
| ICD10CM | M08.89 | Other juvenile arthritis, multiple sites |
| ICD10CM | M08.9 | Juvenile arthritis, unspecified |
| ICD10CM | M08.90 | Juvenile arthritis, unspecified, unspecified site |
| ICD10CM | M08.91 | Juvenile arthritis, unspecified, shoulder |
| ICD10CM | M08.911 | Juvenile arthritis, unspecified, right shoulder |
| ICD10CM | M08.912 | Juvenile arthritis, unspecified, left shoulder |
| ICD10CM | M08.919 | Juvenile arthritis, unspecified, unspecified shoulder |
| ICD10CM | M08.92 | Juvenile arthritis, unspecified, elbow |
| ICD10CM | M08.921 | Juvenile arthritis, unspecified, right elbow |
| ICD10CM | M08.922 | Juvenile arthritis, unspecified, left elbow |
| ICD10CM | M08.929 | Juvenile arthritis, unspecified, unspecified elbow |
| ICD10CM | M08.93 | Juvenile arthritis, unspecified, wrist |
| ICD10CM | M08.931 | Juvenile arthritis, unspecified, right wrist |
| ICD10CM | M08.932 | Juvenile arthritis, unspecified, left wrist |
| ICD10CM | M08.939 | Juvenile arthritis, unspecified, unspecified wrist |
| ICD10CM | M08.94 | Juvenile arthritis, unspecified, hand |
| ICD10CM | M08.941 | Juvenile arthritis, unspecified, right hand |
| ICD10CM | M08.942 | Juvenile arthritis, unspecified, left hand |
| ICD10CM | M08.949 | Juvenile arthritis, unspecified, unspecified hand |
| ICD10CM | M08.95 | Juvenile arthritis, unspecified, hip |
| ICD10CM | M08.951 | Juvenile arthritis, unspecified, right hip |
| ICD10CM | M08.952 | Juvenile arthritis, unspecified, left hip |
| ICD10CM | M08.959 | Juvenile arthritis, unspecified, unspecified hip |
| ICD10CM | M08.96 | Juvenile arthritis, unspecified, knee |
| ICD10CM | M08.961 | Juvenile arthritis, unspecified, right knee |
| ICD10CM | M08.962 | Juvenile arthritis, unspecified, left knee |
| ICD10CM | M08.969 | Juvenile arthritis, unspecified, unspecified knee |
| ICD10CM | M08.97 | Juvenile arthritis, unspecified, ankle and foot |
| ICD10CM | M08.971 | Juvenile arthritis, unspecified, right ankle and foot |
| ICD10CM | M08.972 | Juvenile arthritis, unspecified, left ankle and foot |
| ICD10CM | M08.979 | Juvenile arthritis, unspecified, unspecified ankle and foot |
| ICD10CM | M08.98 | Juvenile arthritis, unspecified, vertebrae |

Continued on next page

**TableS31 – continued from previous page**

| CodeSystem | ConceptCode | ConceptName |
|---|---|---|
| ICD10CM | M08.99 | Juvenile arthritis, unspecified, multiple sites |
| ICD10CM | M30.0 | Polyarteritis nodosa |
| ICD10CM | M30.1 | Polyarteritis with lung involvement [Churg-Strauss] |
| ICD10CM | M30.2 | Juvenile polyarteritis |
| ICD10CM | M30.3 | Mucocutaneous lymph node syndrome [Kawasaki] |
| ICD10CM | M30.8 | Other conditions related to polyarteritis nodosa |
| ICD10CM | M31.30 | Wegener's granulomatosis without renal involvement |
| ICD10CM | M31.31 | Wegener's granulomatosis with renal involvement |
| ICD10CM | M32.0 | Drug-induced systemic lupus erythematosus |
| ICD10CM | M32.1 | Systemic lupus erythematosus with organ or system involvement |
| ICD10CM | M32.10 | Systemic lupus erythematosus, organ or system involvement unspecified |
| ICD10CM | M32.11 | Endocarditis in systemic lupus erythematosus |
| ICD10CM | M32.12 | Pericarditis in systemic lupus erythematosus |
| ICD10CM | M32.13 | Lung involvement in systemic lupus erythematosus |
| ICD10CM | M32.14 | Glomerular disease in systemic lupus erythematosus |
| ICD10CM | M32.15 | Tubulo-interstitial nephropathy in systemic lupus erythematosus |
| ICD10CM | M32.19 | Other organ or system involvement in systemic lupus erythematosus |
| ICD10CM | M32.8 | Other forms of systemic lupus erythematosus |
| ICD10CM | M32.9 | Systemic lupus erythematosus, unspecified |
| ICD10CM | M33.0 | Juvenile dermatomyositis |
| ICD10CM | M33.00 | Juvenile dermatomyositis, organ involvement unspecified |
| ICD10CM | M33.01 | Juvenile dermatomyositis with respiratory involvement |
| ICD10CM | M33.02 | Juvenile dermatomyositis with myopathy |
| ICD10CM | M33.03 | Juvenile dermatomyositis without myopathy |
| ICD10CM | M33.09 | Juvenile dermatomyositis with other organ involvement |
| ICD10CM | M33.1 | Other dermatomyositis |
| ICD10CM | M33.10 | Other dermatomyositis, organ involvement unspecified |
| ICD10CM | M33.11 | Other dermatomyositis with respiratory involvement |
| ICD10CM | M33.12 | Other dermatomyositis with myopathy |
| ICD10CM | M33.13 | Other dermatomyositis without myopathy |
| ICD10CM | M33.19 | Other dermatomyositis with other organ involvement |
| ICD10CM | M33.2 | Polymyositis |
| ICD10CM | M33.20 | Polymyositis, organ involvement unspecified |
| ICD10CM | M33.21 | Polymyositis with respiratory involvement |
| ICD10CM | M33.22 | Polymyositis with myopathy |
| ICD10CM | M33.29 | Polymyositis with other organ involvement |
| ICD10CM | M33.9 | Dermatopolymyositis, unspecified |
| ICD10CM | M33.90 | Dermatopolymyositis, unspecified, organ involvement unspecified |
| ICD10CM | M33.91 | Dermatopolymyositis, unspecified with respiratory involvement |
| ICD10CM | M33.92 | Dermatopolymyositis, unspecified with myopathy |
| ICD10CM | M33.93 | Dermatopolymyositis, unspecified without myopathy |
| ICD10CM | M33.99 | Dermatopolymyositis, unspecified with other organ involvement |
| ICD10CM | M34.0 | Progressive systemic sclerosis |
| ICD10CM | M34.1 | CR(E)ST syndrome |

Continued on next page

**TableS31 – continued from previous page**

| CodeSystem | ConceptCode | ConceptName |
|---|---|---|
| ICD10CM | M34.2 | Systemic sclerosis induced by drug and chemical |
| ICD10CM | M34.8 | Other forms of systemic sclerosis |
| ICD10CM | M34.81 | Systemic sclerosis with lung involvement |
| ICD10CM | M34.82 | Systemic sclerosis with myopathy |
| ICD10CM | M34.83 | Systemic sclerosis with polyneuropathy |
| ICD10CM | M34.89 | Other systemic sclerosis |
| ICD10CM | M34.9 | Systemic sclerosis, unspecified |
| ICD10CM | M46.0 | Spinal enthesopathy |
| ICD10CM | M46.00 | Spinal enthesopathy, site unspecified |
| ICD10CM | M46.01 | Spinal enthesopathy, occipito-atlanto-axial region |
| ICD10CM | M46.02 | Spinal enthesopathy, cervical region |
| ICD10CM | M46.03 | Spinal enthesopathy, cervicothoracic region |
| ICD10CM | M46.04 | Spinal enthesopathy, thoracic region |
| ICD10CM | M46.05 | Spinal enthesopathy, thoracolumbar region |
| ICD10CM | M46.06 | Spinal enthesopathy, lumbar region |
| ICD10CM | M46.07 | Spinal enthesopathy, lumbosacral region |
| ICD10CM | M46.08 | Spinal enthesopathy, sacral and sacrococcygeal region |
| ICD10CM | M46.09 | Spinal enthesopathy, multiple sites in spine |
| ICD10CM | M46.1 | Sacroiliitis, not elsewhere classified |
| ICD10CM | M46.2 | Osteomyelitis of vertebra |
| ICD10CM | M46.20 | Osteomyelitis of vertebra, site unspecified |
| ICD10CM | M46.21 | Osteomyelitis of vertebra, occipito-atlanto-axial region |
| ICD10CM | M46.22 | Osteomyelitis of vertebra, cervical region |
| ICD10CM | M46.23 | Osteomyelitis of vertebra, cervicothoracic region |
| ICD10CM | M46.24 | Osteomyelitis of vertebra, thoracic region |
| ICD10CM | M46.25 | Osteomyelitis of vertebra, thoracolumbar region |
| ICD10CM | M46.26 | Osteomyelitis of vertebra, lumbar region |
| ICD10CM | M46.27 | Osteomyelitis of vertebra, lumbosacral region |
| ICD10CM | M46.28 | Osteomyelitis of vertebra, sacral and sacrococcygeal region |
| ICD10CM | M46.3 | Infection of intervertebral disc (pyogenic) |
| ICD10CM | M46.30 | Infection of intervertebral disc (pyogenic), site unspecified |
| ICD10CM | M46.31 | Infection of intervertebral disc (pyogenic), occipito-atlanto-axial region |
| ICD10CM | M46.32 | Infection of intervertebral disc (pyogenic), cervical region |
| ICD10CM | M46.33 | Infection of intervertebral disc (pyogenic), cervicothoracic region |
| ICD10CM | M46.34 | Infection of intervertebral disc (pyogenic), thoracic region |
| ICD10CM | M46.35 | Infection of intervertebral disc (pyogenic), thoracolumbar region |
| ICD10CM | M46.36 | Infection of intervertebral disc (pyogenic), lumbar region |
| ICD10CM | M46.37 | Infection of intervertebral disc (pyogenic), lumbosacral region |
| ICD10CM | M46.38 | Infection of intervertebral disc (pyogenic), sacral and sacrococcygeal region |
| ICD10CM | M46.39 | Infection of intervertebral disc (pyogenic), multiple sites in spine |
| ICD10CM | M46.4 | Discitis, unspecified |
| ICD10CM | M46.40 | Discitis, unspecified, site unspecified |
| ICD10CM | M46.41 | Discitis, unspecified, occipito-atlanto-axial region |
| ICD10CM | M46.42 | Discitis, unspecified, cervical region |
| ICD10CM | M46.43 | Discitis, unspecified, cervicothoracic region |

Continued on next page

**TableS31 – continued from previous page**

| CodeSystem | ConceptCode | ConceptName |
|---|---|---|
| ICD10CM | M46.44 | Discitis, unspecified, thoracic region |
| ICD10CM | M46.45 | Discitis, unspecified, thoracolumbar region |
| ICD10CM | M46.46 | Discitis, unspecified, lumbar region |
| ICD10CM | M46.47 | Discitis, unspecified, lumbosacral region |
| ICD10CM | M46.48 | Discitis, unspecified, sacral and sacrococcygeal region |
| ICD10CM | M46.49 | Discitis, unspecified, multiple sites in spine |
| ICD10CM | M46.5 | Other infective spondylopathies |
| ICD10CM | M46.50 | Other infective spondylopathies, site unspecified |
| ICD10CM | M46.51 | Other infective spondylopathies, occipito-atlanto-axial region |
| ICD10CM | M46.52 | Other infective spondylopathies, cervical region |
| ICD10CM | M46.53 | Other infective spondylopathies, cervicothoracic region |
| ICD10CM | M46.54 | Other infective spondylopathies, thoracic region |
| ICD10CM | M46.55 | Other infective spondylopathies, thoracolumbar region |
| ICD10CM | M46.56 | Other infective spondylopathies, lumbar region |
| ICD10CM | M46.57 | Other infective spondylopathies, lumbosacral region |
| ICD10CM | M46.58 | Other infective spondylopathies, sacral and sacrococcygeal region |
| ICD10CM | M46.59 | Other infective spondylopathies, multiple sites in spine |
| ICD10CM | M46.8 | Other specified inflammatory spondylopathies |
| ICD10CM | M46.80 | Other specified inflammatory spondylopathies, site unspecified |
| ICD10CM | M46.81 | Other specified inflammatory spondylopathies, occipito-atlanto-axial region |
| ICD10CM | M46.82 | Other specified inflammatory spondylopathies, cervical region |
| ICD10CM | M46.83 | Other specified inflammatory spondylopathies, cervicothoracic region |
| ICD10CM | M46.84 | Other specified inflammatory spondylopathies, thoracic region |
| ICD10CM | M46.85 | Other specified inflammatory spondylopathies, thoracolumbar region |
| ICD10CM | M46.86 | Other specified inflammatory spondylopathies, lumbar region |
| ICD10CM | M46.87 | Other specified inflammatory spondylopathies, lumbosacral region |
| ICD10CM | M46.88 | Other specified inflammatory spondylopathies, sacral and sacrococcygeal region |
| ICD10CM | M46.89 | Other specified inflammatory spondylopathies, multiple sites in spine |
| ICD10CM | M46.9 | Unspecified inflammatory spondylopathy |
| ICD10CM | M46.90 | Unspecified inflammatory spondylopathy, site unspecified |
| ICD10CM | M46.91 | Unspecified inflammatory spondylopathy, occipito-atlanto-axial region |
| ICD10CM | M46.92 | Unspecified inflammatory spondylopathy, cervical region |
| ICD10CM | M46.93 | Unspecified inflammatory spondylopathy, cervicothoracic region |
| ICD10CM | M46.94 | Unspecified inflammatory spondylopathy, thoracic region |
| ICD10CM | M46.95 | Unspecified inflammatory spondylopathy, thoracolumbar region |
| ICD10CM | M46.96 | Unspecified inflammatory spondylopathy, lumbar region |
| ICD10CM | M46.97 | Unspecified inflammatory spondylopathy, lumbosacral region |
| ICD10CM | M46.98 | Unspecified inflammatory spondylopathy, sacral and sacrococcygeal region |

Continued on next page

**TableS31 – continued from previous page**

| CodeSystem | ConceptCode | ConceptName |
|---|---|---|
| ICD10CM | M46.99 | Unspecified inflammatory spondylopathy, multiple sites in spine |
| ICD10CM | T78.40XA | Allergy, unspecified, initial encounter |
| ICD10CM | T78.40XD | Allergy, unspecified, subsequent encounter |
| ICD10CM | T78.40XS | Allergy, unspecified, sequela |
| ICD10CM | T86.30 | Unspecified complication of heart-lung transplant |
| ICD10CM | T86.3 | Complications of heart-lung transplant |
| ICD10CM | T86.0 | Complications of bone marrow transplant |
| ICD10CM | T86.00 | Unspecified complication of bone marrow transplant |
| ICD10CM | T86.01 | Bone marrow transplant rejection |
| ICD10CM | T86.02 | Bone marrow transplant failure |
| ICD10CM | T86.03 | Bone marrow transplant infection |
| ICD10CM | T86.09 | Other complications of bone marrow transplant |
| ICD10CM | T86.1 | Complications of kidney transplant |
| ICD10CM | T86.10 | Unspecified complication of kidney transplant |
| ICD10CM | T86.11 | Kidney transplant rejection |
| ICD10CM | T86.12 | Kidney transplant failure |
| ICD10CM | T86.13 | Kidney transplant infection |
| ICD10CM | T86.19 | Other complication of kidney transplant |
| ICD10CM | T86.2 | Complications of heart transplant |
| ICD10CM | T86.20 | Unspecified complication of heart transplant |
| ICD10CM | T86.21 | Heart transplant rejection |
| ICD10CM | T86.22 | Heart transplant failure |
| ICD10CM | T86.23 | Heart transplant infection |
| ICD10CM | T86.29 | Other complications of heart transplant |
| ICD10CM | T86.290 | Cardiac allograft vasculopathy |
| ICD10CM | T86.298 | Other complications of heart transplant |
| ICD10CM | T86.31 | Heart-lung transplant rejection |
| ICD10CM | T86.32 | Heart-lung transplant failure |
| ICD10CM | T86.33 | Heart-lung transplant infection |
| ICD10CM | T86.39 | Other complications of heart-lung transplant |
| ICD10CM | T86.4 | Complications of liver transplant |
| ICD10CM | T86.40 | Unspecified complication of liver transplant |
| ICD10CM | T86.41 | Liver transplant rejection |
| ICD10CM | T86.42 | Liver transplant failure |
| ICD10CM | T86.43 | Liver transplant infection |
| ICD10CM | T86.49 | Other complications of liver transplant |
| ICD10CM | T86.5 | Complications of stem cell transplant |
| ICD10CM | T86.8 | Complications of other transplanted organs and tissues |
| ICD10CM | T86.81 | Complications of lung transplant |
| ICD10CM | T86.810 | Lung transplant rejection |
| ICD10CM | T86.811 | Lung transplant failure |
| ICD10CM | T86.812 | Lung transplant infection |
| ICD10CM | T86.818 | Other complications of lung transplant |
| ICD10CM | T86.819 | Unspecified complication of lung transplant |
| ICD10CM | T86.82 | Complications of skin graft (allograft) (autograft) |
| ICD10CM | T86.820 | Skin graft (allograft) rejection |
| ICD10CM | T86.821 | Skin graft (allograft) (autograft) failure |

Continued on next page

**TableS31 – continued from previous page**

| CodeSystem | ConceptCode | ConceptName |
|---|---|---|
| ICD10CM | T86.822 | Skin graft (allograft) (autograft) infection |
| ICD10CM | T86.828 | Other complications of skin graft (allograft) (autograft) |
| ICD10CM | T86.829 | Unspecified complication of skin graft (allograft) (autograft) |
| ICD10CM | T86.83 | Complications of bone graft |
| ICD10CM | T86.830 | Bone graft rejection |
| ICD10CM | T86.831 | Bone graft failure |
| ICD10CM | T86.832 | Bone graft infection |
| ICD10CM | T86.838 | Other complications of bone graft |
| ICD10CM | T86.839 | Unspecified complication of bone graft |
| ICD10CM | T86.84 | Complications of corneal transplant |
| ICD10CM | T86.840 | Corneal transplant rejection |
| ICD10CM | T86.841 | Corneal transplant failure |
| ICD10CM | T86.842 | Corneal transplant infection |
| ICD10CM | T86.848 | Other complications of corneal transplant |
| ICD10CM | T86.849 | Unspecified complication of corneal transplant |
| ICD10CM | T86.85 | Complication of intestine transplant |
| ICD10CM | T86.850 | Intestine transplant rejection |
| ICD10CM | T86.851 | Intestine transplant failure |
| ICD10CM | T86.852 | Intestine transplant infection |
| ICD10CM | T86.858 | Other complications of intestine transplant |
| ICD10CM | T86.859 | Unspecified complication of intestine transplant |
| ICD10CM | T86.89 | Complications of other transplanted tissue |
| ICD10CM | T86.890 | Other transplanted tissue rejection |
| ICD10CM | T86.891 | Other transplanted tissue failure |
| ICD10CM | T86.892 | Other transplanted tissue infection |
| ICD10CM | T86.898 | Other complications of other transplanted tissue |
| ICD10CM | T86.899 | Unspecified complication of other transplanted tissue |
| ICD10CM | T86.9 | Complication of unspecified transplanted organ and tissue |
| ICD10CM | T86.90 | Unspecified complication of unspecified transplanted organ and tissue |
| ICD10CM | T86.91 | Unspecified transplanted organ and tissue rejection |
| ICD10CM | T86.92 | Unspecified transplanted organ and tissue failure |
| ICD10CM | T86.93 | Unspecified transplanted organ and tissue infection |
| ICD10CM | T86.99 | Other complications of unspecified transplanted organ and tissue |
| ICD10CM | Z48.21 | Encounter for aftercare following heart transplant |
| ICD10CM | Z48.22 | Encounter for aftercare following kidney transplant |
| ICD10CM | Z48.23 | Encounter for aftercare following liver transplant |
| ICD10CM | Z48.24 | Encounter for aftercare following lung transplant |
| ICD10CM | Z48.28 | Encounter for aftercare following multiple organ transplant |
| ICD10CM | Z48.280 | Encounter for aftercare following heart-lung transplant |
| ICD10CM | Z48.288 | Encounter for aftercare following multiple organ transplant |
| ICD10CM | Z48.29 | Encounter for aftercare following other organ transplant |
| ICD10CM | Z48.290 | Encounter for aftercare following bone marrow transplant |
| ICD10CM | Z48.298 | Encounter for aftercare following other organ transplant |
| ICD10CM | Z51.11 | Encounter for antineoplastic chemotherapy |
| ICD10CM | Z51.12 | Encounter for antineoplastic immunotherapy |
| ICD10CM | Z94.0 | Kidney transplant status |

Continued on next page

**TableS31 – continued from previous page**

| CodeSystem | ConceptCode | ConceptName |
|---|---|---|
| ICD10CM | Z94.1 | Heart transplant status |
| ICD10CM | Z94.2 | Lung transplant status |
| ICD10CM | Z94.3 | Heart and lungs transplant status |
| ICD10CM | Z94.4 | Liver transplant status |
| ICD10CM | Z94.5 | Skin transplant status |
| ICD10CM | Z94.6 | Bone transplant status |
| ICD10CM | Z94.7 | Corneal transplant status |
| ICD10CM | Z94.8 | Other transplanted organ and tissue status |
| ICD10CM | Z94.81 | Bone marrow transplant status |
| ICD10CM | Z94.82 | Intestine transplant status |
| ICD10CM | Z94.83 | Pancreas transplant status |
| ICD10CM | Z94.84 | Stem cells transplant status |
| ICD10CM | Z94.89 | Other transplanted organ and tissue status |
| ICD10CM | Z94.9 | Transplanted organ and tissue status, unspecified |
| ICD10CM | D84.81 | Immunodeficiency due to conditions classified elsewhere |
| ICD10CM | D84.82 | Immunodeficiency due to drugs and external causes |
| ICD10CM | D84.821 | Immunodeficiency due to drugs |
| ICD10CM | D84.822 | Immunodeficiency due to external causes |
| ICD10CM | D84.89 | Other immunodeficiencies |
| ICD10CM | D89.83 | Cytokine release syndrome |
| ICD10CM | D89.831 | Cytokine release syndrome, grade 1 |
| ICD10CM | D89.832 | Cytokine release syndrome, grade 2 |
| ICD10CM | D89.833 | Cytokine release syndrome, grade 3 |
| ICD10CM | D89.834 | Cytokine release syndrome, grade 4 |
| ICD10CM | D89.835 | Cytokine release syndrome, grade 5 |
| ICD10CM | D89.839 | Cytokine release syndrome, grade unspecified |
| ICD10CM | M05.7A | Rheumatoid arthritis with rheumatoid factor of other specified site without organ or systems involvement |
| ICD10CM | M05.8A | Other rheumatoid arthritis with rheumatoid factor of other specified site |
| ICD10CM | M06.0A | Rheumatoid arthritis without rheumatoid factor, other specified site |
| ICD10CM | M06.8A | Other specified rheumatoid arthritis, other specified site |
| ICD10CM | M08.0A | Unspecified juvenile rheumatoid arthritis, other specified site |
| ICD10CM | M08.2A | Juvenile rheumatoid arthritis with systemic onset, other specified site |
| ICD10CM | M08.4A | Pauciarticular juvenile rheumatoid arthritis, other specified site |
| ICD10CM | M08.9A | Juvenile arthritis, unspecified, other specified site |
| ICD10CM | M35.81 | Multisystem inflammatory syndrome (MIS) |
| ICD10CM | M35.89 | Other specified systemic involvement of connective tissue |
| ICD10CM | T86.8401 | Corneal transplant rejection, right eye |
| ICD10CM | T86.8402 | Corneal transplant rejection, left eye |
| ICD10CM | T86.8403 | Corneal transplant rejection, bilateral |
| ICD10CM | T86.8409 | Corneal transplant rejection, unspecified eye |
| ICD10CM | T86.8411 | Corneal transplant failure, right eye |
| ICD10CM | T86.8412 | Corneal transplant failure, left eye |
| ICD10CM | T86.8413 | Corneal transplant failure, bilateral |

Continued on next page

**TableS31 – continued from previous page**

| CodeSystem | ConceptCode | ConceptName |
|---|---|---|
| ICD10CM | T86.8419 | Corneal transplant failure, unspecified eye |
| ICD10CM | T86.8421 | Corneal transplant infection, right eye |
| ICD10CM | T86.8422 | Corneal transplant infection, left eye |
| ICD10CM | T86.8423 | Corneal transplant infection, bilateral |
| ICD10CM | T86.8429 | Corneal transplant infection, unspecified eye |
| ICD10CM | T86.8481 | Other complications of corneal transplant, right eye |
| ICD10CM | T86.8482 | Other complications of corneal transplant, left eye |
| ICD10CM | T86.8483 | Other complications of corneal transplant, bilateral |
| ICD10CM | T86.8489 | Other complications of corneal transplant, unspecified eye |
| ICD10CM | T86.8491 | Unspecified complication of corneal transplant, right eye |
| ICD10CM | T86.8492 | Unspecified complication of corneal transplant, left eye |
| ICD10CM | T86.8493 | Unspecified complication of corneal transplant, bilateral |
| ICD10CM | T86.8499 | Unspecified complication of corneal transplant, unspecified eye |
| ICD10CM | C79.63 | Secondary malignant neoplasm of bilateral ovaries |
| ICD10CM | D89.44 | Hereditary alpha tryptasemia |
| ICD10CM | C84.7A | Anaplastic large cell lymphoma, ALK-negative, breast |
| ICD10CM | C56.3 | Malignant neoplasm of bilateral ovaries |
| SNOMED CT | 263661007 | Complement 5 dysfunction |
| SNOMED CT | 267459007 | Deficiencies of humoral immunity |
| SNOMED CT | 267460002 | Congenital hypogammaglobulinemia |
| SNOMED CT | 267524009 | Constitutional aplastic anemia with malformation |
| SNOMED CT | 267527002 | Aplastic anemia due to drugs |
| SNOMED CT | 267540007 | Neutropenia due to irradiation |
| SNOMED CT | 267543009 | Functional disorders of polymorphonuclear neutrophils |
| SNOMED CT | 269295009 | Transplanted organ failure |
| SNOMED CT | 269475001 | Malignant tumor of lymphoid hemopoietic and related tissue |
| SNOMED CT | 269476000 | Nodular lymphoma |
| SNOMED CT | 274905008 | Malignant lymphoma - lymphocytic, intermediate differentiation |
| SNOMED CT | 275523003 | Pancytopenia-dysmelia |
| SNOMED CT | 275524009 | Immunoproliferative neoplasm |
| SNOMED CT | 276628009 | Chloramphenicol-induced neutropenia |
| SNOMED CT | 276665006 | Congenital acquired immune deficiency syndrome |
| SNOMED CT | 276666007 | Congenital human immunodeficiency virus positive status syndrome |
| SNOMED CT | 276811008 | Gastric lymphoma |
| SNOMED CT | 276815004 | Lymphoma of intestine |
| SNOMED CT | 276836002 | Primary cerebral lymphoma |
| SNOMED CT | 277473004 | B-cell chronic lymphocytic leukemia |
| SNOMED CT | 277474005 | B-cell chronic lymphocytic leukemia variant |
| SNOMED CT | 277543005 | Malignant white blood cell disorder |
| SNOMED CT | 277545003 | T-cell chronic lymphocytic leukemia |
| SNOMED CT | 277549009 | Chronic lymphocytic prolymphocytic leukemia syndrome |
| SNOMED CT | 277551008 | Splenic lymphoma with villous lymphocytes |
| SNOMED CT | 277567002 | T-cell prolymphocytic leukemia |
| SNOMED CT | 277568007 | Hairy cell leukemia variant |
| SNOMED CT | 277569004 | Large granular lymphocytic leukemia |
| SNOMED CT | 277570003 | Lymphoma with spill |

Continued on next page

**TableS31 – continued from previous page**

| CodeSystem | ConceptCode | ConceptName |
|---|---|---|
| SNOMED CT | 277571004 | B-cell acute lymphoblastic leukemia |
| SNOMED CT | 277572006 | Precursor B-cell acute lymphoblastic leukemia |
| SNOMED CT | 277573001 | Common acute lymphoblastic leukemia |
| SNOMED CT | 277574007 | Null cell acute lymphoblastic leukemia |
| SNOMED CT | 277575008 | T-cell acute lymphoblastic leukemia |
| SNOMED CT | 277579002 | Light chain myeloma |
| SNOMED CT | 277580004 | Non-secretory myeloma |
| SNOMED CT | 277587001 | Juvenile chronic myeloid leukemia |
| SNOMED CT | 277589003 | Atypical chronic myeloid leukemia |
| SNOMED CT | 277597005 | Myelodysplastic syndrome with isolated del(5q) |
| SNOMED CT | 277601005 | Acute monoblastic leukemia |
| SNOMED CT | 277602003 | Acute megakaryoblastic leukemia |
| SNOMED CT | 277604002 | Acute eosinophilic leukemia |
| SNOMED CT | 277606000 | Lymphoreticular tumor |
| SNOMED CT | 277613000 | Cutaneous/peripheral T-cell lymphoma |
| SNOMED CT | 277614006 | Prethymic and thymic T-cell lymphoma/leukemia |
| SNOMED CT | 277615007 | Low grade B-cell lymphoma |
| SNOMED CT | 277616008 | Diffuse low grade B-cell lymphoma |
| SNOMED CT | 277617004 | High grade B-cell lymphoma |
| SNOMED CT | 277618009 | Follicular low grade B-cell lymphoma |
| SNOMED CT | 277619001 | B-cell prolymphocytic leukemia |
| SNOMED CT | 277622004 | Mucosa-associated lymphoma |
| SNOMED CT | 277623009 | Monocytoid B-cell lymphoma |
| SNOMED CT | 277624003 | Follicular malignant lymphoma - mixed cell type |
| SNOMED CT | 277625002 | Follicular malignant lymphoma - small cleaved cell |
| SNOMED CT | 277626001 | Diffuse high grade B-cell lymphoma |
| SNOMED CT | 277627005 | Nodular high grade B-cell lymphoma |
| SNOMED CT | 277628000 | Diffuse malignant lymphoma - large cleaved cell |
| SNOMED CT | 277629008 | Diffuse malignant lymphoma - large non-cleaved cell |
| SNOMED CT | 277632006 | Diffuse malignant lymphoma - centroblastic polymorphic |
| SNOMED CT | 277637000 | Large cell anaplastic lymphoma |
| SNOMED CT | 277641001 | Follicular malignant lymphoma - large cell |
| SNOMED CT | 277642008 | Low grade T-cell lymphoma |
| SNOMED CT | 277643003 | High grade T-cell lymphoma |
| SNOMED CT | 277651000 | Peripheral T-cell lymphoma - pleomorphic small cell |
| SNOMED CT | 277653002 | Peripheral T-cell lymphoma - pleomorphic medium and large cell |
| SNOMED CT | 277664004 | Malignant lymphoma of testis |
| SNOMED CT | 278051002 | Malignant lymphoma of thyroid gland |
| SNOMED CT | 278052009 | Malignant lymphoma of breast |
| SNOMED CT | 278189009 | Hypergranular promyelocytic leukemia |
| SNOMED CT | 278256002 | Cord cell transfusion |
| SNOMED CT | 278453007 | Acute biphenotypic leukemia |
| SNOMED CT | 281388009 | HIV-related sclerosing cholangitis |
| SNOMED CT | 285769009 | Acute promyelocytic leukemia - hypogranular variant |
| SNOMED CT | 285776004 | Intermediate grade B-cell lymphoma |
| SNOMED CT | 285839005 | Acute myelomonocytic leukemia - eosinophilic variant |
| SNOMED CT | 287006005 | Rheumatoid arthritis of multiple joints |

Continued on next page

**TableS31 – continued from previous page**

| CodeSystem | ConceptCode | ConceptName |
|---|---|---|
| SNOMED CT | 287007001 | Rheumatoid arthritis - hand joint |
| SNOMED CT | 287008006 | Rheumatoid arthritis - ankle and/or foot |
| SNOMED CT | 288327009 | Granulation anomaly |
| SNOMED CT | 289317009 | Granulocyte granule deficiency |
| SNOMED CT | 302841002 | Malignant lymphoma - small lymphocytic |
| SNOMED CT | 302842009 | Diffuse malignant lymphoma - centroblastic |
| SNOMED CT | 302845006 | Nodular malignant lymphoma, lymphocytic - well differentiated |
| SNOMED CT | 302848008 | Nodular malignant lymphoma, lymphocytic - intermediate differentiation |
| SNOMED CT | 302855005 | Subacute leukemia |
| SNOMED CT | 302856006 | Aleukemic leukemia |
| SNOMED CT | 302874002 | Phagocytic cell dysfunction |
| SNOMED CT | 303017006 | Malignant lymphoma, convoluted cell type |
| SNOMED CT | 303055001 | Malignant lymphoma, follicular center cell |
| SNOMED CT | 303056000 | Malignant lymphoma, follicular center cell, cleaved |
| SNOMED CT | 303057009 | Malignant lymphoma, follicular center cell, non-cleaved |
| SNOMED CT | 304576008 | Metaphyseal chondrodysplasia, McKusick type with associated immunodeficiency |
| SNOMED CT | 306058006 | Aregenerative anemia |
| SNOMED CT | 307340003 | Monosomy 7 syndrome |
| SNOMED CT | 307341004 | Atypical hairy cell leukemia |
| SNOMED CT | 307592006 | Basophilic leukemia |
| SNOMED CT | 307617006 | Neutrophilic leukemia |
| SNOMED CT | 307622006 | Prolymphocytic lymphosarcoma |
| SNOMED CT | 307623001 | Malignant lymphoma - lymphoplasmacytic |
| SNOMED CT | 307624007 | Diffuse malignant lymphoma - centroblastic-centrocytic |
| SNOMED CT | 307625008 | Malignant lymphoma - centrocytic |
| SNOMED CT | 307636001 | Malignant lymphoma, mixed lymphocytic-histiocytic, nodular |
| SNOMED CT | 307637005 | Malignant lymphoma, centroblastic-centrocytic, follicular |
| SNOMED CT | 307646004 | Malignant lymphoma, lymphocytic, poorly differentiated, nodular |
| SNOMED CT | 307647008 | Malignant lymphoma, centroblastic type, follicular |
| SNOMED CT | 307649006 | Primary central nervous system lymphoma |
| SNOMED CT | 307650006 | Histiocytic medullary reticulosis |
| SNOMED CT | 307651005 | Myelosclerosis with myeloid metaplasia |
| SNOMED CT | 308143008 | Seropositive erosive rheumatoid arthritis |
| SNOMED CT | 309762007 | Systemic lupus erythematosus with pericarditis |
| SNOMED CT | 313427003 | Lambda light chain myeloma |
| SNOMED CT | 314922006 | B-cell lymphoma morphology |
| SNOMED CT | 314923001 | Low grade B-cell lymphoma morphology |
| SNOMED CT | 314924007 | Follicular low grade B-cell lymphoma morphology |
| SNOMED CT | 314925008 | High grade B-cell lymphoma morphology |
| SNOMED CT | 314926009 | T-cell lymphoma morphology |
| SNOMED CT | 314927000 | High grade T-cell lymphoma morphology |
| SNOMED CT | 314929002 | Diffuse low grade B-cell lymphoma morphology |
| SNOMED CT | 314930007 | Low grade T-cell lymphoma morphology |
| SNOMED CT | 314931006 | Nodular high grade B-cell lymphoma morphology |

Continued on next page

**TableS31 – continued from previous page**

| CodeSystem | ConceptCode | ConceptName |
|---|---|---|
| SNOMED CT | 314934003 | Diffuse high grade B-cell lymphoma morphology |
| SNOMED CT | 317226009 | Granulocytosis |
| SNOMED CT | 319171004 | Qualitative abnormality of granulocyte |
| SNOMED CT | 351287008 | Reticular dysgenesis with congenital aleukocytosis |
| SNOMED CT | 363009005 | Complement component deficiency |
| SNOMED CT | 363040003 | Congenital immunodeficiency involving the hematopoietic system |
| SNOMED CT | 363138005 | Hereditary disorder of immune system |
| SNOMED CT | 363153002 | Immune system transplantation |
| SNOMED CT | 363265005 | Predominant humoral immune deficiency |
| SNOMED CT | 370388006 | Patient immunocompromised |
| SNOMED CT | 370391006 | Patient immunosuppressed |
| SNOMED CT | 371012000 | Acute lymphoblastic leukemia, transitional pre-B-cell |
| SNOMED CT | 371134001 | Malignant lymphoma, large cell, polymorphous, immunoblastic |
| SNOMED CT | 373168002 | Reticulosarcoma |
| SNOMED CT | 373604002 | Light chain deposition disease |
| SNOMED CT | 387759001 | Chronic granulomatous disease |
| SNOMED CT | 393573009 | Hypereosinophilic syndrome |
| SNOMED CT | 395692003 | Benign paraproteinemia |
| SNOMED CT | 397007003 | Mast cell disorder |
| SNOMED CT | 397008008 | Aggressive lymphadenopathic mastocytosis with eosinophilia |
| SNOMED CT | 397009000 | Mast cell malignancy |
| SNOMED CT | 397011009 | Mast cell malignancy of lymph nodes |
| SNOMED CT | 397015000 | Systemic mastocytosis with associated clonal hematological non-mast cell lineage disease |
| SNOMED CT | 397016004 | Systemic mast cell disease |
| SNOMED CT | 397338009 | Myelodysplastic syndrome with excess blasts-1 |
| SNOMED CT | 397339001 | Myelodysplastic syndrome with excess blasts-2 |
| SNOMED CT | 397340004 | Acute myeloid leukemia with recurrent genetic abnormality |
| SNOMED CT | 397341000 | Acute myeloid leukemia with multilineage dysplasia following a myelodysplastic syndrome or myelodysplastic syndrome/myeloproliferative disorder |
| SNOMED CT | 397342007 | Acute myeloid leukemia with multilineage dysplasia without antecedent myelodysplastic syndrome |
| SNOMED CT | 397343002 | Therapy-related acute myeloid leukemia and myelodysplastic syndrome, alkylating agent-related type |
| SNOMED CT | 397344008 | Therapy-related acute myeloid leukemia and myelodysplastic syndrome, topoisomerase type II inhibitor-related type |
| SNOMED CT | 397345009 | Acute leukemia of ambiguous lineage |
| SNOMED CT | 397349003 | Nodal marginal zone B-cell lymphoma |
| SNOMED CT | 397350003 | Extranodal marginal zone B-cell lymphoma of mucosa-associated lymphoid tissue |
| SNOMED CT | 397352006 | Primary cutaneous anaplastic large cell lymphoma |
| SNOMED CT | 397355008 | Dendritic cell sarcoma, no International Classification of Diseases for Oncology subtype |
| SNOMED CT | 397400006 | Burkitt lymphoma/leukemia |
| SNOMED CT | 397467006 | Follicular lymphoma, cutaneous follicle center sub-type |

Continued on next page

**TableS31 – continued from previous page**

| CodeSystem | ConceptCode | ConceptName |
|---|---|---|
| SNOMED CT | 397468001 | Follicular lymphoma, diffuse follicle center sub-type, grade 1 |
| SNOMED CT | 397469009 | Follicular lymphoma, diffuse follicle center cell sub-type, grade 2 |
| SNOMED CT | 397763006 | Human immunodeficiency virus encephalopathy |
| SNOMED CT | 398055000 | T-lymphocyte deficiency |
| SNOMED CT | 398250003 | Familial hemophagocytic lymphohistiocytosis |
| SNOMED CT | 398271008 | Predominantly T-cell defect |
| SNOMED CT | 398293003 | Cellular immune defect |
| SNOMED CT | 398329009 | Human immunodeficiency virus encephalitis |
| SNOMED CT | 398623004 | Myelodysplastic syndrome with excess blasts |
| SNOMED CT | 398640008 | Rheumatoid arthritis with pneumoconiosis |
| SNOMED CT | 399648005 | Intravascular B-cell lymphoma |
| SNOMED CT | 399923009 | Rheumatoid arteritis |
| SNOMED CT | 400001003 | Primary cutaneous lymphoma |
| SNOMED CT | 400122007 | Primary cutaneous T-cell lymphoma |
| SNOMED CT | 402355000 | Acute graft-versus-host disease |
| SNOMED CT | 402356004 | Chronic graft-versus-host disease |
| SNOMED CT | 402453006 | Amyloid light chain amyloidosis due to multiple myeloma |
| SNOMED CT | 402454000 | Primary systemic amyloidosis associated with occult plasma cell dyscrasia |
| SNOMED CT | 402483002 | Immunodeficiency with multicarboxylase deficiency |
| SNOMED CT | 402791005 | B-lymphocyte immunodeficiency |
| SNOMED CT | 402792003 | T-lymphocyte immunodeficiency |
| SNOMED CT | 402793008 | Heritable disorder of neutrophil production |
| SNOMED CT | 402794002 | Heritable disorder of neutrophil function |
| SNOMED CT | 402879006 | T-cell leukemic infiltration of skin |
| SNOMED CT | 402880009 | Primary cutaneous large T-cell lymphoma |
| SNOMED CT | 402882001 | Hodgkin's disease affecting skin |
| SNOMED CT | 403486000 | Acute systemic lupus erythematosus |
| SNOMED CT | 403487009 | Fulminating systemic lupus erythematosus |
| SNOMED CT | 403488004 | Systemic lupus erythematosus of childhood |
| SNOMED CT | 403836001 | Autosomal recessive hyperimmunoglobulin M syndrome |
| SNOMED CT | 403837005 | Wiskott-Aldrich autosomal dominant variant syndrome |
| SNOMED CT | 403838000 | Jung syndrome |
| SNOMED CT | 403839008 | Siccardi syndrome |
| SNOMED CT | 404107008 | Patch/plaque stage mycosis fungoides |
| SNOMED CT | 404108003 | Poikilodermatous mycosis fungoides |
| SNOMED CT | 404109006 | Follicular mucinosis type mycosis fungoides |
| SNOMED CT | 404110001 | Hypomelanotic mycosis fungoides |
| SNOMED CT | 404111002 | Lymphomatoid papulosis-associated mycosis fungoides |
| SNOMED CT | 404112009 | Granulomatous mycosis fungoides |
| SNOMED CT | 404113004 | Tumor stage mycosis fungoides |
| SNOMED CT | 404114005 | Erythrodermic mycosis fungoides |
| SNOMED CT | 404115006 | Bullous mycosis fungoides |
| SNOMED CT | 404116007 | Mycosis fungoides with systemic infiltration |
| SNOMED CT | 404117003 | Spongiotic mycosis fungoides |
| SNOMED CT | 404118008 | Syringotropic mycosis fungoides |
| SNOMED CT | 404119000 | Pagetoid reticulosis |

Continued on next page

**TableS31 – continued from previous page**

| CodeSystem | ConceptCode | ConceptName |
|---|---|---|
| SNOMED CT | 404120006 | Localized pagetoid reticulosis |
| SNOMED CT | 404121005 | Generalized pagetoid reticulosis |
| SNOMED CT | 404126000 | CD-30 positive pleomorphic large T-cell cutaneous lymphoma |
| SNOMED CT | 404127009 | CD-30 positive T-immunoblastic cutaneous lymphoma |
| SNOMED CT | 404128004 | CD-30 negative cutaneous T-cell lymphoma |
| SNOMED CT | 404129007 | CD-30 negative anaplastic large T-cell cutaneous lymphoma |
| SNOMED CT | 404130002 | CD-30 negative pleomorphic large T-cell cutaneous lymphoma |
| SNOMED CT | 404131003 | CD-30 negative T-immunoblastic cutaneous lymphoma |
| SNOMED CT | 404133000 | Subcutaneous panniculitis-like T-cell lymphoma |
| SNOMED CT | 404134006 | Anaplastic large T-cell systemic malignant lymphoma |
| SNOMED CT | 404137004 | Precursor B-cell lymphoblastic lymphoma involving skin |
| SNOMED CT | 404138009 | Small lymphocytic B-cell lymphoma involving skin |
| SNOMED CT | 404139001 | Leukemic infiltration of skin in hairy-cell leukemia |
| SNOMED CT | 404140004 | Primary cutaneous marginal zone B-cell lymphoma |
| SNOMED CT | 404141000 | Primary cutaneous immunocytoma |
| SNOMED CT | 404142007 | Primary cutaneous plasmacytoma |
| SNOMED CT | 404144008 | Primary cutaneous diffuse large cell B-cell lymphoma |
| SNOMED CT | 404147001 | Follicular center B-cell lymphoma (nodal/systemic with skin involvement) |
| SNOMED CT | 404148006 | Diffuse large B-cell lymphoma (nodal/systemic with skin involvement) |
| SNOMED CT | 404149003 | Lymphoplasmacytic B-cell lymphoma, nodal/systemic with skin involvement |
| SNOMED CT | 404150003 | Mantle cell B-cell lymphoma (nodal/systemic with skin involvement) |
| SNOMED CT | 404151004 | Leukemic infiltration of skin in myeloid leukemia |
| SNOMED CT | 404152006 | Leukemic infiltration of skin in acute myeloid leukemia |
| SNOMED CT | 404153001 | Leukemic infiltration of skin in chronic myeloid leukemia |
| SNOMED CT | 404154007 | Leukemic infiltration of skin in monocytic leukemia |
| SNOMED CT | 404155008 | Granulocytic sarcoma affecting skin |
| SNOMED CT | 404156009 | Leukemic infiltration of skin |
| SNOMED CT | 404160007 | Langerhans cell histiocytosis - Hashimoto-Pritzker type |
| SNOMED CT | 404169008 | Malignant histiocytosis involving skin |
| SNOMED CT | 404171008 | Mastocytoma |
| SNOMED CT | 404172001 | Mast cell leukemia affecting skin |
| SNOMED CT | 405631006 | Pediatric human immunodeficiency virus infection |
| SNOMED CT | 406565005 | CD4 T lymphocyte deficiency |
| SNOMED CT | 413389003 | Accelerated phase chronic myeloid leukemia |
| SNOMED CT | 413440007 | Acute lymphoblastic leukemia - category |
| SNOMED CT | 413441006 | Acute monocytic leukemia |
| SNOMED CT | 413442004 | Acute monocytic/monoblastic leukemia |
| SNOMED CT | 413443009 | Acute myeloid leukemia |
| SNOMED CT | 413537009 | Angioimmunoblastic T-cell lymphoma |
| SNOMED CT | 413565006 | Aplastic anemia associated with metabolic alteration |
| SNOMED CT | 413566007 | Aplastic anemia associated with pancreatitis |
| SNOMED CT | 413567003 | Aplastic anemia associated with pregnancy |
| SNOMED CT | 413587002 | Smoldering myeloma |

Continued on next page

**TableS31 – continued from previous page**

| CodeSystem | ConceptCode | ConceptName |
|---|---|---|
| SNOMED CT | 413656006 | Blastic phase chronic myeloid leukemia |
| SNOMED CT | 413834006 | Chronic disease of immune function |
| SNOMED CT | 413836008 | Chronic eosinophilic leukemia |
| SNOMED CT | 413840004 | Chronic lymphoid leukemia - category |
| SNOMED CT | 413841000 | Chronic myeloid leukemia - category |
| SNOMED CT | 413842007 | Chronic myeloid leukemia in lymphoid blast crisis |
| SNOMED CT | 413843002 | Chronic myeloid leukemia in myeloid blast crisis |
| SNOMED CT | 413847001 | Chronic phase chronic myeloid leukemia |
| SNOMED CT | 413990004 | Diffuse large B-cell lymphoma - category |
| SNOMED CT | 414029004 | Disorder of immune function |
| SNOMED CT | 414031008 | Immunosecretory disorder |
| SNOMED CT | 414395005 | Hereditary white blood cell disorder |
| SNOMED CT | 414553000 | Kappa light chain myeloma |
| SNOMED CT | 414644002 | Malignant hematopoietic neoplasm |
| SNOMED CT | 414645001 | Malignant histiocytic neoplasm |
| SNOMED CT | 414653009 | Mast cell neoplasm |
| SNOMED CT | 414785000 | Multiple solitary plasmacytomas |
| SNOMED CT | 414825006 | Neoplasm of hematopoietic cell type |
| SNOMED CT | 414927004 | Ocular myasthenia with strabismus |
| SNOMED CT | 415005004 | Panleukopenia |
| SNOMED CT | 415110002 | Plasma cell myeloma/plasmacytoma |
| SNOMED CT | 415111003 | Plasma cell neoplasm |
| SNOMED CT | 415112005 | Plasmacytoma |
| SNOMED CT | 415113000 | Plasmacytoma - category |
| SNOMED CT | 415283002 | Myelodysplastic syndrome with excess blasts-1 |
| SNOMED CT | 415284008 | Myelodysplastic syndrome with excess blasts-2 |
| SNOMED CT | 415285009 | Myelodysplastic syndrome with multilineage dysplasia |
| SNOMED CT | 415286005 | Myelodysplastic syndrome with ring sideroblasts and multilineage dysplasia |
| SNOMED CT | 415287001 | Relapsing chronic myeloid leukemia |
| SNOMED CT | 417152008 | Langerhans cell histiocytosis of lung |
| SNOMED CT | 417167007 | Immunoglobulin deficiency |
| SNOMED CT | 417672002 | Granulocytopenic disorder |
| SNOMED CT | 418265009 | Primary cutaneous B-cell lymphoma - category |
| SNOMED CT | 418628003 | Follicular mycosis fungoides |
| SNOMED CT | 418789003 | Primary cutaneous plasmacytoma |
| SNOMED CT | 419018000 | Primary cutaneous large T-cell lymphoma - category |
| SNOMED CT | 419283005 | Primary cutaneous T-cell lymphoma - category |
| SNOMED CT | 419386004 | Pagetoid reticulosis |
| SNOMED CT | 419392005 | Primary cutaneous lymphoma |
| SNOMED CT | 419586003 | Primary cutaneous T-cell lymphoma, large cell, CD30-negative |
| SNOMED CT | 419662008 | Primary cutaneous follicle center cell lymphoma |
| SNOMED CT | 420028002 | Primary cutaneous marginal zone B-cell lymphoma |
| SNOMED CT | 420384005 | Hematopoietic system disease with AIDS (acquired immunodeficiency syndrome) |
| SNOMED CT | 420395004 | Acute endocarditis with AIDS (acquired immunodeficiency syndrome) |

Continued on next page

**TableS31 – continued from previous page**

| CodeSystem | ConceptCode | ConceptName |
|---|---|---|
| SNOMED CT | 420519005 | Malignant lymphoma of the eye region |
| SNOMED CT | 420524008 | Kaposi's sarcoma with AIDS (acquired immunodeficiency syndrome) |
| SNOMED CT | 420788006 | Primary intraocular non-Hodgkin malignant lymphoma |
| SNOMED CT | 420890002 | Precursor T cell lymphoblastic leukemia/lymphoblastic lymphoma |
| SNOMED CT | 421020000 | Microsporidiosis associated with acquired immunodeficiency syndrome |
| SNOMED CT | 421077004 | Disseminated candidiasis with AIDS (acquired immunodeficiency syndrome) |
| SNOMED CT | 421246008 | Precursor T-cell lymphoblastic lymphoma |
| SNOMED CT | 421696004 | Precursor T-cell neoplasm |
| SNOMED CT | 422127002 | Herpes zoster with AIDS (acquired immunodeficiency syndrome) |
| SNOMED CT | 422172005 | T-cell AND/OR NK-cell neoplasm |
| SNOMED CT | 422853008 | Lymphoma of retroperitoneal space |
| SNOMED CT | 425333006 | Myeloproliferative disorder |
| SNOMED CT | 425563003 | Grafting of cord blood to bone marrow |
| SNOMED CT | 425657001 | Osteosclerotic myeloma |
| SNOMED CT | 425688002 | Philadelphia chromosome-positive acute lymphoblastic leukemia |
| SNOMED CT | 425749006 | Subacute myeloid leukemia in remission |
| SNOMED CT | 425835006 | Disorder of immune reconstitution |
| SNOMED CT | 425843001 | Allogeneic peripheral blood stem cell transplant |
| SNOMED CT | 425869007 | Acute promyelocytic leukemia, FAB M3, in remission |
| SNOMED CT | 425941003 | Precursor B-cell acute lymphoblastic leukemia in remission |
| SNOMED CT | 425983008 | Autologous peripheral blood stem cell transplant |
| SNOMED CT | 426071002 | Hodgkin's disease in remission |
| SNOMED CT | 426124006 | Acute myeloid leukemia with maturation, FAB M2, in remission |
| SNOMED CT | 426202004 | Immune reconstitution syndrome |
| SNOMED CT | 426217000 | Aleukaemic leukaemia in remission |
| SNOMED CT | 426248008 | Aleukemic lymphoid leukemia in remission |
| SNOMED CT | 426336007 | Solitary osseous myeloma |
| SNOMED CT | 426370008 | Subacute lymphoid leukemia in remission |
| SNOMED CT | 426425001 | Allograft of bone marrow from sibling donor |
| SNOMED CT | 426598005 | Amyloid light-chain nephropathy |
| SNOMED CT | 426642002 | Erythroleukemia, FAB M6 in remission |
| SNOMED CT | 426768001 | Engraftment syndrome |
| SNOMED CT | 426800001 | Febrile granulocytopenia |
| SNOMED CT | 426885008 | Hodgkin's disease, lymphocytic depletion of lymph nodes of head |
| SNOMED CT | 426955004 | Philadelphia chromosome-positive acute lymphoblastic leukemia |
| SNOMED CT | 427056005 | Subacute leukemia in remission |
| SNOMED CT | 427141003 | Malignant lymphoma in remission |
| SNOMED CT | 427374007 | Immunoproliferative neoplasm in remission |
| SNOMED CT | 427423003 | Allograft of bone marrow from matched unrelated donor |
| SNOMED CT | 427615008 | Allograft of cord blood to bone marrow |
| SNOMED CT | 427642009 | T-cell acute lymphoblastic leukemia in remission |
| SNOMED CT | 427658007 | Acute myelomonocytic leukemia, FAB M4, in remission |
| SNOMED CT | 427770001 | Rheumatoid arthritis of temporomandibular joint |

Continued on next page

**TableS31 – continued from previous page**

| CodeSystem | ConceptCode | ConceptName |
|---|---|---|
| SNOMED CT | 428103008 | Disorder of transplanted bone marrow |
| SNOMED CT | 429054002 | Disorder related to transplantation |
| SNOMED CT | 429192004 | Rheumatoid arthritis of foot |
| SNOMED CT | 429362002 | Short stature associated with bone marrow transplant |
| SNOMED CT | 429450002 | Disorder related to bone marrow transplantation |
| SNOMED CT | 429490004 | Disorder affecting transplanted structure |
| SNOMED CT | 429514007 | Growth hormone deficiency after bone marrow transplant |
| SNOMED CT | 430338009 | Smoldering chronic lymphocytic leukemia |
| SNOMED CT | 430478003 | Macrophage activation syndrome |
| SNOMED CT | 431456004 | Accelerated rejection of bone marrow transplant |
| SNOMED CT | 431506006 | Rejection of pancreas transplant |
| SNOMED CT | 431862001 | Acute rejection of bone marrow transplant |
| SNOMED CT | 431953002 | Chronic rejection of bone marrow transplant |
| SNOMED CT | 432782005 | Hyperacute rejection of bone marrow transplant |
| SNOMED CT | 433592008 | Acute rejection of pancreas transplant |
| SNOMED CT | 433809002 | Accelerated rejection of pancreas transplant |
| SNOMED CT | 434202008 | Hyperacute rejection of pancreas transplant |
| SNOMED CT | 434271002 | Chronic rejection of pancreas transplant |
| SNOMED CT | 440422002 | Asymptomatic multiple myeloma |
| SNOMED CT | 441313008 | Indolent multiple myeloma |
| SNOMED CT | 441559006 | Mantle cell lymphoma of spleen |
| SNOMED CT | 441962003 | Large cell lymphoma of intrapelvic lymph nodes |
| SNOMED CT | 442134007 | Hepatitis B associated with Human immunodeficiency virus infection |
| SNOMED CT | 442459007 | Combined immunodeficiency disease |
| SNOMED CT | 442537007 | Non-Hodgkin lymphoma associated with Human immunodeficiency virus infection |
| SNOMED CT | 442557006 | Grafting of bone marrow using allograft from unmatched unrelated donor |
| SNOMED CT | 443487006 | Mantle cell lymphoma |
| SNOMED CT | 444597005 | Mucosa-associated lymphoid tissue (MALT) lymphoma of stomach |
| SNOMED CT | 444910004 | Primary mediastinal (thymic) large B-cell lymphoma |
| SNOMED CT | 444911000 | Acute myeloid leukemia with t(9:11)(p22;q23); MLLT3-MLL |
| SNOMED CT | 445105005 | Blastic plasmacytoid dendritic cell neoplasm |
| SNOMED CT | 445227008 | Juvenile myelomonocytic leukemia |
| SNOMED CT | 445406001 | Hepatosplenic T-cell lymphoma |
| SNOMED CT | 445448008 | Acute myeloid leukemia with myelodysplasia-related changes |
| SNOMED CT | 445738007 | Myelodysplastic/myeloproliferative disease |
| SNOMED CT | 445757003 | Allogeneic bone marrow transplantation without purging |
| SNOMED CT | 445925008 | Lymphomatous infiltration |
| SNOMED CT | 445945000 | Infectious disease with acquired immune deficiency syndrome |
| SNOMED CT | 445964006 | Monomorphic posttransplant lymphoproliferative disorder |
| SNOMED CT | 446253009 | Allogeneic bone marrow transplantation with purging |
| SNOMED CT | 446643000 | Sarcoma of dendritic cells (accessory cells) |
| SNOMED CT | 446688004 | Leukemic infiltration |
| SNOMED CT | 447100004 | Marginal zone lymphoma |

Continued on next page

**TableS31 – continued from previous page**

| CodeSystem | ConceptCode | ConceptName |
|---|---|---|
| SNOMED CT | 447596005 | Myelodysplastic/myeloproliferative neoplasm, unclassifiable |
| SNOMED CT | 447656001 | Lymphoma of pylorus of stomach |
| SNOMED CT | 447658000 | Lymphoma of fundus of stomach |
| SNOMED CT | 447766003 | Lymphoma of pyloric antrum of stomach |
| SNOMED CT | 447805007 | Lymphoma of greater curvature of stomach |
| SNOMED CT | 447806008 | Lymphoma of cardia of stomach |
| SNOMED CT | 447989004 | Non-Hodgkin's lymphoma of extranodal site |
| SNOMED CT | 448212009 | Anaplastic large cell lymphoma, ALK negative |
| SNOMED CT | 448213004 | Diffuse non-Hodgkin's lymphoma of prostate |
| SNOMED CT | 448217003 | Follicular non-Hodgkin's lymphoma of prostate |
| SNOMED CT | 448220006 | Non-Hodgkin's lymphoma of bone |
| SNOMED CT | 448231003 | Follicular non-Hodgkin's lymphoma of nose |
| SNOMED CT | 448254007 | Non-Hodgkin's lymphoma of central nervous system |
| SNOMED CT | 448269008 | Lymphoma of lesser curvature of stomach |
| SNOMED CT | 448317000 | Follicular non-Hodgkin's lymphoma of soft tissue |
| SNOMED CT | 448319002 | Diffuse non-Hodgkin's lymphoma of nasopharynx |
| SNOMED CT | 448354009 | Non-Hodgkin's lymphoma of intestine |
| SNOMED CT | 448371005 | Non-Hodgkin's lymphoma of nasopharynx |
| SNOMED CT | 448372003 | Non-Hodgkin's lymphoma of lung |
| SNOMED CT | 448376000 | Non-Hodgkin's lymphoma of ovary |
| SNOMED CT | 448384001 | Non-Hodgkin's lymphoma of nose |
| SNOMED CT | 448386004 | Non-Hodgkin's lymphoma of oral cavity |
| SNOMED CT | 448387008 | Non-Hodgkin's lymphoma of testis |
| SNOMED CT | 448447004 | Non-Hodgkin's lymphoma of skin |
| SNOMED CT | 448465000 | Diffuse non-Hodgkin's lymphoma of testis |
| SNOMED CT | 448468003 | Diffuse non-Hodgkin's lymphoma of oral cavity |
| SNOMED CT | 448553002 | Lymphoma of pelvis |
| SNOMED CT | 448555009 | Lymphoma of body of stomach |
| SNOMED CT | 448560008 | Diffuse non-Hodgkin's lymphoma of extranodal site |
| SNOMED CT | 448561007 | Follicular non-Hodgkin's lymphoma of extranodal site |
| SNOMED CT | 448607004 | Diffuse non-Hodgkin's lymphoma of uterine cervix |
| SNOMED CT | 448609001 | Diffuse non-Hodgkin's lymphoma of ovary |
| SNOMED CT | 448663003 | Diffuse non-Hodgkin's lymphoma of stomach |
| SNOMED CT | 448666006 | Follicular non-Hodgkin's lymphoma of bone |
| SNOMED CT | 448672006 | Follicular non-Hodgkin's lymphoma of lung |
| SNOMED CT | 448709005 | Non-Hodgkin's lymphoma of stomach |
| SNOMED CT | 448738008 | Non-Hodgkin's lymphoma of soft tissue |
| SNOMED CT | 448774004 | Non-Hodgkin's lymphoma of uterine cervix |
| SNOMED CT | 448865007 | Follicular non-Hodgkin's lymphoma of skin |
| SNOMED CT | 448867004 | Diffuse non-Hodgkin's lymphoma of lung |
| SNOMED CT | 448995000 | Follicular non-Hodgkin's lymphoma of central nervous system |
| SNOMED CT | 449053004 | Lymphoma of lower esophagus |
| SNOMED CT | 449058008 | Follicular non-Hodgkin's lymphoma of tonsil |
| SNOMED CT | 449059000 | Follicular non-Hodgkin's lymphoma of uterine cervix |
| SNOMED CT | 449063007 | Follicular non-Hodgkin's lymphoma of oral cavity |
| SNOMED CT | 449065000 | Diffuse non-Hodgkin's lymphoma of nose |
| SNOMED CT | 449072004 | Gastrointestinal lymphoma |

Continued on next page

**TableS31 – continued from previous page**

| CodeSystem | ConceptCode | ConceptName |
|---|---|---|
| SNOMED CT | 449074003 | Lymphoma of small bowel |
| SNOMED CT | 449075002 | Lymphoma of cardioesophageal junction |
| SNOMED CT | 449108003 | Philadelphia chromosome positive chronic myelogenous leukemia |
| SNOMED CT | 449173006 | Diffuse non-Hodgkin's lymphoma of tonsil |
| SNOMED CT | 449176003 | Diffuse non-Hodgkin's lymphoma of intestine |
| SNOMED CT | 449177007 | Diffuse non-Hodgkin's lymphoma of bone |
| SNOMED CT | 449187006 | Kappa light chain deficiency |
| SNOMED CT | 449216004 | Diffuse non-Hodgkin's lymphoma of soft tissue |
| SNOMED CT | 449217008 | Diffuse non-Hodgkin's lymphoma of skin |
| SNOMED CT | 449218003 | Lymphoma of sigmoid colon |
| SNOMED CT | 449219006 | Follicular non-Hodgkin's lymphoma of nasopharynx |
| SNOMED CT | 449220000 | Diffuse follicle center lymphoma |
| SNOMED CT | 449221001 | Diffuse non-Hodgkin's lymphoma of central nervous system |
| SNOMED CT | 449222008 | Follicular non-Hodgkin's lymphoma of stomach |
| SNOMED CT | 449292003 | Non-Hodgkin's lymphoma of tonsil |
| SNOMED CT | 449307001 | Follicular non-Hodgkin's lymphoma of ovary |
| SNOMED CT | 449318001 | Non-Hodgkin's lymphoma of prostate |
| SNOMED CT | 449384005 | Lambda light chain deficiency |
| SNOMED CT | 449386007 | Philadelphia chromosome negative chronic myelogenous leukemia |
| SNOMED CT | 449418000 | Follicular non-Hodgkin's lymphoma of testis |
| SNOMED CT | 449419008 | Follicular non-Hodgkin's lymphoma of intestine |
| SNOMED CT | 449784008 | Diffuse infiltrative lymphocytosis syndrome |
| SNOMED CT | 449853003 | Interleukin-12 deficiency |
| SNOMED CT | 450907007 | Hydroa vacciniforme-like lymphoma |
| SNOMED CT | 450908002 | Primary cutaneous gamma-delta T-cell lymphoma |
| SNOMED CT | 450909005 | Plasmablastic lymphoma |
| SNOMED CT | 450910000 | ALK positive large B-cell lymphoma |
| SNOMED CT | 450911001 | Large B-cell lymphoma arising in HHV8 associated multicentric Castleman disease |
| SNOMED CT | 450912008 | Fibroblastic reticular cell tumor |
| SNOMED CT | 450913003 | Mixed phenotype acute leukemia |
| SNOMED CT | 450914009 | Mixed phenotype acute leukemia with t(9;22)(q34;q11.2); BCR-ABL1 |
| SNOMED CT | 450915005 | Mixed phenotype acute leukemia with t(v;11q23); MLL rearranged |
| SNOMED CT | 450916006 | Mixed phenotype acute leukemia B/myeloid |
| SNOMED CT | 450917002 | Mixed phenotype acute leukemia T/myeloid |
| SNOMED CT | 450920005 | B lymphoblastic leukemia / lymphoma - category |
| SNOMED CT | 450928003 | Acute myeloid leukemia with t(6;9)(p23;q34); DEK-NUP214 |
| SNOMED CT | 450929006 | Acute myeloid leukemia with inv(3)(q21q26.2) or t(3;3)(q21;q26.2); RPN1-EVI1 |
| SNOMED CT | 450935006 | Myeloid leukemia associated with Down Syndrome |
| SNOMED CT | 450937003 | Acute myeloid leukemia (megakaryoblastic) with t(1;22)(p13;q13); RBM15-MKL1 |
| SNOMED CT | 450940003 | Myeloid or lymphoid neoplasm with alpha-type platelet-derived growth factor receptor gene rearrangement |

Continued on next page

**TableS31 – continued from previous page**

| CodeSystem | ConceptCode | ConceptName |
|---|---|---|
| SNOMED CT | 450945008 | Refractory cytopenia with unilineage dysplasia |
| SNOMED CT | 450946009 | Refractory neutropenia |
| SNOMED CT | 450947000 | Refractory thrombocytopenia |
| SNOMED CT | 450950002 | B lymphoblastic leukemia lymphoma with t(9;22)(q34;q11.2); BCR-ABL1 |
| SNOMED CT | 450951003 | B lymphoblastic leukemia lymphoma with t(v;11q23); MLL rearranged |
| SNOMED CT | 450952005 | B lymphoblastic leukemia lymphoma with t(12;21)(p13;q22); TEL-AML1 (ETV6-RUNX1) |
| SNOMED CT | 450953000 | B lymphoblastic leukemia lymphoma with hyperdiploidy |
| SNOMED CT | 450954006 | B lymphoblastic leukemia lymphoma with hypodiploidy (Hypodiploid ALL) |
| SNOMED CT | 450955007 | B lymphoblastic leukemia lymphoma with t(5;14)(q31;q32); IL3-IGH |
| SNOMED CT | 450956008 | B lymphoblastic leukemia lymphoma with t(1;19)(q23;p13.3); E2A-PBX1 (TCF3-PBX1) |
| SNOMED CT | 450958009 | Malignant lymphoma, diffuse large B-cell, immunoblastic |
| SNOMED CT | 450959001 | T-cell/histiocyte rich large B-cell lymphoma |
| SNOMED CT | 697922003 | Pulmonary hypertension in Langerhans cell histiocytosis |
| SNOMED CT | 697965002 | Cholangitis with AIDS (acquired immunodeficiency syndrome) |
| SNOMED CT | 698075004 | Syngeneic peripheral blood stem cell transplantation |
| SNOMED CT | 698646006 | Acute monoblastic leukemia in remission |
| SNOMED CT | 698694005 | Systemic lupus erythematosus in remission |
| SNOMED CT | 699293000 | Ischemic myelofibrosis |
| SNOMED CT | 699537002 | Polyostotic sclerosing histiocytosis |
| SNOMED CT | 699657009 | Hepatosplenic gamma-delta cell lymphoma |
| SNOMED CT | 699818003 | T-cell large granular lymphocytosis |
| SNOMED CT | 700053002 | Sepsis with acquired immunodeficiency syndrome |
| SNOMED CT | 702444009 | Autoimmune lymphoproliferative syndrome |
| SNOMED CT | 702446006 | Core binding factor acute myeloid leukemia |
| SNOMED CT | 702476004 | Therapy-related myelodysplastic syndrome |
| SNOMED CT | 702785000 | Large cell (Ki-1 positive) lymphoma |
| SNOMED CT | 702786004 | Follicular non-Hodgkin's lymphoma diffuse follicle center subtype grade 1 |
| SNOMED CT | 702977001 | Follicular non-Hodgkin's lymphoma diffuse follicle center cell sub-type grade 2 |
| SNOMED CT | 703387000 | Cytogenetically normal acute myeloid leukemia |
| SNOMED CT | 703525006 | Anhidrotic ectodermal dysplasia with immune deficiency |
| SNOMED CT | 703540008 | Majeed syndrome |
| SNOMED CT | 703626001 | Anaplastic large cell lymphoma, T/Null cell, primary systemic type |
| SNOMED CT | 703817002 | Myelodysplastic/myeloproliferative neoplasm with ring sideroblasts and thrombocytosis |
| SNOMED CT | 703818007 | Acute monoblastic and monocytic leukemia |
| SNOMED CT | 703819004 | Acute myeloid leukemia with mutation of CEBPA (CCAAT enhancer binding protein alpha) gene |
| SNOMED CT | 703820005 | Acute myeloid leukemia with mutated NPM1 |

Continued on next page

**TableS31 – continued from previous page**

| CodeSystem | ConceptCode | ConceptName |
|---|---|---|
| SNOMED CT | 703821009 | T lymphoblastic leukemia/lymphoma |
| SNOMED CT | 703822002 | Indeterminate dendritic cell tumor |
| SNOMED CT | 705061009 | Childhood myelodysplastic syndrome |
| SNOMED CT | 707152007 | Phagocytic immunodeficiency |
| SNOMED CT | 707431006 | Langerhans cell histiocytosis of diaper area |
| SNOMED CT | 707432004 | Langerhans cell histiocytosis of skin |
| SNOMED CT | 709115004 | Transplantation of autologous hematopoietic stem cell |
| SNOMED CT | 709465004 | Periodontitis co-occurrent with Chédiak-Higashi syndrome |
| SNOMED CT | 709471005 | Periodontitis co-occurrent with leukemia |
| SNOMED CT | 709535007 | Periodontitis co-occurrent with infantile genetic agranulocytosis |
| SNOMED CT | 709608008 | Periodontitis co-occurrent with acquired neutropenia |
| SNOMED CT | 710735009 | Periodontitis co-occurrent with leukocyte adhesion deficiency |
| SNOMED CT | 710926008 | Periodontitis co-occurrent with familial neutropenia |
| SNOMED CT | 710927004 | Periodontitis co-occurrent with cyclical neutropenia |
| SNOMED CT | 711164003 | STING-associated vasculopathy with onset in infancy |
| SNOMED CT | 711429001 | Transplantation of autologous progenitor cell |
| SNOMED CT | 711480000 | Activated PI3K-delta syndrome |
| SNOMED CT | 713260006 | Subacute adenoviral encephalitis co-occurrent with human immunodeficiency virus infection |
| SNOMED CT | 713275003 | Splenomegaly co-occurrent with human immunodeficiency virus infection |
| SNOMED CT | 713278001 | Neuralgia co-occurrent with human immunodeficiency virus infection |
| SNOMED CT | 713297001 | Candidiasis of esophagus co-occurrent with human immunodeficiency virus infection |
| SNOMED CT | 713298006 | Heart disease co-occurrent with human immunodeficiency virus infection |
| SNOMED CT | 713299003 | Disorder of eye proper co-occurrent with human immunodeficiency virus infection |
| SNOMED CT | 713300006 | Disorder of gastrointestinal tract co-occurrent with human immunodeficiency virus infection |
| SNOMED CT | 713316008 | Eruption of skin co-occurrent with human immunodeficiency virus infection |
| SNOMED CT | 713318009 | Myocarditis co-occurrent with human immunodeficiency virus infection |
| SNOMED CT | 713320007 | Radiculitis co-occurrent with human immunodeficiency virus infection |
| SNOMED CT | 713325002 | Primary cerebral lymphoma co-occurrent with human immunodeficiency virus infection |
| SNOMED CT | 713339002 | Infection caused by Strongyloides co-occurrent with human immunodeficiency virus infection |
| SNOMED CT | 713340000 | Disorder of skin co-occurrent with human immunodeficiency virus infection |
| SNOMED CT | 713341001 | Myelitis co-occurrent with human immunodeficiency virus infection |
| SNOMED CT | 713342008 | Infection caused by Salmonella co-occurrent with human immunodeficiency virus infection |

Continued on next page

**TableS31 – continued from previous page**

| CodeSystem | ConceptCode | ConceptName |
|---|---|---|
| SNOMED CT | 713349004 | Anemia co-occurrent with human immunodeficiency virus infection |
| SNOMED CT | 713444005 | Hemophagocytic syndrome with human immunodeficiency virus infection |
| SNOMED CT | 713445006 | Disseminated infection caused by Strongyloides co-occurrent with human immunodeficiency virus infection |
| SNOMED CT | 713446007 | Chronic infection caused by herpes simplex virus co-occurrent with human immunodeficiency virus infection |
| SNOMED CT | 713483007 | Reticulosarcoma co-occurrent with human immunodeficiency virus infection |
| SNOMED CT | 713484001 | Disorder of respiratory system co-occurrent with human immunodeficiency virus infection |
| SNOMED CT | 713487008 | Progressive multifocal leucoencephalopathy co-occurrent with human immunodeficiency virus infection |
| SNOMED CT | 713488003 | Presenile dementia co-occurrent with human immunodeficiency virus infection |
| SNOMED CT | 713489006 | Polyneuropathy co-occurrent with human immunodeficiency virus infection |
| SNOMED CT | 713490002 | Infection caused by Pneumocystis co-occurrent with human immunodeficiency virus infection |
| SNOMED CT | 713491003 | Organic brain syndrome co-occurrent with human immunodeficiency virus infection |
| SNOMED CT | 713497004 | Candidiasis of mouth co-occurrent with human immunodeficiency virus infection |
| SNOMED CT | 713503007 | Disorder of spinal cord co-occurrent with human immunodeficiency virus infection |
| SNOMED CT | 713504001 | Disorder of kidney co-occurrent with human immunodeficiency virus infection |
| SNOMED CT | 713505000 | Gastrointestinal malabsorption syndrome co-occurrent with human immunodeficiency virus infection |
| SNOMED CT | 713506004 | Neuritis co-occurrent with human immunodeficiency virus infection |
| SNOMED CT | 713507008 | Lymphadenopathy co-occurrent with human immunodeficiency virus infection |
| SNOMED CT | 713508003 | Aplastic anemia co-occurrent with human immunodeficiency virus infection |
| SNOMED CT | 713510001 | Enlargement of liver co-occurrent with human immunodeficiency virus infection |
| SNOMED CT | 713511002 | Acute endocarditis co-occurrent with human immunodeficiency virus infection |
| SNOMED CT | 713516007 | Primary effusion lymphoma |
| SNOMED CT | 713523008 | Cardiomyopathy co-occurrent with human immunodeficiency virus infection |
| SNOMED CT | 713526000 | Recurrent bacterial pneumonia co-occurrent with human immunodeficiency virus infection |
| SNOMED CT | 713527009 | Disorder of peripheral nervous system co-occurrent with human immunodeficiency virus infection |

Continued on next page

**TableS31 – continued from previous page**

| CodeSystem | ConceptCode | ConceptName |
|---|---|---|
| SNOMED CT | 713530002 | Agranulocytosis co-occurrent with human immunodeficiency virus infection |
| SNOMED CT | 713531003 | Visual impairment co-occurrent with human immunodeficiency virus infection |
| SNOMED CT | 713532005 | Infective arthritis co-occurrent with human immunodeficiency virus infection |
| SNOMED CT | 713533000 | Acquired hemolytic anemia co-occurrent with human immunodeficiency virus infection |
| SNOMED CT | 713543002 | Demyelinating disease of central nervous system co-occurrent with human immunodeficiency virus infection |
| SNOMED CT | 713544008 | Bacterial pneumonia co-occurrent with human immunodeficiency virus infection |
| SNOMED CT | 713545009 | Infection caused by Nocardia co-occurrent with human immunodeficiency virus infection |
| SNOMED CT | 713546005 | Isosporiasis co-occurrent with human immunodeficiency virus infection |
| SNOMED CT | 713570009 | Infectious gastroenteritis co-occurrent with human immunodeficiency virus infection |
| SNOMED CT | 713571008 | Disorder of central nervous system co-occurrent with human immunodeficiency virus infection |
| SNOMED CT | 713572001 | Malignant neoplastic disease co-occurrent with human immunodeficiency virus infection |
| SNOMED CT | 713695001 | Nephrotic syndrome co-occurrent with human immunodeficiency virus infection |
| SNOMED CT | 713696000 | Renal failure syndrome co-occurrent with human immunodeficiency virus infection |
| SNOMED CT | 713718006 | Diffuse non-Hodgkin immunoblastic lymphoma co-occurrent with human immunodeficiency virus infection |
| SNOMED CT | 713722001 | Infection caused by Cytomegalovirus co-occurrent with human immunodeficiency virus infection |
| SNOMED CT | 713729005 | Coccidiosis co-occurrent with human immunodeficiency virus infection |
| SNOMED CT | 713730000 | Infection caused by herpes simplex virus co-occurrent with human immunodeficiency virus infection |
| SNOMED CT | 713731001 | Pyrexia of unknown origin co-occurrent with human immunodeficiency virus infection |
| SNOMED CT | 713732008 | Aspergillosis co-occurrent with human immunodeficiency virus infection |
| SNOMED CT | 713733003 | Infection caused by herpes zoster virus co-occurrent with human immunodeficiency virus infection |
| SNOMED CT | 713734009 | Dermatophytosis co-occurrent with human immunodeficiency virus infection |
| SNOMED CT | 713742005 | Human immunodeficiency virus antibody positive |
| SNOMED CT | 713844000 | Dementia co-occurrent with human immunodeficiency virus infection |
| SNOMED CT | 713845004 | Infection caused by Cryptosporidium co-occurrent with human immunodeficiency virus infection |

Continued on next page

**TableS31 – continued from previous page**

| CodeSystem | ConceptCode | ConceptName |
|---|---|---|
| SNOMED CT | 713880000 | Opportunistic mycosis co-occurrent with human immunodeficiency virus infection |
| SNOMED CT | 713881001 | Microsporidiosis co-occurrent with human immunodeficiency virus infection |
| SNOMED CT | 713887002 | Focal segmental glomerulosclerosis co-occurrent with human immunodeficiency virus infection |
| SNOMED CT | 713897006 | Burkitt lymphoma co-occurrent with human immunodeficiency virus infection |
| SNOMED CT | 713910008 | Antibody mediated acquired pure red cell aplasia caused by erythropoiesis stimulating agent |
| SNOMED CT | 713964006 | Multidermatomal infection caused by Herpes zoster co-occurrent with human immunodeficiency virus infection |
| SNOMED CT | 713967004 | Disseminated atypical infection caused by Mycobacterium co-occurrent with human immunodeficiency virus infection |
| SNOMED CT | 714083007 | Recurrent salmonella sepsis co-occurrent with human immunodeficiency virus infection |
| SNOMED CT | 714251006 | Philadelphia chromosome-negative precursor B-cell acute lymphoblastic leukemia |
| SNOMED CT | 714463003 | Primary effusion lymphoma co-occurrent with infection caused by Human herpesvirus 8 |
| SNOMED CT | 714464009 | Immune reconstitution inflammatory syndrome caused by human immunodeficiency virus infection |
| SNOMED CT | 715664005 | Interdigitating dendritic cell sarcoma |
| SNOMED CT | 715950008 | ALK-positive large B-cell lymphoma |
| SNOMED CT | 715982006 | Severe combined immunodeficiency due to DCLRE1C deficiency |
| SNOMED CT | 716192009 | Thong Douglas Ferrante syndrome |
| SNOMED CT | 716378008 | Combined immunodeficiency due to ZAP70 deficiency |
| SNOMED CT | 716655008 | Aggressive systemic mastocytosis |
| SNOMED CT | 716788007 | Epstein-Barr virus positive diffuse large B-cell lymphoma of elderly |
| SNOMED CT | 716789004 | Epstein-Barr virus positive diffuse large B-cell lymphoma of elderly |
| SNOMED CT | 716869006 | Mendelian susceptibility to mycobacterial disease due to complete IL12RB1 deficiency |
| SNOMED CT | 716871006 | Severe combined immunodeficiency due to deoxyribonucleic acid dependent protein kinase catalytic subunit deficiency |
| SNOMED CT | 717811007 | Combined immunodeficiency due to CRAC (calcium release activated calcium) channel dysfunction |
| SNOMED CT | 718107000 | T-cell negative B-cell positive severe combined immunodeficiency due to JAK3 deficiency |
| SNOMED CT | 718200007 | Primary pulmonary lymphoma |
| SNOMED CT | 718230004 | Mendelian susceptibility to mycobacterial disease due to complete interferon gamma receptor 1 deficiency |
| SNOMED CT | 718232007 | Pyogenic bacterial infection due to deficiency of myeloid differentiation primary response 88 |
| SNOMED CT | 718614004 | TEMPI syndrome |
| SNOMED CT | 718717004 | Primary immunodeficiency syndrome due to p14 deficiency |

Continued on next page

**TableS31 – continued from previous page**

| CodeSystem | ConceptCode | ConceptName |
|---|---|---|
| SNOMED CT | 718882006 | X-linked severe congenital neutropenia |
| SNOMED CT | 719019000 | WT limb blood syndrome |
| SNOMED CT | 719156006 | X-linked intellectual disability with hypogammaglobulinemia and progressive neurological deterioration syndrome |
| SNOMED CT | 719522009 | Candidiasis of upper respiratory tract co-occurrent with human immunodeficiency virus infection |
| SNOMED CT | 719685004 | Absent thumb with short stature and immunodeficiency syndrome |
| SNOMED CT | 719789000 | Human immunodeficiency virus 2 antibody positive and Human immunodeficiency virus 1 antibody cross-reactivity |
| SNOMED CT | 719810000 | X-linked intellectual disability with seizure and psoriasis syndrome |
| SNOMED CT | 719814009 | X-linked mendelian susceptibility to mycobacterial disease |
| SNOMED CT | 719824001 | Vici syndrome |
| SNOMED CT | 719827008 | X-linked immunoneurologic disorder |
| SNOMED CT | 720520009 | Attenuated Chédiak-Higashi syndrome |
| SNOMED CT | 720853005 | Severe combined immunodeficiency, microcephaly, growth retardation, sensitivity to ionizing radiation syndrome |
| SNOMED CT | 720986005 | Anhidrotic ectodermal dysplasia, immunodeficiency, osteopetrosis, lymphedema syndrome |
| SNOMED CT | 721166000 | Human immunodeficiency virus complicating pregnancy childbirth and the puerperium |
| SNOMED CT | 721301004 | Myeloid and lymphoid neoplasm with fibroblast growth factor receptor 1 abnormality |
| SNOMED CT | 721302006 | Myelodysplastic/myeloproliferative neoplasm with ring sideroblasts and thrombocytosis |
| SNOMED CT | 721303001 | Refractory neutropenia |
| SNOMED CT | 721304007 | Refractory thrombocytopenia |
| SNOMED CT | 721305008 | Acute myeloid leukemia due to recurrent genetic abnormality |
| SNOMED CT | 721306009 | Therapy related acute myeloid leukemia and myelodysplastic syndrome |
| SNOMED CT | 721308005 | Acute leukemia of ambiguous lineage |
| SNOMED CT | 721310007 | Aggressive natural killer-cell leukemia |
| SNOMED CT | 721311006 | Systemic Epstein-Barr virus positive T-cell lymphoproliferative disease of childhood |
| SNOMED CT | 721313009 | Indeterminate dendritic cell neoplasm |
| SNOMED CT | 721314003 | Fibroblastic reticular cell neoplasm |
| SNOMED CT | 721555001 | Follicular lymphoma of small intestine |
| SNOMED CT | 721762007 | Adult T-cell leukemia/lymphoma of skin |
| SNOMED CT | 721876004 | Mendelian susceptibility to mycobacterial disease due to complete interferon gamma receptor 2 deficiency |
| SNOMED CT | 721877008 | Mendelian susceptibility to mycobacterial disease due to complete interleukin 12 subunit beta deficiency |
| SNOMED CT | 721903007 | Microcephaly, hypogammaglobulinemia, abnormal immunity syndrome |
| SNOMED CT | 721977007 | Lung fibrosis, immunodeficiency, 46,XX gonadal dysgenesis syndrome |

Continued on next page

**TableS31 – continued from previous page**

| CodeSystem | ConceptCode | ConceptName |
|---|---|---|
| SNOMED CT | 722067005 | Severe combined immunodeficiency with hypereosinophilia |
| SNOMED CT | 68979007 | Heavy chain disease |
| SNOMED CT | 69077002 | Acute basophilic leukemia |
| SNOMED CT | 69896004 | RhA - Rheumatoid arthritis |
| SNOMED CT | 70958007 | Plasma cell hyperplasia of bone marrow |
| SNOMED CT | 71436005 | Lazy leukocyte syndrome |
| SNOMED CT | 71904008 | SCID due to absent class II HLA antigens |
| SNOMED CT | 71922006 | Immune defect |
| SNOMED CT | 73286009 | SLE glomerulonephritis syndrome, WHO class I |
| SNOMED CT | 74189002 | Hodgkin granuloma [obs] |
| SNOMED CT | 76243000 | Chronic granulomatous disease, type IVA |
| SNOMED CT | 76521009 | SLE glomerulonephritis syndrome, WHO class III |
| SNOMED CT | 77084001 | Immunologic aplastic anemia |
| SNOMED CT | 77330006 | Chronic granulomatous disease, type II |
| SNOMED CT | 77358003 | Congenital leukocyte adherence deficiency |
| SNOMED CT | 77753005 | Lupus disease of the lung |
| SNOMED CT | 78378009 | Isoimmune neutropenia |
| SNOMED CT | 79019005 | Human immunodeficiency virus II infection |
| SNOMED CT | 79369007 | Complication of transplanted pancreas |
| SNOMED CT | 80570006 | Acute panmyelosis with myelofibrosis |
| SNOMED CT | 80976008 | Myasthenic crisis |
| SNOMED CT | 81166004 | Properdin deficiency disease |
| SNOMED CT | 82178003 | Neonatal myasthenia gravis |
| SNOMED CT | 82317007 | Chronic granulomatous disease, type III |
| SNOMED CT | 82546001 | Reactive immunoproliferative disease |
| SNOMED CT | 83980004 | Impaired phagocytosis |
| SNOMED CT | 86219005 | Uveitis-rheumatoid arthritis syndrome |
| SNOMED CT | 86406008 | Human immunodeficiency virus infection |
| SNOMED CT | 87117006 | HIV infection with acute lymphadenitis |
| SNOMED CT | 89454001 | Shwachman syndrome |
| SNOMED CT | 89655007 | Congenital neutropenia |
| SNOMED CT | 90120004 | Mycosis fungoides |
| SNOMED CT | 90414007 | Chronic acquired pure red cell aplasia |
| SNOMED CT | 91637004 | Myasthenia gravis |
| SNOMED CT | 91854005 | Acute leukemia in remission |
| SNOMED CT | 91855006 | Acute leukemia |
| SNOMED CT | 91856007 | Acute lymphoid leukemia in remission |
| SNOMED CT | 91857003 | Acute lymphoid leukemia |
| SNOMED CT | 91858008 | Acute monocytic leukemia in remission |
| SNOMED CT | 91860005 | Acute myeloid leukemia in remission |
| SNOMED CT | 91861009 | Acute myeloid leukemia, disease |
| SNOMED CT | 91947003 | Asymptomatic human immunodeficiency virus infection |
| SNOMED CT | 91948008 | Asymptomatic human immunodeficiency virus infection in pregnancy |
| SNOMED CT | 92811003 | Chronic leukemia in remission |
| SNOMED CT | 92812005 | Chronic leukemia |
| SNOMED CT | 92813000 | Chronic lymphoid leukemia in remission |

Continued on next page

**TableS31 – continued from previous page**

| CodeSystem | ConceptCode | ConceptName |
|---|---|---|
| SNOMED CT | 92814006 | Chronic lymphoid leukemia, disease |
| SNOMED CT | 92817004 | Chronic myeloid leukemia in remission |
| SNOMED CT | 92818009 | Chronic myeloid leukemia |
| SNOMED CT | 93133006 | Letterer-Siwe disease of intra-abdominal lymph nodes |
| SNOMED CT | 93134000 | Letterer-Siwe disease of intrapelvic lymph nodes |
| SNOMED CT | 93135004 | Letterer-Siwe disease of intrathoracic lymph nodes |
| SNOMED CT | 93136003 | Letterer-Siwe disease of lymph nodes of axilla AND/OR upper limb |
| SNOMED CT | 93137007 | Letterer-Siwe disease of lymph nodes of head, face AND/OR neck |
| SNOMED CT | 93138002 | Letterer-Siwe disease of lymph nodes of inguinal region AND/OR lower limb |
| SNOMED CT | 93139005 | Letterer-Siwe disease of lymph nodes of multiple sites |
| SNOMED CT | 93140007 | Letterer-Siwe disease of spleen |
| SNOMED CT | 93141006 | Letterer-Siwe disease of extranodal AND/OR solid organ site |
| SNOMED CT | 93142004 | Leukemia in remission |
| SNOMED CT | 93143009 | Leukemia |
| SNOMED CT | 93144003 | Leukemic reticuloendotheliosis of intra-abdominal lymph nodes |
| SNOMED CT | 93145002 | Leukemic reticuloendotheliosis of intrapelvic lymph nodes |
| SNOMED CT | 93146001 | Leukemic reticuloendotheliosis of intrathoracic lymph nodes |
| SNOMED CT | 93150008 | Leukemic reticuloendotheliosis of lymph nodes of multiple sites |
| SNOMED CT | 93151007 | Hairy cell leukemia of spleen |
| SNOMED CT | 93152000 | Leukemic reticuloendotheliosis of extranodal AND/OR solid organ site |
| SNOMED CT | 93169003 | Lymphoid leukemia in remission |
| SNOMED CT | 93182006 | Malignant histiocytosis of intra-abdominal lymph nodes |
| SNOMED CT | 93183001 | Malignant histiocytosis of intrapelvic lymph nodes |
| SNOMED CT | 93184007 | Malignant histiocytosis of intrathoracic lymph nodes |
| SNOMED CT | 93185008 | Malignant histiocytosis of lymph nodes of axilla AND/OR upper limb |
| SNOMED CT | 93186009 | Malignant histiocytosis of lymph nodes of head, face AND/OR neck |
| SNOMED CT | 93187000 | Malignant histiocytosis of lymph nodes of inguinal region AND/OR lower limb |
| SNOMED CT | 93188005 | Malignant histiocytosis of lymph nodes of multiple sites |
| SNOMED CT | 93189002 | Malignant histiocytosis of spleen |
| SNOMED CT | 93190006 | Malignant histiocytosis of extranodal AND/OR solid organ site |
| SNOMED CT | 93191005 | Malignant lymphoma of intra-abdominal lymph nodes |
| SNOMED CT | 93192003 | Malignant lymphoma of intrapelvic lymph nodes |
| SNOMED CT | 93193008 | Malignant lymphoma of intrathoracic lymph nodes |
| SNOMED CT | 93194002 | Malignant lymphoma of lymph nodes of axilla AND/OR upper limb |
| SNOMED CT | 93195001 | Malignant lymphoma of lymph nodes of head, face AND/OR neck |
| SNOMED CT | 93196000 | Malignant lymphoma of lymph nodes of inguinal region AND/OR lower limb |
| SNOMED CT | 93197009 | Malignant lymphoma of lymph nodes of multiple sites |

Continued on next page

**TableS31 – continued from previous page**

| CodeSystem | ConceptCode | ConceptName |
|---|---|---|
| SNOMED CT | 93198004 | Malignant lymphoma of spleen |
| SNOMED CT | 93199007 | Malignant lymphoma of extranodal AND/OR solid organ site |
| SNOMED CT | 93200005 | Malignant mast cell tumor of intra-abdominal lymph nodes |
| SNOMED CT | 93201009 | Malignant mast cell tumor of intrapelvic lymph nodes |
| SNOMED CT | 93202002 | Malignant mast cell tumor of intrathoracic lymph nodes |
| SNOMED CT | 93203007 | Malignant mast cell tumor of lymph nodes of axilla AND/OR upper limb |
| SNOMED CT | 93204001 | Malignant mast cell tumor of lymph nodes of head, face AND/OR neck |
| SNOMED CT | 93205000 | Malignant mast cell tumor of lymph nodes of inguinal region AND/OR lower limb |
| SNOMED CT | 94148006 | Megakaryocytic leukemia in remission |
| SNOMED CT | 94686001 | Mixed cell type lymphosarcoma of intra-abdominal lymph nodes |
| SNOMED CT | 94687005 | Mixed cell type lymphosarcoma of intrapelvic lymph nodes |
| SNOMED CT | 94688000 | Mixed cell type lymphosarcoma of intrathoracic lymph nodes |
| SNOMED CT | 94690004 | Mixed cell type lymphosarcoma of lymph nodes of head, face, and neck |
| SNOMED CT | 94704006 | Multiple myeloma in remission |
| SNOMED CT | 94707004 | Mycosis fungoides of intra-abdominal lymph nodes |
| SNOMED CT | 94708009 | Mycosis fungoides of intrapelvic lymph nodes |
| SNOMED CT | 94709001 | Mycosis fungoides of intrathoracic lymph nodes |
| SNOMED CT | 94710006 | Mycosis fungoides of lymph nodes of axilla AND/OR upper limb |
| SNOMED CT | 94711005 | Mycosis fungoides of lymph nodes of head, face AND/OR neck |
| SNOMED CT | 94712003 | Mycosis fungoides of lymph nodes of inguinal region AND/OR lower limb |
| SNOMED CT | 94714002 | Mycosis fungoides of spleen |
| SNOMED CT | 94715001 | Mycosis fungoides of extranodal AND/OR solid organ site |
| SNOMED CT | 94716000 | Myeloid leukemia in remission |
| SNOMED CT | 94718004 | Myeloid sarcoma in remission |
| SNOMED CT | 95186006 | Nodular lymphoma of intra-abdominal lymph nodes |
| SNOMED CT | 95187002 | Nodular lymphoma of intrapelvic lymph nodes |
| SNOMED CT | 95188007 | Nodular lymphoma of intrathoracic lymph nodes |
| SNOMED CT | 95192000 | Nodular lymphoma of lymph nodes of multiple sites |
| SNOMED CT | 95193005 | Nodular lymphoma of spleen |
| SNOMED CT | 95194004 | Nodular lymphoma of extranodal AND/OR solid organ site |
| SNOMED CT | 95209008 | Plasma cell leukemia in remission |
| SNOMED CT | 95210003 | Plasma cell leukemia |
| SNOMED CT | 95224004 | Reticulosarcoma of intra-abdominal lymph nodes |
| SNOMED CT | 95225003 | Reticulosarcoma of intrapelvic lymph nodes |
| SNOMED CT | 95226002 | Reticulosarcoma of intrathoracic lymph nodes |
| SNOMED CT | 95230004 | Reticulosarcoma of lymph nodes of multiple sites |
| SNOMED CT | 95231000 | Reticulosarcoma of spleen |
| SNOMED CT | 95260009 | Sézary's disease of lymph nodes of head, face AND/OR neck |
| SNOMED CT | 95263006 | Sézary's disease of spleen |
| SNOMED CT | 95264000 | Sézary's disease of extranodal AND/OR solid organ site |
| SNOMED CT | 95644001 | Systemic lupus erythematosus encephalitis |
| SNOMED CT | 95892003 | Persistent generalized lymphadenopathy |

Continued on next page

**TableS31 – continued from previous page**

| CodeSystem | ConceptCode | ConceptName |
|---|---|---|
| SNOMED CT | 103075007 | Humoral immune defect |
| SNOMED CT | 103077004 | Immunodeficiency secondary to neoplasm |
| SNOMED CT | 103078009 | Immunodeficiency secondary to trauma |
| SNOMED CT | 103079001 | Immunodeficiency secondary to corticosteroid |
| SNOMED CT | 103080003 | Immunodeficiency secondary to radiation therapy |
| SNOMED CT | 103081004 | Immunodeficiency secondary to chemotherapy |
| SNOMED CT | 103682005 | Subcutaneous panniculitis-like T-cell lymphoma |
| SNOMED CT | 103685007 | Hepatosplenic gamma-delta cell lymphoma |
| SNOMED CT | 103688009 | Acute myeloid leukemia with abnormal marrow eosinophils |
| SNOMED CT | 103689001 | Acute myeloid leukemia, minimal differentiation |
| SNOMED CT | 103690005 | Acute myeloid leukemia without maturation |
| SNOMED CT | 103691009 | Acute myeloid leukemia with maturation |
| SNOMED CT | 105600002 | Disorder of neutrophils |
| SNOMED CT | 105603000 | Non-malignant lymphocyte AND/OR plasma cell disorder |
| SNOMED CT | 109958007 | Hematologic neoplasm of uncertain behavior |
| SNOMED CT | 109975001 | T-zone lymphoma (clinical) |
| SNOMED CT | 109976000 | Lymphoepithelioid lymphoma (clinical) |
| SNOMED CT | 109978004 | T-cell lymphoma (clinical) |
| SNOMED CT | 109980005 | Malignant immunoproliferative disease (clinical) |
| SNOMED CT | 109982002 | Alpha heavy chain disease (clinical) |
| SNOMED CT | 109984001 | Gamma heavy chain disease (clinical) |
| SNOMED CT | 109985000 | Immunoproliferative small intestinal disease (clinical) |
| SNOMED CT | 109988003 | Histiocytic sarcoma (clinical) |
| SNOMED CT | 109989006 | Multiple myeloma |
| SNOMED CT | 109991003 | Acute panmyelosis with myelofibrosis |
| SNOMED CT | 109993000 | Chronic myeloproliferative disorder (clinical) |
| SNOMED CT | 109994006 | Essential thrombocythemia |
| SNOMED CT | 109995007 | Myelodysplastic syndrome (clinical) |
| SNOMED CT | 109996008 | Refractory anemia (clinical) |
| SNOMED CT | 109998009 | Refractory anemia with ringed sideroblasts associated with marked thrombocytosis |
| SNOMED CT | 110000005 | Refractory anemia with excess blasts in transformation (clinical) |
| SNOMED CT | 110002002 | Mast cell leukemia (clinical) |
| SNOMED CT | 110006004 | Prolymphocytic leukemia (clinical) |
| SNOMED CT | 110459008 | Malignant lymphoma, metastatic |
| SNOMED CT | 111001004 | Gammopathy |
| SNOMED CT | 111584000 | Reticular dysgenesis |
| SNOMED CT | 111585004 | Neutropenia associated with autoimmune disease |
| SNOMED CT | 111587007 | SCID due to absent IL-2 receptor |
| SNOMED CT | 111880001 | Acute HIV infection |
| SNOMED CT | 112687003 | Hodgkin lymphoma, lymphocyte depletion |
| SNOMED CT | 115244002 | Malignant lymphoma - category |
| SNOMED CT | 115245001 | Malignant lymphoma, follicular AND/OR nodular |
| SNOMED CT | 115246000 | Specified cutaneous AND/OR peripheral T cell lymphoma |
| SNOMED CT | 116133005 | Congenital agammaglobulinemia |
| SNOMED CT | 118607005 | Lymphocyte-rich classical Hodgkin lymphoma |
| SNOMED CT | 118611004 | Sézary's disease (clinical) |

Continued on next page

**TableS31 – continued from previous page**

| CodeSystem | ConceptCode | ConceptName |
|---|---|---|
| SNOMED CT | 118612006 | Malignant histiocytosis (clinical) |
| SNOMED CT | 118615008 | Malignant mast cell tumor (clinical) |
| SNOMED CT | 118618005 | Mycosis fungoides (clinical) |
| SNOMED CT | 119249001 | Agammaglobulinemia |
| SNOMED CT | 119250001 | Hypogammaglobulinemia |
| SNOMED CT | 123313007 | Alpha heavy chain disease, enteric form |
| SNOMED CT | 123777002 | Autoimmune leukopenia |
| SNOMED CT | 123782009 | Dysgammaglobulinemia |
| SNOMED CT | 124950009 | Disorder of immunoglobulin |
| SNOMED CT | 127034005 | Pancytopenia |
| SNOMED CT | 127069007 | Hemophagocytic syndrome |
| SNOMED CT | 127070008 | Malignant histiocytic disorder |
| SNOMED CT | 127220001 | Malignant lymphoma of lymph nodes |
| SNOMED CT | 127225006 | Chronic myelomonocytic leukemia |
| SNOMED CT | 127388009 | Hypergammaglobulinemia |
| SNOMED CT | 128623006 | Myelodysplastic syndrome |
| SNOMED CT | 128798004 | Composite Hodgkin and non-Hodgkin lymphoma |
| SNOMED CT | 128800006 | Primary effusion lymphoma |
| SNOMED CT | 128801005 | Mediastinal large B-cell lymphoma |
| SNOMED CT | 128802003 | Splenic marginal zone B-cell lymphoma |
| SNOMED CT | 128803008 | Marginal zone B-cell lymphoma |
| SNOMED CT | 128806000 | Precursor cell lymphoblastic lymphoma |
| SNOMED CT | 128807009 | Precursor B-cell lymphoblastic lymphoma |
| SNOMED CT | 128808004 | Precursor T-cell lymphoblastic lymphoma |
| SNOMED CT | 128813000 | Histiocytic sarcoma |
| SNOMED CT | 128814006 | Langerhans cell sarcoma |
| SNOMED CT | 128815007 | Interdigitating dendritic cell sarcoma |
| SNOMED CT | 128816008 | Follicular dendritic cell sarcoma |
| SNOMED CT | 128818009 | Acute biphenotypic leukemia |
| SNOMED CT | 128820007 | Prolymphocytic leukemia, B-cell type |
| SNOMED CT | 128822004 | Precursor cell lymphoblastic leukemia |
| SNOMED CT | 128823009 | Precursor B-cell lymphoblastic leukemia |
| SNOMED CT | 128824003 | Precursor T-cell lymphoblastic leukemia |
| SNOMED CT | 128825002 | Chronic myelogenous leukemia, BCR/ABL positive |
| SNOMED CT | 128826001 | Atypical chronic myeloid leukemia, BCR/ABL negative |
| SNOMED CT | 128827005 | Acute myeloid leukemia with myelodysplasia-related changes |
| SNOMED CT | 128828000 | Acute myeloid leukemia, t(8;21) (q22;q22) |
| SNOMED CT | 128829008 | Acute myeloid leukemia, 11q23 abnormalities |
| SNOMED CT | 128830003 | Therapy-related acute myeloid leukemia and myelodysplastic syndrome |
| SNOMED CT | 128832006 | Juvenile myelomonocytic leukemia |
| SNOMED CT | 128834007 | Chronic neutrophilic leukemia |
| SNOMED CT | 128836009 | Myelodysplastic syndrome with multilineage dysplasia |
| SNOMED CT | 128837000 | Myelodysplastic syndrome with isolated del (5q) |
| SNOMED CT | 128838005 | Therapy-related myelodysplastic syndrome |
| SNOMED CT | 128843003 | Myelosclerosis with myeloid metaplasia |
| SNOMED CT | 128845005 | Refractory anemia |

Continued on next page

**TableS31 – continued from previous page**

| CodeSystem | ConceptCode | ConceptName |
|---|---|---|
| SNOMED CT | 128846006 | Refractory anemia with ring sideroblasts associated with marked thrombocytosis |
| SNOMED CT | 128847002 | Myelodysplastic syndrome with excess blasts |
| SNOMED CT | 128848007 | Refractory anemia with excess blasts in transformation |
| SNOMED CT | 128920006 | Malignant histiocytosis |
| SNOMED CT | 128921005 | Plasmacytoma, extramedullary (not occurring in bone) |
| SNOMED CT | 128922003 | Plasma cell leukemia |
| SNOMED CT | 128923008 | Prolymphocytic leukemia |
| SNOMED CT | 128924002 | Mast cell leukemia |
| SNOMED CT | 128931003 | Leukemia (category) |
| SNOMED CT | 128932005 | Acute leukemia (category) |
| SNOMED CT | 128933000 | Chronic leukemia (category) |
| SNOMED CT | 128934006 | Myeloid leukemia (category) |
| SNOMED CT | 128935007 | Lymphoid leukemia (category) |
| SNOMED CT | 129642004 | Chronic idiopathic immunoneutropenia in adult |
| SNOMED CT | 129643009 | Chronic hypoplastic neutropenia |
| SNOMED CT | 129644003 | Myeloperoxidase deficiency syndrome |
| SNOMED CT | 129648000 | Hypoalphaglobulinemia |
| SNOMED CT | 129649008 | Hypobetaglobulinemia |
| SNOMED CT | 165816005 | HIV positive |
| SNOMED CT | 186706006 | Human immunodeficiency virus infection constitutional disease |
| SNOMED CT | 186707002 | Human immunodeficiency virus infection with neurological disease |
| SNOMED CT | 186708007 | Human immunodeficiency virus infection with secondary clinical infectious disease |
| SNOMED CT | 188487008 | Lymphosarcoma and reticulosarcoma |
| SNOMED CT | 188489006 | Reticulosarcoma of lymph nodes of head, face and neck |
| SNOMED CT | 188492005 | Reticulosarcoma of lymph nodes of axilla and upper limb |
| SNOMED CT | 188493000 | Reticulosarcoma of lymph nodes of inguinal region and lower limb |
| SNOMED CT | 188498009 | Lymphosarcoma |
| SNOMED CT | 188500005 | Lymphosarcoma of lymph nodes of head, face and neck |
| SNOMED CT | 188501009 | Lymphosarcoma of intrathoracic lymph nodes |
| SNOMED CT | 188502002 | Lymphosarcoma of intra-abdominal lymph nodes |
| SNOMED CT | 188503007 | Lymphosarcoma of lymph nodes of axilla and upper limb |
| SNOMED CT | 188504001 | Lymphosarcoma of lymph nodes of inguinal region and lower limb |
| SNOMED CT | 188505000 | Lymphosarcoma of intrapelvic lymph nodes |
| SNOMED CT | 188506004 | Lymphosarcoma of spleen |
| SNOMED CT | 188507008 | Lymphosarcoma of lymph nodes of multiple sites |
| SNOMED CT | 188609000 | Nodular lymphoma of lymph nodes of head, face and neck |
| SNOMED CT | 188612002 | Nodular lymphoma of lymph nodes of axilla and upper limb |
| SNOMED CT | 188613007 | Nodular lymphoma of lymph nodes of inguinal region and lower limb |
| SNOMED CT | 188627002 | Mycosis fungoides of lymph nodes of multiple sites |
| SNOMED CT | 188630009 | Sézary's disease of lymph nodes of head, face and neck |
| SNOMED CT | 188631008 | Sézary's disease of intrathoracic lymph nodes |

Continued on next page

**TableS31 – continued from previous page**

| CodeSystem | ConceptCode | ConceptName |
|---|---|---|
| SNOMED CT | 188633006 | Sézary's disease of lymph nodes of axilla AND/OR upper limb |
| SNOMED CT | 188634000 | Sézary's disease of lymph nodes of inguinal region and lower limb |
| SNOMED CT | 188635004 | Sézary's disease of intrapelvic lymph nodes |
| SNOMED CT | 188637007 | Sézary's disease of lymph nodes of multiple sites |
| SNOMED CT | 188640007 | Malignant histiocytosis of lymph nodes of head, face and neck |
| SNOMED CT | 188641006 | Malignant histiocytosis of lymph nodes of axilla and upper limb |
| SNOMED CT | 188642004 | Malignant histiocytosis of lymph nodes of inguinal region and lower limb |
| SNOMED CT | 188645002 | Leukemic reticuloendotheliosis of lymph nodes of head, face and neck |
| SNOMED CT | 188648000 | Leukemic reticuloendotheliosis of lymph nodes of axilla and upper limb |
| SNOMED CT | 188649008 | Leukemic reticuloendotheliosis of lymph nodes of inguinal region and lower limb |
| SNOMED CT | 188660004 | Malignant mast cell tumors |
| SNOMED CT | 188662007 | Mast cell malignancy of lymph nodes of head, face and neck |
| SNOMED CT | 188663002 | Mast cell malignancy of intrathoracic lymph nodes |
| SNOMED CT | 188664008 | Mast cell malignancy of intra-abdominal lymph nodes |
| SNOMED CT | 188665009 | Mast cell malignancy of lymph nodes of axilla and upper limb |
| SNOMED CT | 188666005 | Mast cell malignancy of lymph nodes of inguinal region and lower limb |
| SNOMED CT | 188667001 | Mast cell malignancy of intrapelvic lymph nodes |
| SNOMED CT | 188668006 | Mast cell malignancy of spleen |
| SNOMED CT | 188669003 | Mast cell malignancy of lymph nodes of multiple sites |
| SNOMED CT | 188674006 | Diffuse malignant lymphoma - small non-cleaved cell |
| SNOMED CT | 188675007 | Malignant lymphoma - small cleaved cell |
| SNOMED CT | 188676008 | Malignant lymphoma - mixed small and large cell |
| SNOMED CT | 188691005 | Malignant immunoproliferative small intestinal disease |
| SNOMED CT | 188718006 | Extramedullary plasmacytoma |
| SNOMED CT | 188725004 | Lymphoid leukemia |
| SNOMED CT | 188726003 | Subacute lymphoid leukemia |
| SNOMED CT | 188728002 | Aleukemic lymphoid leukemia |
| SNOMED CT | 188729005 | Adult T-cell leukemia |
| SNOMED CT | 188732008 | Myeloid leukemia |
| SNOMED CT | 188733003 | Chronic eosinophilic leukemia |
| SNOMED CT | 188734009 | Chronic neutrophilic leukemia |
| SNOMED CT | 188736006 | Subacute myeloid leukemia |
| SNOMED CT | 188737002 | Chloroma |
| SNOMED CT | 188738007 | Granulocytic sarcoma |
| SNOMED CT | 188741003 | Aleukemic myeloid leukemia |
| SNOMED CT | 188744006 | Monocytic leukemia |
| SNOMED CT | 188745007 | Chronic monocytic leukemia |
| SNOMED CT | 188746008 | Subacute monocytic leukemia |
| SNOMED CT | 188748009 | Aleukemic monocytic leukemia |
| SNOMED CT | 188754005 | Megakaryocytic leukemia |
| SNOMED CT | 188768003 | Myelomonocytic leukemia |
| SNOMED CT | 188770007 | Subacute myelomonocytic leukemia |

Continued on next page

**TableS31 – continued from previous page**

| CodeSystem | ConceptCode | ConceptName |
|---|---|---|
| SNOMED CT | 189509003 | Refractory anemia without sideroblasts, so stated |
| SNOMED CT | 189962004 | Malignant lymphoma, stem cell type |
| SNOMED CT | 189982000 | Reticulosarcoma morphology |
| SNOMED CT | 189984004 | Reticulosarcoma, pleomorphic cell type |
| SNOMED CT | 189985003 | Reticulosarcoma, nodular |
| SNOMED CT | 190030009 | Compound leukemias |
| SNOMED CT | 190055003 | Eosinophilic leukemia |
| SNOMED CT | 190808009 | Polyclonal hypergammaglobulinemia |
| SNOMED CT | 190817009 | Macroglobulinemia |
| SNOMED CT | 190818004 | Waldenström macroglobulinemia |
| SNOMED CT | 190923000 | Sporadic primary amyloidosis |
| SNOMED CT | 190955000 | Histiocytosis X syndrome |
| SNOMED CT | 190959006 | Haemophagocytic lymphohistiocytosis due to infection |
| SNOMED CT | 190979003 | Selective immunoglobulin A deficiency |
| SNOMED CT | 190980000 | Selective immunoglobulin M deficiency |
| SNOMED CT | 190981001 | Selective immunoglobulin G deficiency |
| SNOMED CT | 190986006 | Antibody deficiency with near-normal immunoglobulins or with hyperimmunoglobulinemia |
| SNOMED CT | 190993005 | Autosomal recessive severe combined immunodeficiency |
| SNOMED CT | 190995003 | Thymic aplasia or dysplasia with immunodeficiency |
| SNOMED CT | 190996002 | Severe combined immunodeficiency with reticular dysgenesis |
| SNOMED CT | 190997006 | Severe combined immunodeficiency with low T- and B-cell numbers |
| SNOMED CT | 190998001 | Severe combined immunodeficiency with low or normal B-cell numbers |
| SNOMED CT | 191001007 | Major histocompatibility complex class I deficiency |
| SNOMED CT | 191002000 | Major histocompatibility complex class II deficiency |
| SNOMED CT | 191008001 | Immunodeficiency following hereditary defective response to Epstein-Barr virus |
| SNOMED CT | 191011000 | Common variable immunodeficiency with predominant abnormalities of B-cell numbers and functions |
| SNOMED CT | 191012007 | Common variable immunodeficiency with predominant immunoregulatory T-cell disorders |
| SNOMED CT | 191013002 | Common variable immunodeficiency with autoantibodies to B- or T-cells |
| SNOMED CT | 191018006 | Lymphocyte function antigen-1 defect |
| SNOMED CT | 191244003 | Aplastic anemia due to chronic disease |
| SNOMED CT | 191246001 | Aplastic anemia due to infection |
| SNOMED CT | 191247005 | Aplastic anemia due to radiation |
| SNOMED CT | 191248000 | Aplastic anemia due to toxic cause |
| SNOMED CT | 191255003 | Transient acquired pure red cell aplasia |
| SNOMED CT | 191256002 | Idiopathic aplastic anemia |
| SNOMED CT | 191338000 | Primary splenic neutropenia |
| SNOMED CT | 191347008 | Cyclical neutropenia |
| SNOMED CT | 193206003 | Persistent neonatal myasthenia gravis |
| SNOMED CT | 193207007 | Juvenile or adult myasthenia gravis |
| SNOMED CT | 196138005 | Lung disease with systemic lupus erythematosus |

Continued on next page

**TableS31 – continued from previous page**

| CodeSystem | ConceptCode | ConceptName |
|---|---|---|
| SNOMED CT | 201764007 | Rheumatoid arthritis of cervical spine |
| SNOMED CT | 201766009 | Rheumatoid arthritis of shoulder |
| SNOMED CT | 201767000 | Rheumatoid arthritis of sternoclavicular joint |
| SNOMED CT | 201768005 | Rheumatoid arthritis of acromioclavicular joint |
| SNOMED CT | 201769002 | Rheumatoid arthritis of elbow |
| SNOMED CT | 201770001 | Rheumatoid arthritis of distal radioulnar joint |
| SNOMED CT | 201771002 | Rheumatoid arthritis of wrist |
| SNOMED CT | 201772009 | Rheumatoid arthritis of metacarpophalangeal joint |
| SNOMED CT | 201773004 | Rheumatoid arthritis of proximal interphalangeal joint of finger |
| SNOMED CT | 201774005 | Rheumatoid arthritis of distal interphalangeal joint of finger |
| SNOMED CT | 201775006 | Rheumatoid arthritis of hip |
| SNOMED CT | 201776007 | Rheumatoid arthritis of sacroiliac joint |
| SNOMED CT | 201777003 | Rheumatoid arthritis of knee |
| SNOMED CT | 201778008 | Rheumatoid arthritis of tibiofibular joint |
| SNOMED CT | 201779000 | Rheumatoid arthritis of ankle |
| SNOMED CT | 201780002 | Rheumatoid arthritis of subtalar joint |
| SNOMED CT | 201781003 | Rheumatoid arthritis of talonavicular joint |
| SNOMED CT | 201783000 | Rheumatoid arthritis of first metatarsophalangeal joint |
| SNOMED CT | 201784006 | Rheumatoid arthritis of lesser metatarsophalangeal joint |
| SNOMED CT | 201785007 | Rheumatoid arthritis of interphalangeal joint of toe |
| SNOMED CT | 201791009 | Flare of rheumatoid arthritis |
| SNOMED CT | 209962000 | Myeloma-associated amyloidosis |
| SNOMED CT | 213148006 | Transplanted organ rejection |
| SNOMED CT | 230180003 | Human immunodeficiency virus leucoencephalopathy |
| SNOMED CT | 230202002 | Vacuolar myelopathy |
| SNOMED CT | 230598008 | Neuropathy due to human immunodeficiency virus |
| SNOMED CT | 230669004 | Genetically determined myasthenia |
| SNOMED CT | 230670003 | Familial infantile myasthenia |
| SNOMED CT | 230671004 | Acetylcholine resynthesis deficiency |
| SNOMED CT | 230672006 | Congenital myasthenic syndrome |
| SNOMED CT | 230673001 | Congenital end-plate acetylcholine receptor deficiency |
| SNOMED CT | 230674007 | Pseudomyopathic myasthenia |
| SNOMED CT | 230676009 | Putative defect in acetylcholine synthesis or packaging |
| SNOMED CT | 230677000 | Congenital end-plate acetylcholinesterase deficiency |
| SNOMED CT | 230678005 | Decrease of motor end-plate potential amplitude without acetylcholine receptor deficiency |
| SNOMED CT | 230679002 | Abnormality of synaptic vesicles |
| SNOMED CT | 230683002 | Transient neonatal myasthenia gravis |
| SNOMED CT | 230684008 | Ocular myasthenia |
| SNOMED CT | 230685009 | Myasthenia gravis associated with thymoma |
| SNOMED CT | 230686005 | Generalized myasthenia |
| SNOMED CT | 230687001 | Myopathy in myasthenia gravis |
| SNOMED CT | 232075002 | Lymphoma of retina |
| SNOMED CT | 234146006 | Hennekam syndrome |
| SNOMED CT | 234331007 | Syngeneic bone marrow transplant |
| SNOMED CT | 234332000 | T-cell depleted allogeneic bone marrow graft |
| SNOMED CT | 234333005 | Imperfect T-cell depleted allogeneic bone marrow graft |

Continued on next page

**TableS31 – continued from previous page**

| CodeSystem | ConceptCode | ConceptName |
|---|---|---|
| SNOMED CT | 234334004 | Allogeneic related bone marrow transplant |
| SNOMED CT | 234335003 | Allogeneic unrelated bone marrow transplant |
| SNOMED CT | 234336002 | Hemopoietic stem cell transplant |
| SNOMED CT | 234367000 | Pancytopenia with pancreatitis |
| SNOMED CT | 234370001 | Pure red cell aplasia, acquired |
| SNOMED CT | 234376007 | Acquired red cell aplasia |
| SNOMED CT | 234414004 | Non-malignant white cell disorder |
| SNOMED CT | 234416002 | X-linked hypogammaglobulinemia |
| SNOMED CT | 234424007 | Metabolic neutropenia |
| SNOMED CT | 234425008 | Autoimmune neutropenia |
| SNOMED CT | 234428005 | Lymphocytoid disorder |
| SNOMED CT | 234429002 | Chemotactic disorder |
| SNOMED CT | 234430007 | CR3-receptor deficiency |
| SNOMED CT | 234431006 | Specific granule deficiency |
| SNOMED CT | 234433009 | Myeloperoxidase deficiency |
| SNOMED CT | 234436001 | Hemolytic erythrophagocytic syndrome |
| SNOMED CT | 234437005 | Hemophagocytic lymphohistiocytosis |
| SNOMED CT | 234519006 | Bone marrow transplant rejection |
| SNOMED CT | 234520000 | Bone marrow transplant failure |
| SNOMED CT | 234522008 | Disease relapse in transplant marrow |
| SNOMED CT | 234532001 | Immunodeficiency disorder |
| SNOMED CT | 234533006 | X-linked agammaglobulinemia with growth hormone deficiency |
| SNOMED CT | 234534000 | Autosomal agammaglobulinemia with absent B-cells |
| SNOMED CT | 234535004 | Selective immunoglobulin dysfunction |
| SNOMED CT | 234539005 | Immunoglobulin heavy chain deficiency |
| SNOMED CT | 234540007 | Selective immunoglobulin E deficiency |
| SNOMED CT | 234541006 | Selective immunoglobulin D deficiency |
| SNOMED CT | 234542004 | Selective immunoglobulin M and immunoglobulin A deficiency |
| SNOMED CT | 234543009 | Immunoglobulin light chain deficiency |
| SNOMED CT | 234544003 | Immunoglobulin subclass deficiency |
| SNOMED CT | 234546001 | Immunoglobulin G2 deficiency |
| SNOMED CT | 234547005 | Combined immunoglobulin G2 and G4 deficiency |
| SNOMED CT | 234548000 | Immunoglobulin G3 deficiency |
| SNOMED CT | 234549008 | Immunoglobulin G4 deficiency |
| SNOMED CT | 234550008 | Immunoglobulin G1 deficiency |
| SNOMED CT | 234551007 | Immunoglobulin A1 deficiency |
| SNOMED CT | 234552000 | Immunoglobulin A2 deficiency |
| SNOMED CT | 234553005 | Immunoglobulin-associated molecule deficiency |
| SNOMED CT | 234554004 | Secretory piece deficiency |
| SNOMED CT | 234555003 | Defective immunoglobulin glycosylation |
| SNOMED CT | 234563002 | Anti-staphylococcal antibody deficiency |
| SNOMED CT | 234564008 | Primary immunoglobulin catabolism abnormality |
| SNOMED CT | 234565009 | Immunoglobulin hypercatabolism |
| SNOMED CT | 234566005 | Familial immunoglobulin hypercatabolism |
| SNOMED CT | 234570002 | Severe combined immunodeficiency with maternofetal engraftment |
| SNOMED CT | 234571003 | Warts, hypogammaglobulinemia, infections, and myelokathexis |

Continued on next page

**TableS31 – continued from previous page**

| CodeSystem | ConceptCode | ConceptName |
|---|---|---|
| SNOMED CT | 234572005 | Benign combined immunodeficiency |
| SNOMED CT | 234573000 | Phagocytic cell defect |
| SNOMED CT | 234574006 | Disorder of phagocytic cell number |
| SNOMED CT | 234576008 | Chronic familial neutropenia |
| SNOMED CT | 234577004 | Lipochrome histiocytosis - familial |
| SNOMED CT | 234578009 | Defective phagocytic cell opsonization |
| SNOMED CT | 234580003 | Defective phagocytic cell chemotaxis |
| SNOMED CT | 234581004 | Defective phagocytic cell adhesion |
| SNOMED CT | 234582006 | Leukocyte adhesion deficiency - type 1 |
| SNOMED CT | 234583001 | Leukocyte adhesion deficiency - type 2 |
| SNOMED CT | 234584007 | Tuftsin deficiency |
| SNOMED CT | 234585008 | Defective phagocytic cell killing |
| SNOMED CT | 234587000 | Neutrophil lactoferrin deficiency |
| SNOMED CT | 234588005 | Neutrophil secondary granule deficiency |
| SNOMED CT | 234591005 | Combined phagocytic defect |
| SNOMED CT | 234593008 | Classical complement pathway abnormality |
| SNOMED CT | 234594002 | Complement 1q deficiency |
| SNOMED CT | 234595001 | Complement 1q beta chain deficiency |
| SNOMED CT | 234596000 | Complement 1q dysfunction |
| SNOMED CT | 234597009 | Complement 1r deficiency |
| SNOMED CT | 234598004 | Complement 1s deficiency |
| SNOMED CT | 234599007 | Complement 2 deficiency |
| SNOMED CT | 234600005 | Complement 4 deficiency |
| SNOMED CT | 234601009 | Complement 4A deficiency |
| SNOMED CT | 234602002 | Complement 4B deficiency |
| SNOMED CT | 234603007 | Complement 3 deficiency |
| SNOMED CT | 234604001 | Alternative pathway deficiency |
| SNOMED CT | 234605000 | Factor B deficiency |
| SNOMED CT | 234607008 | Factor D deficiency |
| SNOMED CT | 234608003 | Terminal component deficiency |
| SNOMED CT | 234609006 | Complement 5 deficiency |
| SNOMED CT | 234611002 | Complement 6 deficiency |
| SNOMED CT | 234612009 | Complement 7 deficiency |
| SNOMED CT | 234613004 | Combined complement 6 and 7 deficiencies |
| SNOMED CT | 234614005 | Complement 8 beta chain deficiency |
| SNOMED CT | 234615006 | Complement 8 beta chain dysfunction |
| SNOMED CT | 234616007 | Complement 8 alpha-gamma deficiency |
| SNOMED CT | 234617003 | Complement 9 deficiency |
| SNOMED CT | 234618008 | Complement regulatory factor defect |
| SNOMED CT | 234621005 | Factor I deficiency |
| SNOMED CT | 234622003 | Factor H deficiency |
| SNOMED CT | 234623008 | Complement 4 binding protein deficiency |
| SNOMED CT | 234624002 | Decay accelerating factor deficiency |
| SNOMED CT | 234625001 | Homologous restriction factor deficiency |
| SNOMED CT | 234626000 | Complement 5a inhibitor deficiency |
| SNOMED CT | 234627009 | Anaphylotoxin inactivator deficiency |
| SNOMED CT | 234628004 | Complement receptor deficiency |

Continued on next page

**TableS31 – continued from previous page**

| CodeSystem | ConceptCode | ConceptName |
|---|---|---|
| SNOMED CT | 234629007 | Complement receptor 1 deficiency |
| SNOMED CT | 234630002 | Complement receptor 3 deficiency |
| SNOMED CT | 234631003 | Immunodeficiency with major anomalies |
| SNOMED CT | 234632005 | Immunodeficiency associated with chromosomal abnormality |
| SNOMED CT | 234633000 | Centromeric instability of chromosomes 1,9 and 16 and immunodeficiency |
| SNOMED CT | 234634006 | Chromosome 18 syndromes and antibody deficiency |
| SNOMED CT | 234635007 | Chromosome 22 abnormalities with hypogammaglobulinemia |
| SNOMED CT | 234636008 | Monosomy 22 and absence of immunoglobulin A |
| SNOMED CT | 234637004 | Deletion of X-chromosome and hypogammaglobulinemia |
| SNOMED CT | 234638009 | Microcephaly, normal intelligence and immunodeficiency |
| SNOMED CT | 234639001 | Triple X syndrome, epilepsy, and hypogammaglobulinemia |
| SNOMED CT | 234640004 | Immunodeficiency associated with 18p syndrome |
| SNOMED CT | 234641000 | Immunodeficiency associated with multiple organ system abnormalities |
| SNOMED CT | 234645009 | Drug-induced immunodeficiency |
| SNOMED CT | 234646005 | Graft versus host disease |
| SNOMED CT | 235009000 | Human immunodeficiency virus-associated periodontitis |
| SNOMED CT | 235726002 | Human immunodeficiency virus enteropathy |
| SNOMED CT | 236406007 | Acquired immune deficiency syndrome-related nephropathy |
| SNOMED CT | 236512004 | Leukemic infiltrate of kidney |
| SNOMED CT | 236513009 | Lymphoma of kidney |
| SNOMED CT | 236585005 | De novo transplant disease |
| SNOMED CT | 239297008 | Lymphomatoid granulomatosis of lung |
| SNOMED CT | 239791005 | Seropositive rheumatoid arthritis |
| SNOMED CT | 239792003 | Seronegative rheumatoid arthritis |
| SNOMED CT | 239795001 | Rheumatoid arthritis with multisystem involvement |
| SNOMED CT | 239886003 | Limited lupus erythematosus |
| SNOMED CT | 239887007 | Systemic lupus erythematosus with organ/system involvement |
| SNOMED CT | 239888002 | Lupus panniculitis |
| SNOMED CT | 239889005 | Bullous systemic lupus erythematosus |
| SNOMED CT | 239890001 | Systemic lupus erythematosus with multisystem involvement |
| SNOMED CT | 239940004 | Lymphomatoid granulomatosis |
| SNOMED CT | 240103002 | Human immunodeficiency virus myopathy |
| SNOMED CT | 240486003 | Parvoviral aplastic crisis |
| SNOMED CT | 247860002 | Familial neutropenia |
| SNOMED CT | 248693006 | Chronic idiopathic neutropenia |
| SNOMED CT | 254067002 | Immuno-osseous dysplasia |
| SNOMED CT | 254290004 | Lymphoproliferative disorder following transplantation |
| SNOMED CT | 254792006 | Proliferating angioendotheliomatosis |
| SNOMED CT | 255048006 | Tumor of lymphoid, hemopoietic and related tissue |
| SNOMED CT | 255102004 | Angioendotheliomatosis |
| SNOMED CT | 255191003 | Localized malignant reticulohistiocytoma |
| SNOMED CT | 835009 | Angioimmunoblastic T-cell lymphoma |
| SNOMED CT | 3239007 | Lymphocyte disorder |
| SNOMED CT | 3439009 | SCID due to absent peripheral T cell maturation |
| SNOMED CT | 183005 | Autoimmune pancytopenia |

Continued on next page

**TableS31 – continued from previous page**

| CodeSystem | ConceptCode | ConceptName |
|---|---|---|
| SNOMED CT | 3902000 | Non dose-related drug-induced neutropenia |
| SNOMED CT | 4434006 | Bloom syndrome |
| SNOMED CT | 4676006 | SLE glomerulonephritis syndrome, WHO class II |
| SNOMED CT | 5876000 | Acquired pancytopenia |
| SNOMED CT | 6398009 | Idiopathic chronic cold agglutinin disease |
| SNOMED CT | 7607008 | Rheumatoid pericarditis |
| SNOMED CT | 7990002 | Immunoglobulinemia with isolated somatotropin deficiency |
| SNOMED CT | 9893005 | Immunodeficiency with thymoma |
| SNOMED CT | 10639003 | Plasmacytoma |
| SNOMED CT | 11013005 | SLE glomerulonephritis syndrome, WHO class VI |
| SNOMED CT | 12814002 | B lymphocyte disorder |
| SNOMED CT | 13048006 | Orbital lymphoma |
| SNOMED CT | 13720003 | Triclonal gammopathy |
| SNOMED CT | 14024008 | Humoral immunologic aplastic anemia |
| SNOMED CT | 15012007 | Granulocyte abnormality due to immune defect |
| SNOMED CT | 16618004 | Polyclonal gammopathy |
| SNOMED CT | 17182001 | Agranulocytosis |
| SNOMED CT | 18827005 | Complement abnormality |
| SNOMED CT | 19340000 | Malignant lymphoma, lymphoplasmacytic |
| SNOMED CT | 19682006 | Lupus hepatitis |
| SNOMED CT | 19944001 | Autologous bone marrow transplant without purging |
| SNOMED CT | 20224008 | Delta heavy chain disease |
| SNOMED CT | 20447006 | Plasma cell dyscrasia with polyneuropathy |
| SNOMED CT | 21043008 | Neutrophil cytomatrix disorder |
| SNOMED CT | 21527007 | Chronic granulomatous disease, type IV |
| SNOMED CT | 22098000 | Chronic idiopathic autoimmune hemolytic anemia |
| SNOMED CT | 22331004 | Acute monocytic leukemia |
| SNOMED CT | 22406001 | SCID due to absent lymphoid stem cells |
| SNOMED CT | 23132008 | AL amyloidosis |
| SNOMED CT | 23238000 | Common variable agammaglobulinemia |
| SNOMED CT | 24072005 | Acute leukemia |
| SNOMED CT | 24181002 | Aplasia of thymus gland with immunodeficiency |
| SNOMED CT | 24419001 | Disorder of complement |
| SNOMED CT | 24743004 | Complement deficiency disease |
| SNOMED CT | 24974008 | Myelokathexis |
| SNOMED CT | 25050002 | Alpha heavy chain disease, respiratory form |
| SNOMED CT | 25109007 | Lymphopenic agammaglobulinemia - short-limbed dwarfism syndrome |
| SNOMED CT | 25380002 | Pericarditis secondary to systemic lupus erythematosus |
| SNOMED CT | 25502009 | Episodic lymphocytopenia |
| SNOMED CT | 26252007 | Chronic granulomatous disease, type IIA |
| SNOMED CT | 28880005 | Rheumatoid carditis |
| SNOMED CT | 28950004 | Acute promyelocytic leukemia |
| SNOMED CT | 28975000 | Constitutional aplastic anemia |
| SNOMED CT | 29260007 | Immunoglobulin A deficiency |
| SNOMED CT | 29272001 | Chronic granulomatous disease, type I |
| SNOMED CT | 30962008 | Acute myelomonocytic leukemia |

Continued on next page

**TableS31 – continued from previous page**

| CodeSystem | ConceptCode | ConceptName |
|---|---|---|
| SNOMED CT | 31323000 | Severe combined immunodeficiency disease |
| SNOMED CT | 31839002 | Myasthenia gravis, adult form |
| SNOMED CT | 32092008 | Toxic neutropenia |
| SNOMED CT | 33286000 | Secondary immune deficiency disorder |
| SNOMED CT | 34916006 | Neutrophil motility disorder |
| SNOMED CT | 35287006 | Myeloid sarcoma |
| SNOMED CT | 36138009 | Congenital immunodeficiency disease |
| SNOMED CT | 36402006 | SLE glomerulonephritis syndrome, WHO class IV |
| SNOMED CT | 36980009 | SCID due to absent adenosine deaminase |
| SNOMED CT | 37465004 | Plasmacytosis |
| SNOMED CT | 37548006 | Hypopigmentation-immunodeficiency disease |
| SNOMED CT | 38970002 | Doan-Wright syndrome |
| SNOMED CT | 39086001 | Hodgkin lymphoma, nodular sclerosis, cellular phase |
| SNOMED CT | 39674000 | Familial C3B inhibitor deficiency syndrome |
| SNOMED CT | 39795003 | Hand-Schüller-Christian disease |
| SNOMED CT | 40145002 | Congenital neutrophil actin dysfunction |
| SNOMED CT | 40197009 | Chronic granulomatous disease, type IA |
| SNOMED CT | 40780007 | Human immunodeficiency virus I infection |
| SNOMED CT | 41529000 | Hodgkin lymphoma, mixed cellularity |
| SNOMED CT | 41814009 | Neutropenia with dysgranulopoiesis |
| SNOMED CT | 43858000 | Secondary aplastic anemia |
| SNOMED CT | 43985008 | Hodgkin lymphoma, nodular sclerosis, grade 2 |
| SNOMED CT | 44306006 | T lymphocyte disorder |
| SNOMED CT | 44371002 | Mixed polyclonal cryoimmunoglobulinemia |
| SNOMED CT | 44940001 | Adenosine deaminase deficiency |
| SNOMED CT | 45390000 | SCID due to absent IL-2 production |
| SNOMED CT | 45572000 | Hodgkin lymphoma, nodular sclerosis, grade 1 |
| SNOMED CT | 45841007 | Kappa light chain disease |
| SNOMED CT | 46280001 | Autologous bone marrow transplant with purging |
| SNOMED CT | 46744002 | Follicular lymphoma, grade 1 |
| SNOMED CT | 46923007 | Hodgkin sarcoma [obs] |
| SNOMED CT | 47318007 | Drug-induced neutropenia |
| SNOMED CT | 48119005 | Dysplasia of thymus gland with immunodeficiency |
| SNOMED CT | 48794007 | HIV infection with infectious mononucleosis-like syndrome |
| SNOMED CT | 49555001 | SCID due to absent T cell receptor |
| SNOMED CT | 50220002 | Cellular immunologic aplastic anemia |
| SNOMED CT | 51092000 | B-cell chronic lymphocytic leukaemia/small lymphocytic lymphoma |
| SNOMED CT | 51319002 | Light chain disease |
| SNOMED CT | 52042003 | SLE glomerulonephritis syndrome, WHO class V |
| SNOMED CT | 52064007 | Biclonal gammopathy |
| SNOMED CT | 52079000 | Congenital human immunodeficiency virus infection |
| SNOMED CT | 52220008 | Acute megakaryoblastic leukemia |
| SNOMED CT | 52248008 | Hodgkin lymphoma, nodular sclerosis |
| SNOMED CT | 52661003 | Extra-articular rheumatoid process |
| SNOMED CT | 52967002 | Myelofibrosis |
| SNOMED CT | 54072008 | Nonbacterial verrucal endocardiosis |

Continued on next page

**TableS31 – continued from previous page**

| CodeSystem | ConceptCode | ConceptName |
|---|---|---|
| SNOMED CT | 54087003 | Hairy cell leukemia variant |
| SNOMED CT | 54097007 | White blood cell disorder |
| SNOMED CT | 55020008 | Follicular lymphoma, grade 2 |
| SNOMED CT | 55051001 | Myasthenia gravis, juvenile form |
| SNOMED CT | 55150002 | Follicular lymphoma |
| SNOMED CT | 55464009 | Systemic lupus erythematosus |
| SNOMED CT | 55907008 | Acquired aplastic anemia |
| SNOMED CT | 55921005 | Multiple myeloma |
| SNOMED CT | 56918001 | Dose-related drug-induced neutropenia |
| SNOMED CT | 57390009 | Mixed cryoimmunoglobulinemia with monoclonal component |
| SNOMED CT | 58034007 | Congenital hypergammaglobulinemia |
| SNOMED CT | 58390007 | Allogeneic bone marrow transplantation |
| SNOMED CT | 58606001 | Primary immune deficiency disorder |
| SNOMED CT | 58648008 | Benign monoclonal gammopathy |
| SNOMED CT | 58776007 | Autologous bone marrow transplant |
| SNOMED CT | 58961005 | Lethal midline granuloma |
| SNOMED CT | 60620005 | Epsilon heavy chain disease |
| SNOMED CT | 60743005 | Purine-nucleoside phosphorylase deficiency |
| SNOMED CT | 61493004 | Mu heavy chain disease |
| SNOMED CT | 62479008 | AIDS |
| SNOMED CT | 63364005 | Chronic myeloid leukemia |
| SNOMED CT | 64575004 | Malignant lymphoma, lymphocytic |
| SNOMED CT | 65623009 | Immune neutropenia |
| SNOMED CT | 66876008 | Lambda light chain disease |
| SNOMED CT | 68504005 | Ataxia-telangiectasia syndrome |
| SNOMED CT | 68712004 | Lymphocytic hypoplasia |
| SNOMED CT | 68815009 | SLE glomerulonephritis syndrome |
| SNOMED CT | 722281001 | Agammaglobulinemia, microcephaly, craniosynostosis, severe dermatitis syndrome |
| SNOMED CT | 722288007 | Autoimmune enteropathy and endocrinopathy with susceptibility to chronic infection syndrome |
| SNOMED CT | 722290008 | Autoimmune lymphoproliferative syndrome with recurrent viral infection |
| SNOMED CT | 722557007 | Parkinsonism due to human immunodeficiency virus infection |
| SNOMED CT | 722795004 | Meningeal leukemia |
| SNOMED CT | 722953004 | B-cell lymphoma unclassifiable with features intermediate between Burkitt lymphoma and diffuse large B-cell lymphoma |
| SNOMED CT | 722954005 | B-cell lymphoma unclassifiable with features intermediate between classical Hodgkin lymphoma and diffuse large B-cell lymphoma |
| SNOMED CT | 722955006 | Chronic lymphoproliferative disorder of natural killer cells |
| SNOMED CT | 722956007 | Early post-transplant lymphoproliferative disorder |
| SNOMED CT | 722957003 | Reactive plasmacytic hyperplasia post-transplant lymphoproliferative disorder |
| SNOMED CT | 722958008 | Infectious mononucleosis-like post-transplant lymphoproliferative disorder |
| SNOMED CT | 723334006 | FADD-related immunodeficiency |

Continued on next page

**TableS31 – continued from previous page**

| CodeSystem | ConceptCode | ConceptName |
|---|---|---|
| SNOMED CT | 723384004 | Mendelian susceptibility to mycobacterial disease due to complete ISG15 deficiency |
| SNOMED CT | 723385003 | Mendelian susceptibility to mycobacterial disease due to partial IRF8 deficiency |
| SNOMED CT | 723386002 | Mendelian susceptibility to mycobacterial disease due to partial STAT1 deficiency |
| SNOMED CT | 723443003 | Neutrophil immunodeficiency syndrome |
| SNOMED CT | 723508002 | RAS-associated autoimmune leukoproliferative disease |
| SNOMED CT | 723512008 | Revesz syndrome |
| SNOMED CT | 723889003 | B lymphoblastic leukemia lymphoma with t(9:22) (q34;q11.2); BCR-ABL 1 |
| SNOMED CT | 723995003 | Schimke immuno-osseous dysplasia |
| SNOMED CT | 724015007 | Pyogenic arthritis, pyoderma gangrenosum, acne syndrome |
| SNOMED CT | 724177005 | LIG4 syndrome |
| SNOMED CT | 724179008 | Laron syndrome with immunodeficiency |
| SNOMED CT | 724275005 | Primary immunodeficiency with natural killer cell deficiency and adrenal insufficiency |
| SNOMED CT | 724276006 | X-linked immune dysregulation, polyendocrinopathy, enteropathy syndrome |
| SNOMED CT | 724361001 | Hepatic veno-occlusive disease with immunodeficiency syndrome |
| SNOMED CT | 724642009 | Myeloid neoplasm associated with beta-type platelet-derived growth factor receptor gene rearrangement |
| SNOMED CT | 724644005 | Myeloid leukemia co-occurrent with Down syndrome |
| SNOMED CT | 724645006 | T-cell histiocyte rich large B-cell lymphoma |
| SNOMED CT | 724647003 | Diffuse large B-cell lymphoma co-occurrent with chronic inflammation caused by Epstein-Barr virus |
| SNOMED CT | 724648008 | Plasmablastic lymphoma |
| SNOMED CT | 724649000 | Langerhans cell sarcoma |
| SNOMED CT | 724650000 | Primary follicular dendritic cell sarcoma |
| SNOMED CT | 724767000 | Chorea co-occurrent and due to systemic lupus erythematosus |
| SNOMED CT | 724781003 | Demyelination of central nervous system co-occurrent and due to systemic lupus erythematosus |
| SNOMED CT | 725135004 | Combined immunodeficiency due to CD3gamma deficiency |
| SNOMED CT | 725136003 | Immunodeficiency by defective expression of human leukocyte antigen class 1 |
| SNOMED CT | 725137007 | Neutropenia, monocytopenia, deafness syndrome |
| SNOMED CT | 725150008 | Autosomal dominant mendelian susceptibility to mycobacterial disease due to partial interferon gamma receptor 1 deficiency |
| SNOMED CT | 725151007 | Autosomal dominant mendelian susceptibility to mycobacterial disease due to partial interferon gamma receptor 2 deficiency |
| SNOMED CT | 725290000 | Combined immunodeficiency due to partial RAG1 deficiency |
| SNOMED CT | 725390002 | Acute myeloid leukemia with t(8;16)(p11;p13) translocation |
| SNOMED CT | 725391003 | Acute myeloid leukemia with t(8;16)(p11;p13) translocation |
| SNOMED CT | 725431001 | Autosomal recessive mendelian susceptibility to mycobacterial disease due to partial interferon gamma receptor 1 deficiency |
| SNOMED CT | 725432008 | Autosomal recessive mendelian susceptibility to mycobacterial disease due to partial interferon gamma receptor 2 deficiency |

Continued on next page

**TableS31 – continued from previous page**

| CodeSystem | ConceptCode | ConceptName |
|---|---|---|
| SNOMED CT | 725437002 | Chronic lymphocytic leukemia genetic mutation variant |
| SNOMED CT | 726721002 | Nodal marginal zone B-cell lymphoma |
| SNOMED CT | 733028000 | Multiple sclerosis, ichthyosis, factor VIII deficiency syndrome |
| SNOMED CT | 733598001 | Acute myeloid leukemia with t(6;9)(p23;q34) translocation |
| SNOMED CT | 733834006 | Invasive carcinoma of uterine cervix co-occurrent with human immunodeficiency virus infection |
| SNOMED CT | 733835007 | Extrapulmonary tuberculosis co-occurrent with human immunodeficiency virus infection |
| SNOMED CT | 733860007 | Pediatric nodal marginal zone lymphoma |
| SNOMED CT | 733895005 | Primary cutaneous CD8 positive aggressive epidermotropic cytotoxic T-cell lymphoma |
| SNOMED CT | 733917002 | Pediatric follicular lymphoma |
| SNOMED CT | 734044003 | Anaplastic lymphoma kinase positive anaplastic large cell lymphoma |
| SNOMED CT | 734066005 | Diffuse large B-cell lymphoma of central nervous system |
| SNOMED CT | 734067001 | Splenic diffuse red pulp small B-cell lymphoma |
| SNOMED CT | 734076008 | Diffuse large B-cell lymphoma associated with chronic inflammation |
| SNOMED CT | 734141009 | Splenic B-cell lymphoma |
| SNOMED CT | 734142002 | B-cell lymphoma with features intermediate between diffuse large B-cell lymphoma and Burkitt lymphoma |
| SNOMED CT | 734522002 | Acute myeloid leukemia with FMS-like tyrosine kinase-3 mutation |
| SNOMED CT | 734524001 | Acute myeloid leukemia with FMS-like tyrosine kinase-3 mutation |
| SNOMED CT | 735332000 | Primary cutaneous diffuse large cell B-cell lymphoma leg type |
| SNOMED CT | 735436001 | Acquired disorder of neutrophil function |
| SNOMED CT | 735437005 | Disorder of neutrophil adhesion |
| SNOMED CT | 735438000 | Disorder of neutrophil chemotaxis |
| SNOMED CT | 735443007 | Acquired lymphocytopenia |
| SNOMED CT | 735521001 | Human immunodeficiency virus World Health Organization 2007 stage 1 co-occurrent with tuberculosis |
| SNOMED CT | 735522008 | Human immunodeficiency virus World Health Organization 2007 stage 1 co-occurrent with malaria |
| SNOMED CT | 735523003 | Human immunodeficiency virus World Health Organization 2007 stage 2 co-occurrent with tuberculosis |
| SNOMED CT | 735524009 | Human immunodeficiency virus World Health Organization 2007 stage 2 co-occurrent with malaria |
| SNOMED CT | 735525005 | Human immunodeficiency virus World Health Organization 2007 stage 3 co-occurrent with tuberculosis |
| SNOMED CT | 735526006 | Human immunodeficiency virus World Health Organization 2007 stage 3 co-occurrent with malaria |
| SNOMED CT | 735527002 | Human immunodeficiency virus World Health Organization 2007 stage 4 co-occurrent with tuberculosis |
| SNOMED CT | 735528007 | Human immunodeficiency virus World Health Organization 2007 stage 4 co-occurrent with malaria |
| SNOMED CT | 735536003 | Adult-onset immunodeficiency |

Continued on next page

**TableS31 – continued from previous page**

| CodeSystem | ConceptCode | ConceptName |
|---|---|---|
| SNOMED CT | 735599007 | Rheumatoid arthritis with erosion of joint |
| SNOMED CT | 735600005 | Rheumatoid arthritis without erosion |
| SNOMED CT | 736024007 | Pancytopenia caused by medication |
| SNOMED CT | 736322001 | Pediatric follicular lymphoma |
| SNOMED CT | 737223000 | Dendritic cell neoplasm |
| SNOMED CT | 737224006 | Histiocytic neoplasm |
| SNOMED CT | 737307003 | Natural-killer cell deficiency |
| SNOMED CT | 737378009 | World Health Organization 2007 Human immunodeficiency virus infection clinical stage 1 |
| SNOMED CT | 737379001 | World Health Organization 2007 Human immunodeficiency virus infection clinical stage 2 |
| SNOMED CT | 737380003 | World Health Organization 2007 Human immunodeficiency virus infection clinical stage 3 |
| SNOMED CT | 737381004 | World Health Organization 2007 Human immunodeficiency virus infection clinical stage 4 |
| SNOMED CT | 738527001 | Myeloid and/or lymphoid neoplasm associated with platelet derived growth factor receptor alpha rearrangement |
| SNOMED CT | 738770003 | Anaplastic lymphoma kinase positive anaplastic large cell lymphoma |
| SNOMED CT | 739301006 | Osteoporosis co-occurrent and due to multiple myeloma |
| SNOMED CT | 762315004 | Therapy related acute myeloid leukemia due to and following administration of antineoplastic agent |
| SNOMED CT | 762316003 | Polymorphic lymphoproliferative disorder following transplant |
| SNOMED CT | 762690000 | Classical Hodgkin lymphoma |
| SNOMED CT | 762691001 | Classical Hodgkin lymphoma |
| SNOMED CT | 763309005 | Acute myeloid leukemia with NPM1 somatic mutation |
| SNOMED CT | 763477007 | Primary lymphoma of conjunctiva |
| SNOMED CT | 763623001 | Severe combined immunodeficiency due to CTPS1 deficiency |
| SNOMED CT | 763666008 | Splenic marginal zone B-cell lymphoma |
| SNOMED CT | 763668009 | Lichtenstein syndrome |
| SNOMED CT | 763713000 | Idiopathic CD4 lymphocytopenia |
| SNOMED CT | 763719001 | Hydroa vacciniforme-like lymphoma |
| SNOMED CT | 763796007 | Megakaryoblastic acute myeloid leukemia with t(1;22)(p13;q13) |
| SNOMED CT | 763884007 | Splenic diffuse red pulp small B-cell lymphoma |
| SNOMED CT | 764855007 | Acute myeloid leukemia with CEBPA somatic mutations |
| SNOMED CT | 764858009 | Isolated agammaglobulinemia |
| SNOMED CT | 764940002 | Inherited acute myeloid leukemia |
| SNOMED CT | 764946008 | Constitutional mismatch repair deficiency syndrome |
| SNOMED CT | 765136002 | Primary cutaneous CD8 positive aggressive epidermotropic cytotoxic T-cell lymphoma |
| SNOMED CT | 765145001 | T cell negative B cell positive severe combined immunodeficiency due to gamma chain deficiency |
| SNOMED CT | 765188009 | Severe combined immunodeficiency due to complete RAG1 and/or RAG2 deficiency |
| SNOMED CT | 765327005 | Congenital sideroblastic anemia, B-cell immunodeficiency, periodic fever, developmental delay syndrome |
| SNOMED CT | 765328000 | Classic mycosis fungoides |

Continued on next page

**TableS31 – continued from previous page**

| CodeSystem | ConceptCode | ConceptName |
|---|---|---|
| SNOMED CT | 765748009 | Adult pure red cell aplasia |
| SNOMED CT | 766045006 | Acute myeloid leukemia and myelodysplastic syndrome related to alkylating agent |
| SNOMED CT | 766046007 | Acute myeloid leukemia and myelodysplastic syndrome related to topoisomerase type 2 inhibitor |
| SNOMED CT | 766048008 | Acute myeloid leukemia and myelodysplastic syndrome related to radiation |
| SNOMED CT | 766705006 | Immunodeficiency due to ficolin 3 deficiency |
| SNOMED CT | 766879006 | Combined immunodeficiency due to OX40 deficiency |
| SNOMED CT | 766935007 | Primary bone lymphoma |
| SNOMED CT | 766983005 | Susceptibility to respiratory infection associated with CD8alpha chain mutation |
| SNOMED CT | 767263007 | 22q11.2 deletion syndrome |
| SNOMED CT | 767658000 | Neutropenia due to and following chemotherapy |
| SNOMED CT | 768556005 | Ataxia pancytopenia syndrome |
| SNOMED CT | 768560008 | MDA5 deficiency |
| SNOMED CT | 770402000 | Aleukemic mast cell leukemia |
| SNOMED CT | 770596007 | Rippling muscle disease with myasthenia gravis |
| SNOMED CT | 770625006 | Combined immunodeficiency with faciooculoskeletal anomalies syndrome |
| SNOMED CT | 770687001 | Vasculitis due to adenosine deaminase 2 deficiency |
| SNOMED CT | 770785002 | T-cell immunodeficiency with epidermodysplasia verruciformis |
| SNOMED CT | 770942003 | Kostmann syndrome |
| SNOMED CT | 770947009 | Autosomal dominant severe congenital neutropenia |
| SNOMED CT | 771073006 | Lymphoproliferative disorder caused by methotrexate |
| SNOMED CT | 771078002 | Immunodeficiency due to MASP-2 deficiency |
| SNOMED CT | 771119002 | Infection caused by Coccidia co-occurrent with acquired immunodeficiency syndrome |
| SNOMED CT | 771126002 | Infection caused by Toxoplasma gondii co-occurrent with acquired immunodeficiency syndrome |
| SNOMED CT | 771127006 | Infection caused by Isospora co-occurrent with acquired immunodeficiency syndrome |
| SNOMED CT | 771309000 | Autosomal recessive lymphoproliferative disease |
| SNOMED CT | 771333006 | Immune dysregulation, inflammatory bowel disease, arthritis, recurrent infection syndrome |
| SNOMED CT | 771443008 | Complement component 3 deficiency |
| SNOMED CT | 771479000 | Combined immunodeficiency due to STK4 deficiency |
| SNOMED CT | 771515001 | Facial dysmorphism, immunodeficiency, livedo, short stature syndrome |
| SNOMED CT | 771517009 | Severe combined immunodeficiency due to CARD11 deficiency |
| SNOMED CT | 772126000 | Poikiloderma with neutropenia |
| SNOMED CT | 773333003 | Autosomal systemic lupus erythematosus |
| SNOMED CT | 773404000 | Roifman syndrome |
| SNOMED CT | 773488000 | Combined immunodeficiency due to MALT1 deficiency |
| SNOMED CT | 773537001 | Differentiation syndrome due to and following chemotherapy co-occurrent with acute promyelocytic leukemia |

Continued on next page

**TableS31 – continued from previous page**

| CodeSystem | ConceptCode | ConceptName |
|---|---|---|
| SNOMED CT | 773646003 | PLCG2-associated antibody deficiency and immune dysregulation |
| SNOMED CT | 773664005 | Deficiency in anterior pituitary function, variable immunodeficiency syndrome |
| SNOMED CT | 773702002 | Sterile multifocal osteomyelitis with periostitis and pustulosis |
| SNOMED CT | 773730002 | Osteopetrosis hypogammaglobulinemia syndrome |
| SNOMED CT | 773995001 | Primary cutaneous anaplastic large cell lymphoma |
| SNOMED CT | 774071007 | Pancytopenia with developmental delay syndrome |
| SNOMED CT | 774211005 | Severe dermatitis, multiple allergies, metabolic wasting syndrome |
| SNOMED CT | 775909002 | Congenital neutropenia, myelofibrosis, nephromegaly syndrome |
| SNOMED CT | 778004006 | Autoinflammation-PLCG2-associated antibody deficiency-immune dysregulation |
| SNOMED CT | 778023004 | Syndromic multisystem autoimmune disease due to ITCH deficiency |
| SNOMED CT | 778024005 | Monocytopenia with susceptibility to infections |
| SNOMED CT | 778027003 | Primary CD59 deficiency |
| SNOMED CT | 778028008 | Immunodeficiency due to CD25 deficiency |
| SNOMED CT | 778045003 | Susceptibility to viral and mycobacterial infection |
| SNOMED CT | 780817000 | Undifferentiated myeloproliferative disease |
| SNOMED CT | 780844005 | Acute myeloid leukemia with inv(3)(q21q26.2) or t(3;3)(q21;q26.2); RPN1-EVI1 |
| SNOMED CT | 782750002 | T-cell receptor alpha-beta-positive T-cell deficiency |
| SNOMED CT | 782751003 | Severe combined immunodeficiency due to IKK2 deficiency |
| SNOMED CT | 782759001 | X-linked dyserythropoietic anemia with abnormal platelets and neutropenia |
| SNOMED CT | 782915004 | Acquired hemophagocytic lymphohistiocytosis associated with malignant disease |
| SNOMED CT | 783007005 | Recurrent Neisseria infection due to factor D deficiency |
| SNOMED CT | 783017000 | Acute myeloid leukemia with BCR-ABL1 |
| SNOMED CT | 783058007 | Autosomal recessive severe congenital neutropenia due to G6PC3 deficiency |
| SNOMED CT | 783099001 | RIDDLE syndrome |
| SNOMED CT | 783142006 | Pancytopenia due to IKZF1 mutations |
| SNOMED CT | 783150002 | Iatrogenic immunodeficiency-associated lymphoproliferative disorder |
| SNOMED CT | 783199003 | Autosomal recessive severe congenital neutropenia due to JAGN1 deficiency |
| SNOMED CT | 783200000 | Autosomal recessive severe congenital neutropenia due to CXCR2 deficiency |
| SNOMED CT | 783201001 | Autosomal recessive severe congenital neutropenia due to CSF3R deficiency |
| SNOMED CT | 783220004 | Burkitt-like lymphoma with 11q aberration |
| SNOMED CT | 783245001 | Primary immunodeficiency with post-measles-mumps-rubella vaccine viral infection |
| SNOMED CT | 783248004 | Hyperimmunoglobulin M syndrome with susceptibility to opportunistic infection |

Continued on next page

**TableS31 – continued from previous page**

| CodeSystem | ConceptCode | ConceptName |
|---|---|---|
| SNOMED CT | 783249007 | Hyperimmunoglobulin M syndrome without susceptibility to opportunistic infection |
| SNOMED CT | 783255002 | Hereditary isolated aplastic anemia |
| SNOMED CT | 783263001 | Acute myeloid leukemia with mutated RUNX1 |
| SNOMED CT | 783541009 | Breast implant-associated anaplastic large-cell lymphoma |
| SNOMED CT | 783565007 | Indolent T-cell lymphoproliferative disorder of gastrointestinal tract |
| SNOMED CT | 783615009 | Erythropoietic uroporphyria associated with myeloid malignancy |
| SNOMED CT | 783617001 | Severe combined immunodeficiency due to LCK deficiency |
| SNOMED CT | 783621008 | Immunodeficiency with factor I anomaly |
| SNOMED CT | 783743009 | Combined immunodeficiency with granulomatosis |
| SNOMED CT | 783744003 | B-lymphoblastic leukemia lymphoma BCR-ABL1-like |
| SNOMED CT | 784339002 | Deficiency of interleukin 36 receptor antagonist |
| SNOMED CT | 784340000 | Combined immunodeficiency due to interleukin 21 receptor deficiency |
| SNOMED CT | 784392009 | Adult chronic idiopathic neutropenia |
| SNOMED CT | 784393004 | Adult-onset immunodeficiency with anti-interferon-gamma autoantibodies |
| SNOMED CT | 784551004 | Follicular T-cell lymphoma |
| SNOMED CT | 785825000 | B lymphoblastic leukemia lymphoma with iAMP21 |
| SNOMED CT | 786909001 | High grade B-cell lymphoma with MYC and BCL2 and/or BCL6 rearrangements |
| SNOMED CT | 786960000 | Large B-cell lymphoma with IRF4 rearrangement |
| SNOMED CT | 787036009 | Monomorphic epitheliotropic intestinal T-cell lymphoma |
| SNOMED CT | 787198005 | Primary cutaneous acral CD8 positive T-cell lymphoma |
| SNOMED CT | 787565006 | Diffuse large B-cell lymphoma activated B-cell subtype |
| SNOMED CT | 787594004 | Diffuse large B-cell lymphoma germinal center B-cell subtype |
| SNOMED CT | 787941004 | Myeloid and/or lymphoid neoplasm with PCM1-JAK2 |
| SNOMED CT | 788674000 | Primary cutaneous CD4 positive small/medium T-cell lymphoproliferative disorder |
| SNOMED CT | 788740009 | Acute myeloid leukemia with biallelic mutation of CEBPA (CCAAT enhancer binding protein alpha) gene |
| SNOMED CT | 788874003 | B-cell prolymphocytic leukemia in remission |
| SNOMED CT | 788972003 | Juvenile myelomonocytic leukemia in remission |
| SNOMED CT | 789435006 | Myelodysplastic syndrome with ring sideroblasts and multilineage dysplasia |
| SNOMED CT | 789689004 | Malignant lymphomatoid granulomatosis grade 3 |
| SNOMED CT | 789690008 | Malignant lymphomatoid granulomatosis grade 3 of lung |
| SNOMED CT | 789777007 | Short-limb skeletal dysplasia with severe combined immunodeficiency |
| SNOMED CT | 829973009 | Autoinflammatory syndrome with pyogenic bacterial infection and amylopectinosis |
| SNOMED CT | 830057003 | Relapsing classical Hodgkin lymphoma |
| SNOMED CT | 836276000 | Hodgkin sarcoma |
| SNOMED CT | 836277009 | Hodgkin granuloma |
| SNOMED CT | 836486002 | Lymphomatous infiltrate of kidney |

Continued on next page

**TableS31 – continued from previous page**

| CodeSystem | ConceptCode | ConceptName |
|---|---|---|
| SNOMED CT | 838340006 | B lymphoblastic leukemia lymphoma with t(5;14)(q31;q32); IL3-IGH |
| SNOMED CT | 838341005 | B lymphoblastic leukemia lymphoma with t(v;11q23); MLL rearranged |
| SNOMED CT | 838342003 | B lymphoblastic leukemia lymphoma with t(12;21) (p13;q22); TEL/AML1 (ETV6-RUNX1) |
| SNOMED CT | 838343008 | B lymphoblastic leukemia lymphoma with t(1;19)(Q23;P13.3); E2A-PBX1 (TCF3/PBX1) |
| SNOMED CT | 838344002 | B lymphoblastic leukemia lymphoma with hypodiploidy |
| SNOMED CT | 838346000 | B lymphoblastic leukemia lymphoma with hyperdiploidy |
| SNOMED CT | 838355002 | Acute myeloid leukemia with inv(16)(p13.1q22) or t(16;16)(p13.1;q22) CBFB-MYH11 |
| SNOMED CT | 838377003 | Chronic hepatitis C co-occurrent with human immunodeficiency virus infection |
| SNOMED CT | 840423002 | Diffuse large B-cell lymphoma of small intestine |
| SNOMED CT | 840424008 | Diffuse large B-cell lymphoma of stomach |
| SNOMED CT | 840442003 | Encephalitis caused by human immunodeficiency virus type 2 |
| SNOMED CT | 840472009 | Deficiency of immunoglobulin A with immunoglobulin G subclass deficiency |
| SNOMED CT | 840498003 | Encephalitis caused by human immunodeficiency virus type 1 |
| SNOMED CT | 866098005 | Large B-cell lymphoma arising in HHV8-associated multicentric Castleman disease |
| SNOMED CT | 866109006 | Lymphocytic hypereosinophilic syndrome |
| SNOMED CT | 871540009 | Post-polycythemia vera myelofibrosis |
| SNOMED CT | 871541008 | Post-essential thrombocythemia myelofibrosis |
| SNOMED CT | 81000119104 | Symptomatic human immunodeficiency virus infection |
| SNOMED CT | 12281000132104 | Relapsing acute myeloid leukemia |
| SNOMED CT | 12291000132102 | Refractory acute myeloid leukemia |
| SNOMED CT | 12301000132103 | Acute lymphoid leukemia relapse |
| SNOMED CT | 12311000132101 | Refractory acute lymphoid leukemia |
| SNOMED CT | 12341000132100 | B-cell lymphoma, unclassifiable, with features intermediate between diffuse large B-cell lymphoma and Burkitt lymphoma |
| SNOMED CT | 12351000132102 | B-cell lymphoma, unclassifiable, with features intermediate between diffuse large B-cell lymphoma and Hodgkin lymphoma |
| SNOMED CT | 12631000119106 | Immunoglobulin G deficiency |
| SNOMED CT | 61291000119103 | Disorder of central nervous system co-occurrent and due to acute lymphoid leukemia in remission |
| SNOMED CT | 61301000119102 | Disorder of central nervous system co-occurrent and due to acute lymphoid leukemia |
| SNOMED CT | 77461000119109 | Myasthenia gravis with exacerbation |
| SNOMED CT | 77471000119103 | Myasthenia gravis without exacerbation |
| SNOMED CT | 80191000119101 | Symptomatic human immunodeficiency virus (HIV) I infection |
| SNOMED CT | 90681000119107 | Asymptomatic human immunodeficiency virus A1 infection |
| SNOMED CT | 90691000119105 | Asymptomatic human immunodeficiency virus A2 infection |
| SNOMED CT | 116691000119101 | Marginal zone lymphoma of spleen |
| SNOMED CT | 116811000119106 | Non-Hodgkin lymphoma of central nervous system metastatic to lymph node of lower limb |

Continued on next page

**TableS31 – continued from previous page**

| CodeSystem | ConceptCode | ConceptName |
|---|---|---|
| SNOMED CT | 116821000119104 | Non-Hodgkin lymphoma of central nervous system metastatic to lymph node of upper limb |
| SNOMED CT | 116841000119105 | Marginal zone lymphoma of lymph nodes of multiple sites |
| SNOMED CT | 116871000119103 | Mantle cell lymphoma of lymph nodes of multiple sites |
| SNOMED CT | 117211000119105 | Peripheral T-cell lymphoma of lymph nodes of multiple sites |
| SNOMED CT | 118791000119106 | Aplastic anemia caused by antineoplastic agent |
| SNOMED CT | 121131000119109 | Chronic graft versus host disease after transplantation of bone marrow |
| SNOMED CT | 122491000119108 | Lymphoproliferative disorder after transplantation of bone marrow |
| SNOMED CT | 133751000119102 | Lymphoma of colon |
| SNOMED CT | 295101000119105 | Nephropathy co-occurrent and due to systemic lupus erythematosus |
| SNOMED CT | 295111000119108 | Nephrotic syndrome co-occurrent and due to systemic lupus erythematosus |
| SNOMED CT | 295121000119101 | Nephrosis co-occurrent and due to systemic lupus erythematosus |
| SNOMED CT | 319081000119109 | Seronegative rheumatoid arthritis of multiple joints |
| SNOMED CT | 319841000119107 | Rheumatoid lung disease with rheumatoid arthritis |
| SNOMED CT | 328301000119102 | Pancytopenia due to antineoplastic chemotherapy |
| SNOMED CT | 328611000119105 | Adult pulmonary Langerhans cell histiocytosis |
| SNOMED CT | 350951000119101 | B-cell lymphoma of intra-abdominal lymph nodes |
| SNOMED CT | 351211000119104 | B-cell lymphoma of lymph nodes of multiple sites |
| SNOMED CT | 352251000119109 | Small lymphocytic B-cell lymphoma of lymph nodes of multiple sites |
| SNOMED CT | 352411000119109 | Small lymphocytic B-cell lymphoma of intra-abdominal lymph nodes |
| SNOMED CT | 352791000119108 | Non-Hodgkin's lymphoma of lymph nodes of multiple sites |
| SNOMED CT | 354851000119101 | Follicular non-Hodgkin's lymphoma of lymph nodes of multiple sites |
| SNOMED CT | 1073731000119100 | Rheumatoid arthritis of left knee |
| SNOMED CT | 1073801000119100 | Rheumatoid arthritis of right hip |
| SNOMED CT | 10755671000119100 | Human immunodeficiency virus in mother complicating childbirth |
| SNOMED CT | 16623961000119100 | Pancytopenia caused by immunosuppressant |
| SNOMED CT | 1929004 | B-cell lymphoma, no ICD-O subtype |
| SNOMED CT | 3172003 | Peripheral T-cell lymphoma, no International Classification of Diseases for Oncology subtype |
| SNOMED CT | 4950009 | Sézary disease |
| SNOMED CT | 14317002 | AML (acute myeloid leukemia), M6 type |
| SNOMED CT | 14537002 | Hodgkin lymphoma, no International Classification of Diseases for Oncology subtype |
| SNOMED CT | 16893006 | [M]Hodgkin's disease, lymphocytic depletion, diffuse fibrosis |
| SNOMED CT | 17788007 | Acute myeloid leukemia, no International Classification of Diseases for Oncology subtype |
| SNOMED CT | 21964009 | Malignant lymphoma, no International Classification of Diseases for Oncology subtype |
| SNOMED CT | 22197008 | Burkitt cell leukaemia |

Continued on next page

**TableS31 – continued from previous page**

| CodeSystem | ConceptCode | ConceptName |
|---|---|---|
| SNOMED CT | 23719005 | Transfusion of bone marrow |
| SNOMED CT | 28054005 | Cutaneous lymphoma |
| SNOMED CT | 30575002 | Pancytopenia-dysmelia syndrome |
| SNOMED CT | 32280000 | Subacute lymphoid leukemia [obs] |
| SNOMED CT | 36070007 | Eczema, thrombocytopenia, immunodeficiency syndrome |
| SNOMED CT | 37810007 | Eosinophilic leukemia |
| SNOMED CT | 40411000 | Malignant lymphoma, lymphocytic, well differentiated, nodular [obs] |
| SNOMED CT | 46732000 | Angiotropic lymphoma |
| SNOMED CT | 46760003 | Estren-Dameshek anaemia |
| SNOMED CT | 50102004 | Malignant lymphoma, centroblastic-centrocytic, diffuse [obs] |
| SNOMED CT | 50926003 | Recurring cold staphylococcal abscesses |
| SNOMED CT | 53237008 | Large cell (Ki-1 positive) lymphoma |
| SNOMED CT | 55602000 | Congenital thymic dysplasia syndrome |
| SNOMED CT | 57160007 | Felty syndrome |
| SNOMED CT | 65399007 | Differentiated progressive histiocytosis |
| SNOMED CT | 65880007 | X linked agammaglobulinemia |
| SNOMED CT | 70600005 | Hodgkin paragranuloma, nodular [obs] |
| SNOMED CT | 71109004 | [M]Hodgkin's disease, lymphocytic depletion, reticular type |
| SNOMED CT | 74654000 | Mantle zone lymphoma [obs] |
| SNOMED CT | 77121009 | Familial fatal Epstein-Barr infection |
| SNOMED CT | 77381001 | Burkitt's tumor |
| SNOMED CT | 77430005 | [M]Adult T-cell leukemia/lymphoma |
| SNOMED CT | 79268002 | Polyneuropathy, organomegaly, endocrinopathy, monoclonal gammopathy, and skin changes syndrome |
| SNOMED CT | 82286005 | Immunodeficiency with IgM hypergammaglobulinemia |
| SNOMED CT | 87163000 | Leukemia, no International Classification of Diseases for Oncology subtype |
| SNOMED CT | 92511007 | Burkitt's tumor of lymph nodes of axilla and upper limb |
| SNOMED CT | 92512000 | Burkitt's tumor of lymph nodes of head, face, and neck |
| SNOMED CT | 92513005 | Burkitt's tumor of lymph nodes of inguinal region and lower limb |
| SNOMED CT | 92516002 | Burkitt's tumor of unspecified, extranodal, or solid organ site |
| SNOMED CT | 93451002 | Erythroleukaemia, FAB M6 |
| SNOMED CT | 93487009 | Hodgkin disease, lymphocytic depletion of lymph nodes of axilla AND/OR upper limb |
| SNOMED CT | 93488004 | Hodgkin disease, lymphocytic depletion of lymph nodes of head, face AND/OR neck |
| SNOMED CT | 93489007 | Hodgkin disease, lymphocytic depletion of lymph nodes of inguinal region AND/OR lower limb |
| SNOMED CT | 93492006 | Hodgkin disease, lymphocytic depletion of extranodal AND/OR solid organ site |
| SNOMED CT | 93493001 | Hodgkin's paragranuloma of intra-abdominal lymph nodes |
| SNOMED CT | 93494007 | Hodgkin disease, lymphocytic-histiocytic predominance of intrapelvic lymph nodes |
| SNOMED CT | 93495008 | Hodgkin disease, lymphocytic-histiocytic predominance of intrathoracic lymph nodes |

Continued on next page

**TableS31 – continued from previous page**

| CodeSystem | ConceptCode | ConceptName |
|---|---|---|
| SNOMED CT | 93496009 | Hodgkin disease, lymphocytic-histiocytic predominance of lymph nodes of axilla AND/OR upper limb |
| SNOMED CT | 93497000 | Hodgkin disease, lymphocytic-histiocytic predominance of lymph nodes of head, face AND/OR neck |
| SNOMED CT | 93498005 | Hodgkin disease, lymphocytic-histiocytic predominance of lymph nodes of inguinal region AND/OR lower limb |
| SNOMED CT | 93500006 | Hodgkin paragranuloma of spleen |
| SNOMED CT | 93501005 | Hodgkin disease, lymphocytic-histiocytic predominance of extranodal AND/OR solid organ site |
| SNOMED CT | 93505001 | Hodgkin disease, mixed cellularity of lymph nodes of axilla AND/OR upper limb |
| SNOMED CT | 93506000 | Hodgkin disease, mixed cellularity of lymph nodes of head, face AND/OR neck |
| SNOMED CT | 93507009 | Hodgkin disease, mixed cellularity of lymph nodes of inguinal region AND/OR lower limb |
| SNOMED CT | 93509007 | Hodgkin disease, mixed cellularity of spleen |
| SNOMED CT | 93510002 | Hodgkin disease, mixed cellularity of extranodal AND/OR solid organ site |
| SNOMED CT | 93514006 | Hodgkin disease, nodular sclerosis of lymph nodes of axilla AND/OR upper limb |
| SNOMED CT | 93515007 | Hodgkin disease, nodular sclerosis of lymph nodes of head, face AND/OR neck |
| SNOMED CT | 93516008 | Hodgkin disease, nodular sclerosis of lymph nodes of inguinal region AND/OR lower limb |
| SNOMED CT | 93518009 | Hodgkin disease, nodular sclerosis of spleen |
| SNOMED CT | 93519001 | Hodgkin disease, nodular sclerosis of extranodal AND/OR solid organ site |
| SNOMED CT | 93520007 | Hodgkin disease of intra-abdominal lymph nodes |
| SNOMED CT | 93521006 | Hodgkin disease of intrapelvic lymph nodes |
| SNOMED CT | 93522004 | Hodgkin disease of intrathoracic lymph nodes |
| SNOMED CT | 93523009 | Hodgkin disease of lymph nodes of axilla AND/OR upper limb |
| SNOMED CT | 93524003 | Hodgkin disease of lymph nodes of head, face AND/OR neck |
| SNOMED CT | 93525002 | Hodgkin disease of lymph nodes of inguinal region AND/OR lower limb |
| SNOMED CT | 93526001 | Hodgkin disease of lymph nodes of multiple sites |
| SNOMED CT | 93527005 | Hodgkin disease of spleen |
| SNOMED CT | 93528000 | Hodgkin disease of extranodal AND/OR solid organ site |
| SNOMED CT | 93530003 | Hodgkin granuloma of intrapelvic lymph nodes |
| SNOMED CT | 93531004 | Hodgkin granuloma of intrathoracic lymph nodes |
| SNOMED CT | 93532006 | Hodgkin granuloma of lymph nodes of axilla AND/OR upper limb |
| SNOMED CT | 93533001 | Hodgkin granuloma of lymph nodes of head, face AND/OR neck |
| SNOMED CT | 93534007 | Hodgkin granuloma of lymph nodes of inguinal region AND/OR lower limb |
| SNOMED CT | 93536009 | Hodgkin granuloma of spleen |
| SNOMED CT | 93537000 | Hodgkin granuloma of extranodal AND/OR solid organ site |

Continued on next page

**TableS31 – continued from previous page**

| CodeSystem | ConceptCode | ConceptName |
|---|---|---|
| SNOMED CT | 93541001 | Hodgkin paragranuloma of lymph nodes of axilla AND/OR upper limb |
| SNOMED CT | 93542008 | Hodgkin paragranuloma of lymph nodes of head, face AND/OR neck |
| SNOMED CT | 93543003 | Hodgkin paragranuloma of lymph nodes of inguinal region AND/OR lower limb |
| SNOMED CT | 93546006 | Hodgkin paragranuloma of extranodal AND/OR solid organ site |
| SNOMED CT | 93547002 | Hodgkin sarcoma of intra-abdominal lymph nodes |
| SNOMED CT | 93548007 | Hodgkin sarcoma of intrapelvic lymph nodes |
| SNOMED CT | 93549004 | Hodgkin sarcoma of intrathoracic lymph nodes |
| SNOMED CT | 93550004 | Hodgkin sarcoma of lymph nodes of axilla AND/OR upper limb |
| SNOMED CT | 93551000 | Hodgkin sarcoma of lymph nodes of head, face AND/OR neck |
| SNOMED CT | 93552007 | Hodgkin sarcoma of lymph nodes of inguinal region AND/OR lower limb |
| SNOMED CT | 93554008 | Hodgkin sarcoma of spleen |
| SNOMED CT | 93555009 | Hodgkin sarcoma of extranodal AND/OR solid organ site |
| SNOMED CT | 94719007 | Granulocytic sarcoma |
| SNOMED CT | 103686008 | Enteropathy associated T-cell lymphoma |
| SNOMED CT | 109391009 | Kaposi sarcoma of lymph nodes |
| SNOMED CT | 109962001 | Nonfollicular lymphoma |
| SNOMED CT | 109964000 | Diffuse non-Hodgkin lymphoma, undifferentiated |
| SNOMED CT | 109965004 | Lymphoblastoma |
| SNOMED CT | 109966003 | Malignant lymphoma - immunoblastic |
| SNOMED CT | 109967007 | Diffuse non-Hodgkin lymphoma, small cleaved cell |
| SNOMED CT | 109968002 | Diffuse non-Hodgkin lymphoma, small cell |
| SNOMED CT | 109969005 | DLBCL - diffuse large B cell lymphoma |
| SNOMED CT | 109970006 | Follicular lymphoma, grade 1 |
| SNOMED CT | 109971005 | Follicular lymphoma grade 2 |
| SNOMED CT | 109972003 | Follicular lymphoma grade 3 |
| SNOMED CT | 109977009 | Peripheral T-cell lymphoma pleomorphic small cell |
| SNOMED CT | 109979007 | B-cell lymphoma |
| SNOMED CT | 109983007 | Monoclonal paraproteinemia |
| SNOMED CT | 110004001 | Acute promyelocytic leukemia (clinical) |
| SNOMED CT | 110005000 | Acute myelomonocytic leukemia (clinical) |
| SNOMED CT | 110007008 | Adult T-cell leukemia |
| SNOMED CT | 111396008 | Hereditary gigantism of cytoplasmic organelles |
| SNOMED CT | 118599009 | Lymphoma, Hodgkins |
| SNOMED CT | 118600007 | Malignant lymphoma (clinical) |
| SNOMED CT | 118601006 | Non-Hodgkin lymphoma |
| SNOMED CT | 118602004 | Hodgkin granuloma |
| SNOMED CT | 118605002 | Hodgkin lymphoma, nodular lymphocyte predominance |
| SNOMED CT | 118606001 | Hodgkin sarcoma |
| SNOMED CT | 118608000 | Hodgkin disease, nodular sclerosis |
| SNOMED CT | 118609008 | Hodgkin mixed cellularity lymphoma |
| SNOMED CT | 118610003 | Hodgkin disease, lymphocytic depletion |
| SNOMED CT | 118613001 | Leukemic reticuloendotheliosis |
| SNOMED CT | 118614007 | Acute infancy reticulosis |

Continued on next page

**TableS31 – continued from previous page**

| CodeSystem | ConceptCode | ConceptName |
|---|---|---|
| SNOMED CT | 118617000 | Burkitt lymphoma |
| SNOMED CT | 128799007 | Classical Hodgkin lymphoma, lymphocyte-rich |
| SNOMED CT | 128805001 | T/NK-cell lymphoma, nasal and nasal type |
| SNOMED CT | 128809007 | Langerhans cell granulomatosis |
| SNOMED CT | 128812005 | Langerhans cell granulomatosis |
| SNOMED CT | 128819001 | T-cell large granular lymphocytic leukaemia |
| SNOMED CT | 128821006 | T-cell prolymphocytic leukemia |
| SNOMED CT | 128833001 | Aggressive natural killer-cell leukemia |
| SNOMED CT | 128875000 | Primary cutaneous CD30 antigen positive large T-cell lymphoma |
| SNOMED CT | 128929007 | Non-Hodgkin lymphoma - category |
| SNOMED CT | 128930002 | Hodgkin lymphoma - category |
| SNOMED CT | 188510001 | Burkitt lymphoma of lymph nodes of head, face and neck |
| SNOMED CT | 188511002 | Burkitt tumor of intrathoracic lymph nodes |
| SNOMED CT | 188512009 | Burkitt tumor of intra-abdominal lymph nodes |
| SNOMED CT | 188513004 | Burkitt lymphoma of lymph nodes of axilla and upper limb |
| SNOMED CT | 188514005 | Burkitt lymphoma of lymph nodes of inguinal region and lower limb |
| SNOMED CT | 188515006 | Burkitt tumor of intrapelvic lymph nodes |
| SNOMED CT | 188516007 | Burkitt tumor of spleen |
| SNOMED CT | 188517003 | Burkitt tumor of lymph nodes of multiple sites |
| SNOMED CT | 188524002 | Hodgkin paragranuloma of intrathoracic lymph nodes |
| SNOMED CT | 188529007 | Hodgkin paragranuloma of intrapelvic lymph nodes |
| SNOMED CT | 188531003 | Hodgkin paragranuloma of lymph nodes of multiple sites |
| SNOMED CT | 188534006 | Hodgkin granuloma of lymph nodes of head, face and neck |
| SNOMED CT | 188536008 | Hodgkin granuloma of intra-abdominal lymph nodes |
| SNOMED CT | 188537004 | Hodgkin granuloma of lymph nodes of axilla and upper limb |
| SNOMED CT | 188538009 | Hodgkin granuloma of lymph nodes of inguinal region and lower limb |
| SNOMED CT | 188541000 | Hodgkin granuloma of lymph nodes of multiple sites |
| SNOMED CT | 188544008 | Hodgkin sarcoma of lymph nodes of head, face and neck |
| SNOMED CT | 188547001 | Hodgkin sarcoma of lymph nodes of axilla and upper limb |
| SNOMED CT | 188548006 | Hodgkin sarcoma of lymph nodes of inguinal region and lower limb |
| SNOMED CT | 188551004 | Hodgkin sarcoma of lymph nodes of multiple sites |
| SNOMED CT | 188554007 | Hodgkin disease, lymphocytic-histiocytic predominance of lymph nodes of head, face and neck |
| SNOMED CT | 188558005 | Hodgkin disease, lymphocytic-histiocytic predominance of lymph nodes of axilla and upper limb |
| SNOMED CT | 188559002 | Hodgkin disease, lymphocytic-histiocytic predominance of lymph nodes of inguinal region and lower limb |
| SNOMED CT | 188562004 | Hodgkin disease, lymphocytic-histiocytic predominance of lymph nodes of multiple sites |
| SNOMED CT | 188565002 | Hodgkin disease, nodular sclerosis of lymph nodes of head, face and neck |
| SNOMED CT | 188566001 | Hodgkin disease, nodular sclerosis of intrathoracic lymph nodes |
| SNOMED CT | 188567005 | Hodgkin disease, nodular sclerosis of intra-abdominal lymph nodes |

Continued on next page

**TableS31 – continued from previous page**

| CodeSystem | ConceptCode | ConceptName |
|---|---|---|
| SNOMED CT | 188568000 | Hodgkin disease, nodular sclerosis of lymph nodes of axilla and upper limb |
| SNOMED CT | 188569008 | Hodgkin disease, nodular sclerosis of lymph nodes of inguinal region and lower limb |
| SNOMED CT | 188570009 | Hodgkin disease, nodular sclerosis of intrapelvic lymph nodes |
| SNOMED CT | 188572001 | Hodgkin disease, nodular sclerosis of lymph nodes of multiple sites |
| SNOMED CT | 188575004 | Hodgkin disease, mixed cellularity of lymph nodes of head, face and neck |
| SNOMED CT | 188576003 | Hodgkin disease, mixed cellularity of intrathoracic lymph nodes |
| SNOMED CT | 188577007 | Hodgkin disease, mixed cellularity of intra-abdominal lymph nodes |
| SNOMED CT | 188578002 | Hodgkin disease, mixed cellularity of lymph nodes of axilla and upper limb |
| SNOMED CT | 188579005 | Hodgkin disease, mixed cellularity of lymph nodes of inguinal region and lower limb |
| SNOMED CT | 188580008 | Hodgkin disease, mixed cellularity of intrapelvic lymph nodes |
| SNOMED CT | 188582000 | Hodgkin disease, mixed cellularity of lymph nodes of multiple sites |
| SNOMED CT | 188585003 | Hodgkin disease, lymphocytic depletion of lymph nodes of head, face and neck |
| SNOMED CT | 188586002 | Hodgkin disease, lymphocytic depletion of intrathoracic lymph nodes |
| SNOMED CT | 188587006 | Hodgkin disease, lymphocytic depletion of intra-abdominal lymph nodes |
| SNOMED CT | 188589009 | Hodgkin disease, lymphocytic depletion of lymph nodes of axilla and upper limb |
| SNOMED CT | 188590000 | Hodgkin disease, lymphocytic depletion of lymph nodes of inguinal region and lower limb |
| SNOMED CT | 188591001 | Hodgkin disease, lymphocytic depletion of intrapelvic lymph nodes |
| SNOMED CT | 188592008 | Hodgkin disease, lymphocytic depletion of spleen |
| SNOMED CT | 188593003 | Hodgkin disease, lymphocytic depletion of lymph nodes of multiple sites |
| SNOMED CT | 188632001 | Lymphoma intra abdominal, Sezarys |
| SNOMED CT | 188672005 | Follicular non-Hodgkin mixed small cleaved and large cell lymphoma |
| SNOMED CT | 188679001 | Diffuse non-Hodgkin lymphoma undifferentiated (diffuse) |
| SNOMED CT | 240531002 | African Burkitt lymphoma |
| SNOMED CT | 277550009 | Richter syndrome |
| SNOMED CT | 277609007 | Hodgkin disease, lymphocytic predominance - diffuse |
| SNOMED CT | 277610002 | Hodgkin disease, nodular sclerosis - lymphocytic predominance |
| SNOMED CT | 277611003 | Hodgkin disease, nodular sclerosis - mixed cellularity |
| SNOMED CT | 277612005 | Hodgkin disease, nodular sclerosis - lymphocytic depletion |
| SNOMED CT | 277654008 | Enteropathy-type T-cell lymphoma |
| SNOMED CT | 278257006 | Peripheral blood stem cell transplantation |
| SNOMED CT | 285420006 | Immunoglobulin A myeloma |

Continued on next page

**TableS31 – continued from previous page**

| CodeSystem | ConceptCode | ConceptName |
|---|---|---|
| SNOMED CT | 285421005 | Immunoglobulin G myeloma |
| SNOMED CT | 285422003 | Immunoglobulin D myeloma |
| SNOMED CT | 307633009 | Hodgkin disease, lymphocytic depletion, diffuse fibrosis |
| SNOMED CT | 307634003 | Hodgkin disease, lymphocytic depletion, reticular type |
| SNOMED CT | 307635002 | Hodgkin disease, nodular sclerosis - cellular phase |
| SNOMED CT | 308121000 | Follicular non-Hodgkin lymphoma |
| SNOMED CT | 315019000 | Human immunodeficiency virus infection with aseptic meningitis |
| SNOMED CT | 350353007 | Immunodeficiency with generalized hematopoietic hypoplasia |
| SNOMED CT | 359631009 | M0 - Acute myeloblastic leukemia - undifferentiated |
| SNOMED CT | 359640008 | Acute myelogenous leukemia without maturation |
| SNOMED CT | 359648001 | M2 - Acute myeloblastic leukemia with maturation |
| SNOMED CT | 362993009 | Autosomal recessive SCID (severe combined immunodeficiency disease) |
| SNOMED CT | 373381004 | Myelodysplastic syndrome, unclassifiable |
| SNOMED CT | 402881008 | Cutaneous B-cell lymphoma |
| SNOMED CT | 403835002 | X-linked hyper-immunoglobulin M syndrome |
| SNOMED CT | 404106004 | Lymphomatoid papulosis with Hodgkin disease |
| SNOMED CT | 404122003 | Leukaemic infiltration of skin (chronic T-cell lymphocytic leukaemia) |
| SNOMED CT | 404123008 | Leukaemic infiltration of skin (T-cell prolymphocytic leukaemia) |
| SNOMED CT | 404124002 | Leukaemic infiltration of skin (T-cell lymphoblastic leukaemia) |
| SNOMED CT | 404135007 | Angiocentric lymphoma involving skin |
| SNOMED CT | 404136008 | Aggressive natural killer-cell leukemia involving skin |
| SNOMED CT | 404143002 | Crosti's lymphoma |
| SNOMED CT | 404157000 | Specific skin infiltration in Hodgkin disease |
| SNOMED CT | 413527004 | Anaplastic large cell lymphoma, T- and null cell primary systemic types |
| SNOMED CT | 414166008 | Extranodal natural killer/T-cell lymphoma, nasal type |
| SNOMED CT | 414780005 | Mucosa-associated lymphoid tissue lymphoma of orbit |
| SNOMED CT | 416729007 | Neutropenia with AIDS (acquired immunodeficiency syndrome) |
| SNOMED CT | 419094004 | Immunodeficiency-associated Burkitt lymphoma |
| SNOMED CT | 419770008 | Endemic Burkitt lymphoma |
| SNOMED CT | 419879004 | Atypical Burkitt lymphoma |
| SNOMED CT | 420063007 | Sporadic Burkitt lymphoma |
| SNOMED CT | 420244003 | Encephalitis with AIDS (acquired immunodeficiency syndrome) |
| SNOMED CT | 420281004 | Skin rash with AIDS (acquired immunodeficiency syndrome) |
| SNOMED CT | 420302007 | Reticulosarcoma with AIDS (acquired immunodeficiency syndrome) |
| SNOMED CT | 420308006 | Retinal vascular changes with AIDS (acquired immunodeficiency syndrome) |
| SNOMED CT | 420321004 | Intestinal malabsorption with AIDS (acquired immunodeficiency syndrome) |
| SNOMED CT | 420403001 | Pneumocystosis with AIDS (acquired immunodeficiency syndrome) |
| SNOMED CT | 420452002 | Myelopathy with AIDS (acquired immunodeficiency syndrome) |
| SNOMED CT | 420543008 | Anemia associated with acquired immunodeficiency syndrome |

Continued on next page

**TableS31 – continued from previous page**

| CodeSystem | ConceptCode | ConceptName |
|---|---|---|
| SNOMED CT | 420544002 | Bacterial pneumonia with AIDS (acquired immunodeficiency syndrome) |
| SNOMED CT | 420549007 | Salivary gland disease with AIDS (acquired immunodeficiency syndrome) |
| SNOMED CT | 420554003 | Progressive multifocal leukoencephalopathy with AIDS (acquired immunodeficiency syndrome) |
| SNOMED CT | 420614009 | Organic dementia with AIDS (acquired immunodeficiency syndrome) |
| SNOMED CT | 420658009 | Radiculitis with AIDS (acquired immunodeficiency syndrome) |
| SNOMED CT | 420687005 | Ill-defined intestinal infection associated with acquired immunodeficiency syndrome |
| SNOMED CT | 420691000 | Nutritional deficiency with AIDS (acquired immunodeficiency syndrome) |
| SNOMED CT | 420718004 | Central nervous system demyelinating disease with AIDS (acquired immunodeficiency syndrome) |
| SNOMED CT | 420721002 | Acquired immunodeficiency syndrome-associated disorder |
| SNOMED CT | 420764009 | Salmonella infection with AIDS (acquired immunodeficiency syndrome) |
| SNOMED CT | 420774007 | Organic brain syndrome with AIDS (acquired immunodeficiency syndrome) |
| SNOMED CT | 420787001 | Pneumococcal pneumonia with AIDS (acquired immunodeficiency syndrome) |
| SNOMED CT | 420801006 | Malaise with AIDS (acquired immunodeficiency syndrome) |
| SNOMED CT | 420818005 | Mycobacteriosis with AIDS (acquired immunodeficiency syndrome) |
| SNOMED CT | 420877009 | Dermatomycosis with AIDS (acquired immunodeficiency syndrome) |
| SNOMED CT | 420900006 | Fatigue with AIDS (acquired immunodeficiency syndrome) |
| SNOMED CT | 420938005 | Subacute endocarditis with AIDS (acquired immunodeficiency syndrome) |
| SNOMED CT | 420945005 | Histoplasmosis with AIDS (acquired immunodeficiency syndrome) |
| SNOMED CT | 421023003 | Presenile dementia with AIDS (acquired immunodeficiency syndrome) |
| SNOMED CT | 421047005 | Candidiasis of lung with AIDS (acquired immunodeficiency syndrome) |
| SNOMED CT | 421102007 | Aplastic anemia with AIDS (acquired immunodeficiency syndrome) |
| SNOMED CT | 421230000 | Hepatomegaly with AIDS (acquired immunodeficiency syndrome) |
| SNOMED CT | 421272004 | Subacute myocarditis with AIDS (acquired immunodeficiency syndrome) |
| SNOMED CT | 421283008 | Primary lymphoma of brain with AIDS (acquired immunodeficiency syndrome) |
| SNOMED CT | 421312009 | Agranulocytosis with AIDS (acquired immunodeficiency syndrome) |
| SNOMED CT | 421315006 | Myelitis with AIDS (acquired immunodeficiency syndrome) |

Continued on next page

**TableS31 – continued from previous page**

| CodeSystem | ConceptCode | ConceptName |
|---|---|---|
| SNOMED CT | 421394009 | Skin disorder with AIDS (acquired immunodeficiency syndrome) |
| SNOMED CT | 421403008 | Cryptococcosis with AIDS (acquired immunodeficiency syndrome) |
| SNOMED CT | 421415007 | Subacute adenoviral encephalitis with AIDS (acquired immunodeficiency syndrome) |
| SNOMED CT | 421418009 | Mature T-cell AND/OR natural killer cell neoplasm |
| SNOMED CT | 421431004 | Nocardiosis with AIDS (acquired immunodeficiency syndrome) |
| SNOMED CT | 421454008 | Infectious gastroenteritis with AIDS (acquired immunodeficiency syndrome) |
| SNOMED CT | 421460008 | Retinopathy with AIDS (acquired immunodeficiency syndrome) |
| SNOMED CT | 421508002 | Viral pneumonia with AIDS (acquired immunodeficiency syndrome) |
| SNOMED CT | 421529006 | Dementia with AIDS (acquired immunodeficiency syndrome) |
| SNOMED CT | 421571007 | Tuberculosis with AIDS (acquired immunodeficiency syndrome) |
| SNOMED CT | 421597001 | Polyneuropathy with AIDS (acquired immunodeficiency syndrome) |
| SNOMED CT | 421660003 | Failure to thrive in infant with AIDS (acquired immunodeficiency syndrome) |
| SNOMED CT | 421671002 | Pneumonia with AIDS (acquired immunodeficiency syndrome) |
| SNOMED CT | 421695000 | Abnormal weight loss associated with acquired immunodeficiency syndrome |
| SNOMED CT | 421706001 | Blindness with AIDS (acquired immunodeficiency syndrome) |
| SNOMED CT | 421708000 | Infective arthritis with AIDS (acquired immunodeficiency syndrome) |
| SNOMED CT | 421710003 | Candidiasis of mouth with AIDS (acquired immunodeficiency syndrome) |
| SNOMED CT | 421766003 | Thrombocytopenia with AIDS (acquired immunodeficiency syndrome) |
| SNOMED CT | 421827003 | Encephalopathy with AIDS (acquired immunodeficiency syndrome) |
| SNOMED CT | 421851008 | Acquired hemolytic anemia associated with acquired immunodeficiency syndrome |
| SNOMED CT | 421874007 | Respiratory disorder with AIDS (acquired immunodeficiency syndrome) |
| SNOMED CT | 421883002 | Strongyloidiasis with AIDS (acquired immunodeficiency syndrome) |
| SNOMED CT | 421929001 | Myocarditis with AIDS (acquired immunodeficiency syndrome) |
| SNOMED CT | 421983003 | Noninfectious gastroenteritis with AIDS (acquired immunodeficiency syndrome) |
| SNOMED CT | 421998001 | Central nervous disorder with AIDS (acquired immunodeficiency syndrome) |
| SNOMED CT | 422003001 | Slim disease |
| SNOMED CT | 422012004 | Neuritis with AIDS (acquired immunodeficiency syndrome) |
| SNOMED CT | 422089004 | Encephalomyelitis with AIDS (acquired immunodeficiency syndrome) |
| SNOMED CT | 422136003 | Neuralgia with AIDS (acquired immunodeficiency syndrome) |
| SNOMED CT | 422177004 | Dyspnea with AIDS (acquired immunodeficiency syndrome) |

Continued on next page

**TableS31 – continued from previous page**

| CodeSystem | ConceptCode | ConceptName |
|---|---|---|
| SNOMED CT | 422189002 | Low vision with AIDS (acquired immunodeficiency syndrome) |
| SNOMED CT | 422194002 | Hyperhidrosis with AIDS (acquired immunodeficiency syndrome) |
| SNOMED CT | 422282000 | Malignant neoplasm with AIDS (acquired immunodeficiency syndrome) |
| SNOMED CT | 422337001 | Coccidioidomycosis with AIDS (acquired immunodeficiency syndrome) |
| SNOMED CT | 423294001 | Idiopathic hypereosinophilic syndrome (HES) |
| SNOMED CT | 445269007 | Extranodal marginal zone B-cell lymphoma of mucosa-associated lymphoid tissue |
| SNOMED CT | 450949002 | B lymphoblastic leukemia lymphoma, no International Classification of Diseases for Oncology subtype |
| SNOMED CT | 720345008 | T-cell immunodeficiency, congenital alopecia, and nail dystrophy |
| SNOMED CT | 733627006 | Primary cutaneous gamma-delta T-cell lymphoma |
| SNOMED CT | 781206002 | Rheumatoid spondylitis |
| SNOMED CT | 783787000 | Retinal vasculopathy with cerebral leucoencephalopathy and systemic manifestations |
| SNOMED CT | 1003381002 | Onycho-tricho-dysplasia neutropenia syndrome |
| SNOMED CT | 1003473002 | B cell lymphocyte aplasia caused by drug |
| SNOMED CT | 1003679009 | Human immunodeficiency virus Centers for Disease Control and Prevention dual infection with HIV-1 and HIV-2 |
| SNOMED CT | 1003680007 | Human immunodeficiency virus Centers for Disease Control and Prevention undifferentiated type |
| SNOMED CT | 1003682004 | Human immunodeficiency virus Centers for Disease Control and Prevention stage 1 |
| SNOMED CT | 1003683009 | Human immunodeficiency virus Centers for Disease Control and Prevention stage 2 |
| SNOMED CT | 1003684003 | Human immunodeficiency virus Centers for Disease Control and Prevention stage 3 |
| SNOMED CT | 1004006009 | World Health Organization human immunodeficiency virus infection clinical stage |
| SNOMED CT | 143291000119104 | Non-Hodgkin's lymphoma in remission |
| SNOMED CT | 318861000119101 | Seropositive rheumatoid arthritis of left hand |
| SNOMED CT | 318931000119100 | Seropositive rheumatoid arthritis of right hand |
| SNOMED CT | 878857009 | Malignant lymphomatoid granulomatosis grade 3 |
| SNOMED CT | 878858004 | Malignant lymphomatoid granulomatosis grade 3 of lung |
| SNOMED CT | 898197001 | Adult T-cell leukemia/lymphoma in remission |
| SNOMED CT | 898198006 | T-cell prolymphocytic leukemia in remission |
| SNOMED CT | 262945004 | Transection of transplanted artery |
| SNOMED CT | 266451002 | Allergic gastroenteritis and colitis |
| SNOMED CT | 267454002 | Acatalasemia |
| SNOMED CT | 267800008 | Benign mucous membrane pemphigoid with no eye involvement |
| SNOMED CT | 272006008 | Gonococcal arthritis dermatitis syndrome |
| SNOMED CT | 274083001 | Tinea kerion of beard |
| SNOMED CT | 275446004 | Gardner-Diamond syndrome |
| SNOMED CT | 276177000 | Thyroid eye disease |
| SNOMED CT | 276229008 | Sepsis-associated gastrointestinal hemorrhage |

Continued on next page

**TableS31 – continued from previous page**

| CodeSystem | ConceptCode | ConceptName |
|---|---|---|
| SNOMED CT | 276288002 | Recurrent salmonella septicemia |
| SNOMED CT | 276575001 | Autoimmune neonatal thrombocytopenia |
| SNOMED CT | 276576000 | Neutropenia of the small for gestational age baby |
| SNOMED CT | 276669000 | Bacterial sepsis of newborn |
| SNOMED CT | 276741000 | Sepsis-associated myocardial dysfunction |
| SNOMED CT | 277010001 | Unexplained episode of renal transplant dysfunction |
| SNOMED CT | 277011002 | Pre-existing disease in renal transplant |
| SNOMED CT | 277475006 | Pemphigus neonatorum |
| SNOMED CT | 277577000 | Monoclonal gammopathy of uncertain significance |
| SNOMED CT | 277638005 | Sepsis-associated left ventricular failure |
| SNOMED CT | 277639002 | Sepsis-associated right ventricular failure |
| SNOMED CT | 277785006 | Penicillin-induced angioedema-urticaria |
| SNOMED CT | 278484009 | Tropical pulmonary eosinophilia |
| SNOMED CT | 282093000 | Allergic reaction to venom |
| SNOMED CT | 282094006 | Allergic reaction to insect venom |
| SNOMED CT | 282095007 | Allergic reaction to bee sting |
| SNOMED CT | 285430002 | Light chain monoclonal gammopathy of uncertain significance |
| SNOMED CT | 286947004 | Chronic rheumatic mitral valvulitis |
| SNOMED CT | 295046003 | Ulcerative proctocolitis |
| SNOMED CT | 297940005 | Drug-induced erythema nodosum |
| SNOMED CT | 297942002 | Drug-induced erythema multiforme |
| SNOMED CT | 298285004 | Systemic sclerosis with limited cutaneous involvement |
| SNOMED CT | 299276009 | Limited systemic sclerosis |
| SNOMED CT | 300909004 | Allergic reaction to wasp sting |
| SNOMED CT | 300985007 | Coliform septicemia |
| SNOMED CT | 300986008 | Proteus septicemia |
| SNOMED CT | 303011007 | Neutropenic disorder |
| SNOMED CT | 307591004 | Malignant mastocytosis |
| SNOMED CT | 309742004 | Drug-induced autoimmune hemolytic anemia |
| SNOMED CT | 310649002 | Vancomycin resistant enterococcal septicemia |
| SNOMED CT | 310669007 | Septicemia due to enterococcus |
| SNOMED CT | 312591002 | Acute rheumatic heart disease |
| SNOMED CT | 312603000 | Lung transplant rejection |
| SNOMED CT | 313437008 | Meningococcal meningitis with acute meningococcal septicemia |
| SNOMED CT | 314002005 | Corneal allograft rejection |
| SNOMED CT | 314130008 | Meningococcal meningitis with meningococcal septicemia |
| SNOMED CT | 314498009 | Contact lens related giant papillary conjunctivitis |
| SNOMED CT | 314551004 | Corneal graft disorder |
| SNOMED CT | 314552006 | Corneal graft astigmatism |
| SNOMED CT | 314553001 | Corneal graft infection |
| SNOMED CT | 314554007 | Corneal graft vascularization |
| SNOMED CT | 314757003 | Ocular cicatricial pemphigoid |
| SNOMED CT | 314760005 | Corneal graft failure |
| SNOMED CT | 317349009 | Vernal keratoconjunctivitis |
| SNOMED CT | 318316003 | Vernal conjunctivitis |
| SNOMED CT | 320150004 | Idiopathic eosinophilia |
| SNOMED CT | 361098001 | Allergic transfusion reaction |

Continued on next page

**TableS31 – continued from previous page**

| CodeSystem | ConceptCode | ConceptName |
|---|---|---|
| SNOMED CT | 361106007 | Transfusion reaction due to allergen in blood |
| SNOMED CT | 361125005 | Subacute autoimmune thyroiditis |
| SNOMED CT | 361212009 | Oral pemphigus vulgaris |
| SNOMED CT | 362985009 | Antibody-mediated activation and inactivation |
| SNOMED CT | 362986005 | Antibody-mediated cytolysis |
| SNOMED CT | 362992004 | Autoimmune connective tissue disorder |
| SNOMED CT | 367498001 | Seasonal allergic rhinitis |
| SNOMED CT | 367542003 | Pulmonary eosinophilia |
| SNOMED CT | 370474006 | Eosinophilic myositis |
| SNOMED CT | 371044001 | Transient respiratory distress with sepsis |
| SNOMED CT | 373674001 | Anaphylactic urticaria |
| SNOMED CT | 389145006 | Allergic asthma |
| SNOMED CT | 397012002 | Cutaneous mastocytosis |
| SNOMED CT | 397013007 | Solitary cutaneous mastocytoma |
| SNOMED CT | 397014001 | Diffuse erythrodermic mastocytosis |
| SNOMED CT | 397173003 | Crohn's disease of intestine |
| SNOMED CT | 397843005 | Sepsis-related gastrointestinal lesions |
| SNOMED CT | 397856003 | Systemic lupus erythematosus-related syndrome |
| SNOMED CT | 398049005 | Mixed collagen vascular disease |
| SNOMED CT | 398726004 | Rheumatoid lung disease |
| SNOMED CT | 398937006 | Cold autoimmune hemolytic anemia |
| SNOMED CT | 399456004 | Plasma cell myeloma/plasmacytoma |
| SNOMED CT | 399946006 | Crohn's disease of skin |
| SNOMED CT | 400054000 | Rheumatoid vasculitis |
| SNOMED CT | 402188000 | Atopic dermatitis aggravated by type 1 immune reaction |
| SNOMED CT | 402244002 | Lichenoid allergic contact reaction due to color developer |
| SNOMED CT | 402245001 | Angry back syndrome |
| SNOMED CT | 402247009 | Lichenoid allergic contact reaction affecting oral mucosa |
| SNOMED CT | 402248004 | Lichenoid allergic contact reaction due to tattoo ink |
| SNOMED CT | 402249007 | Allergic contact dermatitis of eyelid |
| SNOMED CT | 402250007 | Allergic contact cheilitis |
| SNOMED CT | 402251006 | Allergic contact gingivostomatitis |
| SNOMED CT | 402252004 | Allergic contact dermatitis of perianal skin |
| SNOMED CT | 402254003 | Allergic contact dermatitis of hands |
| SNOMED CT | 402255002 | Allergic contact dermatitis of lower leg |
| SNOMED CT | 402256001 | Allergic contact dermatitis of male genitalia |
| SNOMED CT | 402257005 | Allergic contact dermatitis of female genitalia |
| SNOMED CT | 402304007 | Allergic contact urticaria |
| SNOMED CT | 402307000 | Chronic large plaque psoriasis |
| SNOMED CT | 402308005 | Chronic small plaque psoriasis |
| SNOMED CT | 402309002 | Chronic guttate pattern psoriasis |
| SNOMED CT | 402310007 | Chronic stable plaque psoriasis |
| SNOMED CT | 402311006 | Actively extending plaque psoriasis |
| SNOMED CT | 402312004 | Acute guttate psoriasis |
| SNOMED CT | 402313009 | Guttate flare of psoriasis with preexisting plaques |
| SNOMED CT | 402314003 | Psoriasis of vulva |
| SNOMED CT | 402315002 | Psoriasis of penis |

Continued on next page

**TableS31 – continued from previous page**

| CodeSystem | ConceptCode | ConceptName |
|---|---|---|
| SNOMED CT | 402316001 | Psoriasis of perianal skin |
| SNOMED CT | 402317005 | Generalized psoriasis |
| SNOMED CT | 402318000 | Photoaggravated psoriasis |
| SNOMED CT | 402319008 | Psoriasis of scalp margin |
| SNOMED CT | 402320002 | Psoriasis of face |
| SNOMED CT | 402321003 | Non-pustular psoriasis of hands |
| SNOMED CT | 402322005 | Hypertrophic palmar psoriasis |
| SNOMED CT | 402323000 | Hypertrophic palmoplantar psoriasis |
| SNOMED CT | 402324006 | Non-pustular psoriasis of hands and feet |
| SNOMED CT | 402325007 | Drug-exacerbated psoriasis |
| SNOMED CT | 402326008 | Acute generalized pustular psoriasis de novo |
| SNOMED CT | 402327004 | Acute generalized pustular flare of preexisting plaque psoriasis |
| SNOMED CT | 402329001 | Early onset psoriasis type 1 |
| SNOMED CT | 402330006 | Onset of psoriasis in infancy (<1 year) |
| SNOMED CT | 402331005 | Onset of psoriasis in childhood (1-10 years) |
| SNOMED CT | 402332003 | Onset of psoriasis in adolescence (10-20 years) |
| SNOMED CT | 402333008 | Onset of psoriasis in early adulthood (20-40 years) |
| SNOMED CT | 402334002 | Late onset psoriasis type 2 |
| SNOMED CT | 402335001 | Familial psoriasis |
| SNOMED CT | 402336000 | Familial psoriasis with affected first degree relative |
| SNOMED CT | 402337009 | Familial psoriasis without affected first degree relative |
| SNOMED CT | 402357008 | Lichenoid chronic graft-versus-host disease |
| SNOMED CT | 402358003 | Sclerodermoid chronic graft-versus-host disease |
| SNOMED CT | 402360001 | Materno-fetal graft-versus-host disease |
| SNOMED CT | 402375009 | Crohn's disease of vulva |
| SNOMED CT | 402376005 | Crohn's disease of penis |
| SNOMED CT | 402377001 | Crohn's disease of scrotum |
| SNOMED CT | 402384009 | Allergic urticaria due to ingested food |
| SNOMED CT | 402385005 | Allergic urticaria due to bite and/or sting |
| SNOMED CT | 402386006 | Allergic urticaria due to inhaled allergen |
| SNOMED CT | 402387002 | Allergic angioedema |
| SNOMED CT | 402388007 | Allergic angioedema due to ingested food |
| SNOMED CT | 402389004 | Allergic angioedema due to bite and/or sting |
| SNOMED CT | 402390008 | Anaphylaxis due to ingested food |
| SNOMED CT | 402392000 | Allergic urticaria and/or angioedema |
| SNOMED CT | 402397006 | Autoimmune urticaria |
| SNOMED CT | 402398001 | Acute autoimmune urticaria |
| SNOMED CT | 402399009 | Chronic autoimmune urticaria |
| SNOMED CT | 402400002 | Episodic autoimmune urticaria |
| SNOMED CT | 402401003 | Autoimmune angioedema |
| SNOMED CT | 402402005 | Autoimmune urticaria and/or angioedema |
| SNOMED CT | 402404006 | Episodic angioedema with eosinophilia |
| SNOMED CT | 402426007 | Rheumatoid nodulosis |
| SNOMED CT | 402427003 | Accelerated rheumatoid nodulosis |
| SNOMED CT | 402428008 | Accelerated rheumatoid nodulosis caused by methotrexate |
| SNOMED CT | 402431009 | Cutaneous atrophy due to rheumatoid arthritis |
| SNOMED CT | 402432002 | Rheumatoid neutrophilic dermatitis |

Continued on next page

**TableS31 – continued from previous page**

| CodeSystem | ConceptCode | ConceptName |
|---|---|---|
| SNOMED CT | 402433007 | Rheumatoid arthritis with nailfold and finger-pulp infarcts |
| SNOMED CT | 402434001 | Cutaneous complication of rheumatoid disease |
| SNOMED CT | 402435000 | Eosinophilic spongiosis (prebullous pemphigus) |
| SNOMED CT | 402436004 | Eosinophilic spongiosis (prebullous pemphigoid) |
| SNOMED CT | 402437008 | Localized bullous pemphigoid of vulva |
| SNOMED CT | 402438003 | Pemphigoid nodularis |
| SNOMED CT | 402439006 | Childhood bullous pemphigoid |
| SNOMED CT | 402440008 | Oral mucous membrane involvement by bullous pemphigoid |
| SNOMED CT | 402441007 | Oral involvement by mucous membrane pemphigoid |
| SNOMED CT | 402442000 | Brunsting-Perry type cicatricial pemphigoid |
| SNOMED CT | 402444004 | Oral mucosal involvement by dermatitis herpetiformis |
| SNOMED CT | 402447006 | Epidermolysis bullosa acquisita, classical acral type |
| SNOMED CT | 402448001 | Epidermolysis bullosa acquisita, bullous pemphigoid-like |
| SNOMED CT | 402449009 | Epidermolysis bullosa acquisita, cicatricial pemphigoid-like |
| SNOMED CT | 402450009 | Epidermolysis bullosa acquisita, Brunsting-Perry type |
| SNOMED CT | 402451008 | Epidermolysis bullosa acquisita, oral mucosal involvement |
| SNOMED CT | 402576006 | Lichenoid allergic contact reaction |
| SNOMED CT | 402712002 | Occupational scleroderma |
| SNOMED CT | 402717008 | IgA pemphigus |
| SNOMED CT | 402718003 | Pemphigus paraneoplastica |
| SNOMED CT | 402856005 | Acute hemorrhagic edema of childhood |
| SNOMED CT | 402865003 | Systemic lupus erythematosus-associated antiphospholipid syndrome |
| SNOMED CT | 402961006 | Pseudomonas septicemia with skin involvement |
| SNOMED CT | 402968000 | Erythema nodosum caused by Bacteria |
| SNOMED CT | 402969008 | Erythema nodosum caused by Genus Streptococcus |
| SNOMED CT | 402970009 | Erythema nodosum caused by Yersinia enterocolitica |
| SNOMED CT | 402971008 | Erythema multiforme due to mycoplasma infection |
| SNOMED CT | 403211002 | Papular form erythema multiforme |
| SNOMED CT | 403212009 | Vesicobullous form erythema multiforme |
| SNOMED CT | 403213004 | Erythema multiforme due to malignancy |
| SNOMED CT | 403217003 | Subcorneal pustular dermatosis with paraproteinemia |
| SNOMED CT | 403320008 | Circumscribed alopecia areata of scalp |
| SNOMED CT | 403321007 | Circumscribed alopecia areata of eyelashes/eyebrows |
| SNOMED CT | 403322000 | Circumscribed alopecia areata of beard area |
| SNOMED CT | 403323005 | Circumscribed alopecia areata of limbs |
| SNOMED CT | 403324004 | Circumscribed alopecia areata of trunk |
| SNOMED CT | 403434009 | Atopic keratoconjunctivitis |
| SNOMED CT | 403489007 | Subacute cutaneous lupus erythematosus, annular/polycyclic type |
| SNOMED CT | 403490003 | Subacute cutaneous lupus erythematosus, papulosquamous type |
| SNOMED CT | 403491004 | Erythema multiforme-like lupus erythematosus |
| SNOMED CT | 403492006 | Hypertrophic type discoid lupus erythematosus |
| SNOMED CT | 403493001 | Rosaceous type discoid lupus erythematosus |
| SNOMED CT | 403494007 | Discoid lupus erythematosus of mucous membranes |
| SNOMED CT | 403495008 | Discoid lupus erythematosus of oral mucosa |
| SNOMED CT | 403496009 | Discoid lupus erythematosus of genital mucous membranes |

Continued on next page

**TableS31 – continued from previous page**

| CodeSystem | ConceptCode | ConceptName |
|---|---|---|
| SNOMED CT | 403497000 | Discoid lupus erythematosus of scalp |
| SNOMED CT | 403498005 | Discoid lupus erythematosus of face |
| SNOMED CT | 403499002 | Discoid lupus erythematosus of lip |
| SNOMED CT | 403500006 | Discoid lupus erythematosus of hands |
| SNOMED CT | 403501005 | Discoid lupus erythematosus of foot |
| SNOMED CT | 403502003 | Disseminated discoid lupus erythematosus |
| SNOMED CT | 403513000 | Lupus erythematosus-associated nail dystrophy |
| SNOMED CT | 403629000 | Drug-induced bullous pemphigoid |
| SNOMED CT | 403630005 | Drug-induced pemphigus vulgaris |
| SNOMED CT | 403631009 | Drug-induced pemphigus foliaceus |
| SNOMED CT | 403632002 | Drug-induced epidermolysis bullosa acquisita |
| SNOMED CT | 403735006 | Eosinophilia-myalgia syndrome from tryptophan |
| SNOMED CT | 404145009 | Primary cutaneous anaplastic large cell B-cell lymphoma |
| SNOMED CT | 404165002 | Necrobiotic xanthogranuloma with paraproteinemia |
| SNOMED CT | 404170009 | Bullous urticaria pigmentosa |
| SNOMED CT | 404173006 | Familial mastocytosis |
| SNOMED CT | 404804003 | Platinum asthma |
| SNOMED CT | 404805002 | Platinum sensitivity |
| SNOMED CT | 404807005 | Cheese-washers' lung |
| SNOMED CT | 405720007 | Allergic bronchitis |
| SNOMED CT | 405821001 | Allergic otitis externa |
| SNOMED CT | 408335007 | Autoimmune hepatitis |
| SNOMED CT | 408539000 | Insulin autoimmune syndrome |
| SNOMED CT | 409089005 | Febrile neutropenia |
| SNOMED CT | 410482007 | Iritis with psoriatic arthritis |
| SNOMED CT | 410484008 | Iritis with ulcerative colitis |
| SNOMED CT | 410502007 | Juvenile rheumatoid arthritis |
| SNOMED CT | 410796000 | Juvenile seropositive polyarthritis |
| SNOMED CT | 410797009 | Juvenile seronegative polyarthritis |
| SNOMED CT | 410798004 | Juvenile idiopathic arthritis, oligoarthritis |
| SNOMED CT | 410799007 | Juvenile idiopathic arthritis, persistent oligoarthritis |
| SNOMED CT | 410800006 | Juvenile idiopathic arthritis, extended oligoarthritis |
| SNOMED CT | 410801005 | Juvenile idiopathic arthritis, enthesitis related arthritis |
| SNOMED CT | 410802003 | Juvenile idiopathic arthritis, undifferentiated arthritis |
| SNOMED CT | 413603009 | Autoimmune hemolytic anemia |
| SNOMED CT | 414061000 | Drug-induced cicatricial pemphigoid |
| SNOMED CT | 414156000 | Exacerbation of ulcerative colitis |
| SNOMED CT | 414450004 | Idiopathic hypereosinophilic syndrome |
| SNOMED CT | 414850009 | Neutrophilia disorder |
| SNOMED CT | 416093006 | Allergic reaction to drug |
| SNOMED CT | 417389002 | Disorder of basophils |
| SNOMED CT | 417516000 | Anaphylaxis due to substance |
| SNOMED CT | 417967008 | Disorder of eosinophil |
| SNOMED CT | 418176003 | Allergic reaction to plant, except food |
| SNOMED CT | 418282006 | Allergic reaction to cosmetics |
| SNOMED CT | 418325008 | Allergic reaction to adhesive |
| SNOMED CT | 418364006 | Allergic reaction to pollen |

Continued on next page

**TableS31 – continued from previous page**

| CodeSystem | ConceptCode | ConceptName |
|---|---|---|
| SNOMED CT | 418367004 | Allergic reaction to grass pollen |
| SNOMED CT | 418448002 | Allergic reaction to animal |
| SNOMED CT | 418579000 | Allergic reaction to colophony |
| SNOMED CT | 418634005 | Allergic reaction to substance |
| SNOMED CT | 418943003 | Allergic reaction to tree pollen |
| SNOMED CT | 419076005 | Allergic reaction |
| SNOMED CT | 419121001 | Allergic contact dermatitis of pinna |
| SNOMED CT | 419375001 | Allergic reaction to oil |
| SNOMED CT | 419447004 | Allergic reaction to flour dust |
| SNOMED CT | 419452009 | Allergic reaction to food |
| SNOMED CT | 419455006 | Disorder characterized by eosinophilia |
| SNOMED CT | 419515007 | Allergic reaction to detergent |
| SNOMED CT | 419519001 | Allergic reaction to weed pollen |
| SNOMED CT | 419838000 | Allergic reaction to chemical |
| SNOMED CT | 419884005 | Allergic reaction to flea bite |
| SNOMED CT | 419957005 | Allergic contact dermatitis of external auditory canal |
| SNOMED CT | 420198007 | Allergic reaction to platinum |
| SNOMED CT | 420373003 | Allergic disorder of digestive system |
| SNOMED CT | 420505005 | Corneal endothelial allograft rejection |
| SNOMED CT | 420571002 | Corneal subepithelial allograft rejection |
| SNOMED CT | 420881009 | Allergic disorder caused by substance |
| SNOMED CT | 421092003 | Allergic disorder of respiratory system |
| SNOMED CT | 421187003 | Corneal epithelial allograft rejection |
| SNOMED CT | 422290000 | Allergic disorder of skin |
| SNOMED CT | 423486005 | Disseminated eosinophilic collagen disease |
| SNOMED CT | 424643009 | IgE-mediated allergic asthma |
| SNOMED CT | 425011002 | Allergic fungal sinusitis |
| SNOMED CT | 425500002 | Secondary progressive multiple sclerosis |
| SNOMED CT | 426065008 | Hypersensitivity disorder mediated by immune complex |
| SNOMED CT | 426136000 | Delayed renal graft function |
| SNOMED CT | 426364002 | Allergic hepatitis |
| SNOMED CT | 426373005 | Relapsing remitting multiple sclerosis |
| SNOMED CT | 426549001 | Crohn's disease in remission |
| SNOMED CT | 426760008 | Delayed hypersensitivity disorder |
| SNOMED CT | 426871003 | Cell-mediated cytotoxic disorder |
| SNOMED CT | 427213005 | Autoimmune vasculitis |
| SNOMED CT | 427245000 | Febrile leukopenia |
| SNOMED CT | 427435004 | Punctate keratopathy after penetrating keratoplasty |
| SNOMED CT | 427439005 | Immune hypersensitivity disorder by mechanism |
| SNOMED CT | 427833000 | Anaphylaxis due to mollusk |
| SNOMED CT | 427889009 | Hypertension associated with transplantation |
| SNOMED CT | 427903006 | Anaphylaxis due to fish |
| SNOMED CT | 427927008 | Disorder related to cardiac transplantation |
| SNOMED CT | 427928003 | Disorder related to lung transplantation |
| SNOMED CT | 428218004 | Allergic arthritis of spine |
| SNOMED CT | 428470000 | Autoimmune sensorineural hearing loss |
| SNOMED CT | 428575007 | Hypertension secondary to kidney transplant |

Continued on next page

**TableS31 – continued from previous page**

| CodeSystem | ConceptCode | ConceptName |
|---|---|---|
| SNOMED CT | 428700003 | Primary progressive multiple sclerosis |
| SNOMED CT | 428795003 | Anaphylaxis due to vegetable |
| SNOMED CT | 429195002 | Allergic rhinitis due to animal dander |
| SNOMED CT | 429257001 | Disorder of transplanted heart |
| SNOMED CT | 429451003 | Disorder related to renal transplantation |
| SNOMED CT | 429751004 | Anaphylaxis due to fruit |
| SNOMED CT | 430822009 | Basophilic leukaemoid reaction |
| SNOMED CT | 430980000 | Anaphylaxis due to hymenoptera venom |
| SNOMED CT | 431186002 | Accelerated rejection of cardiac transplant |
| SNOMED CT | 431222008 | Acute rejection of liver transplant |
| SNOMED CT | 431223003 | Acute rejection of lung transplant |
| SNOMED CT | 431505005 | Rejection of intestine transplant |
| SNOMED CT | 431507002 | Accelerated rejection of lung transplant |
| SNOMED CT | 431896008 | Chronic rejection of cardiac transplant |
| SNOMED CT | 432265007 | Rejection of corneal graft after penetrating keratoplasty |
| SNOMED CT | 432772009 | Hyperacute rejection of liver transplant |
| SNOMED CT | 432773004 | Hyperacute rejection of cardiac transplant |
| SNOMED CT | 432774005 | Hyperacute rejection of lung transplant |
| SNOMED CT | 432777003 | Accelerated rejection of liver transplant |
| SNOMED CT | 432843002 | Acute rejection of cardiac transplant |
| SNOMED CT | 432908002 | Chronic rejection of liver transplant |
| SNOMED CT | 432958009 | Chronic rejection of lung transplant |
| SNOMED CT | 432989008 | Allergic reaction to tattoo ink |
| SNOMED CT | 433087001 | Primary failure of corneal graft after penetrating keratoplasty |
| SNOMED CT | 433600001 | Hyperacute rejection of intestine transplant |
| SNOMED CT | 433804007 | Accelerated rejection of intestine transplant |
| SNOMED CT | 434238001 | Acute rejection of intestine transplant |
| SNOMED CT | 434270001 | Chronic rejection of intestine transplant |
| SNOMED CT | 438511000 | Benign multiple sclerosis |
| SNOMED CT | 439567002 | Malignant multiple sclerosis |
| SNOMED CT | 441492003 | Anaphylaxis due to seed |
| SNOMED CT | 441495001 | Anaphylaxis due to tree nut |
| SNOMED CT | 441593005 | Anaphylaxis due to latex |
| SNOMED CT | 441971007 | Chronic left-sided ulcerative colitis |
| SNOMED CT | 441978001 | Allergic rhinitis due to food |
| SNOMED CT | 442052005 | Anaphylaxis due to shellfish |
| SNOMED CT | 442159003 | Chronic ulcerative pancolitis |
| SNOMED CT | 443872005 | Systemic sclerosis due to chemical |
| SNOMED CT | 443899007 | Autoimmune inflammation of skeletal muscle |
| SNOMED CT | 443980004 | Neutropenic sepsis |
| SNOMED CT | 444133002 | Progressive systemic sclerosis |
| SNOMED CT | 444546002 | Chronic ulcerative colitis |
| SNOMED CT | 444547006 | Cutaneous graft-versus-host disease |
| SNOMED CT | 444548001 | Ulcerative pancolitis |
| SNOMED CT | 444855007 | Arteriosclerosis of coronary artery bypass graft of transplanted heart |
| SNOMED CT | 445030005 | Blastic plasmacytoid dendritic cell neoplasm |

Continued on next page

**TableS31 – continued from previous page**

| CodeSystem | ConceptCode | ConceptName |
|---|---|---|
| SNOMED CT | 445187004 | Antisynthetase syndrome |
| SNOMED CT | 445243001 | Left sided ulcerative colitis |
| SNOMED CT | 445260006 | Posttransplant diabetes mellitus |
| SNOMED CT | 445261005 | Posttransplant hyperlipidemia |
| SNOMED CT | 446021007 | Limbal vernal conjunctivitis |
| SNOMED CT | 446096008 | Perennial allergic rhinitis |
| SNOMED CT | 446682003 | Pediatric autoimmune neuropsychiatric disorder associated with streptococcal infection |
| SNOMED CT | 447843005 | Severe sepsis with acute organ dysfunction caused by anaerobic bacteria |
| SNOMED CT | 448542008 | Autoimmune pancreatitis |
| SNOMED CT | 448784003 | Sepsis due to herpes simplex |
| SNOMED CT | 449729000 | Allergic rhinitis due to house dust mite |
| SNOMED CT | 449730005 | Autoimmune polyendocrine syndrome type 4 |
| SNOMED CT | 449731009 | Autoimmune polyendocrine syndrome type 3 |
| SNOMED CT | 1197483002 | Functional disorder of polymorphonuclear neutrophil |
| SNOMED CT | 473104002 | Allergic reaction to hair dye |
| SNOMED CT | 473460002 | Allergic conjunctivitis |
| SNOMED CT | 608874000 | Eaton Lambert syndrome with underlying malignancy |
| SNOMED CT | 609329007 | Catastrophic antiphospholipid syndrome |
| SNOMED CT | 609485004 | Induced termination of pregnancy complicated by sepsis |
| SNOMED CT | 609552005 | Allergic reaction to radiocontrast media |
| SNOMED CT | 697969008 | Ulcerative colitis in remission |
| SNOMED CT | 698819004 | Postoperative sepsis |
| SNOMED CT | 700050004 | Overwhelming infection in asplenic patient |
| SNOMED CT | 700051000 | Sepsis in asplenic subject |
| SNOMED CT | 700052007 | Post-splenectomy sepsis |
| SNOMED CT | 700054008 | Sepsis due to fungus |
| SNOMED CT | 700055009 | Sepsis with cutaneous manifestations |
| SNOMED CT | 700057001 | Emberger syndrome |
| SNOMED CT | 700269001 | Discoid lupus erythematosus of lower eyelid |
| SNOMED CT | 700334001 | Discoid lupus erythematosus of upper eyelid |
| SNOMED CT | 700340008 | Allergic contact dermatitis of upper eyelid |
| SNOMED CT | 700341007 | Allergic contact dermatitis of lower eyelid |
| SNOMED CT | 702575003 | Retinocochleocerebral vasculopathy |
| SNOMED CT | 702617007 | Acute generalized exanthematous pustulosis |
| SNOMED CT | 703048006 | Post-renal transplant vesicoureteric reflux |
| SNOMED CT | 703133002 | Lichenoid allergic contact reaction due to amalgam |
| SNOMED CT | 703538003 | Mannose-binding lectin deficiency |
| SNOMED CT | 703802001 | Acquired angioedema due to C1 inhibitor autoantibody |
| SNOMED CT | 707148007 | Recurrent post-transplant renal disease |
| SNOMED CT | 707271004 | Sepsis due to oral infection |
| SNOMED CT | 707301001 | Lupus erythematosus of oral mucous membrane |
| SNOMED CT | 707314000 | Erythema multiforme co-occurrent with ulcer of oral mucous membrane |
| SNOMED CT | 707441009 | Milk-induced pulmonary disease in infant |
| SNOMED CT | 707443007 | Autoimmune pulmonary alveolar proteinosis |

Continued on next page

**TableS31 – continued from previous page**

| CodeSystem | ConceptCode | ConceptName |
|---|---|---|
| SNOMED CT | 707986004 | Mucous membrane pemphigoid with esophageal involvement |
| SNOMED CT | 708093000 | Acute exacerbation of allergic asthma |
| SNOMED CT | 708095007 | Acute severe exacerbation of immunoglobin E-mediated allergic asthma |
| SNOMED CT | 710027002 | Cytokine release syndrome |
| SNOMED CT | 711160007 | Eosinophil peroxidase deficiency |
| SNOMED CT | 711435001 | Chronic migratory panniculitis |
| SNOMED CT | 711526005 | Chronic localized pemphigoid |
| SNOMED CT | 713225000 | Gingival disease due to lupus erythematosus |
| SNOMED CT | 713229006 | Gingival disease due to erythema multiforme |
| SNOMED CT | 713236007 | Allergic contact gingivitis caused by acrylic dental material |
| SNOMED CT | 713654004 | Autoimmune cholangitis |
| SNOMED CT | 713825007 | Transplant renal artery stenosis |
| SNOMED CT | 713854001 | Perinatal sepsis |
| SNOMED CT | 713865006 | Perinatal sepsis caused by Streptococcus agalactiae |
| SNOMED CT | 713866007 | Perinatal sepsis caused by Escherichia coli |
| SNOMED CT | 714148005 | Allergic contact gingivitis caused by nickel |
| SNOMED CT | 714149002 | Allergic contact gingivitis caused by mercury |
| SNOMED CT | 714522006 | Allergic gingival disease |
| SNOMED CT | 715070000 | Sepsis caused by Peptostreptococcus |
| SNOMED CT | 715071001 | Sepsis caused by anaerobic streptococcus |
| SNOMED CT | 715401008 | Reynolds syndrome |
| SNOMED CT | 715770009 | Acute motor axonal neuropathy |
| SNOMED CT | 715840006 | Pemphigus vulgaris of gingival mucous membrane |
| SNOMED CT | 715863001 | Autoimmune necrotizing myopathy |
| SNOMED CT | 715887004 | Anaphylaxis caused by sulfite salt |
| SNOMED CT | 716374005 | Anaphylaxis caused by sulfur dioxide |
| SNOMED CT | 716722005 | Acute motor sensory axonal neuropathy |
| SNOMED CT | 716723000 | Acute inflammatory demyelinating polyradiculoneuropathy |
| SNOMED CT | 717780007 | Monoclonal B-cell lymphocytosis |
| SNOMED CT | 717872008 | Allergic duodenitis |
| SNOMED CT | 718175009 | Macrophagic myofasciitis |
| SNOMED CT | 718471008 | Oral pemphigoid of gingival mucous membrane |
| SNOMED CT | 718716008 | Autoimmune hemolytic anemia mixed type |
| SNOMED CT | 719040004 | Anaphylaxis caused by lupin flour |
| SNOMED CT | 720497002 | Anti-human leukocyte antigen hyperimmunization |
| SNOMED CT | 720587009 | Donor derived melanoma |
| SNOMED CT | 720588004 | Non-immunoglobulin E-mediated drug sensitivity |
| SNOMED CT | 720980004 | Alopecia, psychomotor epilepsy, periodontal pyorrhea, intellectual disability syndrome |
| SNOMED CT | 721093000 | Dianzani autoimmune lymphoproliferative disease |
| SNOMED CT | 721104000 | Sepsis due to urinary tract infection |
| SNOMED CT | 721196005 | Pemphigus of vulva |
| SNOMED CT | 721278008 | Neonatal sepsis caused by Malassezia |
| SNOMED CT | 721307000 | Transient abnormal myelopoiesis |
| SNOMED CT | 721538000 | Psoriasis of anogenital region |
| SNOMED CT | 721572008 | Sepsis caused by Pseudomonas aeruginosa |

Continued on next page

**TableS31 – continued from previous page**

| CodeSystem | ConceptCode | ConceptName |
|---|---|---|
| SNOMED CT | 721686000 | Inflammatory polyps co-occurrent and due to inflammatory bowel disease |
| SNOMED CT | 721702009 | Crohn disease of anal canal |
| SNOMED CT | 721711009 | Autoimmune hepatitis type 1 |
| SNOMED CT | 721712002 | Autoimmune hepatitis type 2 |
| SNOMED CT | 721713007 | Autoimmune hepatitis type 3 |
| SNOMED CT | 69688007 | Chronic erythema nodosum |
| SNOMED CT | 70139002 | Harara |
| SNOMED CT | 70349007 | Pseudoneutrophilia |
| SNOMED CT | 70884002 | Allergic encephalitis |
| SNOMED CT | 70910003 | Indolent systemic mastocytosis |
| SNOMED CT | 71610005 | Neutrophilic leukaemoid reaction |
| SNOMED CT | 71692003 | Leukoerythroblastotic reaction |
| SNOMED CT | 71958004 | Chronic allergic otitis media |
| SNOMED CT | 72272002 | Non malignant mast cell disease |
| SNOMED CT | 72275000 | Relapsing polychondritis |
| SNOMED CT | 72681001 | Allergic jejunitis |
| SNOMED CT | 72986009 | Acute hemorrhagic leukoencephalitis |
| SNOMED CT | 73397007 | Heparin-induced thrombocytopenia |
| SNOMED CT | 73448002 | Fish-meal workers' lung |
| SNOMED CT | 73658009 | Erythema multiforme, epidermal type |
| SNOMED CT | 74326002 | Subacute myeloid leukemia |
| SNOMED CT | 74610006 | Tuberculous erythema nodosum |
| SNOMED CT | 75316000 | Autoimmune hypoparathyroidism |
| SNOMED CT | 75331009 | Evans syndrome |
| SNOMED CT | 76092003 | Benign mucous membrane pemphigoid with ocular involvement |
| SNOMED CT | 76097009 | Erythema nodosum migrans |
| SNOMED CT | 76481008 | Lymphoepithelioid lymphoma |
| SNOMED CT | 76621000 | Urticaria pigmentosa, infantile form |
| SNOMED CT | 76762001 | Eosinophilic myopathy |
| SNOMED CT | 77090002 | Bullous pemphigoid |
| SNOMED CT | 77206006 | Puerperal pelvic sepsis |
| SNOMED CT | 77363004 | Concentric alopecia areata |
| SNOMED CT | 77522006 | Rheumatoid episcleritis |
| SNOMED CT | 78069008 | Chronic rheumatic pericarditis |
| SNOMED CT | 78079005 | Allergic contact dermatitis due to excretions |
| SNOMED CT | 78223000 | Coccidioidomycosis with erythema nodosum |
| SNOMED CT | 78324009 | Mild chronic ulcerative colitis |
| SNOMED CT | 78712000 | Moderate chronic ulcerative colitis |
| SNOMED CT | 78745000 | Urticaria pigmentosa |
| SNOMED CT | 79291003 | Discoid lupus erythematosus of eyelid |
| SNOMED CT | 79336007 | Familial eosinophilia |
| SNOMED CT | 79958002 | Grain fever |
| SNOMED CT | 80255009 | Maternal transfer neutropenia |
| SNOMED CT | 80258006 | Drug-induced lupus erythematosus |
| SNOMED CT | 80369006 | Chronic neutrophilia |
| SNOMED CT | 80988005 | Mixed cryofibrinogenemia |

Continued on next page

**TableS31 – continued from previous page**

| CodeSystem | ConceptCode | ConceptName |
|---|---|---|
| SNOMED CT | 81077008 | Acute rheumatic arthritis |
| SNOMED CT | 81271001 | Localized pustular psoriasis |
| SNOMED CT | 81285006 | Pemphigus vegetans |
| SNOMED CT | 82275008 | Eosinophilic granulomatosis with polyangiitis |
| SNOMED CT | 82939003 | Chronic rheumatoid nodular fibrosis |
| SNOMED CT | 83728000 | Polyglandular autoimmune syndrome, type 2 |
| SNOMED CT | 83839005 | Acrodermatitis continua of Hallopeau |
| SNOMED CT | 83942000 | Acute disseminated encephalomyelitis |
| SNOMED CT | 84479000 | Allergic inhalant dermatitis |
| SNOMED CT | 84730007 | Cryopathy |
| SNOMED CT | 84828003 | Leukopenia |
| SNOMED CT | 84871007 | Acute rheumatic valvulitis |
| SNOMED CT | 85512009 | Contact dermatitis due to jewelry |
| SNOMED CT | 85828009 | Autoimmune disease |
| SNOMED CT | 86081009 | Herpes gestationis |
| SNOMED CT | 86142006 | Pemphigoid |
| SNOMED CT | 86166000 | Alopecia universalis |
| SNOMED CT | 86225009 | Hapten type high affinity hemolytic anemia |
| SNOMED CT | 86563000 | Erythema multiforme, mixed dermal-epidermal type |
| SNOMED CT | 86638007 | Maple-bark strippers' lung |
| SNOMED CT | 88714009 | Transient hypogammaglobulinemia of infancy |
| SNOMED CT | 89099002 | Chronic allergic bronchitis |
| SNOMED CT | 89155008 | Systemic sclerosis |
| SNOMED CT | 89456004 | Chronic rheumatic valvulitis |
| SNOMED CT | 90453003 | Chronic rheumatic mediastinopericarditis |
| SNOMED CT | 91302008 | Sepsis |
| SNOMED CT | 91623008 | Hydroa herpetiformis |
| SNOMED CT | 91924004 | Allergic gingivitis |
| SNOMED CT | 91925003 | Allergic rhinitis due to animals |
| SNOMED CT | 91926002 | Allergic rhinitis due to grass pollen |
| SNOMED CT | 91927006 | Allergic rhinitis due to tree pollen |
| SNOMED CT | 91928001 | Allergic rhinitis due to weed pollen |
| SNOMED CT | 91941002 | Food anaphylaxis |
| SNOMED CT | 92816008 | Chronic monocytic leukemia |
| SNOMED CT | 92831004 | Complication of transplanted embryo |
| SNOMED CT | 95261008 | Sézary's disease of lymph nodes of inguinal region AND/OR lower limb |
| SNOMED CT | 95329006 | Autoimmune skin disease |
| SNOMED CT | 95332009 | Rash of systemic lupus erythematosus |
| SNOMED CT | 95416007 | Eosinophilia myalgia syndrome |
| SNOMED CT | 95609003 | Neonatal lupus erythematosus |
| SNOMED CT | 95624007 | Neonatal lymphocytopenia |
| SNOMED CT | 95643007 | Autoimmune encephalitis |
| SNOMED CT | 95733001 | Eosinophilic keratitis |
| SNOMED CT | 95742008 | Corneal graft rejection |
| SNOMED CT | 105601003 | Quantitative disorder of neutrophils |
| SNOMED CT | 105602005 | Quantitative abnormality of granulocytes |

Continued on next page

**TableS31 – continued from previous page**

| CodeSystem | ConceptCode | ConceptName |
|---|---|---|
| SNOMED CT | 109250009 | Chronic bullous dermatosis of childhood |
| SNOMED CT | 109992005 | Polycythemia vera (clinical) |
| SNOMED CT | 111037009 | Basophilic hyperplasia |
| SNOMED CT | 111196000 | Dermatitis herpetiformis |
| SNOMED CT | 111209006 | Photoallergic dermatitis |
| SNOMED CT | 111583006 | Leukocytosis |
| SNOMED CT | 111588002 | Heparin-induced thrombocytopenia with thrombosis |
| SNOMED CT | 115666004 | Animal handlers' lung |
| SNOMED CT | 123950001 | Rheumatic chorea without heart involvement |
| SNOMED CT | 127049002 | Primary (idiopathic) autoimmune hemolytic anemia |
| SNOMED CT | 127050002 | Secondary autoimmune hemolytic anemia |
| SNOMED CT | 127052005 | Secondary warm autoimmune hemolytic anemia |
| SNOMED CT | 127053000 | Post-infectious cold agglutinin disease |
| SNOMED CT | 127055007 | Chronic cold agglutinin disease |
| SNOMED CT | 127056008 | Chronic cold agglutinin disease associated with B-cell neoplasm |
| SNOMED CT | 127057004 | Paroxysmal cold hemoglobinuria |
| SNOMED CT | 127058009 | Paroxysmal cold hemoglobinuria associated with tertiary syphilis |
| SNOMED CT | 127060006 | Drug-induced immune hemolytic anemia, hapten type |
| SNOMED CT | 127061005 | Autoimmune hemolytic anemia, categorized by antibody class AND/OR complement |
| SNOMED CT | 127066000 | Familial polycythemia vera |
| SNOMED CT | 127067009 | Stress neutrophilia |
| SNOMED CT | 127218004 | Reactive follicular hyperplasia in the elderly |
| SNOMED CT | 128091003 | Primary ITP (immune thrombocytopenia) |
| SNOMED CT | 128092005 | Secondary autoimmune thrombocytopenia |
| SNOMED CT | 128093000 | Alloimmune thrombocytopenia |
| SNOMED CT | 128094006 | Alloimmune platelet transfusion refractoriness |
| SNOMED CT | 128460000 | Systemic sclerosis, diffuse |
| SNOMED CT | 128461001 | Systemic sclerosis sine scleroderma |
| SNOMED CT | 128600008 | Acute ulcerative colitis |
| SNOMED CT | 128831004 | Chronic myelomonocytic leukemia |
| SNOMED CT | 129563009 | Rheumatoid osteoperiostitis |
| SNOMED CT | 129639005 | Hereditary neutrophilia |
| SNOMED CT | 129640007 | Benign granulocytopenia in childhood |
| SNOMED CT | 129641006 | Chronic benign neutropenia of childhood |
| SNOMED CT | 156370009 | Psoriatic arthritis |
| SNOMED CT | 165531008 | Lymphocytosis - absolute |
| SNOMED CT | 186365005 | Acute meningococcemia |
| SNOMED CT | 190454008 | Autoimmune parathyroiditis |
| SNOMED CT | 191210005 | Primary cold-type hemolytic anemia |
| SNOMED CT | 191211009 | Primary warm-type hemolytic anemia |
| SNOMED CT | 191212002 | Secondary cold-type hemolytic anemia |
| SNOMED CT | 191213007 | Secondary warm-type hemolytic anemia |
| SNOMED CT | 191273000 | Leukemoid reaction of the newborn |
| SNOMED CT | 191306005 | Immunoglobulin A vasculitis |
| SNOMED CT | 191345000 | Acquired neutropenia in newborn |
| SNOMED CT | 191358004 | Hereditary eosinophilia |

Continued on next page

**TableS31 – continued from previous page**

| CodeSystem | ConceptCode | ConceptName |
|---|---|---|
| SNOMED CT | 191360002 | Drug-induced eosinophilia |
| SNOMED CT | 192926004 | Multiple sclerosis of the brainstem |
| SNOMED CT | 192927008 | Multiple sclerosis of the spinal cord |
| SNOMED CT | 192929006 | Exacerbation of multiple sclerosis |
| SNOMED CT | 194394004 | Gram positive sepsis |
| SNOMED CT | 194709000 | Acute rheumatic myocarditis |
| SNOMED CT | 195528001 | Acute rheumatic fever |
| SNOMED CT | 195663004 | Allergic pharyngitis |
| SNOMED CT | 195989002 | Pituitary snuff-takers' disease |
| SNOMED CT | 196133001 | Lung disease with systemic sclerosis |
| SNOMED CT | 196578009 | Crohn's disease of oral soft tissues |
| SNOMED CT | 196853004 | Septicemic pasteurellosis |
| SNOMED CT | 196987008 | Ulcerative ileocolitis |
| SNOMED CT | 197024006 | Allergic enterocolitis |
| SNOMED CT | 199127006 | Herpes gestationis - delivered |
| SNOMED CT | 199128001 | Herpes gestationis - delivered with postnatal complication |
| SNOMED CT | 199129009 | Herpes gestationis - not delivered |
| SNOMED CT | 199710008 | Sepsis during labor, delivered |
| SNOMED CT | 199711007 | Sepsis during labor with antenatal problem |
| SNOMED CT | 200196001 | Puerperal sepsis with postnatal complication |
| SNOMED CT | 200842003 | Allergic contact dermatitis due to adhesive |
| SNOMED CT | 200843008 | Allergic contact dermatitis due to cosmetic |
| SNOMED CT | 200844002 | Allergic contact dermatitis due to drug in contact with skin |
| SNOMED CT | 200845001 | Allergic contact dermatitis due to dye |
| SNOMED CT | 200847009 | Allergic contact dermatitis due to food in contact with the skin |
| SNOMED CT | 200907003 | Drug-induced pemphigus |
| SNOMED CT | 200936003 | Lupus erythematosus |
| SNOMED CT | 200937007 | Lupus erythematosus chronicus |
| SNOMED CT | 200938002 | Discoid lupus erythematosus |
| SNOMED CT | 200939005 | Lupus erythematosus migrans |
| SNOMED CT | 200940007 | Lupus erythematosus nodularis |
| SNOMED CT | 200941006 | Lupus erythematosus tumidus |
| SNOMED CT | 200942004 | Lupus erythematosus unguium mutilans |
| SNOMED CT | 200951007 | Erythema marginatum in acute rheumatic fever |
| SNOMED CT | 200956002 | Psoriatic arthritis with spine involvement |
| SNOMED CT | 200962007 | Psoriasis annularis |
| SNOMED CT | 200963002 | Psoriasis circinata |
| SNOMED CT | 200964008 | Psoriasis diffusa |
| SNOMED CT | 200965009 | Plaque psoriasis |
| SNOMED CT | 200966005 | Psoriasis geographica |
| SNOMED CT | 200967001 | Psoriasis gyrata |
| SNOMED CT | 200968006 | Psoriasis inveterata |
| SNOMED CT | 200969003 | Rupioid psoriasis |
| SNOMED CT | 200970002 | Psoriasis palmaris |
| SNOMED CT | 200971003 | Psoriasis plantaris |
| SNOMED CT | 200972005 | Psoriasis punctata |
| SNOMED CT | 200973000 | Pustular psoriasis |

Continued on next page

**TableS31 – continued from previous page**

| CodeSystem | ConceptCode | ConceptName |
|---|---|---|
| SNOMED CT | 200974006 | Psoriasis universalis |
| SNOMED CT | 200975007 | Psoriasis vulgaris |
| SNOMED CT | 200977004 | Erythrodermic psoriasis |
| SNOMED CT | 201436003 | Drug-induced systemic lupus erythematosus |
| SNOMED CT | 201443009 | Systemic sclerosis induced by drugs and chemicals |
| SNOMED CT | 201789001 | Rheumatoid nodule |
| SNOMED CT | 201796004 | Systemic onset juvenile chronic arthritis |
| SNOMED CT | 201805000 | Juvenile arthritis in Crohn disease |
| SNOMED CT | 201807008 | Juvenile arthritis in ulcerative colitis |
| SNOMED CT | 201964002 | Allergic arthritis of the shoulder region |
| SNOMED CT | 201967009 | Allergic arthritis of the hand |
| SNOMED CT | 201968004 | Allergic arthritis of the pelvic region and thigh |
| SNOMED CT | 201970008 | Allergic arthritis of the ankle and/or foot |
| SNOMED CT | 201972000 | Allergic arthritis of multiple sites |
| SNOMED CT | 206376005 | Sepsis of the newborn |
| SNOMED CT | 206380000 | Sepsis of newborn due to anaerobes |
| SNOMED CT | 1208834007 | Alopecia areata of eyelash |
| SNOMED CT | 213085001 | Fracture of bone allograft |
| SNOMED CT | 213086000 | Displacement of bone allograft |
| SNOMED CT | 213088004 | Rejection of bone allograft |
| SNOMED CT | 213127009 | Infection of bone allograft |
| SNOMED CT | 213150003 | Kidney transplant failure and rejection |
| SNOMED CT | 213151004 | Heart transplant failure and rejection |
| SNOMED CT | 213152006 | Heart-lung transplant failure and rejection |
| SNOMED CT | 213153001 | Liver transplant failure and rejection |
| SNOMED CT | 213192008 | Skin graft failure |
| SNOMED CT | 213203004 | Complications of attempted introduction of embryo in embryo transfer |
| SNOMED CT | 213320003 | Anaphylactic shock due to serum |
| SNOMED CT | 230372003 | Acute relapsing multiple sclerosis |
| SNOMED CT | 230591002 | Motor neuropathy with multiple conduction block |
| SNOMED CT | 231854006 | Atopic conjunctivitis |
| SNOMED CT | 231855007 | Seasonal allergic conjunctivitis |
| SNOMED CT | 231856008 | Perennial allergic conjunctivitis |
| SNOMED CT | 231857004 | Giant papillary conjunctivitis |
| SNOMED CT | 232234000 | Acute allergic otitis externa |
| SNOMED CT | 232240007 | Chronic allergic otitis externa |
| SNOMED CT | 232308006 | Autoimmune disorder of inner ear |
| SNOMED CT | 232353008 | Perennial allergic rhinitis with seasonal variation |
| SNOMED CT | 232430006 | Recurrent allergic croup |
| SNOMED CT | 232439007 | Relapsing polychondritis of larynx |
| SNOMED CT | 233634002 | Post-lung transplantation bronchiectasis |
| SNOMED CT | 233683003 | Hay fever with asthma |
| SNOMED CT | 233687002 | Colophony asthma |
| SNOMED CT | 233691007 | Eosinophilic asthma |
| SNOMED CT | 233692000 | Cryptogenic pulmonary eosinophilia |
| SNOMED CT | 233694004 | Dog house disease |

Continued on next page

**TableS31 – continued from previous page**

| CodeSystem | ConceptCode | ConceptName |
|---|---|---|
| SNOMED CT | 233695003 | Dry rot lung |
| SNOMED CT | 233696002 | Lycoperdonosis |
| SNOMED CT | 233697006 | New Guinea lung |
| SNOMED CT | 233698001 | Paprika splitters' lung |
| SNOMED CT | 233699009 | Pyrethrum alveolitis |
| SNOMED CT | 233700005 | Rodent handlers' lung |
| SNOMED CT | 233701009 | Sewage workers' lung |
| SNOMED CT | 233702002 | Summer-type hypersensitivity pneumonitis |
| SNOMED CT | 233774000 | Humidifier fever |
| SNOMED CT | 233844002 | Accelerated coronary artery disease in transplanted heart |
| SNOMED CT | 233932001 | Cardiac transplant disorder |
| SNOMED CT | 233933006 | Cardiac transplant rejection |
| SNOMED CT | 233934000 | Cardiac transplant failure |
| SNOMED CT | 233979001 | Compression of transplanted artery |
| SNOMED CT | 233971003 | Stenosis of transplant artery |
| SNOMED CT | 234001008 | Dissection of transplant artery |
| SNOMED CT | 234004000 | Hemorrhage of transplant artery |
| SNOMED CT | 234008002 | Rupture of transplanted artery |
| SNOMED CT | 234066000 | Stenosis of transplant vein |
| SNOMED CT | 234077006 | Hemorrhage of transplanted vein |
| SNOMED CT | 234079009 | Rupture of transplanted vein |
| SNOMED CT | 234381003 | Maternal autoimmune hemolytic anemia |
| SNOMED CT | 234417006 | T-cell lymphocytosis |
| SNOMED CT | 234418001 | Chronic benign granulocytopenia |
| SNOMED CT | 234423001 | Chronic benign neutropenia |
| SNOMED CT | 234426009 | Corticosteroid-induced neutrophilia |
| SNOMED CT | 234427000 | Monocytoid disorder |
| SNOMED CT | 234434003 | Alders syndrome |
| SNOMED CT | 234435002 | Hereditary hypersegmentation |
| SNOMED CT | 234513007 | Post-splenectomy leukocytosis |
| SNOMED CT | 234538002 | Hyperimmunoglobulin D with periodic fever |
| SNOMED CT | 234556002 | Specific antibody deficiency |
| SNOMED CT | 234557006 | Anti-polysaccharide antibody deficiency |
| SNOMED CT | 234559009 | Anti-pneumococcal polysaccharide antibody deficiency |
| SNOMED CT | 234560004 | Anti-meningococcal polysaccharide A antibody deficiency |
| SNOMED CT | 234561000 | Anti-meningococcal polysaccharide C antibody deficiency |
| SNOMED CT | 234562007 | Anti-protein antibody deficiency |
| SNOMED CT | 234579001 | Mannan-binding protein deficiency |
| SNOMED CT | 234586009 | Leukocyte glucose-6-phosphate dehydrogenase deficiency |
| SNOMED CT | 234589002 | Glutathione synthetase deficiency |
| SNOMED CT | 234590006 | Gluthathione peroxidase deficiency |
| SNOMED CT | 234642007 | Age-related immunodeficiency |
| SNOMED CT | 234643002 | Transient immunodeficiency of infancy |
| SNOMED CT | 234999001 | Crohn's disease of gingivae |
| SNOMED CT | 235607002 | Crohn's disease of esophagus |
| SNOMED CT | 235657005 | Sepsis-related gastritis |
| SNOMED CT | 235714007 | Ulcerative enterocolitis |

Continued on next page

**TableS31 – continued from previous page**

| CodeSystem | ConceptCode | ConceptName |
|---|---|---|
| SNOMED CT | 235728001 | Autoimmune enteropathy |
| SNOMED CT | 235890007 | Autoimmune liver disease |
| SNOMED CT | 235910007 | Liver transplant disorder |
| SNOMED CT | 235911006 | Liver transplant rejection |
| SNOMED CT | 235912004 | Liver transplant failure |
| SNOMED CT | 236436003 | End stage renal failure with renal transplant |
| SNOMED CT | 236502006 | Renal involvement in scleroderma |
| SNOMED CT | 236503001 | Acute scleroderma renal crisis |
| SNOMED CT | 236569000 | Primary non-function of renal transplant |
| SNOMED CT | 236570004 | Renal transplant rejection |
| SNOMED CT | 236571000 | Hyperacute rejection of renal transplant |
| SNOMED CT | 236572007 | Accelerated rejection of renal transplant |
| SNOMED CT | 236573002 | Very mild acute rejection of renal transplant |
| SNOMED CT | 236574008 | Acute rejection of renal transplant |
| SNOMED CT | 236575009 | Acute rejection of renal transplant - grade I |
| SNOMED CT | 236576005 | Acute rejection of renal transplant - grade II |
| SNOMED CT | 236577001 | Acute rejection of renal transplant - grade III |
| SNOMED CT | 236578006 | Chronic rejection of renal transplant |
| SNOMED CT | 236579003 | Chronic rejection of renal transplant - grade I |
| SNOMED CT | 236580000 | Chronic rejection of renal transplant - grade II |
| SNOMED CT | 236581001 | Chronic rejection of renal transplant - grade III |
| SNOMED CT | 236582008 | Acute-on-chronic rejection of renal transplant |
| SNOMED CT | 236583003 | Failed renal transplant |
| SNOMED CT | 236584009 | Perfusion injury of renal transplant |
| SNOMED CT | 236586006 | De novo glomerulonephritis |
| SNOMED CT | 236587002 | Transplant glomerulopathy |
| SNOMED CT | 236588007 | Transplant glomerulopathy - early form |
| SNOMED CT | 236589004 | Transplant glomerulopathy - late form |
| SNOMED CT | 236614007 | Perirenal and periureteric post-transplant lymphocele |
| SNOMED CT | 237499004 | Thyrotoxicosis due to Graves' disease |
| SNOMED CT | 237500008 | Hashitoxicosis - transient |
| SNOMED CT | 237519003 | Autoimmune hypothyroidism |
| SNOMED CT | 237652003 | Insulin resistance - type B |
| SNOMED CT | 237706000 | Autoimmune hypophysitis |
| SNOMED CT | 237790001 | Autoimmune primary ovarian failure |
| SNOMED CT | 237822008 | Autoimmune endocrine disease |
| SNOMED CT | 237824009 | Juvenile Graves' disease |
| SNOMED CT | 238152004 | Sepsis-related gastrointestinal ulceration |
| SNOMED CT | 238428001 | Papular tuberculide |
| SNOMED CT | 238429009 | Nodular tuberculide |
| SNOMED CT | 238476000 | Eosinophilic granuloma of bone |
| SNOMED CT | 238575004 | Allergic contact dermatitis |
| SNOMED CT | 238576003 | Allergic contact dermatitis caused by nickel from implant |
| SNOMED CT | 238577007 | Ingested allergen contact dermatitis |
| SNOMED CT | 238578002 | Occupational allergic contact dermatitis |
| SNOMED CT | 238600001 | Flexural psoriasis |
| SNOMED CT | 238601002 | Unstable psoriasis |

Continued on next page

**TableS31 – continued from previous page**

| CodeSystem | ConceptCode | ConceptName |
|---|---|---|
| SNOMED CT | 238602009 | Psoriasis with eczema |
| SNOMED CT | 238603004 | Eczematized psoriasis |
| SNOMED CT | 238604005 | Psoriasis of nail |
| SNOMED CT | 238605006 | Psoriatic nail dystrophy |
| SNOMED CT | 238606007 | Psoriatic nail pitting |
| SNOMED CT | 238607003 | Psoriatic onycholysis |
| SNOMED CT | 238608008 | Psoriasis of scalp |
| SNOMED CT | 238609000 | Köbner psoriasis |
| SNOMED CT | 238611009 | Lapiere type of psoriasis |
| SNOMED CT | 238613007 | Generalized pustular psoriasis of pregnancy |
| SNOMED CT | 238615000 | Juvenile pustular psoriasis |
| SNOMED CT | 238616004 | Infantile pustular psoriasis |
| SNOMED CT | 238617008 | Circinate and annular pustular psoriasis |
| SNOMED CT | 238672005 | Lichenoid allergic contact dermatitis |
| SNOMED CT | 238675007 | Sarcoidosis-induced erythema nodosum |
| SNOMED CT | 238676008 | Lofgrens syndrome |
| SNOMED CT | 238687000 | Familial cold urticaria |
| SNOMED CT | 238818000 | Non-bullous erythema multiforme |
| SNOMED CT | 238820002 | Erythema multiforme of pregnancy |
| SNOMED CT | 238850005 | Mouth and genital ulcers with inflamed cartilage syndrome |
| SNOMED CT | 238926009 | Lupus erythematosus and erythema multiforme-like syndrome |
| SNOMED CT | 238927000 | Chronic discoid lupus erythematosus |
| SNOMED CT | 238928005 | Chilblain lupus erythematosus |
| SNOMED CT | 238930007 | Sclerodermiform reaction |
| SNOMED CT | 238931006 | Eosinophilic cellulitis |
| SNOMED CT | 238938000 | Prebullous pemphigoid |
| SNOMED CT | 238939008 | Localized pemphigoid |
| SNOMED CT | 238940005 | Pemphigoid vegetans |
| SNOMED CT | 238942002 | Adult linear immunoglobulin A disease |
| SNOMED CT | 238943007 | Eosinophilic spongiosis |
| SNOMED CT | 239098009 | Acropustulosis of infancy |
| SNOMED CT | 239105004 | Transplacental herpes gestationis |
| SNOMED CT | 239147000 | Mast cell nevus |
| SNOMED CT | 239799007 | Early onset polyarticular juvenile chronic arthritis |
| SNOMED CT | 239800006 | Late onset polyarticular juvenile chronic arthritis |
| SNOMED CT | 239802003 | Juvenile psoriatic arthritis |
| SNOMED CT | 239803008 | Juvenile psoriatic arthritis with psoriasis |
| SNOMED CT | 239804002 | Juvenile psoriatic arthritis without psoriasis |
| SNOMED CT | 239806000 | Juvenile spondyloarthropathy |
| SNOMED CT | 239809007 | Juvenile arthritis of inflammatory bowel disease |
| SNOMED CT | 239812005 | Psoriatic arthritis with distal interphalangeal joint involvement |
| SNOMED CT | 239813000 | Psoriatic dactylitis |
| SNOMED CT | 239891002 | Subacute cutaneous lupus erythematosus |
| SNOMED CT | 239892009 | Primary antiphospholipid syndrome |
| SNOMED CT | 239894005 | Primary antiphospholipid syndrome with multisystem involvement |
| SNOMED CT | 239895006 | Secondary antiphospholipid syndrome |

Continued on next page

**TableS31 – continued from previous page**

| CodeSystem | ConceptCode | ConceptName |
|---|---|---|
| SNOMED CT | 239897003 | Secondary antiphospholipid syndrome with multisystem involvement |
| SNOMED CT | 239910001 | Toxic oil syndrome |
| SNOMED CT | 239943002 | Rheumatoid necrotizing vasculitis |
| SNOMED CT | 239946005 | Postinfective immunoglobulin A vasculitis |
| SNOMED CT | 239947001 | Essential mixed cryoglobulinemia |
| SNOMED CT | 240041006 | Rheumatic fever nodule |
| SNOMED CT | 240360007 | Yersinia erythema nodosum |
| SNOMED CT | 240385003 | Septicemic glanders |
| SNOMED CT | 240389009 | Septicemic melioidosis |
| SNOMED CT | 240426001 | Chronic meningococcemia |
| SNOMED CT | 241929008 | Acute allergic reaction |
| SNOMED CT | 241930003 | Anaphylaxis caused by venom |
| SNOMED CT | 241931004 | Bee sting-induced anaphylaxis |
| SNOMED CT | 241932006 | Wasp sting-induced anaphylaxis |
| SNOMED CT | 241933001 | Peanut-induced anaphylaxis |
| SNOMED CT | 241934007 | Seafood-induced anaphylaxis |
| SNOMED CT | 241935008 | Egg white-induced anaphylaxis |
| SNOMED CT | 241936009 | Cow's milk protein-induced anaphylaxis |
| SNOMED CT | 241937000 | Drug-induced anaphylaxis |
| SNOMED CT | 241938005 | Penicillin-induced anaphylaxis |
| SNOMED CT | 241939002 | Insulin-induced anaphylaxis |
| SNOMED CT | 241941001 | Seminal fluid-induced anaphylaxis |
| SNOMED CT | 241952007 | Exercise anaphylaxis |
| SNOMED CT | 241954008 | Idiopathic anaphylaxis |
| SNOMED CT | 241956005 | Venom-induced angioedema-urticaria |
| SNOMED CT | 241957001 | Food-induced angioedema-urticaria |
| SNOMED CT | 241965003 | Latex-induced angioedema-urticaria |
| SNOMED CT | 254289008 | Post-transplant neoplasia |
| SNOMED CT | 255101006 | Sézary disease of skin |
| SNOMED CT | 259097007 | Delayed photoallergic dermatitis |
| SNOMED CT | 1229988003 | Myeloid and/or lymphoid neoplasm with PCM1-JAK2 |
| SNOMED CT | 1204344007 | Myelodysplastic syndrome with single lineage dysplasia |
| SNOMED CT | 16415791000119104 | Myasthenia gravis in remission |
| SNOMED CT | 1204460007 | Myocardial dysfunction with sepsis |
| SNOMED CT | 756003 | Chronic rheumatic myopericarditis |
| SNOMED CT | 961007 | Erythema nodosum, acute form |
| SNOMED CT | 1125006 | Sepsis during labor |
| SNOMED CT | 1824008 | Allergic gastritis |
| SNOMED CT | 2557004 | Chronic leukemia |
| SNOMED CT | 2772003 | Acquired epidermolysis bullosa |
| SNOMED CT | 2858002 | Puerperal sepsis |
| SNOMED CT | 2897005 | Immune thrombocytopenia |
| SNOMED CT | 2990008 | Lymphocytic leukaemoid reaction |
| SNOMED CT | 3533007 | Pustular bacterid |
| SNOMED CT | 3978000 | Warm autoimmune hemolytic anemia |
| SNOMED CT | 4089001 | Meningococcemia |

Continued on next page

**TableS31 – continued from previous page**

| CodeSystem | ConceptCode | ConceptName |
|---|---|---|
| SNOMED CT | 4448006 | Allergic headache |
| SNOMED CT | 5085001 | Gonococcemia |
| SNOMED CT | 1208892009 | Alopecia areata of eyebrow |
| SNOMED CT | 5567000 | Gas gangrene septicemia |
| SNOMED CT | 5860009 | Ophiasis |
| SNOMED CT | 7021009 | Gastrointestinal eosinophilic granuloma |
| SNOMED CT | 7119001 | Cutaneous lupus erythematosus |
| SNOMED CT | 7548000 | Rheumatic pneumonia |
| SNOMED CT | 8090002 | Eosinophilic granuloma of oral mucosa |
| SNOMED CT | 8214000 | Telangiectasia macularis eruptiva perstans |
| SNOMED CT | 8326008 | Acute allergic mucoid otitis media |
| SNOMED CT | 8554002 | Brazilian purpuric fever |
| SNOMED CT | 9012003 | Septicemic plague |
| SNOMED CT | 9014002 | Psoriasis |
| SNOMED CT | 9768003 | Tuberculosis cutis lichenoides |
| SNOMED CT | 10001005 | Bacterial sepsis |
| SNOMED CT | 10294000 | Reactive mastocytosis |
| SNOMED CT | 10713006 | Diffuse interstitial rheumatoid disease of lung |
| SNOMED CT | 10785007 | Vinyard sprayers' lung |
| SNOMED CT | 10803007 | Jarisch Herxheimer reaction |
| SNOMED CT | 10934005 | Cryofibrinogenemia |
| SNOMED CT | 11244009 | Polyglandular autoimmune syndrome, type 1 |
| SNOMED CT | 11352009 | Primary cryoglobulinemia |
| SNOMED CT | 11354005 | Tuberculosis cutis indurativa |
| SNOMED CT | 11373009 | Illegal termination of pregnancy with sepsis |
| SNOMED CT | 11781007 | Autoimmune hemolytic anemia due to complement |
| SNOMED CT | 11944003 | Feather-pickers' disease |
| SNOMED CT | 12088005 | Budgerigar-fanciers' disease |
| SNOMED CT | 12219003 | Aleukemic lymphoid leukemia |
| SNOMED CT | 12263007 | Type 1 hypersensitivity response |
| SNOMED CT | 12761008 | Acute photoallergic dermatitis |
| SNOMED CT | 12834001 | Subacute monocytic leukemia |
| SNOMED CT | 13160009 | Complication of transplanted intestines |
| SNOMED CT | 13172003 | Chronic idiopathic thrombocytopenic purpura |
| SNOMED CT | 13394002 | Suberosis |
| SNOMED CT | 13470001 | Chronic ulcerative ileocolitis |
| SNOMED CT | 14311001 | Severe chronic ulcerative colitis |
| SNOMED CT | 14333004 | Alloimmune neonatal neutropenia |
| SNOMED CT | 14589007 | Phacoanaphylaxis |
| SNOMED CT | 14654002 | Localized anaphylaxis |
| SNOMED CT | 15084002 | Lupus erythematosus profundus |
| SNOMED CT | 15342002 | Chronic ulcerative enterocolitis |
| SNOMED CT | 15470004 | Toxic diffuse goiter with acropachy |
| SNOMED CT | 15676002 | Acute rheumatic pericarditis |
| SNOMED CT | 16623004 | Coffee-workers' lung |
| SNOMED CT | 16751003 | Acute tubulointerstitial nephritis associated with systemic infection |

Continued on next page

**TableS31 – continued from previous page**

| CodeSystem | ConceptCode | ConceptName |
|---|---|---|
| SNOMED CT | 16935003 | Allergic arthritis |
| SNOMED CT | 17866004 | Acute allergic sanguinous otitis media |
| SNOMED CT | 18192007 | Acute rheumatic endocarditis |
| SNOMED CT | 18323000 | Drug-induced immune hemolytic anemia, immune complex type |
| SNOMED CT | 18690003 | Farmers' lung |
| SNOMED CT | 19076009 | Sick building syndrome |
| SNOMED CT | 19087001 | Tinea kerion |
| SNOMED CT | 19267009 | Lupus anticoagulant disorder |
| SNOMED CT | 19274004 | Grain-handlers' disease |
| SNOMED CT | 19307009 | Drug-induced immune thrombocytopenia |
| SNOMED CT | 19514005 | Arthritis mutilans |
| SNOMED CT | 19636003 | Monocytosis |
| SNOMED CT | 20005002 | Recurrent rheumatic heart disease |
| SNOMED CT | 21983002 | Hashimoto thyroiditis |
| SNOMED CT | 22231002 | Allergic enteritis of intestine |
| SNOMED CT | 22310002 | Chronic idiopathic monocytosis |
| SNOMED CT | 22695001 | Aleukemic monocytic leukemia |
| SNOMED CT | 22775002 | Allergic vulvitis |
| SNOMED CT | 22888007 | Drug-induced lupus erythematosus due to procainamide |
| SNOMED CT | 22972008 | Erythema multiforme, dermal type |
| SNOMED CT | 23315001 | Sequoiosis |
| SNOMED CT | 24159007 | Persistent lymphocytosis |
| SNOMED CT | 24363009 | Rheumatic fever without heart involvement |
| SNOMED CT | 24444009 | Failed attempted termination of pregnancy with sepsis |
| SNOMED CT | 24526004 | Inflammatory bowel disease |
| SNOMED CT | 24700007 | Multiple sclerosis |
| SNOMED CT | 24829000 | Eosinophilic ulcerative colitis |
| SNOMED CT | 25042006 | Tracheostomy sepsis |
| SNOMED CT | 25044007 | Neuromyelitis optica |
| SNOMED CT | 25847004 | Seborrheic psoriasis |
| SNOMED CT | 25887009 | Allergic sigmoiditis |
| SNOMED CT | 25897000 | Malt-workers' lung |
| SNOMED CT | 26006005 | Allergic gastroenteritis |
| SNOMED CT | 26045000 | Chronic allergic conjunctivitis |
| SNOMED CT | 26151008 | Graves' disease with acropachy AND with thyrotoxic crisis |
| SNOMED CT | 26169004 | Allergic otitis media |
| SNOMED CT | 26843008 | Antiphospholipid syndrome |
| SNOMED CT | 27040004 | Experimental allergic encephalomyelitis |
| SNOMED CT | 28122003 | Pulmonary eosinophilic granuloma |
| SNOMED CT | 28505005 | Acute idiopathic thrombocytopenic purpura |
| SNOMED CT | 28664002 | Herpes iris |
| SNOMED CT | 28807005 | Secondary cryoglobulinemia |
| SNOMED CT | 28840001 | Generalized pustular psoriasis, exanthematous type |
| SNOMED CT | 29120000 | Eosinophilic colitis |
| SNOMED CT | 29789008 | Monocytic leukemoid reaction |
| SNOMED CT | 30304000 | Allergic colitis |
| SNOMED CT | 30911005 | Cryoglobulinemia |

Continued on next page

**TableS31 – continued from previous page**

| CodeSystem | ConceptCode | ConceptName |
|---|---|---|
| SNOMED CT | 30981000 | Secondary eosinophilia |
| SNOMED CT | 31712002 | Primary biliary cholangitis |
| SNOMED CT | 32273002 | Idiopathic thrombocytopenic purpura |
| SNOMED CT | 32295003 | Allergic ileitis |
| SNOMED CT | 32861005 | Erythema nodosum |
| SNOMED CT | 33167004 | Complication of transplanted liver |
| SNOMED CT | 33339001 | Psoriasis with arthropathy |
| SNOMED CT | 33560006 | Subacute lymphoid leukemia |
| SNOMED CT | 33719002 | Subcutaneous rheumatoid nodule |
| SNOMED CT | 34015007 | Bakers' asthma |
| SNOMED CT | 34250006 | Benign mucous membrane pemphigoid |
| SNOMED CT | 34739009 | Urticaria pigmentosa, adult form |
| SNOMED CT | 35154004 | Pemphigus foliaceus |
| SNOMED CT | 35703006 | Post-viral paroxysmal cold hemoglobinuria |
| SNOMED CT | 35750002 | Aleukemic myeloid leukemia |
| SNOMED CT | 36715001 | Erythema multiforme |
| SNOMED CT | 36739006 | Pemphigus erythematosus |
| SNOMED CT | 37042000 | Guttate psoriasis |
| SNOMED CT | 37471005 | Extrinsic allergic alveolitis |
| SNOMED CT | 37981002 | Allergic bronchopulmonary aspergillosis |
| SNOMED CT | 38064006 | Reactive monocytosis |
| SNOMED CT | 38675009 | Monoclonal cryoimmunoglobulinemia |
| SNOMED CT | 38729007 | Wheat weevil disease |
| SNOMED CT | 38877003 | Rheumatoid aortitis |
| SNOMED CT | 38933006 | Recurrent rheumatic fever |
| SNOMED CT | 39246002 | Legal termination of pregnancy with sepsis |
| SNOMED CT | 39579001 | Anaphylaxis |
| SNOMED CT | 40178009 | Allergic urticaria |
| SNOMED CT | 40712008 | Allergic contact dermatitis caused by Arnica |
| SNOMED CT | 41030002 | Coccidioidomycosis with erythema multiforme |
| SNOMED CT | 41156006 | Tuberculosis papulonecrotica |
| SNOMED CT | 41816006 | Secondary cryofibrinogenemia |
| SNOMED CT | 41864002 | Autoimmune polyendocrinopathy |
| SNOMED CT | 42852008 | Infectious lymphocytosis |
| SNOMED CT | 42939002 | Pyemia |
| SNOMED CT | 43355006 | Eosinopenia |
| SNOMED CT | 44165003 | Complication of transplanted lung |
| SNOMED CT | 44206008 | Hapten type low affinity hemolytic anemia |
| SNOMED CT | 45309006 | Allergic contact dermatitis due to secretions |
| SNOMED CT | 46165000 | Chronic rheumatic endocarditis |
| SNOMED CT | 46359005 | Neutropenia associated with infectious disease |
| SNOMED CT | 46459009 | Brazilian pemphigus foliaceus |
| SNOMED CT | 46586006 | Diffuse alopecia areata |
| SNOMED CT | 46826000 | Rheumatic chorea |
| SNOMED CT | 46848006 | Sepsis following molar AND/OR ectopic pregnancy |
| SNOMED CT | 47144000 | Acute neutrophilia |
| SNOMED CT | 47986005 | Genetic anomaly of leukocyte |

Continued on next page

**TableS31 – continued from previous page**

| CodeSystem | ConceptCode | ConceptName |
|---|---|---|
| SNOMED CT | 48269007 | Allergic parotitis |
| SNOMED CT | 48516007 | Acute infectious lymphocytosis |
| SNOMED CT | 48813009 | Lymphocytopenia |
| SNOMED CT | 49237006 | Allergic diarrhea |
| SNOMED CT | 49420001 | Pemphigus vulgaris |
| SNOMED CT | 50253007 | Secondary paroxysmal cold hemoglobinuria |
| SNOMED CT | 50776006 | Allergic encephalomyelitis |
| SNOMED CT | 51264003 | Limbal AND/OR corneal involvement in vernal conjunctivitis |
| SNOMED CT | 51831006 | Sepsis following infusion, injection, transfusion AND/OR vaccination |
| SNOMED CT | 52168008 | Monocytic leukemia |
| SNOMED CT | 52333004 | Mushroom workers' lung |
| SNOMED CT | 52506002 | Chronic ulcerative rectosigmoiditis |
| SNOMED CT | 54867000 | Rheumatoid fibrosing alveolitis |
| SNOMED CT | 54912002 | Drug-induced lupus erythematosus due to hydralazine |
| SNOMED CT | 55444004 | Transient neonatal neutropenia |
| SNOMED CT | 55807009 | Exophthalmos due to toxic diffuse goiter |
| SNOMED CT | 55985003 | Atopic reaction |
| SNOMED CT | 56478004 | Leukemoid reaction |
| SNOMED CT | 56968009 | Asthma caused by wood dust |
| SNOMED CT | 57803009 | Rheumatic chorea with heart involvement |
| SNOMED CT | 58718002 | Rheumatic fever |
| SNOMED CT | 58797008 | Disorder of transplanted kidney |
| SNOMED CT | 59165007 | Rheumatoid scleritis |
| SNOMED CT | 59275002 | Acute allergic serous otitis media |
| SNOMED CT | 59940009 | Hypersensitivity alveolitis in lungworm infection |
| SNOMED CT | 59957008 | Neonatal Graves' disease |
| SNOMED CT | 60216004 | Toxic diffuse goiter with thyrotoxic crisis |
| SNOMED CT | 60268006 | Toxic diffuse goiter with pretibial myxedema |
| SNOMED CT | 60967006 | Allergic vaginitis |
| SNOMED CT | 61012002 | Chronic rheumatic myocarditis |
| SNOMED CT | 61458006 | Drug-induced lupus erythematosus due to diphenylhydantoin |
| SNOMED CT | 61582004 | Allergic rhinitis |
| SNOMED CT | 61802005 | Primary cryofibrinogenemia |
| SNOMED CT | 62853008 | Anti-glomerular basement membrane tubulointerstitial nephritis |
| SNOMED CT | 62871001 | Idiopathic paroxysmal cold hemoglobinuria |
| SNOMED CT | 63088003 | Allergic asthma without status asthmaticus |
| SNOMED CT | 63175003 | Localized extracutaneous mastocytosis |
| SNOMED CT | 63484008 | Drug-induced neutrophilia |
| SNOMED CT | 64072006 | Acute rheumatic pancarditis |
| SNOMED CT | 64249002 | Allergic eosinophilia |
| SNOMED CT | 64766004 | Ulcerative colitis |
| SNOMED CT | 64936001 | Simple pulmonary eosinophilia |
| SNOMED CT | 65172003 | Pemphigus |
| SNOMED CT | 65394002 | Allergic contact dermatitis caused by urushiol from Chinese lacquer tree |
| SNOMED CT | 65539006 | Impetigo herpetiformis |

Continued on next page

**TableS31 – continued from previous page**

| CodeSystem | ConceptCode | ConceptName |
|---|---|---|
| SNOMED CT | 66291003 | Tuberculous phlyctenular keratoconjunctivitis |
| SNOMED CT | 66628005 | Toxic diffuse goiter with exophthalmos AND with thyrotoxic storm |
| SNOMED CT | 66944004 | Autoimmune thyroiditis |
| SNOMED CT | 67023009 | Lymphocytosis |
| SNOMED CT | 67242002 | Bagassosis |
| SNOMED CT | 67465009 | Miscarriage with sepsis |
| SNOMED CT | 67678004 | Acute atopic conjunctivitis |
| SNOMED CT | 67824002 | Lymphosarcoma |
| SNOMED CT | 67895005 | Phlyctenular keratoconjunctivitis |
| SNOMED CT | 68225006 | Alopecia areata |
| SNOMED CT | 68333005 | Furriers' lung |
| SNOMED CT | 348741000119101 | Corneal infiltrate of left eye following penetrating keratoplasty |
| SNOMED CT | 348291000119100 | Corneal infiltrate of right eye following penetrating keratoplasty |
| SNOMED CT | 1217301006 | Precursor cell lymphoblastic lymphoma |
| SNOMED CT | 722386009 | Celiac disease with epilepsy and cerebral calcification syndrome |
| SNOMED CT | 722401001 | Severe fever with thrombocytopenia syndrome virus |
| SNOMED CT | 722453009 | Hennekam Beemer syndrome |
| SNOMED CT | 722850002 | Crohn's disease of gastrointestinal anastomosis |
| SNOMED CT | 722870008 | IgG4-related sclerosing cholangitis |
| SNOMED CT | 722872000 | Autoimmune pancreatitis type 1 |
| SNOMED CT | 722925004 | Transient neonatal neutropenia due to congenital viral infection |
| SNOMED CT | 722926003 | Transient neonatal neutropenia due to neonatal bacterial sepsis |
| SNOMED CT | 722991004 | Autoimmune myopathy |
| SNOMED CT | 723004005 | Autoimmune bullous dermatosis |
| SNOMED CT | 723022008 | Autoimmune keratitis |
| SNOMED CT | 723674005 | Simple cryoglobulinemia |
| SNOMED CT | 724603009 | Pediatric onset systemic sclerosis |
| SNOMED CT | 724643004 | Transient abnormal myelopoiesis co-occurrent with Down syndrome |
| SNOMED CT | 724778008 | Progressive relapsing multiple sclerosis |
| SNOMED CT | 724782005 | Demyelination of central nervous system co-occurrent and due to Sjogren disease |
| SNOMED CT | 724809006 | Autoimmune retinopathy |
| SNOMED CT | 724825001 | Allergic contact dermatitis caused by hairdressing product |
| SNOMED CT | 724826000 | Allergic contact blepharoconjunctivitis |
| SNOMED CT | 725415009 | House allergic alveolitis |
| SNOMED CT | 726078000 | Sporadic Blau syndrome |
| SNOMED CT | 726101009 | Extraintestinal Crohn's |
| SNOMED CT | 732960002 | Secondary autoimmune hemolytic anemia co-occurrent and due to systemic lupus erythematosus |
| SNOMED CT | 732962005 | Secondary autoimmune hemolytic anemia co-occurrent and due to chronic inflammatory disease |
| SNOMED CT | 732963000 | Secondary autoimmune hemolytic anemia co-occurrent and due to lymphoproliferative disorder |
| SNOMED CT | 732965007 | Secondary autoimmune hemolytic anemia co-occurrent and due to rheumatic disorder |

Continued on next page

**TableS31 – continued from previous page**

| CodeSystem | ConceptCode | ConceptName |
|---|---|---|
| SNOMED CT | 732966008 | Secondary autoimmune hemolytic anemia co-occurrent and due to ulcerative colitis |
| SNOMED CT | 733142005 | Sepsis following obstructed labor |
| SNOMED CT | 733155006 | IgE mediated allergic enteritis of small intestine |
| SNOMED CT | 733157003 | Crohn disease of appendix |
| SNOMED CT | 733207001 | Necrosis of myocutaneous flap |
| SNOMED CT | 735173007 | Shock due to anaphylaxis |
| SNOMED CT | 735434003 | Acquired neutrophilia |
| SNOMED CT | 735435002 | Constitutional neutrophilia |
| SNOMED CT | 735439008 | Constitutional eosinopenia |
| SNOMED CT | 735440005 | Acquired eosinopenia |
| SNOMED CT | 735442002 | Acquired eosinophilia |
| SNOMED CT | 735444001 | Acquired lymphocytosis |
| SNOMED CT | 735445000 | Allergic disorder of respiratory tract |
| SNOMED CT | 735447008 | Anaphylaxis caused by insect venom |
| SNOMED CT | 735448003 | Anaphylaxis due to mast cell disorder |
| SNOMED CT | 735454002 | Allergic bronchospasm caused by dietary substance |
| SNOMED CT | 735588005 | Uncomplicated allergic asthma |
| SNOMED CT | 735637004 | Sepsis of neonate caused by Streptococcus pyogenes |
| SNOMED CT | 735638009 | Sepsis of fetus caused by Streptococcus pyogenes |
| SNOMED CT | 735975001 | Shock co-occurrent and due to anaphylaxis caused by serum |
| SNOMED CT | 737193000 | Allergic enteritis of small intestine |
| SNOMED CT | 737195007 | Crohn disease of upper gastrointestinal tract |
| SNOMED CT | 737249005 | Photodermatitis co-occurrent and due to autoimmune disease |
| SNOMED CT | 737315000 | Food-protein induced enterocolitis syndrome |
| SNOMED CT | 737316004 | Food protein-induced colitis in infant |
| SNOMED CT | 737317008 | Food protein-induced proctitis in infant |
| SNOMED CT | 762282007 | Immunoglobulin G4 related pachymeningitis |
| SNOMED CT | 762303003 | Pediatric onset Sjögren syndrome |
| SNOMED CT | 762375006 | Erythema multiforme due to viral disease |
| SNOMED CT | 762379000 | Allergic contact dermatitis caused by dental material |
| SNOMED CT | 762380002 | Allergic contact dermatitis caused by food flavor |
| SNOMED CT | 762381003 | Allergic contact dermatitis caused by food additive |
| SNOMED CT | 762382005 | Allergic contact dermatitis caused by agrochemical biocide |
| SNOMED CT | 762383000 | Allergic contact dermatitis caused by cutting oil |
| SNOMED CT | 762384006 | Allergic contact dermatitis caused by disinfectant |
| SNOMED CT | 762386008 | Allergic contact dermatitis caused by rubber compound |
| SNOMED CT | 762387004 | Allergic contact dermatitis caused by systemic medicament |
| SNOMED CT | 762455001 | Anaphylaxis caused by low temperature |
| SNOMED CT | 762521001 | Acute allergic asthma |
| SNOMED CT | 762528007 | Sepsis caused by Klebsiella pneumoniae |
| SNOMED CT | 762529004 | Sepsis caused by Acinetobacter baumannii |
| SNOMED CT | 762545002 | Occupational allergic contact dermatitis caused by footwear |
| SNOMED CT | 762548000 | Occupational allergic contact dermatitis caused by cosmetic |
| SNOMED CT | 762549008 | Occupational allergic contact dermatitis caused by fragrance |
| SNOMED CT | 762552000 | Occupational allergic contact dermatitis caused by dental material |

Continued on next page

**TableS31 – continued from previous page**

| CodeSystem | ConceptCode | ConceptName |
|---|---|---|
| SNOMED CT | 762554004 | Occupational allergic contact dermatitis caused by food flavor |
| SNOMED CT | 762555003 | Occupational allergic contact dermatitis caused by food additive |
| SNOMED CT | 762558001 | Occupational allergic contact dermatitis caused by hairdressing product |
| SNOMED CT | 762560004 | Occupational allergic contact dermatitis caused by agrochemical biocide |
| SNOMED CT | 762561000 | Occupational allergic contact dermatitis caused by cutting oil |
| SNOMED CT | 762562007 | Occupational allergic contact dermatitis caused by disinfectant |
| SNOMED CT | 762566005 | Occupational allergic contact dermatitis caused by metal compound |
| SNOMED CT | 762570002 | Occupational allergic contact dermatitis caused by industrial preservative |
| SNOMED CT | 762571003 | Occupational allergic contact dermatitis caused by biocide |
| SNOMED CT | 762574006 | Occupational allergic contact dermatitis caused by rubber compound |
| SNOMED CT | 762576008 | Occupational allergic contact dermatitis caused by systemic drug or medicament |
| SNOMED CT | 762578009 | Occupational allergic contact dermatitis caused by topical medicament |
| SNOMED CT | 762618008 | Bronchiolitis obliterans syndrome due to and after lung transplantation |
| SNOMED CT | 762726008 | Occupational allergic contact dermatitis caused by clothing |
| SNOMED CT | 762754001 | Allergic contact dermatitis caused by fragrance |
| SNOMED CT | 762755000 | Allergic contact dermatitis caused by footwear |
| SNOMED CT | 762757008 | Allergic contact dermatitis caused by clothing |
| SNOMED CT | 763630007 | Satoyoshi syndrome |
| SNOMED CT | 763793004 | Limbic encephalitis with contactin-associated protein-like 2 antibodies |
| SNOMED CT | 763794005 | Limbic encephalitis with leucine-rich glioma-inactivated 1 antibodies |
| SNOMED CT | 763864008 | Persistent polyclonal B-cell lymphocytosis |
| SNOMED CT | 765106006 | Early-onset neonatal sepsis |
| SNOMED CT | 765107002 | Late-onset neonatal sepsis |
| SNOMED CT | 765751002 | Autoimmune encephalopathy with parasomnia and obstructive sleep apnea |
| SNOMED CT | 766049000 | Acute sensory ataxic neuropathy |
| SNOMED CT | 766246000 | Marburg acute multiple sclerosis |
| SNOMED CT | 766252004 | Juvenile overlap myositis |
| SNOMED CT | 766722008 | Paraparetic variant of Guillain-Barré syndrome |
| SNOMED CT | 770349000 | Sepsis caused by virus |
| SNOMED CT | 770545005 | Allergic contact dermatitis caused by iodine |
| SNOMED CT | 770588002 | Allergic contact dermatitis caused by iodophore |
| SNOMED CT | 770589005 | Allergic contact dermatitis caused by povidone iodine |
| SNOMED CT | 771145006 | Herpetiform pemphigus |
| SNOMED CT | 771271000 | Steroid-responsive encephalopathy associated with autoimmune thyroiditis |
| SNOMED CT | 773275000 | Post-transplant acute limbic encephalitis |

Continued on next page

**TableS31 – continued from previous page**

| CodeSystem | ConceptCode | ConceptName |
|---|---|---|
| SNOMED CT | 773662009 | Neonatal inflammatory skin and bowel disease |
| SNOMED CT | 774080007 | Neonatal scleroderma |
| SNOMED CT | 774082004 | Neonatal dermatomyositis |
| SNOMED CT | 774083009 | Neonatal autoimmune hemolytic anemia |
| SNOMED CT | 774084003 | Neonatal antiphospholipid syndrome |
| SNOMED CT | 778050009 | Idiopathic eosinophilic myositis |
| SNOMED CT | 781474001 | Allergic disorder |
| SNOMED CT | 782309008 | Immune-mediated neuropathy |
| SNOMED CT | 782513000 | Acute severe exacerbation of allergic asthma |
| SNOMED CT | 782520007 | Exacerbation of allergic asthma due to infection |
| SNOMED CT | 782866008 | Eosinophilic peritonitis |
| SNOMED CT | 783010003 | Pharyngeal-cervical-brachial variant of Guillain-Barré syndrome |
| SNOMED CT | 783187005 | Aseptic peritoneal eosinophilia after dialysis |
| SNOMED CT | 783205005 | Alopecia antibody deficiency |
| SNOMED CT | 783211008 | Non-chronic lymphocytic leukemia monoclonal B-cell lymphocytosis |
| SNOMED CT | 783244002 | Acute pure sensory neuropathy |
| SNOMED CT | 783566008 | Mucocutaneous ulcer caused by Epstein-Barr virus |
| SNOMED CT | 784327005 | Pustular psoriasis of sole of foot |
| SNOMED CT | 784328000 | Pustular psoriasis of palm of hand |
| SNOMED CT | 786855003 | Monoclonal B-cell lymphocytosis chronic lymphocytic leukemia-type |
| SNOMED CT | 788613004 | Anti-glomerular basement membrane disease |
| SNOMED CT | 788718000 | Iritis co-occurrent with inflammatory bowel disease |
| SNOMED CT | 788803005 | Allergic reaction to galactose-alpha 1,3 galactose |
| SNOMED CT | 789043007 | Sepsis without septic shock |
| SNOMED CT | 789296007 | Phlyctenular keratoconjunctivitis caused by staphylococcus toxin |
| SNOMED CT | 789697006 | Mast cell gastritis |
| SNOMED CT | 792842004 | Coronary arteriosclerosis in artery of transplanted heart |
| SNOMED CT | 816984002 | Progressive multiple sclerosis |
| SNOMED CT | 818950005 | Blau syndrome |
| SNOMED CT | 829971006 | Non-amyloid monoclonal immunoglobulin deposition disease |
| SNOMED CT | 829976001 | Thunderstorm asthma |
| SNOMED CT | 838324008 | Autoimmune optic neuropathy |
| SNOMED CT | 838330008 | Autoimmune ganglionopathy |
| SNOMED CT | 838359008 | Acute cutaneous lupus erythematosus |
| SNOMED CT | 838366009 | Anti-synthetase syndrome due to dermatomyositis |
| SNOMED CT | 838367000 | Allergic rhinosinusitis caused by Aspergillus |
| SNOMED CT | 838551007 | Autoimmune progesterone dermatitis |
| SNOMED CT | 840505007 | Down syndrome co-occurrent with leukemoid reaction associated transient neonatal pustulosis |
| SNOMED CT | 846606007 | Graft versus host disease of liver |
| SNOMED CT | 860800001 | Erythema multiforme due to systemic fungal infection |
| SNOMED CT | 860801002 | Erythema nodosum due to systemic fungal infection |
| SNOMED CT | 860824009 | Eosinophilia due to infectious disease |

Continued on next page

**TableS31 – continued from previous page**

| CodeSystem | ConceptCode | ConceptName |
|---|---|---|
| SNOMED CT | 860825005 | Hypertrophic pachymeningitis due to disorder of immune function |
| SNOMED CT | 860827002 | Eosinopenia due to infectious disease |
| SNOMED CT | 860869003 | Erythema multiforme due to Herpes simplex infection |
| SNOMED CT | 860870002 | Erythema multiforme due to Mycoplasma pneumoniae infection |
| SNOMED CT | 860924005 | Allergic contact dermatitis caused by alkali-metal soap |
| SNOMED CT | 866046008 | Mucous membrane pemphigoid of penile mucosa |
| SNOMED CT | 866084002 | Lymphopenia due to infection |
| SNOMED CT | 866151004 | Lymphocytopenia due to Severe acute respiratory syndrome coronavirus 2 |
| SNOMED CT | 870588003 | Sepsis due to disease caused by severe acute respiratory syndrome coronavirus 2 |
| SNOMED CT | 870749003 | Allergic contact dermatitis due to usnic acid |
| SNOMED CT | 871920005 | Allergic reaction to insect bite |
| SNOMED CT | 871926004 | Anaphylaxis caused by insect bite and/or insect sting |
| SNOMED CT | 871927008 | Allergic reaction caused by insect bite and/or insect sting |
| SNOMED CT | 871930001 | Anaphylaxis after allergen immunotherapy |
| SNOMED CT | 871931002 | Allergic reaction after allergen immunotherapy |
| SNOMED CT | 2701000124105 | Recurrent acute disseminated encephalomyelitis |
| SNOMED CT | 8161000119106 | Arthritis co-occurrent and due to Crohn's disease |
| SNOMED CT | 22241000175107 | Sepsis due to methicillin-sensitive Staphylococcus aureus |
| SNOMED CT | 32751000119100 | Line sepsis associated with dialysis catheter |
| SNOMED CT | 61311000119104 | Acute myeloid leukemia, minimal differentiation, FAB M0 in remission |
| SNOMED CT | 61321000119106 | Acute myeloid leukemia without maturation, FAB M1 in remission |
| SNOMED CT | 68261000119102 | Diffuse non-Hodgkin's lymphoma Lugano stage I |
| SNOMED CT | 68271000119108 | Diffuse non-Hodgkin's lymphoma Lugano stage II |
| SNOMED CT | 68291000119109 | Diffuse non-Hodgkin's lymphoma Lugano stage IV |
| SNOMED CT | 72161000119100 | Antiphospholipid syndrome in pregnancy |
| SNOMED CT | 72631000119101 | Human immunodeficiency virus (HIV) II infection category B2 |
| SNOMED CT | 76981000119106 | Human immunodeficiency virus (HIV) infection category B1 |
| SNOMED CT | 76991000119109 | Human immunodeficiency virus (HIV) infection category B2 |
| SNOMED CT | 84811000119107 | Follicular dendritic sarcoma of intraabdominal lymph node |
| SNOMED CT | 84831000119102 | Follicular dendritic sarcoma of lymph nodes of multiple sites |
| SNOMED CT | 89681000119101 | Glomerulonephritis co-occurrent and due to scleroderma |
| SNOMED CT | 99031000119107 | Acute exacerbation of asthma co-occurrent with allergic rhinitis |
| SNOMED CT | 103781000119103 | Allergic bronchopulmonary mycosis |
| SNOMED CT | 116711000119103 | Marginal zone lymphoma of inguinal lymph node |
| SNOMED CT | 116741000119104 | Marginal zone lymphoma of thoracic lymph node |
| SNOMED CT | 117061000119101 | Peripheral T-cell lymphoma of spleen |
| SNOMED CT | 117091000119108 | Peripheral T-cell lymphoma of axillary lymph node |
| SNOMED CT | 117111000119100 | Peripheral T-cell lymphoma of thoracic lymph node |
| SNOMED CT | 119031000119101 | Myelofibrosis due to another disorder |
| SNOMED CT | 121121000119106 | Acute graft-versus-host reaction following bone marrow transplant |
| SNOMED CT | 121881000119102 | Intractable allergic migraine |

Continued on next page

**TableS31 – continued from previous page**

| CodeSystem | ConceptCode | ConceptName |
|---|---|---|
| SNOMED CT | 122501000119101 | Lymphoproliferative disorder following lung transplant |
| SNOMED CT | 122511000119103 | Lymphoproliferative disorder following heart transplantation |
| SNOMED CT | 122521000119105 | Lymphoproliferative disorder following liver transplant |
| SNOMED CT | 122531000119108 | Lymphoproliferative disorder following kidney transplant |
| SNOMED CT | 122881000119107 | Chronic monocytic leukemia in relapse |
| SNOMED CT | 122901000119109 | Myeloid leukemia in relapse |
| SNOMED CT | 122951000119108 | Lymphoid leukemia in relapse |
| SNOMED CT | 122961000119105 | Chronic lymphoid leukemia in relapse |
| SNOMED CT | 122981000119101 | Plasma cell leukemia in relapse |
| SNOMED CT | 123571000119104 | Erythema multiforme minor |
| SNOMED CT | 123781000119107 | Chronic leukemia in relapse |
| SNOMED CT | 124991000119109 | Severe persistent asthma co-occurrent with allergic rhinitis |
| SNOMED CT | 125001000119103 | Moderate persistent asthma co-occurrent with allergic rhinitis |
| SNOMED CT | 125011000119100 | Mild persistent asthma co-occurrent with allergic rhinitis |
| SNOMED CT | 125021000119107 | Intermittent asthma co-occurrent with allergic rhinitis |
| SNOMED CT | 126811000119102 | Sepsis without acute organ dysfunction caused by Serratia species |
| SNOMED CT | 127121000119101 | Sepsis without acute organ dysfunction caused by Streptococcus pneumoniae |
| SNOMED CT | 127141000119107 | Septic shock co-occurrent with acute organ dysfunction due to anaerobic bacteria |
| SNOMED CT | 127171000119100 | Septic shock co-occurrent with acute organ dysfunction due to Haemophilus influenzae |
| SNOMED CT | 127241000119104 | Septic shock co-occurrent with acute organ dysfunction due to Staphylococcus |
| SNOMED CT | 127751000119108 | Sepsis caused by Staphylococcus without acute organ dysfunction |
| SNOMED CT | 127961000119108 | Aleukemic myeloid leukemia in relapse |
| SNOMED CT | 128631000119109 | Chronic graft-versus-host disease following kidney transplant |
| SNOMED CT | 128971000119101 | Cryoglobulinemia due to chronic hepatitis C |
| SNOMED CT | 129571000119102 | Allergic reaction due to allergy skin test |
| SNOMED CT | 129781000119105 | Neoplasia following lung transplant |
| SNOMED CT | 137421000119106 | Graves' disease in remission |
| SNOMED CT | 143441000119108 | Rheumatoid arthritis in remission |
| SNOMED CT | 145611000119107 | Allergic migraine |
| SNOMED CT | 156051000119109 | Allergic rhinitis due to animal hair and dander |
| SNOMED CT | 182961000119101 | Acute disseminated encephalomyelitis following infectious disease |
| SNOMED CT | 292991000119106 | Eaton Lambert syndrome without underlying malignancy |
| SNOMED CT | 300961000119108 | Juvenile idiopathic arthritis of left hand |
| SNOMED CT | 301051000119101 | Juvenile idiopathic arthritis of right hand |
| SNOMED CT | 318871000119107 | Seropositive rheumatoid arthritis of left hip |
| SNOMED CT | 318881000119105 | Seropositive rheumatoid arthritis of left knee |
| SNOMED CT | 318941000119109 | Seropositive rheumatoid arthritis of right hip |
| SNOMED CT | 318951000119106 | Seropositive rheumatoid arthritis of right knee |
| SNOMED CT | 319031000119108 | Seronegative rheumatoid arthritis of left hand |
| SNOMED CT | 319111000119104 | Seronegative rheumatoid arthritis of right hand |

Continued on next page

**TableS31 – continued from previous page**

| CodeSystem | ConceptCode | ConceptName |
|---|---|---|
| SNOMED CT | 319861000119106 | Rheumatoid nodule of subcutaneous tissue of left elbow |
| SNOMED CT | 319871000119100 | Rheumatoid nodule of subcutaneous tissue of left hand |
| SNOMED CT | 319941000119103 | Rheumatoid nodule of subcutaneous tissue of right elbow |
| SNOMED CT | 319951000119101 | Rheumatoid nodule of subcutaneous tissue of right hand |
| SNOMED CT | 333131000119102 | Phlyctenular keratoconjunctivitis of right eye |
| SNOMED CT | 335981000119102 | Chronic giant papillary conjunctivitis of right eye |
| SNOMED CT | 336601000119106 | Acute atopic conjunctivitis of right eye |
| SNOMED CT | 338741000119102 | Phlyctenular keratoconjunctivitis of left eye |
| SNOMED CT | 341591000119109 | Chronic giant papillary conjunctivitis of left eye |
| SNOMED CT | 342211000119106 | Acute atopic conjunctivitis of left eye |
| SNOMED CT | 342761000119104 | Allergic contact dermatitis of right upper eyelid |
| SNOMED CT | 342771000119105 | Allergic contact dermatitis of right lower eyelid |
| SNOMED CT | 342781000119108 | Allergic contact dermatitis of right eyelid |
| SNOMED CT | 342801000119107 | Allergic contact dermatitis of left lower eyelid |
| SNOMED CT | 342811000119105 | Allergic contact dermatitis of left eyelid |
| SNOMED CT | 344511000119102 | Bilateral phlyctenular keratoconjunctivitis |
| SNOMED CT | 346811000119109 | Bilateral chronic giant papillary conjunctivitis of eyes |
| SNOMED CT | 347371000119108 | Acute atopic conjunctivitis of bilateral eyes |
| SNOMED CT | 428241000124101 | Anaphylaxis caused by human papillomavirus vaccine |
| SNOMED CT | 428281000124107 | Anaphylactic reaction caused by diphtheria and tetanus vaccine |
| SNOMED CT | 428291000124105 | Anaphylaxis caused by tetanus, diphtheria and acellular pertussis vaccine |
| SNOMED CT | 428301000124106 | Anaphylaxis caused by meningococcal vaccine |
| SNOMED CT | 428321000124101 | Anaphylaxis due to Hepatitis B vaccine |
| SNOMED CT | 432451000124103 | Multiphasic acute disseminated encephalomyelitis |
| SNOMED CT | 433621000124101 | Anaphylaxis due to Hemophilus influenzae type b vaccine |
| SNOMED CT | 435181000124108 | Sepsis of newborn due to Streptococcus agalactiae |
| SNOMED CT | 436741000124105 | septicaemia associated with vascular access catheter |
| SNOMED CT | 436751000124107 | Sepsis associated with internal vascular access |
| SNOMED CT | 450521000124109 | Marginal zone lymphoma of axillary lymph node |
| SNOMED CT | 451011000124102 | Allergic conjunctivitis of left eye |
| SNOMED CT | 452281000124106 | Autoimmune encephalitis caused by N-methyl D-aspartate receptor antibody |
| SNOMED CT | 452291000124109 | Relapse multiple myeloma |
| SNOMED CT | 456911000124108 | Diffuse non-Hodgkin's lymphoma Lugano stage III |
| SNOMED CT | 459911000124100 | Rheumatoid factor positive rheumatoid arthritis |
| SNOMED CT | 459921000124108 | Rheumatoid factor positive rheumatoid arthritis of the shoulder |
| SNOMED CT | 461311000124107 | Refractory Hodgkin's lymphoma |
| SNOMED CT | 461351000124108 | Breast implant-associated anaplastic large cell lymphoma |
| SNOMED CT | 679351000119102 | Allergic conjunctivitis of bilateral eyes |
| SNOMED CT | 679371000119106 | Allergic conjunctivitis of right eye |
| SNOMED CT | 1073681000119109 | Rheumatoid arthritis of left ankle |
| SNOMED CT | 1073691000119107 | Rheumatoid arthritis of left elbow |
| SNOMED CT | 1073711000119105 | Rheumatoid arthritis of left hand |
| SNOMED CT | 1073721000119103 | Rheumatoid arthritis of left hip |
| SNOMED CT | 1073741000119109 | Rheumatoid arthritis of left shoulder |
| SNOMED CT | 1073751000119106 | Rheumatoid arthritis of left wrist |

Continued on next page

**TableS31 – continued from previous page**

| CodeSystem | ConceptCode | ConceptName |
|---|---|---|
| SNOMED CT | 1073761000119108 | Rheumatoid arthritis of right ankle |
| SNOMED CT | 1073771000119102 | Rheumatoid arthritis of right elbow |
| SNOMED CT | 1073791000119101 | Rheumatoid arthritis of right hand |
| SNOMED CT | 1073811000119102 | Rheumatoid arthritis of right knee |
| SNOMED CT | 1073821000119109 | Rheumatoid arthritis of right shoulder |
| SNOMED CT | 1073831000119107 | Rheumatoid arthritis of right wrist |
| SNOMED CT | 1073861000119104 | Deformity of right hand co-occurrent and due to rheumatoid arthritis |
| SNOMED CT | 1073871000119105 | Deformity of left hand co-occurrent and due to rheumatoid arthritis |
| SNOMED CT | 1073881000119108 | Deformity of right foot co-occurrent and due to rheumatoid arthritis |
| SNOMED CT | 1073891000119106 | Deformity of left foot co-occurrent and due to rheumatoid arthritis |
| SNOMED CT | 1083041000119104 | Acute recurrent allergic otitis media of bilateral middle ears |
| SNOMED CT | 1083081000119109 | Acute allergic serous otitis media of bilateral middle ears |
| SNOMED CT | 1083191000119108 | Autoimmune disorder of bilateral inner ears |
| SNOMED CT | 1085131000119105 | Abscess of intestine co-occurrent and due to chronic ulcerative pancolitis |
| SNOMED CT | 1085231000119100 | Abscess of intestine co-occurrent and due to chronic ulcerative rectosigmoiditis |
| SNOMED CT | 1085751000119100 | Abscess of intestine co-occurrent and due to Crohn's disease of large intestine |
| SNOMED CT | 1085801000119106 | Abscess of intestine co-occurrent and due to Crohn's disease of small and large intestine |
| SNOMED CT | 1092461000119104 | Sepsis without acute organ dysfunction |
| SNOMED CT | 1092471000119105 | Septic shock co-occurrent with acute organ dysfunction due to Chromobacterium |
| SNOMED CT | 1092481000119108 | Septic shock co-occurrent with acute organ dysfunction due to coagulase-negative Staphylococcus |
| SNOMED CT | 1092491000119106 | Septic shock co-occurrent with acute organ dysfunction due to Enterococcus |
| SNOMED CT | 1092501000119104 | Septic shock co-occurrent with acute organ dysfunction due to Gonococcus |
| SNOMED CT | 1092511000119101 | Septic shock co-occurrent with acute organ dysfunction due to Gram-positive coccus |
| SNOMED CT | 1092531000119106 | Septic shock co-occurrent with acute organ dysfunction due to Group A streptococcus |
| SNOMED CT | 1092541000119102 | Septic shock co-occurrent with acute organ dysfunction due to Group B streptococcus |
| SNOMED CT | 1092571000119109 | Sepsis caused by Chromobacterium |
| SNOMED CT | 1092591000119105 | Sepsis caused by Enterococcus |
| SNOMED CT | 1092601000119103 | Sepsis caused by Gonococcus |
| SNOMED CT | 1092841000119100 | Abscess of intestine co-occurrent and due to ulcerative colitis |
| SNOMED CT | 10628671000119100 | Allergic contact dermatitis caused by latex |
| SNOMED CT | 10628711000119101 | Allergic contact dermatitis caused by plant material |
| SNOMED CT | 10628751000119100 | Allergic contact dermatitis caused by pollen |

Continued on next page

**TableS31 – continued from previous page**

| CodeSystem | ConceptCode | ConceptName |
|---|---|---|
| SNOMED CT | 10629311000119107 | Psoriatic arthritis mutilans |
| SNOMED CT | 10629391000119103 | Allergic rhinitis caused by feathers |
| SNOMED CT | 10629431000119108 | Allergic rhinitis caused by insects |
| SNOMED CT | 10629471000119106 | Allergic rhinitis caused by mold |
| SNOMED CT | 10674711000119105 | Acute severe exacerbation of asthma co-occurrent with allergic rhinitis |
| SNOMED CT | 10674991000119104 | Intermittent allergic asthma |
| SNOMED CT | 10675431000119106 | Severe persistent allergic asthma |
| SNOMED CT | 10675471000119109 | Acute severe exacerbation of severe persistent allergic asthma |
| SNOMED CT | 10675551000119104 | Acute severe exacerbation of severe persistent asthma co-occurrent with allergic rhinitis |
| SNOMED CT | 10675631000119109 | Severe persistent asthma controlled co-occurrent with allergic rhinitis |
| SNOMED CT | 10675711000119106 | Severe persistent asthma uncontrolled co-occurrent with allergic rhinitis |
| SNOMED CT | 10675871000119106 | Mild persistent allergic asthma |
| SNOMED CT | 10675911000119109 | Acute severe exacerbation of mild persistent allergic asthma |
| SNOMED CT | 10675991000119100 | Acute severe exacerbation of mild persistent allergic asthma co-occurrent with allergic rhinitis |
| SNOMED CT | 10676111000119102 | Mild persistent asthma controlled co-occurrent with allergic rhinitis |
| SNOMED CT | 10676231000119102 | Mild persistent asthma uncontrolled co-occurrent with allergic rhinitis |
| SNOMED CT | 10676391000119108 | Moderate persistent allergic asthma |
| SNOMED CT | 10676431000119103 | Acute severe exacerbation of moderate persistent allergic asthma |
| SNOMED CT | 10676511000119109 | Acute severe exacerbation of moderate persistent asthma co-occurrent with allergic rhinitis |
| SNOMED CT | 10676591000119100 | Moderate persistent controlled asthma co-occurrent with allergic rhinitis |
| SNOMED CT | 10676671000119102 | Moderate persistent asthma uncontrolled co-occurrent with allergic rhinitis |
| SNOMED CT | 10676871000119103 | Allergic contact dermatitis caused by chemical |
| SNOMED CT | 10743231000119101 | Crohn's disease of small intestine with stenosis |
| SNOMED CT | 10743271000119103 | Immunoglobulin G4 related systemic disease |
| SNOMED CT | 10746341000119109 | Acquired immune deficiency syndrome complicating childbirth |
| SNOMED CT | 10751351000119104 | Sepsis due to ectopic pregnancy |
| SNOMED CT | 10812001000119100 | Sepsis due to incomplete miscarriage |
| SNOMED CT | 10838971000119103 | Immunodeficiency caused by long term therapeutic use of drug |
| SNOMED CT | 11055151000119108 | Seropositive rheumatoid arthritis of multiple joints |
| SNOMED CT | 13008751000119103 | Allergic proctocolitis caused by food protein |
| SNOMED CT | 15673361000119100 | Bilateral rheumatoid nodule of subcutaneous tissue of elbows |
| SNOMED CT | 15673521000119101 | Bilateral rheumatoid nodule of subcutaneous tissue of hands |
| SNOMED CT | 15679961000119104 | Vernal conjunctivitis of right eye |
| SNOMED CT | 15680001000119101 | Vernal conjunctivitis of left eye |
| SNOMED CT | 15680041000119104 | Vernal conjunctivitis of bilateral eyes |
| SNOMED CT | 15685921000119102 | Rheumatoid arthritis of bilateral temporomandibular joints |
| SNOMED CT | 15685961000119107 | Rheumatoid arthritis of right temporomandibular joint |

Continued on next page

**TableS31 – continued from previous page**

| CodeSystem | ConceptCode | ConceptName |
|---|---|---|
| SNOMED CT | 15686001000119104 | Rheumatoid arthritis of left temporomandibular joint |
| SNOMED CT | 15686281000119101 | Bilateral rheumatoid arthritis of feet |
| SNOMED CT | 15686321000119106 | Rheumatoid arthritis of bilateral elbows |
| SNOMED CT | 15687201000119107 | Bilateral rheumatoid arthritis of knees |
| SNOMED CT | 15687321000119109 | Bilateral rheumatoid arthritis of hands |
| SNOMED CT | 15687841000119108 | Rheumatoid arthritis of bilateral hips |
| SNOMED CT | 15691161000119108 | Seronegative rheumatoid arthritis of bilateral hands |
| SNOMED CT | 15691721000119102 | Rheumatoid arthritis of bilateral ankles |
| SNOMED CT | 15691761000119107 | Rheumatoid arthritis of bilateral wrists |
| SNOMED CT | 15691801000119104 | Seropositive rheumatoid arthritis of bilateral hands |
| SNOMED CT | 15911801000119104 | Allergic reaction caused by penicillin |
| SNOMED CT | 15919441000119101 | Allergic reaction caused by hymenoptera venom |
| SNOMED CT | 15919781000119104 | Allergic reaction caused by antibacterial agent |
| SNOMED CT | 15919821000119109 | Allergic reaction caused by sulfonamide |
| SNOMED CT | 15920041000119105 | Allergic reaction caused by nonsteroidal antiinflammatory agent |
| SNOMED CT | 15920121000119103 | Allergic reaction caused by vaccine product |
| SNOMED CT | 15920161000119108 | Allergic reaction caused by egg protein |
| SNOMED CT | 15920201000119103 | Allergic reaction caused by fish |
| SNOMED CT | 15920361000119106 | Allergic reaction caused by peanut |
| SNOMED CT | 15920401000119102 | Allergic reaction caused by seafood |
| SNOMED CT | 15920481000119105 | Allergic reaction caused by shellfish |
| SNOMED CT | 15920521000119105 | Allergic reaction caused by tree nut |
| SNOMED CT | 15920601000119100 | Allergic reaction caused by latex |
| SNOMED CT | 15928141000119107 | Human immunodeficiency virus infection with cognitive impairment |
| SNOMED CT | 15988671000119100 | Perennial allergic conjunctivitis of bilateral eyes |
| SNOMED CT | 16024431000119108 | Acute polyarticular juvenile rheumatoid arthritis |
| SNOMED CT | 16041671000119109 | Juvenile rheumatoid arthritis of bilateral knees |
| SNOMED CT | 16044751000119106 | Polyarticular juvenile idiopathic arthritis |
| SNOMED CT | 16058671000119103 | Transplant rejection of cornea of left eye |
| SNOMED CT | 16058711000119104 | Transplant failure of cornea of right eye |
| SNOMED CT | 16058831000119102 | Transplant rejection of cornea of right eye |
| SNOMED CT | 16058871000119104 | Transplant failure of cornea of left eye |
| SNOMED CT | 16060271000119109 | Neonatal sepsis caused by Staphylococcus |
| SNOMED CT | 16060311000119109 | Neonatal sepsis caused by Streptococcus |
| SNOMED CT | 16098491000119109 | Chronic autoimmune hepatitis |
| SNOMED CT | 16208561000119106 | Juvenile rheumatoid arthritis of left knee |
| SNOMED CT | 16208721000119109 | Juvenile rheumatoid arthritis of right knee |
| SNOMED CT | 16228891000119105 | Allergic contact dermatitis due to metal |
| SNOMED CT | 16318001000119107 | Hypogammaglobulinemia due to multiple myeloma |
| SNOMED CT | 16697871000119100 | Mast cell activation syndrome |
| SNOMED CT | 674814021000119106 | Acute respiratory distress syndrome due to COVID-19 |
| SNOMED CT | 479003 | Graves' disease with pretibial myxoedema AND with thyrotoxic crisis |
| SNOMED CT | 1089000 | Intrauterine foetal sepsis |
| SNOMED CT | 3815005 | Regional enteritis of rectum |
| SNOMED CT | 5603006 | Autoimmune hemolytic anemia caused by immunoglobulin G |

Continued on next page

**TableS31 – continued from previous page**

| CodeSystem | ConceptCode | ConceptName |
|---|---|---|
| SNOMED CT | 7620006 | Regional enteritis of the large bowel |
| SNOMED CT | 18700001 | Septicemia caused by Chromobacterium |
| SNOMED CT | 19719003 | Jessner's lymphocytic infiltrate |
| SNOMED CT | 21719001 | Hayfever |
| SNOMED CT | 27520001 | Pustular psoriasis of the palms AND/OR soles |
| SNOMED CT | 27538003 | Hashitoxicosis |
| SNOMED CT | 31848007 | CREST - Calcinosis, Raynaud's phenomenon, esophageal dysfunction, sclerodactyly, telangiectasia |
| SNOMED CT | 33491002 | Autoimmune hemolytic anemia caused by IgA plus complement |
| SNOMED CT | 34000006 | Crohns disease |
| SNOMED CT | 38106008 | Crohn disease of ileum |
| SNOMED CT | 40175007 | Allergic dermatitis caused by bite of Ctenocephalides felis |
| SNOMED CT | 40956001 | Guillain Barre syndrome |
| SNOMED CT | 41936006 | Septicemia caused by Erysipelothrix insidiosa |
| SNOMED CT | 48347002 | Air-conditioner and humidifier lung |
| SNOMED CT | 50440006 | Regional colitis |
| SNOMED CT | 50581000 | Pulmonary-renal syndrome |
| SNOMED CT | 51696001 | Subcutaneous lipogranulomatosis |
| SNOMED CT | 52983002 | Kerion caused by Microsporum |
| SNOMED CT | 56287005 | Regional enteritis of the duodenum |
| SNOMED CT | 56689002 | Granulomatous enteritis |
| SNOMED CT | 56989000 | Myasthenic syndrome |
| SNOMED CT | 61424003 | Crohn disease of pylorus |
| SNOMED CT | 62382002 | Calcinosis, Raynaud's phenomenon, sclerodactyly, and telangiectasia syndrome |
| SNOMED CT | 62609001 | Autoimmune hemolytic anemia caused by IgG plus complement |
| SNOMED CT | 69339004 | Pigeon fancier's lung |
| SNOMED CT | 70622003 | Crohn disease of pyloric antrum |
| SNOMED CT | 71832003 | Autoimmune hemolytic anemia caused by immunoglobulin M |
| SNOMED CT | 71833008 | Crohn's disease of small and large intestines |
| SNOMED CT | 73860009 | Kerion caused by Trichophyton |
| SNOMED CT | 76715008 | Autoimmune adrenal atrophy |
| SNOMED CT | 78946008 | Keratoconjunctivitis sicca, in Sjögren syndrome |
| SNOMED CT | 79337003 | Blood product-induced anaphylaxis |
| SNOMED CT | 79587009 | Septicemia caused by Bacteroides |
| SNOMED CT | 83901003 | Sicca syndrome |
| SNOMED CT | 85559002 | Pelger Huet anomaly |
| SNOMED CT | 87442008 | Pericarditis co-occurrent and due to scleroderma |
| SNOMED CT | 91178003 | Allergic dermatitis caused by bite of Ctenocephalides canis |
| SNOMED CT | 91390005 | Crohn disease of jejunum |
| SNOMED CT | 91411007 | Autoimmune hemolytic anemia caused by IgA |
| SNOMED CT | 95330001 | Linear immunoglobulin A dermatosis |
| SNOMED CT | 123583002 | Tuberculids |
| SNOMED CT | 126766000 | Lymphoepithelial sialadenitis of Sjögren syndrome |
| SNOMED CT | 127054006 | Cold agglutinin disease caused by Epstein-Barr virus infection |
| SNOMED CT | 129000002 | Langerhans cell histiocytosis, unifocal |

Continued on next page

**TableS31 – continued from previous page**

| CodeSystem | ConceptCode | ConceptName |
|---|---|---|
| SNOMED CT | 129645002 | Adult glucose-6-phosphate dehydrogenase deficiency of leukocytes syndrome |
| SNOMED CT | 193497004 | Vogt-Koyanagi syndrome |
| SNOMED CT | 196137000 | Lung disease with Sjögren disease |
| SNOMED CT | 196977009 | Crohn disease of terminal ileum |
| SNOMED CT | 206378006 | Sepsis of newborn caused by Staphylococcus aureus |
| SNOMED CT | 206379003 | Sepsis of newborn caused by Escherichia coli |
| SNOMED CT | 234558001 | Anti-hemophilus influenzae B polysaccharide antibody deficiency |
| SNOMED CT | 234619000 | Hereditary angioneurotic edema - type 1 |
| SNOMED CT | 234620006 | Hereditary angioneurotic edema - type 2 |
| SNOMED CT | 235664007 | Crohn disease of stomach |
| SNOMED CT | 235796008 | Perianal Crohn disease |
| SNOMED CT | 237520009 | Hypothyroidism due to Hashimoto thyroiditis |
| SNOMED CT | 237521008 | Hypothyroidism due to thyroid stimulating hormone receptor blocking antibody |
| SNOMED CT | 238612002 | Generalized pustular psoriasis of von Zumbusch |
| SNOMED CT | 238937005 | Pemphigus vegetans (Neumann type) |
| SNOMED CT | 239912009 | Primary Sjögren syndrome |
| SNOMED CT | 239915006 | Secondary Sjögren syndrome |
| SNOMED CT | 285423008 | Immunoglobulin A monoclonal gammopathy of uncertain significance |
| SNOMED CT | 285424002 | Immunoglobulin G monoclonal gammopathy of uncertain significance |
| SNOMED CT | 285426000 | Immunoglobulin M monoclonal gammopathy of uncertain significance |
| SNOMED CT | 285428004 | Immunoglobulin D monoclonal gammopathy of uncertain significance |
| SNOMED CT | 353295004 | Exophthalmic goiter |
| SNOMED CT | 397172008 | Gastrointestinal Crohn disease |
| SNOMED CT | 402378006 | Metastatic Crohn disease of skin |
| SNOMED CT | 402446002 | Oral mucosal involvement by linear immunoglobulin A disease |
| SNOMED CT | 403158001 | Allergic contact dermatitis caused by Bryozoa |
| SNOMED CT | 403633007 | Drug-induced linear immunoglobulin A disease |
| SNOMED CT | 410485009 | Iritis with Crohn disease |
| SNOMED CT | 414153008 | Exacerbation of Crohn disease of large intestine |
| SNOMED CT | 414154002 | Exacerbation of Crohn disease of small intestine |
| SNOMED CT | 421310001 | Non-immunoglobulin E-mediated atopic disorder |
| SNOMED CT | 421871004 | Atopic allergy |
| SNOMED CT | 422076005 | Immediate hypersensitivity disorder |
| SNOMED CT | 422169003 | Nonatopic immunoglobulin E-mediated allergic disorder |
| SNOMED CT | 422339003 | Non-immunoglobulin E-mediated allergic disorder |
| SNOMED CT | 423889005 | Non-immunoglobulin E mediated allergic asthma |
| SNOMED CT | 447684006 | Erysipelothrix sepsis |
| SNOMED CT | 447685007 | Severe sepsis with acute organ dysfunction caused by Haemophilus influenzae |
| SNOMED CT | 447841007 | Sepsis caused by Candida |

Continued on next page

**TableS31 – continued from previous page**

| CodeSystem | ConceptCode | ConceptName |
|---|---|---|
| SNOMED CT | 447894003 | Severe sepsis with acute organ dysfunction caused by Staphylococcus |
| SNOMED CT | 447897005 | Sepsis caused by Acinetobacter |
| SNOMED CT | 447898000 | Sepsis caused by Actinomyces |
| SNOMED CT | 447899008 | Sepsis caused by Escherichia coli |
| SNOMED CT | 448417001 | Sepsis caused by Staphylococcus aureus |
| SNOMED CT | 448418006 | Severe sepsis with acute organ dysfunction caused by Streptococcus |
| SNOMED CT | 448419003 | Severe sepsis with acute organ dysfunction due to Group B streptococcus |
| SNOMED CT | 448420009 | Sepsis caused by Streptococcus group D |
| SNOMED CT | 448421008 | Severe sepsis with acute organ dysfunction caused by Pneumococcus |
| SNOMED CT | 448422001 | Sepsis caused by Streptococcus suis |
| SNOMED CT | 448482003 | Sepsis caused by Bacillus anthracis |
| SNOMED CT | 448483008 | Sepsis caused by Enterobacter |
| SNOMED CT | 448812000 | Severe sepsis with acute organ dysfunction caused by methicillin resistant Staphylococcus aureus |
| SNOMED CT | 448813005 | Severe sepsis with acute organ dysfunction caused by Pseudomonas |
| SNOMED CT | 449082003 | Severe sepsis with acute organ dysfunction caused by Gram-negative bacteria |
| SNOMED CT | 449083008 | Severe sepsis with acute organ dysfunction caused by Salmonella |
| SNOMED CT | 449084002 | Severe sepsis with acute organ dysfunction caused by Serratia |
| SNOMED CT | 449335002 | Listerial sepsis |
| SNOMED CT | 449504009 | Severe sepsis with acute organ dysfunction due to Group A streptococcus |
| SNOMED CT | 449505005 | Severe sepsis with acute organ dysfunction due to coagulase negative Staphylococcus |
| SNOMED CT | 449546000 | Sepsis caused by Haemophilus influenzae type B |
| SNOMED CT | 699861000 | Blau syndrome |
| SNOMED CT | 703903005 | Dermatophagoides farinae allergic rhinitis |
| SNOMED CT | 703937002 | Dermatophagoides pteronyssinus allergic rhinitis |
| SNOMED CT | 703951002 | Allergic conjunctivitis caused by Dermatophagoides farinae |
| SNOMED CT | 703952009 | Allergic conjunctivitis caused by Dermatophagoides pteronyssinus |
| SNOMED CT | 703953004 | Asthma caused by house dust mite, Dermatophagoides pteronyssinus |
| SNOMED CT | 703954005 | Asthma caused by house dust mite, Dermatophagoides farinae |
| SNOMED CT | 762385007 | Allergic contact dermatitis caused by allergenic hapten derived from plants or organic matter |
| SNOMED CT | 72621000119104 | Human immunodeficiency virus |
| SNOMED CT | 127091000119100 | Sepsis of newborn caused by Streptococcus agalactiae |
| SNOMED CT | 127181000119102 | Septic shock co-occurrent with acute organ dysfunction caused by Meningococcus |
| SNOMED CT | 127191000119104 | Septic shock co-occurrent with acute organ dysfunction caused by methicillin susceptible Staphylococcus aureus |

Continued on next page

**TableS31 – continued from previous page**

| CodeSystem | ConceptCode | ConceptName |
|---|---|---|
| SNOMED CT | 127201000119101 | Septic shock co-occurrent with acute organ dysfunction caused by Pseudomonas |
| SNOMED CT | 127231000119108 | Septic shock co-occurrent with acute organ dysfunction caused by Serratia |
| SNOMED CT | 127251000119102 | Septic shock co-occurrent with acute organ dysfunction caused by Streptococcus |
| SNOMED CT | 127261000119100 | Septic shock co-occurrent with acute organ dysfunction caused by Pneumococcus |
| SNOMED CT | 127311000119106 | Sepsis caused by Meningococcus |
| SNOMED CT | 127331000119101 | Sepsis caused by methicillin susceptible Staphylococcus aureus |
| SNOMED CT | 128381000119104 | Septic shock co-occurrent with acute organ dysfunction caused by methicillin resistant Staphylococcus aureus |
| SNOMED CT | 342791000119106 | Contact dermatitis, left upper eyelid |
| SNOMED CT | 428331000124103 | Anaphylaxis caused by rotavirus vaccine |
| SNOMED CT | 1073701000119107 | Rheumatoid arthritis of left foot |
| SNOMED CT | 1073781000119104 | Rheumatoid arthritis of right foot |
| SNOMED CT | 1085851000119105 | Abscess of small intestine due to Crohn's disease |
| SNOMED CT | 1085901000119101 | Abscess of intestine due to Crohn's disease |
| SNOMED CT | 1003756003 | Hevea brasiliensis latex protein-induced angioedema-urticaria |
| SNOMED CT | 1003757007 | Allergic reaction caused by Hevea brasiliensis latex protein |
| SNOMED CT | 1003758002 | Anaphylaxis due to Hevea brasiliensis latex protein |
| SNOMED CT | 1052325005 | Psoriasis exacerbated by human immunodeficiency virus infection |
| SNOMED CT | 1078231000 | Allergic contact dermatitis due to jewelry |
| SNOMED CT | 1082871000119108 | Bilateral acute allergic otitis media of middle ears |
| SNOMED CT | 1083261000119107 | Bilateral chronic allergic otitis media of middle ears |
| SNOMED CT | 1087981000119103 | Acute allergic otitis media of left middle ear |
| SNOMED CT | 1088321000119102 | Autoimmune disorder of left inner ear |
| SNOMED CT | 1090601000119102 | Acute allergic otitis media of right middle ear |
| SNOMED CT | 1090931000119104 | Autoimmune disorder of right inner ear |
| SNOMED CT | 15691241000119101 | Bilateral seronegative rheumatoid arthritis of knees |
| SNOMED CT | 15691321000119101 | Bilateral seronegative rheumatoid arthritis of wrists |
| SNOMED CT | 15691881000119107 | Bilateral seropositive rheumatoid arthritis of knees |
| SNOMED CT | 16050071000119108 | Bilateral seropositive rheumatoid arthritis of feet |
| SNOMED CT | 16318061000119108 | Hypogammaglobulinemia due to monoclonal gammopathy of undetermined significance |
| SNOMED CT | 16606721000119107 | Seropositive rheumatoid arthritis in remission |
| SNOMED CT | 16894671000119102 | Immunoglobulin G deficiency due to monoclonal gammopathy of undetermined significance |
| SNOMED CT | 16894711000119103 | Immunoglobulin G deficiency due to multiple myeloma |
| SNOMED CT | 453720571000119100 | Acute ulcerative rectosigmoiditis |
| SNOMED CT | 63541000119109 | Corneal epithelial defect following penetrating keratoplasty |
| SNOMED CT | 890143000 | Allergic contact dermatitis caused by poison primrose |
| SNOMED CT | 890154005 | Allergic contact dermatitis caused by urushiol from Eastern poison ivy |
| SNOMED CT | 890169006 | Allergic contact dermatitis caused by urushiol from poison sumac |
| SNOMED CT | 890170007 | Allergic contact dermatitis caused by ragweed |

Continued on next page

**TableS31 – continued from previous page**

| CodeSystem | ConceptCode | ConceptName |
|---|---|---|
| SNOMED CT | 890283004 | Allergic contact dermatitis caused by oil |
| SNOMED CT | 890284005 | Allergic contact dermatitis caused by grease |
| SNOMED CT | 890331009 | Allergic contact dermatitis caused by soap |
| SNOMED CT | 966011731000119103 | Acute ulcerative pancolitis |
| SNOMED CT | 969688801000119108 | Acute left-sided ulcerative colitis |
| SNOMED CT | 1073601000119101 | Right foot seropositive rheumatoid arthritis |
| SNOMED CT | 1073611000119103 | Left foot seropositive rheumatoid arthritis |
| SNOMED CT | 1142045004 | Primary human immunodeficiency virus infection |
| SNOMED CT | 1142055000 | HIV modified skin disease |
| SNOMED CT | 1142207008 | Transient neonatal epidermolysis bullosa acquisita |
| SNOMED CT | 1144355009 | Allergic contact stomatitis caused by denture |
| SNOMED CT | 1144805008 | Microscopic polyangiitis |
| SNOMED CT | 1144907001 | Late acute graft-versus-host disease |
| SNOMED CT | 1144908006 | Late chronic graft-versus-host disease |
| SNOMED CT | 1144923001 | Disorder of immune function due to systemic lupus erythematosus |
| SNOMED CT | 1144925008 | Diffuse systemic sclerosis of childhood |
| SNOMED CT | 1144929002 | Secondary agranulocytosis |
| SNOMED CT | 1144941009 | Chronic tubulointerstitial nephritis following renal transplantation |
| SNOMED CT | 1144963000 | Acute on chronic graft-versus-host disease |
| SNOMED CT | 1144974009 | Antisynthetase syndrome due to polymyositis |
| SNOMED CT | 1144980001 | Food-dependent exercise-induced anaphylaxis |
| SNOMED CT | 1148594002 | Chronic rheumatic arthritis |
| SNOMED CT | 1148595001 | Subacute rheumatic arthritis |
| SNOMED CT | 1148597009 | Rheumatic arthritis |
| SNOMED CT | 1148598004 | Rheumatic arthritis of temporomandibular joint |
| SNOMED CT | 1148696006 | Leukopenia caused by drug |
| SNOMED CT | 1148763004 | Acute rheumatic carditis |
| SNOMED CT | 1148765006 | Chronic rheumatic carditis |
| SNOMED CT | 1148845007 | Follicular lymphoma grade 3b |
| SNOMED CT | 1148847004 | Follicular non-Hodgkin's lymphoma in situ |
| SNOMED CT | 1148851002 | Follicular lymphoma grade 3a |
| SNOMED CT | 1148890003 | Hemophagocytic lymphohistiocytosis due to malignant disease |
| SNOMED CT | 1148891004 | Bullous diffuse cutaneous mastocytosis |
| SNOMED CT | 1148906001 | Acute myeloid leukemia with t(8;21)(q22;q22) RUNX1-RUNX1T1 |
| SNOMED CT | 1149066008 | Chronic rheumatic pancarditis |
| SNOMED CT | 1149218009 | Isolated nailfold rheumatoid vasculitis |
| SNOMED CT | 1149219001 | Systemic rheumatoid vasculitis |
| SNOMED CT | 1149481006 | Sepsis caused by Coliform |
| SNOMED CT | 1149482004 | Sepsis caused by Clostridium |
| SNOMED CT | 1149486001 | Sepsis caused by Proteus |
| SNOMED CT | 1149494008 | Sepsis caused by Erysipelothrix rhusiopathiae |
| SNOMED CT | 1153336009 | Pediatric nodal marginal zone B cell lymphoma |
| SNOMED CT | 1153341001 | Myeloproliferative hypereosinophilic syndrome |
| SNOMED CT | 1153344009 | Pseudoxanthomatous nodular cutaneous mastocytosis |

Continued on next page

**TableS31 – continued from previous page**

| CodeSystem | ConceptCode | ConceptName |
|---|---|---|
| SNOMED CT | 1153345005 | Myelodysplastic syndrome with single lineage dysplasia |
| SNOMED CT | 1153346006 | Primary non-Hodgkin malignant lymphoma of uveal tract |
| SNOMED CT | 1153347002 | Primary non-Hodgkin malignant lymphoma of vitreoretinal tract |
| SNOMED CT | 1153348007 | Primary effusion lymphoma due to human immune deficiency virus infection |
| SNOMED CT | 1153349004 | Primary familial polycythemia due to erythropoietin receptor mutation |
| SNOMED CT | 1153354008 | Malignant lymphoma of duodenum |
| SNOMED CT | 1153355009 | Malignant lymphoma of esophagus |
| SNOMED CT | 1153356005 | Lymphoma of anus |
| SNOMED CT | 1153357001 | Lymphoma of appendix |
| SNOMED CT | 1153358006 | Lymphocyte passenger syndrome following organ transplantation |
| SNOMED CT | 1153374001 | Myelofibrosis caused by drug |
| SNOMED CT | 1153377008 | Mixed phenotype acute leukemia with t(9;22) (q34;q11.2); BCR-ABL1 |
| SNOMED CT | 1153379006 | Mixed phenotype acute leukemia with T-cell and myeloid lineage |
| SNOMED CT | 1153381008 | Mixed phenotype acute leukemia with myeloid and B-cell lymphoid phenotypes |
| SNOMED CT | 1153382001 | Lymphoma of lacrimal gland |
| SNOMED CT | 1153383006 | Lymphoma of liver |
| SNOMED CT | 1153397003 | Hydroa vacciniforme-like cutaneous T-cell lymphoma |
| SNOMED CT | 1153417000 | Secondary hemophagocytic lymphohistiocytosis |
| SNOMED CT | 1153534006 | Sepsis caused by vancomycin resistant Enterococcus |
| SNOMED CT | 1155919006 | Acute rheumatic aortitis |
| SNOMED CT | 1155943009 | Allergic contact dermatitis caused by metal and/or metal compound |
| SNOMED CT | 1155956002 | Follicular lymphoma grade 3a |
| SNOMED CT | 1155957006 | Follicular lymphoma grade 3b |
| SNOMED CT | 1156197003 | Complication of embryo transfer |
| SNOMED CT | 1156250009 | Hypereosinophilic syndrome |
| SNOMED CT | 1156296001 | Acquired neutropenia |
| SNOMED CT | 1156300000 | Benign ethnic neutropenia |
| SNOMED CT | 1156403002 | Composite Hodgkin and non-Hodgkin lymphoma |
| SNOMED CT | 1156495004 | Dendritic cell sarcoma |
| SNOMED CT | 1156631003 | Dendritic cell sarcoma |
| SNOMED CT | 1156746003 | Vaccine-induced prothrombotic immune thrombocytopenia |
| SNOMED CT | 1156765006 | Persistent onset antinuclear antibody negative juvenile idiopathic oligoarthritis |
| SNOMED CT | 1156766007 | Persistent onset antinuclear antibody positive juvenile idiopathic oligoarthritis |
| SNOMED CT | 1156774008 | Rheumatoid factor positive and anti-citrullinated protein antibody positive juvenile polyarthritis |
| SNOMED CT | 1156777001 | Rheumatoid factor positive and anti-citrullinated protein antibody negative juvenile polyarthritis |
| SNOMED CT | 1156778006 | Rheumatoid factor negative and anti-citrullinated protein antibody negative juvenile polyarthritis |

Continued on next page

**TableS31 – continued from previous page**

| CodeSystem | ConceptCode | ConceptName |
|---|---|---|
| SNOMED CT | 1156779003 | Rheumatoid factor negative and anti-citrullinated protein antibody positive juvenile polyarthritis |
| SNOMED CT | 1156801007 | Macrophage activation syndrome due to juvenile systemic onset arthritis |
| SNOMED CT | 1156817001 | Extended oligoarticular onset antinuclear antibody positive juvenile idiopathic arthritis |
| SNOMED CT | 1156819003 | Extended oligoarticular onset antinuclear antibody negative juvenile idiopathic arthritis |
| SNOMED CT | 1157157006 | Acute myeloid leukemia with 11q23 abnormality |
| SNOMED CT | 1157158001 | Acute myeloid leukemia with t(9;11)(p22;q23); MLLT3-MLL |
| SNOMED CT | 1157162007 | Intravascular large B-cell lymphoma |
| SNOMED CT | 15691081000119100 | Bilateral seronegative rheumatoid arthritis of feet |
| SNOMED CT | 15691121000119103 | Bilateral seronegative rheumatoid arthritis of elbow joints |
| SNOMED CT | 15691841000119102 | Bilateral seropositive rheumatoid arthritis of shoulder regions |
| SNOMED CT | 15691921000119100 | Bilateral seropositive rheumatoid arthritis of hip joints |
| SNOMED CT | 15691961000119105 | Bilateral seropositive rheumatoid arthritis of ankle joints |
| SNOMED CT | 15692001000119101 | Bilateral seropositive rheumatoid arthritis of joint of elbows |
| SNOMED CT | 15692041000119104 | Bilateral seropositive rheumatoid arthritis of wrist joint regions |
| SNOMED CT | 16606681000119101 | Seronegative rheumatoid arthritis in remission |
| SNOMED CT | 16697831000119103 | Idiopathic mast cell activation syndrome |
| SNOMED CT | 16775311000119107 | Mycosis fungoides of facial lymph node |
| SNOMED CT | 318841000119100 | Left ankle seropositive rheumatoid arthritis |
| SNOMED CT | 318851000119103 | Left elbow seropositive rheumatoid arthritis |
| SNOMED CT | 318891000119108 | Seropositive rheumatoid arthritis of joint of left shoulder region |
| SNOMED CT | 318901000119107 | Left wrist region seropositive rheumatoid arthritis |
| SNOMED CT | 318911000119105 | Right ankle seropositive rheumatoid arthritis |
| SNOMED CT | 318921000119103 | Right elbow seropositive rheumatoid arthritis |
| SNOMED CT | 318961000119108 | Seropositive rheumatoid arthritis of joint of right shoulder region |
| SNOMED CT | 318971000119102 | Right wrist region seropositive rheumatoid arthritis |
| SNOMED CT | 319011000119103 | Seronegative rheumatoid arthritis of joint of left ankle |
| SNOMED CT | 319041000119104 | Left hip joint seronegative rheumatoid arthritis |
| SNOMED CT | 319051000119102 | Seronegative rheumatoid arthritis of left knee joint |
| SNOMED CT | 319061000119100 | Left shoulder region seronegative rheumatoid arthritis |
| SNOMED CT | 319071000119106 | Left wrist region seronegative rheumatoid arthritis |
| SNOMED CT | 319091000119107 | Seronegative rheumatoid arthritis of joint of right ankle |
| SNOMED CT | 319121000119106 | Right hip joint seronegative rheumatoid arthritis |
| SNOMED CT | 319131000119109 | Seronegative rheumatoid arthritis of right knee joint |
| SNOMED CT | 319141000119100 | Right shoulder region seronegative rheumatoid arthritis |
| SNOMED CT | 319151000119103 | Right wrist region seronegative rheumatoid arthritis |
| SNOMED CT | 328371000119107 | Pancytopenia caused by antithyroid drug |
| SNOMED CT | 328381000119105 | Pancytopenia caused by anticonvulsant |
| SNOMED CT | 350691000119103 | Agranulocytosis due to and following administration of antineoplastic agent |
| SNOMED CT | 351251000119103 | Acute myelofibrosis in remission |
| SNOMED CT | 471141000124102 | Anaphylaxis caused by vaccine product containing Streptococcus pneumoniae antigen |

Continued on next page

**TableS31 – continued from previous page**

| CodeSystem | ConceptCode | ConceptName |
|---|---|---|
| SNOMED CT | 471311000124103 | Anaphylaxis caused by vaccine product containing Hepatitis A virus antigen |
| SNOMED CT | 471321000124106 | Anaphylaxis caused by vaccine product containing human poliovirus antigen |
| SNOMED CT | 471331000124109 | Anaphylaxis caused by vaccine product containing Measles morbillivirus and Mumps orthorubulavirus and Rubella virus antigens |
| SNOMED CT | 471341000124104 | Anaphylaxis caused by vaccine containing Human alphaherpesvirus 3 antigen |
| SNOMED CT | 471351000124102 | Anaphylaxis caused by vaccine product |
| SNOMED CT | 471361000124100 | Anaphylaxis caused by vaccine product containing Influenza virus antigen |
| SNOMED CT | 471371000124107 | Anaphylaxis caused by vaccine product containing human alphaherpesvirus 3 recombinant surface glycoprotein E antigen |
| SNOMED CT | 471381000124105 | Anaphylaxis caused by vaccine product containing live attenuated human alphaherpesvirus 3 antigen |
| SNOMED CT | 686981000119108 | Mycosis fungoides of inguinal lymph node |
| SNOMED CT | 686991000119106 | Mycosis fungoides of lower limb lymph node |
| SNOMED CT | 84821000119100 | Primary follicular dendritic cell sarcoma of spleen |
| SNOMED CT | 987840791000119102 | Adenosine deaminase 2 deficiency |
| Truveta | 1215935 | COVID-19 in immunocompromised patient |
| HCPCS | G0267 | BONE MARROW OR PERIPHERAL STEM CELL HARVEST ELIMINATE CELL TYPE(S) (E.G. T-CELLS |
| HCPCS | S2140 | Cord blood harvesting for transplantation, allogeneic |
| HCPCS | S2142 | Cord blood-derived stem-cell transplantation, allogeneic |
| SNOMED CT | 1163005009 | Hodgkin lymphoma |
| SNOMED CT | 1162677006 | Rheumatoid arthritis with rheumatoid lung disease |
| SNOMED CT | 1217476001 | Anaphylactic reaction due to component of vaccine product |
| SNOMED CT | 139841000119108 | Anaphylaxis caused by allergy skin test |
| SNOMED CT | 15919571000119107 | Anaphylaxis caused by antibacterial agent |
| SNOMED CT | 1197476009 | Infantile-onset pulmonary alveolar proteinosis, hypogammaglobulinemia |
| SNOMED CT | 1197594000 | Periodic fever, infantile enterocolitis, autoinflammatory syndrome |
| SNOMED CT | 1197344000 | T-cell lymphoma of small intestine |
| SNOMED CT | 1204420006 | Chronic nonbacterial osteomyelitis/chronic recurrent multifocal osteomyelitis |
| SNOMED CT | 1217669004 | Chronic papillary vernal conjunctivitis |
| SNOMED CT | 1197362009 | STAT3-related early-onset multisystem autoimmune disease |
| SNOMED CT | 353211000119103 | Extramedullary plasmacytoma in remission |
| SNOMED CT | 16058791000119108 | Bilateral corneal transplant rejection |
| SNOMED CT | 1228843008 | Anti-p200 pemphigoid |
| SNOMED CT | 1162336001 | Anti-citrullinated protein antibody positive erosive rheumatoid arthritis |
| SNOMED CT | 1177173001 | Congenital progressive bone marrow failure, B-cell immunodeficiency, skeletal dysplasia syndrome |
| SNOMED CT | 1186855002 | Autoimmune hypopituitarism |

Continued on next page

**TableS31 – continued from previous page**

| CodeSystem | ConceptCode | ConceptName |
|---|---|---|
| SNOMED CT | 1186859008 | Autoimmune male infertility |
| SNOMED CT | 1162928000 | Acute myeloid leukemia |
| SNOMED CT | 1162531007 | Acute myeloid leukemia in complete remission |
| SNOMED CT | 1172892009 | Autosomal recessive mendelian susceptibility to mycobacterial disease due to complete RORgamma receptor mutation |
| SNOMED CT | 1179288008 | Combined immunodeficiency due to TFRC deficiency |
| SNOMED CT | 1162660006 | B lymphoblastic leukemia lymphoma |
| SNOMED CT | 1186720006 | CEBPE-associated autoinflammation, immunodeficiency, neutrophil dysfunction syndrome |
| SNOMED CT | 1179300002 | BENTA disease |
| SNOMED CT | 1172730009 | Breast implant-associated anaplastic large cell lymphoma |
| SNOMED CT | 1177120001 | Bronchiolitis obliterans syndrome due to and following allogeneic stem cell transplant |
| SNOMED CT | 1186712009 | Combined immunodeficiency due to CARMIL2 deficiency |
| SNOMED CT | 1186715006 | Combined immunodeficiency due to CD70 deficiency |
| SNOMED CT | 1179286007 | Combined immunodeficiency due to GINS1 deficiency |
| SNOMED CT | 1186714005 | Combined immunodeficiency due to ITK deficiency |
| SNOMED CT | 1179285006 | Combined immunodeficiency due to moesin deficiency |
| SNOMED CT | 1197732001 | Colorectal Crohn disease |
| SNOMED CT | 1197205005 | Combined immunodeficiency due to DOCK8 deficiency |
| SNOMED CT | 1197477000 | Combined immunodeficiency due to LRBA deficiency |
| SNOMED CT | 1197428008 | Combined immunodeficiency, enteropathy spectrum |
| SNOMED CT | 515623061000119105 | Crohn disease of colon in remission |
| SNOMED CT | 527788091000119106 | Crohn disease of small intestine and large intestine in remission |
| SNOMED CT | 253855721000119107 | Crohn disease of small intestine in remission |
| SNOMED CT | 1197727002 | Crohn's disease of parastomal skin |
| SNOMED CT | 1230290005 | Cytokine release syndrome due to chimeric antigen receptor T-cell immunotherapy |
| SNOMED CT | 1231239003 | Dacryoadenitis due to Graves' disease |
| SNOMED CT | 1197479002 | DOCK2 deficiency |
| SNOMED CT | 1204463009 | Encephalopathy with sepsis |
| SNOMED CT | 1222550009 | Enteropathy-associated T-cell lymphoma |
| SNOMED CT | 1220590003 | Familial chilblain lupus erythematosus |
| SNOMED CT | 1204466001 | Gastritis with sepsis |
| SNOMED CT | 1204464003 | Gastrointestinal hemorrhage with sepsis |
| SNOMED CT | 1204461006 | Gastrointestinal lesion with sepsis |
| SNOMED CT | 1204467005 | Gastrointestinal ulceration due to sepsis |
| SNOMED CT | 1230342001 | Hepatitis B reinfection following liver transplantation |
| SNOMED CT | 1197733006 | Ileojejunal Crohn disease |
| SNOMED CT | 1230414002 | Immune effector cell-associated neurotoxicity syndrome |
| SNOMED CT | 145781000119106 | Infection of transplanted kidney |
| SNOMED CT | 1204462004 | Left ventricular failure with sepsis |
| SNOMED CT | 1204459002 | Lung injury with sepsis |
| SNOMED CT | 15988711000119101 | Perennial allergic conjunctivitis of left eye |
| SNOMED CT | 15988751000119100 | Perennial allergic conjunctivitis of right eye |
| SNOMED CT | 1230291009 | Primary biliary cholangitis and/or primary sclerosing cholangitis and autoimmune hepatitis overlap syndrome |

Continued on next page

**TableS31 – continued from previous page**

| CodeSystem | ConceptCode | ConceptName |
|---|---|---|
| SNOMED CT | 1228842003 | Primary cutaneous plasmacytosis |
| SNOMED CT | 1197478005 | Primary immunodeficiency with multifaceted aberrant lymphoid immunity |
| SNOMED CT | 1197150002 | Recurrent hepatitis C virus induced liver disease following liver transplant |
| SNOMED CT | 1204468000 | Right ventricular failure with sepsis |
| SNOMED CT | 1201760007 | Sepsis caused by Bacteroides |
| SNOMED CT | 1290841000000104 | Sepsis caused by monkeypox virus |
| SNOMED CT | 1204037006 | Sepsis following induced termination of pregnancy |
| SNOMED CT | 1230323000 | Sepsis of premature infant |
| SNOMED CT | 1229942009 | Severe combined immunodeficiency due to CORO1A deficiency |
| SNOMED CT | 1197415001 | Susceptibility to infection due to TYK2 deficiency |
| SNOMED CT | 1197482007 | Susceptibility to localized juvenile periodontitis |
| SNOMED CT | 1197701002 | Type 1 autoimmune pancreatitis with extrapancreatic involvement |
| SNOMED CT | 1197704005 | Autoantibody negative autoimmune hepatitis |
| SNOMED CT | 1208833001 | Autoimmune generalized polymyoclonus |
| SNOMED CT | 1222681008 | Autoimmune hemolytic anemia, autoimmune thrombocytopenia, primary immunodeficiency syndrome |
| SNOMED CT | 1208830003 | Autoimmune interstitial keratitis |
| SNOMED CT | 1222679006 | Autoimmune interstitial lung disease, arthritis syndrome |
| SNOMED CT | 1197361002 | Autoimmune lymphoproliferative syndrome due to CTLA4 haploinsufficiency |
| SNOMED CT | 1197740007 | Autoimmune pancreatitis type 2 |
| SNOMED CT | 1208829008 | Autoimmune retinitis |
| SNOMED CT | 1197366007 | Autosomal recessive primary immunodeficiency with defective spontaneous natural killer cell cytotoxicity |
| SNOMED CT | 1229941002 | Autosomal recessive T-cell negative, B-cell positive severe combined immunodeficiency due to CD45 deficiency |
| SNOMED CT | 1229940001 | Autosomal recessive T-cell negative, B-cell positive severe combined immunodeficiency due to IL-7Ralpha deficiency |
| SNOMED CT | 1196900003 | B cell lymphoma of small intestine |
| SNOMED CT | 1230295000 | B-cell immunodeficiency, limb anomaly, urogenital malformation syndrome |
| SNOMED CT | 1172709000 | Lymphoid leukemia |
| SNOMED CT | 1162263002 | Complement component 8 deficiency |
| SNOMED CT | 1163528001 | Erythema multiforme due to bacterial infection |
| SNOMED CT | 1187129009 | Primary cutaneous B-cell lymphoma |
| SNOMED CT | 1187190002 | Primary cutaneous diffuse large B-cell lymphoma leg type |
| SNOMED CT | 1187135009 | Primary cutaneous large T-cell lymphoma |
| SNOMED CT | 1187136005 | Primary cutaneous T-cell lymphoma |
| SNOMED CT | 1163590002 | Myelomonocytic leukemia |
| SNOMED CT | 1163439000 | Myeloid leukemia |
| SNOMED CT | 16058751000119103 | Bilateral transplant failure of corneas |
| SNOMED CT | 1162588009 | Chronic myeloid leukemia |
| SNOMED CT | 1163404000 | Peripheral T-cell lymphoma |
| SNOMED CT | 1173999006 | IL21-related infantile inflammatory bowel disease |

Continued on next page

**TableS31 – continued from previous page**

| CodeSystem | ConceptCode | ConceptName |
|---|---|---|
| SNOMED CT | 1162973007 | Cutaneous T-cell lymphoma |
| SNOMED CT | 1162993004 | Acute eosinophilic leukemia |
| SNOMED CT | 1186652002 | Inflammatory bowel disease, recurrent sinopulmonary infection syndrome |
| SNOMED CT | 1187123005 | Mixed phenotype acute leukemia |
| SNOMED CT | 1172704005 | High grade B-cell lymphoma with MYC and BCL2 and/or BCL6 rearrangements |
| SNOMED CT | 1172516002 | High-dose chemotherapy with stem cell transplant |
| SNOMED CT | 1186654001 | Immune dysregulation, inflammatory bowel disease, arthritis, recurrent infection, lymphopenia syndrome |
| SNOMED CT | 1187508009 | Immunoglobulin G4 related aortitis |
| SNOMED CT | 1186834004 | Immunoglobulin G4 related hypophysitis |
| SNOMED CT | 1187507004 | Immunoglobulin G4 related kidney disease |
| SNOMED CT | 1187510006 | Immunoglobulin G4 related ophthalmic disease |
| SNOMED CT | 1186721005 | Infantile inflammatory bowel disease with neurological involvement |
| SNOMED CT | 1187000005 | Infectious mononucleosis caused by human immunodeficiency virus |
| SNOMED CT | 1187001009 | Infectious mononucleosis caused by human immunodeficiency virus type I |
| SNOMED CT | 1162768007 | Leukemia |
| SNOMED CT | 1187233008 | Leukocyte adhesion deficiency |
| SNOMED CT | 1169361002 | Limbic encephalitis with neurexin-3 antibodies |
| SNOMED CT | 1186646003 | Malignant histiocytic neoplasm |
| SNOMED CT | 1163043007 | Malignant lymphoma |
| SNOMED CT | 1162264008 | Malignant lymphoma of uveal tract |
| SNOMED CT | 1172895006 | Mendelian susceptibility to mycobacterial disease |
| SNOMED CT | 1172592001 | Non-Hodgkin lymphoma |
| SNOMED CT | 1186728004 | Pediatric multiple sclerosis |
| SNOMED CT | 1187623009 | Phosphoglucomutase 3-related congenital disorder of glycosylation |
| SNOMED CT | 1162576007 | Plasma cell myeloma |
| SNOMED CT | 1187107005 | Plasmacytoma |
| SNOMED CT | 1186719000 | Predisposition to invasive fungal disease due to CARD9 deficiency |
| SNOMED CT | 1172686000 | Protein contact dermatitis |
| SNOMED CT | 1162319009 | Rheumatoid factor and anti-citrullinated protein antibody positive erosive rheumatoid arthritis |
| SNOMED CT | 1162313005 | Rheumatoid factor and anti-citrullinated protein antibody positive rheumatoid arthritis |
| SNOMED CT | 1162303002 | Seronegative erosive rheumatoid arthritis |
| SNOMED CT | 1179284005 | Severe combined immunodeficiency due to LAT deficiency |
| SNOMED CT | 1177175008 | Skeletal dysplasia, T-cell immunodeficiency, developmental delay syndrome |
| SNOMED CT | 1186725001 | Warts, immunodeficiency, lymphedema, anogenital dysplasia syndrome |
| SNOMED CT | 1162828001 | X-linked lymphoproliferative disease due to SH2D1A deficiency |

Continued on next page

**TableS31 – continued from previous page**

| CodeSystem | ConceptCode | ConceptName |
|---|---|---|
| SNOMED CT | 1162830004 | X-linked lymphoproliferative disease due to XIAP deficiency |
| SNOMED CT | 1187193000 | Hairy cell leukemia variant |
| SNOMED CT | 1162505005 | Agranulocytosis caused by antithyroid agent |
| SNOMED CT | 1172729004 | ALK negative anaplastic large cell lymphoma |
| SNOMED CT | 15919911000119108 | Allergic reaction caused by analgesic |
| SNOMED CT | 15919991000119104 | Allergic reaction caused by anesthetic |
| SNOMED CT | 15920641000119103 | Allergic reaction caused by food additive |
| SNOMED CT | 15920241000119101 | Allergic reaction caused by fruit |
| SNOMED CT | 15920441000119100 | Allergic reaction caused by seed |
| SNOMED CT | 1172702009 | Anaplastic diffuse large B-cell lymphoma |
| SNOMED CT | 1172695008 | Diffuse large B cell lymphoma |
| SNOMED CT | 1172701002 | Facial diplegia with paresthesia |
| SNOMED CT | 1162362005 | Felty syndrome with seronegative erosive rheumatoid arthritis |
| SNOMED CT | 1162364006 | Felty syndrome with seronegative rheumatoid arthritis |
| ICD10CM | B20 | Human immunodeficiency virus [HIV] disease |
| ICD10CM | C00 | Malignant neoplasm of lip |
| ICD10CM | C01 | Malignant neoplasm of base of tongue |
| ICD10CM | C02 | Malignant neoplasm of other and unspecified parts of tongue |
| ICD10CM | C03 | Malignant neoplasm of gum |
| ICD10CM | C04 | Malignant neoplasm of floor of mouth |
| ICD10CM | C05 | Malignant neoplasm of palate |
| ICD10CM | C06 | Malignant neoplasm of other and unspecified parts of mouth |
| ICD10CM | C07 | Malignant neoplasm of parotid gland |
| ICD10CM | C08 | Malignant neoplasm of other and unspecified major salivary glands |
| ICD10CM | C09 | Malignant neoplasm of tonsil |
| ICD10CM | C10 | Malignant neoplasm of oropharynx |
| ICD10CM | C11 | Malignant neoplasm of nasopharynx |
| ICD10CM | C12 | Malignant neoplasm of pyriform sinus |
| ICD10CM | C13 | Malignant neoplasm of hypopharynx |
| ICD10CM | C14 | Malignant neoplasm of other and ill-defined sites in the lip, oral cavity and pharynx |
| ICD10CM | C15 | Malignant neoplasm of esophagus |
| ICD10CM | C16 | Malignant neoplasm of stomach |
| ICD10CM | C17 | Malignant neoplasm of small intestine |
| ICD10CM | C18 | Malignant neoplasm of colon |
| ICD10CM | C19 | Malignant neoplasm of rectosigmoid junction |
| ICD10CM | C20 | Malignant neoplasm of rectum |
| ICD10CM | C21 | Malignant neoplasm of anus and anal canal |
| ICD10CM | C22 | Malignant neoplasm of liver and intrahepatic bile ducts |
| ICD10CM | C23 | Malignant neoplasm of gallbladder |
| ICD10CM | C24 | Malignant neoplasm of other and unspecified parts of biliary tract |
| ICD10CM | C25 | Malignant neoplasm of pancreas |
| ICD10CM | C26 | Malignant neoplasm of other and ill-defined digestive organs |
| ICD10CM | C40 | Malignant neoplasm of bone and articular cartilage of limbs |

Continued on next page

**TableS31 – continued from previous page**

| CodeSystem | ConceptCode | ConceptName |
|---|---|---|
| ICD10CM | C41 | Malignant neoplasm of bone and articular cartilage of other and unspecified sites |
| ICD10CM | C43 | Malignant melanoma of skin |
| ICD10CM | C44 | Other and unspecified malignant neoplasm of skin |
| ICD10CM | C45 | Mesothelioma |
| ICD10CM | C46 | Kaposi's sarcoma |
| ICD10CM | C47 | Malignant neoplasm of peripheral nerves and autonomic nervous system |
| ICD10CM | C48 | Malignant neoplasm of retroperitoneum and peritoneum |
| ICD10CM | C49 | Malignant neoplasm of other connective and soft tissue |
| ICD10CM | C50 | Malignant neoplasm of breast |
| ICD10CM | C51 | Malignant neoplasm of vulva |
| ICD10CM | C52 | Malignant neoplasm of vagina |
| ICD10CM | C53 | Malignant neoplasm of cervix uteri |
| ICD10CM | C54 | Malignant neoplasm of corpus uteri |
| ICD10CM | C55 | Malignant neoplasm of uterus, part unspecified |
| ICD10CM | C56 | Malignant neoplasm of ovary |
| ICD10CM | C57 | Malignant neoplasm of other and unspecified female genital organs |
| ICD10CM | C58 | Malignant neoplasm of placenta |
| ICD10CM | C60 | Malignant neoplasm of penis |
| ICD10CM | C61 | Malignant neoplasm of prostate |
| ICD10CM | C62 | Malignant neoplasm of testis |
| ICD10CM | C63 | Malignant neoplasm of other and unspecified male genital organs |
| ICD10CM | C64 | Malignant neoplasm of kidney, except renal pelvis |
| ICD10CM | C65 | Malignant neoplasm of renal pelvis |
| ICD10CM | C66 | Malignant neoplasm of ureter |
| ICD10CM | C67 | Malignant neoplasm of bladder |
| ICD10CM | C68 | Malignant neoplasm of other and unspecified urinary organs |
| ICD10CM | C69 | Malignant neoplasm of eye and adnexa |
| ICD10CM | C70 | Malignant neoplasm of meninges |
| ICD10CM | C71 | Malignant neoplasm of brain |
| ICD10CM | C72 | Malignant neoplasm of spinal cord, cranial nerves and other parts of central nervous system |
| ICD10CM | C73 | Malignant neoplasm of thyroid gland |
| ICD10CM | C74 | Malignant neoplasm of adrenal gland |
| ICD10CM | C75 | Malignant neoplasm of other endocrine glands and related structures |
| ICD10CM | C76 | Malignant neoplasm of other and ill-defined sites |
| ICD10CM | C77 | Secondary and unspecified malignant neoplasm of lymph nodes |
| ICD10CM | C78 | Secondary malignant neoplasm of respiratory and digestive organs |
| ICD10CM | C79 | Secondary malignant neoplasm of other and unspecified sites |
| ICD10CM | C7A | Malignant neuroendocrine tumors |
| ICD10CM | C7B | Secondary neuroendocrine tumors |
| ICD10CM | C80 | Malignant neoplasm without specification of site |
| ICD10CM | C81 | Hodgkin lymphoma |

Continued on next page

**TableS31 – continued from previous page**

| CodeSystem | ConceptCode | ConceptName |
|---|---|---|
| ICD10CM | C82 | Follicular lymphoma |
| ICD10CM | C83 | Non-follicular lymphoma |
| ICD10CM | C84 | Mature T/NK-cell lymphomas |
| ICD10CM | C85 | Other specified and unspecified types of non-Hodgkin lymphoma |
| ICD10CM | C86 | Other specified types of T/NK-cell lymphoma |
| ICD10CM | C88 | Malignant immunoproliferative diseases and certain other B-cell lymphomas |
| ICD10CM | C90 | Multiple myeloma and malignant plasma cell neoplasms |
| ICD10CM | C91 | Lymphoid leukemia |
| ICD10CM | C92 | Myeloid leukemia |
| ICD10CM | C93 | Monocytic leukemia |
| ICD10CM | C94 | Other leukemias of specified cell type |
| ICD10CM | C95 | Leukemia of unspecified cell type |
| ICD10CM | C96 | Other and unspecified malignant neoplasms of lymphoid, hematopoietic and related tissue |
| ICD10CM | D00 | Carcinoma in situ of oral cavity, esophagus and stomach |
| ICD10CM | D01 | Carcinoma in situ of other and unspecified digestive organs |
| ICD10CM | D02 | Carcinoma in situ of middle ear and respiratory system |
| ICD10CM | D03 | Melanoma in situ |
| ICD10CM | D04 | Carcinoma in situ of skin |
| ICD10CM | D05 | Carcinoma in situ of breast |
| ICD10CM | D06 | Carcinoma in situ of cervix uteri |
| ICD10CM | D07 | Carcinoma in situ of other and unspecified genital organs |
| ICD10CM | D09 | Carcinoma in situ of other and unspecified sites |
| ICD10CM | D37 | Neoplasm of uncertain behavior of oral cavity and digestive organs |
| ICD10CM | D38 | Neoplasm of uncertain behavior of middle ear and respiratory and intrathoracic organs |
| ICD10CM | D39 | Neoplasm of uncertain behavior of female genital organs |
| ICD10CM | D40 | Neoplasm of uncertain behavior of male genital organs |
| ICD10CM | D41 | Neoplasm of uncertain behavior of urinary organs |
| ICD10CM | D42 | Neoplasm of uncertain behavior of meninges |
| ICD10CM | D43 | Neoplasm of uncertain behavior of brain and central nervous system |
| ICD10CM | D44 | Neoplasm of uncertain behavior of endocrine glands |
| ICD10CM | D80 | Immunodeficiency with predominantly antibody defects |
| ICD10CM | D81 | Combined immunodeficiencies |
| ICD10CM | D82 | Immunodeficiency associated with other major defects |
| ICD10CM | D83 | Common variable immunodeficiency |
| ICD10CM | D84 | Other immunodeficiencies |
| ICD10CM | D89 | Other disorders involving the immune mechanism, not elsewhere classified |
| ICD10CM | Z94 | Transplanted organ and tissue status |
| ICD10CM | C00.0 | Malignant neoplasm of external upper lip |
| ICD10CM | C00.1 | Malignant neoplasm of external lower lip |
| ICD10CM | C00.2 | Malignant neoplasm of external lip, unspecified |
| ICD10CM | C00.3 | Malignant neoplasm of upper lip, inner aspect |

Continued on next page

**TableS31 – continued from previous page**

| CodeSystem | ConceptCode | ConceptName |
|---|---|---|
| ICD10CM | C00.4 | Malignant neoplasm of lower lip, inner aspect |
| ICD10CM | C00.5 | Malignant neoplasm of lip, unspecified, inner aspect |
| ICD10CM | C00.6 | Malignant neoplasm of commissure of lip, unspecified |
| ICD10CM | C00.8 | Malignant neoplasm of overlapping sites of lip |
| ICD10CM | C00.9 | Malignant neoplasm of lip, unspecified |
| ICD10CM | C02.0 | Malignant neoplasm of dorsal surface of tongue |
| ICD10CM | C02.1 | Malignant neoplasm of border of tongue |
| ICD10CM | C02.2 | Malignant neoplasm of ventral surface of tongue |
| ICD10CM | C02.3 | Malignant neoplasm of anterior two-thirds of tongue, part unspecified |
| ICD10CM | C02.4 | Malignant neoplasm of lingual tonsil |
| ICD10CM | C02.8 | Malignant neoplasm of overlapping sites of tongue |
| ICD10CM | C02.9 | Malignant neoplasm of tongue, unspecified |
| ICD10CM | C03.0 | Malignant neoplasm of upper gum |
| ICD10CM | C03.1 | Malignant neoplasm of lower gum |
| ICD10CM | C03.9 | Malignant neoplasm of gum, unspecified |
| ICD10CM | C04.0 | Malignant neoplasm of anterior floor of mouth |
| ICD10CM | C04.1 | Malignant neoplasm of lateral floor of mouth |
| ICD10CM | C04.8 | Malignant neoplasm of overlapping sites of floor of mouth |
| ICD10CM | C04.9 | Malignant neoplasm of floor of mouth, unspecified |
| ICD10CM | C05.0 | Malignant neoplasm of hard palate |
| ICD10CM | C05.1 | Malignant neoplasm of soft palate |
| ICD10CM | C05.2 | Malignant neoplasm of uvula |
| ICD10CM | C05.8 | Malignant neoplasm of overlapping sites of palate |
| ICD10CM | C05.9 | Malignant neoplasm of palate, unspecified |
| ICD10CM | C06.0 | Malignant neoplasm of cheek mucosa |
| ICD10CM | C06.1 | Malignant neoplasm of vestibule of mouth |
| ICD10CM | C06.2 | Malignant neoplasm of retromolar area |
| ICD10CM | C06.8 | Malignant neoplasm of overlapping sites of other and unspecified parts of mouth |
| ICD10CM | C06.80 | Malignant neoplasm of overlapping sites of unspecified parts of mouth |
| ICD10CM | C06.89 | Malignant neoplasm of overlapping sites of other parts of mouth |
| ICD10CM | C06.9 | Malignant neoplasm of mouth, unspecified |
| ICD10CM | C08.0 | Malignant neoplasm of submandibular gland |
| ICD10CM | C08.1 | Malignant neoplasm of sublingual gland |
| ICD10CM | C08.9 | Malignant neoplasm of major salivary gland, unspecified |
| ICD10CM | C09.0 | Malignant neoplasm of tonsillar fossa |
| ICD10CM | C09.1 | Malignant neoplasm of tonsillar pillar (anterior) (posterior) |
| ICD10CM | C09.8 | Malignant neoplasm of overlapping sites of tonsil |
| ICD10CM | C09.9 | Malignant neoplasm of tonsil, unspecified |
| ICD10CM | C10.0 | Malignant neoplasm of vallecula |
| ICD10CM | C10.1 | Malignant neoplasm of anterior surface of epiglottis |
| ICD10CM | C10.2 | Malignant neoplasm of lateral wall of oropharynx |
| ICD10CM | C10.3 | Malignant neoplasm of posterior wall of oropharynx |
| ICD10CM | C10.4 | Malignant neoplasm of branchial cleft |
| ICD10CM | C10.8 | Malignant neoplasm of overlapping sites of oropharynx |

Continued on next page

**TableS31 – continued from previous page**

| CodeSystem | ConceptCode | ConceptName |
|---|---|---|
| ICD10CM | C10.9 | Malignant neoplasm of oropharynx, unspecified |
| ICD10CM | C11.0 | Malignant neoplasm of superior wall of nasopharynx |
| ICD10CM | C11.1 | Malignant neoplasm of posterior wall of nasopharynx |
| ICD10CM | C11.2 | Malignant neoplasm of lateral wall of nasopharynx |
| ICD10CM | C11.3 | Malignant neoplasm of anterior wall of nasopharynx |
| ICD10CM | C11.8 | Malignant neoplasm of overlapping sites of nasopharynx |
| ICD10CM | C11.9 | Malignant neoplasm of nasopharynx, unspecified |
| ICD10CM | C13.0 | Malignant neoplasm of postcricoid region |
| ICD10CM | C13.1 | Malignant neoplasm of aryepiglottic fold, hypopharyngeal aspect |
| ICD10CM | C13.2 | Malignant neoplasm of posterior wall of hypopharynx |
| ICD10CM | C13.8 | Malignant neoplasm of overlapping sites of hypopharynx |
| ICD10CM | C13.9 | Malignant neoplasm of hypopharynx, unspecified |
| ICD10CM | C14.0 | Malignant neoplasm of pharynx, unspecified |
| ICD10CM | C14.2 | Malignant neoplasm of Waldeyer's ring |
| ICD10CM | C14.8 | Malignant neoplasm of overlapping sites of lip, oral cavity and pharynx |
| ICD10CM | C15.3 | Malignant neoplasm of upper third of esophagus |
| ICD10CM | C15.4 | Malignant neoplasm of middle third of esophagus |
| ICD10CM | C15.5 | Malignant neoplasm of lower third of esophagus |
| ICD10CM | C15.8 | Malignant neoplasm of overlapping sites of esophagus |
| ICD10CM | C15.9 | Malignant neoplasm of esophagus, unspecified |
| ICD10CM | C16.0 | Malignant neoplasm of cardia |
| ICD10CM | C16.1 | Malignant neoplasm of fundus of stomach |
| ICD10CM | C16.2 | Malignant neoplasm of body of stomach |
| ICD10CM | C16.3 | Malignant neoplasm of pyloric antrum |
| ICD10CM | C16.4 | Malignant neoplasm of pylorus |
| ICD10CM | C16.5 | Malignant neoplasm of lesser curvature of stomach, unspecified |
| ICD10CM | C16.6 | Malignant neoplasm of greater curvature of stomach, unspecified |
| ICD10CM | C16.8 | Malignant neoplasm of overlapping sites of stomach |
| ICD10CM | C16.9 | Malignant neoplasm of stomach, unspecified |
| ICD10CM | C17.0 | Malignant neoplasm of duodenum |
| ICD10CM | C17.1 | Malignant neoplasm of jejunum |
| ICD10CM | C17.2 | Malignant neoplasm of ileum |
| ICD10CM | C17.3 | Meckel's diverticulum, malignant |
| ICD10CM | C17.8 | Malignant neoplasm of overlapping sites of small intestine |
| ICD10CM | C17.9 | Malignant neoplasm of small intestine, unspecified |
| ICD10CM | C18.0 | Malignant neoplasm of cecum |
| ICD10CM | C18.1 | Malignant neoplasm of appendix |
| ICD10CM | C18.2 | Malignant neoplasm of ascending colon |
| ICD10CM | C18.3 | Malignant neoplasm of hepatic flexure |
| ICD10CM | C18.4 | Malignant neoplasm of transverse colon |
| ICD10CM | C18.5 | Malignant neoplasm of splenic flexure |
| ICD10CM | C18.6 | Malignant neoplasm of descending colon |
| ICD10CM | C18.7 | Malignant neoplasm of sigmoid colon |
| ICD10CM | C18.8 | Malignant neoplasm of overlapping sites of colon |
| ICD10CM | C18.9 | Malignant neoplasm of colon, unspecified |
| ICD10CM | C21.0 | Malignant neoplasm of anus, unspecified |

Continued on next page

**TableS31 – continued from previous page**

| CodeSystem | ConceptCode | ConceptName |
|---|---|---|
| ICD10CM | C21.1 | Malignant neoplasm of anal canal |
| ICD10CM | C21.2 | Malignant neoplasm of cloacogenic zone |
| ICD10CM | C21.8 | Malignant neoplasm of overlapping sites of rectum, anus and anal canal |
| ICD10CM | C22.0 | Liver cell carcinoma |
| ICD10CM | C22.1 | Intrahepatic bile duct carcinoma |
| ICD10CM | C22.2 | Hepatoblastoma |
| ICD10CM | C22.3 | Angiosarcoma of liver |
| ICD10CM | C22.4 | Other sarcomas of liver |
| ICD10CM | C22.7 | Other specified carcinomas of liver |
| ICD10CM | C22.8 | Malignant neoplasm of liver, primary, unspecified as to type |
| ICD10CM | C22.9 | Malignant neoplasm of liver, not specified as primary or secondary |
| ICD10CM | C24.0 | Malignant neoplasm of extrahepatic bile duct |
| ICD10CM | C24.1 | Malignant neoplasm of ampulla of Vater |
| ICD10CM | C24.8 | Malignant neoplasm of overlapping sites of biliary tract |
| ICD10CM | C24.9 | Malignant neoplasm of biliary tract, unspecified |
| ICD10CM | C25.0 | Malignant neoplasm of head of pancreas |
| ICD10CM | C25.1 | Malignant neoplasm of body of pancreas |
| ICD10CM | C25.2 | Malignant neoplasm of tail of pancreas |
| ICD10CM | C25.3 | Malignant neoplasm of pancreatic duct |
| ICD10CM | C25.4 | Malignant neoplasm of endocrine pancreas |
| ICD10CM | C25.7 | Malignant neoplasm of other parts of pancreas |
| ICD10CM | C25.8 | Malignant neoplasm of overlapping sites of pancreas |
| ICD10CM | C25.9 | Malignant neoplasm of pancreas, unspecified |
| ICD10CM | C26.0 | Malignant neoplasm of intestinal tract, part unspecified |
| ICD10CM | C26.1 | Malignant neoplasm of spleen |
| ICD10CM | C26.9 | Malignant neoplasm of ill-defined sites within the digestive system |
| ICD10CM | C40.0 | Malignant neoplasm of scapula and long bones of upper limb |
| ICD10CM | C40.00 | Malignant neoplasm of scapula and long bones of unspecified upper limb |
| ICD10CM | C40.01 | Malignant neoplasm of scapula and long bones of right upper limb |
| ICD10CM | C40.02 | Malignant neoplasm of scapula and long bones of left upper limb |
| ICD10CM | C40.1 | Malignant neoplasm of short bones of upper limb |
| ICD10CM | C40.10 | Malignant neoplasm of short bones of unspecified upper limb |
| ICD10CM | C40.11 | Malignant neoplasm of short bones of right upper limb |
| ICD10CM | C40.12 | Malignant neoplasm of short bones of left upper limb |
| ICD10CM | C40.2 | Malignant neoplasm of long bones of lower limb |
| ICD10CM | C40.20 | Malignant neoplasm of long bones of unspecified lower limb |
| ICD10CM | C40.21 | Malignant neoplasm of long bones of right lower limb |
| ICD10CM | C40.22 | Malignant neoplasm of long bones of left lower limb |
| ICD10CM | C40.3 | Malignant neoplasm of short bones of lower limb |
| ICD10CM | C40.30 | Malignant neoplasm of short bones of unspecified lower limb |
| ICD10CM | C40.31 | Malignant neoplasm of short bones of right lower limb |
| ICD10CM | C40.32 | Malignant neoplasm of short bones of left lower limb |

Continued on next page

**TableS31 – continued from previous page**

| CodeSystem | ConceptCode | ConceptName |
|---|---|---|
| ICD10CM | C40.8 | Malignant neoplasm of overlapping sites of bone and articular cartilage of limb |
| ICD10CM | C40.80 | Malignant neoplasm of overlapping sites of bone and articular cartilage of unspecified limb |
| ICD10CM | C40.81 | Malignant neoplasm of overlapping sites of bone and articular cartilage of right limb |
| ICD10CM | C40.82 | Malignant neoplasm of overlapping sites of bone and articular cartilage of left limb |
| ICD10CM | C40.9 | Malignant neoplasm of unspecified bones and articular cartilage of limb |
| ICD10CM | C40.90 | Malignant neoplasm of unspecified bones and articular cartilage of unspecified limb |
| ICD10CM | C40.91 | Malignant neoplasm of unspecified bones and articular cartilage of right limb |
| ICD10CM | C40.92 | Malignant neoplasm of unspecified bones and articular cartilage of left limb |
| ICD10CM | C41.0 | Malignant neoplasm of bones of skull and face |
| ICD10CM | C41.1 | Malignant neoplasm of mandible |
| ICD10CM | C41.2 | Malignant neoplasm of vertebral column |
| ICD10CM | C41.3 | Malignant neoplasm of ribs, sternum and clavicle |
| ICD10CM | C41.4 | Malignant neoplasm of pelvic bones, sacrum and coccyx |
| ICD10CM | C41.9 | Malignant neoplasm of bone and articular cartilage, unspecified |
| ICD10CM | C43.0 | Malignant melanoma of lip |
| ICD10CM | C43.1 | Malignant melanoma of eyelid, including canthus |
| ICD10CM | C43.10 | Malignant melanoma of unspecified eyelid, including canthus |
| ICD10CM | C43.11 | Malignant melanoma of right eyelid, including canthus |
| ICD10CM | C43.111 | Malignant melanoma of right upper eyelid, including canthus |
| ICD10CM | C43.112 | Malignant melanoma of right lower eyelid, including canthus |
| ICD10CM | C43.12 | Malignant melanoma of left eyelid, including canthus |
| ICD10CM | C43.121 | Malignant melanoma of left upper eyelid, including canthus |
| ICD10CM | C43.122 | Malignant melanoma of left lower eyelid, including canthus |
| ICD10CM | C43.2 | Malignant melanoma of ear and external auricular canal |
| ICD10CM | C43.20 | Malignant melanoma of unspecified ear and external auricular canal |
| ICD10CM | C43.21 | Malignant melanoma of right ear and external auricular canal |
| ICD10CM | C43.22 | Malignant melanoma of left ear and external auricular canal |
| ICD10CM | C43.3 | Malignant melanoma of other and unspecified parts of face |
| ICD10CM | C43.30 | Malignant melanoma of unspecified part of face |
| ICD10CM | C43.31 | Malignant melanoma of nose |
| ICD10CM | C43.39 | Malignant melanoma of other parts of face |
| ICD10CM | C43.4 | Malignant melanoma of scalp and neck |
| ICD10CM | C43.5 | Malignant melanoma of trunk |
| ICD10CM | C43.51 | Malignant melanoma of anal skin |
| ICD10CM | C43.52 | Malignant melanoma of skin of breast |
| ICD10CM | C43.59 | Malignant melanoma of other part of trunk |
| ICD10CM | C43.6 | Malignant melanoma of upper limb, including shoulder |

Continued on next page

**TableS31 – continued from previous page**

| CodeSystem | ConceptCode | ConceptName |
|---|---|---|
| ICD10CM | C43.60 | Malignant melanoma of unspecified upper limb, including shoulder |
| ICD10CM | C43.61 | Malignant melanoma of right upper limb, including shoulder |
| ICD10CM | C43.62 | Malignant melanoma of left upper limb, including shoulder |
| ICD10CM | C43.7 | Malignant melanoma of lower limb, including hip |
| ICD10CM | C43.70 | Malignant melanoma of unspecified lower limb, including hip |
| ICD10CM | C43.71 | Malignant melanoma of right lower limb, including hip |
| ICD10CM | C43.72 | Malignant melanoma of left lower limb, including hip |
| ICD10CM | C43.8 | Malignant melanoma of overlapping sites of skin |
| ICD10CM | C43.9 | Malignant melanoma of skin, unspecified |
| ICD10CM | C44.0 | Other and unspecified malignant neoplasm of skin of lip |
| ICD10CM | C44.00 | Unspecified malignant neoplasm of skin of lip |
| ICD10CM | C44.01 | Basal cell carcinoma of skin of lip |
| ICD10CM | C44.02 | Squamous cell carcinoma of skin of lip |
| ICD10CM | C44.09 | Other specified malignant neoplasm of skin of lip |
| ICD10CM | C44.1 | Other and unspecified malignant neoplasm of skin of eyelid, including canthus |
| ICD10CM | C44.10 | Unspecified malignant neoplasm of skin of eyelid, including canthus |
| ICD10CM | C44.101 | Unspecified malignant neoplasm of skin of unspecified eyelid, including canthus |
| ICD10CM | C44.102 | Unspecified malignant neoplasm of skin of right eyelid, including canthus |
| ICD10CM | C44.1021 | Unspecified malignant neoplasm of skin of right upper eyelid, including canthus |
| ICD10CM | C44.1022 | Unspecified malignant neoplasm of skin of right lower eyelid, including canthus |
| ICD10CM | C44.109 | Unspecified malignant neoplasm of skin of left eyelid, including canthus |
| ICD10CM | C44.1091 | Unspecified malignant neoplasm of skin of left upper eyelid, including canthus |
| ICD10CM | C44.1092 | Unspecified malignant neoplasm of skin of left lower eyelid, including canthus |
| ICD10CM | C44.11 | Basal cell carcinoma of skin of eyelid, including canthus |
| ICD10CM | C44.111 | Basal cell carcinoma of skin of unspecified eyelid, including canthus |
| ICD10CM | C44.112 | Basal cell carcinoma of skin of right eyelid, including canthus |
| ICD10CM | C44.1121 | Basal cell carcinoma of skin of right upper eyelid, including canthus |
| ICD10CM | C44.1122 | Basal cell carcinoma of skin of right lower eyelid, including canthus |
| ICD10CM | C44.119 | Basal cell carcinoma of skin of left eyelid, including canthus |
| ICD10CM | C44.1191 | Basal cell carcinoma of skin of left upper eyelid, including canthus |
| ICD10CM | C44.1192 | Basal cell carcinoma of skin of left lower eyelid, including canthus |
| ICD10CM | C44.12 | Squamous cell carcinoma of skin of eyelid, including canthus |

Continued on next page

**TableS31 – continued from previous page**

| CodeSystem | ConceptCode | ConceptName |
|---|---|---|
| ICD10CM | C44.121 | Squamous cell carcinoma of skin of unspecified eyelid, including canthus |
| ICD10CM | C44.122 | Squamous cell carcinoma of skin of right eyelid, including canthus |
| ICD10CM | C44.1221 | Squamous cell carcinoma of skin of right upper eyelid, including canthus |
| ICD10CM | C44.1222 | Squamous cell carcinoma of skin of right lower eyelid, including canthus |
| ICD10CM | C44.129 | Squamous cell carcinoma of skin of left eyelid, including canthus |
| ICD10CM | C44.1291 | Squamous cell carcinoma of skin of left upper eyelid, including canthus |
| ICD10CM | C44.1292 | Squamous cell carcinoma of skin of left lower eyelid, including canthus |
| ICD10CM | C44.13 | Sebaceous cell carcinoma of skin of eyelid, including canthus |
| ICD10CM | C44.131 | Sebaceous cell carcinoma of skin of unspecified eyelid, including canthus |
| ICD10CM | C44.132 | Sebaceous cell carcinoma of skin of right eyelid, including canthus |
| ICD10CM | C44.1321 | Sebaceous cell carcinoma of skin of right upper eyelid, including canthus |
| ICD10CM | C44.1322 | Sebaceous cell carcinoma of skin of right lower eyelid, including canthus |
| ICD10CM | C44.139 | Sebaceous cell carcinoma of skin of left eyelid, including canthus |
| ICD10CM | C44.1391 | Sebaceous cell carcinoma of skin of left upper eyelid, including canthus |
| ICD10CM | C44.1392 | Sebaceous cell carcinoma of skin of left lower eyelid, including canthus |
| ICD10CM | C44.19 | Other specified malignant neoplasm of skin of eyelid, including canthus |
| ICD10CM | C44.191 | Other specified malignant neoplasm of skin of unspecified eyelid, including canthus |
| ICD10CM | C44.192 | Other specified malignant neoplasm of skin of right eyelid, including canthus |
| ICD10CM | C44.1921 | Other specified malignant neoplasm of skin of right upper eyelid, including canthus |
| ICD10CM | C44.1922 | Other specified malignant neoplasm of skin of right lower eyelid, including canthus |
| ICD10CM | C44.199 | Other specified malignant neoplasm of skin of left eyelid, including canthus |
| ICD10CM | C44.1991 | Other specified malignant neoplasm of skin of left upper eyelid, including canthus |
| ICD10CM | C44.1992 | Other specified malignant neoplasm of skin of left lower eyelid, including canthus |
| ICD10CM | C44.2 | Other and unspecified malignant neoplasm of skin of ear and external auricular canal |
| ICD10CM | C44.20 | Unspecified malignant neoplasm of skin of ear and external auricular canal |

Continued on next page

**TableS31 – continued from previous page**

| CodeSystem | ConceptCode | ConceptName |
|---|---|---|
| ICD10CM | C44.201 | Unspecified malignant neoplasm of skin of unspecified ear and external auricular canal |
| ICD10CM | C44.202 | Unspecified malignant neoplasm of skin of right ear and external auricular canal |
| ICD10CM | C44.209 | Unspecified malignant neoplasm of skin of left ear and external auricular canal |
| ICD10CM | C44.21 | Basal cell carcinoma of skin of ear and external auricular canal |
| ICD10CM | C44.211 | Basal cell carcinoma of skin of unspecified ear and external auricular canal |
| ICD10CM | C44.212 | Basal cell carcinoma of skin of right ear and external auricular canal |
| ICD10CM | C44.219 | Basal cell carcinoma of skin of left ear and external auricular canal |
| ICD10CM | C44.22 | Squamous cell carcinoma of skin of ear and external auricular canal |
| ICD10CM | C44.221 | Squamous cell carcinoma of skin of unspecified ear and external auricular canal |
| ICD10CM | C44.222 | Squamous cell carcinoma of skin of right ear and external auricular canal |
| ICD10CM | C44.229 | Squamous cell carcinoma of skin of left ear and external auricular canal |
| ICD10CM | C44.29 | Other specified malignant neoplasm of skin of ear and external auricular canal |
| ICD10CM | C44.291 | Other specified malignant neoplasm of skin of unspecified ear and external auricular canal |
| ICD10CM | C44.292 | Other specified malignant neoplasm of skin of right ear and external auricular canal |
| ICD10CM | C44.299 | Other specified malignant neoplasm of skin of left ear and external auricular canal |
| ICD10CM | C44.3 | Other and unspecified malignant neoplasm of skin of other and unspecified parts of face |
| ICD10CM | C44.30 | Unspecified malignant neoplasm of skin of other and unspecified parts of face |
| ICD10CM | C44.300 | Unspecified malignant neoplasm of skin of unspecified part of face |
| ICD10CM | C44.301 | Unspecified malignant neoplasm of skin of nose |
| ICD10CM | C44.309 | Unspecified malignant neoplasm of skin of other parts of face |
| ICD10CM | C44.31 | Basal cell carcinoma of skin of other and unspecified parts of face |
| ICD10CM | C44.310 | Basal cell carcinoma of skin of unspecified parts of face |
| ICD10CM | C44.311 | Basal cell carcinoma of skin of nose |
| ICD10CM | C44.319 | Basal cell carcinoma of skin of other parts of face |
| ICD10CM | C44.32 | Squamous cell carcinoma of skin of other and unspecified parts of face |
| ICD10CM | C44.320 | Squamous cell carcinoma of skin of unspecified parts of face |
| ICD10CM | C44.321 | Squamous cell carcinoma of skin of nose |
| ICD10CM | C44.329 | Squamous cell carcinoma of skin of other parts of face |

Continued on next page

**TableS31 – continued from previous page**

| CodeSystem | ConceptCode | ConceptName |
|---|---|---|
| ICD10CM | C44.39 | Other specified malignant neoplasm of skin of other and unspecified parts of face |
| ICD10CM | C44.390 | Other specified malignant neoplasm of skin of unspecified parts of face |
| ICD10CM | C44.391 | Other specified malignant neoplasm of skin of nose |
| ICD10CM | C44.399 | Other specified malignant neoplasm of skin of other parts of face |
| ICD10CM | C44.4 | Other and unspecified malignant neoplasm of skin of scalp and neck |
| ICD10CM | C44.40 | Unspecified malignant neoplasm of skin of scalp and neck |
| ICD10CM | C44.41 | Basal cell carcinoma of skin of scalp and neck |
| ICD10CM | C44.42 | Squamous cell carcinoma of skin of scalp and neck |
| ICD10CM | C44.49 | Other specified malignant neoplasm of skin of scalp and neck |
| ICD10CM | C44.5 | Other and unspecified malignant neoplasm of skin of trunk |
| ICD10CM | C44.50 | Unspecified malignant neoplasm of skin of trunk |
| ICD10CM | C44.500 | Unspecified malignant neoplasm of anal skin |
| ICD10CM | C44.501 | Unspecified malignant neoplasm of skin of breast |
| ICD10CM | C44.509 | Unspecified malignant neoplasm of skin of other part of trunk |
| ICD10CM | C44.51 | Basal cell carcinoma of skin of trunk |
| ICD10CM | C44.510 | Basal cell carcinoma of anal skin |
| ICD10CM | C44.511 | Basal cell carcinoma of skin of breast |
| ICD10CM | C44.519 | Basal cell carcinoma of skin of other part of trunk |
| ICD10CM | C44.52 | Squamous cell carcinoma of skin of trunk |
| ICD10CM | C44.520 | Squamous cell carcinoma of anal skin |
| ICD10CM | C44.521 | Squamous cell carcinoma of skin of breast |
| ICD10CM | C44.529 | Squamous cell carcinoma of skin of other part of trunk |
| ICD10CM | C44.59 | Other specified malignant neoplasm of skin of trunk |
| ICD10CM | C44.590 | Other specified malignant neoplasm of anal skin |
| ICD10CM | C44.591 | Other specified malignant neoplasm of skin of breast |
| ICD10CM | C44.599 | Other specified malignant neoplasm of skin of other part of trunk |
| ICD10CM | C44.6 | Other and unspecified malignant neoplasm of skin of upper limb, including shoulder |
| ICD10CM | C44.60 | Unspecified malignant neoplasm of skin of upper limb, including shoulder |
| ICD10CM | C44.601 | Unspecified malignant neoplasm of skin of unspecified upper limb, including shoulder |
| ICD10CM | C44.602 | Unspecified malignant neoplasm of skin of right upper limb, including shoulder |
| ICD10CM | C44.609 | Unspecified malignant neoplasm of skin of left upper limb, including shoulder |
| ICD10CM | C44.61 | Basal cell carcinoma of skin of upper limb, including shoulder |
| ICD10CM | C44.611 | Basal cell carcinoma of skin of unspecified upper limb, including shoulder |
| ICD10CM | C44.612 | Basal cell carcinoma of skin of right upper limb, including shoulder |
| ICD10CM | C44.619 | Basal cell carcinoma of skin of left upper limb, including shoulder |
| ICD10CM | C44.62 | Squamous cell carcinoma of skin of upper limb, including shoulder |

Continued on next page

**TableS31 – continued from previous page**

| CodeSystem | ConceptCode | ConceptName |
|---|---|---|
| ICD10CM | C44.621 | Squamous cell carcinoma of skin of unspecified upper limb, including shoulder |
| ICD10CM | C44.622 | Squamous cell carcinoma of skin of right upper limb, including shoulder |
| ICD10CM | C44.629 | Squamous cell carcinoma of skin of left upper limb, including shoulder |
| ICD10CM | C44.69 | Other specified malignant neoplasm of skin of upper limb, including shoulder |
| ICD10CM | C44.691 | Other specified malignant neoplasm of skin of unspecified upper limb, including shoulder |
| ICD10CM | C44.692 | Other specified malignant neoplasm of skin of right upper limb, including shoulder |
| ICD10CM | C44.699 | Other specified malignant neoplasm of skin of left upper limb, including shoulder |
| ICD10CM | C44.7 | Other and unspecified malignant neoplasm of skin of lower limb, including hip |
| ICD10CM | C44.70 | Unspecified malignant neoplasm of skin of lower limb, including hip |
| ICD10CM | C44.701 | Unspecified malignant neoplasm of skin of unspecified lower limb, including hip |
| ICD10CM | C44.702 | Unspecified malignant neoplasm of skin of right lower limb, including hip |
| ICD10CM | C44.709 | Unspecified malignant neoplasm of skin of left lower limb, including hip |
| ICD10CM | C44.71 | Basal cell carcinoma of skin of lower limb, including hip |
| ICD10CM | C44.711 | Basal cell carcinoma of skin of unspecified lower limb, including hip |
| ICD10CM | C44.712 | Basal cell carcinoma of skin of right lower limb, including hip |
| ICD10CM | C44.719 | Basal cell carcinoma of skin of left lower limb, including hip |
| ICD10CM | C44.72 | Squamous cell carcinoma of skin of lower limb, including hip |
| ICD10CM | C44.721 | Squamous cell carcinoma of skin of unspecified lower limb, including hip |
| ICD10CM | C44.722 | Squamous cell carcinoma of skin of right lower limb, including hip |
| ICD10CM | C44.729 | Squamous cell carcinoma of skin of left lower limb, including hip |
| ICD10CM | C44.79 | Other specified malignant neoplasm of skin of lower limb, including hip |
| ICD10CM | C44.791 | Other specified malignant neoplasm of skin of unspecified lower limb, including hip |
| ICD10CM | C44.792 | Other specified malignant neoplasm of skin of right lower limb, including hip |
| ICD10CM | C44.799 | Other specified malignant neoplasm of skin of left lower limb, including hip |
| ICD10CM | C44.8 | Other and unspecified malignant neoplasm of overlapping sites of skin |
| ICD10CM | C44.80 | Unspecified malignant neoplasm of overlapping sites of skin |
| ICD10CM | C44.81 | Basal cell carcinoma of overlapping sites of skin |

Continued on next page

**TableS31 – continued from previous page**

| CodeSystem | ConceptCode | ConceptName |
|---|---|---|
| ICD10CM | C44.82 | Squamous cell carcinoma of overlapping sites of skin |
| ICD10CM | C44.89 | Other specified malignant neoplasm of overlapping sites of skin |
| ICD10CM | C44.9 | Other and unspecified malignant neoplasm of skin, unspecified |
| ICD10CM | C44.90 | Unspecified malignant neoplasm of skin, unspecified |
| ICD10CM | C44.91 | Basal cell carcinoma of skin, unspecified |
| ICD10CM | C44.92 | Squamous cell carcinoma of skin, unspecified |
| ICD10CM | C44.99 | Other specified malignant neoplasm of skin, unspecified |
| ICD10CM | C45.0 | Mesothelioma of pleura |
| ICD10CM | C45.1 | Mesothelioma of peritoneum |
| ICD10CM | C45.2 | Mesothelioma of pericardium |
| ICD10CM | C45.7 | Mesothelioma of other sites |
| ICD10CM | C45.9 | Mesothelioma, unspecified |
| ICD10CM | C46.0 | Kaposi's sarcoma of skin |
| ICD10CM | C46.1 | Kaposi's sarcoma of soft tissue |
| ICD10CM | C46.2 | Kaposi's sarcoma of palate |
| ICD10CM | C46.3 | Kaposi's sarcoma of lymph nodes |
| ICD10CM | C46.4 | Kaposi's sarcoma of gastrointestinal sites |
| ICD10CM | C46.5 | Kaposi's sarcoma of lung |
| ICD10CM | C46.50 | Kaposi's sarcoma of unspecified lung |
| ICD10CM | C46.51 | Kaposi's sarcoma of right lung |
| ICD10CM | C46.52 | Kaposi's sarcoma of left lung |
| ICD10CM | C46.7 | Kaposi's sarcoma of other sites |
| ICD10CM | C46.9 | Kaposi's sarcoma, unspecified |
| ICD10CM | C47.0 | Malignant neoplasm of peripheral nerves of head, face and neck |
| ICD10CM | C47.1 | Malignant neoplasm of peripheral nerves of upper limb, including shoulder |
| ICD10CM | C47.10 | Malignant neoplasm of peripheral nerves of unspecified upper limb, including shoulder |
| ICD10CM | C47.11 | Malignant neoplasm of peripheral nerves of right upper limb, including shoulder |
| ICD10CM | C47.12 | Malignant neoplasm of peripheral nerves of left upper limb, including shoulder |
| ICD10CM | C47.2 | Malignant neoplasm of peripheral nerves of lower limb, including hip |
| ICD10CM | C47.20 | Malignant neoplasm of peripheral nerves of unspecified lower limb, including hip |
| ICD10CM | C47.21 | Malignant neoplasm of peripheral nerves of right lower limb, including hip |
| ICD10CM | C47.22 | Malignant neoplasm of peripheral nerves of left lower limb, including hip |
| ICD10CM | C47.3 | Malignant neoplasm of peripheral nerves of thorax |
| ICD10CM | C47.4 | Malignant neoplasm of peripheral nerves of abdomen |
| ICD10CM | C47.5 | Malignant neoplasm of peripheral nerves of pelvis |
| ICD10CM | C47.6 | Malignant neoplasm of peripheral nerves of trunk, unspecified |
| ICD10CM | C47.8 | Malignant neoplasm of overlapping sites of peripheral nerves and autonomic nervous system |

Continued on next page

**TableS31 – continued from previous page**

| CodeSystem | ConceptCode | ConceptName |
|---|---|---|
| ICD10CM | C47.9 | Malignant neoplasm of peripheral nerves and autonomic nervous system, unspecified |
| ICD10CM | C48.0 | Malignant neoplasm of retroperitoneum |
| ICD10CM | C48.1 | Malignant neoplasm of specified parts of peritoneum |
| ICD10CM | C48.2 | Malignant neoplasm of peritoneum, unspecified |
| ICD10CM | C48.8 | Malignant neoplasm of overlapping sites of retroperitoneum and peritoneum |
| ICD10CM | C49.0 | Malignant neoplasm of connective and soft tissue of head, face and neck |
| ICD10CM | C49.1 | Malignant neoplasm of connective and soft tissue of upper limb, including shoulder |
| ICD10CM | C49.10 | Malignant neoplasm of connective and soft tissue of unspecified upper limb, including shoulder |
| ICD10CM | C49.11 | Malignant neoplasm of connective and soft tissue of right upper limb, including shoulder |
| ICD10CM | C49.12 | Malignant neoplasm of connective and soft tissue of left upper limb, including shoulder |
| ICD10CM | C49.2 | Malignant neoplasm of connective and soft tissue of lower limb, including hip |
| ICD10CM | C49.20 | Malignant neoplasm of connective and soft tissue of unspecified lower limb, including hip |
| ICD10CM | C49.21 | Malignant neoplasm of connective and soft tissue of right lower limb, including hip |
| ICD10CM | C49.22 | Malignant neoplasm of connective and soft tissue of left lower limb, including hip |
| ICD10CM | C49.3 | Malignant neoplasm of connective and soft tissue of thorax |
| ICD10CM | C49.4 | Malignant neoplasm of connective and soft tissue of abdomen |
| ICD10CM | C49.5 | Malignant neoplasm of connective and soft tissue of pelvis |
| ICD10CM | C49.6 | Malignant neoplasm of connective and soft tissue of trunk, unspecified |
| ICD10CM | C49.8 | Malignant neoplasm of overlapping sites of connective and soft tissue |
| ICD10CM | C49.9 | Malignant neoplasm of connective and soft tissue, unspecified |
| ICD10CM | C49.A | Gastrointestinal stromal tumor |
| ICD10CM | C49.A0 | Gastrointestinal stromal tumor, unspecified site |
| ICD10CM | C49.A1 | Gastrointestinal stromal tumor of esophagus |
| ICD10CM | C49.A2 | Gastrointestinal stromal tumor of stomach |
| ICD10CM | C49.A3 | Gastrointestinal stromal tumor of small intestine |
| ICD10CM | C49.A4 | Gastrointestinal stromal tumor of large intestine |
| ICD10CM | C49.A5 | Gastrointestinal stromal tumor of rectum |
| ICD10CM | C49.A9 | Gastrointestinal stromal tumor of other sites |
| ICD10CM | C50.0 | Malignant neoplasm of nipple and areola |
| ICD10CM | C50.01 | Malignant neoplasm of nipple and areola, female |
| ICD10CM | C50.011 | Malignant neoplasm of nipple and areola, right female breast |
| ICD10CM | C50.012 | Malignant neoplasm of nipple and areola, left female breast |
| ICD10CM | C50.019 | Malignant neoplasm of nipple and areola, unspecified female breast |

Continued on next page

**TableS31 – continued from previous page**

| CodeSystem | ConceptCode | ConceptName |
|---|---|---|
| ICD10CM | C50.02 | Malignant neoplasm of nipple and areola, male |
| ICD10CM | C50.021 | Malignant neoplasm of nipple and areola, right male breast |
| ICD10CM | C50.022 | Malignant neoplasm of nipple and areola, left male breast |
| ICD10CM | C50.029 | Malignant neoplasm of nipple and areola, unspecified male breast |
| ICD10CM | C50.1 | Malignant neoplasm of central portion of breast |
| ICD10CM | C50.11 | Malignant neoplasm of central portion of breast, female |
| ICD10CM | C50.111 | Malignant neoplasm of central portion of right female breast |
| ICD10CM | C50.112 | Malignant neoplasm of central portion of left female breast |
| ICD10CM | C50.119 | Malignant neoplasm of central portion of unspecified female breast |
| ICD10CM | C50.12 | Malignant neoplasm of central portion of breast, male |
| ICD10CM | C50.121 | Malignant neoplasm of central portion of right male breast |
| ICD10CM | C50.122 | Malignant neoplasm of central portion of left male breast |
| ICD10CM | C50.129 | Malignant neoplasm of central portion of unspecified male breast |
| ICD10CM | C50.2 | Malignant neoplasm of upper-inner quadrant of breast |
| ICD10CM | C50.21 | Malignant neoplasm of upper-inner quadrant of breast, female |
| ICD10CM | C50.211 | Malignant neoplasm of upper-inner quadrant of right female breast |
| ICD10CM | C50.212 | Malignant neoplasm of upper-inner quadrant of left female breast |
| ICD10CM | C50.219 | Malignant neoplasm of upper-inner quadrant of unspecified female breast |
| ICD10CM | C50.22 | Malignant neoplasm of upper-inner quadrant of breast, male |
| ICD10CM | C50.221 | Malignant neoplasm of upper-inner quadrant of right male breast |
| ICD10CM | C50.222 | Malignant neoplasm of upper-inner quadrant of left male breast |
| ICD10CM | C50.229 | Malignant neoplasm of upper-inner quadrant of unspecified male breast |
| ICD10CM | C50.3 | Malignant neoplasm of lower-inner quadrant of breast |
| ICD10CM | C50.31 | Malignant neoplasm of lower-inner quadrant of breast, female |
| ICD10CM | C50.311 | Malignant neoplasm of lower-inner quadrant of right female breast |
| ICD10CM | C50.312 | Malignant neoplasm of lower-inner quadrant of left female breast |
| ICD10CM | C50.319 | Malignant neoplasm of lower-inner quadrant of unspecified female breast |
| ICD10CM | C50.32 | Malignant neoplasm of lower-inner quadrant of breast, male |
| ICD10CM | C50.321 | Malignant neoplasm of lower-inner quadrant of right male breast |
| ICD10CM | C50.322 | Malignant neoplasm of lower-inner quadrant of left male breast |
| ICD10CM | C50.329 | Malignant neoplasm of lower-inner quadrant of unspecified male breast |
| ICD10CM | C50.4 | Malignant neoplasm of upper-outer quadrant of breast |
| ICD10CM | C50.41 | Malignant neoplasm of upper-outer quadrant of breast, female |
| ICD10CM | C50.411 | Malignant neoplasm of upper-outer quadrant of right female breast |
| ICD10CM | C50.412 | Malignant neoplasm of upper-outer quadrant of left female breast |
| ICD10CM | C50.419 | Malignant neoplasm of upper-outer quadrant of unspecified female breast |
| ICD10CM | C50.42 | Malignant neoplasm of upper-outer quadrant of breast, male |
| ICD10CM | C50.421 | Malignant neoplasm of upper-outer quadrant of right male breast |

Continued on next page

**TableS31 – continued from previous page**

| CodeSystem | ConceptCode | ConceptName |
|---|---|---|
| ICD10CM | C50.422 | Malignant neoplasm of upper-outer quadrant of left male breast |
| ICD10CM | C50.429 | Malignant neoplasm of upper-outer quadrant of unspecified male breast |
| ICD10CM | C50.5 | Malignant neoplasm of lower-outer quadrant of breast |
| ICD10CM | C50.51 | Malignant neoplasm of lower-outer quadrant of breast, female |
| ICD10CM | C50.511 | Malignant neoplasm of lower-outer quadrant of right female breast |
| ICD10CM | C50.512 | Malignant neoplasm of lower-outer quadrant of left female breast |
| ICD10CM | C50.519 | Malignant neoplasm of lower-outer quadrant of unspecified female breast |
| ICD10CM | C50.52 | Malignant neoplasm of lower-outer quadrant of breast, male |
| ICD10CM | C50.521 | Malignant neoplasm of lower-outer quadrant of right male breast |
| ICD10CM | C50.522 | Malignant neoplasm of lower-outer quadrant of left male breast |
| ICD10CM | C50.529 | Malignant neoplasm of lower-outer quadrant of unspecified male breast |
| ICD10CM | C50.6 | Malignant neoplasm of axillary tail of breast |
| ICD10CM | C50.61 | Malignant neoplasm of axillary tail of breast, female |
| ICD10CM | C50.611 | Malignant neoplasm of axillary tail of right female breast |
| ICD10CM | C50.612 | Malignant neoplasm of axillary tail of left female breast |
| ICD10CM | C50.619 | Malignant neoplasm of axillary tail of unspecified female breast |
| ICD10CM | C50.62 | Malignant neoplasm of axillary tail of breast, male |
| ICD10CM | C50.621 | Malignant neoplasm of axillary tail of right male breast |
| ICD10CM | C50.622 | Malignant neoplasm of axillary tail of left male breast |
| ICD10CM | C50.629 | Malignant neoplasm of axillary tail of unspecified male breast |
| ICD10CM | C50.8 | Malignant neoplasm of overlapping sites of breast |
| ICD10CM | C50.81 | Malignant neoplasm of overlapping sites of breast, female |
| ICD10CM | C50.811 | Malignant neoplasm of overlapping sites of right female breast |
| ICD10CM | C50.812 | Malignant neoplasm of overlapping sites of left female breast |
| ICD10CM | C50.819 | Malignant neoplasm of overlapping sites of unspecified female breast |
| ICD10CM | C50.82 | Malignant neoplasm of overlapping sites of breast, male |
| ICD10CM | C50.821 | Malignant neoplasm of overlapping sites of right male breast |
| ICD10CM | C50.822 | Malignant neoplasm of overlapping sites of left male breast |
| ICD10CM | C50.829 | Malignant neoplasm of overlapping sites of unspecified male breast |
| ICD10CM | C50.9 | Malignant neoplasm of breast of unspecified site |
| ICD10CM | C50.91 | Malignant neoplasm of breast of unspecified site, female |
| ICD10CM | C50.911 | Malignant neoplasm of unspecified site of right female breast |
| ICD10CM | C50.912 | Malignant neoplasm of unspecified site of left female breast |
| ICD10CM | C50.919 | Malignant neoplasm of unspecified site of unspecified female breast |
| ICD10CM | C50.92 | Malignant neoplasm of breast of unspecified site, male |
| ICD10CM | C50.921 | Malignant neoplasm of unspecified site of right male breast |
| ICD10CM | C50.922 | Malignant neoplasm of unspecified site of left male breast |
| ICD10CM | C50.929 | Malignant neoplasm of unspecified site of unspecified male breast |
| ICD10CM | C51.0 | Malignant neoplasm of labium majus |
| ICD10CM | C51.1 | Malignant neoplasm of labium minus |

Continued on next page

**TableS31 – continued from previous page**

| CodeSystem | ConceptCode | ConceptName |
|---|---|---|
| ICD10CM | C51.2 | Malignant neoplasm of clitoris |
| ICD10CM | C51.8 | Malignant neoplasm of overlapping sites of vulva |
| ICD10CM | C51.9 | Malignant neoplasm of vulva, unspecified |
| ICD10CM | C53.0 | Malignant neoplasm of endocervix |
| ICD10CM | C53.1 | Malignant neoplasm of exocervix |
| ICD10CM | C53.8 | Malignant neoplasm of overlapping sites of cervix uteri |
| ICD10CM | C53.9 | Malignant neoplasm of cervix uteri, unspecified |
| ICD10CM | C54.0 | Malignant neoplasm of isthmus uteri |
| ICD10CM | C54.1 | Malignant neoplasm of endometrium |
| ICD10CM | C54.2 | Malignant neoplasm of myometrium |
| ICD10CM | C54.3 | Malignant neoplasm of fundus uteri |
| ICD10CM | C54.8 | Malignant neoplasm of overlapping sites of corpus uteri |
| ICD10CM | C54.9 | Malignant neoplasm of corpus uteri, unspecified |
| ICD10CM | C56.1 | Malignant neoplasm of right ovary |
| ICD10CM | C56.2 | Malignant neoplasm of left ovary |
| ICD10CM | C56.9 | Malignant neoplasm of unspecified ovary |
| ICD10CM | C57.0 | Malignant neoplasm of fallopian tube |
| ICD10CM | C57.00 | Malignant neoplasm of unspecified fallopian tube |
| ICD10CM | C57.01 | Malignant neoplasm of right fallopian tube |
| ICD10CM | C57.02 | Malignant neoplasm of left fallopian tube |
| ICD10CM | C57.1 | Malignant neoplasm of broad ligament |
| ICD10CM | C57.10 | Malignant neoplasm of unspecified broad ligament |
| ICD10CM | C57.11 | Malignant neoplasm of right broad ligament |
| ICD10CM | C57.12 | Malignant neoplasm of left broad ligament |
| ICD10CM | C57.2 | Malignant neoplasm of round ligament |
| ICD10CM | C57.20 | Malignant neoplasm of unspecified round ligament |
| ICD10CM | C57.21 | Malignant neoplasm of right round ligament |
| ICD10CM | C57.22 | Malignant neoplasm of left round ligament |
| ICD10CM | C57.3 | Malignant neoplasm of parametrium |
| ICD10CM | C57.4 | Malignant neoplasm of uterine adnexa, unspecified |
| ICD10CM | C57.7 | Malignant neoplasm of other specified female genital organs |
| ICD10CM | C57.8 | Malignant neoplasm of overlapping sites of female genital organs |
| ICD10CM | C57.9 | Malignant neoplasm of female genital organ, unspecified |
| ICD10CM | C60.0 | Malignant neoplasm of prepuce |
| ICD10CM | C60.1 | Malignant neoplasm of glans penis |
| ICD10CM | C60.2 | Malignant neoplasm of body of penis |
| ICD10CM | C60.8 | Malignant neoplasm of overlapping sites of penis |
| ICD10CM | C60.9 | Malignant neoplasm of penis, unspecified |
| ICD10CM | C62.0 | Malignant neoplasm of undescended testis |
| ICD10CM | C62.00 | Malignant neoplasm of unspecified undescended testis |
| ICD10CM | C62.01 | Malignant neoplasm of undescended right testis |
| ICD10CM | C62.02 | Malignant neoplasm of undescended left testis |
| ICD10CM | C62.1 | Malignant neoplasm of descended testis |
| ICD10CM | C62.10 | Malignant neoplasm of unspecified descended testis |
| ICD10CM | C62.11 | Malignant neoplasm of descended right testis |
| ICD10CM | C62.12 | Malignant neoplasm of descended left testis |

Continued on next page

**TableS31 – continued from previous page**

| CodeSystem | ConceptCode | ConceptName |
|---|---|---|
| ICD10CM | C62.9 | Malignant neoplasm of testis, unspecified whether descended or undescended |
| ICD10CM | C62.90 | Malignant neoplasm of unspecified testis, unspecified whether descended or undescended |
| ICD10CM | C62.91 | Malignant neoplasm of right testis, unspecified whether descended or undescended |
| ICD10CM | C62.92 | Malignant neoplasm of left testis, unspecified whether descended or undescended |
| ICD10CM | C63.0 | Malignant neoplasm of epididymis |
| ICD10CM | C63.00 | Malignant neoplasm of unspecified epididymis |
| ICD10CM | C63.01 | Malignant neoplasm of right epididymis |
| ICD10CM | C63.02 | Malignant neoplasm of left epididymis |
| ICD10CM | C63.1 | Malignant neoplasm of spermatic cord |
| ICD10CM | C63.10 | Malignant neoplasm of unspecified spermatic cord |
| ICD10CM | C63.11 | Malignant neoplasm of right spermatic cord |
| ICD10CM | C63.12 | Malignant neoplasm of left spermatic cord |
| ICD10CM | C63.2 | Malignant neoplasm of scrotum |
| ICD10CM | C63.7 | Malignant neoplasm of other specified male genital organs |
| ICD10CM | C63.8 | Malignant neoplasm of overlapping sites of male genital organs |
| ICD10CM | C63.9 | Malignant neoplasm of male genital organ, unspecified |
| ICD10CM | C64.1 | Malignant neoplasm of right kidney, except renal pelvis |
| ICD10CM | C64.2 | Malignant neoplasm of left kidney, except renal pelvis |
| ICD10CM | C64.9 | Malignant neoplasm of unspecified kidney, except renal pelvis |
| ICD10CM | C65.1 | Malignant neoplasm of right renal pelvis |
| ICD10CM | C65.2 | Malignant neoplasm of left renal pelvis |
| ICD10CM | C65.9 | Malignant neoplasm of unspecified renal pelvis |
| ICD10CM | C66.1 | Malignant neoplasm of right ureter |
| ICD10CM | C66.2 | Malignant neoplasm of left ureter |
| ICD10CM | C66.9 | Malignant neoplasm of unspecified ureter |
| ICD10CM | C67.0 | Malignant neoplasm of trigone of bladder |
| ICD10CM | C67.1 | Malignant neoplasm of dome of bladder |
| ICD10CM | C67.2 | Malignant neoplasm of lateral wall of bladder |
| ICD10CM | C67.3 | Malignant neoplasm of anterior wall of bladder |
| ICD10CM | C67.4 | Malignant neoplasm of posterior wall of bladder |
| ICD10CM | C67.5 | Malignant neoplasm of bladder neck |
| ICD10CM | C67.6 | Malignant neoplasm of ureteric orifice |
| ICD10CM | C67.7 | Malignant neoplasm of urachus |
| ICD10CM | C67.8 | Malignant neoplasm of overlapping sites of bladder |
| ICD10CM | C67.9 | Malignant neoplasm of bladder, unspecified |
| ICD10CM | C68.0 | Malignant neoplasm of urethra |
| ICD10CM | C68.1 | Malignant neoplasm of paraurethral glands |
| ICD10CM | C68.8 | Malignant neoplasm of overlapping sites of urinary organs |
| ICD10CM | C68.9 | Malignant neoplasm of urinary organ, unspecified |
| ICD10CM | C69.0 | Malignant neoplasm of conjunctiva |
| ICD10CM | C69.00 | Malignant neoplasm of unspecified conjunctiva |
| ICD10CM | C69.01 | Malignant neoplasm of right conjunctiva |
| ICD10CM | C69.02 | Malignant neoplasm of left conjunctiva |

Continued on next page

**TableS31 – continued from previous page**

| CodeSystem | ConceptCode | ConceptName |
|---|---|---|
| ICD10CM | C69.1 | Malignant neoplasm of cornea |
| ICD10CM | C69.10 | Malignant neoplasm of unspecified cornea |
| ICD10CM | C69.11 | Malignant neoplasm of right cornea |
| ICD10CM | C69.12 | Malignant neoplasm of left cornea |
| ICD10CM | C69.2 | Malignant neoplasm of retina |
| ICD10CM | C69.20 | Malignant neoplasm of unspecified retina |
| ICD10CM | C69.21 | Malignant neoplasm of right retina |
| ICD10CM | C69.22 | Malignant neoplasm of left retina |
| ICD10CM | C69.3 | Malignant neoplasm of choroid |
| ICD10CM | C69.30 | Malignant neoplasm of unspecified choroid |
| ICD10CM | C69.31 | Malignant neoplasm of right choroid |
| ICD10CM | C69.32 | Malignant neoplasm of left choroid |
| ICD10CM | C69.4 | Malignant neoplasm of ciliary body |
| ICD10CM | C69.40 | Malignant neoplasm of unspecified ciliary body |
| ICD10CM | C69.41 | Malignant neoplasm of right ciliary body |
| ICD10CM | C69.42 | Malignant neoplasm of left ciliary body |
| ICD10CM | C69.5 | Malignant neoplasm of lacrimal gland and duct |
| ICD10CM | C69.50 | Malignant neoplasm of unspecified lacrimal gland and duct |
| ICD10CM | C69.51 | Malignant neoplasm of right lacrimal gland and duct |
| ICD10CM | C69.52 | Malignant neoplasm of left lacrimal gland and duct |
| ICD10CM | C69.6 | Malignant neoplasm of orbit |
| ICD10CM | C69.60 | Malignant neoplasm of unspecified orbit |
| ICD10CM | C69.61 | Malignant neoplasm of right orbit |
| ICD10CM | C69.62 | Malignant neoplasm of left orbit |
| ICD10CM | C69.8 | Malignant neoplasm of overlapping sites of eye and adnexa |
| ICD10CM | C69.80 | Malignant neoplasm of overlapping sites of unspecified eye and adnexa |
| ICD10CM | C69.81 | Malignant neoplasm of overlapping sites of right eye and adnexa |
| ICD10CM | C69.82 | Malignant neoplasm of overlapping sites of left eye and adnexa |
| ICD10CM | C69.9 | Malignant neoplasm of unspecified site of eye |
| ICD10CM | C69.90 | Malignant neoplasm of unspecified site of unspecified eye |
| ICD10CM | C69.91 | Malignant neoplasm of unspecified site of right eye |
| ICD10CM | C69.92 | Malignant neoplasm of unspecified site of left eye |
| ICD10CM | C70.0 | Malignant neoplasm of cerebral meninges |
| ICD10CM | C70.1 | Malignant neoplasm of spinal meninges |
| ICD10CM | C70.9 | Malignant neoplasm of meninges, unspecified |
| ICD10CM | C71.0 | Malignant neoplasm of cerebrum, except lobes and ventricles |
| ICD10CM | C71.1 | Malignant neoplasm of frontal lobe |
| ICD10CM | C71.2 | Malignant neoplasm of temporal lobe |
| ICD10CM | C71.3 | Malignant neoplasm of parietal lobe |
| ICD10CM | C71.4 | Malignant neoplasm of occipital lobe |
| ICD10CM | C71.5 | Malignant neoplasm of cerebral ventricle |
| ICD10CM | C71.6 | Malignant neoplasm of cerebellum |
| ICD10CM | C71.7 | Malignant neoplasm of brain stem |
| ICD10CM | C71.8 | Malignant neoplasm of overlapping sites of brain |
| ICD10CM | C71.9 | Malignant neoplasm of brain, unspecified |
| ICD10CM | C72.0 | Malignant neoplasm of spinal cord |

Continued on next page

**TableS31 – continued from previous page**

| CodeSystem | ConceptCode | ConceptName |
|---|---|---|
| ICD10CM | C72.1 | Malignant neoplasm of cauda equina |
| ICD10CM | C72.2 | Malignant neoplasm of olfactory nerve |
| ICD10CM | C72.20 | Malignant neoplasm of unspecified olfactory nerve |
| ICD10CM | C72.21 | Malignant neoplasm of right olfactory nerve |
| ICD10CM | C72.22 | Malignant neoplasm of left olfactory nerve |
| ICD10CM | C72.3 | Malignant neoplasm of optic nerve |
| ICD10CM | C72.30 | Malignant neoplasm of unspecified optic nerve |
| ICD10CM | C72.31 | Malignant neoplasm of right optic nerve |
| ICD10CM | C72.32 | Malignant neoplasm of left optic nerve |
| ICD10CM | C72.4 | Malignant neoplasm of acoustic nerve |
| ICD10CM | C72.40 | Malignant neoplasm of unspecified acoustic nerve |
| ICD10CM | C72.41 | Malignant neoplasm of right acoustic nerve |
| ICD10CM | C72.42 | Malignant neoplasm of left acoustic nerve |
| ICD10CM | C72.5 | Malignant neoplasm of other and unspecified cranial nerves |
| ICD10CM | C72.50 | Malignant neoplasm of unspecified cranial nerve |
| ICD10CM | C72.59 | Malignant neoplasm of other cranial nerves |
| ICD10CM | C72.9 | Malignant neoplasm of central nervous system, unspecified |
| ICD10CM | C74.0 | Malignant neoplasm of cortex of adrenal gland |
| ICD10CM | C74.00 | Malignant neoplasm of cortex of unspecified adrenal gland |
| ICD10CM | C74.01 | Malignant neoplasm of cortex of right adrenal gland |
| ICD10CM | C74.02 | Malignant neoplasm of cortex of left adrenal gland |
| ICD10CM | C74.1 | Malignant neoplasm of medulla of adrenal gland |
| ICD10CM | C74.10 | Malignant neoplasm of medulla of unspecified adrenal gland |
| ICD10CM | C74.11 | Malignant neoplasm of medulla of right adrenal gland |
| ICD10CM | C74.12 | Malignant neoplasm of medulla of left adrenal gland |
| ICD10CM | C74.9 | Malignant neoplasm of unspecified part of adrenal gland |
| ICD10CM | C74.90 | Malignant neoplasm of unspecified part of unspecified adrenal gland |
| ICD10CM | C74.91 | Malignant neoplasm of unspecified part of right adrenal gland |
| ICD10CM | C74.92 | Malignant neoplasm of unspecified part of left adrenal gland |
| ICD10CM | C75.0 | Malignant neoplasm of parathyroid gland |
| ICD10CM | C75.1 | Malignant neoplasm of pituitary gland |
| ICD10CM | C75.2 | Malignant neoplasm of craniopharyngeal duct |
| ICD10CM | C75.3 | Malignant neoplasm of pineal gland |
| ICD10CM | C75.4 | Malignant neoplasm of carotid body |
| ICD10CM | C75.5 | Malignant neoplasm of aortic body and other paraganglia |
| ICD10CM | C75.8 | Malignant neoplasm with pluriglandular involvement, unspecified |
| ICD10CM | C75.9 | Malignant neoplasm of endocrine gland, unspecified |
| ICD10CM | C76.0 | Malignant neoplasm of head, face and neck |
| ICD10CM | C76.1 | Malignant neoplasm of thorax |
| ICD10CM | C76.2 | Malignant neoplasm of abdomen |
| ICD10CM | C76.3 | Malignant neoplasm of pelvis |
| ICD10CM | C76.4 | Malignant neoplasm of upper limb |
| ICD10CM | C76.40 | Malignant neoplasm of unspecified upper limb |
| ICD10CM | C76.41 | Malignant neoplasm of right upper limb |
| ICD10CM | C76.42 | Malignant neoplasm of left upper limb |

Continued on next page

**TableS31 – continued from previous page**

| CodeSystem | ConceptCode | ConceptName |
|---|---|---|
| ICD10CM | C76.5 | Malignant neoplasm of lower limb |
| ICD10CM | C76.50 | Malignant neoplasm of unspecified lower limb |
| ICD10CM | C76.51 | Malignant neoplasm of right lower limb |
| ICD10CM | C76.52 | Malignant neoplasm of left lower limb |
| ICD10CM | C76.8 | Malignant neoplasm of other specified ill-defined sites |
| ICD10CM | C77.0 | Secondary and unspecified malignant neoplasm of lymph nodes of head, face and neck |
| ICD10CM | C77.1 | Secondary and unspecified malignant neoplasm of intrathoracic lymph nodes |
| ICD10CM | C77.2 | Secondary and unspecified malignant neoplasm of intra-abdominal lymph nodes |
| ICD10CM | C77.3 | Secondary and unspecified malignant neoplasm of axilla and upper limb lymph nodes |
| ICD10CM | C77.4 | Secondary and unspecified malignant neoplasm of inguinal and lower limb lymph nodes |
| ICD10CM | C77.5 | Secondary and unspecified malignant neoplasm of intrapelvic lymph nodes |
| ICD10CM | C77.8 | Secondary and unspecified malignant neoplasm of lymph nodes of multiple regions |
| ICD10CM | C77.9 | Secondary and unspecified malignant neoplasm of lymph node, unspecified |
| ICD10CM | C78.0 | Secondary malignant neoplasm of lung |
| ICD10CM | C78.00 | Secondary malignant neoplasm of unspecified lung |
| ICD10CM | C78.01 | Secondary malignant neoplasm of right lung |
| ICD10CM | C78.02 | Secondary malignant neoplasm of left lung |
| ICD10CM | C78.1 | Secondary malignant neoplasm of mediastinum |
| ICD10CM | C78.2 | Secondary malignant neoplasm of pleura |
| ICD10CM | C78.3 | Secondary malignant neoplasm of other and unspecified respiratory organs |
| ICD10CM | C78.30 | Secondary malignant neoplasm of unspecified respiratory organ |
| ICD10CM | C78.39 | Secondary malignant neoplasm of other respiratory organs |
| ICD10CM | C78.4 | Secondary malignant neoplasm of small intestine |
| ICD10CM | C78.5 | Secondary malignant neoplasm of large intestine and rectum |
| ICD10CM | C78.6 | Secondary malignant neoplasm of retroperitoneum and peritoneum |
| ICD10CM | C78.7 | Secondary malignant neoplasm of liver and intrahepatic bile duct |
| ICD10CM | C78.8 | Secondary malignant neoplasm of other and unspecified digestive organs |
| ICD10CM | C78.80 | Secondary malignant neoplasm of unspecified digestive organ |
| ICD10CM | C78.89 | Secondary malignant neoplasm of other digestive organs |
| ICD10CM | C79.0 | Secondary malignant neoplasm of kidney and renal pelvis |
| ICD10CM | C79.00 | Secondary malignant neoplasm of unspecified kidney and renal pelvis |
| ICD10CM | C79.01 | Secondary malignant neoplasm of right kidney and renal pelvis |
| ICD10CM | C79.02 | Secondary malignant neoplasm of left kidney and renal pelvis |
| ICD10CM | C79.1 | Secondary malignant neoplasm of bladder and other and unspecified urinary organs |

Continued on next page

**TableS31 – continued from previous page**

| CodeSystem | ConceptCode | ConceptName |
|---|---|---|
| ICD10CM | C79.10 | Secondary malignant neoplasm of unspecified urinary organs |
| ICD10CM | C79.11 | Secondary malignant neoplasm of bladder |
| ICD10CM | C79.19 | Secondary malignant neoplasm of other urinary organs |
| ICD10CM | C79.2 | Secondary malignant neoplasm of skin |
| ICD10CM | C79.3 | Secondary malignant neoplasm of brain and cerebral meninges |
| ICD10CM | C79.31 | Secondary malignant neoplasm of brain |
| ICD10CM | C79.32 | Secondary malignant neoplasm of cerebral meninges |
| ICD10CM | C79.4 | Secondary malignant neoplasm of other and unspecified parts of nervous system |
| ICD10CM | C79.40 | Secondary malignant neoplasm of unspecified part of nervous system |
| ICD10CM | C79.49 | Secondary malignant neoplasm of other parts of nervous system |
| ICD10CM | C79.5 | Secondary malignant neoplasm of bone and bone marrow |
| ICD10CM | C79.51 | Secondary malignant neoplasm of bone |
| ICD10CM | C79.52 | Secondary malignant neoplasm of bone marrow |
| ICD10CM | C79.6 | Secondary malignant neoplasm of ovary |
| ICD10CM | C79.60 | Secondary malignant neoplasm of unspecified ovary |
| ICD10CM | C79.61 | Secondary malignant neoplasm of right ovary |
| ICD10CM | C79.62 | Secondary malignant neoplasm of left ovary |
| ICD10CM | C79.7 | Secondary malignant neoplasm of adrenal gland |
| ICD10CM | C79.70 | Secondary malignant neoplasm of unspecified adrenal gland |
| ICD10CM | C79.71 | Secondary malignant neoplasm of right adrenal gland |
| ICD10CM | C79.72 | Secondary malignant neoplasm of left adrenal gland |
| ICD10CM | C79.8 | Secondary malignant neoplasm of other specified sites |
| ICD10CM | C79.81 | Secondary malignant neoplasm of breast |
| ICD10CM | C79.82 | Secondary malignant neoplasm of genital organs |
| ICD10CM | C79.89 | Secondary malignant neoplasm of other specified sites |
| ICD10CM | C79.9 | Secondary malignant neoplasm of unspecified site |
| ICD10CM | C7A.0 | Malignant carcinoid tumors |
| ICD10CM | C7A.00 | Malignant carcinoid tumor of unspecified site |
| ICD10CM | C7A.01 | Malignant carcinoid tumors of the small intestine |
| ICD10CM | C7A.010 | Malignant carcinoid tumor of the duodenum |
| ICD10CM | C7A.011 | Malignant carcinoid tumor of the jejunum |
| ICD10CM | C7A.012 | Malignant carcinoid tumor of the ileum |
| ICD10CM | C7A.019 | Malignant carcinoid tumor of the small intestine, unspecified portion |
| ICD10CM | C7A.02 | Malignant carcinoid tumors of the appendix, large intestine, and rectum |
| ICD10CM | C7A.020 | Malignant carcinoid tumor of the appendix |
| ICD10CM | C7A.021 | Malignant carcinoid tumor of the cecum |
| ICD10CM | C7A.022 | Malignant carcinoid tumor of the ascending colon |
| ICD10CM | C7A.023 | Malignant carcinoid tumor of the transverse colon |
| ICD10CM | C7A.024 | Malignant carcinoid tumor of the descending colon |
| ICD10CM | C7A.025 | Malignant carcinoid tumor of the sigmoid colon |
| ICD10CM | C7A.026 | Malignant carcinoid tumor of the rectum |
| ICD10CM | C7A.029 | Malignant carcinoid tumor of the large intestine, unspecified portion |

Continued on next page

**TableS31 – continued from previous page**

| CodeSystem | ConceptCode | ConceptName |
|---|---|---|
| ICD10CM | C7A.09 | Malignant carcinoid tumors of other sites |
| ICD10CM | C7A.090 | Malignant carcinoid tumor of the bronchus and lung |
| ICD10CM | C7A.091 | Malignant carcinoid tumor of the thymus |
| ICD10CM | C7A.092 | Malignant carcinoid tumor of the stomach |
| ICD10CM | C7A.093 | Malignant carcinoid tumor of the kidney |
| ICD10CM | C7A.094 | Malignant carcinoid tumor of the foregut, unspecified |
| ICD10CM | C7A.095 | Malignant carcinoid tumor of the midgut, unspecified |
| ICD10CM | C7A.096 | Malignant carcinoid tumor of the hindgut, unspecified |
| ICD10CM | C7A.098 | Malignant carcinoid tumors of other sites |
| ICD10CM | C7A.1 | Malignant poorly differentiated neuroendocrine tumors |
| ICD10CM | C7A.8 | Other malignant neuroendocrine tumors |
| ICD10CM | C7B.0 | Secondary carcinoid tumors |
| ICD10CM | C7B.00 | Secondary carcinoid tumors, unspecified site |
| ICD10CM | C7B.01 | Secondary carcinoid tumors of distant lymph nodes |
| ICD10CM | C7B.02 | Secondary carcinoid tumors of liver |
| ICD10CM | C7B.03 | Secondary carcinoid tumors of bone |
| ICD10CM | C7B.04 | Secondary carcinoid tumors of peritoneum |
| ICD10CM | C7B.09 | Secondary carcinoid tumors of other sites |
| ICD10CM | C7B.1 | Secondary Merkel cell carcinoma |
| ICD10CM | C7B.8 | Other secondary neuroendocrine tumors |
| ICD10CM | C80.0 | Disseminated malignant neoplasm, unspecified |
| ICD10CM | C80.1 | Malignant (primary) neoplasm, unspecified |
| ICD10CM | C80.2 | Malignant neoplasm associated with transplanted organ |
| ICD10CM | C81.0 | Nodular lymphocyte predominant Hodgkin lymphoma |
| ICD10CM | C81.00 | Nodular lymphocyte predominant Hodgkin lymphoma, unspecified site |
| ICD10CM | C81.01 | Nodular lymphocyte predominant Hodgkin lymphoma, lymph nodes of head, face, and neck |
| ICD10CM | C81.02 | Nodular lymphocyte predominant Hodgkin lymphoma, intrathoracic lymph nodes |
| ICD10CM | C81.03 | Nodular lymphocyte predominant Hodgkin lymphoma, intra-abdominal lymph nodes |
| ICD10CM | C81.04 | Nodular lymphocyte predominant Hodgkin lymphoma, lymph nodes of axilla and upper limb |
| ICD10CM | C81.05 | Nodular lymphocyte predominant Hodgkin lymphoma, lymph nodes of inguinal region and lower limb |
| ICD10CM | C81.06 | Nodular lymphocyte predominant Hodgkin lymphoma, intrapelvic lymph nodes |
| ICD10CM | C81.07 | Nodular lymphocyte predominant Hodgkin lymphoma, spleen |
| ICD10CM | C81.08 | Nodular lymphocyte predominant Hodgkin lymphoma, lymph nodes of multiple sites |
| ICD10CM | C81.09 | Nodular lymphocyte predominant Hodgkin lymphoma, extranodal and solid organ sites |
| ICD10CM | C81.1 | Nodular sclerosis Hodgkin lymphoma |
| ICD10CM | C81.10 | Nodular sclerosis Hodgkin lymphoma, unspecified site |
| ICD10CM | C81.11 | Nodular sclerosis Hodgkin lymphoma, lymph nodes of head, face, and neck |

Continued on next page

**TableS31 – continued from previous page**

| CodeSystem | ConceptCode | ConceptName |
|---|---|---|
| ICD10CM | C81.12 | Nodular sclerosis Hodgkin lymphoma, intrathoracic lymph nodes |
| ICD10CM | C81.13 | Nodular sclerosis Hodgkin lymphoma, intra-abdominal lymph nodes |
| ICD10CM | C81.14 | Nodular sclerosis Hodgkin lymphoma, lymph nodes of axilla and upper limb |
| ICD10CM | C81.15 | Nodular sclerosis Hodgkin lymphoma, lymph nodes of inguinal region and lower limb |
| ICD10CM | C81.16 | Nodular sclerosis Hodgkin lymphoma, intrapelvic lymph nodes |
| ICD10CM | C81.17 | Nodular sclerosis Hodgkin lymphoma, spleen |
| ICD10CM | C81.18 | Nodular sclerosis Hodgkin lymphoma, lymph nodes of multiple sites |
| ICD10CM | C81.19 | Nodular sclerosis Hodgkin lymphoma, extranodal and solid organ sites |
| ICD10CM | C81.2 | Mixed cellularity Hodgkin lymphoma |
| ICD10CM | C81.20 | Mixed cellularity Hodgkin lymphoma, unspecified site |
| ICD10CM | C81.21 | Mixed cellularity Hodgkin lymphoma, lymph nodes of head, face, and neck |
| ICD10CM | C81.22 | Mixed cellularity Hodgkin lymphoma, intrathoracic lymph nodes |
| ICD10CM | C81.23 | Mixed cellularity Hodgkin lymphoma, intra-abdominal lymph nodes |
| ICD10CM | C81.24 | Mixed cellularity Hodgkin lymphoma, lymph nodes of axilla and upper limb |
| ICD10CM | C81.25 | Mixed cellularity Hodgkin lymphoma, lymph nodes of inguinal region and lower limb |
| ICD10CM | C81.26 | Mixed cellularity Hodgkin lymphoma, intrapelvic lymph nodes |
| ICD10CM | C81.27 | Mixed cellularity Hodgkin lymphoma, spleen |
| ICD10CM | C81.28 | Mixed cellularity Hodgkin lymphoma, lymph nodes of multiple sites |
| ICD10CM | C81.29 | Mixed cellularity Hodgkin lymphoma, extranodal and solid organ sites |
| ICD10CM | C81.3 | Lymphocyte depleted Hodgkin lymphoma |
| ICD10CM | C81.30 | Lymphocyte depleted Hodgkin lymphoma, unspecified site |
| ICD10CM | C81.31 | Lymphocyte depleted Hodgkin lymphoma, lymph nodes of head, face, and neck |
| ICD10CM | C81.32 | Lymphocyte depleted Hodgkin lymphoma, intrathoracic lymph nodes |
| ICD10CM | C81.33 | Lymphocyte depleted Hodgkin lymphoma, intra-abdominal lymph nodes |
| ICD10CM | C81.34 | Lymphocyte depleted Hodgkin lymphoma, lymph nodes of axilla and upper limb |
| ICD10CM | C81.35 | Lymphocyte depleted Hodgkin lymphoma, lymph nodes of inguinal region and lower limb |
| ICD10CM | C81.36 | Lymphocyte depleted Hodgkin lymphoma, intrapelvic lymph nodes |
| ICD10CM | C81.37 | Lymphocyte depleted Hodgkin lymphoma, spleen |
| ICD10CM | C81.38 | Lymphocyte depleted Hodgkin lymphoma, lymph nodes of multiple sites |

Continued on next page

**TableS31 – continued from previous page**

| CodeSystem | ConceptCode | ConceptName |
|---|---|---|
| ICD10CM | C81.39 | Lymphocyte depleted Hodgkin lymphoma, extranodal and solid organ sites |
| ICD10CM | C81.4 | Lymphocyte-rich Hodgkin lymphoma |
| ICD10CM | C81.40 | Lymphocyte-rich Hodgkin lymphoma, unspecified site |
| ICD10CM | C81.41 | Lymphocyte-rich Hodgkin lymphoma, lymph nodes of head, face, and neck |
| ICD10CM | C81.42 | Lymphocyte-rich Hodgkin lymphoma, intrathoracic lymph nodes |
| ICD10CM | C81.43 | Lymphocyte-rich Hodgkin lymphoma, intra-abdominal lymph nodes |
| ICD10CM | C81.44 | Lymphocyte-rich Hodgkin lymphoma, lymph nodes of axilla and upper limb |
| ICD10CM | C81.45 | Lymphocyte-rich Hodgkin lymphoma, lymph nodes of inguinal region and lower limb |
| ICD10CM | C81.46 | Lymphocyte-rich Hodgkin lymphoma, intrapelvic lymph nodes |
| ICD10CM | C81.47 | Lymphocyte-rich Hodgkin lymphoma, spleen |
| ICD10CM | C81.48 | Lymphocyte-rich Hodgkin lymphoma, lymph nodes of multiple sites |
| ICD10CM | C81.49 | Lymphocyte-rich Hodgkin lymphoma, extranodal and solid organ sites |
| ICD10CM | C81.7 | Other Hodgkin lymphoma |
| ICD10CM | C81.70 | Other Hodgkin lymphoma, unspecified site |
| ICD10CM | C81.71 | Other Hodgkin lymphoma, lymph nodes of head, face, and neck |
| ICD10CM | C81.72 | Other Hodgkin lymphoma, intrathoracic lymph nodes |
| ICD10CM | C81.73 | Other Hodgkin lymphoma, intra-abdominal lymph nodes |
| ICD10CM | C81.74 | Other Hodgkin lymphoma, lymph nodes of axilla and upper limb |
| ICD10CM | C81.75 | Other Hodgkin lymphoma, lymph nodes of inguinal region and lower limb |
| ICD10CM | C81.76 | Other Hodgkin lymphoma, intrapelvic lymph nodes |
| ICD10CM | C81.77 | Other Hodgkin lymphoma, spleen |
| ICD10CM | C81.78 | Other Hodgkin lymphoma, lymph nodes of multiple sites |
| ICD10CM | C81.79 | Other Hodgkin lymphoma, extranodal and solid organ sites |
| ICD10CM | C81.9 | Hodgkin lymphoma, unspecified |
| ICD10CM | C81.90 | Hodgkin lymphoma, unspecified, unspecified site |
| ICD10CM | C81.91 | Hodgkin lymphoma, unspecified, lymph nodes of head, face, and neck |
| ICD10CM | C81.92 | Hodgkin lymphoma, unspecified, intrathoracic lymph nodes |
| ICD10CM | C81.93 | Hodgkin lymphoma, unspecified, intra-abdominal lymph nodes |
| ICD10CM | C81.94 | Hodgkin lymphoma, unspecified, lymph nodes of axilla and upper limb |
| ICD10CM | C81.95 | Hodgkin lymphoma, unspecified, lymph nodes of inguinal region and lower limb |
| ICD10CM | C81.96 | Hodgkin lymphoma, unspecified, intrapelvic lymph nodes |
| ICD10CM | C81.97 | Hodgkin lymphoma, unspecified, spleen |
| ICD10CM | C81.98 | Hodgkin lymphoma, unspecified, lymph nodes of multiple sites |
| ICD10CM | C81.99 | Hodgkin lymphoma, unspecified, extranodal and solid organ sites |
| ICD10CM | C82.0 | Follicular lymphoma grade I |

Continued on next page

**TableS31 – continued from previous page**

| CodeSystem | ConceptCode | ConceptName |
|---|---|---|
| ICD10CM | C82.00 | Follicular lymphoma grade I, unspecified site |
| ICD10CM | C82.01 | Follicular lymphoma grade I, lymph nodes of head, face, and neck |
| ICD10CM | C82.02 | Follicular lymphoma grade I, intrathoracic lymph nodes |
| ICD10CM | C82.03 | Follicular lymphoma grade I, intra-abdominal lymph nodes |
| ICD10CM | C82.04 | Follicular lymphoma grade I, lymph nodes of axilla and upper limb |
| ICD10CM | C82.05 | Follicular lymphoma grade I, lymph nodes of inguinal region and lower limb |
| ICD10CM | C82.06 | Follicular lymphoma grade I, intrapelvic lymph nodes |
| ICD10CM | C82.07 | Follicular lymphoma grade I, spleen |
| ICD10CM | C82.08 | Follicular lymphoma grade I, lymph nodes of multiple sites |
| ICD10CM | C82.09 | Follicular lymphoma grade I, extranodal and solid organ sites |
| ICD10CM | C82.1 | Follicular lymphoma grade II |
| ICD10CM | C82.10 | Follicular lymphoma grade II, unspecified site |
| ICD10CM | C82.11 | Follicular lymphoma grade II, lymph nodes of head, face, and neck |
| ICD10CM | C82.12 | Follicular lymphoma grade II, intrathoracic lymph nodes |
| ICD10CM | C82.13 | Follicular lymphoma grade II, intra-abdominal lymph nodes |
| ICD10CM | C82.14 | Follicular lymphoma grade II, lymph nodes of axilla and upper limb |
| ICD10CM | C82.15 | Follicular lymphoma grade II, lymph nodes of inguinal region and lower limb |
| ICD10CM | C82.16 | Follicular lymphoma grade II, intrapelvic lymph nodes |
| ICD10CM | C82.17 | Follicular lymphoma grade II, spleen |
| ICD10CM | C82.18 | Follicular lymphoma grade II, lymph nodes of multiple sites |
| ICD10CM | C82.19 | Follicular lymphoma grade II, extranodal and solid organ sites |
| ICD10CM | C82.2 | Follicular lymphoma grade III, unspecified |
| ICD10CM | C82.20 | Follicular lymphoma grade III, unspecified, unspecified site |
| ICD10CM | C82.21 | Follicular lymphoma grade III, unspecified, lymph nodes of head, face, and neck |
| ICD10CM | C82.22 | Follicular lymphoma grade III, unspecified, intrathoracic lymph nodes |
| ICD10CM | C82.23 | Follicular lymphoma grade III, unspecified, intra-abdominal lymph nodes |
| ICD10CM | C82.24 | Follicular lymphoma grade III, unspecified, lymph nodes of axilla and upper limb |
| ICD10CM | C82.25 | Follicular lymphoma grade III, unspecified, lymph nodes of inguinal region and lower limb |
| ICD10CM | C82.26 | Follicular lymphoma grade III, unspecified, intrapelvic lymph nodes |
| ICD10CM | C82.27 | Follicular lymphoma grade III, unspecified, spleen |
| ICD10CM | C82.28 | Follicular lymphoma grade III, unspecified, lymph nodes of multiple sites |
| ICD10CM | C82.29 | Follicular lymphoma grade III, unspecified, extranodal and solid organ sites |
| ICD10CM | C82.3 | Follicular lymphoma grade IIIa |

Continued on next page

**TableS31 – continued from previous page**

| CodeSystem | ConceptCode | ConceptName |
|---|---|---|
| ICD10CM | C82.30 | Follicular lymphoma grade IIIa, unspecified site |
| ICD10CM | C82.31 | Follicular lymphoma grade IIIa, lymph nodes of head, face, and neck |
| ICD10CM | C82.32 | Follicular lymphoma grade IIIa, intrathoracic lymph nodes |
| ICD10CM | C82.33 | Follicular lymphoma grade IIIa, intra-abdominal lymph nodes |
| ICD10CM | C82.34 | Follicular lymphoma grade IIIa, lymph nodes of axilla and upper limb |
| ICD10CM | C82.35 | Follicular lymphoma grade IIIa, lymph nodes of inguinal region and lower limb |
| ICD10CM | C82.36 | Follicular lymphoma grade IIIa, intrapelvic lymph nodes |
| ICD10CM | C82.37 | Follicular lymphoma grade IIIa, spleen |
| ICD10CM | C82.38 | Follicular lymphoma grade IIIa, lymph nodes of multiple sites |
| ICD10CM | C82.39 | Follicular lymphoma grade IIIa, extranodal and solid organ sites |
| ICD10CM | C82.4 | Follicular lymphoma grade IIIb |
| ICD10CM | C82.40 | Follicular lymphoma grade IIIb, unspecified site |
| ICD10CM | C82.41 | Follicular lymphoma grade IIIb, lymph nodes of head, face, and neck |
| ICD10CM | C82.42 | Follicular lymphoma grade IIIb, intrathoracic lymph nodes |
| ICD10CM | C82.43 | Follicular lymphoma grade IIIb, intra-abdominal lymph nodes |
| ICD10CM | C82.44 | Follicular lymphoma grade IIIb, lymph nodes of axilla and upper limb |
| ICD10CM | C82.45 | Follicular lymphoma grade IIIb, lymph nodes of inguinal region and lower limb |
| ICD10CM | C82.46 | Follicular lymphoma grade IIIb, intrapelvic lymph nodes |
| ICD10CM | C82.47 | Follicular lymphoma grade IIIb, spleen |
| ICD10CM | C82.48 | Follicular lymphoma grade IIIb, lymph nodes of multiple sites |
| ICD10CM | C82.49 | Follicular lymphoma grade IIIb, extranodal and solid organ sites |
| ICD10CM | C82.5 | Diffuse follicle center lymphoma |
| ICD10CM | C82.50 | Diffuse follicle center lymphoma, unspecified site |
| ICD10CM | C82.51 | Diffuse follicle center lymphoma, lymph nodes of head, face, and neck |
| ICD10CM | C82.52 | Diffuse follicle center lymphoma, intrathoracic lymph nodes |
| ICD10CM | C82.53 | Diffuse follicle center lymphoma, intra-abdominal lymph nodes |
| ICD10CM | C82.54 | Diffuse follicle center lymphoma, lymph nodes of axilla and upper limb |
| ICD10CM | C82.55 | Diffuse follicle center lymphoma, lymph nodes of inguinal region and lower limb |
| ICD10CM | C82.56 | Diffuse follicle center lymphoma, intrapelvic lymph nodes |
| ICD10CM | C82.57 | Diffuse follicle center lymphoma, spleen |
| ICD10CM | C82.58 | Diffuse follicle center lymphoma, lymph nodes of multiple sites |
| ICD10CM | C82.59 | Diffuse follicle center lymphoma, extranodal and solid organ sites |
| ICD10CM | C82.6 | Cutaneous follicle center lymphoma |
| ICD10CM | C82.60 | Cutaneous follicle center lymphoma, unspecified site |
| ICD10CM | C82.61 | Cutaneous follicle center lymphoma, lymph nodes of head, face, and neck |
| ICD10CM | C82.62 | Cutaneous follicle center lymphoma, intrathoracic lymph nodes |

Continued on next page

**TableS31 – continued from previous page**

| CodeSystem | ConceptCode | ConceptName |
|---|---|---|
| ICD10CM | C82.63 | Cutaneous follicle center lymphoma, intra-abdominal lymph nodes |
| ICD10CM | C82.64 | Cutaneous follicle center lymphoma, lymph nodes of axilla and upper limb |
| ICD10CM | C82.65 | Cutaneous follicle center lymphoma, lymph nodes of inguinal region and lower limb |
| ICD10CM | C82.66 | Cutaneous follicle center lymphoma, intrapelvic lymph nodes |
| ICD10CM | C82.67 | Cutaneous follicle center lymphoma, spleen |
| ICD10CM | C82.68 | Cutaneous follicle center lymphoma, lymph nodes of multiple sites |
| ICD10CM | C82.69 | Cutaneous follicle center lymphoma, extranodal and solid organ sites |
| ICD10CM | C82.8 | Other types of follicular lymphoma |
| ICD10CM | C82.80 | Other types of follicular lymphoma, unspecified site |
| ICD10CM | C82.81 | Other types of follicular lymphoma, lymph nodes of head, face, and neck |
| ICD10CM | C82.82 | Other types of follicular lymphoma, intrathoracic lymph nodes |
| ICD10CM | C82.83 | Other types of follicular lymphoma, intra-abdominal lymph nodes |
| ICD10CM | C82.84 | Other types of follicular lymphoma, lymph nodes of axilla and upper limb |
| ICD10CM | C82.85 | Other types of follicular lymphoma, lymph nodes of inguinal region and lower limb |
| ICD10CM | C82.86 | Other types of follicular lymphoma, intrapelvic lymph nodes |
| ICD10CM | C82.87 | Other types of follicular lymphoma, spleen |
| ICD10CM | C82.88 | Other types of follicular lymphoma, lymph nodes of multiple sites |
| ICD10CM | C82.89 | Other types of follicular lymphoma, extranodal and solid organ sites |
| ICD10CM | C82.9 | Follicular lymphoma, unspecified |
| ICD10CM | C82.90 | Follicular lymphoma, unspecified, unspecified site |
| ICD10CM | C82.91 | Follicular lymphoma, unspecified, lymph nodes of head, face, and neck |
| ICD10CM | C82.92 | Follicular lymphoma, unspecified, intrathoracic lymph nodes |
| ICD10CM | C82.93 | Follicular lymphoma, unspecified, intra-abdominal lymph nodes |
| ICD10CM | C82.94 | Follicular lymphoma, unspecified, lymph nodes of axilla and upper limb |
| ICD10CM | C82.95 | Follicular lymphoma, unspecified, lymph nodes of inguinal region and lower limb |
| ICD10CM | C82.96 | Follicular lymphoma, unspecified, intrapelvic lymph nodes |
| ICD10CM | C82.97 | Follicular lymphoma, unspecified, spleen |
| ICD10CM | C82.98 | Follicular lymphoma, unspecified, lymph nodes of multiple sites |
| ICD10CM | C82.99 | Follicular lymphoma, unspecified, extranodal and solid organ sites |
| ICD10CM | C83.0 | Small cell B-cell lymphoma |
| ICD10CM | C83.00 | Small cell B-cell lymphoma, unspecified site |
| ICD10CM | C83.01 | Small cell B-cell lymphoma, lymph nodes of head, face, and neck |

Continued on next page

**TableS31 – continued from previous page**

| CodeSystem | ConceptCode | ConceptName |
|---|---|---|
| ICD10CM | C83.02 | Small cell B-cell lymphoma, intrathoracic lymph nodes |
| ICD10CM | C83.03 | Small cell B-cell lymphoma, intra-abdominal lymph nodes |
| ICD10CM | C83.04 | Small cell B-cell lymphoma, lymph nodes of axilla and upper limb |
| ICD10CM | C83.05 | Small cell B-cell lymphoma, lymph nodes of inguinal region and lower limb |
| ICD10CM | C83.06 | Small cell B-cell lymphoma, intrapelvic lymph nodes |
| ICD10CM | C83.07 | Small cell B-cell lymphoma, spleen |
| ICD10CM | C83.08 | Small cell B-cell lymphoma, lymph nodes of multiple sites |
| ICD10CM | C83.09 | Small cell B-cell lymphoma, extranodal and solid organ sites |
| ICD10CM | C83.1 | Mantle cell lymphoma |
| ICD10CM | C83.10 | Mantle cell lymphoma, unspecified site |
| ICD10CM | C83.11 | Mantle cell lymphoma, lymph nodes of head, face, and neck |
| ICD10CM | C83.12 | Mantle cell lymphoma, intrathoracic lymph nodes |
| ICD10CM | C83.13 | Mantle cell lymphoma, intra-abdominal lymph nodes |
| ICD10CM | C83.14 | Mantle cell lymphoma, lymph nodes of axilla and upper limb |
| ICD10CM | C83.15 | Mantle cell lymphoma, lymph nodes of inguinal region and lower limb |
| ICD10CM | C83.16 | Mantle cell lymphoma, intrapelvic lymph nodes |
| ICD10CM | C83.17 | Mantle cell lymphoma, spleen |
| ICD10CM | C83.18 | Mantle cell lymphoma, lymph nodes of multiple sites |
| ICD10CM | C83.19 | Mantle cell lymphoma, extranodal and solid organ sites |
| ICD10CM | C83.3 | Diffuse large B-cell lymphoma |
| ICD10CM | C83.30 | Diffuse large B-cell lymphoma, unspecified site |
| ICD10CM | C83.31 | Diffuse large B-cell lymphoma, lymph nodes of head, face, and neck |
| ICD10CM | C83.32 | Diffuse large B-cell lymphoma, intrathoracic lymph nodes |
| ICD10CM | C83.33 | Diffuse large B-cell lymphoma, intra-abdominal lymph nodes |
| ICD10CM | C83.34 | Diffuse large B-cell lymphoma, lymph nodes of axilla and upper limb |
| ICD10CM | C83.35 | Diffuse large B-cell lymphoma, lymph nodes of inguinal region and lower limb |
| ICD10CM | C83.36 | Diffuse large B-cell lymphoma, intrapelvic lymph nodes |
| ICD10CM | C83.37 | Diffuse large B-cell lymphoma, spleen |
| ICD10CM | C83.38 | Diffuse large B-cell lymphoma, lymph nodes of multiple sites |
| ICD10CM | C83.39 | Diffuse large B-cell lymphoma, extranodal and solid organ sites |
| ICD10CM | C83.5 | Lymphoblastic (diffuse) lymphoma |
| ICD10CM | C83.50 | Lymphoblastic (diffuse) lymphoma, unspecified site |
| ICD10CM | C83.51 | Lymphoblastic (diffuse) lymphoma, lymph nodes of head, face, and neck |
| ICD10CM | C83.52 | Lymphoblastic (diffuse) lymphoma, intrathoracic lymph nodes |
| ICD10CM | C83.53 | Lymphoblastic (diffuse) lymphoma, intra-abdominal lymph nodes |
| ICD10CM | C83.54 | Lymphoblastic (diffuse) lymphoma, lymph nodes of axilla and upper limb |
| ICD10CM | C83.55 | Lymphoblastic (diffuse) lymphoma, lymph nodes of inguinal region and lower limb |

Continued on next page

**TableS31 – continued from previous page**

| CodeSystem | ConceptCode | ConceptName |
|---|---|---|
| ICD10CM | C83.56 | Lymphoblastic (diffuse) lymphoma, intrapelvic lymph nodes |
| ICD10CM | C83.57 | Lymphoblastic (diffuse) lymphoma, spleen |
| ICD10CM | C83.58 | Lymphoblastic (diffuse) lymphoma, lymph nodes of multiple sites |
| ICD10CM | C83.59 | Lymphoblastic (diffuse) lymphoma, extranodal and solid organ sites |
| ICD10CM | C83.7 | Burkitt lymphoma |
| ICD10CM | C83.70 | Burkitt lymphoma, unspecified site |
| ICD10CM | C83.71 | Burkitt lymphoma, lymph nodes of head, face, and neck |
| ICD10CM | C83.72 | Burkitt lymphoma, intrathoracic lymph nodes |
| ICD10CM | C83.73 | Burkitt lymphoma, intra-abdominal lymph nodes |
| ICD10CM | C83.74 | Burkitt lymphoma, lymph nodes of axilla and upper limb |
| ICD10CM | C83.75 | Burkitt lymphoma, lymph nodes of inguinal region and lower limb |
| ICD10CM | C83.76 | Burkitt lymphoma, intrapelvic lymph nodes |
| ICD10CM | C83.77 | Burkitt lymphoma, spleen |
| ICD10CM | C83.78 | Burkitt lymphoma, lymph nodes of multiple sites |
| ICD10CM | C83.79 | Burkitt lymphoma, extranodal and solid organ sites |
| ICD10CM | C83.8 | Other non-follicular lymphoma |
| ICD10CM | C83.80 | Other non-follicular lymphoma, unspecified site |
| ICD10CM | C83.81 | Other non-follicular lymphoma, lymph nodes of head, face, and neck |
| ICD10CM | C83.82 | Other non-follicular lymphoma, intrathoracic lymph nodes |
| ICD10CM | C83.83 | Other non-follicular lymphoma, intra-abdominal lymph nodes |
| ICD10CM | C83.84 | Other non-follicular lymphoma, lymph nodes of axilla and upper limb |
| ICD10CM | C83.85 | Other non-follicular lymphoma, lymph nodes of inguinal region and lower limb |
| ICD10CM | C83.86 | Other non-follicular lymphoma, intrapelvic lymph nodes |
| ICD10CM | C83.87 | Other non-follicular lymphoma, spleen |
| ICD10CM | C83.88 | Other non-follicular lymphoma, lymph nodes of multiple sites |
| ICD10CM | C83.89 | Other non-follicular lymphoma, extranodal and solid organ sites |
| ICD10CM | C83.9 | Non-follicular (diffuse) lymphoma, unspecified |
| ICD10CM | C83.90 | Non-follicular (diffuse) lymphoma, unspecified, unspecified site |
| ICD10CM | C83.91 | Non-follicular (diffuse) lymphoma, unspecified, lymph nodes of head, face, and neck |
| ICD10CM | C83.92 | Non-follicular (diffuse) lymphoma, unspecified, intrathoracic lymph nodes |
| ICD10CM | C83.93 | Non-follicular (diffuse) lymphoma, unspecified, intra-abdominal lymph nodes |
| ICD10CM | C83.94 | Non-follicular (diffuse) lymphoma, unspecified, lymph nodes of axilla and upper limb |
| ICD10CM | C83.95 | Non-follicular (diffuse) lymphoma, unspecified, lymph nodes of inguinal region and lower limb |
| ICD10CM | C83.96 | Non-follicular (diffuse) lymphoma, unspecified, intrapelvic lymph nodes |
| ICD10CM | C83.97 | Non-follicular (diffuse) lymphoma, unspecified, spleen |

Continued on next page

**TableS31 – continued from previous page**

| CodeSystem | ConceptCode | ConceptName |
|---|---|---|
| ICD10CM | C83.98 | Non-follicular (diffuse) lymphoma, unspecified, lymph nodes of multiple sites |
| ICD10CM | C83.99 | Non-follicular (diffuse) lymphoma, unspecified, extranodal and solid organ sites |
| ICD10CM | C84.0 | Mycosis fungoides |
| ICD10CM | C84.00 | Mycosis fungoides, unspecified site |
| ICD10CM | C84.01 | Mycosis fungoides, lymph nodes of head, face, and neck |
| ICD10CM | C84.02 | Mycosis fungoides, intrathoracic lymph nodes |
| ICD10CM | C84.03 | Mycosis fungoides, intra-abdominal lymph nodes |
| ICD10CM | C84.04 | Mycosis fungoides, lymph nodes of axilla and upper limb |
| ICD10CM | C84.05 | Mycosis fungoides, lymph nodes of inguinal region and lower limb |
| ICD10CM | C84.06 | Mycosis fungoides, intrapelvic lymph nodes |
| ICD10CM | C84.07 | Mycosis fungoides, spleen |
| ICD10CM | C84.08 | Mycosis fungoides, lymph nodes of multiple sites |
| ICD10CM | C84.09 | Mycosis fungoides, extranodal and solid organ sites |
| ICD10CM | C84.1 | Sezary disease |
| ICD10CM | C84.10 | Sezary disease, unspecified site |
| ICD10CM | C84.11 | Sezary disease, lymph nodes of head, face, and neck |
| ICD10CM | C84.12 | Sezary disease, intrathoracic lymph nodes |
| ICD10CM | C84.13 | Sezary disease, intra-abdominal lymph nodes |
| ICD10CM | C84.14 | Sezary disease, lymph nodes of axilla and upper limb |
| ICD10CM | C84.15 | Sezary disease, lymph nodes of inguinal region and lower limb |
| ICD10CM | C84.16 | Sezary disease, intrapelvic lymph nodes |
| ICD10CM | C84.17 | Sezary disease, spleen |
| ICD10CM | C84.18 | Sezary disease, lymph nodes of multiple sites |
| ICD10CM | C84.19 | Sezary disease, extranodal and solid organ sites |
| ICD10CM | C84.4 | Peripheral T-cell lymphoma, not classified |
| ICD10CM | C84.40 | Peripheral T-cell lymphoma, not classified, unspecified site |
| ICD10CM | C84.41 | Peripheral T-cell lymphoma, not classified, lymph nodes of head, face, and neck |
| ICD10CM | C84.42 | Peripheral T-cell lymphoma, not classified, intrathoracic lymph nodes |
| ICD10CM | C84.43 | Peripheral T-cell lymphoma, not classified, intra-abdominal lymph nodes |
| ICD10CM | C84.44 | Peripheral T-cell lymphoma, not classified, lymph nodes of axilla and upper limb |
| ICD10CM | C84.45 | Peripheral T-cell lymphoma, not classified, lymph nodes of inguinal region and lower limb |
| ICD10CM | C84.46 | Peripheral T-cell lymphoma, not classified, intrapelvic lymph nodes |
| ICD10CM | C84.47 | Peripheral T-cell lymphoma, not classified, spleen |
| ICD10CM | C84.48 | Peripheral T-cell lymphoma, not classified, lymph nodes of multiple sites |
| ICD10CM | C84.49 | Peripheral T-cell lymphoma, not classified, extranodal and solid organ sites |
| ICD10CM | C84.6 | Anaplastic large cell lymphoma, ALK-positive |
| ICD10CM | C84.60 | Anaplastic large cell lymphoma, ALK-positive, unspecified site |

Continued on next page

**TableS31 – continued from previous page**

| CodeSystem | ConceptCode | ConceptName |
|---|---|---|
| ICD10CM | C84.61 | Anaplastic large cell lymphoma, ALK-positive, lymph nodes of head, face, and neck |
| ICD10CM | C84.62 | Anaplastic large cell lymphoma, ALK-positive, intrathoracic lymph nodes |
| ICD10CM | C84.63 | Anaplastic large cell lymphoma, ALK-positive, intra-abdominal lymph nodes |
| ICD10CM | C84.64 | Anaplastic large cell lymphoma, ALK-positive, lymph nodes of axilla and upper limb |
| ICD10CM | C84.65 | Anaplastic large cell lymphoma, ALK-positive, lymph nodes of inguinal region and lower limb |
| ICD10CM | C84.66 | Anaplastic large cell lymphoma, ALK-positive, intrapelvic lymph nodes |
| ICD10CM | C84.67 | Anaplastic large cell lymphoma, ALK-positive, spleen |
| ICD10CM | C84.68 | Anaplastic large cell lymphoma, ALK-positive, lymph nodes of multiple sites |
| ICD10CM | C84.69 | Anaplastic large cell lymphoma, ALK-positive, extranodal and solid organ sites |
| ICD10CM | C84.7 | Anaplastic large cell lymphoma, ALK-negative |
| ICD10CM | C84.70 | Anaplastic large cell lymphoma, ALK-negative, unspecified site |
| ICD10CM | C84.71 | Anaplastic large cell lymphoma, ALK-negative, lymph nodes of head, face, and neck |
| ICD10CM | C84.72 | Anaplastic large cell lymphoma, ALK-negative, intrathoracic lymph nodes |
| ICD10CM | C84.73 | Anaplastic large cell lymphoma, ALK-negative, intra-abdominal lymph nodes |
| ICD10CM | C84.74 | Anaplastic large cell lymphoma, ALK-negative, lymph nodes of axilla and upper limb |
| ICD10CM | C84.75 | Anaplastic large cell lymphoma, ALK-negative, lymph nodes of inguinal region and lower limb |
| ICD10CM | C84.76 | Anaplastic large cell lymphoma, ALK-negative, intrapelvic lymph nodes |
| ICD10CM | C84.77 | Anaplastic large cell lymphoma, ALK-negative, spleen |
| ICD10CM | C84.78 | Anaplastic large cell lymphoma, ALK-negative, lymph nodes of multiple sites |
| ICD10CM | C84.79 | Anaplastic large cell lymphoma, ALK-negative, extranodal and solid organ sites |
| ICD10CM | C84.9 | Mature T/NK-cell lymphomas, unspecified |
| ICD10CM | C84.90 | Mature T/NK-cell lymphomas, unspecified, unspecified site |
| ICD10CM | C84.91 | Mature T/NK-cell lymphomas, unspecified, lymph nodes of head, face, and neck |
| ICD10CM | C84.92 | Mature T/NK-cell lymphomas, unspecified, intrathoracic lymph nodes |
| ICD10CM | C84.93 | Mature T/NK-cell lymphomas, unspecified, intra-abdominal lymph nodes |
| ICD10CM | C84.94 | Mature T/NK-cell lymphomas, unspecified, lymph nodes of axilla and upper limb |

Continued on next page

**TableS31 – continued from previous page**

| CodeSystem | ConceptCode | ConceptName |
|---|---|---|
| ICD10CM | C84.95 | Mature T/NK-cell lymphomas, unspecified, lymph nodes of inguinal region and lower limb |
| ICD10CM | C84.96 | Mature T/NK-cell lymphomas, unspecified, intrapelvic lymph nodes |
| ICD10CM | C84.97 | Mature T/NK-cell lymphomas, unspecified, spleen |
| ICD10CM | C84.98 | Mature T/NK-cell lymphomas, unspecified, lymph nodes of multiple sites |
| ICD10CM | C84.99 | Mature T/NK-cell lymphomas, unspecified, extranodal and solid organ sites |
| ICD10CM | C84.A | Cutaneous T-cell lymphoma, unspecified |
| ICD10CM | C84.A0 | Cutaneous T-cell lymphoma, unspecified, unspecified site |
| ICD10CM | C84.A1 | Cutaneous T-cell lymphoma, unspecified lymph nodes of head, face, and neck |
| ICD10CM | C84.A2 | Cutaneous T-cell lymphoma, unspecified, intrathoracic lymph nodes |
| ICD10CM | C84.A3 | Cutaneous T-cell lymphoma, unspecified, intra-abdominal lymph nodes |
| ICD10CM | C84.A4 | Cutaneous T-cell lymphoma, unspecified, lymph nodes of axilla and upper limb |
| ICD10CM | C84.A5 | Cutaneous T-cell lymphoma, unspecified, lymph nodes of inguinal region and lower limb |
| ICD10CM | C84.A6 | Cutaneous T-cell lymphoma, unspecified, intrapelvic lymph nodes |
| ICD10CM | C84.A7 | Cutaneous T-cell lymphoma, unspecified, spleen |
| ICD10CM | C84.A8 | Cutaneous T-cell lymphoma, unspecified, lymph nodes of multiple sites |
| ICD10CM | C84.A9 | Cutaneous T-cell lymphoma, unspecified, extranodal and solid organ sites |
| ICD10CM | C84.Z | Other mature T/NK-cell lymphomas |
| ICD10CM | C84.Z0 | Other mature T/NK-cell lymphomas, unspecified site |
| ICD10CM | C84.Z1 | Other mature T/NK-cell lymphomas, lymph nodes of head, face, and neck |
| ICD10CM | C84.Z2 | Other mature T/NK-cell lymphomas, intrathoracic lymph nodes |
| ICD10CM | C84.Z3 | Other mature T/NK-cell lymphomas, intra-abdominal lymph nodes |
| ICD10CM | C84.Z4 | Other mature T/NK-cell lymphomas, lymph nodes of axilla and upper limb |
| ICD10CM | C84.Z5 | Other mature T/NK-cell lymphomas, lymph nodes of inguinal region and lower limb |
| ICD10CM | C84.Z6 | Other mature T/NK-cell lymphomas, intrapelvic lymph nodes |
| ICD10CM | C84.Z7 | Other mature T/NK-cell lymphomas, spleen |
| ICD10CM | C84.Z8 | Other mature T/NK-cell lymphomas, lymph nodes of multiple sites |
| ICD10CM | C84.Z9 | Other mature T/NK-cell lymphomas, extranodal and solid organ sites |
| ICD10CM | C85.1 | Unspecified B-cell lymphoma |
| ICD10CM | C85.10 | Unspecified B-cell lymphoma, unspecified site |

Continued on next page

**TableS31 – continued from previous page**

| CodeSystem | ConceptCode | ConceptName |
|---|---|---|
| ICD10CM | C85.11 | Unspecified B-cell lymphoma, lymph nodes of head, face, and neck |
| ICD10CM | C85.12 | Unspecified B-cell lymphoma, intrathoracic lymph nodes |
| ICD10CM | C85.13 | Unspecified B-cell lymphoma, intra-abdominal lymph nodes |
| ICD10CM | C85.14 | Unspecified B-cell lymphoma, lymph nodes of axilla and upper limb |
| ICD10CM | C85.15 | Unspecified B-cell lymphoma, lymph nodes of inguinal region and lower limb |
| ICD10CM | C85.16 | Unspecified B-cell lymphoma, intrapelvic lymph nodes |
| ICD10CM | C85.17 | Unspecified B-cell lymphoma, spleen |
| ICD10CM | C85.18 | Unspecified B-cell lymphoma, lymph nodes of multiple sites |
| ICD10CM | C85.19 | Unspecified B-cell lymphoma, extranodal and solid organ sites |
| ICD10CM | C85.2 | Mediastinal (thymic) large B-cell lymphoma |
| ICD10CM | C85.20 | Mediastinal (thymic) large B-cell lymphoma, unspecified site |
| ICD10CM | C85.21 | Mediastinal (thymic) large B-cell lymphoma, lymph nodes of head, face, and neck |
| ICD10CM | C85.22 | Mediastinal (thymic) large B-cell lymphoma, intrathoracic lymph nodes |
| ICD10CM | C85.23 | Mediastinal (thymic) large B-cell lymphoma, intra-abdominal lymph nodes |
| ICD10CM | C85.24 | Mediastinal (thymic) large B-cell lymphoma, lymph nodes of axilla and upper limb |
| ICD10CM | C85.25 | Mediastinal (thymic) large B-cell lymphoma, lymph nodes of inguinal region and lower limb |
| ICD10CM | C85.26 | Mediastinal (thymic) large B-cell lymphoma, intrapelvic lymph nodes |
| ICD10CM | C85.27 | Mediastinal (thymic) large B-cell lymphoma, spleen |
| ICD10CM | C85.28 | Mediastinal (thymic) large B-cell lymphoma, lymph nodes of multiple sites |
| ICD10CM | C85.29 | Mediastinal (thymic) large B-cell lymphoma, extranodal and solid organ sites |
| ICD10CM | C85.8 | Other specified types of non-Hodgkin lymphoma |
| ICD10CM | C85.80 | Other specified types of non-Hodgkin lymphoma, unspecified site |
| ICD10CM | C85.81 | Other specified types of non-Hodgkin lymphoma, lymph nodes of head, face, and neck |
| ICD10CM | C85.82 | Other specified types of non-Hodgkin lymphoma, intrathoracic lymph nodes |
| ICD10CM | C85.83 | Other specified types of non-Hodgkin lymphoma, intra-abdominal lymph nodes |
| ICD10CM | C85.84 | Other specified types of non-Hodgkin lymphoma, lymph nodes of axilla and upper limb |
| ICD10CM | C85.85 | Other specified types of non-Hodgkin lymphoma, lymph nodes of inguinal region and lower limb |
| ICD10CM | C85.86 | Other specified types of non-Hodgkin lymphoma, intrapelvic lymph nodes |
| ICD10CM | C85.87 | Other specified types of non-Hodgkin lymphoma, spleen |

Continued on next page

**TableS31 – continued from previous page**

| CodeSystem | ConceptCode | ConceptName |
|---|---|---|
| ICD10CM | C85.88 | Other specified types of non-Hodgkin lymphoma, lymph nodes of multiple sites |
| ICD10CM | C85.89 | Other specified types of non-Hodgkin lymphoma, extranodal and solid organ sites |
| ICD10CM | C85.9 | Non-Hodgkin lymphoma, unspecified |
| ICD10CM | C85.90 | Non-Hodgkin lymphoma, unspecified, unspecified site |
| ICD10CM | C85.91 | Non-Hodgkin lymphoma, unspecified, lymph nodes of head, face, and neck |
| ICD10CM | C85.92 | Non-Hodgkin lymphoma, unspecified, intrathoracic lymph nodes |
| ICD10CM | C85.93 | Non-Hodgkin lymphoma, unspecified, intra-abdominal lymph nodes |
| ICD10CM | C85.94 | Non-Hodgkin lymphoma, unspecified, lymph nodes of axilla and upper limb |
| ICD10CM | C85.95 | Non-Hodgkin lymphoma, unspecified, lymph nodes of inguinal region and lower limb |
| ICD10CM | C85.96 | Non-Hodgkin lymphoma, unspecified, intrapelvic lymph nodes |
| ICD10CM | C85.97 | Non-Hodgkin lymphoma, unspecified, spleen |
| ICD10CM | C85.98 | Non-Hodgkin lymphoma, unspecified, lymph nodes of multiple sites |
| ICD10CM | C85.99 | Non-Hodgkin lymphoma, unspecified, extranodal and solid organ sites |
| ICD10CM | C86.0 | Extranodal NK/T-cell lymphoma, nasal type |
| ICD10CM | C86.1 | Hepatosplenic T-cell lymphoma |
| ICD10CM | C86.2 | Enteropathy-type (intestinal) T-cell lymphoma |
| ICD10CM | C86.3 | Subcutaneous panniculitis-like T-cell lymphoma |
| ICD10CM | C86.4 | Blastic NK-cell lymphoma |
| ICD10CM | C86.5 | Angioimmunoblastic T-cell lymphoma |
| ICD10CM | C86.6 | Primary cutaneous CD30-positive T-cell proliferations |
| ICD10CM | C88.0 | Waldenstrom macroglobulinemia |
| ICD10CM | C88.2 | Heavy chain disease |
| ICD10CM | C88.3 | Immunoproliferative small intestinal disease |
| ICD10CM | C88.4 | Extranodal marginal zone B-cell lymphoma of mucosa-associated lymphoid tissue [MALT-lymphoma] |
| ICD10CM | C88.8 | Other malignant immunoproliferative diseases |
| ICD10CM | C88.9 | Malignant immunoproliferative disease, unspecified |
| ICD10CM | C90.0 | Multiple myeloma |
| ICD10CM | C90.00 | Multiple myeloma not having achieved remission |
| ICD10CM | C90.01 | Multiple myeloma in remission |
| ICD10CM | C90.02 | Multiple myeloma in relapse |
| ICD10CM | C90.1 | Plasma cell leukemia |
| ICD10CM | C90.10 | Plasma cell leukemia not having achieved remission |
| ICD10CM | C90.11 | Plasma cell leukemia in remission |
| ICD10CM | C90.12 | Plasma cell leukemia in relapse |
| ICD10CM | C90.2 | Extramedullary plasmacytoma |
| ICD10CM | C90.20 | Extramedullary plasmacytoma not having achieved remission |
| ICD10CM | C90.21 | Extramedullary plasmacytoma in remission |
| ICD10CM | C90.22 | Extramedullary plasmacytoma in relapse |

Continued on next page

**TableS31 – continued from previous page**

| CodeSystem | ConceptCode | ConceptName |
|---|---|---|
| ICD10CM | C90.3 | Solitary plasmacytoma |
| ICD10CM | C90.30 | Solitary plasmacytoma not having achieved remission |
| ICD10CM | C90.31 | Solitary plasmacytoma in remission |
| ICD10CM | C90.32 | Solitary plasmacytoma in relapse |
| ICD10CM | C91.0 | Acute lymphoblastic leukemia [ALL] |
| ICD10CM | C91.00 | Acute lymphoblastic leukemia not having achieved remission |
| ICD10CM | C91.01 | Acute lymphoblastic leukemia, in remission |
| ICD10CM | C91.02 | Acute lymphoblastic leukemia, in relapse |
| ICD10CM | C91.1 | Chronic lymphocytic leukemia of B-cell type |
| ICD10CM | C91.10 | Chronic lymphocytic leukemia of B-cell type not having achieved remission |
| ICD10CM | C91.11 | Chronic lymphocytic leukemia of B-cell type in remission |
| ICD10CM | C91.12 | Chronic lymphocytic leukemia of B-cell type in relapse |
| ICD10CM | C91.3 | Prolymphocytic leukemia of B-cell type |
| ICD10CM | C91.30 | Prolymphocytic leukemia of B-cell type not having achieved remission |
| ICD10CM | C91.31 | Prolymphocytic leukemia of B-cell type, in remission |
| ICD10CM | C91.32 | Prolymphocytic leukemia of B-cell type, in relapse |
| ICD10CM | C91.4 | Hairy cell leukemia |
| ICD10CM | C91.40 | Hairy cell leukemia not having achieved remission |
| ICD10CM | C91.41 | Hairy cell leukemia, in remission |
| ICD10CM | C91.42 | Hairy cell leukemia, in relapse |
| ICD10CM | C91.5 | Adult T-cell lymphoma/leukemia (HTLV-1-associated) |
| ICD10CM | C91.50 | Adult T-cell lymphoma/leukemia (HTLV-1-associated) not having achieved remission |
| ICD10CM | C91.51 | Adult T-cell lymphoma/leukemia (HTLV-1-associated), in remission |
| ICD10CM | C91.52 | Adult T-cell lymphoma/leukemia (HTLV-1-associated), in relapse |
| ICD10CM | C91.6 | Prolymphocytic leukemia of T-cell type |
| ICD10CM | C91.60 | Prolymphocytic leukemia of T-cell type not having achieved remission |
| ICD10CM | C91.61 | Prolymphocytic leukemia of T-cell type, in remission |
| ICD10CM | C91.62 | Prolymphocytic leukemia of T-cell type, in relapse |
| ICD10CM | C91.9 | Lymphoid leukemia, unspecified |
| ICD10CM | C91.90 | Lymphoid leukemia, unspecified not having achieved remission |
| ICD10CM | C91.91 | Lymphoid leukemia, unspecified, in remission |
| ICD10CM | C91.92 | Lymphoid leukemia, unspecified, in relapse |
| ICD10CM | C91.A | Mature B-cell leukemia Burkitt-type |
| ICD10CM | C91.A0 | Mature B-cell leukemia Burkitt-type not having achieved remission |
| ICD10CM | C91.A1 | Mature B-cell leukemia Burkitt-type, in remission |
| ICD10CM | C91.A2 | Mature B-cell leukemia Burkitt-type, in relapse |
| ICD10CM | C91.Z | Other lymphoid leukemia |
| ICD10CM | C91.Z0 | Other lymphoid leukemia not having achieved remission |
| ICD10CM | C91.Z1 | Other lymphoid leukemia, in remission |
| ICD10CM | C91.Z2 | Other lymphoid leukemia, in relapse |

Continued on next page

**TableS31 – continued from previous page**

| CodeSystem | ConceptCode | ConceptName |
|---|---|---|
| ICD10CM | C92.0 | Acute myeloblastic leukemia |
| ICD10CM | C92.00 | Acute myeloblastic leukemia, not having achieved remission |
| ICD10CM | C92.01 | Acute myeloblastic leukemia, in remission |
| ICD10CM | C92.02 | Acute myeloblastic leukemia, in relapse |
| ICD10CM | C92.1 | Chronic myeloid leukemia, BCR/ABL-positive |
| ICD10CM | C92.10 | Chronic myeloid leukemia, BCR/ABL-positive, not having achieved remission |
| ICD10CM | C92.11 | Chronic myeloid leukemia, BCR/ABL-positive, in remission |
| ICD10CM | C92.12 | Chronic myeloid leukemia, BCR/ABL-positive, in relapse |
| ICD10CM | C92.2 | Atypical chronic myeloid leukemia, BCR/ABL-negative |
| ICD10CM | C92.20 | Atypical chronic myeloid leukemia, BCR/ABL-negative, not having achieved remission |
| ICD10CM | C92.21 | Atypical chronic myeloid leukemia, BCR/ABL-negative, in remission |
| ICD10CM | C92.22 | Atypical chronic myeloid leukemia, BCR/ABL-negative, in relapse |
| ICD10CM | C92.3 | Myeloid sarcoma |
| ICD10CM | C92.30 | Myeloid sarcoma, not having achieved remission |
| ICD10CM | C92.31 | Myeloid sarcoma, in remission |
| ICD10CM | C92.32 | Myeloid sarcoma, in relapse |
| ICD10CM | C92.4 | Acute promyelocytic leukemia |
| ICD10CM | C92.40 | Acute promyelocytic leukemia, not having achieved remission |
| ICD10CM | C92.41 | Acute promyelocytic leukemia, in remission |
| ICD10CM | C92.42 | Acute promyelocytic leukemia, in relapse |
| ICD10CM | C92.5 | Acute myelomonocytic leukemia |
| ICD10CM | C92.50 | Acute myelomonocytic leukemia, not having achieved remission |
| ICD10CM | C92.51 | Acute myelomonocytic leukemia, in remission |
| ICD10CM | C92.52 | Acute myelomonocytic leukemia, in relapse |
| ICD10CM | C92.6 | Acute myeloid leukemia with 11q23-abnormality |
| ICD10CM | C92.60 | Acute myeloid leukemia with 11q23-abnormality not having achieved remission |
| ICD10CM | C92.61 | Acute myeloid leukemia with 11q23-abnormality in remission |
| ICD10CM | C92.62 | Acute myeloid leukemia with 11q23-abnormality in relapse |
| ICD10CM | C92.9 | Myeloid leukemia, unspecified |
| ICD10CM | C92.90 | Myeloid leukemia, unspecified, not having achieved remission |
| ICD10CM | C92.91 | Myeloid leukemia, unspecified in remission |
| ICD10CM | C92.92 | Myeloid leukemia, unspecified in relapse |
| ICD10CM | C92.A | Acute myeloid leukemia with multilineage dysplasia |
| ICD10CM | C92.A0 | Acute myeloid leukemia with multilineage dysplasia, not having achieved remission |
| ICD10CM | C92.A1 | Acute myeloid leukemia with multilineage dysplasia, in remission |
| ICD10CM | C92.A2 | Acute myeloid leukemia with multilineage dysplasia, in relapse |
| ICD10CM | C92.Z | Other myeloid leukemia |
| ICD10CM | C92.Z0 | Other myeloid leukemia not having achieved remission |
| ICD10CM | C92.Z1 | Other myeloid leukemia, in remission |
| ICD10CM | C92.Z2 | Other myeloid leukemia, in relapse |
| ICD10CM | C93.0 | Acute monoblastic/monocytic leukemia |

Continued on next page

**TableS31 – continued from previous page**

| CodeSystem | ConceptCode | ConceptName |
|---|---|---|
| ICD10CM | C93.00 | Acute monoblastic/monocytic leukemia, not having achieved remission |
| ICD10CM | C93.01 | Acute monoblastic/monocytic leukemia, in remission |
| ICD10CM | C93.02 | Acute monoblastic/monocytic leukemia, in relapse |
| ICD10CM | C93.1 | Chronic myelomonocytic leukemia |
| ICD10CM | C93.10 | Chronic myelomonocytic leukemia not having achieved remission |
| ICD10CM | C93.11 | Chronic myelomonocytic leukemia, in remission |
| ICD10CM | C93.12 | Chronic myelomonocytic leukemia, in relapse |
| ICD10CM | C93.3 | Juvenile myelomonocytic leukemia |
| ICD10CM | C93.30 | Juvenile myelomonocytic leukemia, not having achieved remission |
| ICD10CM | C93.31 | Juvenile myelomonocytic leukemia, in remission |
| ICD10CM | C93.32 | Juvenile myelomonocytic leukemia, in relapse |
| ICD10CM | C93.9 | Monocytic leukemia, unspecified |
| ICD10CM | C93.90 | Monocytic leukemia, unspecified, not having achieved remission |
| ICD10CM | C93.91 | Monocytic leukemia, unspecified in remission |
| ICD10CM | C93.92 | Monocytic leukemia, unspecified in relapse |
| ICD10CM | C93.Z | Other monocytic leukemia |
| ICD10CM | C93.Z0 | Other monocytic leukemia, not having achieved remission |
| ICD10CM | C93.Z1 | Other monocytic leukemia, in remission |
| ICD10CM | C93.Z2 | Other monocytic leukemia, in relapse |
| ICD10CM | C94.0 | Acute erythroid leukemia |
| ICD10CM | C94.00 | Acute erythroid leukemia, not having achieved remission |
| ICD10CM | C94.01 | Acute erythroid leukemia, in remission |
| ICD10CM | C94.02 | Acute erythroid leukemia, in relapse |
| ICD10CM | C94.2 | Acute megakaryoblastic leukemia |
| ICD10CM | C94.20 | Acute megakaryoblastic leukemia not having achieved remission |
| ICD10CM | C94.21 | Acute megakaryoblastic leukemia, in remission |
| ICD10CM | C94.22 | Acute megakaryoblastic leukemia, in relapse |
| ICD10CM | C94.3 | Mast cell leukemia |
| ICD10CM | C94.30 | Mast cell leukemia not having achieved remission |
| ICD10CM | C94.31 | Mast cell leukemia, in remission |
| ICD10CM | C94.32 | Mast cell leukemia, in relapse |
| ICD10CM | C94.4 | Acute panmyelosis with myelofibrosis |
| ICD10CM | C94.40 | Acute panmyelosis with myelofibrosis not having achieved remission |
| ICD10CM | C94.41 | Acute panmyelosis with myelofibrosis, in remission |
| ICD10CM | C94.42 | Acute panmyelosis with myelofibrosis, in relapse |
| ICD10CM | C94.6 | Myelodysplastic disease, not classified |
| ICD10CM | C94.8 | Other specified leukemias |
| ICD10CM | C94.80 | Other specified leukemias not having achieved remission |
| ICD10CM | C94.81 | Other specified leukemias, in remission |
| ICD10CM | C94.82 | Other specified leukemias, in relapse |
| ICD10CM | C95.0 | Acute leukemia of unspecified cell type |
| ICD10CM | C95.00 | Acute leukemia of unspecified cell type not having achieved remission |
| ICD10CM | C95.01 | Acute leukemia of unspecified cell type, in remission |

Continued on next page

**TableS31 – continued from previous page**

| CodeSystem | ConceptCode | ConceptName |
|---|---|---|
| ICD10CM | C95.02 | Acute leukemia of unspecified cell type, in relapse |
| ICD10CM | C95.1 | Chronic leukemia of unspecified cell type |
| ICD10CM | C95.10 | Chronic leukemia of unspecified cell type not having achieved remission |
| ICD10CM | C95.11 | Chronic leukemia of unspecified cell type, in remission |
| ICD10CM | C95.12 | Chronic leukemia of unspecified cell type, in relapse |
| ICD10CM | C95.9 | Leukemia, unspecified |
| ICD10CM | C95.90 | Leukemia, unspecified not having achieved remission |
| ICD10CM | C95.91 | Leukemia, unspecified, in remission |
| ICD10CM | C95.92 | Leukemia, unspecified, in relapse |
| ICD10CM | C96.0 | Multifocal and multisystemic (disseminated) Langerhans-cell histiocytosis |
| ICD10CM | C96.2 | Malignant mast cell neoplasm |
| ICD10CM | C96.20 | Malignant mast cell neoplasm, unspecified |
| ICD10CM | C96.21 | Aggressive systemic mastocytosis |
| ICD10CM | C96.22 | Mast cell sarcoma |
| ICD10CM | C96.29 | Other malignant mast cell neoplasm |
| ICD10CM | C96.4 | Sarcoma of dendritic cells (accessory cells) |
| ICD10CM | C96.5 | Multifocal and unisystemic Langerhans-cell histiocytosis |
| ICD10CM | C96.6 | Unifocal Langerhans-cell histiocytosis |
| ICD10CM | C96.9 | Malignant neoplasm of lymphoid, hematopoietic and related tissue, unspecified |
| ICD10CM | C96.A | Histiocytic sarcoma |
| ICD10CM | C96.Z | Other specified malignant neoplasms of lymphoid, hematopoietic and related tissue |
| ICD10CM | D00.0 | Carcinoma in situ of lip, oral cavity and pharynx |
| ICD10CM | D00.00 | Carcinoma in situ of oral cavity, unspecified site |
| ICD10CM | D00.01 | Carcinoma in situ of labial mucosa and vermilion border |
| ICD10CM | D00.02 | Carcinoma in situ of buccal mucosa |
| ICD10CM | D00.03 | Carcinoma in situ of gingiva and edentulous alveolar ridge |
| ICD10CM | D00.04 | Carcinoma in situ of soft palate |
| ICD10CM | D00.05 | Carcinoma in situ of hard palate |
| ICD10CM | D00.06 | Carcinoma in situ of floor of mouth |
| ICD10CM | D00.07 | Carcinoma in situ of tongue |
| ICD10CM | D00.08 | Carcinoma in situ of pharynx |
| ICD10CM | D00.1 | Carcinoma in situ of esophagus |
| ICD10CM | D00.2 | Carcinoma in situ of stomach |
| ICD10CM | D01.0 | Carcinoma in situ of colon |
| ICD10CM | D01.1 | Carcinoma in situ of rectosigmoid junction |
| ICD10CM | D01.2 | Carcinoma in situ of rectum |
| ICD10CM | D01.3 | Carcinoma in situ of anus and anal canal |
| ICD10CM | D01.4 | Carcinoma in situ of other and unspecified parts of intestine |
| ICD10CM | D01.40 | Carcinoma in situ of unspecified part of intestine |
| ICD10CM | D01.49 | Carcinoma in situ of other parts of intestine |
| ICD10CM | D01.5 | Carcinoma in situ of liver, gallbladder and bile ducts |
| ICD10CM | D01.7 | Carcinoma in situ of other specified digestive organs |
| ICD10CM | D01.9 | Carcinoma in situ of digestive organ, unspecified |

Continued on next page

**TableS31 – continued from previous page**

| CodeSystem | ConceptCode | ConceptName |
|---|---|---|
| ICD10CM | D02.0 | Carcinoma in situ of larynx |
| ICD10CM | D02.1 | Carcinoma in situ of trachea |
| ICD10CM | D02.2 | Carcinoma in situ of bronchus and lung |
| ICD10CM | D02.20 | Carcinoma in situ of unspecified bronchus and lung |
| ICD10CM | D02.21 | Carcinoma in situ of right bronchus and lung |
| ICD10CM | D02.22 | Carcinoma in situ of left bronchus and lung |
| ICD10CM | D02.3 | Carcinoma in situ of other parts of respiratory system |
| ICD10CM | D02.4 | Carcinoma in situ of respiratory system, unspecified |
| ICD10CM | D03.0 | Melanoma in situ of lip |
| ICD10CM | D03.1 | Melanoma in situ of eyelid, including canthus |
| ICD10CM | D03.10 | Melanoma in situ of unspecified eyelid, including canthus |
| ICD10CM | D03.11 | Melanoma in situ of right eyelid, including canthus |
| ICD10CM | D03.111 | Melanoma in situ of right upper eyelid, including canthus |
| ICD10CM | D03.112 | Melanoma in situ of right lower eyelid, including canthus |
| ICD10CM | D03.12 | Melanoma in situ of left eyelid, including canthus |
| ICD10CM | D03.121 | Melanoma in situ of left upper eyelid, including canthus |
| ICD10CM | D03.122 | Melanoma in situ of left lower eyelid, including canthus |
| ICD10CM | D03.2 | Melanoma in situ of ear and external auricular canal |
| ICD10CM | D03.20 | Melanoma in situ of unspecified ear and external auricular canal |
| ICD10CM | D03.21 | Melanoma in situ of right ear and external auricular canal |
| ICD10CM | D03.22 | Melanoma in situ of left ear and external auricular canal |
| ICD10CM | D03.3 | Melanoma in situ of other and unspecified parts of face |
| ICD10CM | D03.30 | Melanoma in situ of unspecified part of face |
| ICD10CM | D03.39 | Melanoma in situ of other parts of face |
| ICD10CM | D03.4 | Melanoma in situ of scalp and neck |
| ICD10CM | D03.5 | Melanoma in situ of trunk |
| ICD10CM | D03.51 | Melanoma in situ of anal skin |
| ICD10CM | D03.52 | Melanoma in situ of breast (skin) (soft tissue) |
| ICD10CM | D03.59 | Melanoma in situ of other part of trunk |
| ICD10CM | D03.6 | Melanoma in situ of upper limb, including shoulder |
| ICD10CM | D03.60 | Melanoma in situ of unspecified upper limb, including shoulder |
| ICD10CM | D03.61 | Melanoma in situ of right upper limb, including shoulder |
| ICD10CM | D03.62 | Melanoma in situ of left upper limb, including shoulder |
| ICD10CM | D03.7 | Melanoma in situ of lower limb, including hip |
| ICD10CM | D03.70 | Melanoma in situ of unspecified lower limb, including hip |
| ICD10CM | D03.71 | Melanoma in situ of right lower limb, including hip |
| ICD10CM | D03.72 | Melanoma in situ of left lower limb, including hip |
| ICD10CM | D03.8 | Melanoma in situ of other sites |
| ICD10CM | D03.9 | Melanoma in situ, unspecified |
| ICD10CM | D04.0 | Carcinoma in situ of skin of lip |
| ICD10CM | D04.1 | Carcinoma in situ of skin of eyelid, including canthus |
| ICD10CM | D04.10 | Carcinoma in situ of skin of unspecified eyelid, including canthus |
| ICD10CM | D04.11 | Carcinoma in situ of skin of right eyelid, including canthus |
| ICD10CM | D04.111 | Carcinoma in situ of skin of right upper eyelid, including canthus |
| ICD10CM | D04.112 | Carcinoma in situ of skin of right lower eyelid, including canthus |
| ICD10CM | D04.12 | Carcinoma in situ of skin of left eyelid, including canthus |
| ICD10CM | D04.121 | Carcinoma in situ of skin of left upper eyelid, including canthus |

Continued on next page

**TableS31 – continued from previous page**

| CodeSystem | ConceptCode | ConceptName |
|---|---|---|
| ICD10CM | D04.122 | Carcinoma in situ of skin of left lower eyelid, including canthus |
| ICD10CM | D04.2 | Carcinoma in situ of skin of ear and external auricular canal |
| ICD10CM | D04.20 | Carcinoma in situ of skin of unspecified ear and external auricular canal |
| ICD10CM | D04.21 | Carcinoma in situ of skin of right ear and external auricular canal |
| ICD10CM | D04.22 | Carcinoma in situ of skin of left ear and external auricular canal |
| ICD10CM | D04.3 | Carcinoma in situ of skin of other and unspecified parts of face |
| ICD10CM | D04.30 | Carcinoma in situ of skin of unspecified part of face |
| ICD10CM | D04.39 | Carcinoma in situ of skin of other parts of face |
| ICD10CM | D04.4 | Carcinoma in situ of skin of scalp and neck |
| ICD10CM | D04.5 | Carcinoma in situ of skin of trunk |
| ICD10CM | D04.6 | Carcinoma in situ of skin of upper limb, including shoulder |
| ICD10CM | D04.60 | Carcinoma in situ of skin of unspecified upper limb, including shoulder |
| ICD10CM | D04.61 | Carcinoma in situ of skin of right upper limb, including shoulder |
| ICD10CM | D04.62 | Carcinoma in situ of skin of left upper limb, including shoulder |
| ICD10CM | D04.7 | Carcinoma in situ of skin of lower limb, including hip |
| ICD10CM | D04.70 | Carcinoma in situ of skin of unspecified lower limb, including hip |
| ICD10CM | D04.71 | Carcinoma in situ of skin of right lower limb, including hip |
| ICD10CM | D04.72 | Carcinoma in situ of skin of left lower limb, including hip |
| ICD10CM | D04.8 | Carcinoma in situ of skin of other sites |
| ICD10CM | D04.9 | Carcinoma in situ of skin, unspecified |
| ICD10CM | D05.0 | Lobular carcinoma in situ of breast |
| ICD10CM | D05.00 | Lobular carcinoma in situ of unspecified breast |
| ICD10CM | D05.01 | Lobular carcinoma in situ of right breast |
| ICD10CM | D05.02 | Lobular carcinoma in situ of left breast |
| ICD10CM | D05.1 | Intraductal carcinoma in situ of breast |
| ICD10CM | D05.10 | Intraductal carcinoma in situ of unspecified breast |
| ICD10CM | D05.11 | Intraductal carcinoma in situ of right breast |
| ICD10CM | D05.12 | Intraductal carcinoma in situ of left breast |
| ICD10CM | D05.8 | Other specified type of carcinoma in situ of breast |
| ICD10CM | D05.80 | Other specified type of carcinoma in situ of unspecified breast |
| ICD10CM | D05.81 | Other specified type of carcinoma in situ of right breast |
| ICD10CM | D05.82 | Other specified type of carcinoma in situ of left breast |
| ICD10CM | D05.9 | Unspecified type of carcinoma in situ of breast |
| ICD10CM | D05.90 | Unspecified type of carcinoma in situ of unspecified breast |
| ICD10CM | D05.91 | Unspecified type of carcinoma in situ of right breast |
| ICD10CM | D05.92 | Unspecified type of carcinoma in situ of left breast |
| ICD10CM | D06.0 | Carcinoma in situ of endocervix |
| ICD10CM | D06.1 | Carcinoma in situ of exocervix |
| ICD10CM | D06.7 | Carcinoma in situ of other parts of cervix |
| ICD10CM | D06.9 | Carcinoma in situ of cervix, unspecified |
| ICD10CM | D07.0 | Carcinoma in situ of endometrium |
| ICD10CM | D07.1 | Carcinoma in situ of vulva |
| ICD10CM | D07.2 | Carcinoma in situ of vagina |
| ICD10CM | D07.3 | Carcinoma in situ of other and unspecified female genital organs |
| ICD10CM | D07.30 | Carcinoma in situ of unspecified female genital organs |

Continued on next page

**TableS31 – continued from previous page**

| CodeSystem | ConceptCode | ConceptName |
|---|---|---|
| ICD10CM | D07.39 | Carcinoma in situ of other female genital organs |
| ICD10CM | D07.4 | Carcinoma in situ of penis |
| ICD10CM | D07.5 | Carcinoma in situ of prostate |
| ICD10CM | D07.6 | Carcinoma in situ of other and unspecified male genital organs |
| ICD10CM | D07.60 | Carcinoma in situ of unspecified male genital organs |
| ICD10CM | D07.61 | Carcinoma in situ of scrotum |
| ICD10CM | D07.69 | Carcinoma in situ of other male genital organs |
| ICD10CM | D09.0 | Carcinoma in situ of bladder |
| ICD10CM | D09.1 | Carcinoma in situ of other and unspecified urinary organs |
| ICD10CM | D09.10 | Carcinoma in situ of unspecified urinary organ |
| ICD10CM | D09.19 | Carcinoma in situ of other urinary organs |
| ICD10CM | D09.2 | Carcinoma in situ of eye |
| ICD10CM | D09.20 | Carcinoma in situ of unspecified eye |
| ICD10CM | D09.21 | Carcinoma in situ of right eye |
| ICD10CM | D09.22 | Carcinoma in situ of left eye |
| ICD10CM | D09.3 | Carcinoma in situ of thyroid and other endocrine glands |
| ICD10CM | D09.8 | Carcinoma in situ of other specified sites |
| ICD10CM | D09.9 | Carcinoma in situ, unspecified |
| ICD10CM | D37.0 | Neoplasm of uncertain behavior of lip, oral cavity and pharynx |
| ICD10CM | D37.01 | Neoplasm of uncertain behavior of lip |
| ICD10CM | D37.02 | Neoplasm of uncertain behavior of tongue |
| ICD10CM | D37.03 | Neoplasm of uncertain behavior of the major salivary glands |
| ICD10CM | D37.030 | Neoplasm of uncertain behavior of the parotid salivary glands |
| ICD10CM | D37.031 | Neoplasm of uncertain behavior of the sublingual salivary glands |
| ICD10CM | D37.032 | Neoplasm of uncertain behavior of the submandibular salivary glands |
| ICD10CM | D37.039 | Neoplasm of uncertain behavior of the major salivary glands, unspecified |
| ICD10CM | D37.04 | Neoplasm of uncertain behavior of the minor salivary glands |
| ICD10CM | D37.05 | Neoplasm of uncertain behavior of pharynx |
| ICD10CM | D37.09 | Neoplasm of uncertain behavior of other specified sites of the oral cavity |
| ICD10CM | D37.1 | Neoplasm of uncertain behavior of stomach |
| ICD10CM | D37.2 | Neoplasm of uncertain behavior of small intestine |
| ICD10CM | D37.3 | Neoplasm of uncertain behavior of appendix |
| ICD10CM | D37.4 | Neoplasm of uncertain behavior of colon |
| ICD10CM | D37.5 | Neoplasm of uncertain behavior of rectum |
| ICD10CM | D37.6 | Neoplasm of uncertain behavior of liver, gallbladder and bile ducts |
| ICD10CM | D37.8 | Neoplasm of uncertain behavior of other specified digestive organs |
| ICD10CM | D37.9 | Neoplasm of uncertain behavior of digestive organ, unspecified |
| ICD10CM | D38.0 | Neoplasm of uncertain behavior of larynx |
| ICD10CM | D38.1 | Neoplasm of uncertain behavior of trachea, bronchus and lung |
| ICD10CM | D38.2 | Neoplasm of uncertain behavior of pleura |
| ICD10CM | D38.3 | Neoplasm of uncertain behavior of mediastinum |
| ICD10CM | D38.4 | Neoplasm of uncertain behavior of thymus |

Continued on next page

**TableS31 – continued from previous page**

| CodeSystem | ConceptCode | ConceptName |
|---|---|---|
| ICD10CM | D38.5 | Neoplasm of uncertain behavior of other respiratory organs |
| ICD10CM | D38.6 | Neoplasm of uncertain behavior of respiratory organ, unspecified |
| ICD10CM | D39.0 | Neoplasm of uncertain behavior of uterus |
| ICD10CM | D39.1 | Neoplasm of uncertain behavior of ovary |
| ICD10CM | D39.10 | Neoplasm of uncertain behavior of unspecified ovary |
| ICD10CM | D39.11 | Neoplasm of uncertain behavior of right ovary |
| ICD10CM | D39.12 | Neoplasm of uncertain behavior of left ovary |
| ICD10CM | D39.2 | Neoplasm of uncertain behavior of placenta |
| ICD10CM | D39.8 | Neoplasm of uncertain behavior of other specified female genital organs |
| ICD10CM | D39.9 | Neoplasm of uncertain behavior of female genital organ, unspecified |
| ICD10CM | D40.0 | Neoplasm of uncertain behavior of prostate |
| ICD10CM | D40.1 | Neoplasm of uncertain behavior of testis |
| ICD10CM | D40.10 | Neoplasm of uncertain behavior of unspecified testis |
| ICD10CM | D40.11 | Neoplasm of uncertain behavior of right testis |
| ICD10CM | D40.12 | Neoplasm of uncertain behavior of left testis |
| ICD10CM | D40.8 | Neoplasm of uncertain behavior of other specified male genital organs |
| ICD10CM | D40.9 | Neoplasm of uncertain behavior of male genital organ, unspecified |
| ICD10CM | D41.0 | Neoplasm of uncertain behavior of kidney |
| ICD10CM | D41.00 | Neoplasm of uncertain behavior of unspecified kidney |
| ICD10CM | D41.01 | Neoplasm of uncertain behavior of right kidney |
| ICD10CM | D41.02 | Neoplasm of uncertain behavior of left kidney |
| ICD10CM | D41.1 | Neoplasm of uncertain behavior of renal pelvis |
| ICD10CM | D41.10 | Neoplasm of uncertain behavior of unspecified renal pelvis |
| ICD10CM | D41.11 | Neoplasm of uncertain behavior of right renal pelvis |
| ICD10CM | D41.12 | Neoplasm of uncertain behavior of left renal pelvis |
| ICD10CM | D41.2 | Neoplasm of uncertain behavior of ureter |
| ICD10CM | D41.20 | Neoplasm of uncertain behavior of unspecified ureter |
| ICD10CM | D41.21 | Neoplasm of uncertain behavior of right ureter |
| ICD10CM | D41.22 | Neoplasm of uncertain behavior of left ureter |
| ICD10CM | D41.3 | Neoplasm of uncertain behavior of urethra |
| ICD10CM | D41.4 | Neoplasm of uncertain behavior of bladder |
| ICD10CM | D41.8 | Neoplasm of uncertain behavior of other specified urinary organs |
| ICD10CM | D41.9 | Neoplasm of uncertain behavior of unspecified urinary organ |
| ICD10CM | D42.0 | Neoplasm of uncertain behavior of cerebral meninges |
| ICD10CM | D42.1 | Neoplasm of uncertain behavior of spinal meninges |
| ICD10CM | D42.9 | Neoplasm of uncertain behavior of meninges, unspecified |
| ICD10CM | D43.0 | Neoplasm of uncertain behavior of brain, supratentorial |
| ICD10CM | D43.1 | Neoplasm of uncertain behavior of brain, infratentorial |
| ICD10CM | D43.2 | Neoplasm of uncertain behavior of brain, unspecified |
| ICD10CM | D43.3 | Neoplasm of uncertain behavior of cranial nerves |
| ICD10CM | D43.4 | Neoplasm of uncertain behavior of spinal cord |
| ICD10CM | D43.8 | Neoplasm of uncertain behavior of other specified parts of central nervous system |

Continued on next page

**TableS31 – continued from previous page**

| CodeSystem | ConceptCode | ConceptName |
|---|---|---|
| ICD10CM | D43.9 | Neoplasm of uncertain behavior of central nervous system, unspecified |
| ICD10CM | D44.0 | Neoplasm of uncertain behavior of thyroid gland |
| ICD10CM | D44.1 | Neoplasm of uncertain behavior of adrenal gland |
| ICD10CM | D44.10 | Neoplasm of uncertain behavior of unspecified adrenal gland |
| ICD10CM | D44.11 | Neoplasm of uncertain behavior of right adrenal gland |
| ICD10CM | D44.12 | Neoplasm of uncertain behavior of left adrenal gland |
| ICD10CM | D44.2 | Neoplasm of uncertain behavior of parathyroid gland |
| ICD10CM | D44.3 | Neoplasm of uncertain behavior of pituitary gland |
| ICD10CM | D44.4 | Neoplasm of uncertain behavior of craniopharyngeal duct |
| ICD10CM | D44.5 | Neoplasm of uncertain behavior of pineal gland |
| ICD10CM | D44.6 | Neoplasm of uncertain behavior of carotid body |
| ICD10CM | D44.7 | Neoplasm of uncertain behavior of aortic body and other paraganglia |
| ICD10CM | D44.9 | Neoplasm of uncertain behavior of unspecified endocrine gland |
| ICD10CM | D80.0 | Hereditary hypogammaglobulinemia |
| ICD10CM | D80.1 | Nonfamilial hypogammaglobulinemia |
| ICD10CM | D80.2 | Selective deficiency of immunoglobulin A [IgA] |
| ICD10CM | D80.3 | Selective deficiency of immunoglobulin G [IgG] subclasses |
| ICD10CM | D80.4 | Selective deficiency of immunoglobulin M [IgM] |
| ICD10CM | D80.5 | Immunodeficiency with increased immunoglobulin M [IgM] |
| ICD10CM | D80.6 | Antibody deficiency with near-normal immunoglobulins or with hyperimmunoglobulinemia |
| ICD10CM | D80.7 | Transient hypogammaglobulinemia of infancy |
| ICD10CM | D80.8 | Other immunodeficiencies with predominantly antibody defects |
| ICD10CM | D80.9 | Immunodeficiency with predominantly antibody defects, unspecified |
| ICD10CM | D81.0 | Severe combined immunodeficiency [SCID] with reticular dysgenesis |
| ICD10CM | D81.1 | Severe combined immunodeficiency [SCID] with low T- and B-cell numbers |
| ICD10CM | D81.2 | Severe combined immunodeficiency [SCID] with low or normal B-cell numbers |
| ICD10CM | D81.3 | Adenosine deaminase [ADA] deficiency |
| ICD10CM | D81.30 | Adenosine deaminase deficiency, unspecified |
| ICD10CM | D81.31 | Severe combined immunodeficiency due to adenosine deaminase deficiency |
| ICD10CM | D81.32 | Adenosine deaminase 2 deficiency |
| ICD10CM | D81.39 | Other adenosine deaminase deficiency |
| ICD10CM | D81.4 | Nezelof's syndrome |
| ICD10CM | D81.5 | Purine nucleoside phosphorylase [PNP] deficiency |
| ICD10CM | D81.6 | Major histocompatibility complex class I deficiency |
| ICD10CM | D81.7 | Major histocompatibility complex class II deficiency |
| ICD10CM | D81.8 | Other combined immunodeficiencies |
| ICD10CM | D81.81 | Biotin-dependent carboxylase deficiency |
| ICD10CM | D81.810 | Biotinidase deficiency |
| ICD10CM | D81.818 | Other biotin-dependent carboxylase deficiency |

Continued on next page

**TableS31 – continued from previous page**

| CodeSystem | ConceptCode | ConceptName |
|---|---|---|
| ICD10CM | D81.819 | Biotin-dependent carboxylase deficiency, unspecified |
| ICD10CM | D81.89 | Other combined immunodeficiencies |
| ICD10CM | D81.9 | Combined immunodeficiency, unspecified |
| ICD10CM | D82.0 | Wiskott-Aldrich syndrome |
| ICD10CM | D82.1 | Di George's syndrome |
| ICD10CM | D82.2 | Immunodeficiency with short-limbed stature |
| ICD10CM | D82.3 | Immunodeficiency following hereditary defective response to Epstein-Barr virus |
| ICD10CM | D82.4 | Hyperimmunoglobulin E [IgE] syndrome |
| ICD10CM | D82.8 | Immunodeficiency associated with other specified major defects |
| ICD10CM | D82.9 | Immunodeficiency associated with major defect, unspecified |
| ICD10CM | D89.4 | Mast cell activation syndrome and related disorders |
| ICD10CM | D83.0 | Common variable immunodeficiency with predominant abnormalities of B-cell numbers and function |
| ICD10CM | D83.1 | Common variable immunodeficiency with predominant immunoregulatory T-cell disorders |
| ICD10CM | D83.2 | Common variable immunodeficiency with autoantibodies to B- or T-cells |
| ICD10CM | D83.8 | Other common variable immunodeficiencies |
| ICD10CM | D83.9 | Common variable immunodeficiency, unspecified |
| ICD10CM | D84.0 | Lymphocyte function antigen-1 [LFA-1] defect |
| ICD10CM | D84.1 | Defects in the complement system |
| ICD10CM | D84.8 | Other specified immunodeficiencies |
| ICD10CM | D84.9 | Immunodeficiency, unspecified |
| ICD10CM | D89.0 | Polyclonal hypergammaglobulinemia |
| ICD10CM | D89.1 | Cryoglobulinemia |
| ICD10CM | D89.2 | Hypergammaglobulinemia, unspecified |
| ICD10CM | D89.3 | Immune reconstitution syndrome |
| ICD10CM | D89.40 | Mast cell activation, unspecified |
| ICD10CM | D89.41 | Monoclonal mast cell activation syndrome |
| ICD10CM | D89.42 | Idiopathic mast cell activation syndrome |
| ICD10CM | D89.43 | Secondary mast cell activation |
| ICD10CM | D89.49 | Other mast cell activation disorder |
| ICD10CM | D89.8 | Other specified disorders involving the immune mechanism, not elsewhere classified |
| ICD10CM | D89.81 | Graft-versus-host disease |
| ICD10CM | D89.810 | Acute graft-versus-host disease |
| ICD10CM | D89.811 | Chronic graft-versus-host disease |
| ICD10CM | D89.812 | Acute on chronic graft-versus-host disease |
| ICD10CM | D89.813 | Graft-versus-host disease, unspecified |
| ICD10CM | D89.82 | Autoimmune lymphoproliferative syndrome [ALPS] |
| ICD10CM | D89.89 | Other specified disorders involving the immune mechanism, not elsewhere classified |
| ICD10CM | D89.9 | Disorder involving the immune mechanism, unspecified |
| ICD10CM | Z94.0 | Kidney transplant status |
| ICD10CM | Z94.1 | Heart transplant status |
| ICD10CM | Z94.2 | Lung transplant status |

Continued on next page

**TableS31 – continued from previous page**

| CodeSystem | ConceptCode | ConceptName |
|---|---|---|
| ICD10CM | Z94.3 | Heart and lungs transplant status |
| ICD10CM | Z94.4 | Liver transplant status |
| ICD10CM | Z94.5 | Skin transplant status |
| ICD10CM | Z94.6 | Bone transplant status |
| ICD10CM | Z94.7 | Corneal transplant status |
| ICD10CM | Z94.8 | Other transplanted organ and tissue status |
| ICD10CM | Z94.81 | Bone marrow transplant status |
| ICD10CM | Z94.82 | Intestine transplant status |
| ICD10CM | Z94.83 | Pancreas transplant status |
| ICD10CM | Z94.84 | Stem cells transplant status |
| ICD10CM | Z94.89 | Other transplanted organ and tissue status |
| ICD10CM | Z94.9 | Transplanted organ and tissue status, unspecified |
| ICD10CM | D84.81 | Immunodeficiency due to conditions classified elsewhere |
| ICD10CM | D84.82 | Immunodeficiency due to drugs and external causes |
| ICD10CM | D84.821 | Immunodeficiency due to drugs |
| ICD10CM | D84.822 | Immunodeficiency due to external causes |
| ICD10CM | D84.89 | Other immunodeficiencies |
| ICD10CM | D89.83 | Cytokine release syndrome |
| ICD10CM | D89.831 | Cytokine release syndrome, grade 1 |
| ICD10CM | D89.832 | Cytokine release syndrome, grade 2 |
| ICD10CM | D89.833 | Cytokine release syndrome, grade 3 |
| ICD10CM | D89.834 | Cytokine release syndrome, grade 4 |
| ICD10CM | D89.835 | Cytokine release syndrome, grade 5 |
| ICD10CM | D89.839 | Cytokine release syndrome, grade unspecified |
| ICD10CM | C79.63 | Secondary malignant neoplasm of bilateral ovaries |
| ICD10CM | D89.44 | Hereditary alpha tryptasemia |
| ICD10CM | C84.7A | Anaplastic large cell lymphoma, ALK-negative, breast |
| ICD10CM | C56.3 | Malignant neoplasm of bilateral ovaries |

#### 5.11 Peripheral artery disease

Table S32: Concept codes used to identify if a patient was diagnosed with PAD.

| CodeSystem | ConceptCode | ConceptName |
|---|---|---|
| ICD10CM | E08.5 | Diabetes mellitus due to underlying condition with circulatory complications |
| ICD10CM | E09.5 | Drug or chemical induced diabetes mellitus with circulatory complications |
| ICD10CM | E10.5 | Type 1 diabetes mellitus with circulatory complications |
| ICD10CM | E11.5 | Type 2 diabetes mellitus with circulatory complications |
| ICD10CM | E13.5 | Other specified diabetes mellitus with circulatory complications |
| ICD10CM | I70.20 | Unspecified atherosclerosis of native arteries of extremities |
| ICD10CM | I70.21 | Atherosclerosis of native arteries of extremities with intermittent claudication |

Continued on next page

**TableS32 – continued from previous page**

| CodeSystem | ConceptCode | ConceptName |
|---|---|---|
| ICD10CM | I70.22 | Atherosclerosis of native arteries of extremities with rest pain |
| ICD10CM | I70.23 | Atherosclerosis of native arteries of right leg with ulceration |
| ICD10CM | I70.24 | Atherosclerosis of native arteries of left leg with ulceration |
| ICD10CM | I70.25 | Atherosclerosis of native arteries of other extremities with ulceration |
| ICD10CM | I70.26 | Atherosclerosis of native arteries of extremities with gangrene |
| ICD10CM | I70.29 | Other atherosclerosis of native arteries of extremities |
| ICD10CM | I70.30 | Unspecified atherosclerosis of unspecified type of bypass graft(s) of the extremities |
| ICD10CM | I70.31 | Atherosclerosis of unspecified type of bypass graft(s) of the extremities with intermittent claudication |
| ICD10CM | I70.32 | Atherosclerosis of unspecified type of bypass graft(s) of the extremities with rest pain |
| ICD10CM | I70.33 | Atherosclerosis of unspecified type of bypass graft(s) of the right leg with ulceration |
| ICD10CM | I70.34 | Atherosclerosis of unspecified type of bypass graft(s) of the left leg with ulceration |
| ICD10CM | I70.35 | Atherosclerosis of unspecified type of bypass graft(s) of other extremity with ulceration |
| ICD10CM | I70.4 | Atherosclerosis of autologous vein bypass graft(s) of the extremities |
| ICD10CM | I70.5 | Atherosclerosis of nonautologous biological bypass graft(s) of the extremities |
| ICD10CM | I70.6 | Atherosclerosis of nonbiological bypass graft(s) of the extremities |
| ICD10CM | I70.7 | Atherosclerosis of other type of bypass graft(s) of the extremities |
| ICD10CM | I70.90 | Unspecified atherosclerosis |
| ICD10CM | I70.91 | Generalized atherosclerosis |
| ICD10CM | I70.92 | Chronic total occlusion of artery of the extremities |
| ICD10CM | I73.89 | Other specified peripheral vascular diseases |
| ICD10CM | I73.9 | Peripheral vascular disease, unspecified |
| ICD10CM | E08.51 | Diabetes mellitus due to underlying condition with diabetic peripheral angiopathy without gangrene |
| ICD10CM | E08.52 | Diabetes mellitus due to underlying condition with diabetic peripheral angiopathy with gangrene |
| ICD10CM | E08.59 | Diabetes mellitus due to underlying condition with other circulatory complications |
| ICD10CM | E09.51 | Drug or chemical induced diabetes mellitus with diabetic peripheral angiopathy without gangrene |
| ICD10CM | E09.52 | Drug or chemical induced diabetes mellitus with diabetic peripheral angiopathy with gangrene |
| ICD10CM | E09.59 | Drug or chemical induced diabetes mellitus with other circulatory complications |
| ICD10CM | E10.51 | Type 1 diabetes mellitus with diabetic peripheral angiopathy without gangrene |
| ICD10CM | E10.52 | Type 1 diabetes mellitus with diabetic peripheral angiopathy with gangrene |
| ICD10CM | E10.59 | Type 1 diabetes mellitus with other circulatory complications |

Continued on next page

**TableS32 – continued from previous page**

| CodeSystem | ConceptCode | ConceptName |
|---|---|---|
| ICD10CM | E11.51 | Type 2 diabetes mellitus with diabetic peripheral angiopathy without gangrene |
| ICD10CM | E11.52 | Type 2 diabetes mellitus with diabetic peripheral angiopathy with gangrene |
| ICD10CM | E11.59 | Type 2 diabetes mellitus with other circulatory complications |
| ICD10CM | E13.51 | Other specified diabetes mellitus with diabetic peripheral angiopathy without gangrene |
| ICD10CM | E13.52 | Other specified diabetes mellitus with diabetic peripheral angiopathy with gangrene |
| ICD10CM | E13.59 | Other specified diabetes mellitus with other circulatory complications |
| ICD10CM | I70.201 | Unspecified atherosclerosis of native arteries of extremities, right leg |
| ICD10CM | I70.202 | Unspecified atherosclerosis of native arteries of extremities, left leg |
| ICD10CM | I70.203 | Unspecified atherosclerosis of native arteries of extremities, bilateral legs |
| ICD10CM | I70.208 | Unspecified atherosclerosis of native arteries of extremities, other extremity |
| ICD10CM | I70.209 | Unspecified atherosclerosis of native arteries of extremities, unspecified extremity |
| ICD10CM | I70.211 | Atherosclerosis of native arteries of extremities with intermittent claudication, right leg |
| ICD10CM | I70.212 | Atherosclerosis of native arteries of extremities with intermittent claudication, left leg |
| ICD10CM | I70.213 | Atherosclerosis of native arteries of extremities with intermittent claudication, bilateral legs |
| ICD10CM | I70.218 | Atherosclerosis of native arteries of extremities with intermittent claudication, other extremity |
| ICD10CM | I70.219 | Atherosclerosis of native arteries of extremities with intermittent claudication, unspecified extremity |
| ICD10CM | I70.221 | Atherosclerosis of native arteries of extremities with rest pain, right leg |
| ICD10CM | I70.222 | Atherosclerosis of native arteries of extremities with rest pain, left leg |
| ICD10CM | I70.223 | Atherosclerosis of native arteries of extremities with rest pain, bilateral legs |
| ICD10CM | I70.228 | Atherosclerosis of native arteries of extremities with rest pain, other extremity |
| ICD10CM | I70.229 | Atherosclerosis of native arteries of extremities with rest pain, unspecified extremity |
| ICD10CM | I70.231 | Atherosclerosis of native arteries of right leg with ulceration of thigh |
| ICD10CM | I70.232 | Atherosclerosis of native arteries of right leg with ulceration of calf |
| ICD10CM | I70.233 | Atherosclerosis of native arteries of right leg with ulceration of ankle |

Continued on next page

**TableS32 – continued from previous page**

| CodeSystem | ConceptCode | ConceptName |
|---|---|---|
| ICD10CM | I70.234 | Atherosclerosis of native arteries of right leg with ulceration of heel and midfoot |
| ICD10CM | I70.235 | Atherosclerosis of native arteries of right leg with ulceration of other part of foot |
| ICD10CM | I70.238 | Atherosclerosis of native arteries of right leg with ulceration of other part of lower leg |
| ICD10CM | I70.239 | Atherosclerosis of native arteries of right leg with ulceration of unspecified site |
| ICD10CM | I70.241 | Atherosclerosis of native arteries of left leg with ulceration of thigh |
| ICD10CM | I70.242 | Atherosclerosis of native arteries of left leg with ulceration of calf |
| ICD10CM | I70.243 | Atherosclerosis of native arteries of left leg with ulceration of ankle |
| ICD10CM | I70.244 | Atherosclerosis of native arteries of left leg with ulceration of heel and midfoot |
| ICD10CM | I70.245 | Atherosclerosis of native arteries of left leg with ulceration of other part of foot |
| ICD10CM | I70.248 | Atherosclerosis of native arteries of left leg with ulceration of other part of lower leg |
| ICD10CM | I70.249 | Atherosclerosis of native arteries of left leg with ulceration of unspecified site |
| ICD10CM | I70.261 | Atherosclerosis of native arteries of extremities with gangrene, right leg |
| ICD10CM | I70.262 | Atherosclerosis of native arteries of extremities with gangrene, left leg |
| ICD10CM | I70.263 | Atherosclerosis of native arteries of extremities with gangrene, bilateral legs |
| ICD10CM | I70.268 | Atherosclerosis of native arteries of extremities with gangrene, other extremity |
| ICD10CM | I70.269 | Atherosclerosis of native arteries of extremities with gangrene, unspecified extremity |
| ICD10CM | I70.291 | Other atherosclerosis of native arteries of extremities, right leg |
| ICD10CM | I70.292 | Other atherosclerosis of native arteries of extremities, left leg |
| ICD10CM | I70.293 | Other atherosclerosis of native arteries of extremities, bilateral legs |
| ICD10CM | I70.298 | Other atherosclerosis of native arteries of extremities, other extremity |
| ICD10CM | I70.299 | Other atherosclerosis of native arteries of extremities, unspecified extremity |
| ICD10CM | I70.301 | Unspecified atherosclerosis of unspecified type of bypass graft(s) of the extremities, right leg |
| ICD10CM | I70.302 | Unspecified atherosclerosis of unspecified type of bypass graft(s) of the extremities, left leg |
| ICD10CM | I70.303 | Unspecified atherosclerosis of unspecified type of bypass graft(s) of the extremities, bilateral legs |
| ICD10CM | I70.308 | Unspecified atherosclerosis of unspecified type of bypass graft(s) of the extremities, other extremity |

Continued on next page

**TableS32 – continued from previous page**

| CodeSystem | ConceptCode | ConceptName |
|---|---|---|
| ICD10CM | I70.309 | Unspecified atherosclerosis of unspecified type of bypass graft(s) of the extremities, unspecified extremity |
| ICD10CM | I70.311 | Atherosclerosis of unspecified type of bypass graft(s) of the extremities with intermittent claudication, right leg |
| ICD10CM | I70.312 | Atherosclerosis of unspecified type of bypass graft(s) of the extremities with intermittent claudication, left leg |
| ICD10CM | I70.313 | Atherosclerosis of unspecified type of bypass graft(s) of the extremities with intermittent claudication, bilateral legs |
| ICD10CM | I70.318 | Atherosclerosis of unspecified type of bypass graft(s) of the extremities with intermittent claudication, other extremity |
| ICD10CM | I70.319 | Atherosclerosis of unspecified type of bypass graft(s) of the extremities with intermittent claudication, unspecified extremity |
| ICD10CM | I70.321 | Atherosclerosis of unspecified type of bypass graft(s) of the extremities with rest pain, right leg |
| ICD10CM | I70.322 | Atherosclerosis of unspecified type of bypass graft(s) of the extremities with rest pain, left leg |
| ICD10CM | I70.323 | Atherosclerosis of unspecified type of bypass graft(s) of the extremities with rest pain, bilateral legs |
| ICD10CM | I70.328 | Atherosclerosis of unspecified type of bypass graft(s) of the extremities with rest pain, other extremity |
| ICD10CM | I70.329 | Atherosclerosis of unspecified type of bypass graft(s) of the extremities with rest pain, unspecified extremity |
| ICD10CM | I70.331 | Atherosclerosis of unspecified type of bypass graft(s) of the right leg with ulceration of thigh |
| ICD10CM | I70.332 | Atherosclerosis of unspecified type of bypass graft(s) of the right leg with ulceration of calf |
| ICD10CM | I70.333 | Atherosclerosis of unspecified type of bypass graft(s) of the right leg with ulceration of ankle |
| ICD10CM | I70.334 | Atherosclerosis of unspecified type of bypass graft(s) of the right leg with ulceration of heel and midfoot |
| ICD10CM | I70.335 | Atherosclerosis of unspecified type of bypass graft(s) of the right leg with ulceration of other part of foot |
| ICD10CM | I70.338 | Atherosclerosis of unspecified type of bypass graft(s) of the right leg with ulceration of other part of lower leg |
| ICD10CM | I70.339 | Atherosclerosis of unspecified type of bypass graft(s) of the right leg with ulceration of unspecified site |
| ICD10CM | I70.341 | Atherosclerosis of unspecified type of bypass graft(s) of the left leg with ulceration of thigh |
| ICD10CM | I70.342 | Atherosclerosis of unspecified type of bypass graft(s) of the left leg with ulceration of calf |
| ICD10CM | I70.343 | Atherosclerosis of unspecified type of bypass graft(s) of the left leg with ulceration of ankle |
| ICD10CM | I70.344 | Atherosclerosis of unspecified type of bypass graft(s) of the left leg with ulceration of heel and midfoot |
| ICD10CM | I70.345 | Atherosclerosis of unspecified type of bypass graft(s) of the left leg with ulceration of other part of foot |

Continued on next page

**TableS32 – continued from previous page**

| CodeSystem | ConceptCode | ConceptName |
|---|---|---|
| ICD10CM | I70.348 | Atherosclerosis of unspecified type of bypass graft(s) of the left leg with ulceration of other part of lower leg |
| ICD10CM | I70.349 | Atherosclerosis of unspecified type of bypass graft(s) of the left leg with ulceration of unspecified site |
| ICD10CM | I70.40 | Unspecified atherosclerosis of autologous vein bypass graft(s) of the extremities |
| ICD10CM | I70.401 | Unspecified atherosclerosis of autologous vein bypass graft(s) of the extremities, right leg |
| ICD10CM | I70.402 | Unspecified atherosclerosis of autologous vein bypass graft(s) of the extremities, left leg |
| ICD10CM | I70.403 | Unspecified atherosclerosis of autologous vein bypass graft(s) of the extremities, bilateral legs |
| ICD10CM | I70.408 | Unspecified atherosclerosis of autologous vein bypass graft(s) of the extremities, other extremity |
| ICD10CM | I70.409 | Unspecified atherosclerosis of autologous vein bypass graft(s) of the extremities, unspecified extremity |
| ICD10CM | I70.41 | Atherosclerosis of autologous vein bypass graft(s) of the extremities with intermittent claudication |
| ICD10CM | I70.411 | Atherosclerosis of autologous vein bypass graft(s) of the extremities with intermittent claudication, right leg |
| ICD10CM | I70.412 | Atherosclerosis of autologous vein bypass graft(s) of the extremities with intermittent claudication, left leg |
| ICD10CM | I70.413 | Atherosclerosis of autologous vein bypass graft(s) of the extremities with intermittent claudication, bilateral legs |
| ICD10CM | I70.418 | Atherosclerosis of autologous vein bypass graft(s) of the extremities with intermittent claudication, other extremity |
| ICD10CM | I70.419 | Atherosclerosis of autologous vein bypass graft(s) of the extremities with intermittent claudication, unspecified extremity |
| ICD10CM | I70.42 | Atherosclerosis of autologous vein bypass graft(s) of the extremities with rest pain |
| ICD10CM | I70.421 | Atherosclerosis of autologous vein bypass graft(s) of the extremities with rest pain, right leg |
| ICD10CM | I70.422 | Atherosclerosis of autologous vein bypass graft(s) of the extremities with rest pain, left leg |
| ICD10CM | I70.423 | Atherosclerosis of autologous vein bypass graft(s) of the extremities with rest pain, bilateral legs |
| ICD10CM | I70.428 | Atherosclerosis of autologous vein bypass graft(s) of the extremities with rest pain, other extremity |
| ICD10CM | I70.429 | Atherosclerosis of autologous vein bypass graft(s) of the extremities with rest pain, unspecified extremity |
| ICD10CM | I70.43 | Atherosclerosis of autologous vein bypass graft(s) of the right leg with ulceration |
| ICD10CM | I70.431 | Atherosclerosis of autologous vein bypass graft(s) of the right leg with ulceration of thigh |
| ICD10CM | I70.432 | Atherosclerosis of autologous vein bypass graft(s) of the right leg with ulceration of calf |

Continued on next page

**TableS32 – continued from previous page**

| CodeSystem | ConceptCode | ConceptName |
|---|---|---|
| ICD10CM | I70.433 | Atherosclerosis of autologous vein bypass graft(s) of the right leg with ulceration of ankle |
| ICD10CM | I70.434 | Atherosclerosis of autologous vein bypass graft(s) of the right leg with ulceration of heel and midfoot |
| ICD10CM | I70.435 | Atherosclerosis of autologous vein bypass graft(s) of the right leg with ulceration of other part of foot |
| ICD10CM | I70.438 | Atherosclerosis of autologous vein bypass graft(s) of the right leg with ulceration of other part of lower leg |
| ICD10CM | I70.439 | Atherosclerosis of autologous vein bypass graft(s) of the right leg with ulceration of unspecified site |
| ICD10CM | I70.44 | Atherosclerosis of autologous vein bypass graft(s) of the left leg with ulceration |
| ICD10CM | I70.441 | Atherosclerosis of autologous vein bypass graft(s) of the left leg with ulceration of thigh |
| ICD10CM | I70.442 | Atherosclerosis of autologous vein bypass graft(s) of the left leg with ulceration of calf |
| ICD10CM | I70.443 | Atherosclerosis of autologous vein bypass graft(s) of the left leg with ulceration of ankle |
| ICD10CM | I70.444 | Atherosclerosis of autologous vein bypass graft(s) of the left leg with ulceration of heel and midfoot |
| ICD10CM | I70.445 | Atherosclerosis of autologous vein bypass graft(s) of the left leg with ulceration of other part of foot |
| ICD10CM | I70.448 | Atherosclerosis of autologous vein bypass graft(s) of the left leg with ulceration of other part of lower leg |
| ICD10CM | I70.449 | Atherosclerosis of autologous vein bypass graft(s) of the left leg with ulceration of unspecified site |
| ICD10CM | I70.45 | Atherosclerosis of autologous vein bypass graft(s) of other extremity with ulceration |
| ICD10CM | I70.46 | Atherosclerosis of autologous vein bypass graft(s) of the extremities with gangrene |
| ICD10CM | I70.461 | Atherosclerosis of autologous vein bypass graft(s) of the extremities with gangrene, right leg |
| ICD10CM | I70.462 | Atherosclerosis of autologous vein bypass graft(s) of the extremities with gangrene, left leg |
| ICD10CM | I70.463 | Atherosclerosis of autologous vein bypass graft(s) of the extremities with gangrene, bilateral legs |
| ICD10CM | I70.468 | Atherosclerosis of autologous vein bypass graft(s) of the extremities with gangrene, other extremity |
| ICD10CM | I70.469 | Atherosclerosis of autologous vein bypass graft(s) of the extremities with gangrene, unspecified extremity |
| ICD10CM | I70.49 | Other atherosclerosis of autologous vein bypass graft(s) of the extremities |
| ICD10CM | I70.491 | Other atherosclerosis of autologous vein bypass graft(s) of the extremities, right leg |
| ICD10CM | I70.492 | Other atherosclerosis of autologous vein bypass graft(s) of the extremities, left leg |

Continued on next page

**TableS32 – continued from previous page**

| CodeSystem | ConceptCode | ConceptName |
|---|---|---|
| ICD10CM | I70.493 | Other atherosclerosis of autologous vein bypass graft(s) of the extremities, bilateral legs |
| ICD10CM | I70.498 | Other atherosclerosis of autologous vein bypass graft(s) of the extremities, other extremity |
| ICD10CM | I70.499 | Other atherosclerosis of autologous vein bypass graft(s) of the extremities, unspecified extremity |
| ICD10CM | I70.50 | Unspecified atherosclerosis of nonautologous biological bypass graft(s) of the extremities |
| ICD10CM | I70.501 | Unspecified atherosclerosis of nonautologous biological bypass graft(s) of the extremities, right leg |
| ICD10CM | I70.502 | Unspecified atherosclerosis of nonautologous biological bypass graft(s) of the extremities, left leg |
| ICD10CM | I70.503 | Unspecified atherosclerosis of nonautologous biological bypass graft(s) of the extremities, bilateral legs |
| ICD10CM | I70.508 | Unspecified atherosclerosis of nonautologous biological bypass graft(s) of the extremities, other extremity |
| ICD10CM | I70.509 | Unspecified atherosclerosis of nonautologous biological bypass graft(s) of the extremities, unspecified extremity |
| ICD10CM | I70.51 | Atherosclerosis of nonautologous biological bypass graft(s) of the extremities intermittent claudication |
| ICD10CM | I70.511 | Atherosclerosis of nonautologous biological bypass graft(s) of the extremities with intermittent claudication, right leg |
| ICD10CM | I70.512 | Atherosclerosis of nonautologous biological bypass graft(s) of the extremities with intermittent claudication, left leg |
| ICD10CM | I70.513 | Atherosclerosis of nonautologous biological bypass graft(s) of the extremities with intermittent claudication, bilateral legs |
| ICD10CM | I70.518 | Atherosclerosis of nonautologous biological bypass graft(s) of the extremities with intermittent claudication, other extremity |
| ICD10CM | I70.519 | Atherosclerosis of nonautologous biological bypass graft(s) of the extremities with intermittent claudication, unspecified extremity |
| ICD10CM | I70.52 | Atherosclerosis of nonautologous biological bypass graft(s) of the extremities with rest pain |
| ICD10CM | I70.521 | Atherosclerosis of nonautologous biological bypass graft(s) of the extremities with rest pain, right leg |
| ICD10CM | I70.522 | Atherosclerosis of nonautologous biological bypass graft(s) of the extremities with rest pain, left leg |
| ICD10CM | I70.523 | Atherosclerosis of nonautologous biological bypass graft(s) of the extremities with rest pain, bilateral legs |
| ICD10CM | I70.528 | Atherosclerosis of nonautologous biological bypass graft(s) of the extremities with rest pain, other extremity |
| ICD10CM | I70.529 | Atherosclerosis of nonautologous biological bypass graft(s) of the extremities with rest pain, unspecified extremity |
| ICD10CM | I70.53 | Atherosclerosis of nonautologous biological bypass graft(s) of the right leg with ulceration |
| ICD10CM | I70.531 | Atherosclerosis of nonautologous biological bypass graft(s) of the right leg with ulceration of thigh |

Continued on next page

**TableS32 – continued from previous page**

| CodeSystem | ConceptCode | ConceptName |
|---|---|---|
| ICD10CM | I70.532 | Atherosclerosis of nonautologous biological bypass graft(s) of the right leg with ulceration of calf |
| ICD10CM | I70.533 | Atherosclerosis of nonautologous biological bypass graft(s) of the right leg with ulceration of ankle |
| ICD10CM | I70.534 | Atherosclerosis of nonautologous biological bypass graft(s) of the right leg with ulceration of heel and midfoot |
| ICD10CM | I70.535 | Atherosclerosis of nonautologous biological bypass graft(s) of the right leg with ulceration of other part of foot |
| ICD10CM | I70.538 | Atherosclerosis of nonautologous biological bypass graft(s) of the right leg with ulceration of other part of lower leg |
| ICD10CM | I70.539 | Atherosclerosis of nonautologous biological bypass graft(s) of the right leg with ulceration of unspecified site |
| ICD10CM | I70.54 | Atherosclerosis of nonautologous biological bypass graft(s) of the left leg with ulceration |
| ICD10CM | I70.541 | Atherosclerosis of nonautologous biological bypass graft(s) of the left leg with ulceration of thigh |
| ICD10CM | I70.542 | Atherosclerosis of nonautologous biological bypass graft(s) of the left leg with ulceration of calf |
| ICD10CM | I70.543 | Atherosclerosis of nonautologous biological bypass graft(s) of the left leg with ulceration of ankle |
| ICD10CM | I70.544 | Atherosclerosis of nonautologous biological bypass graft(s) of the left leg with ulceration of heel and midfoot |
| ICD10CM | I70.545 | Atherosclerosis of nonautologous biological bypass graft(s) of the left leg with ulceration of other part of foot |
| ICD10CM | I70.548 | Atherosclerosis of nonautologous biological bypass graft(s) of the left leg with ulceration of other part of lower leg |
| ICD10CM | I70.549 | Atherosclerosis of nonautologous biological bypass graft(s) of the left leg with ulceration of unspecified site |
| ICD10CM | I70.55 | Atherosclerosis of nonautologous biological bypass graft(s) of other extremity with ulceration |
| ICD10CM | I70.56 | Atherosclerosis of nonautologous biological bypass graft(s) of the extremities with gangrene |
| ICD10CM | I70.561 | Atherosclerosis of nonautologous biological bypass graft(s) of the extremities with gangrene, right leg |
| ICD10CM | I70.562 | Atherosclerosis of nonautologous biological bypass graft(s) of the extremities with gangrene, left leg |
| ICD10CM | I70.563 | Atherosclerosis of nonautologous biological bypass graft(s) of the extremities with gangrene, bilateral legs |
| ICD10CM | I70.568 | Atherosclerosis of nonautologous biological bypass graft(s) of the extremities with gangrene, other extremity |
| ICD10CM | I70.569 | Atherosclerosis of nonautologous biological bypass graft(s) of the extremities with gangrene, unspecified extremity |
| ICD10CM | I70.59 | Other atherosclerosis of nonautologous biological bypass graft(s) of the extremities |
| ICD10CM | I70.591 | Other atherosclerosis of nonautologous biological bypass graft(s) of the extremities, right leg |

Continued on next page

**TableS32 – continued from previous page**

| CodeSystem | ConceptCode | ConceptName |
|---|---|---|
| ICD10CM | I70.592 | Other atherosclerosis of nonautologous biological bypass graft(s) of the extremities, left leg |
| ICD10CM | I70.593 | Other atherosclerosis of nonautologous biological bypass graft(s) of the extremities, bilateral legs |
| ICD10CM | I70.598 | Other atherosclerosis of nonautologous biological bypass graft(s) of the extremities, other extremity |
| ICD10CM | I70.599 | Other atherosclerosis of nonautologous biological bypass graft(s) of the extremities, unspecified extremity |
| ICD10CM | I70.60 | Unspecified atherosclerosis of nonbiological bypass graft(s) of the extremities |
| ICD10CM | I70.601 | Unspecified atherosclerosis of nonbiological bypass graft(s) of the extremities, right leg |
| ICD10CM | I70.602 | Unspecified atherosclerosis of nonbiological bypass graft(s) of the extremities, left leg |
| ICD10CM | I70.603 | Unspecified atherosclerosis of nonbiological bypass graft(s) of the extremities, bilateral legs |
| ICD10CM | I70.608 | Unspecified atherosclerosis of nonbiological bypass graft(s) of the extremities, other extremity |
| ICD10CM | I70.609 | Unspecified atherosclerosis of nonbiological bypass graft(s) of the extremities, unspecified extremity |
| ICD10CM | I70.61 | Atherosclerosis of nonbiological bypass graft(s) of the extremities with intermittent claudication |
| ICD10CM | I70.611 | Atherosclerosis of nonbiological bypass graft(s) of the extremities with intermittent claudication, right leg |
| ICD10CM | I70.612 | Atherosclerosis of nonbiological bypass graft(s) of the extremities with intermittent claudication, left leg |
| ICD10CM | I70.613 | Atherosclerosis of nonbiological bypass graft(s) of the extremities with intermittent claudication, bilateral legs |
| ICD10CM | I70.618 | Atherosclerosis of nonbiological bypass graft(s) of the extremities with intermittent claudication, other extremity |
| ICD10CM | I70.619 | Atherosclerosis of nonbiological bypass graft(s) of the extremities with intermittent claudication, unspecified extremity |
| ICD10CM | I70.62 | Atherosclerosis of nonbiological bypass graft(s) of the extremities with rest pain |
| ICD10CM | I70.621 | Atherosclerosis of nonbiological bypass graft(s) of the extremities with rest pain, right leg |
| ICD10CM | I70.622 | Atherosclerosis of nonbiological bypass graft(s) of the extremities with rest pain, left leg |
| ICD10CM | I70.623 | Atherosclerosis of nonbiological bypass graft(s) of the extremities with rest pain, bilateral legs |
| ICD10CM | I70.628 | Atherosclerosis of nonbiological bypass graft(s) of the extremities with rest pain, other extremity |
| ICD10CM | I70.629 | Atherosclerosis of nonbiological bypass graft(s) of the extremities with rest pain, unspecified extremity |
| ICD10CM | I70.63 | Atherosclerosis of nonbiological bypass graft(s) of the right leg with ulceration |

Continued on next page

**TableS32 – continued from previous page**

| CodeSystem | ConceptCode | ConceptName |
|---|---|---|
| ICD10CM | I70.631 | Atherosclerosis of nonbiological bypass graft(s) of the right leg with ulceration of thigh |
| ICD10CM | I70.632 | Atherosclerosis of nonbiological bypass graft(s) of the right leg with ulceration of calf |
| ICD10CM | I70.633 | Atherosclerosis of nonbiological bypass graft(s) of the right leg with ulceration of ankle |
| ICD10CM | I70.634 | Atherosclerosis of nonbiological bypass graft(s) of the right leg with ulceration of heel and midfoot |
| ICD10CM | I70.635 | Atherosclerosis of nonbiological bypass graft(s) of the right leg with ulceration of other part of foot |
| ICD10CM | I70.638 | Atherosclerosis of nonbiological bypass graft(s) of the right leg with ulceration of other part of lower leg |
| ICD10CM | I70.639 | Atherosclerosis of nonbiological bypass graft(s) of the right leg with ulceration of unspecified site |
| ICD10CM | I70.64 | Atherosclerosis of nonbiological bypass graft(s) of the left leg with ulceration |
| ICD10CM | I70.641 | Atherosclerosis of nonbiological bypass graft(s) of the left leg with ulceration of thigh |
| ICD10CM | I70.642 | Atherosclerosis of nonbiological bypass graft(s) of the left leg with ulceration of calf |
| ICD10CM | I70.643 | Atherosclerosis of nonbiological bypass graft(s) of the left leg with ulceration of ankle |
| ICD10CM | I70.644 | Atherosclerosis of nonbiological bypass graft(s) of the left leg with ulceration of heel and midfoot |
| ICD10CM | I70.645 | Atherosclerosis of nonbiological bypass graft(s) of the left leg with ulceration of other part of foot |
| ICD10CM | I70.648 | Atherosclerosis of nonbiological bypass graft(s) of the left leg with ulceration of other part of lower leg |
| ICD10CM | I70.649 | Atherosclerosis of nonbiological bypass graft(s) of the left leg with ulceration of unspecified site |
| ICD10CM | I70.65 | Atherosclerosis of nonbiological bypass graft(s) of other extremity with ulceration |
| ICD10CM | I70.66 | Atherosclerosis of nonbiological bypass graft(s) of the extremities with gangrene |
| ICD10CM | I70.661 | Atherosclerosis of nonbiological bypass graft(s) of the extremities with gangrene, right leg |
| ICD10CM | I70.662 | Atherosclerosis of nonbiological bypass graft(s) of the extremities with gangrene, left leg |
| ICD10CM | I70.663 | Atherosclerosis of nonbiological bypass graft(s) of the extremities with gangrene, bilateral legs |
| ICD10CM | I70.668 | Atherosclerosis of nonbiological bypass graft(s) of the extremities with gangrene, other extremity |
| ICD10CM | I70.669 | Atherosclerosis of nonbiological bypass graft(s) of the extremities with gangrene, unspecified extremity |
| ICD10CM | I70.69 | Other atherosclerosis of nonbiological bypass graft(s) of the extremities |

Continued on next page

**TableS32 – continued from previous page**

| CodeSystem | ConceptCode | ConceptName |
|---|---|---|
| ICD10CM | I70.691 | Other atherosclerosis of nonbiological bypass graft(s) of the extremities, right leg |
| ICD10CM | I70.692 | Other atherosclerosis of nonbiological bypass graft(s) of the extremities, left leg |
| ICD10CM | I70.693 | Other atherosclerosis of nonbiological bypass graft(s) of the extremities, bilateral legs |
| ICD10CM | I70.698 | Other atherosclerosis of nonbiological bypass graft(s) of the extremities, other extremity |
| ICD10CM | I70.699 | Other atherosclerosis of nonbiological bypass graft(s) of the extremities, unspecified extremity |
| ICD10CM | I70.70 | Unspecified atherosclerosis of other type of bypass graft(s) of the extremities |
| ICD10CM | I70.701 | Unspecified atherosclerosis of other type of bypass graft(s) of the extremities, right leg |
| ICD10CM | I70.702 | Unspecified atherosclerosis of other type of bypass graft(s) of the extremities, left leg |
| ICD10CM | I70.703 | Unspecified atherosclerosis of other type of bypass graft(s) of the extremities, bilateral legs |
| ICD10CM | I70.708 | Unspecified atherosclerosis of other type of bypass graft(s) of the extremities, other extremity |
| ICD10CM | I70.709 | Unspecified atherosclerosis of other type of bypass graft(s) of the extremities, unspecified extremity |
| ICD10CM | I70.71 | Atherosclerosis of other type of bypass graft(s) of the extremities with intermittent claudication |
| ICD10CM | I70.711 | Atherosclerosis of other type of bypass graft(s) of the extremities with intermittent claudication, right leg |
| ICD10CM | I70.712 | Atherosclerosis of other type of bypass graft(s) of the extremities with intermittent claudication, left leg |
| ICD10CM | I70.713 | Atherosclerosis of other type of bypass graft(s) of the extremities with intermittent claudication, bilateral legs |
| ICD10CM | I70.718 | Atherosclerosis of other type of bypass graft(s) of the extremities with intermittent claudication, other extremity |
| ICD10CM | I70.719 | Atherosclerosis of other type of bypass graft(s) of the extremities with intermittent claudication, unspecified extremity |
| ICD10CM | I70.72 | Atherosclerosis of other type of bypass graft(s) of the extremities with rest pain |
| ICD10CM | I70.721 | Atherosclerosis of other type of bypass graft(s) of the extremities with rest pain, right leg |
| ICD10CM | I70.722 | Atherosclerosis of other type of bypass graft(s) of the extremities with rest pain, left leg |
| ICD10CM | I70.723 | Atherosclerosis of other type of bypass graft(s) of the extremities with rest pain, bilateral legs |
| ICD10CM | I70.728 | Atherosclerosis of other type of bypass graft(s) of the extremities with rest pain, other extremity |
| ICD10CM | I70.729 | Atherosclerosis of other type of bypass graft(s) of the extremities with rest pain, unspecified extremity |

Continued on next page

**TableS32 – continued from previous page**

| CodeSystem | ConceptCode | ConceptName |
|---|---|---|
| ICD10CM | I70.73 | Atherosclerosis of other type of bypass graft(s) of the right leg with ulceration |
| ICD10CM | I70.731 | Atherosclerosis of other type of bypass graft(s) of the right leg with ulceration of thigh |
| ICD10CM | I70.732 | Atherosclerosis of other type of bypass graft(s) of the right leg with ulceration of calf |
| ICD10CM | I70.733 | Atherosclerosis of other type of bypass graft(s) of the right leg with ulceration of ankle |
| ICD10CM | I70.734 | Atherosclerosis of other type of bypass graft(s) of the right leg with ulceration of heel and midfoot |
| ICD10CM | I70.735 | Atherosclerosis of other type of bypass graft(s) of the right leg with ulceration of other part of foot |
| ICD10CM | I70.738 | Atherosclerosis of other type of bypass graft(s) of the right leg with ulceration of other part of lower leg |
| ICD10CM | I70.739 | Atherosclerosis of other type of bypass graft(s) of the right leg with ulceration of unspecified site |
| ICD10CM | I70.74 | Atherosclerosis of other type of bypass graft(s) of the left leg with ulceration |
| ICD10CM | I70.741 | Atherosclerosis of other type of bypass graft(s) of the left leg with ulceration of thigh |
| ICD10CM | I70.742 | Atherosclerosis of other type of bypass graft(s) of the left leg with ulceration of calf |
| ICD10CM | I70.743 | Atherosclerosis of other type of bypass graft(s) of the left leg with ulceration of ankle |
| ICD10CM | I70.744 | Atherosclerosis of other type of bypass graft(s) of the left leg with ulceration of heel and midfoot |
| ICD10CM | I70.745 | Atherosclerosis of other type of bypass graft(s) of the left leg with ulceration of other part of foot |
| ICD10CM | I70.748 | Atherosclerosis of other type of bypass graft(s) of the left leg with ulceration of other part of lower leg |
| ICD10CM | I70.749 | Atherosclerosis of other type of bypass graft(s) of the left leg with ulceration of unspecified site |
| ICD10CM | I70.75 | Atherosclerosis of other type of bypass graft(s) of other extremity with ulceration |
| ICD10CM | I70.76 | Atherosclerosis of other type of bypass graft(s) of the extremities with gangrene |
| ICD10CM | I70.761 | Atherosclerosis of other type of bypass graft(s) of the extremities with gangrene, right leg |
| ICD10CM | I70.762 | Atherosclerosis of other type of bypass graft(s) of the extremities with gangrene, left leg |
| ICD10CM | I70.763 | Atherosclerosis of other type of bypass graft(s) of the extremities with gangrene, bilateral legs |
| ICD10CM | I70.768 | Atherosclerosis of other type of bypass graft(s) of the extremities with gangrene, other extremity |
| ICD10CM | I70.769 | Atherosclerosis of other type of bypass graft(s) of the extremities with gangrene, unspecified extremity |

Continued on next page

**TableS32 – continued from previous page**

| CodeSystem | ConceptCode | ConceptName |
|---|---|---|
| ICD10CM | I70.79 | Other atherosclerosis of other type of bypass graft(s) of the extremities |
| ICD10CM | I70.791 | Other atherosclerosis of other type of bypass graft(s) of the extremities, right leg |
| ICD10CM | I70.792 | Other atherosclerosis of other type of bypass graft(s) of the extremities, left leg |
| ICD10CM | I70.793 | Other atherosclerosis of other type of bypass graft(s) of the extremities, bilateral legs |
| ICD10CM | I70.798 | Other atherosclerosis of other type of bypass graft(s) of the extremities, other extremity |
| ICD10CM | I70.799 | Other atherosclerosis of other type of bypass graft(s) of the extremities, unspecified extremity |

### 6 Demographics and other patient descriptors

#### 6.1 Influenza vaccine

Table S33: Concept codes used to identify if a patient had a influenza vaccination event.

| CodeSystem | ConceptCode | ConceptName |
|---|---|---|
| CVX | 135 | influenza, high dose seasonal, preservative-free |
| CVX | 140 | Influenza, seasonal, injectable, preservative free |
| CVX | 141 | Influenza, seasonal, injectable |
| CVX | 144 | seasonal influenza, intradermal, preservative free |
| CVX | 149 | influenza, live, intranasal, quadrivalent |
| CVX | 150 | Influenza, injectable, quadrivalent, preservative free |
| CVX | 155 | Seasonal, trivalent, recombinant, injectable influenza vaccine, preservative free |
| CVX | 158 | influenza, injectable, quadrivalent, contains preservative |
| CVX | 161 | Influenza, injectable,quadrivalent, preservative free, pediatric |
| CVX | 166 | influenza, intradermal, quadrivalent, preservative free, injectable |
| CVX | 168 | Seasonal trivalent influenza vaccine, adjuvanted, preservative free |
| CVX | 171 | Influenza, injectable, Madin Darby Canine Kidney, preservative free, quadrivalent |
| CVX | 185 | Seasonal, quadrivalent, recombinant, injectable influenza vaccine, preservative free |
| CVX | 186 | Influenza, injectable, Madin Darby Canine Kidney, quadrivalent with preservative |
| CVX | 197 | influenza, high-dose seasonal, quadrivalent, .7mL dose, preservative free |
| CVX | 205 | influenza, seasonal vaccine, quadrivalent, adjuvanted, .5mL dose, preservative free |

Continued on next page

**TableS33 – continued from previous page**

| CodeSystem | ConceptCode | ConceptName |
|---|---|---|
| CVX | 88 | influenza virus vaccine, unspecified formulation |

#### 6.2 Smoking

Table S34: Concept codes used to identify if a patient can be identified as a smoker.

| CodeSystem | ConceptCode | ConceptName |
|---|---|---|
| ICD10CM | F17 | Nicotine dependence |
| ICD10CM | O99.33 | Tobacco use disorder complicating pregnancy, childbirth, and the puerperium |
| ICD10CM | Z53.01 | Procedure and treatment not carried out due to patient smoking |
| ICD10CM | Z72.0 | Tobacco use |
| ICD10CM | Z87.891 | Personal history of nicotine dependence |
| ICD10CM | F17.2 | Nicotine dependence |
| ICD10CM | F17.20 | Nicotine dependence, unspecified |
| ICD10CM | F17.200 | Nicotine dependence, unspecified, uncomplicated |
| ICD10CM | F17.201 | Nicotine dependence, unspecified, in remission |
| ICD10CM | F17.203 | Nicotine dependence unspecified, with withdrawal |
| ICD10CM | F17.208 | Nicotine dependence, unspecified, with other nicotine-induced disorders |
| ICD10CM | F17.209 | Nicotine dependence, unspecified, with unspecified nicotine-induced disorders |
| ICD10CM | F17.21 | Nicotine dependence, cigarettes |
| ICD10CM | F17.210 | Nicotine dependence, cigarettes, uncomplicated |
| ICD10CM | F17.211 | Nicotine dependence, cigarettes, in remission |
| ICD10CM | F17.213 | Nicotine dependence, cigarettes, with withdrawal |
| ICD10CM | F17.218 | Nicotine dependence, cigarettes, with other nicotine-induced disorders |
| ICD10CM | F17.219 | Nicotine dependence, cigarettes, with unspecified nicotine-induced disorders |
| ICD10CM | F17.22 | Nicotine dependence, chewing tobacco |
| ICD10CM | F17.220 | Nicotine dependence, chewing tobacco, uncomplicated |
| ICD10CM | F17.221 | Nicotine dependence, chewing tobacco, in remission |
| ICD10CM | F17.223 | Nicotine dependence, chewing tobacco, with withdrawal |
| ICD10CM | F17.228 | Nicotine dependence, chewing tobacco, with other nicotine-induced disorders |
| ICD10CM | F17.229 | Nicotine dependence, chewing tobacco, with unspecified nicotine-induced disorders |
| ICD10CM | F17.29 | Nicotine dependence, other tobacco product |
| ICD10CM | F17.290 | Nicotine dependence, other tobacco product, uncomplicated |
| ICD10CM | F17.291 | Nicotine dependence, other tobacco product, in remission |
| ICD10CM | F17.293 | Nicotine dependence, other tobacco product, with withdrawal |
| ICD10CM | F17.298 | Nicotine dependence, other tobacco product, with other nicotine-induced disorders |

Continued on next page

**TableS34 – continued from previous page**

| CodeSystem | ConceptCode | ConceptName |
|---|---|---|
| ICD10CM | F17.299 | Nicotine dependence, other tobacco product, with unspecified nicotine-induced disorders |
| ICD10CM | O99.330 | Smoking (tobacco) complicating pregnancy, unspecified trimester |
| ICD10CM | O99.331 | Smoking (tobacco) complicating pregnancy, first trimester |
| ICD10CM | O99.332 | Smoking (tobacco) complicating pregnancy, second trimester |
| ICD10CM | O99.333 | Smoking (tobacco) complicating pregnancy, third trimester |
| ICD10CM | O99.334 | Smoking (tobacco) complicating childbirth |
| ICD10CM | O99.335 | Smoking (tobacco) complicating the puerperium |
| SNOMED CT | 77176002 | Smoking |
| SNOMED CT | 266920004 | Trivial cigarette smoker (less than one cigarette/day) |
| SNOMED CT | 266929003 | Smoking started |
| SNOMED CT | 308438006 | Smoking restarted |
| SNOMED CT | 82302008 | Pipe smoker |
| SNOMED CT | 160603005 | Light cigarette smoker (1-9 cigs/day) |
| SNOMED CT | 160604004 | Moderate cigarette smoker (10-19 cigs/day) |
| SNOMED CT | 160605003 | Heavy cigarette smoker (20-39 cigs/day) |
| SNOMED CT | 160606002 | Very heavy cigarette smoker (40+ cigs/day) |
| SNOMED CT | 160619003 | Rolls own cigarettes |
| SNOMED CT | 230059006 | Occasional cigarette smoker |
| SNOMED CT | 230060001 | Light cigarette smoker |
| SNOMED CT | 230062009 | Moderate cigarette smoker |
| SNOMED CT | 230063004 | Heavy cigarette smoker |
| SNOMED CT | 230064005 | Very heavy cigarette smoker |
| SNOMED CT | 230065006 | Chain smoker |
| SNOMED CT | 56578002 | Moderate smoker (20 or less per day) |
| SNOMED CT | 56771006 | Heavy smoker (over 20 per day) |
| SNOMED CT | 59978006 | Cigar smoker |
| SNOMED CT | 65568007 | Cigarette smoker |
| SNOMED CT | 428061000124105 | Light tobacco smoker |
| SNOMED CT | 428071000124103 | Heavy tobacco smoker |
| SNOMED CT | 450811000124104 | Cigarette smoker (5-9 cigarettes/day) |
| SNOMED CT | 450821000124107 | Cigarette smoker (1-4 cigarettes/day) |
| SNOMED CT | 428041000124106 | Occasional tobacco smoker |

### 7 Sensitivity analyses

We performed two sensitivity analyses to determine the effect of certain design decisions made during out analyses. We performed these analyses for both definitions of Long COVID considered in the main text.

#### 7.1 Remove patients with zero follow-up time

We analyzed the effect of excluding all patients who had effectively 0 follow-up time after 28 days post COVID-19 infection. This group of patients represent individuals where their positive COVID-19 test which forms our inclusion criterion represents their last recorded electronic health record event. By including them in our main analyses we are assuming that these patients did not die within 28 days of their infection and can be right-censored at approximately T0 (28 days after a positive COVID-19 test).

This removal of patients with effectively zero follow-up time is the only difference in methods compared to the analysis of the effect of vaccination modeled as a time-dependent covariate presented in the main text.

##### 7.1.1 Long COVID symptoms

We had a total of 290,277 people in this analysis. This sample size is smaller than the analyses presented in the main text because here we excluded individuals with effectively zero follow-up time.

Estimated survival functions for the time from T0 till development of Long COVID symptoms with vaccination state treated as a time-dependent covariate (Fig. S1) have a similar pattern to the survival curves from the full analysis presented in the main text. We see obvious differences in the time till outcome between persons with the unvaccinated state compared those with the vaccinated or vaccinated and boosted states (Fig. S1).

The pattern from the Kaplan-Meier curves is effectively retained when time till development of Long COVID symptoms is further conditioned on the comorbidities and patient descriptive features described above, as evidenced by the hazard ratios estimated as part of our Cox regression model (Fig.S2, Table S35). We find that patients who are vaccinated are at a lower risk of developing Long COVID symptoms than those who are unvaccinated (Fig. S2, Table S35). Similarly, those who are vaccinated and boosted are at a lower risk of developing Long COVID symptoms than those who are unvaccinated. Finally, we also find no evidence of a difference in risk of developing Long COVID symptoms between patients vaccinated and boosted versus those patients who are vaccinated (Fig. S2, Table S35).

| Comparison | Hazard Ratio [95% CI] |
|---|---|
| vaccinated vs unvaccinated | 0.91 [0.88, 0.95] |
| vaccinated and boosted vs unvaccinated | 0.88 [0.85, 0.91] |
| vaccinated and boosted vs vaccinated | 0.96 [0.92, 1.01] |

Table S35: Estimated hazard ratios for risk of developing Long COVID symptoms associated with vaccination status where vaccination status is modeled as time-dependent covariates. Hazard ratios are presented with 95% confidence intervals.

##### 7.1.2 Diagnosis with Long COVID

We had a total of 288,860 people in this analysis. This sample size is smaller than the full analysis presented in the main text because here we excluded individuals with effectively zero follow-up time.

As with the main analysis of diagnosis of Long COVID as our response of interest along with treating vaccination status as a time-dependent covariate, the results from our analysis removing all patients with effectively zero follow-up time are nearly flat over time (Fig. S3). Whatever difference exists between the unvaccinated, vaccinated, and vaccinated and boosted populations they are extremely small in absolute effect on time till diagnosis.

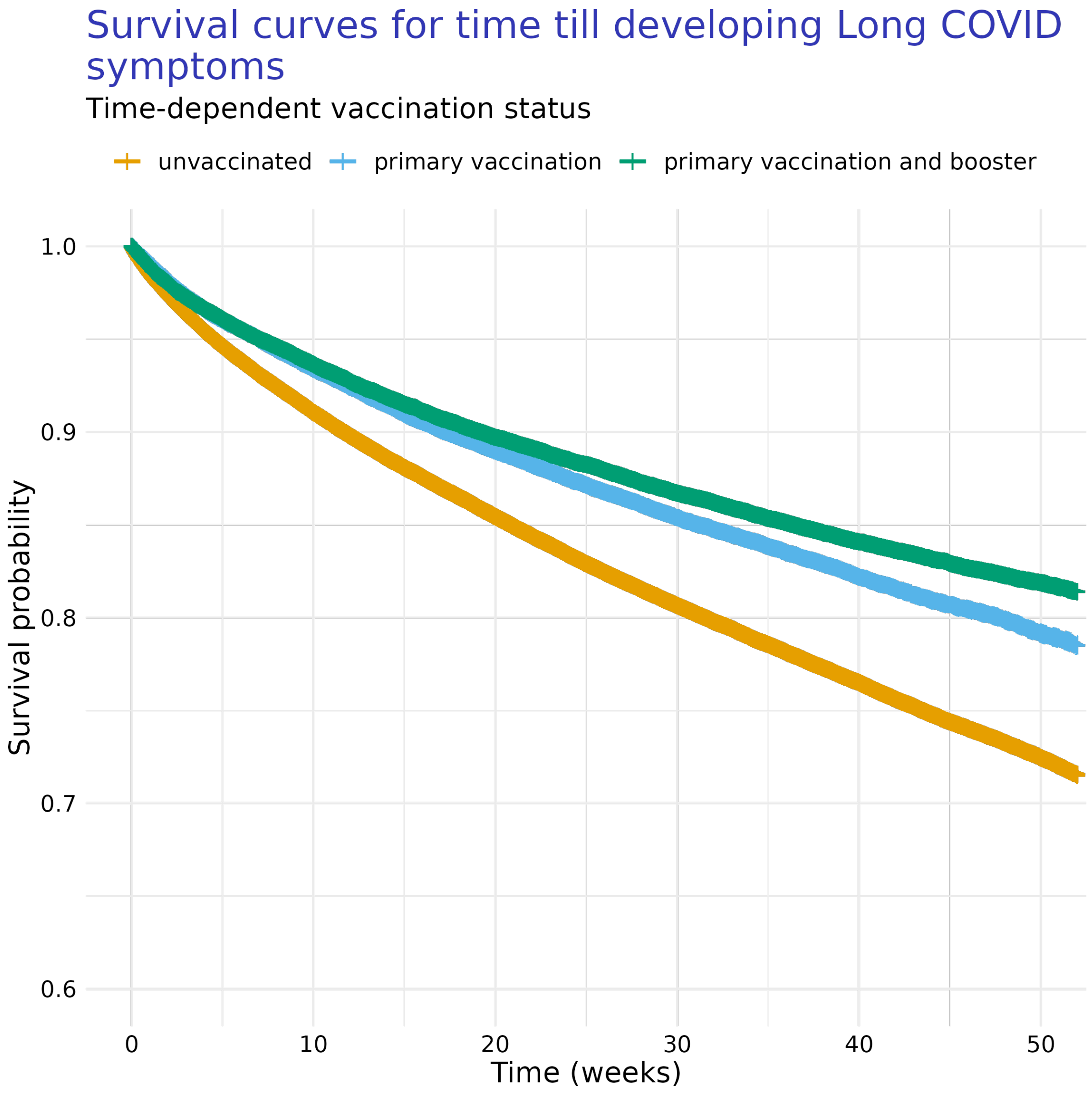

Figure S1: Kaplan-Meier curves of time till development of Long COVID symptoms depending on patient COVID-19 vaccination status (i.e., unvaccinated, completed primary vaccination sequence, and vaccinated plus having received a booster doses). For this analysis, we considered vaccination and booster states as time-dependent covariates, meaning they can happen at any point relative to T0.

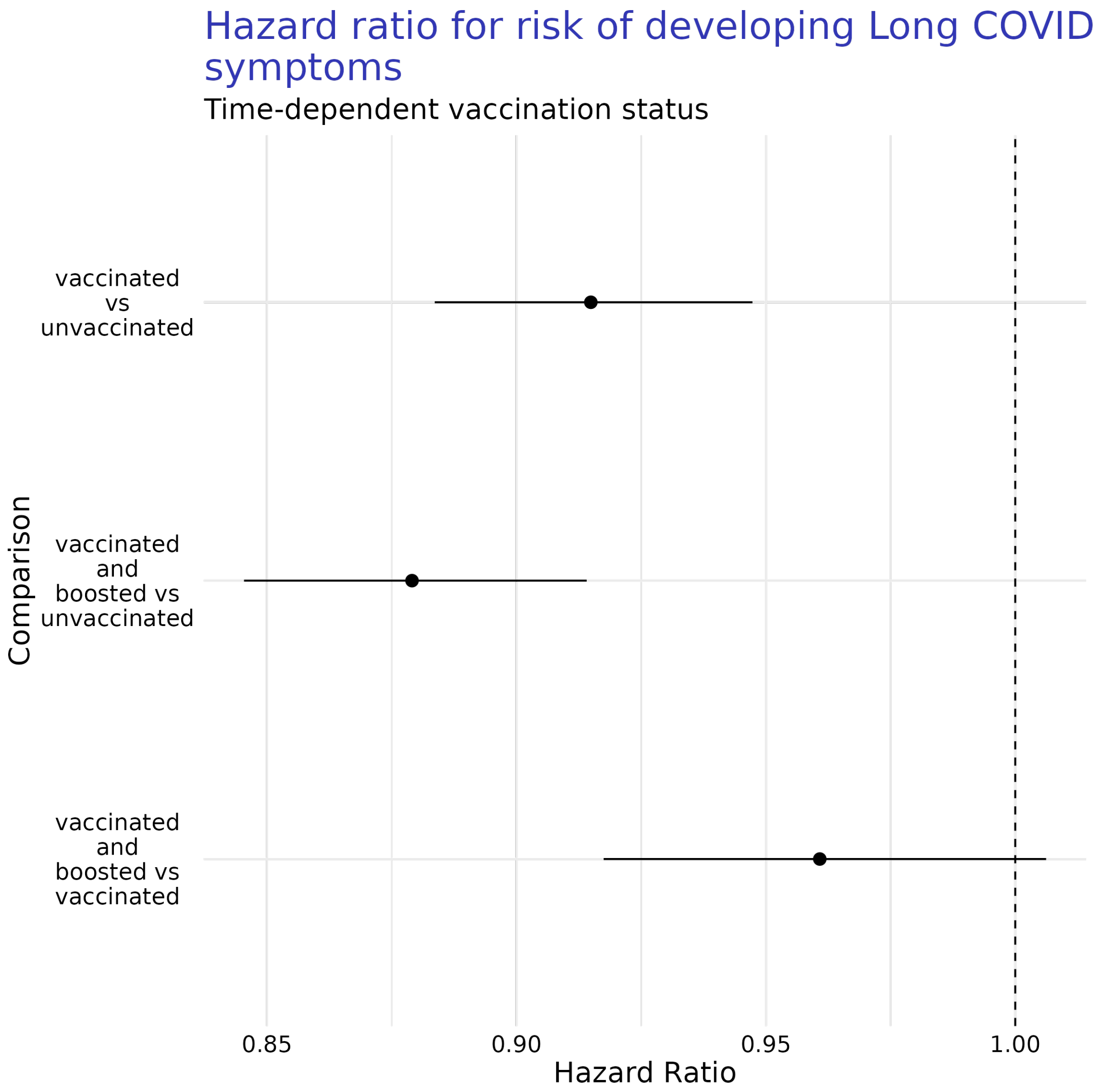

Figure S2: Comparison of estimated hazard ratios for risk of developing Long COVID symptoms after COVID-19 infection based on a person's vaccination state at time of COVID-19 infection (i.e., unvaccinated, completed primary vaccination sequence, and vaccinated plus having received a booster doses). Estimates are presented with 95% confidence intervals. For this analysis, we considered vaccination and booster states as time-dependent covariates, meaning they can happen at any point relative to T0.

The pattern from the Kaplan-Meier curves is effectively retained when time till development of Long COVID symptoms is further conditioned on the comorbidities and patient descriptive features described above, as evidenced by the hazard ratios estimated as part of our Cox regression model (Fig.S4, Table S36). These results are consistent with the comparable analysis presented in main text.

We find that, when vaccination and booster status are modeled as time-dependent covariates, patients who are vaccinated are at a lower risk of being diagnosed with Long COVID than those who are unvaccinated (Fig. S4). Similarly, we find that patients who are vaccinated and boosted have a lower risk of being diagnosed with Long COVID over time than the unvaccinated. Again, this result is consistent with the analysis presented in the main text.

| Comparison | Hazard Ratio [95% CI] |
|---|---|
| vaccinated vs unvaccinated | 0.86 [0.75, 0.99] |
| vaccinated and boosted vs unvaccinated | 0.77 [0.67, 0.9] |
| vaccinated and boosted vs vaccinated | 0.89 [0.76, 1.06] |

Table S36: Estimated hazard ratios for risk of being diagnosed with Long COVID associated with vaccination status where vaccination status is modeled as time-dependent covariates. Hazard ratios are presented with 95% confidence intervals.

#### 7.2 Only vaccination events after COVID-19 infection

We also analyzed the sub-population of patients who were all unvaccinated at time of COVID-19 infection and included any vaccination events which occurred after COVID-19 infection, such as completing a primary COVID-19 vaccination sequence or receiving a booster, as time-dependent covariates.

##### 7.2.1 Long COVID symptoms

There were a total of 357,855 patients included in this analysis. This sample size is smaller than the analyses presented in the main text because here we excluded individuals who completed a primary vaccination sequence prior to their COVID-19 infection were removed, which precludes individuals who received a booster dose prior prior to their COVID-19 infection.

The estimated survival functions for time till development of Long COVID symptoms with vaccination state treated as time-dependent covariates are markedly different from those from previous analyses (Fig. S5). There is little distinguishable difference between the survival curve for unvaccinated patients and those patients who completed a primary vaccination sequence after COVID-19 infection. Similarly, the survival curve for patients who had a primary vaccination and a booster dose exhibits very wide confidence intervals due to a low sample size within that strata.

The uncertainty and lack of differences between the vaccination states exhibited by the Kaplan-Meier curves is retained when time till development of Long COVID symptoms is further conditioned on patient comorbidities and demographic features as evidenced by the hazard ratios estimated as part of our Cox regression model (Fig. S6, Table S37). This analysis yields no evidence that vaccination events after COVID-19 infection have a protective effect against the risk of developing Long COVID symptoms.

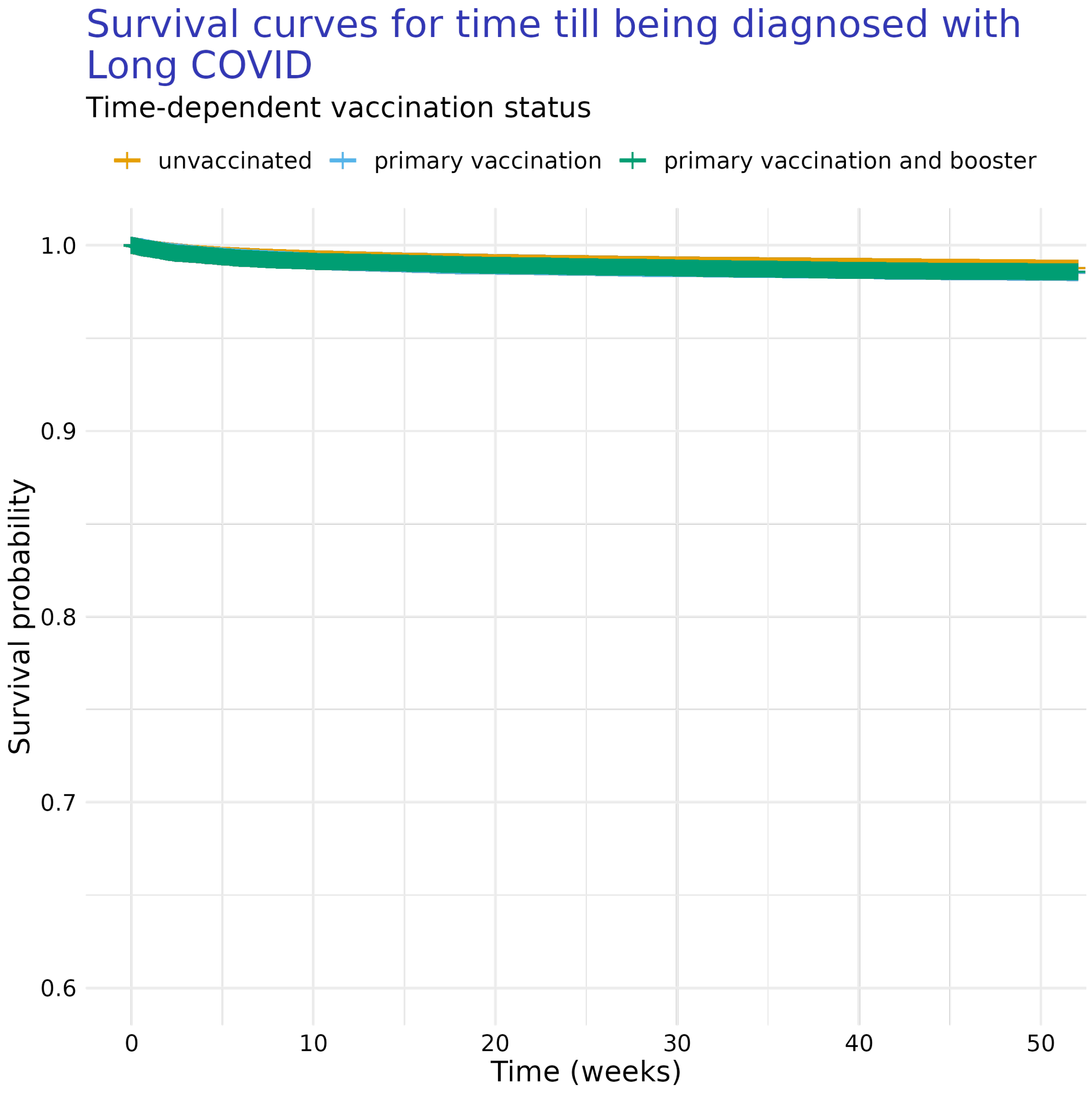

Figure S3: Kaplan-Meier curves for time till diagnosis with Long COVID depending on a patient's COVID-19 vaccination states (i.e., unvaccinated, completed primary vaccination sequence, and vaccinated plus having received a booster doses). For this analysis, we considered vaccination and booster states as time-dependent covariates, meaning they can happen at any point relative to T0.

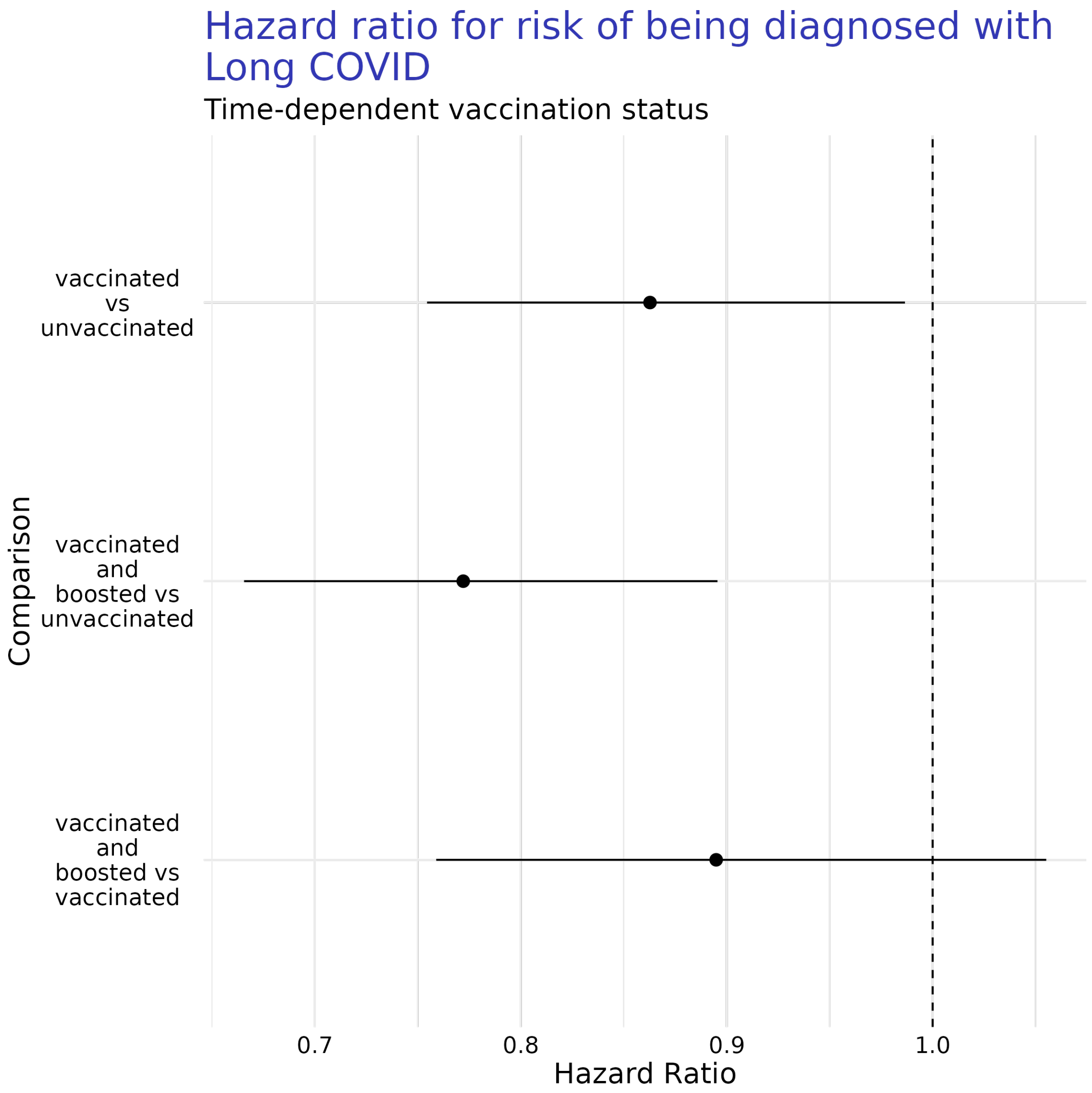

Figure S4: Comparison of estimated hazard ratios for risk of diagnosis with Long COVID symptoms after COVID-19 infection based on a person's vaccination state at time of COVID-19 infection (i.e., unvaccinated, completed primary vaccination sequence, and vaccinated plus having received a booster doses). Estimates are presented with 95% confidence intervals. For this analysis, we considered vaccination and booster states as time-dependent covariates, meaning they can happen at any point relative to T0.

A particularly notable aspect of these results are the large confidence intervals on all estimates involving the effect of booster doses (Fig. S6, Table S37). These confidence intervals are similar to those present in the Kaplan-Meier curves presented earlier (Fig. S5), and are indicative of a very small sample size of individuals who completed both a primary vaccination sequence and received a booster dose within a year after their COVID-19 infection.

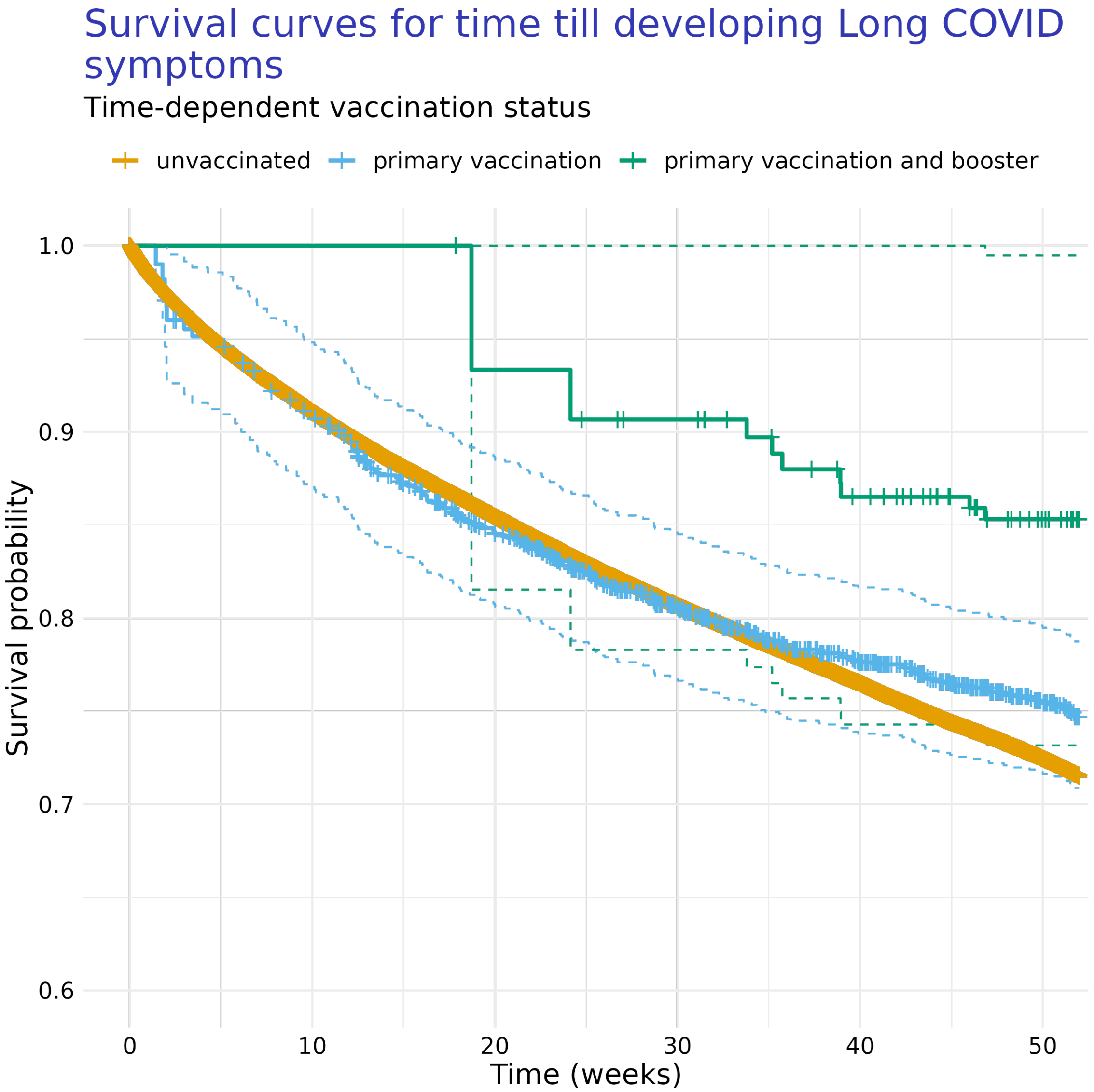

Figure S5: after

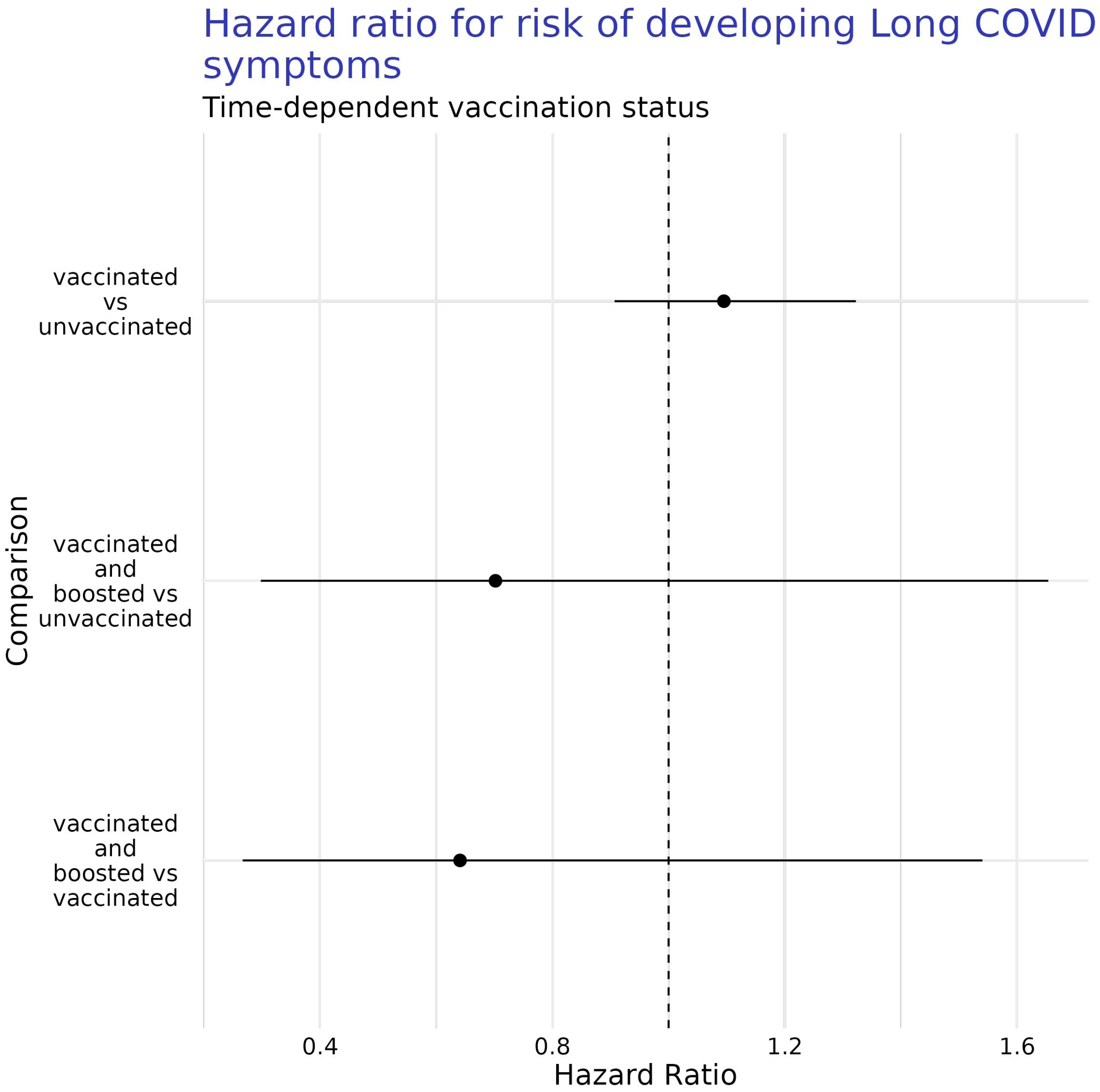

Figure S6: after

| Comparison | Hazard Ratio [95% CI] |
|---|---|
| vaccinated vs unvaccinated | 1.1 [0.91, 1.32] |
| vaccinated and boosted vs unvaccinated | 0.7 [0.3, 1.65] |
| vaccinated and boosted vs vaccinated | 0.64 [0.27, 1.54] |

Table S37: Estimated hazard ratios for risk of developing Long COVID symptoms associated with vaccination status where vaccination status is modeled as time-dependent covariates. Hazard ratios are presented with 95% confidence intervals.

##### 7.2.2 Diagnosis with Long COVID

There were a total of 357,851 patients included in this analysis. This sample size is smaller than the analyses presented in the main text because here we excluded individuals who completed a primary vaccination sequence prior to their COVID-19 infection were removed, which precludes individuals who received a booster dose prior prior to their COVID-19 infection.

The estimated survival functions for time till development of Long COVID symptoms with vaccination state treated as time-dependent covariates are generally consistent with previous analyses of time till diagnosis with Long COVID, as the curves are markedly flat over time (Fig. S5). There is little distinguishable difference between the survival curve for our three patient categories.

The Cox regression model fit to this data failed to converge, as there were too combinations of patients who had received a booster dose and had received a diagnosis with Long COVID. This leads to complete separation of our estimates, an effectively flat likelihood surface, and infinitely wide confidence intervals (Fig. S8, Table S38). Thus, these results mostly indicate this deficiency in our data, and we won't be interpreting them as meaningful. Future analyses may be able to adequately address this scenario as more people receive booster doses.

| Comparison | Hazard Ratio [95% CI] |
|---|---|
| vaccinated vs unvaccinated | 1.54 [0.8, 2.95] |
| vaccinated and boosted vs unvaccinated | 0 [0, Inf] |
| vaccinated and boosted vs vaccinated | 0 [0, Inf] |

Table S38: Estimated hazard ratios for risk of being diagnosed with Long COVID associated with vaccination status where vaccination status is modeled as time-dependent covariates. Hazard ratios are presented with 95% confidence intervals.

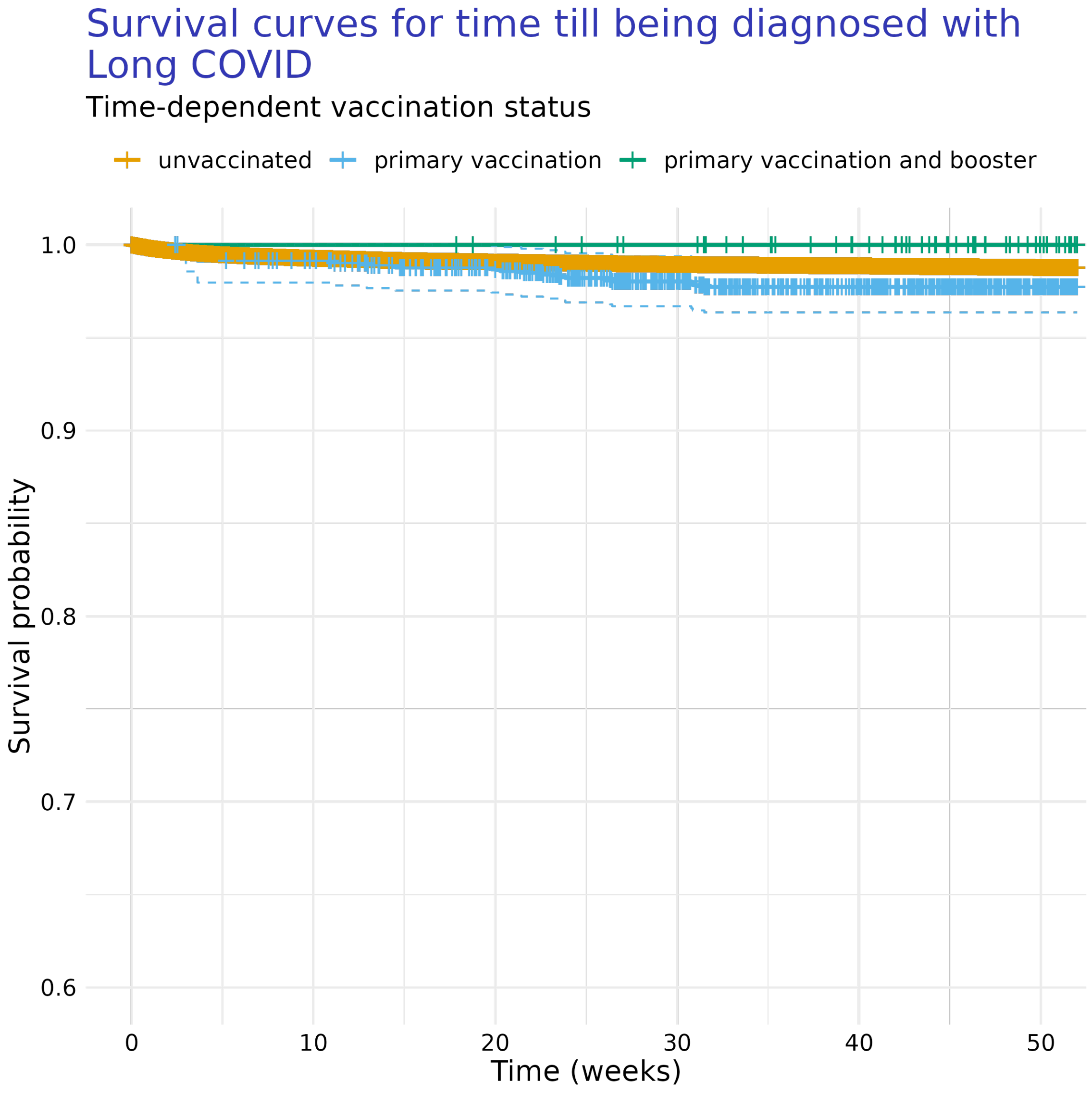

Figure S7: after

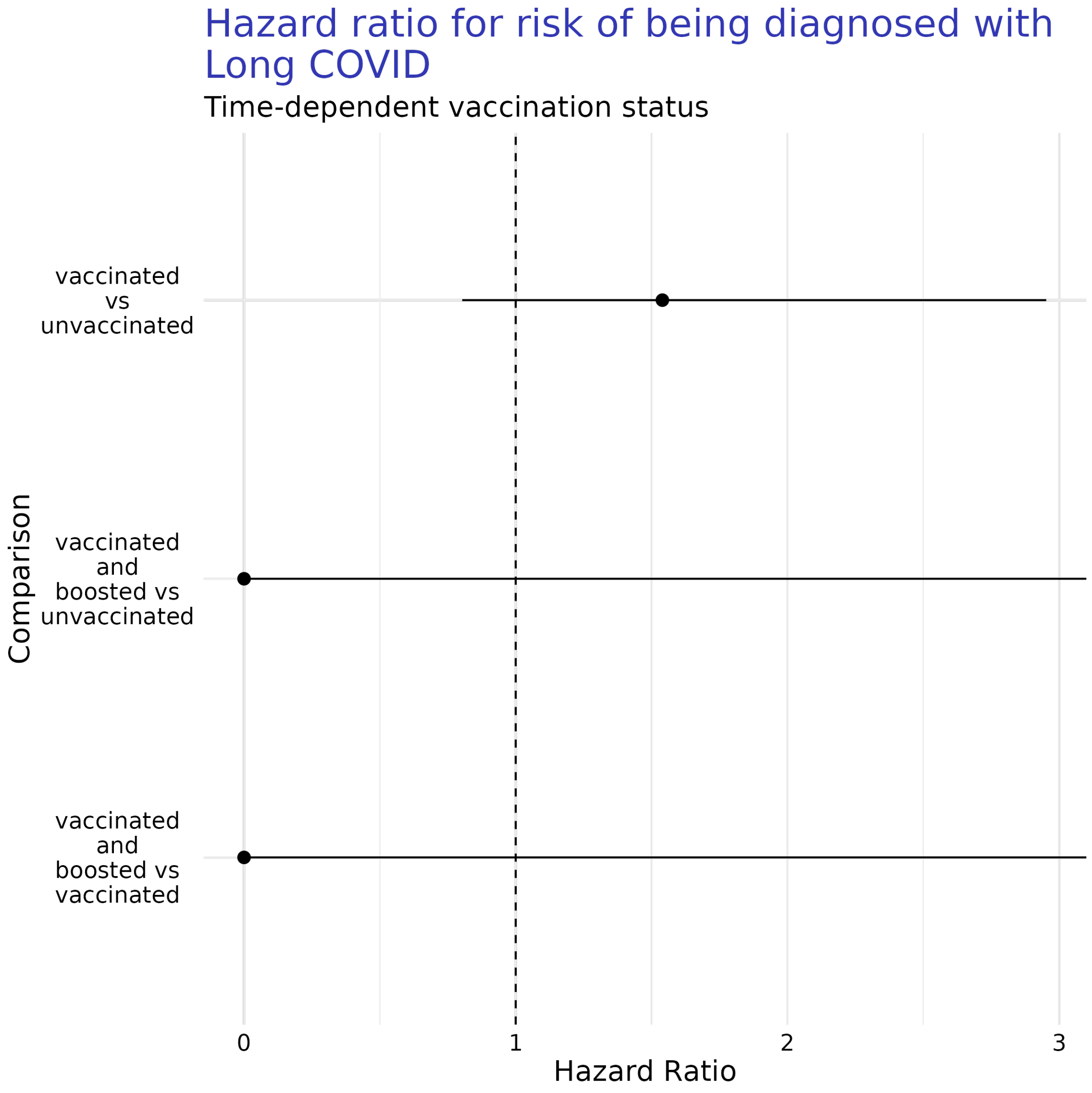

Figure S8: after
